# Aggregating data to accelerate personalized therapy in heart failure (ADAPT-HF)

**DOI:** 10.64898/2026.07.13.26357501

**Authors:** Chris Roeder, Carsten Gorg, Atefeh Talebi, Laura Stevens, Denise Scholtens, Laura Rasmussen-Torvik, Laura M Alagna, Sanjiv Shah, Jennifer Hall, Amar Das, Pardeep Jhund, David Kao

## Abstract

**Background:** Increased public access to data from disparate sources provides opportunities to study and validate predictive and subphenotype models in heterogeneous disease conditions using aggregated individual patient data. Robust, explicit, and transparent harmonization of data elements is critical to ensure interpretability, reproducibility, and generalizability of secondary and retrospective analyses.

**Methods & Results:** We designed and implemented ADAPT (Aggregating Data to Accelerate Personalized Therapy), a scalable framework using multiple software packages (R, SQL, BigQuery) that enables rapid, explicit harmonization of structured data elements from randomized trials and observational studies using a standard spreadsheet interface. User-specified criteria are applied to primary study data to produce harmonized longitudinal datasets comprised of demographics, medical history, quantitative observations, repeated measures, and clinical outcomes. We demonstrate this functionality using 26 clinical studies found in the National Heart, Lung, and Blood Institute’s BioLINCC resource. We illustrate the scalability of ADAPT to the order of billions of datapoints using administrative clinical data in a cloud-computing platform. We also present examples of collaborators using ADAPT for independent harmonization tasks for secondary analyses and democratization of publicly available data.

**Conclusion:** ADAPT is a disease-agnostic, extensible, and scalable platform to support robust, transparent harmonization of structured research data using interfaces accessible to a variety of researchers regardless of programming ability. It extends FAIR principles beyond research data to also represent harmonization analyses by improving <u>F</u>indability of harmonization decisions, <u>A</u>ccessibility of methods to other stakeholders, <u>I</u>nteroperability with independent analyses and datasets, and <u>R</u>eusability through efficient implementation in a variety of analysis environments.

## BACKGROUND

Biomedical research has been revolutionized by the development of high-throughput molecular experimental methods and robust techniques available for analyzing large, complex translational datasets. Analogously, large amounts of data are generated during clinical research and routine care, and there is growing interest in establishing frameworks for harmonizing multiple clinical datasets.^1–6^ Despite increasingly robust conceptual frameworks, practical curation and computational methods for leveraging secondary data analysis in clinical research remain comparatively underdeveloped. Aggregation of structured study data has been limited, in part due to uncertainty regarding comparability of data between sources, lack of consensus regarding clinical definitions, and inter-study differences in harmonization decisions. For example, if one study defines a chronic disease like heart failure (HF) based on self-reported history, another uses a set of clinical criteria such as those from the Framingham Heart Study and yet another uses administrative codes, patients identified as having HF in each study may not be truly comparable (**Figure 1**). Recommendations and consensus statements that articulate frameworks for harmonizing disparate data describe the key components in the process of codifying relationships between potentially similar data fields, but there remains no widely used platform for completing these tasks in a discoverable and reusable way. Uncertainty and difficulty in scaling clinical data harmonization have a number of consequences including lost opportunity for discovery, unnecessary effort and resource utilization through redundant harmonization of datasets for different projects, lack of transparency regarding reusability and comparability of different harmonization projects, and uncertainty regarding generalizability of any research findings to specific populations.

**Figure 1.**
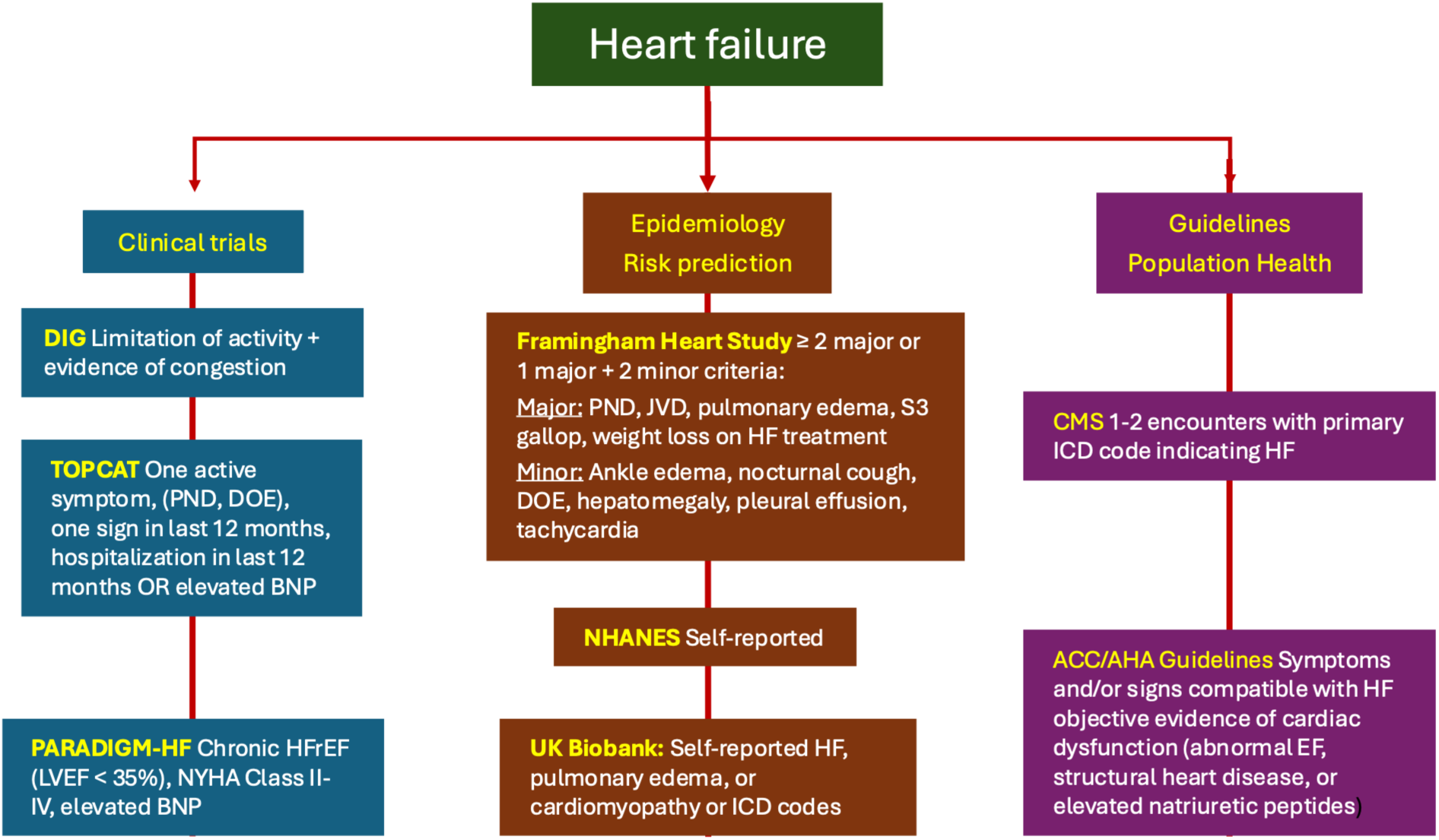
Divergent, context diagnostic criteria for heart failure.

One commonly cited motivation for analysis of harmonized data from multiple studies, also called individual patient data meta-analysis, is the ability to study small but important patient populations. Robust analysis of these subgroups can be difficult due to challenges in explicit, portable classification criteria and small sample sizes, particularly in clinical trials. Evidence from clinical trials has been treated as equally applicable to all patients meeting a trial’s inclusion criteria. In reality, phenotypes are often heterogeneous among patients with chronic conditions, which may result in variable prognoses and treatment responses.^7^ For example, guideline-directed medical therapy for HF is recommended based on a single measurement of heart function, the left ventricular ejection fraction (LVEF). However, there are dozens of known HF etiologies that could affect long-term prognosis and treatment response, such as coronary artery disease, toxic exposures, and genetic abnormalities affecting muscle contraction.^8^ This diversity of phenotypes means that even evidence-based interventions may have beneficial, neutral, or harmful effects in any given subpopulation.

To overcome limited sample sizes, characterization of disease sub-populations can be accomplished through individual patient-level data harmonization and analysis,^5,9^ wherein patients with similar subphenotypes can be aggregated from multiple studies. Utility, credibility, and generalizability of secondary analysis findings from aggregated individual patient data analyses from multiple studies, however, depend on the accuracy and consistency of data harmonization between relevant studies as well as the ability for other investigators and clinicians to examine the harmonization mappings. Historically, reconciling these differences has been accomplished by examining different statistical scripts for each unique analysis. For example, the fields ‘sex’ = “Male” in Study A, ‘gender’ = “1” in Study B, and ‘male’ = “Yes” in Study C could all denote male subjects, but the mapping definitions can only be discerned by reading the primary code (e.g. R, SAS, Stata, Python). Due to the complexity of clinical data, harmonizing the wide range of elements needed for aggregate analysis is often labor-intensive and difficult to replicate. Systematic uncertainty is introduced by the limited ability to identify specific harmonization decisions within programming code and to apply those definitions to an independent dataset or clinical implementation system. This uncertainty compounds the impacts on generalizability that arise from aggregate analyses from multiple clinical studies based on study enrollment criteria. Despite the complexity of aggregating these data and potential impacts on the utility of resulting findings, there is little, if any, documentation of harmonization or variable mapping between studies used for aggregate analysis.

The principles of <u>F</u>indability, <u>A</u>ccessibility, Interoperability, and <u>R</u>eusability (FAIR) are frequently used to guide collection, annotation, and storage of biomedical data.^10^ We propose that FAIR principles are also applicable to data harmonization mappings and pipelines. We further propose that the construction of standardized frameworks to codify and implement harmonization specifications will facilitate explicit, reusable, and generalizable aggregation of clinical data.

### Objective

Develop and demonstrate a scalable, reusable framework that enables rapid, flexible, and transparent harmonization of clinical data to produce aggregate harmonized datasets from disparate sources.

## METHODS

### Requirements

Our framework addresses the following properties of analyses using harmonized data:

i. Clinical data may originate from different sources in different formats.
ii. Acquisition of new clinical data or discovery of new knowledge may necessitate revision of concept mapping or phenotype definitions.
iii. A clinical concept may have different definitions in different contexts (e.g. DM)
iv. Analyses may require an arbitrarily large number of harmonized data elements (e. g. artificial intelligence/machine learning).
v. Analyses and software may require data outputs in a variety of formats.

The ADAPT (Aggregating Data to Accelerate Personalized Therapy) framework applies user-specified harmonization criteria to patient-level data from disparate sources to create a single harmonized dataset. Inputs are raw clinical data from the primary study, such as medical history, medications, laboratory values, and imaging findings; domain knowledge from the researcher, such as disease definitions, pathophysiology, study design, and time intervals; and data format requirements for the desired analysis such as normalized, entity-attribute-value (EAV) and repeated-measures tables. Unique harmonization criteria can be created by a researcher for a given project, or definitions from previous analyses can be reused from prior analyses. Criteria may be based on formal disease definitions (e.g. HF with reduced ejection fraction = clinical HF with LVEF < 40%), widely accepted clinical knowledge (e.g. chronic kidney disease stage 3a = estimated glomerular filtration rate = 15-29 ml/min), or they can be customized for specific needs (e.g. baseline B-type natriuretic peptide level > median of all subjects enrolled in study A). Outputs of ADAPT are harmonization specifications in tabular format and aggregated patient-centered datasets that can be formatted for many purposes such as disease-association, decision support, or quality assurance analysis. Herein we describe our framework and apply it to datasets and concepts related to HF as our initial use case.

### Data sources

We obtained publicly available, deidentified data from two primary sources to develop and test the scalability of ADAPT. Use of these data is therefore not considered human subjects research, but we did conduct this work with review and approval of the Colorado Multi-Institutional Research Board as required by agencies providing the data. A full list of data sets, total subjects in each, and the source data agency are found in the **Supplement**.

#### BioLINCC

Data for 20 clinical HF trials (n=32,442 subjects) and 5 observational cohorts (n=54,085) were obtained from the National Heart, Lung, and Blood Institute’s Biological Specimen and Data Repository Information Coordinating Center (BioLINCC, https://biolincc.nhlbi.nih.gov/home/).^1111^Primary data were received as *.csv (comma-separated values), *.sas7bdat (SAS) or *.dta (Stata) files. Data dictionaries were obtained from BioLINCC and study sites (e.g. ARIC: https://aric.cscc.unc.edu/) in PDF, word-processing document, and spreadsheet formats.

#### Administrative hospital discharge data

Deidentified records for 20 states between 1994 and 2023 totaling 288,341,132 hospital discharges were obtained from individual state public health agencies as available and/or the AHRQ Healthcare Utilization Project Statewide Inpatient Databases.^14^ Diagnoses, external injuries, and procedures performed were reported using both International Classification of Disease (ICD) version 9 (up to 2015 quarter 3) and version 10 (2015 quarter 4 and after).

#### Data management software

Data were aggregated, harmonized and analyzed in multiple platforms including Protégé-OWL 3.3.1 (Stanford University, Palo Alto, CA, https://protege.stanford.edu/),^12,13^ RStudio (version 2025.09.1, Posit Software, Boston, MA, https://posit.co/products/open-source/rstudio), the R statistical package (version 4.5.0, R Foundation for Statistical Computing, Vienna, Austria, https://www.r-project.org/), Stata Statistical Software (Release 18, StataCorp LLC), MySQL (version 8.0.27, Oracle Corporation. https://www.mysql.com/), PostgreSQL (version 9.6, PostgreSQL Global Development Group, https://www.postgresql.org), and Google BigQuery (Google LLC, https://cloud.google.com/bigquery).

### ADAPT Framework

To harmonize data, domain-specific knowledge asserting functional equivalency between data elements must first be encoded by the user. Four primary types of information are required: 1) primary clinical data such as tests, measurements, physical examination or patient self-reported history; 2) harmonization criteria (e.g., a patient has anemia if their hemoglobin level is < 13 g/dL for men and < 12 g/dL for women); 3) domain knowledge regarding concepts, phenotypes, or disease processes relevant to the current project (e.g. studying the impact of anemia in HF); and 4) the analysis being performed and data format needed to execute that analysis (e.g. survival analysis vs. random forest prediction vs. growth curve analysis).

**Figure 1** shows a schematic of relationships between subjects, data, and harmonized phenotype concepts as linked by a set of simple relationships. The key differentiator of ADAPT is the prioritization of rapid, transparent and scalable harmonization. Although not exhaustive, we have found a wide variety of analyses can be accomplished using a combination of the following archetypal methods: 1) “identity” or “exact value” (harmonized value = original value), 2) “exact match” (harmonized value = x if original value = y), 3) “ordinal” (harmonized value = x if any of a set of specified values are present), and 4) “scalar” (harmonized = x if original value is “greater than,” “less than,” or “between” user-specified values). Ordinal variables may be handled either using exact string matching or using a scalar comparison (e.g. ‘> 12^th^ grade education’), so we elected not to create a distinct method for ordinal variables. Specific harmonization criteria may represent accepted clinical definitions (e.g., patients with a body mass index > 30 kg/m2, or a waist circumference ≥ 94 cm in men or ≥ 80 cm in women indicates obesity), or they may be analysis- or dataset-specific (e.g., lab value greater or less than the median of a specified dataset). Specification of harmonization criteria or rules is accomplished in a standard spreadsheet format, eliminating the requirement for programming knowledge and allowing use of convenience tools like copy/paste. Importantly, these assertions can be reused between multiple datasets, making subsequent harmonization tasks more efficient and, where desired, standardized.

The process of transforming raw data into many harmonized values can be broken down into the three primary steps: 1) collation of raw data from disparate sources into a single table of primary or raw data points with contextual annotation, such as the study, source table, table, field, and units for each datum; 2) application of scalar comparisons, string-matching, or assertions of equivalence (i.e. identity) comparing individual patient data and user-defined criteria (e.g., obesity = body mass index > 30 kg/m^2^) to coalesce source data into common clinical concepts and values, and 3) formatting and export of harmonized results for analysis (e.g. descriptive statistics, hypothesis testing, machine learning in R, Stata, or SAS). Using a denormalized data structure, transparent harmonization specifications, and a modular pipeline, ADAPT enables more rapid ingestion of new datasets than standard methods as well as simplified reuse of harmonization definitions and analytic methods from prior projects on data acquired from disparate sources.

**Figure 2.**
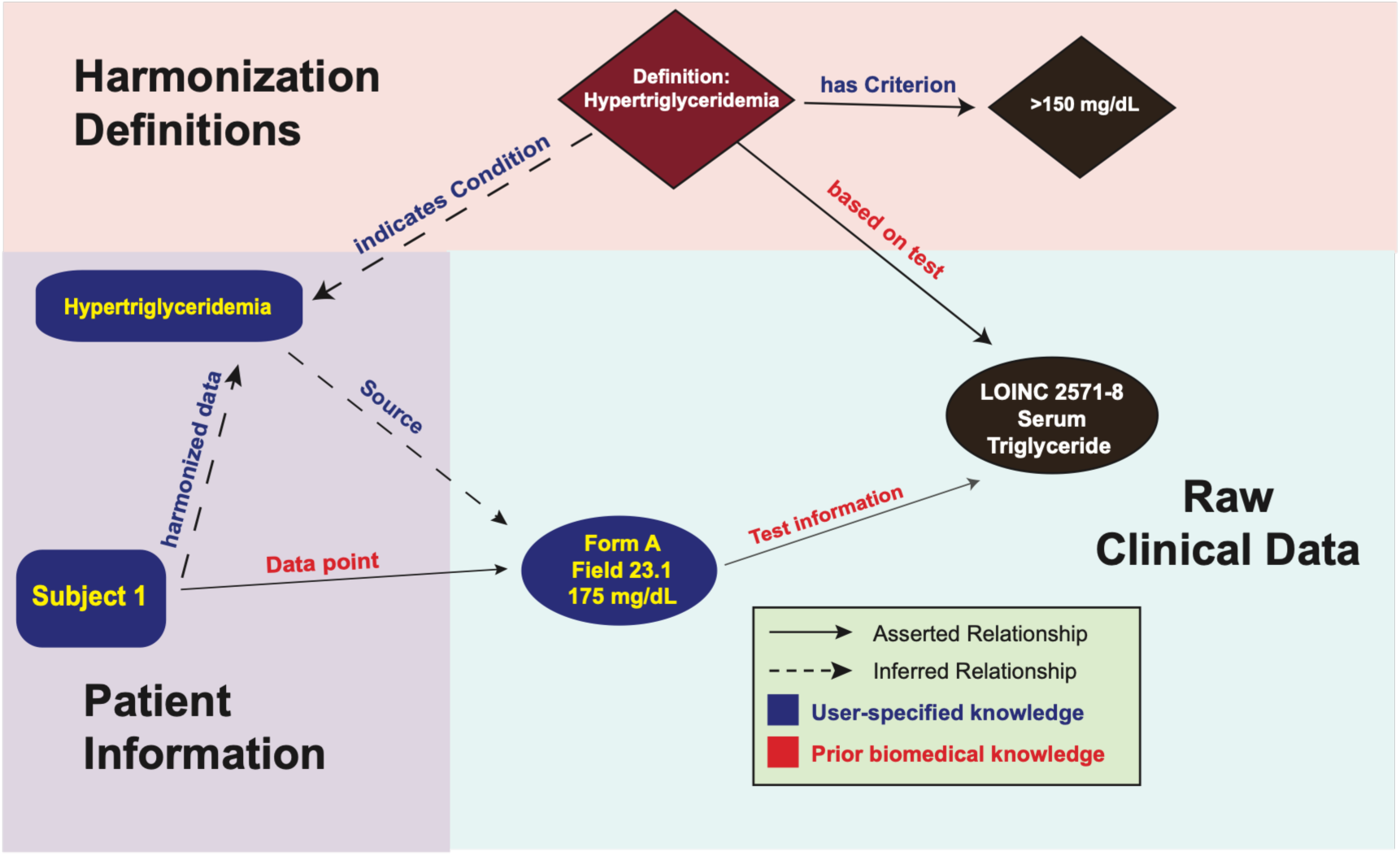
Conceptual ADAPT harmonization framework.

### Implementation

The flow of data through ADAPT is shown in **Figure 3**. Primary data from sources of interest are then transformed and assembled into a single denormalized data repository using either a performance-optimized custom schema or an established Common Data Model such as that of the Observational Medical Outcomes Partnership. In the resulting repository, each line represents a single raw observation about one patient as recorded in the primary data source, e.g. Framingham subject 2345 (patient) -> Field *AB12* (sex) -> 1 (male). A unique index is assigned to each primary datapoint in order to maintain the provenance of harmonized values. Harmonized datapoints are produced by applying community-defined or user-specified criteria to the denormalized primary data using code that applies one of the harmonization method archetypes described above: *exact matching* (e.g. study field demo.gender = ‘M’ -> concept:Sex = value:Male), *scalar comparisons* (study field demo.age < 18 -> concept:Adult = ‘Yes’), or *identity* (e.g. study field demo.agey = 18 -> concept:age_yrs = 18). We implemented the ability to use phenotype-contingent criteria (e.g. race- and sex-specific ranges) if desired.

**Figure 3.**
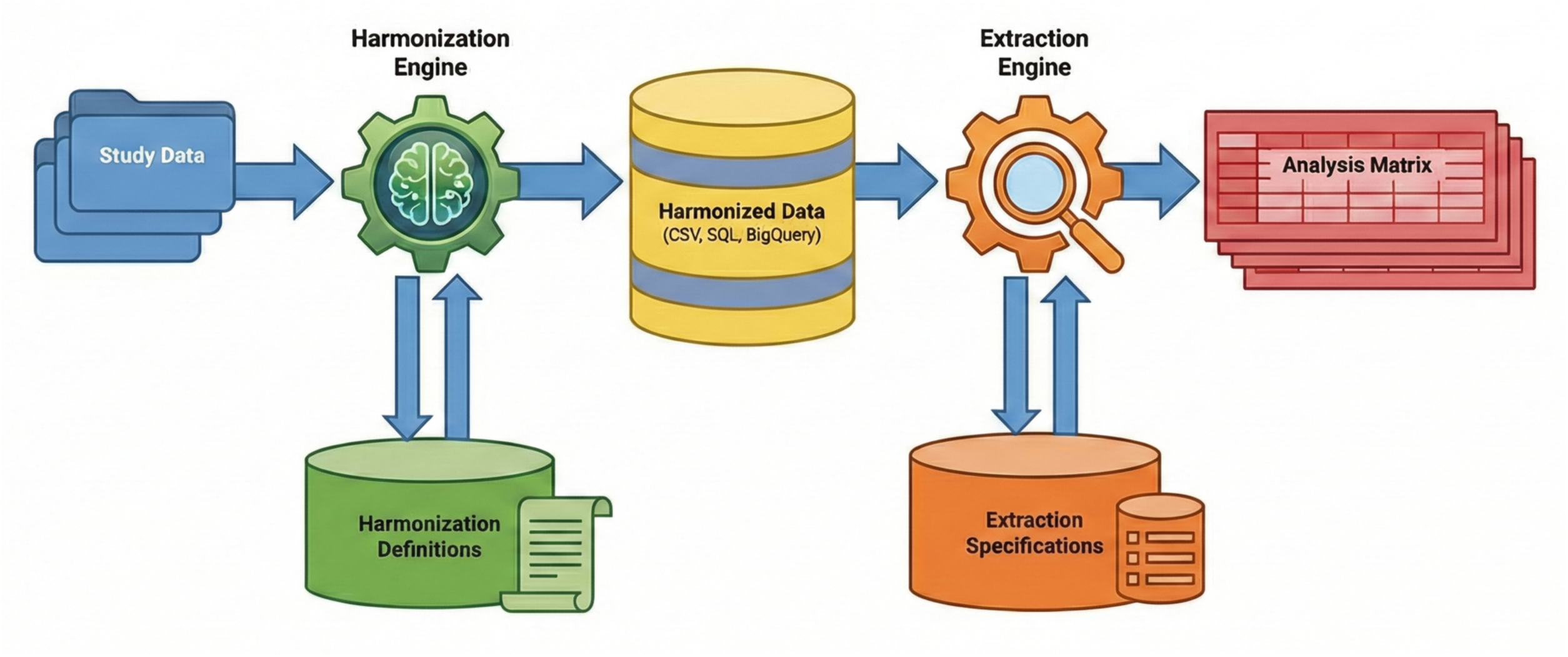
ADAPT harmonization pipeline components.

Application of harmonization specifications is performed using conditional joins or table merges based on user specifications. Example code blocks using R, SQL, and BigQuery are provided in **Table 1**. All versions of the ADAPT pipeline utilize the same simple tabular format to specify harmonization criteria through standard spreadsheet applications and as a harmonization criteria repository that may be readily searched and explored for existing harmonization rules (**Table 2**). This table can be manipulated easily by domain experts even if they lack coding expertise. The resulting file is used directly to execute the harmonization code, eliminating the possibility of discordance between the harmonized data produced and the repository of definitions. The product is a second denormalized EAV repository where each line represents one harmonized observation about one subject: e.g., Framingham subject FHS2345 (patient) -> common concept = “Sex” -> harmonized value = “Male” using datapoint F87654. Comprehensive BioLINCC harmonization mappings for BioLINCC data as well as harmonization code for the R statistical package and Google BigQuery are available at https://github.com/dkao42/ADAPT-HF.

**Table 1.** Example code for exact string match harmonization.

| R | BigQuery | MySQL (variant legacy SQL) |
| --- | --- | --- |
| `harmonized_data <- unique( merge( alldata[, c("patientid", "study", "visit_yr", "form", "field", "value", "datapoint")], criteria_match[, c("study", "form", "field", "phenotype", "match_string", "harmonized_val_numeric", "harmonized_val_string", "rule_number")], by.x=c("study", "form", "field", "value"), by.y=c("study", "form", "field", "match_string"))]` | `create or replace table harmonized_data as ( select distinct study, patientid, visit_yr, phenotype, harmonized_val_numeric harmonized_val_string, rule_num, datapoint, from adapt.alldata as d join adapt.criteria_match as c on (d.study = c.study, d.form = c.form, d.field = c.field, d.value = c.match_string)` | `insert into harmonized_data select distinct d.study, d.patientid, d.visit_yr, c.phenotype, c.harmonized_val_numeric c.harmonized_val_string, c.rule_num, d.datapoint, from criteria_match as c, join alldata as d using (field) where and c.study = d.study and c.form = d.form and c.field = d.field and c.match_string = d.value` |

**Table 2.** Example harmonization criteria table.

| Study | Form | Field | Concept | Lower bound | Upper bound | Match string | Sex-specific? | Harmonized value | Harmonized string | Rule type | Criteria source |
| --- | --- | --- | --- | --- | --- | --- | --- | --- | --- | --- | --- |
| DOSE | a_visitsumm | LL_HMG | Anemia | 13 |  |  | Male | 1 | No | range | WHO Iron Deficiency Anemia Programme |
| DOSE | a_visitsumm | LL_HMG | Anemia | 12 |  |  | Female | 1 | No | range | WHO Iron Deficiency Anemia Programme |
| DOSE | a_visitsumm | LL_HMG | Anemia |  | 13 |  | Male | 2 | Yes | range | WHO Iron Deficiency Anemia Programme |
| DOSE | a_visitsumm | LL_HMG | Anemia |  | 12 |  | Female | 2 | Yes | range | WHO Iron Deficiency Anemia Programme |
| ESCAPE | lab | 3 | Anemia | 41 |  |  | Male | 1 | No | range | WHO Iron Deficiency Anemia Programme |
| ESCAPE | lab | 3 | Anemia | 36 |  |  | Female | 1 | No | range | WHO Iron Deficiency Anemia Programme |
| ESCAPE | lab | 3 | Anemia |  | 41 |  | Male | 2 | Yes | range | WHO Iron Deficiency Anemia Programme |
| ESCAPE | lab | 3 | Anemia |  | 36 |  | Female | 2 | Yes | range | WHO Iron Deficiency Anemia Programme |
| SCD-HeFT | baseline_new | gender | Sex |  |  | 1 |  | 1 | Male | match |  |
| SCD-HeFT | baseline_new | gender | Sex |  |  | 2 |  | 2 | Female | match |  |
| SOLVD | SEF | SEF9 | Sex |  |  | M |  | 1 | Male | match |  |
| SOLVD | SEF | SEF9 | Sex |  |  | F |  | 2 | Female | match |  |

When a harmonized value is created, it is annotated with the index of the primary datum used to create it (e.g., Subject FHS2345 from Clinical Trial A’s concept:Sex has the harmonized value “Male” based on datapoint F87654 (Clinical Trial A, Subject 2345, form *a_demo*, field gender = 1). This index allows datum metadata such as the source table, field, or timepoint, to be retrieved. An example of the harmonization criteria table and application to compiled raw data is provided in **Figure 4**. Using this approach, researchers can choose to treat measurements using different methods as equivalent (e.g. left ventricular ejection fraction via echocardiography can be compared to same measurement via magnetic resonance imaging) and represent this equivalence explicitly in harmonization criteria while retaining the ability to audit the source data in the event of unexpected findings.

**Figure 4:**
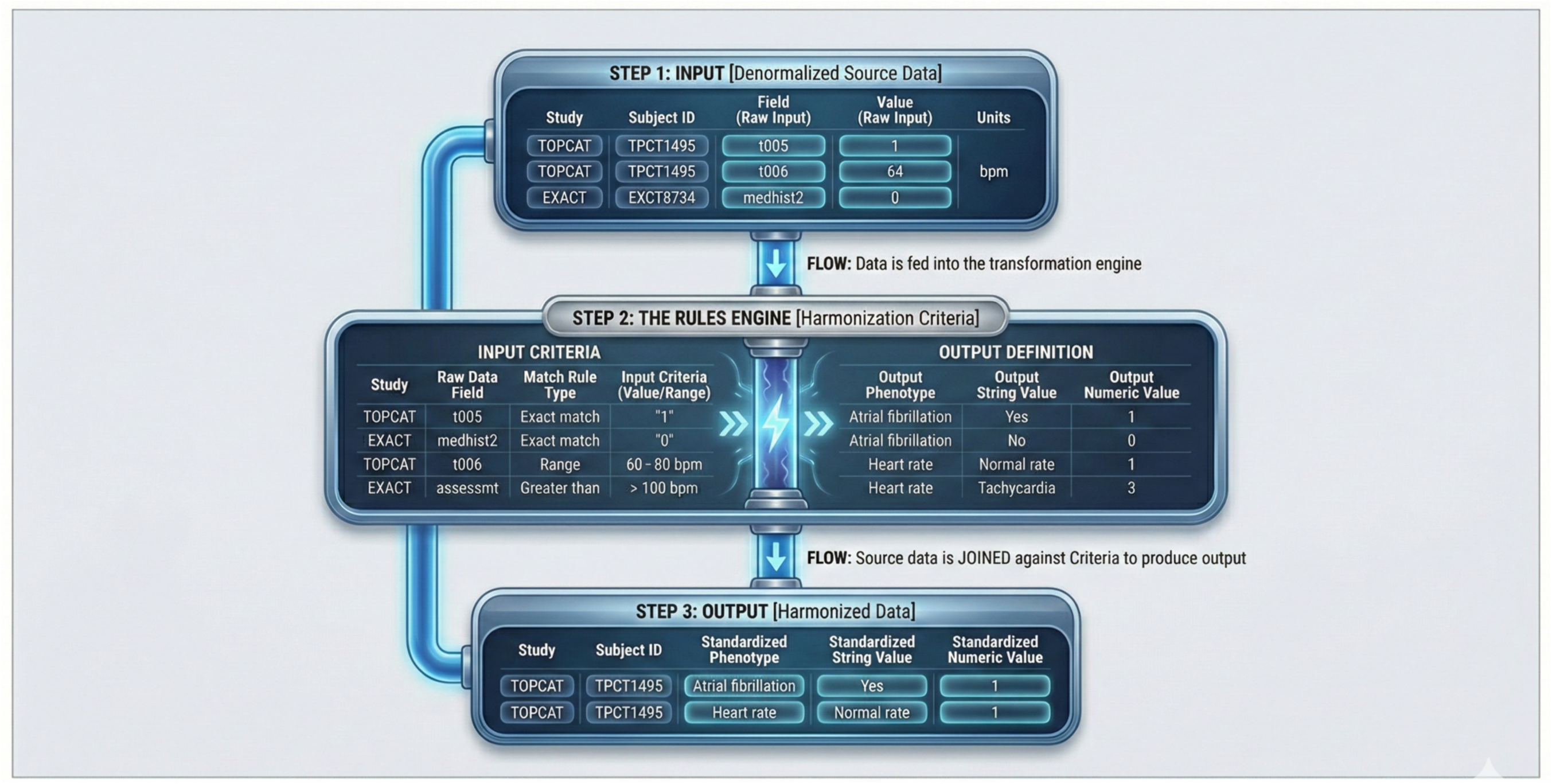
Application of ADAPT harmonization criteria.

The final result is a patient-centered graph-like structure for each subject that is a greatly expanded version of **Figure 1**. These graphs contain all available primary data relating to an individual, harmonized observations created from primary clinical data according to the user-specified phenotype definitions, and explicit representation of the data point(s) used to generate each harmonized value. Queries from any standard statistical package can be used to produce specific output formats for specific data analysis packages and algorithms. We have used this approach to conduct analyses ranging from simple logistic regression to unsupervised machine learning.

### Example 1: Secondary analysis of clinical HF studies

We have used ADAPT to produce harmonized datasets comprised of all clinical randomized HF trials in BioLINCC as well as the ARIC, CHS, Framingham (Original, Offspring, Offspring Spouse, Generation III, Omni 1-2), JHS, MESA observational cohorts (**Table 3**). Taken together, we harmonized > 7519 study variables to 692 harmonized concepts from both study groups (>85,000 subjects, > 140 million raw data points, > 30 million harmonized data points using ∼17,800 individual harmonization statements. This comprised a wide range of variables including demographics, comorbidities, medications, physical findings, test results, survey instruments, and clinical outcomes. Harmonized baseline characteristics are provided in **Table S1**. We implemented and tested ADAPT in multiple software environments to produce different degrees of computational performance, user accessibility, expressivity, and scalability. Listed roughly in order of maximal expressivity to maximal scalability, we used the R statistical package, local and cloud-based Structured Query Language (SQL) relational databases (MySQL, PostgreSQL), and the cloud-based Google BigQuery. Code for implementation of ADAPT in these systems is available from the authors on request. We have demonstrated the ability to implement ADAPT in web-based workbook environments including the American Heart Association’s Precision Medicine Platform and the University of Colorado Health Data Compass Eureka environment. The latter enables access to deidentified EHR data for our health system using the Observational Medical Outcomes Partnership (OMOP) Common Data Model.

**Table 3.** Summary of data from BioLINCC HF studies harmonized using ADAPT.

| Trial | Subjects | Study fields | Harmonized concepts | Time points | Raw data processed | Harmonized observations |
| --- | --- | --- | --- | --- | --- | --- |
| ATHENA | 360 | 134 | 102 | 7 | 114,969 | 69,181 |
| BEST | 2708 | 88 | 107 | 10 | 1,606,613 | 693,385 |
| CARRESS | 188 | 113 | 129 | 10 | 60,897 | 59,416 |
| DIG | 7788 | 48 | 66 | 4 | 533,021 | 379,327 |
| DOSE-AHF | 308 | 110 | 124 | 9 | 88,770 | 90,963 |
| ESCAPE | 433 | 146 | 175 | 13 | 476,688 | 203,892 |
| EXACT-HF | 253 | 160 | 24 | 4 | 137,445 | 83,050 |
| FIGHT | 300 | 171 | 188 | 4 | 220,931 | 99,188 |
| GUIDE-IT | 894 | 107 | 182 | 11 | 796,552 | 352,177 |
| HF-ACTION | 2130 | 123 | 122 | 8 | 1,451,701 | 487,769 |
| INDIE | 105 | 131 | 144 | 5 | 55,935 | 27,552 |
| IRONOUT | 225 | 116 | 146 | 4 | 94,449 | 50,703 |
| LIFE | 335 | 107 | 113 | 10 | 313,067 | 135,275 |
| MOCHA | 345 | 66 | 112 | 7 | 224,271 | 94,059 |
| NEAT-HFpEF | 110 | 153 | 171 | 3 | 60,677 | 25,639 |
| RELAX | 216 | 156 | 185 | 3 | 103,487 | 50,724 |
| ROSE-AHF | 360 | 108 | 116 | 5 | 134,248 | 93,325 |
| SCD-HEFT | 2521 | 123 | 96 | 23 | 4,822,498 | 902,582 |
| SOLVD | 6797 | 86 | 79 | 18 | 5,885,741 | 2,122,563 |
| STICH | 2621 | 143 | 175 | 22 | 2,361,186 | 834,017 |
| TOPCAT | 3445 | 128 | 167 | 16 | 3,086,502 | 1,660,995 |
| Total | 32442 | 2517 | 3252 |  | 22,629,648 | 8,515,782 |
| ARIC | 15004 | 477 | 299 | 28 | 4,424,380 | 7,149,645 |
| CHS | 5888 | 1841 | 249 | 23 | 30,613,027 | 5,040,353 |
| FHS | 15126 | 1423 | 288 | 36 | 50,334,253 | 5,191,332 |
| JHS | 3883 | 281 | 216 | 18 | 6,670,066 | 1,164,141 |
| MESA | 6814 | 515 | 207 | 5 | 25,416,682 | 3,293,729 |
| SOLVD Registry | 6270 | 74 | 104 | 1 | 771,044 | 392,335 |
| Total | 52985 | 4537 | 524 | 48 | 118,229,452 | 22,231,535 |
| Combined | 85427 | 7519 | 692 |  | 140,859,100 | 30,747,317 |

### Example 2: Administrative hospital discharge data

After establishing key workflow components and testing harmonization pipeline integrity on BioLINCC data, we migrated the same data schema, workflow, and code to Google BigQuery to assess scalability. We used ADAPT to harmonize and analyze statewide administrative hospital discharge from data (**Table 4**, **Supplement Table S2**) from 20 states. Slight modifications were incorporated to accommodate unique features of administrative hospital discharge records (e.g. diagnosis present on admission, designation of primary diagnosis or procedure). Harmonization specifications were derived using a combination of state-specific documentation, standardized case report forms, and International Classification of Disease (ICD) version 9 and 10 codes.^15,16^ To illustrate adaptability to independently derived harmonization schema, we also harmonized diagnosis data to phecodes, ^17,18^ which are curated, standardized sets of ICD codes used in phenome-wide association analysis (PheWAS). There were total of ∼9.0 billion demographic, administrative, and outcomes-oriented primary data points ∼2.1 billion diagnosis ICD codes, ∼461.7 million procedure ICD codes, and ∼45 million external injury codes in the primary raw dataset. We then used > 15,800 demographic/administrative/outcomes criteria, > 7,900 diagnosis codes (ICD-9 and -10), and > 84,500 phecode specifications to produce a harmonized dataset comprising 8.1 billion administrative data points, 1.1 billion diagnoses, 109 million procedures, 45.1 million external injuries, and 1.2 billion phecodes (total ∼10.5 billion datapoints).

**Table 4.**
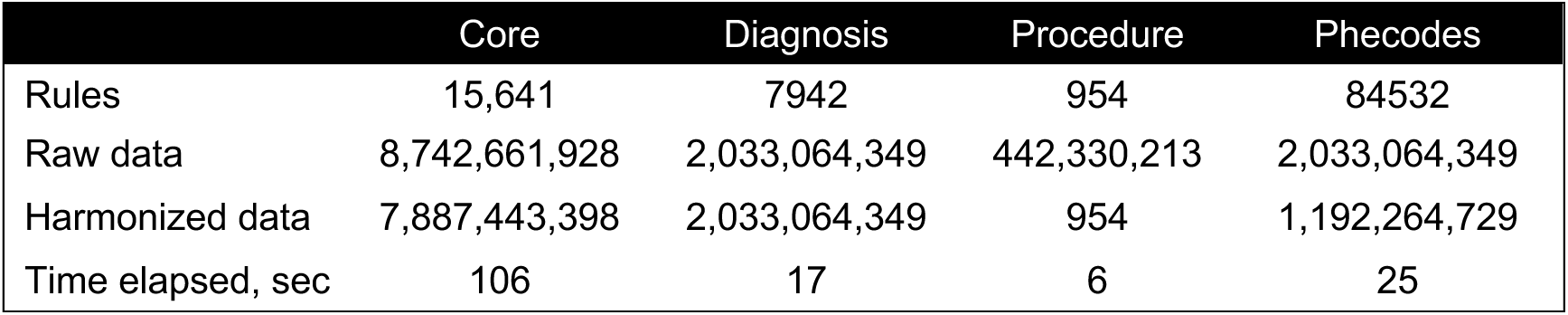
Administrative hospital discharge data.

|  | Core | Diagnosis | Procedure | Phecodes |
| --- | --- | --- | --- | --- |
| Rules | 15,641 | 7942 | 954 | 84532 |
| Raw data | 8,742,661,928 | 2,033,064,349 | 442,330,213 | 2,033,064,349 |
| Harmonized data | 7,887,443,398 | 2,033,064,349 | 954 | 1,192,264,729 |
| Time elapsed, sec | 106 | 17 | 6 | 25 |

### Clinical research productivity (**Supplemental Table S3a-b**)

Using these data, we have published 16 peer-reviewed manuscripts (46 distinct coauthors and presented 49 abstracts at national and international conferences (55 distinct coauthors). We have found these data to be particularly helpful for trainees. Among 27 young investigators using these data, 23 presented abstracts at national or international conferences and 13 produced at least 1 peer-reviewed publication. Topics have ranged from association between coffee intake and heart failure risk to impact of climate and air quality on heart failure, and in-hospital outcomes of necrotizing fasciitis. We have also used ADAPT to create interactive R Shiny-based worksheets for self-directed exploration and cluster analysis of HF clinical trial data, a project which was chosen as a Selected Solution for the NHLBI Big Data Analysis Heart Failure Challenge. ^19^

### Portability

Two groups of collaborators have successfully used ADAPT to harmonize clinical trial data independently from the development team. First, HF researchers at the University of Glasgow have harmonized data from industry-sponsored clinical trials, which we have merged with BioLINCC data. Second, we collaborated with the AMP HF/HeartShare network to create^20^ a large harmonized dataset for use by independent researchers comprised of all publicly available HF trials from BioLINCC HF trials and several observational cohorts. This helped achieve an important part of the network’s mission to democratize publicly available clinical trial data. Harmonized data are now published on BioData Catalyst (https://www.nhlbi.nih.gov/science/biodata-catalyst).

## DISCUSSION

Using a variety of widely-used free and low-cost software platforms, we created a flexible, scalable, and transparent system for encoding, applying, and retaining harmonization criteria for complex integration of clinical research data. In order to make the platform useful to a wide range of investigators and analytic needs, we chose to support multiple harmonization definitions for any given concept rather than limiting mappings to singular definitions of complex traits such as HF. We focused instead on efficient capture and representation of harmonization criteria based on the categories of harmonization (e.g. original value, exact matching, scalar comparison). ADAPT can support consensus disease definitions as easily as project-specific criteria, even within a single project. The explicit transparency achieved by using a familiar spreadsheet format enables investigators to easily find and compare harmonization criteria between projects and to identify easily the raw datum used for each harmonized datapoint. Maintenance of the harmonization repository is simplified by using the same physical files for encoding, querying, execution, and sharing. ADAPT can be deployed locally or in the cloud, and we have found this architecture to be scalable up to tens of billions of datapoints and thousands of harmonization rules by using high performance cloud-based data stores. We have found ADAPT to be a broadly effective tool for our work in a wide range of research applications across a several types of datasets, and it has been used by other research groups for similar tasks. We believe that systems such as ADAPT will be pivotal to realizing the research potential of the ever-increasing number of clinical and translational datasets.

Rapid accumulation of clinical and research datasets with a focus on data sharing democratization has prompted increasing discussion of strategies for harmonizing structured data.^1,2,4–6,21^ Recommendations often include standardized data collection templates and terminologies, common data elements and data models, and validation of harmonized data. While these recommendations are central to improving data interoperability, comparability, and efficiency of data sharing, practical guidance and infrastructure regarding implementation of these principles are lacking. ADAPT represents a concrete step towards implementing those community recommendations, in part by focusing on the technical workflows to integrate data from disparate sources rather than curation of specific biomedical concepts.

An important decision in design of a practical harmonization framework is whether to use a reductionist philosophy, wherein the goal is to establish a singular definition for each concept to be used by all investigators. Such an approach provides high confidence in comparability of data from multiple sources. However, it can be difficult to establish and confidently apply formal, specific definitions to a large number of datapoints in diverse contexts from different sources, restricting the amount of data that may be aggregated and the purposes for which they may be used. Alternatively, the harmonization framework may focus on maximizing expressivity in defining harmonization terms without requiring a ground-truth definition for each concept. In practice, this approach is used frequently in both research and clinical care, albeit often in an ad hoc manner. For example, the left ventricular ejection fraction (LVEF) is a critical measurement in HF research and can be measured by echocardiogram, computed tomography, or magnetic resonance imaging, each with different strengths and weaknesses. Precision and accuracy vary between modalities, and the reported LVEF is usually not numerically identical between modalities. However, classification by LVEF ranges of < 40% (HF with reduced ejection fraction or HFrEF), 40-49% (HF with mildly reduced ejection fraction or HFmrEF) and ≥ 50% (HF with preserved ejection fraction or HFpEF) are often sufficient for an analysis. In such cases, the advantages of comparing less precise LVEFs over a larger population may be larger than the impact of inter-modality variation. However, other scenarios which focus on quantitative correlation between LVEF and, for example, exposure, may require higher precision. These sorts of decisions are quite common in clinical medicine, but are often implicit in statistical analysis. With ADAPT, it is easy to implement explicitly the decision to use imaging modalities interchangeably if appropriate for a given analysis or to be more restrictive for others. Furthermore, it is easy for a subsequent user (e.g. researcher or clinician) to ascertain and evaluate this choice for their needs.

We elected to develop a flexible framework because a) it can still be used to implement fundamental or consensus concept definitions if desired and b) it can be adapted for different communities and requirements to harmonize their data. Importantly, we believe that this strategy is only possible with a very high degree of transparency into harmonization decisions, a quality that is often lost with manual or project-specific harmonization, for example in data collection or statistical code. Full transparency allows investigators to a) evaluate the credibility of prior decisions, b) assess comparability of harmonized datasets and generalizability of related findings, and c) determine the relevance of existing libraries of criteria to new projects and whether the prior content may be reused. Flexibility in harmonization parameters is important to maximize utility for research, but we believe a robust harmonization framework must also support integration with community-oriented resources such as standardized definitions and controlled terminologies. After using our framework in a number of settings, we have concluded that the FAIR principles can be applied to harmonization processes themselves in addition to primary data and their metadata. Using FAIR principles in harmonization projects would facilitate collaboration, support robust validation, and increase the reusability of any given set of harmonization specifications. We propose that FAIR principles may be applied to data harmonization as follows using ADAPT as an example:

### Findability

Users may wish to identify harmonization criteria by the clinical concept used (e.g. LVEF), the primary data element (e.g. lvef_baseline), the study (e.g. Framingham Heart Study Original Cohort) or the harmonized value (e.g. “< 40%”). ADAPT captures user-designated harmonization criteria in a generic tabular format (CSV), which can be explored using any spreadsheet application and all statistical packages (**Figure 3**). Because the statistical code contains no criteria directly but rather extracts and applies the criteria from the primary harmonization dictionary, reviewing primary statistical code to recreate harmonization decisions is rendered unnecessary. Use of a familiar user interface enables non-coding subject matter experts to participate directly in the harmonization process and makes available features such as copying/pasting, searching, sorting and filtering to manipulate the research criteria. Differences between concept definitions (e.g. WHO vs. ADA definition of diabetes vs. self-reported history) can be compared given that the harmonization criteria are clearly and explicitly stated. Additionally, use of the same file for both creating criteria, accessing them subsequently AND applying criteria to the data of interest reduces the need for maintenance and dramatically reduces the possibility of unexpected discordant harmonization criteria.

### Accessibility

The highly readable and portable harmonization dictionary format used by ADAPT improves researchers’ ability to identify and reuse harmonization schemata when trying to validate findings or reuse definitions in independent datasets. Large data aggregation projects have historically required considerable technical proficiency in statistical programming, which frequently limits the participation of clinical or translational subject matter experts who lack those skills. ADAPT allows clear and unambiguous examination of these harmonization decisions. Consequently, investigators with the most domain knowledge often have the least access to creating and reviewing mapping criteria. Because it is unnecessary to review the statistical code the simplicity and familiarity of the spreadsheet interface can enable investigators and subject matter experts to select appropriate harmonization criteria from prior projects or if none exist, encode new criteria without statistical programming ability. Criteria can also be revised and directly applied to data by the same investigator iteratively (e.g. quantile sizes). These advantages therefore enable investigators who could not previously directly interact with the harmonization process to evaluate existing rules or specify new ones fully if they wish.

### Interoperability

ADAPT is built upon common functionalities present in a variety of analytic and programming environments. The harmonization criteria components are stored in comma separated value text format, making the core content usable in any computing environment. It is operating-system agnostic and was developed entirely using free software, although it can also be implemented in commercial environments (e.g. Stata, BigQuery). It can be run on local hardware, on-premises servers, or in cloud-based resources. This provides access to ADAPT in most settings upon translation of the core algorithmic routines which we already provide for R, Stata, SQL, and BigQuery. This enables robust reuse and reapplication to independent datasets as well as the ability to critically evaluate published research and the definitions utilized in any given analysis that uses ADAPT. Finally, the ability to collate data sources without limit within a given harmonization specification allows easy comparison of fields from the data source, and transformation across many studies. The ability to understand the details and specificity of definitions can greatly improve (or reduce) confidence in veracity and generalizability of findings. Furthermore, the ability to implement ADAPT quickly in a new environment simplifies harmonization and sharing where raw data is subject to access restrictions.

### Reusability

<u>R</u>eusability is facilitated by the streamlined, clinical domain-agnostic structure which can be applied to new electronic clinical data requiring minimal programming background for new harmonization projects. Reliance on a simple tab-delimited format to transfer harmonization specification reduces the time required to ingest new datasets for a collaborative project. Additionally, the explicit knowledge of the harmonization decisions allows statistical analysis code to be run on many datasets with little customization. The ability to rapidly inspect and select harmonization rules from prior work promotes consistency through time-savings for subsequent investigators. In other words, it is often easier to use an existing representation of a condition than to create a new one. As a library of harmonization rules grows, the time needed to incorporate new datasets will decrease.

### Limitations

A key limitation of ADAPT presently is the inability to express complex mathematical transformations. This is often a core requirement of harmonizing certain datasets, but implementing an integrated mathematical equation platform was beyond the scope of this effort. Currently, complex formulas can be applied prior to harmonization or can be executed using harmonized data, which still simplifies the analysis process. However, a more expressive computation would be an important incremental feature. Although we have made the harmonization activity quite streamlined by reducing code requirements, the process does require manual review of data dictionaries to determine whether variables of interest exist. Automated abstraction of data dictionaries and coding manuals could help make that review more efficient, and ADAPT would allow investigators to express rules of interest. Although we have created thousands of rules for hundreds of clinical concepts, the focus of these rules is still our use case domain of HF. Several of the advantages of ADAPT such as reusability and interoperability would grow rapidly with implementation of common clinical concepts that could then be reused in other fields of study. Ideally, harmonization libraries will be created by subject matter experts in different clinical domains to optimize quality and utility, but it could take some time to achieve widespread use through reusability.

### Future directions

The use cases described here harmonized structured data from clinical studies or systematic administrative data. One of the largest barriers to model testing and implementation in clinical care is the difference in quality and veracity between research or administrative data and a primary electronic health record. We believe ADAPT has potential utility in translating certain computational tools developed in research data to active clinical data. We planned for this by establishing the ability to use ADAPT on data stored in the OMOP common data model, which is a widely used clinical data warehouse framework. Although not real-time, OMOP-formatted data nevertheless demonstrates many of the weaknesses of real-world clinical data. We are currently exploring use of ADAPT in real-world data and to determine if and how it could be promoted to a real-time framework.

We believe that a growing user community may ultimately improve standardization, scope, and quality of harmonization libraries. There are no specific constraints on harmonization rules currently, and any user can implement any rule they require. However, it will likely prove easier for researchers to leverage definitions for common features that have already been encoded. Since the foundation of those definitions is easy to interrogate, we believe that commonly used concepts will ultimately attain relatively stable definitions based on the viewpoints of the larger community. Widespread reuse of specific rules across many users would effectively result in standardization within harmonization projects, and ultimately a formal strategy for designating standardization could prove useful.

## CONCLUSION

Validation, sharing, and scaling applications using data from disparate sources requires confidence in comparability of data between sources. The almost limitless variety of uses for these data means that singular definitions for any one concept are unlikely to meet all users’ needs. We have created and demonstrated application of a streamlined, platform-agnostic pipeline that enables rapid, explicit, and transparent definition of harmonization decisions that, using cloud computing platforms, can be scaled at minimum to the order of billions of data points. Using tools like ADAPT, we believe that applying the concepts of Findability, Accessibility, Interoperability, and Reusability to harmonization operations in addition to the data sources will ultimately improve confidence in and performance of algorithms based on aggregated and independent clinical data.

## Supporting information

Dataset references

Harmonization scripts

Harmonization definitions

## Data Availability

All data used are available at the sites listed above and in the supplemental material. Here are the links again:
NHLBI's BioLINCC: (biolincc.nhlbi.nih.gov)
Agency for Healthcare Research and Quality Healthcare Utilization Project: (https://hcup-us.ahrq.gov/)
California (https://hcai.ca.gov/data/request-data/research-data-request-information/)
Colorado (https://cha.com/center-for-health-information-and-data-analytics/)
Delaware (https://dhss.delaware.gov/dph/hp/hosp_dis_data/)
Louisiana (https://ldh.la.gov/bureau-of-health-informatics/lahidd),
New Hampshire (https://www.dhhs.nh.gov/programs-services/population-health/health-statistics-informatics/hospital-discharge-data)
New Jersey (https://www.nj.gov/health/healthcarequality/health-care-professionals/njddcs/)
Nevada (https://www.dhs.nv.gov/Programs/dhhs-office-of-analytics/),
New York (http://www.health.ny.gov/statistics/sparcs/)
Oklahoma (https://oklahoma.gov/health/health-education/data-and-statistics/center-for-health-statistics/health-care-information/hospital-discharge-and-outpatient-asc-surgery-data.html)
Texas (https://www.dshs.texas.gov/center-health-statistics/texas-health-care-information-collection/download-and-purchase-data/texas-inpatient-public-use-data-file-pudf)
Vermont (https://gmcboard.vermont.gov/data-release-0/vermont-uniform-hospital-discharge-data-system-vuhdds)
West Virginia: (https://hca.wv.gov/fdhome/HospInpatientData/Pages/default.aspx)

https://biolincc.nhlbi.nih.gov

https://hcup-us.ahrq.gov/

https://hcai.ca.gov/data/request-data/research-data-request-information/

https://cha.com/center-for-health-information-and-data-analytics/

https://dhss.delaware.gov/dph/hp/hosp_dis_data/

https://ldh.la.gov/bureau-of-health-informatics/lahidd

https://www.dhhs.nh.gov/programs-services/population-health/health-statistics-informatics/hospital-discharge-data

https://www.nj.gov/health/healthcarequality/health-care-professionals/njddcs/

https://www.dhs.nv.gov/Programs/dhhs-office-of-analytics/

http://www.health.ny.gov/statistics/sparcs/

## ACKNOWLEDGEMENTS

The authors wish to thank Martin O’Connor, Mark Musen, John Graybeal, Mike Bristow, John McMurray, James Lewsey, and JoAnn Lindenfeld for their advice and support.

---

**Table S1:**
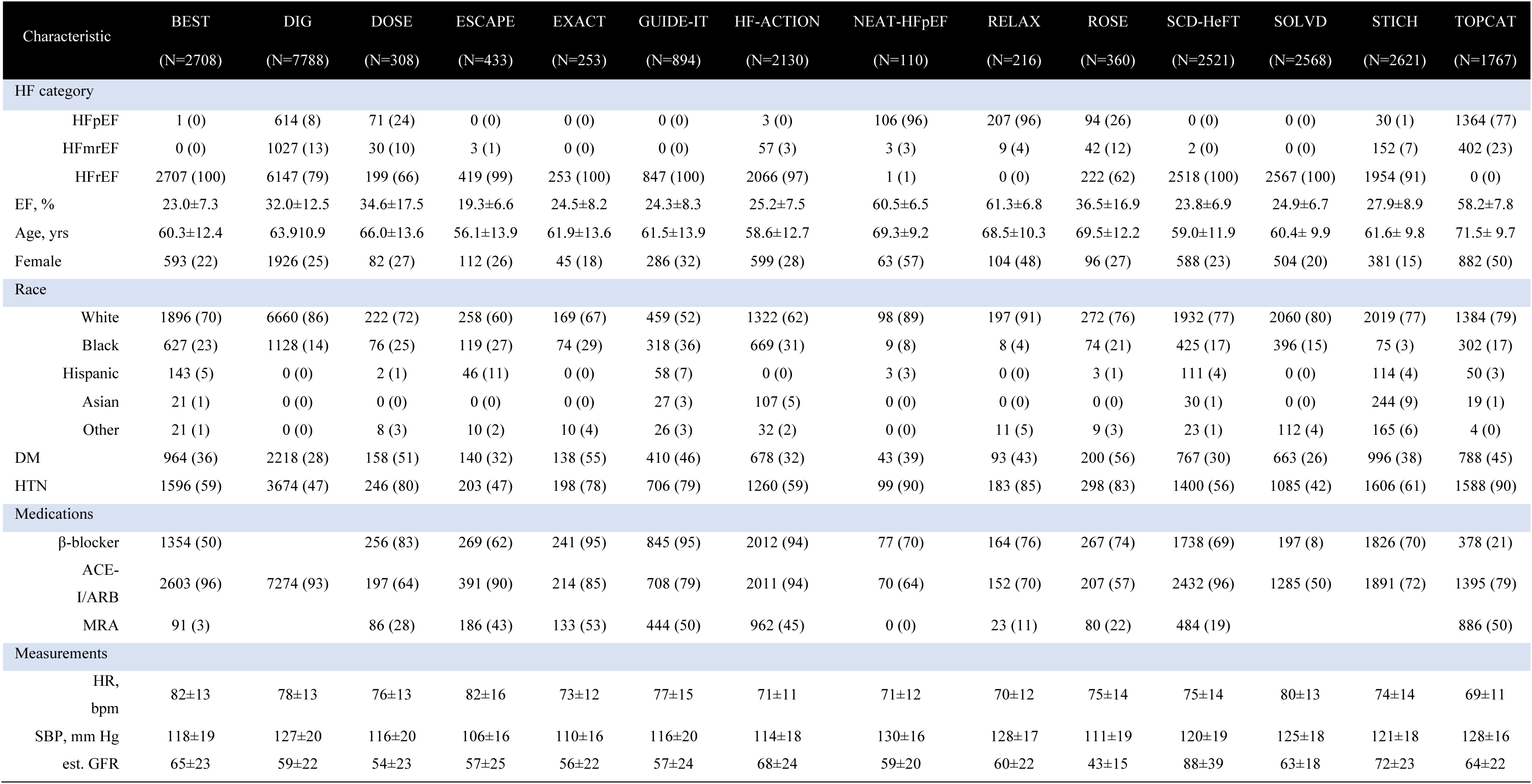
Baseline characteristics of BioLINCC heart failure trials using harmonized dataset.

**Table S2:**
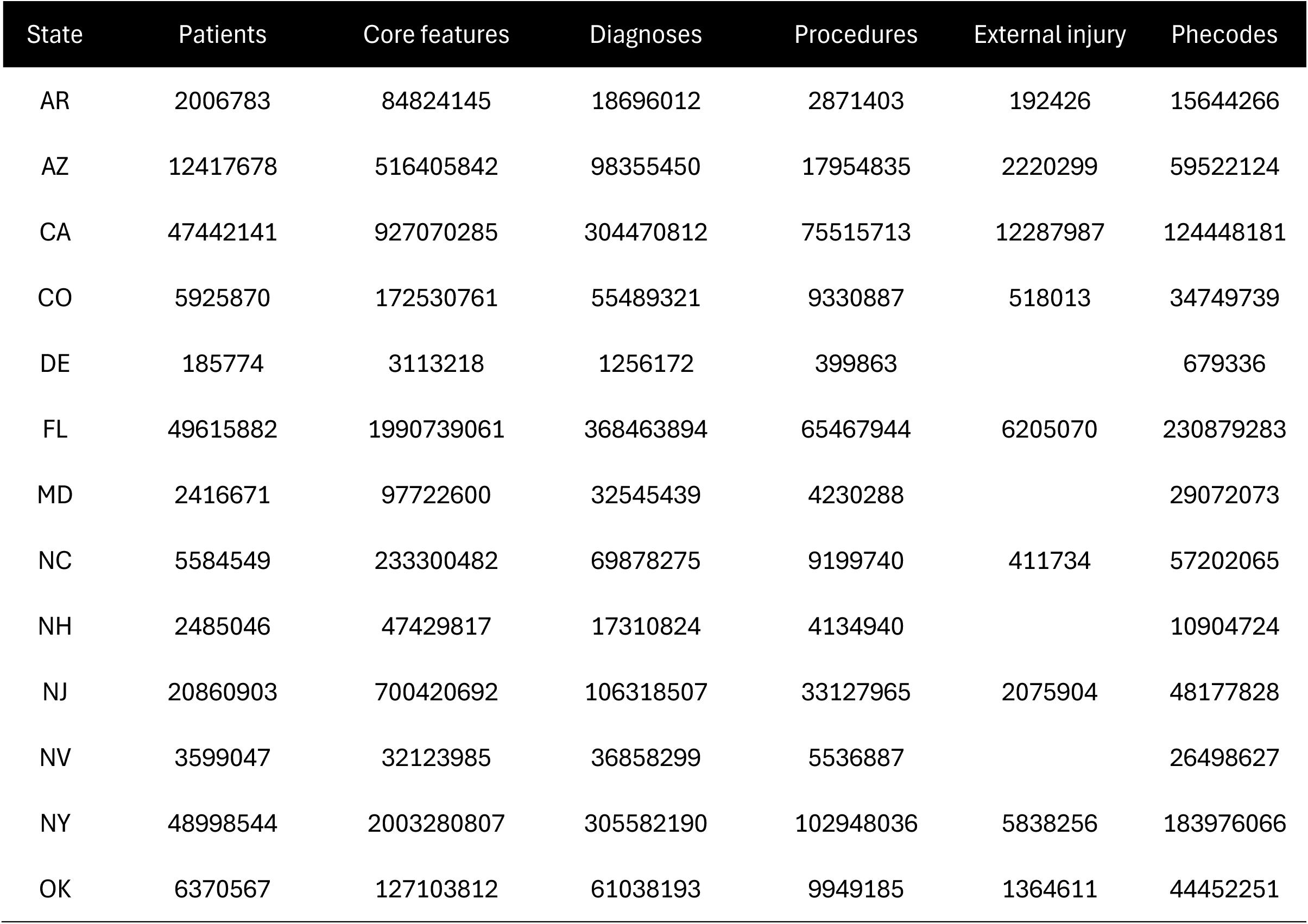
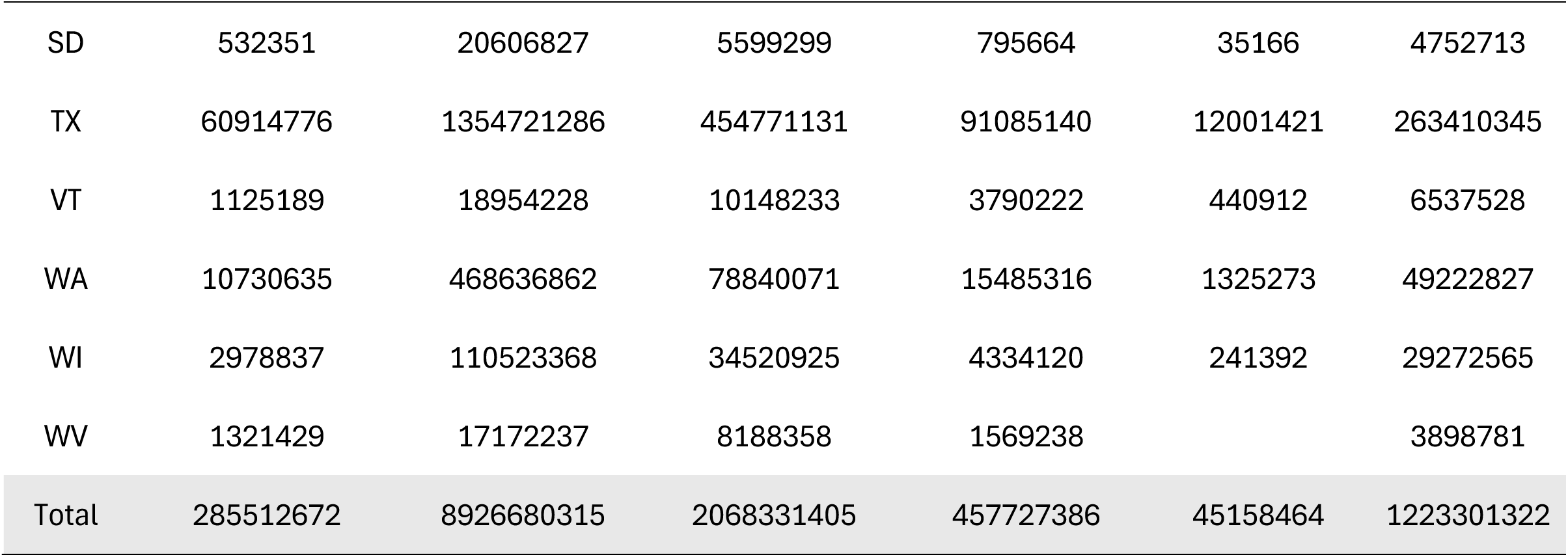
Number of hospital records harmonized using ADAPT.

**Table S3a:**
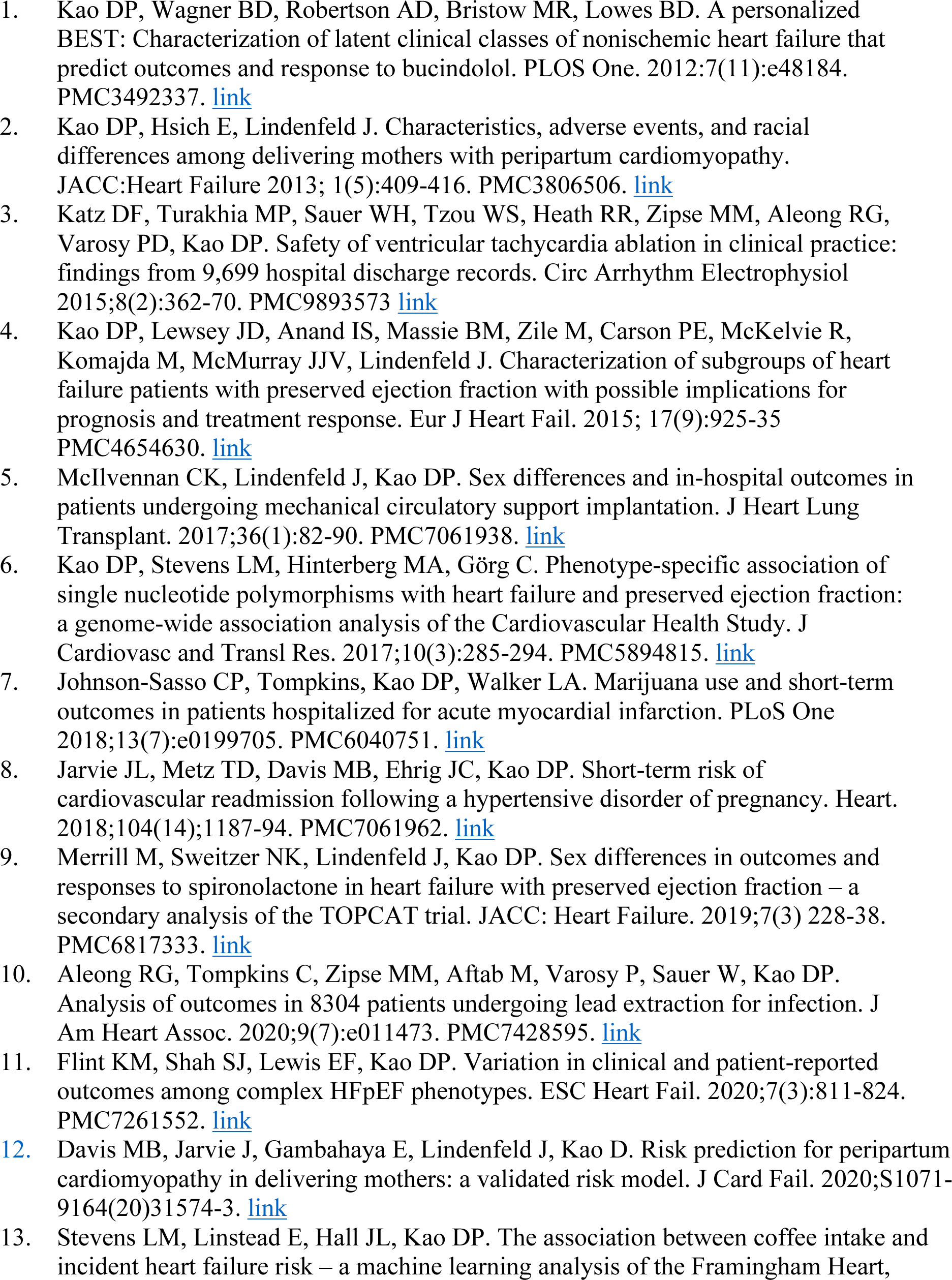
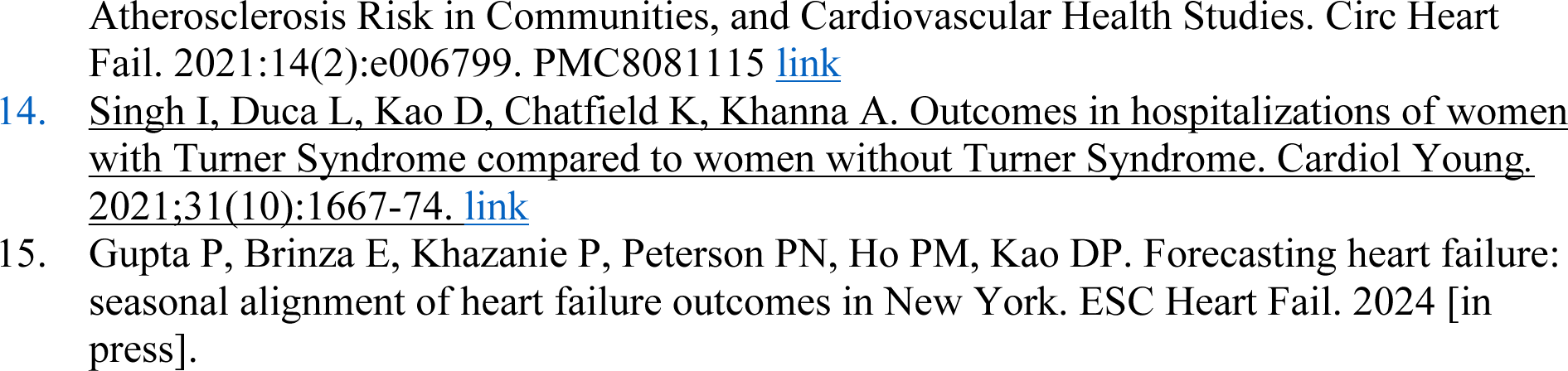
Peer-reviewed publications produced using ADAPT-generated datasets:

**Table S3b:**
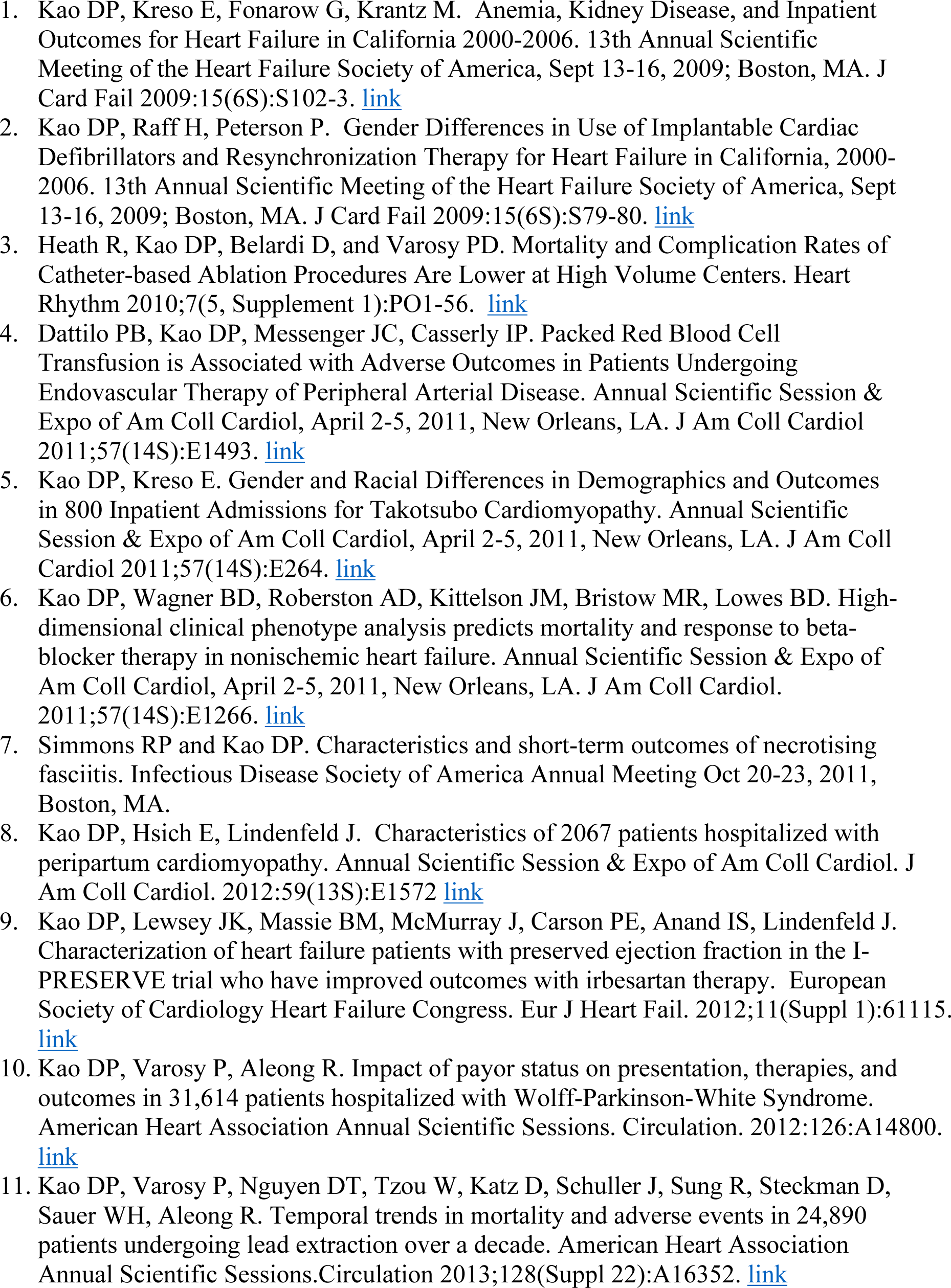
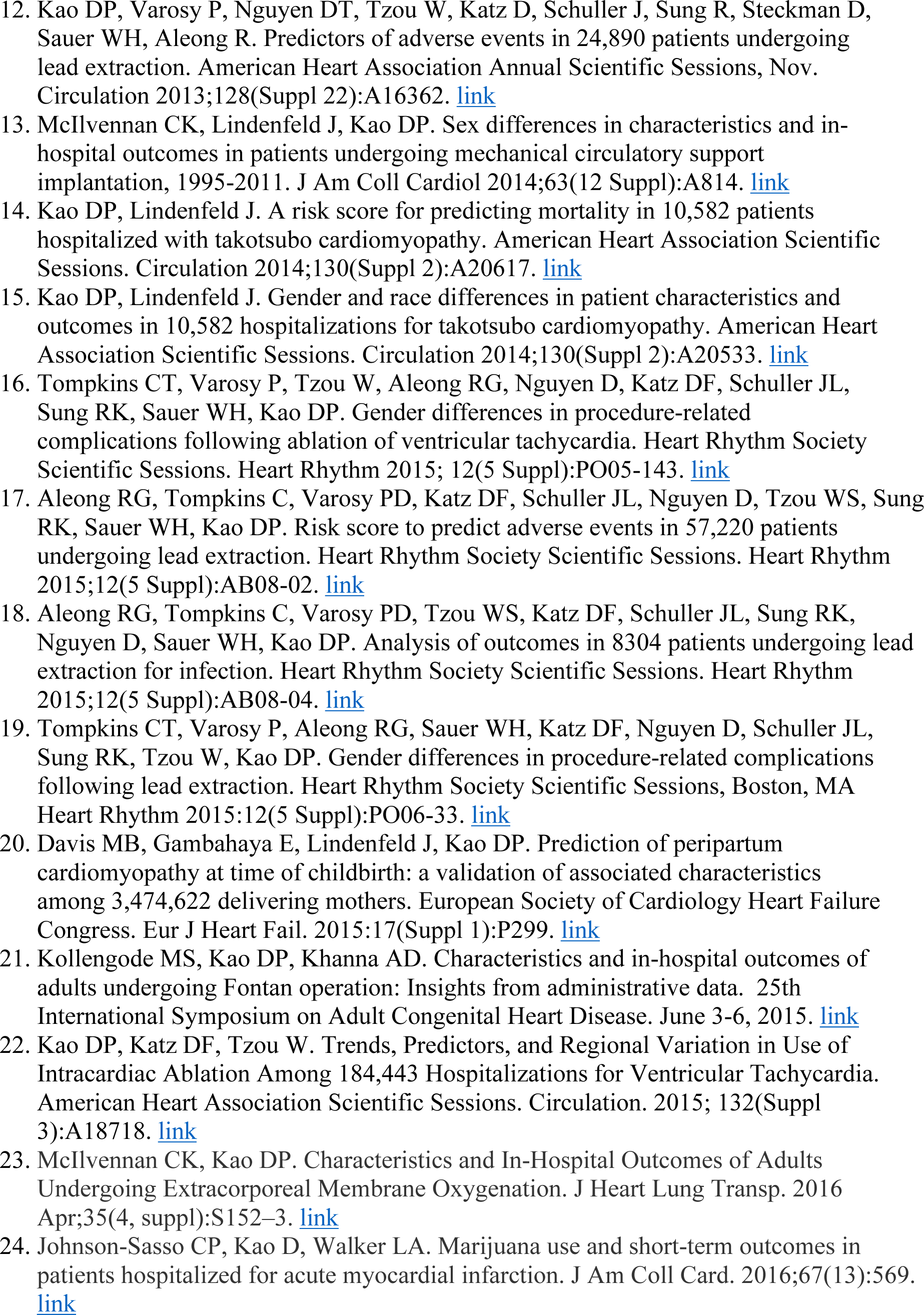
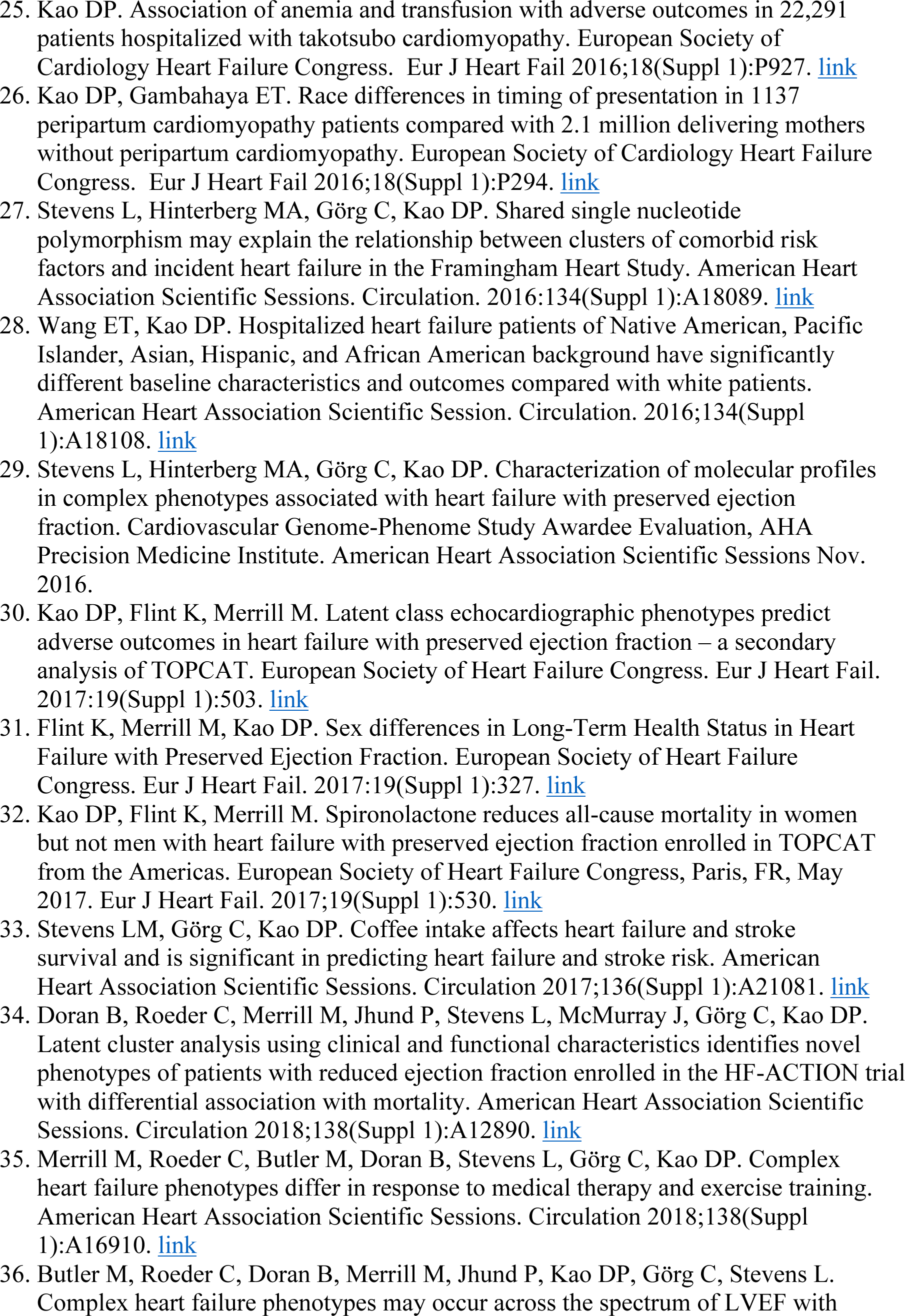
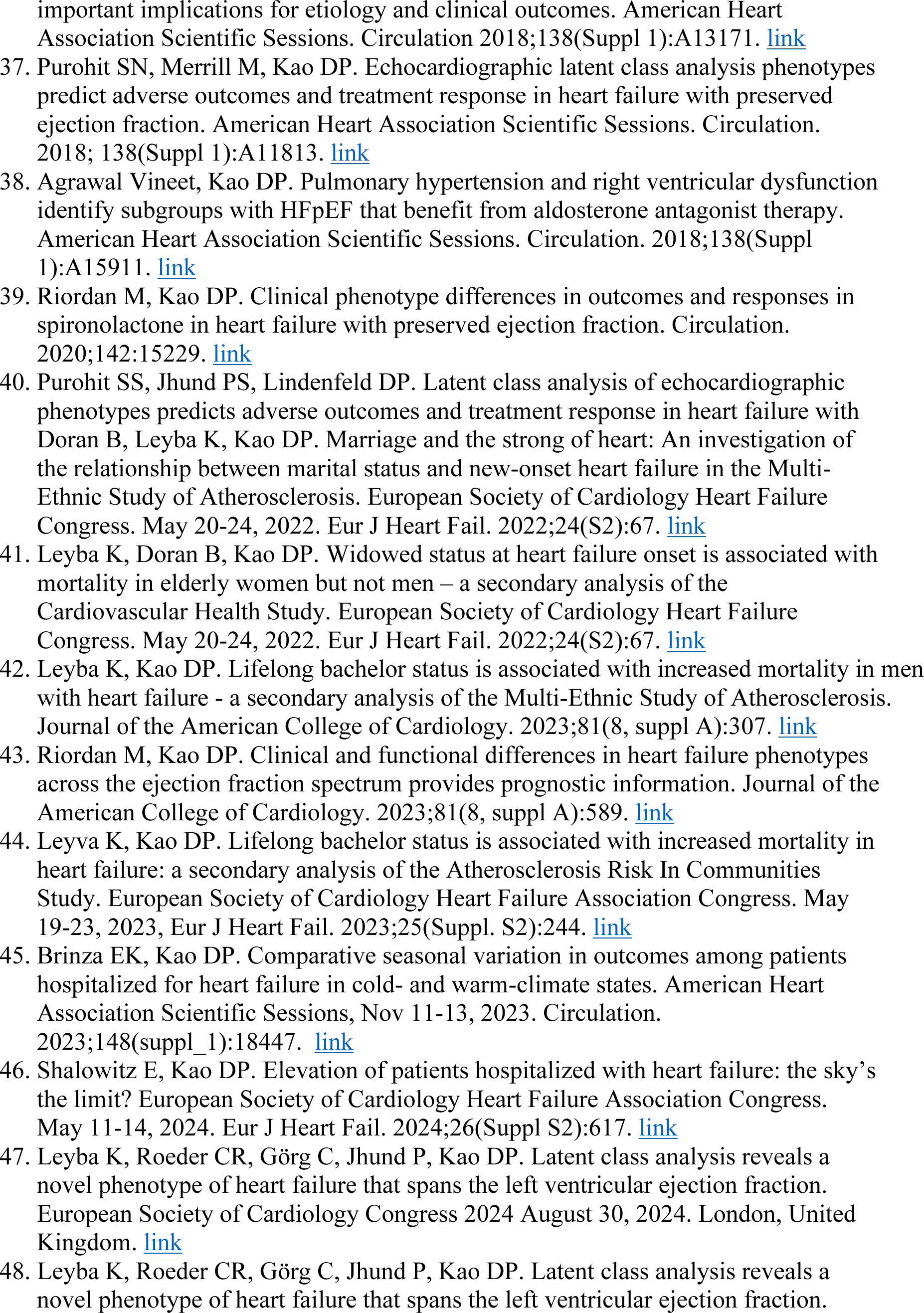
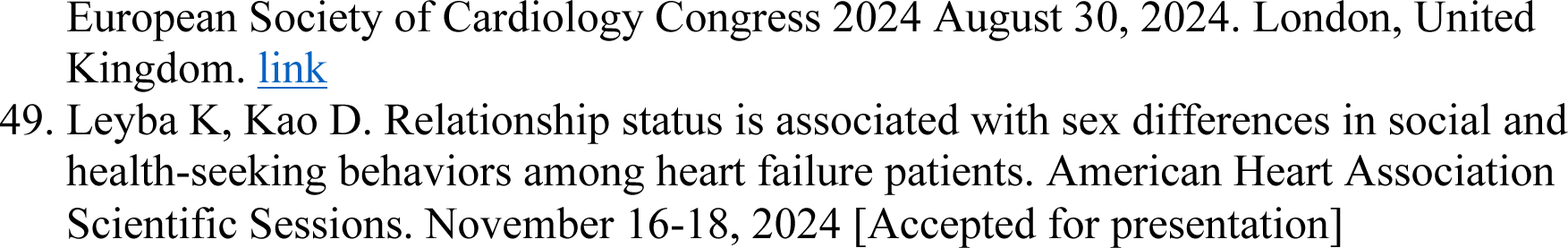
Peer-reviewed abstracts made using ADAPT-generated datasets (n=49):

