## Supplementary material for "Aggregating data to accelerate personalized therapy in heart failure (ADAPT-HF)": Dataset references

**Randomized clinical trials (n=32,442)**

ATHENA-HF (Aldosterone Targeted Neurohormonal Combined with Natriuresis Therapy in Heart Failure trial, n=360): Butler J, Anstrom KJ, Felker GM, Givertz MM, Kalogeropoulos AP, Konstam MA, et al. Efficacy and safety of spironolactone in acute heart failure: The ATHENA-HF randomized clinical trial. JAMA Cardiology. 2017;2(9):950–8.

BEST (The Beta-blocker Evaluation of Survival Trial, n=2708): The Beta-Blocker Evaluation of Survival Trial Investigators. A trial of the beta-blocker bucindolol in patients with advanced chronic heart failure. New England Journal of Medicine. 2001;344(22):1659–67.

CARRESS-HF (Cardiorenal Rescue Study in Acute Decompensated Heart Failure, n=188): Bart BA, Goldsmith SR, Lee KL, Givertz MM, O'Connor CM, Bull DA, et al. Ultrafiltration in Decompensated Heart Failure with Cardiorenal Syndrome. New England Journal of Medicine 2012 Dec 13;367(24):2296–304.

DIG (Digitalis Investigation Group, n=7788). The Digitalis Investigation Group. The effect of digoxin on mortality and morbidity in patients with heart failure. New England Journal of Medicine. 1997;336(8):525–33.

DOSE-AHF (Diuretic Optimization Strategies Evaluation, n=308): Felker GM, Lee KL, Bull DA, Redfield MM, Stevenson LW, Goldsmith SR, et al. Diuretic Strategies in Patients with Acute Decompensated Heart Failure. New England Journal of Medicine. 2011 Mar 3;364(9):797–805.

ESCAPE (Evaluation Study of Congestive Heart Failure and Pulmonary Artery Catheterization Effectiveness, n=433): The ESCAPE Investigators and ESCAPE Study Coordinators. Evaluation study of congestive heart failure and pulmonary artery catheterization effectiveness: the ESCAPE trial. JAMA. 2005 Aug;294(13):1625–33.

EXACT-HF (Xanthine Oxidase Inhibition for Hyperuricemic Heart Failure, n=253): Givertz MM, Anstrom KJ, Redfield MM, Deswal A, Haddad H, Butler J, et al. Effects of xanthine oxidase inhibition in hyperuricemic heart failure patients: The xanthine oxidase inhibition for hyperuricemic heart failure patients (EXACT-HF) study. Circulation. 2015;131(20):1763–71.

FIGHT (Functional Impact of GLP-1 for Heart Failure Treatment, n=300): Margulies KB, Hernandez AF, Redfield MM, Givertz MM, Oliveira GH, Cole R, et al. Effects of Liraglutide on Clinical Stability Among Patients With Advanced Heart Failure and Reduced Ejection Fraction: A Randomized Clinical Trial. JAMA. 2016;316(5):500–8.

GUIDE-IT (Guiding Evidence Based Therapy Using Biomarker Intensified Treatment in Heart Failure trial, n=894): Felker GM, Anstrom KJ, Adams KF, Ezekowitz JA, Fiuzat M, Houston-miller N, et al. Effect of Natriuretic Peptide–Guided Therapy on Hospitalization or Cardiovascular Mortality in High-Risk Patients With Heart Failure and Reduced Ejection Fraction A Randomized Clinical Trial. JAMA. 2017;318(8):713–20.

HF-ACTION (Heart Failure: A Controlled Trial Investigating Outcomes of Exercise Training trial, n=2130): O'Connor CM, Whellan DJ, Lee KL, Keteyian SJ, Cooper LS, Ellis SJ, et al. Efficacy and safety of exercise training in patients with chronic heart failure: HF-ACTION randomized controlled trial. JAMA. 2009;301(14):1439–50.

INDIE-HFpEF (Inorganic Nitrite Delivery to Improve Exercise Capacity in Heart Failure With Preserved Ejection Fraction trial, n=105): Borlaug BA, Anstrom KJ, Lewis GD, Shah SJ, Levine JA, Koepp GA, et al. Effect of Inorganic Nitrite vs Placebo on Exercise Capacity among Patients with Heart Failure with Preserved Ejection Fraction: The INDIE-HFpEF Randomized Clinical Trial. JAMA. 2018 Nov 6;320(17):1764–73.

IRONOUT-HF (Iron Repletion Effects on Oxygen Uptake in Heart Failure, n=225): Lewis GD, Malhotra R, Hernandez AF, McNulty SE, Smith A, Michael Felker G, et al. Effect of Oral Iron Repletion on Exercise Capacity in Patients With Heart Failure With Reduced Ejection Fraction and Iron Deficiency: The IRONOUT HF Randomized Clinical Trial. JAMA. 2017;317(19):1958–66.

LIFE (LCZ696 in Advanced Heart Failure trial, n=335): Mann DL, Givertz MM, Vader JM, Starling RC, Shah P, McNulty SE, et al. Effect of Treatment with Sacubitril/Valsartan in Patients with Advanced Heart Failure and Reduced Ejection Fraction: A Randomized Clinical Trial. JAMA Cardiology. 2022 Jan 1;7(1):17–25.

NEAT-HFpEF (Nitrate's Effect on Activity Tolerance in Heart Failure with Preserved Ejection Fraction trial, n=110): Redfield MM, Anstrom KJ, Levine JA, Koepp GA, Borlaug BA, Chen HH, et al. Isosorbide Mononitrate in Heart Failure with Preserved Ejection Fraction. New England Journal of Medicine. 2015;373:2314–24.

RELAX (Phosphodiesterase-5 Inhibition to Improve Clinical Status and Exercise Capacity in Heart Failure with Preserved Ejection Fraction trial, n=216): Redfield MM, Chen HH, Borlaug BA, Semigran MJ, Lee KL, Lewis G, et al. Effect of phosphodiesterase-5 inhibition on exercise capacity and clinical status in heart failure with preserved ejection fraction: A randomized clinical trial. JAMA. 2013;309(12):1268–77.

ROSE-AHF (Renal Optimization Strategies Evaluation Trial, n=360): Chen HH, Anstrom KJ, Givertz MM, Stevenson LW, Semigran MJ, Goldsmith SR, et al. Low-Dose Dopamine or Low-Dose Nesiritide in Acute Heart Failure With Renal Dysfunction - The ROSE Acute Heart Failure Randomized Trial. JAMA. 2013;310(23):2533–43.

SCD-HEFT (Sudden Cardiac Death in Heart Failure Trial, n=2521): Bardy GH, Lee KL, Mark DB, Poole JE, Packer DL, Boineau R, et al. Amiodarone or an implantable cardioverter-defibrillator for congestive heart failure. New England Journal of Medicine. 2005;351(15):1493–501.

SOLVD-Treat (Studies of Left Ventricular Dysfunction, n=2569): The SOLVD Investigators. Effect of enalapril on survival in patients with reduced left ventricular ejection fractions and congestive heart failure. New England Journal of Medicine. 1991;325(5):293–302.

SOLVD-Prevent (Studies of Left Ventricular Dysfunction, n=4228): The SOLVD Investigators. Effect of enalapril on mortality and the development of heart failure in asymptomatic patients with reduced left ventricular ejection Fractions. New England Journal of Medicine. 1992;327(10):685–91.

STICH (Surgical Treatment for Ischemic Heart Failure, n=1212): Velazquez EJ, Lee KL, Deja MA, Jain A, Sopko G, Marchenko A, et al. Coronary-Artery Bypass Surgery in Patients with Left Ventricular Dysfunction. New England Journal of Medicine. 2011;364(17):1607–16.

TOPCAT (Treatment of Preserved Cardiac Function Heart Failure with an Aldosterone Antagonist, n=3445): Pitt B, Pfeffer MA, Assmann SF, Boineau R, Anand IS, Claggett B, et al. Spironolactone for heart failure with preserved ejection fraction. New England Journal of Medicine. 2014;370(15):1383–92.

Observational cohorts (n=54085)

ARIC (Atherosclerosis Risk in Communities, n=15,792): The ARIC investigators. The Atherosclerosis Risk in Communities (ARIC) Study: design and objectives. American Journal of Epidemiology. 1989 Apr;129(4):687–702.

CHS (Cardiovascular Health Study, n=5888): Fried LP, Borhani NO, Enright P, Furberg CD, Gardin JM, Kronmal RA, et al. The Cardiovascular Health Study: design and rationale. Annals of Epidemiology. 1991 Feb;1(3):263–76.

FHS (Framingham Heart Study): Tsao CW, Vasan RS. Cohort Profile: The Framingham Heart Study (FHS): Overview of milestones in cardiovascular epidemiology. International Journal of Epidemiology. 2015 Dec 1;44(6):1800–13.

*Original* (n=5209): Dawber TR, Meadors GF, Moore FE Jr. Epidemiological approaches to heart disease: the Framingham Study. Am J Public Health Nations Health 1951;41:279–81.

*Offspring* (n=5214): Kannel WB, Feinleib M, McNamara PM et al. An investigation of coronary heart disease in families. The Framingham offspring study. Am J Epidemiol 1979;110:281–90

*Generation 3* (n=4095), Offspring Spouse (n=103): Splansky GL, Corey D, Yang Q et al. The Third Generation Cohort of the NHLBI's Framingham Heart Study: design, recruitment, and initial examination. Am J Epidemiol 2007;165:1328–35.

*Omni 1* (n=507): Kannel WB, Feinleib M, McNamara PM, et al. "Overview of the Framingham Heart Study's design and the Omni Cohort, with baseline characteristics." Ethn Dis. 2000;10(2):220-226

*Omni 2* (n=410): Tighe AP, D'Agostino RB, Demissie S, et al. "Cohort Profile: The Framingham Heart Study (FHS): overview of the Omni Cohorts." International Journal of Epidemiology. 2015;44(6):1800–1813

JHS - Jackson Heart Study (n=3883): Taylor HA, Wilson JG, Jones DW, Sarpong DF, Srinivasan A, Garrison RJ, et al. Toward resolution of cardiovascular health disparities in African Americans: design and methods of the Jackson Heart Study. Ethnicity & disease. 2005;15(4 Suppl 6):S6-4–17.

MESA (Multiethnic Study of Atherosclerosis, n=6814): Bild DE, Bluemke DA, Burke GL, Detrano R, Diez Roux AV, Folsom AR, et al. "Multi-Ethnic Study of Atherosclerosis: Objectives and Design." American Journal of Epidemiology. 2002;156(9):871-881

SOLVD Registry (Studies of LV Dysfunction, n=6273): Bourassa MG, Gurné O, Bangdiwala SI, Ghali JK, Young JB, Rousseau M, et al. Natural history and patterns of current practice in heart failure. The Studies of Left Ventricular Dysfunction (SOLVD) Investigators. Journal of the American College of Cardiology. 1993 Oct;22(4 Suppl A):14A-19A.

**Hospital discharge data (1994-2023, n=288,341,132)**

Healthcare Cost and Utilization Project. Statewide Inpatient Databases. Agency for Healthcare Research and Quality; Rockville, MD. [cited 2025 Nov 2]. Available from: www.hcup-us.ahrq.gov/sidoverview.jsp

- Arizona (2000-2019, n=12,417,678)
- Arkansas (2015-2019, n=2,006,783)
- Colorado (2015-2019, n=2,396,793)
- Florida (2000-2019, n=49,615,882)
- Maryland (2016-2019, n=2,416,671)
- North Carolina (2015-2019, n=5,584,549)
- New York (2015-2019, n=11,706,162)
- South Dakota (2015-2019, n=532,351)
- Washington (2001-2019, n=10,730,635)
- Wisconsin (2015-2019, n=2,978,837)

California (2000-2011, n=47,442,141): Patient Discharge Data (PDD). California Department of Health Care Access and Information. [cited 2025 Nov 2]. Available from: https://hcai.ca.gov/data/request-data/research-data-request-information/

Colorado (2006-2013, n=3,529,077) 1.
Hospital Discharge Data. Center for Health Information and Data Analytics, Colorado Hospital Association. Denver, CO. [cited 2025 Nov 2]. Available from: https://cha.com/center-for-health-information-and-data-analytics/.

Delaware (2010-2020, n=185,774): Hospital Discharge Data. Health and Social Services, Delaware Division of Public Health. Dover, DE [cited 2025 Nov 2]. Available from: https://dhss.delaware.gov/dph/hp/hosp_dis_data/

Louisiana (2010-2023, n=2,828,460): Louisiana Hospital Inpatient Discharge Database. Bureau of Health Informatics, Louisiana Department of Health. Baton Rouge, LA; [cited 2025 Nov 2]. Available from: https://ldh.la.gov/bureau-of-health-informatics/lahidd

New Hampshire (1999-2021, n=2,485,056): Hospital Discharge Data. New Hampshire Department of Health and Human Services, Concord, NH [cited 2025 Nov 2]. Available from: https://www.dhhs.nh.gov/programs-services/population-health/health-statistics-informatics/hospital-discharge-data

New Jersey (1997-2011, n=20,860,903): NJ Hospital Discharge Data Collection System. Office of Health Care Quality Assessment, New Jersey Department of Health, Trenton, NJ [cited 2025 Nov 2]. Available from: https://www.nj.gov/health/healthcarequality/health-care-professionals/njddcs/

Nevada (2010-2020, n=3,599,047): Inpatient Admission Data [Internet]. Office of Analytics, Nevada Department of Human Services, Carson City, NV. [cited 2025 Nov 2]. Available from: https://www.dhs.nv.gov/Programs/dhhs-office-of-analytics/

New York (1994-2007, n=37,292,382): Statewide Planning and Research Cooperative System (SPARCS). New York Department of Health; Albany, NY. [cited 2025 Nov 2]. Available from: http://www.health.ny.gov/statistics/sparcs/

Oklahoma (2010-2023, n=6,370,567): Hospital Discharge Data. Oklahoma State Department of Health, Oklahoma City, OK [cited 2025 Nov 2]. Available from: https://oklahoma.gov/health/health-education/data-and-statistics/center-for-health-statistics/health-care-information/hospital-discharge-and-outpatient-asc-surgery-data.html

Texas (1999-2019, n=60,914,776): Texas Inpatient Public Use Data File. Texas Department of State Health Services, Austin, TX [cited 2025 Nov 2]. Available from: https://www.dshs.texas.gov/center-health-statistics/texas-health-care-information-collection/download-and-purchase-data/texas-inpatient-public-use-data-file-pudf

Vermont (2002-2022, n=1,125,189): Vermont Uniform Hospital Discharge Data System. Green Mountain Care Board, Montpelier VT [cited 2025 Nov 2]. Available from: https://gmcboard.vermont.gov/data-release-0/vermont-uniform-hospital-discharge-data-system-vuhdds

West Virginia (2003-2007, n=1,321,429): West Virginia Hospital Inpatient Data System. West Virginia Health Care Authority, Charleston, WV [cited 2025 Nov 2]. Available from: https://hca.wv.gov/fdhome/HospInpatientData/Pages/default.aspx
