## Supplementary material for "Aggregating data to accelerate personalized therapy in heart failure (ADAPT-HF)": Harmonization scripts: Cohort dates amd outcomes.R

```
library(sqldf)
library(Hmisc)
library(plotrix)
library(reshape2)
library(descr)
library(dplyr)
library(relper)
library(stringr)
library(bigrquery)
library(haven)
library(googleCloudStorageR)
library(arrow)
library(data.table)
library(parquetize)

source("~/Dropbox/R scripts/Misc clinical scripts.R")
source("~/Dropbox/R scripts/echo scripts.R")


visit_yrs <- read.csv('~/Dropbox/ADAPT-HF/Master HDCP files/Cohort visit yrs.csv')

data_fields <- 
  c("study",
    "patientid",
    "visit_yr",
    "visitdays",
    "form",
    "variable",
    "value",
    "datapoint",
    "study_field",
    "cohort",
    "cohort_name",
    "age_obs")


outcomes_list <- c("dth_status","dth_dt",
                   "cvdth_status","cvdth_dt",
                   "hfdth_status","hfdth_dt",
                   "hfhosp_ef_val","hfhosp_ef_cat",
                   "noncvdth_status","noncvdth_dt",
                   "hosp_status","hosp_dt",
                   "cadhosp_status","cadhosp_dt",
                   "cvhosp_status","cvhosp_dt",
                   "hfhosp_status","hfhosp_dt",
                   "noncvhosp_status","noncvhosp_dt")

dates_long_fields <-
  c("study",
    "cohort",
    "cohort_name",
    "patientid",
    "sex",
    "visit_yr",
    "visitdays",
    "age_obs")

######### ---- OUTCOMES ---- #######

# ..%%%%%%%..%%.....%%.%%%%%%%%..%%%%%%...%%%%%%%..%%.....%%.%%%%%%%%..%%%%%%.
# .%%.....%%.%%.....%%....%%....%%....%%.%%.....%%.%%%...%%%.%%.......%%....%%
# .%%.....%%.%%.....%%....%%....%%.......%%.....%%.%%%%.%%%%.%%.......%%......
# .%%.....%%.%%.....%%....%%....%%.......%%.....%%.%%.%%%.%%.%%%%%%....%%%%%%.
# .%%.....%%.%%.....%%....%%....%%.......%%.....%%.%%.....%%.%%.............%%
# .%%.....%%.%%.....%%....%%....%%....%%.%%.....%%.%%.....%%.%%.......%%....%%
# ..%%%%%%%...%%%%%%%.....%%.....%%%%%%...%%%%%%%..%%.....%%.%%%%%%%%..%%%%%%.

#### FHS ####

fhs1_chfinit <- fread('~/Dropbox/BioLINCC files/Framingham cohort/datasets/CSV/vr_chfinit_2013_a_0828d.csv',na.strings=c("NA","","NULL"))
fhs1_survcvd <- fread('~/Dropbox/BioLINCC files/Framingham cohort/datasets/CSV/vr_survcvd_2018_a_1267d.csv',na.strings=c("NA","","NULL"))
fhs1_survdth <- fread('~/Dropbox/BioLINCC files/Framingham cohort/datasets/CSV/vr_survdth_2018_a_1268d.csv',na.strings=c("NA","","NULL"))
fhs1_survstk <- fread('~/Dropbox/BioLINCC files/Framingham cohort/datasets/CSV/vr_svstk_2018_a_1269d.csv',na.strings=c("NA","","NULL"))
fhs1_soechf <- fread('~/Dropbox/BioLINCC files/Framingham cohort/datasets/CSV/VR_SOECHF_2016_a_1070d.csv',na.strings=c("NA","","NULL"))
fhs1_afcum <- fread('~/Dropbox/BioLINCC files/Framingham cohort/datasets/CSV/VR_AFCUM_2019_a_1186d_v1.csv',na.strings=c("NA","","NULL")) # Cumulative Atrial fibrillation
fhs1_survaf <- fread('~/Dropbox/BioLINCC files/Framingham cohort/datasets/CSV/VR_SURVAF_2018_a_1272D.csv',na.strings=c("NA","","NULL")) # Cumulative Atrial fibrillation
fhs1_cancer <- fread('~/Dropbox/BioLINCC files/Framingham cohort/datasets/CSV/VR_CANCER_2019_A_1162D.csv',na.strings=c("NA","","NULL")) # Incident cancer
fhs1_diab <- fread('~/Dropbox/BioLINCC files/Framingham cohort/datasets/CSV/VR_DIAB_EX28_0_0601D.csv',na.strings=c("NA","","NULL")) # Diabetes status by cycle
fhs1_soe <- fread('~/Dropbox/BioLINCC files/Framingham cohort/datasets/CSV/VR_SOE_2019_A_1217D.csv',na.strings=c("NA","","NULL"))

names(fhs1_chfinit) <- toupper(names(fhs1_chfinit))
names(fhs1_survcvd) <- toupper(names(fhs1_survcvd))
names(fhs1_survdth) <- toupper(names(fhs1_survdth))
names(fhs1_soechf) <- toupper(names(fhs1_soechf))
names(fhs1_afcum) <- toupper(names(fhs1_afcum))
names(fhs1_survaf) <- toupper(names(fhs1_survaf))
names(fhs1_cancer) <- toupper(names(fhs1_cancer))
names(fhs1_diab) <- toupper(names(fhs1_diab))
names(fhs1_soe) <- toupper(names(fhs1_soe))


fhs2_chfinit <- fread('~/Dropbox/BioLINCC files/Framingham offspring/datasets/CSV/vr_chfinit_2013_a_0828d.csv',na.strings=c("NA","","NULL"))
fhs2_survcvd <- fread('~/Dropbox/BioLINCC files/Framingham offspring/datasets/CSV/vr_survcvd_2021_a_1453d.csv',na.strings=c("NA","","NULL"))
fhs2_survdth <- fread('~/Dropbox/BioLINCC files/Framingham offspring/datasets/CSV/vr_survdth_2021_a_1452d.csv',na.strings=c("NA","","NULL"))
fhs2_survstk <- fread('~/Dropbox/BioLINCC files/Framingham offspring/datasets/CSV/vr_svstk_2021_a_1455d.csv',na.strings=c("NA","","NULL"))
fhs2_soechf <- fread('~/Dropbox/BioLINCC files/Framingham offspring/datasets/CSV/VR_SOECHF_2016_A_1070D.csv',na.strings=c("NA","","NULL"))
fhs2_afcum <- fread('~/Dropbox/BioLINCC files/Framingham offspring/datasets/CSV/VR_AFCUM_2022_A_1412D.csv',na.strings=c("NA","","NULL")) # Cumulative Atrial fibrillation
fhs2_survaf <- fread('~/Dropbox/BioLINCC files/Framingham offspring/datasets/CSV/VR_SURVAF_2021_A_1456D.csv',na.strings=c("NA","","NULL")) # Cumulative Atrial fibrillation
fhs2_cancer <- fread('~/Dropbox/BioLINCC files/Framingham offspring/datasets/CSV/VR_CANCER_2019_A_1162D.csv',na.strings=c("NA","","NULL")) # Incident cancer
fhs2_diab <- fread('~/Dropbox/BioLINCC files/Framingham offspring/datasets/CSV/vr_diab_ex10_1b_1489d.csv',na.strings=c("NA","","NULL")) # Diabetes status by cycle
fhs2_soe <- fread('~/Dropbox/BioLINCC files/Framingham offspring/datasets/CSV/VR_SOE_2022_A_1424D.csv',na.strings=c("NA","","NULL"))

names(fhs2_chfinit) <- toupper(names(fhs2_chfinit))
names(fhs2_survcvd) <- toupper(names(fhs2_survcvd))
names(fhs2_survdth) <- toupper(names(fhs2_survdth))
names(fhs2_survstk) <- toupper(names(fhs2_survstk))
names(fhs2_soechf) <- toupper(names(fhs2_soechf))
names(fhs2_afcum) <- toupper(names(fhs2_afcum))
names(fhs2_survaf) <- toupper(names(fhs2_survaf))
names(fhs2_cancer) <- toupper(names(fhs2_cancer))
names(fhs2_diab) <- toupper(names(fhs2_diab))
names(fhs2_soe) <- toupper(names(fhs2_soe))

fhs2_afcum$ORIGIN_SOURCE <- NA

fhs3_chfinit <- fread('~/Dropbox/BioLINCC files/Framingham third generation/datasets/CSV/vr_chfinit_2013_a_0828d.csv',na.strings=c("NA","","NULL"))
fhs3_survcvd <- fread('~/Dropbox/BioLINCC files/Framingham third generation/datasets/CSV/vr_survcvd_2019_a_1334d.csv',na.strings=c("NA","","NULL"))
fhs3_survdth <- fread('~/Dropbox/BioLINCC files/Framingham third generation/datasets/CSV/vr_survdth_2019_a_1337d.csv',na.strings=c("NA","","NULL"))
fhs3_survstk <- fread('~/Dropbox/BioLINCC files/Framingham third generation/datasets/CSV/vr_svstk_2019_a_1335d.csv',na.strings=c("NA","","NULL"))
fhs3_soechf <- fread('~/Dropbox/BioLINCC files/Framingham third generation/datasets/CSV/VR_SOECHF_2016_a_1070D.csv',na.strings=c("NA","","NULL"))
fhs3_afcum <- fread('~/Dropbox/BioLINCC files/Framingham third generation/datasets/CSV/VR_AFCUM_2021_A_1366D.csv',na.strings=c("NA","","NULL")) # Cumulative Atrial fibrillation
fhs3_survaf <- fread('~/Dropbox/BioLINCC files/Framingham third generation/datasets/CSV/VR_SURVAF_2019_A_1338D.csv',na.strings=c("NA","","NULL")) # Cumulative Atrial fibrillation
fhs3_cancer <- fread('~/Dropbox/BioLINCC files/Framingham third generation/datasets/CSV/VR_CANCER_2019_A_1162D.csv',na.strings=c("NA","","NULL")) # Incident cancer
fhs3_diab <- fread('~/Dropbox/BioLINCC files/Framingham third generation/datasets/CSV/VR_DIAB_EX03_3b_1312D.csv',na.strings=c("NA","","NULL")) # Diabetes status by cycle
fhs3_soe <- fread('~/Dropbox/BioLINCC files/Framingham third generation/datasets/CSV/VR_SOE_2020_A_1340D.csv',na.strings=c("NA","","NULL"))

fhs3_afcum$ORIGIN_SOURCE <- NA


dm_names <- unique(c(names(fhs1_diab),names(fhs2_diab),names(fhs3_diab)))

fhs1_diab[,dm_names[!dm_names %in% names(fhs1_diab)]] <- NA
fhs2_diab[,dm_names[!dm_names %in% names(fhs2_diab)]] <- NA
fhs3_diab[,dm_names[!dm_names %in% names(fhs3_diab)]] <- NA


fhs_chfinit <- rbind(fhs1_chfinit,fhs2_chfinit,fhs3_chfinit)
fhs_survcvd <- rbind(fhs1_survcvd,fhs2_survcvd,fhs3_survcvd)
fhs_survdth <- rbind(fhs1_survdth,fhs2_survdth,fhs3_survdth)
fhs_survstk <- rbind(fhs1_survstk,fhs2_survstk,fhs3_survstk)
fhs_survaf <- rbind(fhs1_survaf,fhs2_survaf,fhs3_survaf)
fhs_afcum <- rbind(fhs1_afcum,fhs2_afcum,fhs3_afcum)
fhs_soechf <- rbind(fhs1_soechf,fhs2_soechf,fhs3_soechf)
fhs_soe <- rbind(fhs1_soe,fhs2_soe,fhs3_soe)
fhs_cancer <- rbind(fhs1_cancer,fhs2_cancer,fhs3_cancer)
fhs_diab <- rbind(fhs1_diab,fhs2_diab,fhs3_diab)

fhs_survca <- subset(fhs_cancer,(!TOPO==173)|(TOPO==173&HIST>=8720&HIST<=8790)|HIST==8247)

fhs_survca <- 
  fhs_cancer %>%
  group_by(IDTYPE,PID) %>%
  filter(D_DATE == min(D_DATE,na.rm=T))

fhs_survca_first_any <- 
  as.data.table(sqldf("select distinct PID,IDTYPE,D_DATE
        from fhs_survca"))

fhs_survdth$dth_status <- 0 
fhs_survdth$dth_status[!is.na(fhs_survdth$DATEDTH)] <- 1 
fhs_survdth$dth_dt <- fhs_survdth$DATEDTH
fhs_survdth$dth_dt[fhs_survdth$dth_status==0] <- fhs_survdth$LASTCON[fhs_survdth$dth_status==0]

fhs_survdth$cvdth_status <- 0 
fhs_survdth$cvdth_status[fhs_survdth$CVDDEATH==1] <- 1
fhs_survdth$cvdth_dt <- fhs_survdth$dth_dt

fhs_survdth$noncvdth_status <- 0 
fhs_survdth$noncvdth_status[fhs_survdth$CVDDEATH==0] <- 1
fhs_survdth$noncvdth_dt <- fhs_survdth$dth_dt

names(fhs_survcvd)[c(3,4,5,6,7,8)] <- 
  c("cadhosp_status",
    "cadhosp_dt",
    "hfhosp_status",
    "hfhosp_dt",
    "cvhosp_status",
    "cvhosp_dt")

names(fhs_survstk)[3:6] <-
  c("cvahosp_status",
    "cva_type",
    "cva_lacunar",
    "cvahosp_dt")


fhs_outcomes <- merge(fhs_survdth[,c("PID","IDTYPE",
                                     "dth_status","dth_dt",
                                     "cvdth_status","cvdth_dt",
                                     "noncvdth_status","noncvdth_dt")],
                      fhs_survcvd[,c("PID","IDTYPE",
                                     "cadhosp_status","cadhosp_dt",
                                     "cvhosp_status","cvhosp_dt",
                                     "hfhosp_status","hfhosp_dt")])

fhs_outcomes <-
  merge(fhs_outcomes,
        fhs_survstk[,c("PID","IDTYPE","cvahosp_status","cva_type","cvahosp_dt")],
        by=c("PID","IDTYPE"),
        all.x=T)


## Ischemic stroke

fhs_outcomes$ischem_cvahosp_status <- 0
fhs_outcomes$ischem_cvahosp_status[
  fhs_outcomes$cva_type %in% 
    c(11,13)] <- 1


fhs_outcomes$ischem_cvahosp_dt[
  fhs_outcomes$ischem_cvahosp_status==1] <-
  fhs_outcomes$cvahosp_dt[
    fhs_outcomes$ischem_cvahosp_status==1]

fhs_outcomes$ischem_cvahosp_dt[
  fhs_outcomes$ischem_cvahosp_status==0] <-
  fhs_outcomes$dth_dt[
    fhs_outcomes$ischem_cvahosp_status==0]


fhs_outcomes[
  fhs_outcomes$cva_type %in% 
    c(10,16,17,19),
  c("ischem_cvahosp_status",
    "ischem_cvahosp_dt")] <- NA


## Hemorrhagic

fhs_outcomes$hemo_cvahosp_status <- 0
fhs_outcomes$hemo_cvahosp_status[
  fhs_outcomes$cva_type %in% 
    c(14,15)] <- 1


fhs_outcomes$hemo_cvahosp_dt[
  fhs_outcomes$hemo_cvahosp_status==1] <-
  fhs_outcomes$cvahosp_dt[
    fhs_outcomes$hemo_cvahosp_status==1]

fhs_outcomes$hemo_cvahosp_dt[
  fhs_outcomes$hemo_cvahosp_status==0] <-
  fhs_outcomes$dth_dt[
    fhs_outcomes$hemo_cvahosp_status==0]

fhs_outcomes[
  fhs_outcomes$cva_type %in% 
    c(10,16,17,19),
  c("hemo_cvahosp_status",
    "hemo_cvahosp_dt")] <- NA


fhs_outcomes <- merge(fhs_outcomes,
                      fhs_chfinit[,c("PID",
                                     "IDTYPE",
                                     "CHF011D",
                                     "CHF017",
                                     "EFLT50")],
                      by.x=c("PID","IDTYPE","hfhosp_dt"),
                      by.y=c("PID","IDTYPE","CHF011D"),
                      all.x=T)


fhs_outcomes$hfhosp_ef_cat[fhs_outcomes$CHF017==0] <- "Normal"
fhs_outcomes$hfhosp_ef_cat[fhs_outcomes$CHF017==3] <- "Mildly reduced"
fhs_outcomes$hfhosp_ef_cat[fhs_outcomes$CHF017==2] <- "Moderately reduced"
fhs_outcomes$hfhosp_ef_cat[fhs_outcomes$CHF017==1] <- "Severely reduced"


fhs_outcomes$study <- "FHS"

names(fhs_outcomes)[1:2] <- 
  c("patientid","cohort")

fhs_outcomes$patientid <- as.character(fhs_outcomes$patientid)

outcomes_to_fill <- outcomes_list[!outcomes_list %in% names(fhs_outcomes)]
outcomes_to_fill <- outcomes_to_fill[!is.na(outcomes_to_fill)]
fhs_outcomes[,outcomes_to_fill] <- NA


write_parquet(fhs_outcomes,
              "~/Dropbox/ADAPT-HF/Master HDCP files/Cohort data/fhs_outcomes.parquet")

if (bq_table_exists("harmonization-286013.cohorts.fhs_outcomes"))
{bq_table_delete("harmonization-286013.cohorts.fhs_outcomes")}
bq_table_upload(x="harmonization-286013.cohorts.fhs_outcomes",
                fhs_outcomes,quiet=T,
                create_disposition="CREATE_IF_NEEDED",
                write_disposition="WRITE_TRUNCATE")


#### ARIC Cohort ####

aric_incps <- fread('~/Dropbox/BioLINCC files/ARIC/Main_Study/cohort_incident_recurrent/csv/incps19.csv',na.strings=c("NA","","NULL"))
aric_hfcchips <- fread('~/Dropbox/BioLINCC files/ARIC/Main_Study/cohort_HF/csv/hfcchips19.csv',na.strings=c("NA","","NULL"))

aric_ccelps19 <- fread('~/Dropbox/BioLINCC files/ARIC/Main_Study/cohort_HF/csv/hfccelps19.csv',na.strings=c("NA","","NULL"))


aric_outcomes <- as.data.table(sqldf('select distinct ID_C, 
                        C7_INCHF_P_V5 as hf_after_exam5_status, 
                        C7_FT_INCHF_P_V5 as hf_after_exam5_dys,
                        ISP19 as cadhosp_status,
                        C7_FUTIMEA as cadhosp_dt,
                        PRVCHD05 as baseline_chd,
                        INCHF19 as hfhosp_status, 
                        C7_FUTIMEHF as hfhosp_dt,
                        PREVHF01 as baseline_hf,
                        DEAD19 as dth_status, 
                        FUDTH19 as dth_dt,
                        UCOD as dth_cause
                       from aric_incps'))


### CV death was defined as death with ICD‐9 code 401‐459 or ICD‐10 code I10‐I99 
# PMC4710457
# PMC6343542
# PMC8109763

aric_outcomes$cvdth_status <- 0
aric_outcomes$cvdth_status[between(as.numeric(substr(aric_outcomes$dth_cause,1,3)),401,459)|
                             substr(aric_outcomes$dth_cause,1,1)=="I"] <- 1

aric_outcomes$noncvdth_status[aric_outcomes$dth_status==0|aric_outcomes$cvdth_status==1] <- 0
aric_outcomes$noncvdth_status[aric_outcomes$cvdth_status==0&aric_outcomes$dth_status==1] <- 1

aric_outcomes$cvdth_dt <- aric_outcomes$dth_dt
aric_outcomes$noncvdth_dt <- aric_outcomes$dth_dt

aric_outcomes$hfhosp_status[aric_outcomes$baseline_hf==1&!is.na(aric_outcomes$baseline_hf)] <- 1
aric_outcomes$hfhosp_dt[aric_outcomes$baseline_hf==1&!is.na(aric_outcomes$baseline_hf)] <- 0

aric_hf_cohort <- fread('~/Dropbox/BioLINCC files/ARIC/Main_Study/cohort_HF/csv/hfcoccps19.csv',na.strings=c("NA","","NULL"))
aric_hf_cohort$visit_yr <- floor(aric_hf_cohort$HFDAYS/365)
aric_hfchfaps <- fread('~/Dropbox/BioLINCC files/ARIC/Main_Study/cohort_HF/csv/hfchfaps19.csv',na.strings=c("NA","","NULL"))

aric_hf_cohort <- 
  merge(aric_hf_cohort,
        aric_outcomes[,c("ID_C","cvdth_status","noncvdth_status")],
        by="ID_C",
        all.x=T)

aric_hfchfaps$dispo[aric_hfchfaps$HFAA0D=="D"] <- "Deceased"
aric_hfchfaps$dispo[aric_hfchfaps$HFAA0D1=="Y"] <- "Home"
aric_hfchfaps$dispo[aric_hfchfaps$HFAA0D2=="Y"] <- "Home"
aric_hfchfaps$dispo[aric_hfchfaps$HFAA0D3=="Y"] <- "Short term care"
aric_hfchfaps$dispo[aric_hfchfaps$HFAA0D4=="Y"] <- "Home"
aric_hfchfaps$dispo[aric_hfchfaps$HFAA0D5=="Y"] <- "Long term care"
aric_hfchfaps$dispo[aric_hfchfaps$HFAA0D6=="Y"] <- "Hospice"
aric_hfchfaps$dispo[aric_hfchfaps$HFAA0D7=="Y"] <- "Left AMA"

aric_hfchfaps$adm_wt_kg[aric_hfchfaps$HFAA20A1=="K"&!is.na(aric_hfchfaps$HFAA20A1)] <- 
  aric_hfchfaps$HFAA20A[aric_hfchfaps$HFAA20A1=="K"&!is.na(aric_hfchfaps$HFAA20A1)] 

aric_hfchfaps$adm_wt_kg[aric_hfchfaps$HFAA20A1=="L"&!is.na(aric_hfchfaps$HFAA20A1)] <- 
  aric_hfchfaps$HFAA20A[aric_hfchfaps$HFAA20A1=="L"&!is.na(aric_hfchfaps$HFAA20A1)] / 2.2 

aric_hfchfaps$dc_wt_kg[aric_hfchfaps$HFAA20B1=="K"&!is.na(aric_hfchfaps$HFAA20B1)] <- 
  aric_hfchfaps$HFAA20B[aric_hfchfaps$HFAA20B1=="K"&!is.na(aric_hfchfaps$HFAA20B1)] 

aric_hfchfaps$dc_wt_kg[aric_hfchfaps$HFAA20B1=="L"&!is.na(aric_hfchfaps$HFAA20B1)] <- 
  aric_hfchfaps$HFAA20B[aric_hfchfaps$HFAA20B1=="L"&!is.na(aric_hfchfaps$HFAA20B1)] / 2.2 

aric_hfchfaps$wt_chg <- 
  aric_hfchfaps$dc_wt_kg-aric_hfchfaps$adm_wt_kg

aric_hfchfaps$wt_chg_pct <- 
  (aric_hfchfaps$dc_wt_kg-aric_hfchfaps$adm_wt_kg)/aric_hfchfaps$adm_wt_kg


aric_hfchfaps$lvh_aric_grade[!aric_hfchfaps$HFAA29D1 %in% c("N","P")] <-
  aric_hfchfaps$HFAA29D1[!aric_hfchfaps$HFAA29D1 %in% c("N","P")]

aric_hfchfaps$lvh_aric_present[aric_hfchfaps$HFAA29D1 %in% c("E")] <- "No"
aric_hfchfaps$lvh_aric_present[aric_hfchfaps$HFAA29D1 %in% c("D","E","M","P","S")] <- "Yes"

aric_hfchfaps$HFAA29C1[aric_hfchfaps$ID_S0 %in% c("S111312",
                                                  "S113765",
                                                  "S114430",
                                                  "S115152",
                                                  "S116317",
                                                  "S116365",
                                                  "S117549",
                                                  "S118803",
                                                  "S120190",
                                                  "S120281",
                                                  "S122791",
                                                  "S124361",
                                                  "S125222",
                                                  "S125919",
                                                  "S127584",
                                                  "S129313",
                                                  "S130485",
                                                  "S132063",
                                                  "S135940",
                                                  "S136416",
                                                  "S136850",
                                                  "S138443",
                                                  "S138586",
                                                  "S140721",
                                                  "S141731",
                                                  "S143363",
                                                  "S143410",
                                                  "S143485",
                                                  "S144028",
                                                  "S144323",
                                                  "S144348",
                                                  "S145732",
                                                  "S146215",
                                                  "S147470",
                                                  "S148454",
                                                  "S150389",
                                                  "S152207",
                                                  "S173877",
                                                  "S185468",
                                                  "S196087",
                                                  "S196969",
                                                  "S199559",
                                                  "S203483",
                                                  "S206479")] <- 1


aric_hfchfaps$HFAA29C1[aric_hfchfaps$ID_S0 %in%  c("S112486",
                                                   "S113665",
                                                   "S114916",
                                                   "S115209",
                                                   "S124266",
                                                   "S131451",
                                                   "S132554",
                                                   "S137531",
                                                   "S140461",
                                                   "S146420",
                                                   "S150799",
                                                   "S194585",
                                                   "S204599",
                                                   "S217931",
                                                   "S218887")] <- 2


aric_hfchfaps$HFAA29C1[aric_hfchfaps$ID_S0 %in% c("S127574",
                                                  "S136289",
                                                  "S155314",
                                                  "S155682",
                                                  "S212030")] <- 3


aric_hfchfaps$ivs_cm[aric_hfchfaps$HFAA29C1==1&!is.na(aric_hfchfaps$HFAA29C1)] <-
  aric_hfchfaps$HFAA29C[aric_hfchfaps$HFAA29C1==1&!is.na(aric_hfchfaps$HFAA29C1)]

aric_hfchfaps$ivs_cm[aric_hfchfaps$HFAA29C1==2&!is.na(aric_hfchfaps$HFAA29C1)] <-
  aric_hfchfaps$HFAA29C[aric_hfchfaps$HFAA29C1==2&!is.na(aric_hfchfaps$HFAA29C1)]/10

aric_hfchfaps$ivs_cm[aric_hfchfaps$HFAA29C1==3&!is.na(aric_hfchfaps$HFAA29C1)] <-
  aric_hfchfaps$HFAA29C[aric_hfchfaps$HFAA29C1==3&!is.na(aric_hfchfaps$HFAA29C1)]/100


aric_hfchfaps$HFAA29C3[aric_hfchfaps$ID_S0 %in%  c("S111312",
                                                   "S111340",
                                                   "S113765",
                                                   "S114430",
                                                   "S114649",
                                                   "S115152",
                                                   "S116317",
                                                   "S116365",
                                                   "S116866",
                                                   "S117549",
                                                   "S117788",
                                                   "S118803",
                                                   "S119432",
                                                   "S120190",
                                                   "S120281",
                                                   "S121711",
                                                   "S122791",
                                                   "S124361",
                                                   "S124890",
                                                   "S125222",
                                                   "S125919",
                                                   "S126733",
                                                   "S127584",
                                                   "S128895",
                                                   "S129313",
                                                   "S130485",
                                                   "S132505",
                                                   "S133402",
                                                   "S134453",
                                                   "S134634",
                                                   "S135940",
                                                   "S136416",
                                                   "S136616",
                                                   "S136850",
                                                   "S138443",
                                                   "S138586",
                                                   "S140721",
                                                   "S141731",
                                                   "S142015",
                                                   "S143363",
                                                   "S143410",
                                                   "S143485",
                                                   "S144028",
                                                   "S144056",
                                                   "S144323",
                                                   "S144348",
                                                   "S145732",
                                                   "S146215",
                                                   "S147470",
                                                   "S148454",
                                                   "S148785",
                                                   "S150389",
                                                   "S152207",
                                                   "S154133",
                                                   "S154852",
                                                   "S158042",
                                                   "S169510",
                                                   "S172049",
                                                   "S181658",
                                                   "S185468",
                                                   "S185548",
                                                   "S193553",
                                                   "S201989",
                                                   "S203050",
                                                   "S203483")] <- 1


aric_hfchfaps$HFAA29C3[aric_hfchfaps$ID_S0 %in% c("S112486",
                                                  "S114916",
                                                  "S115209",
                                                  "S115893",
                                                  "S128703",
                                                  "S131451",
                                                  "S217931",
                                                  "S208111")] <- 2


aric_hfchfaps$HFAA29C3[aric_hfchfaps$ID_S0 %in% c("S127574",
                                                  "S136289",
                                                  "S155314",
                                                  "S212030")] <- 3


aric_hfchfaps$HFAA29C3[aric_hfchfaps$ID_S0 %in% c("S169510",
                                                  "S172049",
                                                  "S181658",
                                                  "S154133")] <- 4


aric_hfchfaps$lvpwd_cm[aric_hfchfaps$HFAA29C3==1&!is.na(aric_hfchfaps$HFAA29C3)] <-
  aric_hfchfaps$HFAA29C2[aric_hfchfaps$HFAA29C3==1&!is.na(aric_hfchfaps$HFAA29C3)]

aric_hfchfaps$lvpwd_cm[aric_hfchfaps$HFAA29C3==2&!is.na(aric_hfchfaps$HFAA29C3)] <-
  aric_hfchfaps$HFAA29C2[aric_hfchfaps$HFAA29C3==2&!is.na(aric_hfchfaps$HFAA29C3)]/10

aric_hfchfaps$lvpwd_cm[aric_hfchfaps$HFAA29C3==3&!is.na(aric_hfchfaps$HFAA29C3)] <-
  aric_hfchfaps$HFAA29C2[aric_hfchfaps$HFAA29C3==3&!is.na(aric_hfchfaps$HFAA29C3)]/100

aric_hfchfaps$lvpwd_cm[aric_hfchfaps$HFAA29C3==4&!is.na(aric_hfchfaps$HFAA29C3)] <-
  aric_hfchfaps$HFAA29C2[aric_hfchfaps$HFAA29C3==4&!is.na(aric_hfchfaps$HFAA29C3)] * 10


aric_hf_cohort <- merge(aric_hf_cohort, 
                        aric_hfchfaps[,c("ID_S0",
                                         "dispo",
                                         "HFAA1A",
                                         "HFAA1B",
                                         "HFAA1C",
                                         "HFAA1D",
                                         "HFAA1E",
                                         "HFAA6A",
                                         "HFAA6B",
                                         "HFAA6JNEW",
                                         "HFAA7A",
                                         "HFAA7B",
                                         "HFAA7C",
                                         "HFAA8A",
                                         "HFAA8A1",
                                         "HFAA11B1",
                                         "HFAA11B3",
                                         "HFAA11E4",
                                         "HFAA11E5",
                                         "HFAA11G",
                                         "HFAA11H",
                                         "HFAA16A",
                                         "HFAA16B",
                                         "HFAA16D",
                                         "HFAA16E",
                                         "HFAA17A",
                                         "HFAA18A",
                                         "adm_wt_kg",
                                         "dc_wt_kg",
                                         "wt_chg",
                                         "wt_chg_pct",
                                         "HFAA29B",
                                         "ivs_cm",
                                         "lvpwd_cm",
                                         "HFAA29D1",
                                         "HFAA29D2",
                                         "HFAA29D3",
                                         "HFAA29D4",
                                         "HFAA29D5",
                                         "HFAA29D6",
                                         "HFAA29D7",
                                         "HFAA29D8",
                                         "HFAA29D9",
                                         "HFAA29D9A",
                                         "HFAA29D10",
                                         "HFAA29D12",
                                         "HFAA29D13",
                                         "HFAA29D14",
                                         "HFAA32",
                                         "HFAA32B1",
                                         "HFAA32B2A",
                                         "HFAA32B2B",
                                         "HFAA32B2C",
                                         "HFAA32B2D",
                                         "HFAA32B2E",
                                         "HFAA32B3",
                                         "HFAA32B3A",
                                         "HFAA37A",
                                         "HFAA39A",
                                         "HFAA39B",
                                         "HFAA40A",
                                         "HFAA40B",
                                         "HFAA41A",
                                         "HFAA41B",
                                         "HFAA42A",
                                         "HFAA42B",
                                         "HFAA43A",
                                         "HFAA43B",
                                         "HFAA44A",
                                         "HFAA44B",
                                         "HFAA45A",
                                         "HFAA45B",
                                         "HFAA59",
                                         "HFAA59A",
                                         "HFAA60",
                                         "HFAA60A",
                                         "HFAA61A",
                                         "HFAA61A1",
                                         "HFAA61B",
                                         "HFAA61B1",
                                         "HFAA62",
                                         "HFAA62A",
                                         "HFAA64A",
                                         "HFAA64A1",
                                         "HFAA64B",
                                         "HFAA64B1",
                                         "HFAA65",
                                         "HFAA65A",
                                         "HFAA66",
                                         "HFAA66A",
                                         "HFAA71",
                                         "HFAA71A",
                                         "HFAA72",
                                         "HFAA72A")], 
                        by="ID_S0", 
                        all.x=T)


aric_hf_cohort$adm_wt_kg[aric_hf_cohort$wt_chg < -20|aric_hf_cohort$wt_chg > 20] <- NA
aric_hf_cohort$dc_wt_kg[aric_hf_cohort$wt_chg < -20|aric_hf_cohort$wt_chg > 20] <- NA
aric_hf_cohort$wt_chg[aric_hf_cohort$wt_chg < -20|aric_hf_cohort$wt_chg > 20] <- NA

aric_hf_cohort$HFAA29D2[aric_hf_cohort$HFAA29D2=="N"] <- NA
aric_hf_cohort$HFAA29D2[aric_hf_cohort$HFAA29D2==""] <- NA

aric_hf_cohort$HFAA29D3[aric_hf_cohort$HFAA29D3=="N"] <- NA
aric_hf_cohort$HFAA29D3[aric_hf_cohort$HFAA29D3==""] <- NA

aric_hf_cohort$ef_type <- aric_hf_cohort$LVEF_CUR_LOW
aric_hf_cohort$ef_type[is.na(aric_hf_cohort$ef_type)] <-
  aric_hf_cohort$LVEF_PRE_LOW[is.na(aric_hf_cohort$ef_type)]

aric_hf_cohort$ef_val <- aric_hf_cohort$LVEF_CUR
aric_hf_cohort$ef_val[is.na(aric_hf_cohort$ef_val)] <-
  aric_hf_cohort$LVEF_PRE[is.na(aric_hf_cohort$ef_val)]


aric_hf_cohort_first <- 
  as.data.table(sqldf('select * 
        from aric_hf_cohort 
        group by id_c 
        having min(HFDAYS)'))


aric_outcomes <- merge(aric_outcomes,
                       aric_hf_cohort[,c("ID_C",
                                         "HFDAYS",
                                         "ef_type",
                                         "ef_val",
                                         "HFAA29D2",
                                         "HFAA29B")],
                       by.x=c("ID_C",
                              "hfhosp_dt"),
                       by.y=c("ID_C",
                              "HFDAYS"),
                       all.x=T,
                       all.y=F)

aric_outcomes <- as.data.table(sqldf("select distinct * from aric_outcomes"))

aric_outcomes$hfhosp_ef_cat[aric_outcomes$HFAA29D2=='E'] <- 'Normal'
aric_outcomes$hfhosp_ef_cat[aric_outcomes$HFAA29D2=='M'] <- 'Mildly reduced'
aric_outcomes$hfhosp_ef_cat[aric_outcomes$HFAA29D2=='D'] <- 'Moderately reduced'
aric_outcomes$hfhosp_ef_cat[aric_outcomes$HFAA29D2=='S'] <- 'Severely reduced'
aric_outcomes$hfhosp_ef_val <- aric_outcomes$HFAA29B


aric_hfccelps <- fread('~/Dropbox/BioLINCC files/ARIC/Main_study/cohort_HF/csv/hfccelps19.csv')

names(aric_outcomes)[1] <- "patientid"
aric_outcomes$study <- "ARIC"

outcomes_to_fill <- outcomes_list[!outcomes_list %in% names(aric_outcomes)]
outcomes_to_fill <- outcomes_to_fill[!is.na(outcomes_to_fill)]
aric_outcomes[,outcomes_to_fill] <- NA


if (bq_table_exists("harmonization-286013.cohorts.aric_outcomes")) {
  bq_table_delete("harmonization-286013.cohorts.aric_outcomes")
  message("Table deleted.")
} 

bq_table_upload(x="harmonization-286013.cohorts.aric_outcomes",
                aric_outcomes,quiet=T,
                create_disposition="CREATE_IF_NEEDED",
                write_disposition="WRITE_TRUNCATE")


write_parquet(aric_outcomes,
              "~/Dropbox/ADAPT-HF/Master HDCP files/Cohort data/aric_outcomes.parquet")

#### ARIC community ####

aric_hf_comm <- fread('~/Dropbox/BioLINCC files/aric/Main_Study/comm_HF/csv/hfsoccps14.csv',na.strings=c("NA","","NULL"))
aric_hf_comm_first <- as.data.table(sqldf('select * from aric_hf_comm group by id_c having min(HFDAYS)'))
aric_hf_comm$visit_yr <- floor(aric_hf_comm$HFDAYS/365)


aric_sdthps <- fread('~/Dropbox/BioLINCC files/ARIC/Main_Study/comm_CHD/csv/sdthps14.csv',na.strings=c("NA","","NULL"))
aric_hfschips <- fread('~/Dropbox/BioLINCC files/ARIC/Main_Study/comm_HF/csv/hfschips14.csv',na.strings=c("NA","","NULL"))

aric_sdthps$Type[
  substr(aric_sdthps$DTH2Z18,1,1) %in% c("A","B","C","D")] <- "CV" 

aric_sdthps$Type[
  substr(aric_sdthps$DTH2Z18,1,1) %in% c("E","F","G","K","L")] <- "Non-CV" 


aric_hfshfaps <- fread('~/Dropbox/BioLINCC files/ARIC/Main_Study/comm_HF/csv/hfshfaps14.csv',na.strings=c("NA","","NULL"))

aric_hfshfaps$dispo[aric_hfshfaps$HFAA0D=="D"] <- "Deceased"
aric_hfshfaps$dispo[aric_hfshfaps$HFAA0D1=="Y"] <- "Home"
aric_hfshfaps$dispo[aric_hfshfaps$HFAA0D2=="Y"] <- "Home"
aric_hfshfaps$dispo[aric_hfshfaps$HFAA0D3=="Y"] <- "Short term care"
aric_hfshfaps$dispo[aric_hfshfaps$HFAA0D4=="Y"] <- "Home"
aric_hfshfaps$dispo[aric_hfshfaps$HFAA0D5=="Y"] <- "Long term care"
aric_hfshfaps$dispo[aric_hfshfaps$HFAA0D6=="Y"] <- "Hospice"
aric_hfshfaps$dispo[aric_hfshfaps$HFAA0D7=="Y"] <- "Left AMA"


aric_hfshfaps$adm_wt_kg[aric_hfshfaps$HFAA20A1=="K"&!is.na(aric_hfshfaps$HFAA20A1)] <- 
  aric_hfshfaps$HFAA20A[aric_hfshfaps$HFAA20A1=="K"&!is.na(aric_hfshfaps$HFAA20A1)] 


aric_hfshfaps$adm_wt_kg[aric_hfshfaps$HFAA20A1=="L"&!is.na(aric_hfshfaps$HFAA20A1)] <- 
  aric_hfshfaps$HFAA20A[aric_hfshfaps$HFAA20A1=="L"&!is.na(aric_hfshfaps$HFAA20A1)] / 2.2 


aric_hfshfaps$dc_wt_kg[aric_hfshfaps$HFAA20B1=="K"&!is.na(aric_hfshfaps$HFAA20B1)] <- 
  aric_hfshfaps$HFAA20B[aric_hfshfaps$HFAA20B1=="K"&!is.na(aric_hfshfaps$HFAA20B1)] 

aric_hfshfaps$dc_wt_kg[aric_hfshfaps$HFAA20B1=="L"&!is.na(aric_hfshfaps$HFAA20B1)] <- 
  aric_hfshfaps$HFAA20B[aric_hfshfaps$HFAA20B1=="L"&!is.na(aric_hfshfaps$HFAA20B1)] / 2.2 

aric_hfshfaps$wt_chg <- 
  aric_hfshfaps$dc_wt_kg-aric_hfshfaps$adm_wt_kg

aric_hfshfaps$wt_chg_pct <- 
  (aric_hfshfaps$dc_wt_kg-aric_hfshfaps$adm_wt_kg)/aric_hfshfaps$adm_wt_kg

aric_hfshfaps$lvh_aric_grade[!aric_hfshfaps$HFAA29D1 %in% c("N","P")] <-
  aric_hfshfaps$HFAA29D1[!aric_hfshfaps$HFAA29D1 %in% c("N","P")]

aric_hfshfaps$lvh_aric_present[aric_hfshfaps$HFAA29D1 %in% c("E")] <- "No"
aric_hfshfaps$lvh_aric_present[aric_hfshfaps$HFAA29D1 %in% c("D","E","M","P","S")] <- "Yes"


## Fixing unit errors
## Units codes: 1=cm, 2=mm

aric_hfshfaps$HFAA29C1[
  aric_hfshfaps$ID_S0 %in% 
    subset(aric_hfshfaps, 
           HFAA29C<5&
             HFAA29C1==2)$ID_S0] <- 1

aric_hfshfaps$HFAA29C1[
  aric_hfshfaps$ID_S0 %in% 
    subset(aric_hfshfaps, 
           HFAA29C<5&
             HFAA29C1==1)$ID_S0] <- 2

aric_hfshfaps$ivs_cm[aric_hfshfaps$HFAA29C1==1&!is.na(aric_hfshfaps$HFAA29C1)] <-
  aric_hfshfaps$HFAA29C[aric_hfshfaps$HFAA29C1==1&!is.na(aric_hfshfaps$HFAA29C1)]

aric_hfshfaps$ivs_cm[aric_hfshfaps$HFAA29C1==2&!is.na(aric_hfshfaps$HFAA29C1)] <-
  aric_hfshfaps$HFAA29C[aric_hfshfaps$HFAA29C1==2&!is.na(aric_hfshfaps$HFAA29C1)]/10

aric_hfshfaps$ivs_cm[aric_hfshfaps$HFAA29C1==3&!is.na(aric_hfshfaps$HFAA29C1)] <-
  aric_hfshfaps$HFAA29C[aric_hfshfaps$HFAA29C1==3&!is.na(aric_hfshfaps$HFAA29C1)]/100


aric_hfshfaps$HFAA29C3[
  aric_hfshfaps$ID_S0 %in% 
    subset(aric_hfshfaps, HFAA29C2 <5&HFAA29C1==2)[,c("ID_S0")]] <- 1


aric_hfshfaps$HFAA29C3[
  aric_hfshfaps$ID_S0 %in% 
    subset(aric_hfshfaps, HFAA29C2 >7&HFAA29C1==1)[,c("ID_S0")]] <- 2


aric_hfshfaps$lvpwd_cm[aric_hfshfaps$HFAA29C3==1&!is.na(aric_hfshfaps$HFAA29C3)] <-
  aric_hfshfaps$HFAA29C2[aric_hfshfaps$HFAA29C3==1&!is.na(aric_hfshfaps$HFAA29C3)]

aric_hfshfaps$lvpwd_cm[aric_hfshfaps$HFAA29C3==2&!is.na(aric_hfshfaps$HFAA29C3)] <-
  aric_hfshfaps$HFAA29C2[aric_hfshfaps$HFAA29C3==2&!is.na(aric_hfshfaps$HFAA29C3)]/10


aric_hf_comm <-
  aric_hfshfaps[,c("ID_S0",
                   "dispo",
                   "HFAA1A",
                   "HFAA1B",
                   "HFAA1C",
                   "HFAA1D",
                   "HFAA1E",
                   "HFAA6A",
                   "HFAA6B",
                   "HFAA6JNEW",
                   "HFAA7A",
                   "HFAA7B",
                   "HFAA7C",
                   "HFAA8A",
                   "HFAA8A1",
                   "HFAA11B1",
                   "HFAA11B3",
                   "HFAA11E4",
                   "HFAA11E5",
                   "HFAA11G",
                   "HFAA11H",
                   "HFAA16A",
                   "HFAA16B",
                   "HFAA16D",
                   "HFAA16E",
                   "HFAA17A",
                   "HFAA18A",
                   "adm_wt_kg",
                   "dc_wt_kg",
                   "wt_chg",
                   "wt_chg_pct",
                   "HFAA29B",
                   "ivs_cm",
                   "lvpwd_cm",
                   "HFAA29D1",
                   "HFAA29D2",
                   "HFAA29D3",
                   "HFAA29D4",
                   "HFAA29D5",
                   "HFAA29D6",
                   "HFAA29D7",
                   "HFAA29D8",
                   "HFAA29D9",
                   "HFAA29D9A",
                   "HFAA29D10",
                   "HFAA29D12",
                   "HFAA29D13",
                   "HFAA29D14",
                   "HFAA32",
                   "HFAA32B1",
                   "HFAA32B2A",
                   "HFAA32B2B",
                   "HFAA32B2C",
                   "HFAA32B2D",
                   "HFAA32B2E",
                   "HFAA32B3",
                   "HFAA32B3A",
                   "HFAA37A",
                   "HFAA39A",
                   "HFAA39B",
                   "HFAA40A",
                   "HFAA40B",
                   "HFAA41A",
                   "HFAA41B",
                   "HFAA42A",
                   "HFAA42B",
                   "HFAA43A",
                   "HFAA43B",
                   "HFAA44A",
                   "HFAA44B",
                   "HFAA45A",
                   "HFAA45B",
                   "HFAA59",
                   "HFAA59A",
                   "HFAA60",
                   "HFAA60A",
                   "HFAA61A",
                   "HFAA61A1",
                   "HFAA61B",
                   "HFAA61B1",
                   "HFAA62",
                   "HFAA62A",
                   "HFAA64A",
                   "HFAA64A1",
                   "HFAA64B",
                   "HFAA64B1",
                   "HFAA65",
                   "HFAA65A",
                   "HFAA66",
                   "HFAA66A",
                   "HFAA71",
                   "HFAA71A",
                   "HFAA72",
                   "HFAA72A")]

aric_hf_comm$adm_wt_kg[aric_hf_comm$wt_chg < -20|aric_hf_comm$wt_chg > 20] <- NA
aric_hf_comm$dc_wt_kg[aric_hf_comm$wt_chg < -20|aric_hf_comm$wt_chg > 20] <- NA
aric_hf_comm$wt_chg[aric_hf_comm$wt_chg < -20|aric_hf_comm$wt_chg > 20] <- NA

aric_hf_comm$HFAA29D2[aric_hf_comm$HFAA29D2=="N"] <- NA
aric_hf_comm$HFAA29D2[aric_hf_comm$HFAA29D2==""] <- NA

aric_hf_comm$HFAA29D3[aric_hf_comm$HFAA29D3=="N"] <- NA
aric_hf_comm$HFAA29D3[aric_hf_comm$HFAA29D3==""] <- NA


aric_hfccelps <- 
  read.csv('~/Dropbox/BioLINCC files/ARIC/Main_study/cohort_HF/csv/hfccelps19.csv',
           na.strings=c("",NA))


rm(list=ls(pattern="\\baric_hf.*$"))


#### CHS ####

chs_events <- read.csv("~/Dropbox/BioLINCC files/CHS/Main_Study/Events/events.csv")
chs_drhosp11 <- read.csv("~/Dropbox/BioLINCC files/CHS/Main_Study/ICD9/drhosp16.csv")

chs_censor <- sqldf('select distinct newid,
                    DEATH as dth_status,
                    CENSTIME dth_dt from chs_events')

chs_fatalevents <- sqldf("select distinct 
      newid, 
      censtime,
      cause80,
      mechan80
      from chs_events
      where fatal == 1")

chs_fatalevents$cvdth_status[chs_fatalevents$cause80 %in% c(1,2,3,4)] <- 1
chs_fatalevents$noncvdth_status[chs_fatalevents$cause80 %in% c(5)] <- 1
chs_fatalevents$hfdth_status[chs_fatalevents$cause80 %in% c(3)] <- 1

chs_fatalevents$cvdth_status[is.na(chs_fatalevents$cvdth_status)] <- 0
chs_fatalevents$noncvdth_status[is.na(chs_fatalevents$noncvdth_status)] <- 0
chs_fatalevents$hfdth_status[is.na(chs_fatalevents$hfdth_status)] <- 0

chs_fatalevents$cvdth_dt <- chs_fatalevents$censtime
chs_fatalevents$noncvdth_dt <- chs_fatalevents$censtime
chs_fatalevents$hfdth_dt <- chs_fatalevents$censtime


chs_incidenthf <- sqldf("select distinct newid, 
                        1 as hfhosp_status,
                        TTOEVENT as hfhosp_dt
                        from chs_events
                        where EVTYPE = 4
                        group by newid 
                        having min(hfhosp_dt)")

chs_incidentcad <- sqldf("select distinct newid, 
                        1 as cadhosp_status,
                        TTOEVENT as cadhosp_dt
                        from chs_events
                        where EVTYPE in (1,2,7,8,10,11)
                        group by newid 
                        having min(cadhosp_dt)")


chs_incidentcva <- sqldf("select distinct newid, 
                        1 as cvahosp_status,
                        TTOEVENT as cvahosp_dt
                        from chs_events
                        where EVTYPE in (3)
                        group by newid 
                        having min(cvahosp_dt)")

chs_outcomes <- merge(chs_censor,chs_fatalevents,by="newid",all.x=T)
chs_outcomes <- merge(chs_outcomes,chs_incidenthf,by="newid",all.x=T)
chs_outcomes <- merge(chs_outcomes,chs_incidentcad,by="newid",all.x=T)
chs_outcomes <- merge(chs_outcomes,chs_incidentcva,by="newid",all.x=T)

chs_outcomes$time_hfhosp_to_eos <- chs_outcomes$censtime-chs_outcomes$hfhosp_dt
chs_outcomes$hfhosp_status[is.na(chs_outcomes$hfhosp_status)] <- 0
chs_outcomes$hfhosp_dt[chs_outcomes$hfhosp_status==0] <- chs_outcomes$censtime[chs_outcomes$hfhosp_status==0]

chs_outcomes$time_cad_to_eos <- chs_outcomes$censtime - chs_outcomes$cadhosp_dt
chs_outcomes$cadhosp_status[is.na(chs_outcomes$cadhosp_status)] <- 0
chs_outcomes$cadhosp_dt[chs_outcomes$cadhosp_status==0] <- chs_outcomes$censtime[chs_outcomes$cadhosp_status==0]


chs_outcomes$study <- "CHS"

names(chs_outcomes)[1] <- "patientid"
outcomes_to_fill <- outcomes_list[!outcomes_list %in% names(chs_outcomes)]
outcomes_to_fill <- outcomes_to_fill[!is.na(outcomes_to_fill)]
chs_outcomes[,outcomes_to_fill] <- NA

chs_outcomes$patientid <-
  as.character(chs_outcomes$patientid)

bq_table_upload(x="harmonization-286013.cohorts.chs_outcomes",
                chs_outcomes,quiet=T,
                create_disposition="CREATE_IF_NEEDED",
                write_disposition="WRITE_TRUNCATE")

write_parquet(chs_outcomes,
              "~/Dropbox/ADAPT-HF/Master HDCP files/Cohort data/chs_outcomes.parquet")


#### MESA ####


mesa_hfevents <- read.csv("~/Dropbox/BioLINCC files/MESA/Primary/Events/CVD/Data/mesaevefthru2015_drepos_20200330.csv")
mesa_afevents <- read.csv("~/Dropbox/BioLINCC files/MESA/Primary/Events/CVD/Data/mesaevaffu10_drepos_2015416.csv")
mesa_cvevents <- read.csv("~/Dropbox/BioLINCC files/MESA/Primary/Events/CVD/Data/mesaevthr2020_drepos_20241120.csv")
mesa_noncvevents <- read.csv("~/Dropbox/BioLINCC files/MESA/Primary/Events/NonCVD/Data/mesaevnoncvddres06192012.csv")

mesa_cvevents$dth_status <- mesa_cvevents$dth
mesa_cvevents$dth_dt <- mesa_cvevents$dthtt

mesa_cvevents$cvdth_status[mesa_cvevents$dthtype %in% c(1,2,3,4)] <- 1
mesa_cvevents$cvdth_status[is.na(mesa_cvevents$cvdth_status)] <- 0
mesa_cvevents$cvdth_dt <- mesa_cvevents$dthtt

mesa_cvevents$noncvdth_status[mesa_cvevents$dthtype %in% c(5, 
                                                           9
)] <- 1
mesa_cvevents$noncvdth_status[is.na(mesa_cvevents$noncvdth_status)] <- 0
mesa_cvevents$noncvdth_dt <- mesa_cvevents$dthtt

mesa_cvevents$cvhosp_status <- mesa_cvevents$cvda
mesa_cvevents$cvhosp_dt <- mesa_cvevents$cvdatt

mesa_cvevents$hfhosp_status <- mesa_cvevents$chf
mesa_cvevents$hfhosp_dt <- mesa_cvevents$chftt

mesa_cvevents$cadhosp_status <- mesa_cvevents$chda
mesa_cvevents$cadhosp_dt <- mesa_cvevents$chdatt

mesa_noncvevents$noncvhosp_dt <- 
  apply(mesa_noncvevents[,c(2,
                            4,
                            6,
                            8,
                            10,
                            12,
                            14,
                            16,
                            18,
                            20,
                            22,
                            24,
                            26,
                            28,
                            30)],
        MARGIN=1,
        FUN=min,
        na.rm=T)


mesa_noncvevents$noncvhosp_status[mesa_noncvevents$noncvhosp_dt==mesa_noncvevents$fuptt] <- 0
mesa_noncvevents$noncvhosp_status[!mesa_noncvevents$noncvhosp_dt==mesa_noncvevents$fuptt] <- 1


mesa_outcomes <- merge(
  mesa_cvevents[,c("MESAID",
                   "dth_status","dth_dt",
                   "cvdth_status","cvdth_dt",
                   "noncvdth_status","noncvdth_dt",
                   "cvhosp_status","cvhosp_dt",
                   "cadhosp_status","cadhosp_dt",
                   "hfhosp_status","hfhosp_dt")],
  mesa_noncvevents[,c("MESAID",
                      "noncvhosp_status",
                      "noncvhosp_dt")],
  by="MESAID",
  all.x=T
)

mesa_outcomes <- merge(mesa_outcomes,
                       mesa_hfevents[,c("MESAID","TTCHF","EFMEAS","EFCLASS")],
                       by.x=c("MESAID","hfhosp_dt"),
                       by.y=c("MESAID","TTCHF"),
                       all.x=T)

mesa_outcomes$hfhosp_ef_val <- mesa_outcomes$EFMEAS

mesa_outcomes$hfhosp_ef_cat[mesa_outcomes$hfhosp_ef_val <= 35] <- "Severely reduced"
mesa_outcomes$hfhosp_ef_cat[mesa_outcomes$hfhosp_ef_val > 35&
                              mesa_outcomes$hfhosp_ef_val <= 45] <- "Moderately reduced"
mesa_outcomes$hfhosp_ef_cat[mesa_outcomes$hfhosp_ef_val > 45&
                              mesa_outcomes$hfhosp_ef_val <= 55] <- "Mildly reduced"

mesa_outcomes$hfhosp_ef_cat[mesa_outcomes$hfhosp_ef_val > 55] <- "Normal"

mesa_outcomes$hosp_status <- 0
mesa_outcomes$hosp_dt <- mesa_outcomes$dth_dt
mesa_outcomes$hosp_status[mesa_outcomes$cvhosp_status==1|
                            mesa_outcomes$noncvhosp_status==1] <- 1

mesa_outcomes$hosp_dt[mesa_outcomes$hosp_status==1] <-
  min(mesa_outcomes$cvhosp_dt[mesa_outcomes$hosp_status==1],
      mesa_outcomes$noncvhosp_dt[mesa_outcomes$hosp_status==1])

mesa_outcomes$study <- "MESA"

names(mesa_outcomes)[1] <- "patientid"
mesa_outcomes$patientid <- as.character(mesa_outcomes$patientid)

outcomes_to_fill <- outcomes_list[!outcomes_list %in% names(mesa_outcomes)]
outcomes_to_fill <- outcomes_to_fill[!is.na(outcomes_to_fill)]
mesa_outcomes[,outcomes_to_fill] <- NA

write_parquet(mesa_outcomes,
              '~/Dropbox/ADAPT-HF/Master HDCP files/Cohort data/mesa_outcomes.parquet')

bq_table_upload(x="harmonization-286013.cohorts.mesa_outcomes",
                mesa_outcomes,quiet=T,
                create_disposition="CREATE_IF_NEEDED",
                write_disposition="WRITE_TRUNCATE")

#### JHS ####

jhs_allevtchd <- read.csv("~/Dropbox/BioLINCC files/JHS/Data/Events/csv/allevtchd.csv")
jhs_allevthf <- read.csv("~/Dropbox/BioLINCC files/JHS/Data/Events/csv/allevthf.csv")
jhs_allevtstroke <- read.csv("~/Dropbox/BioLINCC files/JHS/Data/Events/csv/allevtstroke.csv")
jhs_incevtchd <- read.csv("~/Dropbox/BioLINCC files/JHS/Data/Events/csv/incevtchd.csv")
jhs_incevthfder <- read.csv("~/Dropbox/BioLINCC files/JHS/Data/Events/csv/incevthfder.csv")
jhs_incevtstroke <- read.csv("~/Dropbox/BioLINCC files/JHS/Data/Events/csv/incevtstroke.csv")

jhs_allevtchd <- merge(jhs_allevtchd,
                       jhs_incevtchd[,c("newid","V1date")],
                       by="newid")

jhs_allevtchd$days_to_event <- as.numeric(as.Date(jhs_allevtchd$eventDate, format="%m/%d/%Y") - 
                                            as.Date(jhs_allevtchd$V1date, 
                                                    format="%m/%d/%Y"))

jhs_incevthfder$HF[jhs_incevthfder$examdate==""] <- 0
jhs_incevthfder$days[jhs_incevthfder$examdate==""] <- 0
jhs_incevthfder$examdate[jhs_incevthfder$examdate==""] <- 
  jhs_incevthfder$V1date[jhs_incevthfder$examdate==""]

jhs_allevthf <- merge(jhs_allevthf,
                      jhs_incevthfder[,c("newid","examdate")],
                      by="newid")

jhs_allevthf$days_to_event <- as.numeric(as.Date(jhs_allevthf$eventDate, format="%m/%d/%Y") - 
                                           as.Date(jhs_allevthf$examdate, format="%m/%d/%Y"))

jhs_allevtstroke <- merge(jhs_allevtstroke,
                          jhs_incevthfder[,c("newid","V1date")],
                          by="newid")

jhs_allevtstroke$days_to_event <- as.numeric(as.Date(jhs_allevtstroke$eventdate, format="%m/%d/%Y") - 
                                               as.Date(jhs_allevtstroke$V1date, format="%m/%d/%Y"))

jhs_outcomes_hf <- sqldf("select newid,
                      days as hfhosp_dt,
                      HF as hfhosp_status
                      from jhs_incevthfder")

jhs_outcomes_cad <- sqldf("select newid,
                      days as cadhosp_dt,
                      CHD as cadhosp_status
                      from jhs_incevtchd")

jhs_outcomes <- merge(jhs_outcomes_hf,
                      jhs_outcomes_cad,
                      by="newid",
                      all.x=T)


jhs_outcomes$study <- "JHS"


names(jhs_outcomes)[1] <- "patientid"

jhs_outcomes$patientid <-
  as.character(jhs_outcomes$patientid)

outcomes_to_fill <- outcomes_list[!outcomes_list %in% names(jhs_outcomes)]
outcomes_to_fill <- outcomes_to_fill[!is.na(outcomes_to_fill)]
jhs_outcomes[,outcomes_to_fill] <- NA

outcomes_list_2 <- c("patientid","study",outcomes_list)


write_parquet(jhs_outcomes,
              '~/Dropbox/ADAPT-HF/Master HDCP files/Cohort data/jhs_outcomes.parquet')

bq_table_upload(x="harmonization-286013.cohorts.jhs_outcomes",
                jhs_outcomes,quiet=T,
                create_disposition="CREATE_IF_NEEDED",
                write_disposition="WRITE_TRUNCATE")

#### Assemble all outcomes ####

all_outcomes <-
  rbind(
    as.data.frame(aric_outcomes[,..outcomes_list_2]),
    as.data.frame(chs_outcomes[,outcomes_list_2]),
    as.data.frame(fhs_outcomes[,..outcomes_list_2]),
    as.data.frame(jhs_outcomes[,outcomes_list_2]),
    as.data.frame(mesa_outcomes[,outcomes_list_2]))


all_outcomes$patientid <-
  as.character(all_outcomes$patientid)

bq_table_upload(x="harmonization-286013.cohorts.outcomes",
                all_outcomes,quiet=T,
                create_disposition="CREATE_IF_NEEDED",
                write_disposition="WRITE_TRUNCATE")

write_parquet(all_outcomes,
              "~/Dropbox/ADAPT-HF/Master HDCP files/Cohort data/all_outcomes.parquet")


########### ---- DATES ---- ##########

# .%%%%%%%%.....%%%....%%%%%%%%.%%%%%%%%..%%%%%%.
# .%%.....%%...%%.%%......%%....%%.......%%....%%
# .%%.....%%..%%...%%.....%%....%%.......%%......
# .%%.....%%.%%.....%%....%%....%%%%%%....%%%%%%.
# .%%.....%%.%%%%%%%%%....%%....%%.............%%
# .%%.....%%.%%.....%%....%%....%%.......%%....%%
# .%%%%%%%%..%%.....%%....%%....%%%%%%%%..%%%%%%.


#### FHS ####

fhs1_dates <- read.csv('~/Dropbox/BioLINCC files/Framingham cohort/datasets/CSV/vr_dates_2014_a_0912d.csv',na.strings=c("NA","","NULL"))
fhs1_wkthru <- read.csv('~/Dropbox/BioLINCC files/Framingham cohort/datasets/CSV/vr_wkthru_ex32_0_0997d.csv',na.strings=c("NA","","NULL"))

fhs1_melt_sex <- sqldf("select PID,
                       IDTYPE,
                       1 as visit,
                       'sex' as variable,
                       SEX as value,
                       'fhs1_dates' as form
                       from fhs1_dates")

names(fhs1_dates) <- toupper(names(fhs1_dates))
names(fhs1_wkthru) <- toupper(names(fhs1_wkthru))

for (v in 1:32) {
  fhs1_wkthru[,paste("BMIC",v,sep="")] <- 
    round(calc_bmi(dat=fhs1_wkthru,
                   weight=paste("WGT",v,sep=""),
                   height="HGT1",
                   metric=F),2)
}

for (whr in c(19:23)) {
  fhs1_wkthru[,paste("WHR",whr,sep="")] <-
    round(fhs1_wkthru[,paste("WAIST",whr,sep="")]/fhs1_wkthru[,paste("HIP",whr,sep="")],3)
}

fhs2_omni1_wkthru <- read.csv('~/Dropbox/BioLINCC files/Framingham offspring/datasets/CSV/vr_wkthru_ex04_7_0917d.csv',na.strings=c("NA","","NULL"))
fhs2_offspring_wkthru <- read.csv('~/Dropbox/BioLINCC files/Framingham offspring/datasets/CSV/vr_wkthru_ex09_1_1001d.csv',na.strings=c("NA","","NULL"))
fhs2_dates <- read.csv('~/Dropbox/BioLINCC files/Framingham offspring/datasets/CSV/vr_dates_2019_a_1175d.csv',na.strings=c("NA","","NULL"))

fhs2_race_1 <- read.csv('~/Dropbox/BioLINCC files/Framingham offspring/datasets/CSV/race_1.csv',na.strings=c("NA","","NULL")) # Physical exam questionnaire, exam 5
names(fhs2_race_1) <- toupper(names(fhs2_race_1))
fhs2_race_1$RACE[fhs2_race_1$ETHNICITY=="Hisp"] <- "Hisp" 
fhs2_race_1$variable <- "race"
fhs2_race_1$value <- fhs2_race_1$RACE
fhs2_race_1$form <- "fhs2_race_1"
fhs2_race_1$visit <- 1

fhs2_melt_sex <- sqldf("select PID,
                       IDTYPE as IDTYPE,
                       1 as visit,
                       'sex' as variable,
                       SEX as value,
                       'fhs2_dates' as form
                       from fhs2_dates")


names(fhs2_dates) <- toupper(names(fhs2_dates))
names(fhs2_omni1_wkthru) <- toupper(names(fhs2_omni1_wkthru))
names(fhs2_offspring_wkthru) <- toupper(names(fhs2_offspring_wkthru))

for (whr in c(4,5,6,7,9)) {
  fhs2_offspring_wkthru[,paste("WHR",whr,sep="")] <-
    round(fhs2_offspring_wkthru[,paste("WAIST",whr,sep="")]/fhs2_offspring_wkthru[,paste("HIP",whr,sep="")],3)
}

fhs2_offspring_wkthru$RACE <- 1

for (i in c(2,5,6,7,8,9)) {
  fhs2_offspring_wkthru[,paste("EGFR_MDRD",i,sep="")] <-
    calc_MDRD4(fhs2_offspring_wkthru,
               age=paste("AGE",i,sep=""),
               cr=paste("CREAT",i,sep=""),
               sex="SEX",
               race="RACE")
}


for (whr in c(1,2,4)) {
  fhs2_omni1_wkthru[,paste("WHR",whr,sep="")] <-
    round(fhs2_omni1_wkthru[,paste("WAIST",whr,sep="")]/fhs2_omni1_wkthru[,paste("HIP",whr,sep="")],3)
}


fhs2_omni1_wkthru <-
  merge(fhs2_omni1_wkthru,
        fhs2_race_1[,c("PID","IDTYPE","RACE")],
        by=c("PID","IDTYPE"),
        all.x=T)

for (i in c(2,3,4)) {
  fhs2_omni1_wkthru[,paste("EGFR_MDRD",i,sep="")] <-
    calc_MDRD4(fhs2_omni1_wkthru,
               age=paste("AGE",i,sep=""),
               cr=paste("CREAT",i,sep=""),
               sex="SEX",
               race="RACE",
               black="B")
}


fhs3_wkthru <- read.csv('~/Dropbox/BioLINCC files/Framingham third generation/datasets/CSV/vr_wkthru_ex03_3b_1191d.csv',na.strings=c("NA","","NULL"))
fhs3_dates <- read.csv('~/Dropbox/BioLINCC files/Framingham third generation/datasets/CSV/vr_dates_2019_a_1175d.csv',na.strings=c("NA","","NULL"))
names(fhs3_dates) <- toupper(names(fhs3_dates))
names(fhs3_wkthru) <- toupper(names(fhs3_wkthru))

fhs3_melt_sex <- sqldf("select PID,
                       IDTYPE,
                       1 as visit,
                       'sex' as variable,
                       SEX as value,
                       'fhs3_dates' as form
                       from fhs3_dates")

fhs_dates_fields <- names(fhs1_dates)
fhs2_dates[,fhs_dates_fields[!fhs_dates_fields %in% names(fhs2_dates)]] <- NA
fhs3_dates[,fhs_dates_fields[!fhs_dates_fields %in% names(fhs3_dates)]] <- NA


fhs_dates <- rbind(fhs1_dates[fhs_dates_fields],
                   fhs2_dates[fhs_dates_fields],
                   fhs3_dates[fhs_dates_fields])


fhs_dates <- merge(fhs_dates,
                   fhs_survcvd,
                   by=c("IDTYPE","PID"),
                   all.x=T)
fhs_dates$DATE1 <- 0

for (i in 2:20) {
  thisexamdate <- paste("DATE",i,sep="")
  fhs_dates[(fhs_dates$hfhosp_status==1&
               fhs_dates$hfhosp_dt > fhs_dates[,thisexamdate]&
               !is.na(fhs_dates$hfhosp_dt)&
               !is.na(fhs_dates[,thisexamdate])),'last_exam_before_hf']  <- i
}

age_cols <- names(fhs_dates)[grep("AGE", names(fhs_dates))]
date_cols <- names(fhs_dates)[grep("DATE", names(fhs_dates))]

fhs_dates_long_age <- melt(fhs_dates[,c("PID","IDTYPE","SEX",age_cols)],
                           id.vars=c("PID","IDTYPE","SEX"),
                           na.rm=T,
                           factorsAsStrings = T)

fhs_dates_long_age$variable <- as.character(fhs_dates_long_age$variable)

fhs_dates_long_age$visit <- 
  substring(fhs_dates_long_age$variable,
            str_locate(fhs_dates_long_age$variable,
                       "\\d")[1],length(fhs_dates_long_age$variable))

fhs_dates_long_age$variable <- 
  gsub("\\d","",fhs_dates_long_age$variable)

fhs_dates_long_date <- 
  melt(fhs_dates[,c("PID","IDTYPE",date_cols)],
       id.vars=c("PID","IDTYPE"),
       na.rm=T,
       factorsAsStrings = T)

fhs_dates_long_date$variable <- as.character(fhs_dates_long_date$variable)

fhs_dates_long_date <- subset(fhs_dates_long_date,!variable=="CHFDATE")

fhs_dates_long_date$visit <- 
  substring(fhs_dates_long_date$variable,
            str_locate(fhs_dates_long_date$variable,
                       "\\d")[1],length(fhs_dates_long_date$variable))

fhs_dates_long_date$variable <- gsub("\\d","",fhs_dates_long_date$variable)

names(fhs_dates_long_date)[names(fhs_dates_long_date)=="value"] <- "visitdays"
names(fhs_dates_long_age)[names(fhs_dates_long_age)=="value"] <- "age_obs"

fhs_dates_long <- merge(fhs_dates_long_age[,c("PID","IDTYPE","SEX","visit","age_obs")],
                        fhs_dates_long_date[,c("PID","IDTYPE","visit","visitdays")],
                        by=c("PID","IDTYPE","visit"))

fhs_dates_long$study <- "FHS"

names(fhs_dates_long)[2] <- "cohort"

fhs_dates_long <- merge(fhs_dates_long,
                        visit_yrs[visit_yrs$study=="FHS",
                                  c("cohort","visit","visit_yr")])

fhs_dates_long$cohort_name[fhs_dates_long$cohort==0] <- "Original"
fhs_dates_long$cohort_name[fhs_dates_long$cohort==1] <- "Offspring"
fhs_dates_long$cohort_name[fhs_dates_long$cohort==7] <- "OMNI 1"
fhs_dates_long$cohort_name[fhs_dates_long$cohort==3] <- "Gen III"
fhs_dates_long$cohort_name[fhs_dates_long$cohort==2] <- "Offspring spouse"
fhs_dates_long$cohort_name[fhs_dates_long$cohort==72] <- "OMNI 2"

fhs_dates_long$patientid <- fhs_dates_long$PID
fhs_dates_long$sex[fhs_dates_long$SEX==1] <- "Male"
fhs_dates_long$sex[fhs_dates_long$SEX==2] <- "Female"

rm(fhs_dates_long_age,
   fhs_dates_long_date)


#### ARIC ####


aric_derive1 <- read.csv('~/Dropbox/BioLINCC files/ARIC/Main_Study/V1/csv/derive13.csv',na.strings=c("NA","","NULL"))
aric_derive2 <- read.csv('~/Dropbox/BioLINCC files/ARIC/Main_Study/V2/csv/derive2_10.csv',na.strings=c("NA","","NULL"))
aric_derive3 <- read.csv('~/Dropbox/BioLINCC files/ARIC/Main_Study/V3/csv/derive37.csv',na.strings=c("NA","","NULL"))
aric_derive4 <- read.csv('~/Dropbox/BioLINCC files/ARIC/Main_Study/V4/csv/derive47.csv',na.strings=c("NA","","NULL"))
aric_derive5 <- read.csv('~/Dropbox/BioLINCC files/ARIC/Main_Study/V5/csv/derive51.csv',na.strings=c("NA","","NULL"))
aric_derive6 <- read.csv('~/Dropbox/BioLINCC files/ARIC/Main_Study/V6/csv/derive61.csv',na.strings=c("NA","","NULL"))
aric_derive7 <- read.csv('~/Dropbox/BioLINCC files/ARIC/Main_Study/V7/csv/derive71.csv',na.strings=c("NA","","NULL"))

aric_dates <- merge(aric_derive1[,c("ID_C","CENTERID","ENROLL_YR","PREVHF01","V1AGE01")],
                    aric_derive2[,c("ID_C","V2DAYS","V2AGE22")],all.x=T)
aric_dates <- merge(aric_dates,aric_derive3[,c("ID_C","V3DAYS","V3AGE31")],all.x=T)
aric_dates <- merge(aric_dates,aric_derive4[,c("ID_C","V4DAYS","V4AGE41")],all.x=T)
aric_dates <- merge(aric_dates,aric_derive5[,c("ID_C","V5DATE51_DAYS","V5AGE51")],all.x=T)
aric_dates <- merge(aric_dates,aric_derive6[,c("ID_C","V6DATE61_DAYS","V6AGE61")],all.x=T)
aric_dates <- merge(aric_dates,aric_derive7[,c("ID_C","V7DATE71_FOLLOWUPDAYS","V7AGE72")],all.x=T)

aric_dates$cohort <- aric_dates$CENTERID

aric_dates$cohort_name[aric_dates$cohort=="A"] <- "MN"
aric_dates$cohort_name[aric_dates$cohort=="B"] <- "MD"
aric_dates$cohort_name[aric_dates$cohort=="C"] <- "MI"
aric_dates$cohort_name[aric_dates$cohort=="D"] <- "NC"


aric_dates1_long <- sqldf("select ID_C,
                          1 as visit,
                          0 as visitdays,
                          V1AGE01 as age_obs
                          from aric_derive1")

aric_dates2_long <- sqldf("select ID_C,
                          2 as visit,
                          V2DAYS as visitdays,
                          V2AGE22 as age_obs
                          from aric_derive2")

aric_dates3_long <- sqldf("select ID_C,
                          3 as visit,
                          V3DAYS as visitdays,
                          V3AGE31 as age_obs
                          from aric_derive3")

aric_dates4_long <- sqldf("select ID_C,
                          4 as visit,
                          V4DAYS as visitdays,
                          V4AGE41 as age_obs
                          from aric_derive4")

aric_dates5_long <- sqldf("select ID_C,
                          5 as visit,
                          V5DATE51_DAYS as visitdays,
                          V5AGE51 as age_obs
                          from aric_derive5")

aric_dates6_long <- sqldf("select ID_C,
                          6 as visit,
                          V6DATE61_DAYS as visitdays,
                          V6AGE61 as age_obs
                          from aric_derive6")

aric_dates7_long <- sqldf("select ID_C,
                          7 as visit,
                          V7DATE71_FOLLOWUPDAYS as visitdays,
                          V7AGE72 as age_obs
                          from aric_derive7")


aric_dates_long <- rbind(aric_dates1_long,
                         aric_dates2_long,
                         aric_dates3_long,
                         aric_dates4_long,
                         aric_dates5_long,
                         aric_dates6_long,
                         aric_dates7_long)


rm(aric_dates1_long,
   aric_dates2_long,
   aric_dates3_long,
   aric_dates4_long,
   aric_dates5_long,
   aric_dates6_long,
   aric_dates7_long)


aric_dates_long <- merge(aric_dates_long,
                         aric_derive1[,c("ID_C",
                                         "CENTERID",
                                         "GENDER")],
                         by="ID_C",
                         all.x=T)

aric_dates_long <- merge(aric_dates_long,
                         subset(visit_yrs,
                                study=="ARIC"&
                                  cohort=="A"))

aric_dates_long$cohort <- aric_dates_long$CENTERID

aric_dates_long$cohort_name[aric_dates_long$cohort=="A"] <- "MN"
aric_dates_long$cohort_name[aric_dates_long$cohort=="B"] <- "MD"
aric_dates_long$cohort_name[aric_dates_long$cohort=="C"] <- "MI"
aric_dates_long$cohort_name[aric_dates_long$cohort=="D"] <- "NC"

aric_dates_long$study <- "ARIC"


#### CHS ####

#### Baseboth ###

chs_baseboth <- read.csv("~/Dropbox/BioLINCC files/CHS_2019a/BASELINE/basebothfinal.csv",na.strings=c("NA","","NULL"))
names(chs_baseboth) <- toupper(names(chs_baseboth))

chs_baseboth$agey <- 2*(chs_baseboth$AGE2-1)+64

chs_melt_baseboth <- unique(chs_baseboth[,c("IDNO","PERSTAT")])
names(chs_melt_baseboth)[1:2] <- c("patientid","COHORT")
chs_melt_baseboth$visitdays <- 0
chs_melt_baseboth$visit <- 1


#### CHS Year 3 ###

chs_yr3 <- read.csv("~/Dropbox/BioLINCC files/CHS_2019a/YEAR3/yr3final.csv",na.strings=c("NA","","NULL"))
names(chs_yr3) <- toupper(names(chs_yr3))

chs_yr3$COHORT <- 1
chs_melt_yr3 <- 
  unique(chs_yr3[
    !is.na(chs_yr3$STDYTIME),
    c("IDNO","STDYTIME","COHORT")])
names(chs_melt_yr3)[1:2] <- c("patientid","visitdays")
chs_melt_yr3$visit <- 3


#-----------------------------------------------------------------------------------#

#### CHS Year 4 ###

chs_yr4 <- read.csv("~/Dropbox/BioLINCC files/CHS_2019a/YEAR4/yr4final.csv",na.strings=c("NA","","NULL"))
names(chs_yr4) <- toupper(names(chs_yr4))

chs_yr4$COHORT <- 1
chs_melt_yr4 <- 
  unique(chs_yr4[
    !is.na(chs_yr4$STDYTIME),
    c("IDNO","STDYTIME","COHORT")])

names(chs_melt_yr4)[1:2] <- c("patientid","visitdays")
chs_melt_yr4$visit <- 4


#----------------------------------------------------------------#

#### CHS Year 5 - AA cohort ###

chs_yr5new <- read.csv("~/Dropbox/BioLINCC files/CHS_2019a/YEAR5/yr5newfinal.csv",
                       na.strings=c("NA","","NULL"))
names(chs_yr5new) <- toupper(names(chs_yr5new))

chs_yr5new$STDYTIME <- 0

chs_melt_yr5new <- 
  unique(chs_yr5new[
    !is.na(chs_yr5new$STDYTIME),
    c("IDNO","STDYTIME","COHORT")])

names(chs_melt_yr5new)[1:2] <- c("patientid","visitdays")
chs_melt_yr5new$visit <- 1


#### CHS Year 5 - Original cohort ###

chs_yr5old <- read.csv("~/Dropbox/BioLINCC files/CHS_2019a/YEAR5/yr5oldfinal.csv",na.strings=c("NA","","NULL"))
names(chs_yr5old) <- toupper(names(chs_yr5old))


chs_melt_yr5old <- 
  unique(chs_yr5old[
    !is.na(chs_yr5old$STDYTIME),
    c("IDNO","STDYTIME","COHORT")])

names(chs_melt_yr5old)[1:2] <- c("patientid","visitdays")
chs_melt_yr5old$visit <- 5


#### CHS Year 6 ###


chs_yr6 <- read.csv("~/Dropbox/BioLINCC files/CHS_2019a/YEAR6/yr6final.csv",na.strings=c("NA","","NULL"))
names(chs_yr6) <- toupper(names(chs_yr6))


chs_melt_yr6 <- 
  unique(chs_yr6[
    !is.na(chs_yr6$STDYTIME),
    c("IDNO","STDYTIME","COHORT")])

names(chs_melt_yr6)[1:2] <- c("patientid","visitdays")
chs_melt_yr6$visit <- 6


#### CHS Year 7 ###


chs_yr7 <- read.csv("~/Dropbox/BioLINCC files/CHS_2019a/YEAR7/yr7final.csv",na.strings=c("NA","","NULL"))
names(chs_yr7) <- toupper(names(chs_yr7))

chs_melt_yr7 <- 
  unique(chs_yr7[
    !is.na(chs_yr7$STDYTIME),
    c("IDNO","STDYTIME","COHORT")])

names(chs_melt_yr7)[1:2] <- c("patientid","visitdays")
chs_melt_yr7$visit <- 7


#----------------------------------------------------------------------------------------------------------------------#

#### CHS Year 8 ###

chs_yr8 <- read.csv("~/Dropbox/BioLINCC files/CHS_2019a/YEAR8/yr8final.csv",na.strings=c("NA","","NULL"))
names(chs_yr8) <- toupper(names(chs_yr8))


chs_melt_yr8 <- 
  unique(chs_yr8[
    !is.na(chs_yr8$STDYTIME),
    c("IDNO","STDYTIME","COHORT")])

names(chs_melt_yr8)[1:2] <- c("patientid","visitdays")
chs_melt_yr8$visit <- 8


#----------------------------------------------------------------------------------------------------------------------#

#### CHS Year 9 ###

chs_yr9 <- read.csv("~/Dropbox/BioLINCC files/CHS_2019a/YEAR9/yr9final.csv",na.strings=c("NA","","NULL"))
names(chs_yr9) <- toupper(names(chs_yr9))

chs_melt_yr9 <- 
  unique(chs_yr9[
    !is.na(chs_yr9$STDYTIME),
    c("IDNO","STDYTIME","COHORT")])

names(chs_melt_yr9)[1:2] <- c("patientid","visitdays")
chs_melt_yr9$visit <- 9


#----------------------------------------------------------------------------------------------------------------------#

#### CHS Year 10 ###

chs_yr10 <- read.csv("~/Dropbox/BioLINCC files/CHS_2019a/YEAR10/yr10final.csv",na.strings=c("NA","","NULL"))
names(chs_yr10) <- toupper(names(chs_yr10))

chs_melt_yr10 <- 
  unique(chs_yr10[
    !is.na(chs_yr10$STDYTIME),
    c("IDNO","STDYTIME","COHORT")])

names(chs_melt_yr10)[1:2] <- c("patientid","visitdays")
chs_melt_yr10$visit <- 10

chs_melt_yr10$form <- "yr10final"


#----------------------------------------------------------------------------------------------------------------------#

#### CHS Year 11 ###

chs_yr11<- read.csv("~/Dropbox/BioLINCC files/CHS_2019a/YEAR11/yr11final.csv",na.strings=c("NA","","NULL"))
names(chs_yr11) <- toupper(names(chs_yr11))


chs_melt_yr11 <- 
  unique(chs_yr11[
    !is.na(chs_yr11$STDYTIME),
    c("IDNO","STDYTIME","COHORT")])

names(chs_melt_yr11)[1:2] <- c("patientid","visitdays")
chs_melt_yr11$visit <- 11


#----------------------------------------------------------------------------------------------------------------------#

#### CHS Year 12 ###

chs_yr12 <- read.csv("~/Dropbox/BioLINCC files/CHS_2019a/YEAR12/year12annualfinal.csv",na.strings=c("NA","","NULL"))
chs_yr12$COHORT <- chs_baseboth$COHORT


names(chs_yr12) <- toupper(names(chs_yr12))


chs_melt_yr12 <- 
  unique(chs_yr12[
    !is.na(chs_yr12$STDYTIME),
    c("IDNO","STDYTIME","COHORT")])

names(chs_melt_yr12)[1:2] <- c("patientid","visitdays")

chs_melt_yr12$visit <- 12


#----------------------------------------------------------------------------------------------------------------------#

#### CHS Year 13 ###

chs_yr13 <- read.csv("~/Dropbox/BioLINCC files/CHS_2019a/YEAR13/year13annualfinal.csv",na.strings=c("NA","","NULL"))
names(chs_yr13) <- toupper(names(chs_yr13))

chs_yr13$COHORT <- chs_baseboth$COHORT

chs_melt_yr13 <- 
  unique(chs_yr13[
    !is.na(chs_yr13$STDYTIME),
    c("IDNO","STDYTIME","COHORT")])

names(chs_melt_yr13)[1:2] <- c("patientid","visitdays")
chs_melt_yr13$visit <- 13


#----------------------------------------------------------------------------------------------------------------------#

#### CHS year 14 ###

chs_yr14 <- read.csv("~/Dropbox/BioLINCC files/CHS_2019a/YEAR14/year14annualfinal.csv",na.strings=c("NA","","NULL"))
names(chs_yr14) <- toupper(names(chs_yr14))
chs_yr14$COHORT <- chs_baseboth$COHORT


chs_melt_yr14 <- 
  unique(chs_yr14[
    !is.na(chs_yr14$STDYTIME),
    c("IDNO","STDYTIME","COHORT")])

names(chs_melt_yr14)[1:2] <- c("patientid","visitdays")
chs_melt_yr14$visit <- 14


#----------------------------------------------------------------------------------------------------------------------#

#### CHS year 15 ###


chs_yr15 <- read.csv("~/Dropbox/BioLINCC files/CHS_2019a/YEAR15/year15annualfinal.csv",na.strings=c("NA","","NULL"))
chs_yr15$COHORT <- chs_baseboth$COHORT

names(chs_yr15) <- toupper(names(chs_yr15))


chs_melt_yr15 <- 
  unique(chs_yr15[
    !is.na(chs_yr15$STDYTIME),
    c("IDNO","STDYTIME","COHORT")])

names(chs_melt_yr15)[1:2] <- c("patientid","visitdays")
chs_melt_yr15$visit <- 15


#----------------------------------------------------------------------------------------------------------------------#


#### CHS year 16 ###

chs_yr16 <- read.csv("~/Dropbox/BioLINCC files/CHS_2019a/YEAR16/year16annualfinal.csv",na.strings=c("NA","","NULL"))
names(chs_yr16) <- toupper(names(chs_yr16))
chs_yr16$COHORT <- chs_baseboth$COHORT


chs_melt_yr16 <- 
  unique(chs_yr16[
    !is.na(chs_yr16$STDYTIME),
    c("IDNO","STDYTIME","COHORT")])

names(chs_melt_yr16)[1:2] <- c("patientid","visitdays")
chs_melt_yr16$visit <- 16


#----------------------------------------------------------------------------------------------------------------------#

#### CHS year 17 ###


chs_yr17 <- read.csv("~/Dropbox/BioLINCC files/CHS_2019a/YEAR17/year17annualfinal.csv",na.strings=c("NA","","NULL"))
names(chs_yr17) <- toupper(names(chs_yr17))
chs_yr17$COHORT <- chs_baseboth$COHORT


chs_melt_yr17 <- 
  unique(chs_yr17[
    !is.na(chs_yr17$STDYTIME),
    c("IDNO","STDYTIME","COHORT")])

names(chs_melt_yr17)[1:2] <- c("patientid","visitdays")
chs_melt_yr17$visit <- 17


#----------------------------------------------------------------------------------------------------------------------#

#### CHS year 18 ###

chs_yr18_phone <- read.csv("~/Dropbox/BioLINCC files/CHS_2019a/YEAR18/year18phonefinal.csv",na.strings=c("NA","","NULL"))
names(chs_yr18_phone) <- toupper(names(chs_yr18_phone))
chs_yr18_phone$COHORT <- chs_baseboth$COHORT

chs_melt_phone_yr18 <- 
  unique(chs_yr18_phone[
    !is.na(chs_yr18_phone$STDYTIME),
    c("IDNO","STDYTIME","COHORT")])

names(chs_melt_phone_yr18)[1:2] <- c("patientid","visitdays")
chs_melt_phone_yr18$visit <- 18


#----------------------------------------------------------------------------------------------------------------------#

#### CHS year 19 ###

chs_yr19_phone <- read.csv("~/Dropbox/BioLINCC files/CHS_2019a/YEAR19/year19phonefinal.csv",na.strings=c("NA","","NULL"))
names(chs_yr19_phone) <- toupper(names(chs_yr19_phone))
chs_yr19_phone$COHORT <- chs_baseboth$COHORT


chs_melt_phone_yr19 <- 
  unique(chs_yr19_phone[
    !is.na(chs_yr19_phone$STDYTIME),
    c("IDNO","STDYTIME","COHORT")])

names(chs_melt_phone_yr19)[1:2] <- c("patientid","visitdays")

chs_melt_phone_yr19$visit <- 19


#----------------------------------------------------------------------------------------------------------------------#

#### CHS year 20 ###

chs_yr20_phone <- read.csv("~/Dropbox/BioLINCC files/CHS_2019a/YEAR20/year20phonefinal.csv",na.strings=c("NA","","NULL"))
names(chs_yr20_phone) <- toupper(names(chs_yr20_phone))
chs_yr20_phone$COHORT <- chs_baseboth$COHORT

chs_melt_phone_yr20 <- 
  unique(chs_yr20_phone[
    !is.na(chs_yr20_phone$STDYTIME),
    c("IDNO","STDYTIME","COHORT")])

names(chs_melt_phone_yr20)[1:2] <- c("patientid","visitdays")
chs_melt_phone_yr20$visit <- 20


#----------------------------------------------------------------------------------------------------------------------#

#### CHS year 21 ###

chs_yr21_phone <- read.csv("~/Dropbox/BioLINCC files/CHS_2019a/YEAR21/year21phonefinal.csv",na.strings=c("NA","","NULL"))
names(chs_yr21_phone) <- toupper(names(chs_yr21_phone))
chs_yr21_phone$COHORT <- chs_baseboth$COHORT

chs_melt_phone_yr21 <- 
  unique(chs_yr21_phone[
    !is.na(chs_yr21_phone$STDYTIME),
    c("IDNO","STDYTIME","COHORT")])

names(chs_melt_phone_yr21)[1:2] <- c("patientid","visitdays")
chs_melt_phone_yr21$visit <- 21


#----------------------------------------------------------------------------------------------------------------------#

### CHS year 22 ###

chs_yr22_phone <- read.csv("~/Dropbox/BioLINCC files/CHS_2019a/YEAR22/year22phonefinal.csv",na.strings=c("NA","","NULL"))
names(chs_yr22_phone) <- toupper(names(chs_yr22_phone))
chs_yr22_phone$COHORT <- chs_baseboth$COHORT

chs_melt_phone_yr22 <- 
  unique(chs_yr22_phone[
    !is.na(chs_yr22_phone$STDYTIME),
    c("IDNO","STDYTIME","COHORT")])

names(chs_melt_phone_yr22)[1:2] <- c("patientid","visitdays")
chs_melt_phone_yr22$visit <- 22

chs_melt_phone_yr22$form <- "yr22_phone"


#----------------------------------------------------------------------------------------------------------------------#

### CHS year 23 ###

chs_yr23_phone <- read.csv("~/Dropbox/BioLINCC files/CHS_2019a/YEAR23/year23phonefinal.csv",na.strings=c("NA","","NULL"))
chs_yr23_phone$COHORT <- chs_baseboth$COHORT

chs_melt_phone_yr23 <- 
  unique(chs_yr23_phone[
    !is.na(chs_yr23_phone$STDYTIME),
    c("IDNO","STDYTIME","COHORT")])

names(chs_melt_phone_yr23)[1:2] <- c("patientid","visitdays")
chs_melt_phone_yr23$visit <- 23


#----------------------------------------------------------------------------------------------------------------------#

### CHS year 24 ###

chs_yr24 <- read.csv("~/Dropbox/BioLINCC files/CHS_2019a/YEAR24/year24annualfinal.csv",na.strings=c("NA","","NULL"))
names(chs_yr24) <- toupper(names(chs_yr24))
chs_yr24$COHORT <- chs_baseboth$COHORT

chs_melt_phone_yr24 <- 
  unique(chs_yr24_phone[
    !is.na(chs_yr24_phone$STDYTIME),
    c("IDNO","STDYTIME","COHORT")])

names(chs_melt_phone_yr24)[1:2] <- c("patientid","visitdays")
chs_melt_phone_yr24$visit <- 24


#----------------------------------------------------------------------------------------------------------------------#

### *** CHS final assembly *** ###

chs_melt_all <- rbind(
  chs_melt_baseboth[,c("patientid","COHORT","visitdays","visit")],
  chs_melt_yr3[,c("patientid","COHORT","visitdays","visit")],
  chs_melt_yr4[,c("patientid","COHORT","visitdays","visit")],
  chs_melt_yr5old[,c("patientid","COHORT","visitdays","visit")],
  chs_melt_yr6[,c("patientid","COHORT","visitdays","visit")],
  chs_melt_yr7[,c("patientid","COHORT","visitdays","visit")],
  chs_melt_yr8[,c("patientid","COHORT","visitdays","visit")],
  chs_melt_yr9[,c("patientid","COHORT","visitdays","visit")],
  chs_melt_yr10[,c("patientid","COHORT","visitdays","visit")],
  chs_melt_yr11[,c("patientid","COHORT","visitdays","visit")],
  chs_melt_yr12[,c("patientid","COHORT","visitdays","visit")],
  chs_melt_yr13[,c("patientid","COHORT","visitdays","visit")],
  chs_melt_yr14[,c("patientid","COHORT","visitdays","visit")],
  chs_melt_yr15[,c("patientid","COHORT","visitdays","visit")],
  chs_melt_yr16[,c("patientid","COHORT","visitdays","visit")],
  chs_melt_yr17[,c("patientid","COHORT","visitdays","visit")],
  chs_melt_phone_yr18[,c("patientid","COHORT","visitdays","visit")],
  chs_melt_phone_yr19[,c("patientid","COHORT","visitdays","visit")],
  chs_melt_phone_yr20[,c("patientid","COHORT","visitdays","visit")],
  chs_melt_phone_yr21[,c("patientid","COHORT","visitdays","visit")],
  chs_melt_phone_yr22[,c("patientid","COHORT","visitdays","visit")],
  chs_melt_phone_yr23[,c("patientid","COHORT","visitdays","visit")],
  chs_melt_phone_yr24[,c("patientid","COHORT","visitdays","visit")]
)

chs_melt_all <-
  merge(chs_melt_all,
        chs_baseboth[,c("IDNO","agey")],
        by.x=c("patientid"),
        by.y="IDNO",
        all.x=T)

chs_melt_all$age_obs <-
  floor(chs_melt_all$agey+chs_melt_all$visitdays/365)

chs_melt_all$study <- "CHS"

chs_melt_all <-
  merge(chs_melt_all,
        visit_yrs,
        by.x=c("study","COHORT","visit"),
        by.y=c("study","cohort","visit"),
        all.x=T)

chs_dates_long <-
  sqldf("select distinct
        patientid,
        age_obs,
        visitdays,
        visit_yr,
        study,
        cohort,
        cohort_name
        from chs_melt_all
        ")

#### MESA ####


mesa_exam1 <- read.csv("~/Dropbox/BioLINCC files/MESA/Primary/Exam1/Data/mesae1dres06192012.csv",na.strings=c("NA","","NULL"))
mesa_exam2 <- read.csv("~/Dropbox/BioLINCC files/MESA/Primary/Exam2/Data/mesae2dres06222012.csv",na.strings=c("NA","","NULL"))
mesa_exam3 <- read.csv("~/Dropbox/BioLINCC files/MESA/Primary/Exam3/Data/mesae3dres06222012.csv",na.strings=c("NA","","NULL"))
mesa_exam4 <- read.csv("~/Dropbox/BioLINCC files/MESA/Primary/Exam4/Data/mesae4dres06222012.csv",na.strings=c("NA","","NULL"))
mesa_exam5 <- read.csv("~/Dropbox/BioLINCC files/MESA/Primary/Exam5/Data/mesae5_drepos_20210920.csv",na.strings=c("NA","","NULL"))

names(mesa_exam1) <- tolower(names(mesa_exam1))
names(mesa_exam2) <- tolower(names(mesa_exam2))
names(mesa_exam3) <- tolower(names(mesa_exam3))
names(mesa_exam4) <- tolower(names(mesa_exam4))
names(mesa_exam5) <- tolower(names(mesa_exam5))


mesa_efevents <- read.csv("~/Dropbox/BioLINCC files/MESA/Primary/Events/CVD/Data/mesaevefthru2015_drepos_20200330.csv",na.strings=c("NA","","NULL"))
mesa_events <- read.csv("~/Dropbox/BioLINCC files/MESA/Primary/Events/CVD/Data/mesaevthr2015_drepos_20200330.csv",na.strings=c("NA","","NULL"))
mesa_site <- read.csv("~/Dropbox/BioLINCC files/MESA/Primary/Site/mesa_site_drepos_20181106.csv")

mesa_site$cohort_name[mesa_site$site1c=="A"] <- "MD" 
mesa_site$cohort_name[mesa_site$site1c=="B"] <- "MN" 
mesa_site$cohort_name[mesa_site$site1c=="C"] <- "IL" 
mesa_site$cohort_name[mesa_site$site1c=="D"] <- "CA" 
mesa_site$cohort_name[mesa_site$site1c=="E"] <- "NC" 
mesa_site$cohort_name[mesa_site$site1c=="F"] <- "NY" 

names(mesa_efevents) <- tolower(names(mesa_efevents))
names(mesa_events) <- tolower(names(mesa_events))

mesa_dates <- mesa_exam1[,c("mesaid","age1c")]
mesa_dates <- merge(mesa_dates, mesa_exam2[,c("mesaid","age2c","e12dyc")],all.x=T)
mesa_dates <- merge(mesa_dates, mesa_exam3[,c("mesaid","age3c","e13dyc")],all.x=T)
mesa_dates <- merge(mesa_dates, mesa_exam4[,c("mesaid","age4c","e14dyc")],all.x=T)
mesa_dates <- merge(mesa_dates, mesa_exam5[,c("mesaid","age5c","e15dyc")],all.x=T)

mesa_sex <- mesa_exam1[,c("mesaid","gender1")]

mesa_dates_1 <- mesa_exam1[,c("mesaid","age1c")]
names(mesa_dates_1)[names(mesa_dates_1)=="age1c"] <- "age_obs"
mesa_dates_1$visitdays <- 0
mesa_dates_1$visit <- 1
mesa_dates_1$visit_yr <- 0

mesa_dates_2 <-mesa_exam2[,c("mesaid","age2c","e12dyc")]
names(mesa_dates_2)[names(mesa_dates_2)=="age2c"] <- "age_obs"
names(mesa_dates_2)[names(mesa_dates_2)=="e12dyc"] <- "visitdays"
mesa_dates_2$visit_yr <- 3
mesa_dates_2$visit <- 2

mesa_dates_3 <-mesa_exam3[,c("mesaid","age3c","e13dyc")]
names(mesa_dates_3)[names(mesa_dates_3)=="age3c"] <- "age_obs"
names(mesa_dates_3)[names(mesa_dates_3)=="e13dyc"] <- "visitdays"
mesa_dates_3$visit <- 3
mesa_dates_3$visit_yr <- 5

mesa_dates_4 <-mesa_exam4[,c("mesaid","age4c","e14dyc")]
names(mesa_dates_4)[names(mesa_dates_4)=="age4c"] <- "age_obs"
names(mesa_dates_4)[names(mesa_dates_4)=="e14dyc"] <- "visitdays"
mesa_dates_4$visit <- 4
mesa_dates_4$visit_yr <- 6

mesa_dates_5 <-mesa_exam5[,c("mesaid","age5c","e15dyc")]
names(mesa_dates_5)[names(mesa_dates_5)=="age5c"] <- "age_obs"
names(mesa_dates_5)[names(mesa_dates_5)=="e15dyc"] <- "visitdays"
mesa_dates_5$visit <- 5
mesa_dates_5$visit_yr <- 10

mesa_dates_long <- rbind(mesa_dates_1,
                         mesa_dates_2,
                         mesa_dates_3,
                         mesa_dates_4,
                         mesa_dates_5)

mesa_dates_long <- subset(mesa_dates_long,!is.na(visitdays))

mesa_dates_long <- merge(mesa_dates_long,
                         mesa_sex,
                         by="mesaid")

mesa_dates <- merge(mesa_dates,mesa_events,by="mesaid")
mesa_dates <- merge(mesa_dates,
                    mesa_site[,c("MESAID","site1c","cohort_name")],
                    by.x="mesaid",
                    by.y="MESAID",
                    all.x=T)

mesa_dates_long <- merge(mesa_dates_long,
                         mesa_site[,c("MESAID","site1c","cohort_name")],
                         by.x="mesaid",
                         by.y="MESAID",
                         all.x=T)

mesa_dates_long$study <- "MESA"

names(mesa_dates_long)[names(mesa_dates_long)=="site1c"] <- "cohort"


mesa_dates$age_hf <- round(mesa_dates$age1c+mesa_dates$chftt/365)

mesa_dates$last_exam_before_hf[mesa_dates$chf==1&mesa_dates$chftt>0] <- 1
mesa_dates$last_exam_before_hf[mesa_dates$chf==1&mesa_dates$chftt>=mesa_dates$e12dyc] <- 3
mesa_dates$last_exam_before_hf[mesa_dates$chf==1&mesa_dates$chftt>=mesa_dates$e13dyc] <- 5
mesa_dates$last_exam_before_hf[mesa_dates$chf==1&mesa_dates$chftt>=mesa_dates$e14dyc] <- 6
mesa_dates$last_exam_before_hf[mesa_dates$chf==1&mesa_dates$chftt>=mesa_dates$e15dyc] <- 10

mesa_dates$last_exam_date_before_hf[mesa_dates$last_exam_before_hf==1&!is.na(mesa_dates$last_exam_before_hf)] <-  0
mesa_dates$last_exam_date_before_hf[mesa_dates$last_exam_before_hf==2&!is.na(mesa_dates$last_exam_before_hf)] <-  mesa_dates$e12dyc[mesa_dates$last_exam_before_hf==2&!is.na(mesa_dates$last_exam_before_hf)]
mesa_dates$last_exam_date_before_hf[mesa_dates$last_exam_before_hf==3&!is.na(mesa_dates$last_exam_before_hf)] <-  mesa_dates$e13dyc[mesa_dates$last_exam_before_hf==3&!is.na(mesa_dates$last_exam_before_hf)]
mesa_dates$last_exam_date_before_hf[mesa_dates$last_exam_before_hf==4&!is.na(mesa_dates$last_exam_before_hf)] <-  mesa_dates$e14dyc[mesa_dates$last_exam_before_hf==4&!is.na(mesa_dates$last_exam_before_hf)]
mesa_dates$last_exam_date_before_hf[mesa_dates$last_exam_before_hf==5&!is.na(mesa_dates$last_exam_before_hf)] <-  mesa_dates$e15dyc[mesa_dates$last_exam_before_hf==5&!is.na(mesa_dates$last_exam_before_hf)]

mesa_dates$hf_dys_last_exam <- mesa_dates$chftt-mesa_dates$last_exam_before_hf
mesa_dates$time_hf_to_dth <- mesa_dates$dthtt-mesa_dates$chftt


#### JHS ####


jhs_analysis1 <- read.csv("~/Dropbox/BioLINCC files/JHS/Data/Analysis_Data/CSV/analysis1.csv",na.strings=c("","NA","NULL"))
jhs_analysis2 <- read.csv("~/Dropbox/BioLINCC files/JHS/Data/Analysis_Data/CSV/analysis2.csv",na.strings=c("","NA","NULL"))
jhs_analysis3 <- read.csv("~/Dropbox/BioLINCC files/JHS/Data/Analysis_Data/CSV/analysis3.csv",na.strings=c("","NA","NULL"))

jhs_v1date <- 
  sqldf('select newid as patientid, 
        0 as visitdays, 
        1 as visit, 
        0 as visit_yr, 
        age as age_obs,
        ARIC 
        from jhs_analysis1')

jhs_v2date <- 
  sqldf('select newid as patientid,
        DaysFromV1 as visitdays, 
        2 as visit, 
        6 as visit_yr, 
        age as age_obs 
        from jhs_analysis2')


jhs_v3date <- 
  sqldf('select newid as patientid, 
        DaysFromV1 as visit_days, 
        3 as visit, 
        9 as visit_yr, 
        age as age_obs 
        from jhs_analysis3')


## Create a dataset starting at a set age for all patients who survive to that age by adding the observation day to baseline age.
## From there,  choose the most recent value available for each patient/variable pair going all the way back to baseline.
## This will be accomplished by subsetting on age at observation (age_obs) being
## Will use cutoff of age_obs <= specified baseline age to capture baseline observations (visitdays = 0, so age_obs==baseline age)
## Then will subset again to select highest available age for each patient/variable pair (i.e. most recent)
## Need to check on methdologic issues here - should be a sub-analysis until validity/robustness proven.


jhs_dates$cohort <- jhs_dates$ARIC

jhs_dates$cohort_name[jhs_dates$cohort==1] <- "ARIC"
jhs_dates$cohort_name[jhs_dates$cohort==0] <- "Not ARIC"


jhs_melt_analysis1 <- melt(jhs_analysis1,c(id.vars=c("newid","visit")),na.rm=T,stringsAsFactor=T)
jhs_melt_analysis1$visitdays <- 0
jhs_melt_analysis1$visit_yr <- 0


jhs_melt_analysis2 <- 
  unique(jhs_analysis2[,c("newid","visit","DaysFromV1")])
names(jhs_melt_analysis2)[names(jhs_melt_analysis2)=="DaysFromV1"] <- "visitdays"
jhs_melt_analysis2$visit_yr <- 6


jhs_melt_analysis3 <- melt(jhs_analysis3,c(id.vars=c("newid","visit","DaysFromV1")),na.rm=T,stringsAsFactor=T)
names(jhs_melt_analysis3)[names(jhs_melt_analysis3)=="DaysFromV1"] <- "visitdays"
jhs_melt_analysis3$form <- "analysis3"
jhs_melt_analysis3$visit_yr <- 9


# ==============================================================#
####             *** JHS longitudinal visits ***             ####
# ==============================================================#

## Loading and preparing annual longitudinal visits here
## Currently Visit 1 = baseline, Visit 2  = 6 years, Visit 3 = 9 years.
## afulong visit assigned by taking time since baseline rounded to nearest year (jhs_afulong$time) +1
## 1 added because Baseline visit = Visit 1
## Then remove any afulong record with assigned visit 1,6,or 9 to eliminate clashing with main study visits.


jhs_afulong <- read.csv("~/Dropbox/BioLINCC files/JHS/Data/AFU/CSV/afulong.csv",na.strings=c("","NA","NULL"))

jhs_afulong$visit_yr <- 
  round(jhs_afulong$time)

jhs_afulong$visitdays <- 
  as.numeric(
    difftime(
      as.Date(jhs_afulong$date,"%m/%d/%Y"),
      as.Date(jhs_afulong$v1date,"%m/%d/%Y"),
      units="days"))

jhs_afulong$visit2days <- 
  as.numeric(
    difftime(
      as.Date(jhs_afulong$date,"%m/%d/%Y"),
      as.Date(jhs_afulong$V2date,"%m/%d/%Y"),
      units="days"))

jhs_afulong$visit2years <- round(jhs_afulong$visit2days/365,0)+1

jhs_afulong$visit3days <- 
  as.numeric(
    difftime(
      as.Date(jhs_afulong$date,"%m/%d/%Y"),
      as.Date(jhs_afulong$V3date,"%m/%d/%Y"),
      units="days"))

jhs_afulong$visit3years <- round(jhs_afulong$visit3days/365,0)+1

jhs_afulong <- subset(jhs_afulong,
                      !visit_yr %in% c(-2,-1,0,6,9)&
                        abs(days_from_v2)>180&
                        abs(days_from_v3)&
                        abs(visitdays)>180)

## This statement handles a situation where multiple studies round to the same visit, resulting in multiple separate lines for that visit.
## This solution takes the MAX (or latest) observation - this can be changed to the earliest, etc.

jhs_afulong <- sqldf("select * 
                      from jhs_afulong
                      group by newid,visit_yr
                      having max(visitdays)")

jhs_melt_afulong <- 
  unique(jhs_afulong[,c("newid",
                        "visit_yr",
                        "visitdays")])
         
jhs_melt_afulong$form <-"afulong"


# ==============================================================--#
####                ** Assemble all JHS tables **              ####
# ==============================================================--#

jhs_melt_all <- rbind(jhs_melt_analysis1[,c("newid","visit_yr","visitdays","variable","value","form")],
                      jhs_melt_analysis2[,c("newid","visit_yr","visitdays","variable","value","form")],
                      jhs_melt_analysis3[,c("newid","visit_yr","visitdays","variable","value","form")],
                      jhs_melt_afulong[,c("newid","visit_yr","visitdays","variable","value","form")])


names(jhs_melt_all)[1] <- "patientid"
jhs_melt_all$study <- "JHS"

jhs_melt_all$study_field <- paste(jhs_melt_all$study,
                                  jhs_melt_all$form,
                                  jhs_melt_all$variable,sep="_")

##  Add age at observation for assembling cohorts of same starting age

jhs_melt_all <- merge(jhs_melt_all,jhs_dates[,c("patientid","enroll_yr","age")], by="patientid")

names(jhs_melt_all)[ncol(jhs_melt_all)] <- "baseline_age"

jhs_melt_all$age_obs <- floor(jhs_melt_all$baseline_age+jhs_melt_all$visitdays/365)

jhs_dates_long <- sqldf("select distinct
                        patientid,
                        age_obs,
                        visitdays,
                        visit_yr,
                        study
                        from jhs_melt_all")


jhs_dates_long <- merge(jhs_dates_long,
                        jhs_dates[,c("patientid","cohort","cohort_name","sex")],
                        by="patientid")
```
