## Supplementary material for "Aggregating data to accelerate personalized therapy in heart failure (ADAPT-HF)": Harmonization scripts: cohort outcomes.R

```
library(sqldf)
library(Hmisc)
library(plotrix)
library(reshape2)
library(descr)
library(dplyr)
library(relper)
library(stringr)
library(bigrquery)
library(haven)
library(googleCloudStorageR)

source("~/Dropbox/R scripts/Misc clinical scripts.R")
source("~/Dropbox/R scripts/echo scripts.R")

f <- function(input,output) {
  write.csv(input,file=output, row.names=F, na="")
}

visit_yrs <- read.csv('~/Dropbox/ADAPT-HF/Master HDCP files/Cohort visit yrs.csv')

data_fields <- 
  c("study",
    "patientid",
    "visit_yr",
    "visitdays",
    "form",
    "variable",
    "value",
    "datapoint",
    "study_field",
    "cohort",
    "cohort_name",
    "age_obs")

outcomes_list <- c("dth_status","dth_dt",
                   "cvdth_status","cvdth_dt",
                   "hfdth_status","hfdth_dt",
                   "hfhosp_ef_val","hfhosp_ef_cat",
                   "noncvdth_status","noncvdth_dt",
                   "hosp_status","hosp_dt",
                   "cadhosp_status","cadhosp_dt",
                   "cvhosp_status","cvhosp_dt",
                   "hfhosp_status","hfhosp_dt",
                   "noncvhosp_status","noncvhosp_dt")


#### FHS Outcomes ####

fhs1_chfinit <- read.csv('~/Dropbox/BioLINCC files/Framingham cohort/datasets/CSV/vr_chfinit_2013_a_0828d.csv',na.strings=c("NA","","NULL"))
fhs1_survcvd <- read.csv('~/Dropbox/BioLINCC files/Framingham cohort/datasets/CSV/vr_survcvd_2018_a_1267d.csv',na.strings=c("NA","","NULL"))
fhs1_survdth <- read.csv('~/Dropbox/BioLINCC files/Framingham cohort/datasets/CSV/vr_survdth_2018_a_1268d.csv',na.strings=c("NA","","NULL"))
fhs1_survstk <- read.csv('~/Dropbox/BioLINCC files/Framingham cohort/datasets/CSV/vr_svstk_2018_a_1269d.csv',na.strings=c("NA","","NULL"))
fhs1_soechf <- read.csv('~/Dropbox/BioLINCC files/Framingham cohort/datasets/CSV/VR_SOECHF_2016_a_1070d.csv',na.strings=c("NA","","NULL"))
fhs1_afcum <- read.csv('~/Dropbox/BioLINCC files/Framingham cohort/datasets/CSV/VR_AFCUM_2019_a_1186d_v1.csv',na.strings=c("NA","","NULL")) # Cumulative Atrial fibrillation
fhs1_survaf <- read.csv('~/Dropbox/BioLINCC files/Framingham cohort/datasets/CSV/VR_SURVAF_2018_a_1272D.csv',na.strings=c("NA","","NULL")) # Cumulative Atrial fibrillation
fhs1_cancer <- read.csv('~/Dropbox/BioLINCC files/Framingham cohort/datasets/CSV/VR_CANCER_2019_A_1162D.csv',na.strings=c("NA","","NULL")) # Incident cancer
fhs1_diab <- read.csv('~/Dropbox/BioLINCC files/Framingham cohort/datasets/CSV/VR_DIAB_EX28_0_0601D.csv',na.strings=c("NA","","NULL")) # Diabetes status by cycle
fhs1_soe <- read.csv('~/Dropbox/BioLINCC files/Framingham cohort/datasets/CSV/VR_SOE_2019_A_1217D.csv',na.strings=c("NA","","NULL"))

names(fhs1_chfinit) <- toupper(names(fhs1_chfinit))
names(fhs1_survcvd) <- toupper(names(fhs1_survcvd))
names(fhs1_survdth) <- toupper(names(fhs1_survdth))
names(fhs1_soechf) <- toupper(names(fhs1_soechf))
names(fhs1_afcum) <- toupper(names(fhs1_afcum))
names(fhs1_survaf) <- toupper(names(fhs1_survaf))
names(fhs1_cancer) <- toupper(names(fhs1_cancer))
names(fhs1_diab) <- toupper(names(fhs1_diab))
names(fhs1_soe) <- toupper(names(fhs1_soe))


fhs2_chfinit <- read.csv('~/Dropbox/BioLINCC files/Framingham offspring/datasets/CSV/vr_chfinit_2013_a_0828d.csv',na.strings=c("NA","","NULL"))
fhs2_survcvd <- read.csv('~/Dropbox/BioLINCC files/Framingham offspring/datasets/CSV/vr_survcvd_2019_a_1334d.csv',na.strings=c("NA","","NULL"))
fhs2_survdth <- read.csv('~/Dropbox/BioLINCC files/Framingham offspring/datasets/CSV/vr_survdth_2019_a_1337d.csv',na.strings=c("NA","","NULL"))
fhs2_survstk <- read.csv('~/Dropbox/BioLINCC files/Framingham offspring/datasets/CSV/vr_svstk_2019_a_1335d.csv',na.strings=c("NA","","NULL"))
fhs2_soechf <- read.csv('~/Dropbox/BioLINCC files/Framingham offspring/datasets/CSV/VR_SOECHF_2016_A_1070D.csv',na.strings=c("NA","","NULL"))
fhs2_afcum <- read.csv('~/Dropbox/BioLINCC files/Framingham offspring/datasets/CSV/VR_AFCUM_2021_A_1366D.csv',na.strings=c("NA","","NULL")) # Cumulative Atrial fibrillation
fhs2_survaf <- read.csv('~/Dropbox/BioLINCC files/Framingham offspring/datasets/CSV/VR_SURVAF_2019_A_1338D.csv',na.strings=c("NA","","NULL")) # Cumulative Atrial fibrillation
fhs2_cancer <- read.csv('~/Dropbox/BioLINCC files/Framingham offspring/datasets/CSV/VR_CANCER_2019_A_1162D.csv',na.strings=c("NA","","NULL")) # Incident cancer
fhs2_diab <- read.csv('~/Dropbox/BioLINCC files/Framingham offspring/datasets/CSV/vr_diab_ex09_1_1002d.csv',na.strings=c("NA","","NULL")) # Diabetes status by cycle
fhs2_soe <- read.csv('~/Dropbox/BioLINCC files/Framingham offspring/datasets/CSV/VR_SOE_2020_A_1340D.csv',na.strings=c("NA","","NULL"))

names(fhs2_chfinit) <- toupper(names(fhs2_chfinit))
names(fhs2_survcvd) <- toupper(names(fhs2_survcvd))
names(fhs2_survdth) <- toupper(names(fhs2_survdth))
names(fhs2_survstk) <- toupper(names(fhs2_survstk))
names(fhs2_soechf) <- toupper(names(fhs2_soechf))
names(fhs2_afcum) <- toupper(names(fhs2_afcum))
names(fhs2_survaf) <- toupper(names(fhs2_survaf))
names(fhs2_cancer) <- toupper(names(fhs2_cancer))
names(fhs2_diab) <- toupper(names(fhs2_diab))
names(fhs2_soe) <- toupper(names(fhs2_soe))

fhs2_afcum$ORIGIN_SOURCE <- NA

fhs3_chfinit <- read.csv('~/Dropbox/BioLINCC files/Framingham third generation/datasets/CSV/vr_chfinit_2013_a_0828d.csv',na.strings=c("NA","","NULL"))
fhs3_survcvd <- read.csv('~/Dropbox/BioLINCC files/Framingham third generation/datasets/CSV/vr_survcvd_2019_a_1334d.csv',na.strings=c("NA","","NULL"))
fhs3_survdth <- read.csv('~/Dropbox/BioLINCC files/Framingham third generation/datasets/CSV/vr_survdth_2019_a_1337d.csv',na.strings=c("NA","","NULL"))
fhs3_survstk <- read.csv('~/Dropbox/BioLINCC files/Framingham third generation/datasets/CSV/vr_svstk_2019_a_1335d.csv',na.strings=c("NA","","NULL"))
fhs3_soechf <- read.csv('~/Dropbox/BioLINCC files/Framingham third generation/datasets/CSV/VR_SOECHF_2016_a_1070D.csv',na.strings=c("NA","","NULL"))
fhs3_afcum <- read.csv('~/Dropbox/BioLINCC files/Framingham third generation/datasets/CSV/VR_AFCUM_2021_A_1366D.csv',na.strings=c("NA","","NULL")) # Cumulative Atrial fibrillation
fhs3_survaf <- read.csv('~/Dropbox/BioLINCC files/Framingham third generation/datasets/CSV/VR_SURVAF_2019_A_1338D.csv',na.strings=c("NA","","NULL")) # Cumulative Atrial fibrillation
fhs3_cancer <- read.csv('~/Dropbox/BioLINCC files/Framingham third generation/datasets/CSV/VR_CANCER_2019_A_1162D.csv',na.strings=c("NA","","NULL")) # Incident cancer
fhs3_diab <- read.csv('~/Dropbox/BioLINCC files/Framingham third generation/datasets/CSV/VR_DIAB_EX03_3b_1312D.csv',na.strings=c("NA","","NULL")) # Diabetes status by cycle
fhs3_soe <- read.csv('~/Dropbox/BioLINCC files/Framingham third generation/datasets/CSV/VR_SOE_2020_A_1340D.csv',na.strings=c("NA","","NULL"))

fhs3_afcum$ORIGIN_SOURCE <- NA


dm_names <- unique(c(names(fhs1_diab),names(fhs2_diab),names(fhs3_diab)))

fhs1_diab[,dm_names[!dm_names %in% names(fhs1_diab)]] <- NA
fhs2_diab[,dm_names[!dm_names %in% names(fhs2_diab)]] <- NA
fhs3_diab[,dm_names[!dm_names %in% names(fhs3_diab)]] <- NA


fhs_chfinit <- rbind(fhs1_chfinit,fhs2_chfinit,fhs3_chfinit)
fhs_survcvd <- rbind(fhs1_survcvd,fhs2_survcvd,fhs3_survcvd)
fhs_survdth <- rbind(fhs1_survdth,fhs2_survdth,fhs3_survdth)
fhs_survstk <- rbind(fhs1_survstk,fhs2_survstk,fhs3_survstk)
fhs_survaf <- rbind(fhs1_survaf,fhs2_survaf,fhs3_survaf)
fhs_afcum <- rbind(fhs1_afcum,fhs2_afcum,fhs3_afcum)
fhs_soechf <- rbind(fhs1_soechf,fhs2_soechf,fhs3_soechf)
fhs_soe <- rbind(fhs1_soe,fhs2_soe,fhs3_soe)
fhs_cancer <- rbind(fhs1_cancer,fhs2_cancer,fhs3_cancer)
fhs_diab <- rbind(fhs1_diab,fhs2_diab,fhs3_diab)

fhs_survca <- subset(fhs_cancer,(!TOPO==173)|(TOPO==173&HIST>=8720&HIST<=8790)|HIST==8247)

fhs_survca <- 
  fhs_cancer %>%
  group_by(IDTYPE,PID) %>%
  filter(D_DATE == min(D_DATE,na.rm=T))

fhs_survca_first_any <- 
  sqldf("select distinct PID,IDTYPE,D_DATE
        from fhs_survca")

fhs_survdth$dth_status <- 0 
fhs_survdth$dth_status[!is.na(fhs_survdth$DATEDTH)] <- 1 
fhs_survdth$dth_dt <- fhs_survdth$DATEDTH
fhs_survdth$dth_dt[fhs_survdth$dth_status==0] <- fhs_survdth$LASTCON[fhs_survdth$dth_status==0]

fhs_survdth$cvdth_status <- 0 
fhs_survdth$cvdth_status[fhs_survdth$CVDDEATH==1] <- 1
fhs_survdth$cvdth_dt <- fhs_survdth$dth_dt

fhs_survdth$noncvdth_status <- 0 
fhs_survdth$noncvdth_status[fhs_survdth$CVDDEATH==0] <- 1
fhs_survdth$noncvdth_dt <- fhs_survdth$dth_dt

names(fhs_survcvd)[c(3,4,5,6,7,8)] <- 
  c("cadhosp_status",
    "cadhosp_dt",
    "hfhosp_status",
    "hfhosp_dt",
    "cvhosp_status",
    "cvhosp_dt")

fhs_outcomes <- merge(fhs_survdth[,c("PID","IDTYPE",
                                     "dth_status","dth_dt",
                                     "cvdth_status","cvdth_dt",
                                     "noncvdth_status","noncvdth_dt")],
                      fhs_survcvd[,c("PID","IDTYPE",
                                     "cadhosp_status","cadhosp_dt",
                                     "cvhosp_status","cvhosp_dt",
                                     "hfhosp_status","hfhosp_dt")])


fhs_outcomes <- merge(fhs_outcomes,
                      fhs_chfinit[,c("PID",
                                     "IDTYPE",
                                     "CHF011D",
                                     "CHF017")],
                      by.x=c("PID","IDTYPE","hfhosp_dt"),
                      by.y=c("PID","IDTYPE","CHF011D"),
                      all.x=T)


fhs_outcomes$hfhosp_ef_cat[fhs_outcomes$CHF017==0] <- "Normal"
fhs_outcomes$hfhosp_ef_cat[fhs_outcomes$CHF017==3] <- "Mildly reduced"
fhs_outcomes$hfhosp_ef_cat[fhs_outcomes$CHF017==2] <- "Moderately reduced"
fhs_outcomes$hfhosp_ef_cat[fhs_outcomes$CHF017==1] <- "Severely reduced"

fhs_outcomes$study <- "FHS"
fhs_outcomes$patientid <- fhs_outcomes$PID

fhs_outcomes[,outcomes_list[!outcomes_list %in% names(fhs_outcomes)]] <- NA


if (bq_table_exists("harmonization-286013.cohorts.fhs_outcomes")) {
  bq_table_delete("harmonization-286013.cohorts.fhs_outcomes") }

bq_table_upload(x="harmonization-286013.cohorts.fhs_outcomes",
                fhs_outcomes,quiet=T,
                create_disposition="CREATE_IF_NEEDED",
                write_disposition="WRITE_TRUNCATE")


write.csv(fhs_outcomes,
          "~/Dropbox/ADAPT-HF/Master HDCP files/Cohort data/fhs_outcomes.csv",
          row.names=F,
          na="")


gcs_auth("~/Dropbox/ADAPT-HF/Master HDCP files/harmonization-286013-39f492122f69.json")
gcs_upload(file=fhs_outcomes, 
           bucket="master_hdcp_files",
           name="fhs_outcomes.csv",
           object_function = f)


#### ARIC Outcomes ####

aric_incps <- read.csv('~/Dropbox/BioLINCC files/ARIC/cohort_incident/csv/incps18.csv',na.strings=c("NA","","NULL"))
aric_hfschips <- read.csv('~/Dropbox/BioLINCC files/ARIC/cohort_HF/csv/hfcchips18.csv',na.strings=c("NA","","NULL"))


aric_outcomes <- sqldf('select distinct ID_C, 
                        C7_INCHF_P_V5 as hf_after_exam5_status, 
                        C7_FT_INCHF_P_V5 as hf_after_exam5_dys,
                        ISP18 as cadhosp_status,
                        C7_FUTIMEA as cadhosp_dt,
                        PRVCHD05 as baseline_chd,
                        INCHF18 as hfhosp_status, 
                        C7_FUTIMEHF as hfhosp_dt,
                        PREVHF01 as baseline_hf,
                        DEAD18 as dth_status, 
                        FUDTH18 as dth_dt
                       from aric_incps')

aric_outcomes$hfhosp_status[aric_outcomes$baseline_hf==1&!is.na(aric_outcomes$baseline_hf)] <- 1
aric_outcomes$hfhosp_dt[aric_outcomes$baseline_hf==1&!is.na(aric_outcomes$baseline_hf)] <- 0

aric_hf_cohort <- read.csv('~/Dropbox/BioLINCC files/ARIC/cohort_HF/csv/hfcoccps18.csv',na.strings=c("NA","","NULL"))

aric_hf_cohort$visit_yr <- floor(aric_hf_cohort$HFDAYS/365)

aric_hfchfaps <- read.csv('~/Dropbox/BioLINCC files/ARIC/cohort_HF/csv/hfchfaps18.csv')


aric_hfchfaps$dispo[aric_hfchfaps$HFAA0D=="D"] <- "Deceased"
aric_hfchfaps$dispo[aric_hfchfaps$HFAA0D1=="Y"] <- "Home"
aric_hfchfaps$dispo[aric_hfchfaps$HFAA0D2=="Y"] <- "Home health"
aric_hfchfaps$dispo[aric_hfchfaps$HFAA0D3=="Y"] <- "Short term care"
aric_hfchfaps$dispo[aric_hfchfaps$HFAA0D4=="Y"] <- "Home PT"
aric_hfchfaps$dispo[aric_hfchfaps$HFAA0D5=="Y"] <- "Long term care"
aric_hfchfaps$dispo[aric_hfchfaps$HFAA0D6=="Y"] <- "Hospice"
aric_hfchfaps$dispo[aric_hfchfaps$HFAA0D7=="Y"] <- "Left AMA"
aric_hfchfaps$dispo[aric_hfchfaps$HFAA0D8=="Y"] <- "Not stated"


aric_hfchfaps$adm_wt_kg[aric_hfchfaps$HFAA20A1=="K"] <- 
  aric_hfchfaps$HFAA20A[aric_hfchfaps$HFAA20A1=="K"] 

aric_hfchfaps$adm_wt_kg[aric_hfchfaps$HFAA20A1=="L"] <- 
  aric_hfchfaps$HFAA20A[aric_hfchfaps$HFAA20A1=="L"] / 2.2 


aric_hfchfaps$dc_wt_kg[aric_hfchfaps$HFAA20B1=="K"] <- 
  aric_hfchfaps$HFAA20B[aric_hfchfaps$HFAA20B1=="K"] 

aric_hfchfaps$dc_wt_kg[aric_hfchfaps$HFAA20B1=="L"] <- 
  aric_hfchfaps$HFAA20B[aric_hfchfaps$HFAA20B1=="L"] / 2.2 

aric_hfchfaps$wt_chg <- aric_hfchfaps$dc_wt_kg-aric_hfchfaps$adm_wt_kg


aric_hfchfaps$lvh_aric_grade[!aric_hfchfaps$HFAA29D1 %in% c("N","P")] <-
  aric_hfchfaps$HFAA29D1[!aric_hfchfaps$HFAA29D1 %in% c("N","P")]
aric_hfchfaps$lvh_aric_present[aric_hfchfaps$HFAA29D1 %in% c("E")] <- "No"
aric_hfchfaps$lvh_aric_present[aric_hfchfaps$HFAA29D1 %in% c("D","E","M","P","S")] <- "Yes"

aric_hfchfaps$HFAA29C1[aric_hfchfaps$ID_S0 %in% c("S111312",
                                                  "S113765",
                                                  "S114430",
                                                  "S115152",
                                                  "S116317",
                                                  "S116365",
                                                  "S117549",
                                                  "S118803",
                                                  "S120190",
                                                  "S120281",
                                                  "S122791",
                                                  "S124361",
                                                  "S125222",
                                                  "S125919",
                                                  "S127584",
                                                  "S129313",
                                                  "S130485",
                                                  "S132063",
                                                  "S135940",
                                                  "S136416",
                                                  "S136850",
                                                  "S138443",
                                                  "S138586",
                                                  "S140721",
                                                  "S141731",
                                                  "S143363",
                                                  "S143410",
                                                  "S143485",
                                                  "S144028",
                                                  "S144323",
                                                  "S144348",
                                                  "S145732",
                                                  "S146215",
                                                  "S147470",
                                                  "S148454",
                                                  "S150389",
                                                  "S152207",
                                                  "S173877",
                                                  "S185468",
                                                  "S196087",
                                                  "S196969",
                                                  "S199559",
                                                  "S203483",
                                                  "S206479")] <- 1


aric_hfchfaps$HFAA29C1[aric_hfchfaps$ID_S0 %in%  c("S112486",
                                                   "S113665",
                                                   "S114916",
                                                   "S115209",
                                                   "S124266",
                                                   "S131451",
                                                   "S132554",
                                                   "S137531",
                                                   "S140461",
                                                   "S146420",
                                                   "S150799",
                                                   "S194585",
                                                   "S204599",
                                                   "S217931",
                                                   "S218887")] <- 2


aric_hfchfaps$HFAA29C1[aric_hfchfaps$ID_S0 %in% c("S127574",
                                                  "S136289",
                                                  "S155314",
                                                  "S155682",
                                                  "S212030")] <- 3


aric_hfchfaps$ivs_cm[aric_hfchfaps$HFAA29C1==1] <-
  aric_hfchfaps$HFAA29C[aric_hfchfaps$HFAA29C1==1]

aric_hfchfaps$ivs_cm[aric_hfchfaps$HFAA29C1==2] <-
  aric_hfchfaps$HFAA29C[aric_hfchfaps$HFAA29C1==2]/10

aric_hfchfaps$ivs_cm[aric_hfchfaps$HFAA29C1==3] <-
  aric_hfchfaps$HFAA29C[aric_hfchfaps$HFAA29C1==3]/100


aric_hfchfaps$HFAA29C3[aric_hfchfaps$ID_S0 %in%  c("S111312",
                                                   "S111340",
                                                   "S113765",
                                                   "S114430",
                                                   "S114649",
                                                   "S115152",
                                                   "S116317",
                                                   "S116365",
                                                   "S116866",
                                                   "S117549",
                                                   "S117788",
                                                   "S118803",
                                                   "S119432",
                                                   "S120190",
                                                   "S120281",
                                                   "S121711",
                                                   "S122791",
                                                   "S124361",
                                                   "S124890",
                                                   "S125222",
                                                   "S125919",
                                                   "S126733",
                                                   "S127584",
                                                   "S128895",
                                                   "S129313",
                                                   "S130485",
                                                   "S132505",
                                                   "S133402",
                                                   "S134453",
                                                   "S134634",
                                                   "S135940",
                                                   "S136416",
                                                   "S136616",
                                                   "S136850",
                                                   "S138443",
                                                   "S138586",
                                                   "S140721",
                                                   "S141731",
                                                   "S142015",
                                                   "S143363",
                                                   "S143410",
                                                   "S143485",
                                                   "S144028",
                                                   "S144056",
                                                   "S144323",
                                                   "S144348",
                                                   "S145732",
                                                   "S146215",
                                                   "S147470",
                                                   "S148454",
                                                   "S148785",
                                                   "S150389",
                                                   "S152207",
                                                   "S154133",
                                                   "S154852",
                                                   "S158042",
                                                   "S169510",
                                                   "S172049",
                                                   "S181658",
                                                   "S185468",
                                                   "S185548",
                                                   "S193553",
                                                   "S201989",
                                                   "S203050",
                                                   "S203483")] <- 1


aric_hfchfaps$HFAA29C3[aric_hfchfaps$ID_S0 %in% c("S112486",
                                                  "S114916",
                                                  "S115209",
                                                  "S115893",
                                                  "S128703",
                                                  "S131451",
                                                  "S217931",
                                                  "S208111")] <- 2


aric_hfchfaps$HFAA29C3[aric_hfchfaps$ID_S0 %in% c("S127574",
                                                  "S136289",
                                                  "S155314",
                                                  "S212030")] <- 3


aric_hfchfaps$HFAA29C3[aric_hfchfaps$ID_S0 %in% c("S169510",
                                                  "S172049",
                                                  "S181658",
                                                  "S154133")] <- 4


aric_hfchfaps$lvpwd_cm[aric_hfchfaps$HFAA29C3==1] <-
  aric_hfchfaps$HFAA29C2[aric_hfchfaps$HFAA29C3==1]

aric_hfchfaps$lvpwd_cm[aric_hfchfaps$HFAA29C3==2] <-
  aric_hfchfaps$HFAA29C2[aric_hfchfaps$HFAA29C3==2]/10

aric_hfchfaps$lvpwd_cm[aric_hfchfaps$HFAA29C3==3] <-
  aric_hfchfaps$HFAA29C2[aric_hfchfaps$HFAA29C3==3]/100

aric_hfchfaps$lvpwd_cm[aric_hfchfaps$HFAA29C3==4] <-
  aric_hfchfaps$HFAA29C2[aric_hfchfaps$HFAA29C3==4] * 10


aric_hf_cohort <- merge(aric_hf_cohort, 
                        aric_hfchfaps[,c("ID_S0",
                                         "dispo",
                                         "HFAA1A",
                                         "HFAA1B",
                                         "HFAA1C",
                                         "HFAA1D",
                                         "HFAA1E",
                                         "HFAA6A",
                                         "HFAA6B",
                                         "HFAA6JNEW",
                                         "HFAA7A",
                                         "HFAA7B",
                                         "HFAA7C",
                                         "HFAA8A",
                                         "HFAA8A1",
                                         "HFAA11B1",
                                         "HFAA11B3",
                                         "HFAA11E4",
                                         "HFAA11E5",
                                         "HFAA11G",
                                         "HFAA11H",
                                         "HFAA16A",
                                         "HFAA16B",
                                         "HFAA16D",
                                         "HFAA16E",
                                         "HFAA17A",
                                         "HFAA18A",
                                         "adm_wt_kg",
                                         "dc_wt_kg",
                                         "wt_chg",
                                         "HFAA29B",
                                         "ivs_cm",
                                         "lvpwd_cm",
                                         "HFAA29D1",
                                         "HFAA29D2",
                                         "HFAA29D3",
                                         "HFAA29D4",
                                         "HFAA29D5",
                                         "HFAA29D6",
                                         "HFAA29D7",
                                         "HFAA29D8",
                                         "HFAA29D9",
                                         "HFAA29D9A",
                                         "HFAA29D10",
                                         "HFAA29D12",
                                         "HFAA29D13",
                                         "HFAA29D14",
                                         "HFAA32",
                                         "HFAA32B1",
                                         "HFAA32B2A",
                                         "HFAA32B2B",
                                         "HFAA32B2C",
                                         "HFAA32B2D",
                                         "HFAA32B2E",
                                         "HFAA32B3",
                                         "HFAA32B3A",
                                         "HFAA37A",
                                         "HFAA39A",
                                         "HFAA39B",
                                         "HFAA40A",
                                         "HFAA40B",
                                         "HFAA41A",
                                         "HFAA41B",
                                         "HFAA42A",
                                         "HFAA42B",
                                         "HFAA43A",
                                         "HFAA43B",
                                         "HFAA44A",
                                         "HFAA44B",
                                         "HFAA45A",
                                         "HFAA45B",
                                         "HFAA59",
                                         "HFAA59A",
                                         "HFAA60",
                                         "HFAA60A",
                                         "HFAA61A",
                                         "HFAA61A1",
                                         "HFAA61B",
                                         "HFAA61B1",
                                         "HFAA62",
                                         "HFAA62A",
                                         "HFAA64A",
                                         "HFAA64A1",
                                         "HFAA64B",
                                         "HFAA64B1",
                                         "HFAA65",
                                         "HFAA65A",
                                         "HFAA66",
                                         "HFAA66A",
                                         "HFAA71",
                                         "HFAA71A",
                                         "HFAA72",
                                         "HFAA72A")], 
                        by="ID_S0", 
                        all.x=T)


aric_hf_cohort[aric_hf_cohort==""] <- NA


aric_hf_cohort$adm_wt_kg[aric_hf_cohort$wt_chg < -20|aric_hf_cohort$wt_chg > 20] <- NA
aric_hf_cohort$dc_wt_kg[aric_hf_cohort$wt_chg < -20|aric_hf_cohort$wt_chg > 20] <- NA
aric_hf_cohort$wt_chg[aric_hf_cohort$wt_chg < -20|aric_hf_cohort$wt_chg > 20] <- NA

aric_hf_cohort$HFAA29D2[aric_hf_cohort$HFAA29D2=="N"] <- NA
aric_hf_cohort$HFAA29D2[aric_hf_cohort$HFAA29D2==""] <- NA

aric_hf_cohort$HFAA29D3[aric_hf_cohort$HFAA29D3=="N"] <- NA
aric_hf_cohort$HFAA29D3[aric_hf_cohort$HFAA29D3==""] <- NA

aric_hf_cohort$ef_type <- aric_hf_cohort$LVEF_CUR_LOW
aric_hf_cohort$ef_type[is.na(aric_hf_cohort$ef_type)] <-
  aric_hf_cohort$LVEF_PRE_LOW[is.na(aric_hf_cohort$ef_type)]

aric_hf_cohort$ef_val <- aric_hf_cohort$LVEF_CUR
aric_hf_cohort$ef_val[is.na(aric_hf_cohort$ef_val)] <-
  aric_hf_cohort$LVEF_PRE[is.na(aric_hf_cohort$ef_val)]


aric_hf_cohort_first <- sqldf('select * from aric_hf_cohort group by id_c having min(HFDAYS)')


aric_outcomes <- merge(aric_outcomes,
                       aric_hf_cohort[,c("ID_C",
                                         "HFDAYS",
                                         "ef_type",
                                         "ef_val",
                                         "HFAA29D2",
                                         "HFAA29B")],
                       by.x=c("ID_C",
                              "hfhosp_dt"),
                       by.y=c("ID_C",
                              "HFDAYS"),
                       all.x=T,
                       all.y=F)

aric_outcomes <- sqldf("select distinct * from aric_outcomes")

aric_outcomes$hfhosp_ef_cat[aric_outcomes$HFAA29D2=='E'] <- 'Normal'
aric_outcomes$hfhosp_ef_cat[aric_outcomes$HFAA29D2=='M'] <- 'Mildly reduced'
aric_outcomes$hfhosp_ef_cat[aric_outcomes$HFAA29D2=='D'] <- 'Moderately reduced'
aric_outcomes$hfhosp_ef_cat[aric_outcomes$HFAA29D2=='S'] <- 'Severely reduced'
aric_outcomes$hfhosp_ef_val <- aric_outcomes$HFAA29B


aric_hfccelps <- read.csv('~/Dropbox/BioLINCC files/ARIC/cohort_HF/csv/hfccelps18.csv')
aric_hfcchips <- read.csv('~/Dropbox/BioLINCC files/ARIC/cohort_HF/csv/hfcchips18.csv')
aric_hfchdxps <- read.csv('~/Dropbox/BioLINCC files/ARIC/cohort_HF/csv/hfchdxps18.csv')
aric_hfccelps <- read.csv('~/Dropbox/BioLINCC files/ARIC/cohort_HF/csv/hfccelps18.csv')


aric_hf_comm <- read.csv('~/Dropbox/BioLINCC files/aric/comm_HF/csv/hfsoccps14.csv',na.strings=c("NA","","NULL"))
aric_hf_comm_first <- sqldf('select * from aric_hf_comm group by id_c having min(HFDAYS)')


names(aric_outcomes)[1] <- "patientid"
aric_outcomes$study <- "ARIC"

aric_outcomes[,outcomes_list[!outcomes_list %in% names(aric_outcomes)]] <- NA

if (bq_table_exists("harmonization-286013.cohorts.aric_outcomes")) {
  bq_table_delete("harmonization-286013.cohorts.aric_outcomes") }

bq_table_upload(x="harmonization-286013.cohorts.aric_outcomes",
                aric_outcomes,quiet=T,
                create_disposition="CREATE_IF_NEEDED",
                write_disposition="WRITE_TRUNCATE")

write.csv(aric_outcomes,
          "~/Dropbox/ADAPT-HF/Master HDCP files/Cohort data/aric_outcomes.csv", 
          row.names = F)

gcs_auth("~/Dropbox/ADAPT-HF/Master HDCP files/harmonization-286013-39f492122f69.json")
gcs_upload(file=aric_outcomes, 
           bucket="master_hdcp_files",
           name="aric_outcomes.csv",
           object_function = f)


rm(list=ls(pattern="\\baric_hf.*$"))


#### CHS Outcomes ####


chs_events <- read.csv("~/Dropbox/BioLINCC files/CHS_2019a/EVENTS/events.csv")
chs_drhosp11 <- read.csv("~/Dropbox/BioLINCC files/CHS_2019a/ICD9/drhosp11.csv")

chs_censor <- sqldf('select distinct IDNO,
                    DEATH as dth_status,
                    CENSTIME dth_dt from chs_events')

chs_fatalevents <- sqldf("select distinct IDNO, 
      CENSTIME,
      CAUSE80,
      MECHAN80
      from chs_events
      where FATAL == 1")

chs_fatalevents$cvdth_status[chs_fatalevents$CAUSE80 %in% c(1,2,3,4)] <- 1
chs_fatalevents$noncvdth_status[chs_fatalevents$CAUSE80 %in% c(5)] <- 1
chs_fatalevents$hfdth_status[chs_fatalevents$MECHAN80 %in% c(3)] <- 1

chs_fatalevents$cvdth_status[is.na(chs_fatalevents$cvdth_status)] <- 0
chs_fatalevents$noncvdth_status[is.na(chs_fatalevents$noncvdth_status)] <- 0
chs_fatalevents$hfdth_status[is.na(chs_fatalevents$hfdth_status)] <- 0

chs_fatalevents$cvdth_dt <- chs_fatalevents$CENSTIME
chs_fatalevents$noncvdth_dt <- chs_fatalevents$CENSTIME
chs_fatalevents$hfdth_dt <- chs_fatalevents$CENSTIME


chs_incidenthf <- sqldf("select distinct IDNO, 
                        1 as hfhosp_status,
                        TTOEVENT as hfhosp_dt
                        from chs_events
                        where EVTYPE = 4
                        group by IDNO 
                        having min(hfhosp_dt)")

chs_incidentcad <- sqldf("select distinct IDNO, 
                        1 as cadhosp_status,
                        TTOEVENT as cadhosp_dt
                        from chs_events
                        where EVTYPE in (1,2,7,8,10,11)
                        group by IDNO 
                        having min(cadhosp_dt)")


chs_incidentcva <- sqldf("select distinct IDNO, 
                        1 as cvahosp_status,
                        TTOEVENT as cvahosp_dt
                        from chs_events
                        where EVTYPE in (3)
                        group by IDNO 
                        having min(cvahosp_dt)")

chs_outcomes <- merge(chs_censor,chs_fatalevents,by="IDNO",all.x=T)
chs_outcomes <- merge(chs_outcomes,chs_incidenthf,by="IDNO",all.x=T)
chs_outcomes <- merge(chs_outcomes,chs_incidentcad,by="IDNO",all.x=T)
chs_outcomes <- merge(chs_outcomes,chs_incidentcva,by="IDNO",all.x=T)

chs_outcomes$time_hfhosp_to_eos <- chs_outcomes$CENSTIME-chs_outcomes$hfhosp_dt
chs_outcomes$hfhosp_status[is.na(chs_outcomes$hfhosp_status)] <- 0
chs_outcomes$hfhosp_dt[chs_outcomes$hfhosp_status==0] <- chs_outcomes$CENSTIME[chs_outcomes$hfhosp_status==0]

chs_outcomes$time_cad_to_eos <- chs_outcomes$CENSTIME - chs_outcomes$cadhosp_dt
chs_outcomes$cadhosp_status[is.na(chs_outcomes$cadhosp_status)] <- 0
chs_outcomes$cadhosp_dt[chs_outcomes$cadhosp_status==0] <- chs_outcomes$CENSTIME[chs_outcomes$cadhosp_status==0]


chs_outcomes$study <- "CHS"

names(chs_outcomes)[1] <- "patientid"
outcomes_to_fill <- outcomes_list[!outcomes_list %in% names(chs_outcomes)]
outcomes_to_fill <- outcomes_to_fill[!is.na(outcomes_to_fill)]
chs_outcomes[,outcomes_to_fill] <- NA


if (bq_table_exists("harmonization-286013.cohorts.chs_outcomes")) {
  bq_table_delete("harmonization-286013.cohorts.chs_outcomes") }

bq_table_upload(x="harmonization-286013.cohorts.chs_outcomes",
                chs_outcomes,quiet=T,
                create_disposition="CREATE_IF_NEEDED",
                write_disposition="WRITE_TRUNCATE")

write.csv(chs_outcomes,
          "~/Dropbox/ADAPT-HF/Master HDCP files/Cohort data/chs_outcomes.csv",
          row.names=F,
          na="")


gcs_auth("~/Dropbox/ADAPT-HF/Master HDCP files/harmonization-286013-39f492122f69.json")
gcs_upload(file=chs_outcomes, 
           bucket="master_hdcp_files",
           name="chs_outcomes.csv",
           object_function = f)


#### MESA Outcomes ####


mesa_hfevents <- read.csv("~/Dropbox/BioLINCC files/MESA/Primary/Events/CVD/Data/mesaevefthru2015_drepos_20200330.csv")
mesa_afevents <- read.csv("~/Dropbox/BioLINCC files/MESA/Primary/Events/CVD/Data/mesaevaffu10_drepos_2015416.csv")
mesa_cvevents <- read.csv("~/Dropbox/BioLINCC files/MESA/Primary/Events/CVD/Data/mesaevthr2015_drepos_20200330.csv")
mesa_noncvevents <- read.csv("~/Dropbox/BioLINCC files/MESA/Primary/Events/NonCVD/Data/mesaevnoncvddres06192012.csv")

mesa_cvevents$dth_status <- mesa_cvevents$DTH
mesa_cvevents$dth_dt <- mesa_cvevents$DTHTT

mesa_cvevents$cvdth_status[mesa_cvevents$DTHTYPE %in% c(1,2,3,4)] <- 1
mesa_cvevents$cvdth_status[is.na(mesa_cvevents$cvdth_status)] <- 0
mesa_cvevents$cvdth_dt <- mesa_cvevents$DTHTT

mesa_cvevents$noncvdth_status[mesa_cvevents$DTHTYPE %in% c(5, 
                                                           9
)] <- 1
mesa_cvevents$noncvdth_status[is.na(mesa_cvevents$noncvdth_status)] <- 0
mesa_cvevents$noncvdth_dt <- mesa_cvevents$DTHTT

mesa_cvevents$cvhosp_status <- mesa_cvevents$CVDA
mesa_cvevents$cvhosp_dt <- mesa_cvevents$CVDATT

mesa_cvevents$hfhosp_status <- mesa_cvevents$CHF
mesa_cvevents$hfhosp_dt <- mesa_cvevents$CHFTT

mesa_cvevents$cadhosp_status <- mesa_cvevents$CHDA
mesa_cvevents$cadhosp_dt <- mesa_cvevents$CHDATT

mesa_noncvevents$noncvhosp_dt <- apply(mesa_noncvevents[,c(2,
                                                           4,
                                                           6,
                                                           8,
                                                           10,
                                                           12,
                                                           14,
                                                           16,
                                                           18,
                                                           20,
                                                           22,
                                                           24,
                                                           26,
                                                           28,
                                                           30)],
                                       MARGIN=1,
                                       FUN=min,
                                       na.rm=T)


mesa_noncvevents$noncvhosp_status[mesa_noncvevents$noncvhosp_dt==mesa_noncvevents$fuptt] <- 0
mesa_noncvevents$noncvhosp_status[!mesa_noncvevents$noncvhosp_dt==mesa_noncvevents$fuptt] <- 1


mesa_outcomes <- merge(
  mesa_cvevents[,c("MESAID",
                   "dth_status","dth_dt",
                   "cvdth_status","cvdth_dt",
                   "noncvdth_status","noncvdth_dt",
                   "cvhosp_status","cvhosp_dt",
                   "cadhosp_status","cadhosp_dt",
                   "hfhosp_status","hfhosp_dt")],
  mesa_noncvevents[,c("MESAID",
                      "noncvhosp_status",
                      "noncvhosp_dt")],
  by="MESAID",
  all.x=T
)

mesa_outcomes <- merge(mesa_outcomes,
                       mesa_hfevents[,c("MESAID","TTCHF","EFMEAS","EFCLASS")],
                       by.x=c("MESAID","hfhosp_dt"),
                       by.y=c("MESAID","TTCHF"),
                       all.x=T)

mesa_outcomes$hfhosp_ef_val <- mesa_outcomes$EFMEAS

mesa_outcomes$hfhosp_ef_cat[mesa_outcomes$hfhosp_ef_val <= 35] <- "Severely reduced"
mesa_outcomes$hfhosp_ef_cat[mesa_outcomes$hfhosp_ef_val > 35&
                              mesa_outcomes$hfhosp_ef_val <= 45] <- "Moderately reduced"
mesa_outcomes$hfhosp_ef_cat[mesa_outcomes$hfhosp_ef_val > 45&
                              mesa_outcomes$hfhosp_ef_val <= 55] <- "Mildly reduced"

mesa_outcomes$hfhosp_ef_cat[mesa_outcomes$hfhosp_ef_val > 55] <- "Normal"

mesa_outcomes$hosp_status <- 0
mesa_outcomes$hosp_dt <- mesa_outcomes$dth_dt
mesa_outcomes$hosp_status[mesa_outcomes$cvhosp_status==1|
                            mesa_outcomes$noncvhosp_status==1] <- 1

mesa_outcomes$hosp_dt[mesa_outcomes$hosp_status==1] <-
  min(mesa_outcomes$cvhosp_dt[mesa_outcomes$hosp_status==1],
      mesa_outcomes$noncvhosp_dt[mesa_outcomes$hosp_status==1])

mesa_outcomes$study <- "MESA"

names(mesa_outcomes)[1] <- "patientid"
outcomes_to_fill <- outcomes_list[!outcomes_list %in% names(mesa_outcomes)]
outcomes_to_fill <- outcomes_to_fill[!is.na(outcomes_to_fill)]
mesa_outcomes[,outcomes_to_fill] <- NA

write.csv(mesa_outcomes,
          '~/Dropbox/ADAPT-HF/Master HDCP files/Cohort data/mesa_outcomes.csv',
          row.names=F,
          na="")

if (bq_table_exists("harmonization-286013.cohorts.mesa_outcomes")) {
  bq_table_delete("harmonization-286013.cohorts.mesa_outcomes") }


bq_table_upload(x="harmonization-286013.cohorts.mesa_outcomes",
                mesa_outcomes,quiet=T,
                create_disposition="CREATE_IF_NEEDED",
                write_disposition="WRITE_TRUNCATE")

gcs_auth("~/Dropbox/ADAPT-HF/Master HDCP files/harmonization-286013-39f492122f69.json")
gcs_upload(file=mesa_outcomes, 
           bucket="master_hdcp_files",
           name="mesa_outcomes.csv",
           object_function = f)


#### JHS Outcomes ####


jhs_allevtchd <- read.csv("~/Dropbox/BioLINCC files/JHS/Data/Events/csv/allevtchd.csv")
jhs_allevthf <- read.csv("~/Dropbox/BioLINCC files/JHS/Data/Events/csv/allevthf.csv")
jhs_allevtstroke <- read.csv("~/Dropbox/BioLINCC files/JHS/Data/Events/csv/allevtstroke.csv")
jhs_incevtchd <- read.csv("~/Dropbox/BioLINCC files/JHS/Data/Events/csv/incevtchd.csv")
jhs_incevthfder <- read.csv("~/Dropbox/BioLINCC files/JHS/Data/Events/csv/incevthfder.csv")
jhs_incevtstroke <- read.csv("~/Dropbox/BioLINCC files/JHS/Data/Events/csv/incevtstroke.csv")

jhs_allevtchd <- merge(jhs_allevtchd,
                       jhs_incevtchd[,c("newid","V1date")],
                       by="newid")

jhs_allevtchd$days_to_event <- as.numeric(as.Date(jhs_allevtchd$eventDate, format="%m/%d/%Y") - 
                                            as.Date(jhs_allevtchd$V1date, 
                                                    format="%m/%d/%Y"))

jhs_incevthfder$HF[jhs_incevthfder$examdate==""] <- 0
jhs_incevthfder$days[jhs_incevthfder$examdate==""] <- 0
jhs_incevthfder$examdate[jhs_incevthfder$examdate==""] <- 
  jhs_incevthfder$V1date[jhs_incevthfder$examdate==""]

jhs_allevthf <- merge(jhs_allevthf,
                      jhs_incevthfder[,c("newid","examdate")],
                      by="newid")

jhs_allevthf$days_to_event <- as.numeric(as.Date(jhs_allevthf$eventDate, format="%m/%d/%Y") - 
                                           as.Date(jhs_allevthf$examdate, format="%m/%d/%Y"))

jhs_allevtstroke <- merge(jhs_allevtstroke,
                          jhs_incevthfder[,c("newid","V1date")],
                          by="newid")

jhs_allevtstroke$days_to_event <- as.numeric(as.Date(jhs_allevtstroke$eventdate, format="%m/%d/%Y") - 
                                               as.Date(jhs_allevtstroke$V1date, format="%m/%d/%Y"))

jhs_outcomes_hf <- sqldf("select newid,
                      days as hfhosp_dt,
                      HF as hfhosp_status
                      from jhs_incevthfder")

jhs_outcomes_cad <- sqldf("select newid,
                      days as cadhosp_dt,
                      CHD as cadhosp_status
                      from jhs_incevtchd")

jhs_outcomes <- merge(jhs_outcomes_hf,
                      jhs_outcomes_cad,
                      by="newid",
                      all.x=T)


jhs_outcomes$study <- "JHS"


names(jhs_outcomes)[1] <- "patientid"
outcomes_to_fill <- outcomes_list[!outcomes_list %in% names(jhs_outcomes)]
outcomes_to_fill <- outcomes_to_fill[!is.na(outcomes_to_fill)]
jhs_outcomes[,outcomes_to_fill] <- NA


outcomes_list_2 <- c("patientid","study",outcomes_list)

write.csv(jhs_outcomes,
          "~/Dropbox/ADAPT-HF/Master HDCP files/Cohort data/jhs_outcomes.csv",
          row.names=F,
          na="")

if (bq_table_exists("harmonization-286013.cohorts.jhs_outcomes")) {
  bq_table_delete("harmonization-286013.cohorts.jhs_outcomes") }

bq_table_upload(x="harmonization-286013.cohorts.jhs_outcomes",
                jhs_outcomes,quiet=T,
                create_disposition="CREATE_IF_NEEDED",
                write_disposition="WRITE_TRUNCATE")

gcs_auth("~/Dropbox/ADAPT-HF/Master HDCP files/harmonization-286013-39f492122f69.json")
gcs_upload(file=jhs_outcomes, 
           bucket="master_hdcp_files",
           name="jhs_outcomes.csv",
           object_function = f)

all_outcomes <- 
  rbind(aric_outcomes[,outcomes_list_2],
        fhs_outcomes[,outcomes_list_2],
        chs_outcomes[,outcomes_list_2],
        jhs_outcomes[,outcomes_list_2],
        mesa_outcomes[,outcomes_list_2])

write.csv(all_outcomes,
          "~/Dropbox/ADAPT-HF/Master HDCP files/Cohort data/all_outcomes.csv",
          row.names=F,
          na="")

if (bq_table_exists("harmonization-286013.cohorts.outcomes")) {
  bq_table_delete("harmonization-286013.cohorts.outcomes") }

bq_table_upload(x="harmonization-286013.cohorts.outcomes",
                all_outcomes,quiet=T,
                create_disposition="CREATE_IF_NEEDED",
                write_disposition="WRITE_TRUNCATE")

gcs_auth("~/Dropbox/ADAPT-HF/Master HDCP files/harmonization-286013-39f492122f69.json")
gcs_upload(file=all_outcomes, 
           bucket="master_hdcp_files",
           name="all_outcomes.csv",
           object_function = f)
```
