## Supplementary material for "Aggregating data to accelerate personalized therapy in heart failure (ADAPT-HF)": Harmonization scripts: Compile ARIC.R

```
library(Hmisc)
library(plotrix)
library(descr)
library(dplyr)
library(relper)
library(stringr)
library(bigrquery)
library(haven)
library(googleCloudStorageR)
library(data.table)
library(arrow)
library(parquetize)
library(bigQueryR)

source("~/Dropbox/R scripts/Misc clinical scripts.R")
source("~/Dropbox/R scripts/echo scripts.R")

f <- function(input,output) {
  write.csv(input,file=output, row.names=F, na="")
}

visit_yrs <- fread('~/Dropbox/ADAPT-HF/Master HDCP files/Cohort visit yrs.csv')

data_fields <- 
  c("study",
    "patientid",
    "visit_yr",
    "visitdays",
    "form",
    "variable",
    "value",
    "datapoint",
    "study_field",
    "cohort",
    "cohort_name",
    "age_obs")

outcomes_list <- c("dth_status","dth_dt",
                   "cvdth_status","cvdth_dt",
                   "hfdth_status","hfdth_dt",
                   "hfhosp_ef_val","hfhosp_ef_cat",
                   "noncvdth_status","noncvdth_dt",
                   "hosp_status","hosp_dt",
                   "cadhosp_status","cadhosp_dt",
                   "cvhosp_status","cvhosp_dt",
                   "hfhosp_status","hfhosp_dt",
                   "noncvhosp_status","noncvhosp_dt")

dates_long_fields <-
  c("study",
    "cohort",
    "cohort_name",
    "patientid",
    "sex",
    "visit_yr",
    "visitdays",
    "age_obs")


#### ************************ ARIC ************************ #### 

# ....%%%....%%%%%%%%..%%%%..%%%%%%.
# ...%%.%%...%%.....%%..%%..%%....%%
# ..%%...%%..%%.....%%..%%..%%......
# .%%.....%%.%%%%%%%%...%%..%%......
# .%%%%%%%%%.%%...%%....%%..%%......
# .%%.....%%.%%....%%...%%..%%....%%
# .%%.....%%.%%.....%%.%%%%..%%%%%%.


#===============================================================##
####                      ARIC dates                          ####
#===============================================================##

aric_derive1 <- fread('~/Dropbox/BioLINCC files/ARIC/Main_Study/v1/csv/derive13.csv',na.strings=c("NA","","NULL"))
aric_derive2 <- fread('~/Dropbox/BioLINCC files/ARIC/Main_Study/v2/csv/derive2_10.csv',na.strings=c("NA","","NULL"))
aric_derive3 <- fread('~/Dropbox/BioLINCC files/ARIC/Main_Study/v3/csv/derive37.csv',na.strings=c("NA","","NULL"))
aric_derive4 <- fread('~/Dropbox/BioLINCC files/ARIC/Main_Study/v4/csv/derive47.csv',na.strings=c("NA","","NULL"))
aric_derive5 <- fread('~/Dropbox/BioLINCC files/ARIC/Main_Study/v5/csv/derive51.csv',na.strings=c("NA","","NULL"))
aric_derive6 <- fread('~/Dropbox/BioLINCC files/ARIC/Main_Study/V6/csv/derive61.csv',na.strings=c("NA","","NULL"))
aric_derive7 <- fread('~/Dropbox/BioLINCC files/ARIC/Main_Study/V7/csv/derive71.csv',na.strings=c("NA","","NULL"))
aric_derive8 <- fread('~/Dropbox/BioLINCC files/ARIC/Main_Study/V8T/csv/derive8t1_np.csv',na.strings=c("NA","","NULL"))

aric_dates <- merge(aric_derive1[,c("ID_C","CENTERID","ENROLL_YR","PREVHF01","V1AGE01")],
                    aric_derive2[,c("ID_C","V2DAYS","V2AGE22")],all.x=T)
aric_dates <- merge(aric_dates,aric_derive3[,c("ID_C","V3DAYS","V3AGE31")],all.x=T)
aric_dates <- merge(aric_dates,aric_derive4[,c("ID_C","V4DAYS","V4AGE41")],all.x=T)
aric_dates <- merge(aric_dates,aric_derive5[,c("ID_C","V5DATE51_DAYS","V5AGE51")],all.x=T)
aric_dates <- merge(aric_dates,aric_derive6[,c("ID_C","V6DATE61_DAYS","V6AGE61")],all.x=T)
aric_dates <- merge(aric_dates,aric_derive7[,c("ID_C","V7DATE71_FOLLOWUPDAYS","V7AGE72")],all.x=T)
aric_dates <- merge(aric_dates,aric_derive8[,c("ID_C","V8TDATE8T1_FOLLOWUPDAYS","V8TAGE8T2")],all.x=T)

aric_dates$cohort <- aric_dates$CENTERID

aric_dates$cohort_name[aric_dates$cohort=="A"] <- "MN"
aric_dates$cohort_name[aric_dates$cohort=="B"] <- "MD"
aric_dates$cohort_name[aric_dates$cohort=="C"] <- "MI"
aric_dates$cohort_name[aric_dates$cohort=="D"] <- "NC"


# aric_dates1_long <- as.data.table(sqldf("select ID_C,
#                           1 as visit,
#                           0 as visitdays,
#                           V1AGE01 as age_obs
#                           from aric_derive1"))

aric_dates1_long <- 
  aric_derive1[
  ,
  .(
    ID_C,
    visit     = 1L,
    visitdays = 0L,
    age_obs   = V1AGE01
  )
]

aric_dates2_long <- 
  aric_derive2[,.(ID_C,
                  visit = 2L,
                  visitdays = V2DAYS,
                  age_obs = V2AGE22)]

aric_dates3_long <- 
  aric_derive3[,.(ID_C,
                  visit = 3L,
                  visitdays = V3DAYS,
                  age_obs = V3AGE31)]

aric_dates4_long <- 
  aric_derive4[,.(ID_C,
                  visit = 4L,
                  visitdays = V4DAYS,
                  age_obs = V4AGE41)]

aric_dates5_long <- 
  aric_derive5[,.(ID_C,
                  visit = 5L,
                  visitdays = V5DATE51_DAYS,
                  age_obs = V5AGE51)]

aric_dates6_long <- 
  aric_derive6[,.(ID_C,
                  visit = 6L,
                  visitdays = V6DATE61_DAYS,
                  age_obs = V6AGE61)]

aric_dates7_long <- 
  aric_derive7[,.(ID_C,
                  visit = 7L,
                  visitdays = V7DATE71_FOLLOWUPDAYS,
                  age_obs = V7AGE72)]

aric_dates8_long <- 
  aric_derive8[,.(ID_C,
                  visit = 8L,
                  visitdays = V8TDATE8T1_FOLLOWUPDAYS,
                  age_obs = V8TAGE8T2)]


aric_dates_long <- rbindlist(
  list(aric_dates1_long,
       aric_dates2_long,
       aric_dates3_long,
       aric_dates4_long,
       aric_dates5_long,
       aric_dates6_long,
       aric_dates7_long,
       aric_dates8_long))


rm(aric_dates1_long,
   aric_dates2_long,
   aric_dates3_long,
   aric_dates4_long,
   aric_dates5_long,
   aric_dates6_long,
   aric_dates7_long,
   aric_dates8_long)


aric_dates_long <- 
  aric_dates_long[
    aric_derive1[,c("ID_C","GENDER", "CENTERID")],
    on=.(ID_C)]


aric_dates_long <- 
  visit_yrs[
    study=="ARIC"&
      cohort=="A",][aric_dates_long,
                    on=.(visit)]

aric_dates_long[,cohort := CENTERID]

aric_dates_long[cohort=="A", cohort_name := "MN"]
aric_dates_long[cohort=="B", cohort_name := "MD"]
aric_dates_long[cohort=="C", cohort_name := "MI"]
aric_dates_long[cohort=="D", cohort_name := "NC"]

aric_dates_long$study <- "ARIC"


# ==============================================================#
####                      ARIC Outcomes                      ####
# ==============================================================#


### Cohort

# Incident events, all outcomes
aric_incps <- fread('~/Dropbox/BioLINCC files/ARIC/Main_Study/cohort_incident_recurrent/csv/incps19.csv',na.strings=c("NA","","NULL"))


aric_hfcchips <- fread('~/Dropbox/BioLINCC files/ARIC/Main_Study/cohort_HF/csv/hfcchips19.csv',na.strings=c("NA","","NULL"))

# Cohort event eligibility
aric_ccelps19 <- fread('~/Dropbox/BioLINCC files/ARIC/Main_Study/cohort_HF/csv/hfccelps19.csv',na.strings=c("NA","","NULL"))


aric_outcomes <- 
  unique(aric_incps[,.(
    ID_C, 
    hf_after_exam5_status =C7_INCHF_P_V5, 
    hf_after_exam5_dys = C7_FT_INCHF_P_V5,
    cadhosp_status = ISP19,
    cadhosp_dt = C7_FUTIMEA,
    baseline_chd  = PRVCHD05,
    hfhosp_status = INCHF19, 
    hfhosp_dt = C7_FUTIMEHF,
    baseline_hf = PREVHF01,
    dth_status = DEAD19, 
    dth_dt = FUDTH19,
    dth_cause = UCOD)])

### CV death was defined as death with ICD‐9 code 401‐459 or ICD‐10 code I10‐I99 
# PMC4710457
# PMC6343542
# PMC8109763

aric_outcomes[,cvdth_status := 0]
aric_outcomes[between(as.numeric(substr(dth_cause,1,3)),401,459)|
                             substr(dth_cause,1,1)=="I",cvdth_status :=  1]

aric_outcomes[cvdth_status==1|dth_status==1,noncvdth_status := 0]
aric_outcomes[cvdth_status==0&dth_status==1, noncvdth_status := 1]

aric_outcomes[,cvdth_dt := dth_dt]
aric_outcomes[,noncvdth_dt := dth_dt]

aric_outcomes[baseline_hf==1&!is.na(baseline_hf), hfhosp_status := 1]
aric_outcomes[baseline_hf==1&!is.na(baseline_hf), hfhosp_dt := 0]


aric_hf_cohort <- # Cohort HF occurences
  fread('~/Dropbox/BioLINCC files/ARIC/Main_Study/cohort_HF/csv/hfcoccps19.csv',na.strings=c("NA","","NULL"))
aric_hf_cohort[,visit_yr := floor(HFDAYS/365)]

aric_hfchfaps <- # 
  fread('~/Dropbox/BioLINCC files/ARIC/Main_Study/cohort_HF/csv/hfchfaps19.csv',na.strings=c("NA","","NULL"))

aric_hf_cohort <- 
  aric_hf_cohort[
        aric_outcomes[,c("ID_C","cvdth_status","noncvdth_status")],
        on=.(ID_C)]

aric_hfchfaps[HFAA0D=="D", dispo := "Deceased"]
aric_hfchfaps[HFAA0D1=="Y", dispo := "Home"]
aric_hfchfaps[HFAA0D2=="Y", dispo := "Home"]
aric_hfchfaps[HFAA0D3=="Y", dispo := "Short term care"]
aric_hfchfaps[HFAA0D4=="Y", dispo := "Home"]
aric_hfchfaps[HFAA0D5=="Y", dispo := "Long term care"]
aric_hfchfaps[HFAA0D6=="Y", dispo := "Hospice"]
aric_hfchfaps[HFAA0D7=="Y", dispo:= "Left AMA"]

aric_hfchfaps[HFAA20A1=="K"&!is.na(HFAA20A1), adm_wt_kg := HFAA20A] 

aric_hfchfaps[HFAA20A1=="L"&!is.na(HFAA20A1), adm_wt_kg := HFAA20A/2.2]  

aric_hfchfaps[HFAA20B1=="K"&!is.na(HFAA20B1), dc_wt_kg := HFAA20B] 

aric_hfchfaps[HFAA20B1=="L"&!is.na(HFAA20B1), dc_wt_kg := HFAA20B / 2.2] 

aric_hfchfaps[,wt_chg := dc_wt_kg-adm_wt_kg]

aric_hfchfaps[,wt_chg_pct := (dc_wt_kg-adm_wt_kg)/adm_wt_kg]


aric_hfchfaps[!HFAA29D1 %in% c("N","P"), lvh_aric_grade := HFAA29D1]

aric_hfchfaps[HFAA29D1 %in% c("E"), lvh_aric_present := "No"]
aric_hfchfaps[HFAA29D1 %in% c("D","E","M","P","S"), lvh_aric_present := "Yes"]

aric_hfchfaps[ID_S0 %in% c("S111312",
                           "S113765",
                           "S114430",
                           "S115152",
                           "S116317",
                           "S116365",
                           "S117549",
                           "S118803",
                           "S120190",
                           "S120281",
                           "S122791",
                           "S124361",
                           "S125222",
                           "S125919",
                           "S127584",
                           "S129313",
                           "S130485",
                           "S132063",
                           "S135940",
                           "S136416",
                           "S136850",
                           "S138443",
                           "S138586",
                           "S140721",
                           "S141731",
                           "S143363",
                           "S143410",
                           "S143485",
                           "S144028",
                           "S144323",
                           "S144348",
                           "S145732",
                           "S146215",
                           "S147470",
                           "S148454",
                           "S150389",
                           "S152207",
                           "S173877",
                           "S185468",
                           "S196087",
                           "S196969",
                           "S199559",
                           "S203483",
                           "S206479"),
              HFAA29C1 := 1]


aric_hfchfaps[ID_S0 %in%  c("S112486",
                            "S113665",
                            "S114916",
                            "S115209",
                            "S124266",
                            "S131451",
                            "S132554",
                            "S137531",
                            "S140461",
                            "S146420",
                            "S150799",
                            "S194585",
                            "S204599",
                            "S217931",
                            "S218887"),
              HFAA29C1 := 2]


aric_hfchfaps[ID_S0 %in% 
                c("S127574",
                  "S136289",
                  "S155314",
                  "S155682",
                  "S212030"), 
              HFAA29C1 := 3]


aric_hfchfaps[HFAA29C1==1&!is.na(HFAA29C1), ivs_cm := HFAA29C]

aric_hfchfaps[HFAA29C1==2&!is.na(HFAA29C1), ivs_cm := HFAA29C/10]

aric_hfchfaps[HFAA29C1==3&!is.na(HFAA29C1), ivs_cm := HFAA29C/100]


aric_hfchfaps[ID_S0 %in%  
                c("S111312",
                  "S111340",
                  "S113765",
                  "S114430",
                  "S114649",
                  "S115152",
                  "S116317",
                  "S116365",
                  "S116866",
                  "S117549",
                  "S117788",
                  "S118803",
                  "S119432",
                  "S120190",
                  "S120281",
                  "S121711",
                  "S122791",
                  "S124361",
                  "S124890",
                  "S125222",
                  "S125919",
                  "S126733",
                  "S127584",
                  "S128895",
                  "S129313",
                  "S130485",
                  "S132505",
                  "S133402",
                  "S134453",
                  "S134634",
                  "S135940",
                  "S136416",
                  "S136616",
                  "S136850",
                  "S138443",
                  "S138586",
                  "S140721",
                  "S141731",
                  "S142015",
                  "S143363",
                  "S143410",
                  "S143485",
                  "S144028",
                  "S144056",
                  "S144323",
                  "S144348",
                  "S145732",
                  "S146215",
                  "S147470",
                  "S148454",
                  "S148785",
                  "S150389",
                  "S152207",
                  "S154133",
                  "S154852",
                  "S158042",
                  "S169510",
                  "S172049",
                  "S181658",
                  "S185468",
                  "S185548",
                  "S193553",
                  "S201989",
                  "S203050",
                  "S203483"), 
              HFAA29C3 := 1]


aric_hfchfaps[ID_S0 %in% 
                c("S112486",
                  "S114916",
                  "S115209",
                  "S115893",
                  "S128703",
                  "S131451",
                  "S217931",
                  "S208111"), 
              HFAA29C3 := 2]


aric_hfchfaps[ID_S0 %in% 
                c("S127574",
                  "S136289",
                  "S155314",
                  "S212030"), 
              HFAA29C3 := 3]


aric_hfchfaps[ID_S0 %in% 
                c("S169510",
                  "S172049",
                  "S181658",
                  "S154133"), 
              HFAA29C3 := 4]


aric_hfchfaps[HFAA29C3==1&!is.na(HFAA29C3), lvpwd_cm := HFAA29C2]

aric_hfchfaps[HFAA29C3==2&!is.na(HFAA29C3), lvpwd_cm := HFAA29C2/10]

aric_hfchfaps[HFAA29C3==3&!is.na(HFAA29C3), lvpwd_cm := HFAA29C2/100]

aric_hfchfaps[HFAA29C3==4&!is.na(HFAA29C3), lvpwd_cm := HFAA29C2 * 10]

aric_hf_cohort <-
    aric_hfchfaps[,c("ID_S0",
                     "dispo",
                     "HFAA1A",
                     "HFAA1B",
                     "HFAA1C",
                     "HFAA1D",
                     "HFAA1E",
                     "HFAA6A",
                     "HFAA6B",
                     "HFAA6JNEW",
                     "HFAA7A",
                     "HFAA7B",
                     "HFAA7C",
                     "HFAA8A",
                     "HFAA8A1",
                     "HFAA11B1",
                     "HFAA11B3",
                     "HFAA11E4",
                     "HFAA11E5",
                     "HFAA11G",
                     "HFAA11H",
                     "HFAA16A",
                     "HFAA16B",
                     "HFAA16D",
                     "HFAA16E",
                     "HFAA17A",
                     "HFAA18A",
                     "adm_wt_kg",
                     "dc_wt_kg",
                     "wt_chg",
                     "wt_chg_pct",
                     "HFAA29B",
                     "ivs_cm",
                     "lvpwd_cm",
                     "HFAA29D1",
                     "HFAA29D2",
                     "HFAA29D3",
                     "HFAA29D4",
                     "HFAA29D5",
                     "HFAA29D6",
                     "HFAA29D7",
                     "HFAA29D8",
                     "HFAA29D9",
                     "HFAA29D9A",
                     "HFAA29D10",
                     "HFAA29D12",
                     "HFAA29D13",
                     "HFAA29D14",
                     "HFAA32",
                     "HFAA32B1",
                     "HFAA32B2A",
                     "HFAA32B2B",
                     "HFAA32B2C",
                     "HFAA32B2D",
                     "HFAA32B2E",
                     "HFAA32B3",
                     "HFAA32B3A",
                     "HFAA37A",
                     "HFAA39A",
                     "HFAA39B",
                     "HFAA40A",
                     "HFAA40B",
                     "HFAA41A",
                     "HFAA41B",
                     "HFAA42A",
                     "HFAA42B",
                     "HFAA43A",
                     "HFAA43B",
                     "HFAA44A",
                     "HFAA44B",
                     "HFAA45A",
                     "HFAA45B",
                     "HFAA59",
                     "HFAA59A",
                     "HFAA60",
                     "HFAA60A",
                     "HFAA61A",
                     "HFAA61A1",
                     "HFAA61B",
                     "HFAA61B1",
                     "HFAA62",
                     "HFAA62A",
                     "HFAA64A",
                     "HFAA64A1",
                     "HFAA64B",
                     "HFAA64B1",
                     "HFAA65",
                     "HFAA65A",
                     "HFAA66",
                     "HFAA66A",
                     "HFAA71",
                     "HFAA71A",
                     "HFAA72",
                     "HFAA72A")][aric_hf_cohort, 
    on="ID_S0"]

aric_hf_cohort[wt_chg < -20|wt_chg > 20, adm_wt_kg := NA]
aric_hf_cohort[wt_chg < -20|wt_chg > 20, dc_wt_kg := NA]
aric_hf_cohort[wt_chg < -20|wt_chg > 20, wt_chg := NA]

aric_hf_cohort[HFAA29D2 %in% c("N",""), HFAA29D2 := NA]

aric_hf_cohort[HFAA29D3 %in% c("N",""), HFAA29D3 := NA]

aric_hf_cohort[,ef_type := LVEF_CUR_LOW]
aric_hf_cohort[is.na(ef_type), ef_type := LVEF_PRE_LOW]

aric_hf_cohort[,ef_val := LVEF_CUR]
aric_hf_cohort[is.na(ef_val), ef_val := LVEF_PRE]

aric_hf_cohort_first <- 
  aric_hf_cohort[
  order(HFDAYS)][, .SD[1], by = ID_C]

aric_hf_cohort$hfhosp_dt <- aric_hf_cohort$HFDAYS


  
aric_outcomes <-
    aric_hf_cohort[,.(ID_C,
                      hfhosp_dt,
                      ef_type,
                      ef_val,
                      HFAA29D2,
                      HFAA29B)][aric_outcomes,
    on = .(ID_C, hfhosp_dt)]


aric_outcomes[HFAA29D2=='E', hfhosp_ef_cat := 'Normal']
aric_outcomes[HFAA29D2=='M', hfhosp_ef_cat := 'Mildly reduced']
aric_outcomes[HFAA29D2=='D', hfhosp_ef_cat := 'Moderately reduced']
aric_outcomes[HFAA29D2=='S', hfhosp_ef_cat := 'Severely reduced']
aric_outcomes[,hfhosp_ef_val := HFAA29B]


aric_hfccelps <- fread('~/Dropbox/BioLINCC files/ARIC/Main_study/cohort_HF/csv/hfccelps19.csv')
names(aric_outcomes)[1] <- "patientid"
aric_outcomes[,study := "ARIC"]

aric_outcomes <- aric_outcomes[!patientid==1,]

write_parquet(aric_outcomes,
              "~/Dropbox/ADAPT-HF/Master HDCP files/Cohort data/aric_outcomes.parquet")

#### Community ####

aric_hf_comm <- fread('~/Dropbox/BioLINCC files/aric/Main_Study/comm_HF/csv/hfsoccps14.csv',na.strings=c("NA","","NULL"))
aric_hf_comm_first <- aric_hf_comm[
  order(HFDAYS)
][
  , .SD[1], by = ID_C
]
aric_hf_comm[,visit_yr := floor(HFDAYS/365)]


aric_sdthps <- fread('~/Dropbox/BioLINCC files/ARIC/Main_Study/comm_CHD/csv/sdthps14.csv',na.strings=c("NA","","NULL"))
aric_hfschips <- fread('~/Dropbox/BioLINCC files/ARIC/Main_Study/comm_HF/csv/hfschips14.csv',na.strings=c("NA","","NULL"))

aric_sdthps[
  substr(DTH2Z18,1,1) %in% c("A","B","C","D"), Type := "CV"] 

aric_sdthps[
  substr(DTH2Z18,1,1) %in% c("E","F","G","K","L"), Type := "Non-CV"] 


aric_hfshfaps <- fread('~/Dropbox/BioLINCC files/ARIC/Main_Study/comm_HF/csv/hfshfaps14.csv',na.strings=c("NA","","NULL"))

aric_hfshfaps[HFAA0D=="D", dispo := "Deceased"]
aric_hfshfaps[HFAA0D1=="Y", dispo := "Home"]
aric_hfshfaps[HFAA0D2=="Y", dispo := "Home"]
aric_hfshfaps[HFAA0D3=="Y", dispo := "Short term care"]
aric_hfshfaps[HFAA0D4=="Y", dispo := "Home"]
aric_hfshfaps[HFAA0D5=="Y", dispo := "Long term care"]
aric_hfshfaps[HFAA0D6=="Y", dispo := "Hospice"]
aric_hfshfaps[HFAA0D7=="Y", dispo := "Left AMA"]

aric_hfshfaps[HFAA20A1=="K"&!is.na(HFAA20A1), 
              adm_wt_kg := HFAA20A]
              
aric_hfshfaps[HFAA20A1=="L"&!is.na(HFAA20A1), 
              adm_wt_kg := HFAA20A / 2.2] 


aric_hfshfaps[HFAA20B1=="K"&!is.na(HFAA20B1), 
              dc_wt_kg := HFAA20B] 

aric_hfshfaps[HFAA20B1=="L"&!is.na(HFAA20B1), 
              dc_wt_kg := HFAA20B / 2.2] 

aric_hfshfaps[,wt_chg := dc_wt_kg-adm_wt_kg]

aric_hfshfaps[,wt_chg_pct := wt_chg/adm_wt_kg]

aric_hfshfaps[!HFAA29D1 %in% c("N","P"), 
               lvh_aric_grade := HFAA29D1]

aric_hfshfaps[HFAA29D1 %in% c("E"), lvh_aric_present := "No"]
aric_hfshfaps[HFAA29D1 %in% c("D","E","M","P","S"), lvh_aric_present := "Yes"]


## Fixing unit errors
## Units codes: 1=cm, 2=mm

aric_hfshfaps[
  ID_S0 %in% aric_hfshfaps[ 
    HFAA29C<5&
      HFAA29C1==2, .(ID_S0)], 
  HFAA29C1 := 1]

aric_hfshfaps[
  ID_S0 %in% aric_hfshfaps[HFAA29C<5&HFAA29C1==1, .(ID_S0)], 
  HFAA29C1 := 2]

aric_hfshfaps[HFAA29C1==1&!is.na(HFAA29C1),
              ivs_cm := HFAA29C1]

aric_hfshfaps[HFAA29C1==2&!is.na(HFAA29C1), 
              ivs_cm := HFAA29C1/10]

aric_hfshfaps[HFAA29C1==3&!is.na(HFAA29C1),
  ivs_cm := HFAA29C1/100]


aric_hfshfaps[ID_S0 %in% 
    aric_hfshfaps[HFAA29C2 <5&HFAA29C1==2,.(ID_S0)], 
    HFAA29C3 := 1]

aric_hfshfaps[ID_S0 %in% 
                aric_hfshfaps[HFAA29C2 <5&HFAA29C1==1,.(ID_S0)], 
              HFAA29C3 := 2]


aric_hfshfaps[HFAA29C3==1&!is.na(HFAA29C3), 
  lvpwd_cm := HFAA29C2]

aric_hfshfaps[HFAA29C3==2&!is.na(HFAA29C3), 
              lvpwd_cm := HFAA29C2/10]

aric_hf_comm <-
  aric_hfshfaps[,c("ID_S0",
                   "dispo",
                   "HFAA1A",
                   "HFAA1B",
                   "HFAA1C",
                   "HFAA1D",
                   "HFAA1E",
                   "HFAA6A",
                   "HFAA6B",
                   "HFAA6JNEW",
                   "HFAA7A",
                   "HFAA7B",
                   "HFAA7C",
                   "HFAA8A",
                   "HFAA8A1",
                   "HFAA11B1",
                   "HFAA11B3",
                   "HFAA11E4",
                   "HFAA11E5",
                   "HFAA11G",
                   "HFAA11H",
                   "HFAA16A",
                   "HFAA16B",
                   "HFAA16D",
                   "HFAA16E",
                   "HFAA17A",
                   "HFAA18A",
                   "adm_wt_kg",
                   "dc_wt_kg",
                   "wt_chg",
                   "wt_chg_pct",
                   "HFAA29B",
                   "ivs_cm",
                   "lvpwd_cm",
                   "HFAA29D1",
                   "HFAA29D2",
                   "HFAA29D3",
                   "HFAA29D4",
                   "HFAA29D5",
                   "HFAA29D6",
                   "HFAA29D7",
                   "HFAA29D8",
                   "HFAA29D9",
                   "HFAA29D9A",
                   "HFAA29D10",
                   "HFAA29D12",
                   "HFAA29D13",
                   "HFAA29D14",
                   "HFAA32",
                   "HFAA32B1",
                   "HFAA32B2A",
                   "HFAA32B2B",
                   "HFAA32B2C",
                   "HFAA32B2D",
                   "HFAA32B2E",
                   "HFAA32B3",
                   "HFAA32B3A",
                   "HFAA37A",
                   "HFAA39A",
                   "HFAA39B",
                   "HFAA40A",
                   "HFAA40B",
                   "HFAA41A",
                   "HFAA41B",
                   "HFAA42A",
                   "HFAA42B",
                   "HFAA43A",
                   "HFAA43B",
                   "HFAA44A",
                   "HFAA44B",
                   "HFAA45A",
                   "HFAA45B",
                   "HFAA59",
                   "HFAA59A",
                   "HFAA60",
                   "HFAA60A",
                   "HFAA61A",
                   "HFAA61A1",
                   "HFAA61B",
                   "HFAA61B1",
                   "HFAA62",
                   "HFAA62A",
                   "HFAA64A",
                   "HFAA64A1",
                   "HFAA64B",
                   "HFAA64B1",
                   "HFAA65",
                   "HFAA65A",
                   "HFAA66",
                   "HFAA66A",
                   "HFAA71",
                   "HFAA71A",
                   "HFAA72",
                   "HFAA72A")]

aric_hf_comm[wt_chg < -20|wt_chg > 20, adm_wt_kg := NA]
aric_hf_comm[wt_chg < -20|wt_chg > 20, dc_wt_kg :=  NA]
aric_hf_comm[wt_chg < -20|wt_chg > 20, wt_chg := NA]

aric_hf_comm[HFAA29D2=="N", HFAA29D2 := NA]
aric_hf_comm[HFAA29D2=="", HFAA29D2 := NA]

aric_hf_comm[HFAA29D3=="N", HFAA29D3 := NA]
aric_hf_comm[HFAA29D3=="", HFAA29D3 := NA]

# aric_hf_comm[,ef_type := LVEF_CUR_LOW]
# aric_hf_comm[is.na(ef_type)], ef_type := LVEF_PRE_LOW]
# 
# aric_hf_comm[,ef_val := LVEF_CUR]
# aric_hf_comm[is.na(ef_val)], ef_val := LVEF_PRE]


aric_hf_comm[HFAA29D2=='E', hfhosp_ef_cat := 'Normal']
aric_hf_comm[HFAA29D2=='M', hfhosp_ef_cat := 'Mildly reduced']
aric_hf_comm[HFAA29D2=='D', hfhosp_ef_cat := 'Moderately reduced']
aric_hf_comm[HFAA29D2=='S', hfhosp_ef_cat := 'Severely reduced']
aric_hf_comm[,hfhosp_ef_val := HFAA29B]

write_parquet(aric_hf_comm,
              "~/Dropbox/ADAPT-HF/Master HDCP files/Cohort data/aric_hf_comm.parquet")

rm(list=ls(pattern="\\baric_hf.*$"))


#===============================================================#
####             Semi-annual follow-up forms                 ####
#===============================================================#


### GEN

aric_genps1 <- fread('~/Dropbox/BioLINCC files/ARIC/Main_Study/AFU/csv/genps1.csv',
                     na.strings=c("NA","","NULL"))

aric_melt_genps1 <- melt(aric_genps1,
                         id.vars=c("ID_C",
                                   "GEN_FUDAYS",
                                   "CONTYR"),
                         na.rm=T,
                         stringsAsFactors=T)
aric_melt_genps1[,form := "genps"]
names(aric_melt_genps1)[1:3] <- c("patientid","visitdays","visit_yr")

### GNB

aric_gnbps1 <- fread('~/Dropbox/BioLINCC files/ARIC/Main_Study/AFU/csv/gnbps1.csv',
                     na.strings=c("NA","","NULL"))
aric_gnbps1[,CONTYR := substr(EVENTNAME,3,4)]

aric_melt_gnbps1 <- melt(aric_gnbps1[,-c(3,38,40)],
                         id.vars=c("ID_C",
                                   "GNB_FUDAYS",
                                   "CONTYR"),
                         na.rm=T,
                         stringsAsFactors=T)
aric_melt_gnbps1[,form := "gnbps"]
names(aric_melt_gnbps1)[1:3] <- c("patientid","visitdays","visit_yr")


### GNC

aric_gncps1 <- fread('~/Dropbox/BioLINCC files/ARIC/Main_Study/AFU/csv/gncps1.csv',
                     na.strings=c("NA","","NULL"))
aric_gncps1[,CONTYR := substr(EVENTNAME,3,4)]


aric_gncps1[,gh1 := GNC1]
aric_gncps1[,pf02 := GNC2A]
aric_gncps1[,pf04 := GNC2B]
aric_gncps1[GNC3A %in% c(1,2,3),rp2 := 1]
aric_gncps1[GNC3A %in% c(4,5), rp2 := 0] 

aric_gncps1[GNC3B %in% c(1,2,3), rp3 := 1]
aric_gncps1[GNC3B %in% c(4,5), rp3:= 0]

aric_gncps1[GNC4A %in% c(1,2,3), re2 := 1]
aric_gncps1[GNC4A %in% c(4,5), re2 := 0]

aric_gncps1[GNC4B %in% c(1,2,3), re3 := 1]
aric_gncps1[GNC4B %in% c(4,5), re3 := 0]

aric_gncps1[,bp2 := GNC5]

aric_gncps1[,mh3 := GNC6A]
aric_gncps1[,vt2 := GNC6B]
aric_gncps1[,mh4 := GNC6C]

aric_gncps1[,sf2 := GNC7]

# aric_gncps1 <- cbind(aric_gncps1,sf12(aric_gncps1[,30:41]))

aric_melt_gncps1 <- melt(aric_gncps1[,-c(3,28)],
                         id.vars=c("ID_C",
                                   "GNC_FUDAYS",
                                   "CONTYR"),
                         na.rm=T,
                         stringsAsFactors=T)
aric_melt_gncps1[,form := "gncps"]
names(aric_melt_gncps1)[1:3] <- c("patientid","visitdays","visit_yr")


### GND


aric_gndps1 <- fread('~/Dropbox/BioLINCC files/ARIC/Main_Study/AFU/csv/gndps1.csv',
                     na.strings=c("NA","","NULL"))

aric_gndps1[,CONTYR := substr(EVENTNAME,4,5)]

aric_melt_gndps1 <- 
  melt(aric_gndps1[,-c(2,25)],
       id.vars=c("ID_C",
                 "GND_FUDAYS",
                 "CONTYR"),
       na.rm=T,
       stringsAsFactors=T)


aric_melt_gndps1[,form := "gndps"]
names(aric_melt_gndps1)[1:3] <- c("patientid","visitdays","visit_yr")


### GNE


aric_gneps1 <- fread('~/Dropbox/BioLINCC files/ARIC/Main_Study/AFU/csv/gneps1.csv',
                     na.strings=c("NA","","NULL"))
aric_gneps1[,CONTYR := substr(EVENTNAME,4,5)]

aric_gneps1[,forgetful := 0]

aric_gneps1[GNE13==1|
              GNE14==1|
              GNE15==1|
              GNE16==1|
              GNE17==1|
              GNE18==1|
              GNE19==1|
              GNE20==1|
              GNE21==1|
              GNE22==1, forgetful := 1]


aric_melt_gneps1 <- 
  melt(aric_gneps1[,-c(2,32)],
       id.vars=c("ID_C",
                 "GNE_FUDAYS",
                 "CONTYR"),
       na.rm=T,
       stringsAsFactors=T)

aric_melt_gneps1[,form := "gneps"]
names(aric_melt_gneps1)[1:3] <- c("patientid","visitdays","visit_yr")


### Assemble GN

aric_melt_gnps <- rbindlist(
  list(aric_melt_genps1,
       aric_melt_gnbps1,
       aric_melt_gncps1,
       aric_melt_gndps1,
       aric_melt_gneps1)
)

rm(list=ls(pattern="\\baric_melt_g.*\\ps1$"))

#### ARIC analytes, all visits ####


aric_analytes <- fread('~/Dropbox/BioLINCC files/ARIC/Main_Study/Longitudinal/csv/v1_v5_analytes.csv',na.strings=c("NA","","NULL"))

aric_analytes_long <- 
  melt(aric_analytes,
       id.vars=c("ID_C"),
       na.rm=T,
       factorsAsStrings = T)

aric_analytes_long[,variable := as.character(variable)]

aric_analytes_long[substring(variable,nchar(variable))==6, visit := 6]
aric_analytes_long[substring(variable,nchar(variable))==5, visit := 5]
aric_analytes_long[substring(variable,nchar(variable))==4, visit := 4]
aric_analytes_long[substring(variable,nchar(variable))==3, visit := 3]
aric_analytes_long[substring(variable,nchar(variable))==2, visit := 2]
aric_analytes_long[substring(variable,nchar(variable))==1, visit := 1]

analyte_points <- names(aric_analytes)[grep("_V",names(aric_analytes))]

aric_analytes_long[
  variable %in% analyte_points, 
  variable := 
    substring(variable,1, 
              str_locate(variable[variable %in% analyte_points],"_")[,"start"]-1)]

aric_analytes_long[
  variable=="PRO", 
  variable := "PRO_BNP"]

aric_analytes_long <-
  aric_analytes_long[!variable=="FORPROFIT"]

visit_yrs <- 
  visit_yrs[study=="ARIC"&cohort=="A",.(visit,visit_yr)]

aric_analytes_long <-
  visit_yrs[aric_analytes_long,on=.(visit)]

aric_analytes_long[,form := "analytes"]
setnames(aric_analytes_long,"visit_yr","CONTYR")
aric_analytes_long[,visitdays := CONTYR * 365]

#===============================================================#
####                       ARIC Exam 1                       ####
#===============================================================#

aric_derive1[,drinks_wk  := ETHANL03/14]

aric_anta_v1 <- fread('~/Dropbox/BioLINCC files/ARIC/Main_Study/V1/csv/anta.csv',na.strings=c("NA","","NULL"))
aric_dtia_v1 <- fread('~/Dropbox/BioLINCC files/ARIC/Main_Study/V1/csv/dtia.csv',na.strings=c("NA","","NULL"))

aric_anta_v1[,weight_kg :=  as.numeric(ANTA04)/2.2046226218]

aric_ecgma03_v1 <- fread('~/Dropbox/BioLINCC files/ARIC/Main_Study/V1/csv/ecgma03.csv',na.strings=c("NA","","NULL"))

aric_rpaa_v1 <- fread('~/Dropbox/BioLINCC files/ARIC/Main_Study/V1/csv/rpaa02.csv',na.strings=c("NA","","NULL"))
aric_anut2_v1 <- fread('~/Dropbox/BioLINCC files/ARIC/Main_Study/V1/csv/anut2.csv',na.strings=c("NA","","NULL"))
aric_totnutx_v1 <- fread('~/Dropbox/BioLINCC files/ARIC/Main_Study/V1/csv/totnutx.csv',na.strings=c("NA","","NULL"))
aric_atrfib11_v1 <- fread('~/Dropbox/BioLINCC files/ARIC/Main_Study/V1/csv/atrfib11.csv',na.strings=c("NA","","NULL"))
aric_chma_v1 <- fread('~/Dropbox/BioLINCC files/ARIC/Main_Study/V1/csv/chma.csv',na.strings=c("NA","","NULL"))
aric_sbpa02_v1 <- fread('~/Dropbox/BioLINCC files/ARIC/Main_Study/V1/csv/sbpa02.csv',na.strings=c("NA","","NULL"))
aric_atrfib11_v1[,AFIBFL := 0]
aric_atrfib11_v1[AF==1|AFL==1, AFIBFL := 1]
aric_ftra02_v1 <- fread('~/Dropbox/BioLINCC files/ARIC/Main_Study/V1/csv/ftra02.csv',na.strings=c("NA","","NULL"))
aric_hom_v1 <- fread('~/Dropbox/BioLINCC files/ARIC/Main_Study/V1/csv/hom.csv',na.strings=c("NA","","NULL"))
aric_msra_v1 <- fread('~/Dropbox/BioLINCC files/ARIC/Main_Study/V1/csv/msra.csv',na.strings=c("NA","","NULL"))
aric_msrcod07_v1 <- fread('~/Dropbox/BioLINCC files/ARIC/Main_Study/V1/csv/msrcod07.csv',na.strings=c("NA","","NULL"))
aric_mhxa_v1 <- fread('~/Dropbox/BioLINCC files/ARIC/Main_Study/V1/csv/mhxa02.csv',na.strings=c("NA","","NULL"))
aric_hmta_v1 <- fread('~/Dropbox/BioLINCC files/ARIC/Main_Study/V1/csv/hmta.csv',na.strings=c("NA","","NULL"))
aric_pulm_v1_ref <- fread('~/Dropbox/BioLINCC files/ARIC/Main_Study/V1/csv/pulm.csv',na.strings=c("NA","","NULL"))
aric_pfta_v1 <- fread('~/Dropbox/BioLINCC files/ARIC/Main_Study/V1/csv/pfta.csv',na.strings=c("NA","","NULL"))
aric_phea_v1 <- fread('~/Dropbox/BioLINCC files/ARIC/Main_Study/V1/csv/phea.csv',na.strings=c("NA","","NULL"))
aric_sbpa_v1 <- fread('~/Dropbox/BioLINCC files/ARIC/Main_Study/V1/csv/sbpa02.csv',na.strings=c("NA","","NULL"))

aric_pfta_ref_v1 <- aric_pulm_v1_ref[aric_pfta_v1, on="ID_C"]
aric_pfta_ref_v1[, FEV1PP := PFTA26/FEV_101]


aric_hom_v1[HOM54<=12&!is.na(HOM54), ed_years :=  HOM54]
aric_hom_v1[HOM54==13, ed_years := 12]
aric_hom_v1[HOM54==14|HOM54==17, ed_years := 13] 
aric_hom_v1[HOM54==15|HOM54==18, ed_years := 14]
aric_hom_v1[HOM54==16|HOM54==19, ed_years := 15]
aric_hom_v1[HOM54==20, ed_years := 16]
aric_hom_v1[HOM54==21, ed_years := 20]

aric_lipa_v1 <- fread('~/Dropbox/BioLINCC files/ARIC/Main_Study/V1/csv/lipa.csv',na.strings=c("NA","","NULL"))
aric_rhxa_v1 <- fread('~/Dropbox/BioLINCC files/ARIC/Main_Study/V1/csv/rhxa.csv',na.strings=c("NA","","NULL"), colClasses = "character")


string_to_replace <- "A"
columns_to_modify <- names(aric_rhxa_v1)[4:52]

aric_rhxa_v1 <- aric_rhxa_v1 %>%
  mutate(across(all_of(columns_to_modify), 
                ~ na_if(., string_to_replace)))

aric_rhxa_v1[,RHXA01 := as.numeric(RHXA01)]

aric_rhxa_v1[,RHXA08 := as.numeric(RHXA08)]

aric_rhxa_v1[,RHXA15 := as.numeric(RHXA15)]

aric_rhxa_v1[,RHXA22 := as.numeric(RHXA22)]

aric_rhxa_v1[,RHXA29 := as.numeric(RHXA29)]

aric_rhxa_v1[,RHXA36 := as.numeric(RHXA36)]

aric_rhxa_v1[,RHXA43 := as.numeric(RHXA43)]


aric_rhxa_v1[,hrt_agent1_yrs := RHXA22]

aric_rhxa_v1[,hrt_agent2_yrs := RHXA29]

aric_rhxa_v1[,hrt_agent3_yrs := RHXA36]

aric_rhxa_v1[,hrt_agent4_yrs := RHXA43]

aric_rhxa_v1[,years_on_ocp := RHXA15]

aric_hrt_yrs <-
  aric_rhxa_v1[,c("ID_C",
                  "hrt_agent1_yrs",
                  "hrt_agent2_yrs",
                  "hrt_agent3_yrs",
                  "hrt_agent4_yrs")]


aric_msrcod07_v1 <- fread('~/Dropbox/BioLINCC files/ARIC/Main_Study/V1/csv/msrcod07.csv',na.strings=c("NA","","NULL"))
aric_msrcod07_codes <- aric_msrcod07_v1[,c(1,19:35)]

aric_melt_msrcod07_codes <- melt(aric_msrcod07_codes,id.vars="ID_C",na.rm=T,factorsAsStrings=T)
aric_melt_msrcod07_codes[,variable := "medcode"]
aric_melt_msrcod07_codes[,form := "msrcod"]
aric_melt_msrcod07_codes <-  unique(aric_melt_msrcod07_codes)

aric_melt_derive1_v1 <- melt(aric_derive1,id.vars=c("ID_C"),na.rm=T,factorsAsStrings=T)
aric_melt_derive1_v1[,form := "derive1"]
aric_melt_anta_v1 <- melt(aric_anta_v1,id.vars=c("ID_C"),na.rm=T,factorsAsStrings=T)
aric_melt_anta_v1[,form := "anta"]
aric_melt_anut2_v1 <- melt(aric_anut2_v1,id.vars=c("ID_C"),na.rm=T,factorsAsStrings=T)
aric_melt_anut2_v1[,form := "anut2"]
aric_melt_totnux_v1 <- melt(aric_totnutx_v1,id.vars=c("ID_C"),na.rm=T,factorsAsStrings=T)
aric_melt_totnux_v1[,form := "totnux"]
aric_melt_dtia_v1 <- melt(aric_dtia_v1,id.vars=c("ID_C"),na.rm=T,factorsAsStrings=T)
aric_melt_dtia_v1[,form := "dtia"]
aric_melt_ecgma03_v1 <- melt(aric_ecgma03_v1,id.vars=c("ID_C"),na.rm=T,factorsAsStrings=T)
aric_melt_ecgma03_v1[,form := "ecgma03"]
aric_melt_rpaa_v1 <- melt(aric_rpaa_v1,id.vars=c("ID_C"),na.rm=T,factorsAsStrings=T)
aric_melt_rpaa_v1[,form := "rpaa"]
aric_melt_atrfib11_v1 <- melt(aric_atrfib11_v1,id.vars=c("ID_C"),na.rm=T,factorsAsStrings=T)
aric_melt_atrfib11_v1[,form := "atrfib11"]
aric_melt_chma_v1 <- melt(aric_chma_v1,id.vars=c("ID_C"),na.rm=T,factorsAsStrings=T)
aric_melt_chma_v1[,form := "chma"]
aric_melt_hmta_v1 <- melt(aric_hmta_v1,id.vars=c("ID_C"),na.rm=T,factorsAsStrings=T)
aric_melt_hmta_v1[,form := "hmta"]
aric_melt_mhxa_v1 <- melt(aric_mhxa_v1,id.vars=c("ID_C"),na.rm=T,factorsAsStrings=T)
aric_melt_mhxa_v1[,form := "mhxa"]
aric_melt_pfta_ref_v1 <- melt(aric_pfta_ref_v1,id.vars=c("ID_C"),na.rm=T,factorsAsStrings=T)
aric_melt_pfta_ref_v1[,form := "pfta"]
aric_melt_phea_v1 <- melt(aric_phea_v1,id.vars=c("ID_C"),na.rm=T,factorsAsStrings=T)
aric_melt_phea_v1[,form := "phea"]
aric_melt_ftra02_v1 <- melt(aric_ftra02_v1,id.vars=c("ID_C"),na.rm=T,factorsAsStrings=T)
aric_melt_ftra02_v1[,form := "ftra"]
aric_melt_hom_v1 <- melt(aric_hom_v1,id.vars=c("ID_C"),na.rm=T,factorsAsStrings=T)
aric_melt_hom_v1[,form := "hom"]
aric_melt_lipa_v1 <- melt(aric_lipa_v1,id.vars=c("ID_C"),na.rm=T,factorsAsStrings=T)
aric_melt_lipa_v1[,form := "lipa"]
aric_melt_pulm_v1 <- melt(aric_pulm_v1_ref,id.vars=c("ID_C"),na.rm=T,factorsAsStrings=T)
aric_melt_pulm_v1[,form := "pulm"]
aric_melt_rhxa_v1 <- melt(aric_rhxa_v1,id.vars=c("ID_C"),na.rm=T,factorsAsStrings=T)
aric_melt_rhxa_v1[,form := "rhxa"]
aric_melt_sbpa_v1 <- melt(aric_sbpa02_v1,id.vars=c("ID_C"),na.rm=T,factorsAsStrings=T)
aric_melt_sbpa_v1[,form := "sbpa"]


aric_melt_v1_all <- 
  rbindlist(
    list(
  aric_melt_derive1_v1,
  aric_melt_anta_v1,
  aric_melt_anut2_v1,
  aric_melt_atrfib11_v1,
  aric_melt_chma_v1,
  aric_melt_dtia_v1,
  aric_melt_ecgma03_v1,
  aric_melt_ftra02_v1,
  aric_melt_hmta_v1,
  aric_melt_lipa_v1,
  aric_melt_hom_v1,
  aric_melt_mhxa_v1,
  aric_melt_pfta_ref_v1,
  aric_melt_phea_v1,
  aric_melt_pulm_v1,
  aric_melt_rhxa_v1,
  aric_melt_rpaa_v1,
  aric_melt_sbpa_v1,
  aric_melt_totnux_v1))


aric_melt_v1_all[,visit := 1]
aric_melt_v1_all[,visitdays := 0]
aric_melt_v1_all[,CONTYR := 0]

rm(list=ls(pattern="\\baric_.*\\_v1$"))


#===============================================================#
####                       ARIC Exam 2                       ####
#===============================================================#


aric_derive2[,drinks_wk := ETHANL24/14]

aric_antb_v2 <- fread('~/Dropbox/BioLINCC files/ARIC/Main_Study/V2/csv/antb.csv',na.strings=c("NA","","NULL"))
aric_antb_v2[,weight_kg := as.numeric(ANTB01)/2.2046226218]

aric_ecgmb22_v2 <- fread('~/Dropbox/BioLINCC files/ARIC/Main_Study/V2/csv/ecgmb22.csv',na.strings=c("NA","","NULL"))
aric_fhxa_v2 <- fread('~/Dropbox/BioLINCC files/ARIC/Main_Study/V2/csv/fhxa.csv',na.strings=c("NA","","NULL"))
aric_chmb_v2 <- fread('~/Dropbox/BioLINCC files/ARIC/Main_Study/V2/csv/chmb.csv',na.strings=c("NA","","NULL"))
aric_dtib_v2 <- fread('~/Dropbox/BioLINCC files/ARIC/Main_Study/V2/csv/dtib.csv',na.strings=c("NA","","NULL"))
aric_hhxb_v2 <- fread('~/Dropbox/BioLINCC files/ARIC/Main_Study/V2/csv/hhxb.csv',na.strings=c("NA","","NULL"),colClasses = "character")

aric_hhxb_v2[HHXB01A=="A", HHXB01A := NA]
aric_hhxb_v2[HHXB01B=="A", HHXB01B := NA]
aric_hhxb_v2[,last_doc_months := 
               as.numeric(HHXB01A)*12+
               as.numeric(HHXB01B)]

string_to_replace <- "A"
columns_to_modify <- names(aric_hhxb_v2)[4:96]

aric_hhxb_v2 <- 
  aric_hhxb_v2 %>%
  mutate(across(all_of(columns_to_modify), ~ na_if(., string_to_replace)))

aric_hhxb_v2[,hrt_agent5_yrs :=
  as.numeric(HHXB28A) +
  as.numeric(HHXB28B)/12]


aric_hhxb_v2[,hrt_agent6_yrs :=
  as.numeric(HHXB35A) +
  as.numeric(HHXB35B)/12]

aric_hrt_v2 <- aric_hhxb_v2[, .(ID_C, hrt_agent5_yrs, hrt_agent6_yrs)]

aric_hrt_yrs <- 
    aric_hrt_v2[aric_hrt_yrs,
    on = .(ID_C)
  ]


aric_hmtb_v2 <- fread('~/Dropbox/BioLINCC files/ARIC/Main_Study/V2/csv/hmtb.csv',na.strings=c("NA","","NULL"))
aric_hpaa_v2 <- fread('~/Dropbox/BioLINCC files/ARIC/Main_Study/V2/csv/hpaa.csv',na.strings=c("NA","","NULL"))
aric_hpba_v2 <- fread('~/Dropbox/BioLINCC files/ARIC/Main_Study/V2/csv/hpba.csv',na.strings=c("NA","","NULL"))


for (i in 3:23) {
  set(aric_hpba_v2, which(aric_hpba_v2[[i]] == "Y" & !is.na(aric_hpba_v2[[i]])), i, 2)
  set(aric_hpba_v2, which(aric_hpba_v2[[i]] == "D" & !is.na(aric_hpba_v2[[i]])), i, 1)
  set(aric_hpba_v2, which(aric_hpba_v2[[i]] == "N" & !is.na(aric_hpba_v2[[i]])), i, 0)
}  

aric_hpba_v2[,maas_vit_exhaustm :=
  apply(aric_hpba_v2[,3:23],
        MARGIN=1,
        FUN=function(x) sum(as.numeric(x)
                            )
        )
  ]

aric_hpca_v2 <- fread('~/Dropbox/BioLINCC files/ARIC/Main_Study/V2/csv/hpca.csv',na.strings=c("NA","","NULL"))

for (i in 4:13) {
  set(aric_hpca_v2, which(aric_hpca_v2[[i]]=="A"&!is.na(aric_hpca_v2[[i]])),i,1)
  set(aric_hpca_v2, which(aric_hpca_v2[[i]]=="B"&!is.na(aric_hpca_v2[[i]])),i,2)
  set(aric_hpca_v2, which(aric_hpca_v2[[i]]=="C"&!is.na(aric_hpca_v2[[i]])),i,3)
  set(aric_hpca_v2, which(aric_hpca_v2[[i]]=="D"&!is.na(aric_hpca_v2[[i]])),i,4) 
  set(aric_hpca_v2, which(aric_hpca_v2[[i]]=="E"&!is.na(aric_hpca_v2[[i]])),i,NA)
}  

aric_hpca_v2[,anger_spielberger :=
  apply(aric_hpca_v2[,4:13],
        MARGIN=1,
        FUN=function(x) sum(as.numeric(x)))]

aric_lipb_v2 <- fread('~/Dropbox/BioLINCC files/ARIC/Main_Study/V2/csv/lipb.csv',na.strings=c("NA","","NULL"))
aric_nutv2_v2 <- fread('~/Dropbox/BioLINCC files/ARIC/Main_Study/V2/csv/nutv2.csv',na.strings=c("NA","","NULL"))
aric_pheb_v2 <- fread('~/Dropbox/BioLINCC files/ARIC/Main_Study/V2/csv/pheb.csv',na.strings=c("NA","","NULL"))
aric_sbpb_v2 <- fread('~/Dropbox/BioLINCC files/ARIC/Main_Study/V2/csv/sbpb02.csv',na.strings=c("NA","","NULL"))
aric_atrfib21_v2 <- fread('~/Dropbox/BioLINCC files/ARIC/Main_Study/V2/csv/atrfib21.csv',na.strings=c("NA","","NULL"))
aric_atrfib21_v2[,AFIBFL := 0]
aric_atrfib21_v2[AF==1|AFL==1, AFIBFL := 1]

aric_pftb_v2 <- fread('~/Dropbox/BioLINCC files/ARIC/Main_Study/V2/csv/pftb.csv',na.strings=c("NA","","NULL"))
aric_pftb_v2_ref <- aric_pulm_v1_ref[aric_pftb_v2, on=.(ID_C)]
aric_pftb_v2_ref[,FEV1PP := PFTB26/FEV_101]

#### Hypertension


#### Diabetes


#### COPD


#### Asthma


#### Arthritis


#### CKD


#### CHF


aric_derive2 <- 
aric_outcomes[, .(patientid, hfhosp_status, hfhosp_dt)][aric_derive2,
    on = .(patientid = ID_C)
  ]


aric_derive2[!is.na(hfhosp_status), chf_frail := 0]
aric_derive2[!is.na(hfhosp_status)&
                         hfhosp_status==1&
                         hfhosp_dt <= V2DAYS, 
                       chf_frail := 1]

#### CAD


#### Stroke


#### Cancer

aric_msrcod26_v2 <- fread('~/Dropbox/BioLINCC files/ARIC/Main_Study/V2/csv/msrcod26.csv',na.strings=c("NA","","NULL"))
aric_msrcod26_codes <- aric_msrcod26_v2[,c(1,19:35)]

aric_melt_msrcod26_codes <- 
  melt(aric_msrcod26_codes,
       id.vars="ID_C",
       na.rm=T,
       factorsAsStrings=T)

aric_melt_msrcod26_codes[,variable := "medcode"]
aric_melt_msrcod26_codes[,form := "msrcod"]
aric_melt_msrcod26_codes <- unique(aric_melt_msrcod26_codes)

names(aric_derive2)[names(aric_derive2)=="patientid"] <- "ID_C"

aric_melt_derive2_v2 <- melt(aric_derive2,id.vars=c("ID_C"),na.rm=T,factorsAsStrings=T)
aric_melt_derive2_v2[, form := "derive2"]
aric_melt_antb_v2 <- melt(aric_antb_v2,id.vars=c("ID_C"),na.rm=T,factorsAsStrings=T)
aric_melt_antb_v2[, form := "antb"]
aric_melt_chmb_v2 <- melt(aric_chmb_v2,id.vars=c("ID_C"),na.rm=T,factorsAsStrings=T)
aric_melt_chmb_v2[,form := "chmb"]
aric_melt_dtib_v2 <- melt(aric_dtib_v2,id.vars=c("ID_C"),na.rm=T,factorsAsStrings=T)
aric_melt_dtib_v2[, form := "dtib"]
aric_melt_ecgmb22_v2 <- melt(aric_ecgmb22_v2,id.vars=c("ID_C"),na.rm=T,factorsAsStrings=T)
aric_melt_ecgmb22_v2[,form := "ecgmb22"]
aric_melt_fhxa_v2 <- melt(aric_fhxa_v2,id.vars=c("ID_C"),na.rm=T,factorsAsStrings=T)
aric_melt_fhxa_v2[, form := "fhxa"]
aric_melt_hhxb_v2 <- melt(aric_hhxb_v2,id.vars=c("ID_C"),na.rm=T,factorsAsStrings=T)
aric_melt_hhxb_v2[,form := "hhxb"]
aric_melt_hmtb_v2 <- melt(aric_hmtb_v2,id.vars=c("ID_C"),na.rm=T,factorsAsStrings=T)
aric_melt_hmtb_v2[,form := "hmtb"]
aric_melt_hpaa_v2 <- melt(aric_hpaa_v2,id.vars=c("ID_C"),na.rm=T,factorsAsStrings=T)
aric_melt_hpaa_v2[,form := "hpaa"]
aric_melt_hpba_v2 <- melt(aric_hpba_v2,id.vars=c("ID_C"),na.rm=T,factorsAsStrings=T)
aric_melt_hpba_v2[,form := "hpba"]
aric_melt_hpca_v2 <- melt(aric_hpca_v2,id.vars=c("ID_C"),na.rm=T,factorsAsStrings=T)
aric_melt_hpca_v2[,form := "hpca"]
aric_melt_lipb_v2 <- melt(aric_lipb_v2,id.vars=c("ID_C"),na.rm=T,factorsAsStrings=T)
aric_melt_lipb_v2[,form := "lipb"]
aric_melt_nutv2_v2 <- melt(aric_nutv2_v2,id.vars=c("ID_C"),na.rm=T,factorsAsStrings=T)
aric_melt_nutv2_v2[,form := "nutv2"]
aric_melt_pftb_ref_v2 <- melt(aric_pftb_v2_ref,id.vars=c("ID_C"),na.rm=T,factorsAsStrings=T)
aric_melt_pftb_ref_v2[,form := "pftb"]
aric_melt_pheb_v2 <- melt(aric_pheb_v2,id.vars=c("ID_C"),na.rm=T,factorsAsStrings=T)
aric_melt_pheb_v2[,form := "pheb"]

aric_melt_atrfib21_v2 <- melt(aric_atrfib21_v2,id.vars=c("ID_C"),na.rm=T,factorsAsStrings=T)
aric_melt_atrfib21_v2[,form := "atrfib21"]
aric_melt_sbpb_v2 <- melt(aric_sbpb_v2,id.vars=c("ID_C"),na.rm=T,factorsAsStrings=T)
aric_melt_sbpb_v2[,form := "sbpb"]

aric_melt_v2_all <- 
  rbindlist(
    list(aric_melt_derive2_v2,
         aric_melt_antb_v2,
         aric_melt_chmb_v2,
         aric_melt_dtib_v2,
         aric_melt_ecgmb22_v2,
         aric_melt_fhxa_v2,
         aric_melt_hhxb_v2,
         aric_melt_hmtb_v2,
         aric_melt_hpaa_v2,
         aric_melt_hpba_v2,
         aric_melt_hpca_v2,
         aric_melt_lipb_v2,
         aric_melt_nutv2_v2,
         aric_melt_pheb_v2,
         aric_melt_pftb_ref_v2,
         aric_melt_sbpb_v2,
         aric_melt_atrfib21_v2
    )
  )

aric_v2_tempdays <- 
  aric_dates[!is.na(V2DAYS)
             ,
             .(ID_C,
               visitdays = V2DAYS)
  ]


aric_melt_v2_all[,visit := 2]

aric_melt_v2_all <- 
  aric_v2_tempdays[aric_melt_v2_all,
                          on=.(ID_C)]

aric_melt_v2_all[,CONTYR := 4]

rm(list=ls(pattern="\\baric_.*\\_v2$"))


#===============================================================#
####                       ARIC Exam 3                       ####
#===============================================================#

aric_derive3 <- fread('~/Dropbox/BioLINCC files/ARIC/Main_Study/v3/csv/derive37.csv',na.strings=c("NA","","NULL"))

aric_derive3[,drinks_wk := ETHANL32/14]

aric_antc04_v3 <- fread('~/Dropbox/BioLINCC files/ARIC/Main_Study/V3/csv/antc04.csv',na.strings=c("NA","","NULL"))
aric_amha02_v3 <- fread('~/Dropbox/BioLINCC files/ARIC/Main_Study/V3/csv/amha02.csv',na.strings=c("NA","","NULL"))
aric_antc04_v3$weight_kg <- as.numeric(aric_antc04_v3$ANTC2)/2.2046226218
aric_atrfib31_v3 <- fread('~/Dropbox/BioLINCC files/ARIC/Main_Study/V3/csv/atrfib31.csv',na.strings=c("NA","","NULL"))
aric_atrfib31_v3$AFIBFL <- 0
aric_atrfib31_v3$AFIBFL[aric_atrfib31_v3$AF==1|aric_atrfib31_v3$AFL==1] <- 1

aric_dtic04_v3 <- fread('~/Dropbox/BioLINCC files/ARIC/Main_Study/V3/csv/dtic04.csv',na.strings=c("NA","","NULL"))
aric_ecgmc35_v3 <- fread('~/Dropbox/BioLINCC files/ARIC/Main_Study/V3/csv/ecgmc35.csv',na.strings=c("NA","","NULL"))
aric_echa_v3 <- fread('~/Dropbox/BioLINCC files/ARIC/Main_Study/V3/csv/echa04.csv',na.strings=c("NA","","NULL"))
aric_echa_v3$lvmass_ix <- aric_echa_v3$ECHA58/aric_echa_v3$ECHA6
aric_hhxc_v3 <- fread('~/Dropbox/BioLINCC files/ARIC/Main_Study/V3/csv/hhxc04.csv',na.strings=c("NA","","NULL"))
aric_hmtcv301_v3 <- fread('~/Dropbox/BioLINCC files/ARIC/Main_Study/V3/csv/hmtcv301.csv',na.strings=c("NA","","NULL"))
aric_lipc04_v3 <- fread('~/Dropbox/BioLINCC files/ARIC/Main_Study/V3/csv/lipc04.csv',na.strings=c("NA","","NULL"))
aric_nutv3_v3 <- fread('~/Dropbox/BioLINCC files/ARIC/Main_Study/V3/csv/nutv3.csv',na.strings=c("NA","","NULL"))
aric_nutv3x_v3 <- fread('~/Dropbox/BioLINCC files/ARIC/Main_Study/V3/csv/nutv3.csv',na.strings=c("NA","","NULL"))
aric_phxa04_v3 <- fread('~/Dropbox/BioLINCC files/ARIC/Main_Study/V3/csv/phxa04.csv',na.strings=c("NA","","NULL"))
aric_rhxb_v3 <- fread('~/Dropbox/BioLINCC files/ARIC/Main_Study/V3/csv/rhxb04.csv',na.strings=c("NA","","NULL"))
aric_rpac04_v3 <- fread('~/Dropbox/BioLINCC files/ARIC/Main_Study/V3/csv/rpac04.csv',na.strings=c("NA","","NULL"))
aric_sbpc_v3 <- fread('~/Dropbox/BioLINCC files/ARIC/Main_Study/V3/csv/sbpc04_02.csv',na.strings=c("NA","","NULL"))
aric_vitnut01_v3 <- fread('~/Dropbox/BioLINCC files/ARIC/Main_Study/V3/csv/vitnut01.csv',na.strings=c("NA","","NULL"))

#### Hypertension


#### Diabetes


#### COPD


#### Asthma


#### Arthritis


#### CKD


#### Reproductive history 

## Calculate total time using hormone replacement agents
## Note these are only since last ARIC visit


aric_rhxb_v3[,hrt_agent7_yrs := RHXB16A + RHXB16B/12]

aric_rhxb_v3[,hrt_agent8_yrs :=
  RHXB28A + RHXB28B/12]


aric_hrt_yrs <-
        aric_rhxb_v3[,c("ID_C",
                        "hrt_agent7_yrs",
                        "hrt_agent8_yrs")][aric_hrt_yrs,
        on=.(ID_C)]

#### CHF

aric_outcomes$ID_C <- aric_outcomes$patientid

aric_derive3 <- 
  aric_outcomes[, .(ID_C, hfhosp_status, hfhosp_dt)][aric_derive3,
    on = .(ID_C)
  ]

aric_derive3[!is.na(hfhosp_status), chf_frail := 0]
aric_derive3[!is.na(hfhosp_status)&
               hfhosp_status==1&
               hfhosp_dt <= V3DAYS, chf_frail := 1]


#### CAD


#### Stroke


#### Cancer

### Echo calcs

aric_echa_v3 <-
  merge(aric_echa_v3,
        aric_antc04_v3[,c("ID_C","ANTC1","weight_kg")],
        on="ID_C",
        all.x=T)

aric_echa_v3 <-
  aric_derive1[,.(ID_C,GENDER)][aric_echa_v3,on=.(ID_C)]

aric_echa_v3 <- setDT(calc_hypertrophy_type(aric_echa_v3,
                      id = "ID_C",
                      sex="GENDER",
                      male="M",
                      female="F",
                      lvedd_cm = "ECHA51",
                      ivsd_cm = "ECHA40",
                      lvpwtd_cm = "ECHA54",
                      height_cm = "ANTC1",
                      weight_kg = "weight_kg"))


aric_msrcod36_v3 <- fread('~/Dropbox/BioLINCC files/ARIC/Main_Study/V3/csv/msrcod36.csv',na.strings=c("NA","","NULL"))
aric_msrcod36_codes <- aric_msrcod36_v3[,c(1,19:35)]
aric_melt_msrcod36_codes <- melt(aric_msrcod36_codes,id.vars="ID_C",na.rm=T,factorsAsStrings=T)
aric_melt_msrcod36_codes[,variable := "medcode"]
aric_melt_msrcod36_codes[,form := "msrcod"]
aric_melt_msrcod36_codes <- unique(aric_melt_msrcod36_codes)

aric_melt_derive3_v3 <- melt(aric_derive3,id.vars=c("ID_C"),na.rm=T,factorsAsStrings=T)
aric_melt_derive3_v3[,form := "derive3"]
aric_melt_antc_v3 <- melt(aric_antc04_v3,id.vars=c("ID_C"),na.rm=T,factorsAsStrings=T)
aric_melt_antc_v3[,form := "antc"]
aric_melt_atrfib31_v3 <- melt(aric_atrfib31_v3,id.vars=c("ID_C"),na.rm=T,factorsAsStrings=T)
aric_melt_atrfib31_v3[,form := "atrfib31"]
aric_melt_dtic04_v3 <- melt(aric_dtic04_v3,id.vars=c("ID_C"),na.rm=T,factorsAsStrings=T)
aric_melt_dtic04_v3[,form := "dtic04"]
aric_melt_ecgmc35_v3 <- melt(aric_ecgmc35_v3,id.vars=c("ID_C"),na.rm=T,factorsAsStrings=T)
aric_melt_ecgmc35_v3[,form := "ecgmc35"]
aric_melt_echa_v3 <- setDT(melt(aric_echa_v3,id.vars=c("ID_C"),na.rm=T,factorsAsStrings=T))
aric_melt_echa_v3[,form := "echa"]
aric_melt_hhxc_v3 <- melt(aric_hhxc_v3,id.vars=c("ID_C"),na.rm=T,factorsAsStrings=T)
aric_melt_hhxc_v3[,form := "hhxc"]
aric_melt_hmtcv301_v3 <- melt(aric_hmtcv301_v3,id.vars=c("ID_C"),na.rm=T,factorsAsStrings=T)
aric_melt_hmtcv301_v3[,form := "hmtcv301"]
aric_melt_lipc04_v3 <- melt(aric_lipc04_v3,id.vars=c("ID_C"),na.rm=T,factorsAsStrings=T)
aric_melt_lipc04_v3[,form := "lipc04"]
aric_melt_nutv3_v3 <- melt(aric_nutv3_v3,id.vars=c("ID_C"),na.rm=T,factorsAsStrings=T)
aric_melt_nutv3_v3[,form := "nutv3"]
aric_melt_phxa04_v3 <- melt(aric_phxa04_v3,id.vars=c("ID_C"),na.rm=T,factorsAsStrings=T)
aric_melt_phxa04_v3[,form := "phxa"]
aric_melt_rhxb_v3 <- melt(aric_rhxb_v3,id.vars=c("ID_C"),na.rm=T,factorsAsStrings=T)
aric_melt_rhxb_v3[,form := "rhxb"]
aric_melt_sbpc_v3 <- melt(aric_sbpc_v3,id.vars=c("ID_C"),na.rm=T,factorsAsStrings=T)
aric_melt_sbpc_v3[,form := "sbpc"]
aric_melt_vitnut01_v3 <- melt(aric_vitnut01_v3,id.vars=c("ID_C"),na.rm=T,factorsAsStrings=T)
aric_melt_vitnut01_v3[,form := "vitnut01"]

aric_melt_amha02_v3 <- melt(aric_amha02_v3,id.vars="ID_C",na.rm=T, factorsAsStrings=T)
aric_melt_amha02_v3[,form := "amha"]
aric_melt_v3_all <- 
  rbindlist(
    list(
      aric_melt_derive3_v3,
      aric_melt_antc_v3,
      aric_melt_atrfib31_v3,
      aric_melt_dtic04_v3,
      aric_melt_ecgmc35_v3,
      aric_melt_echa_v3,
      aric_melt_amha02_v3,
      aric_melt_hhxc_v3,
      aric_melt_hmtcv301_v3,
      aric_melt_lipc04_v3,
      aric_melt_nutv3_v3,
      aric_melt_phxa04_v3,
      aric_melt_rhxb_v3,
      aric_melt_sbpc_v3,
      aric_melt_vitnut01_v3))

aric_melt_v3_all[,variable := as.character(variable)]

aric_v3_tempdays <- 
  aric_dates[!is.na(V3DAYS)
             ,
             .(
               ID_C,
               visitdays = V3DAYS
             )
  ]


aric_melt_v3_all[,visit := 3]
aric_melt_v3_all <- 
  aric_v3_tempdays[aric_melt_v3_all,
                          on=.(ID_C)]

aric_melt_v3_all[,CONTYR := 7]

rm(list=ls(pattern="\\baric_.*\\_v3$"))


#===============================================================#
####                       ARIC Exam 4                       ####
#===============================================================#

aric_derive4 <- fread('~/Dropbox/BioLINCC files/ARIC/Main_Study/v4/csv/derive47.csv',na.strings=c("NA","","NULL"))

aric_derive4[,drinks_wk := ETHANL41/14]


#### CHF

aric_derive4 <-
    aric_outcomes[, .(ID_C, hfhosp_status, hfhosp_dt)][aric_derive4,
    on = .(ID_C)
  ]

aric_derive4[!is.na(hfhosp_status), chf_frail :=  0]
aric_derive4[!is.na(hfhosp_status)&
               hfhosp_status==1&
               hfhosp_dt <= V4DAYS, chf_frail := 1]

#### CAD

aric_antd05_v4 <- fread('~/Dropbox/BioLINCC files/ARIC/Main_Study/V4/csv/antd05.csv',na.strings=c("NA","","NULL"))
aric_antd05_v4[,weight_kg := ANTD2/2.2046226218]
aric_atrfib41_v4 <- fread('~/Dropbox/BioLINCC files/ARIC/Main_Study/V4/csv/atrfib41.csv',na.strings=c("NA","","NULL"))
aric_atrfib41_v4[, AFIBFL := 0]
aric_atrfib41_v4[AF==1|AFL==1, AFIBFL := 1]
aric_bpub04_v4 <- fread('~/Dropbox/BioLINCC files/ARIC/Main_Study/V4/csv/bpub04.csv',na.strings=c("NA","","NULL"))
aric_dhsa_v4 <- fread('~/Dropbox/BioLINCC files/ARIC/Main_Study/V4/csv/dhsa04.csv',na.strings=c("NA","","NULL"))

### For FIFE dental score, using history of tooth loss due to cavities, gum disease, or accident and presence of false teeh

aric_dhsa_v4[!is.na(DHSA2A)&
               !is.na(DHSA2B)&
               !is.na(DHSA2C)&
               !is.na(DHSA3), 
             mouth_frail := 0]

aric_dhsa_v4[DHSA2A=="Y"|
               DHSA2B=="Y"|
               DHSA2C=="Y"|
               DHSA3=="Y", 
             mouth_frail := 1]


aric_ecgmd41_v4 <- fread('~/Dropbox/BioLINCC files/ARIC/Main_Study/V4/csv/ecgmd41.csv',na.strings=c("NA","","NULL"))
aric_hhxd04_v4 <- fread('~/Dropbox/BioLINCC files/ARIC/Main_Study/V4/csv/hhxd04.csv',na.strings=c("NA","","NULL"))
aric_hmtcv401_v4 <- fread('~/Dropbox/BioLINCC files/ARIC/Main_Study/V4/csv/hmtcv401.csv',na.strings=c("NA","","NULL"))
aric_infa04_v4 <- fread('~/Dropbox/BioLINCC files/ARIC/Main_Study/V4/csv/infa04.csv',na.strings=c("NA","","NULL"))
aric_mhqa04_v4 <- fread('~/Dropbox/BioLINCC files/ARIC/Main_Study/V4/csv/mhqa04.csv',na.strings=c("NA","","NULL"))
aric_lipd04_v4 <- fread('~/Dropbox/BioLINCC files/ARIC/Main_Study/V4/csv/lipd04.csv',na.strings=c("NA","","NULL"))
aric_phxb04_v4 <- fread('~/Dropbox/BioLINCC files/ARIC/Main_Study/V4/csv/phxb04.csv',na.strings=c("NA","","NULL"))
aric_sbpd_v4 <- fread('~/Dropbox/BioLINCC files/ARIC/Main_Study/V4/csv/sbpd04_02.csv',na.strings=c("NA","","NULL"))

aric_phxb04_v4[PHXB2A=="Y"|
                 PHXB2B=="Y"|
                 PHXB2C=="Y"|
                 PHXB2D=="Y", 
               health_coverage := "Y"]

aric_phxb04_v4[PHXB2A=="N"|
                 PHXB2B=="N"|
                 PHXB2C=="N"|
                 PHXB2D=="N", 
               health_coverage := "N"]

aric_rhxc04_v4 <- fread('~/Dropbox/BioLINCC files/ARIC/Main_Study/V4/csv/rhxc04.csv',na.strings=c("NA","","NULL"))

#### Reproductive history 

## Calculate total time using hormone replacement agents
## Note these are only since last ARIC visit


aric_rhxc04_v4[, hrt_agent9_yrs := RHXC16A + RHXC16B/12]

aric_rhxc04_v4[, hrt_agent10_yrs := RHXC28A + RHXC28B/12]

aric_hrt_yrs <-
  aric_hrt_yrs[
        aric_rhxc04_v4[,c("ID_C",
                          "hrt_agent9_yrs",
                          "hrt_agent10_yrs")],
        on=.(ID_C)]


aric_hrt_yrs[,hrt_years_sum :=
  apply(aric_hrt_yrs[,2:11],MARGIN=1,FUN=function(x) sum(x,na.rm=T))]

aric_sesa04_v4 <- fread('~/Dropbox/BioLINCC files/ARIC/Main_Study/V4/csv/sesa04.csv',na.strings=c("NA","","NULL"))


aric_msrcod43_v4 <- fread('~/Dropbox/BioLINCC files/ARIC/Main_Study/V4/csv/msrcod43.csv',na.strings=c("NA","","NULL"))
aric_msrcod43_codes <- aric_msrcod43_v4[,c(1,19:35)]
aric_melt_msrcod43_codes <- melt(aric_msrcod43_codes,id.vars="ID_C",na.rm=T,factorsAsStrings=T)
aric_melt_msrcod43_codes[,variable := "medcode"]
aric_melt_msrcod43_codes[,form := "msrcod"]
aric_melt_msrcod43_codes <- unique(aric_melt_msrcod43_codes)

aric_melt_derive4_v4 <- melt(aric_derive4,id.vars=c("ID_C"),na.rm=T,factorsAsStrings=T)
aric_melt_derive4_v4[,form := "derive4"]

aric_melt_antd05_v4 <- melt(aric_antd05_v4,id.vars=c("ID_C"),na.rm=T,factorsAsStrings=T)
aric_melt_antd05_v4[,form := "antd"]

aric_melt_atrfib41_v4 <- melt(aric_atrfib41_v4,id.vars=c("ID_C"),na.rm=T,factorsAsStrings=T)
aric_melt_atrfib41_v4[,form := "atrfib41"]
aric_melt_ecgmd41_v4 <- melt(aric_ecgmd41_v4,id.vars=c("ID_C"),na.rm=T,factorsAsStrings=T)
aric_melt_ecgmd41_v4[,form := "ecgmd41"]
aric_melt_hhxd04_v4 <- melt(aric_hhxd04_v4,id.vars=c("ID_C"),na.rm=T,factorsAsStrings=T)
aric_melt_hhxd04_v4[,form := "hhxd04"]
aric_melt_hmtcv401_v4 <- melt(aric_hmtcv401_v4,id.vars=c("ID_C"),na.rm=T,factorsAsStrings=T)
aric_melt_hmtcv401_v4[,form := "hmtcv401"]
aric_melt_lipd04_v4 <- melt(aric_lipd04_v4,id.vars=c("ID_C"),na.rm=T,factorsAsStrings=T)
aric_melt_lipd04_v4[,form := "lipd04"]
aric_melt_phxb04_v4 <- melt(aric_phxb04_v4,id.vars=c("ID_C"),na.rm=T,factorsAsStrings=T)
aric_melt_phxb04_v4[,form := "phxb04"]
aric_melt_rhxc04_v4 <- melt(aric_rhxc04_v4,id.vars=c("ID_C"),na.rm=T,factorsAsStrings=T)
aric_melt_rhxc04_v4[,form := "rhxc"]
aric_melt_sesa04_v4 <- melt(aric_sesa04_v4,id.vars=c("ID_C"),na.rm=T,factorsAsStrings=T)
aric_melt_sesa04_v4[,form := "sesa"]
aric_melt_sbpd04_v4 <- melt(aric_sbpd_v4,id.vars=c("ID_C"),na.rm=T,factorsAsStrings=T)
aric_melt_sbpd04_v4[,form := "sbpd"]

aric_melt_hrt_yrs <- aric_hrt_yrs[,.(ID_C,hrt_years_sum)]
aric_melt_hrt_yrs[,variable := "hrt_years_total"]
aric_melt_hrt_yrs[, value := hrt_years_sum]
aric_melt_hrt_yrs[,form := "hrt_yrs"]


aric_melt_v4_all <- 
  rbindlist(
    list(aric_melt_derive4_v4,
         aric_melt_antd05_v4,
         aric_melt_atrfib41_v4,
         aric_melt_ecgmd41_v4,
         aric_melt_hhxd04_v4,
         aric_melt_hmtcv401_v4,
         aric_melt_lipd04_v4,
         aric_melt_phxb04_v4,
         aric_melt_rhxc04_v4,
         aric_melt_sesa04_v4,
         aric_melt_sbpd04_v4,
         aric_melt_hrt_yrs[,c("ID_C","variable","value","form")])
  )


aric_v4_tempdays <- 
  unique(aric_dates[!is.na(V4DAYS),
             .(ID_C, visitdays = V4DAYS)])

aric_melt_v4_all[,visit := 4]

aric_melt_v4_all <-
  aric_v4_tempdays[aric_melt_v4_all,
  on = .(ID_C)
]

aric_melt_v4_all[,CONTYR := 10]

rm(list=ls(pattern="\\baric_.*\\_v4$"))


#===============================================================#
####                       ARIC Exam 5                       ####
#===============================================================#


aric_derive5[,drinks_wk := ETHANL51/14]
aric_derive5[,fev1fvc_fraction := FEV1FVC51/100]
aric_derive5 <-
  aric_derive5[unique(aric_dates_long[,.(ID_C,GENDER)]),
               on=.(ID_C)]


aric_alc_v5 <- fread('~/Dropbox/BioLINCC files/ARIC/Main_Study/V5/csv/alc.csv',na.strings=c("NA","","NULL"))
aric_ant_v5 <- fread('~/Dropbox/BioLINCC files/ARIC/Main_Study/V5/csv/ant.csv',na.strings=c("NA","","NULL"))
aric_cbc_v5 <- fread('~/Dropbox/BioLINCC files/ARIC/Main_Study/V5/csv/cbc.csv',na.strings=c("NA","","NULL"))
aric_cdi_v5 <- fread('~/Dropbox/BioLINCC files/ARIC/Main_Study/V5/csv/cdi.csv',na.strings=c("NA","","NULL"))
aric_ces_v5 <- fread('~/Dropbox/BioLINCC files/ARIC/Main_Study/V5/csv/ces.csv',na.strings=c("NA","","NULL"))
aric_chm_v5 <- fread('~/Dropbox/BioLINCC files/ARIC/Main_Study/V5/csv/chm.csv',na.strings=c("NA","","NULL"))
aric_dcm_v5 <- fread('~/Dropbox/BioLINCC files/ARIC/Main_Study/V5/csv/dcm.csv',na.strings=c("NA","","NULL"))
aric_ecg_v5 <- fread('~/Dropbox/BioLINCC files/ARIC/Main_Study/V5/csv/ecg.csv',na.strings=c("NA","","NULL"))
aric_ech_v5 <- fread('~/Dropbox/BioLINCC files/ARIC/Main_Study/V5/csv/ech.csv',na.strings=c("NA","","NULL"))
aric_eco_v5 <- fread('~/Dropbox/BioLINCC files/ARIC/Main_Study/V5/csv/eco.csv',na.strings=c("NA","","NULL"))
aric_lip_v5 <- fread('~/Dropbox/BioLINCC files/ARIC/Main_Study/V5/csv/lip.csv',na.strings=c("NA","","NULL"))
aric_nfh_v5 <- fread('~/Dropbox/BioLINCC files/ARIC/Main_Study/V5/csv/nfh.csv',na.strings=c("NA","","NULL"))
aric_nhx_v5 <- fread('~/Dropbox/BioLINCC files/ARIC/Main_Study/V5/csv/nhx.csv',na.strings=c("NA","","NULL"))
aric_pac_v5 <- fread('~/Dropbox/BioLINCC files/ARIC/Main_Study/V5/csv/pac.csv',na.strings=c("NA","","NULL"))
aric_phx_v5 <- fread('~/Dropbox/BioLINCC files/ARIC/Main_Study/V5/csv/phx.csv',na.strings=c("NA","","NULL"))
aric_rse_v5 <- fread('~/Dropbox/BioLINCC files/ARIC/Main_Study/V5/csv/rse.csv',na.strings=c("NA","","NULL"))
aric_aqc_v5 <- fread('~/Dropbox/BioLINCC files/ARIC/Main_Study/V5/csv/aqc.csv',na.strings=c("NA","","NULL"))
aric_sbp_v5 <- fread('~/Dropbox/BioLINCC files/ARIC/Main_Study/V5/csv/sbp.csv',na.strings=c("NA","","NULL"))
aric_sfe_v5 <- fread('~/Dropbox/BioLINCC files/ARIC/Main_Study/V5/csv/sfe.csv',na.strings=c("NA","","NULL"))
aric_status51_v5 <- fread('~/Dropbox/BioLINCC files/ARIC/Main_Study/V5/csv/status51.csv',na.strings=c("NA","","NULL"))
aric_hom_v1 <- fread('~/Dropbox/BioLINCC files/ARIC/Main_Study/V1/csv/hom.csv',na.strings=c("NA","","NULL"))


aric_status51_v5[,AGE_MENOPAUSE_EX05 :=
  apply(aric_status51_v5[,c("AGENATMENOPAUSEF","AGESRGMENOPAUSEF")],
        MARGIN=1,
        FUN=function(x) min(x,na.rm=T))]

aric_status51_v5[AGE_MENOPAUSE_EX05=="Inf", AGE_MENOPAUSE_EX05 := NA]


### Exam 5 FRAIL Score ###

## Fatigue

aric_derive5[,fatigue_frail := EXHAUSTCOMP]

## Resistance

##### Used SFE question 2b ('Does your health now limit you in climing several flights of stairs?)
##### 1: Yes, limited a lot
##### 2: Yes limited a little
##### 3: No, not limited at all

# aric_genps1$GEN10
# aric_gnbps1$GNB5
# aric_gncps1$GNC2B
# aric_gneps1$GNE4
# aric_paq[,c("PAQ2","PAQ2A")]
# aric_paqa04[,c("PAQA2")]
# aric_sfe$SFE2B

aric_derive5 <- 
  aric_sfe_v5[,.(ID_C, SFE2B)][aric_derive5, on=.(ID_C)]


aric_derive5[SFE2B %in% c(1,2), resistance_frail :=  1]
aric_derive5[SFE2B == 3, resistance_frail := 0]


## Ambulate

aric_derive5 <- 
  aric_genps1[,.(ID_C,GEN9)][aric_derive5, on=.(ID_C)]
  


aric_derive5 <- 
  aric_rse_v5[,.(ID_C,RSE7,RSE8,RSE9)][aric_derive5,
    on=.(ID_C)]


aric_derive5[RSE8=="Y"|
               RSE9=="Y"|
               GEN9 %in% c("Much Difficulty",
                           "Some Difficulty",
                           "Unable to Do"), 
             ambulate_frail := 1]

aric_derive5[RSE8=="N"&
               RSE9=="N"&
               GEN9 %in% c("No Difficulty"), 
             ambulate_frail := 0]


## Illness (composite)

#### Hypertension

aric_derive5[,htn_frail := HYPERT54]

#### Diabetes

aric_derive5[!DIABTS54=="T", dm_frail := as.numeric(DIABTS54)]


#### COPD

aric_derive5 <- 
    aric_status51_v5[,.(ID_C,INCSELFREPLUNG51)][aric_derive5,
    on=.(ID_C)
  ]

aric_derive5 <- 
  aric_rse_v5[,.(ID_C,RSE11,RSE11B)][aric_derive5,on=.(ID_C)]


aric_derive5 <- 
    aric_hom_v1[,.(ID_C,HOM10G)][aric_derive5,
    on=.(ID_C)]
  

aric_derive5[INCSELFREPLUNG51==1|
               RSE11=="Y"|
               RSE11B=="Y"|
               HOM10G=="Y" , copd_frail := 1]

aric_derive5[is.na(copd_frail), copd_frail := 0]

#### Asthma

aric_derive5 <- 
    aric_status51_v5[,.(ID_C,INCSELFREPAST51)][aric_derive5,
    on=.(ID_C)
  ]


aric_derive5 <- 
    aric_rse_v5[,.(ID_C,RSE14,RSE14C)][aric_derive5,
    on=.(ID_C)
  ]


aric_derive5 <- 
    aric_hom_v1[,.(ID_C,HOM10H)][aric_derive5,
    on=.(ID_C)
  ]

aric_derive5[INCSELFREPAST51==1|
                 RSE14=="Y"|
                 RSE14C=="Y"|
                 HOM10H=="Y", 
               asthma_frail := 1]


  aric_derive5[is.na(asthma_frail), 
               asthma_frail := 0]


#### Arthritis

aric_infa04_v4 <- fread('~/Dropbox/BioLINCC files/ARIC/Main_Study/V4/csv/infa04.csv',na.strings=c("NA","","NULL"))

aric_derive5 <- 
  aric_infa04_v4[
    aric_derive5,
    on=.(ID_C)
  ]

aric_derive5[INFA3A=="Y", djd_frail := 1]
aric_derive5[INFA3A=="N", djd_frail := 0]

#### CKD

aric_derive5 <- 
    aric_analytes[,.(ID_C, EGFRSCR_V5)][aric_derive5,
    on=.(ID_C)
  ]

aric_derive5[EGFRSCR_V5>=60, ckd_frail := 0]
aric_derive5[EGFRSCR_V5 <60, ckd_frail := 1]


#### CHF

aric_derive5 <-
  aric_derive5[, chf_frail := PREVDEFHF51]


#### CAD

aric_derive5 <-
  aric_derive5[,chd_frail := PRVCHD51]


#### Stroke

aric_derive5 <-
  aric_derive5[, stroke_frail := PRVSTR51]


#### Cancer

aric_derive5 <-
aric_derive5[,cancer_frail := V5CANCER51]

aric_derive5[,conditions_frail := 
               dm_frail+
               htn_frail+
               copd_frail+
               asthma_frail+
               djd_frail+
               ckd_frail+
               chf_frail+
               chd_frail+
               stroke_frail+
               cancer_frail]


aric_derive5[conditions_frail <= 4, illness_frail :=  0]
aric_derive5[conditions_frail > 4, illness_frail := 1]

aric_derive5[,wtloss_frail := WTLOSSCOMP10]

aric_derive5[,total_frail := 
  fatigue_frail+
  resistance_frail+
  ambulate_frail+
  illness_frail+
  wtloss_frail]


# -------------------------------------------------- #


######### FRIED

##### Walking speed


aric_derive5 <- 
    aric_ant_v5[,.(ID_C, ANT3)][aric_derive5,,
    on = .(ID_C)
  ]

# Set default to 0

aric_derive5[!is.na(WALK4M51), gaitspeed_fried := 0] 

# Set to 1 for male criteria

#### Men:
# Height ≤ 173 cm (68.1 in):  ≤0.96 m/s      
# Height >173 cm (68.1 in):   ≤1.04  m/s	
# 


aric_derive5[GENDER=="M"&
               ANT3>173&
               WALK4M51 >= 6, 
             gaitspeed_fried := 1]

aric_derive5[GENDER=="M"&
               ANT3 <= 173&
               WALK4M51 >=7, gaitspeed_fried := 1]

### Women
# Height ≤ 159 cm (62.6 in):  ≤0.90m/s
# Height >159 cm (62.6 in):   ≤1.02 m/s


aric_derive5[GENDER=="F"&
               ANT3>159&
               WALK4M51 >= 6, gaitspeed_fried := 1]

aric_derive5[GENDER=="F"&
               ANT3 <= 159&
               WALK4M51 >= 7, gaitspeed_fried := 1]

########### Grip (kg)

# Trials for each hand

## Uses BMI for normalization

##### Men
# BMI ≤ 24.0:     ≤ 29 kg
# BMI 24.1-26.0:  ≤ 30 kg
# BMI 26.1-28.0:  ≤ 30 kg
# BMI > 28.0:     ≤ 32 kg

##### Women
# BMI ≤ 23.0:     ≤ 17
# BMI 23.1-26.0:  ≤ 17.3 kg
# BMI 26.1-29.0:  ≤ 18 kg
# BMI > 29.0:     ≤ 21 kg

aric_derive5[,grip_avg := GRIPMEAN51]
aric_derive5[,grip_max := GRIPBEST51]

aric_derive5[!is.na(grip_max), grip_fried := 0] 

# Men

aric_derive5[
  BMI51<=24&
    grip_max <= 29&
    GENDER=="M", 
  grip_fried := 1] 

aric_derive5[
  BMI51>24&BMI51<=26&
    grip_max <= 30&
    GENDER=="M",
  grip_fried := 1] 

aric_derive5[BMI51>26&
               BMI51<=28&
               grip_max <= 30&
               GENDER=="M", grip_fried := 1] 

aric_derive5[BMI51>28&
               grip_max <= 32&
               GENDER=="M", grip_fried := 1] 

# Women

aric_derive5[BMI51<=23&
               grip_max <= 17&
               GENDER=="F", 
             grip_fried := 1] 

aric_derive5[BMI51>23&
               BMI51<=26&
               grip_max <= 17.3&
               GENDER=="F", 
             grip_fried := 1] 

aric_derive5[BMI51>26&
               BMI51<=29&
               grip_max <= 18&
               GENDER=="F", 
             grip_fried := 1] 

aric_derive5[BMI51>29&
               grip_max <= 21&
               GENDER=="F", 
             grip_fried := 1] 


aric_derive5[,activity_fried := PACCOMP20]

aric_derive5[,wtloss_fried := WTLOSSCOMP10]

aric_derive5[,fatigue_fried := EXHAUSTCOMP]


aric_derive5[,total_fried := 
  wtloss_fried+
  grip_fried+
  fatigue_fried+
  gaitspeed_fried+
  activity_fried]


### CES-D

aric_ces_v5[,cesd_11 := 
              apply(aric_ces_v5[,2:12],
                    MARGIN=1,
                    function(x)
                      sum(x))]

## Echo E/e'

aric_ech_v5[,e_eprime_lateral := ECH20/ECH24]
aric_ech_v5[,e_eprime_septal := ECH20/ECH26]
aric_ech_v5[,e_eprime_avg := (e_eprime_lateral+e_eprime_septal)/2]

# RVEDA index
aric_ech_v5[,bsa := ECH11/ECH12]
aric_ech_v5[,rveda_ix := ECH36/bsa]

# LVH

aric_ech_v5 <-
  merge(aric_ech_v5,
        aric_ant_v5[,c("ID_C","ANT3","ANT4")],
        on="ID_C",
        all.x=T)

aric_ech_v5 <-
  aric_derive1[,.(ID_C,GENDER)][aric_ech_v5,on=.(ID_C)]

aric_ech_v5 <- setDT(calc_hypertrophy_type(aric_ech_v5,
                                      id = "ID_C",
                                      sex="GENDER",
                                      male="M",
                                      female="F",
                                      lvedd_cm = "ECH4",
                                      ivsd_cm = "ECH6",
                                      lvpwtd_cm = "ECH7",
                                      height_cm = "ANT3",
                                      weight_kg = "ANT4"))


# -------------------------------------------------- #

## SF-12

### Physical functioning scale

aric_sfe_v5[,SF12PF51 := 100*(((SFE2A+SFE2B)-2)/4)]

aric_sfe_v5[,SF12PFZ51 :=
  (SF12PF51 -81.18122)/29.10588]


### Role Physical Scale

aric_sfe_v5[,SF12RP51 :=
  100*(((SFE3A+SFE3B)-2)/8)]

aric_sfe_v5[,SF12RPZ51 :=
  (SF12RP51 - 80.52856)/27.13526]


### Bodily Pain scale

aric_sfe_v5[,SF12BP51 :=
  100*(((6+SFE5)-1)/4)]

aric_sfe_v5[,SF12BPZ51 :=
  (SF12BP51 - 81.74015)/24.53019]


### General Health Scale

aric_sfe_v5[SFE1 %in% c(1,4,5), 
            SFE1_ren := 6-SFE1]

aric_sfe_v5[SFE1 %in% c(2,3), 
            SFE1_ren := (6-SFE1)+0.4]

aric_sfe_v5[,SF12GH51 := 100*((SFE1_ren=1)/4)]

aric_sfe_v5[,SF12GHZ51 := (SF12GH51-55.9090)/24.84380]


### Vitality scale

aric_sfe_v5[,SF12VT51 := 100*(((6-SFE6B)-1)/4)]

aric_sfe_v5[,SF12VTZ51 := (SF12VT51 - 55.59090)/24.84380]


### Social functioning scale

aric_sfe_v5[,SF12SF51 := 
  100*(((6-SFE7)-1)/4)]

aric_sfe_v5[,SF12SFZ51 := 
  (SF12SF51 - 86.41051)/22.35543]


### Role Emotional Scale

aric_sfe_v5[,SF12RE51 := 
  100*(((SFE4A+
         SFE4B)-2)/8)]

aric_sfe_v5[,SF12REZ51 := 
  (SF12RE51 - 86.41051)/22.35543]


### Mental health scale

aric_sfe_v5[,SF12MH51 := 
  100*((((6-SFE6A)+SFE6C)-2)/8)]

aric_sfe_v5[,SF12MHZ51 := 
  (SF12MH51 - 70.18217)/20.50597] 


### Aggregate Mental Health Score

aric_sfe_v5[,SF12AGGMENT51 := 
  (SF12PFZ51* (-0.22999)) +
  (SF12RPZ51* (-0.12329)) +
  (SF12BPZ51* (-0.09731)) +
  (SF12GHZ51* (-0.01571)) +
  (SF12VTZ51* 0.23534) +
  (SF12SFZ51* 0.26876) +
  (SF12REZ51* 0.43407) +
  (SF12MHZ51* 0.48581)]


### Aggregate Physical Health Score

aric_sfe_v5[,SF12AGGPHYS51 :=
  (SF12PFZ51 * 0.42402) +
  (SF12RPZ51* 0.35119) +
  (SF12BPZ51* 0.31754) +
  (SF12GHZ51* 0.24954) +
  (SF12VTZ51* 0.02877) +
  (SF12SFZ51* (-0.00753)) +
  (SF12REZ51* (-0.19206)) +
  (SF12MHZ51* (-0.22069))] 

# -------------------------------------------------- #

## Medications

aric_msrcod51_v5 <- fread('~/Dropbox/BioLINCC files/ARIC/Main_Study/V5/csv/msrcod51.csv',na.strings=c("NA","","NULL"))
aric_msrcod51_codes <- aric_msrcod51_v5[,c(1,27:51)]
aric_melt_msrcod51_codes <- melt(aric_msrcod51_codes,id.vars="ID_C",na.rm=T,factorsAsStrings=T)
aric_melt_msrcod51_codes[,variable := "medcode"]
aric_melt_msrcod51_codes[,form := "msrcod"]
aric_melt_msrcod51_codes <- unique(aric_melt_msrcod51_codes)

aric_melt_derive5_v5 <- melt(aric_derive5,id.vars=c("ID_C"),na.rm=T,factorsAsStrings=T)
aric_melt_derive5_v5[,form := "derive5"]
aric_melt_ant_v5 <- melt(aric_ant_v5,id.vars=c("ID_C"),na.rm=T,factorsAsStrings=T)
aric_melt_ant_v5[,form := "ant"]
aric_melt_cbc_v5 <- melt(aric_cbc_v5,id.vars=c("ID_C"),na.rm=T,factorsAsStrings=T)
aric_melt_cbc_v5[,form := "cbc"]
aric_melt_cdi_v5 <- melt(aric_cdi_v5,id.vars=c("ID_C"),na.rm=T,factorsAsStrings=T)
aric_melt_cdi_v5[,form := "cdi"]
aric_melt_ces_v5 <- melt(aric_ces_v5,id.vars=c("ID_C"),na.rm=T,factorsAsStrings=T)
aric_melt_ces_v5[,form := "ces"]
aric_melt_chm_v5 <- melt(aric_chm_v5,id.vars=c("ID_C"),na.rm=T,factorsAsStrings=T)
aric_melt_chm_v5[,form := "chm"]
aric_melt_dcm_v5 <- melt(aric_dcm_v5,id.vars=c("ID_C"),na.rm=T,factorsAsStrings=T)
aric_melt_dcm_v5[,form := "dcm"]
aric_melt_ech_v5 <- melt(aric_ech_v5,id.vars=c("ID_C"),na.rm=T,factorsAsStrings=T)
aric_melt_ech_v5[,form := "ech"]
aric_melt_eco_v5 <- melt(aric_eco_v5,id.vars=c("ID_C"),na.rm=T,factorsAsStrings=T)
aric_melt_eco_v5[,form := "eco"]
aric_melt_ecg_v5 <- melt(aric_ecg_v5,id.vars=c("ID_C"),na.rm=T,factorsAsStrings=T)
aric_melt_ecg_v5[,form := "ecg"]
aric_melt_lip_v5 <- melt(aric_lip_v5,id.vars=c("ID_C"),na.rm=T,factorsAsStrings=T)
aric_melt_lip_v5[,form := "lip"]

aric_melt_nfh_v5 <- melt(aric_nfh_v5,id.vars=c("ID_C"),na.rm=T,factorsAsStrings=T)
aric_melt_nfh_v5[,form := "nfh"]
aric_melt_nhx_v5 <- melt(aric_nhx_v5,id.vars=c("ID_C"),na.rm=T,factorsAsStrings=T)
aric_melt_nhx_v5[,form := "nhx"]

aric_melt_pac_v5 <- melt(aric_pac_v5,id.vars=c("ID_C"),na.rm=T,factorsAsStrings=T)
aric_melt_pac_v5[,form := "pac"]

aric_melt_rse_v5 <- melt(aric_rse_v5,id.vars=c("ID_C"),na.rm=T,factorsAsStrings=T)
aric_melt_rse_v5[,form := "rse"]

aric_melt_sfe_v5 <- melt(aric_sfe_v5,id.vars=c("ID_C"),na.rm=T,factorsAsStrings=T)
aric_melt_sfe_v5[,form := "sfe"]

aric_melt_status51_v5 <- melt(aric_status51_v5,id.vars=c("ID_C"),na.rm=T,factorsAsStrings=T)
aric_melt_status51_v5[,form := "status51"]


aric_melt_v5_all <- 
  rbindlist(
    list(aric_melt_derive5_v5,
         # aric_melt_sbp_v5,
         aric_melt_ant_v5,
         aric_melt_cbc_v5,
         aric_melt_ces_v5,
         aric_melt_chm_v5,
         aric_melt_dcm_v5,
         aric_melt_ecg_v5,
         aric_melt_ech_v5,
         aric_melt_eco_v5,
         aric_melt_lip_v5,
         aric_melt_nfh_v5,
         aric_melt_nhx_v5,
         aric_melt_pac_v5,
         aric_melt_rse_v5,
         aric_melt_sfe_v5,
         aric_melt_status51_v5))


aric_v5_tempdays <-
  aric_dates[!is.na(V5DATE51_DAYS),.(ID_C,visitdays = V5DATE51_DAYS)]

aric_melt_v5_all[,visit := 5]

aric_melt_v5_all <-
    aric_v5_tempdays[
      aric_melt_v5_all,
    on="ID_C"]

aric_melt_v5_all[,CONTYR := 23]

rm(list=ls(pattern="\\baric_.*\\_v5$"))

#===============================================================#
####                       ARIC Exam 6                       ####
#===============================================================#

aric_derive6 <- fread('~/Dropbox/BioLINCC files/ARIC/Main_Study/V6/csv/derive61.csv',na.strings=c("NA","","NULL"))


aric_derive6[,drinks_wk := ETHANL61/14]
aric_derive6[, hdl_mgdl := HDLSIU61 * 38.67]


aric_alc_v6 <- fread('~/Dropbox/BioLINCC files/ARIC/Main_Study/V6/csv/alc.csv',na.strings=c("NA","","NULL"))
aric_ant_v6 <- fread('~/Dropbox/BioLINCC files/ARIC/Main_Study/V6/csv/ant.csv',na.strings=c("NA","","NULL"))
aric_cdi_v6 <- fread('~/Dropbox/BioLINCC files/ARIC/Main_Study/V6/csv/cdi.csv',na.strings=c("NA","","NULL"))
aric_ces_v6 <- fread('~/Dropbox/BioLINCC files/ARIC/Main_Study/V6/csv/ces.csv',na.strings=c("NA","","NULL"))
aric_chm_v6 <- fread('~/Dropbox/BioLINCC files/ARIC/Main_Study/V6/csv/chem2.csv',na.strings=c("NA","","NULL"))
aric_egfr_v6 <- fread('~/Dropbox/BioLINCC files/ARIC/Main_Study/V6/csv/egfr.csv',na.strings=c("NA","","NULL"))
aric_ess_v6 <- fread('~/Dropbox/BioLINCC files/ARIC/Main_Study/V6/csv/ess.csv',na.strings=c("NA","","NULL"))
aric_lip_v6 <- fread('~/Dropbox/BioLINCC files/ARIC/Main_Study/V6/csv/lipf.csv',na.strings=c("NA","","NULL"))
aric_nhx_v6 <- fread('~/Dropbox/BioLINCC files/ARIC/Main_Study/V6/csv/nhx.csv',na.strings=c("NA","","NULL"))
aric_pac_v6 <- fread('~/Dropbox/BioLINCC files/ARIC/Main_Study/V6/csv/pac.csv',na.strings=c("NA","","NULL"))
aric_paq_v6 <- fread('~/Dropbox/BioLINCC files/ARIC/Main_Study/V6/csv/paq.csv',na.strings=c("NA","","NULL"))
aric_pfx_v6 <- fread('~/Dropbox/BioLINCC files/ARIC/Main_Study/V6/csv/pfx.csv',na.strings=c("NA","","NULL"))
aric_phx_v6 <- fread('~/Dropbox/BioLINCC files/ARIC/Main_Study/V6/csv/phx.csv',na.strings=c("NA","","NULL"))
aric_sbp_v6 <- fread('~/Dropbox/BioLINCC files/ARIC/Main_Study/V6/csv/sbp.csv',na.strings=c("NA","","NULL"))
aric_sfe_v6 <- fread('~/Dropbox/BioLINCC files/ARIC/Main_Study/V6/csv/sfe.csv',na.strings=c("NA","","NULL"))
aric_status61_v6 <- fread('~/Dropbox/BioLINCC files/ARIC/Main_Study/V6/csv/status61.csv',na.strings=c("NA","","NULL"))
aric_tmw_v6 <- fread('~/Dropbox/BioLINCC files/ARIC/Main_Study/V6/csv/tmw.csv',na.strings=c("NA","","NULL"))

aric_status61_v6[,AGE_MENOPAUSE_EX06  := apply(aric_status61_v6[,.(AGENATMENOPAUSEF,AGESRGMENOPAUSEF)],
        MARGIN=1,
        FUN=function(x) min(x,na.rm=T))]

aric_status61_v6[AGE_MENOPAUSE_EX06=="Inf",AGE_MENOPAUSE_EX06 := NA]

### FRAIL Score ###

## Fatigue

aric_derive6[,fatigue_frail := EXHAUST61]

## Resistance

##### Used SFE question 2b ('Does your health now limit you in climing several flights of stairs?)
##### 1: Yes, limited a lot
##### 2: Yes limited a little
##### 3: No, not limited at all

# aric_genps1$GEN10
# aric_gnbps1$GNB5
# aric_gncps1$GNC2B
# aric_gneps1$GNE4
# aric_paq[,c("PAQ2","PAQ2A")]
# aric_paqa04[,c("PAQA2")]
# aric_sfe$SFE2B


aric_derive6 <- 
  aric_derive6[
    aric_paq_v6[,.(ID_C,PAQ2)],
    on=.(ID_C)]

aric_derive6[PAQ2 %in% c('B','C'),resistance_frail := 1]
aric_derive6[PAQ2=='A', resistance_frail :=0 ]


## Ambulate

aric_derive6 <- 
  aric_paq_v6[,.(ID_C,PAQ1)][aric_derive6,
                             on=.(ID_C)]

aric_derive6[PAQ1 %in% c('B','C'), ambulate_frail := 1]
aric_derive6[PAQ1=='A', ambulate_frail := 0]


## Illness (composite)

#### Hypertension

aric_derive6[!HYPERT64=='T', htn_frail := HYPERT64]

#### Diabetes


aric_derive6[!DIABTS64=="T", dm_frail := as.numeric(DIABTS64)]


#### COPD

aric_derive6 <- 
    aric_status61_v6[,c("ID_C","INCSELFREPLUNG61")][aric_derive6,
    on=.(ID_C)]

aric_derive6 <- 
  aric_derive5[,c("ID_C","copd_frail")][aric_derive6, on=.(ID_C)]


aric_derive6[INCSELFREPLUNG61==1, copd_frail := 1]


aric_derive6[is.na(copd_frail)|
               INCSELFREPLUNG61=='T', copd_frail := 0]

#### Asthma

aric_derive6 <- 
  aric_derive6[aric_status61_v6[,c("ID_C","INCSELFREPAST61")],
                      on=.(ID_C)]

aric_derive6 <- 
    aric_derive5[,c("ID_C","asthma_frail")][aric_derive6,
    on=.(ID_C)]

aric_derive6[INCSELFREPAST61==1, asthma_frail := 1]


aric_derive6[is.na(asthma_frail)|INCSELFREPAST61=='T', asthma_frail := 0]

#### Arthritis

aric_derive6 <- 
    aric_derive5[,c('ID_C','djd_frail')][aric_derive6,
    on=.(ID_C)]

#### CKD

aric_derive6[EGFRCR61>=60, ckd_frail := 0]
aric_derive6[EGFRCR61 <60, ckd_frail := 1]


#### CHF


aric_derive6[,chf_frail := PREVDEFHF61]


#### CAD

aric_derive6[!PRVCHD61=='T', chd_frail := PRVCHD61]


#### Stroke

aric_derive6[!PRVSTR61=='T', stroke_frail := PRVSTR61]


#### Cancer

aric_derive6[, cancer_frail := V6CANCER61]

aric_derive6[,conditions_frail :=
  as.numeric(dm_frail)+
  as.numeric(htn_frail)+
  as.numeric(copd_frail)+
  as.numeric(asthma_frail)+
  as.numeric(djd_frail)+
  as.numeric(ckd_frail)+
  as.numeric(chf_frail)+
  as.numeric(chd_frail)+
  as.numeric(stroke_frail)+
  as.numeric(cancer_frail)]

aric_derive6[conditions_frail <= 4, illness_frail := 0]
aric_derive6[conditions_frail > 4, illness_frail := 1]

aric_derive6[,wtloss_frail := WTLOSSCOMPA61]

aric_derive6[,total_frail :=
  fatigue_frail+
  resistance_frail+
  ambulate_frail+
  illness_frail+
  wtloss_frail]


######### FRIED

aric_ant_v5 <- fread('~/Dropbox/BioLINCC files/ARIC/Main_Study/V5/csv/ant.csv',na.strings=c("NA","","NULL"))
aric_derive6 <- 
  aric_derive6[
aric_ant_v5[,c("ID_C","ANT3")],
                      on=.(ID_C)]

aric_derive6[!is.na(WALK4M61), gaitspeed := 0] 


##### Walking speed

#### Men:
# Height ≤ 173 cm (68.1 in):  ≤0.96 m/s      
# Height >173 cm (68.1 in):   ≤1.04  m/s	
# 


aric_derive6[GENDER=="M"&
               ANT3>173&
               WALK4M61 >= 6, 
             gaitspeed_fried := 1]

aric_derive6[GENDER=="M"&
             ANT3 <= 173&
             WALK4M61 >=7, 
             gaitspeed_fried := 1]


### Women
# Height ≤ 159 cm (62.6 in):  ≤0.90m/s
# Height >159 cm (62.6 in):   ≤1.02 m/s


aric_derive6[GENDER=="F"&
                               ANT3>159&
                               WALK4M61 >= 6, gaitspeed_fried := 1]

aric_derive6[GENDER=="F"&
               ANT3 <= 159&
               WALK4M61 >= 7, 
             gaitspeed_fried := 1]

########### Grip (kg)

# Trials for each hand

## Uses BMI for normalization

##### Men
# BMI ≤ 24.0:     ≤ 29 kg
# BMI 24.1-26.0:  ≤ 30 kg
# BMI 26.1-28.0:  ≤ 30 kg
# BMI > 28.0:     ≤ 32 kg

##### Women
# BMI ≤ 23.0:     ≤ 17
# BMI 23.1-26.0:  ≤ 17.3 kg
# BMI 26.1-29.0:  ≤ 18 kg
# BMI > 29.0:     ≤ 21 kg

aric_derive6 <- 
    aric_pfx_v6[,c("ID_C",
                   "PFX11B",
                   "PFX11C")][aric_derive6,
    on=.(ID_C)]

aric_derive6[,grip_avg := apply(aric_derive6[,c("PFX11B","PFX11C")],MARGIN=1,mean)]
aric_derive6[,grip_max := apply(aric_derive6[,c("PFX11B","PFX11C")],MARGIN=1,max)]

aric_derive6[!is.na(grip_max),grip_fried := 0] 

# Men

aric_derive6[BMI61<=24&
               grip_max <= 29&
               GENDER=="M", 
             grip_fried := 1] 

aric_derive6[BMI61>24&
               BMI61<=26&
               grip_max <= 30&
               GENDER=="M", 
             grip_fried := 1] 

aric_derive6[BMI61>26&
               BMI61<=28&
               grip_max <= 30&
               GENDER=="M", 
             grip_friend := 1] 

aric_derive6[BMI61>28&
               grip_max <= 32&
               GENDER=="M", 
             grip_fried := 1] 

# Women

aric_derive6[BMI61<=23&
               grip_max <= 17&
               GENDER=="F", 
             grip_fried := 1] 

aric_derive6[BMI61>23&
               BMI61<=26&
               grip_max <= 17.3&
               GENDER=="F", 
             grip_fried := 1]

aric_derive6[BMI61>26&
               BMI61<=29&
               grip_max <= 18&
               GENDER=="F", 
             grip_fried := 1] 

aric_derive6[BMI61>29&
               grip_max <= 21&
               GENDER=="F", 
             grip_fried := 1] 


aric_derive6[,activity_fried := LOWENERGYCOMP61]

aric_derive6[,wtloss_fried := WTLOSSCOMPA61]

aric_derive6[,fatigue_fried := EXHAUST61]

aric_derive6[,total_fried := 
  wtloss_fried+
  grip_fried+
  fatigue_fried+
  gaitspeed_fried+
  activity_fried]


# -------------------------------------------------- #


aric_msrcod61_v6 <- fread('~/Dropbox/BioLINCC files/ARIC/Main_Study/V6/csv/msrcod61.csv',na.strings=c("NA","","NULL"))

aric_msrcod61_codes <- aric_msrcod61_v6[,c(1,27:51)]
aric_melt_msrcod61_codes <- melt(aric_msrcod61_codes,id.vars="ID_C",na.rm=T,factorsAsStrings=T)
aric_melt_msrcod61_codes[,variable := "medcode"]
aric_melt_msrcod61_codes[,form := "msrcod"]
aric_melt_msrcod61_codes <- unique(aric_melt_msrcod61_codes)

aric_melt_derive6_v6 <- melt(aric_derive6,id.vars=c("ID_C"),na.rm=T,factorsAsStrings=T)
aric_melt_derive6_v6[,form := "derive6"]
aric_melt_ant_v6 <- melt(aric_ant_v6,id.vars=c("ID_C"),na.rm=T,factorsAsStrings=T)
aric_melt_ant_v6[,form := "ant"]
aric_melt_sbp_v6 <- melt(aric_sbp_v6,id.vars=c("ID_C"),na.rm=T,factorsAsStrings=T)
aric_melt_sbp_v6[,form := "sbp"]
aric_melt_cdi_v6 <- melt(aric_cdi_v6,id.vars=c("ID_C"),na.rm=T,factorsAsStrings=T)
aric_melt_cdi_v6[,form := "cdi"]
aric_melt_ces_v6 <- melt(aric_ces_v6,id.vars=c("ID_C"),na.rm=T,factorsAsStrings=T)
aric_melt_ces_v6[,form := "ces"]
aric_melt_chm_v6 <- melt(aric_chm_v6,id.vars=c("ID_C"),na.rm=T,factorsAsStrings=T)
aric_melt_chm_v6[,form := "chem2"]
aric_melt_egfr_v6 <- melt(aric_egfr_v6,id.vars=c("ID_C"),na.rm=T,factorsAsStrings=T)
aric_melt_egfr_v6[,form := "egfr"]


aric_ess_v6[, ess_total := rowSums(.SD), .SDcols=3:10]

aric_melt_ess_v6 <- melt(aric_ess_v6,id.vars=c("ID_C"),na.rm=T,factorsAsStrings=T)
aric_melt_ess_v6[,form := "ess"]
aric_melt_lip_v6 <- melt(aric_lip_v6,id.vars=c("ID_C"),na.rm=T,factorsAsStrings=T)
aric_melt_lip_v6[,form := "lipf"]
aric_melt_nhx_v6 <- melt(aric_nhx_v6,id.vars=c("ID_C"),na.rm=T,factorsAsStrings=T)
aric_melt_nhx_v6[,form := "nhx"]
aric_melt_pac_v6 <- melt(aric_pac_v6,id.vars=c("ID_C"),na.rm=T,factorsAsStrings=T)
aric_melt_pac_v6[,form := "pac"]
aric_melt_paq_v6 <- melt(aric_paq_v6,id.vars=c("ID_C"),na.rm=T,factorsAsStrings=T)
aric_melt_paq_v6[,form := "paq"]
aric_melt_pfx_v6 <- melt(aric_pfx_v6,id.vars=c("ID_C"),na.rm=T,factorsAsStrings=T)
aric_melt_pfx_v6[,form := "pfx"]
aric_melt_phx_v6 <- melt(aric_phx_v6,id.vars=c("ID_C"),na.rm=T,factorsAsStrings=T)
aric_melt_phx_v6[,form := "phx"]
aric_melt_sfe_v6 <- melt(aric_sfe_v6,id.vars=c("ID_C"),na.rm=T,factorsAsStrings=T)
aric_melt_sfe_v6[,form := "sfe"]
aric_melt_status61_v6 <- melt(aric_status61_v6,id.vars=c("ID_C"),na.rm=T,factorsAsStrings=T)
aric_melt_status61_v6[,form := "status6"]
aric_melt_tmw_v6 <- melt(aric_tmw_v6,id.vars=c("ID_C"),na.rm=T,factorsAsStrings=T)
aric_melt_tmw_v6[,form := "tmw"]

aric_melt_v6_all <- 
  rbindlist(
    list(aric_melt_derive6_v6,
         aric_melt_ant_v6,
         aric_melt_ces_v6,
         aric_melt_chm_v6,
         aric_melt_egfr_v6,
         aric_melt_ess_v6,
         aric_melt_lip_v6,
         aric_melt_nhx_v6,
         aric_melt_pac_v6,
         aric_melt_paq_v6,
         aric_melt_pfx_v6,
         aric_melt_phx_v6,
         aric_melt_sfe_v6,
         aric_melt_status61_v6,
         aric_melt_tmw_v6))

aric_v6_tempdays <- 
  aric_dates[!is.na(V6DATE61_DAYS), .(ID_C, visitdays = V6DATE61_DAYS)]
  

aric_melt_v6_all[,visit := 6]
aric_melt_v6_all <- 
  aric_v6_tempdays[aric_melt_v6_all,on=.(ID_C)]

aric_melt_v6_all[,CONTYR := 28]

rm(list=ls(pattern="\\baric_.*\\_v6$"))

#===============================================================#
####                       ARIC Exam 7                       ####
#===============================================================#

aric_derive7 <- fread('~/Dropbox/BioLINCC files/ARIC/Main_Study/V7/csv/derive71.csv',na.strings=c("NA","","NULL"))

aric_derive7[,drinks_wk := ETHANL71/14]
aric_derive7[,hdl_mgdl := HDLSIU71 * 38.67]


aric_alc_v7 <- fread('~/Dropbox/BioLINCC files/ARIC/Main_Study/V7/csv/alc.csv',na.strings=c("NA","","NULL"))
aric_ant_v7 <- fread('~/Dropbox/BioLINCC files/ARIC/Main_Study/V7/csv/ant.csv',na.strings=c("NA","","NULL"))
aric_cdi_v7 <- fread('~/Dropbox/BioLINCC files/ARIC/Main_Study/V7/csv/cdi.csv',na.strings=c("NA","","NULL"))
aric_cdp_v7 <- fread('~/Dropbox/BioLINCC files/ARIC/Main_Study/V7/csv/cdp.csv',na.strings=c("NA","","NULL"))
aric_cds_v7 <- fread('~/Dropbox/BioLINCC files/ARIC/Main_Study/V7/csv/cds.csv',na.strings=c("NA","","NULL"))
aric_ces_v7 <- fread('~/Dropbox/BioLINCC files/ARIC/Main_Study/V7/csv/ces.csv',na.strings=c("NA","","NULL"))
aric_chm_v7 <- fread('~/Dropbox/BioLINCC files/ARIC/Main_Study/V7/csv/chem2.csv',na.strings=c("NA","","NULL"))
aric_ess_v7 <- fread('~/Dropbox/BioLINCC files/ARIC/Main_Study/V7/csv/ess.csv',na.strings=c("NA","","NULL"))
aric_hne_v7 <- fread('~/Dropbox/BioLINCC files/ARIC/Main_Study/V7/csv/hne.csv',na.strings=c("NA","","NULL"))
aric_lip_v7 <- fread('~/Dropbox/BioLINCC files/ARIC/Main_Study/V7/csv/lipf.csv',na.strings=c("NA","","NULL"))
aric_mme6_v7 <- fread('~/Dropbox/BioLINCC files/ARIC/Main_Study/V7/csv/mme6.csv',na.strings=c("NA","","NULL"))
aric_ncs_v7 <- fread('~/Dropbox/BioLINCC files/ARIC/Main_Study/V7/csv/ncs.csv',na.strings=c("NA","","NULL"))
aric_nhx_v7 <- fread('~/Dropbox/BioLINCC files/ARIC/Main_Study/V7/csv/nhx.csv',na.strings=c("NA","","NULL"))
aric_npi_v7 <- fread('~/Dropbox/BioLINCC files/ARIC/Main_Study/V7/csv/npi.csv',na.strings=c("NA","","NULL"))
aric_pac_v7 <- fread('~/Dropbox/BioLINCC files/ARIC/Main_Study/V7/csv/pfx.csv',na.strings=c("NA","","NULL"))
aric_pfx_v7 <- fread('~/Dropbox/BioLINCC files/ARIC/Main_Study/V7/csv/pfx.csv',na.strings=c("NA","","NULL"))
aric_sbp_v7 <- fread('~/Dropbox/BioLINCC files/ARIC/Main_Study/V7/csv/sbp.csv',na.strings=c("NA","","NULL"))
aric_six7_v7 <- fread('~/Dropbox/BioLINCC files/ARIC/Main_Study/V7/csv/six7.csv',na.strings=c("NA","","NULL"))
aric_status71_v7 <- fread('~/Dropbox/BioLINCC files/ARIC/Main_Study/V7/csv/status71.csv',na.strings=c("NA","","NULL"))
aric_msrcod71_v7 <- fread('~/Dropbox/BioLINCC files/ARIC/Main_Study/V7/csv/msrcod71.csv',na.strings=c("NA","","NULL"))
aric_msrcod71_codes <- aric_msrcod71_v7[,c(1,27:51)]

aric_status71_v7[,AGE_MENOPAUSE_EX07 :=
  apply(aric_status71_v7[,c("AGENATMENOPAUSEF","AGESRGMENOPAUSEF")],
        MARGIN=1,
        FUN=function(x) min(x,na.rm=T))]

aric_status71_v7[AGE_MENOPAUSE_EX07=="Inf", AGE_MENOPAUSE_EX07 := NA]

### Exam 7 FRAIL Score 

## Fatigue

aric_derive7[,fatigue_frail := EXHAUST71]

## Resistance

##### Used SFE question 2b ('Does your health now limit you in climing several flights of stairs?)
##### 1: Yes, limited a lot
##### 2: Yes limited a little
##### 3: No, not limited at all

# aric_genps1$GEN10
# aric_gnbps1$GNB5
# aric_gncps1$GNC2B
# aric_gneps1$GNE4
# aric_paq[,c("PAQ2","PAQ2A")]
# aric_paqa04[,c("PAQA2")]
# aric_sfe$SFE2B

aric_paq_v6 <- fread('~/Dropbox/BioLINCC files/ARIC/Main_Study/V6/csv/paq.csv',na.strings=c("NA","","NULL"))

aric_derive7 <- 
    aric_paq_v6[,c('ID_C','PAQ2')][aric_derive7,
    on=.(ID_C)]

aric_derive7[PAQ2 %in% c('B','C'), resistance_frail := 1]
aric_derive7[PAQ2=='A', resistance_frail := 0]


## Ambulate

aric_derive7 <- 
  aric_gneps1[,c('ID_C','GNE3')][aric_derive7, on=.(ID_C)]

aric_derive7[GNE3 == 0, ambulate_frail := 1]
aric_derive7[GNE3 == 1, ambulate_frail := 0]


# Illness (composite)

#### Hypertension

aric_derive7[aric_derive7$HYPERT74=="T", HYPERT74 := NA]

aric_derive7[,htn_frail := as.numeric(HYPERT74)]

#### Diabetes


aric_derive7[DIABTS74=="T", dm_frail := as.numeric(DIABTS74[!DIABTS74=="T"])]


#### COPD

aric_derive7 <- 
    aric_status71_v7[,c("ID_C","INCSELFREPLUNG71")][aric_derive7,
    on=.(ID_C)]


aric_derive7 <- 
    aric_derive6[,c("ID_C","copd_frail")][aric_derive7,
    on=.(ID_C)]


aric_derive7[INCSELFREPLUNG71==1, copd_frail := 1]


#### Asthma

aric_derive7 <- 
    aric_status71_v7[,c("ID_C","INCSELFREPAST71")][aric_derive7,
    on=.(ID_C)]

aric_derive7 <- 
    aric_derive6[,c("ID_C","asthma_frail")][aric_derive7,
    on=.(ID_C)]


aric_derive7[INCSELFREPAST71==1, asthma_frail := 1]

aric_derive7[is.na(asthma_frail), asthma_frail := 0]


#### Arthritis

aric_derive7 <- 
    aric_derive6[,c('ID_C','djd_frail')][aric_derive7,
    on=.(ID_C)]

#### CKD

aric_derive7[EGFRCR71>=60, ckd_frail := 0]
aric_derive7[EGFRCR71 <60, ckd_frail := 1]


#### CHF


aric_derive7[,chf_frail := PREVDEFHF71]


#### CAD

aric_derive7[!PRVCHD71 == "T", chd_frail := PRVCHD71]

aric_derive7[,chd_frail := as.numeric(chd_frail)]

#### Stroke

aric_derive7[!PRVSTR71=="T", stroke_frail := PRVSTR71]

aric_derive7[, stroke_frail := as.numeric(stroke_frail)]

#### Cancer

aric_derive7 <- 
    aric_derive6[,c("ID_C","V6CANCER61")][aric_derive7,
               on=.(ID_C)]

aric_derive7[,cancer_frail := V6CANCER61]

aric_derive7[,conditions_frail :=
  dm_frail+
  htn_frail+
  copd_frail+
  asthma_frail+
  djd_frail+
  ckd_frail+
  chf_frail+
  chd_frail+
  stroke_frail+
  cancer_frail]

aric_derive7[conditions_frail <= 4, illess_frail := 0]
aric_derive7[conditions_frail > 4, illness_frail := 1]

aric_derive7[,wtloss_frail := WTLOSSCOMPA71]

aric_derive7[,total_frail :=
  fatigue_frail+
  resistance_frail+
  ambulate_frail+
  illness_frail+
  wtloss_frail]


######### FRIED

aric_derive7 <- 
    aric_ant_v5[,c("ID_C","ANT3")][aric_derive7,
    on=.(ID_C)]

aric_derive7[!is.na(WALK4M71), gaitspeed_fried := 0] 


##### Walking speed

#### Men:
# Height ≤ 173 cm (68.1 in):  ≤0.96 m/s      
# Height >173 cm (68.1 in):   ≤1.04  m/s	
# 


aric_derive7[GENDER=="M"&
               ANT3>173&
               WALK4M71 >= 6, 
             gaitspeed_fried := 1]

aric_derive7[GENDER=="M"&
               ANT3 <= 173&
               WALK4M71 >=7, 
             gaitspeed_fried := 1]


### Women
# Height ≤ 159 cm (62.6 in):  ≤0.90m/s
# Height >159 cm (62.6 in):   ≤1.02 m/s


aric_derive7[GENDER=="F"&
               ANT3>159&
               WALK4M71 >= 6, 
             gaitspeed_fried := 1]

aric_derive7[GENDER=="F"&
               ANT3 <= 159&
               WALK4M71 >= 7, 
             gaitspeed_fried := 1]

########### Grip (kg)

# Trials for each hand

## Uses BMI for normalization

##### Men
# BMI ≤ 24.0:     ≤ 29 kg
# BMI 24.1-26.0:  ≤ 30 kg
# BMI 26.1-28.0:  ≤ 30 kg
# BMI > 28.0:     ≤ 32 kg

##### Women
# BMI ≤ 23.0:     ≤ 17
# BMI 23.1-26.0:  ≤ 17.3 kg
# BMI 26.1-29.0:  ≤ 18 kg
# BMI > 29.0:     ≤ 21 kg

aric_derive7 <- 
    aric_pfx_v7[,c("ID_C",
                   "PFX11B",
                   "PFX11C")][aric_derive7,
    on=.(ID_C)]

aric_derive7[,grip_avg := apply(aric_derive7[,c("PFX11B","PFX11C")],MARGIN=1,mean)]
aric_derive7[,grip_max := apply(aric_derive7[,c("PFX11B","PFX11C")],MARGIN=1,max)]

aric_derive7[!is.na(grip_max), grip_fried := 0] 


# Men

aric_derive7[BMI71<=24&
               grip_max <= 29&
               GENDER=="M", 
             grip_fried := 1] 

aric_derive7[BMI71>24&
               BMI71<=26&
               grip_max <= 30&
               GENDER=="M", 
             grip_fried := 1] 

aric_derive7[BMI71>26&
               BMI71<=28&
               grip_max <= 30&
               GENDER=="M", 
             grip_fried := 1] 

aric_derive7[BMI71>28&
               grip_max <= 32&
               GENDER=="M", 
             grip_fried := 1]
# Women

aric_derive7[BMI71<=23&
               grip_max <= 17&
               GENDER=="F", 
             grip_fried := 1] 

aric_derive7[BMI71>23&
               BMI71<=26&
               grip_max <= 17.3&
               GENDER=="F", 
             grip_fried := 1]

aric_derive7[BMI71>26&
               BMI71<=29&
               grip_max <= 18&
               GENDER=="F", 
             grip_fried := 1]

aric_derive7[BMI71>29&
               grip_max <= 21&
               GENDER=="F", 
             grip_fried := 1]


aric_derive7[,activity_fried := LOWENERGYCOMP71]

aric_derive7[,wtloss_fried := WTLOSSCOMPA71]

aric_derive7[,fatigue_fried := EXHAUST71]

aric_derive7[,total_fried := 
  wtloss_fried+
  grip_fried+
  fatigue_fried+
  gaitspeed_fried+
  activity_fried]


# -------------------------------------------- #


aric_melt_msrcod71_codes <- melt(aric_msrcod71_codes,id.vars="ID_C",na.rm=T,factorsAsStrings=T)
aric_melt_msrcod71_codes[,variable := "medcode"]
aric_melt_msrcod71_codes[,form := "msrcod"]
aric_melt_msrcod71_codes <- unique(aric_melt_msrcod71_codes)
aric_melt_derive7_v7 <- melt(aric_derive7,id.vars=c("ID_C"),na.rm=T,factorsAsStrings=T)
aric_melt_derive7_v7[,form := "derive7"]
aric_melt_alc_v7 <- melt(aric_alc_v7,id.vars=c("ID_C"),na.rm=T,factorsAsStrings=T)
aric_melt_alc_v7[,form := "alc"]
aric_melt_ant_v7 <- melt(aric_ant_v7,id.vars=c("ID_C"),na.rm=T,factorsAsStrings=T)
aric_melt_ant_v7[,form := "ant"]
aric_melt_cdi_v7 <- melt(aric_cdi_v7,id.vars=c("ID_C"),na.rm=T,factorsAsStrings=T)
aric_melt_cdi_v7[,form := "cdi"]
aric_melt_cdp_v7 <- melt(aric_cdp_v7,id.vars=c("ID_C"),na.rm=T,factorsAsStrings=T)
aric_melt_cdp_v7[,form := "cdp"]
aric_melt_cds_v7 <- melt(aric_cds_v7,id.vars=c("ID_C"),na.rm=T,factorsAsStrings=T)
aric_melt_cds_v7[,form := "cds"]
aric_melt_ces_v7 <- melt(aric_ces_v7,id.vars=c("ID_C"),na.rm=T,factorsAsStrings=T)
aric_melt_ces_v7[,form := "ces"]
aric_melt_chm_v7 <- melt(aric_chm_v7,id.vars=c("ID_C"),na.rm=T,factorsAsStrings=T)
aric_melt_chm_v7[,form := "chem2"]
aric_melt_ess_v7 <- melt(aric_ess_v7,id.vars=c("ID_C"),na.rm=T,factorsAsStrings=T)
aric_melt_ess_v7[,form := "ess"]
aric_melt_lip_v7 <- melt(aric_lip_v7,id.vars=c("ID_C"),na.rm=T,factorsAsStrings=T)
aric_melt_lip_v7[,form := "lipf"]
aric_melt_mme6_v7 <- melt(aric_mme6_v7,id.vars=c("ID_C"),na.rm=T,factorsAsStrings=T)
aric_melt_mme6_v7[,form := "mme6"]
aric_melt_ncs_v7 <- melt(aric_ncs_v7,id.vars=c("ID_C"),na.rm=T,factorsAsStrings=T)
aric_melt_ncs_v7[,form := "ncs"]
aric_melt_nhx_v7 <- melt(aric_nhx_v7,id.vars=c("ID_C"),na.rm=T,factorsAsStrings=T)
aric_melt_nhx_v7[,form := "nhx"]
aric_melt_npi_v7 <- melt(aric_npi_v7,id.vars=c("ID_C"),na.rm=T,factorsAsStrings=T)
aric_melt_npi_v7[,form := "npi"]
aric_melt_pfx_v7 <- melt(aric_pfx_v7,id.vars=c("ID_C"),na.rm=T,factorsAsStrings=T)
aric_melt_pfx_v7[,form := "pfx"]
aric_melt_sbp_v7 <- melt(aric_sbp_v7,id.vars=c("ID_C"),na.rm=T,factorsAsStrings=T)
aric_melt_sbp_v7[,form := "sbp"]
aric_melt_six7_v7 <- melt(aric_six7_v7,id.vars=c("ID_C"),na.rm=T,factorsAsStrings=T)
aric_melt_six7_v7[,form := "sfe"]
aric_melt_status71_v7 <- melt(aric_status71_v7,id.vars=c("ID_C"),na.rm=T,factorsAsStrings=T)
aric_melt_status71_v7[,form := "status71"]

aric_melt_v7_all <- 
  rbindlist(
    list(aric_melt_derive7_v7,
                          aric_melt_alc_v7,
                          aric_melt_ant_v7,
                          aric_melt_cdi_v7,
                          aric_melt_cdp_v7,
                          aric_melt_cds_v7,
                          aric_melt_ces_v7,
                          aric_melt_chm_v7,
                          aric_melt_ess_v7,
                          aric_melt_lip_v7,
                          aric_melt_mme6_v7,
                          aric_melt_ncs_v7,
                          aric_melt_nhx_v7,
                          aric_melt_npi_v7,
                          aric_melt_pfx_v7,
                          aric_melt_six7_v7,
                          aric_melt_status71_v7))

aric_v7_tempdays <- 
  aric_dates[!is.na(V7DATE71_FOLLOWUPDAYS),.(ID_C, visitdays = V7DATE71_FOLLOWUPDAYS)]

aric_melt_v7_all[,visit := 7]
aric_melt_v7_all <- aric_v7_tempdays[aric_melt_v7_all, on=.(ID_C)]

aric_melt_v7_all[,CONTYR := 30]

rm(list=ls(pattern="\\baric_.*\\_v7$"))
rm(list=ls(pattern="\\baric_.*\\_codes$"))
rm(list=ls(pattern="\\baric_.*\\_tempdays$"))
rm(list=ls(pattern="\\baric_.*\\_ref$"))


#===============================================================#
####                       ARIC Exam 8T                       ####
#===============================================================#

aric_derive8 <- fread('~/Dropbox/BioLINCC files/ARIC/Main_Study/V8T/csv/derive8t1_np.csv',na.strings=c("NA","","NULL"))


aric_cdi_v8 <- fread('~/Dropbox/BioLINCC files/ARIC/Main_Study/V8T/csv/cdi_np.csv',na.strings=c("NA","","NULL"))
aric_cdpt_v8 <- fread('~/Dropbox/BioLINCC files/ARIC/Main_Study/V8T/csv/cdpt_np.csv',na.strings=c("NA","","NULL"))
aric_cds_v8 <- fread('~/Dropbox/BioLINCC files/ARIC/Main_Study/V8T/csv/cds_np.csv',na.strings=c("NA","","NULL"))
aric_cest_v8 <- fread('~/Dropbox/BioLINCC files/ARIC/Main_Study/V8T/csv/cest_np.csv',na.strings=c("NA","","NULL"))
aric_dcf8t_v8 <- fread('~/Dropbox/BioLINCC files/ARIC/Main_Study/V8T/csv/dcf8t_np.csv',na.strings=c("NA","","NULL"))
aric_esut_v8 <- fread('~/Dropbox/BioLINCC files/ARIC/Main_Study/V8T/csv/esut_np.csv',na.strings=c("NA","","NULL"))
aric_mcht_v8 <- fread('~/Dropbox/BioLINCC files/ARIC/Main_Study/V8T/csv/mcht_np.csv',na.strings=c("NA","","NULL"))
aric_ncst_v8 <- fread('~/Dropbox/BioLINCC files/ARIC/Main_Study/V8T/csv/ncst_np.csv',na.strings=c("NA","","NULL"))
aric_ncstderv8t1_v8 <- fread('~/Dropbox/BioLINCC files/ARIC/Main_Study/V8T/csv/ncstderv8t1_np.csv',na.strings=c("NA","","NULL"))
aric_npi_v8 <- fread('~/Dropbox/BioLINCC files/ARIC/Main_Study/V8T/csv/npi_np.csv',na.strings=c("NA","","NULL"))
aric_psq_v8 <- fread('~/Dropbox/BioLINCC files/ARIC/Main_Study/V8T/csv/psq_np.csv',na.strings=c("NA","","NULL"))
aric_pwp_v8 <- fread('~/Dropbox/BioLINCC files/ARIC/Main_Study/V8T/csv/pwp_np.csv',na.strings=c("NA","","NULL"))
aric_pwx_v8 <- fread('~/Dropbox/BioLINCC files/ARIC/Main_Study/V8T/csv/pwx_np.csv',na.strings=c("NA","","NULL"))
aric_status81_v8 <- fread('~/Dropbox/BioLINCC files/ARIC/Main_Study/V8T/csv/status81_np.csv',na.strings=c("NA","","NULL"))
aric_v2_v8cnf_v8 <- fread('~/Dropbox/BioLINCC files/ARIC/Main_Study/V8T/csv/v2_v8_cnfa_np.csv',na.strings=c("NA","","NULL"))

aric_status81_v8[,AGE_MENOPAUSE_EX08 :=
  apply(aric_status81_v8[,c("AGENATMENOPAUSEF","AGESRGMENOPAUSEF")],
        MARGIN=1,
        FUN=function(x) min(x,na.rm=T))]

aric_status81_v8[AGE_MENOPAUSE_EX08=="Inf",AGE_MENOPAUSE_EX08 := NA]

aric_melt_derive8_v8 <- melt(aric_derive8,id.vars=c("ID_C"),na.rm=T,factorsAsStrings=T)
aric_melt_derive8_v8[,form := "derive8"]
aric_melt_cdi_v8 <- melt(aric_cdi_v8,id.vars=c("ID_C"),na.rm=T,factorsAsStrings=T)
aric_melt_cdi_v8[,form := "cdi"]
aric_melt_cdpt_v8 <- melt(aric_cdpt_v8,id.vars="ID_C",na.rm=T,factorsAsStrings=T)
aric_melt_cdpt_v8[,form := "cdpt"]
aric_melt_cds_v8 <- melt(aric_cds_v8,id.vars="ID_C",na.rm=T,factorsAsStrings=T)
aric_melt_cds_v8[,form := "cds"]
aric_melt_cest_v8 <- melt(aric_cest_v8,id.vars="ID_C",na.rm=T,factorsAsStrings=T)
aric_melt_cest_v8[,form := "cest"]
aric_melt_dcf8t_v8 <- melt(aric_dcf8t_v8,id.vars="ID_C",na.rm=T,factorsAsStrings=T)
aric_melt_dcf8t_v8[,form := "dcf8t"]
aric_melt_dcf8t_v8 <- melt(aric_dcf8t_v8,id.vars="ID_C",na.rm=T,factorsAsStrings=T)
aric_melt_dcf8t_v8[,form := "dcf8t"]
aric_melt_esut_v8 <- melt(aric_esut_v8,id.vars="ID_C",na.rm=T,factorsAsStrings=T)
aric_melt_esut_v8[,form := "esut"]
aric_melt_mcht_v8 <- melt(aric_mcht_v8,id.vars="ID_C",na.rm=T,factorsAsStrings=T)
aric_melt_mcht_v8[,form := "mcht"]
aric_melt_ncst_v8 <- melt(aric_ncst_v8,id.vars="ID_C",na.rm=T,factorsAsStrings=T)
aric_melt_ncst_v8[,form := "ncst"]
aric_melt_ncstderv8t1_v8 <- melt(aric_ncstderv8t1_v8,id.vars="ID_C",na.rm=T,factorsAsStrings=T)
aric_melt_ncstderv8t1_v8[,form := "ncstderv8t1"]
aric_melt_npi_v8 <- melt(aric_npi_v8,id.vars="ID_C",na.rm=T,factorsAsStrings=T)
aric_melt_npi_v8[,form := "npi"]
aric_melt_psq_v8 <- melt(aric_psq_v8,id.vars="ID_C",na.rm=T,factorsAsStrings=T)
aric_melt_psq_v8[,form := "psq"]
aric_melt_pwp_v8 <- melt(aric_pwp_v8,id.vars="ID_C",na.rm=T,factorsAsStrings=T)
aric_melt_pwp_v8[,form := "pwp"]
aric_melt_pwx_v8 <- melt(aric_pwx_v8,id.vars="ID_C",na.rm=T,factorsAsStrings=T)
aric_melt_pwx_v8[,form := "pwx"]
aric_melt_status81_v8 <- melt(aric_status81_v8,id.vars="ID_C",na.rm=T,factorsAsStrings=T)
aric_melt_status81_v8[,form := "status81"]
aric_melt_status81_v8 <-
  aric_melt_status81_v8[ID_C %in% aric_derive8$ID_C,]
aric_melt_v2_v8cnf_v8 <- melt(aric_v2_v8cnf_v8,id.vars="ID_C",na.rm=T,factorsAsStrings=T)
aric_melt_v2_v8cnf_v8[,form := "v2_v8cnf"]
aric_melt_v2_v8cnf_v8 <-
  aric_melt_v2_v8cnf_v8[ID_C %in% aric_derive8$ID_C,]

aric_melt_v8_all <- 
  rbindlist(
    list(aric_melt_derive8_v8,
         aric_melt_cdi_v8,
         aric_melt_cdpt_v8,
         aric_melt_cds_v8,
         aric_melt_cest_v8,
         aric_melt_dcf8t_v8,
         aric_melt_esut_v8,
         aric_melt_mcht_v8,
         aric_melt_ncst_v8,
         aric_melt_ncstderv8t1_v8,
         aric_melt_npi_v8,
         aric_melt_psq_v8,
         aric_melt_pwp_v8,
         aric_melt_pwx_v8,
         aric_melt_status81_v8,
         aric_melt_v2_v8cnf_v8))

aric_v8_tempdays <- 
  aric_derive8[!is.na(V8TDATE8T1_FOLLOWUPDAYS),.(ID_C,V8TDATE8T1_FOLLOWUPDAYS)]
names(aric_v8_tempdays)[2] <- "visitdays"

aric_melt_v8_all[,visit := 8]
aric_melt_v8_all <- 
  aric_v8_tempdays[aric_melt_v8_all,
                   on=.(ID_C)]

aric_melt_v8_all[,CONTYR := 33]

rm(list=ls(pattern="\\baric_.*\\_v8$"))
rm(list=ls(pattern="\\baric_.*\\_codes$"))
rm(list=ls(pattern="\\baric_.*\\_tempdays$"))
rm(list=ls(pattern="\\baric_.*\\_ref$"))


# ==========================================================================#
##                                                                         ##
####                            ARIC FOLLOW-UP                           ####
##                                                                         ##
# ==========================================================================#

aric_afu <- fread('~/Dropbox/BioLINCC files/ARIC/Main_Study/AFU/csv/afucomposite32ps1.csv',
                  na.strings=c("NA","","NULL"))

aric_afu[,nas := apply(aric_afu,MARGIN=1,FUN=function(x) sum(is.na(x)))]

aric_afu_comp <- 
  aric_afu[ , .SD[which.min(nas)], by = .(ID_C, AGE,AFUCOMP1_A_YEAR,CONTYR)]

aric_melt_afu <- melt(aric_afu_comp,
                      id.vars=c("ID_C",
                                "AFUCOMP1_A_FOLLOWUPDAYS",
                                "CONTYR",
                                "AGE"),
                      na.rm=T,
                      stringsAsFactors=T)


aric_melt_afu[,form := "afu"]

aric_melt_marital_gap <- 
  aric_melt_afu[
    variable=="AFUCOMP40A_D"&
      CONTYR %in% c(4,7,23,28,30),]

aric_melt_afu <- 
  aric_melt_afu[!CONTYR %in% c(4,7,10,23,28,30),]


aric_melt_afu <- 
  rbindlist(
    list(aric_melt_afu,
                       aric_melt_marital_gap))

names(aric_melt_afu)[1:7] <- 
  c("patientid","visitdays","CONTYR","age_obs","variable","value","form")

rm(aric_afu,
   aric_afu_comp)

#===============================================================#
####                   Combine ARIC exams                    ####
#===============================================================#

aric_melt_all <- 
  rbindlist(
    list(aric_melt_v1_all[,c("ID_C","CONTYR","visit","visitdays","variable","value","form")],
         aric_melt_v2_all[,c("ID_C","CONTYR","visit","visitdays","variable","value","form")],
         aric_melt_v3_all[,c("ID_C","CONTYR","visit","visitdays","variable","value","form")],
         aric_melt_v4_all[,c("ID_C","CONTYR","visit","visitdays","variable","value","form")],
         aric_melt_v5_all[,c("ID_C","CONTYR","visit","visitdays","variable","value","form")],
         aric_melt_v6_all[,c("ID_C","CONTYR","visit","visitdays","variable","value","form")],
         aric_melt_v7_all[,c("ID_C","CONTYR","visit","visitdays","variable","value","form")],
         aric_melt_v8_all[,c("ID_C","CONTYR","visit","visitdays","variable","value","form")],
         aric_analytes_long[,c("ID_C","CONTYR","visit","visitdays","variable","value","form")]))

names(aric_melt_all)[1] <- "patientid"

aric_melt_all[,variable := as.character(variable)]

aric_bl_age <- aric_derive1[,.(ID_C,V1AGE01,GENDER)]
setnames(aric_bl_age,c("ID_C","V1AGE01","GENDER"),c("patientid","bl_age","sex"))

aric_melt_all <-
  aric_melt_all[!variable=="FORPROFIT",]

setnames(aric_melt_all,"CONTYR","visit_yr")
setnames(aric_melt_afu,"CONTYR","visit_yr")

aric_melt_all <- 
  rbindlist(
    list(aric_melt_all[,c("patientid","visitdays","visit_yr","form","variable","value")],
         aric_melt_afu[,c("patientid","visitdays","visit_yr","form","variable","value")],
         aric_melt_gnps[,c("patientid","visitdays","visit_yr","form","variable","value")]))

aric_melt_all[,study := "ARIC"]

aric_melt_all[,variable := as.character(variable)]


# Age menopause

aric_melt_menopause_parts <-
  aric_melt_all[
         variable %in% c("RHXA01",             # menarche
                         "RHXA08",              # Visit 1 answer
                         "HHXB19",              # Visit 2
                         "RHXB7",               # Visit 3
                         "RHXC7",               # Visit 4
                         "AGE_MENOPAUSE_EX05",  # Visit 5
                         "AGE_MENOPAUSE_EX06",  # Visit 6
                         "AGE_MENOPAUSE_EX07",   # Visit 7
                         "AGE_MENOPAUSE_EX08",   # Visit 8
                         "RHXA07", # Reported menopause at Visit 1
                         "HHXB18", # Reported at visit 2
                         "RHXB6",  # Reported at visit 3
                         "RHXC6" # Reported at visit 4
         ),
         ]

aric_menopause_parts <- 
  dcast(aric_melt_menopause_parts,
        patientid~variable,
        value.var="value")

# Menopause reported

aric_menopause_parts[,meno_yn := "No"]
aric_menopause_parts[RHXA07=="Y",meno_yn := "Yes"]
aric_menopause_parts[HHXB18=="Y",meno_yn := "Yes"]
aric_menopause_parts[RHXB6=="Y",meno_yn := "Yes"]
aric_menopause_parts[RHXC6=="Y",meno_yn := "Yes"]
aric_menopause_parts[!is.na(AGE_MENOPAUSE_EX05),meno_yn := "Yes"]
aric_menopause_parts[!is.na(AGE_MENOPAUSE_EX06),meno_yn := "Yes"]
aric_menopause_parts[!is.na(AGE_MENOPAUSE_EX07),meno_yn := "Yes"]
aric_menopause_parts[!is.na(AGE_MENOPAUSE_EX08),meno_yn := "Yes"]


aric_menopause_parts[,RHXA01 := as.numeric(RHXA01)]
aric_menopause_parts[,RHXA08 := as.numeric(RHXA08)]
aric_menopause_parts[,HHXB19 := as.numeric(HHXB19)]
aric_menopause_parts[,RHXB7 := as.numeric(RHXB7)]
aric_menopause_parts[,RHXC7 := as.numeric(RHXC7)]
aric_menopause_parts[,AGE_MENOPAUSE_EX05 := as.numeric(AGE_MENOPAUSE_EX05)]
aric_menopause_parts[,AGE_MENOPAUSE_EX06 := as.numeric(AGE_MENOPAUSE_EX06)]
aric_menopause_parts[,AGE_MENOPAUSE_EX07 := as.numeric(AGE_MENOPAUSE_EX07)]
aric_menopause_parts[,AGE_MENOPAUSE_EX07 := as.numeric(AGE_MENOPAUSE_EX08)]

aric_menopause_parts[,age_menopause := AGE_MENOPAUSE_EX08]
aric_menopause_parts[,age_menopause := AGE_MENOPAUSE_EX07]
aric_menopause_parts[is.na(age_menopause), age_menopause :=  AGE_MENOPAUSE_EX06]
aric_menopause_parts[is.na(age_menopause), age_menopause := AGE_MENOPAUSE_EX05]
aric_menopause_parts[is.na(age_menopause), age_menopause := RHXC7]
aric_menopause_parts[is.na(age_menopause), age_menopause := RHXB7]
aric_menopause_parts[is.na(age_menopause), age_menopause := HHXB19]
aric_menopause_parts[is.na(age_menopause), age_menopause := RHXA08]

aric_menopause_parts[
  age_menopause > (RHXA01+3)&!is.na(age_menopause)&!is.na(RHXA01), 
  years_menses := age_menopause - RHXA01]

aric_hrt <- 
  aric_melt_all[
    variable %in% 
      c("RHXA19","RHXA21",  # Visit 1 Hormone 1
        "RHXA26","RHXA28", # Visit 1 Hormone 2
        "RHXA33","RHXA35", # Visit 1 Hormone 3
        "RHXA40","RHXA42", # Visit 1 Hormone 4
        "HHXB25","HHXB27", # Visit 2 Hormone 1
        "HHXB32","HHXB34", # Visit 2 Hormone 2
        "RHXB13","RHXB15", # VISIT 3 Hormone 1
        "RHXB25","RHXB27", # Visit 3 Hormone 2
        "RHXC13","RHXC15", # Visit 4 Hormone 1
        "RHXC25","RHXC27"),  # Visit 4 Hormone 2
  ]


aric_hrt <- 
  dcast(aric_hrt,
        study+patientid~variable,
        value.var="value")

aric_hrt[, (3:22) := lapply(.SD, as.numeric), .SDcols=3:22]

cols <- c("RHXA19","RHXA26","RHXA33","RHXA40","HHXB25","HHXB32",
          "RHXB13","RHXB25","RHXC13","RHXC25")

aric_hrt[, hrt_start := do.call(pmin, c(.SD, list(na.rm = TRUE))), .SDcols = cols]

cols <- c("RHXA21",
          "RHXA28",
          "RHXA35",
          "RHXA42",
          "HHXB27",
          "HHXB34",
          "RHXB15",
          "RHXB27",
          "RHXC15",
          "RHXC27")

aric_hrt[, hrt_stop := do.call(pmax, c(.SD, list(na.rm = TRUE))), .SDcols = cols]


aric_hrt[hrt_stop == -Inf, hrt_stop := NA]

aric_hrt <- 
  aric_bl_age[aric_hrt, on = .(patientid)]

# setnames(aric_hrt,"ID_C","patientid")

aric_hrt <-
    aric_outcomes[,.(patientid,dth_dt)][aric_hrt, 
    on = .(patientid)]

aric_hrt[,age_eos := floor(dth_dt/365) + bl_age]

aric_hrt[!is.na(hrt_start)&is.na(hrt_stop), hrt_stop := age_eos]

aric_hrt[,years_hrt := hrt_stop-hrt_start]

aric_hrt <- aric_hrt[years_hrt >= 0,]

aric_menopause_parts <-
  aric_hrt[aric_menopause_parts,
        on=.(patientid)]

aric_menopause_parts[RHXA19 < age_menopause-3, RHXA19 := NA]
aric_menopause_parts[RHXA26 < age_menopause-3, RHXA26 := NA]
aric_menopause_parts[RHXA33 < age_menopause-3, RHXA33 := NA]
aric_menopause_parts[RHXA40 < age_menopause-3, RHXA40 := NA]
aric_menopause_parts[HHXB25 < age_menopause-3, HHXB25 := NA]
aric_menopause_parts[HHXB32 < age_menopause-3, HHXB32 := NA]
aric_menopause_parts[RHXB13 < age_menopause-3, RHXB13 := NA]
aric_menopause_parts[RHXB25 < age_menopause-3, RHXB25 := NA]
aric_menopause_parts[RHXC13 < age_menopause-3, RHXC13 := NA]
aric_menopause_parts[RHXC25 < age_menopause-3, RHXC25 := NA]
aric_menopause_parts[RHXA21 < age_menopause-3, RHXA21 := NA]
aric_menopause_parts[RHXA28 < age_menopause-3, RHXA28 := NA]
aric_menopause_parts[RHXA35 < age_menopause-3, RHXA35 := NA]
aric_menopause_parts[RHXA42 < age_menopause-3, RHXA42 := NA]
aric_menopause_parts[HHXB27 < age_menopause-3, HHXB27 := NA]
aric_menopause_parts[HHXB34 < age_menopause-3, HHXB34 := NA]
aric_menopause_parts[RHXB15 < age_menopause-3, RHXB15 := NA]
aric_menopause_parts[RHXB27 < age_menopause-3, RHXB27 := NA]
aric_menopause_parts[RHXC15 < age_menopause-3, RHXC15 := NA]
aric_menopause_parts[RHXC27 < age_menopause-3, RHXC27 := NA]


aric_melt_menses <-
  melt(aric_menopause_parts[,
                            c("patientid",
                              "meno_yn",
                              "age_menopause",
                              "years_menses",
                              "hrt_start",
                              "hrt_stop",
                              "years_hrt")],
       id.vars=c("patientid"))

aric_melt_menses[,visit := 1]
aric_melt_menses[,form := "aric_menses"]
aric_melt_menses[,visitdays := 0]
aric_melt_menses[,visit_yr := 0]

aric_melt_all <-
  rbindlist(
    list(aric_melt_all[,c("patientid","visit_yr","visitdays","variable","value","form")],
        aric_melt_menses[,c("patientid","visit_yr","visitdays","variable","value","form")]))


aric_melt_all[,study := "ARIC"]

gc()

aric_melt_all[,form := toupper(form)]
aric_melt_all[,variable := toupper(variable)]

aric_melt_all[,study_field := 
                paste(study,
                      form,
                      variable,sep="_")]
              
# rm(aric_melt_afu,
#    aric_melt_gnps,
#    aric_analytes_long)


# aric_dates_long <- 
#   read_parquet("~/Dropbox/ADAPT-HF/Master HDCP files/Cohort data/ptlist.parquet")

aric_dates_long[,study:="ARIC"]

setnames(aric_dates_long,'ID_C','patientid')

aric_melt_all[,visit_yr := as.numeric(visit_yr)]
aric_melt_all <-
  aric_bl_age[aric_melt_all,on=.(patientid)]

aric_melt_all[,age_obs := bl_age + visit_yr]

# aric_melt_all <-
#         aric_dates_long[,c("patientid",
#                            "visit_yr",
#                            "age_obs",
#                            "cohort",
#                            "cohort_name")][aric_melt_all,
#         on=.(patientid,visit_yr)]

aric_melt_all[,datapoint :=
  paste("ARIC",row.names(aric_melt_all),sep="")]

cohort_name <- unique(aric_dates_long[,.(patientid, cohort, cohort_name)])

aric_melt_all <-
  cohort_name[aric_melt_all,on=.(patientid)]

aric_data_fields <- unique(aric_melt_all$study_field)

write_parquet(aric_melt_all[,..data_fields],
              "~/Dropbox/ADAPT-HF/Master HDCP files/Cohort data/aric_melt_all.parquet")

gcs_auth("~/Dropbox/ADAPT-HF/Master HDCP files/harmonization-286013-39f492122f69.json")
gcs_upload(aric_melt_all[,..data_fields], 
           bucket="master_hdcp_files",
           name="aric_melt_all.parquet",
           object_function = f)

rm(list=ls(pattern="\\baric."))

gc()
```
