## Supplementary material for "Aggregating data to accelerate personalized therapy in heart failure (ADAPT-HF)": Harmonization scripts: Compile CHS data table.R

```
library(Hmisc)
library(plotrix)
library(descr)
library(dplyr)
library(relper)
library(stringr)
library(bigrquery)
library(haven)
library(googleCloudStorageR)
library(data.table)
library(arrow)
library(parquetize)
library(bigQueryR)

source("~/Dropbox/R scripts/Misc clinical scripts.R")
source("~/Dropbox/R scripts/echo scripts.R")


outcomes_list <- c("dth_status","dth_dt",
                   "cvdth_status","cvdth_dt",
                   "hfdth_status","hfdth_dt",
                   "hfhosp_ef_val","hfhosp_ef_cat",
                   "noncvdth_status","noncvdth_dt",
                   "hosp_status","hosp_dt",
                   "cadhosp_status","cadhosp_dt",
                   "cvhosp_status","cvhosp_dt",
                   "hfhosp_status","hfhosp_dt",
                   "noncvhosp_status","noncvhosp_dt")


visit_yrs <- fread('~/Dropbox/ADAPT-HF/Master HDCP files/Cohort visit yrs.csv')

data_fields <- 
  c("study",
    "patientid",
    "visit_yr",
    "visitdays",
    "form",
    "variable",
    "value",
    "datapoint",
    "study_field",
    "cohort",
    "cohort_name",
    "age_obs")

outcomes_list <- c("dth_status","dth_dt",
                   "cvdth_status","cvdth_dt",
                   "hfdth_status","hfdth_dt",
                   "hfhosp_ef_val","hfhosp_ef_cat",
                   "noncvdth_status","noncvdth_dt",
                   "hosp_status","hosp_dt",
                   "cadhosp_status","cadhosp_dt",
                   "cvhosp_status","cvhosp_dt",
                   "hfhosp_status","hfhosp_dt",
                   "noncvhosp_status","noncvhosp_dt")

dates_long_fields <-
  c("study",
    "cohort",
    "cohort_name",
    "patientid",
    "sex",
    "visit_yr",
    "visitdays",
    "age_obs")


#### ************************ CHS ************************ #### 

# ..%%%%%%..%%.....%%..%%%%%%.
# .%%....%%.%%.....%%.%%....%%
# .%%.......%%.....%%.%%......
# .%%.......%%%%%%%%%..%%%%%%.
# .%%.......%%.....%%.......%%
# .%%....%%.%%.....%%.%%....%%
# ..%%%%%%..%%.....%%..%%%%%%.


# ==============================================================##
####                       CHS Outcomes                       ####
# ==============================================================##


chs_events <- fread("~/Dropbox/BioLINCC files/CHS/Main_Study/EVENTS/events.csv")
chs_drhosp11 <- fread("~/Dropbox/BioLINCC files/CHS/Main_Study/ICD9/drhosp16.csv")

chs_drhosp11[chs_drhosp11==""] <- NA

chs_censor <- 
  unique(chs_events[, .(newid, dth_status = death, dth_dt = censtime)])


chs_fatalevents <- 
  unique(chs_events[fatal==1, .(newid, censtime , cause80, mechan80)])

chs_fatalevents[cause80 %in% c(1,2,3,4), cvdth_status := 1]
chs_fatalevents[cause80 %in% c(5), noncvdth_status := 1]
chs_fatalevents[mechan80 %in% c(3), hfdth_status := 1]

chs_fatalevents[is.na(cvdth_status), cvdth_status := 0]
chs_fatalevents[is.na(noncvdth_status)] <- 0
chs_fatalevents[is.na(hfdth_status), hfdth_status := 0]

chs_fatalevents <-
  chs_fatalevents[,.(newid,
                   cvdth_status,
                   cvdth_dt = censtime, 
                   noncvdth_status,
                   noncvdth_dt = censtime,
                   hfdth_status,
                   hfdth_dt = censtime)] 


chs_incidenthf <- 
  chs_events[evtype == 4, 
             .(hfhosp_status = 1, 
               hfhosp_dt = min(ttoevent)), 
             by = newid]


chs_incidentcad <- 
  chs_events[evtype %in% c(1,2,7,8,10,11), 
             .(cadhosp_status = 1, 
               cadhosp_dt = min(ttoevent)), 
             by = newid]


chs_incidentcva <- 
  chs_events[evtype == (3), 
             .(cvahosp_status = 1, 
               cvahosp_dt = min(ttoevent)), 
             by = newid]


chs_outcomes <- chs_censor[chs_fatalevents,on=.(newid)]
chs_outcomes <- chs_outcomes[chs_incidenthf,on=.(newid)]
chs_outcomes <- chs_outcomes[chs_incidentcad,on=.(newid)]
chs_outcomes <- chs_outcomes[chs_incidentcva,on=.(newid)]

chs_outcomes[is.na(hfhosp_status), hfhosp_status := 0]

chs_outcomes[chs_outcomes$hfhosp_status==1, 
             time_hfhosp_to_eos := 
               dth_dt-hfhosp_dt]
             
chs_outcomes[hfhosp_status==0,hfhosp_dt := dth_dt]

chs_outcomes[is.na(cadhosp_status), cadhosp_status := 0]
chs_outcomes[cadhosp_status==1,
             time_cad_to_eos := dth_dt - cadhosp_dt]

chs_outcomes[cadhosp_status==0,cadhosp_dt :=  dth_dt]

chs_outcomes[is.na(cvahosp_status), cvahosp_status := 0]
chs_outcomes[cvahosp_status==1, time_cva_to_eos := dth_dt - cvahosp_dt]
chs_outcomes[cvahosp_status==0, cvahosp_dt := dth_dt]


chs_outcomes[,study := "CHS"]

names(chs_outcomes)[1] <- "patientid"
outcomes_to_fill <- outcomes_list[!outcomes_list %in% names(chs_outcomes)]
chs_outcomes[,outcomes_to_fill] <- NA
chs_outcomes[, (20:27) := lapply(.SD, as.numeric), .SDcols = 20:27]

write_parquet(chs_outcomes,
              "~/Dropbox/ADAPT-HF/Master HDCP files/Cohort data/chs_outcomes.parquet")

#===============================================================#
####                   CHS baseline tables                   ####
#===============================================================#


chs_base1 <- fread("~/Dropbox/BioLINCC files/CHS/Main_Study/Baseline/base1.csv",na.strings=c("NA","","NULL"))
names(chs_base1) <- toupper(names(chs_base1))
chs_base2 <- fread("~/Dropbox/BioLINCC files/CHS/Main_Study/Baseline/base2.csv",na.strings=c("NA","","NULL"))
names(chs_base2) <- toupper(names(chs_base2))
chs_baseboth <- fread("~/Dropbox/BioLINCC files/CHS/Main_Study/Baseline/baseboth.csv",na.strings=c("NA","","NULL"))
names(chs_baseboth) <- toupper(names(chs_baseboth))

chs_base1_2019 <- fread("~/Dropbox/BioLINCC files/CHS/Main_Study/Baseline/base1_2019.csv",na.strings=c("NA","","NULL"))

chs_hisp <- chs_base1_2019[,
                           c("IDNO","HISP01")]

names(chs_hisp)[1] <- "NEWID"

chs_ntbnp <- 
  fread("~/Dropbox/BioLINCC files/CHS/ANCILLARY_STUDIES/DeFilippi_(ProBNP)/ntprobnpdatafinal.csv",
        na.strings=c("NA","","NULL"))

chs_base1 <- chs_hisp[chs_base1,on=.(NEWID)]

chs_baseboth[,agey := 2*(AGE2-1)+64]
chs_baseboth <-chs_hisp[chs_baseboth,on=.(NEWID)]
chs_baseboth[HISP01==1, RACE01 := 8]
chs_baseboth[,weight_kg := WEIGHT/2.2046226218]


chs_base1[,lsns_dep := HLPSHP03]
chs_base1[chs_base1$RELYON03==5,lsns_dep := 5]


chs_illness_bl <-
  chs_baseboth[,c("NEWID",
                  "PERSTAT",
                  "GEND01",
                  "RACE01",
                  "STHT",
                  "ANBLMOD",
                  "CHBLMOD",
                  "CHDBLMOD",
                  "MIBLMOD",
                  "STBLMOD",
                  "TIBLMOD",
                  "DIABADA",
                  "HYPERADJ",  ## Changed name
                  "COPDBL",
                  "CYSGFRBL",  ## Changed name
                  "ARTH01",
                  "ASTHCUR", ## Changed name
                  "DIAG01")]


names(chs_illness_bl)[3:18] <- 
  c("gender_ref",
    "race_ref",
    "height_cm",
    "ang_bl",
    "chf_bl",
    "cad_bl",
    "mi_bl",
    "cva_bl",
    "tia_bl",
    "dm_bl",
    "htn_bl",
    "copd_bl",
    "gfr_bl",
    "arth_bl",
    "asthma_bl",
    "cancer_bl") 

### Base 1 FRAIL

####### FRAIL score

## Fatigue, Resistance, Ambulation encoded in 'criteriaset'

chs_base1[EFFORT05 %in% c(2,3)|GETGO05 %in% c(2,3), fatigue_frail := 1]
chs_base1[EFFORT05 %in% c(0,1)|GETGO05 %in% c(0,1), fatigue_frail := 0]

## Resistance

chs_base1[,resistance_frail := STEPS09]

## Ambulate

chs_base1[,ambulate_frail := WHMILE09]

# FRAIL Illness score counts presence of HTN, DM, cancer, chronic lung disease, MI, CHD, angina, asthma, arthritis, stroke, and ckd
# Scores for 0-4 conditions, score = 0, for ≥ 5, score=1


####### FRAIL illness

# DM


chs_base1[DIABADA==3, dm_frail := 1]
chs_base1[DIABADA %in% c(1,2), dm_frail := 0]

# HTN

chs_base1[HYPER %in% c(1,2), htn_frail := 0]
chs_base1[HYPER %in% c(3), htn_frail := 1]

###------------###

# COPD (chronic lung disease)

chs_base1[,copd_frail := EMPHYSEM]


###------------###

# Asthma

chs_base1[,asthma_frail := ASTHCUR]


###------------###

# DJD (arthritis)

chs_base1[,djd_frail := ARTH01]


###------------###

# CKD (renal disease)

chs_base1 <- 
  chs_baseboth[,c("NEWID","CYSGFRBL")][chs_base1, on=.(NEWID)]

chs_base1[CYSGFRBL >= 60, ckd_frail := 0]
chs_base1[CYSGFRBL < 60, ckd_frail := 1]


###------------###

# CHF

chs_base1[, chf_frail := CHBLMOD]

###------------###

# CAD/MI

chs_base1[MIBLMOD==0&CHDBLMOD==0, chd_frail := 0]
chs_base1[MIBLMOD==1|CHDBLMOD==1, chd_frail := 1]

###------------###

# Stroke

chs_base1[,stroke_frail := STBLMOD]


###------------###

# Cancer


chs_base1[,cancer_frail := DIAG01]


###------------###

# Compile chronic illness

chs_base1[,conditions_frail := 
  dm_frail+
  htn_frail+
  copd_frail+
  asthma_frail+
  djd_frail+
  ckd_frail+
  chf_frail+
  chd_frail+
  stroke_frail+
  cancer_frail]

chs_base1[conditions_frail <= 4, illness_frail := 0]
chs_base1[conditions_frail > 4, illness_frail := 1]

chs_base1[WEIGHT08==1, wtloss_frail := 1]
chs_base1[WEIGHT08 %in% c(2,3,4), wtloss_frail := 0]

chs_base1[,total_frail := 
  fatigue_frail+
  resistance_frail+
  ambulate_frail+
  illness_frail+
  wtloss_frail]


######### FRIED

##### Walking speed

#### Men:
# Height ≤ 173 cm (68.1 in):  ≤0.96 m/s      
# Height >173 cm (68.1 in):   ≤1.04  m/s	
# 


chs_base1[!is.na(TIME17),gaitspeed_fried := 0] 
chs_base1[GEND01==1&
            STHT>173&
            TIME17 >= 6, gaitspeed_fried := 1]

chs_base1[GEND01==1&
                  STHT <= 173&
                  TIME17 >=7, gaitspeed_fried := 1]


### Women
# Height ≤ 159 cm (62.6 in):  ≤0.90m/s
# Height >159 cm (62.6 in):   ≤1.02 m/s


chs_base1[GEND01==0&
            STHT>159&
            TIME17 >= 6, gaitspeed_fried := 1]

chs_base1[GEND01==0&
            STHT <= 159&
            TIME17 >= 7, gaitspeed_fried := 1]

########### Grip (kg)

# Trials for each hand

## Uses BMI for normalization

##### Men
# BMI ≤ 24.0:     ≤ 29 kg
# BMI 24.1-26.0:  ≤ 30 kg
# BMI 26.1-28.0:  ≤ 30 kg
# BMI > 28.0:     ≤ 32 kg

##### Women
# BMI ≤ 23.0:     ≤ 17
# BMI 23.1-26.0:  ≤ 17.3 kg
# BMI 26.1-29.0:  ≤ 18 kg
# BMI > 29.0:     ≤ 21 kg

chs_base1[,grip_avg := round(apply(chs_base1[,c(385:387,390:392)],MARGIN=1,mean),1)]
chs_base1[,grip_max := round(apply(chs_base1[,c(385:387,390:392)],MARGIN=1,max),1)]

chs_base1[!is.na(grip_max), grip_fried := 0]

# Men

chs_base1[BMI<=24&
            grip_max <= 29&
            GEND01==1, grip_fried := 1] 

chs_base1[BMI>24&BMI<=26&
            grip_max <= 30&
            GEND01==1, grip_fried := 1] 

chs_base1[BMI>26&BMI<=28&
            grip_max <= 30&
            GEND01==1, grip_fried := 1] 

chs_base1[BMI>28&
            grip_max <= 32&
            GEND01==1, grip_fried := 1]

# Women

chs_base1[BMI<=23&
            grip_max <= 17&
            GEND01==0, grip_fried := 1]

chs_base1[BMI>23&BMI<=26&
            grip_max <= 17.3&
            GEND01==0, grip_fried := 1]

chs_base1[BMI>26&BMI<=29&
            grip_max <= 18&
            GEND01==0, grip_fried := 1]

chs_base1[BMI>29&
            grip_max <= 21&
            GEND01==0, grip_fried := 1]


chs_base1[GEND01==1&KCAL < 383, activity_fried := 1]
chs_base1[GEND01==1&KCAL >= 383, activity_fried := 0]

chs_base1[GEND01==0&KCAL < 270, activity_fried := 1]
chs_base1[GEND01==0&KCAL >= 270, activity_fried := 0]

chs_base1[chs_base1$WEIGHT08==1, wtloss_fried := 1]
chs_base1[chs_base1$WEIGHT08 %in% c(2,3,4)] <- 0

chs_base1[EFFORT05 %in% c(2,3)|GETGO05 %in% c(2,3), fatigue_fried := 1]
chs_base1[EFFORT05 %in% c(0,1)|GETGO05 %in% c(0,1), fatigue_fried := 0]


chs_base1[,total_fried := 
  wtloss_fried+
  grip_fried+
  fatigue_fried+
  gaitspeed_fried+
  activity_fried]

#####. Baseline echo


chs_base2[,earatio := DPMEI43/DPMAI43]

chs_base2[,STHT := chs_base1$STHT]
chs_base2[,WEIGHT_kg := chs_base1$WEIGHT]
chs_base2[,GEND01 := chs_base1$GEND01]

chs_base2 <- 
  as.data.table(
    calc_hypertrophy_type(
      df=chs_base2,
      sex="GEND01",
      male="1",
      female="0",
      lvedd="MMLVDD43",
      ivsd="MMVSTD43",
      lvpwtd="MMLVWD43",
      height="STHT",
      weight="WEIGHT_kg"))

chs_melt_base1 <- 
  melt(chs_base1,id.vars=c("NEWID"),
                       na.rm=T,
                       factorsAsStrings = T,
                       variable.factor=F)
setnames(chs_melt_base1,"NEWID","patientid")
chs_melt_base1[,visitdays := 0]
chs_melt_base1[,visit := 1]
chs_melt_base1[,form := "base1"]

chs_melt_base2 <- 
  melt(chs_base2,
       id.vars=c("NEWID"),
       na.rm=T,
       factorsAsStrings = T,
       variable.factor=F)

setnames(chs_melt_base2,"NEWID","patientid")
chs_melt_base2[,visitdays := 0]
chs_melt_base2[,visit := 1]
chs_melt_base2[,form := "base2"]

chs_melt_baseboth <- 
  melt(chs_baseboth,
       id.vars=c("NEWID"),
       na.rm=T,
       factorsAsStrings = T,
       variable.factor=F)

setnames(chs_melt_baseboth,"NEWID","patientid")
chs_melt_baseboth[,visitdays := 0]
chs_melt_baseboth[,visit := 1]
chs_melt_baseboth[,form := "baseboth"]


#-----------------------------------------------------------------------------------#

#### CHS Year 3 ####

chs_yr3 <- 
  fread("~/Dropbox/BioLINCC files/CHS/Main_Study/YEAR3/yr3.csv",
           na.strings=c("NA","","NULL"))

names(chs_yr3) <- toupper(names(chs_yr3))

chs_yr3 <-
  chs_baseboth[,.(NEWID, 
                  agebl=agey)][chs_yr3,
                               on=.(NEWID)]

chs_yr3[,agey := agebl+STDYTIME/365]

chs_yr3 <- 
  chs_illness_bl[chs_yr3,
                 on=.(NEWID)]

chs_yr3 <- 
  chs_outcomes[chs_yr3,
                 on=.(patientid=NEWID)]


chs_yr3[,lsns_dep := HLPSHP03]
chs_yr3[RELYON03==5, lsns_dep := 5]

####### FRAIL score

## Fatigue, Resistance, Ambulation encoded in 'criteriaset'

chs_yr3[EFFORT05 %in% c(2,3)|GETGO05 %in% c(2,3),fatigue_frail := 1]
chs_yr3[EFFORT05 %in% c(0,1)|GETGO05 %in% c(0,1), fatigue_frail := 0]

## Resistance

chs_yr3[STEPS09 %in% c(0,1), resistance_frail := STEPS09]

## Ambulate

chs_yr3[WHMILE09 %in% c(0,1), ambulate_frail := WHMILE09]


# FRAIL Illness score counts presence of HTN, DM, cancer, chronic lung disease, MI, CHD, angina, asthma, arthritis, stroke, and ckd
# Scores for 0-4 conditions, score = 0, for ≥ 5, score=1

# DM

chs_yr3[!is.na(STDYTIME), dm_frail := 0]
chs_yr3[DIABET37==1, dm_frail := 1]
chs_yr3[dm_bl %in% c(3,4)&!is.na(STDYTIME), dm_frail := 1]

# HTN

chs_yr3[!is.na(STDYTIME), htn_frail := htn_bl]
chs_yr3[HYPER %in% c(0,1), htn_frail := 0]
chs_yr3[HYPER %in% c(2), htn_frail := 1]

###------------###

# COPD (chronic lung disease)

chs_yr3[,copd_frail := copd_bl]


###------------###

# Asthma
chs_yr3[,asthma_frail := asthma_bl]


###------------###

# DJD (arthritis)

chs_yr3[,djd_frail := 0]
chs_yr3[arth_bl==1|
          ARTHND37==1|
          ARTSHD37==1|
          ARTHIP37==1|
          ARTTRT37==1,
        djd_frail := 1]


###------------###

# CKD (renal disease)

chs_yr3[gfr_bl >= 60, ckd_frail := 0]
chs_yr3[gfr_bl < 60, ckd_frail := 1]


###------------###

# CHF/MI

chs_yr3[!is.na(STDYTIME), chf_frail := chf_bl]
chs_yr3[hfhosp_status==1&
          hfhosp_dt <= STDYTIME, chf_frail := 1]

###------------###

# CAD

chs_yr3[!is.na(STDYTIME), chd_frail := cad_bl]
chs_yr3[cadhosp_status==1&
          cadhosp_dt <= STDYTIME, chf_frail := 1]

###------------###

# Stroke

chs_yr3[!is.na(STDYTIME), stroke_frail := cva_bl]
chs_yr3[cvahosp_status==1&
          cvahosp_dt <= STDYTIME, 
        stroke_frail := 1]


###------------###

# Cancer

chs_yr3[!is.na(STDYTIME), cancer_frail := cancer_bl]

chs_yr3[CANCER37==1, cancer_frail := 1]


###------------###

# Compile chronic illness

chs_yr3[,conditions_frail := 
  dm_frail+
  htn_frail+
  copd_frail+
  asthma_frail+
  djd_frail+
  ckd_frail+
  chf_frail+
  chd_frail+
  stroke_frail+
  cancer_frail]

chs_yr3[conditions_frail <= 4, illness_frail := 0]
chs_yr3[conditions_frail > 4, illness_frail := 1]

chs_yr3[WEIGHT38==1, wtloss_frail := 1]
chs_yr3[WEIGHT38 %in% c(2,3,4), wtloss_frail := 0]


chs_yr3[,total_frail := 
  fatigue_frail+
  resistance_frail+
  ambulate_frail+
  illness_frail+
  wtloss_frail]


chs_yr3[PERSTAT==1, visit := 3]

chs_melt_yr3 <- melt(subset(chs_yr3,!is.na(STDYTIME)),
                     id.vars=c("patientid","STDYTIME","PERSTAT","visit"),
                     na.rm=T,
                     factorsAsStrings = T,
                     variable.factor = F)
setnames(chs_melt_yr3,c("STDYTIME","PERSTAT"),c("visitdays","cohort"))

chs_melt_yr3$form <- "yr3final"

#-----------------------------------------------------------------------------------#

#### CHS Year 4 ####

chs_yr4 <- 
  fread("~/Dropbox/BioLINCC files/CHS/Main_Study/YEAR4/yr4.csv",
        na.strings=c("NA","","NULL"))

names(chs_yr4) <- toupper(names(chs_yr4))

chs_yr4 <-
  chs_baseboth[,.(NEWID, agebl=agey)][chs_yr4,
        on=.(NEWID)]

chs_yr4[,agey := agebl+STDYTIME/365]

chs_yr4[,hrt_years := EPMO39/12]

chs_yr4 <- chs_illness_bl[chs_yr4,
                 on=.(NEWID)]

chs_yr4 <- chs_outcomes[chs_yr4,
                 on=.(patientid = NEWID)]


chs_yr4[,WT_CHG := round(WEIGHT-WEIGHT,1)]
chs_yr4[,weight_kg := WEIGHT/2.54]
chs_yr4[,BMI := calc_bmi(chs_yr4)]

chs_yr4[,lsns_dep := HLPSHP03]
chs_yr4[RELYON03==5,lsns_dep := 5]


####### FRAIL score

## Fatigue, Resistance, Ambulation encoded in 'criteriaset'

## Fatigue

chs_yr4[EFFORT05 %in% c(2,3)|
                        GETGO05 %in% c(2,3), fatigue_frail := 1]

chs_yr4[EFFORT05 %in% c(0,1)|
                        GETGO05 %in% c(0,1), fatigue_frail := 0]

## Resistance

chs_yr4[STEPS09 %in% c(0,1), resistance_frail := STEPS09]

## Ambulate

chs_yr4[WHMILE09 %in% c(0,1), ambulate_frail := WHMILE09]


# FRAIL Illness score counts presence of HTN, DM, cancer, chronic lung disease, MI, CHD, angina, asthma, arthritis, stroke, and ckd
# Scores for 0-4 conditions, score = 0, for ≥ 5, score=1


#----------------------#

# DM

chs_yr4[!is.na(STDYTIME), dm_frail := chs_yr3[!is.na(chs_yr4$STDYTIME),dm_frail]]
chs_yr4[DIABET39==1, dm_frail := 1]


#----------------------#

# HTN

chs_yr4[!is.na(STDYTIME), htn_frail := chs_yr3[!is.na(chs_yr4$STDYTIME), htn_frail]]
chs_yr4[HYPER %in% c(0,1), htn_frail := 0]
chs_yr4[HYPER %in% c(2), htn_frail := 1]


#----------------------#

# COPD (chronic lung disease)

chs_yr4[!is.na(STDYTIME), copd_frail := chs_yr3[!is.na(chs_yr4$STDYTIME), copd_frail]]


#----------------------#

# Asthma

chs_yr4[!is.na(STDYTIME), asthma_frail := chs_yr3[!is.na(chs_yr4$STDYTIME),asthma_frail]]


#----------------------#

# DJD (arthritis)

chs_yr4[!is.na(STDYTIME), djd_frail := chs_yr3$djd_frail[!is.na(chs_yr4$STDYTIME)]]


#----------------------#

# CKD (renal disease)

chs_yr4[!is.na(STDYTIME), 
                  ckd_frail := chs_yr3[!is.na(chs_yr4$STDYTIME), ckd_frail]]


#----------------------#

# CHF/MI

chs_yr4[!is.na(STDYTIME), chf_frail :=  chs_yr3$chf_frail[!is.na(chs_yr4$STDYTIME)]]
chs_yr4[hfhosp_status==1&hfhosp_dt <= STDYTIME, chf_frail := 1]

#----------------------#

# CAD

chs_yr4[!is.na(STDYTIME), chd_frail := chs_yr3$chd_frail[!is.na(chs_yr4$STDYTIME)]]
chs_yr4[cadhosp_status==1&cadhosp_dt <= STDYTIME, chd_frail := 1]

#----------------------#

# Stroke

chs_yr4[!is.na(STDYTIME), stroke_frail :=  chs_yr3[!is.na(chs_yr4$STDYTIME), stroke_frail]]
chs_yr4[cvahosp_status==1&
          cvahosp_dt <= STDYTIME, stroke_frail := 1]


#----------------------#

# Cancer

chs_yr4[!is.na(STDYTIME), cancer_frail := chs_yr3[!is.na(chs_yr4$STDYTIME), cancer_frail]]


#----------------------#

# Compile chronic illness

chs_yr4[,conditions_frail := 
  dm_frail+
  htn_frail+
  copd_frail+
  asthma_frail+
  djd_frail+
  ckd_frail+
  chf_frail+
  chd_frail+
  stroke_frail+
  cancer_frail]

chs_yr4[conditions_frail <= 4, illness_frail := 0]
chs_yr4[conditions_frail > 4, illness_frail := 1]

## Weight loss

chs_yr4[WT_CHG < -10, wtloss_frail := 1]
chs_yr4[WT_CHG >= -10, wtloss_frail := 0]

## FRAIL score

chs_yr4[,total_frail := 
          fatigue_frail+
          resistance_frail+
          ambulate_frail+
          illness_frail+
          wtloss_frail]


#-----------------------------------------------------------------------#

chs_yr4[,visit := 4]

### Melt ###

chs_melt_yr4 <- melt(subset(chs_yr4,!is.na(STDYTIME)),
                     id.vars=c("patientid","STDYTIME","PERSTAT","visit"),
                     na.rm=T,
                     factorsAsStrings = T,
                     variable.factor=F)
setnames(chs_melt_yr4,c("STDYTIME","PERSTAT"),c("visitdays","cohort"))
chs_melt_yr4[,form := "yr4final"]


#----------------------------------------------------------------#

#### CHS Year 5 - AA cohort ####

chs_yr5new <- fread("~/Dropbox/BioLINCC files/CHS/Main_Study/YEAR5/yr5new.csv",
                    na.strings=c("NA","","NULL"))

names(chs_yr5new) <- toupper(names(chs_yr5new))

chs_yr5new <- 
  chs_illness_bl[PERSTAT==2,][chs_yr5new,
                    on=.(NEWID,PERSTAT)]

chs_yr5new <- chs_outcomes[chs_yr5new,
                    on=.(patientid=NEWID)]

chs_yr5new[,weight_kg := WEIGHT/2.2]


#----------------------#

####### FRAIL score

## Fatigue, Resistance, Ambulation encoded in 'criteriaset'


## Fatigue

## Fatigue

chs_yr5new[EFFORT05 %in% c(2,3)|GETGO05 %in% c(2,3), fatigue_frail := 1]
chs_yr5new[EFFORT05 %in% c(0,1)|GETGO05 %in% c(0,1), fatigue_frail := 0]

## Resistance

chs_yr5new[STEPS09 %in% c(0,1), 
           resistance_frail :=  STEPS09]

## Ambulate

chs_yr5new[WHMILE09 %in% c(0,1), ambulate_frail := WHMILE09]


# FRAIL Illness score counts presence of HTN, DM, cancer, chronic lung disease, MI, CHD, angina, asthma, arthritis, stroke, and ckd
# Scores for 0-4 conditions, score = 0, for ≥ 5, score=1


#----------------------#

# DM

chs_yr5new[DIABADA %in% c(1,2), dm_frail := 0]
chs_yr5new[DIABADA %in% c(3,4), dm_frail := 1]


#----------------------#

# HTN

chs_yr5new[HYPER %in% c(0,1), htn_frail := 0]
chs_yr5new[HYPER %in% c(2), htn_frail := 1]

#----------------------#

# COPD (chronic lung disease)

chs_yr5new[copd_bl==0&EMPHYSEM==0, copd_frail := 0]
chs_yr5new[copd_bl==1|EMPHYSEM==1, copd_frail := 1]


#----------------------#

# Asthma

chs_yr5new[,asthma_frail := ASTHMA]


#----------------------#

# DJD (arthritis)

chs_yr5new[,djd_frail := ARTH01]


#----------------------#

# CKD (renal disease)

chs_yr5new[CYSGFR5 >= 60, ckd_frail := 0]
chs_yr5new[CYSGFR5 < 60, ckd_frail := 1]


#----------------------#

# CHF/MI

chs_yr5new[,chf_frail := CHBLMOD]

#----------------------#

# CAD

chs_yr5new[,chd_frail := chs_yr5new$CHDBLMOD]

#----------------------#

# Stroke

chs_yr5new[,stroke_frail := STBLMOD]


#----------------------#

# Cancer

chs_yr5new[,cancer_frail := DIAG01]


#----------------------#

# Compile chronic illness

chs_yr5new[,conditions_frail := 
  dm_frail+
  htn_frail+
  copd_frail+
  asthma_frail+
  djd_frail+
  ckd_frail+
  chf_frail+
  chd_frail+
  stroke_frail+
  cancer_frail]

chs_yr5new[conditions_frail <= 4, illness_frail := 0]
chs_yr5new[conditions_frail > 4, illness_frail := 1]

## Weight loss

chs_yr5new[WEIGHT58==1, wtloss_frail := 1]
chs_yr5new[WEIGHT58 %in% c(2,3,4), wtloss_frail := 0]

## FRAIL score

chs_yr5new[,total_frail := 
             fatigue_frail+
             resistance_frail+
             ambulate_frail+
             illness_frail+
             wtloss_frail]


#-----------------------------------------------------------------------------------#

######### FRIED

##### Walking speed

#### Men:
# Height ≤ 173 cm (68.1 in):  ≤0.96 m/s      
# Height >173 cm (68.1 in):   ≤1.04  m/s	
# 


chs_yr5new[!is.na(TIME27), gaitspeed_fried := 0] 
chs_yr5new[gender_ref==1&
             height_cm>173&
             TIME27 >= 6, gaitspeed_fried := 1]

chs_yr5new[gender_ref==1&
             height_cm <= 173&
             TIME27 >=7, gaitspeed_fried := 1]


### Women
# Height ≤ 159 cm (62.6 in):  ≤0.90m/s
# Height >159 cm (62.6 in):   ≤1.02 m/s


chs_yr5new[gender_ref==0&
             height_cm>159&
             TIME27 >= 6, gaitspeed_fried := 1]

chs_yr5new[gender_ref==0&
             height_cm <= 159&
             TIME27 >= 7, gaitspeed_fried := 1]

########### Grip (kg)

# Trials for each hand

## Uses BMI for normalization

##### Men
# BMI ≤ 24.0:     ≤ 29 kg
# BMI 24.1-26.0:  ≤ 30 kg
# BMI 26.1-28.0:  ≤ 30 kg
# BMI > 28.0:     ≤ 32 kg

##### Women
# BMI ≤ 23.0:     ≤ 17
# BMI 23.1-26.0:  ≤ 17.3 kg
# BMI 26.1-29.0:  ≤ 18 kg
# BMI > 29.0:     ≤ 21 kg

chs_yr5new$grip_avg <- round(apply(chs_yr5new[,.(TRY127,TRY227,TRY327, TRY21I27,TRY22I27,TRY23I27)],MARGIN=1,mean),1)
chs_yr5new$grip_max <- round(apply(chs_yr5new[,.(TRY127,TRY227,TRY327, TRY21I27,TRY22I27,TRY23I27)],MARGIN=1,max),1)

chs_yr5new$grip_fried[!is.na(chs_yr5new$grip_max)] <- 0 

# Men

chs_yr5new[BMI<=24&
             grip_max <= 29&
             gender_ref==1, grip_fried := 1] 

chs_yr5new[BMI>24&BMI<=26&
             grip_max <= 30&
             gender_ref==1, grip_fried := 1]  

chs_yr5new[BMI>26&BMI<=28&
             grip_max <= 30&
             gender_ref==1, grip_fried := 1]  

chs_yr5new[BMI>28&
             grip_max <= 32&
             gender_ref==1, grip_fried := 1] 

# Women

chs_yr5new[BMI<=23&
             grip_max <= 17&
             gender_ref==0, grip_fried := 1]  

chs_yr5new[BMI>23&BMI<=26&
             grip_max <= 17.3&
             gender_ref==0, grip_fried := 1] 

chs_yr5new[BMI>26&BMI<=29&
             grip_max <= 18&
             gender_ref==0, grip_fried := 1] 

chs_yr5new[BMI>29&
             grip_max <= 21&
             gender_ref==0, grip_fried := 1] 


chs_yr5new[GEND01==1&KCAL < 383, activity_fried := 1]
chs_yr5new[GEND01==1&KCAL >= 383, activity_fried := 0]

chs_yr5new[GEND01==0&KCAL < 270, activity_fried := 1]
chs_yr5new[GEND01==0&KCAL >= 270, activity_fried := 0]

chs_yr5new[WEIGHT58==1, wtloss_fried := 1]
chs_yr5new[WEIGHT58 %in% c(2,3,4), wtloss_fried := 0]

chs_yr5new[EFFORT05 %in% c(2,3)|GETGO05 %in% c(2,3), fatigue_fried := 1]
chs_yr5new[EFFORT05 %in% c(0,1)|GETGO05 %in% c(0,1), fatigue_fried := 0]


chs_yr5new[,total_fried := 
  wtloss_fried+
  grip_fried+
  fatigue_fried+
  gaitspeed_fried+
  activity_fried]


chs_yr5new[,STDYTIME := 0]

chs_yr5new[,visit := 1]


chs_melt_yr5new <- melt(chs_yr5new,
                        id.vars=c("patientid","STDYTIME","PERSTAT","visit"),
                        na.rm=T,
                        factorsAsStrings = T,
                        variable.factor = F)


setnames(chs_melt_yr5new,
         c("STDYTIME","PERSTAT"),
         c("visitdays","cohort"))

chs_melt_yr5new[,form := "yr5final_new"]


#-----------------------------------------------------------------------------------#

#### CHS Year 5 - Original cohort ####

chs_yr5old <- 
  fread("~/Dropbox/BioLINCC files/CHS/Main_Study/YEAR5/yr5old.csv",
        na.strings=c("NA","","NULL"))

names(chs_yr5old) <- toupper(names(chs_yr5old))

chs_yr5old <-
  chs_baseboth[,.(NEWID, agebl=agey,race=RACE01,sex=GEND01)][
    chs_yr5old, on=.(NEWID)]

chs_yr5old <- chs_outcomes[chs_yr5old,
                        on=.(patientid = NEWID)]


chs_yr5old[,agey := floor(agebl+(STDYTIME/365))]

chs_yr5old[,lsns_dep := HLPSHP03]
chs_yr5old[RELYON03==5,lsns_dep := 5]

chs_illness_bl$patientid <- chs_illness_bl$NEWID
chs_yr5old <- 
  chs_illness_bl[PERSTAT==1,.(patientid,gender_ref,race_ref,height_cm)][
    chs_yr5old,
        on=.(patientid)]


chs_yr5old <- 
  chs_yr4[,c("patientid",
             "dm_frail",
             "htn_frail",
             "djd_frail",
             "asthma_frail",
             "copd_frail",
             "chf_frail",
             "chd_frail",
             "stroke_frail",
             "cancer_frail")][chs_yr5old,on=.(patientid)]


chs_yr5old[,weight_kg := WEIGHT/2.2]


#----------------------#

####### FRAIL score

## Fatigue, Resistance, Ambulation encoded in 'criteriaset'

## Fatigue

chs_yr5old[EFFORT05 %in% c(2,3)|GETGO05 %in% c(2,3), fatigue_frail := 1]
chs_yr5old[EFFORT05 %in% c(0,1)|GETGO05 %in% c(0,1), fatigue_frail := 0]

## Resistance

chs_yr5old[STEPS09 %in% c(0,1), 
           resistance_frail := STEPS09]

## Ambulate

chs_yr5old[WHMILE09 %in% c(0,1), ambulate_frail := 
  WHMILE09]


# FRAIL Illness score counts presence of HTN, DM, cancer, chronic lung disease, MI, CHD, angina, asthma, arthritis, stroke, and ckd
# Scores for 0-4 conditions, score = 0, for ≥ 5, score=1


#----------------------#

# DM

chs_yr5old[DIABADA %in% c(3), dm_frail := 1]
chs_yr5old[is.na(STDYTIME), dm_frail :=NA]


#----------------------#

# HTN

chs_yr5old[HYPER %in% c(0,1), htn_frail := 0]
chs_yr5old[HYPER %in% c(2), htn_frail := 1]
chs_yr5old[is.na(STDYTIME), htn_frail := NA]


#----------------------#

# COPD (chronic lung disease)

chs_yr5old[!is.na(STDYTIME)&patientid %in% chs_yr4[copd_frail == 1,patientid], 
           copd_frail := 1]
chs_yr5old[!is.na(STDYTIME)&patientid %in% chs_yr4[copd_frail == 0,patientid], 
           copd_frail := 0]
chs_yr5old[is.na(STDYTIME), copd_frail := NA]


#----------------------#

# Asthma

chs_yr5old[is.na(STDYTIME), asthma_frail := NA]


#----------------------#

# DJD (arthritis)

chs_yr5old[is.na(STDYTIME), djd_frail := NA]

chs_yr5old[ARTHND29==1|
                       ARTSHD29==1|
                       ARTHIP29==1|
                       ARTTRT29==1, djd_frail := 1]


#----------------------#

# CKD (renal disease)

chs_yr5old[CYSGFR5 >= 60, ckd_frail := 0]
chs_yr5old[CYSGFR5 < 60, ckd_frail := 1]


#----------------------#

# CHF/MI

chs_yr5old[is.na(STDYTIME), chf_frail := NA]
chs_yr5old[hfhosp_status==1&
                       hfhosp_dt <= STDYTIME, chf_frail := 1]

#----------------------#

# CAD

chs_yr5old[is.na(STDYTIME), chd_frail := NA]
chs_yr5old[cadhosp_status==1&
                       cadhosp_dt <= STDYTIME, chd_frail :=  1]

#----------------------#

# Stroke

chs_yr5old[is.na(STDYTIME), stroke_frail := NA]
chs_yr5old[cvahosp_status==1&
                          cvahosp_dt <= STDYTIME, stroke_frail :=  1]


#----------------------#

# Cancer

chs_yr5old[is.na(STDYTIME), cancer_frail := NA]

chs_yr5old[CANCER29==1&
                          !is.na(STDYTIME), cancer_frail := 1]


#----------------------#

# Compile chronic illness

chs_yr5old[,conditions_frail := 
  dm_frail+
  htn_frail+
  copd_frail+
  asthma_frail+
  djd_frail+
  ckd_frail+
  chf_frail+
  chd_frail+
  stroke_frail+
  cancer_frail]

chs_yr5old[conditions_frail <= 4, illness_frail := 0]
chs_yr5old[conditions_frail > 4, illness_frail := 1]

## Weight loss

chs_yr5old[WEIGHT29==1, wtloss_frail := 1]
chs_yr5old[WEIGHT29 %in% c(2,3,4), wtloss_frail := 0]

## FRAIL score

chs_yr5old[,total_frail := 
  fatigue_frail+
  resistance_frail+
  ambulate_frail+
  illness_frail+
  wtloss_frail]


#-----------------------------------------------------------------------------------#

######### FRIED

##### Walking speed

#### Men:
# Height ≤ 173 cm (68.1 in):  ≤0.96 m/s      
# Height >173 cm (68.1 in):   ≤1.04  m/s	
# 


chs_yr5old[!is.na(TIME27), gaitspeed_fried := 0] 

chs_yr5old[GEND01==1&
             height_cm > 173&
             TIME27 >= 6, gaitspeed_fried := 1]

chs_yr5old[GEND01==1&
             height_cm <= 173&
             TIME27 >= 7, gaitspeed_fried := 1]


### Women
# Height ≤ 159 cm (62.6 in):  ≤0.90m/s
# Height >159 cm (62.6 in):   ≤1.02 m/s


chs_yr5old[GEND01==0&
             height_cm>159&
             TIME27 >= 6, gaitspeed_fried := 1]

chs_yr5old[GEND01==0&
             height_cm <= 159&
             TIME27 >= 7, gaitspeed_fried := 1]

########### Grip (kg)

# Trials for each hand

## Uses BMI for normalization

##### Men
# BMI ≤ 24.0:     ≤ 29 kg
# BMI 24.1-26.0:  ≤ 30 kg
# BMI 26.1-28.0:  ≤ 30 kg
# BMI > 28.0:     ≤ 32 kg

##### Women
# BMI ≤ 23.0:     ≤ 17
# BMI 23.1-26.0:  ≤ 17.3 kg
# BMI 26.1-29.0:  ≤ 18 kg
# BMI > 29.0:     ≤ 21 kg

chs_yr5old[, grip_avg := round(apply(chs_yr5old[,.(TRY127,TRY227,TRY327, TRY21I27,TRY22I27,TRY23I27)],MARGIN=1,mean),1)]
chs_yr5old[, grip_max := round(apply(chs_yr5old[,.(TRY127,TRY227,TRY327, TRY21I27,TRY22I27,TRY23I27)],MARGIN=1,max),1)]

chs_yr5old[!is.na(grip_max), grip_fried := 0] 

# Men

chs_yr5old[BMI<=24&
             grip_max <= 29&
             gender_ref==1, grip_fried := 1]

chs_yr5old[BMI>24&BMI<=26&
             grip_max <= 30&
             gender_ref==1, grip_fried := 1]

chs_yr5old[BMI>26&BMI<=28&
             grip_max <= 30&
             gender_ref==1, grip_fried := 1]

chs_yr5old[BMI>28&
             grip_max <= 32&
             gender_ref==1, grip_fried := 1]

# Women

chs_yr5old[BMI<=23&
             grip_max <= 17&
             gender_ref==0, grip_fried := 1]

chs_yr5old[BMI>23&BMI<=26&
             grip_max <= 17.3&
             gender_ref==0, grip_fried := 1]

chs_yr5old[BMI>26&BMI<=29&
             grip_max <= 18&
             gender_ref==0, grip_fried := 1]

chs_yr5old[BMI>29&
             grip_max <= 21&
             gender_ref==0, grip_fried := 1]


chs_yr5old[GEND01==1&KCAL < 383, activity_fried := 1]
chs_yr5old[GEND01==1&KCAL >= 383, activity_fried := 0]

chs_yr5old[GEND01==0&KCAL < 270, activity_fried := 1]
chs_yr5old[GEND01==0&KCAL >= 270, activity_fried := 0]

chs_yr5old[WEIGHT29==1, wtloss_fried := 1]
chs_yr5old[WEIGHT29 %in% c(2,3,4), wtloss_fried := 0]

chs_yr5old[EFFORT05 %in% c(2,3)|GETGO05 %in% c(2,3), fatigue_fried := 1]
chs_yr5old[EFFORT05 %in% c(0,1)|GETGO05 %in% c(0,1), fatigue_fried := 0]


chs_yr5old[,total_fried := 
             wtloss_fried+
             grip_fried+
             fatigue_fried+
             gaitspeed_fried+
             activity_fried]


chs_yr5new[,COHORT := 2]
chs_yr5old[,COHORT := 1]


chs_illness_yr5 <- 
  rbindlist(
    list(chs_yr5new[,c("patientid",
                       "COHORT",
                       "STDYTIME",
                       "dm_frail",
                       "htn_frail",
                       "copd_frail",
                       "asthma_frail",
                       "djd_frail",
                       "ckd_frail",
                       "chf_frail",
                       "chd_frail",
                       "stroke_frail",
                       "cancer_frail")],
         chs_yr5old[,c("patientid",
                       "COHORT",
                       "STDYTIME",
                       "dm_frail",
                       "htn_frail",
                       "copd_frail",
                       "asthma_frail",
                       "djd_frail",
                       "ckd_frail",
                       "chf_frail",
                       "chd_frail",
                       "stroke_frail",
                       "cancer_frail")]))

chs_yr5old[,visit := 5]

chs_yr5_crcl <-
  rbind(chs_yr5old[,c("patientid","CYSGFR5")],
        chs_yr5new[,c("patientid","CYSGFR5")])

chs_melt_yr5old <- 
  melt(subset(chs_yr5old,!is.na(STDYTIME)),
                        id.vars=c("patientid","STDYTIME","COHORT","visit"),
                        na.rm=T,
                        factorsAsStrings = T,
                        variable.factor=F)

setnames(chs_melt_yr5old,c("STDYTIME","COHORT"),c("visitdays","cohort"))
chs_melt_yr5old[,form := "yr5final_old"]


#### CHS Year 6 ####


chs_yr6 <- 
  fread("~/Dropbox/BioLINCC files/CHS/Main_Study/YEAR6/yr6.csv",
        na.strings=c("NA","","NULL"))


names(chs_yr6) <- toupper(names(chs_yr6))

chs_yr6 <-
  chs_baseboth[,.(NEWID, agebl=agey)][chs_yr6,
                                      on=.(NEWID)]

setnames(chs_yr6,"NEWID","patientid")

chs_yr6 <-
  chs_yr5_crcl[chs_yr6, on=.(patientid)]

chs_yr6[,agey := floor(agebl+(STDYTIME/365))]


chs_yr6 <- 
        chs_illness_yr5[,!("STDYTIME"), with = FALSE][chs_yr6,on=.(patientid)]


chs_yr6 <-
  chs_outcomes[chs_yr6,on=.(patientid)]


chs_yr6[,weight_kg := WEIGHT/2.2]
chs_yr6[,height_cm := chs_illness_bl$height_cm]
chs_yr6[,gender_ref := chs_illness_bl$gender_ref]
chs_yr6[,BMI := calc_bmi(chs_yr6)]


chs_yr6[,lsns_dep := HLPSHP03]
chs_yr6[RELYON03==5, lsns_dep := 5]


## Fatigue

chs_yr6[EFFORT05 %in% c(2,3)|GETGO05 %in% c(2,3), fatigue_frail := 1]
chs_yr6[EFFORT05 %in% c(0,1)|GETGO05 %in% c(0,1), fatigue_frail := 0]

## Resistance

chs_yr6[STEPS09 %in% c(0,1), resistance_frail := 
  STEPS09]

## Ambulate

chs_yr6[WHMILE09 %in% c(0,1), ambulate_frail := 
  WHMILE09]


#------------------------------#

# DM

# chs_yr6 <-
#   as.data.frame(chs_yr6)

chs_yr6[is.na(dm_frail), dm_frail := NA]
chs_yr6[DIABET59 %in% c(2,3), dm_frail := 1]

#------------------------------#

# HTN

chs_yr6[is.na(STDYTIME), htn_frail := NA]
chs_yr6[HYPER %in% c(0,1), htn_frail := 0]
chs_yr6[HYPER %in% c(2), htn_frail := 1]

#------------------------------#

# COPD (chronic lung disease)

chs_yr6[is.na(STDYTIME), copd_frail := NA]


###------------###

# Asthma

chs_yr6[is.na(STDYTIME), asthma_frail := NA]
chs_yr6[ASTHMA56 == 1, asthma_frail := 1]


###------------###

# DJD (arthritis)

chs_yr6[is.na(STDYTIME), djd_frail := NA]
chs_yr6[ARTHND59==1|
                    ARTSHD59==1|
                    ARTHIP59==1|
                    ARTTRT59==1, djd_frail := 1]  


chs_yr6[ARTHND59==0&
                    ARTSHD59==0&
                    ARTHIP59==0&
                    ARTTRT59==0, djd_frail := 1]  


###------------###

# CKD (renal disease)


chs_yr6[CYSGFR5 >= 60, ckd_frail := 0]
chs_yr6[CYSGFR5 < 60, ckd_frail := 1]

chs_yr6[is.na(STDYTIME), ckd_frail := 0]


###------------###

# CHF/MI

chs_yr6[hfhosp_status==1&
                    hfhosp_dt <= STDYTIME, chf_frail := 1]

###------------###

# CAD

chs_yr6[cadhosp_status==1&
                    cadhosp_dt <= STDYTIME, chd_frail := 1]

###------------###

# Stroke

chs_yr6[cvahosp_status==1&
                       cvahosp_dt <= STDYTIME, stroke_frail := 1]


###------------###

# Cancer

chs_yr6[is.na(STDYTIME), cancer_frail := NA]
chs_yr6[,cancer_frail := CANCER59]


###------------###

# Compile chronic illness

chs_yr6[,conditions_frail := 
  dm_frail+
  htn_frail+
  copd_frail+
  asthma_frail+
  djd_frail+
  ckd_frail+
  chf_frail+
  chd_frail+
  stroke_frail+
  cancer_frail]

chs_yr6[conditions_frail <= 4, illness_frail := 0]
chs_yr6[conditions_frail > 4, illness_frail := 1]

## Weight loss

chs_yr6[WEIGHT59==1, wtloss_frail := 1]
chs_yr6[WEIGHT59 %in% c(2,3,4), wtloss_frail := 0]


## Add up FRAIL score

chs_yr6[,total_frail := 
  fatigue_frail+
  resistance_frail+
  ambulate_frail+
  illness_frail+
  wtloss_frail]


chs_yr6 <-
  as.data.table(chs_yr6)


#-----------------------------------------------------------------------------------#

## Melt


chs_melt_yr6 <- melt(subset(chs_yr6,!is.na(STDYTIME)),
                     id.vars=c("patientid","STDYTIME","COHORT"),
                     na.rm=T,
                     factorsAsStrings = T,
                     variable.factor=F)
setnames(chs_melt_yr6,c("STDYTIME","COHORT"),c("visitdays","cohort"))
chs_melt_yr6[,visit := 6]
chs_melt_yr6[,form := "yr6final"]


#### CHS Year 7 ####


chs_yr7 <- 
  fread("~/Dropbox/BioLINCC files/CHS/Main_Study/YEAR7/yr7.csv",
        na.strings=c("NA","","NULL"))

names(chs_yr7) <- toupper(names(chs_yr7))

chs_yr7 <-
  merge(chs_yr7,
        chs_baseboth[,.(NEWID, agebl=agey)],
        by="NEWID",
        all.x=T)

chs_yr7[,agey := 
  floor(chs_baseboth$agebl+
          (STDYTIME/365))]

chs_yr7 <- chs_outcomes[chs_yr7, 
                 on=.(patientid=NEWID)]

chs_yr7[,gender_ref := chs_illness_bl$gender_ref]

chs_yr7[,weight_kg := WEIGHT/2.2]
chs_yr7[,height_cm := chs_illness_bl$height_cm]

chs_yr7[,BMI := calc_bmi(chs_yr7)]

## Fatigue

chs_yr7[EFFORT05 %in% c(2,3)|GETGO05 %in% c(2,3), fatigue_frail := 1]
chs_yr7[EFFORT05 %in% c(0,1)|GETGO05 %in% c(0,1), fatigue_frail := 0]

## Resistance

chs_yr7[,resistance_frail := STEPS09]

## Ambulate

chs_yr7[,ambulate_frail := WHMILE09]


#--------------------------#

# DM

chs_yr7[!is.na(STDYTIME), dm_frail := chs_yr6$dm_frail[!is.na(chs_yr7$STDYTIME)]]
chs_yr7[DIABET59 %in% c(2,3), dm_frail := 1]


#--------------------------#

# HTN

chs_yr7[!is.na(STDYTIME), htn_frail := chs_yr6$htn_frail[!is.na(chs_yr7$STDYTIME)]]
chs_yr7[HYPER %in% c(0,1), htn_frail := 0] 
chs_yr7[HYPER %in% c(2), htn_frail := 1]


#----------------------------#

# COPD

chs_yr7[!is.na(STDYTIME), copd_frail := chs_yr6$copd_frail[!is.na(chs_yr7$STDYTIME)]]


#----------------------------#

# Asthma

chs_yr7[!is.na(STDYTIME), asthma_frail := chs_yr6$asthma_frail[!is.na(chs_yr7$STDYTIME)]]


###------------###

# DJD (arthritis)

chs_yr7[!is.na(chs_yr7$STDYTIME), djd_frail := chs_yr6$djd_frail[!is.na(chs_yr7$STDYTIME)]]
chs_yr7[ARTHND59==1|
                    ARTSHD59==1|
                    ARTHIP59==1|
                    ARTTRT59==1, djd_frail := 1]  


###------------###

# CKD (renal disease)

chs_yr7[!is.na(STDYTIME), ckd_frail := 
  chs_yr6$ckd_frail[!is.na(chs_yr7$STDYTIME)]]


###------------###

# CHF/MI

chs_yr7[!is.na(STDYTIME), chf_frail :=  
  chs_yr6$chf_frail[!is.na(chs_yr7$STDYTIME)]]
  
chs_yr7[hfhosp_status==1&
                    hfhosp_dt <= STDYTIME, chf_frail := 1]

###------------###

# CAD

chs_yr7[!is.na(STDYTIME), chd_frail :=  
  chs_yr6$chd_frail[!is.na(chs_yr7$STDYTIME)]]
  
chs_yr7[cadhosp_status==1&
                    cadhosp_dt <= STDYTIME, chd_frail :=  1]

###------------###

# Stroke

chs_yr7[!is.na(STDYTIME), stroke_frail := 
  chs_yr6$stroke_frail[!is.na(chs_yr7$STDYTIME)]]

chs_yr7[cvahosp_status==1&
                       cvahosp_dt <= STDYTIME, stroke_frail := 1]


###------------###

# Cancer

chs_yr7[!is.na(STDYTIME), cancer_frail :=  
  chs_yr6$cancer_frail[!is.na(chs_yr7$STDYTIME)]]

# Compile chronic illness

chs_yr7[,conditions_frail := 
  dm_frail+
  htn_frail+
  copd_frail+
  asthma_frail+
  djd_frail+
  ckd_frail+
  chf_frail+
  chd_frail+
  stroke_frail+
  cancer_frail]

chs_yr7[conditions_frail <= 4, illness_frail := 0]
chs_yr7[conditions_frail > 4, illness_frail := 1]


## Weight loss

chs_yr7[WEIGHT59==1, wtloss_frail := 1]
chs_yr7[WEIGHT59 %in% c(2,3,4), wtloss_frail := 0]

## Add up FRAIL score

chs_yr7[,total_frail := 
  fatigue_frail+
  resistance_frail+
  ambulate_frail+
  illness_frail+
  wtloss_frail]


#-----------------------------------------------------------------------------------#


# Echo

chs_yr7[,earatio := DPMEP43/DPMAP43]

chs_yr7 <- 
  as.data.table(
    calc_hypertrophy_type(
      df=chs_yr7,
      sex="GEND01",
      male="1",
      female="0",
      lvedd="MMLVDD43",
      ivsd="MMVSTD43",
      lvpwtd="MMLVWD43",
      height="height_cm",
      weight="weight_kg"))


chs_yr7[,visit := 7]

chs_melt_yr7 <- melt(subset(chs_yr7,!is.na(STDYTIME)),
                     id.vars=c("patientid","STDYTIME","PERSTAT","visit"),
                     na.rm=T,
                     factorsAsStrings = T,
                     variable.factor = F)

setnames(chs_melt_yr7,c("STDYTIME","PERSTAT"),c("visitdays","cohort"))
chs_melt_yr7[,form := "yr7final"]


#----------------------------------------------------------------------------------------------------------------------#

#### CHS Year 8 ####

chs_yr8 <- 
  fread("~/Dropbox/BioLINCC files/CHS/Main_Study/YEAR8/yr8.csv",
        na.strings=c("NA","","NULL"))

names(chs_yr8) <- toupper(names(chs_yr8))

chs_yr8[,agey := 
  floor(chs_baseboth$agebl+
          (STDYTIME/365))]

chs_yr8 <- chs_outcomes[chs_yr8, 
                 on=.(patientid=NEWID)]


chs_yr8[,weight_kg := WEIGHT/2.2]
chs_yr8[,height_cm := chs_illness_bl$height_cm]
chs_yr8[,gender_ref := chs_illness_bl$gender_ref]

chs_yr8[,BMI := calc_bmi(chs_yr8)]

## Fatigue

chs_yr8[chs_yr8$EFFORT05 %in% c(2,3)|chs_yr8$GETGO05 %in% c(2,3), fatigue_frail := 1]
chs_yr8[chs_yr8$EFFORT05 %in% c(0,1)|chs_yr8$GETGO05 %in% c(0,1), fatigue_frail := 0]

## Resistance

chs_yr8[,resistance_frail := STEPS09]

## Ambulate

chs_yr8[,ambulate_frail := WHMILE09]


#----------------------------#

# DM

chs_yr8[!is.na(chs_yr8$STDYTIME), dm_frail :=  
  chs_yr7$dm_frail[!is.na(chs_yr8$STDYTIME)]]
  
chs_yr8[chs_yr8$DIABET59 %in% c(2,3), dm_frail := 1]


#----------------------------#

# HTN

chs_yr8[!is.na(chs_yr8$STDYTIME), htn_frail := chs_yr7$htn_frail[!is.na(chs_yr8$STDYTIME)]]


#----------------------------#

# COPD

chs_yr8[!is.na(chs_yr8$STDYTIME), copd_frail := chs_yr7$copd_frail[!is.na(chs_yr8$STDYTIME)]]


#----------------------------#

# Asthma

chs_yr8[!is.na(chs_yr8$STDYTIME), asthma_frail := chs_yr7$asthma_frail[!is.na(chs_yr8$STDYTIME)]]


###------------###

# DJD (arthritis)

chs_yr8[!is.na(chs_yr8$STDYTIME), dj_frail := chs_yr7$djd_frail[!is.na(chs_yr8$STDYTIME)]]
chs_yr8[chs_yr8$ARTHND59==1|
                    chs_yr8$ARTSHD59==1|
                    chs_yr8$ARTHIP59==1|
                    chs_yr8$ARTTRT59==1, djd_frail := 1]  


###------------###

# CKD (renal disease)

chs_yr8[!is.na(chs_yr8$STDYTIME), ckd_frail := chs_yr7$ckd_frail[!is.na(chs_yr8$STDYTIME)]]


###------------###

# CHF/MI

chs_yr8[!is.na(chs_yr8$STDYTIME), chf_frail :=  
  chs_yr7$chf_frail[!is.na(chs_yr8$STDYTIME)]]

chs_yr8[chs_yr8$hfhosp_status==1&
                    chs_yr8$hfhosp_dt <= chs_yr8$STDYTIME, chf_frail := 1]

###------------###

# CAD

chs_yr8[!is.na(chs_yr8$STDYTIME), chd_frail :=  
  chs_yr7$chd_frail[!is.na(chs_yr8$STDYTIME)]]

chs_yr8[chs_yr8$cadhosp_status==1&
                    chs_yr8$cadhosp_dt <= chs_yr8$STDYTIME, chd_frail := 1]

###------------###

# Stroke

chs_yr8[!is.na(chs_yr8$STDYTIME), stroke_frail :=  
  chs_yr7$stroke_frail[!is.na(chs_yr8$STDYTIME)]]
chs_yr8[chs_yr8$cvahosp_status==1&
                       chs_yr8$cvahosp_dt <= chs_yr8$STDYTIME, stroke_frail := 1]


###------------###

# Cancer

chs_yr8[!is.na(chs_yr8$STDYTIME), cancer_frail :=  
  chs_yr7$cancer_frail[!is.na(chs_yr8$STDYTIME)]]


# Compile chronic illness

chs_yr8[,conditions_frail := 
  dm_frail+
  htn_frail+
  copd_frail+
  asthma_frail+
  djd_frail+
  ckd_frail+
  chf_frail+
  chd_frail+
  stroke_frail+
  cancer_frail]

chs_yr8[conditions_frail <= 4, illness_frail := 0]
chs_yr8[conditions_frail > 4, illness_frail := 1]

## Weight loss

chs_yr8[WEIGHT59==1, wtloss_frail := 1]
chs_yr8[WEIGHT59 %in% c(2,3,4), wtloss_frail := 0]

chs_yr8[,total_frail := 
  fatigue_frail+
  resistance_frail+
  ambulate_frail+
  illness_frail+
  wtloss_frail]


#-----------------------------------------------------------------------------------#

######### FRIED

##### Walking speed

#### Men:
# Height ≤ 173 cm (68.1 in):  ≤0.96 m/s      
# Height >173 cm (68.1 in):   ≤1.04  m/s	
# 


chs_yr8$gaitspeed_fried[!is.na(chs_yr8$TIME27)] <- 0 

chs_yr8$gaitspeed_fried[chs_yr8$gender_ref==1&
                          chs_yr8$height_cm > 173&
                          chs_yr8$TIME27 >= 6] <- 1

chs_yr8$gaitspeed_fried[chs_yr8$gender_ref==1&
                          chs_yr8$height_cm <= 173&
                          chs_yr8$TIME27 >= 7] <- 1


### Women
# Height ≤ 159 cm (62.6 in):  ≤0.90m/s
# Height >159 cm (62.6 in):   ≤1.02 m/s


chs_yr8$gaitspeed_fried[chs_yr8$gender_ref==0&
                          chs_yr8$height_cm>159&
                          chs_yr8$TIME27 >= 6] <- 1

chs_yr8$gaitspeed_fried[chs_yr8$gender_ref==0&
                          chs_yr8$height_cm <= 159&
                          chs_yr8$TIME27 >= 7] <- 1

########### Grip (kg)

# Trials for each hand

## Uses BMI for normalization

##### Men
# BMI ≤ 24.0:     ≤ 29 kg
# BMI 24.1-26.0:  ≤ 30 kg
# BMI 26.1-28.0:  ≤ 30 kg
# BMI > 28.0:     ≤ 32 kg

##### Women
# BMI ≤ 23.0:     ≤ 17
# BMI 23.1-26.0:  ≤ 17.3 kg
# BMI 26.1-29.0:  ≤ 18 kg
# BMI > 29.0:     ≤ 21 kg

chs_yr8$grip_avg <- round(apply(chs_yr8[,c(57:59,61:63)],MARGIN=1,mean),1)
chs_yr8$grip_max <- round(apply(chs_yr8[,c(57:59,61:63)],MARGIN=1,max),1)

chs_yr8$grip_fried[!is.na(chs_yr8$grip_max)] <- 0 

# Men

chs_yr8$grip_fried[chs_yr8$BMI<=24&
                     chs_yr8$grip_max <= 29&
                     chs_yr8$gender_ref==1] <- 1 

chs_yr8$grip_fried[chs_yr8$BMI>24&chs_yr8$BMI<=26&
                     chs_yr8$grip_max <= 30&
                     chs_yr8$gender_ref==1] <- 1 

chs_yr8$grip_fried[chs_yr8$BMI>26&chs_yr8$BMI<=28&
                     chs_yr8$grip_max <= 30&
                     chs_yr8$gender_ref==1] <- 1 

chs_yr8$grip_fried[chs_yr8$BMI>28&
                     chs_yr8$grip_max <= 32&
                     chs_yr8$gender_ref==1] <- 1 

# Women

chs_yr8$grip_fried[chs_yr8$BMI<=23&
                     chs_yr8$grip_max <= 17&
                     chs_yr8$gender_ref==0] <- 1 

chs_yr8$grip_fried[chs_yr8$BMI>23&chs_yr8$BMI<=26&
                     chs_yr8$grip_max <= 17.3&
                     chs_yr8$gender_ref==0] <- 1 

chs_yr8$grip_fried[chs_yr8$BMI>26&chs_yr8$BMI<=29&
                     chs_yr8$grip_max <= 18&
                     chs_yr8$gender_ref==0] <- 1 

chs_yr8$grip_fried[chs_yr8$BMI>29&
                     chs_yr8$grip_max <= 21&
                     chs_yr8$gender_ref==0] <- 1 


chs_yr8$activity_fried[chs_yr8$GEND01==1&chs_yr8$CAL65 < 383] <- 1
chs_yr8$activity_fried[chs_yr8$GEND01==1&chs_yr8$CAL65 >= 383] <- 0

chs_yr8$activity_fried[chs_yr8$GEND01==0&chs_yr8$CAL65 < 270] <- 1
chs_yr8$activity_fried[chs_yr8$GEND01==0&chs_yr8$CAL65 >= 270] <- 0

chs_yr8$wtloss_fried[chs_yr8$WEIGHT59==1] <- 1
chs_yr8$wtloss_fried[chs_yr8$WEIGHT59 %in% c(2,3,4)] <- 0

chs_yr8$fatigue_fried[chs_yr8$EFFORT05 %in% c(2,3)|chs_yr8$GETGO05 %in% c(2,3)] <- 1
chs_yr8$fatigue_fried[chs_yr8$EFFORT05 %in% c(0,1)|chs_yr8$GETGO05 %in% c(0,1)] <- 0


chs_yr8$total_fried <- 
  chs_yr8$wtloss_fried+
  chs_yr8$grip_fried+
  chs_yr8$fatigue_fried+
  chs_yr8$gaitspeed_fried+
  chs_yr8$activity_fried


chs_yr8$visit <- 8


chs_melt_yr8 <- melt(subset(chs_yr8,!is.na(STDYTIME)),
                     id.vars=c("patientid","STDYTIME","PERSTAT","visit"),
                     na.rm=T,
                     factorsAsStrings = T,
                     variable.factor=F)

setnames(chs_melt_yr8, c("STDYTIME","PERSTAT"),c("visitdays","cohort"))

chs_melt_yr8$form <- "yr8final"


#----------------------------------------------------------------------------------------------------------------------#

#### CHS Year 9 ####

chs_yr9 <- fread("~/Dropbox/BioLINCC files/CHS/Main_Study/YEAR9/yr9.csv",na.strings=c("NA","","NULL"))
names(chs_yr9) <- toupper(names(chs_yr9))

chs_yr9$agey <- 
  floor(chs_baseboth$agebl+
          (chs_yr9$STDYTIME/365))

chs_yr9 <- merge(chs_yr9,
                 chs_outcomes,
                 by.x="NEWID",
                 by.y="patientid",
                 all.x=T,
                 all.y=F)


chs_yr9$weight_kg <- chs_yr9$WEIGHT/2.2  
chs_yr9$height_cm <- chs_yr8$height_cm  
chs_yr9$gender_ref <- chs_yr8$gender_ref
chs_yr9$BMI <- calc_bmi(chs_yr9)

#----------------------------#

####### FRAIL score

## Fatigue, Resistance, Ambulation encoded in 'criteriaset'

## Fatigue

chs_yr9$fatigue_frail[chs_yr9$EFFORT05 %in% c(2,3)|chs_yr9$GETGO05 %in% c(2,3)] <- 1
chs_yr9$fatigue_frail[chs_yr9$EFFORT05 %in% c(0,1)|chs_yr9$GETGO05 %in% c(0,1)] <- 0

## Resistance

chs_yr9$resistance_frail[chs_yr9$STEPS09 %in% c(0,1)] <- 
  chs_yr9$STEPS09[chs_yr9$STEPS09 %in% c(0,1)]

## Ambulate

chs_yr9$ambulate_frail[chs_yr9$WHMILE09 %in% c(0,1)] <- 
  chs_yr9$WHMILE09[chs_yr9$WHMILE09 %in% c(0,1)]

# FRAIL Illness score counts presence of HTN, DM, cancer, chronic lung disease, MI, CHD, angina, asthma, arthritis, stroke, and ckd
# Scores for 0-4 conditions, score = 0, for ≥ 5, score=1


#----------------------------#

# DM

chs_yr9$dm_frail[!is.na(chs_yr9$STDYTIME)] <- chs_yr8$dm_frail[!is.na(chs_yr9$STDYTIME)]
chs_yr9$dm_frail[chs_yr9$DIABADA %in% c(1,2)] <- 0
chs_yr9$dm_frail[chs_yr9$DIABADA %in% c(3)] <- 1


#----------------------------#

# HTN

chs_yr9$htn_frail[!is.na(chs_yr9$STDYTIME)] <- chs_yr8$htn_frail[!is.na(chs_yr9$STDYTIME)]
chs_yr9$htn_frail[chs_yr9$HYPER %in% c(3)] <- 1

###------------###

# COPD (chronic lung disease)

chs_yr9$copd_frail[!is.na(chs_yr9$STDYTIME)] <- chs_yr8$copd_frail[!is.na(chs_yr9$STDYTIME)]
chs_yr9$copd_frail[chs_yr9$EMPHYS59 %in% c(2,3)] <- 1


###------------###

# Asthma

chs_yr9$asthma_frail[!is.na(chs_yr9$STDYTIME)] <- chs_yr8$asthma_frail[!is.na(chs_yr9$STDYTIME)]
chs_yr9$asthma_frail[chs_yr9$ASTHMA59 %in% c(2,3)] <- 1


###------------###

# DJD (arthritis)

chs_yr9$djd_frail[!is.na(chs_yr9$STDYTIME)] <- chs_yr8$djd_frail[!is.na(chs_yr9$STDYTIME)]
chs_yr9$djd_frail[chs_yr9$ARTHND59==1|
                    chs_yr9$ARTSHD59==1|
                    chs_yr9$ARTHIP59==1|
                    chs_yr9$ARTTRT59==1] <- 1  


###------------###

# CKD (renal disease)


chs_yr9$ckd_frail[!is.na(chs_yr9$STDYTIME)] <- chs_yr8$ckd_frail[!is.na(chs_yr9$STDYTIME)]
chs_yr9$ckd_frail[chs_yr9$MDRD44CLB9 < 60] <- 1


###------------###

# CHF/MI

chs_yr9$chf_frail[!is.na(chs_yr9$STDYTIME)] <- chs_yr8$chf_frail[!is.na(chs_yr9$STDYTIME)]
chs_yr9$chf_frail[chs_yr9$hfhosp_status==1&
                    chs_yr9$hfhosp_dt <= chs_yr9$STDYTIME] <- 1

###------------###

# CAD

chs_yr9$chd_frail[!is.na(chs_yr9$STDYTIME)] <- chs_yr8$chd_frail[!is.na(chs_yr9$STDYTIME)]
chs_yr9$chd_frail[chs_yr9$cadhosp_status==1&
                    chs_yr9$cadhosp_dt <= chs_yr9$STDYTIME] <- 1

###------------###

# Stroke

chs_yr9$stroke_frail[!is.na(chs_yr9$STDYTIME)] <- chs_yr8$stroke_frail[!is.na(chs_yr9$STDYTIME)]
chs_yr9$stroke_frail[chs_yr9$cvahosp_status==1&
                       chs_yr9$cvahosp_dt <= chs_yr9$STDYTIME] <- 1


###------------###

# Cancer

chs_yr9$cancer_frail[!is.na(chs_yr9$STDYTIME)] <- chs_yr8$cancer_frail[!is.na(chs_yr9$STDYTIME)]

###------------###

# Compile chronic illness

chs_yr9$conditions_frail <- 
  chs_yr9$dm_frail+
  chs_yr9$htn_frail+
  chs_yr9$copd_frail+
  chs_yr9$asthma_frail+
  chs_yr9$djd_frail+
  chs_yr9$ckd_frail+
  chs_yr9$chf_frail+
  chs_yr9$chd_frail+
  chs_yr9$stroke_frail+
  chs_yr9$cancer_frail

chs_yr9$illness_frail[chs_yr9$conditions_frail <= 4] <- 0
chs_yr9$illness_frail[chs_yr9$conditions_frail > 4] <- 1

## Weight loss

chs_yr9$wtloss_frail[chs_yr9$WEIGHT59==1] <- 1
chs_yr9$wtloss_frail[chs_yr9$WEIGHT59 %in% c(2,3,4)] <- 0


chs_yr9$total_frail <- chs_yr9$fatigue_frail+
  chs_yr9$resistance_frail+
  chs_yr9$ambulate_frail+
  chs_yr9$illness_frail+
  chs_yr9$wtloss_frail


#-----------------------------------------------------------------------------------#

######### FRIED

##### Walking speed

#### Men:
# Height ≤ 173 cm (68.1 in):  ≤0.96 m/s      
# Height >173 cm (68.1 in):   ≤1.04  m/s	
# 


chs_yr9$gaitspeed_fried[!is.na(chs_yr9$TIME27)] <- 0 
chs_yr9$gaitspeed_fried[chs_yr9$gender_ref==1&
                          chs_yr9$height_cm>173&
                          chs_yr9$TIME27 >= 6] <- 1

chs_yr9$gaitspeed_fried[chs_yr9$gender_ref==1&
                          chs_yr9$height_cm <= 173&
                          chs_yr9$TIME27 >=7] <- 1


### Women
# Height ≤ 159 cm (62.6 in):  ≤0.90m/s
# Height >159 cm (62.6 in):   ≤1.02 m/s


chs_yr9$gaitspeed_fried[chs_yr9$gender_ref==0&
                          chs_yr9$height_cm>159&
                          chs_yr9$TIME27 >= 6] <- 1

chs_yr9$gaitspeed_fried[chs_yr9$gender_ref==0&
                          chs_yr9$height_cm <= 159&
                          chs_yr9$TIME27 >= 7] <- 1

########### Grip (kg)

# Trials for each hand

## Uses BMI for normalization

##### Men
# BMI ≤ 24.0:     ≤ 29 kg
# BMI 24.1-26.0:  ≤ 30 kg
# BMI 26.1-28.0:  ≤ 30 kg
# BMI > 28.0:     ≤ 32 kg

##### Women
# BMI ≤ 23.0:     ≤ 17
# BMI 23.1-26.0:  ≤ 17.3 kg
# BMI 26.1-29.0:  ≤ 18 kg
# BMI > 29.0:     ≤ 21 kg

chs_yr9$grip_avg <- round(apply(chs_yr9[,c(339:341,343:345)],MARGIN=1,mean),1)
chs_yr9$grip_max <- round(apply(chs_yr9[,c(339:341,343:345)],MARGIN=1,max),1)

chs_yr9$grip_fried[!is.na(chs_yr9$grip_max)] <- 0 

# Men

chs_yr9$grip_fried[chs_yr9$BMI<=24&
                     chs_yr9$grip_max <= 29&
                     chs_yr9$gender_ref==1] <- 1 

chs_yr9$grip_fried[chs_yr9$BMI>24&chs_yr9$BMI<=26&
                     chs_yr9$grip_max <= 30&
                     chs_yr9$gender_ref==1] <- 1 

chs_yr9$grip_fried[chs_yr9$BMI>26&chs_yr9$BMI<=28&
                     chs_yr9$grip_max <= 30&
                     chs_yr9$gender_ref==1] <- 1 

chs_yr9$grip_fried[chs_yr9$BMI>28&
                     chs_yr9$grip_max <= 32&
                     chs_yr9$gender_ref==1] <- 1 

# Women

chs_yr9$grip_fried[chs_yr9$BMI<=23&
                     chs_yr9$grip_max <= 17&
                     chs_yr9$gender_ref==0] <- 1 

chs_yr9$grip_fried[chs_yr9$BMI>23&chs_yr9$BMI<=26&
                     chs_yr9$grip_max <= 17.3&
                     chs_yr9$gender_ref==0] <- 1 

chs_yr9$grip_fried[chs_yr9$BMI>26&chs_yr9$BMI<=29&
                     chs_yr9$grip_max <= 18&
                     chs_yr9$gender_ref==0] <- 1 

chs_yr9$grip_fried[chs_yr9$BMI>29&
                     chs_yr9$grip_max <= 21&
                     chs_yr9$gender_ref==0] <- 1 


chs_yr9$activity_fried[chs_yr9$GEND01==1&chs_yr9$KCAL < 383] <- 1
chs_yr9$activity_fried[chs_yr9$GEND01==1&chs_yr9$KCAL >= 383] <- 0

chs_yr9$activity_fried[chs_yr9$GEND01==0&chs_yr9$KCAL < 270] <- 1
chs_yr9$activity_fried[chs_yr9$GEND01==0&chs_yr9$KCAL >= 270] <- 0

chs_yr9$wtloss_fried[chs_yr9$WEIGHT59==1] <- 1
chs_yr9$wtloss_fried[chs_yr9$WEIGHT59 %in% c(2,3,4)] <- 0

chs_yr9$fatigue_fried[chs_yr9$EFFORT05 %in% c(2,3)|chs_yr9$GETGO05 %in% c(2,3)] <- 1
chs_yr9$fatigue_fried[chs_yr9$EFFORT05 %in% c(0,1)|chs_yr9$GETGO05 %in% c(0,1)] <- 0


chs_yr9$total_fried <- 
  chs_yr9$wtloss_fried+
  chs_yr9$grip_fried+
  chs_yr9$fatigue_fried+
  chs_yr9$gaitspeed_fried+
  chs_yr9$activity_fried


chs_yr9$visit <- 9


chs_melt_yr9 <- melt(subset(chs_yr9,!is.na(STDYTIME)),
                     id.vars=c("NEWID","STDYTIME","PERSTAT","visit"),
                     na.rm=T,
                     factorsAsStrings = T,
                     variable.factor=F)
names(chs_melt_yr9)[1:3] <- c("patientid","visitdays","cohort")
chs_melt_yr9$visit <- 9
chs_melt_yr9$form <- "yr9final"


#----------------------------------------------------------------------------------------------------------------------#

#### CHS Year 10 ####

chs_yr10 <- fread("~/Dropbox/BioLINCC files/CHS/Main_Study/YEAR10/yr10.csv",na.strings=c("NA","","NULL"))
names(chs_yr10) <- toupper(names(chs_yr10))

chs_yr10$agey <- 
  floor(chs_baseboth$agebl+
          (chs_yr10$STDYTIME/365))


####### FRAIL score

## Fatigue, Resistance, Ambulation encoded in 'criteriaset'

# FRAIL Illness score counts presence of HTN, DM, cancer, chronic lung disease, MI, CHD, angina, asthma, arthritis, stroke, and ckd
# Scores for 0-4 conditions, score = 0, for ≥ 5, score=1


## Fatigue

chs_yr10$fatigue_frail[chs_yr10$EFFORT05 %in% c(2,3)|chs_yr10$GETGO05 %in% c(2,3)] <- 1
chs_yr10$fatigue_frail[chs_yr10$EFFORT05 %in% c(0,1)|chs_yr10$GETGO05 %in% c(0,1)] <- 0

## Resistance

chs_yr10$resistance_frail <- chs_yr10$STEPS09

## Ambulate

chs_yr10$ambulate_frail <- chs_yr10$WHMILE09


#----------------------------#

# DM

chs_yr10$dm_frail[!is.na(chs_yr10$STDYTIME)] <- chs_yr9$dm_frail[!is.na(chs_yr10$STDYTIME)]
chs_yr10$dm_frail[chs_yr10$DIABADA %in% c(1,2)] <- 0
chs_yr10$dm_frail[chs_yr10$DIABADA %in% c(3)] <- 1


#----------------------------#

# HTN

chs_yr10$htn_frail[!is.na(chs_yr10$STDYTIME)] <- chs_yr9$htn_frail[!is.na(chs_yr10$STDYTIME)]
chs_yr10$htn_frail[chs_yr10$HYPER %in% c(3)] <- 1

###------------###

# COPD (chronic lung disease)

chs_yr10$copd_frail[!is.na(chs_yr10$STDYTIME)] <- chs_yr9$copd_frail[!is.na(chs_yr10$STDYTIME)]
chs_yr10$copd_frail[chs_yr10$EMPHYS59 %in% c(2,3)] <- 1


###------------###

# Asthma

chs_yr10$asthma_frail[!is.na(chs_yr10$STDYTIME)] <- chs_yr9$asthma_frail[!is.na(chs_yr10$STDYTIME)]
chs_yr10$asthma_frail[chs_yr10$ASTHMA59 %in% c(2,3)] <- 1


###------------###

# DJD (arthritis)

chs_yr10$djd_frail[!is.na(chs_yr10$STDYTIME)] <- chs_yr9$djd_frail[!is.na(chs_yr10$STDYTIME)]
chs_yr10$djd_frail[chs_yr10$ARTHND59==1|
                     chs_yr10$ARTSHD59==1|
                     chs_yr10$ARTHIP59==1|
                     chs_yr10$ARTTRT59==1] <- 1  


###------------###

# CKD (renal disease)


chs_yr10$ckd_frail[!is.na(chs_yr10$STDYTIME)] <- chs_yr9$ckd_frail[!is.na(chs_yr10$STDYTIME)]


###------------###

# CHF/MI

chs_yr10$chf_frail[!is.na(chs_yr10$STDYTIME)] <- chs_yr9$chf_frail[!is.na(chs_yr10$STDYTIME)]
chs_yr10$chf_frail[chs_yr10$hfhosp_status==1&
                     chs_yr10$hfhosp_dt <= chs_yr10$STDYTIME] <- 1

###------------###

# CAD

chs_yr10$chd_frail[!is.na(chs_yr10$STDYTIME)] <- chs_yr9$chd_frail[!is.na(chs_yr10$STDYTIME)]
chs_yr10$chd_frail[chs_yr10$cadhosp_status==1&
                     chs_yr10$cadhosp_dt <= chs_yr10$STDYTIME] <- 1

###------------###

# Stroke

chs_yr10$stroke_frail[!is.na(chs_yr10$STDYTIME)] <- chs_yr9$stroke_frail[!is.na(chs_yr10$STDYTIME)]
chs_yr10$stroke_frail[chs_yr10$cvahosp_status==1&
                        chs_yr10$cvahosp_dt <= chs_yr10$STDYTIME] <- 1


###------------###

# Cancer

chs_yr10$cancer_frail[!is.na(chs_yr10$STDYTIME)] <- chs_yr9$cancer_frail[!is.na(chs_yr10$STDYTIME)]

###------------###

# Compile chronic illness

chs_yr10$conditions_frail <- 
  chs_yr10$dm_frail+
  chs_yr10$htn_frail+
  chs_yr10$copd_frail+
  chs_yr10$asthma_frail+
  chs_yr10$djd_frail+
  chs_yr10$ckd_frail+
  chs_yr10$chf_frail+
  chs_yr10$chd_frail+
  chs_yr10$stroke_frail+
  chs_yr10$cancer_frail

chs_yr10$illness_frail[chs_yr10$conditions_frail <= 4] <- 0
chs_yr10$illness_frail[chs_yr10$conditions_frail > 4] <- 1


## Weight loss

chs_yr10$wtloss_frail[chs_yr10$WEIGHT59==1] <- 1
chs_yr10$wtloss_frail[chs_yr10$WEIGHT59 %in% c(2,3,4)] <- 0


chs_yr10$total_frail <- chs_yr10$fatigue_frail+
  chs_yr10$resistance_frail+
  chs_yr10$ambulate_frail+
  chs_yr10$illness_frail+
  chs_yr10$wtloss_frail


chs_melt_yr10 <- melt(subset(chs_yr10,!is.na(STDYTIME)),
                      id.vars=c("NEWID","STDYTIME","PERSTAT"),
                      na.rm=T,
                      factorsAsStrings = T,
                      variable.factor=F)
names(chs_melt_yr10)[1:3] <- c("patientid","visitdays","cohort")
chs_melt_yr10$visit <- 10

chs_melt_yr10$form <- "yr10final"


#----------------------------------------------------------------------------------------------------------------------#

#### CHS Year 11 ####

chs_yr11<- fread("~/Dropbox/BioLINCC files/CHS/Main_Study/YEAR11/yr11.csv",na.strings=c("NA","","NULL"))
names(chs_yr11) <- toupper(names(chs_yr11))

chs_yr11$agey <- floor(chs_baseboth$agebl+(chs_yr11$STDYTIME/365))


####### FRAIL score

## Fatigue, Resistance, Ambulation encoded in 'criteriaset'

# FRAIL Illness score counts presence of HTN, DM, cancer, chronic lung disease, MI, CHD, angina, asthma, arthritis, stroke, and ckd
# Scores for 0-4 conditions, score = 0, for ≥ 5, score=1


## Fatigue

chs_yr11$fatigue_frail[chs_yr11$EFFORT05 %in% c(2,3)|chs_yr11$GETGO05 %in% c(2,3)] <- 1
chs_yr11$fatigue_frail[chs_yr11$EFFORT05 %in% c(0,1)|chs_yr11$GETGO05 %in% c(0,1)] <- 0

## Resistance

chs_yr11$resistance_frail[chs_yr11$STEPS09 %in% c(0,1)] <- 
  chs_yr11$STEPS09[chs_yr11$STEPS09 %in% c(0,1)]

## Ambulate

chs_yr11$ambulate_frail[chs_yr11$WHMILE09 %in% c(0,1)] <- 
  chs_yr11$WHMILE09[chs_yr11$WHMILE09 %in% c(0,1)]


#----------------------------#

# DM

chs_yr11$dm_frail[!is.na(chs_yr11$STDYTIME)] <- 
  chs_yr10$dm_frail[!is.na(chs_yr11$STDYTIME)]
chs_yr11$dm_frail[chs_yr11$DIABADA %in% c(1,2)] <- 0
chs_yr11$dm_frail[chs_yr11$DIABADA %in% c(3)] <- 1


#----------------------------#

# HTN

chs_yr11$htn_frail[!is.na(chs_yr11$STDYTIME)] <- chs_yr10$htn_frail[!is.na(chs_yr11$STDYTIME)]
chs_yr11$htn_frail[chs_yr11$HYPER %in% c(3)] <- 1

###------------###

# COPD (chronic lung disease)

chs_yr11$copd_frail[!is.na(chs_yr11$STDYTIME)] <- chs_yr10$copd_frail[!is.na(chs_yr11$STDYTIME)]
chs_yr11$copd_frail[chs_yr11$EMPHYS59 %in% c(2,3)] <- 1


###------------###

# Asthma

chs_yr11$asthma_frail[!is.na(chs_yr11$STDYTIME)] <- chs_yr10$asthma_frail[!is.na(chs_yr11$STDYTIME)]
chs_yr11$asthma_frail[chs_yr11$ASTHMA59 %in% c(2,3)] <- 1


###------------###

# DJD (arthritis)

chs_yr11$djd_frail[!is.na(chs_yr11$STDYTIME)] <- chs_yr10$djd_frail[!is.na(chs_yr11$STDYTIME)]
chs_yr11$djd_frail[chs_yr11$ARTHND59==1|
                     chs_yr11$ARTSHD59==1|
                     chs_yr11$ARTHIP59==1|
                     chs_yr11$ARTTRT59==1] <- 1  


###------------###

# CKD (renal disease)


chs_yr11$ckd_frail[!is.na(chs_yr11$STDYTIME)] <- chs_yr10$ckd_frail[!is.na(chs_yr11$STDYTIME)]


###------------###

# CHF/MI

chs_yr11$chf_frail[!is.na(chs_yr11$STDYTIME)] <- chs_yr10$chf_frail[!is.na(chs_yr11$STDYTIME)]
chs_yr11$chf_frail[chs_yr11$hfhosp_status==1&
                     chs_yr11$hfhosp_dt <= chs_yr11$STDYTIME] <- 1

###------------###

# CAD

chs_yr11$chd_frail[!is.na(chs_yr11$STDYTIME)] <- chs_yr10$chd_frail[!is.na(chs_yr11$STDYTIME)]
chs_yr11$chd_frail[chs_yr11$cadhosp_status==1&
                     chs_yr11$cadhosp_dt <= chs_yr11$STDYTIME] <- 1

###------------###

# Stroke

chs_yr11$stroke_frail[!is.na(chs_yr11$STDYTIME)] <- chs_yr10$stroke_frail[!is.na(chs_yr11$STDYTIME)]
chs_yr11$stroke_frail[chs_yr11$cvahosp_status==1&
                        chs_yr11$cvahosp_dt <= chs_yr11$STDYTIME] <- 1


###------------###

# Cancer

chs_yr11$cancer_frail[!is.na(chs_yr11$STDYTIME)] <- chs_yr10$cancer_frail[!is.na(chs_yr11$STDYTIME)]

###------------###

# Compile chronic illness

chs_yr11$conditions_frail <- 
  chs_yr11$dm_frail+
  chs_yr11$htn_frail+
  chs_yr11$copd_frail+
  chs_yr11$asthma_frail+
  chs_yr11$djd_frail+
  chs_yr11$ckd_frail+
  chs_yr11$chf_frail+
  chs_yr11$chd_frail+
  chs_yr11$stroke_frail+
  chs_yr11$cancer_frail

chs_yr11$illness_frail[chs_yr11$conditions_frail <= 4] <- 0
chs_yr11$illness_frail[chs_yr11$conditions_frail > 4] <- 1


## Weight loss

chs_yr11$wtloss_frail[chs_yr11$WEIGHT59==1] <- 1
chs_yr11$wtloss_frail[chs_yr11$WEIGHT59 %in% c(2,3,4)] <- 0


chs_yr11$total_frail <- chs_yr11$fatigue_frail+
  chs_yr11$resistance_frail+
  chs_yr11$ambulate_frail+
  chs_yr11$illness_frail+
  chs_yr11$wtloss_frail


chs_yr11$visit <- 11


chs_melt_yr11 <- melt(subset(chs_yr11,!is.na(STDYTIME)),
                      id.vars=c("NEWID","STDYTIME","PERSTAT","visit"),
                      na.rm=T,
                      factorsAsStrings = T,
                      variable.factor=F)
names(chs_melt_yr11)[1:3] <- c("patientid","visitdays","cohort")

chs_melt_yr11$form <- "yr11final"


#----------------------------------------------------------------------------------------------------------------------#

#### CHS Year 12 ####

chs_yr12 <- fread("~/Dropbox/BioLINCC files/CHS/Main_Study/YEAR12/yr12ann.csv",na.strings=c("NA","","NULL"))
chs_yr12_phone <- fread("~/Dropbox/BioLINCC files/CHS/Main_Study/YEAR12/yr12ph.csv",na.strings=c("NA","","NULL"))
# chs_yr12 <- merge(chs_yr12,chs_yr11[,c("NEWID","WHMILE09","STEPS09")],by = "NEWID", all.x=T)

names(chs_yr12) <- toupper(names(chs_yr12))
names(chs_yr12_phone) <- toupper(names(chs_yr12_phone))

chs_yr12$agey <- floor(chs_baseboth$agebl+(chs_yr12$STDYTIME/365))


chs_melt_yr12 <- melt(subset(chs_yr12,!is.na(STDYTIME)),
                      id.vars=c("NEWID","STDYTIME","PERSTAT"),
                      na.rm=T,
                      factorsAsStrings = T,
                      variable.factor=F)


chs_melt_yr12$form <- "yr12final"
chs_melt_yr12$visit <- 12


names(chs_yr12_phone) <- toupper(names(chs_yr12_phone))


chs_melt_phone_yr12 <- melt(subset(chs_yr12_phone,!is.na(STDYTIME)),
                            id.vars=c("NEWID","STDYTIME","PERSTAT"),
                            na.rm=T,
                            factorsAsStrings = T)

names(chs_melt_yr12)[1:3] <- c("patientid","visitdays","cohort")
names(chs_melt_phone_yr12)[1:3] <- c("patientid","visitdays","cohort")
chs_melt_phone_yr12$visit <- 12

chs_melt_phone_yr12$form <- "yr12_phone"


#----------------------------------------------------------------------------------------------------------------------#

#### CHS Year 13 ####

chs_yr13 <- fread("~/Dropbox/BioLINCC files/CHS/Main_Study/YEAR13/yr13ann.csv",na.strings=c("NA","","NULL"))
chs_yr13_phone <- fread("~/Dropbox/BioLINCC files/CHS/Main_Study/YEAR13/yr13ph.csv",na.strings=c("NA","","NULL"))
names(chs_yr13) <- toupper(names(chs_yr13))

chs_yr13$agey <- floor(chs_baseboth$agebl+(chs_yr13$STDYTIME/365))

chs_melt_yr13 <- melt(subset(chs_yr13,!is.na(STDYTIME)),
                      id.vars=c("NEWID","STDYTIME","PERSTAT"),
                      na.rm=T,
                      factorsAsStrings = T,
                      variable.factor=F)


chs_melt_yr13$form <- "yr13final"
chs_melt_yr13$visit <- 13


names(chs_yr13_phone) <- toupper(names(chs_yr13_phone))


chs_melt_phone_yr13 <- melt(subset(chs_yr13_phone,!is.na(STDYTIME)),
                            id.vars=c("NEWID","STDYTIME","PERSTAT"),
                            na.rm=T,
                            factorsAsStrings = T,
                            variable.factor=F)

names(chs_melt_yr13)[1:3] <- c("patientid","visitdays","cohort")
names(chs_melt_phone_yr13)[1:3] <- c("patientid","visitdays","cohort")
chs_melt_phone_yr13$visit <- 13

chs_melt_phone_yr13$form <- "yr13_phone"


#----------------------------------------------------------------------------------------------------------------------#

#### CHS year 14 ####

chs_yr14 <- fread("~/Dropbox/BioLINCC files/CHS/Main_Study/YEAR14/yr14ann.csv",na.strings=c("NA","","NULL"))
chs_yr14_phone <- fread("~/Dropbox/BioLINCC files/CHS/Main_Study/YEAR14/yr14ph.csv",na.strings=c("NA","","NULL"))
names(chs_yr14) <- toupper(names(chs_yr14))
names(chs_yr14_phone) <- toupper(names(chs_yr14_phone))

chs_yr14$agey <- floor(chs_baseboth$agebl+(chs_yr14$STDYTIME/365))

chs_melt_yr14 <- melt(subset(chs_yr14,!is.na(STDYTIME)),
                      id.vars=c("NEWID","STDYTIME","PERSTAT"),
                      na.rm=T,
                      factorsAsStrings = T,
                      variable.factor=F)

chs_melt_yr14$form <- "yr14final"
chs_melt_yr14$visit <- 14

chs_melt_phone_yr14 <- melt(subset(chs_yr14_phone,!is.na(STDYTIME)),
                            id.vars=c("NEWID","STDYTIME","PERSTAT"),
                            na.rm=T,
                            factorsAsStrings = T,
                            variable.factor=F)

names(chs_melt_yr14)[1:3] <- c("patientid","visitdays","cohort")
names(chs_melt_phone_yr14)[1:3] <- c("patientid","visitdays","cohort")

chs_melt_phone_yr14$visit <- 14
chs_melt_phone_yr14$form <- "yr14_phone"


#----------------------------------------------------------------------------------------------------------------------#

#### CHS year 15 ####


chs_yr15 <- fread("~/Dropbox/BioLINCC files/CHS/Main_Study/YEAR15/yr15ann.csv",na.strings=c("NA","","NULL"))
chs_yr15_phone <- fread("~/Dropbox/BioLINCC files/CHS/Main_Study/YEAR15/yr15ph.csv",na.strings=c("NA","","NULL"))
names(chs_yr15) <- toupper(names(chs_yr15))


chs_yr15$agey <- floor(chs_baseboth$agebl+(chs_yr15$STDYTIME/365))

chs_melt_yr15 <- melt(subset(chs_yr15,!is.na(STDYTIME)),
                      id.vars=c("NEWID","STDYTIME","PERSTAT"),
                      na.rm=T,
                      factorsAsStrings = T,
                      variable.factor=F)


chs_melt_yr15$form <- "yr15final"
chs_melt_yr15$visit <- 15


names(chs_yr15_phone) <- toupper(names(chs_yr15_phone))


chs_melt_phone_yr15 <- melt(subset(chs_yr15_phone,!is.na(STDYTIME)),
                            id.vars=c("NEWID","STDYTIME","PERSTAT"),
                            na.rm=T,
                            factorsAsStrings = T,
                            variable.factor=F)

names(chs_melt_yr15)[1:3] <- c("patientid","visitdays","cohort")
names(chs_melt_phone_yr15)[1:3] <- c("patientid","visitdays","cohort")
chs_melt_phone_yr15$visit <- 15

chs_melt_phone_yr15$form <- "yr15_phone"


#----------------------------------------------------------------------------------------------------------------------#

#### CHS year 16 ####

chs_yr16 <- fread("~/Dropbox/BioLINCC files/CHS/Main_Study/YEAR16/yr16ann.csv",na.strings=c("NA","","NULL"))
chs_yr16_phone <- fread("~/Dropbox/BioLINCC files/CHS/Main_Study/YEAR16/yr16ph.csv",na.strings=c("NA","","NULL"))

names(chs_yr16) <- toupper(names(chs_yr16))

chs_yr16$agey <- floor(chs_baseboth$agebl+(chs_yr16$STDYTIME/365))

chs_melt_yr16 <- melt(subset(chs_yr16,!is.na(STDYTIME)),
                      id.vars=c("NEWID","STDYTIME","PERSTAT"),
                      na.rm=T,
                      factorsAsStrings = T,
                      variable.factor=F)


chs_melt_yr16$form <- "yr16final"
chs_melt_yr16$visit <- 16


names(chs_yr16_phone) <- toupper(names(chs_yr16_phone))

chs_melt_phone_yr16 <- melt(subset(chs_yr16_phone,!is.na(STDYTIME)),
                            id.vars=c("NEWID","STDYTIME","PERSTAT"),
                            na.rm=T,
                            factorsAsStrings = T,
                            variable.factor=F)

names(chs_melt_yr16)[1:3] <- c("patientid","visitdays","cohort")
names(chs_melt_phone_yr16)[1:3] <- c("patientid","visitdays","cohort")
chs_melt_phone_yr16$visit <- 16

chs_melt_phone_yr16$form <- "yr16_phone"


#----------------------------------------------------------------------------------------------------------------------#

#### CHS year 17 ####


chs_yr17 <- fread("~/Dropbox/BioLINCC files/CHS/Main_Study/YEAR17/yr17ann.csv",na.strings=c("NA","","NULL"))
names(chs_yr17) <- toupper(names(chs_yr17))

chs_yr17$agey <- floor(chs_baseboth$agebl+(chs_yr17$STDYTIME/365))

## For some reason there were inconsistencies in coding in the YR17 visit that affected a smaller number of patients
## This corrects those, erring on the side of 'more frail'.

chs_yr17$TLTDIF[!is.na(chs_yr17$STDYTIME)&is.na(chs_yr17$TLTDIF)] <- 0
chs_yr17$BEDDIF[!is.na(chs_yr17$STDYTIME)&is.na(chs_yr17$BEDDIF)] <- 0
chs_yr17$BTHDIF[!is.na(chs_yr17$STDYTIME)&is.na(chs_yr17$BTHDIF)] <- 0
chs_yr17$DRSDIF[!is.na(chs_yr17$STDYTIME)&is.na(chs_yr17$DRSDIF)] <- 0
chs_yr17$EATDIF[!is.na(chs_yr17$STDYTIME)&is.na(chs_yr17$EATDIF)] <- 0
chs_yr17$STPDIF[!is.na(chs_yr17$STDYTIME)&is.na(chs_yr17$STPDIF)] <- 0
chs_yr17$WHODIF[!is.na(chs_yr17$STDYTIME)&is.na(chs_yr17$WHODIF)] <- 0


chs_melt_yr17 <- melt(subset(chs_yr17,!is.na(STDYTIME)),
                      id.vars=c("NEWID","STDYTIME","PERSTAT"),
                      na.rm=T,
                      factorsAsStrings = T,
                      variable.factor=F)

names(chs_melt_yr17)[1:3] <- c("patientid","visitdays","cohort")
chs_melt_yr17$visit <- 17
chs_melt_yr17$form <- "yr17final"

chs_yr17_phone <- fread("~/Dropbox/BioLINCC files/CHS/Main_Study/YEAR17/yr17ph.csv",na.strings=c("NA","","NULL"))
names(chs_yr17_phone) <- toupper(names(chs_yr17_phone))
chs_melt_phone_yr17 <- melt(subset(chs_yr17_phone,!is.na(STDYTIME)),
                            id.vars=c("NEWID","STDYTIME","PERSTAT"),
                            na.rm=T,
                            factorsAsStrings = T,
                            variable.factor=F)

names(chs_melt_phone_yr17)[1:3] <- c("patientid","visitdays","cohort")
chs_melt_phone_yr17$visit <- 17
chs_melt_phone_yr17$form <- "yr17_phone"


#----------------------------------------------------------------------------------------------------------------------#

#### CHS year 18 ####

chs_yr18_phone <- fread("~/Dropbox/BioLINCC files/CHS/Main_Study/YEAR18/yr18ph.csv",na.strings=c("NA","","NULL"))
names(chs_yr18_phone) <- toupper(names(chs_yr18_phone))

chs_melt_phone_yr18 <- melt(subset(chs_yr18_phone,!is.na(STDYTIME)),
                            id.vars=c("NEWID","STDYTIME","PERSTAT"),
                            na.rm=T,
                            factorsAsStrings = T,
                            variable.factor=F)

names(chs_melt_phone_yr18)[1:2] <- c("patientid","visitdays","cohort")
chs_melt_phone_yr18$visit <- 18
chs_melt_phone_yr18$form <- "yr18_phone"

chs_yr18_phone$agey <- floor(chs_baseboth$agebl+(chs_yr18_phone$STDYTIME/365))


chs_yr18 <- fread("~/Dropbox/BioLINCC files/CHS/Main_Study/YEAR18/yr18ann.csv",na.strings=c("NA","","NULL"))
names(chs_yr18) <- toupper(names(chs_yr18))

chs_yr18 <- cbind(chs_yr18,
                  chs_baseboth[,c("RACE01",
                                  "GEND01")])

chs_yr18$agey <- floor(chs_yr18$agey+chs_yr18$STDYTIME/365) 

chs_yr18$height_in <- chs_yr18$HEIGHT/2.54

chs_yr18$gfr_calc <- calc_MDRD4(chs_yr18,cr="CREATININE", age="agey",sex="GEND01",race="RACE01")

chs_yr18$TIME27[chs_yr18$DISTWALK==2&!is.na(chs_yr18$DISTWALK)] <- chs_yr18$WLKTIME[chs_yr18$DISTWALK==2&!is.na(chs_yr18$DISTWALK)]
chs_yr18$TIME27[chs_yr18$DISTWALK==1&!is.na(chs_yr18$DISTWALK)] <- chs_yr18$WLKTIME[chs_yr18$DISTWALK==1&!is.na(chs_yr18$DISTWALK)] * 1.524

####### FRAIL score

## Fatigue, Resistance, Ambulation encoded in 'criteriaset'

# FRAIL Illness score counts presence of HTN, DM, cancer, chronic lung disease, MI, CHD, angina, asthma, arthritis, stroke, and ckd
# Scores for 0-4 conditions, score = 0, for ≥ 5, score=1


chs_yr18 <- merge(chs_yr18, chs_yr9[,c("NEWID",
                                       "dm_frail",
                                       "htn_frail",
                                       "copd_frail",
                                       "asthma_frail",
                                       "djd_frail",
                                       "ckd_frail",
                                       "chf_frail",
                                       "chd_frail",
                                       "stroke_frail",
                                       "cancer_frail")])

chs_yr18[is.na(chs_yr18$STDYTIME),
         c("dm_frail",
           "htn_frail",
           "copd_frail",
           "asthma_frail",
           "djd_frail",
           "ckd_frail",
           "chf_frail",
           "chd_frail",
           "stroke_frail",
           "cancer_frail")] <- NA


#----------------------------#

## Fatigue

chs_yr18$fatigue_frail[chs_yr18$EFFORT %in% c(2,3)|chs_yr18$GETGO %in% c(2,3)] <- 1
chs_yr18$fatigue_frail[chs_yr18$EFFORT %in% c(0,1)|chs_yr18$GETGO %in% c(0,1)] <- 0

## Resistance

chs_yr18$resistance_frail[chs_yr18$STEPS %in% c(0,1)] <- 
  chs_yr18$STEPS[chs_yr18$STEPS %in% c(0,1)]

## Ambulate

chs_yr18$ambulate_frail[chs_yr18$WHMILE %in% c(0,1)] <- 
  chs_yr18$WHMILE[chs_yr18$WHMILE %in% c(0,1)]


#----------------------------#

# DM

chs_yr18$dm_frail[chs_yr18$DIABETES %in% c(0)] <- 0
chs_yr18$dm_frail[chs_yr18$DIABETES %in% c(1,2)] <- 1


#----------------------------#

# HTN


chs_yr18$htn_frail[chs_yr18$HYPER %in% c(3)] <- 1

###------------###

# COPD (chronic lung disease)

chs_yr18$copd_frail[chs_yr18$EMPHYS59 %in% c(2,3)] <- 1


###------------###

# Asthma

chs_yr18$asthma_frail[chs_yr18$ASTHMA59 %in% c(2,3)] <- 1


###------------###

# DJD (arthritis)

chs_yr18$djd_frail[chs_yr18$ARTHND59==1|
                     chs_yr18$ARTSHD59==1|
                     chs_yr18$ARTHIP59==1|
                     chs_yr18$ARTTRT59==1] <- 1  


###------------###

# CKD (renal disease)

chs_yr18$ckd_frail[chs_yr18$crcl < 60] <- 1


###------------###

# CHF/MI

chs_yr18$chf_frail[chs_yr18$hfhosp_status==1&
                     chs_yr18$hfhosp_dt <= chs_yr18$STDYTIME] <- 1

###------------###

# CAD

chs_yr18$chd_frail[chs_yr18$cadhosp_status==1&
                     chs_yr18$cadhosp_dt <= chs_yr18$STDYTIME] <- 1

###------------###

# Stroke

chs_yr18$stroke_frail[!is.na(chs_yr18$STDYTIME)] <- chs_yr9$stroke_frail[!is.na(chs_yr18$STDYTIME)]

chs_yr18$stroke_frail[chs_yr18$cvahosp_status==1&
                        chs_yr18$cvahosp_dt <= chs_yr18$STDYTIME] <- 1


###------------###

# Compile chronic illness

chs_yr18$conditions_frail <- 
  chs_yr18$dm_frail+
  chs_yr18$htn_frail+
  chs_yr18$copd_frail+
  chs_yr18$asthma_frail+
  chs_yr18$djd_frail+
  chs_yr18$ckd_frail+
  chs_yr18$chf_frail+
  chs_yr18$chd_frail+
  chs_yr18$stroke_frail+
  chs_yr18$cancer_frail

chs_yr18$illness_frail[chs_yr18$conditions_frail <= 4] <- 0
chs_yr18$illness_frail[chs_yr18$conditions_frail > 4] <- 1


## Weight loss

chs_yr18$wtloss_frail[chs_yr18$WEIGHT10==1] <- 1
chs_yr18$wtloss_frail[chs_yr18$WEIGHT10 %in% c(2,3,4)] <- 0


chs_yr18$total_frail <- chs_yr18$fatigue_frail+
  chs_yr18$resistance_frail+
  chs_yr18$ambulate_frail+
  chs_yr18$illness_frail+
  chs_yr18$wtloss_frail

###----###

chs_melt_yr18 <- melt(subset(chs_yr18,!is.na(STDYTIME)),
                      id.vars=c("NEWID","STDYTIME"),
                      na.rm=T,
                      factorsAsStrings = T,
                      variable.factor=F)

names(chs_melt_yr18)[1:2] <- c("patientid","visitdays")
chs_melt_yr18$visit <- 18
chs_melt_yr18$form <- "yr18_allstarsfinal"


#----------------------------------------------------------------------------------------------------------------------#

#### CHS year 19 ####

chs_yr19_phone <- fread("~/Dropbox/BioLINCC files/CHS/Main_Study/YEAR19/yr19ph.csv",na.strings=c("NA","","NULL"))
names(chs_yr19_phone) <- toupper(names(chs_yr19_phone))

chs_yr19_phone$agey <- floor(chs_baseboth$agey+(chs_yr19_phone$STDYTIME/365))


chs_melt_phone_yr19 <- melt(subset(chs_yr19_phone,!is.na(STDYTIME)),
                            id.vars=c("NEWID","STDYTIME","PERSTAT"),
                            na.rm=T,
                            factorsAsStrings = T)
names(chs_melt_phone_yr19)[1:3] <- c("patientid","visitdays","cohort")

chs_melt_phone_yr19$visit <- 19
chs_melt_phone_yr19$form <- "yr19_phone"

chs_yr19 <- 
  fread("~/Dropbox/BioLINCC files/CHS/Main_Study/YEAR19/yr19ann.csv",
        na.strings=c("NA","","NULL"))
names(chs_yr19) <- toupper(names(chs_yr19))


chs_yr19 <- 
  merge(chs_yr19, 
        chs_yr18[,c("NEWID",
                    "dm_frail",
                    "htn_frail",
                    "copd_frail",
                    "asthma_frail",
                    "djd_frail",
                    "ckd_frail",
                    "chf_frail",
                    "chd_frail",
                    "stroke_frail",
                    "cancer_frail")])

chs_yr19[is.na(chs_yr19$STDYTIME),
         c("dm_frail",
           "htn_frail",
           "copd_frail",
           "asthma_frail",
           "djd_frail",
           "ckd_frail",
           "chf_frail",
           "chd_frail",
           "stroke_frail",
           "cancer_frail")] <- NA


chs_melt_yr19 <- melt(subset(chs_yr19,!is.na(STDYTIME)),
                      id.vars=c("NEWID","STDYTIME","PERSTAT"),
                      na.rm=T,
                      factorsAsStrings = T)
names(chs_melt_yr19)[1:3] <- c("patientid","visitdays","cohort")
chs_melt_yr19$visit <- 19
chs_melt_yr19$form <- "yr19final"


#----------------------------------------------------------------------------------------------------------------------#

#### CHS year 20 ####

chs_yr20_phone <- fread("~/Dropbox/BioLINCC files/CHS/Main_Study/YEAR20/yr20ph.csv",na.strings=c("NA","","NULL"))
names(chs_yr20_phone) <- toupper(names(chs_yr20_phone))

chs_yr20_phone$agey <- floor(chs_baseboth$agey+(chs_yr20_phone$STDYTIME/365))

chs_melt_phone_yr20 <- melt(subset(chs_yr20_phone,!is.na(STDYTIME)),
                            id.vars=c("NEWID","STDYTIME","PERSTAT"),
                            na.rm=T,
                            variable.factor=F,
                            factorsAsStrings = T)
names(chs_melt_phone_yr20)[1:3] <- c("patientid","visitdays","cohort")

chs_melt_phone_yr20$visit <- 20
chs_melt_phone_yr20$form <- "yr20_phone"

chs_yr20 <- 
  fread("~/Dropbox/BioLINCC files/CHS/Main_Study/YEAR20/yr20ann.csv",
        na.strings=c("NA","","NULL"))
names(chs_yr20) <- toupper(names(chs_yr20))

chs_yr20 <- merge(chs_yr20, chs_yr19[,c("NEWID",
                                        "dm_frail",
                                        "htn_frail",
                                        "copd_frail",
                                        "asthma_frail",
                                        "djd_frail",
                                        "ckd_frail",
                                        "chf_frail",
                                        "chd_frail",
                                        "stroke_frail",
                                        "cancer_frail")])

chs_yr20[is.na(chs_yr20$STDYTIME),
         c("dm_frail",
           "htn_frail",
           "copd_frail",
           "asthma_frail",
           "djd_frail",
           "ckd_frail",
           "chf_frail",
           "chd_frail",
           "stroke_frail",
           "cancer_frail")] <- NA


chs_melt_yr20 <- melt(subset(chs_yr20,!is.na(STDYTIME)),
                      id.vars=c("NEWID","STDYTIME","PERSTAT"),
                      na.rm=T,
                      factorsAsStrings = T,
                      variable.factor=F)
names(chs_melt_yr20)[1:3] <- c("patientid","visitdays","cohort")
chs_melt_yr20$visit <- 20
chs_melt_yr20$form <- "yr20final"


#----------------------------------------------------------------------------------------------------------------------#

#### CHS year 21 ####

chs_yr21_phone <- fread("~/Dropbox/BioLINCC files/CHS/Main_Study/YEAR21/yr21ph.csv",na.strings=c("NA","","NULL"))
names(chs_yr21_phone) <- toupper(names(chs_yr21_phone))

chs_yr21_phone$agey <- floor(chs_baseboth$agey+(chs_yr21_phone$STDYTIME/365))


chs_melt_phone_yr21 <- melt(subset(chs_yr21_phone,!is.na(STDYTIME)),
                            id.vars=c("NEWID","STDYTIME","PERSTAT"),
                            na.rm=T,
                            factorsAsStrings = T,
                            variable.factor = F)
names(chs_melt_phone_yr21)[1:3] <- c("patientid","visitdays","cohort")
chs_melt_phone_yr21$visit <- 21
chs_melt_phone_yr21$form <- "yr21_phone"

chs_yr21 <- fread("~/Dropbox/BioLINCC files/CHS/Main_Study/YEAR21/yr21ann.csv",na.strings=c("NA","","NULL"))
names(chs_yr21) <- toupper(names(chs_yr21))

chs_yr21 <- merge(chs_yr21, chs_yr20[,c("NEWID",
                                        "dm_frail",
                                        "htn_frail",
                                        "copd_frail",
                                        "asthma_frail",
                                        "djd_frail",
                                        "ckd_frail",
                                        "chf_frail",
                                        "chd_frail",
                                        "stroke_frail",
                                        "cancer_frail")])

chs_yr21[is.na(chs_yr21$STDYTIME),
         c("dm_frail",
           "htn_frail",
           "copd_frail",
           "asthma_frail",
           "djd_frail",
           "ckd_frail",
           "chf_frail",
           "chd_frail",
           "stroke_frail",
           "cancer_frail")] <- NA


chs_melt_yr21 <- melt(subset(chs_yr21,!is.na(STDYTIME)),
                      id.vars=c("NEWID","STDYTIME","PERSTAT"),
                      na.rm=T,
                      factorsAsStrings = T,
                      variable.factor = F)
names(chs_melt_yr21)[1:3] <- c("patientid","visitdays","cohort")
chs_melt_yr21$visit <- 21
chs_melt_yr21$form <- "yr21final"


#----------------------------------------------------------------------------------------------------------------------#

#### CHS year 22 ####

chs_yr22_phone <- fread("~/Dropbox/BioLINCC files/CHS/Main_Study/YEAR22/yr22ph.csv",na.strings=c("NA","","NULL"))
names(chs_yr22_phone) <- toupper(names(chs_yr22_phone))

chs_melt_phone_yr22 <- melt(subset(chs_yr22_phone,!is.na(STDYTIME)),
                            id.vars=c("NEWID","STDYTIME","PERSTAT"),
                            na.rm=T,
                            factorsAsStrings = T,
                            variable.factor=F)
names(chs_melt_phone_yr22)[1:3] <- c("patientid","visitdays","cohort")
chs_melt_phone_yr22$visit <- 22
chs_melt_phone_yr22$form <- "yr22_phone"

chs_yr22 <- fread("~/Dropbox/BioLINCC files/CHS/Main_Study/YEAR22/yr22ann.csv",na.strings=c("NA","","NULL"))
names(chs_yr22) <- toupper(names(chs_yr22))

chs_yr22$agey <- floor(chs_baseboth$agey+(chs_yr22$STDYTIME/365))


chs_yr22 <- merge(chs_yr22, chs_yr21[,c("NEWID",
                                        "dm_frail",
                                        "htn_frail",
                                        "copd_frail",
                                        "asthma_frail",
                                        "djd_frail",
                                        "ckd_frail",
                                        "chf_frail",
                                        "chd_frail",
                                        "stroke_frail",
                                        "cancer_frail")])

chs_yr22[is.na(chs_yr22$STDYTIME),
         c("dm_frail",
           "htn_frail",
           "copd_frail",
           "asthma_frail",
           "djd_frail",
           "ckd_frail",
           "chf_frail",
           "chd_frail",
           "stroke_frail",
           "cancer_frail")] <- NA


chs_melt_yr22 <- melt(subset(chs_yr22,!is.na(STDYTIME)),
                      id.vars=c("NEWID","STDYTIME","PERSTAT"),
                      na.rm=T,
                      factorsAsStrings = T,
                      variable.factor=F)
names(chs_melt_yr22)[1:3] <- c("patientid","visitdays","cohort")
chs_melt_yr22$visit <- 22
chs_melt_yr22$form <- "yr22final"


#----------------------------------------------------------------------------------------------------------------------#

#### CHS year 23 ####

chs_yr23_phone <- fread("~/Dropbox/BioLINCC files/CHS/Main_Study/YEAR23/yr23ph.csv",na.strings=c("NA","","NULL"))
names(chs_yr23_phone) <- toupper(names(chs_yr23_phone))

chs_melt_phone_yr23 <- melt(subset(chs_yr23_phone,!is.na(STDYTIME)),
                            id.vars=c("NEWID","STDYTIME","PERSTAT"),
                            na.rm=T,
                            factorsAsStrings = T,
                            variable.factor=F)
names(chs_melt_phone_yr23)[1:3] <- c("patientid","visitdays","cohort")
chs_melt_phone_yr23$visit <- 23
chs_melt_phone_yr23$form <- "yr23_phone"

chs_yr23 <- fread("~/Dropbox/BioLINCC files/CHS/Main_Study/YEAR23/yr23ann.csv",na.strings=c("NA","","NULL"))
names(chs_yr23) <- toupper(names(chs_yr23))

chs_yr23$agey <- floor(chs_baseboth$agey+(chs_yr23$STDYTIME/365))


chs_melt_yr23 <- melt(subset(chs_yr23,!is.na(STDYTIME)),
                      id.vars=c("NEWID","STDYTIME","PERSTAT"),
                      na.rm=T,
                      factorsAsStrings = T,
                      variable.factor=F)
names(chs_melt_yr23)[1:3] <- c("patientid","visitdays","cohort")

chs_yr23 <- merge(chs_yr23, chs_yr22[,c("NEWID",
                                        "dm_frail",
                                        "htn_frail",
                                        "copd_frail",
                                        "asthma_frail",
                                        "djd_frail",
                                        "ckd_frail",
                                        "chf_frail",
                                        "chd_frail",
                                        "stroke_frail",
                                        "cancer_frail")])

chs_yr23[is.na(chs_yr23$STDYTIME),
         c("dm_frail",
           "htn_frail",
           "copd_frail",
           "asthma_frail",
           "djd_frail",
           "ckd_frail",
           "chf_frail",
           "chd_frail",
           "stroke_frail",
           "cancer_frail")] <- NA


chs_melt_yr23$visit <- 23
chs_melt_yr23$form <- "yr23final"


#----------------------------------------------------------------------------------------------------------------------#

#### CHS year 24 ####

chs_yr24 <- fread("~/Dropbox/BioLINCC files/CHS/Main_Study/YEAR24/yr24ann.csv",na.strings=c("NA","","NULL"))
names(chs_yr24) <- toupper(names(chs_yr24))

chs_yr24$agey <- floor(chs_baseboth$agey+(chs_yr24$STDYTIME/365))


chs_melt_yr24 <- melt(subset(chs_yr24,!is.na(STDYTIME)),
                      id.vars=c("NEWID","STDYTIME","PERSTAT"),
                      na.rm=T,
                      factorsAsStrings = T,
                      variable.factor=F)

names(chs_melt_yr24)[1:3] <- c("patientid","visitdays","cohort")
chs_melt_yr24$visit <- 24

chs_yr24 <- merge(chs_yr24, chs_yr23[,c("NEWID",
                                        "dm_frail",
                                        "htn_frail",
                                        "copd_frail",
                                        "asthma_frail",
                                        "djd_frail",
                                        "ckd_frail",
                                        "chf_frail",
                                        "chd_frail",
                                        "stroke_frail",
                                        "cancer_frail")])

chs_yr24[is.na(chs_yr24$STDYTIME),
         c("dm_frail",
           "htn_frail",
           "copd_frail",
           "asthma_frail",
           "djd_frail",
           "ckd_frail",
           "chf_frail",
           "chd_frail",
           "stroke_frail",
           "cancer_frail")] <- NA


chs_melt_yr24$form <- "yr24final"

chs_yr24_phone <- fread("~/Dropbox/BioLINCC files/CHS/Main_Study/YEAR24/yr24ph.csv",na.strings=c("NA","","NULL"))
names(chs_yr24_phone) <- toupper(names(chs_yr24_phone))
chs_melt_phone_yr24 <- melt(subset(chs_yr24_phone,!is.na(STDYTIME)),
                            id.vars=c("NEWID","STDYTIME","PERSTAT"),
                            na.rm=T,
                            factorsAsStrings = T,
                            variable.factor=F)

names(chs_melt_phone_yr24)[1:3] <- c("patientid","visitdays","cohort")
chs_melt_phone_yr24$visit <- 24
chs_melt_phone_yr24$form <- "yr24_phone"


#----------------------------------------------------------------------------------------------------------------------#


#### CHS year 25 ####

chs_yr25 <- fread("~/Dropbox/BioLINCC files/CHS/Main_Study/YEAR25/yr25ann.csv",na.strings=c("NA","","NULL"))
names(chs_yr25) <- toupper(names(chs_yr25))

chs_yr25$agey <- floor(chs_baseboth$agey+(chs_yr25$STDYTIME/365))


chs_melt_yr25 <- melt(subset(chs_yr25,!is.na(STDYTIME)),
                      id.vars=c("NEWID","STDYTIME","PERSTAT"),
                      na.rm=T,
                      factorsAsStrings = T,
                      variable.factor=F)

names(chs_melt_yr25)[1:3] <- c("patientid","visitdays","cohort")
chs_melt_yr25$visit <- 25

chs_melt_yr25$cohort <- as.character(chs_melt_yr25$cohort)

chs_yr25 <- merge(chs_yr25, chs_yr23[,c("NEWID",
                                        "dm_frail",
                                        "htn_frail",
                                        "copd_frail",
                                        "asthma_frail",
                                        "djd_frail",
                                        "ckd_frail",
                                        "chf_frail",
                                        "chd_frail",
                                        "stroke_frail",
                                        "cancer_frail")])

chs_yr25[is.na(chs_yr25$STDYTIME),
         c("dm_frail",
           "htn_frail",
           "copd_frail",
           "asthma_frail",
           "djd_frail",
           "ckd_frail",
           "chf_frail",
           "chd_frail",
           "stroke_frail",
           "cancer_frail")] <- NA

chs_melt_yr25$form <- "yr25final"

chs_yr25_phone <- fread("~/Dropbox/BioLINCC files/CHS/Main_Study/YEAR25/yr25ph.csv",na.strings=c("NA","","NULL"))
names(chs_yr25_phone) <- toupper(names(chs_yr25_phone))
chs_melt_phone_yr25 <- melt(subset(chs_yr25_phone,!is.na(STDYTIME)),
                            id.vars=c("NEWID","STDYTIME","PERSTAT"),
                            na.rm=T,
                            factorsAsStrings = T,
                            variable.factor=F)

names(chs_melt_phone_yr25)[1:3] <- c("patientid","visitdays","cohort")
chs_melt_phone_yr25$visit <- 25
chs_melt_phone_yr25$form <- "yr25_phone"


#----------------------------------------------------------------------------------------------------------------------#


#### CHS year 26 ####

chs_yr26 <- fread("~/Dropbox/BioLINCC files/CHS/Main_Study/YEAR26/yr26ann.csv",na.strings=c("NA","","NULL"))
names(chs_yr26) <- toupper(names(chs_yr26))

chs_yr26$agey <- floor(chs_baseboth$agey+(chs_yr26$STDYTIME/365))


chs_melt_yr26 <- melt(subset(chs_yr26,!is.na(STDYTIME)),
                      id.vars=c("NEWID","STDYTIME","PERSTAT"),
                      na.rm=T,
                      factorsAsStrings = T,
                      variable.factor=F)

names(chs_melt_yr26)[1:3] <- c("patientid","visitdays","cohort")
chs_melt_yr26$visit <- 26

chs_yr26 <- merge(chs_yr26, chs_yr23[,c("NEWID",
                                        "dm_frail",
                                        "htn_frail",
                                        "copd_frail",
                                        "asthma_frail",
                                        "djd_frail",
                                        "ckd_frail",
                                        "chf_frail",
                                        "chd_frail",
                                        "stroke_frail",
                                        "cancer_frail")])

chs_yr26[is.na(chs_yr26$STDYTIME),
         c("dm_frail",
           "htn_frail",
           "copd_frail",
           "asthma_frail",
           "djd_frail",
           "ckd_frail",
           "chf_frail",
           "chd_frail",
           "stroke_frail",
           "cancer_frail")] <- NA

chs_melt_yr26$form <- "yr26final"

chs_yr26_phone <- fread("~/Dropbox/BioLINCC files/CHS/Main_Study/YEAR26/yr26ph.csv",na.strings=c("NA","","NULL"))
names(chs_yr26_phone) <- toupper(names(chs_yr26_phone))
chs_melt_phone_yr26 <- melt(subset(chs_yr26_phone,!is.na(STDYTIME)),
                            id.vars=c("NEWID","STDYTIME","PERSTAT"),
                            na.rm=T,
                            factorsAsStrings = T,
                            variable.factor=F)

names(chs_melt_phone_yr26)[1:3] <- c("patientid","visitdays","cohort")
chs_melt_phone_yr26$visit <- 26
chs_melt_phone_yr26$form <- "yr26_phone"


#----------------------------------------------------------------------------------------------------------------------#


#### CHS year 27 ####

chs_yr27 <- fread("~/Dropbox/BioLINCC files/CHS/Main_Study/YEAR27/yr27ann.csv",na.strings=c("NA","","NULL"))
names(chs_yr27) <- toupper(names(chs_yr27))

chs_yr27$agey <- floor(chs_baseboth$agey+(chs_yr27$STDYTIME/365))


chs_melt_yr27 <- melt(subset(chs_yr27,!is.na(STDYTIME)),
                      id.vars=c("NEWID","STDYTIME","PERSTAT"),
                      na.rm=T,
                      factorsAsStrings = T,
                      variable.factor=F)

names(chs_melt_yr27)[1:3] <- c("patientid","visitdays","cohort")
chs_melt_yr27$visit <- 27

chs_yr27 <- merge(chs_yr27, chs_yr23[,c("NEWID",
                                        "dm_frail",
                                        "htn_frail",
                                        "copd_frail",
                                        "asthma_frail",
                                        "djd_frail",
                                        "ckd_frail",
                                        "chf_frail",
                                        "chd_frail",
                                        "stroke_frail",
                                        "cancer_frail")])

chs_yr27[is.na(chs_yr27$STDYTIME),
         c("dm_frail",
           "htn_frail",
           "copd_frail",
           "asthma_frail",
           "djd_frail",
           "ckd_frail",
           "chf_frail",
           "chd_frail",
           "stroke_frail",
           "cancer_frail")] <- NA

chs_melt_yr27$form <- "yr27final"

chs_yr27_phone <- fread("~/Dropbox/BioLINCC files/CHS/Main_Study/YEAR27/yr27ph.csv",na.strings=c("NA","","NULL"))
names(chs_yr27_phone) <- toupper(names(chs_yr27_phone))
chs_melt_phone_yr27 <- melt(subset(chs_yr27_phone,!is.na(STDYTIME)),
                            id.vars=c("NEWID","STDYTIME","PERSTAT"),
                            na.rm=T,
                            factorsAsStrings = T,
                            variable.factor=F)

names(chs_melt_phone_yr27)[1:3] <- c("patientid","visitdays","cohort")
chs_melt_phone_yr27$visit <- 27
chs_melt_phone_yr27$form <- "yr27_phone"


#----------------------------------------------------------------------------------------------------------------------#


#### CHS year 28 ####

chs_yr28 <- fread("~/Dropbox/BioLINCC files/CHS/Main_Study/YEAR28/yr28ann.csv",na.strings=c("NA","","NULL"))
names(chs_yr28) <- toupper(names(chs_yr28))

chs_yr28$agey <- floor(chs_baseboth$agey+(chs_yr28$STDYTIME/365))


chs_melt_yr28 <- melt(subset(chs_yr28,!is.na(STDYTIME)),
                      id.vars=c("NEWID","STDYTIME","PERSTAT"),
                      na.rm=T,
                      factorsAsStrings = T,
                      variable.factor=F)

names(chs_melt_yr28)[1:3] <- c("patientid","visitdays","cohort")
chs_melt_yr28$visit <- 28

chs_yr28 <- merge(chs_yr28, chs_yr23[,c("NEWID",
                                        "dm_frail",
                                        "htn_frail",
                                        "copd_frail",
                                        "asthma_frail",
                                        "djd_frail",
                                        "ckd_frail",
                                        "chf_frail",
                                        "chd_frail",
                                        "stroke_frail",
                                        "cancer_frail")])

chs_yr28[is.na(chs_yr28$STDYTIME),
         c("dm_frail",
           "htn_frail",
           "copd_frail",
           "asthma_frail",
           "djd_frail",
           "ckd_frail",
           "chf_frail",
           "chd_frail",
           "stroke_frail",
           "cancer_frail")] <- NA

chs_melt_yr28$form <- "yr28final"

chs_yr28_phone <- fread("~/Dropbox/BioLINCC files/CHS/Main_Study/YEAR28/yr28ph.csv",na.strings=c("NA","","NULL"))
names(chs_yr28_phone) <- toupper(names(chs_yr28_phone))
chs_melt_phone_yr28 <- melt(subset(chs_yr28_phone,!is.na(STDYTIME)),
                            id.vars=c("NEWID","STDYTIME","PERSTAT"),
                            na.rm=T,
                            factorsAsStrings = T,
                            variable.factor=F)

names(chs_melt_phone_yr28)[1:3] <- c("patientid","visitdays","cohort")
chs_melt_phone_yr28$visit <- 28
chs_melt_phone_yr28$form <- "yr28_phone"


#----------------------------------------------------------------------------------------------------------------------#


#### CHS year 29 ####

chs_yr29 <- fread("~/Dropbox/BioLINCC files/CHS/Main_Study/YEAR29/yr29ann.csv",na.strings=c("NA","","NULL"))
names(chs_yr29) <- toupper(names(chs_yr29))

chs_yr29$agey <- floor(chs_baseboth$agey+(chs_yr29$STDYTIME/365))


chs_melt_yr29 <- melt(subset(chs_yr29,!is.na(STDYTIME)),
                      id.vars=c("NEWID","STDYTIME","PERSTAT"),
                      na.rm=T,
                      factorsAsStrings = T,
                      variable.factor=F)

names(chs_melt_yr29)[1:3] <- c("patientid","visitdays","cohort")
chs_melt_yr29$visit <- 29

chs_yr29 <- merge(chs_yr29, chs_yr23[,c("NEWID",
                                        "dm_frail",
                                        "htn_frail",
                                        "copd_frail",
                                        "asthma_frail",
                                        "djd_frail",
                                        "ckd_frail",
                                        "chf_frail",
                                        "chd_frail",
                                        "stroke_frail",
                                        "cancer_frail")])

chs_yr29[is.na(chs_yr29$STDYTIME),
         c("dm_frail",
           "htn_frail",
           "copd_frail",
           "asthma_frail",
           "djd_frail",
           "ckd_frail",
           "chf_frail",
           "chd_frail",
           "stroke_frail",
           "cancer_frail")] <- NA

chs_melt_yr29$form <- "yr29final"

chs_yr29_phone <- fread("~/Dropbox/BioLINCC files/CHS/Main_Study/YEAR29/yr29ph.csv",na.strings=c("NA","","NULL"))
names(chs_yr29_phone) <- toupper(names(chs_yr29_phone))
chs_melt_phone_yr29 <- melt(subset(chs_yr29_phone,!is.na(STDYTIME)),
                            id.vars=c("NEWID","STDYTIME","PERSTAT"),
                            na.rm=T,
                            factorsAsStrings = T,
                            variable.factor=F)

names(chs_melt_phone_yr29)[1:3] <- c("patientid","visitdays","cohort")
chs_melt_phone_yr29$visit <- 29
chs_melt_phone_yr29$form <- "yr29_phone"


#----------------------------------------------------------------------------------------------------------------------#


#### CHS year 30 ####

chs_yr30 <- fread("~/Dropbox/BioLINCC files/CHS/Main_Study/YEAR30/yr30ann.csv",na.strings=c("NA","","NULL"))
names(chs_yr30) <- toupper(names(chs_yr30))

chs_yr30$agey <- floor(chs_baseboth$agey+(chs_yr30$STDYTIME/365))


chs_melt_yr30 <- melt(subset(chs_yr30,!is.na(STDYTIME)),
                      id.vars=c("NEWID","STDYTIME","PERSTAT"),
                      na.rm=T,
                      factorsAsStrings = T,
                      variable.factor=F)

names(chs_melt_yr30)[1:3] <- c("patientid","visitdays","cohort")
chs_melt_yr30$visit <- 30

chs_yr30 <- merge(chs_yr30, chs_yr23[,c("NEWID",
                                        "dm_frail",
                                        "htn_frail",
                                        "copd_frail",
                                        "asthma_frail",
                                        "djd_frail",
                                        "ckd_frail",
                                        "chf_frail",
                                        "chd_frail",
                                        "stroke_frail",
                                        "cancer_frail")])

chs_yr30[is.na(chs_yr30$STDYTIME),
         c("dm_frail",
           "htn_frail",
           "copd_frail",
           "asthma_frail",
           "djd_frail",
           "ckd_frail",
           "chf_frail",
           "chd_frail",
           "stroke_frail",
           "cancer_frail")] <- NA

chs_melt_yr30$form <- "yr30final"

chs_yr30_phone <- fread("~/Dropbox/BioLINCC files/CHS/Main_Study/YEAR30/yr30ph.csv",na.strings=c("NA","","NULL"))
names(chs_yr30_phone) <- toupper(names(chs_yr30_phone))
chs_melt_phone_yr30 <- melt(subset(chs_yr30_phone,!is.na(STDYTIME)),
                            id.vars=c("NEWID","STDYTIME","PERSTAT"),
                            na.rm=T,
                            factorsAsStrings = T,
                            variable.factor=F)

names(chs_melt_phone_yr30)[1:3] <- c("patientid","visitdays","cohort")
chs_melt_phone_yr30$visit <- 30
chs_melt_phone_yr30$form <- "yr30_phone"


#----------------------------------------------------------------------------------------------------------------------#


#### CHS year 31 ####

chs_yr31 <- fread("~/Dropbox/BioLINCC files/CHS/Main_Study/YEAR31/yr31ann.csv",na.strings=c("NA","","NULL"))
names(chs_yr31) <- toupper(names(chs_yr31))

chs_yr31$agey <- floor(chs_baseboth$agey+(chs_yr31$STDYTIME/365))


chs_melt_yr31 <- melt(subset(chs_yr31,!is.na(STDYTIME)),
                      id.vars=c("NEWID","STDYTIME","PERSTAT"),
                      na.rm=T,
                      factorsAsStrings = T,
                      variable.factor=F)

names(chs_melt_yr31)[1:3] <- c("patientid","visitdays","cohort")
chs_melt_yr31$visit <- 31

chs_yr31 <- merge(chs_yr31, chs_yr23[,c("NEWID",
                                        "dm_frail",
                                        "htn_frail",
                                        "copd_frail",
                                        "asthma_frail",
                                        "djd_frail",
                                        "ckd_frail",
                                        "chf_frail",
                                        "chd_frail",
                                        "stroke_frail",
                                        "cancer_frail")])

chs_yr31[is.na(chs_yr31$STDYTIME),
         c("dm_frail",
           "htn_frail",
           "copd_frail",
           "asthma_frail",
           "djd_frail",
           "ckd_frail",
           "chf_frail",
           "chd_frail",
           "stroke_frail",
           "cancer_frail")] <- NA

chs_melt_yr31$form <- "yr31final"

chs_yr31_phone <- fread("~/Dropbox/BioLINCC files/CHS/Main_Study/YEAR31/yr31ph.csv",na.strings=c("NA","","NULL"))
names(chs_yr31_phone) <- toupper(names(chs_yr31_phone))
chs_melt_phone_yr31 <- melt(subset(chs_yr31_phone,!is.na(STDYTIME)),
                            id.vars=c("NEWID","STDYTIME","PERSTAT"),
                            na.rm=T,
                            factorsAsStrings = T,
                            variable.factor=F)

names(chs_melt_phone_yr31)[1:3] <- c("patientid","visitdays","cohort")
chs_melt_phone_yr31$visit <- 31
chs_melt_phone_yr31$form <- "yr31_phone"


#----------------------------------------------------------------------------------------------------------------------#


#### CHS year 32 ####

chs_yr32 <- fread("~/Dropbox/BioLINCC files/CHS/Main_Study/YEAR32/yr32ann.csv",na.strings=c("NA","","NULL"))
names(chs_yr32) <- toupper(names(chs_yr32))

chs_yr32$agey <- floor(chs_baseboth$agey+(chs_yr32$STDYTIME/365))


chs_melt_yr32 <- melt(subset(chs_yr32,!is.na(STDYTIME)),
                      id.vars=c("NEWID","STDYTIME","PERSTAT"),
                      na.rm=T,
                      factorsAsStrings = T,
                      variable.factor=F)

names(chs_melt_yr32)[1:3] <- c("patientid","visitdays","cohort")
chs_melt_yr32$visit <- 32

chs_yr32 <- merge(chs_yr32, chs_yr23[,c("NEWID",
                                        "dm_frail",
                                        "htn_frail",
                                        "copd_frail",
                                        "asthma_frail",
                                        "djd_frail",
                                        "ckd_frail",
                                        "chf_frail",
                                        "chd_frail",
                                        "stroke_frail",
                                        "cancer_frail")])

chs_yr32[is.na(chs_yr32$STDYTIME),
         c("dm_frail",
           "htn_frail",
           "copd_frail",
           "asthma_frail",
           "djd_frail",
           "ckd_frail",
           "chf_frail",
           "chd_frail",
           "stroke_frail",
           "cancer_frail")] <- NA

chs_melt_yr32$form <- "yr32final"

chs_yr32_phone <- fread("~/Dropbox/BioLINCC files/CHS/Main_Study/YEAR32/yr32ph.csv",na.strings=c("NA","","NULL"))
names(chs_yr32_phone) <- toupper(names(chs_yr32_phone))
chs_melt_phone_yr32 <- melt(subset(chs_yr32_phone,!is.na(STDYTIME)),
                            id.vars=c("NEWID","STDYTIME","PERSTAT"),
                            na.rm=T,
                            factorsAsStrings = T,
                            variable.factor=F)

names(chs_melt_phone_yr32)[1:3] <- c("patientid","visitdays","cohort")
chs_melt_phone_yr32$visit <- 32
chs_melt_phone_yr32$form <- "yr32_phone"


#----------------------------------------------------------------------------------------------------------------------#


#### CHS year 33 ####

chs_yr33 <- fread("~/Dropbox/BioLINCC files/CHS/Main_Study/YEAR33/yr33ann.csv",na.strings=c("NA","","NULL"))
names(chs_yr33) <- toupper(names(chs_yr33))

chs_yr33$agey <- floor(chs_baseboth$agey+(chs_yr33$STDYTIME/365))


chs_melt_yr33 <- melt(subset(chs_yr33,!is.na(STDYTIME)),
                      id.vars=c("NEWID","STDYTIME","PERSTAT"),
                      na.rm=T,
                      factorsAsStrings = T,
                      variable.factor=F)

names(chs_melt_yr33)[1:3] <- c("patientid","visitdays","cohort")
chs_melt_yr33$visit <- 33

chs_yr33 <- merge(chs_yr33, chs_yr23[,c("NEWID",
                                        "dm_frail",
                                        "htn_frail",
                                        "copd_frail",
                                        "asthma_frail",
                                        "djd_frail",
                                        "ckd_frail",
                                        "chf_frail",
                                        "chd_frail",
                                        "stroke_frail",
                                        "cancer_frail")])

chs_yr33[is.na(chs_yr33$STDYTIME),
         c("dm_frail",
           "htn_frail",
           "copd_frail",
           "asthma_frail",
           "djd_frail",
           "ckd_frail",
           "chf_frail",
           "chd_frail",
           "stroke_frail",
           "cancer_frail")] <- NA

chs_melt_yr33$form <- "yr33final"

chs_yr33_phone <- fread("~/Dropbox/BioLINCC files/CHS/Main_Study/YEAR33/yr33ph.csv",na.strings=c("NA","","NULL"))
names(chs_yr33_phone) <- toupper(names(chs_yr33_phone))
chs_melt_phone_yr33 <- melt(subset(chs_yr33_phone,!is.na(STDYTIME)),
                            id.vars=c("NEWID","STDYTIME","PERSTAT"),
                            na.rm=T,
                            factorsAsStrings = T,
                            variable.factor=F)

names(chs_melt_phone_yr33)[1:3] <- c("patientid","visitdays","cohort")
chs_melt_phone_yr33$visit <- 33
chs_melt_phone_yr33$form <- "yr33_phone"


#----------------------------------------------------------------------------------------------------------------------#


#### CHS year 34 ####

chs_yr34 <- fread("~/Dropbox/BioLINCC files/CHS/Main_Study/YEAR34/yr34ann.csv",na.strings=c("NA","","NULL"))
names(chs_yr34) <- toupper(names(chs_yr34))

chs_yr34$agey <- floor(chs_baseboth$agey+(chs_yr34$STDYTIME/365))


chs_melt_yr34 <- melt(subset(chs_yr34,!is.na(STDYTIME)),
                      id.vars=c("NEWID","STDYTIME","PERSTAT"),
                      na.rm=T,
                      factorsAsStrings = T,
                      variable.factor=F)

names(chs_melt_yr34)[1:3] <- c("patientid","visitdays","cohort")
chs_melt_yr34$visit <- 34

chs_yr34 <- merge(chs_yr34, chs_yr23[,c("NEWID",
                                        "dm_frail",
                                        "htn_frail",
                                        "copd_frail",
                                        "asthma_frail",
                                        "djd_frail",
                                        "ckd_frail",
                                        "chf_frail",
                                        "chd_frail",
                                        "stroke_frail",
                                        "cancer_frail")])

chs_yr34[is.na(chs_yr34$STDYTIME),
         c("dm_frail",
           "htn_frail",
           "copd_frail",
           "asthma_frail",
           "djd_frail",
           "ckd_frail",
           "chf_frail",
           "chd_frail",
           "stroke_frail",
           "cancer_frail")] <- NA

chs_melt_yr34$form <- "yr34final"

chs_yr34_phone <- fread("~/Dropbox/BioLINCC files/CHS/Main_Study/YEAR34/yr34ph.csv",na.strings=c("NA","","NULL"))
names(chs_yr34_phone) <- toupper(names(chs_yr34_phone))
chs_melt_phone_yr34 <- melt(subset(chs_yr34_phone,!is.na(STDYTIME)),
                            id.vars=c("NEWID","STDYTIME","PERSTAT"),
                            na.rm=T,
                            factorsAsStrings = T,
                            variable.factor=F)

names(chs_melt_phone_yr34)[1:3] <- c("patientid","visitdays","cohort")
chs_melt_phone_yr34$visit <- 34
chs_melt_phone_yr34$form <- "yr34_phone"


#----------------------------------------------------------------------------------------------------------------------#


#### CHS year 35 ####

chs_yr35 <- fread("~/Dropbox/BioLINCC files/CHS/Main_Study/YEAR35/yr35ann.csv",na.strings=c("NA","","NULL"))
names(chs_yr35) <- toupper(names(chs_yr35))

chs_yr35$agey <- floor(chs_baseboth$agey+(chs_yr35$STDYTIME/365))


chs_melt_yr35 <- melt(subset(chs_yr35,!is.na(STDYTIME)),
                      id.vars=c("NEWID","STDYTIME","PERSTAT"),
                      na.rm=T,
                      factorsAsStrings = T,
                      variable.factor=F)

names(chs_melt_yr35)[1:3] <- c("patientid","visitdays","cohort")
chs_melt_yr35$visit <- 35

chs_yr35 <- merge(chs_yr35, chs_yr23[,c("NEWID",
                                        "dm_frail",
                                        "htn_frail",
                                        "copd_frail",
                                        "asthma_frail",
                                        "djd_frail",
                                        "ckd_frail",
                                        "chf_frail",
                                        "chd_frail",
                                        "stroke_frail",
                                        "cancer_frail")])

chs_yr35[is.na(chs_yr35$STDYTIME),
         c("dm_frail",
           "htn_frail",
           "copd_frail",
           "asthma_frail",
           "djd_frail",
           "ckd_frail",
           "chf_frail",
           "chd_frail",
           "stroke_frail",
           "cancer_frail")] <- NA

chs_melt_yr35$form <- "yr35final"


# ==============================================================#
####        CHS ancillary estrogens, yr 7                    ####
# ==============================================================#

chs_as447 <- fread("~/Dropbox/BioLINCC files/CHS/Ancillary_Studies/AS_447_Njoroge_Estrogens/as447.csv",na.strings=c("NA","","NULL"))
names(chs_as447) <- toupper(names(chs_as447))

chs_melt_as447 <- melt(chs_as447,
                       id.vars=c("NEWID"),
                       na.rm=T,
                       factorsAsStrings = T,
                       variable.factor=F)
names(chs_melt_as447)[1] <- c("patientid")
chs_melt_as447$visit <- 7
chs_melt_as447$form <- "estrogens"
chs_melt_as447$visitdays <- NA


# ==============================================================#
####        CHS ancillary tau protein, yr 5                  ####
# ==============================================================#

chs_as415 <- fread("~/Dropbox/BioLINCC files/CHS/Ancillary_Studies/AS_415_Fohner_pTau181/as415.csv",na.strings=c("NA","","NULL"))
names(chs_as415) <- toupper(names(chs_as415))

chs_melt_as415 <- melt(chs_as415,
                       id.vars=c("NEWID"),
                       na.rm=T,
                       factorsAsStrings = T,
                       variable.factor=F)

names(chs_melt_as415)[1] <- c("patientid")
chs_melt_as415$visit <- 5
chs_melt_as415$form <- "ptau181"
chs_melt_as415$visitdays <- NA


# ==============================================================#
####        CHS ancillary proteomics, yr 5                  ####
# ==============================================================#

library(haven)

chs_as354 <- read_sas("~/Dropbox/BioLINCC files/CHS/Ancillary_Studies/AS_354_Psaty_Proteomics/chs_values_5k_7k_scaled.sas7bdat")

names(chs_as354) <- toupper(names(chs_as354))

chs_as354 <- as.data.table(chs_as354)

chs_melt_as354 <- melt(chs_as354,
                       id.vars=c("NEWID"),
                       na.rm=T,
                       factorsAsStrings = T,
                       variable.factor=F)

names(chs_melt_as354)[1] <- c("patientid")
chs_melt_as354$visit <- 5
chs_melt_as354$form <- "proteomics"
chs_melt_as354$visitdays <- NA


# ==============================================================#
####        CHS ancillary HRV, yr 2, 7                       ####
# ==============================================================#

chs_as68y2 <- 
  fread("~/Dropbox/BioLINCC files/CHS/Ancillary_Studies/AS_068_Stein_HRV/as68y2.csv",
                    na.strings=c("NA","","NULL"))
names(chs_as68y2) <- toupper(names(chs_as68y2))

chs_as68y2 <- subset(chs_as68y2,EXCLUDE == 0)

chs_as68y2$cohort <- 1

chs_melt_as68y2 <- melt(chs_as68y2,
                        id.vars=c("NEWID","cohort"),
                        na.rm=T,
                        factorsAsStrings = T,
                        variable.factor=F)

names(chs_melt_as68y2)[1] <- c("patientid")
chs_melt_as68y2$form <- "hrv"
chs_melt_as68y2$visit <- 1
chs_melt_as68y2$visitdays <- 0

# Year 7

chs_as68y7 <- 
  fread("~/Dropbox/BioLINCC files/CHS/Ancillary_Studies/AS_068_Stein_HRV/as68y7.csv",
                    na.strings=c("NA","","NULL"))
names(chs_as68y7) <- toupper(names(chs_as68y7))

#### Remove records that indicated atrial fibrillation, atrial pacing, or 
#### other irregularity that that limit characterization 

chs_as68y7 <- subset(chs_as68y7,EXCLUDE7 == 0)

chs_as68y7 <-
  merge(chs_as68y7,
        chs_yr7[,c("patientid","PERSTAT","STDYTIME")],
        by.x="NEWID",
        by.y="patientid")

names(chs_as68y7)[c(128:129)] <- c("cohort","visitdays")

chs_melt_as68y7 <- melt(chs_as68y7,
                        id.vars=c("NEWID","cohort","visitdays"),
                        na.rm=T,
                        factorsAsStrings = T,
                        variable.factor=F)

names(chs_melt_as68y7)[1] <- c("patientid")
chs_melt_as68y7$visit <- 7
chs_melt_as68y7$form <- "hrv"

chs_melt_hrv <-
  rbind(chs_melt_as68y2,
        chs_melt_as68y7)

# ==============================================================#
####        CHS ancillary testosterone                      ####
# ==============================================================#

chs_as200 <- fread("~/Dropbox/BioLINCC files/CHS/Ancillary_Studies/AS_200_Shores_Testosterone/as200.csv",
                   na.strings=c("NA","","NULL"))
names(chs_as200) <- toupper(names(chs_as200))

chs_melt_as200 <- melt(chs_as200,
                       id.vars=c("NEWID","YEAR"),
                       na.rm=T,
                       factorsAsStrings = T,
                       variable.factor=F)

names(chs_melt_as200)[1] <- c("patientid")
chs_melt_as200$form <- "testosterone"
chs_melt_as200$visitdays <- NA
chs_melt_as200$visit <- chs_melt_as200$YEAR


# ==============================================================#
####        CHS ancillary aging proteins, yr 7                ####
# ==============================================================#

chs_as348 <- fread("~/Dropbox/BioLINCC files/CHS/Ancillary_Studies/AS_348_Cummings_CARGO/as348.csv",
                   na.strings=c("NA","","NULL"))
names(chs_as348) <- toupper(names(chs_as348))

chs_as348 <-
  merge(chs_as348,
        chs_yr7[,c("patientid","STDYTIME","PERSTAT")],
        by.x="NEWID",
        by.y="patientid",
        all.x=T)

names(chs_as348)[9:10] <- c("visitdays","cohort")

chs_melt_as348 <- melt(chs_as348,
                       id.vars=c("NEWID","visitdays","cohort"),
                       na.rm=T,
                       factorsAsStrings = T,
                       variable.factor=F)
names(chs_melt_as348)[1] <- c("patientid")
chs_melt_as348$visit <- 7
chs_melt_as348$form <- "aging_proteins"

# ==============================================================#
####        CHS ancillary diastolic function SNPS            ####
# ==============================================================#

chs_ddsnp <- fread("~/Dropbox/BioLINCC files/CHS/Ancillary_Studies/AS_161_Daniel_SNPs/danielsnp.csv",
                   na.strings=c("NA","","NULL"))
names(chs_ddsnp) <- toupper(names(chs_ddsnp))

chs_melt_ddsnp <- melt(chs_ddsnp,
                       id.vars=c("NEWID"),
                       na.rm=T,
                       factorsAsStrings = T,
                       variable.factor=F)
names(chs_melt_ddsnp)[1] <- c("patientid")
chs_melt_ddsnp$visit <- 1
chs_melt_ddsnp$form <- "dd_snps"
chs_melt_ddsnp$visitdays <- 0

# ==============================================================#
####        CHS ancillary cardiac mechanics, yr 2,7          ####
# ==============================================================#

chs_as320 <- fread("~/Dropbox/BioLINCC files/CHS/Ancillary_Studies/AS_320_Shah_Cardiac_Mechanics/as320.csv",
                   na.strings=c("NA","","NULL"))
names(chs_as320) <- toupper(names(chs_as320))

chs_as320[,LVGLS := -1*LVGLS]
chs_as320[,LVGLS_Y7 := -1*LVGLS_Y7]


mech_fields <-
  c("NEWID",
    "SITE",
    "READER",
    "A4CQ",
    "LVQ",
    "LAQ",
    "RVQ",
    "RV_CUTOFF",
    "HR",
    "LVEDV",
    "LVESV",
    "LVSV",
    "LVEF",
    "LVMASS",
    "LVLS_AVG",
    "LVGLS",
    "LV_EDSR",
    "ESEPTAL",
    "ELATERAL",
    "LAEDV",
    "LAESV",
    "LASV",
    "LAEF",
    "LAMASS",
    "LALS_AVG",
    "RVEDV",
    "RVESV",
    "RVSV",
    "RVEF",
    "RVMASS",
    "RVLS_AVG",
    "RVGLS",
    "RVFW")

mech_fields_y7 <-
  paste(mech_fields,
        "_Y7",
        sep="")

chs_as320_y2 <-
  chs_as320[,..mech_fields]

chs_as320_y2 <-
  merge(chs_as320_y2,
        chs_base2[,c("NEWID",
                     "DPMEP43")],
        by="NEWID",
        all.x=T)

chs_as320_y2$eeprime_sep <-   ## Add to criteriaset
  chs_as320_y2$DPMEP43*100/chs_as320_y2$ESEPTAL

chs_as320_y2$eeprime_lat <-   ## Add to criteriaset
  chs_as320_y2$DPMEP43*100/chs_as320_y2$ELATERAL

mech_fields_y7[1] <- "NEWID"
chs_as320_y7 <-
  chs_as320[,..mech_fields_y7]

chs_as320_y7$eeprime_sep <-   ## Add to criteriaset
  chs_as320_y2$DPMEP43*100/chs_as320_y2$ESEPTAL

chs_as320_y7$eeprime_lat <-   ## Add to criteriaset
  chs_as320_y7$DPMEP43*100/chs_as320_y7$ELATERAL

chs_as320_y7[, eeprime_avg := rowMeans(.SD, na.rm = TRUE),
         .SDcols = c("eeprime_lat", "eeprime_sep")]

chs_melt_as320_y2 <- melt(chs_as320_y2,
                          id.vars=c("NEWID"),
                          na.rm=T,
                          factorsAsStrings = T,
                          variable.factor=F)

names(chs_melt_as320_y2)[1] <- c("patientid")
chs_melt_as320_y2$visit <- 2
chs_melt_as320_y2$form <- "cardiac_mechanics"
chs_melt_as320_y2$visitdays <- NA

chs_melt_as320_y7 <- melt(chs_as320_y7,
                          id.vars=c("NEWID"),
                          na.rm=T,
                          factorsAsStrings = T,
                          variable.factor=F)
names(chs_melt_as320_y7)[1] <- c("patientid")
chs_melt_as320_y7$visit <- 2
chs_melt_as320_y7$form <- "cardiac_mechanics"
chs_melt_as320_y7$visitdays <- NA


# ==============================================================#
####        CHS ancillary telomeres, yr 5                  ####
# ==============================================================#

chs_as156 <- fread("~/Dropbox/BioLINCC files/CHS/Ancillary_Studies/AS_156_Fitpatrick_Telomere/as156.csv",
                   na.strings=c("NA","","NULL"))
names(chs_as156) <- toupper(names(chs_as156))

chs_melt_as156 <- melt(chs_as156,
                       id.vars=c("NEWID"),
                       na.rm=T,
                       factorsAsStrings = T,
                       variable.factor=F)

chs_melt_as156$visit[chs_melt_as156$variable %in% c("trf1","TRFmean_y5")] <- 5
chs_melt_as156$visit[chs_melt_as156$variable %in% c("trf2","TRFmean_y10")] <- 10

chs_melt_as156$variable[chs_melt_as156$variable %in% c("trf1","trf2")]  <- "trf"
chs_melt_as156$variable[chs_melt_as156$variable %in% c("TRFmean_y5","TRFmean_y10")]  <- "TRFmean"

chs_melt_as156 <-
  subset(chs_melt_as156,!is.na(value)&value > 0)

names(chs_melt_as156)[1] <- c("patientid")
chs_melt_as156$visit <- 5
chs_melt_as156$form <- "telomeres"
chs_melt_as156$visitdays <- NA


#----------------------------------------------------------------------------------------------------------------------#

#### *** CHS final assembly *** ####

chs_melt_all <- rbind(
  chs_melt_base1[,c("patientid","variable","value","visitdays","visit","form")],
  chs_melt_base2[,c("patientid","variable","value","visitdays","visit","form")],
  chs_melt_baseboth[,c("patientid","variable","value","visitdays","visit","form")],
  chs_melt_yr3[,c("patientid","variable","value","visitdays","visit","form")],
  chs_melt_yr4[,c("patientid","variable","value","visitdays","visit","form")],
  chs_melt_yr5new[,c("patientid","variable","value","visitdays","visit","form")],
  chs_melt_yr5old[,c("patientid","variable","value","visitdays","visit","form")],
  chs_melt_yr6[,c("patientid","variable","value","visitdays","visit","form")],
  chs_melt_yr7[,c("patientid","variable","value","visitdays","visit","form")],
  chs_melt_yr8[,c("patientid","variable","value","visitdays","visit","form")],
  chs_melt_yr9[,c("patientid","variable","value","visitdays","visit","form")],
  chs_melt_yr10[,c("patientid","variable","value","visitdays","visit","form")],
  chs_melt_yr11[,c("patientid","variable","value","visitdays","visit","form")],
  chs_melt_yr12[,c("patientid","variable","value","visitdays","visit","form")],
  chs_melt_phone_yr12[,c("patientid","variable","value","visitdays","visit","form")],
  chs_melt_yr13[,c("patientid","variable","value","visitdays","visit","form")],
  chs_melt_phone_yr13[,c("patientid","variable","value","visitdays","visit","form")],
  chs_melt_yr14[,c("patientid","variable","value","visitdays","visit","form")],
  chs_melt_phone_yr14[,c("patientid","variable","value","visitdays","visit","form")],
  chs_melt_yr15[,c("patientid","variable","value","visitdays","visit","form")],
  chs_melt_phone_yr15[,c("patientid","variable","value","visitdays","visit","form")],
  chs_melt_yr16[,c("patientid","variable","value","visitdays","visit","form")],
  chs_melt_phone_yr16[,c("patientid","variable","value","visitdays","visit","form")],
  chs_melt_yr17[,c("patientid","variable","value","visitdays","visit","form")],
  chs_melt_phone_yr17[,c("patientid","variable","value","visitdays","visit","form")],
  chs_melt_yr18[,c("patientid","variable","value","visitdays","visit","form")],
  chs_melt_phone_yr18[,c("patientid","variable","value","visitdays","visit","form")],
  chs_melt_yr19[,c("patientid","variable","value","visitdays","visit","form")],
  chs_melt_phone_yr19[,c("patientid","variable","value","visitdays","visit","form")],
  chs_melt_yr20[,c("patientid","variable","value","visitdays","visit","form")],
  chs_melt_phone_yr20[,c("patientid","variable","value","visitdays","visit","form")],
  chs_melt_yr21[,c("patientid","variable","value","visitdays","visit","form")],
  chs_melt_phone_yr21[,c("patientid","variable","value","visitdays","visit","form")],
  chs_melt_yr22[,c("patientid","variable","value","visitdays","visit","form")],
  chs_melt_phone_yr22[,c("patientid","variable","value","visitdays","visit","form")],
  chs_melt_yr23[,c("patientid","variable","value","visitdays","visit","form")],
  chs_melt_phone_yr23[,c("patientid","variable","value","visitdays","visit","form")],
  chs_melt_yr24[,c("patientid","variable","value","visitdays","visit","form")],
  chs_melt_phone_yr24[,c("patientid","variable","value","visitdays","visit","form")],
  chs_melt_yr25[,c("patientid","variable","value","visitdays","visit","form")],
  chs_melt_phone_yr25[,c("patientid","variable","value","visitdays","visit","form")],
  chs_melt_yr26[,c("patientid","variable","value","visitdays","visit","form")],
  chs_melt_phone_yr26[,c("patientid","variable","value","visitdays","visit","form")],
  chs_melt_yr27[,c("patientid","variable","value","visitdays","visit","form")],
  chs_melt_phone_yr27[,c("patientid","variable","value","visitdays","visit","form")],
  chs_melt_yr28[,c("patientid","variable","value","visitdays","visit","form")],
  chs_melt_phone_yr28[,c("patientid","variable","value","visitdays","visit","form")],
  chs_melt_yr29[,c("patientid","variable","value","visitdays","visit","form")],
  chs_melt_phone_yr29[,c("patientid","variable","value","visitdays","visit","form")],
  chs_melt_yr30[,c("patientid","variable","value","visitdays","visit","form")],
  chs_melt_phone_yr30[,c("patientid","variable","value","visitdays","visit","form")],
  chs_melt_yr31[,c("patientid","variable","value","visitdays","visit","form")],
  chs_melt_phone_yr31[,c("patientid","variable","value","visitdays","visit","form")],
  chs_melt_yr32[,c("patientid","variable","value","visitdays","visit","form")],
  chs_melt_phone_yr32[,c("patientid","variable","value","visitdays","visit","form")],
  chs_melt_yr33[,c("patientid","variable","value","visitdays","visit","form")],
  chs_melt_phone_yr33[,c("patientid","variable","value","visitdays","visit","form")],
  chs_melt_yr34[,c("patientid","variable","value","visitdays","visit","form")],
  chs_melt_phone_yr34[,c("patientid","variable","value","visitdays","visit","form")],
  chs_melt_yr35[,c("patientid","variable","value","visitdays","visit","form")],
  chs_melt_as156[,c("patientid","variable","value","visitdays","visit","form")],
  chs_melt_as200[,c("patientid","variable","value","visitdays","visit","form")],
  chs_melt_as320_y2[,c("patientid","variable","value","visitdays","visit","form")],
  chs_melt_as320_y7[,c("patientid","variable","value","visitdays","visit","form")],
  chs_melt_as348[,c("patientid","variable","value","visitdays","visit","form")],
#  chs_melt_as354[,c("patientid","variable","value","visitdays","visit","form")],
  chs_melt_as415[,c("patientid","variable","value","visitdays","visit","form")],
  chs_melt_as447[,c("patientid","variable","value","visitdays","visit","form")],
  chs_melt_as68y2[,c("patientid","variable","value","visitdays","visit","form")],
  chs_melt_as68y7[,c("patientid","variable","value","visitdays","visit","form")]
)

chs_melt_all$variable <- as.character(chs_melt_all$variable)
chs_melt_all$variable <- toupper(chs_melt_all$variable)
chs_melt_all$form <- toupper(chs_melt_all$form)
chs_melt_all$study <- "CHS"

chs_melt_all$study_field <- paste(chs_melt_all$study,
                                  chs_melt_all$form,
                                  chs_melt_all$variable,
                                  sep="_")


chs_melt_all <- merge(chs_melt_all,
                      chs_baseboth[,c("NEWID","agey","PERSTAT")],
                      by.x=c("patientid"),
                      by.y=c("NEWID"))


names(chs_melt_all)[ncol(chs_melt_all)] <- "cohort"

chs_melt_all$age_obs <- round(chs_melt_all$agey+chs_melt_all$visitdays/365,0)

k <- 
  c("visit","cohort","cohort_name","visit_yr")

chs_melt_all$cohort <- as.character(chs_melt_all$cohort)

chs_melt_all <- merge(chs_melt_all,
                      visit_yrs[study=="CHS",..k],
                      by=c("visit","cohort"),all.x=T)

chs_data_fields <- unique(chs_melt_all$study_field)

chs_melt_all$datapoint <-
  paste("CHS",row.names(chs_melt_all),sep="")

write_parquet(chs_melt_all[,..data_fields],
              "~/Dropbox/ADAPT-HF/Master HDCP files/Cohort data/chs_melt_all.parquet")

f <- function(input,output) {
  write.csv(input,file=output, row.names=F, na="")
}

gcs_auth("~/Dropbox/ADAPT-HF/Master HDCP files/harmonization-286013-39f492122f69.json")
gcs_upload(chs_melt_all[,..data_fields], 
           bucket="master_hdcp_files",
           name="chs_melt_all.parquet",
           object_function = f)

rm(list=ls(pattern="\\bchs."))

gc()
```
