## Supplementary material for "Aggregating data to accelerate personalized therapy in heart failure (ADAPT-HF)": Harmonization scripts: Compile cohort dates.R

```
# The primary difference between v5 and v6 is using datatables 
# instead of data frames to manipulate raw data.

# ==============================================================#
####           **** Load R packages ****                    ####
# ==============================================================#


library(sqldf)
library(Hmisc)
library(plotrix)
library(descr)
library(dplyr)
library(relper)
library(stringr)
library(bigrquery)
library(haven)
library(googleCloudStorageR)
library(data.table)
library(arrow)
library(parquetize)
library(bigQueryR)

source("~/Dropbox/R scripts/Misc clinical scripts.R")
source("~/Dropbox/R scripts/echo scripts.R")

f <- function(input,output) {
  write.csv(input,file=output, row.names=F, na="")
}

visit_yrs <- fread('~/Dropbox/ADAPT-HF/Master HDCP files/Cohort visit yrs.csv')

data_fields <- 
  c("study",
    "patientid",
    "visit_yr",
    "visitdays",
    "form",
    "variable",
    "value",
    "datapoint",
    "study_field",
    "cohort",
    "cohort_name",
    "age_obs")

outcomes_list <- c("dth_status","dth_dt",
                   "cvdth_status","cvdth_dt",
                   "hfdth_status","hfdth_dt",
                   "hfhosp_ef_val","hfhosp_ef_cat",
                   "noncvdth_status","noncvdth_dt",
                   "hosp_status","hosp_dt",
                   "cadhosp_status","cadhosp_dt",
                   "cvhosp_status","cvhosp_dt",
                   "hfhosp_status","hfhosp_dt",
                   "noncvhosp_status","noncvhosp_dt")

dates_long_fields <-
  c("study",
    "cohort",
    "cohort_name",
    "patientid",
    "sex",
    "visit_yr",
    "visitdays",
    "age_obs")

#=========================================================================#
####            **** Load phenotyping criteria ****                    ####
#=========================================================================#


bq_auth(path="harmonization-286013-39f492122f69.json")

my_bq <- dbConnect(
  bigrquery::bigquery(),
  project="harmonization",
  dataset="cohorts",
  billing="harmonization"
)


hdcp_cohort_criteria <- 
  fread("~/Dropbox/ADAPT-HF/Master HDCP files/Criteriaset_cohort study 5.0.csv",
        na.strings=c("NA","","NULL","\\N"),
        stringsAsFactors = F)

hdcp_cohort_criteria$rule_num <-
  rownames(hdcp_cohort_criteria)


phenotypes_cohort <- 
  fread("~/Dropbox/ADAPT-HF/Master HDCP files/phenotypelist_cohort.csv",
        na.strings=c("NA","","NULL","\\N"))

phenos <- unique(hdcp_cohort_criteria$phenotype)

miss_phenos <- phenos[!phenos %in% phenotypes_cohort$phenotype]

hdcp_cohort_criteria$string_value <- str_replace(hdcp_cohort_criteria$string_value,"\xb2 ","≤ ")
hdcp_cohort_criteria$string_value <- str_replace(hdcp_cohort_criteria$string_value,"\xb3","≥ ")
hdcp_cohort_criteria$rule_num <- rownames(hdcp_cohort_criteria)

hdcp_cohort_criteria$lower_bound[is.na(hdcp_cohort_criteria$lower_bound)&hdcp_cohort_criteria$rule_type=="range"] <- -1000000
hdcp_cohort_criteria$upper_bound[is.na(hdcp_cohort_criteria$upper_bound)&hdcp_cohort_criteria$rule_type=="range"] <- 1000000

hdcp_cohort_criteria$form <- toupper(hdcp_cohort_criteria$form)

hdcp_cohort_criteria$field <- toupper(hdcp_cohort_criteria$field)

unique(hdcp_cohort_criteria$field[hdcp_cohort_criteria$study=="JHS"])

hdcp_cohort_criteria$study_field <-
  paste(hdcp_cohort_criteria$study,
        hdcp_cohort_criteria$form,
        hdcp_cohort_criteria$field,
        sep="_")

if (bq_table_exists("harmonization-286013.pipeline.cohort_criteria"))
{bq_table_delete("harmonization-286013.pipeline.cohort_criteria")}


bq_table_upload(x="harmonization-286013.pipeline.cohort_criteria",
                hdcp_cohort_criteria,quiet=T,
                create_disposition="CREATE_IF_NEEDED",
                write_disposition="WRITE_TRUNCATE")


if (bq_table_exists("harmonization-286013.pipeline.phenotypes_cohort"))
{bq_table_delete("harmonization-286013.pipeline.phenotypes_cohort")}
bq_table_upload(x="harmonization-286013.pipeline.phenotypes_cohort",
                phenotypes_cohort,
                quiet=T,
                create_disposition="CREATE_IF_NEEDED",
                write_disposition="WRITE_TRUNCATE")


#### ************************ FRAMINGHAM ************************ #### 

# .%%%%%%%%.%%%%%%%%.....%%%....%%.....%%.%%%%.%%....%%..%%%%%%...%%.....%%....%%%....%%.....%%
# .%%.......%%.....%%...%%.%%...%%%...%%%..%%..%%%...%%.%%....%%..%%.....%%...%%.%%...%%%...%%%
# .%%.......%%.....%%..%%...%%..%%%%.%%%%..%%..%%%%..%%.%%........%%.....%%..%%...%%..%%%%.%%%%
# .%%%%%%...%%%%%%%%..%%.....%%.%%.%%%.%%..%%..%%.%%.%%.%%...%%%%.%%%%%%%%%.%%.....%%.%%.%%%.%%
# .%%.......%%...%%...%%%%%%%%%.%%.....%%..%%..%%..%%%%.%%....%%..%%.....%%.%%%%%%%%%.%%.....%%
# .%%.......%%....%%..%%.....%%.%%.....%%..%%..%%...%%%.%%....%%..%%.....%%.%%.....%%.%%.....%%
# .%%.......%%.....%%.%%.....%%.%%.....%%.%%%%.%%....%%..%%%%%%...%%.....%%.%%.....%%.%%.....%%


#^^^^^^^^^^^^^^^^^^^^^^^^^^^^^^^^^^^^^^^^^^^^^^^^^^^^^^^^^^^^^^##
####                       FHS Outcomes                      ####
#^^^^^^^^^^^^^^^^^^^^^^^^^^^^^^^^^^^^^^^^^^^^^^^^^^^^^^^^^^^^^^##


fhs1_chfinit <- fread('~/Dropbox/BioLINCC files/Framingham cohort/datasets/CSV/vr_chfinit_2013_a_0828d.csv',na.strings=c("NA","","NULL"))
fhs1_survcvd <- fread('~/Dropbox/BioLINCC files/Framingham cohort/datasets/CSV/vr_survcvd_2018_a_1267d.csv',na.strings=c("NA","","NULL"))
fhs1_survdth <- fread('~/Dropbox/BioLINCC files/Framingham cohort/datasets/CSV/vr_survdth_2018_a_1268d.csv',na.strings=c("NA","","NULL"))
fhs1_survstk <- fread('~/Dropbox/BioLINCC files/Framingham cohort/datasets/CSV/vr_svstk_2018_a_1269d.csv',na.strings=c("NA","","NULL"))
fhs1_soechf <- fread('~/Dropbox/BioLINCC files/Framingham cohort/datasets/CSV/VR_SOECHF_2016_a_1070d.csv',na.strings=c("NA","","NULL"))
fhs1_afcum <- fread('~/Dropbox/BioLINCC files/Framingham cohort/datasets/CSV/VR_AFCUM_2019_a_1186d_v1.csv',na.strings=c("NA","","NULL")) # Cumulative Atrial fibrillation
fhs1_survaf <- fread('~/Dropbox/BioLINCC files/Framingham cohort/datasets/CSV/VR_SURVAF_2018_a_1272D.csv',na.strings=c("NA","","NULL")) # Cumulative Atrial fibrillation
fhs1_cancer <- fread('~/Dropbox/BioLINCC files/Framingham cohort/datasets/CSV/VR_CANCER_2019_A_1162D.csv',na.strings=c("NA","","NULL")) # Incident cancer
fhs1_diab <- fread('~/Dropbox/BioLINCC files/Framingham cohort/datasets/CSV/VR_DIAB_EX28_0_0601D.csv',na.strings=c("NA","","NULL")) # Diabetes status by cycle
fhs1_soe <- fread('~/Dropbox/BioLINCC files/Framingham cohort/datasets/CSV/VR_SOE_2019_A_1217D.csv',na.strings=c("NA","","NULL"))

names(fhs1_chfinit) <- toupper(names(fhs1_chfinit))
names(fhs1_survcvd) <- toupper(names(fhs1_survcvd))
names(fhs1_survdth) <- toupper(names(fhs1_survdth))
names(fhs1_soechf) <- toupper(names(fhs1_soechf))
names(fhs1_afcum) <- toupper(names(fhs1_afcum))
names(fhs1_survaf) <- toupper(names(fhs1_survaf))
names(fhs1_cancer) <- toupper(names(fhs1_cancer))
names(fhs1_diab) <- toupper(names(fhs1_diab))
names(fhs1_soe) <- toupper(names(fhs1_soe))


fhs2_chfinit <- fread('~/Dropbox/BioLINCC files/Framingham offspring/datasets/CSV/vr_chfinit_2013_a_0828d.csv',na.strings=c("NA","","NULL"))
fhs2_survcvd <- fread('~/Dropbox/BioLINCC files/Framingham offspring/datasets/CSV/vr_survcvd_2021_a_1453d.csv',na.strings=c("NA","","NULL"))
fhs2_survdth <- fread('~/Dropbox/BioLINCC files/Framingham offspring/datasets/CSV/vr_survdth_2021_a_1452d.csv',na.strings=c("NA","","NULL"))
fhs2_survstk <- fread('~/Dropbox/BioLINCC files/Framingham offspring/datasets/CSV/vr_svstk_2021_a_1455d.csv',na.strings=c("NA","","NULL"))
fhs2_soechf <- fread('~/Dropbox/BioLINCC files/Framingham offspring/datasets/CSV/VR_SOECHF_2016_A_1070D.csv',na.strings=c("NA","","NULL"))
fhs2_afcum <- fread('~/Dropbox/BioLINCC files/Framingham offspring/datasets/CSV/VR_AFCUM_2022_A_1412D.csv',na.strings=c("NA","","NULL")) # Cumulative Atrial fibrillation
fhs2_survaf <- fread('~/Dropbox/BioLINCC files/Framingham offspring/datasets/CSV/VR_SURVAF_2021_A_1456D.csv',na.strings=c("NA","","NULL")) # Cumulative Atrial fibrillation
fhs2_cancer <- fread('~/Dropbox/BioLINCC files/Framingham offspring/datasets/CSV/VR_CANCER_2019_A_1162D.csv',na.strings=c("NA","","NULL")) # Incident cancer
fhs2_diab <- fread('~/Dropbox/BioLINCC files/Framingham offspring/datasets/CSV/vr_diab_ex10_1b_1489d.csv',na.strings=c("NA","","NULL")) # Diabetes status by cycle
fhs2_soe <- fread('~/Dropbox/BioLINCC files/Framingham offspring/datasets/CSV/VR_SOE_2022_A_1424D.csv',na.strings=c("NA","","NULL"))

names(fhs2_chfinit) <- toupper(names(fhs2_chfinit))
names(fhs2_survcvd) <- toupper(names(fhs2_survcvd))
names(fhs2_survdth) <- toupper(names(fhs2_survdth))
names(fhs2_survstk) <- toupper(names(fhs2_survstk))
names(fhs2_soechf) <- toupper(names(fhs2_soechf))
names(fhs2_afcum) <- toupper(names(fhs2_afcum))
names(fhs2_survaf) <- toupper(names(fhs2_survaf))
names(fhs2_cancer) <- toupper(names(fhs2_cancer))
names(fhs2_diab) <- toupper(names(fhs2_diab))
names(fhs2_soe) <- toupper(names(fhs2_soe))

fhs2_afcum$ORIGIN_SOURCE <- NA

fhs3_chfinit <- fread('~/Dropbox/BioLINCC files/Framingham third generation/datasets/CSV/vr_chfinit_2013_a_0828d.csv',na.strings=c("NA","","NULL"))
fhs3_survcvd <- fread('~/Dropbox/BioLINCC files/Framingham third generation/datasets/CSV/vr_survcvd_2019_a_1334d.csv',na.strings=c("NA","","NULL"))
fhs3_survdth <- fread('~/Dropbox/BioLINCC files/Framingham third generation/datasets/CSV/vr_survdth_2019_a_1337d.csv',na.strings=c("NA","","NULL"))
fhs3_survstk <- fread('~/Dropbox/BioLINCC files/Framingham third generation/datasets/CSV/vr_svstk_2019_a_1335d.csv',na.strings=c("NA","","NULL"))
fhs3_soechf <- fread('~/Dropbox/BioLINCC files/Framingham third generation/datasets/CSV/VR_SOECHF_2016_a_1070D.csv',na.strings=c("NA","","NULL"))
fhs3_afcum <- fread('~/Dropbox/BioLINCC files/Framingham third generation/datasets/CSV/VR_AFCUM_2021_A_1366D.csv',na.strings=c("NA","","NULL")) # Cumulative Atrial fibrillation
fhs3_survaf <- fread('~/Dropbox/BioLINCC files/Framingham third generation/datasets/CSV/VR_SURVAF_2019_A_1338D.csv',na.strings=c("NA","","NULL")) # Cumulative Atrial fibrillation
fhs3_cancer <- fread('~/Dropbox/BioLINCC files/Framingham third generation/datasets/CSV/VR_CANCER_2019_A_1162D.csv',na.strings=c("NA","","NULL")) # Incident cancer
fhs3_diab <- fread('~/Dropbox/BioLINCC files/Framingham third generation/datasets/CSV/VR_DIAB_EX03_3b_1312D.csv',na.strings=c("NA","","NULL")) # Diabetes status by cycle
fhs3_soe <- fread('~/Dropbox/BioLINCC files/Framingham third generation/datasets/CSV/VR_SOE_2020_A_1340D.csv',na.strings=c("NA","","NULL"))

fhs3_afcum$ORIGIN_SOURCE <- NA


dm_names <- unique(c(names(fhs1_diab),names(fhs2_diab),names(fhs3_diab)))

fhs1_diab[,dm_names[!dm_names %in% names(fhs1_diab)]] <- NA
fhs2_diab[,dm_names[!dm_names %in% names(fhs2_diab)]] <- NA
fhs3_diab[,dm_names[!dm_names %in% names(fhs3_diab)]] <- NA


fhs_chfinit <- rbind(fhs1_chfinit,fhs2_chfinit,fhs3_chfinit)
fhs_survcvd <- rbind(fhs1_survcvd,fhs2_survcvd,fhs3_survcvd)
fhs_survdth <- rbind(fhs1_survdth,fhs2_survdth,fhs3_survdth)
fhs_survstk <- rbind(fhs1_survstk,fhs2_survstk,fhs3_survstk)
fhs_survaf <- rbind(fhs1_survaf,fhs2_survaf,fhs3_survaf)
fhs_afcum <- rbind(fhs1_afcum,fhs2_afcum,fhs3_afcum)
fhs_soechf <- rbind(fhs1_soechf,fhs2_soechf,fhs3_soechf)
fhs_soe <- rbind(fhs1_soe,fhs2_soe,fhs3_soe)
fhs_cancer <- rbind(fhs1_cancer,fhs2_cancer,fhs3_cancer)
fhs_diab <- rbind(fhs1_diab,fhs2_diab,fhs3_diab)

fhs_survca <- subset(fhs_cancer,(!TOPO==173)|(TOPO==173&HIST>=8720&HIST<=8790)|HIST==8247)

fhs_survca <- 
  fhs_cancer %>%
  group_by(IDTYPE,PID) %>%
  filter(D_DATE == min(D_DATE,na.rm=T))

fhs_survca_first_any <- 
 as.data.table(sqldf("select distinct PID,IDTYPE,D_DATE
        from fhs_survca"))

fhs_survdth$dth_status <- 0 
fhs_survdth$dth_status[!is.na(fhs_survdth$DATEDTH)] <- 1 
fhs_survdth$dth_dt <- fhs_survdth$DATEDTH
fhs_survdth$dth_dt[fhs_survdth$dth_status==0] <- fhs_survdth$LASTCON[fhs_survdth$dth_status==0]

fhs_survdth$cvdth_status <- 0 
fhs_survdth$cvdth_status[fhs_survdth$CVDDEATH==1] <- 1
fhs_survdth$cvdth_dt <- fhs_survdth$dth_dt

fhs_survdth$noncvdth_status <- 0 
fhs_survdth$noncvdth_status[fhs_survdth$CVDDEATH==0] <- 1
fhs_survdth$noncvdth_dt <- fhs_survdth$dth_dt

names(fhs_survcvd)[c(3,4,5,6,7,8)] <- 
  c("cadhosp_status",
    "cadhosp_dt",
    "hfhosp_status",
    "hfhosp_dt",
    "cvhosp_status",
    "cvhosp_dt")

names(fhs_survstk)[3:6] <-
  c("cvahosp_status",
    "cva_type",
    "cva_lacunar",
    "cvahosp_dt")


fhs_outcomes <- merge(fhs_survdth[,c("PID","IDTYPE",
                                     "dth_status","dth_dt",
                                     "cvdth_status","cvdth_dt",
                                     "noncvdth_status","noncvdth_dt")],
                      fhs_survcvd[,c("PID","IDTYPE",
                                     "cadhosp_status","cadhosp_dt",
                                     "cvhosp_status","cvhosp_dt",
                                     "hfhosp_status","hfhosp_dt")])

fhs_outcomes <-
  merge(fhs_outcomes,
        fhs_survstk[,c("PID","IDTYPE","cvahosp_status","cva_type","cvahosp_dt")],
        by=c("PID","IDTYPE"),
        all.x=T)


## Ischemic stroke

fhs_outcomes$ischem_cvahosp_status <- 0
fhs_outcomes$ischem_cvahosp_status[
  fhs_outcomes$cva_type %in% 
    c(11,13)] <- 1


fhs_outcomes$ischem_cvahosp_dt[
  fhs_outcomes$ischem_cvahosp_status==1] <-
  fhs_outcomes$cvahosp_dt[
    fhs_outcomes$ischem_cvahosp_status==1]

fhs_outcomes$ischem_cvahosp_dt[
  fhs_outcomes$ischem_cvahosp_status==0] <-
  fhs_outcomes$dth_dt[
    fhs_outcomes$ischem_cvahosp_status==0]


fhs_outcomes[
  fhs_outcomes$cva_type %in% 
    c(10,16,17,19),
  c("ischem_cvahosp_status",
    "ischem_cvahosp_dt")] <- NA


## Hemorrhagic

fhs_outcomes$hemo_cvahosp_status <- 0
fhs_outcomes$hemo_cvahosp_status[
  fhs_outcomes$cva_type %in% 
    c(14,15)] <- 1


fhs_outcomes$hemo_cvahosp_dt[
  fhs_outcomes$hemo_cvahosp_status==1] <-
  fhs_outcomes$cvahosp_dt[
    fhs_outcomes$hemo_cvahosp_status==1]

fhs_outcomes$hemo_cvahosp_dt[
  fhs_outcomes$hemo_cvahosp_status==0] <-
  fhs_outcomes$dth_dt[
    fhs_outcomes$hemo_cvahosp_status==0]

fhs_outcomes[
  fhs_outcomes$cva_type %in% 
    c(10,16,17,19),
  c("hemo_cvahosp_status",
    "hemo_cvahosp_dt")] <- NA


fhs_outcomes <- merge(fhs_outcomes,
                      fhs_chfinit[,c("PID",
                                     "IDTYPE",
                                     "CHF011D",
                                     "CHF017")],
                      by.x=c("PID","IDTYPE","hfhosp_dt"),
                      by.y=c("PID","IDTYPE","CHF011D"),
                      all.x=T)


fhs_outcomes$hfhosp_ef_cat[fhs_outcomes$CHF017==0] <- "Normal"
fhs_outcomes$hfhosp_ef_cat[fhs_outcomes$CHF017==3] <- "Mildly reduced"
fhs_outcomes$hfhosp_ef_cat[fhs_outcomes$CHF017==2] <- "Moderately reduced"
fhs_outcomes$hfhosp_ef_cat[fhs_outcomes$CHF017==1] <- "Severely reduced"


fhs_outcomes$study <- "FHS"

write_parquet(fhs_outcomes,
              "~/Dropbox/ADAPT-HF/Master HDCP files/Cohort data/fhs_outcomes.parquet")

#=========================================================================#
####                    ** Combine FHS Dates **                        ####
#=========================================================================#

# ==============================================================#
####       Framingham Original cohort dates & outcomes       ####
# ==============================================================#

fhs1_dates <- fread('~/Dropbox/BioLINCC files/Framingham cohort/datasets/CSV/vr_dates_2014_a_0912d.csv',na.strings=c("NA","","NULL"))
fhs1_wkthru <- fread('~/Dropbox/BioLINCC files/Framingham cohort/datasets/CSV/vr_wkthru_ex32_0_0997d.csv',na.strings=c("NA","","NULL"))

fhs1_melt_sex <-as.data.table(sqldf("select PID,
                       IDTYPE,
                       1 as visit,
                       'sex' as variable,
                       SEX as value,
                       'fhs1_dates' as form
                       from fhs1_dates"))

names(fhs1_dates) <- toupper(names(fhs1_dates))
names(fhs1_wkthru) <- toupper(names(fhs1_wkthru))

for (v in 1:32) {
  fhs1_wkthru[,paste("BMIC",v,sep="")] <- 
    round(calc_bmi(dat=as.data.frame(fhs1_wkthru),
                   weight=paste("WGT",v,sep=""),
                   height="HGT1",
                   metric=F),2)
}

for (whr in c(19:23)) {
  fhs1_wkthru[,paste("WHR",whr,sep="")] <-
    round(as.data.frame(fhs1_wkthru)[,paste("WAIST",whr,sep="")]/as.data.frame(fhs1_wkthru)[,paste("HIP",whr,sep="")],3)
}

### FHS 2 Walkthru

fhs2_offspring_wkthru <- fread('~/Dropbox/BioLINCC files/Framingham offspring/datasets/CSV/vr_wkthru_ex10_1b_1488d.csv',na.strings=c("NA","","NULL"))

fhs2_offspring_wkthru <-
  fhs2_offspring_wkthru %>%
  select(-att5_status,
         -att10_status,
         -date5_fu,
         -date10_fu)

fhs2_dates <- fread('~/Dropbox/BioLINCC files/Framingham offspring/datasets/CSV/vr_dates_2022_a_1487d.csv',na.strings=c("NA","","NULL"))

fhs2_race_1 <- fread('~/Dropbox/BioLINCC files/Framingham offspring/datasets/CSV/race_1.csv',na.strings=c("NA","","NULL")) # Physical exam questionnaire, exam 5
names(fhs2_race_1) <- toupper(names(fhs2_race_1))

fhs2_race_1$RACE[fhs2_race_1$ETHNICITY=="Hisp"] <- "Hisp" 
fhs2_race_1$RACE[is.na(fhs2_race_1$RACE)] <- "W"

fhs2_melt_race <-
  melt(fhs2_race_1,
       id.vars=c("PID","IDTYPE"),
       na.rm=T)

fhs2_offspring_wkthru <-
  merge(fhs2_offspring_wkthru,
        fhs2_race_1,
        by=c("PID","IDTYPE"),
        all.x=T)


fhs2_melt_race$form <- "fhs2_race_1"
fhs2_melt_race$visit <- 1


fhs2_melt_sex <-as.data.table(sqldf("select PID,
                       IDTYPE as IDTYPE,
                       1 as visit,
                       'sex' as variable,
                       SEX as value,
                       'fhs2_dates' as form
                       from fhs2_dates"))


names(fhs2_dates) <- toupper(names(fhs2_dates))
names(fhs2_offspring_wkthru) <- toupper(names(fhs2_offspring_wkthru))

for (whr in c(1,2,4,5,6,7,9,10)) {
  this_waist <- paste("WAIST",whr,sep="")
  this_hip <- paste("HIP",whr,sep="")
  fhs2_offspring_wkthru[,paste("WHR",whr,sep="")] <-
    round(fhs2_offspring_wkthru[,..this_waist]/fhs2_offspring_wkthru[,..this_hip],3)
}


for (i in c(2:10)) {
  fhs2_offspring_wkthru[,paste("EGFR_MDRD",i,sep="")] <-
    calc_MDRD4(as.data.frame(fhs2_offspring_wkthru),
               age=paste("AGE",i,sep=""),
               cr=paste("CREAT",i,sep=""),
               sex="SEX",
               race="RACE",
               black="B")
}


#### FHS 3 


fhs3_wkthru <- fread('~/Dropbox/BioLINCC files/Framingham third generation/datasets/CSV/vr_wkthru_ex03_3b_1191d.csv',na.strings=c("NA","","NULL"))
fhs3_dates <- fread('~/Dropbox/BioLINCC files/Framingham third generation/datasets/CSV/vr_dates_2019_a_1175d.csv',na.strings=c("NA","","NULL"))
names(fhs3_dates) <- toupper(names(fhs3_dates))
names(fhs3_wkthru) <- toupper(names(fhs3_wkthru))

fhs3_melt_sex <-as.data.table(sqldf("select PID,
                       IDTYPE,
                       1 as visit,
                       'sex' as variable,
                       SEX as value,
                       'fhs3_dates' as form
                       from fhs3_dates"))

fhs3_black <-
  fread('~/Dropbox/BioLINCC files/Framingham third generation/datasets/CSV/e_exam_ex01_3_0086d.csv')

fhs3_black <-
  fhs3_black[,c("PID",
                "IDTYPE",
                "G3A485")]

fhs_spouse_black <-
  fread('~/Dropbox/BioLINCC files/Framingham third generation/datasets/CSV/e_exam_ex01_2_0813d.csv')

fhs_spouse_black <-    
  fhs_spouse_black[,
                   c("PID",
                     "IDTYPE",
                     "G3A485")]

fhs_omni2_black <-
  fread('~/Dropbox/BioLINCC files/Framingham third generation/datasets/CSV/e_exam_ex01_72_0652d.csv')

fhs_omni2_black <-    
  fhs_omni2_black[, 
                  c("PID",
                    "IDTYPE",
                    "G3A485")]

fhs3_black_all <-
  rbind(fhs3_black,
        fhs_spouse_black,
        fhs_omni2_black)

fhs3_wkthru <-
  merge(fhs3_wkthru,
        fhs3_black_all,
        by=c("PID","IDTYPE"))

names(fhs3_wkthru)[ncol(fhs3_wkthru)] <- "BLACK"


for (i in c(1:3)) {
  fhs3_wkthru[,paste("EGFR_MDRD",i,sep="")] <-
    calc_MDRD4(as.data.frame(fhs3_wkthru),
               age=paste("AGE",i,sep=""),
               cr=paste("CREAT",i,sep=""),
               sex="SEX",
               race="BLACK",
               black="1")
}


#======================================================#
####  Assemble Framingham dates and outcomes files  ####
#======================================================#


fhs_dates_fields <- names(fhs1_dates)
fhs2_dates[,fhs_dates_fields[!fhs_dates_fields %in% names(fhs2_dates)]] <- NA
fhs3_dates[,fhs_dates_fields[!fhs_dates_fields %in% names(fhs3_dates)]] <- NA


fhs_dates <- 
  rbindlist(
    list(fhs1_dates[,..fhs_dates_fields],
         fhs2_dates[,..fhs_dates_fields],
         fhs3_dates[,..fhs_dates_fields]))


fhs_dates <- merge(fhs_dates,
                   fhs_survcvd,
                   by=c("IDTYPE","PID"),
                   all.x=T)
fhs_dates$DATE1 <- 0

for (i in 2:20) {
  thisexamdate <- paste0("DATE", i)
  fhs_dates[
    hfhosp_status == 1 & 
      hfhosp_dt > get(thisexamdate) & 
      !is.na(hfhosp_dt) & 
      !is.na(get(thisexamdate)),
    last_exam_before_hf := i
  ]
}

age_cols <- names(fhs_dates)[grep("AGE", names(fhs_dates))]
date_cols <- names(fhs_dates)[grep("DATE", names(fhs_dates))]

col_age <- c("PID","IDTYPE","SEX",age_cols)

fhs_dates_long_age <- melt(fhs_dates[,..col_age],
                           id.vars=c("PID","IDTYPE","SEX"),
                           na.rm=T,
                           factorsAsStrings = T)

fhs_dates_long_age$variable <- as.character(fhs_dates_long_age$variable)

fhs_dates_long_age$visit <- 
  substring(fhs_dates_long_age$variable,
            str_locate(fhs_dates_long_age$variable,
                       "\\d")[1],length(fhs_dates_long_age$variable))

fhs_dates_long_age$variable <- 
  gsub("\\d","",fhs_dates_long_age$variable)

###########################3

col_dates <- 
  c("PID","IDTYPE",date_cols)

fhs_dates_long_date <- 
  melt(fhs_dates[,..col_dates],
       id.vars=c("PID","IDTYPE"),
       na.rm=T,
       factorsAsStrings = T)

fhs_dates_long_date$variable <- as.character(fhs_dates_long_date$variable)

fhs_dates_long_date <- subset(fhs_dates_long_date,!variable=="CHFDATE")

fhs_dates_long_date$visit <- 
  substring(fhs_dates_long_date$variable,
            str_locate(fhs_dates_long_date$variable,
                       "\\d")[1],length(fhs_dates_long_date$variable))

fhs_dates_long_date$variable <- gsub("\\d","",fhs_dates_long_date$variable)

names(fhs_dates_long_date)[names(fhs_dates_long_date)=="value"] <- "visitdays"
names(fhs_dates_long_age)[names(fhs_dates_long_age)=="value"] <- "age_obs"

fhs_dates_long <- merge(fhs_dates_long_age[,c("PID","IDTYPE","SEX","visit","age_obs")],
                        fhs_dates_long_date[,c("PID","IDTYPE","visit","visitdays")],
                        by=c("PID","IDTYPE","visit"))

fhs_dates_long$study <- "FHS"

names(fhs_dates_long)[2] <- "cohort"

fhs_dates_long$visit <- as.integer(fhs_dates_long$visit)
fhs_dates_long$cohort <- as.character(fhs_dates_long$cohort)

fhs_dates_long <- merge(fhs_dates_long,
                        visit_yrs[visit_yrs$study=="FHS",
                                  c("cohort","study","visit","visit_yr")],
                        by=c("visit","study","cohort"))

fhs_dates_long$cohort_name[fhs_dates_long$cohort==0] <- "Original"
fhs_dates_long$cohort_name[fhs_dates_long$cohort==1] <- "Offspring"
fhs_dates_long$cohort_name[fhs_dates_long$cohort==7] <- "OMNI 1"
fhs_dates_long$cohort_name[fhs_dates_long$cohort==3] <- "Gen III"
fhs_dates_long$cohort_name[fhs_dates_long$cohort==2] <- "Offspring spouse"
fhs_dates_long$cohort_name[fhs_dates_long$cohort==72] <- "OMNI 2"

fhs_dates_long$patientid <- fhs_dates_long$PID
fhs_dates_long$sex[fhs_dates_long$SEX==1] <- "Male"
fhs_dates_long$sex[fhs_dates_long$SEX==2] <- "Female"

rm(fhs_dates_long_age,
   fhs_dates_long_date)

#====================================================#
####  Breakdown Framingham Original wkthru table  ####
#====================================================#


fhs1_melt_wkthru <- melt(fhs1_wkthru,
                         id.vars=c("PID","IDTYPE","SEX"),
                         na.rm=T,
                         factorsAsStrings=T)

fhs1_melt_wkthru$variable <-  as.character(fhs1_melt_wkthru$variable)
fhs1_melt_wkthru$visit  <- 
  substr(fhs1_melt_wkthru$variable,
         str_locate(fhs1_melt_wkthru$variable,"\\d")[,1],
         nchar(fhs1_melt_wkthru$variable))

fhs1_melt_wkthru$variable <- 
  substr(fhs1_melt_wkthru$variable,
         1, str_locate(fhs1_melt_wkthru$variable,"\\d")[,1]-1)

fhs1_melt_wkthru$value[fhs1_melt_wkthru$variable=="WAIST"] <- fhs1_melt_wkthru$value[fhs1_melt_wkthru$variable=="WAIST"] * 2.54
fhs1_melt_wkthru_kg <- subset(fhs1_melt_wkthru,variable=="WGT")
fhs1_melt_wkthru_kg$value <- fhs1_melt_wkthru_kg$value/2.20462
fhs1_melt_wkthru_kg$variable <- "WGT_KG"
fhs1_melt_wkthru <- rbind(fhs1_melt_wkthru,fhs1_melt_wkthru_kg)
fhs1_melt_wkthru$form <- "fhs1_wkthru"


# ============================================================== #
####                  Assemble Framingham DM                  ####
# ============================================================== #


fhs1_vr_diab <- fread('~/Dropbox/BioLINCC files/Framingham cohort/datasets/CSV/VR_DIAB_EX28_0_0601D.csv',
                      na.strings=c("NA","","NULL"),
                      stringsAsFactors=F)

names(fhs1_vr_diab) <- toupper(names(fhs1_vr_diab))

fhs1_melt_vr_diab <- melt(fhs1_vr_diab[,c(1,25,82:109)],
                          id.vars=c("PID",
                                    "IDTYPE"),
                          na.rm=T,
                          factorsAsStrings=T)

fhs1_melt_vr_diab$variable <- as.character(fhs1_melt_vr_diab$variable)

fhs1_melt_vr_diab$visit <- str_select(fhs1_melt_vr_diab$variable,
                                      after="BG200_HX_DIAB")

fhs1_melt_vr_diab$variable <- "BG200_HX_DIAB"

fhs1_melt_vr_diab$form <- "fhs1_vr_diab"


#### Diabetes status by cycle

fhs2_vr_diab <- 
  fread('~/Dropbox/BioLINCC files/Framingham offspring/datasets/CSV/vr_diab_ex10_1b_1489d.csv',
        na.strings=c("NA","","NULL"),
        stringsAsFactors=F)
names(fhs2_vr_diab) <- toupper(names(fhs2_vr_diab))

diab_cols <-
  c("PID",
    "IDTYPE",
    "HX_DIAB1",
    "HX_DIAB2",
    "HX_DIAB3",
    "HX_DIAB4",
    "HX_DIAB5",
    "HX_DIAB6",
    "HX_DIAB7",
    "HX_DIAB8",
    "HX_DIAB9",
    "HX_DIAB10")

fhs2_melt_vr_diab <- melt(fhs2_vr_diab[,..diab_cols],
                          id.vars=c("PID",
                                    "IDTYPE"),
                          na.rm=T,
                          factorsAsStrings=T)

fhs2_melt_vr_diab$variable <- as.character(fhs2_melt_vr_diab$variable)

fhs2_melt_vr_diab$visit <- str_select(fhs2_melt_vr_diab$variable,
                                      after="HX_DIAB")

fhs2_melt_vr_diab$variable <- "HX_DIAB"

fhs2_melt_vr_diab$form <- "fhs2_vr_diab"


####

fhs3_vr_diab <- fread('~/Dropbox/BioLINCC files/Framingham third generation/datasets/CSV/VR_DIAB_EX03_3b_1312D.csv',na.strings=c("NA","","NULL")) # Diabetes status by cycle
names(fhs3_vr_diab) <- toupper(names(fhs3_vr_diab))

fhs3_melt_vr_diab <- melt(fhs3_vr_diab[,c("PID","IDTYPE","HX_DIAB1","HX_DIAB2")],
                          id.vars=c("PID",
                                    "IDTYPE"),
                          na.rm=T,
                          factorsAsStrings=T)

fhs3_melt_vr_diab$variable <- as.character(fhs3_melt_vr_diab$variable)

fhs3_melt_vr_diab$visit <- str_select(fhs3_melt_vr_diab$variable,
                                      after="HX_DIAB")

fhs3_melt_vr_diab$variable <- "HX_DIAB"

fhs3_melt_vr_diab$form <- "fhs3_vr_diab"

## For some reason FHS Omni did not was not included in vr_diab files.  
## Rederived baseline DM status per documentation (BG > 200 or fasting BG ≥ 126 or DM tx)
## Did not do Omni1 visits 2-4 due-to missing variables. 


fhs_vr_diab <- rbind(fhs1_melt_vr_diab,
                     fhs2_melt_vr_diab,
                     fhs3_melt_vr_diab)

names(fhs_vr_diab)[2] <- "cohort"

fhs_vr_diab$cohort <- as.character(fhs_vr_diab$cohort)

fhs_vr_diab$visit <- as.integer(fhs_vr_diab$visit)

fhs_vr_diab <- merge(fhs_vr_diab,
                     fhs_dates_long[,c("PID","cohort","visit","visitdays")],
                     by=c("PID","cohort","visit"))

fhs_vr_diab$study <- "FHS"

rm(fhs1_melt_vr_diab,
   fhs2_melt_vr_diab,
   fhs3_melt_vr_diab)


#=========================================================================#
#                                                                         #
####          ******** FRAMINGHAM ORIGINAL DATA ********               ####
#                                                                         #
#=========================================================================#

fhs1_ecgalld <- fread('~/Dropbox/BioLINCC files/Framingham cohort/datasets/CSV/ECG_ALLD.csv',na.strings=c("NA","","NULL"))
fhs1_ecgalld$visit <- 11
fhs1_melt_ecgalld <- melt(fhs1_ecgalld,id.vars=c("PID","IDTYPE","visit"),na.rm=T,factorsAsStrings=T)
fhs1_melt_ecgalld$form <- "fhs1_ecgall"

# ==================================================================##
####                    FHS original, Exam 1-7                    ####
# ==================================================================##

fhs1_ex0_7d <- fread('~/Dropbox/BioLINCC files/Framingham cohort/datasets/CSV/EX0_7D.csv',na.strings=c("NA","","NULL"))  # Exams 1-7

fhs1_ex0_7d$height_cm <- fhs1_ex0_7d$MF67*2.54
fhs1_ex0_7d$height_m <- fhs1_ex0_7d$height_cm/100

fhs1_ex0_7d$MF69_kg <- fhs1_ex0_7d$MF69/2.2
fhs1_ex0_7d$MF166_kg <- fhs1_ex0_7d$MF166/2.2
fhs1_ex0_7d$MF180_kg <- fhs1_ex0_7d$MF180/2.2
fhs1_ex0_7d$MF216_kg <- fhs1_ex0_7d$MF216/2.2
fhs1_ex0_7d$MF292_kg <- fhs1_ex0_7d$MF292/2.2
fhs1_ex0_7d$MF380_kg <- fhs1_ex0_7d$MF380/2.2
fhs1_ex0_7d$MF470_kg <- fhs1_ex0_7d$MF470/2.2

fhs1_exam1_calc_vars <- "MF69_kg"
fhs1_exam2_calc_vars <- "MF166_kg"
fhs1_exam3_calc_vars <- "MF180_kg"
fhs1_exam4_calc_vars <- "MF216_kg"
fhs1_exam5_calc_vars <- "MF292_kg"
fhs1_exam6_calc_vars <- "MF380_kg"
fhs1_exam7_calc_vars <- "MF470_kg"

fhs1_m2 <- fhs1_ex0_7d[,c("PID","IDTYPE","height_m","height_cm")]
fhs1_m2$height_m2 <- fhs1_m2$height_m^2

fhs1_ex0_7d$MF78_wk <- fhs1_ex0_7d$MF78/4.354
fhs1_ex0_7d$MF253_wk <- fhs1_ex0_7d$MF253/4.354
fhs1_ex0_7d$MF667_wk <- fhs1_ex0_7d$MF667/4.354

fhs1_exam2_calc_vars <- c(fhs1_exam2_calc_vars,"MF78_wk")
fhs1_exam4_calc_vars <- c(fhs1_exam4_calc_vars,"MF253_wk")
fhs1_exam7_calc_vars <- c(fhs1_exam7_calc_vars,"MF667_wk")

fhs1_ex0_7d$MF63_gdL <- fhs1_ex0_7d$MF63/10
fhs1_ex0_7d$MF161_gdL <- fhs1_ex0_7d$MF161/10
fhs1_ex0_7d$MF168_gdL <- fhs1_ex0_7d$MF168/10
fhs1_ex0_7d$MF209_gdL <- fhs1_ex0_7d$MF209/10
fhs1_ex0_7d$MF312_gdL <- fhs1_ex0_7d$MF312/10
fhs1_ex0_7d$MF392_gdL <- fhs1_ex0_7d$MF392/10

fhs1_exam1_calc_vars <- c(fhs1_exam1_calc_vars,"MF63_gdL")
fhs1_exam2_calc_vars <- c(fhs1_exam2_calc_vars,"MF161_gdL")
fhs1_exam3_calc_vars <- c(fhs1_exam3_calc_vars,"MF168_gdL")
fhs1_exam4_calc_vars <- c(fhs1_exam4_calc_vars,"MF209_gdL")
fhs1_exam5_calc_vars <- c(fhs1_exam5_calc_vars,"MF312_gdL")
fhs1_exam6_calc_vars <- c(fhs1_exam6_calc_vars,"MF392_gdL")

fhs1_ex0_7d$MF223_cm <- fhs1_ex0_7d$MF223/10

fhs1_exam4_calc_vars <- c(fhs1_exam4_calc_vars,"MF223_cm")

fhs1_ex0_7d$MF302_ratio <- fhs1_ex0_7d$MF302/100 # FEV1/FVC
fhs1_ex0_7d$MF299_L <- fhs1_ex0_7d$MF299/10
fhs1_ex0_7d$FEV1_ex5 <- fhs1_ex0_7d$MF302_ratio*fhs1_ex0_7d$MF299_L

fhs1_exam5_calc_vars <- c(fhs1_exam5_calc_vars,"FEV1_ex5")
fhs1_exam5_calc_vars <- c(fhs1_exam5_calc_vars,"MF302_ratio")
fhs1_exam5_calc_vars <- c(fhs1_exam5_calc_vars,"MF299_L")


fhs1_ex0_7d$MF451_dth[fhs1_ex0_7d$MF452==0&!is.na(fhs1_ex0_7d$MF452)] <- 
  fhs1_ex0_7d$MF451[fhs1_ex0_7d$MF452==0&!is.na(fhs1_ex0_7d$MF452)]

fhs1_exam6_calc_vars <- c(fhs1_exam6_calc_vars,"MF451_dth")


fhs1_ex0_7d$MF451_alive[fhs1_ex0_7d$MF452==1&!is.na(fhs1_ex0_7d$MF452)] <- 
  fhs1_ex0_7d$MF451[fhs1_ex0_7d$MF452==1&!is.na(fhs1_ex0_7d$MF452)]

fhs1_exam6_calc_vars <- c(fhs1_exam6_calc_vars,"MF451_alive")

fhs1_ex0_7d$MF453_dth[fhs1_ex0_7d$MF454==0&!is.na(fhs1_ex0_7d$MF454)] <- 
  fhs1_ex0_7d$MF453[fhs1_ex0_7d$MF454==0&!is.na(fhs1_ex0_7d$MF454)]

fhs1_exam6_calc_vars <- c(fhs1_exam6_calc_vars,"MF453_dth")

fhs1_ex0_7d$MF453_alive[fhs1_ex0_7d$MF454==1&!is.na(fhs1_ex0_7d$MF454)] <- 
  fhs1_ex0_7d$MF453[fhs1_ex0_7d$MF454==1&!is.na(fhs1_ex0_7d$MF454)]

fhs1_exam6_calc_vars <- c(fhs1_exam6_calc_vars,"MF453_alive")


exam1_names <- colnames(fhs1_ex0_7d)[c(1:71,73:74,540,541:542,549)]
exam1_names <- c(exam1_names,fhs1_exam1_calc_vars,"height_cm","height_m")
exam2_names <- colnames(fhs1_ex0_7d)[c(72,75:154,422:427,543,546,550)]
exam2_names <- c(exam2_names,fhs1_exam2_calc_vars)
exam3_names <- colnames(fhs1_ex0_7d)[c(155:192,428:431,544,547,551)]
exam3_names <- c(exam3_names,fhs1_exam3_calc_vars)
exam4_names <- colnames(fhs1_ex0_7d)[c(193:265,432:436,552,539,545,548,552)]
exam4_names <- c(exam4_names,fhs1_exam4_calc_vars)
exam5_names <- colnames(fhs1_ex0_7d)[c(266:350,553)]
exam5_names <- c(exam5_names,fhs1_exam5_calc_vars)
exam6_names <- colnames(fhs1_ex0_7d)[c(351:421,554)]
exam6_names <- c(exam6_names,fhs1_exam6_calc_vars)
exam7_names <- colnames(fhs1_ex0_7d)[c(437:538,555:556)]
exam7_names <- c(exam7_names,fhs1_exam7_calc_vars)

fhs1_melt_ex0_7d <- melt(fhs1_ex0_7d,id.vars=c("PID","IDTYPE"),na.rm=T,factorsAsStrings=T)
fhs1_melt_ex0_7d$form <- "fhs1_ex0_7"

fhs1_melt_ex0_7d$visit[fhs1_melt_ex0_7d$variable %in% exam1_names] <- 1
fhs1_melt_ex0_7d$visit[fhs1_melt_ex0_7d$variable %in% exam2_names] <- 2
fhs1_melt_ex0_7d$visit[fhs1_melt_ex0_7d$variable %in% exam3_names] <- 3
fhs1_melt_ex0_7d$visit[fhs1_melt_ex0_7d$variable %in% exam4_names] <- 4
fhs1_melt_ex0_7d$visit[fhs1_melt_ex0_7d$variable %in% exam5_names] <- 5
fhs1_melt_ex0_7d$visit[fhs1_melt_ex0_7d$variable %in% exam6_names] <- 6
fhs1_melt_ex0_7d$visit[fhs1_melt_ex0_7d$variable %in% exam7_names] <- 7


rm(fhs1_ex0_7d,
   fhs1_exam1_calc_vars,
   fhs1_exam2_calc_vars,
   fhs1_exam3_calc_vars,
   fhs1_exam4_calc_vars,
   fhs1_exam5_calc_vars,
   fhs1_exam6_calc_vars,
   fhs1_exam7_calc_vars)

rm(list=ls(pattern="\\bexam\\d."))

# ==============================================================##
####                     FHS original, Exam 8                 ####
# ==============================================================##

fhs1_ex0_8d <- fread('~/Dropbox/BioLINCC files/Framingham cohort/datasets/CSV/EX0_8D_V1.csv',na.strings=c("NA","","NULL")) # Exam 8
fhs1_ex0_8d$visit <- 8

fhs1_ex0_8d[fhs1_ex0_8d==2000] <- NA
fhs1_ex0_8d[fhs1_ex0_8d==9999] <- NA
fhs1_ex0_8d[fhs1_ex0_8d==1999] <- NA

fhs1_melt_ex0_8d <- melt(fhs1_ex0_8d,id.vars=c("PID","IDTYPE","visit"),na.rm=T,factorsAsStrings=T)
fhs1_melt_ex0_8d$form <- "fhs1_ex0_8"

rm(fhs1_ex0_8d)

# ==============================================================##
####                     FHS original, Exam 9                 ####
# ==============================================================##

fhs1_ex0_9d <- fread('~/Dropbox/BioLINCC files/Framingham cohort/datasets/CSV/EX0_9D.csv',na.strings=c("NA","","NULL")) # Exam 9
fhs1_ex0_9d$visit <- 9
fhs1_melt_ex0_9d <- melt(fhs1_ex0_9d,id.vars=c("PID","IDTYPE","visit"),na.rm=T,factorsAsStrings=T)
fhs1_melt_ex0_9d$form <- "fhs1_ex0_9"

rm(fhs1_ex0_9d)


# ==============================================================#
####                    FHS original, Exam 10                ####
# ==============================================================#

fhs1_ex0_10d <- fread('~/Dropbox/BioLINCC files/Framingham cohort/datasets/CSV/EX0_10D_V1.csv',na.strings=c("NA","","NULL")) # Exam 10
fhs1_ex0_10d$visit <- 10
fhs1_melt_ex0_10d <- melt(fhs1_ex0_10d,id.vars=c("PID","IDTYPE","visit"),na.rm=T,factorsAsStrings=T)
fhs1_melt_ex0_10d$form <- "fhs1_ex0_10"

rm(fhs1_ex0_10d)


# ==============================================================#
####                    FHS original, Exam 11                ####
# ==============================================================#

fhs1_ex0_11d <- fread('~/Dropbox/BioLINCC files/Framingham cohort/datasets/CSV/EX0_11D.csv',na.strings=c("NA","","NULL")) # Exam 11
fhs1_ex0_11d$visit <- 11
fhs1_ex0_11d[fhs1_ex0_11d==99] <- NA

fhs1_ex0_11d$fhs_pai <- 
  calc_framingham_pai(as.data.frame(fhs1_ex0_11d),
                      slp_hrs="FD58",
                      sed_hrs="FD60",
                      slgt_hrs="FD61",
                      mod_hrs="FD62",
                      hvy_hrs="FD63")

fhs1_melt_ex0_11d <- melt(fhs1_ex0_11d,id.vars=c("PID","IDTYPE","visit"),na.rm=T,factorsAsStrings=T)
fhs1_melt_ex0_11d$form <- "fhs1_ex0_11"

rm(fhs1_ex0_11d)


# ===============================================================#
####                    FHS original, Exam 12                 ####
# ===============================================================#

fhs1_ex0_12d <- fread('~/Dropbox/BioLINCC files/Framingham cohort/datasets/CSV/EX0_12D.csv',na.strings=c("NA","","NULL")) # Exam 12
fhs1_ex0_12d$visit <- 12

fhs1_ex0_12d$FE127[fhs1_ex0_12d$FE127==99] <- NA 
fhs1_ex0_12d$FE128[fhs1_ex0_12d$FE128==99] <- NA 
fhs1_ex0_12d$FE129[fhs1_ex0_12d$FE129==99] <- NA 
fhs1_ex0_12d$FE186[fhs1_ex0_12d$FE186==99] <- NA 

fhs1_ex0_12d$fhs_pai <- 
  calc_framingham_pai(as.data.frame(fhs1_ex0_12d),
                      slp_hrs="FE186",
                      sed_hrs="FE187",
                      slgt_hrs="FE188",
                      mod_hrs="FE189",
                      hvy_hrs="FE190")

fhs1_ex0_12d$drinks_wk <- fhs1_ex0_12d$FE127+fhs1_ex0_12d$FE128+fhs1_ex0_12d$FE129

fhs1_melt_ex0_12d <- melt(fhs1_ex0_12d,id.vars=c("PID","IDTYPE","visit"),na.rm=T,factorsAsStrings=T)
fhs1_melt_ex0_12d$form <- "fhs1_ex0_12"

rm(fhs1_ex0_12d)


# ==============================================================##
##                 FHS original, lipids exam 12                 ##
# ==============================================================##

fhs1_l_lipids_ex12 <- fread('~/Dropbox/BioLINCC files/Framingham cohort/datasets/CSV/L_LIPIDS_EX12_0_0342D.csv',na.strings=c("NA","","NULL"))  # Standard lipid panel
fhs1_l_lipids_ex12$visit <- 12

fhs1_melt_l_lipids_ex12 <- melt(fhs1_l_lipids_ex12,id.vars=c("PID","IDTYPE","visit"),na.rm=T,factorsAsStrings=T)
fhs1_melt_l_lipids_ex12$form <- "fhs1_lipids"

rm(fhs1_l_lipids_ex12)


# ==================================================================#
####                    FHS original, Exam 13                    ####
# ==================================================================#

fhs1_ex0_13d <- fread('~/Dropbox/BioLINCC files/Framingham cohort/datasets/CSV/EX0_13D.csv',na.strings=c("NA","","NULL")) # Exam 13
fhs1_ex0_13d$visit <- 13

fhs1_ex0_13d[fhs1_ex0_13d=="."] <- NA

fhs1_ex0_13d$drinks_wk <- fhs1_ex0_13d$FF125+fhs1_ex0_13d$FF126+fhs1_ex0_13d$FF127


fhs1_melt_ex0_13d <- melt(fhs1_ex0_13d,id.vars=c("PID","IDTYPE","visit"),na.rm=T,factorsAsStrings=T)
fhs1_melt_ex0_13d$form <- "fhs1_ex0_13"

rm(fhs1_ex0_13d)


# ===============================================================#
####                    FHS original, Exam 14                 ####
# ===============================================================#

fhs1_ex0_14d <- fread('~/Dropbox/BioLINCC files/Framingham cohort/datasets/CSV/EX0_14D.csv',na.strings=c("NA","","NULL")) # Exam 14
fhs1_ex0_14d$visit <- 14

fhs1_meno <- fread('~/Dropbox/BioLINCC files/Framingham cohort/datasets/CSV/vr_meno_ex14_0_0153d.csv',na.strings=c("NA","","NULL")) # Exam 14
fhs1_meno$visit <- 14

fhs1_meno$menopause[fhs1_meno$AM2==0] <- "N"
fhs1_meno$menopause[between(fhs1_meno$AM2,18,59)] <- "Y"

fhs1_ex0_14d[fhs1_ex0_14d=="."] <- NA

fhs1_ex0_14d$drinks_wk <- fhs1_ex0_14d$FG118+fhs1_ex0_14d$FG119+fhs1_ex0_14d$FG120

fhs1_ex0_14d$FG62_kg <- fhs1_ex0_14d$FG62/2.2
fhs1_ex0_14d <- merge(fhs1_ex0_14d,fhs1_m2[,c("PID","height_m2")],by=c("PID"),all.x=T)
fhs1_ex0_14d$FG62_bmi <- fhs1_ex0_14d$FG62_kg/fhs1_ex0_14d$height_m2


fhs1_melt_ex0_14d <- melt(fhs1_ex0_14d,id.vars=c("PID","IDTYPE","visit"),na.rm=T,factorsAsStrings=T)
fhs1_melt_ex0_14d$form <- "fhs1_ex0_14"


rm(fhs1_ex0_14d)


# ==============================================================#
####                    FHS original, Exam 15                ####
# ==============================================================#


fhs1_ex0_15d <- fread('~/Dropbox/BioLINCC files/Framingham cohort/datasets/CSV/EX0_15D.csv',na.strings=c("NA","","NULL")) # Exam 15
fhs1_ex0_15d$visit <- 15

fhs1_ex0_15d[fhs1_ex0_15d=="."] <- NA

fhs1_ex0_15d$drinks_wk <- fhs1_ex0_15d$FH115+fhs1_ex0_15d$FH116+fhs1_ex0_15d$FH117


fhs1_melt_ex0_15d <- melt(fhs1_ex0_15d,id.vars=c("PID","IDTYPE","visit"),na.rm=T,factorsAsStrings=T)
fhs1_melt_ex0_15d$form <- "fhs1_ex0_15"

rm(fhs1_ex0_15d)


# ===============================================================#
####                    FHS original, Exam 16                 ####
# ===============================================================#

fhs1_ex0_16d <-fread('~/Dropbox/BioLINCC files/Framingham cohort/datasets/CSV/EX0_16D_V2.csv',na.strings=c("NA","","NULL")) # Exam 16
fhs1_ex0_16d$visit <- 16

fhs1_ex0_16d <- merge(fhs1_ex0_16d,fhs1_dates[,c("PID","IDTYPE","SEX")],all.x=T)

fhs1_ex0_16d$FI297_cm  <- fhs1_ex0_16d$FI297/10
fhs1_ex0_16d$FI302_cm  <- fhs1_ex0_16d$FI302/10
fhs1_ex0_16d$FI306_cm  <- fhs1_ex0_16d$FI306/10
fhs1_ex0_16d$rwt <- fhs1_ex0_16d$FI311/100
fhs1_ex0_16d$FI15_cm <- fhs1_ex0_16d$FI15*2.54
fhs1_ex0_16d$FI15_m <- fhs1_ex0_16d$FI15_cm/100
fhs1_ex0_16d$FI14_kg <- fhs1_ex0_16d$FI14/2.2

fhs1_ex0_16d$BSA2 <- 0.20247*((fhs1_ex0_16d$FI14_kg)^0.425)*((fhs1_ex0_16d$FI15_m)^0.725)

fhs1_ex0_16d <- 
  as.data.table(
    calc_hypertrophy_type(
      df=as.data.frame(fhs1_ex0_16d),
      sex="SEX",
      male="1",
      female="2",
      lvedd="FI306_cm",
      ivsd="FI297_cm",
      lvpwtd="FI302_cm",
      height="FI15_cm",
      weight="FI14_kg"))


fhs1_melt_ex0_16d <- melt(fhs1_ex0_16d,id.vars=c("PID","IDTYPE","visit"),na.rm=T,factorsAsStrings=T)
fhs1_melt_ex0_16d$form <- "fhs1_ex0_16"

rm(fhs1_ex0_16d)


# ===============================================================#
####                    FHS original, Exam 17                 ####
# ===============================================================#

fhs1_ex0_17d <-fread('~/Dropbox/BioLINCC files/Framingham cohort/datasets/CSV/EX0_17D.csv',na.strings=c("NA","","NULL")) # Exam 17
fhs1_ex0_17d$visit <- 17

fhs1_ex0_17d$drinks_wk <- fhs1_ex0_17d$FJ59+fhs1_ex0_17d$FJ60+fhs1_ex0_17d$FJ61


####### FRAIL score

## Fatigue, Resistance, Ambulation encoded in 'criteriaset'

fhs1_ex0_17d$fatigue_frail[fhs1_ex0_17d$FJ191==1|fhs1_ex0_17d$FJ192==1] <- 1
fhs1_ex0_17d$fatigue_frail[fhs1_ex0_17d$FJ191 %in% c(0,2)|fhs1_ex0_17d$FJ192 %in% c(0,2)] <- 0

## Resistance

fhs1_ex0_17d$resistance_frail[fhs1_ex0_17d$FJ175==1] <- 0
fhs1_ex0_17d$resistance_frail[fhs1_ex0_17d$FJ175==0] <- 1

## Ambulate

fhs1_ex0_17d$ambulate_frail[fhs1_ex0_17d$FJ176==1] <- 0
fhs1_ex0_17d$ambulate_frail[fhs1_ex0_17d$FJ176==0] <- 1

# FRAIL Illness score counts presence of HTN, DM, cancer, chronic lung disease, MI, CHD, angina, asthma, arthritis, stroke, and ckd
# Scores for 0-4 conditions, score = 0, for ≥ 5, score=1

fhs1_ex0_17d <- merge(fhs1_ex0_17d,fhs_dates[,c("PID","IDTYPE","DATE17")],by=c("PID","IDTYPE"),all.x=T)
fhs1_ex0_17d <- merge(fhs1_ex0_17d,fhs_outcomes,by=c("PID","IDTYPE"),all.x=T)
fhs1_ex0_17d <- merge(fhs1_ex0_17d,fhs_survca_first_any[,c("PID","IDTYPE","D_DATE")],by=c("PID","IDTYPE"),all.x=T)
fhs1_ex0_17d <- merge(fhs1_ex0_17d,fhs1_wkthru[,c("PID","IDTYPE","WGT16","WGT17")],by=c("PID","IDTYPE"),all.x=T)

# DM

fhs1_ex0_17d <- merge(fhs1_ex0_17d,
                      fhs_diab[,c("PID","BG200_HX_DIAB17")],
                      by="PID",
                      all.x=T)

fhs1_ex0_17d$dm_frail <- fhs1_ex0_17d$BG200_HX_DIAB17

# HTN

fhs1_ex0_17d$htn_frail[fhs1_ex0_17d$FJ318==0] <- 0
fhs1_ex0_17d$htn_frail[fhs1_ex0_17d$FJ318==1] <- 1

###------------###

# COPD (chronic lung disease)

fhs1_ex0_17d$copd_frail[fhs1_ex0_17d$FJ348==0] <- 0
fhs1_ex0_17d$copd_frail[fhs1_ex0_17d$FJ348==1] <- 1


###------------###


# Asthma

fhs1_ex0_17d$asthma_frail[fhs1_ex0_17d$FJ71==0] <- 0
fhs1_ex0_17d$asthma_frail[fhs1_ex0_17d$FJ71==1] <- 1

###------------###

# DJD (arthritis)

fhs1_ex0_17d$djd_frail[fhs1_ex0_17d$FJ351==0] <- 0
fhs1_ex0_17d$djd_frail[fhs1_ex0_17d$FJ351==1] <- 1


###------------###

# CKD (renal disease)


fhs1_ex0_17d$ckd_frail[fhs1_ex0_17d$FJ346==0] <- 0
fhs1_ex0_17d$ckd_frail[fhs1_ex0_17d$FJ346==1] <- 1

###------------###

# CHF

fhs1_ex0_17d$chf_frail[fhs1_ex0_17d$hfhosp_status==0|(fhs1_ex0_17d$hfhosp_status==1&fhs1_ex0_17d$DATE17 < fhs1_ex0_17d$hfhosp_dt)] <- 0
fhs1_ex0_17d$chf_frail[fhs1_ex0_17d$hfhosp_status==1&fhs1_ex0_17d$DATE17 >= fhs1_ex0_17d$hfhosp_dt] <- 1

###------------###

# CAD/MI

fhs1_ex0_17d$chd_frail[fhs1_ex0_17d$cadhosp_status==0|(fhs1_ex0_17d$cadhosp_status==1&fhs1_ex0_17d$DATE17 < fhs1_ex0_17d$cadhosp_dt)] <- 0
fhs1_ex0_17d$chd_frail[fhs1_ex0_17d$cadhosp_status==1&fhs1_ex0_17d$DATE17 >= fhs1_ex0_17d$cadhosp_dt] <- 1

###------------###


# Stroke

fhs1_ex0_17d$stroke_frail[fhs1_ex0_17d$cvahosp_status==0|(fhs1_ex0_17d$cvahosp_status==1&fhs1_ex0_17d$DATE17 < fhs1_ex0_17d$cvahosp_dt)] <- 0
fhs1_ex0_17d$stroke_frail[fhs1_ex0_17d$cvahosp_status==1&fhs1_ex0_17d$cvahosp_dt <= fhs1_ex0_17d$DATE17] <- 1


###------------###

# Cancer

fhs1_ex0_17d$cancer_frail <- 0
fhs1_ex0_17d$cancer_frail[fhs1_ex0_17d$D_DATE <= fhs1_ex0_17d$DATE17] <- 1

###------------###

# Compile chronic illness

fhs1_ex0_17d$conditions_frail <- 
  fhs1_ex0_17d$dm_frail+
  fhs1_ex0_17d$htn_frail+
  fhs1_ex0_17d$copd_frail+
  fhs1_ex0_17d$asthma_frail+
  fhs1_ex0_17d$djd_frail+
  fhs1_ex0_17d$ckd_frail+
  fhs1_ex0_17d$chf_frail+
  fhs1_ex0_17d$chd_frail+
  fhs1_ex0_17d$stroke_frail+
  fhs1_ex0_17d$cancer_frail

fhs1_ex0_17d$illness_frail[fhs1_ex0_17d$conditions_frail <= 4] <- 0
fhs1_ex0_17d$illness_frail[fhs1_ex0_17d$conditions_frail > 4] <- 1

fhs1_ex0_17d$wtloss_frail[fhs1_ex0_17d$WGT17-fhs1_ex0_17d$WGT16 <= -15] <- 1
fhs1_ex0_17d$wtloss_frail[fhs1_ex0_17d$WGT17-fhs1_ex0_17d$WGT16 > -15] <- 0

fhs1_ex0_17d$total_frail <- 
  fhs1_ex0_17d$fatigue_frail+
  fhs1_ex0_17d$resistance_frail+
  fhs1_ex0_17d$ambulate_frail+
  fhs1_ex0_17d$illness_frail+
  fhs1_ex0_17d$wtloss_frail

#### ADL mod

fhs1_ex0_17d$FJ177[fhs1_ex0_17d$FJ177 > 3] <- 0
fhs1_ex0_17d$FJ178[fhs1_ex0_17d$FJ178 > 3] <- 0
fhs1_ex0_17d$FJ179[fhs1_ex0_17d$FJ179 > 3] <- 0
fhs1_ex0_17d$FJ180[fhs1_ex0_17d$FJ180 > 3] <- 0
fhs1_ex0_17d$FJ181[fhs1_ex0_17d$FJ181 > 3] <- 0
fhs1_ex0_17d$FJ182[fhs1_ex0_17d$FJ182 > 3] <- 0


fhs1_melt_ex0_17d <- melt(fhs1_ex0_17d,id.vars=c("PID","IDTYPE","visit"),na.rm=T,factorsAsStrings=T)
fhs1_melt_ex0_17d$form <- "fhs1_ex0_17"

rm(fhs1_ex0_17d)

# ==============================================================#
####                    FHS original, Exam 18                ####
# ==============================================================#

fhs1_ex0_18d <-fread('~/Dropbox/BioLINCC files/Framingham cohort/datasets/CSV/EX0_18D_V2.csv',na.strings=c("NA","","NULL")) # Exam 18
fhs1_ex0_18d$visit <- 18
fhs1_melt_ex0_18d <- melt(fhs1_ex0_18d,id.vars=c("PID","IDTYPE","visit"),na.rm=T,factorsAsStrings=T)
fhs1_melt_ex0_18d$form <- "fhs1_ex0_18"

rm(fhs1_ex0_18d)


# ==============================================================#
####                    FHS original, Exam 19                ####
# ==============================================================#

fhs1_ex0_19d <-fread('~/Dropbox/BioLINCC files/Framingham cohort/datasets/CSV/EX0_19D.csv',na.strings=c("NA","","NULL")) # Exam 19
fhs1_ex0_19d$visit <- 19

fhs1_ex0_19d$fhs_pai <- calc_framingham_pai(as.data.frame(fhs1_ex0_19d),
                                            slp_hrs="FL089",
                                            sed_hrs="FL090",
                                            slgt_hrs="FL091",
                                            mod_hrs="FL092",
                                            hvy_hrs="FL093")


fhs1_melt_ex0_19d <- melt(fhs1_ex0_19d,id.vars=c("PID","IDTYPE","visit"),na.rm=T,factorsAsStrings=T)
fhs1_melt_ex0_19d$form <- "fhs1_ex0_19"

rm(fhs1_ex0_19d)


# ===============================================================#
####                  FHS original, Exam 20                   ####
# ===============================================================#

fhs1_ex0_20d <-fread('~/Dropbox/BioLINCC files/Framingham cohort/datasets/CSV/EX0_20D_V1.csv',na.strings=c("NA","","NULL")) # Exam 20
fhs1_ex0_20d$visit <- 20

fhs1_ex0_20d$fhs_pai <- 
  calc_framingham_pai(
    as.data.frame(fhs1_ex0_20d),
    slp_hrs="FM94",
    sed_hrs="FM95",
    slgt_hrs="FM96",
    mod_hrs="FM97",
    hvy_hrs="FM98")

fhs1_meno <- 
  merge(fhs1_meno,
        fhs1_ex0_20d[,c("PID","FM201")],
        all.x=T)


fhs1_melt_ex0_20d <- melt(fhs1_ex0_20d,id.vars=c("PID","IDTYPE","visit"),na.rm=T,factorsAsStrings=T)
fhs1_melt_ex0_20d$form <- "fhs1_ex0_20"

rm(fhs1_ex0_20d)


# ==============================================================#
#                     FHS original, Lab exam 20                 #
# ==============================================================#

fhs1_l_fhslab1_ex20 <-fread('~/Dropbox/BioLINCC files/Framingham cohort/datasets/CSV/L_FHSLAB1_EX20_0494D.csv',na.strings=c("NA","","NULL")) # Standard labs
fhs1_l_fhslab1_ex20$visit <- 20

fhs1_ex20_demo <- 
  fhs1_wkthru[,c("PID",
                 "IDTYPE",
                 "SEX",
                 "AGE20")]

fhs1_l_fhslab1_ex20 <- 
  merge(fhs1_l_fhslab1_ex20,
        fhs1_ex20_demo,
        by=c("PID","IDTYPE"),
        all.x=T)

fhs1_l_fhslab1_ex20$race <- 1

fhs1_l_fhslab1_ex20$gfr_mdrd <- calc_MDRD4(dat=as.data.frame(fhs1_l_fhslab1_ex20),
                                           cr="CREAT",
                                           age="AGE20",
                                           sex="SEX",
                                           race="race")

fhs1_melt_l_fhslab1_ex20 <- melt(fhs1_l_fhslab1_ex20,id.vars=c("PID","IDTYPE","visit"),na.rm=T,factorsAsStrings=T)
fhs1_melt_l_fhslab1_ex20$form <- "fhs1_fhslab1"

rm(fhs1_l_fhslab1_ex20)


#================================================================##
#             FHS original, echo with doppler, exam 20            #
#================================================================##

fhs1_t_echo_ex20 <- fread('~/Dropbox/BioLINCC files/Framingham cohort/datasets/CSV/T_ECHO_EX20_0_0576D.csv',na.strings=c("NA","","NULL")) # Echo dimensions
fhs1_t_echo_ex20$visit <- 20

fhs1_t_echo_ex20 <- merge(fhs1_t_echo_ex20,
                          fhs1_wkthru[,c("PID","IDTYPE","SEX","HGT1","WGT20")])

fhs1_t_echo_ex20$height_cm <- fhs1_t_echo_ex20$HGT1*2.54
fhs1_t_echo_ex20$weight_kg <- fhs1_t_echo_ex20$WGT20/2.2

fhs1_t_echo_ex20 <- 
  as.data.table(
    calc_hypertrophy_type(
      df=as.data.frame(fhs1_t_echo_ex20),
      sex="SEX",
      male="1",
      female="2",
      lvedd="LVDD20",
      ivsd="IVSD20",
      lvpwtd="LVPD20",
      height="height_cm",
      weight="weight_kg"))


fhs1_melt_t_echo_ex20 <- melt(fhs1_t_echo_ex20,id.vars=c("PID","IDTYPE","visit"),na.rm=T,factorsAsStrings=T)
fhs1_melt_t_echo_ex20$form <- "fhs1_echo"


fhs1_t_echodop_ex20 <- fread('~/Dropbox/BioLINCC files/Framingham cohort/datasets/CSV/T_ECHODOP_EX20_0_0886D.csv',na.strings=c("NA","","NULL")) # Echo doppler
fhs1_t_echodop_ex20$visit <- 20
fhs1_t_echodop_ex20$DEC_MM_msec <- fhs1_t_echodop_ex20$DEC_MM*1000

fhs1_melt_t_echodop_ex20 <- melt(fhs1_t_echodop_ex20,id.vars=c("PID","IDTYPE","visit"),na.rm=T,factorsAsStrings=T)

fhs1_melt_t_echodop_ex20$form <- "fhs1_echodop"


rm(fhs1_t_echo_ex20,
   fhs1_t_echodop_ex20)

# ==================================================================#
#              FHS original, HR variability, exam 20                #
# ==================================================================#

fhs1_t_hrv_ex20 <- fread('~/Dropbox/BioLINCC files/Framingham cohort/datasets/CSV/T_HRV_1987_M_0118D.csv',na.strings=c("NA","","NULL")) # Heart rate variability
fhs1_t_hrv_ex20$visit <- 20
fhs1_melt_t_hrv_ex20 <- melt(fhs1_t_hrv_ex20,id.vars=c("PID","IDTYPE","visit"),na.rm=T,factorsAsStrings=T)
fhs1_melt_t_hrv_ex20$form <- "fhs1_hrv"

rm(fhs1_t_hrv_ex20)


#=============================================#
#     FHS original food frequency exam 20     #
#=============================================#

fhs1_vr_ffreq_ex20 <- fread('~/Dropbox/BioLINCC files/Framingham cohort/datasets/CSV/VR_FFREQ_EX20_0_0572D_v1.csv',na.strings=c("NA","","NULL")) # Food intake exam 20
fhs1_vr_ffreq_ex20$visit <- 20
fhs1_vr_ffreq_ex20$SCORE33_wk <- fhs1_vr_ffreq_ex20$SCORE33/14*7
fhs1_melt_vr_ffreq_ex20 <- melt(fhs1_vr_ffreq_ex20,id.vars=c("PID","IDTYPE","visit"),na.rm=T,factorsAsStrings=T)
fhs1_melt_vr_ffreq_ex20$form <- "fhs1_vr_ffreq"

rm(fhs1_vr_ffreq_ex20)


# ==============================================================#
####                    FHS original, Exam 21                ####
# ==============================================================#

fhs1_ex0_21d <-fread('~/Dropbox/BioLINCC files/Framingham cohort/datasets/CSV/EX0_21D_V2.csv',na.strings=c("NA","","NULL")) # Exam 21
fhs1_ex0_21d$visit <- 21

fhs1_ex0_21d$FN150[!between(fhs1_ex0_21d$FN150,23,87)] <- NA

fhs1_meno <- 
  merge(fhs1_meno,
        fhs1_ex0_21d[,c("PID","FN151")],
        all.x=T)

fhs1_melt_ex0_21d <- melt(fhs1_ex0_21d,id.vars=c("PID","IDTYPE","visit"),na.rm=T,factorsAsStrings=T)
fhs1_melt_ex0_21d$form <- "fhs1_ex0_21"

rm(fhs1_ex0_21d)


#=============================================#
#     FHS original food frequency exam 21     #
#=============================================#

fhs1_vr_ffreq_ex21 <- fread('~/Dropbox/BioLINCC files/Framingham cohort/datasets/CSV/VR_FFREQ_EX21_0_0573D_v1.csv',na.strings=c("NA","","NULL")) # Food intake exam 21
fhs1_vr_ffreq_ex21$visit <- 21
fhs1_vr_ffreq_ex21$SCORE33_wk <- fhs1_vr_ffreq_ex21$SCORE33/14*7
fhs1_melt_vr_ffreq_ex21 <- melt(fhs1_vr_ffreq_ex21,id.vars=c("PID","IDTYPE","visit"),na.rm=T,factorsAsStrings=T)
fhs1_melt_vr_ffreq_ex21$form <- "fhs1_vr_ffreq"

rm(fhs1_vr_ffreq_ex21)


# ==============================================================#
####                    FHS original, Exam 22                ####
# ==============================================================#

fhs1_ex0_22d <-fread('~/Dropbox/BioLINCC files/Framingham cohort/datasets/CSV/EX0_22D_V3.csv',na.strings=c("NA","","NULL")) # Exam 21
fhs1_ex0_22d$visit <- 22

fhs1_meno <- 
  merge(fhs1_meno,
        fhs1_ex0_22d[,c("PID","FO149")],
        all.x=T)

fhs1_meno$ovaries_removed <-
  apply(fhs1_meno[,c("AM4","FM201","FN151","FO149")],
        MARGIN=1,
        FUN=function(x) max(x, na.rm=T))

fhs1_meno$ovaries_removed[fhs1_meno$ovaries_removed %in% c("-Inf","7")] <- NA

fhs1_ex0_22d$CESD20_missing <- rowSums(is.na(fhs1_ex0_22d[,370:389]))
fhs1_ex0_22d$CESD20 <- rowSums(fhs1_ex0_22d[,370:389])
fhs1_ex0_22d$CESD20[fhs1_ex0_22d$CESD20_missing >= 4] <- NA

fhs1_ex0_22d$barthel_hygiene <- 0

fhs1_ex0_22d$FO148[!between(fhs1_ex0_22d$FO148,20,87)]<- NA
fhs1_ex0_22d$FO150[!between(fhs1_ex0_22d$FO150,0,9)]<- NA


fhs1_melt_ex0_22d <- melt(fhs1_ex0_22d,id.vars=c("PID","IDTYPE","visit"),na.rm=T,factorsAsStrings=T)
fhs1_melt_ex0_22d$form <- "fhs1_ex0_22"

fhs1_melt_meno <- melt(fhs1_meno,id.vars=c("PID","IDTYPE","visit"),na.rm=T,factorsAsStrings=T)
fhs1_melt_meno$form <- "fhs1_meno"


rm(fhs1_ex0_22d)

#=============================================#
#    FHS original food frequency exam 22     #
#=============================================#

fhs1_vr_ffreq_ex22 <- fread('~/Dropbox/BioLINCC files/Framingham cohort/datasets/CSV/VR_FFREQ_EX22_0_0574D_v1.csv',na.strings=c("NA","","NULL")) # Food intake exam 22
fhs1_vr_ffreq_ex22$visit <- 22
fhs1_vr_ffreq_ex22$SCORE33_wk <- fhs1_vr_ffreq_ex22$SCORE33/14*7
fhs1_melt_vr_ffreq_ex22 <- melt(fhs1_vr_ffreq_ex22,id.vars=c("PID","IDTYPE","visit"),na.rm=T,factorsAsStrings=T)
fhs1_melt_vr_ffreq_ex22$form <- "fhs1_vr_ffreq"

rm(fhs1_vr_ffreq_ex22)


# ==============================================================#
####                   FHS original, Exam 23                 ####
# ==============================================================#

fhs1_ex0_23d <-fread('~/Dropbox/BioLINCC files/Framingham cohort/datasets/CSV/EX0_23D_V3.csv',na.strings=c("NA","","NULL")) # Exam 23
fhs1_ex0_23d$visit <- 23

fhs1_ex0_23d$CESD20_missing <- rowSums(is.na(fhs1_ex0_23d[,390:409]))
fhs1_ex0_23d$CESD20 <- rowSums(fhs1_ex0_23d[,390:409])
fhs1_ex0_23d$CESD20[fhs1_ex0_23d$CESD20_missing >= 4] <- NA

fhs1_ex0_23d$barthel_hygiene <- 0


fhs1_melt_ex0_23d <- melt(fhs1_ex0_23d,id.vars=c("PID","IDTYPE","visit"),na.rm=T,factorsAsStrings=T)
fhs1_melt_ex0_23d$form <- "fhs1_ex0_23"

rm(fhs1_ex0_23d)


# ==============================================================#
####                    FHS original, Exam 24                ####
# ==============================================================#

fhs1_ex0_24d <-fread('~/Dropbox/BioLINCC files/Framingham cohort/datasets/CSV/EX0_24D_V2.csv',na.strings=c("NA","","NULL")) # Exam 24
fhs1_ex0_24d$visit <- 24

fhs1_ex0_24d$barthel_hygiene <- 0


fhs1_melt_ex0_24d <- melt(fhs1_ex0_24d,id.vars=c("PID","IDTYPE","visit"),na.rm=T,factorsAsStrings=T)
fhs1_melt_ex0_24d$form <- "fhs1_ex0_24"

rm(fhs1_ex0_24d)


# ==============================================================#
####                 FHS original, Exam 25                   ####
# ==============================================================#

fhs1_ex0_25d <-fread('~/Dropbox/BioLINCC files/Framingham cohort/datasets/CSV/EX0_25D_V1.csv',na.strings=c("NA","","NULL")) # Exam 25
fhs1_ex0_25d$visit <- 25

fhs1_ex0_25d$CESD20_missing <- rowSums(is.na(fhs1_ex0_25d[,89:108]))
fhs1_ex0_25d$CESD20 <- rowSums(fhs1_ex0_25d[,89:108])
fhs1_ex0_25d$CESD20[fhs1_ex0_25d$CESD20_missing >= 4] <- NA

fhs1_ex0_25d$barthel_hygiene <- 0
fhs1_ex0_25d$essi_chores <- 0

fhs1_melt_ex0_25d <- melt(fhs1_ex0_25d,id.vars=c("PID","IDTYPE","visit"),na.rm=T,factorsAsStrings=T)
fhs1_melt_ex0_25d$form <- "fhs1_ex0_25"

rm(fhs1_ex0_25d)


# ==============================================================#
####                    FHS original, Exam 26                ####
# ==============================================================#

## Uses method described here https://www.ncbi.nlm.nih.gov/pmc/articles/PMC5005887/

fhs1_ex0_26d <-fread('~/Dropbox/BioLINCC files/Framingham cohort/datasets/CSV/EX0_26D_V1.csv',na.strings=c("NA","","NULL")) # Exam 26
fhs1_ex0_26d$visit <- 26
fhs1_ex0_26d[fhs1_ex0_26d==99] <- NA

fhs1_ex0_26d$CESD20_missing <- rowSums(is.na(fhs1_ex0_26d[,107:126]))
fhs1_ex0_26d$CESD20 <- rowSums(fhs1_ex0_26d[,107:126])
fhs1_ex0_26d$CESD20[fhs1_ex0_26d$CESD20_missing >= 4] <- NA

fhs1_ex0_26d$barthel_hygiene <- 0
fhs1_ex0_26d$essi_chores <- 0


fhs1_ex0_26d <- 
  merge(fhs1_ex0_26d,fhs1_wkthru[,c("PID",
                                    "SEX",
                                    "AGE26",
                                    "HGT1",
                                    "BMIC26",
                                    "DATE26",
                                    "WGT26",
                                    "WGT25",
                                    "DMRX26",
                                    "HRX26",
                                    "CREAT26")],
        by="PID",
        all.x=T)


fhs1_ex0_26d$essi_chores <- 0


##### Walking speed (sec/15 ft)

fhs1_ex0_26d$walktime_1 <- fhs1_ex0_26d$FS213+fhs1_ex0_26d$FS214/100
fhs1_ex0_26d$walktime_2 <- fhs1_ex0_26d$FS216+fhs1_ex0_26d$FS217/100

fhs1_ex0_26d$walktime_avg <- round(apply(fhs1_ex0_26d[,c("walktime_1","walktime_2")],MARGIN=1,mean,na.rm=T),3)
fhs1_ex0_26d$walktime_avg[fhs1_ex0_26d$walktime_avg=="NaN"] <- NA
fhs1_ex0_26d$walktime_min <- round(apply(fhs1_ex0_26d[,c("walktime_1","walktime_2")],MARGIN=1,min,na.rm=T),3)
fhs1_ex0_26d$walktime_min[fhs1_ex0_26d$walktime_min=="-Inf"] <- NA


########### Grip (kg)

# Produces highest average by hand (assumes dominant hand is stronger)

fhs1_ex0_26d$grip_avg_right <- round(apply(fhs1_ex0_26d[,141:143],MARGIN=1,mean),1)
fhs1_ex0_26d$grip_avg_left <- round(apply(fhs1_ex0_26d[,144:146],MARGIN=1,mean),1)

fhs1_ex0_26d$grip_max <- 
  round(
    apply(
      fhs1_ex0_26d[,c("grip_avg_right","grip_avg_left")],
      MARGIN=1,max),1)


##### Exhaustion

fhs1_ex0_26d$exhaustion_fried[fhs1_ex0_26d$FS163==3|fhs1_ex0_26d$FS176==3] <- 1
fhs1_ex0_26d$exhaustion_fried[fhs1_ex0_26d$FS163<3&fhs1_ex0_26d$FS176<3] <- 0

fhs1_ex0_26d$exhaustion_fried[fhs1_ex0_26d$FS163==3|fhs1_ex0_26d$FS176==3] <- 1
fhs1_ex0_26d$exhaustion_fried[fhs1_ex0_26d$FS163<3&fhs1_ex0_26d$FS176<3] <- 0


#### (Weight) Loss

fhs1_ex0_26d$wtloss_fried[(fhs1_ex0_26d$WGT26-fhs1_ex0_26d$WGT25) >= -10] <- 0
fhs1_ex0_26d$wtloss_fried[(fhs1_ex0_26d$WGT26-fhs1_ex0_26d$WGT25) < -10] <- 1


######### FRAIL Score


##### Fatigue

fhs1_ex0_26d$fatigue_frail[fhs1_ex0_26d$FS163 >=2|fhs1_ex0_26d$FS176 >= 2] <- 1
fhs1_ex0_26d$fatigue_frail[fhs1_ex0_26d$FS163<2&fhs1_ex0_26d$FS176<2] <- 0


#### Resistance

fhs1_ex0_26d$resistance_frail[fhs1_ex0_26d$FS072==0] <- 0
fhs1_ex0_26d$resistance_frail[fhs1_ex0_26d$FS072 %in% c(1,2,3)] <- 1

#### Ambulation

fhs1_ex0_26d$ambulate_frail[fhs1_ex0_26d$FS071==1] <- 0
fhs1_ex0_26d$ambulate_frail[fhs1_ex0_26d$FS071 %in% c(0,2)] <- 1


# FRAIL Illness score counts presence of HTN, DM, cancer, chronic lung disease, MI, CHD, angina, asthma, arthritis, stroke, and ckd
# Scores for 0-4 conditions, score = 0, for ≥ 5, score=1


# COPD (chronic lung disease)

fhs1_ex0_26d$copd_frail[fhs1_ex0_26d$FS522==2] <- 0
fhs1_ex0_26d$copd_frail[fhs1_ex0_26d$FS522==1] <- 1
fhs1_ex0_26d$copd_frail[fhs1_ex0_26d$FS522==0] <- 0

# asthma

fhs1_ex0_26d$asthma_frail[fhs1_ex0_26d$FS525==2] <- 0
fhs1_ex0_26d$asthma_frail[fhs1_ex0_26d$FS525==1] <- 1
fhs1_ex0_26d$asthma_frail[fhs1_ex0_26d$FS525==0] <- 0

# DJD (arthritis)

fhs1_ex0_26d$djd_frail[fhs1_ex0_26d$FS528==2] <- 0
fhs1_ex0_26d$djd_frail[fhs1_ex0_26d$FS528==1] <- 1
fhs1_ex0_26d$djd_frail[fhs1_ex0_26d$FS528==0] <- 0

# CKD (renal disease)

fhs1_ex0_26d$RACE <- 1
fhs1_ex0_26d$crcl <- calc_MDRD4(as.data.frame(fhs1_ex0_26d),cr="CREAT26",age="AGE26")

fhs1_ex0_26d$ckd_frail[fhs1_ex0_26d$FS521==2] <- 0
fhs1_ex0_26d$ckd_frail[fhs1_ex0_26d$FS521==1|fhs1_ex0_26d$crcl < 60] <- 1
fhs1_ex0_26d$ckd_frail[fhs1_ex0_26d$FS521==0] <- 0

# CHF/CAD

fhs1_ex0_26d <- merge(fhs1_ex0_26d,fhs_outcomes,by=c("PID","IDTYPE"))

fhs1_ex0_26d$chf_frail <- 0
fhs1_ex0_26d$chf_frail[fhs1_ex0_26d$hfhosp_status==1&fhs1_ex0_26d$hfhosp_dt <= fhs1_ex0_26d$DATE26] <- 1

fhs1_ex0_26d$chd_frail <- 0
fhs1_ex0_26d$chd_frail[fhs1_ex0_26d$cadhosp_status==1&fhs1_ex0_26d$cadhosp_dt <= fhs1_ex0_26d$DATE26] <- 1

# Stroke

fhs1_ex0_26d$stroke_frail <- 0
fhs1_ex0_26d$stroke_frail[fhs1_ex0_26d$cvahosp_status==1&fhs1_ex0_26d$cvahosp_dt <= fhs1_ex0_26d$DATE26] <- 1

# Cancer

fhs1_ex0_26d <- merge(fhs1_ex0_26d,
                      fhs_survca_first_any[,c("PID","IDTYPE","D_DATE")],by=c("PID","IDTYPE"),all.x=T)

fhs1_ex0_26d$cancer_frail <- 0
fhs1_ex0_26d$cancer_frail[fhs1_ex0_26d$D_DATE <= fhs1_ex0_26d$DATE26] <- 1

fhs1_ex0_26d$conditions_frail <- fhs1_ex0_26d$HRX26+
  fhs1_ex0_26d$DMRX26+
  fhs1_ex0_26d$copd_frail+
  fhs1_ex0_26d$asthma_frail+
  fhs1_ex0_26d$djd_frail+
  fhs1_ex0_26d$ckd_frail+
  fhs1_ex0_26d$chf_frail+
  fhs1_ex0_26d$chd_frail+
  fhs1_ex0_26d$stroke_frail+
  fhs1_ex0_26d$cancer_frail

fhs1_ex0_26d$illness_frail[fhs1_ex0_26d$conditions_frail <= 4] <- 0
fhs1_ex0_26d$illness_frail[fhs1_ex0_26d$conditions_frail > 4] <- 1


#### (Weight) Loss

fhs1_ex0_26d$wgt26_25_delta <- fhs1_ex0_26d$WGT26-fhs1_ex0_26d$WGT25


fhs1_ex0_26d$wtloss_frail[(fhs1_ex0_26d$WGT26-fhs1_ex0_26d$WGT25) > -10] <- 0
fhs1_ex0_26d$wtloss_frail[(fhs1_ex0_26d$WGT26-fhs1_ex0_26d$WGT25) <= -10] <- 1


#### Calculate FRAIL score

fhs1_ex0_26d$total_frail <- fhs1_ex0_26d$fatigue_frail+
  fhs1_ex0_26d$resistance_frail+
  fhs1_ex0_26d$ambulate_frail+
  fhs1_ex0_26d$illness_frail+
  fhs1_ex0_26d$wtloss_frail

fhs1_ex0_26d <- fhs1_ex0_26d[-c(354,356),]

fhs1_melt_ex0_26d <- melt(fhs1_ex0_26d,id.vars=c("PID","IDTYPE","visit"),na.rm=T,factorsAsStrings=T)
fhs1_melt_ex0_26d$form <- "fhs1_ex0_26"


rm(fhs1_ex0_26d)


# ==============================================================#
####                  FHS original, Exam 27                  ####
# ==============================================================#

fhs1_ex0_27d <-fread('~/Dropbox/BioLINCC files/Framingham cohort/datasets/CSV/E_EXAM_EX27_0_0075D.csv',na.strings=c("NA","","NULL")) # Exam 27
fhs1_ex0_27d$visit <- 27
fhs1_ex0_27d <- 
  merge(fhs1_ex0_27d,fhs1_wkthru[,c("PID",
                                    "SEX",
                                    "HGT1",
                                    "DATE27",
                                    "BMIC27",
                                    "WGT27",
                                    "WGT26",
                                    "DMRX27",
                                    "HRX27",
                                    "CREAT27",
                                    "AGE27")],
        by="PID",all.x=T)


fhs1_ex0_27d$essi_chores <- 0

## Walking speed

fhs1_ex0_27d$walktime_1 <- fhs1_ex0_27d$FT225+fhs1_ex0_27d$FT226/100
fhs1_ex0_27d$walktime_2 <- fhs1_ex0_27d$FT229+fhs1_ex0_27d$FT230/100

fhs1_ex0_27d$walktime_avg <- round(apply(fhs1_ex0_27d[,c("walktime_1","walktime_2")],MARGIN=1,mean,na.rm=T),3)
fhs1_ex0_27d$walktime_avg[fhs1_ex0_27d$walktime_avg=="NaN"] <- NA
fhs1_ex0_27d$walktime_min <- round(apply(fhs1_ex0_27d[,c("walktime_1","walktime_2")],MARGIN=1,min,na.rm=T),3)
fhs1_ex0_27d$walktime_min[fhs1_ex0_27d$walktime_min=="-Inf"] <- NA


########### Grip (kg)

# Produces highest average by hand (assumes dominant hand is stronger)

fhs1_ex0_27d$grip_avg_right <- round(apply(fhs1_ex0_27d[,99:101],MARGIN=1,mean),1)
fhs1_ex0_27d$grip_avg_left <- round(apply(fhs1_ex0_27d[,102:104],MARGIN=1,mean),1)

fhs1_ex0_27d$grip_max <- round(apply(fhs1_ex0_27d[,c("grip_avg_right","grip_avg_left")],MARGIN=1,max),1)


#### Exhaustion

fhs1_ex0_27d$exhaustion_fried[fhs1_ex0_27d$FT032==3|fhs1_ex0_27d$FT045==3] <- 1
fhs1_ex0_27d$exhaustion_fried[fhs1_ex0_27d$FT032<3&fhs1_ex0_27d$FT045<3] <- 0


#### (Weight) Loss

fhs1_ex0_27d$wtloss_fried[(fhs1_ex0_27d$WGT27-fhs1_ex0_27d$WGT26) <= - 10] <- 1
fhs1_ex0_27d$wtloss_fried[(fhs1_ex0_27d$WGT27-fhs1_ex0_27d$WGT26) > 10] <- 0


######### FRAIL Score

#### Fatigue

fhs1_ex0_27d$fatigue_frail[fhs1_ex0_27d$FT032==3|fhs1_ex0_27d$FT045==3] <- 1
fhs1_ex0_27d$fatigue_frail[fhs1_ex0_27d$FT032<3&fhs1_ex0_27d$FT045<3] <- 0

#### Resistance

fhs1_ex0_27d$resistance_frail[fhs1_ex0_27d$FT095==0] <- 0
fhs1_ex0_27d$resistance_frail[fhs1_ex0_27d$FT095 %in% c(1,2,3)] <- 1

#### Ambulation

fhs1_ex0_27d$ambulate_frail[fhs1_ex0_27d$FT136==1] <- 0
fhs1_ex0_27d$ambulate_frail[fhs1_ex0_27d$FT136 %in% c(0,2)] <- 1


# FRAIL Illness score counts presence of HTN, DM, cancer, chronic lung disease, MI, CHD, angina, asthma, arthritis, stroke, and ckd
# Scores for 0-4 conditions, score = 0, for ≥ 5, score=1


# COPD (chronic lung disease)

fhs1_ex0_27d$copd_frail[fhs1_ex0_27d$FT527==2] <- 0
fhs1_ex0_27d$copd_frail[fhs1_ex0_27d$FT527==1] <- 1
fhs1_ex0_27d$copd_frail[fhs1_ex0_27d$FT527==0] <- 0

# asthma

fhs1_ex0_27d$asthma_frail[fhs1_ex0_27d$FT530==2] <- 0
fhs1_ex0_27d$asthma_frail[fhs1_ex0_27d$FT530==1] <- 1
fhs1_ex0_27d$asthma_frail[fhs1_ex0_27d$FT530==0] <- 0

# DJD (arthritis)

fhs1_ex0_27d$djd_frail[fhs1_ex0_27d$FT533==2] <- 0
fhs1_ex0_27d$djd_frail[fhs1_ex0_27d$FT533==1] <- 1
fhs1_ex0_27d$djd_frail[fhs1_ex0_27d$FT533==0] <- 0

# CKD (renal disease)

fhs1_ex0_27d$RACE <- 1
fhs1_ex0_27d$crcl <- calc_MDRD4(as.data.frame(fhs1_ex0_27d),
                                cr="CREAT27",
                                age="AGE27")


fhs1_ex0_27d$ckd_frail[fhs1_ex0_27d$FT526==2] <- 0
fhs1_ex0_27d$ckd_frail[fhs1_ex0_27d$FT526==1|fhs1_ex0_27d$crcl < 60] <- 1
fhs1_ex0_27d$ckd_frail[fhs1_ex0_27d$FT526==0] <- 0

# CHF/MI

fhs1_ex0_27d <- merge(fhs1_ex0_27d,fhs_outcomes,by=c("PID","IDTYPE"))

fhs1_ex0_27d$chf_frail <- 0
fhs1_ex0_27d$chf_frail[fhs1_ex0_27d$hfhosp_status==1&fhs1_ex0_27d$hfhosp_dt <= fhs1_ex0_27d$DATE27] <- 1

fhs1_ex0_27d$chd_frail <- 0
fhs1_ex0_27d$chd_frail[fhs1_ex0_27d$hfhosp_status==1&fhs1_ex0_27d$hfhosp_dt <= fhs1_ex0_27d$DATE27] <- 1

# Stroke

fhs1_ex0_27d$stroke_frail <- 0
fhs1_ex0_27d$stroke_frail[fhs1_ex0_27d$cvahosp_status==1&fhs1_ex0_27d$cvahosp_dt <= fhs1_ex0_27d$DATE27] <- 1

# Cancer

fhs1_ex0_27d <- merge(fhs1_ex0_27d,
                      fhs_survca_first_any[,c("PID","IDTYPE","D_DATE")],
                      by=c("PID","IDTYPE"),
                      all.x=T)

fhs1_ex0_27d$cancer_frail <- 0
fhs1_ex0_27d$cancer_frail[fhs1_ex0_27d$D_DATE <= fhs1_ex0_27d$DATE27] <- 1

fhs1_ex0_27d$conditions_frail <- fhs1_ex0_27d$HRX27+
  fhs1_ex0_27d$DMRX27+
  fhs1_ex0_27d$copd_frail+
  fhs1_ex0_27d$asthma_frail+
  fhs1_ex0_27d$djd_frail+
  fhs1_ex0_27d$ckd_frail+
  fhs1_ex0_27d$chf_frail+
  fhs1_ex0_27d$chd_frail+
  fhs1_ex0_27d$stroke_frail+
  fhs1_ex0_27d$cancer_frail

fhs1_ex0_27d$illness_frail[fhs1_ex0_27d$conditions_frail <= 4] <- 0
fhs1_ex0_27d$illness_frail[fhs1_ex0_27d$conditions_frail > 4] <- 1


#### (Weight) Loss

fhs1_ex0_27d$wgt27_26_delta <- fhs1_ex0_27d$WGT27-fhs1_ex0_27d$WGT26

fhs1_ex0_27d$wtloss_frail[(fhs1_ex0_27d$WGT27-fhs1_ex0_27d$WGT26) >= 10] <- 1
fhs1_ex0_27d$wtloss_frail[(fhs1_ex0_27d$WGT27-fhs1_ex0_27d$WGT26) < 10] <- 0


#### Calculate FRAIL score

fhs1_ex0_27d$total_frail <- fhs1_ex0_27d$fatigue_frail +
  fhs1_ex0_27d$resistance_frail+
  fhs1_ex0_27d$ambulate_frail+
  fhs1_ex0_27d$illness_frail+
  fhs1_ex0_27d$wtloss_frail

fhs1_ex0_27d$barthel_hygiene <- 0


fhs1_melt_ex0_27d <- melt(fhs1_ex0_27d,id.vars=c("PID","IDTYPE","visit"),na.rm=T,factorsAsStrings=T)
fhs1_melt_ex0_27d$form <- "fhs1_ex0_27"

rm(fhs1_ex0_27d)


# ==============================================================#
####                    FHS original, Exam 28                ####
# ==============================================================#

fhs1_ex0_28d <-
  fread('~/Dropbox/BioLINCC files/Framingham cohort/datasets/CSV/E_EXAM_EX28_0_0256D.csv',
        na.strings=c("NA","","NULL")) # Exam 28

fhs1_ex0_28d$visit <- 28
fhs1_ex0_28d <- merge(fhs1_ex0_28d,fhs1_wkthru[,c("PID",
                                                  "DATE28",
                                                  "SEX",
                                                  "HGT1",
                                                  "BMIC28",
                                                  "WGT28",
                                                  "WGT27",
                                                  "DMRX28",
                                                  "HRX28",
                                                  "AGE28",
                                                  "CREAT28"
)],by="PID",all.x=T)


fhs1_ex0_28d$barthel_hygiene <- 0
fhs1_ex0_28d$essi_chores <- 0

###### Walking speed

fhs1_ex0_28d$walktime_1 <- fhs1_ex0_28d$FU542
fhs1_ex0_28d$walktime_2 <- fhs1_ex0_28d$FU545

fhs1_ex0_28d$walktime_avg <- round(apply(fhs1_ex0_28d[,c("walktime_1","walktime_2")],MARGIN=1,mean,na.rm=T),3)
fhs1_ex0_28d$walktime_avg[fhs1_ex0_28d$walktime_avg=="NaN"] <- NA
fhs1_ex0_28d$walktime_min <- round(apply(fhs1_ex0_28d[,c("walktime_1","walktime_2")],MARGIN=1,min,na.rm=T),3)
fhs1_ex0_28d$walktime_min[fhs1_ex0_28d$walktime_min=="-Inf"] <- NA


########### Grip (kg)

# Produces highest average by hand (assumes dominant hand is stronger)

fhs1_ex0_28d$grip_avg_right <- round(apply(fhs1_ex0_28d[,99:101],MARGIN=1,mean),1)
fhs1_ex0_28d$grip_avg_left <- round(apply(fhs1_ex0_28d[,102:104],MARGIN=1,mean),1)

fhs1_ex0_28d$grip_max <- round(apply(fhs1_ex0_28d[,c("grip_avg_right","grip_avg_left")],MARGIN=1,max),1)


#### Exhaustion


fhs1_ex0_28d$exhaustion_fried[fhs1_ex0_28d$FU367==3|fhs1_ex0_28d$FU380==3] <- 1
fhs1_ex0_28d$exhaustion_fried[fhs1_ex0_28d$FU367<3&fhs1_ex0_28d$FU380<3] <- 0


#### (Weight) Loss

fhs1_ex0_28d$wtloss_fried[(fhs1_ex0_28d$WGT28-fhs1_ex0_28d$WGT27) >= 10] <- 1
fhs1_ex0_28d$wtloss_fried[(fhs1_ex0_28d$WGT28-fhs1_ex0_28d$WGT27) < 10] <- 0


######### FRAIL Score

#### Fatigue

fhs1_ex0_28d$fatigue_frail[fhs1_ex0_28d$FU367==3|fhs1_ex0_28d$FU380==3] <- 1
fhs1_ex0_28d$fatigue_frail[fhs1_ex0_28d$FU367<3&fhs1_ex0_28d$FU380<3] <- 0


#### Resistance

fhs1_ex0_28d$resistance_frail[fhs1_ex0_28d$FU403==0] <- 0
fhs1_ex0_28d$resistance_frail[fhs1_ex0_28d$FU403 %in% c(1,2,3)] <- 1


#### Ambulation

fhs1_ex0_28d$ambulate_frail[fhs1_ex0_28d$FU444==1] <- 0
fhs1_ex0_28d$ambulate_frail[fhs1_ex0_28d$FU444 %in% c(0,2)] <- 1


# FRAIL Illness score counts presence of HTN, DM, cancer, chronic lung disease, MI, CHD, angina, asthma, arthritis, stroke, and ckd
# Scores for 0-4 conditions, score = 0, for ≥ 5, score=1

# DM and HTN

# COPD (chronic lung disease)

fhs1_ex0_28d$copd_frail[fhs1_ex0_28d$FU239==2] <- 0
fhs1_ex0_28d$copd_frail[fhs1_ex0_28d$FU239==1] <- 1
fhs1_ex0_28d$copd_frail[fhs1_ex0_28d$FU239==0] <- 0

# asthma

fhs1_ex0_28d$asthma_frail[fhs1_ex0_28d$FU242==2] <- 0
fhs1_ex0_28d$asthma_frail[fhs1_ex0_28d$FU242==1] <- 1
fhs1_ex0_28d$asthma_frail[fhs1_ex0_28d$FU242==0] <- 0

# DJD (arthritis)

fhs1_ex0_28d$djd_frail[fhs1_ex0_28d$FU245==2] <- 0
fhs1_ex0_28d$djd_frail[fhs1_ex0_28d$FU245==1] <- 1
fhs1_ex0_28d$djd_frail[fhs1_ex0_28d$FU245==0] <- 0

# CKD (renal disease)

fhs1_ex0_28d$RACE <- 1
fhs1_ex0_28d$crcl <- 
  calc_MDRD4(
    as.data.frame(fhs1_ex0_28d),cr="CREAT28",age="AGE28")

fhs1_ex0_28d$ckd_frail[fhs1_ex0_28d$FU238==2] <- 0
fhs1_ex0_28d$ckd_frail[fhs1_ex0_28d$FU238==1|fhs1_ex0_28d$crcl < 60] <- 1
fhs1_ex0_28d$ckd_frail[fhs1_ex0_28d$FU238==0] <- 0

# CHF/MI

fhs1_ex0_28d <- merge(fhs1_ex0_28d,fhs_outcomes,by=c("PID","IDTYPE"))

fhs1_ex0_28d$chf_frail <- 0
fhs1_ex0_28d$chf_frail[fhs1_ex0_28d$hfhosp_status==1&fhs1_ex0_28d$hfhosp_dt <= fhs1_ex0_28d$DATE28] <- 1

fhs1_ex0_28d$chd_frail <- 0
fhs1_ex0_28d$chd_frail[fhs1_ex0_28d$cadhosp_status==1&fhs1_ex0_28d$cadhosp_dt <= fhs1_ex0_28d$DATE28] <- 1

# Stroke

fhs1_ex0_28d$stroke_frail <- 0
fhs1_ex0_28d$stroke_frail[fhs1_ex0_28d$cvahosp_status==1&fhs1_ex0_28d$cvahosp_dt <= fhs1_ex0_28d$DATE28] <- 1

# Cancer

fhs1_ex0_28d <- merge(fhs1_ex0_28d,
                      fhs_survca_first_any[,c("PID","IDTYPE","D_DATE")],
                      by=c("PID","IDTYPE"),
                      all.x=T)

fhs1_ex0_28d$cancer_frail <- 0
fhs1_ex0_28d$cancer_frail[fhs1_ex0_28d$D_DATE <= fhs1_ex0_28d$DATE28] <- 1

fhs1_ex0_28d$conditions_frail <- 
  fhs1_ex0_28d$HRX28+
  fhs1_ex0_28d$DMRX28+
  fhs1_ex0_28d$copd_frail+
  fhs1_ex0_28d$asthma_frail+
  fhs1_ex0_28d$djd_frail+
  fhs1_ex0_28d$ckd_frail+
  fhs1_ex0_28d$chf_frail+
  fhs1_ex0_28d$chd_frail+
  fhs1_ex0_28d$stroke_frail+
  fhs1_ex0_28d$cancer_frail

fhs1_ex0_28d$illness_frail[fhs1_ex0_28d$conditions_frail <= 4] <- 0
fhs1_ex0_28d$illness_frail[fhs1_ex0_28d$conditions_frail > 4] <- 1

#### (Weight) Loss

fhs1_ex0_28d$wtloss_frail[(fhs1_ex0_28d$WGT28-fhs1_ex0_28d$WGT27) >= 10] <- 1
fhs1_ex0_28d$wtloss_frail[(fhs1_ex0_28d$WGT28-fhs1_ex0_28d$WGT27) < 10] <- 0


#### Calculate FRAIL score

fhs1_ex0_28d$total_frail <- fhs1_ex0_28d$fatigue_frail+
  fhs1_ex0_28d$resistance_frail+
  fhs1_ex0_28d$ambulate_frail+
  fhs1_ex0_28d$illness_frail+
  fhs1_ex0_28d$wtloss_frail


fhs1_melt_ex0_28d <- 
  melt(fhs1_ex0_28d,
       id.vars=c("PID","IDTYPE","visit"),
       na.rm=T,
       factorsAsStrings=T)

fhs1_melt_ex0_28d$form <- "fhs1_ex0_28"

rm(fhs1_ex0_28d)


# ============================================================#
###          FHS original, Physical function, Exam 28       ###
# ============================================================#

fhs1_physf_2005 <-
  fread('~/Dropbox/BioLINCC files/Framingham cohort/datasets/CSV/t_physf_2005_m_0162d_v2.csv',
        na.strings=c("NA","","NULL")) # Exam 28

names(fhs1_physf_2005) <-
  toupper(names(fhs1_physf_2005))

fhs1_physf_2005$visit <- 28

fhs1_melt_physf_2005 <- 
  melt(fhs1_physf_2005,
       id.vars=c("PID","IDTYPE","visit"),
       na.rm=T,
       factorsAsStrings=T)

fhs1_melt_physf_2005$form <- "fhs1_physf_2005"

rm(fhs1_physf_2005)


# ==============================================================#
####                FHS original, Exam 29                    ####
# ==============================================================#

fhs1_ex0_29d <-
  fread('~/Dropbox/BioLINCC files/Framingham cohort/datasets/CSV/E_EXAM_EX29_0_0210D.csv',
        na.strings=c("NA","","NULL")) # Exam 29

fhs1_ex0_29d$visit <- 29
fhs1_ex0_29d <- merge(fhs1_ex0_29d,fhs1_wkthru[,c("PID",
                                                  "SEX",
                                                  "DATE29",
                                                  "HGT1",
                                                  "BMIC29",
                                                  "WGT29",
                                                  "WGT28",
                                                  "DMRX29",
                                                  "HRX29")],
                      by="PID",
                      all.x=T)


fhs1_ex0_29d$barthel_hygiene <- 0
fhs1_ex0_29d$essi_chores <- 0


######### FRIED

##### Walking speed 

## Walking speed

fhs1_ex0_29d$walktime_1 <- fhs1_ex0_29d$FV315
fhs1_ex0_29d$walktime_2 <- fhs1_ex0_29d$FV319

fhs1_ex0_29d$walktime_avg <- round(apply(fhs1_ex0_29d[,c("walktime_1","walktime_2")],MARGIN=1,mean,na.rm=T),3)
fhs1_ex0_29d$walktime_avg[fhs1_ex0_29d$walktime_avg=="NaN"] <- NA
fhs1_ex0_29d$walktime_min <- round(apply(fhs1_ex0_29d[,c("walktime_1","walktime_2")],MARGIN=1,min,na.rm=T),3)
fhs1_ex0_29d$walktime_min[fhs1_ex0_29d$walktime_min=="-Inf"] <- NA


########### Grip (kg)

# Produces highest average by hand (assumes dominant hand is stronger)

fhs1_ex0_29d$grip_avg_right <- round(apply(fhs1_ex0_29d[,177:179],MARGIN=1,mean),1)
fhs1_ex0_29d$grip_avg_left <- round(apply(fhs1_ex0_29d[,180:182],MARGIN=1,mean),1)

fhs1_ex0_29d$grip_max <- round(apply(fhs1_ex0_29d[,c("grip_avg_right","grip_avg_left")],MARGIN=1,max),1)


##### Exhaustion - Fried

fhs1_ex0_29d$exhaustion_fried[fhs1_ex0_29d$FV526==3|fhs1_ex0_29d$FV539==3] <- 1
fhs1_ex0_29d$exhaustion_fried[fhs1_ex0_29d$FV526<3&fhs1_ex0_29d$FV539<3] <- 0

#### (Weight) Loss

fhs1_ex0_29d$wtloss_fried[(fhs1_ex0_29d$WGT29-fhs1_ex0_29d$WGT28) >= 10] <- 1
fhs1_ex0_29d$wtloss_fried[(fhs1_ex0_29d$WGT29-fhs1_ex0_29d$WGT28) < 10] <- 0


######### FRAIL Score

#### Fatigue

fhs1_ex0_29d$fatigue_frail[fhs1_ex0_29d$FV526==3|fhs1_ex0_29d$FV539==3] <- 1
fhs1_ex0_29d$fatigue_frail[fhs1_ex0_29d$FV526<3&fhs1_ex0_29d$FV539<3] <- 0


#### Resistance

fhs1_ex0_29d$resistance_frail[fhs1_ex0_29d$FV399==0] <- 0
fhs1_ex0_29d$resistance_frail[fhs1_ex0_29d$FV399 %in% c(1,2,3)] <- 1

#### Ambulation

fhs1_ex0_29d$ambulate_frail[fhs1_ex0_29d$FV386==1] <- 0
fhs1_ex0_29d$ambulate_frail[fhs1_ex0_29d$FV386==0] <- 1


# FRAIL Illness score counts presence of HTN, DM, cancer, 
# chronic lung disease, MI, CHD, angina, asthma, arthritis, stroke, and ckd
# Scores for 0-4 conditions, score = 0, for ≥ 5, score=1

# DM and HTN


# COPD (chronic lung disease)

fhs1_ex0_29d$copd_frail[fhs1_ex0_29d$FV241==2] <- 0
fhs1_ex0_29d$copd_frail[fhs1_ex0_29d$FV241==1] <- 1
fhs1_ex0_29d$copd_frail[fhs1_ex0_29d$FV241==0] <- 0

# asthma

fhs1_ex0_29d$asthma_frail[fhs1_ex0_29d$FV244==2] <- 0
fhs1_ex0_29d$asthma_frail[fhs1_ex0_29d$FV244==1] <- 1
fhs1_ex0_29d$asthma_frail[fhs1_ex0_29d$FV244==0] <- 0

# DJD (arthritis)

fhs1_ex0_29d$djd_frail[fhs1_ex0_29d$FV247==2] <- 0
fhs1_ex0_29d$djd_frail[fhs1_ex0_29d$FV247==1] <- 1
fhs1_ex0_29d$djd_frail[fhs1_ex0_29d$FV247==0] <- 0

# CKD (renal disease)

fhs1_ex0_29d$ckd_frail[fhs1_ex0_29d$FV240==2] <- 0
fhs1_ex0_29d$ckd_frail[fhs1_ex0_29d$FV240==1] <- 1
fhs1_ex0_29d$ckd_frail[fhs1_ex0_29d$FV240==0] <- 0

# CHF/MI

fhs1_ex0_29d <- merge(fhs1_ex0_29d,fhs_outcomes,by=c("PID","IDTYPE"))

fhs1_ex0_29d$chf_frail <- 0
fhs1_ex0_29d$chf_frail[fhs1_ex0_29d$hfhosp_status==1&fhs1_ex0_29d$hfhosp_dt <= fhs1_ex0_29d$DATE29] <- 1

fhs1_ex0_29d$chd_frail <- 0
fhs1_ex0_29d$chd_frail[fhs1_ex0_29d$cadhosp_status==1&fhs1_ex0_29d$cadhosp_dt <= fhs1_ex0_29d$DATE29] <- 1

# Stroke

fhs1_ex0_29d$stroke_frail <- 0
fhs1_ex0_29d$stroke_frail[fhs1_ex0_29d$cvahosp_status==1&fhs1_ex0_29d$cvahosp_dt <= fhs1_ex0_29d$DATE29] <- 1

# Cancer

fhs1_ex0_29d <- 
  merge(fhs1_ex0_29d,
        fhs_survca_first_any[,c("PID","IDTYPE","D_DATE")],
        by=c("PID","IDTYPE"),
        all.x=T)

fhs1_ex0_29d$cancer_frail <- 0
fhs1_ex0_29d$cancer_frail[fhs1_ex0_29d$D_DATE <= fhs1_ex0_29d$DATE29] <- 1

fhs1_ex0_29d$conditions_frail <- fhs1_ex0_29d$HRX29+
  fhs1_ex0_29d$DMRX29+
  fhs1_ex0_29d$copd_frail+
  fhs1_ex0_29d$asthma_frail+
  fhs1_ex0_29d$djd_frail+
  fhs1_ex0_29d$ckd_frail+
  fhs1_ex0_29d$chf_frail+
  fhs1_ex0_29d$chd_frail+
  fhs1_ex0_29d$stroke_frail+
  fhs1_ex0_29d$cancer_frail

fhs1_ex0_29d$illness_frail[fhs1_ex0_29d$conditions_frail <= 4] <- 0
fhs1_ex0_29d$illness_frail[fhs1_ex0_29d$conditions_frail > 4] <- 1

#### (Weight) Loss

fhs1_ex0_29d$wtloss_frail[(fhs1_ex0_29d$WGT29-fhs1_ex0_29d$WGT28) >= 10] <- 1
fhs1_ex0_29d$wtloss_frail[(fhs1_ex0_29d$WGT29-fhs1_ex0_29d$WGT28) < 10] <- 0

#### Calculate FRAIL score

fhs1_ex0_29d$total_frail <- fhs1_ex0_29d$fatigue_frail+
  fhs1_ex0_29d$resistance_frail+
  fhs1_ex0_29d$ambulate_frail+
  fhs1_ex0_29d$illness_frail+
  fhs1_ex0_29d$wtloss_frail


fhs1_melt_ex0_29d <- melt(fhs1_ex0_29d,id.vars=c("PID","IDTYPE","visit"),na.rm=T,factorsAsStrings=T)
fhs1_melt_ex0_29d$form <- "fhs1_ex0_29"

rm(fhs1_ex0_29d)

# =============================================================#
###         FHS original, Exam 29 Physical function          ###
# =============================================================#

fhs1_t_physfunc_2010 <-
  fread('~/Dropbox/BioLINCC files/Framingham cohort/datasets/CSV/t_physfunc_2010_m_0634d.csv',
        na.strings=c("NA","","NULL")) # Exam 29

fhs1_t_physfunc_2010$visit <- 29
fhs1_melt_t_physfunc_2010 <- melt(fhs1_t_physfunc_2010,id.vars=c("PID","IDTYPE","visit"),na.rm=T,factorsAsStrings=T)
fhs1_melt_t_physfunc_2010$form <- "fhs1_physfunc_2010"

rm(fhs1_t_physfunc_2010)


# ===============================================================#
####                    FHS original, Exam 30                 ####
# ===============================================================#

fhs1_ex0_30d <-fread('~/Dropbox/BioLINCC files/Framingham cohort/datasets/CSV/E_EXAM_EX30_0_0274D.csv',na.strings=c("NA","","NULL")) # Exam 30
fhs1_ex0_30d$visit <- 30
fhs1_ex0_30d <- merge(fhs1_ex0_30d,
                      fhs1_wkthru[,c("PID",
                                     "SEX",
                                     "AGE30",
                                     "DATE30",
                                     "HGT1",
                                     "BMIC30",
                                     "WGT30",
                                     "WGT29",
                                     "HRX30",
                                     "DMRX30")],by="PID",all.x=T)


fhs1_ex0_30d$barthel_hygiene <- 0
fhs1_ex0_30d$essi_chores <- 0


## Walking speed

fhs1_ex0_30d$walktime_1 <- fhs1_ex0_30d$FW367
fhs1_ex0_30d$walktime_2 <- fhs1_ex0_30d$FW372

fhs1_ex0_30d$walktime_avg <- round(apply(fhs1_ex0_30d[,c("walktime_1","walktime_2")],MARGIN=1,mean,na.rm=T),3)
fhs1_ex0_30d$walktime_avg[fhs1_ex0_30d$walktime_avg=="NaN"] <- NA
fhs1_ex0_30d$walktime_min <- round(apply(fhs1_ex0_30d[,c("walktime_1","walktime_2")],MARGIN=1,min,na.rm=T),3)
fhs1_ex0_30d$walktime_min[fhs1_ex0_30d$walktime_min=="-Inf"] <- NA


########### Grip (kg)

# Produces highest average by hand (assumes dominant hand is stronger)

fhs1_ex0_30d$grip_avg_right <- round(apply(fhs1_ex0_30d[,173:175],MARGIN=1,mean),1)
fhs1_ex0_30d$grip_avg_left <- round(apply(fhs1_ex0_30d[,176:178],MARGIN=1,mean),1)

fhs1_ex0_30d$grip_max <- round(apply(fhs1_ex0_30d[,c("grip_avg_right","grip_avg_left")],MARGIN=1,max),1)


#### Exhaustion

fhs1_ex0_30d$exhaustion_fried[fhs1_ex0_30d$FW528==3|fhs1_ex0_30d$FW541==3] <- 1
fhs1_ex0_30d$exhaustion_fried[fhs1_ex0_30d$FW528<3&fhs1_ex0_30d$FW541<3] <- 0


#### (Weight) Loss

fhs1_ex0_30d$wtloss_fried[(fhs1_ex0_30d$WGT30-fhs1_ex0_30d$WGT29)>=10] <- 1
fhs1_ex0_30d$wtloss_fried[(fhs1_ex0_30d$WGT30-fhs1_ex0_30d$WGT29) < 10] <- 0


##### CES-D

fhs1_ex0_30d$CESD20_missing <- rowSums(is.na(fhs1_ex0_30d[,300:319]))
fhs1_ex0_30d$CESD20 <- rowSums(fhs1_ex0_30d[,300:319])
fhs1_ex0_30d$CESD20[fhs1_ex0_30d$CESD20_missing >= 4] <- NA


######### FRAIL Score

#### Fatigue

fhs1_ex0_30d$fatigue_frail[fhs1_ex0_30d$FW528==3|fhs1_ex0_30d$FW541==3] <- 1
fhs1_ex0_30d$fatigue_frail[fhs1_ex0_30d$FW528<3&fhs1_ex0_30d$FW541<3] <- 0


#### Resistance

fhs1_ex0_30d$resistance_frail[fhs1_ex0_30d$FW448 %in% c(0,1)] <- fhs1_ex0_30d$FW448[fhs1_ex0_30d$FW448 %in% c(0,1)]

#### Ambulation

fhs1_ex0_30d$ambulate_frail[fhs1_ex0_30d$FW446 %in% c(0,1)] <- fhs1_ex0_30d$FW446[fhs1_ex0_30d$FW446 %in% c(0,1)]


# FRAIL Illness score counts presence of HTN, DM, cancer, chronic lung disease, MI, CHD, angina, asthma, arthritis, stroke, and ckd
# Scores for 0-4 conditions, score = 0, for ≥ 5, score=1

# DM and HTN

# COPD (chronic lung disease)

fhs1_ex0_30d$copd_frail[fhs1_ex0_30d$FW283==2] <- 0
fhs1_ex0_30d$copd_frail[fhs1_ex0_30d$FW283==1] <- 1
fhs1_ex0_30d$copd_frail[fhs1_ex0_30d$FW283==0] <- 0

# asthma

fhs1_ex0_30d$asthma_frail[fhs1_ex0_30d$FW286==2] <- 0
fhs1_ex0_30d$asthma_frail[fhs1_ex0_30d$FW286==1] <- 1
fhs1_ex0_30d$asthma_frail[fhs1_ex0_30d$FW286==0] <- 0

# DJD (arthritis)

fhs1_ex0_30d$djd_frail[fhs1_ex0_30d$FW289==2] <- 0
fhs1_ex0_30d$djd_frail[fhs1_ex0_30d$FW289==1] <- 1
fhs1_ex0_30d$djd_frail[fhs1_ex0_30d$FW289==0] <- 0

# CKD (renal disease)

fhs1_ex0_30d$ckd_frail[fhs1_ex0_30d$FW281==2] <- 0
fhs1_ex0_30d$ckd_frail[fhs1_ex0_30d$FW281==1] <- 1
fhs1_ex0_30d$ckd_frail[fhs1_ex0_30d$FW281==0] <- 0

# CHF/MI

fhs1_ex0_30d <- merge(fhs1_ex0_30d,fhs_outcomes,by=c("PID","IDTYPE"))

fhs1_ex0_30d$chf_frail <- 0
fhs1_ex0_30d$chf_frail[fhs1_ex0_30d$hfhosp_status==1&fhs1_ex0_30d$hfhosp_dt <= fhs1_ex0_30d$DATE30] <- 1

fhs1_ex0_30d$chd_frail <- 0
fhs1_ex0_30d$chd_frail[fhs1_ex0_30d$cadhosp_status==1&fhs1_ex0_30d$cadhosp_dt <= fhs1_ex0_30d$DATE30] <- 1

# Stroke

fhs1_ex0_30d$stroke_frail <- 0
fhs1_ex0_30d$stroke_frail[
  fhs1_ex0_30d$cvahosp_status==1&
    fhs1_ex0_30d$cvahosp_dt <= fhs1_ex0_30d$DATE30] <- 1

# Cancer

fhs1_ex0_30d <- 
  merge(fhs1_ex0_30d,
        fhs_survca_first_any[,c("PID","IDTYPE","D_DATE")],
        by=c("PID","IDTYPE"),all.x=T)

fhs1_ex0_30d$cancer_frail <- 0
fhs1_ex0_30d$cancer_frail[fhs1_ex0_30d$D_DATE <= fhs1_ex0_30d$DATE30] <- 1

fhs1_ex0_30d$conditions_frail <- fhs1_ex0_30d$HRX30+
  fhs1_ex0_30d$DMRX30+
  fhs1_ex0_30d$copd_frail+
  fhs1_ex0_30d$asthma_frail+
  fhs1_ex0_30d$djd_frail+
  fhs1_ex0_30d$ckd_frail+
  fhs1_ex0_30d$chf_frail+
  fhs1_ex0_30d$chd_frail+
  fhs1_ex0_30d$stroke_frail+
  fhs1_ex0_30d$cancer_frail

fhs1_ex0_30d$illness_frail[fhs1_ex0_30d$conditions_frail <= 4] <- 0
fhs1_ex0_30d$illness_frail[fhs1_ex0_30d$conditions_frail > 4] <- 1

#### (Weight) Loss

fhs1_ex0_30d$wtloss_frail[(fhs1_ex0_30d$WGT30-fhs1_ex0_30d$WGT29) <= -10] <- 1
fhs1_ex0_30d$wtloss_frail[(fhs1_ex0_30d$WGT30-fhs1_ex0_30d$WGT29) > -10] <- 0


#### Calculate FRAIL score

fhs1_ex0_30d$total_frail <- fhs1_ex0_30d$fatigue_frail+
  fhs1_ex0_30d$resistance_frail+
  fhs1_ex0_30d$ambulate_frail+
  fhs1_ex0_30d$illness_frail+
  fhs1_ex0_30d$wtloss_frail


fhs1_melt_ex0_30d <- melt(fhs1_ex0_30d,id.vars=c("PID","IDTYPE","visit"),na.rm=T,factorsAsStrings=T)
fhs1_melt_ex0_30d$form <- "fhs1_ex0_30"

rm(fhs1_ex0_30d)


# =============================================================#
###      FHS original, Berkman questionnaire Exam 30         ###
# =============================================================#

fhs1_bsni_2009 <-
  fread('~/Dropbox/BioLINCC files/Framingham cohort/datasets/CSV/q_berkman_2009_m_0635d.csv',
        na.strings=c("NA","","NULL")) # Exam 30

names(fhs1_bsni_2009) <- toupper(names(fhs1_bsni_2009))

fhs1_bsni_2009$visit <- 30

fhs1_melt_bsni_2009 <- 
  melt(fhs1_bsni_2009,
       id.vars=c("PID","IDTYPE","visit"),
       na.rm=T,
       factorsAsStrings=T)

fhs1_melt_bsni_2009$form <- "fhs1_bsni_2009"

rm(fhs1_bsni_2009)


# ==============================================================#
####                   FHS original, Exam 31                 ####
# ==============================================================#


fhs1_ex0_31d <-
  fread('~/Dropbox/BioLINCC files/Framingham cohort/datasets/CSV/E_EXAM_EX31_0_0738D.csv',
        na.strings=c("NA","","NULL")) # Exam 31

fhs1_ex0_31d$visit <- 31
fhs1_ex0_31d <- merge(fhs1_ex0_31d,fhs1_wkthru[,c("PID",
                                                  "SEX",
                                                  "HGT1",
                                                  "BMIC31",
                                                  "WGT31",
                                                  "WGT30",
                                                  "HRX31",
                                                  "DMRX31",
                                                  "DATE31")],by="PID",all.x=T)

fhs1_ex0_31d$barthel_hygiene <- 0
fhs1_ex0_31d$essi_chores <- 0


## Walking speed

fhs1_ex0_31d$walktime_1 <- fhs1_ex0_31d$FX367
fhs1_ex0_31d$walktime_2 <- fhs1_ex0_31d$FX372

fhs1_ex0_31d$walktime_avg <- round(apply(fhs1_ex0_31d[,c("walktime_1","walktime_2")],MARGIN=1,mean,na.rm=T),3)
fhs1_ex0_31d$walktime_avg[fhs1_ex0_31d$walktime_avg=="NaN"] <- NA
fhs1_ex0_31d$walktime_min <- round(apply(fhs1_ex0_31d[,c("walktime_1","walktime_2")],MARGIN=1,min,na.rm=T),3)
fhs1_ex0_31d$walktime_min[fhs1_ex0_31d$walktime_min=="-Inf"] <- NA


########### Grip (kg)

# Produces highest average by hand (assumes dominant hand is stronger)

fhs1_ex0_31d$grip_avg_right <- round(apply(fhs1_ex0_31d[,179:181],MARGIN=1,mean),1)
fhs1_ex0_31d$grip_avg_left <- round(apply(fhs1_ex0_31d[,182:184],MARGIN=1,mean),1)

fhs1_ex0_31d$grip_max <- round(apply(fhs1_ex0_31d[,c("grip_avg_right","grip_avg_left")],MARGIN=1,max),1)


#### Exhaustion

fhs1_ex0_31d$exhaustion_fried[fhs1_ex0_31d$FX528==3|fhs1_ex0_31d$FX541==3] <- 1
fhs1_ex0_31d$exhaustion_fried[fhs1_ex0_31d$FX528<3&fhs1_ex0_31d$FX541<3] <- 0


#### (Weight) Loss

fhs1_ex0_31d$wtloss_fried[(fhs1_ex0_31d$WGT31-fhs1_ex0_31d$WGT30)>=10] <- 1
fhs1_ex0_31d$wtloss_fried[(fhs1_ex0_31d$WGT31-fhs1_ex0_31d$WGT30) < 10] <- 0


######### FRAIL Score

fhs1_ex0_31d$fatigue_frail[fhs1_ex0_31d$FX528==3|fhs1_ex0_31d$FX541==3] <- 1
fhs1_ex0_31d$fatigue_frail[fhs1_ex0_31d$FX528<3&fhs1_ex0_31d$FX541<3] <- 0


#### Resistance

fhs1_ex0_31d$resistance_frail[fhs1_ex0_31d$FX448 %in% c(0,2)] <- 1
fhs1_ex0_31d$resistance_frail[fhs1_ex0_31d$FX448==1] <- 0

#### Ambulation

fhs1_ex0_31d$ambulate_frail[fhs1_ex0_31d$FX446 %in% c(0,2)] <-1
fhs1_ex0_31d$ambulate_frail[fhs1_ex0_31d$FX446==1] <-0


# FRAIL Illness score counts presence of HTN, DM, cancer, chronic lung disease, MI, CHD, angina, asthma, arthritis, stroke, and ckd
# Scores for 0-4 conditions, score = 0, for ≥ 5, score=1


# COPD (chronic lung disease)

fhs1_ex0_31d$copd_frail[fhs1_ex0_31d$FX283==2] <- 0
fhs1_ex0_31d$copd_frail[fhs1_ex0_31d$FX283==1] <- 1
fhs1_ex0_31d$copd_frail[fhs1_ex0_31d$FX283==0] <- 0

# asthma

fhs1_ex0_31d$asthma_frail[fhs1_ex0_31d$FX286==2] <- 0
fhs1_ex0_31d$asthma_frail[fhs1_ex0_31d$FX286==1] <- 1
fhs1_ex0_31d$asthma_frail[fhs1_ex0_31d$FX286==0] <- 0

# DJD (arthritis)

fhs1_ex0_31d$djd_frail[fhs1_ex0_31d$FX289==2] <- 0
fhs1_ex0_31d$djd_frail[fhs1_ex0_31d$FX289==1] <- 1
fhs1_ex0_31d$djd_frail[fhs1_ex0_31d$FX289==0] <- 0

# CKD (renal disease)

fhs1_ex0_31d$ckd_frail[fhs1_ex0_31d$FX281==2] <- 0
fhs1_ex0_31d$ckd_frail[fhs1_ex0_31d$FX281==1] <- 1
fhs1_ex0_31d$ckd_frail[fhs1_ex0_31d$FX281==0] <- 0

# CHF/MI

fhs1_ex0_31d <- merge(fhs1_ex0_31d,fhs_outcomes,by=c("PID","IDTYPE"))

fhs1_ex0_31d$chf_frail <- 0
fhs1_ex0_31d$chf_frail[fhs1_ex0_31d$hfhosp_status==1&fhs1_ex0_31d$hfhosp_dt <= fhs1_ex0_31d$DATE31] <- 1

fhs1_ex0_31d$chd_frail <- 0
fhs1_ex0_31d$chd_frail[fhs1_ex0_31d$cadhosp_status==1&fhs1_ex0_31d$cadhosp_dt <= fhs1_ex0_31d$DATE31] <- 1

# Stroke

fhs1_ex0_31d$stroke_frail <- 0
fhs1_ex0_31d$stroke_frail[fhs1_ex0_31d$cvahosp_status==1&fhs1_ex0_31d$cvahosp_dt <= fhs1_ex0_31d$DATE31] <- 1

# Cancer

fhs1_ex0_31d <- 
  merge(fhs1_ex0_31d,
        fhs_survca_first_any[,c("PID","IDTYPE","D_DATE")],
        by=c("PID","IDTYPE"),
        all.x=T)

fhs1_ex0_31d$cancer_frail <- 0
fhs1_ex0_31d$cancer_frail[fhs1_ex0_31d$D_DATE <= fhs1_ex0_31d$DATE31] <- 1

fhs1_ex0_31d$conditions_frail <- fhs1_ex0_31d$HRX31+
  fhs1_ex0_31d$DMRX31+
  fhs1_ex0_31d$copd_frail+
  fhs1_ex0_31d$asthma_frail+
  fhs1_ex0_31d$djd_frail+
  fhs1_ex0_31d$ckd_frail+
  fhs1_ex0_31d$chf_frail+
  fhs1_ex0_31d$chd_frail+
  fhs1_ex0_31d$stroke_frail+
  fhs1_ex0_31d$cancer_frail

fhs1_ex0_31d$illness_frail[fhs1_ex0_31d$conditions_frail <= 4] <- 0
fhs1_ex0_31d$illness_frail[fhs1_ex0_31d$conditions_frail > 4] <- 1


#### (Weight) Loss

fhs1_ex0_31d$wtloss_frail[(fhs1_ex0_31d$WGT31-fhs1_ex0_31d$WGT30)>=10] <- 1
fhs1_ex0_31d$wtloss_frail[(fhs1_ex0_31d$WGT31-fhs1_ex0_31d$WGT30) < 10] <- 0


#### Calculate FRAIL score

fhs1_ex0_31d$total_frail <- fhs1_ex0_31d$fatigue_frail+
  fhs1_ex0_31d$resistance_frail+
  fhs1_ex0_31d$ambulate_frail+
  fhs1_ex0_31d$illness_frail+
  fhs1_ex0_31d$wtloss_frail


##### CES-D

fhs1_ex0_31d$CESD20_missing <- rowSums(is.na(fhs1_ex0_31d[,306:325]))
fhs1_ex0_31d$CESD20 <- rowSums(fhs1_ex0_31d[,306:325])
fhs1_ex0_31d$CESD20[fhs1_ex0_31d$CESD20_missing >= 4] <- NA


####

fhs1_melt_ex0_31d <- melt(fhs1_ex0_31d,id.vars=c("PID","IDTYPE","visit"),na.rm=T,factorsAsStrings=T)
fhs1_melt_ex0_31d$form <- "fhs1_ex0_31"

rm(fhs1_ex0_31d)


# ==============================================================#
####                    FHS original, Exam 32                ####
# ==============================================================#

fhs1_ex0_32d <-
  fread('~/Dropbox/BioLINCC files/Framingham cohort/datasets/CSV/E_EXAM_EX32_0_0939D.csv',
        na.strings=c("NA","","NULL")) # Exam 32

fhs1_ex0_32d$visit <- 32
fhs1_ex0_32d <- merge(fhs1_ex0_32d,fhs1_wkthru[,c("PID",
                                                  "SEX",
                                                  "HGT1",
                                                  "BMIC32",
                                                  "DATE32",
                                                  "WGT32",
                                                  "WGT31",
                                                  "DMRX32",
                                                  "HRX32")],by="PID",all.x=T)

fhs1_ex0_32d$barthel_hygiene <- 0


## Walking speed

fhs1_ex0_32d$walktime_1 <- fhs1_ex0_32d$FY367
fhs1_ex0_32d$walktime_2 <- fhs1_ex0_32d$FY372

fhs1_ex0_32d$walktime_avg <- round(apply(fhs1_ex0_32d[,c("walktime_1","walktime_2")],MARGIN=1,mean,na.rm=T),3)
fhs1_ex0_32d$walktime_avg[fhs1_ex0_32d$walktime_avg=="NaN"] <- NA
fhs1_ex0_32d$walktime_min <- round(apply(fhs1_ex0_32d[,c("walktime_1","walktime_2")],MARGIN=1,min,na.rm=T),3)
fhs1_ex0_32d$walktime_min[fhs1_ex0_32d$walktime_min=="-Inf"] <- NA

########### Grip (kg)


# Produces highest average by hand (assumes dominant hand is stronger)

fhs1_ex0_32d$grip_avg_right <- round(apply(fhs1_ex0_32d[,182:184],MARGIN=1,mean),1)
fhs1_ex0_32d$grip_avg_left <- round(apply(fhs1_ex0_32d[,185:187],MARGIN=1,mean),1)

fhs1_ex0_32d$grip_max <- round(apply(fhs1_ex0_32d[,c("grip_avg_right","grip_avg_left")],MARGIN=1,max),1)


#### Exhaustion

fhs1_ex0_32d$exhaustion_fried[fhs1_ex0_32d$FY528==3|fhs1_ex0_32d$FY541==3] <- 1
fhs1_ex0_32d$exhaustion_fried[fhs1_ex0_32d$FY528<3&fhs1_ex0_32d$FY541<3] <- 0


#### (Weight) Loss

fhs1_ex0_32d$wtloss_fried[(fhs1_ex0_32d$WGT32-fhs1_ex0_32d$WGT31)>=10] <- 1
fhs1_ex0_32d$wtloss_fried[(fhs1_ex0_32d$WGT32-fhs1_ex0_32d$WGT31) < 10] <- 0


#=============================================================================#

######### FRAIL Score


#### Fatigue

fhs1_ex0_32d$fatigue_frail[fhs1_ex0_32d$FY528==3|fhs1_ex0_32d$FY541==3] <- 1
fhs1_ex0_32d$fatigue_frail[fhs1_ex0_32d$FY528<3&fhs1_ex0_32d$FY541<3] <- 0


#### Resistance

fhs1_ex0_32d$resistance_frail[fhs1_ex0_32d$FY463==0] <- 1
fhs1_ex0_32d$resistance_frail[fhs1_ex0_32d$FY463==1] <- 0

#### Ambulation

fhs1_ex0_32d$ambulate_frail[fhs1_ex0_32d$FY462==0] <-1
fhs1_ex0_32d$ambulate_frail[fhs1_ex0_32d$FY462==1] <-0


# FRAIL Illness score counts presence of HTN, DM, cancer, chronic lung disease, MI, CHD, angina, asthma, arthritis, stroke, and ckd
# Scores for 0-4 conditions, score = 0, for ≥ 5, score=1


# COPD (chronic lung disease)

fhs1_ex0_32d$copd_frail[fhs1_ex0_32d$FY283==2] <- 0
fhs1_ex0_32d$copd_frail[fhs1_ex0_32d$FY283==1] <- 1
fhs1_ex0_32d$copd_frail[fhs1_ex0_32d$FY283==0] <- 0

# asthma

fhs1_ex0_32d$asthma_frail[fhs1_ex0_32d$FY286==2] <- 0
fhs1_ex0_32d$asthma_frail[fhs1_ex0_32d$FY286==1] <- 1
fhs1_ex0_32d$asthma_frail[fhs1_ex0_32d$FY286==0] <- 0

# DJD (arthritis)

fhs1_ex0_32d$djd_frail[fhs1_ex0_32d$FY289==2] <- 0
fhs1_ex0_32d$djd_frail[fhs1_ex0_32d$FY289==1] <- 1
fhs1_ex0_32d$djd_frail[fhs1_ex0_32d$FY289==0] <- 0

# CKD (renal disease)

fhs1_ex0_32d$ckd_frail[fhs1_ex0_32d$FY281==2] <- 0
fhs1_ex0_32d$ckd_frail[fhs1_ex0_32d$FY281==1] <- 1
fhs1_ex0_32d$ckd_frail[fhs1_ex0_32d$FY281==0] <- 0

# CHF/MI

fhs1_ex0_32d <- merge(fhs1_ex0_32d,fhs_outcomes,by=c("PID","IDTYPE"))

fhs1_ex0_32d$chf_frail <- 0
fhs1_ex0_32d$chf_frail[fhs1_ex0_32d$hfhosp_status==1&fhs1_ex0_32d$hfhosp_dt <= fhs1_ex0_32d$DATE32] <- 1

fhs1_ex0_32d$chd_frail <- 0
fhs1_ex0_32d$chd_frail[fhs1_ex0_32d$cadhosp_status==1&fhs1_ex0_32d$cadhosp_dt <= fhs1_ex0_32d$DATE32] <- 1

# Stroke

fhs1_ex0_32d <- merge(fhs1_ex0_32d,fhs_survstk,by=c("PID","IDTYPE"))

fhs1_ex0_32d$stroke_frail <- 0
fhs1_ex0_32d$stroke_frail[fhs1_ex0_32d$cvahosp_status==1&fhs1_ex0_32d$cvahosp_dt <= fhs1_ex0_32d$DATE32] <- 1

# Cancer

fhs1_ex0_32d <- merge(fhs1_ex0_32d,
                      fhs_survca_first_any[,c("PID","IDTYPE","D_DATE")],
                      by=c("PID","IDTYPE"),
                      all.x=T)

fhs1_ex0_32d$cancer_frail <- 0
fhs1_ex0_32d$cancer_frail[fhs1_ex0_32d$D_DATE <= fhs1_ex0_32d$DATE32] <- 1

fhs1_ex0_32d$conditions_frail <- fhs1_ex0_32d$HRX32+
  fhs1_ex0_32d$DMRX32+
  fhs1_ex0_32d$copd_frail+
  fhs1_ex0_32d$asthma_frail+
  fhs1_ex0_32d$djd_frail+
  fhs1_ex0_32d$ckd_frail+
  fhs1_ex0_32d$chf_frail+
  fhs1_ex0_32d$chd_frail+
  fhs1_ex0_32d$stroke_frail+
  fhs1_ex0_32d$cancer_frail

fhs1_ex0_32d$illness_frail[fhs1_ex0_32d$conditions_frail <= 4] <- 0
fhs1_ex0_32d$illness_frail[fhs1_ex0_32d$conditions_frail > 4] <- 1


#### (Weight) Loss

fhs1_ex0_32d$wtloss_frail[(fhs1_ex0_32d$WGT32-fhs1_ex0_32d$WGT31)>=10] <- 1
fhs1_ex0_32d$wtloss_frail[(fhs1_ex0_32d$WGT32-fhs1_ex0_32d$WGT31) < 10] <- 0


#### Calculate FRAIL score

fhs1_ex0_32d$total_frail <- 
  fhs1_ex0_32d$fatigue_frail+
  fhs1_ex0_32d$resistance_frail+
  fhs1_ex0_32d$ambulate_frail+
  fhs1_ex0_32d$illness_frail+
  fhs1_ex0_32d$wtloss_frail


##### CES-D


fhs1_ex0_32d$CESD20_missing <- rowSums(is.na(fhs1_ex0_32d[,295:304]))
fhs1_ex0_32d$CESD20 <- rowSums(fhs1_ex0_32d[,295:304])
fhs1_ex0_32d$CESD20[fhs1_ex0_32d$CESD20_missing >= 4] <- NA

### 

fhs1_melt_ex0_32d <- melt(fhs1_ex0_32d,
                          id.vars=c("PID","IDTYPE","visit"),
                          na.rm=T,
                          factorsAsStrings=T)

fhs1_melt_ex0_32d$form <- "fhs1_ex0_32"

rm(fhs1_ex0_32d)


# ===============================================================##
####              FHS original, HA1c, exam 19,23               ---#
# ===============================================================##

fhs1_l_hba1c_ex23 <-
  fread('~/Dropbox/BioLINCC files/Framingham cohort/datasets/CSV/L_HBA1C_EX23_0_0110D.csv',
        na.strings=c("NA","","NULL")) # HbA1c

fhs1_l_hba1c_ex19 <- fhs1_l_hba1c_ex23[,c("PID","IDTYPE","DATE1920","HBA1920")]
fhs1_l_hba1c_ex19$visit <- 19
fhs1_l_hba1c_ex19$form <- "fhs1_hba1c"
fhs1_l_hba1c_ex19$variable <- "HB1920"
names(fhs1_l_hba1c_ex19) <- c("PID","IDTYPE","DATE","value","visit","form","variable")
fhs1_melt_l_hba1c_ex19 <- fhs1_l_hba1c_ex19


fhs1_l_hba1c_ex22 <- fhs1_l_hba1c_ex23[,c("PID","IDTYPE","DATE22","HBA22")]
fhs1_l_hba1c_ex22$visit <- 22
fhs1_l_hba1c_ex22$form <- "fhs1_hba1c"
fhs1_l_hba1c_ex22$variable <- "HBA22"
names(fhs1_l_hba1c_ex22) <- c("PID","IDTYPE","DATE","value","visit","form","variable")
fhs1_melt_l_hba1c_ex22 <- fhs1_l_hba1c_ex22

fhs1_melt_l_hba1c <- rbind(fhs1_melt_l_hba1c_ex19,
                           fhs1_melt_l_hba1c_ex22)

fhs1_melt_l_hba1c <- subset(fhs1_melt_l_hba1c,!is.na(value))

rm(fhs1_l_hba1c_ex23,
   fhs1_l_hba1c_ex19,
   fhs1_l_hba1c_ex22,
   fhs1_melt_l_hba1c_ex19,
   fhs1_melt_l_hba1c_ex22
)


# ==============================================================#
##                 FHS original, PFTs, exam 19                 ##
# ==============================================================#

fhs1_t_pft_ex19 <- 
  fread('~/Dropbox/BioLINCC files/Framingham cohort/datasets/CSV/T_PFT_EX19_0_0169D.csv',
        na.strings=c("NA","","NULL")) # Pulmonary function tests

fhs1_t_pft_ex19$visit <- 19

fhs1_melt_t_pft_ex19 <- 
  melt(fhs1_t_pft_ex19,
       id.vars=c("PID","IDTYPE","visit"),
       na.rm=T,
       factorsAsStrings=T)

fhs1_melt_t_pft_ex19$form <- "fhs1_t_pft_ex19"

rm(fhs1_t_pft_ex19)

# ==============================================================#
##          FHS original, sex hormones, exam 17-21             ##
# ==============================================================#

fhs1_shorm0 <- 
  fread('~/Dropbox/BioLINCC files/Framingham cohort/datasets/CSV/shorm0_21d.csv',
        na.strings=c("NA","","NULL")) 

fhs1_melt_shorm <- 
  melt(fhs1_shorm0,
       na.rm=T,
       factorsAsStrings=T,
       id.vars=c("PID","IDTYPE"))

fhs1_melt_shorm$var <-
  substr(fhs1_melt_shorm$variable,
         1,
         str_locate(fhs1_melt_shorm$variable,"_")-1)

fhs1_melt_shorm$visit <-
  substr(fhs1_melt_shorm$variable,
         str_locate(fhs1_melt_shorm$variable,"_")+1,
         str_length(fhs1_melt_shorm$variable))

fhs1_melt_shorm$variable <- fhs1_melt_shorm$var

fhs1_melt_shorm$form <- "fhs1_shorm0"

# =============================================================#
####       * FHS original, Assemble all analysis data *     ####
# =============================================================#

fhs1_melt_all <- rbind(fhs1_melt_ex0_7d[,c("PID","IDTYPE","visit","variable","value","form")],
                       fhs1_melt_ex0_8d[,c("PID","IDTYPE","visit","variable","value","form")],
                       fhs1_melt_ex0_9d[,c("PID","IDTYPE","visit","variable","value","form")],
                       fhs1_melt_ex0_10d[,c("PID","IDTYPE","visit","variable","value","form")],
                       fhs1_melt_ex0_11d[,c("PID","IDTYPE","visit","variable","value","form")],
                       fhs1_melt_ex0_12d[,c("PID","IDTYPE","visit","variable","value","form")],
                       fhs1_melt_ex0_13d[,c("PID","IDTYPE","visit","variable","value","form")],
                       fhs1_melt_ex0_14d[,c("PID","IDTYPE","visit","variable","value","form")],
                       fhs1_melt_ex0_15d[,c("PID","IDTYPE","visit","variable","value","form")],
                       fhs1_melt_ex0_16d[,c("PID","IDTYPE","visit","variable","value","form")],
                       fhs1_melt_ex0_17d[,c("PID","IDTYPE","visit","variable","value","form")],
                       fhs1_melt_ex0_18d[,c("PID","IDTYPE","visit","variable","value","form")],
                       fhs1_melt_ex0_19d[,c("PID","IDTYPE","visit","variable","value","form")],
                       fhs1_melt_ex0_20d[,c("PID","IDTYPE","visit","variable","value","form")],
                       fhs1_melt_ex0_21d[,c("PID","IDTYPE","visit","variable","value","form")],
                       fhs1_melt_ex0_22d[,c("PID","IDTYPE","visit","variable","value","form")],
                       fhs1_melt_ex0_23d[,c("PID","IDTYPE","visit","variable","value","form")],
                       fhs1_melt_ex0_24d[,c("PID","IDTYPE","visit","variable","value","form")],
                       fhs1_melt_ex0_25d[,c("PID","IDTYPE","visit","variable","value","form")],
                       fhs1_melt_ex0_26d[,c("PID","IDTYPE","visit","variable","value","form")],
                       fhs1_melt_ex0_27d[,c("PID","IDTYPE","visit","variable","value","form")],
                       fhs1_melt_ex0_28d[,c("PID","IDTYPE","visit","variable","value","form")],
                       fhs1_melt_ex0_29d[,c("PID","IDTYPE","visit","variable","value","form")],
                       fhs1_melt_ex0_30d[,c("PID","IDTYPE","visit","variable","value","form")],
                       fhs1_melt_ex0_31d[,c("PID","IDTYPE","visit","variable","value","form")],
                       fhs1_melt_ex0_32d[,c("PID","IDTYPE","visit","variable","value","form")],
                       fhs1_melt_l_fhslab1_ex20[,c("PID","IDTYPE","visit","variable","value","form")],
                       fhs1_melt_l_hba1c[,c("PID","IDTYPE","visit","variable","value","form")],
                       fhs1_melt_l_lipids_ex12[,c("PID","IDTYPE","visit","variable","value","form")],
                       fhs1_melt_t_echo_ex20[,c("PID","IDTYPE","visit","variable","value","form")],
                       fhs1_melt_t_echodop_ex20[,c("PID","IDTYPE","visit","variable","value","form")],
                       fhs1_melt_t_hrv_ex20[,c("PID","IDTYPE","visit","variable","value","form")],
                       fhs1_melt_t_pft_ex19[,c("PID","IDTYPE","visit","variable","value","form")],
                       fhs1_melt_vr_ffreq_ex20[,c("PID","IDTYPE","visit","variable","value","form")],
                       fhs1_melt_vr_ffreq_ex21[,c("PID","IDTYPE","visit","variable","value","form")],
                       fhs1_melt_vr_ffreq_ex22[,c("PID","IDTYPE","visit","variable","value","form")],
                       fhs1_melt_wkthru[,c("PID","IDTYPE","visit","variable","value","form")],
                       fhs1_melt_sex[,c("PID","IDTYPE","visit","variable","value","form")],
                       fhs1_melt_meno[,c("PID","IDTYPE","visit","variable","value","form")],
                       fhs1_melt_shorm[,c("PID","IDTYPE","visit","variable","value","form")],
                       fhs1_melt_bsni_2009[,c("PID","IDTYPE","visit","variable","value","form")],
                       fhs1_melt_physf_2005[,c("PID","IDTYPE","visit","variable","value","form")],
                       fhs1_melt_t_physfunc_2010[,c("PID","IDTYPE","visit","variable","value","form")])


fhs1_no_attend <- 
sqldf("select PID,
      visit, 
      value as att 
      from fhs1_melt_wkthru 
      where variable = 'ATT'"))

fhs1_melt_all <- merge(fhs1_melt_all,fhs1_no_attend,by=c("PID","visit"))
fhs1_melt_all <- merge(fhs1_melt_all,fhs1_wkthru[,c("PID","SEX")],by=c("PID"))

fhs1_melt_all <- subset(fhs1_melt_all,att==1)

fhs1_melt_all$variable <- as.character(fhs1_melt_all$variable)

#=========================================================================#
#                                                                         #
####            *** FRAMINGHAM Offspring/Omni 1 DATA ***               ####
#                                                                         #
#=========================================================================#


# ================================================================#
####  ** Framingham Offspring + OMNI 1 dates & outcomes**      ####
# ================================================================#

names(fhs2_dates) <- toupper(names(fhs2_dates))
names(fhs2_offspring_wkthru) <- toupper(names(fhs2_offspring_wkthru))

fhs2_melt_offspring_wkthru <- 
  # as.data.frame(
  melt(fhs2_offspring_wkthru,
       id.vars=c("PID","IDTYPE","SEX"),
       na.rm=T,
       factorsAsStrings=T)

fhs2_melt_offspring_wkthru$variable <-  as.character(fhs2_melt_offspring_wkthru$variable)

fhs2_melt_offspring_wkthru$variable[fhs2_melt_offspring_wkthru$variable=="RACE"] <-
  paste(fhs2_melt_offspring_wkthru$variable[fhs2_melt_offspring_wkthru$variable=="RACE"],"1",sep="")

fhs2_melt_offspring_wkthru$variable[fhs2_melt_offspring_wkthru$variable=="ETHNICITY"] <-
  paste(fhs2_melt_offspring_wkthru$variable[fhs2_melt_offspring_wkthru$variable=="ETHNICITY"],"1",sep="")


fhs2_melt_offspring_wkthru$visit  <- 
  substr(fhs2_melt_offspring_wkthru$variable,
         str_locate(fhs2_melt_offspring_wkthru$variable,"\\d")[,1],
         nchar(fhs2_melt_offspring_wkthru$variable))

fhs2_melt_offspring_wkthru$variable  <- 
  substr(fhs2_melt_offspring_wkthru$variable,
         1,str_locate(fhs2_melt_offspring_wkthru$variable,"\\d")[,1]-1)


fhs2_melt_wkthru <- as.data.frame(fhs2_melt_offspring_wkthru)

fhs2_melt_wkthru$value[fhs2_melt_wkthru$variable=="WAIST"] <- 
  as.character(as.numeric(fhs2_melt_wkthru$value[fhs2_melt_wkthru$variable=="WAIST"]) * 2.54)

fhs2_melt_wkthru_kg <- subset(fhs2_melt_wkthru,variable=="WGT")
fhs2_melt_wkthru_kg$value <- as.numeric(fhs2_melt_wkthru_kg$value)/2.20462
fhs2_melt_wkthru_kg$variable <- "WGT_KG"
fhs2_melt_wkthru <- 
  as.data.table(
    rbind(fhs2_melt_wkthru,
          fhs2_melt_wkthru_kg))


fhs2_wkthru <- dcast(fhs2_melt_wkthru,...~variable)


fhs2_melt_wkthru$form <- "fhs2_wkthru"


#================================#
#### *** Offspring exam 1 *** ####
#================================#

fhs2_ex1_1d <- fread('~/Dropbox/BioLINCC files/Framingham offspring/datasets/CSV/ex1_1d_v4.csv',na.strings=c("NA","","NULL"))  # Offspring, Exam 1
fhs2_ex1_1d$visitdays <- 0
fhs2_ex1_1d$visit <- 1
names(fhs2_ex1_1d)[names(fhs2_ex1_1d)=="idtype"] <- "IDTYPE"

fhs2_ex1_1d$drinks_wk <- 
  fhs2_ex1_1d$A111+
  fhs2_ex1_1d$A112+
  fhs2_ex1_1d$A113

fhs2_ex1_1d$A89[!between(fhs2_ex1_1d$A89,18,55)] <- NA

fhs2_ex1_1d$A89


fhs2_melt_ex1_1d <- 
  melt(fhs2_ex1_1d,
       id.vars=c("PID",
                 "IDTYPE",
                 "visit",
                 "visitdays"), 
       na.rm=T, 
       factorsAsStrings=T)

fhs2_melt_ex1_1d$form <- "fhs2_offspring_ex01"

rm(fhs2_ex1_1d)

#==========================================#
#.    Renin activity, Offspring Exam 1     #
#==========================================#

fhs2_renin1 <- fread('~/Dropbox/BioLINCC files/Framingham offspring/datasets/CSV/renin1_6d.csv',na.strings=c("NA","","NULL"))
names(fhs2_renin1)[names(fhs2_renin1)=="idtype"] <- "IDTYPE"
fhs2_renin1$visit <- 1
fhs2_melt_renin1 <- melt(fhs2_renin1,id.vars=c("PID","IDTYPE","visit"), na.rm=T, factorsAsStrings=T)
fhs2_melt_renin1$form <- "fhs2_renin1"

rm(fhs2_renin1)


#==================================#
####  *** Offspring exam 2 ***  ####
#==================================#

fhs2_ex1_2d <- 
  fread('~/Dropbox/BioLINCC files/Framingham offspring/datasets/CSV/ex1_2d_v4.csv',
        na.strings=c("NA","","NULL"))  # Offspring, Exam 2

names(fhs2_ex1_2d)[names(fhs2_ex1_2d)=="idtype"] <- "IDTYPE"
fhs2_ex1_2d$visit <- 2
fhs2_ex1_2d <- 
  merge(fhs2_ex1_2d,
        fhs2_dates[,c("PID","IDTYPE","SEX")],
        by=c("PID","IDTYPE"))

fhs2_ex1_2d$B439_cm <- fhs2_ex1_2d$B439/10  # IVS
fhs2_ex1_2d$B443_cm <- fhs2_ex1_2d$B443/10  # LVPWd
fhs2_ex1_2d$B448_cm <- fhs2_ex1_2d$B448/10  # LVIDD


fhs2_ex1_2d <- 
  as.data.table(
    calc_hypertrophy_type(
      as.data.frame(fhs2_ex1_2d), 
      id = "PID",
      sex="SEX",
      male="1",
      female="2",
      lvedd="B448_cm",
      ivsd="B439_cm",
      lvpwtd="B443_cm",
      height="B14",
      weight="B13"))

fhs2_ex1_2d$fhs_pai <- 
  calc_framingham_pai(
    as.data.frame(fhs2_ex1_2d),
    slp_hrs = "B104",
    sed_hrs = "B105",
    slgt_hrs = "B106",
    mod_hrs = "B107",
    hvy_hrs = "B108")

fhs2_ex1_2d$drinks_wk <- fhs2_ex1_2d$B117+
  fhs2_ex1_2d$B118+
  fhs2_ex1_2d$B119

fhs2_ex1_2d$B71[!between(fhs2_ex1_2d$B71,19,58)] <- NA
fhs2_ex1_2d$B75[!between(fhs2_ex1_2d$B75,7,22)] <- NA
fhs2_ex1_2d$B76[!between(fhs2_ex1_2d$B76,1,11)] <- NA
fhs2_ex1_2d$B77[!between(fhs2_ex1_2d$B77,20,53)] <- NA
fhs2_ex1_2d$B80[!between(fhs2_ex1_2d$B80,1,16)] <- NA
fhs2_ex1_2d$B82[!between(fhs2_ex1_2d$B82,1,23)] <- NA


fhs2_melt_ex1_2d <- 
  as.data.table(
    melt(fhs2_ex1_2d,
         id.vars=c("PID","IDTYPE","visit"), 
         na.rm=T, 
         factorsAsStrings=T))

fhs2_melt_ex1_2d$form <- "fhs2_offspring_ex02"

rm(fhs2_ex1_2d)

#========================#
####    CRP, Exam 2      #
#========================#


fhs2_l_crp_ex02 <- 
  fread('~/Dropbox/BioLINCC files/Framingham offspring/datasets/CSV/l_crp_ex02_1_0089d.csv',
        na.strings=c("NA","","NULL"))

names(fhs2_l_crp_ex02)[names(fhs2_l_crp_ex02)=="idtype"] <- "IDTYPE"
fhs2_l_crp_ex02$visit <- 2
fhs2_melt_l_crp_ex02 <- 
  melt(fhs2_l_crp_ex02,
       id.vars=c("PID","IDTYPE","visit"), 
       na.rm=T, 
       factorsAsStrings=T)
fhs2_melt_l_crp_ex02$form <- "fhs2_crp_ex02"


rm(fhs2_l_crp_ex02)


#================================#
#### *** Offspring exam 3 *** ####
#================================#


fhs2_ex1_3d <- 
  fread('~/Dropbox/BioLINCC files/Framingham offspring/datasets/CSV/ex1_3d_v1.csv',
        na.strings=c("NA","","NULL"))  # Offspring, Exam 3

names(fhs2_ex1_3d)[names(fhs2_ex1_3d)=="idtype"] <- "IDTYPE"

fhs2_ex1_3d$visit <- 3

fhs2_ex1_3d$drinks_wk <- fhs2_ex1_3d$C83+
  fhs2_ex1_3d$C86+
  fhs2_ex1_3d$C89

fhs2_ex1_3d$C48[!between(fhs2_ex1_3d$C48,19,60)] <- NA
fhs2_ex1_3d$C50[!between(fhs2_ex1_3d$C50,20,57)] <- NA
fhs2_ex1_3d$C52[!between(fhs2_ex1_3d$C52,1,11)] <- NA
fhs2_ex1_3d$C53[!between(fhs2_ex1_3d$C53,20,51)] <- NA

fhs2_melt_ex1_3d <- 
  melt(fhs2_ex1_3d,
       id.vars=c("PID","IDTYPE","visit"), 
       na.rm=T, 
       factorsAsStrings=T)

fhs2_melt_ex1_3d$form <- "fhs2_offspring_ex03"

rm(fhs2_ex1_3d)

#=========================================================#
####  Food frequency questionnaire, Offspring, Exam 3. ---#
#=========================================================#

fhs2_q_ffreq_ex03 <- fread('~/Dropbox/BioLINCC files/Framingham offspring/datasets/CSV/q_ffreq_ex03_1_0302d.csv',na.strings=c("NA","","NULL"))
names(fhs2_q_ffreq_ex03)[names(fhs2_q_ffreq_ex03)=="idtype"] <- "IDTYPE"
fhs2_q_ffreq_ex03$visit <- 3

fhs2_melt_q_ffreq_ex03 <- 
  melt(fhs2_q_ffreq_ex03,
       id.vars=c("PID","IDTYPE","visit"), 
       na.rm=T, 
       factorsAsStrings=T)

fhs2_melt_q_ffreq_ex03$form <- "fhs2_q_ffreq"

rm(fhs2_q_ffreq_ex03)

#================================================#
####  Psych questionnaire, Offspring, Exam 3  ---#
#================================================#

fhs2_q_psych_ex03<- 
  fread('~/Dropbox/BioLINCC files/Framingham offspring/datasets/CSV/q_psych_ex03_1_0167d.csv',
        na.strings=c("NA","","NULL"))

names(fhs2_q_psych_ex03)[names(fhs2_q_psych_ex03)=="idtype"] <- "IDTYPE"

fhs2_q_psych_ex03$PY109[is.na(fhs2_q_psych_ex03$PY109)] <- 0

fhs2_q_psych_ex03$visit <- 3
fhs2_melt_q_psych_ex03 <- 
  melt(fhs2_q_psych_ex03,
       id.vars=c("PID","IDTYPE","visit"), 
       na.rm=T, 
       factorsAsStrings=T)

fhs2_melt_q_psych_ex03$form <- "fhs2_q_psych_ex03"

rm(fhs2_q_psych_ex03)


#====================================================#
####  Heart rate variability, Offspring - Exam 3  ---#
#====================================================#

fhs2_t_hrv <- fread('~/Dropbox/BioLINCC files/Framingham offspring/datasets/CSV/t_hrv_1987_m_0118d.csv',na.strings=c("NA","","NULL"))
names(fhs2_t_hrv) <- toupper(names(fhs2_t_hrv))
fhs2_t_hrv$visit <- fhs2_t_hrv$HRV_EXAM
fhs2_melt_t_hrv <- melt(fhs2_t_hrv,id.vars=c("PID","IDTYPE","visit"), na.rm=T, factorsAsStrings=T)
fhs2_melt_t_hrv$form <- "fhs2_t_hrv"

rm(fhs2_t_hrv)


#================================================================#
####          Sex hormones, Offspring, Exam 3                 ---#
#================================================================#

fhs2_shorm_ex03 <- fread('~/Dropbox/BioLINCC files/Framingham Offspring/datasets/CSV/l_shorm_ex03_1_0177d.csv')
fhs2_melt_shorm_ex03 <-  melt(fhs2_shorm_ex03,id.vars=c("PID","IDTYPE"),na.rm=T,factorsAsStrings = T)
fhs2_melt_shorm_ex03$visit <- 3
fhs2_melt_shorm_ex03$form <- "fhs2_shorm_ex03"

rm(fhs2_shorm_ex03)


#==================================#
####  *** Offspring exam 4 ***  ####
#==================================#

fhs2_ex1_4d <- fread('~/Dropbox/BioLINCC files/Framingham offspring/datasets/CSV/ex1_4d_v1.csv',na.strings=c("NA","","NULL"))  # Offspring, Exam 4
names(fhs2_ex1_4d)[names(fhs2_ex1_4d)=="idtype"] <- "IDTYPE"
fhs2_ex1_4d$visit <- 4

fhs2_ex1_4d$drinks_wk <- fhs2_ex1_4d$D082+
  fhs2_ex1_4d$D086+
  fhs2_ex1_4d$D088

fhs2_ex1_4d$fhs_pai <- 
  calc_framingham_pai(
    as.data.frame(fhs2_ex1_4d),
    slp_hrs = "D426",
    sed_hrs = "D427",
    slgt_hrs = "D428",
    mod_hrs = "D429",
    hvy_hrs = "D430")


fhs2_ex1_4d$D054[!between(fhs2_ex1_4d$D054,23,75)] <- NA
fhs2_ex1_4d$D056[!between(fhs2_ex1_4d$D056,19,81)] <- NA
fhs2_ex1_4d$D058[!between(fhs2_ex1_4d$D058,1,11)] <- NA
fhs2_ex1_4d$D059[!between(fhs2_ex1_4d$D059,19,61)] <- NA


fhs2_melt_ex1_4d <- 
  as.data.table(
    melt(fhs2_ex1_4d,
         id.vars=c("PID","IDTYPE","visit"),
         na.rm=T, 
         factorsAsStrings=T))

fhs2_melt_ex1_4d$form <- "fhs2_offspring_ex04"

rm(fhs2_ex1_4d)

#==========================#
####    Echo, Exam 4    ---#
#==========================#

fhs2_t_echo_ex04 <- fread('~/Dropbox/BioLINCC files/Framingham offspring/datasets/CSV/t_echo_ex04_1_0101d.csv',na.strings=c("NA","","NULL"))
names(fhs2_t_echo_ex04)[names(fhs2_t_echo_ex04)=="idtype"] <- "IDTYPE"
fhs2_t_echo_ex04$visit <- 4

fhs2_meas4 <-as.data.table(sqldf("select PID,
                   IDTYPE,
                   SEX,
                   HGT1*2.54 as HEIGHT,
                   WGT4/2.2 as WEIGHT
                   from fhs2_offspring_wkthru"))

fhs2_t_echo_ex04 <- 
  merge(fhs2_t_echo_ex04,
        fhs2_meas4,
        by=c("PID","IDTYPE"),
        all.x=T)

fhs2_t_echo_ex04 <- 
  as.data.table(
    calc_hypertrophy_type(
      as.data.frame(fhs2_t_echo_ex04), 
      id = "PID",
      sex="SEX",
      lvedd="LVDD4",
      ivsd="IVSD4",
      lvpwtd="LVPD4",
      height="HEIGHT",
      weight="WEIGHT"))

fhs2_melt_t_echo_ex04 <- 
  as.data.table(
    melt(fhs2_t_echo_ex04,
         id.vars=c("PID","IDTYPE","visit"), 
         na.rm=T, 
         factorsAsStrings=T))

fhs2_melt_t_echo_ex04$form <- "fhs2_echo_ex04"

rm(fhs2_t_echo_ex04,
   fhs2_meas4)


#================================================================#
####          Sex hormones, Offspring, Exam 4                 ---#
#================================================================#

fhs2_shorm_ex04 <- fread('~/Dropbox/BioLINCC files/Framingham Offspring/datasets/CSV/l_shorm_ex04_1_0178d.csv')
fhs2_melt_shorm_ex04 <-  
  melt(fhs2_shorm_ex04,
       id.vars=c("PID","IDTYPE"),
       na.rm=T,
       factorsAsStrings = T)
fhs2_melt_shorm_ex04$visit <- 4
fhs2_melt_shorm_ex04$form <- "fhs2_shorm_ex04"

rm(fhs2_shorm_ex04)


#=================================#
####  *** Offspring exam 5 *** ####
#=================================#

fhs2_ex1_5d <- fread('~/Dropbox/BioLINCC files/Framingham offspring/datasets/CSV/ex1_5d_v1.csv',na.strings=c("NA","","NULL"))  # Offspring, Exam 5
names(fhs2_ex1_5d)[names(fhs2_ex1_5d)=="idtype"] <- "IDTYPE"
fhs2_ex1_5d$visit <- 5

fhs2_ex1_5d$drinks_wk <- 
  fhs2_ex1_5d$E310+
  fhs2_ex1_5d$E313+
  fhs2_ex1_5d$E316

fhs2_ex1_5d$E274[!between(fhs2_ex1_5d$E274,19,61)] <- NA
fhs2_ex1_5d$E276[!between(fhs2_ex1_5d$E276,19,67)] <- NA
fhs2_ex1_5d$E278[!between(fhs2_ex1_5d$E278,0,11)] <- NA
fhs2_ex1_5d$E279[!between(fhs2_ex1_5d$E279,19,51)] <- NA

fhs2_melt_ex1_5d <- 
  melt(fhs2_ex1_5d,
       id.vars=c("PID","IDTYPE","visit"), 
       na.rm=T, 
       factorsAsStrings=T)

fhs2_melt_ex1_5d$form <- "fhs2_offspring_ex05"

rm(fhs2_ex1_5d)


#=============================#
####   Lab panel, exam 5   ---#
#=============================#

fhs2_l_fhslab_ex05 <- 
  fread('~/Dropbox/BioLINCC files/Framingham offspring/datasets/CSV/l_fhslaba_ex05_1_0578d.csv',
        na.strings=c("NA","","NULL"))

names(fhs2_l_fhslab_ex05)[names(fhs2_l_fhslab_ex05)=="idtype"] <- "IDTYPE"

fhs2_l_fhslab_ex05$visit <- 5

fhs2_melt_l_fhslab_ex05 <- 
  melt(fhs2_l_fhslab_ex05,
       id.vars=c("PID","IDTYPE","visit"), 
       na.rm=T, 
       factorsAsStrings=T)

fhs2_melt_l_fhslab_ex05$form <- "fhs2_fhslab_ex05"

rm(fhs2_l_fhslab_ex05)


# ============================================##
####     Physical activity, exam 5          ---#
# ============================================##

fhs2_act1_5d <- 
  fread('~/Dropbox/BioLINCC files/Framingham offspring/datasets/CSV/act1_5d_v1.csv',
        na.strings=c("NA","","NULL")) # Physical exam questionnaire, exam 5

names(fhs2_act1_5d)[3] <- "visit"
fhs2_act1_5d$visit <- 5

fhs2_act1_5d$totkcd <- fhs2_act1_5d$TOTKCW/7

fhs2_act1_5d$fhs_pai <- 
  calc_framingham_pai(
    as.data.frame(fhs2_act1_5d),
    slp_hrs="EH_10A",
    sed_hrs="EH_10B",
    slgt_hrs="EH_10C",
    mod_hrs="EH_10D",
    hvy_hrs="EH_10E")

fhs2_melt_act1_5d <- melt(fhs2_act1_5d,
                          id.vars=c("PID","IDTYPE","visit"),
                          na.rm=T,
                          factorsAsStrings=T)

fhs2_melt_act1_5d$form <- "fhs2_act1_5"

rm(fhs2_act1_5d)


#===========================#
####     Echo, Exam 5    ---#
#===========================#

fhs2_t_echo_ex05 <- 
  fread('~/Dropbox/BioLINCC files/Framingham offspring/datasets/CSV/t_echo_ex05_1_0206d.csv',
        na.strings=c("NA","","NULL"))

names(fhs2_t_echo_ex05)[names(fhs2_t_echo_ex05)=="idtype"] <- "IDTYPE"
fhs2_t_echo_ex05$visit <- 5


fhs2_meas5 <- 
  as.data.table(sqldf("select PID,
                   IDTYPE,
                   SEX,
                   HGT1*2.54 as HEIGHT,
                   WGT5/2.2 as WEIGHT
                   from fhs2_offspring_wkthru"))

fhs2_t_echo_ex05 <- merge(fhs2_t_echo_ex05,fhs2_meas5,by=c("PID","IDTYPE"),all.x=T)

fhs2_t_echo_ex05 <- 
  as.data.table(
    calc_hypertrophy_type(
      as.data.frame(fhs2_t_echo_ex05), 
      id = "PID",
      sex="SEX",
      lvedd="LVDD5",
      ivsd="IVSD5",
      lvpwtd="LVPD5",
      height="HEIGHT",
      weight="WEIGHT"))

fhs2_melt_t_echo_ex05 <- 
  as.data.table(
    melt(
      fhs2_t_echo_ex05,
      id.vars=c("PID","IDTYPE","visit"), 
      na.rm=T, 
      factorsAsStrings=T))

fhs2_melt_t_echo_ex05$form <- "fhs2_echo_ex05"

rm(fhs2_t_echo_ex05,
   fhs2_meas5)


#=============================================#
####  Echo mitral valve prolapse, Exam 5   ---#
#=============================================#

fhs2_t_echomvp_ex05 <- fread('~/Dropbox/BioLINCC files/Framingham offspring/datasets/CSV/t_echomvp_ex05_1_0753d.csv',na.strings=c("NA","","NULL"))
names(fhs2_t_echomvp_ex05)[names(fhs2_t_echomvp_ex05)=="idtype"] <- "IDTYPE"
fhs2_t_echomvp_ex05$visit <- 5
fhs2_melt_t_echomvp_ex05 <- melt(fhs2_t_echomvp_ex05,id.vars=c("PID","IDTYPE","visit"), na.rm=T, factorsAsStrings=T)
fhs2_melt_t_echomvp_ex05$form <- "fhs2_t_echomvp_ex05"

rm(fhs2_t_echomvp_ex05)


#=================================================#
####    Validated food frequency data, Exam 5  ---#
#=================================================#

fhs2_vr_ffreq_ex05 <- fread('~/Dropbox/BioLINCC files/Framingham offspring/datasets/CSV/vr_ffreq_ex05_1_0575d.csv',na.strings=c("NA","","NULL"))
names(fhs2_vr_ffreq_ex05)<- toupper(names(fhs2_vr_ffreq_ex05))
fhs2_vr_ffreq_ex05$visit <- 5

fhs2_melt_vr_ffreq_ex05 <- melt(fhs2_vr_ffreq_ex05,id.vars=c("PID","IDTYPE","visit"), na.rm=T, factorsAsStrings=T)
fhs2_melt_vr_ffreq_ex05$form <- "fhs2_vr_ffreq"

rm(fhs2_vr_ffreq_ex05)


#================================#
#### *** Offspring exam 6 *** ####
#================================#

fhs2_ex1_6d <- 
  fread('~/Dropbox/BioLINCC files/Framingham offspring/datasets/CSV/ex1_6d_v1.csv',
        na.strings=c("NA","","NULL"))  # Offspring, Exam 6

names(fhs2_ex1_6d)[names(fhs2_ex1_6d)=="idtype"] <- "IDTYPE"
fhs2_ex1_6d$visit <- 6

fhs2_ex1_6d$drinks_wk <- fhs2_ex1_6d$F276+
  fhs2_ex1_6d$F279+
  fhs2_ex1_6d$F285

fhs2_ex1_6d$CESD20_missing <- rowSums(is.na(fhs2_ex1_6d[,67:86]))
fhs2_ex1_6d$CESD20 <- rowSums(fhs2_ex1_6d[,67:86])
fhs2_ex1_6d$CESD20[fhs2_ex1_6d$CESD20_missing >= 4] <- NA

fhs2_ex1_6d$F238[!between(fhs2_ex1_6d$F238,20,67)] <- NA
fhs2_ex1_6d$F242[!between(fhs2_ex1_6d$F242,20,81)] <- NA
fhs2_ex1_6d$F244[!between(fhs2_ex1_6d$F242,1,11)] <- NA
fhs2_ex1_6d$F245[!between(fhs2_ex1_6d$F245,19,51)] <- NA


fhs2_melt_ex1_6d <- melt(fhs2_ex1_6d,id.vars=c("PID","IDTYPE","visit"), na.rm=T, factorsAsStrings=T)
fhs2_melt_ex1_6d$form <- "fhs2_offspring_ex06"

rm(fhs2_ex1_6d)

#==============================#
####  Childbirth, Exam 6    ---#
#==============================#

fhs2_ex1_bwgt <- 
  fread('~/Dropbox/BioLINCC files/Framingham offspring/datasets/CSV/bwgt1_6d_v1.csv',
        na.strings=c("NA","","NULL"))  
fhs2_ex1_bwgt$visit <- 6

fhs2_ex1_bwgt$age_lastbirth <- 
  fhs2_ex1_bwgt$BW79

fhs2_ex1_bwgt$age_lastbirth[is.na(fhs2_ex1_bwgt$age_lastbirth)] <- 
  fhs2_ex1_bwgt$BW79[is.na(fhs2_ex1_bwgt$age_lastbirth)]

fhs2_ex1_bwgt$age_lastbirth[is.na(fhs2_ex1_bwgt$age_lastbirth)] <- 
  fhs2_ex1_bwgt$BW66[is.na(fhs2_ex1_bwgt$age_lastbirth)]

fhs2_ex1_bwgt$age_lastbirth[is.na(fhs2_ex1_bwgt$age_lastbirth)] <- 
  fhs2_ex1_bwgt$BW53[is.na(fhs2_ex1_bwgt$age_lastbirth)]

fhs2_ex1_bwgt$age_lastbirth[is.na(fhs2_ex1_bwgt$age_lastbirth)] <- 
  fhs2_ex1_bwgt$BW40[is.na(fhs2_ex1_bwgt$age_lastbirth)]

fhs2_ex1_bwgt$age_lastbirth[is.na(fhs2_ex1_bwgt$age_lastbirth)] <- 
  fhs2_ex1_bwgt$BW27[is.na(fhs2_ex1_bwgt$age_lastbirth)]


fhs2_melt_ex1_bwgt <- melt(fhs2_ex1_bwgt,id.vars=c("PID","IDTYPE","visit"), na.rm=T, factorsAsStrings=T)
fhs2_melt_ex1_bwgt$form <- "fhs2_offspring_bwgt"

rm(fhs2_ex1_bwgt)


#==============================#
####  Aldosterone, Exam 6   ---#
#==============================#

fhs2_l_aldost_ex06 <- fread('~/Dropbox/BioLINCC files/Framingham offspring/datasets/CSV/l_aldost_ex06_1b_0046d_v1.csv',na.strings=c("NA","","NULL"))
names(fhs2_l_aldost_ex06)[names(fhs2_l_aldost_ex06)=="idtype"] <- "IDTYPE"
fhs2_l_aldost_ex06$visit <- 6
fhs2_melt_l_aldost_ex06 <- melt(fhs2_l_aldost_ex06,id.vars=c("PID","IDTYPE","visit"), na.rm=T, factorsAsStrings=T)
fhs2_melt_l_aldost_ex06$form <- "fhs2_offspring_aldost_ex06"

rm(fhs2_l_aldost_ex06)

# ========================================##
####    Physical Activity, Exam 6       ---#
# ========================================##

fhs2_act1_6d <- fread('~/Dropbox/BioLINCC files/Framingham offspring/datasets/CSV/act1_6d_v1.csv',na.strings=c("NA","","NULL")) # Physical exam questionnaire, exam 6
fhs2_act1_6d$visit <- 6
fhs2_melt_act1_6d <- melt(fhs2_act1_6d,
                          id.vars=c("PID",
                                    "IDTYPE",
                                    "visit"),
                          na.rm=T,
                          factorsAsStrings=T)
fhs2_melt_act1_6d$form <- "fhs2_act1_6"

rm(fhs2_act1_6d)


#=============================#
####   Lab Panel, Exam 6   ---#
#=============================#

fhs2_l_fhslab_ex06 <- fread('~/Dropbox/BioLINCC files/Framingham offspring/datasets/CSV/l_fhslaba_ex06_1_0579d.csv',na.strings=c("NA","","NULL"))
names(fhs2_l_fhslab_ex06)[names(fhs2_l_fhslab_ex06)=="idtype"] <- "IDTYPE"
fhs2_l_fhslab_ex06$visit <- 6
fhs2_melt_l_fhslab_ex06 <- melt(fhs2_l_fhslab_ex06,id.vars=c("PID","IDTYPE","visit"), na.rm=T, factorsAsStrings=T)
fhs2_melt_l_fhslab_ex06$form <- "fhs2_fhslab_ex06"

rm(fhs2_l_fhslab_ex06)

#=======================================#
####  Natriuretic peptide, Exam 6    ---#
#=======================================#

fhs2_natpep1 <- fread('~/Dropbox/BioLINCC files/Framingham offspring/datasets/CSV/natpep1_6d_v1.csv',na.strings=c("NA","","NULL"))
names(fhs2_natpep1)[names(fhs2_natpep1)=="idtype"] <- "IDTYPE"
fhs2_natpep1$visit <- 6
fhs2_melt_natpep1 <- melt(fhs2_natpep1,id.vars=c("PID","IDTYPE","visit"), na.rm=T, factorsAsStrings=T)
fhs2_melt_natpep1$form <- "fhs2_natpep1_6"


rm(fhs2_natpep1)

# ================================================#
####               Galectin Exam 6             ---#
# ================================================#


fhs2_l_gal3_ex06 <- fread('~/Dropbox/BioLINCC files/Framingham offspring/datasets/CSV/l_gal3_ex06_1_0623d.csv',na.strings=c("NA","","NULL"))
fhs2_l_gal3_ex06$visit <- 6

fhs2_melt_l_gal3_ex06 <- melt(fhs2_l_gal3_ex06,id.vars=c("PID","IDTYPE","visit"), na.rm=T, factorsAsStrings=T)
fhs2_melt_l_gal3_ex06$form <- 'fhs2_gal3_ex06'

rm(fhs2_l_gal3_ex06)

#==================================#
####  CKD biomarkers, Exam 6    ---#
#==================================#

fhs2_ckd_ex06_1 <- fread('~/Dropbox/BioLINCC files/Framingham offspring/datasets/CSV/l_ckd_ex06_1_0536d.csv',na.strings=c("NA","","NULL"))
names(fhs2_ckd_ex06_1)[names(fhs2_ckd_ex06_1)=="idtype"] <- "IDTYPE"
fhs2_ckd_ex06_1$visit <- 6
fhs2_melt_l_ckd_ex06 <- melt(fhs2_ckd_ex06_1,id.vars=c("PID","IDTYPE","visit"), na.rm=T, factorsAsStrings=T)
fhs2_melt_l_ckd_ex06$form <- "fhs2_ckd_ex06"

rm(fhs2_ckd_ex06_1)

#================================#
####  Echo, Exam 6            ---#
#================================#

fhs2_t_echo_ex06 <- fread('~/Dropbox/BioLINCC files/Framingham offspring/datasets/CSV/t_echo_ex06_1b_0207d_v2.csv',na.strings=c("NA","","NULL"))
names(fhs2_t_echo_ex06)[names(fhs2_t_echo_ex06)=="idtype"] <- "IDTYPE"
fhs2_t_echo_ex06$visit <- 6


fhs2_meas6 <- 
  as.data.table(
    sqldf("select PID,
                   IDTYPE,
                   SEX,
                   HGT1*2.54 as HEIGHT,
                   WGT6/2.2 as WEIGHT
                   from fhs2_offspring_wkthru"))

fhs2_t_echo_ex06 <- merge(fhs2_t_echo_ex06,fhs2_meas6,by=c("PID","IDTYPE"),all.x=T)

fhs2_t_echo_ex06 <- 
  as.data.table(
    calc_hypertrophy_type(
      fhs2_t_echo_ex06, 
      id = "PID",
      sex="SEX",
      lvedd="LMLVD_DV",
      ivsd="LMIVS_DV",
      lvpwtd="LMLVP_DV",
      height="HEIGHT",
      weight="WEIGHT"))

fhs2_melt_t_echo_ex06 <- melt(fhs2_t_echo_ex06,id.vars=c("PID","IDTYPE","visit"), na.rm=T, factorsAsStrings=T)
fhs2_melt_t_echo_ex06$form <- "fhs2_echo_ex06"

rm(fhs2_t_echo_ex06,
   fhs2_meas6)


#==============================#
####       CRP, Exam 6      ---#
#==============================#


fhs2_l_crp_ex06 <- fread('~/Dropbox/BioLINCC files/Framingham offspring/datasets/CSV/l_crp_ex06_1_0091d.csv',na.strings=c("NA","","NULL"))
names(fhs2_l_crp_ex06)[names(fhs2_l_crp_ex06)=="idtype"] <- "IDTYPE"
fhs2_l_crp_ex06$visit <- 6
fhs2_melt_l_crp_ex06 <- melt(fhs2_l_crp_ex06,id.vars=c("PID","IDTYPE","visit"), na.rm=T, factorsAsStrings=T)
fhs2_melt_l_crp_ex06$form <- "fhs2_crp_ex06"

rm(fhs2_l_crp_ex06)


#=================================================#
####  Validated food frequency data, Exam 6    ---#
#=================================================#

fhs2_vr_ffreq_ex06 <- fread('~/Dropbox/BioLINCC files/Framingham offspring/datasets/CSV/vr_ffreq_ex06_1_0506d.csv',na.strings=c("NA","","NULL"))
names(fhs2_vr_ffreq_ex06)[names(fhs2_vr_ffreq_ex06)=="idtype"] <- "IDTYPE"
fhs2_vr_ffreq_ex06$visit <- 6

fhs2_melt_vr_ffreq_ex06 <- melt(fhs2_vr_ffreq_ex06,id.vars=c("PID","IDTYPE","visit"), na.rm=T, factorsAsStrings=T)
fhs2_melt_vr_ffreq_ex06$form <- "fhs2_vr_ffreq"

rm(fhs2_vr_ffreq_ex06)


#================================#
#### *** Offspring exam 7 *** ####
#================================#

fhs2_ex1_7d <- fread('~/Dropbox/BioLINCC files/Framingham offspring/datasets/CSV/ex1_7d_v2.csv',na.strings=c("NA","","NULL"))   # Offspring, Exam 7
names(fhs2_ex1_7d)[names(fhs2_ex1_7d)=="idtype"] <- "IDTYPE"


fhs2_obsper_2005 <- fread('~/Dropbox/BioLINCC files/Framingham offspring/datasets/CSV/t_physf_2005_m_0162d_v2.csv',na.strings=c("NA","","NULL"))  # Omni 1, Exam 1
names(fhs2_obsper_2005)[names(fhs2_obsper_2005)=="idtype"] <- "IDTYPE"

offspring_30 <- fhs_dates_long[fhs_dates_long$cohort_name=="Offspring"&fhs_dates_long$visit_yr==30,c("PID","visitdays")]
offspring_35 <- fhs_dates_long[fhs_dates_long$cohort_name=="Offspring"&fhs_dates_long$visit_yr==35,c("PID","visitdays")]

names(offspring_30)[2] <- "visitdays_30"
names(offspring_35)[2] <- "visitdays_35"

fhs2_obsper_2005 <- 
  merge(fhs2_obsper_2005,
        offspring_30,
        by="PID",
        all.x=T)

fhs2_obsper_2005 <- 
  merge(fhs2_obsper_2005,
        offspring_35,
        by="PID",
        all.x=T)


fhs2_obsper_2005_ex30 <- 
  as.data.table(sqldf("select * 
        from fhs2_obsper_2005
        where mridate >= visitdays_30
        and mridate < visitdays_35
        group by PID,IDTYPE
        having min(mridate)"))


fhs2_ex1_7d <- merge(fhs2_ex1_7d,
                     fhs2_obsper_2005_ex30,
                     by=c("PID","IDTYPE"),
                     all.x=T)


fhs2_survca_first <- 
as.data.table(
sqldf("select PID, IDTYPE, D_DATE
                           from fhs2_cancer
                           where TOPO <> 173
                           group by PID, IDTYPE
                           having MIN(D_DATE)"))


fhs2_ex1_7d$drinks_wk <- 
  fhs2_ex1_7d$G104+
  fhs2_ex1_7d$G107+
  fhs2_ex1_7d$G110+
  fhs2_ex1_7d$G113

fhs2_ex1_7d$bsnq_total <- 
  fhs2_ex1_7d$G645+
  fhs2_ex1_7d$G646+
  fhs2_ex1_7d$G647+
  fhs2_ex1_7d$G648+
  fhs2_ex1_7d$G649+
  fhs2_ex1_7d$G650+
  fhs2_ex1_7d$G651+
  fhs2_ex1_7d$G652+
  fhs2_ex1_7d$G653+
  fhs2_ex1_7d$G654+
  fhs2_ex1_7d$G655+
  fhs2_ex1_7d$G656+
  fhs2_ex1_7d$G657

fhs2_ex1_7d$G078[!between(fhs2_ex1_7d$G078,22,62)] <- NA
fhs2_ex1_7d$G083[!between(fhs2_ex1_7d$G083,20,83)] <- NA
fhs2_ex1_7d$G085[!between(fhs2_ex1_7d$G085,1,11)] <- NA
fhs2_ex1_7d$G086[!between(fhs2_ex1_7d$G086,21,47)] <- NA

fhs2_ex1_7d$G092[between(fhs2_ex1_7d$G086,1,40)&!is.na(fhs2_ex1_7d$G086)] <-
  fhs2_ex1_7d$G092[between(fhs2_ex1_7d$G086,1,40)&!is.na(fhs2_ex1_7d$G086)]*12


fhs2_ex1_7d <- merge(fhs2_ex1_7d,
                     fhs2_offspring_wkthru[,c("PID",
                                              "AGE7",
                                              "SEX",
                                              "DMRX7",
                                              "HRX7",
                                              "DATE7",
                                              "HGT1",
                                              "WGT7",
                                              "BMI7",
                                              "CREAT7",
                                              "WGT6")],
                     by=c("PID") )


fhs2_ex1_7d <- merge(fhs2_ex1_7d, 
                     fhs_survcvd,
                     by=c("PID","IDTYPE"),
                     all.x=T)

fhs2_ex1_7d <- merge(fhs2_ex1_7d, 
                     fhs_survstk,
                     by=c("PID","IDTYPE"),
                     all.x=T)

fhs2_ex1_7d <- merge(fhs2_ex1_7d,
                     fhs2_survca_first,
                     by=c("PID","IDTYPE"),
                     all.x=T)


fhs2_ex1_7d$CESD20_missing <- rowSums(is.na(fhs2_ex1_7d[,448:467]))
fhs2_ex1_7d$CESD20 <- rowSums(fhs2_ex1_7d[,448:467])
fhs2_ex1_7d$CESD20[fhs2_ex1_7d$CESD20_missing >= 4] <- NA


####### FRIED frailty score

## Walking speed

# Uses greater of 2 speeds (PMID 26695510)

### Men:
# Height ≤ 173 cm (68.1 in):  ≤0.96 m/s
# Height >173 cm (68.1 in):   ≤1.04  m/s


fhs2_ex1_7d$gaitspeed_1 <- 4/(fhs2_ex1_7d$mri339)
fhs2_ex1_7d$gaitspeed_2 <- 4/(fhs2_ex1_7d$mri341)

fhs2_ex1_7d$gaitspeed_avg <- round(apply(fhs2_ex1_7d[,c("gaitspeed_1","gaitspeed_2")],
                                         MARGIN=1,
                                         mean,
                                         na.rm=T),3)

fhs2_ex1_7d$gaitspeed_avg[fhs2_ex1_7d$gaitspeed_avg=="NaN"] <- NA

fhs2_ex1_7d$gaitspeed_max <- round(apply(fhs2_ex1_7d[,c("gaitspeed_1","gaitspeed_2")],
                                         MARGIN=1,max),3)
fhs2_ex1_7d$gaitspeed_max[fhs2_ex1_7d$gaitspeed_max=="-Inf"] <- NA

fhs2_ex1_7d$gaitspeed_fried[!is.na(fhs2_ex1_7d$gaitspeed_1)|
                              !is.na(fhs2_ex1_7d$gaitspeed_2)] <- 0

fhs2_ex1_7d$gaitspeed_fried[fhs2_ex1_7d$SEX==1&
                              fhs2_ex1_7d$HGT1 >68.1&
                              fhs2_ex1_7d$gaitspeed_max <=1.04] <- 1

fhs2_ex1_7d$gaitspeed_fried[fhs2_ex1_7d$SEX==1&
                              fhs2_ex1_7d$HGT1 <= 68.1&
                              fhs2_ex1_7d$gaitspeed_max <= 0.96] <- 1


### Women
# Height ≤ 159 cm (62.6 in):  ≤0.90m/s
# Height >159 cm (62.6 in):   ≤1.02 m/s


fhs2_ex1_7d$gaitspeed_fried[fhs2_ex1_7d$SEX==2&
                              fhs2_ex1_7d$HGT1 >62.6&
                              fhs2_ex1_7d$gaitspeed_max <= 1.02] <- 1

fhs2_ex1_7d$gaitspeed_fried[fhs2_ex1_7d$SEX==2&
                              fhs2_ex1_7d$HGT1 <= 62.6&
                              fhs2_ex1_7d$gaitspeed_max <= 0.90] <- 1


########### Grip (kg)

# Used max of all trials per visit  (PMID 26695510)


## Uses BMI for normalization

##### Men
# BMI ≤ 24.0:     ≤ 29 kg
# BMI 24.1-26.0:  ≤ 30 kg
# BMI 26.1-28.0:  ≤ 30 kg
# BMI > 28.0:     ≤ 32 kg

##### Women
# BMI ≤ 23.0:     ≤ 17
# BMI 23.1-26.0:  ≤ 17.3 kg
# BMI 26.1-29.0:  ≤ 18 kg
# BMI > 29.0:     ≤ 21 kg

fhs2_ex1_7d$grip_avg <- round(apply(fhs2_ex1_7d[,c("mri356","mri357","mri358","mri360","mri361","mri362")],MARGIN=1,mean),1)
fhs2_ex1_7d$grip_max <- round(apply(fhs2_ex1_7d[,c("mri359","mri363")],MARGIN=1,max),1)

fhs2_ex1_7d$grip_fried[!is.na(fhs2_ex1_7d$grip_max)] <- 0


##### Men

fhs2_ex1_7d$grip_fried[fhs2_ex1_7d$BMI7<=24&
                         fhs2_ex1_7d$grip_max <= 29&
                         fhs2_ex1_7d$SEX==1] <- 1

fhs2_ex1_7d$grip_fried[fhs2_ex1_7d$BMI7>24&
                         fhs2_ex1_7d$BMI7<=26&
                         fhs2_ex1_7d$grip_max <= 30&
                         fhs2_ex1_7d$SEX==1] <- 1

fhs2_ex1_7d$grip_fried[fhs2_ex1_7d$BMI7>26&
                         fhs2_ex1_7d$BMI7<=28&
                         fhs2_ex1_7d$grip_max <= 30&
                         fhs2_ex1_7d$SEX==1] <- 1

fhs2_ex1_7d$grip_fried[fhs2_ex1_7d$BMI7>28&
                         fhs2_ex1_7d$grip_max <= 32&
                         fhs2_ex1_7d$SEX==1] <- 1

##### Women

fhs2_ex1_7d$grip_fried[fhs2_ex1_7d$BMI7<=23&
                         fhs2_ex1_7d$grip_max <= 17&
                         fhs2_ex1_7d$SEX==2] <- 1

fhs2_ex1_7d$grip_fried[fhs2_ex1_7d$BMI7>23&
                         fhs2_ex1_7d$BMI7<=26&
                         fhs2_ex1_7d$grip_max <= 17.3&
                         fhs2_ex1_7d$SEX==2] <- 1

fhs2_ex1_7d$grip_fried[fhs2_ex1_7d$BMI7>26&
                         fhs2_ex1_7d$BMI7<=29&
                         fhs2_ex1_7d$grip_max <= 18&
                         fhs2_ex1_7d$SEX==2] <- 1

fhs2_ex1_7d$grip_fried[fhs2_ex1_7d$BMI7>29&
                         fhs2_ex1_7d$grip_max <= 21&
                         fhs2_ex1_7d$SEX==2] <- 1

fhs2_ex1_7d$exhaustion_fried[fhs2_ex1_7d$G606 %in% c(0,1)|fhs2_ex1_7d$G593 %in% c(0,1)] <- 0
fhs2_ex1_7d$exhaustion_fried[fhs2_ex1_7d$G606 %in% c(2,3)|fhs2_ex1_7d$G593 %in% c(2,3)] <- 1


#### Weight loss - Fried

fhs2_ex1_7d$wt_loss_pct <- round((fhs2_ex1_7d$WGT6 - fhs2_ex1_7d$WGT7)/fhs2_ex1_7d$WGT6*100,1)
fhs2_ex1_7d$wtloss_fried[fhs2_ex1_7d$wt_loss_pct >= 5] <- 1
fhs2_ex1_7d$wtloss_fried[fhs2_ex1_7d$wt_loss_pct < 5] <- 0


#### Physical activity - Fried

fhs2_ex1_7d$fhs_pai <- 
  calc_framingham_pai(as.data.frame(fhs2_ex1_7d),
                      slp_hrs = "G689",
                      sed_hrs = "G690",
                      slgt_hrs = "G691",
                      mod_hrs = "G692",
                      hvy_hrs = "G693")

fhs2_ex1_7d$activity_fried[fhs2_ex1_7d$SEX==1&
                             fhs2_ex1_7d$fhs_pai < 30.3] <- 1

fhs2_ex1_7d$activity_fried[fhs2_ex1_7d$SEX==1&
                             fhs2_ex1_7d$fhs_pai >= 30.3] <- 0


fhs2_ex1_7d$activity_fried[fhs2_ex1_7d$SEX==2&
                             fhs2_ex1_7d$fhs_pai < 30.1] <- 1

fhs2_ex1_7d$activity_fried[fhs2_ex1_7d$SEX==2&
                             fhs2_ex1_7d$fhs_pai >= 30.1] <- 0


#### Calculate Fried score

fhs2_ex1_7d$total_fried <-
  fhs2_ex1_7d$gaitspeed_fried+
  fhs2_ex1_7d$grip_fried+
  fhs2_ex1_7d$exhaustion_fried+
  fhs2_ex1_7d$wtloss_fried+
  fhs2_ex1_7d$activity_fried


######### FRAIL Score

#### Fatigue

fhs2_ex1_7d$fatigue_frail[fhs2_ex1_7d$G606 %in% c(0,1)|fhs2_ex1_7d$G593 %in% c(0,1)] <- 0
fhs2_ex1_7d$fatigue_frail[fhs2_ex1_7d$G606 %in% c(2,3)|fhs2_ex1_7d$G593 %in% c(2,3)] <- 1


#### Resistance

fhs2_ex1_7d$resistance_frail[fhs2_ex1_7d$G525==0] <- 0
fhs2_ex1_7d$resistance_frail[fhs2_ex1_7d$G525 %in% c(1,2,3,4)] <- 1


#### Ambulation

fhs2_ex1_7d$ambulate_frail[fhs2_ex1_7d$G548==1] <- 0
fhs2_ex1_7d$ambulate_frail[fhs2_ex1_7d$G548==0] <- 1


# FRAIL Illness score counts presence of HTN, DM, cancer, chronic lung disease, MI, CHD, angina, asthma, arthritis, stroke, and ckd
# Scores for 0-4 conditions, score = 0, for ≥ 5, score=1

# HTN

fhs2_ex1_7d$htn_frail[fhs2_ex1_7d$HRX7==0] <- 0
fhs2_ex1_7d$htn_frail[fhs2_ex1_7d$HRX7==1] <- 1


# COPD (chronic lung disease)

fhs2_ex1_7d$copd_frail[fhs2_ex1_7d$G415 %in% c(0,2)] <- 0
fhs2_ex1_7d$copd_frail[fhs2_ex1_7d$G415==1] <- 1

# Asthma

fhs2_ex1_7d$asthma_frail[fhs2_ex1_7d$G418 %in% c(0,2)] <- 0
fhs2_ex1_7d$asthma_frail[fhs2_ex1_7d$G418==1] <- 1


# DJD (arthritis)

fhs2_ex1_7d$djd_frail[fhs2_ex1_7d$G421 %in% c(0,2)] <- 0
fhs2_ex1_7d$djd_frail[fhs2_ex1_7d$G421==1] <- 1


# CKD (renal disease)

fhs2_ex1_7d <-
  merge(fhs2_ex1_7d,
        fhs2_race_1,
        by=c("PID","IDTYPE"),
        all.x=T)

fhs2_ex1_7d$crcl <- 
  calc_MDRD4(dat=as.data.frame(fhs2_ex1_7d),
             age="AGE7",
             cr="CREAT7",
             black="B")

fhs2_ex1_7d$ckd_frail[fhs2_ex1_7d$crcl >= 60] <- 0
fhs2_ex1_7d$ckd_frail[fhs2_ex1_7d$crcl < 60] <- 1


# CHF/MI

fhs2_ex1_7d <- merge(fhs2_ex1_7d,
                     fhs2_survcvd,
                     by=c("PID",
                          "IDTYPE"))

fhs2_ex1_7d$chf_frail <- 0
fhs2_ex1_7d$chf_frail[fhs2_ex1_7d$hfhosp_status==1&fhs2_ex1_7d$hfhosp_dt <= fhs2_ex1_7d$DATE7] <- 1

fhs2_ex1_7d$chd_frail <- 0
fhs2_ex1_7d$chd_frail[fhs2_ex1_7d$cadhosp_status==1&fhs2_ex1_7d$cadhosp_dt <= fhs2_ex1_7d$DATE7] <- 1


# Stroke

fhs2_ex1_7d$stroke_frail <- 0
fhs2_ex1_7d$stroke_frail[fhs2_ex1_7d$STROKE==1&fhs2_ex1_7d$STROKEDATE <= fhs2_ex1_7d$DATE7] <- 1


# Cancer

fhs2_ex1_7d$cancer_frail <- 0 
fhs2_ex1_7d$cancer_frail[fhs2_ex1_7d$D_DATE <= fhs2_ex1_7d$DATE7] <- 1


# Sum conditions

fhs2_ex1_7d$conditions_frail <-
  fhs2_ex1_7d$htn_frail+
  fhs2_ex1_7d$DMRX7+
  fhs2_ex1_7d$copd_frail+
  fhs2_ex1_7d$asthma_frail+
  fhs2_ex1_7d$djd_frail+
  fhs2_ex1_7d$ckd_frail+
  fhs2_ex1_7d$chf_frail+
  fhs2_ex1_7d$chd_frail+
  fhs2_ex1_7d$stroke_frail+
  fhs2_ex1_7d$cancer_frail

fhs2_ex1_7d$illness_frail[fhs2_ex1_7d$conditions_frail <= 4] <- 0
fhs2_ex1_7d$illness_frail[fhs2_ex1_7d$conditions_frail > 4] <- 1

#### (Weight) Loss

fhs2_ex1_7d$wtloss_frail[fhs2_ex1_7d$wt_loss_pct >= 5] <- 1
fhs2_ex1_7d$wtloss_frail[fhs2_ex1_7d$wt_loss_pct < 5] <- 0


#### Calculate FRAIL score

fhs2_ex1_7d$total_frail <- 
  fhs2_ex1_7d$fatigue_frail+
  fhs2_ex1_7d$resistance_frail+
  fhs2_ex1_7d$ambulate_frail+
  fhs2_ex1_7d$illness_frail+
  fhs2_ex1_7d$wtloss_frail


#### 

fhs2_melt_ex1_7d <- melt(fhs2_ex1_7d,id.vars=c("PID","IDTYPE"), na.rm=T, factorsAsStrings=T)
fhs2_melt_ex1_7d$form <- "fhs2_offspring_ex07"

fhs2_melt_ex1_7d$visit <- 7

rm(fhs2_ex1_7d)


#=================================#
####     Cardiac MRI exam 7    ---#
#=================================#

fhs2_cmrlvh1 <- fread('~/Dropbox/BioLINCC files/Framingham offspring/datasets/CSV/t_mrcdlvh_2006_1b_0907d_v1.csv',na.strings=c("NA","","NULL")) # Offspring cardiac MRI LVH 
fhs2_cmrlvh1$visit <- 7

fhs2_cmrlvh1$CMR03_cm <- fhs2_cmrlvh1$CMR03/10
fhs2_cmrlvh1$CMR04_cm <- fhs2_cmrlvh1$CMR04/10
fhs2_cmrlvh1$CMR05_cm <- fhs2_cmrlvh1$CMR05/10

fhs2_cmrlvh1$rwt <- 2*fhs2_cmrlvh1$CMR04/fhs2_cmrlvh1$CMR05


fhs2_meas7 <- fhs2_offspring_wkthru[,c("PID","IDTYPE","SEX","HGT1","WGT7")]

fhs2_meas7$weight_kg <- fhs2_meas7$WGT7/2.2
fhs2_meas7$height_cm <- fhs2_meas7$HGT1*2.54


fhs2_meas7$BSA <- calc_bsa(dat=fhs2_meas7)

fhs2_cmrlvh1 <- merge(fhs2_cmrlvh1,
                      fhs2_meas7,
                      by=c("PID","IDTYPE"),
                      all.x=T)

fhs2_cmrlvh1$lvmass_ix <- fhs2_cmrlvh1$CMR22/fhs2_cmrlvh1$BSA

fhs2_cmrlvh1$lvh_type[fhs2_cmrlvh1$SEX==1&fhs2_cmrlvh1$lvmass_ix>115&fhs2_cmrlvh1$rwt<=0.42&!is.na(fhs2_cmrlvh1$lvmass_ix)&!is.na(fhs2_cmrlvh1$rwt)] <- "Eccentric hypertropy"
fhs2_cmrlvh1$lvh_type[fhs2_cmrlvh1$SEX==2&fhs2_cmrlvh1$lvmass_ix>95&fhs2_cmrlvh1$rwt<=0.42&!is.na(fhs2_cmrlvh1$lvmass_ix)&!is.na(fhs2_cmrlvh1$rwt)] <- "Eccentric hypertropy"

fhs2_cmrlvh1$lvh_type[fhs2_cmrlvh1$SEX==1&fhs2_cmrlvh1$lvmass_ix>115&fhs2_cmrlvh1$rwt>0.42&!is.na(fhs2_cmrlvh1$lvmass_ix)&!is.na(fhs2_cmrlvh1$rwt)] <- "Concentric hypertropy"
fhs2_cmrlvh1$lvh_type[fhs2_cmrlvh1$SEX==2&fhs2_cmrlvh1$lvmass_ix>95&fhs2_cmrlvh1$rwt>0.42&!is.na(fhs2_cmrlvh1$lvmass_ix)&!is.na(fhs2_cmrlvh1$rwt)] <- "Concentric hypertropy"

fhs2_cmrlvh1$lvh_type[fhs2_cmrlvh1$SEX==1&fhs2_cmrlvh1$lvmass_ix<=115&fhs2_cmrlvh1$rwt<=0.42&!is.na(fhs2_cmrlvh1$lvmass_ix)&!is.na(fhs2_cmrlvh1$rwt)] <- "Normal geometry"
fhs2_cmrlvh1$lvh_type[fhs2_cmrlvh1$SEX==2&fhs2_cmrlvh1$lvmass_ix<=95&fhs2_cmrlvh1$rwt<=0.42&!is.na(fhs2_cmrlvh1$lvmass_ix)&!is.na(fhs2_cmrlvh1$rwt)] <- "Normal geometry"

fhs2_cmrlvh1$lvh_type[fhs2_cmrlvh1$SEX==1&fhs2_cmrlvh1$lvmass_ix<=115&fhs2_cmrlvh1$rwt>0.42&!is.na(fhs2_cmrlvh1$lvmass_ix)&!is.na(fhs2_cmrlvh1$rwt)] <- "Concentric remodeling"
fhs2_cmrlvh1$lvh_type[fhs2_cmrlvh1$SEX==2&fhs2_cmrlvh1$lvmass_ix<=95&fhs2_cmrlvh1$rwt>0.42&!is.na(fhs2_cmrlvh1$lvmass_ix)&!is.na(fhs2_cmrlvh1$rwt)] <- "Concentric remodeling"

fhs2_melt_cmrlvh1 <- 
  melt(fhs2_cmrlvh1,id.vars=c("PID",
                              "IDTYPE",
                              "visit"),
       na.rm=T,
       factorsAsStrings=T)

fhs2_melt_cmrlvh1$form <- "fhs2_cmrlvh1"


rm(fhs2_cmrlvh1)


### MRI wall motion abnormalities

fhs2_cmrwma1 <- fread('~/Dropbox/BioLINCC files/Framingham offspring/datasets/CSV/t_mrcdwma_2006_1b_0909d_v1.csv',na.strings=c("NA","","NULL"))  # Offspring cardiac MRI wall motion abnormalities
names(fhs2_cmrwma1)[names(fhs2_cmrwma1)=="idtype"] <- "IDTYPE"
fhs2_cmrwma1$visit <- 7
fhs2_melt_cmrwma1 <- melt(fhs2_cmrwma1,id.vars=c("PID","IDTYPE","visit"),na.rm=T,factorsAsStrings=T)
fhs2_melt_cmrwma1$form <- "fhs2_cmrwma1"

rm(fhs2_cmrwma1)


#=========================#
####  DM labs, Exam 7  ---#
#=========================#

fhs2_l_dbtlab_ex07 <- fread('~/Dropbox/BioLINCC files/Framingham offspring/datasets/CSV/l_dbtlab_ex07_1b_1237d_v1.csv',na.strings=c("NA","","NULL"))
names(fhs2_l_dbtlab_ex07)[names(fhs2_l_dbtlab_ex07)=="idtype"] <- "IDTYPE"
fhs2_l_dbtlab_ex07$visit <- fhs2_l_dbtlab_ex07$EXAM
fhs2_melt_l_dbtlab_ex07 <- melt(fhs2_l_dbtlab_ex07,id.vars=c("PID","IDTYPE","visit"), na.rm=T, factorsAsStrings=T)
fhs2_melt_l_dbtlab_ex07$form <- "fhs2_dbtlab_ex07"

rm(fhs2_l_dbtlab_ex07)


#======================================#
####      Lab panel, Exam 7         ---#
#======================================#

fhs2_l_fhslab_ex07 <- fread('~/Dropbox/BioLINCC files/Framingham offspring/datasets/CSV/l_fhslaba_ex07_1_0284d.csv',na.strings=c("NA","","NULL"))
names(fhs2_l_fhslab_ex07)[names(fhs2_l_fhslab_ex07)=="idtype"] <- "IDTYPE"
fhs2_l_fhslab_ex07$visit <- 6
fhs2_melt_l_fhslab_ex07 <- melt(fhs2_l_fhslab_ex07,id.vars=c("PID","IDTYPE","visit"), na.rm=T, factorsAsStrings=T)
fhs2_melt_l_fhslab_ex07$form <- "fhs2_fhslab_ex07"

rm(fhs2_l_fhslab_ex07)

#=======================================#
####  Creatinine/cysteine, Exam 7    ---#
#=======================================#

fhs2_creacys_ex07_1 <- fread('~/Dropbox/BioLINCC files/Framingham offspring/datasets/CSV/l_creacys_ex07_1_0092d.csv',na.strings=c("NA","","NULL"))
names(fhs2_creacys_ex07_1)[names(fhs2_creacys_ex07_1)=="idtype"] <- "IDTYPE"
fhs2_creacys_ex07_1$visit <- 7
fhs2_melt_l_creacys_ex07 <- melt(fhs2_creacys_ex07_1,id.vars=c("PID","IDTYPE","visit"), na.rm=T, factorsAsStrings=T)
fhs2_melt_l_creacys_ex07$form <- "fhs2_creacys_ex07"

rm(fhs2_creacys_ex07_1)


#====================================#
####      Walk test, Exam 7       ---#
#====================================#

fhs2_t_wktest_07 <- fread('~/Dropbox/BioLINCC files/Framingham offspring/datasets/CSV/t_wktest_ex07_1_0016d.csv',na.strings=c("NA","","NULL"))
names(fhs2_t_wktest_07)[names(fhs2_t_wktest_07)=="idtype"] <- "IDTYPE"
fhs2_t_wktest_07$visit <- 7
fhs2_melt_t_wktest_07 <- melt(fhs2_t_wktest_07,id.vars=c("PID","IDTYPE","visit"), na.rm=T, factorsAsStrings=T)
fhs2_melt_t_wktest_07$form <- "fhs2_t_wktest"

rm(fhs2_t_wktest_07)


#================================================================#
####  Estrogen levels, Framingham 3, Exam 1/Omni 2, Exam 1    ---#
#================================================================#

fhs2_estr_ex7 <- fread('~/Dropbox/BioLINCC files/Framingham offspring/datasets/CSV/l_estr_2005_m_0604d.csv')
fhs2_melt_estr_ex7 <-  melt(fhs2_estr_ex7,id.vars=c("PID","IDTYPE"),na.rm=T,factorsAsStrings = T)
fhs2_melt_estr_ex7$visit <- 7
fhs2_melt_estr_ex7$form <- "fhs2_estr_07"

rm(fhs2_estr_ex7)


#==============================#
####       CRP, Exam 7      ---#
#==============================#


fhs2_l_inflamm_ex07 <- 
  fread('~/Dropbox/BioLINCC files/Framingham offspring/datasets/CSV/l_inflamm_ex07_1b_1105d.csv',
        na.strings=c("NA","","NULL"))

names(fhs2_l_inflamm_ex07) <- toupper(names(fhs2_l_inflamm_ex07))

fhs2_l_inflamm_ex07$VISIT[fhs2_l_inflamm_ex07$IDTYPE==1] <- 7
fhs2_l_inflamm_ex07$VISIT[fhs2_l_inflamm_ex07$IDTYPE==7] <- 2

fhs2_melt_inflamm_ex07 <- 
  melt(fhs2_l_inflamm_ex07,
       id.vars=c("PID","IDTYPE","VISIT"), 
       na.rm=T, 
       factorsAsStrings=T)

names(fhs2_melt_inflamm_ex07)[3] <- "visit"

fhs2_melt_inflamm_ex07$form <- "fhs2_inflamm_ex07"

rm(fhs2_l_inflamm_ex07)


#====================================================#
####  Validated food frequency data, exam 7       ---#
#====================================================#

fhs2_vr_ffreq_ex07 <- fread('~/Dropbox/BioLINCC files/Framingham offspring/datasets/CSV/vr_ffreq_ex07_1_0460d.csv',na.strings=c("NA","","NULL"))
names(fhs2_vr_ffreq_ex07)[names(fhs2_vr_ffreq_ex07)=="idtype"] <- "IDTYPE"
fhs2_vr_ffreq_ex07$visit <- 7
fhs2_melt_vr_ffreq_ex07 <- melt(fhs2_vr_ffreq_ex07,id.vars=c("PID","IDTYPE","visit"), na.rm=T, factorsAsStrings=T)
fhs2_melt_vr_ffreq_ex07$form <- "fhs2_vr_ffreq"

rm(fhs2_vr_ffreq_ex07)


#================================================================#
####   Male hormones in females, Offspring, Exam 7            ---#
#================================================================#

fhs2_fhorm_ex07 <- fread('~/Dropbox/BioLINCC files/Framingham offspring/datasets/CSV/l_fhorm_2005_m_0493d.csv')
fhs2_melt_fhorm_ex07 <-  melt(fhs2_fhorm_ex07,id.vars=c("PID","IDTYPE"),na.rm=T,factorsAsStrings = T)
fhs2_melt_fhorm_ex07$visit <- 1
fhs2_melt_fhorm_ex07$form <- "fhs2_fhorm_ex07"

rm(fhs2_fhorm_ex07)

#================================================================#
####  Male hormones, Offspring, Exam 7      ---#
#================================================================#

fhs2_mhorm_ex07 <- fread('~/Dropbox/BioLINCC files/Framingham offspring/datasets/CSV/l_mhorm_2005_m_0490d_v1.csv')
fhs2_melt_mhorm_ex07 <-  melt(fhs2_mhorm_ex07,id.vars=c("PID","IDTYPE"),na.rm=T,factorsAsStrings = T)
fhs2_melt_mhorm_ex07$visit <- 1
fhs2_melt_mhorm_ex07$form <- "fhs2_mhorm_ex07"

rm(fhs2_mhorm_ex07)


#==================================#
####  *** Offspring exam 8 ***  ####
#==================================#

fhs2_ex1_8d <- fread('~/Dropbox/BioLINCC files/Framingham offspring/datasets/CSV/e_exam_ex08_1_0005d.csv',na.strings=c("NA","","NULL")) # Omni 1, Exam 8
names(fhs2_ex1_8d)[names(fhs2_ex1_8d)=="idtype"] <- "IDTYPE"
fhs2_ex1_8d$visit <- 8
fhs2_ex1_8d <- merge(fhs2_ex1_8d,
                     fhs2_offspring_wkthru[,c("PID",
                                              "AGE8",
                                              "SEX",
                                              "DMRX8",
                                              "HRX8",
                                              "DATE8",
                                              "HGT1",
                                              "WGT8",
                                              "CREAT8",
                                              "WGT7")],
                     by=c("PID") )

fhs2_ex1_8d$H033[!between(fhs2_ex1_8d$H033,22,83)] <- NA
fhs2_ex1_8d$H046[!between(fhs2_ex1_8d$H046,21,77)] <- NA


fhs2_ex1_8d$hrt_years_total[between(fhs2_ex1_8d$H047,1,42)&!is.na(fhs2_ex1_8d$H047)] <- 
  fhs2_ex1_8d$H047[between(fhs2_ex1_8d$H047,1,42)&!is.na(fhs2_ex1_8d$H047)] 

fhs2_ex1_8d$hrt_years_total[between(fhs2_ex1_8d$H048,1,10)&!is.na(fhs2_ex1_8d$H048)] <- 
  fhs2_ex1_8d$hrt_years_total[between(fhs2_ex1_8d$H048,1,10)&!is.na(fhs2_ex1_8d$H048)] + 
  fhs2_ex1_8d$H048[between(fhs2_ex1_8d$H048,1,10)&!is.na(fhs2_ex1_8d$H048)]/12

fhs2_ex1_8d$H037[fhs2_ex1_8d$H037==888]

####### FRIED frailty score

## Walking speed

# Uses greater of 2 speeds (PMID 26695510)

fhs2_ex1_8d$gaitspeed_1 <- 4/(fhs2_ex1_8d$H617)
fhs2_ex1_8d$gaitspeed_2 <- 4/(fhs2_ex1_8d$H620)

fhs2_ex1_8d$gaitspeed_avg <- round(apply(fhs2_ex1_8d[,c("gaitspeed_1","gaitspeed_2")],
                                         MARGIN=1,
                                         mean,
                                         na.rm=T),3)

fhs2_ex1_8d$gaitspeed_avg[fhs2_ex1_8d$gaitspeed_avg=="NaN"] <- NA

fhs2_ex1_8d$gaitspeed_max <- round(apply(fhs2_ex1_8d[,c("gaitspeed_1","gaitspeed_2")],
                                         MARGIN=1,max),3)
fhs2_ex1_8d$gaitspeed_max[fhs2_ex1_8d$gaitspeed_max=="-Inf"] <- NA

fhs2_ex1_8d$gaitspeed_fried[!is.na(fhs2_ex1_8d$gaitspeed_1)|
                              !is.na(fhs2_ex1_8d$gaitspeed_2)] <- 0

### Men:

# Height ≤ 173 cm (68.1 in):  ≤0.96 m/s
# Height >173 cm (68.1 in):   ≤1.04  m/s

fhs2_ex1_8d$gaitspeed_fried[fhs2_ex1_8d$SEX==1&
                              fhs2_ex1_8d$HGT1 >68.1&
                              fhs2_ex1_8d$gaitspeed_max <=1.04] <- 1

fhs2_ex1_8d$gaitspeed_fried[fhs2_ex1_8d$SEX==1&
                              fhs2_ex1_8d$HGT1 <= 68.1&
                              fhs2_ex1_8d$gaitspeed_max <= 0.96] <- 1

### Women
# Height ≤ 159 cm (62.6 in):  ≤0.90m/s
# Height >159 cm (62.6 in):   ≤1.02 m/s


fhs2_ex1_8d$gaitspeed_fried[fhs2_ex1_8d$SEX==2&
                              fhs2_ex1_8d$HGT1 >62.6&
                              fhs2_ex1_8d$gaitspeed_max <= 1.02] <- 1

fhs2_ex1_8d$gaitspeed_fried[fhs2_ex1_8d$SEX==2&
                              fhs2_ex1_8d$HGT1 <= 62.6&
                              fhs2_ex1_8d$gaitspeed_max <= 0.90] <- 1


########### Grip (kg)

# Used max of all trials per visit  (PMID 26695510)


## Uses BMI for normalization

##### Men
# BMI ≤ 24.0:     ≤ 29 kg
# BMI 24.1-26.0:  ≤ 30 kg
# BMI 26.1-28.0:  ≤ 30 kg
# BMI > 28.0:     ≤ 32 kg

##### Women
# BMI ≤ 23.0:     ≤ 17
# BMI 23.1-26.0:  ≤ 17.3 kg
# BMI 26.1-29.0:  ≤ 18 kg
# BMI > 29.0:     ≤ 21 kg

fhs2_ex1_8d$grip_avg <- round(apply(fhs2_ex1_8d[,c("H605","H606","H607","H608","H609","H610")],MARGIN=1,mean),1)
fhs2_ex1_8d$grip_max <- round(apply(fhs2_ex1_8d[,c("H605","H606","H607","H608","H609","H610")],MARGIN=1,max),1)

fhs2_ex1_8d$grip_fried[!is.na(fhs2_ex1_8d$grip_max)] <- 0


##### Men

fhs2_ex1_8d$grip_fried[fhs2_ex1_8d$BMI7<=24&
                         fhs2_ex1_8d$grip_max <= 29&
                         fhs2_ex1_8d$SEX==1] <- 1

fhs2_ex1_8d$grip_fried[fhs2_ex1_8d$BMI7>24&
                         fhs2_ex1_8d$BMI7<=26&
                         fhs2_ex1_8d$grip_max <= 30&
                         fhs2_ex1_8d$SEX==1] <- 1

fhs2_ex1_8d$grip_fried[fhs2_ex1_8d$BMI7>26&
                         fhs2_ex1_8d$BMI7<=28&
                         fhs2_ex1_8d$grip_max <= 30&
                         fhs2_ex1_8d$SEX==1] <- 1

fhs2_ex1_8d$grip_fried[fhs2_ex1_8d$BMI7>28&
                         fhs2_ex1_8d$grip_max <= 32&
                         fhs2_ex1_8d$SEX==1] <- 1

##### Women

fhs2_ex1_8d$grip_fried[fhs2_ex1_8d$BMI7<=23&
                         fhs2_ex1_8d$grip_max <= 17&
                         fhs2_ex1_8d$SEX==2] <- 1

fhs2_ex1_8d$grip_fried[fhs2_ex1_8d$BMI7>23&
                         fhs2_ex1_8d$BMI7<=26&
                         fhs2_ex1_8d$grip_max <= 17.3&
                         fhs2_ex1_8d$SEX==2] <- 1

fhs2_ex1_8d$grip_fried[fhs2_ex1_8d$BMI7>26&
                         fhs2_ex1_8d$BMI7<=29&
                         fhs2_ex1_8d$grip_max <= 18&
                         fhs2_ex1_8d$SEX==2] <- 1

fhs2_ex1_8d$grip_fried[fhs2_ex1_8d$BMI7>29&
                         fhs2_ex1_8d$grip_max <= 21&
                         fhs2_ex1_8d$SEX==2] <- 1

### Fried exhaustion

fhs2_ex1_8d$exhaustion_fried[fhs2_ex1_8d$H471 %in% c(0,1)&fhs2_ex1_8d$H472 %in% c(0,1)] <- 0
fhs2_ex1_8d$exhaustion_fried[fhs2_ex1_8d$H471 %in% c(2,3)|fhs2_ex1_8d$H472 %in% c(2,3)] <- 1


#### Weight loss - Fried

fhs2_ex1_8d$wt_loss_pct <- round((fhs2_ex1_8d$WGT7 - fhs2_ex1_8d$WGT8)/fhs2_ex1_8d$WGT7*100,1)
fhs2_ex1_8d$wtloss_fried[fhs2_ex1_8d$wt_loss_pct >= 5] <- 1
fhs2_ex1_8d$wtloss_fried[fhs2_ex1_8d$wt_loss_pct < 5] <- 0


#### Physical activity - Fried

fhs2_ex1_8d$fhs_pai <- 
  calc_framingham_pai(
    as.data.frame(fhs2_ex1_8d),
    slp_hrs = "H480",
    sed_hrs = "H481",
    slgt_hrs = "H482",
    mod_hrs = "H483",
    hvy_hrs = "H484")

fhs2_ex1_8d$activity_fried[fhs2_ex1_8d$SEX==1&
                             fhs2_ex1_8d$fhs_pai < 30.3] <- 1

fhs2_ex1_8d$activity_fried[fhs2_ex1_8d$SEX==1&
                             fhs2_ex1_8d$fhs_pai >= 30.3] <- 0


fhs2_ex1_8d$activity_fried[fhs2_ex1_8d$SEX==2&
                             fhs2_ex1_8d$fhs_pai < 30.1] <- 1

fhs2_ex1_8d$activity_fried[fhs2_ex1_8d$SEX==2&
                             fhs2_ex1_8d$fhs_pai >= 30.1] <- 0


#### Calculate Fried score

fhs2_ex1_8d$total_fried <-
  fhs2_ex1_8d$gaitspeed_fried+
  fhs2_ex1_8d$grip_fried+
  fhs2_ex1_8d$exhaustion_fried+
  fhs2_ex1_8d$wtloss_fried+
  fhs2_ex1_8d$activity_fried


fhs2_ex1_8d$BMIC8 <- 
  calc_bmi(dat=as.data.frame(fhs2_ex1_8d), 
           weight="WGT8",
           height="HGT1",
           metric=F)

fhs2_ex1_8d <- merge(fhs2_ex1_8d, 
                     fhs_survcvd,
                     by=c("PID","IDTYPE"),
                     all.x=T)

fhs2_ex1_8d <- merge(fhs2_ex1_8d, 
                     fhs_survstk,
                     by=c("PID","IDTYPE"),
                     all.x=T)


fhs2_ex1_8d <- merge(fhs2_ex1_8d,
                     fhs_survca_first_any[,c("PID","IDTYPE","D_DATE")],
                     by=c("PID","IDTYPE"),
                     all.x=T)

fhs2_ex1_8d$drinks_wk <- fhs2_ex1_8d$H072+
  fhs2_ex1_8d$H075+
  fhs2_ex1_8d$H078

fhs2_ex1_8d$epworth_ss_nas <- 
  is.na(fhs2_ex1_8d$H734)+
  is.na(fhs2_ex1_8d$H735)+
  is.na(fhs2_ex1_8d$H736)+
  is.na(fhs2_ex1_8d$H737)+
  is.na(fhs2_ex1_8d$H738)+
  is.na(fhs2_ex1_8d$H739)+
  is.na(fhs2_ex1_8d$H740)+
  is.na(fhs2_ex1_8d$H741)

fhs2_ex1_8d$epworth_ss <- 
  fhs2_ex1_8d$H734+
  fhs2_ex1_8d$H735+
  fhs2_ex1_8d$H736+
  fhs2_ex1_8d$H737+
  fhs2_ex1_8d$H738+
  fhs2_ex1_8d$H739+
  fhs2_ex1_8d$H740+
  fhs2_ex1_8d$H741


## Denormalize

fhs2_melt_ex1_8d <- melt(fhs2_ex1_8d,
                         id.vars=c("PID","IDTYPE","visit"), 
                         na.rm=T, 
                         factorsAsStrings=T)

fhs2_melt_ex1_8d$form <- "fhs2_offspring_ex08"

rm(fhs2_ex1_8d)


#================================#
####     Lab panel, Exam 8    ---#
#================================#

fhs2_l_fhslab_ex08 <- fread('~/Dropbox/BioLINCC files/Framingham offspring/datasets/CSV/l_fhslab_ex08_1_0257d.csv',na.strings=c("NA","","NULL"))
names(fhs2_l_fhslab_ex08)[names(fhs2_l_fhslab_ex08)=="idtype"] <- "IDTYPE"
fhs2_l_fhslab_ex08$visit <- 8
fhs2_melt_l_fhslab_ex08 <- melt(fhs2_l_fhslab_ex08,id.vars=c("PID","IDTYPE","visit"), na.rm=T, factorsAsStrings=T)
fhs2_melt_l_fhslab_ex08$form <- "fhs2_fhslab_ex08"

rm(fhs2_l_fhslab_ex08)


#==========================================#
####     Echo cardiac strain, Exam 8    ---#
#==========================================#

fhs2_t_echocs_ex08 <- fread('~/Dropbox/BioLINCC files/Framingham offspring/datasets/CSV/t_echocs_ex08_1_0705d.csv',na.strings=c("NA","","NULL"))
names(fhs2_t_echocs_ex08)[names(fhs2_t_echocs_ex08)=="idtype"] <- "IDTYPE"

names(fhs2_t_echocs_ex08) <-
  sub("^_","",names(fhs2_t_echocs_ex08))

fhs2_t_echocs_ex08$visit <- 8
fhs2_melt_t_echocs_ex08 <- melt(fhs2_t_echocs_ex08,id.vars=c("PID","IDTYPE","visit"), na.rm=T, factorsAsStrings=T)
fhs2_melt_t_echocs_ex08$form <- "fhs2_echocs_ex08"

rm(fhs2_t_echocs_ex08)


#==========================================#
####     Echo RV, Exam 8                ---#
#==========================================#

fhs2_t_echorv_ex08 <- fread('~/Dropbox/BioLINCC files/Framingham offspring/datasets/CSV/t_echorv_ex08_1b_0830d_v1.csv',na.strings=c("NA","","NULL"))
names(fhs2_t_echorv_ex08)[names(fhs2_t_echorv_ex08)=="idtype"] <- "IDTYPE"


fhs2_t_echorv_ex08$visit[fhs2_t_echorv_ex08$IDTYPE==1] <- 8
fhs2_t_echorv_ex08$visit[fhs2_t_echorv_ex08$IDTYPE==7] <- 3

fhs2_melt_t_echorv_ex08 <- 
  melt(fhs2_t_echorv_ex08,id.vars=c("PID","IDTYPE","visit"), 
       na.rm=T, 
       factorsAsStrings=T)

fhs2_melt_t_echorv_ex08$form <- "fhs2_echorv_ex08"

rm(fhs2_t_echorv_ex08)


#=========================================================#
####  Validated food frequency data, Exam 8            ---#
#=========================================================#

fhs2_vr_ffreq_ex08 <- fread('~/Dropbox/BioLINCC files/Framingham offspring/datasets/CSV/vr_ffreq_ex08_1_0615d_v1.csv',na.strings=c("NA","","NULL"))
names(fhs2_vr_ffreq_ex08)[names(fhs2_vr_ffreq_ex08)=="idtype"] <- "IDTYPE"
fhs2_vr_ffreq_ex08$visit <- 8
fhs2_melt_vr_ffreq_ex08 <- melt(fhs2_vr_ffreq_ex08,id.vars=c("PID","IDTYPE","visit"), na.rm=T, factorsAsStrings=T)
fhs2_melt_vr_ffreq_ex08$form <- "fhs2_vr_ffreq"

rm(fhs2_vr_ffreq_ex08)


#================================#
####  *** OMNI 1, exam 1 ***  ####
#================================#

fhs2_omni1_ex01 <- fread('~/Dropbox/BioLINCC files/Framingham offspring/datasets/CSV/e_exam_ex01_7_0020d.csv',na.strings=c("NA","","NULL"))  # Omni 1, Exam 1
names(fhs2_omni1_ex01)[names(fhs2_omni1_ex01)=="idtype"] <- "IDTYPE"

names(fhs2_omni1_ex01) <- toupper(names(fhs2_omni1_ex01))

fhs2_omni1_ex01$drinks_wk <- fhs2_omni1_ex01$E310+
  fhs2_omni1_ex01$E313+
  fhs2_omni1_ex01$E316

fhs2_omni1_ex01$visit <- 1
fhs2_melt_omni1_ex01 <- melt(fhs2_omni1_ex01,id.vars=c("PID","IDTYPE","visit"), na.rm=T, factorsAsStrings=T)
fhs2_melt_omni1_ex01$form <- "fhs2_omni1_ex01"

rm(fhs2_omni1_ex01)


#===================================#
####   Lab panel, Exam 1         ---#
#===================================#

fhs2_l_fhslab_ex01 <- fread('~/Dropbox/BioLINCC files/Framingham offspring/datasets/CSV/l_fhslab_ex01_7_0270d_v1.csv',na.strings=c("NA","","NULL"))
names(fhs2_l_fhslab_ex01)[names(fhs2_l_fhslab_ex01)=="idtype"] <- "IDTYPE"
fhs2_l_fhslab_ex01$visit <- 1
fhs2_melt_l_fhslab_ex01 <- melt(fhs2_l_fhslab_ex01,id.vars=c("PID","IDTYPE","visit"), na.rm=T, factorsAsStrings=T)
fhs2_melt_l_fhslab_ex01$form <- "fhs2_fhslab_ex01_7"

rm(fhs2_l_fhslab_ex01)


#==============================#
#### *** OMNI 1, exam 2 *** ####
#==============================#

fhs2_omni1_ex02 <- fread('~/Dropbox/BioLINCC files/Framingham offspring/datasets/CSV/e_exam_ex02_7_0003d.csv',na.strings=c("NA","","NULL"))  # Omni 1, Exam 2
names(fhs2_omni1_ex02) <- toupper(names(fhs2_omni1_ex02))

fhs2_omni1_ex02 <- merge(fhs2_omni1_ex02,
                         fhs2_race_1[,c('PID',
                                        'IDTYPE',
                                        'RACE',
                                        'ETHNICITY')],
                         by=c("PID","IDTYPE"),
                         all.x=T)

fhs2_omni1_ex02$drinks_wk <- 
  fhs2_omni1_ex02$G104+
  fhs2_omni1_ex02$G107+
  fhs2_omni1_ex02$G110+
  fhs2_omni1_ex02$G113


fhs2_omni1_ex02$bsnq_total <- 
  fhs2_omni1_ex02$G645+
  fhs2_omni1_ex02$G646+
  fhs2_omni1_ex02$G647+
  fhs2_omni1_ex02$G648+
  fhs2_omni1_ex02$G649+
  fhs2_omni1_ex02$G650+
  fhs2_omni1_ex02$G651+
  fhs2_omni1_ex02$G652+
  fhs2_omni1_ex02$G653+
  fhs2_omni1_ex02$G654+
  fhs2_omni1_ex02$G655+
  fhs2_omni1_ex02$G656+
  fhs2_omni1_ex02$G657


fhs2_omni1_ex02$CESD20_missing <- rowSums(is.na(fhs2_omni1_ex02[,443:462]))
fhs2_omni1_ex02$CESD20 <- rowSums(fhs2_omni1_ex02[,443:462])
fhs2_omni1_ex02$CESD20[fhs2_omni1_ex02$CESD20_missing >= 4] <- NA

fhs2_obsper_2005 <- 
  fread('~/Dropbox/BioLINCC files/Framingham offspring/datasets/CSV/t_physf_2005_m_0162d_v2.csv',
        na.strings=c("NA","","NULL"))  # Omni 1, Exam 1
names(fhs2_obsper_2005)[names(fhs2_obsper_2005)=="idtype"] <- "IDTYPE"

omni1_5 <- fhs_dates_long[fhs_dates_long$cohort_name=="OMNI 1"&fhs_dates_long$visit_yr==5,c("PID","visitdays")]
omni1_10 <- fhs_dates_long[fhs_dates_long$cohort_name=="OMNI 1"&fhs_dates_long$visit_yr==10,c("PID","visitdays")]

names(omni1_5)[2] <- "visitdays_omni1_5"
names(omni1_10)[2] <- "visitdays_omni1_10"

fhs2_obsper_2005 <- merge(fhs2_obsper_2005,
                          omni1_5,
                          by="PID",
                          all.x=T)

fhs2_obsper_2005 <- merge(fhs2_obsper_2005,
                          omni1_10,
                          by="PID",
                          all.x=T)


fhs2_obsper_2005_omni1_ex02 <- 
  as.data.table(sqldf("select * 
        from fhs2_obsper_2005
        where mridate >= visitdays_omni1_5
        and mridate < visitdays_omni1_10
        group by PID,IDTYPE
        having min(mridate)"))

fhs2_omni1_ex02 <- merge(fhs2_omni1_ex02,
                         fhs2_obsper_2005_omni1_ex02,
                         by=c("PID","IDTYPE"),
                         all.x=T)

fhs2_omni1_ex02 <- merge(fhs2_omni1_ex02,
                         fhs2_offspring_wkthru[,c("PID",
                                                  "IDTYPE",
                                                  "AGE2",
                                                  "SEX",
                                                  "DMRX2",
                                                  "HRX2",
                                                  "DATE2",
                                                  "HGT1",
                                                  "WGT2",
                                                  "CREAT2",
                                                  "WGT1")],
                         by=c("PID","IDTYPE") )

fhs2_omni1_ex02$BMI2 <- 
  calc_bmi(dat=as.data.frame(fhs2_omni1_ex02), 
           weight="WGT2",
           height="HGT1",
           metric=F)


fhs2_omni1_ex02 <- merge(fhs2_omni1_ex02, 
                         fhs_survcvd,
                         by=c("PID","IDTYPE"),
                         all.x=T)

fhs2_omni1_ex02 <- merge(fhs2_omni1_ex02, 
                         fhs_survstk,
                         by=c("PID","IDTYPE"),
                         all.x=T)


fhs2_omni1_ex02 <- merge(fhs2_omni1_ex02,
                         fhs2_survca_first,
                         by=c("PID","IDTYPE"),
                         all.x=T)


fhs2_omni1_ex02$CESD20_missing <- rowSums(is.na(fhs2_omni1_ex02[,448:467]))
fhs2_omni1_ex02$CESD20 <- rowSums(fhs2_omni1_ex02[,448:467])
fhs2_omni1_ex02$CESD20[fhs2_omni1_ex02$CESD20_missing >= 4] <- NA


####### FRIED frailty score

## Walking speed

# Uses greater of 2 speeds (PMID 26695510)

### Men:
# Height ≤ 173 cm (68.1 in):  ≤0.96 m/s
# Height >173 cm (68.1 in):   ≤1.04  m/s


fhs2_omni1_ex02$gaitspeed_1 <- 4/(fhs2_omni1_ex02$mri339)
fhs2_omni1_ex02$gaitspeed_2 <- 4/(fhs2_omni1_ex02$mri341)

fhs2_omni1_ex02$gaitspeed_avg <- 
  round(apply(fhs2_omni1_ex02[,c("gaitspeed_1","gaitspeed_2")],
              MARGIN=1,
              mean,
              na.rm=T),3)

fhs2_omni1_ex02$gaitspeed_avg[
  fhs2_omni1_ex02$gaitspeed_avg=="NaN"] <- NA

fhs2_omni1_ex02$gaitspeed_max <- 
  round(
    apply(
      fhs2_omni1_ex02[,c("gaitspeed_1","gaitspeed_2")],
      MARGIN=1,max),3)

fhs2_omni1_ex02$gaitspeed_max[
  fhs2_omni1_ex02$gaitspeed_max=="-Inf"] <- NA

fhs2_omni1_ex02$gaitspeed_fried[
  !is.na(fhs2_omni1_ex02$gaitspeed_1)|
    !is.na(fhs2_omni1_ex02$gaitspeed_2)] <- 0

fhs2_omni1_ex02$gaitspeed_fried[
  fhs2_omni1_ex02$SEX==1&
    fhs2_omni1_ex02$HGT1>68.1&
    fhs2_omni1_ex02$gaitspeed_max <=1.04] <- 1

fhs2_omni1_ex02$gaitspeed_fried[
  fhs2_omni1_ex02$SEX==1&
    fhs2_omni1_ex02$HGT1 <= 68.1&
    fhs2_omni1_ex02$gaitspeed_max <= 0.96] <- 1


### Women
# Height ≤ 159 cm (62.6 in):  ≤0.90m/s
# Height >159 cm (62.6 in):   ≤1.02 m/s


fhs2_omni1_ex02$gaitspeed_fried[fhs2_omni1_ex02$SEX==2&
                                  fhs2_omni1_ex02$HGT1>62.6&
                                  fhs2_omni1_ex02$gaitspeed_max <= 1.02] <- 1

fhs2_omni1_ex02$gaitspeed_fried[fhs2_omni1_ex02$SEX==2&
                                  fhs2_omni1_ex02$HGT1 <= 62.6&
                                  fhs2_omni1_ex02$gaitspeed_max <= 0.90] <- 1


########### Grip (kg)

# Used max of all trials per visit  (PMID 26695510)


## Uses BMI for normalization

##### Men
# BMI ≤ 24.0:     ≤ 29 kg
# BMI 24.1-26.0:  ≤ 30 kg
# BMI 26.1-28.0:  ≤ 30 kg
# BMI > 28.0:     ≤ 32 kg

##### Women
# BMI ≤ 23.0:     ≤ 17
# BMI 23.1-26.0:  ≤ 17.3 kg
# BMI 26.1-29.0:  ≤ 18 kg
# BMI > 29.0:     ≤ 21 kg

fhs2_omni1_ex02$grip_avg <- round(apply(fhs2_omni1_ex02[,c("mri356","mri357","mri358","mri360","mri361","mri362")],MARGIN=1,mean),1)
fhs2_omni1_ex02$grip_max <- round(apply(fhs2_omni1_ex02[,c("mri359","mri363")],MARGIN=1,max),1)

fhs2_omni1_ex02$grip_fried[!is.na(fhs2_omni1_ex02$grip_max)] <- 0


##### Men

fhs2_omni1_ex02$grip_fried[fhs2_omni1_ex02$BMI2<=24&
                             fhs2_omni1_ex02$grip_max <= 29&
                             fhs2_omni1_ex02$SEX==1] <- 1

fhs2_omni1_ex02$grip_fried[fhs2_omni1_ex02$BMI2>24&
                             fhs2_omni1_ex02$BMI2<=26&
                             fhs2_omni1_ex02$grip_max <= 30&
                             fhs2_omni1_ex02$SEX==1] <- 1

fhs2_omni1_ex02$grip_fried[fhs2_omni1_ex02$BMI2>26&
                             fhs2_omni1_ex02$BMI2<=28&
                             fhs2_omni1_ex02$grip_max <= 30&
                             fhs2_omni1_ex02$SEX==1] <- 1

fhs2_omni1_ex02$grip_fried[fhs2_omni1_ex02$BMI2>28&
                             fhs2_omni1_ex02$grip_max <= 32&
                             fhs2_omni1_ex02$SEX==1] <- 1

##### Women

fhs2_omni1_ex02$grip_fried[fhs2_omni1_ex02$BMI2<=23&
                             fhs2_omni1_ex02$grip_max <= 17&
                             fhs2_omni1_ex02$SEX==2] <- 1

fhs2_omni1_ex02$grip_fried[fhs2_omni1_ex02$BMI2>23&
                             fhs2_omni1_ex02$BMI2<=26&
                             fhs2_omni1_ex02$grip_max <= 17.3&
                             fhs2_omni1_ex02$SEX==2] <- 1

fhs2_omni1_ex02$grip_fried[fhs2_omni1_ex02$BMI2>26&
                             fhs2_omni1_ex02$BMI2<=29&
                             fhs2_omni1_ex02$grip_max <= 18&
                             fhs2_omni1_ex02$SEX==2] <- 1

fhs2_omni1_ex02$grip_fried[fhs2_omni1_ex02$BMI2>29&
                             fhs2_omni1_ex02$grip_max <= 21&
                             fhs2_omni1_ex02$SEX==2] <- 1

fhs2_omni1_ex02$exhaustion_fried[fhs2_omni1_ex02$G606 %in% c(0,1)|fhs2_omni1_ex02$G593 %in% c(0,1)] <- 0
fhs2_omni1_ex02$exhaustion_fried[fhs2_omni1_ex02$G606 %in% c(2,3)|fhs2_omni1_ex02$G593 %in% c(2,3)] <- 1


#### Weight loss - Fried

fhs2_omni1_ex02$wt_loss_pct <- round((fhs2_omni1_ex02$WGT1 - fhs2_omni1_ex02$WGT2)/fhs2_omni1_ex02$WGT2*100,1)
fhs2_omni1_ex02$wtloss_fried[fhs2_omni1_ex02$wt_loss_pct >= 5] <- 1
fhs2_omni1_ex02$wtloss_fried[fhs2_omni1_ex02$wt_loss_pct < 5] <- 0


#### Physical activity - Fried

fhs2_omni1_ex02$fhs_pai <- 
  calc_framingham_pai(
    as.data.frame(fhs2_omni1_ex02),
    slp_hrs = "G689",
    sed_hrs = "G690",
    slgt_hrs = "G691",
    mod_hrs = "G692",
    hvy_hrs = "G693")

fhs2_omni1_ex02$activity_fried[fhs2_omni1_ex02$SEX==1&
                                 fhs2_omni1_ex02$fhs_pai < 30.3] <- 1

fhs2_omni1_ex02$activity_fried[fhs2_omni1_ex02$SEX==1&
                                 fhs2_omni1_ex02$fhs_pai >= 30.3] <- 0


fhs2_omni1_ex02$activity_fried[fhs2_omni1_ex02$SEX==2&
                                 fhs2_omni1_ex02$fhs_pai < 30.1] <- 1

fhs2_omni1_ex02$activity_fried[fhs2_omni1_ex02$SEX==2&
                                 fhs2_omni1_ex02$fhs_pai >= 30.1] <- 0


#### Calculate Fried score

fhs2_omni1_ex02$total_fried <-
  fhs2_omni1_ex02$gaitspeed_fried+
  fhs2_omni1_ex02$grip_fried+
  fhs2_omni1_ex02$exhaustion_fried+
  fhs2_omni1_ex02$wtloss_fried+
  fhs2_omni1_ex02$activity_fried


######### FRAIL Score

#### Fatigue

fhs2_omni1_ex02$fatigue_frail[fhs2_omni1_ex02$G606 %in% c(0,1)|fhs2_omni1_ex02$G593 %in% c(0,1)] <- 0
fhs2_omni1_ex02$fatigue_frail[fhs2_omni1_ex02$G606 %in% c(2,3)|fhs2_omni1_ex02$G593 %in% c(2,3)] <- 1


#### Resistance

fhs2_omni1_ex02$resistance_frail[fhs2_omni1_ex02$G525==0] <- 0
fhs2_omni1_ex02$resistance_frail[fhs2_omni1_ex02$G525 %in% c(1,2,3,4)] <- 1


#### Ambulation

fhs2_omni1_ex02$ambulate_frail[fhs2_omni1_ex02$G548==1] <- 0
fhs2_omni1_ex02$ambulate_frail[fhs2_omni1_ex02$G548==0] <- 1


# FRAIL Illness score counts presence of HTN, DM, cancer, chronic lung disease, MI, CHD, angina, asthma, arthritis, stroke, and ckd
# Scores for 0-4 conditions, score = 0, for ≥ 5, score=1

# HTN

fhs2_omni1_ex02$htn_frail[fhs2_omni1_ex02$HRX2==0] <- 0
fhs2_omni1_ex02$htn_frail[fhs2_omni1_ex02$HRX2==1] <- 1

# COPD (chronic lung disease)

fhs2_omni1_ex02$copd_frail[fhs2_omni1_ex02$G415  %in% c(0,2)] <- 0
fhs2_omni1_ex02$copd_frail[fhs2_omni1_ex02$G415==1] <- 1

# Asthma

fhs2_omni1_ex02$asthma_frail[fhs2_omni1_ex02$G418 %in% c(0,2)] <- 0
fhs2_omni1_ex02$asthma_frail[fhs2_omni1_ex02$G418==1] <- 1

# DJD (arthritis)

fhs2_omni1_ex02$djd_frail[fhs2_omni1_ex02$G421 %in% c(0,2)] <- 0
fhs2_omni1_ex02$djd_frail[fhs2_omni1_ex02$G421==1] <- 1


# CKD (renal disease)


fhs2_omni1_ex02$crcl <- 
  calc_MDRD4(
    dat=as.data.frame(fhs2_omni1_ex02),
    age="AGE2",
    cr="CREAT2",
    race="RACE",
    black="B")

fhs2_omni1_ex02$ckd_frail[fhs2_omni1_ex02$crcl >= 60] <- 0
fhs2_omni1_ex02$ckd_frail[fhs2_omni1_ex02$crcl < 60] <- 1


# CHF/MI

fhs2_omni1_ex02 <- merge(fhs2_omni1_ex02,
                         fhs2_survcvd,
                         by=c("PID",
                              "IDTYPE"))
fhs2_omni1_ex02$chf_frail <- 0
fhs2_omni1_ex02$chf_frail[fhs2_omni1_ex02$hfhosp_status==1&fhs2_omni1_ex02$hfhosp_dt <= fhs2_omni1_ex02$DATE7] <- 1

fhs2_omni1_ex02$chd_frail <- 0
fhs2_omni1_ex02$chd_frail[fhs2_omni1_ex02$cadhosp_status==1&fhs2_omni1_ex02$cadhosp_dt <= fhs2_omni1_ex02$DATE7] <- 1


# Stroke

fhs2_omni1_ex02$stroke_frail <- 0
fhs2_omni1_ex02$stroke_frail[fhs2_omni1_ex02$STROKE==1&fhs2_omni1_ex02$STROKEDATE <= fhs2_omni1_ex02$DATE7] <- 1


# Cancer

fhs2_omni1_ex02$cancer_frail <- 0 
fhs2_omni1_ex02$cancer_frail[fhs2_omni1_ex02$D_DATE <= fhs2_omni1_ex02$DATE7] <- 1


# Sum conditions

fhs2_omni1_ex02$conditions_frail <-
  fhs2_omni1_ex02$htn_frail+
  fhs2_omni1_ex02$DMRX2+
  fhs2_omni1_ex02$copd_frail+
  fhs2_omni1_ex02$asthma_frail+
  fhs2_omni1_ex02$djd_frail+
  fhs2_omni1_ex02$ckd_frail+
  fhs2_omni1_ex02$chf_frail+
  fhs2_omni1_ex02$chd_frail+
  fhs2_omni1_ex02$stroke_frail+
  fhs2_omni1_ex02$cancer_frail

fhs2_omni1_ex02$illness_frail[fhs2_omni1_ex02$conditions_frail <= 4] <- 0
fhs2_omni1_ex02$illness_frail[fhs2_omni1_ex02$conditions_frail > 4] <- 1

#### (Weight) Loss

fhs2_omni1_ex02$wtloss_frail[fhs2_omni1_ex02$wt_loss_pct >= 5] <- 1
fhs2_omni1_ex02$wtloss_frail[fhs2_omni1_ex02$wt_loss_pct < 5] <- 0

#### Calculate FRAIL score

fhs2_omni1_ex02$total_frail <- 
  fhs2_omni1_ex02$fatigue_frail+
  fhs2_omni1_ex02$resistance_frail+
  fhs2_omni1_ex02$ambulate_frail+
  fhs2_omni1_ex02$illness_frail+
  fhs2_omni1_ex02$wtloss_frail


fhs2_omni1_ex02$visit <- 2

fhs2_melt_omni1_ex02 <- 
  melt(
    fhs2_omni1_ex02,
    id.vars=c("PID","IDTYPE","visit"), 
    na.rm=T, 
    factorsAsStrings=T)

fhs2_melt_omni1_ex02$form <- "fhs2_omni1_ex02"

rm(fhs2_omni1_ex02)


#===================================#
####     Lab panel, Exam 2       ---#
#===================================#

fhs2_l_fhslab_ex02 <- fread('~/Dropbox/BioLINCC files/Framingham offspring/datasets/CSV/l_fhslab_ex02_7_0561d.csv',na.strings=c("NA","","NULL"))
names(fhs2_l_fhslab_ex02)[names(fhs2_l_fhslab_ex02)=="idtype"] <- "IDTYPE"
fhs2_l_fhslab_ex02$visit <- 2
fhs2_melt_l_fhslab_ex02 <- melt(fhs2_l_fhslab_ex02,id.vars=c("PID","IDTYPE","visit"), na.rm=T, factorsAsStrings=T)
fhs2_melt_l_fhslab_ex02$form <- "fhs2_fhslab_ex02_7"

rm(fhs2_l_fhslab_ex02)


#===================================#
####     Food freq, Exam 2       ---#
#===================================#

fhs2_vr_ffreq_omni1_ex02 <- fread('~/Dropbox/BioLINCC files/Framingham offspring/datasets/CSV/vr_ffreq_ex02_7_1301d.csv',na.strings=c("NA","","NULL"))
names(fhs2_vr_ffreq_omni1_ex02)[names(fhs2_vr_ffreq_omni1_ex02)=="IDType"] <- "IDTYPE"
fhs2_vr_ffreq_omni1_ex02$visit <- 2
fhs2_melt_vr_ffreq_omni1_ex02 <- melt(fhs2_vr_ffreq_omni1_ex02,id.vars=c("PID","IDTYPE","visit"), na.rm=T, factorsAsStrings=T)
fhs2_melt_vr_ffreq_omni1_ex02$form <- "fhs2_vr_ffreq"

rm(fhs2_vr_ffreq_omni1_ex02)


#===============================#
#### *** OMNI 1, exam 3 ***  ####
#===============================#

fhs2_omni1_ex03 <- fread('~/Dropbox/BioLINCC files/Framingham offspring/datasets/CSV/e_exam_ex03_7_0426d.csv',na.strings=c("NA","","NULL")) # Omni 1, Exam 3
names(fhs2_omni1_ex03) <- toupper(names(fhs2_omni1_ex03))
fhs2_omni1_ex03$visit <- 3

fhs2_omni1_ex03 <- 
  merge(
    fhs2_omni1_ex03,
    fhs2_offspring_wkthru[,c("PID",
                             "DMRX3",
                             "HRX3",
                             "DATE3",
                             "AGE3",
                             "WGT3",
                             "WGT2",
                             "SEX",
                             "CREAT3",
                             "BMI3")],
    by=c("PID"),
    all.x=T)


fhs2_omni1_ex03$drinks_wk <- 
  fhs2_omni1_ex03$H072+
  fhs2_omni1_ex03$H075+
  fhs2_omni1_ex03$H078

fhs2_omni1_ex03$epworth_ss_nas <- 
  is.na(fhs2_omni1_ex03$H734)+
  is.na(fhs2_omni1_ex03$H735)+
  is.na(fhs2_omni1_ex03$H736)+
  is.na(fhs2_omni1_ex03$H737)+
  is.na(fhs2_omni1_ex03$H738)+
  is.na(fhs2_omni1_ex03$H739)+
  is.na(fhs2_omni1_ex03$H740)+
  is.na(fhs2_omni1_ex03$H741)

fhs2_omni1_ex03$H033[!between(fhs2_omni1_ex03$H033,22,83)] <- NA
fhs2_omni1_ex03$H037[!between(fhs2_omni1_ex03$H037,18,83)] <- NA
fhs2_omni1_ex03$H046[!between(fhs2_omni1_ex03$H046,21,77)] <- NA

fhs2_omni1_ex03$hrt_years_total[between(fhs2_omni1_ex03$H047,1,42)&!is.na(fhs2_omni1_ex03$H047)] <- 
  fhs2_omni1_ex03$H047[between(fhs2_omni1_ex03$H047,1,42)&!is.na(fhs2_omni1_ex03$H047)]

fhs2_omni1_ex03$hrt_years_total[between(fhs2_omni1_ex03$H048,1,10)&!is.na(fhs2_omni1_ex03$H048)] <- 
  fhs2_omni1_ex03$hrt_years_total[between(fhs2_omni1_ex03$H048,1,10)&!is.na(fhs2_omni1_ex03$H048)] + 
  fhs2_omni1_ex03$H048[between(fhs2_omni1_ex03$H048,1,10)&!is.na(fhs2_omni1_ex03$H048)]/12


fhs2_omni1_ex03$epworth_ss <- 
  fhs2_omni1_ex03$H734+
  fhs2_omni1_ex03$H735+
  fhs2_omni1_ex03$H736+
  fhs2_omni1_ex03$H737+
  fhs2_omni1_ex03$H738+
  fhs2_omni1_ex03$H739+
  fhs2_omni1_ex03$H740+
  fhs2_omni1_ex03$H741


########### FRIED

## Walking speed

# Uses greater of 2 speeds (PMID 26695510)


## Walking speed

fhs2_omni1_ex03$walktime_1 <- fhs2_omni1_ex03$H617
fhs2_omni1_ex03$walktime_2 <- fhs2_omni1_ex03$H620

fhs2_omni1_ex03$walktime_avg <- round(apply(fhs2_omni1_ex03[,c("walktime_1","walktime_2")],
                                            MARGIN=1,
                                            mean,
                                            na.rm=T),3)
fhs2_omni1_ex03$walktime_avg[fhs2_omni1_ex03$walktime_avg=="NaN"] <- NA

fhs2_omni1_ex03$walktime_min <- round(apply(fhs2_omni1_ex03[,c("walktime_1","walktime_2")],
                                            MARGIN=1,min),3)
fhs2_omni1_ex03$walktime_min[fhs2_omni1_ex03$walktime_min=="-Inf"] <- NA

fhs2_omni1_ex03$walktime_min[!is.na(fhs2_omni1_ex03$walktime_1)|
                               !is.na(fhs2_omni1_ex03$walktime_2)] <- 0 


########### Grip (kg)

# Produces highest average by hand (assumes dominant hand is stronger)

fhs2_omni1_ex03$grip_avg_right <- round(apply(fhs2_omni1_ex03[,407:409],MARGIN=1,mean),1)
fhs2_omni1_ex03$grip_avg_left <- round(apply(fhs2_omni1_ex03[,410:412],MARGIN=1,mean),1)

fhs2_omni1_ex03$grip_max <- round(apply(fhs2_omni1_ex03[,c("grip_avg_right","grip_avg_left")],MARGIN=1,max),1)


#### Exhaustion

fhs2_omni1_ex03$exhaustion_fried[fhs2_omni1_ex03$H471==3|fhs2_omni1_ex03$H472==3] <- 1
fhs2_omni1_ex03$exhaustion_fried[fhs2_omni1_ex03$H471<3&fhs2_omni1_ex03$H472<3] <- 0


#### Weight loss

fhs2_omni1_ex03$wtloss_fried[fhs2_omni1_ex03$H401==1] <- 1
fhs2_omni1_ex03$wtloss_fried[fhs2_omni1_ex03$H401 %in% c(0,2)] <- 1


#### Physical activity

fhs2_omni1_ex03$fhs_pai <- 
  calc_framingham_pai(
    as.data.frame(fhs2_omni1_ex03),
    slp_hrs="H480",
    sed_hrs="H481",
    slgt_hrs="H482",
    mod_hrs="H483",
    hvy_hrs="H484")

fhs2_omni1_ex03$physical_activity_fried[fhs2_omni1_ex03$SEX==1&
                                          fhs2_omni1_ex03$fhs_pai < 30.3] <- 1

fhs2_omni1_ex03$physical_activity_fried[fhs2_omni1_ex03$SEX==1&
                                          fhs2_omni1_ex03$fhs_pai >= 30.3] <- 0


fhs2_omni1_ex03$physical_activity_fried[fhs2_omni1_ex03$SEX==2&
                                          fhs2_omni1_ex03$fhs_pai < 30.1] <- 1

fhs2_omni1_ex03$physical_activity_fried[fhs2_omni1_ex03$SEX==2&
                                          fhs2_omni1_ex03$fhs_pai >= 30.1] <- 0


######### FRAIL Score

#### Fatigue

fhs2_omni1_ex03$fatigue_frail[fhs2_omni1_ex03$H723 <=3] <- 0
fhs2_omni1_ex03$fatigue_frail[fhs2_omni1_ex03$H723 > 3] <- 1


#### Resistance

fhs2_omni1_ex03$resistance_frail[fhs2_omni1_ex03$H470==1] <- 0
fhs2_omni1_ex03$resistance_frail[fhs2_omni1_ex03$H470==0] <- 1

#### Ambulation

fhs2_omni1_ex03$ambulate_frail[fhs2_omni1_ex03$H469==1] <- 0
fhs2_omni1_ex03$ambulate_frail[fhs2_omni1_ex03$H469==0] <- 1


# FRAIL Illness score counts presence of HTN, DM, cancer, chronic lung disease, MI, CHD, angina, asthma, arthritis, stroke, and ckd
# Scores for 0-4 conditions, score = 0, for ≥ 5, score=1


# HTN

fhs2_omni1_ex03$htn_frail[fhs2_omni1_ex03$H014 %in% c(0,2)] <- 0
fhs2_omni1_ex03$htn_frail[fhs2_omni1_ex03$H014==1] <- 1

# COPD (chronic lung disease)

fhs2_omni1_ex03$copd_frail[fhs2_omni1_ex03$H357 %in% c(0,2)] <- 0
fhs2_omni1_ex03$copd_frail[fhs2_omni1_ex03$H357==1] <- 1

# asthma

fhs2_omni1_ex03$asthma_frail[fhs2_omni1_ex03$H359 %in% c(0,2)] <- 0
fhs2_omni1_ex03$asthma_frail[fhs2_omni1_ex03$H359==1] <- 1

# DJD (arthritis)

fhs2_omni1_ex03$djd_frail[fhs2_omni1_ex03$H362 %in% c(0,2)] <- 0
fhs2_omni1_ex03$djd_frail[fhs2_omni1_ex03$H362==1] <- 1

# CKD (renal disease)

fhs2_omni1_ex03$RACE <- 2

fhs2_omni1_ex03$crcl <- 
  calc_MDRD4(
    dat=as.data.frame(fhs2_omni1_ex03),
    cr="CREAT3",
    age="AGE3")

fhs2_omni1_ex03$ckd_frail[fhs2_omni1_ex03$H354 %in% c(0,2)] <- 0
fhs2_omni1_ex03$ckd_frail[fhs2_omni1_ex03$H354==1|fhs2_omni1_ex03$crcl < 60] <- 1

# CHF/MI

fhs2_omni1_ex03 <- merge(fhs2_omni1_ex03,fhs_survcvd,by=c("PID","IDTYPE"))

fhs2_omni1_ex03$chf_frail <- 0
fhs2_omni1_ex03$chf_frail[fhs2_omni1_ex03$hfhosp_status==1&fhs2_omni1_ex03$hfhosp_dt <= fhs2_omni1_ex03$DATE3] <- 1

fhs2_omni1_ex03$chd_frail <- 0
fhs2_omni1_ex03$chd_frail[fhs2_omni1_ex03$cadhosp_status==1&fhs2_omni1_ex03$cadhosp_dt <= fhs2_omni1_ex03$DATE3] <- 1

# Stroke

fhs2_omni1_ex03 <- merge(fhs2_omni1_ex03,fhs_survstk,by=c("PID","IDTYPE"))

fhs2_omni1_ex03$stroke_frail <- 0
fhs2_omni1_ex03$stroke_frail[fhs2_omni1_ex03$STROKE==1&fhs2_omni1_ex03$STROKEDATE <= fhs2_omni1_ex03$DATE3] <- 1

# Cancer

fhs2_omni1_ex03 <- merge(fhs2_omni1_ex03,
                         fhs2_survca_first[,c("PID","IDTYPE","D_DATE")],
                         by=c("PID","IDTYPE"),
                         all.x=T)

fhs2_omni1_ex03$cancer_frail <- 0
fhs2_omni1_ex03$cancer_frail[fhs2_omni1_ex03$D_DATE <= fhs2_omni1_ex03$DATE3] <- 1

fhs2_omni1_ex03$conditions_frail <- 
  fhs2_omni1_ex03$htn_frail+
  fhs2_omni1_ex03$DMRX3+
  fhs2_omni1_ex03$copd_frail+
  fhs2_omni1_ex03$asthma_frail+
  fhs2_omni1_ex03$djd_frail+
  fhs2_omni1_ex03$ckd_frail+
  fhs2_omni1_ex03$chf_frail+
  fhs2_omni1_ex03$chd_frail+
  fhs2_omni1_ex03$stroke_frail+
  fhs2_omni1_ex03$cancer_frail

fhs2_omni1_ex03$illness_frail[fhs2_omni1_ex03$conditions_frail <= 4] <- 0
fhs2_omni1_ex03$illness_frail[fhs2_omni1_ex03$conditions_frail > 4] <- 1

#### (Weight) Loss

fhs2_omni1_ex03$wtloss_frail[fhs2_omni1_ex03$H401==1] <- 1
fhs2_omni1_ex03$wtloss_frail[fhs2_omni1_ex03$H401 %in% c(0,2)] <- 0

fhs2_omni1_ex03$wgt_delta <- fhs2_omni1_ex03$WGT3 - fhs2_omni1_ex03$WGT2


fhs2_melt_omni1_ex03 <- melt(fhs2_omni1_ex03,id.vars=c("PID","IDTYPE","visit"), na.rm=T, factorsAsStrings=T)
fhs2_melt_omni1_ex03$form <- "fhs2_omni1_ex03"

rm(fhs2_omni1_ex03)

#===================================#
####     Lab Panel, Exam 3       ---#
#===================================#

fhs2_l_fhslab_ex03 <- fread('~/Dropbox/BioLINCC files/Framingham offspring/datasets/CSV/l_fhslab_ex03_7_0267d.csv',na.strings=c("NA","","NULL"))
names(fhs2_l_fhslab_ex03)[names(fhs2_l_fhslab_ex03)=="idtype"] <- "IDTYPE"
fhs2_l_fhslab_ex03$visit <- 3
fhs2_melt_l_fhslab_ex03 <- melt(fhs2_l_fhslab_ex03,id.vars=c("PID","IDTYPE","visit"), na.rm=T, factorsAsStrings=T)
fhs2_melt_l_fhslab_ex03$form <- "fhs2_fhslab_ex03_7"

rm(fhs2_l_fhslab_ex03)

#===================================#
####     Food freq, Exam 3       ---#
#===================================#

fhs2_vr_ffreq_omni1_ex03 <- fread('~/Dropbox/BioLINCC files/Framingham offspring/datasets/CSV/vr_ffreq_ex03_7_0973d.csv',na.strings=c("NA","","NULL"))
names(fhs2_vr_ffreq_omni1_ex03)[names(fhs2_vr_ffreq_omni1_ex03)=="idtype"] <- "IDTYPE"
fhs2_vr_ffreq_omni1_ex03$visit <- 3
fhs2_melt_vr_ffreq_omni1_ex03 <- melt(fhs2_vr_ffreq_omni1_ex03,id.vars=c("PID","IDTYPE","visit"), na.rm=T, factorsAsStrings=T)
fhs2_melt_vr_ffreq_omni1_ex03$form <- "fhs2_vr_ffreq"

rm(fhs2_vr_ffreq_omni1_ex03)


# ===============================================##
####  CRP, Offspring Exam 8, Omni 1 Exam 3     ---#
# ===============================================##

fhs2_inflamm_ex08 <- fread('~/Dropbox/BioLINCC files/Framingham offspring/datasets/CSV/l_inflamm_ex08_1b_1106d.csv',na.strings=c("NA","","NULL"))

names(fhs2_inflamm_ex08) <- toupper(names(fhs2_inflamm_ex08))


fhs2_inflamm_ex08$visit[fhs2_inflamm_ex08$IDTYPE==1] <- 8
fhs2_inflamm_ex08$visit[fhs2_inflamm_ex08$IDTYPE==7] <- 3

fhs2_melt_inflamm_ex08 <- 
  melt(fhs2_inflamm_ex08,
       id.vars=c("PID","IDTYPE","visit"), 
       na.rm=T, 
       factorsAsStrings=T)

fhs2_melt_inflamm_ex08$form <- "fhs2_inflamm_ex08"

rm(fhs2_inflamm_ex08)


#==============================================#
####  CES-D Offspring exam 8, OMNI1 exam 3  ---#
#==============================================#


fhs2_q_cesd_ex09 <- fread('~/Dropbox/BioLINCC files/Framingham offspring/datasets/CSV/q_cesd_2009_m_0570sd.csv',na.strings=c("NA","","NULL"))
names(fhs2_q_cesd_ex09)[names(fhs2_q_cesd_ex09)=="idtype"] <- "IDTYPE"
fhs2_q_cesd_ex09$visit[fhs2_q_cesd_ex09$IDTYPE==1] <- 8
fhs2_q_cesd_ex09$visit[fhs2_q_cesd_ex09$IDTYPE==7] <- 3
fhs2_q_cesd_ex09$CESD20_missing <- rowSums(is.na(fhs2_q_cesd_ex09[,2:21]))
fhs2_q_cesd_ex09$CESD20 <- rowSums(fhs2_q_cesd_ex09[,2:21])
fhs2_q_cesd_ex09$CESD20[fhs2_q_cesd_ex09$CESD20_missing >= 4] <- NA

fhs2_q_cesd_ex09 <- 
  as.data.table(
    sqldf("select * 
          from fhs2_q_cesd_ex09
          where CESD20 is not null
          group by PID, IDTYPE, VISIT
          having min(testdate)"))


fhs2_melt_q_cesd_ex09 <- 
  melt(fhs2_q_cesd_ex09,
       id.vars=c("PID","IDTYPE","visit"), 
       na.rm=T, 
       factorsAsStrings=T)

fhs2_melt_q_cesd_ex09$form <- "fhs2_q_cesd"

rm(fhs2_q_cesd_ex09)


#========================================================#
####  Echo Doppler - Offspring Exam 8, OMNI 1 Exam 3  ---#
#========================================================#

fhs2_t_doppvasc <- fread('~/Dropbox/BioLINCC files/Framingham offspring/datasets/CSV/t_doppvasc_2008_m_0756d.csv',na.strings=c("NA","","NULL"))
names(fhs2_t_doppvasc)[names(fhs2_t_doppvasc)=="idtype"] <- "IDTYPE"
fhs2_t_doppvasc$visit[fhs2_t_doppvasc$IDTYPE==1] <- 8
fhs2_t_doppvasc$visit[fhs2_t_doppvasc$IDTYPE==7] <- 3

fhs2_t_doppvasc$earatio <- fhs2_t_doppvasc$MEMAX/fhs2_t_doppvasc$MAMAX
fhs2_t_doppvasc$e_eprime_lateral <- fhs2_t_doppvasc$MEMAX/fhs2_t_doppvasc$DEMAX

fhs2_melt_t_doppvasc <- melt(fhs2_t_doppvasc,id.vars=c("PID","IDTYPE","visit"), na.rm=T, factorsAsStrings=T)
fhs2_melt_t_doppvasc$form <- "fhs_doppvasc_2008"

rm(fhs2_t_doppvasc)


#================================================#
####  Echo - Offspring Exam 8, OMNI 1 Exam 3  ---#
#=========s=======================================#

fhs2_t_echo_2008 <- fread('~/Dropbox/BioLINCC files/Framingham offspring 2022a/datasets/CSV/t_echo_2008_m_0549d_v1.csv',na.strings=c("NA","","NULL"))
names(fhs2_t_echo_2008)[names(fhs2_t_echo_2008)=="idtype"] <-"IDTYPE"
names(fhs2_t_echo_2008) <- toupper(names(fhs2_t_echo_2008))
names(fhs2_t_echo_2008)[names(fhs2_t_echo_2008)=="EXAM"] <-"visit"

fhs2_t_echo_2008$visit <- as.character(fhs2_t_echo_2008$visit)

fhs2_t_echo_2008 <- merge(fhs2_t_echo_2008,
                          fhs2_wkthru[,c("PID","IDTYPE","SEX","visit","WGT","HGT")],
                          by=c("PID","IDTYPE","visit"), all.x=T)

fhs2_t_echo_2008$HEIGHT <- as.numeric(fhs2_t_echo_2008$HGT)*2.54
fhs2_t_echo_2008$WEIGHT <- as.numeric(fhs2_t_echo_2008$WGT)/2.2

fhs2_t_echo_2008 <- 
  as.data.table(
    calc_hypertrophy_type(
      as.data.frame(fhs2_t_echo_2008), 
      id = "PID",
      sex="SEX",
      lvedd="X76",
      ivsd="X62",
      lvpwtd="X69",
      height="HEIGHT",
      weight="WEIGHT"))

fhs2_melt_t_echo_2008 <- melt(fhs2_t_echo_2008,id.vars=c("PID","IDTYPE","visit"), na.rm=T, factorsAsStrings=T)

fhs2_melt_t_echo_2008$form <- "fhs2_echo_2008"

rm(fhs2_t_echo_2008)


#==========================================#
####     Echo cardiac strain, Omni 3    ---#
#==========================================#

fhs2_t_echocs_ex03 <- 
  fread('~/Dropbox/BioLINCC files/Framingham offspring/datasets/CSV/t_echocs_ex03_7_0901d.csv',
        na.strings=c("NA","","NULL"))

names(fhs2_t_echocs_ex03)[names(fhs2_t_echocs_ex03)=="idtype"] <- "IDTYPE"
fhs2_t_echocs_ex03$visit <- 3

names(fhs2_t_echocs_ex03) <-
  sub("^_","",names(fhs2_t_echocs_ex03))

fhs2_melt_t_echocs_ex03 <- melt(fhs2_t_echocs_ex03,id.vars=c("PID","IDTYPE","visit"), 
                                na.rm=T, factorsAsStrings=T)

fhs2_melt_t_echocs_ex03$form <- "fhs2_omni1_echocs_ex03"

rm(fhs2_t_echocs_ex03)


#=================================================#
####  *** Offspring exam 9, Omni 1 exam 4 ***  ####
#=================================================#

fhs2_offspring_ex09_omni1_ex04 <- 
  fread('~/Dropbox/BioLINCC files/Framingham offspring/datasets/CSV/e_exam_ex09_1b_0844d.csv',
        na.strings=c("NA","","NULL")) # Omni1, Exam 9

names(fhs2_offspring_ex09_omni1_ex04) <- 
  toupper(names(fhs2_offspring_ex09_omni1_ex04))

fhs2_offspring_ex09_omni1_ex04$CESD20_missing <- 
  rowSums(is.na(fhs2_offspring_ex09_omni1_ex04[,427:446]))
fhs2_offspring_ex09_omni1_ex04$CESD20 <- 
  rowSums(fhs2_offspring_ex09_omni1_ex04[,427:446])

fhs2_offspring_ex09_omni1_ex04$CESD20[
  fhs2_offspring_ex09_omni1_ex04$CESD20_missing >= 4] <- NA

fhs2_offspring_ex09_omni1_ex04$visit[fhs2_offspring_ex09_omni1_ex04$IDTYPE==1] <- 9
fhs2_offspring_ex09_omni1_ex04$visit[fhs2_offspring_ex09_omni1_ex04$IDTYPE==7] <- 4

fhs2_offspring_ex09_omni1_ex04$drinks_wk <- fhs2_offspring_ex09_omni1_ex04$J075+
  fhs2_offspring_ex09_omni1_ex04$J077+
  fhs2_offspring_ex09_omni1_ex04$J079

fhs2_offspring_ex09_omni1_ex04$J055[!between(fhs2_offspring_ex09_omni1_ex04$J055,39,79)] <- NA


fhs2_offspring_ex09_omni1_ex04 <- 
  merge(fhs2_offspring_ex09_omni1_ex04,
        fhs2_wkthru[fhs2_wkthru$visit==1,
                    c("PID",
                      "IDTYPE",
                      "SEX",
                      "HGT")],
        by=c("PID","IDTYPE"),
        all.x=T)

fhs2_omni1_ex04_wgt3 <- 
  fhs2_wkthru[fhs2_wkthru$visit==3&
                fhs2_wkthru$IDTYPE==7,
              c("PID",
                "IDTYPE",
                "WGT")]

fhs2_offspring_ex09_wgt8 <- 
  fhs2_wkthru[fhs2_wkthru$visit==8&
                fhs2_wkthru$IDTYPE==1,
              c("PID",
                "IDTYPE",
                "WGT")]

fhs2_offspring_ex09_omni1_ex04_lastwgt <- 
  rbind(fhs2_omni1_ex04_wgt3,
        fhs2_offspring_ex09_wgt8)

names(fhs2_offspring_ex09_omni1_ex04_lastwgt)[3] <- "last_wgt"

fhs2_omni1_ex04_wgt4 <- 
  fhs2_wkthru[fhs2_wkthru$visit==4&
                fhs2_wkthru$IDTYPE==7,
              c("PID",
                "IDTYPE",
                "WGT")]

fhs2_offspring_ex09_wgt9 <- 
  fhs2_wkthru[fhs2_wkthru$visit==9&
                fhs2_wkthru$IDTYPE==1,
              c("PID",
                "IDTYPE",
                "WGT")]


fhs2_offspring_ex09_omni1_ex04_wgt <- 
  rbind(fhs2_omni1_ex04_wgt4,
        fhs2_offspring_ex09_wgt9)

fhs2_offspring_ex09_omni1_ex04 <- 
  merge(fhs2_offspring_ex09_omni1_ex04,
        fhs2_offspring_ex09_omni1_ex04_wgt,
        by=c("PID","IDTYPE"),
        all.x=T)

fhs2_offspring_ex09_omni1_ex04 <- 
  merge(fhs2_offspring_ex09_omni1_ex04,
        fhs2_offspring_ex09_omni1_ex04_lastwgt,
        by=c("PID","IDTYPE"),
        all.x=T)


fhs2_offspring_ex09_bmi <- 
  fhs2_wkthru[fhs2_wkthru$visit==9&
                fhs2_wkthru$IDTYPE==1&
                !is.na(fhs2_wkthru$BMI),
              c("PID",
                "IDTYPE",
                "BMI",
                "CREAT",
                "DMRX",
                "HRX",
                "AGE")]

fhs2_omni1_ex04_bmi <- 
  fhs2_wkthru[fhs2_wkthru$visit==4&
                fhs2_wkthru$IDTYPE==7&
                !is.na(fhs2_wkthru$BMI),
              c("PID",
                "IDTYPE",
                "BMI",
                "CREAT",
                "DMRX",
                "HRX",
                "AGE")]

fhs2_off9_omni4_bmi <- 
  rbind(fhs2_offspring_ex09_bmi,
        fhs2_omni1_ex04_bmi)

fhs2_offspring_ex09_omni1_ex04 <- 
  merge(fhs2_offspring_ex09_omni1_ex04,
        fhs2_off9_omni4_bmi,
        by=c("PID","IDTYPE"),
        all.x=T)


fhs2_offspring_ex09_omni1_ex04 <- 
  merge(fhs2_offspring_ex09_omni1_ex04,
        fhs2_survcvd,
        by=c("PID",
             "IDTYPE"))


fhs2_offspring_ex09_omni1_ex04 <- 
  merge(fhs2_offspring_ex09_omni1_ex04,
        fhs2_survstk,
        by=c("PID",
             "IDTYPE"))


fhs2_offspring_ex09_omni1_ex04 <- 
  merge(fhs2_offspring_ex09_omni1_ex04,
        fhs_survca_first_any[,c("PID",
                                "IDTYPE",
                                "D_DATE")],
        by=c("PID",
             "IDTYPE"),
        all.x=T)


## Walking speed - Fried

# Uses greater of 2 speeds (PMID 26695510)

fhs2_offspring_ex09_omni1_ex04$walktime_1 <- fhs2_offspring_ex09_omni1_ex04$J785
fhs2_offspring_ex09_omni1_ex04$walktime_2 <- fhs2_offspring_ex09_omni1_ex04$J790

fhs2_offspring_ex09_omni1_ex04$walktime_avg <- round(apply(fhs2_offspring_ex09_omni1_ex04[,c("walktime_1","walktime_2")],
                                                           MARGIN=1,
                                                           mean,
                                                           na.rm=T),3)
fhs2_offspring_ex09_omni1_ex04$walktime_avg[fhs2_offspring_ex09_omni1_ex04$walktime_avg=="NaN"] <- NA

fhs2_offspring_ex09_omni1_ex04$walktime_min <- round(apply(fhs2_offspring_ex09_omni1_ex04[,c("walktime_1","walktime_2")],
                                                           MARGIN=1,min),3)
fhs2_offspring_ex09_omni1_ex04$walktime_min[fhs2_offspring_ex09_omni1_ex04$walktime_min=="-Inf"] <- NA

fhs2_offspring_ex09_omni1_ex04$walktime_min[!is.na(fhs2_offspring_ex09_omni1_ex04$walktime_1)|
                                              !is.na(fhs2_offspring_ex09_omni1_ex04$walktime_2)] <- 0 


########### Grip (kg)

# Produces highest average by hand (assumes dominant hand is stronger)

fhs2_offspring_ex09_omni1_ex04$grip_avg_right <- round(apply(fhs2_offspring_ex09_omni1_ex04[,451:453],MARGIN=1,mean),1)
fhs2_offspring_ex09_omni1_ex04$grip_avg_left <- round(apply(fhs2_offspring_ex09_omni1_ex04[,454:456],MARGIN=1,mean),1)

fhs2_offspring_ex09_omni1_ex04$grip_max <- round(apply(fhs2_offspring_ex09_omni1_ex04[,c("grip_avg_right","grip_avg_left")],MARGIN=1,max),1)


#### Physical activity index - Fried

fhs2_offspring_ex09_omni1_ex04$fhs_pai <- 
  calc_framingham_pai(
    as.data.frame(fhs2_offspring_ex09_omni1_ex04),
    slp_hrs="J628",
    sed_hrs="J629",
    slgt_hrs="J630",
    mod_hrs="J631",
    hvy_hrs="J632")

fhs2_offspring_ex09_omni1_ex04$physical_activity_fried[fhs2_offspring_ex09_omni1_ex04$SEX==1&
                                                         fhs2_offspring_ex09_omni1_ex04$fhs_pai < 30.3] <- 1

fhs2_offspring_ex09_omni1_ex04$physical_activity_fried[fhs2_offspring_ex09_omni1_ex04$SEX==1&
                                                         fhs2_offspring_ex09_omni1_ex04$fhs_pai >= 30.3] <- 0


fhs2_offspring_ex09_omni1_ex04$physical_activity_fried[fhs2_offspring_ex09_omni1_ex04$SEX==2&
                                                         fhs2_offspring_ex09_omni1_ex04$fhs_pai < 30.1] <- 1

fhs2_offspring_ex09_omni1_ex04$physical_activity_fried[fhs2_offspring_ex09_omni1_ex04$SEX==2&
                                                         fhs2_offspring_ex09_omni1_ex04$fhs_pai >= 30.1] <- 0


#### Exhaustion - Fried

fhs2_offspring_ex09_omni1_ex04$exhaustion_fried[fhs2_offspring_ex09_omni1_ex04$J733==3|
                                                  fhs2_offspring_ex09_omni1_ex04$J746==3] <- 1
fhs2_offspring_ex09_omni1_ex04$exhaustion_fried[fhs2_offspring_ex09_omni1_ex04$J733<3&
                                                  fhs2_offspring_ex09_omni1_ex04$J746<3] <- 0

#### Weight loss - Fried

fhs2_offspring_ex09_omni1_ex04$wgt_delta <-
  as.numeric(fhs2_offspring_ex09_omni1_ex04$WGT) - 
  as.numeric(fhs2_offspring_ex09_omni1_ex04$last_wgt)

fhs2_offspring_ex09_omni1_ex04$wtloss_fried[fhs2_offspring_ex09_omni1_ex04$wgt_delta <= -10] <- 1
fhs2_offspring_ex09_omni1_ex04$wtloss_fried[fhs2_offspring_ex09_omni1_ex04$wgt_delta > -10] <- 0


######### FRAIL Score

#### Fatigue

fhs2_offspring_ex09_omni1_ex04$fatigue_frail[fhs2_offspring_ex09_omni1_ex04$J900 <=3] <- 0
fhs2_offspring_ex09_omni1_ex04$fatigue_frail[fhs2_offspring_ex09_omni1_ex04$J900 > 3] <- 1


#### Resistance

fhs2_offspring_ex09_omni1_ex04$resistance_frail[fhs2_offspring_ex09_omni1_ex04$J611==1] <- 0
fhs2_offspring_ex09_omni1_ex04$resistance_frail[fhs2_offspring_ex09_omni1_ex04$J611==0] <- 1

#### Ambulation

fhs2_offspring_ex09_omni1_ex04$ambulate_frail[fhs2_offspring_ex09_omni1_ex04$J610==0] <- 1
fhs2_offspring_ex09_omni1_ex04$ambulate_frail[fhs2_offspring_ex09_omni1_ex04$J610==1] <- 0


# FRAIL Illness score counts presence of HTN, DM, cancer, chronic lung disease, MI, CHD, angina, asthma, arthritis, stroke, and ckd
# Scores for 0-4 conditions, score = 0, for ≥ 5, score=1


# COPD (chronic lung disease)

fhs2_offspring_ex09_omni1_ex04$copd_frail[fhs2_offspring_ex09_omni1_ex04$J419 %in% c(0,2)] <- 0
fhs2_offspring_ex09_omni1_ex04$copd_frail[fhs2_offspring_ex09_omni1_ex04$J419==1] <- 1

# asthma

fhs2_offspring_ex09_omni1_ex04$asthma_frail[fhs2_offspring_ex09_omni1_ex04$J421 %in% c(0,2)] <- 0
fhs2_offspring_ex09_omni1_ex04$asthma_frail[fhs2_offspring_ex09_omni1_ex04$J421==1] <- 1

# DJD (arthritis)

fhs2_offspring_ex09_omni1_ex04$djd_frail[fhs2_offspring_ex09_omni1_ex04$J426 %in% c(0,2)] <- 0
fhs2_offspring_ex09_omni1_ex04$djd_frail[fhs2_offspring_ex09_omni1_ex04$J426==1] <- 1

# CKD (renal disease)

fhs2_offspring_ex09_omni1_ex04$RACE[fhs2_offspring_ex09_omni1_ex04$IDTYPE==1] <- 1
fhs2_offspring_ex09_omni1_ex04$RACE[fhs2_offspring_ex09_omni1_ex04$IDTYPE==7] <- 2

fhs2_offspring_ex09_omni1_ex04$crcl <- 
  calc_MDRD4(
    dat=as.data.frame(fhs2_offspring_ex09_omni1_ex04),
    cr="CREAT")

fhs2_offspring_ex09_omni1_ex04$ckd_frail[fhs2_offspring_ex09_omni1_ex04$J414 %in% c(0,2)] <- 0
fhs2_offspring_ex09_omni1_ex04$ckd_frail[fhs2_offspring_ex09_omni1_ex04$J414==1] <- 1

# CHF/MI


fhs2_offspring_ex09_omni1_ex04$chf_frail <- 0
fhs2_offspring_ex09_omni1_ex04$chf_frail[fhs2_offspring_ex09_omni1_ex04$hfhosp_status==1&
                                           fhs2_offspring_ex09_omni1_ex04$hfhosp_dt <= fhs2_offspring_ex09_omni1_ex04$DATE] <- 1

fhs2_offspring_ex09_omni1_ex04$chd_frail <- 0
fhs2_offspring_ex09_omni1_ex04$chd_frail[fhs2_offspring_ex09_omni1_ex04$cadhosp_status==1&
                                           fhs2_offspring_ex09_omni1_ex04$cadhosp_dt <= fhs2_offspring_ex09_omni1_ex04$DATE] <- 1

# Stroke

fhs2_offspring_ex09_omni1_ex04$stroke_frail <- 0
fhs2_offspring_ex09_omni1_ex04$stroke_frail[fhs2_offspring_ex09_omni1_ex04$STROKE==1&fhs2_offspring_ex09_omni1_ex04$STROKEDATE <= fhs2_offspring_ex09_omni1_ex04$DATE] <- 1

# Cancer

fhs2_offspring_ex09_omni1_ex04$cancer_frail <- 0
fhs2_offspring_ex09_omni1_ex04$cancer_frail[fhs2_offspring_ex09_omni1_ex04$D_DATE <= fhs2_offspring_ex09_omni1_ex04$DATE] <- 1


#### Calculate FRAIL illness score
fhs2_offspring_ex09_omni1_ex04$conditions_frail <- 
  as.numeric(fhs2_offspring_ex09_omni1_ex04$HRX)+
  as.numeric(fhs2_offspring_ex09_omni1_ex04$DMRX)+
  fhs2_offspring_ex09_omni1_ex04$copd_frail+
  fhs2_offspring_ex09_omni1_ex04$asthma_frail+
  fhs2_offspring_ex09_omni1_ex04$djd_frail+
  fhs2_offspring_ex09_omni1_ex04$ckd_frail+
  fhs2_offspring_ex09_omni1_ex04$chf_frail+
  fhs2_offspring_ex09_omni1_ex04$chd_frail+
  fhs2_offspring_ex09_omni1_ex04$stroke_frail+
  fhs2_offspring_ex09_omni1_ex04$cancer_frail

fhs2_offspring_ex09_omni1_ex04$illness_frail[fhs2_offspring_ex09_omni1_ex04$conditions_frail <= 4] <- 0
fhs2_offspring_ex09_omni1_ex04$illness_frail[fhs2_offspring_ex09_omni1_ex04$conditions_frail > 4] <- 1

#### (Weight) Loss

fhs2_offspring_ex09_omni1_ex04$wtloss_frail[fhs2_offspring_ex09_omni1_ex04$wgt_delta <= -10] <- 1
fhs2_offspring_ex09_omni1_ex04$wtloss_frail[fhs2_offspring_ex09_omni1_ex04$wgt_delta > -10] <- 0


######### Epworth Sleepiness scale. 

fhs2_offspring_ex09_omni1_ex04$epworth_ss_nas <- 
  is.na(fhs2_offspring_ex09_omni1_ex04$J904)+
  is.na(fhs2_offspring_ex09_omni1_ex04$J905)+
  is.na(fhs2_offspring_ex09_omni1_ex04$J906)+
  is.na(fhs2_offspring_ex09_omni1_ex04$J907)+
  is.na(fhs2_offspring_ex09_omni1_ex04$J908)+
  is.na(fhs2_offspring_ex09_omni1_ex04$J909)+
  is.na(fhs2_offspring_ex09_omni1_ex04$J910)+
  is.na(fhs2_offspring_ex09_omni1_ex04$J911)

fhs2_offspring_ex09_omni1_ex04$epworth_ss <- 
  fhs2_offspring_ex09_omni1_ex04$J904+
  fhs2_offspring_ex09_omni1_ex04$J905+
  fhs2_offspring_ex09_omni1_ex04$J906+
  fhs2_offspring_ex09_omni1_ex04$J907+
  fhs2_offspring_ex09_omni1_ex04$J908+
  fhs2_offspring_ex09_omni1_ex04$J909+
  fhs2_offspring_ex09_omni1_ex04$J910+
  fhs2_offspring_ex09_omni1_ex04$J911

## DENORMALIZE

fhs2_melt_offspring_ex09_omni1_ex04 <- melt(fhs2_offspring_ex09_omni1_ex04,id.vars=c("PID","IDTYPE","visit"), na.rm=T, factorsAsStrings=T)

fhs2_melt_offspring_ex09_omni1_ex04$form <- "fhs2_offspring_ex09_omni1_ex04"

rm(fhs2_offspring_ex09_omni1_ex04)


#======================================================================#
####  Actigraphy, Framingham Offspring Exam 9 - Omni 1, Exam 4      ---#
#======================================================================#

act_fields <-
  c("zmin_vdt",
    "zmin_vdtd",
    "zmin_vdtm",
    "zmin_vdtwd",
    "zmin_vdtwdd",
    "zmin_vdtwdm",
    "zmin_vdtwe",
    "zmin_vdtwed",
    "zmin_vdtwedm",
    "wstep_vdt",
    "wstep_vdtd",
    "wstep_vdtm",
    "wstep_vdtwd",
    "wstep_vdtwdd",
    "wstep_vdtwdm",
    "wstep_vdtwe",
    "wstep_vdtwed",
    "wstep_vdtwedm",
    "zb_vdt",
    "zb_vdtd",
    "zb_vdtmin",
    "zb_vdtwd",
    "zb_vdtwdmin",
    "zb_vdtwe",
    "zb_vdtwed",
    "zb_vdtwedd",
    "zb_vdtwemin",
    "vigstepvdt",
    "vigstepvdtd",
    "vigstepvdtwd",
    "vigstepvdtwdd",
    "vigstepvdtwe",
    "vigstepvdtwed",
    "vigminvdt",
    "vigminvdtd",
    "vigminvdtwd",
    "vigminvdtwdd",
    "vigminvdtwe",
    "vigminvdtwed",
    "vigcntvdt",
    "vigcntvdtd",
    "vigcntvdtwd",
    "vigcntvdtwdd",
    "vigcntvdtwe",
    "vigcntvdtwed",
    "sedcntvdt",
    "sedcntvdtd",
    "sedcntvdtwd",
    "sedcntvdtwdd",
    "sedcntvdtwe",
    "sedcntvdtwed",
    "sedminvdt",
    "sedminvdtd",
    "sedminvdtwd",
    "sedminvdtwdd",
    "sedminvdtwe",
    "sedminvdtwed",
    "sedminwed",
    "bgt_litcntvdt",
    "bgt_litcntvdtd",
    "bgt_litcntvdtwd",
    "bgt_litcntvdtwdd",
    "bgt_litcntvdtwe",
    "bgt_litcntvdtwed",
    "bgt_litminvdt",
    "bgt_litminvdtd",
    "bgt_litminvdtwd",
    "bgt_litminvdtwdd",
    "bgt_litminvdtwe",
    "bgt_litminvdtwed",
    "bgt_litstepvdt",
    "bgt_litstepvdtd",
    "bgt_litstepvdtwd",
    "bgt_litstepvdtwdd",
    "bgt_litstepvdtwe",
    "bgt_litstepvdtwed",
    "bgt_modcntvdt",
    "bgt_modcntvdtd",
    "bgt_modcntvdtwd",
    "bgt_modcntvdtwdd",
    "bgt_modcntvdtwe",
    "bgt_modcntvdtwed",
    "bgt_modminvdt",
    "bgt_modminvdtd",
    "bgt_modminvdtwd",
    "bgt_modminvdtwdd",
    "bgt_modminvdtwe",
    "bgt_modminvdtwed",
    "bgt_modstepvdt",
    "bgt_modstepvdtd",
    "bgt_modstepvdtwd",
    "bgt_modstepvdtwdd",
    "bgt_modstepvdtwe",
    "bgt_modstepvdtwed",
    "bgt_mvpcntvdt",
    "bgt_mvpcntvdtd",
    "bgt_mvpcntvdtwd",
    "bgt_mvpcntvdtwdd",
    "bgt_mvpcntvdtwe",
    "bgt_mvpcntvdtwed",
    "bgt_mvpminvdt",
    "bgt_mvpminvdtd",
    "bgt_mvpminvdtwd",
    "bgt_mvpminvdtwdd",
    "bgt_mvpminvdtwe",
    "bgt_mvpminvdtwed",
    "bgt_mvpstepvdt",
    "bgt_mvpstepvdtd",
    "bgt_mvpstepvdtwd",
    "bgt_mvpstepvdtwdd",
    "bgt_mvpstepvdtwe",
    "bgt_mvpstepvdtwed",
    "bgt_sedcntvdt",
    "bgt_sedcntvdtd",
    "bgt_sedcntvdtwd",
    "bgt_sedcntvdtwdd",
    "bgt_sedcntvdtwe",
    "bgt_sedcntvdtwed",
    "bgt_sedminvdt",
    "bgt_sedminvdtd",
    "bgt_sedminvdtwd",
    "bgt_sedminvdtwdd",
    "bgt_sedminvdtwe",
    "bgt_sedminvdtwed",
    "bgt_sedstepvdt",
    "bgt_sedstepvdtd",
    "bgt_sedstepvdtwd",
    "bgt_sedstepvdtwdd",
    "bgt_sedstepvdtwe",
    "bgt_sedstepvdtwed",
    "bgt_vigcntvdt",
    "bgt_vigcntvdtd",
    "bgt_vigcntvdtwd",
    "bgt_vigcntvdtwdd",
    "bgt_vigcntvdtwe",
    "bgt_vigcntvdtwed",
    "bgt_vigminvdt",
    "bgt_vigminvdtd",
    "bgt_vigminvdtwd",
    "bgt_vigminvdtwdd",
    "bgt_vigminvdtwe",
    "bgt_vigminvdtwed",
    "bgt_vigstepvdt",
    "bgt_vigstepvdtd",
    "bgt_vigstepvdtwd",
    "bgt_vigstepvdtwdd",
    "bgt_vigstepvdtwe",
    "bgt_vigstepvdtwed",
    "litcntvdt",
    "litcntvdtd",
    "litcntvdtwd",
    "litcntvdtwdd",
    "litcntvdtwe",
    "litcntvdtwed",
    "litminvdt",
    "litminvdtd",
    "litminvdtwd",
    "litminvdtwdd",
    "litminvdtwe",
    "litminvdtwed",
    "litstepvdt",
    "litstepvdtd",
    "litstepvdtwd",
    "litstepvdtwdd",
    "litstepvdtwe",
    "litstepvdtwed",
    "modcntvdt",
    "modcntvdtd",
    "modcntvdtwd",
    "modcntvdtwdd",
    "modcntvdtwe",
    "modcntvdtwed",
    "modminvdt",
    "modminvdtd",
    "modminvdtwd",
    "modminvdtwdd",
    "modminvdtwe",
    "modminvdtwed",
    "modstepvdt",
    "modstepvdtd",
    "modstepvdtwd",
    "modstepvdtwdd",
    "modstepvdtwe",
    "modstepvdtwed",
    "motcntvdt",
    "motcntvdtd",
    "motcntvdtwd",
    "motcntvdtwdd",
    "motcntvdtwe",
    "motcntvdtwed",
    "motminvdt",
    "motminvdtd",
    "motminvdtwd",
    "motminvdtwdd",
    "motminvdtwe",
    "motminvdtwed",
    "motstepvdt",
    "motstepvdtd",
    "motstepvdtwd",
    "motstepvdtwdd",
    "motstepvdtwe",
    "motstepvdtwed",
    "mvpcntvdt",
    "mvpcntvdtd",
    "mvpcntvdtwd",
    "mvpcntvdtwdd",
    "mvpcntvdtwe",
    "mvpcntvdtwed",
    "mvpminvdt",
    "mvpminvdtd",
    "mvpminvdtwd",
    "mvpminvdtwdd",
    "mvpminvdtwe",
    "mvpminvdtwed",
    "mvpstepvdt",
    "mvpstepvdtd",
    "mvpstepvdtwd",
    "mvpstepvdtwdd",
    "mvpstepvdtwe",
    "mvpstepvdtwed",
    "s_litcntvdt",
    "s_litcntvdtd",
    "s_litcntvdtwd",
    "s_litcntvdtwdd",
    "s_litcntvdtwe",
    "s_litcntvdtwed",
    "s_litminvdt",
    "s_litminvdtd",
    "s_litminvdtwd",
    "s_litminvdtwdd",
    "s_litminvdtwe",
    "s_litminvdtwed",
    "s_litstepvdt",
    "s_litstepvdtd",
    "s_litstepvdtwd",
    "s_litstepvdtwdd",
    "s_litstepvdtwe",
    "s_litstepvdtwed",
    "s_modcntvdt",
    "s_modcntvdtd",
    "s_modcntvdtwd",
    "s_modcntvdtwdd",
    "s_modcntvdtwe",
    "s_modcntvdtwed",
    "s_modminvdt",
    "s_modminvdtd",
    "s_modminvdtwd",
    "s_modminvdtwdd",
    "s_modminvdtwe",
    "s_modminvdtwed",
    "s_modstepvdt",
    "s_modstepvdtd",
    "s_modstepvdtwd",
    "s_modstepvdtwdd",
    "s_modstepvdtwe",
    "s_modstepvdtwed",
    "s_mvpcntvdt",
    "s_mvpcntvdtd",
    "s_mvpcntvdtwd",
    "s_mvpcntvdtwdd",
    "s_mvpcntvdtwe",
    "s_mvpcntvdtwed",
    "s_mvpminvdt",
    "s_mvpminvdtd",
    "s_mvpminvdtwd",
    "s_mvpminvdtwdd",
    "s_mvpminvdtwe",
    "s_mvpminvdtwed",
    "s_mvpstepvdt",
    "s_mvpstepvdtd",
    "s_mvpstepvdtwd",
    "s_mvpstepvdtwdd",
    "s_mvpstepvdtwe",
    "s_mvpstepvdtwed",
    "s_sedcntvdt",
    "s_sedcntvdtd",
    "s_sedcntvdtwd",
    "s_sedcntvdtwdd",
    "s_sedcntvdtwe",
    "s_sedcntvdtwed",
    "s_sedminvdt",
    "s_sedminvdtd",
    "s_sedminvdtwd",
    "s_sedminvdtwdd",
    "s_sedminvdtwe",
    "s_sedminvdtwed",
    "s_sedstepvdt",
    "s_sedstepvdtd",
    "s_sedstepvdtwd",
    "s_sedstepvdtwdd",
    "s_sedstepvdtwe",
    "s_sedstepvdtwed",
    "s_vigcntvdt",
    "s_vigcntvdtd",
    "s_vigcntvdtwd",
    "s_vigcntvdtwdd",
    "s_vigcntvdtwe",
    "s_vigcntvdtwed",
    "s_vigminvdt",
    "s_vigminvdtd",
    "s_vigminvdtwd",
    "s_vigminvdtwdd",
    "s_vigminvdtwe",
    "s_vigminvdtwed",
    "s_vigstepvdt",
    "s_vigstepvdtd",
    "s_vigstepvdtwd",
    "s_vigstepvdtwdd",
    "s_vigstepvdtwe",
    "s_vigstepvdtwed",
    "zstep_vdt",
    "zstep_vdtd",
    "zstep_vdtm",
    "zstep_vdtwd",
    "zstep_vdtwdd",
    "zstep_vdtwdm",
    "zstep_vdtwe",
    "zstep_vdtwed",
    "zstep_vdtwedm",
    "zcnt_vdt",
    "zcnt_vdtd",
    "zcnt_vdtm",
    "zcnt_vdtwd",
    "zcnt_vdtwdd",
    "zcnt_vdtwdm",
    "zcnt_vdtwe",
    "zcnt_vdtwed",
    "zcnt_vdtwedm",
    "zb_vdt",
    "zb_vdtd",
    "zb_vdtmin",
    "zb_vdtwd",
    "zb_vdtwdmin",
    "zb_vdtwe",
    "zb_vdtwed",
    "zb_vdtwedd",
    "zb_vdtwemin",
    "wb_vdt",
    "wb_vdtd",
    "wb_vdtmin",
    "wb_vdtwd",
    "wb_vdtwdmin",
    "wb_vdtwe",
    "wb_vdtwed",
    "wb_vdtwedd",
    "wb_vdtwemin",
    "wcnt_vdt",
    "wcnt_vdtd",
    "wcnt_vdtm",
    "wcnt_vdtwd",
    "wcnt_vdtwdd",
    "wcnt_vdtwdm",
    "wcnt_vdtwe",
    "wcnt_vdtwed",
    "wcnt_vdtwedm",
    "wmin_vdt",
    "wmin_vdtd",
    "wmin_vdtm",
    "wmin_vdtwd",
    "wmin_vdtwdd",
    "wmin_vdtwdm",
    "wmin_vdtwe",
    "wmin_vdtwed",
    "wmin_vdtwedm",
    "wstep_vdt",
    "wstep_vdtd",
    "wstep_vdtm",
    "wstep_vdtwd",
    "wstep_vdtwdd",
    "wstep_vdtwdm",
    "wstep_vdtwe",
    "wstep_vdtwed",
    "wstep_vdtwedm")

act_fields <- toupper(act_fields)

fhs2_actigraphy_offspring_ex09_omni1_ex04 <- 
  as.data.table(
    read_sas('~/Dropbox/BioLINCC files/Framingham Offspring/datasets/t_physactf_ex09_1b_0833d.sas7bdat'))


names(fhs2_actigraphy_offspring_ex09_omni1_ex04) <- toupper(names(fhs2_actigraphy_offspring_ex09_omni1_ex04))

fhs2_melt_actigraphy_offspring_ex09_omni1_ex04 <-  
  melt(fhs2_actigraphy_offspring_ex09_omni1_ex04,id.vars=c("PID","IDTYPE"),na.rm=T,factorsAsStrings = T)

fhs2_melt_actigraphy_offspring_ex09_omni1_ex04$visit[fhs2_melt_actigraphy_offspring_ex09_omni1_ex04$IDTYPE==1] <- 9
fhs2_melt_actigraphy_offspring_ex09_omni1_ex04$visit[fhs2_melt_actigraphy_offspring_ex09_omni1_ex04$IDTYPE==7] <- 4

fhs2_melt_actigraphy_offspring_ex09_omni1_ex04$form <- "fhs2_actigraphy_ex09_ex04"

fhs2_melt_actigraphy_offspring_ex09_omni1_ex04 <- 
  subset(fhs2_melt_actigraphy_offspring_ex09_omni1_ex04,variable %in% act_fields)

rm(fhs2_actigraphy_offspring_ex09_omni1_ex04)


#=========================================================#
####  Validated food frequency data, exam 9            ---#
#=========================================================#

fhs2_vr_ffreq_offspring_ex09_omni1_ex04 <- fread('~/Dropbox/BioLINCC files/Framingham offspring/datasets/CSV/vr_ffreq_ex09_1b_1636d_v1.csv',na.strings=c("NA","","NULL"))
fhs2_vr_ffreq_offspring_ex09_omni1_ex04$visit[fhs2_vr_ffreq_offspring_ex09_omni1_ex04$IDTYPE==1] <- 9
fhs2_vr_ffreq_offspring_ex09_omni1_ex04$visit[fhs2_vr_ffreq_offspring_ex09_omni1_ex04$IDTYPE==7] <- 4

fhs2_melt_vr_ffreq_offspring_ex09_omni1_ex04 <- melt(fhs2_vr_ffreq_offspring_ex09_omni1_ex04,
                                                     id.vars=c("PID","IDTYPE","visit"),
                                                     na.rm=T, 
                                                     factorsAsStrings=T)
fhs2_melt_vr_ffreq_offspring_ex09_omni1_ex04$form <- "fhs2_vr_ffreq"

rm(fhs2_vr_ffreq_offspring_ex09_omni1_ex04)


#==============================#
####  Lab panel, Exam 9     ---#
#==============================#

fhs2_l_fhslab_ex09 <- fread('~/Dropbox/BioLINCC files/Framingham offspring/datasets/CSV/l_fhslab_ex09_1b_0658d.csv',na.strings=c("NA","","NULL"))
names(fhs2_l_fhslab_ex09)[names(fhs2_l_fhslab_ex09)=="idtype"] <- "IDTYPE"
fhs2_l_fhslab_ex09$visit[fhs2_l_fhslab_ex09$IDTYPE==1] <- 9
fhs2_l_fhslab_ex09$visit[fhs2_l_fhslab_ex09$IDTYPE==7] <- 4
fhs2_melt_l_fhslab_ex09 <- melt(fhs2_l_fhslab_ex09,id.vars=c("PID","IDTYPE","visit"), na.rm=T, factorsAsStrings=T)
fhs2_melt_l_fhslab_ex09$form <- "fhs2_fhslab_ex09"

rm(fhs2_l_fhslab_ex09)


#================================================#
####  Menses                                  ---#
#================================================#

fhs2_menarche <- fread('~/Dropbox/BioLINCC files/Framingham offspring/datasets/CSV/menarche1_7d.csv',na.strings=c("NA","","NULL"))

names(fhs2_menarche) <- toupper(names(fhs2_menarche))

fhs2_offspring_menopause <- fread('~/Dropbox/BioLINCC files/Framingham offspring/datasets/CSV/meno1_8d.csv',
                                  na.strings=c("NA","","NULL"))


fhs2_offspring_menopause$num_ovaries_removed <-
  apply(fhs2_offspring_menopause[,3:9],
        MARGIN=1,
        function(x) max(x, na.rm=T))

fhs2_offspring_menopause$num_ovaries_removed[fhs2_offspring_menopause$num_ovaries_removed=="-Inf"] <- NA

fhs2_omni1_menopause <- fread('~/Dropbox/BioLINCC files/Framingham offspring/datasets/CSV/vr_meno_ex03_7_0916d.csv',
                              na.strings=c("NA","","NULL"))

fhs2_omni1_menopause$num_ovaries_removed <-
  apply(fhs2_omni1_menopause[,11:13],
        MARGIN=1,
        function(x) max(x, na.rm=T))

fhs2_omni1_menopause$num_ovaries_removed[fhs2_omni1_menopause$num_ovaries_removed=="-Inf"] <- NA

fhs2_offspring_menopause$STOPAGE <- fhs2_offspring_menopause$STOP_AGE

fhs2_menopause <- 
  rbind(fhs2_offspring_menopause[,c("PID","IDTYPE","STOPAGE","CAUSE","num_ovaries_removed")],
        fhs2_omni1_menopause[,c("PID","IDTYPE","STOPAGE","CAUSE","num_ovaries_removed")])

fhs2_menopause$STOPAGE[fhs2_menopause$STOPAGE==0] <- NA

fhs2_menses <- merge(fhs2_menarche,
                     fhs2_menopause,
                     by=c("PID","IDTYPE"),
                     all=T)

fhs2_menses$years_menses <- fhs2_menses$STOPAGE-fhs2_menses$IVQ1AGE

fhs2_melt_menses <- melt(fhs2_menses,id.vars=c("PID","IDTYPE"), na.rm=T, factorsAsStrings=T)

fhs2_melt_menses$form <- "fhs2_menses"
fhs2_melt_menses$visit <- 8

rm(fhs2_menses)

#===============================================#
#### *** Offspring exam 10/Omni 1 exam 5 *** ####
#===============================================#

fhs2_offspring_ex10_omni1_ex05 <- 
  fread('~/Dropbox/BioLINCC files/Framingham offspring/datasets/CSV/e_exam_ex10_1b_1409d.csv',
        na.strings=c("NA","","NULL")) # Omni1 5  Offspring Exam 10

names(fhs2_offspring_ex10_omni1_ex05) <- 
  toupper(names(fhs2_offspring_ex10_omni1_ex05))

fhs2_offspring_ex10_omni1_ex05 <-
  merge(fhs2_offspring_ex10_omni1_ex05,
        fhs_dates[fhs_dates$IDTYPE==1,c("PID","DATE10")],
        by="PID",
        all.x=T)

fhs2_offspring_ex10_omni1_ex05 <-
  merge(fhs2_offspring_ex10_omni1_ex05,
        fhs_dates[fhs_dates$IDTYPE==7,c("PID","DATE5")],
        by="PID",
        all.x=T)

fhs2_offspring_ex10_omni1_ex05$DATE[
  fhs2_offspring_ex10_omni1_ex05$IDTYPE==7] <-
  fhs2_offspring_ex10_omni1_ex05$DATE5[
    fhs2_offspring_ex10_omni1_ex05$IDTYPE==7]

fhs2_offspring_ex10_omni1_ex05$DATE[
  fhs2_offspring_ex10_omni1_ex05$IDTYPE==1] <-
  fhs2_offspring_ex10_omni1_ex05$DATE10[
    fhs2_offspring_ex10_omni1_ex05$IDTYPE==1]


fhs2_offspring_ex10_omni1_ex05$CESD20_missing <- 
  rowSums(is.na(fhs2_offspring_ex10_omni1_ex05[,268:287]))
fhs2_offspring_ex10_omni1_ex05$CESD20 <- 
  rowSums(fhs2_offspring_ex10_omni1_ex05[,268:287])

fhs2_offspring_ex10_omni1_ex05$CESD20[
  fhs2_offspring_ex10_omni1_ex05$CESD20_missing >= 4] <- NA

fhs2_offspring_ex10_omni1_ex05$visit[fhs2_offspring_ex10_omni1_ex05$IDTYPE==1] <- 10
fhs2_offspring_ex10_omni1_ex05$visit[fhs2_offspring_ex10_omni1_ex05$IDTYPE==7] <- 5

fhs2_offspring_ex10_omni1_ex05$drinks_wk <- fhs2_offspring_ex10_omni1_ex05$K0321+
  fhs2_offspring_ex10_omni1_ex05$K0325+
  fhs2_offspring_ex10_omni1_ex05$K0329

fhs2_offspring_ex10_omni1_ex05$K0295[
  !between(fhs2_offspring_ex10_omni1_ex05$K0295,37,80)] <- NA


fhs2_offspring_ex10_omni1_ex05 <- 
  merge(fhs2_offspring_ex10_omni1_ex05,
        fhs2_wkthru[fhs2_wkthru$visit==1,
                    c("PID",
                      "IDTYPE",
                      "SEX",
                      "HGT")],
        by=c("PID","IDTYPE"),
        all.x=T)

fhs2_omni1_ex05_wgt4 <- 
  fhs2_wkthru[fhs2_wkthru$visit==4&
                fhs2_wkthru$IDTYPE==7,
              c("PID",
                "IDTYPE",
                "WGT")]

fhs2_offspring_ex10_wgt9 <- 
  fhs2_wkthru[fhs2_wkthru$visit==9&
                fhs2_wkthru$IDTYPE==1,
              c("PID",
                "IDTYPE",
                "WGT")]

fhs2_offspring_ex10_omni1_ex05_lastwgt <- 
  rbind(fhs2_omni1_ex05_wgt4,
        fhs2_offspring_ex10_wgt9)

names(fhs2_offspring_ex10_omni1_ex05_lastwgt)[3] <- "last_wgt"

fhs2_offspring_ex10_omni1_ex05 <- 
  merge(fhs2_offspring_ex10_omni1_ex05,
        fhs2_offspring_ex10_omni1_ex05_lastwgt,
        by=c("PID","IDTYPE"),
        all.x=T)

fhs2_offspring_ex10_bmi <- 
  fhs2_wkthru[fhs2_wkthru$visit==10&
                fhs2_wkthru$IDTYPE==1&
                !is.na(fhs2_wkthru$BMI),
              c("PID",
                "IDTYPE",
                "BMI",
                "CREAT",
                "DMRX",
                "HRX",
                "AGE")]

fhs2_omni1_ex05_bmi <- 
  fhs2_wkthru[fhs2_wkthru$visit==5&
                fhs2_wkthru$IDTYPE==7&
                !is.na(fhs2_wkthru$BMI),
              c("PID",
                "IDTYPE",
                "BMI",
                "CREAT",
                "DMRX",
                "HRX",
                "AGE")]

fhs2_off10_omni5_bmi <- 
  rbind(fhs2_offspring_ex10_bmi,
        fhs2_omni1_ex05_bmi)

fhs2_offspring_ex10_omni1_ex05 <- 
  merge(fhs2_offspring_ex10_omni1_ex05,
        fhs2_off10_omni5_bmi,
        by=c("PID","IDTYPE"),
        all.x=T)


fhs2_offspring_ex10_omni1_ex05 <- 
  merge(fhs2_offspring_ex10_omni1_ex05,
        fhs_outcomes,
        by=c("PID",
             "IDTYPE"))


fhs2_offspring_ex10_omni1_ex05 <- 
  merge(fhs2_offspring_ex10_omni1_ex05,
        fhs_survca_first_any[,c("PID",
                                "IDTYPE",
                                "D_DATE")],
        by=c("PID",
             "IDTYPE"),
        all.x=T)


## Walking speed - Fried

# Uses greater of 2 speeds (PMID 26695510)

fhs2_offspring_ex10_omni1_ex05$walktime_1 <- fhs2_offspring_ex10_omni1_ex05$K1593
fhs2_offspring_ex10_omni1_ex05$walktime_2 <- fhs2_offspring_ex10_omni1_ex05$K1595

fhs2_offspring_ex10_omni1_ex05$walktime_avg <- 
  round(apply(fhs2_offspring_ex10_omni1_ex05[,c("walktime_1","walktime_2")],
              MARGIN=1,
              mean,
              na.rm=T),3)

fhs2_offspring_ex10_omni1_ex05$walktime_avg[
  fhs2_offspring_ex10_omni1_ex05$walktime_avg=="NaN"] <- NA

fhs2_offspring_ex10_omni1_ex05$walktime_min <-
  round(apply(fhs2_offspring_ex10_omni1_ex05[,c("walktime_1","walktime_2")],
              MARGIN=1,min),3)

fhs2_offspring_ex10_omni1_ex05$walktime_min[
  fhs2_offspring_ex10_omni1_ex05$walktime_min=="-Inf"] <- NA

fhs2_offspring_ex10_omni1_ex05$walktime_min[
  !is.na(fhs2_offspring_ex10_omni1_ex05$walktime_1)|
    !is.na(fhs2_offspring_ex10_omni1_ex05$walktime_2)] <- 0 


########### Grip (kg)

# Produces highest average by hand (assumes dominant hand is stronger)

fhs2_offspring_ex10_omni1_ex05$grip_avg_right <- 
  round(apply(fhs2_offspring_ex10_omni1_ex05[,260:262],MARGIN=1,mean),1)

fhs2_offspring_ex10_omni1_ex05$grip_avg_left <- 
  round(apply(fhs2_offspring_ex10_omni1_ex05[,263:265],MARGIN=1,mean),1)

fhs2_offspring_ex10_omni1_ex05$grip_max <- 
  round(apply(fhs2_offspring_ex10_omni1_ex05[,c("grip_avg_right","grip_avg_left")],MARGIN=1,max),1)


#### Physical activity index - Fried

fhs2_offspring_ex10_omni1_ex05$fhs_pai <- 
  calc_framingham_pai(
    as.data.frame(fhs2_offspring_ex10_omni1_ex05),
    slp_hrs="K1234",
    sed_hrs="K1235",
    slgt_hrs="K1236",
    mod_hrs="K1237",
    hvy_hrs="K1238")

fhs2_offspring_ex10_omni1_ex05$physical_activity_fried[
  fhs2_offspring_ex10_omni1_ex05$SEX==1&
    fhs2_offspring_ex10_omni1_ex05$fhs_pai < 30.3] <- 1

fhs2_offspring_ex10_omni1_ex05$physical_activity_fried[
  fhs2_offspring_ex10_omni1_ex05$SEX==1&
    fhs2_offspring_ex10_omni1_ex05$fhs_pai >= 30.3] <- 0


fhs2_offspring_ex10_omni1_ex05$physical_activity_fried[
  fhs2_offspring_ex10_omni1_ex05$SEX==2&
    fhs2_offspring_ex10_omni1_ex05$fhs_pai < 30.1] <- 1

fhs2_offspring_ex10_omni1_ex05$physical_activity_fried[
  fhs2_offspring_ex10_omni1_ex05$SEX==2&
    fhs2_offspring_ex10_omni1_ex05$fhs_pai >= 30.1] <- 0


#### Exhaustion - Fried

fhs2_offspring_ex10_omni1_ex05$exhaustion_fried[
  fhs2_offspring_ex10_omni1_ex05$K1203==3|
    fhs2_offspring_ex10_omni1_ex05$K1216==3] <- 1
fhs2_offspring_ex10_omni1_ex05$exhaustion_fried[
  fhs2_offspring_ex10_omni1_ex05$K1203<3&
    fhs2_offspring_ex10_omni1_ex05$K1216<3] <- 0

#### Weight loss - Fried

fhs2_offspring_ex10_omni1_ex05$wgt_delta <-
  as.numeric(fhs2_offspring_ex10_omni1_ex05$WGT) - 
  as.numeric(fhs2_offspring_ex10_omni1_ex05$last_wgt)

fhs2_offspring_ex10_omni1_ex05$wtloss_fried[
  fhs2_offspring_ex10_omni1_ex05$wgt_delta <= -10] <- 1

fhs2_offspring_ex10_omni1_ex05$wtloss_fried[
  fhs2_offspring_ex10_omni1_ex05$wgt_delta > -10] <- 0


######### FRAIL Score

#### Fatigue

fhs2_offspring_ex10_omni1_ex05$fatigue_frail[
  fhs2_offspring_ex10_omni1_ex05$K1739 <=3] <- 0

fhs2_offspring_ex10_omni1_ex05$fatigue_frail[
  fhs2_offspring_ex10_omni1_ex05$K1739 > 3] <- 1


#### Resistance

fhs2_offspring_ex10_omni1_ex05$resistance_frail[fhs2_offspring_ex10_omni1_ex05$K1224==1] <- 0
fhs2_offspring_ex10_omni1_ex05$resistance_frail[fhs2_offspring_ex10_omni1_ex05$K1224==0] <- 1

#### Ambulation

fhs2_offspring_ex10_omni1_ex05$ambulate_frail[fhs2_offspring_ex10_omni1_ex05$K1222==0] <- 1
fhs2_offspring_ex10_omni1_ex05$ambulate_frail[fhs2_offspring_ex10_omni1_ex05$K1222==1] <- 0


# FRAIL Illness score counts presence of HTN, DM, cancer, chronic lung disease, MI, CHD, angina, asthma, arthritis, stroke, and ckd
# Scores for 0-4 conditions, score = 0, for ≥ 5, score=1


# COPD (chronic lung disease)

fhs2_offspring_ex10_omni1_ex05$copd_frail[fhs2_offspring_ex10_omni1_ex05$K0995 %in% c(0,2)] <- 0
fhs2_offspring_ex10_omni1_ex05$copd_frail[fhs2_offspring_ex10_omni1_ex05$K0995==1] <- 1

# asthma

fhs2_offspring_ex10_omni1_ex05$asthma_frail[fhs2_offspring_ex10_omni1_ex05$K0997 %in% c(0,2)] <- 0
fhs2_offspring_ex10_omni1_ex05$asthma_frail[fhs2_offspring_ex10_omni1_ex05$K0997==1] <- 1

# DJD (arthritis)

fhs2_offspring_ex10_omni1_ex05$djd_frail[fhs2_offspring_ex10_omni1_ex05$K1003 %in% c(0,2)] <- 0
fhs2_offspring_ex10_omni1_ex05$djd_frail[fhs2_offspring_ex10_omni1_ex05$K1003==1] <- 1

# CKD (renal disease)

fhs2_offspring_ex10_omni1_ex05$RACE[fhs2_offspring_ex10_omni1_ex05$IDTYPE==1] <- 1
fhs2_offspring_ex10_omni1_ex05$RACE[fhs2_offspring_ex10_omni1_ex05$IDTYPE==7] <- 2

fhs2_offspring_ex10_omni1_ex05$crcl <- 
  calc_MDRD4(
    dat=as.data.frame(fhs2_offspring_ex10_omni1_ex05),
    cr="CREAT")

fhs2_offspring_ex10_omni1_ex05$ckd_frail[fhs2_offspring_ex10_omni1_ex05$K0990 %in% c(0,2)] <- 0
fhs2_offspring_ex10_omni1_ex05$ckd_frail[fhs2_offspring_ex10_omni1_ex05$K0990==1] <- 1

# CHF/MI


fhs2_offspring_ex10_omni1_ex05$chf_frail <- 0
fhs2_offspring_ex10_omni1_ex05$chf_frail[fhs2_offspring_ex10_omni1_ex05$hfhosp_status==1&
                                           fhs2_offspring_ex10_omni1_ex05$hfhosp_dt <= fhs2_offspring_ex10_omni1_ex05$DATE] <- 1

fhs2_offspring_ex10_omni1_ex05$chd_frail <- 0
fhs2_offspring_ex10_omni1_ex05$chd_frail[fhs2_offspring_ex10_omni1_ex05$cadhosp_status==1&
                                           fhs2_offspring_ex10_omni1_ex05$cadhosp_dt <= fhs2_offspring_ex10_omni1_ex05$DATE] <- 1

# Stroke

fhs2_offspring_ex10_omni1_ex05$stroke_frail <- 0
fhs2_offspring_ex10_omni1_ex05$stroke_frail[
  fhs2_offspring_ex10_omni1_ex05$cvahosp_status==1&
    fhs2_offspring_ex10_omni1_ex05$cvahosp_dt <= fhs2_offspring_ex10_omni1_ex05$DATE] <- 1

# Cancer

fhs2_offspring_ex10_omni1_ex05$cancer_frail <- 0
fhs2_offspring_ex10_omni1_ex05$cancer_frail[fhs2_offspring_ex10_omni1_ex05$D_DATE <= fhs2_offspring_ex10_omni1_ex05$DATE] <- 1


#### Calculate FRAIL illness score
fhs2_offspring_ex10_omni1_ex05$conditions_frail <- 
  as.numeric(fhs2_offspring_ex10_omni1_ex05$HRX)+
  as.numeric(fhs2_offspring_ex10_omni1_ex05$DMRX)+
  fhs2_offspring_ex10_omni1_ex05$copd_frail+
  fhs2_offspring_ex10_omni1_ex05$asthma_frail+
  fhs2_offspring_ex10_omni1_ex05$djd_frail+
  fhs2_offspring_ex10_omni1_ex05$ckd_frail+
  fhs2_offspring_ex10_omni1_ex05$chf_frail+
  fhs2_offspring_ex10_omni1_ex05$chd_frail+
  fhs2_offspring_ex10_omni1_ex05$stroke_frail+
  fhs2_offspring_ex10_omni1_ex05$cancer_frail

fhs2_offspring_ex10_omni1_ex05$illness_frail[fhs2_offspring_ex10_omni1_ex05$conditions_frail <= 4] <- 0
fhs2_offspring_ex10_omni1_ex05$illness_frail[fhs2_offspring_ex10_omni1_ex05$conditions_frail > 4] <- 1

#### (Weight) Loss

fhs2_offspring_ex10_omni1_ex05$wtloss_frail[fhs2_offspring_ex10_omni1_ex05$wgt_delta <= -10] <- 1
fhs2_offspring_ex10_omni1_ex05$wtloss_frail[fhs2_offspring_ex10_omni1_ex05$wgt_delta > -10] <- 0


######### Epworth Sleepiness scale. 

fhs2_offspring_ex10_omni1_ex05$epworth_ss_nas <- 
  is.na(fhs2_offspring_ex10_omni1_ex05$J904)+
  is.na(fhs2_offspring_ex10_omni1_ex05$J905)+
  is.na(fhs2_offspring_ex10_omni1_ex05$J906)+
  is.na(fhs2_offspring_ex10_omni1_ex05$J907)+
  is.na(fhs2_offspring_ex10_omni1_ex05$J908)+
  is.na(fhs2_offspring_ex10_omni1_ex05$J909)+
  is.na(fhs2_offspring_ex10_omni1_ex05$J910)+
  is.na(fhs2_offspring_ex10_omni1_ex05$J911)

fhs2_offspring_ex10_omni1_ex05$epworth_ss <- 
  fhs2_offspring_ex10_omni1_ex05$J904+
  fhs2_offspring_ex10_omni1_ex05$J905+
  fhs2_offspring_ex10_omni1_ex05$J906+
  fhs2_offspring_ex10_omni1_ex05$J907+
  fhs2_offspring_ex10_omni1_ex05$J908+
  fhs2_offspring_ex10_omni1_ex05$J909+
  fhs2_offspring_ex10_omni1_ex05$J910+
  fhs2_offspring_ex10_omni1_ex05$J911

## DENORMALIZE

fhs2_melt_offspring_ex10_omni1_ex05 <- 
  melt(fhs2_offspring_ex10_omni1_ex05,
       id.vars=c("PID","IDTYPE","visit"), 
       na.rm=T, 
       factorsAsStrings=T)

fhs2_melt_offspring_ex10_omni1_ex05$form <- "fhs2_offspring_ex10_omni1_ex05"

rm(fhs2_offspring_ex10_omni1_ex05)


#==============================#
####  Lab panel, Exam 10     ---#
#==============================#

fhs2_l_fhslab_ex10 <- 
  fread('~/Dropbox/BioLINCC files/Framingham offspring/datasets/CSV/l_fhslab_ex10_1b_1401d.csv',na.strings=c("NA","","NULL"))

names(fhs2_l_fhslab_ex10)[names(fhs2_l_fhslab_ex10)=="idtype"] <- "IDTYPE"
fhs2_l_fhslab_ex10$visit[fhs2_l_fhslab_ex10$IDTYPE==1] <- 10
fhs2_l_fhslab_ex10$visit[fhs2_l_fhslab_ex10$IDTYPE==7] <- 5
fhs2_melt_l_fhslab_ex10 <- melt(fhs2_l_fhslab_ex10,id.vars=c("PID","IDTYPE","visit"), na.rm=T, factorsAsStrings=T)
fhs2_melt_l_fhslab_ex10$form <- "fhs2_fhslab_ex10"

rm(fhs2_l_fhslab_ex10)


#=================================#
####  MMSE, Offspring/Omni     ---#
#=================================#

fhs2_vr_mmse_ex10 <- 
  fread('~/Dropbox/BioLINCC files/Framingham offspring/datasets/CSV/vr_mmse_ex10_1b_1395d.csv',
        na.strings=c("NA","","NULL"))

names(fhs2_vr_mmse_ex10) <- toupper(names(fhs2_vr_mmse_ex10))
names(fhs2_vr_mmse_ex10)[2] <- "visit"

fhs2_vr_mmse_ex10$COGSCR_diff <- 
  fhs2_vr_mmse_ex10$MAXCOG-fhs2_vr_mmse_ex10$COGSCR

fhs2_melt_vr_mmse_ex10 <- 
  melt(fhs2_vr_mmse_ex10,
       id.vars=c("PID","IDTYPE","visit"), 
       na.rm=T, 
       factorsAsStrings=T)

fhs2_melt_vr_mmse_ex10$form <- "fhs2_mmse_ex10"

rm(fhs2_vr_mmse_ex10)


#======================================#
####  Fibroscan, Offspring 10/Omni 5     ---#
#======================================#

fhs2_t_livrvcte_ex10 <- 
  fread('~/Dropbox/BioLINCC files/Framingham offspring/datasets/CSV/t_livrvcte_ex10_1b_1413d.csv',
        na.strings=c("NA","","NULL"))

names(fhs2_t_livrvcte_ex10) <- toupper(names(fhs2_t_livrvcte_ex10))
fhs2_t_livrvcte_ex10$visit[fhs2_t_livrvcte_ex10$IDTYPE==1] <- 10
fhs2_t_livrvcte_ex10$visit[fhs2_t_livrvcte_ex10$IDTYPE==7] <- 5

fhs2_melt_t_livrvcte_ex10 <- 
  melt(fhs2_t_livrvcte_ex10,
       id.vars=c("PID","IDTYPE","visit"), 
       na.rm=T, 
       factorsAsStrings=T)

fhs2_melt_t_livrvcte_ex10$form <- "fhs2_livrvcte_ex10"

rm(fhs2_t_livrvcte_ex10)


# ==============================================================##
####               Assemble Offspring and Omni 1 data         ####
# ==============================================================##


fhs2_melt_all <- rbind(fhs2_melt_act1_5d[,c("PID","IDTYPE","visit","variable","value","form")],
                       fhs2_melt_act1_6d[,c("PID","IDTYPE","visit","variable","value","form")],
                       fhs2_melt_cmrlvh1[,c("PID","IDTYPE","visit","variable","value","form")],
                       fhs2_melt_cmrwma1[,c("PID","IDTYPE","visit","variable","value","form")],
                       fhs2_melt_ex1_1d[,c("PID","IDTYPE","visit","variable","value","form")],
                       fhs2_melt_ex1_2d[,c("PID","IDTYPE","visit","variable","value","form")],
                       fhs2_melt_ex1_3d[,c("PID","IDTYPE","visit","variable","value","form")],
                       fhs2_melt_ex1_4d[,c("PID","IDTYPE","visit","variable","value","form")],
                       fhs2_melt_ex1_5d[,c("PID","IDTYPE","visit","variable","value","form")],
                       fhs2_melt_ex1_6d[,c("PID","IDTYPE","visit","variable","value","form")],
                       fhs2_melt_ex1_7d[,c("PID","IDTYPE","visit","variable","value","form")],
                       fhs2_melt_ex1_8d[,c("PID","IDTYPE","visit","variable","value","form")],
                       fhs2_melt_offspring_ex09_omni1_ex04[,c("PID","IDTYPE","visit","variable","value","form")],
                       fhs2_melt_offspring_ex10_omni1_ex05[,c("PID","IDTYPE","visit","variable","value","form")],
                       fhs2_melt_l_gal3_ex06[,c("PID","IDTYPE","visit","variable","value","form")],
                       fhs2_melt_l_aldost_ex06[,c("PID","IDTYPE","visit","variable","value","form")],
                       fhs2_melt_l_creacys_ex07[,c("PID","IDTYPE","visit","variable","value","form")],
                       fhs2_melt_l_ckd_ex06[,c("PID","IDTYPE","visit","variable","value","form")],
                       fhs2_melt_l_fhslab_ex01[,c("PID","IDTYPE","visit","variable","value","form")],  
                       fhs2_melt_l_fhslab_ex02[,c("PID","IDTYPE","visit","variable","value","form")],
                       fhs2_melt_l_fhslab_ex03[,c("PID","IDTYPE","visit","variable","value","form")],
                       fhs2_melt_l_fhslab_ex05[,c("PID","IDTYPE","visit","variable","value","form")],
                       fhs2_melt_l_fhslab_ex06[,c("PID","IDTYPE","visit","variable","value","form")],
                       fhs2_melt_l_fhslab_ex07[,c("PID","IDTYPE","visit","variable","value","form")], 
                       fhs2_melt_l_fhslab_ex08[,c("PID","IDTYPE","visit","variable","value","form")],
                       fhs2_melt_l_fhslab_ex09[,c("PID","IDTYPE","visit","variable","value","form")],
                       fhs2_melt_l_fhslab_ex10[,c("PID","IDTYPE","visit","variable","value","form")],
                       fhs2_melt_natpep1[,c("PID","IDTYPE","visit","variable","value","form")],
                       fhs2_melt_omni1_ex01[,c("PID","IDTYPE","visit","variable","value","form")],
                       fhs2_melt_omni1_ex02[,c("PID","IDTYPE","visit","variable","value","form")],    
                       fhs2_melt_omni1_ex03[,c("PID","IDTYPE","visit","variable","value","form")],
                       fhs2_melt_q_cesd_ex09[,c("PID","IDTYPE","visit","variable","value","form")],
                       fhs2_melt_q_psych_ex03[,c("PID","IDTYPE","visit","variable","value","form")],
                       fhs2_melt_renin1[,c("PID","IDTYPE","visit","variable","value","form")],
                       fhs2_melt_t_echo_2008[,c("PID","IDTYPE","visit","variable","value","form")],
                       fhs2_melt_t_echo_ex04[,c("PID","IDTYPE","visit","variable","value","form")],
                       fhs2_melt_t_echo_ex05[,c("PID","IDTYPE","visit","variable","value","form")],    
                       fhs2_melt_t_echo_ex06[,c("PID","IDTYPE","visit","variable","value","form")],       
                       fhs2_melt_t_echocs_ex03[,c("PID","IDTYPE","visit","variable","value","form")],
                       fhs2_melt_t_echocs_ex08[,c("PID","IDTYPE","visit","variable","value","form")],
                       fhs2_melt_t_echomvp_ex05[,c("PID","IDTYPE","visit","variable","value","form")],
                       fhs2_melt_t_echorv_ex08[,c("PID","IDTYPE","visit","variable","value","form")],
                       fhs2_melt_t_doppvasc[,c("PID","IDTYPE","visit","variable","value","form")],     
                       fhs2_melt_t_wktest_07[,c("PID","IDTYPE","visit","variable","value","form")],
                       fhs2_melt_q_ffreq_ex03[,c("PID","IDTYPE","visit","variable","value","form")],
                       fhs2_melt_vr_ffreq_ex05[,c("PID","IDTYPE","visit","variable","value","form")],
                       fhs2_melt_vr_ffreq_ex06[,c("PID","IDTYPE","visit","variable","value","form")],
                       fhs2_melt_vr_ffreq_ex07[,c("PID","IDTYPE","visit","variable","value","form")],
                       fhs2_melt_vr_ffreq_ex08[,c("PID","IDTYPE","visit","variable","value","form")],
                       fhs2_melt_vr_ffreq_omni1_ex02[,c("PID","IDTYPE","visit","variable","value","form")],
                       fhs2_melt_vr_ffreq_omni1_ex03[,c("PID","IDTYPE","visit","variable","value","form")],
                       fhs2_melt_vr_ffreq_offspring_ex09_omni1_ex04[,c("PID","IDTYPE","visit","variable","value","form")],
                       fhs2_melt_wkthru[,c("PID","IDTYPE","visit","variable","value","form")],
                       fhs2_melt_l_crp_ex02[,c("PID","IDTYPE","visit","variable","value","form")],
                       fhs2_melt_l_crp_ex06[,c("PID","IDTYPE","visit","variable","value","form")],
                       fhs2_melt_inflamm_ex07[,c("PID","IDTYPE","visit","variable","value","form")],
                       fhs2_melt_inflamm_ex08[,c("PID","IDTYPE","visit","variable","value","form")],
                       fhs2_melt_l_dbtlab_ex07[,c("PID","IDTYPE","visit","variable","value","form")],
                       fhs2_melt_sex[,c("PID","IDTYPE","visit","variable","value","form")],
                       fhs2_melt_race[,c("PID","IDTYPE","visit","variable","value","form")],
                       fhs2_melt_menses[,c("PID","IDTYPE","visit","variable","value","form")],
                       fhs2_melt_actigraphy_offspring_ex09_omni1_ex04[,c("PID","IDTYPE","visit","variable","value","form")],
                       fhs2_melt_fhorm_ex07[,c("PID","IDTYPE","visit","variable","value","form")],
                       fhs2_melt_shorm_ex03[,c("PID","IDTYPE","visit","variable","value","form")],
                       fhs2_melt_shorm_ex04[,c("PID","IDTYPE","visit","variable","value","form")],
                       fhs2_melt_vr_mmse_ex10[,c("PID","IDTYPE","visit","variable","value","form")],
                       fhs2_melt_t_livrvcte_ex10[,c("PID","IDTYPE","visit","variable","value","form")])


fhs2_melt_all <- merge(fhs2_melt_all,
                       fhs2_wkthru[,c("PID",
                                      "visit",
                                      "ATT",
                                      "SEX")],
                       by=c("PID","visit"),
                       all.x=T)


fhs2_melt_all <- subset(fhs2_melt_all,
                        ATT==1)

fhs2_melt_all$variable <- as.character(fhs2_melt_all$variable)

# ============================================================ #
####                  Assemble Framingham DM                ####
# ============================================================ #


fhs1_vr_diab <- fread('~/Dropbox/BioLINCC files/Framingham cohort/datasets/CSV/VR_DIAB_EX28_0_0601D.csv',
                      na.strings=c("NA","","NULL"),
                      stringsAsFactors=F)

names(fhs1_vr_diab) <- toupper(names(fhs1_vr_diab))

fhs1_melt_vr_diab <- melt(fhs1_vr_diab[,c(1,25,82:109)],
                          id.vars=c("PID",
                                    "IDTYPE"),
                          na.rm=T,
                          factorsAsStrings=T)

fhs1_melt_vr_diab$variable <- as.character(fhs1_melt_vr_diab$variable)

fhs1_melt_vr_diab$visit <- str_select(fhs1_melt_vr_diab$variable,
                                      after="BG200_HX_DIAB")

fhs1_melt_vr_diab$variable <- "BG200_HX_DIAB"

fhs1_melt_vr_diab$form <- "fhs1_vr_diab"

rm(fhs1_vr_diab)

#### Diabetes status by cycle

fhs2_vr_diab <- fread('~/Dropbox/BioLINCC files/Framingham offspring/datasets/CSV/VR_DIAB_EX10_1b_1489D.csv',
                      na.strings=c("NA","","NULL"),
                      stringsAsFactors=F)

names(fhs2_vr_diab) <- toupper(names(fhs2_vr_diab))

fhs2_vr_diab <- 
  fhs2_vr_diab[,c(1,12:22)]

fhs2_melt_vr_diab <- melt(fhs2_vr_diab,
                          id.vars=c("PID",
                                    "IDTYPE"),
                          na.rm=T,
                          factorsAsStrings=T)

fhs2_melt_vr_diab$variable <- as.character(fhs2_melt_vr_diab$variable)

fhs2_melt_vr_diab$visit <- str_select(fhs2_melt_vr_diab$variable,
                                      after="HX_DIAB")

fhs2_melt_vr_diab$variable <- "HX_DIAB"

fhs2_melt_vr_diab$form <- "fhs2_vr_diab"

rm(fhs2_vr_diab)

####

fhs3_vr_diab <- 
  fread('~/Dropbox/BioLINCC files/Framingham third generation/datasets/CSV/VR_DIAB_EX03_3b_1312D.csv',
        na.strings=c("NA","","NULL")) # Diabetes status by cycle


names(fhs3_vr_diab) <- toupper(names(fhs3_vr_diab))

fhs3_melt_vr_diab <- melt(fhs3_vr_diab[,c("PID","IDTYPE","HX_DIAB1","HX_DIAB2")],
                          id.vars=c("PID",
                                    "IDTYPE"),
                          na.rm=T,
                          factorsAsStrings=T)

fhs3_melt_vr_diab$variable <- as.character(fhs3_melt_vr_diab$variable)

fhs3_melt_vr_diab$visit <- str_select(fhs3_melt_vr_diab$variable,
                                      after="HX_DIAB")

fhs3_melt_vr_diab$variable <- "HX_DIAB"

fhs3_melt_vr_diab$form <- "fhs3_vr_diab"

rm(fhs3_vr_diab)

fhs_vr_diab <- rbind(fhs1_melt_vr_diab,
                     fhs2_melt_vr_diab,
                     fhs3_melt_vr_diab)

names(fhs_vr_diab)[2] <- "cohort"
fhs_vr_diab$cohort <- as.character(fhs_vr_diab$cohort)
fhs_vr_diab$visit <- as.integer(fhs_vr_diab$visit)

fhs_vr_diab <- merge(fhs_vr_diab,
                     fhs_dates_long[,c("PID","cohort","visit","visitdays")],
                     by=c("PID","cohort","visit"))

fhs_vr_diab$study <- "FHS"

rm(fhs1_melt_vr_diab,
   fhs2_melt_vr_diab,
   fhs3_melt_vr_diab)


# ==========================================================================#
##                                                                         ##
####                  ******** FRAMINGHAM GEN III ********               ####
##                                                                         ##
# ==========================================================================#


#=======================================#
####    *** FHS3: Gen 3 - Exam 1 *** ####
#=======================================#

fhs3_e_exam_ex01_id3 <- 
  fread('~/Dropbox/BioLINCC files/Framingham third generation/datasets/CSV/E_EXAM_EX01_3_0086D.csv')

## Calculate drinks/week by adding beers/wk + white wine/week + red wine/week + liquor/week + other/week

fhs3_e_exam_ex01_id3$drinks_wk <- 
  fhs3_e_exam_ex01_id3$G3A115+
  fhs3_e_exam_ex01_id3$G3A119+
  fhs3_e_exam_ex01_id3$G3A123+
  fhs3_e_exam_ex01_id3$G3A127+
  fhs3_e_exam_ex01_id3$G3A131

fhs3_e_exam_ex01_id3$CESD20_missing <- 
  rowSums(is.na(fhs3_e_exam_ex01_id3[,402:421]))

fhs3_e_exam_ex01_id3$CESD20 <- 
  rowSums(fhs3_e_exam_ex01_id3[,402:421])

fhs3_e_exam_ex01_id3$CESD20[fhs3_e_exam_ex01_id3$CESD20_missing >= 4] <- NA

fhs3_e_exam_ex01_id3$ever_smoke <- 0
fhs3_e_exam_ex01_id3$ever_smoke[fhs3_e_exam_ex01_id3$G3A070==1|    # Cigarettes
                                  fhs3_e_exam_ex01_id3$G3A079==1| # Pipes
                                  fhs3_e_exam_ex01_id3$G3A088==1] <- 1  # Cigars

fhs3_e_exam_ex01_id3$G3A023[!between(fhs3_e_exam_ex01_id3$G3A023,7,25)] <- NA
fhs3_e_exam_ex01_id3$G3A032[!between(fhs3_e_exam_ex01_id3$G3A032,1,30)] <- NA
fhs3_e_exam_ex01_id3$G3A034[!between(fhs3_e_exam_ex01_id3$G3A034,1,11)] <- NA
fhs3_e_exam_ex01_id3$G3A035[!between(fhs3_e_exam_ex01_id3$G3A035,1,10)] <- NA
fhs3_e_exam_ex01_id3$G3A036[!between(fhs3_e_exam_ex01_id3$G3A036,15,43)] <- NA
fhs3_e_exam_ex01_id3$G3A037[!between(fhs3_e_exam_ex01_id3$G3A037,16,45)] <- NA
fhs3_e_exam_ex01_id3$G3A040[!between(fhs3_e_exam_ex01_id3$G3A040,19,57)] <- NA
fhs3_e_exam_ex01_id3$G3A044[!between(fhs3_e_exam_ex01_id3$G3A044,9,57)] <- NA
fhs3_e_exam_ex01_id3$G3A051[!between(fhs3_e_exam_ex01_id3$G3A051,16,51)] <- NA
fhs3_e_exam_ex01_id3$G3A055[!between(fhs3_e_exam_ex01_id3$G3A055,1,32)] <- NA
fhs3_e_exam_ex01_id3$G3A056[!between(fhs3_e_exam_ex01_id3$G3A056,1,12)] <- NA

fhs3_e_exam_ex01_id3$fhs_pai <- 
  calc_framingham_pai(fhs3_e_exam_ex01_id3,
                      slp_hrs="G3A596",
                      sed_hrs="G3A597",
                      slgt_hrs="G3A598",
                      mod_hrs="G3A599",
                      hvy_hrs="G3A600")


fhs3_e_exam_ex01_id3$hrt_years_total <- 
  fhs3_e_exam_ex01_id3$G3A055 +
  fhs3_e_exam_ex01_id3$G3A056/12

fhs3_e_exam_ex01_id3$race[fhs3_e_exam_ex01_id3$G3A483==1] <- "W"
fhs3_e_exam_ex01_id3$race[fhs3_e_exam_ex01_id3$G3A484==1] <- "Hisp"
fhs3_e_exam_ex01_id3$race[fhs3_e_exam_ex01_id3$G3A485==1] <- "B"
fhs3_e_exam_ex01_id3$race[fhs3_e_exam_ex01_id3$G3A486==1] <- "A"
fhs3_e_exam_ex01_id3$race[fhs3_e_exam_ex01_id3$G3A488==1] <- "N"
fhs3_e_exam_ex01_id3$race[fhs3_e_exam_ex01_id3$G3A489==1] <- "O"


fhs3_meno_ex01_id3 <- 
  fhs3_e_exam_ex01_id3[fhs3_e_exam_ex01_id3$G3A033 %in% c(0,1),c("PID",
                                                                 "IDTYPE",
                                                                 "G3A023",
                                                                 "G3A045",
                                                                 "G3A051")]


fhs3_melt_e_exam_ex01_id3 <- melt(fhs3_e_exam_ex01_id3,id.vars=c("PID","IDTYPE"),na.rm=T,factorsAsStrings=T)
fhs3_melt_e_exam_ex01_id3$visit <- 1
fhs3_melt_e_exam_ex01_id3$form <- "fhs3_e_ex01"


#===================================#
####    FHS3: Omni 2 - Exam 1    ---#
#===================================#

fhs3_e_exam_ex01_id72 <- fread('~/Dropbox/BioLINCC files/Framingham third generation/datasets/CSV/E_EXAM_EX01_72_0652D.csv')

## Calculate drinks/week by adding beers/wk + white wine/week + red wine/week + liquor/week + other/week

fhs3_e_exam_ex01_id72$drinks_wk <- 
  fhs3_e_exam_ex01_id72$G3A115+
  fhs3_e_exam_ex01_id72$G3A119+
  fhs3_e_exam_ex01_id72$G3A123+
  fhs3_e_exam_ex01_id72$G3A127+
  fhs3_e_exam_ex01_id72$G3A131

fhs3_e_exam_ex01_id72$CESD20_missing <- rowSums(is.na(fhs3_e_exam_ex01_id72[,402:421]))
fhs3_e_exam_ex01_id72$CESD20 <- rowSums(fhs3_e_exam_ex01_id72[,402:421])
fhs3_e_exam_ex01_id72$CESD20[fhs3_e_exam_ex01_id72$CESD20_missing >= 4] <- NA

fhs3_e_exam_ex01_id72$ever_smoke <- 0
fhs3_e_exam_ex01_id72$ever_smoke[fhs3_e_exam_ex01_id72$G3A070==1|    # Cigarettes
                                   fhs3_e_exam_ex01_id72$G3A079==1| # Pipes
                                   fhs3_e_exam_ex01_id72$G3A088==1] <- 1  # Cigars


fhs3_e_exam_ex01_id72$fhs_pai <- 
  calc_framingham_pai(fhs3_e_exam_ex01_id72,
                      slp_hrs="G3A596",
                      sed_hrs="G3A597",
                      slgt_hrs="G3A598",
                      mod_hrs="G3A599",
                      hvy_hrs="G3A600")


fhs3_e_exam_ex01_id72$race[fhs3_e_exam_ex01_id72$G3A483==1] <- "W"
fhs3_e_exam_ex01_id72$race[fhs3_e_exam_ex01_id72$G3A484==1] <- "Hisp"
fhs3_e_exam_ex01_id72$race[fhs3_e_exam_ex01_id72$G3A485==1] <- "B"
fhs3_e_exam_ex01_id72$race[fhs3_e_exam_ex01_id72$G3A486==1] <- "A"
fhs3_e_exam_ex01_id72$race[fhs3_e_exam_ex01_id72$G3A488==1] <- "N"
fhs3_e_exam_ex01_id72$race[fhs3_e_exam_ex01_id72$G3A489==1] <- "O"


fhs3_meno_ex01_id72 <- 
  fhs3_e_exam_ex01_id72[fhs3_e_exam_ex01_id72$G3A033 %in% c(0,1),c("PID",
                                                                   "IDTYPE",
                                                                   "G3A023",
                                                                   "G3A045",
                                                                   "G3A051")]


fhs3_melt_e_exam_ex01_id72 <- 
  melt(fhs3_e_exam_ex01_id72,
       id.vars=c("PID","IDTYPE"),
       na.rm=T,
       factorsAsStrings=T)

fhs3_melt_e_exam_ex01_id72$visit <- 1
fhs3_melt_e_exam_ex01_id72$form <- "fhs3_e_ex01"


#=======================================#
####  FHS Offspring Spouse - Exam 1  ---#
#=======================================#

fhs3_e_exam_ex01_id2 <- fread('~/Dropbox/BioLINCC files/Framingham third generation/datasets/CSV/E_EXAM_EX01_2_0813D.csv')

## Calculate drinks/week by adding beers/wk + white wine/week + red wine/week + liquor/week + other/week

fhs3_e_exam_ex01_id2$drinks_wk <- 
  fhs3_e_exam_ex01_id2$G3A115+
  fhs3_e_exam_ex01_id2$G3A119+
  fhs3_e_exam_ex01_id2$G3A123+
  fhs3_e_exam_ex01_id2$G3A127+
  fhs3_e_exam_ex01_id2$G3A131

fhs3_e_exam_ex01_id2$CESD20_missing <- rowSums(is.na(fhs3_e_exam_ex01_id2[,398:417]))
fhs3_e_exam_ex01_id2$CESD20 <- rowSums(fhs3_e_exam_ex01_id2[,398:417])
fhs3_e_exam_ex01_id2$CESD20[fhs3_e_exam_ex01_id2$CESD20_missing >= 4] <- NA

fhs3_e_exam_ex01_id2$ever_smoke <- 0
fhs3_e_exam_ex01_id2$ever_smoke[fhs3_e_exam_ex01_id2$G3A070==1|    # Cigarettes
                                  fhs3_e_exam_ex01_id2$G3A079==1| # Pipes
                                  fhs3_e_exam_ex01_id2$G3A088==1] <- 1  # Cigars

fhs3_e_exam_ex01_id2$fhs_pai <- 
  calc_framingham_pai(fhs3_e_exam_ex01_id2,
                      slp_hrs="G3A596",
                      sed_hrs="G3A597",
                      slgt_hrs="G3A598",
                      mod_hrs="G3A599",
                      hvy_hrs="G3A600")


fhs3_e_exam_ex01_id2$race[fhs3_e_exam_ex01_id2$G3A483==1] <- "W"
fhs3_e_exam_ex01_id2$race[fhs3_e_exam_ex01_id2$G3A484==1] <- "Hisp"
fhs3_e_exam_ex01_id2$race[fhs3_e_exam_ex01_id2$G3A485==1] <- "B"
fhs3_e_exam_ex01_id2$race[fhs3_e_exam_ex01_id2$G3A486==1] <- "A"
fhs3_e_exam_ex01_id2$race[fhs3_e_exam_ex01_id2$G3A488==1] <- "N"
fhs3_e_exam_ex01_id2$race[fhs3_e_exam_ex01_id2$G3A489==1] <- "O"


fhs3_meno_ex01_id2 <- 
  fhs3_e_exam_ex01_id2[fhs3_e_exam_ex01_id2$G3A033 %in% c(0,1),
                       c("PID",
                         "IDTYPE",
                         "G3A023",
                         "G3A045",
                         "G3A051")]

fhs3_meno_ex01 <- 
  rbind(fhs3_meno_ex01_id2,
        fhs3_meno_ex01_id3,
        fhs3_meno_ex01_id72)


fhs3_melt_e_exam_ex01_id2 <- melt(fhs3_e_exam_ex01_id2,id.vars=c("PID","IDTYPE"),na.rm=T,factorsAsStrings=T)
fhs3_melt_e_exam_ex01_id2$visit <- 1
fhs3_melt_e_exam_ex01_id2$form <- "fhs3_e_ex01"


#======================================================#
####  FHS3: Gen 3, Offspring Spouse, Omni 2 - Race  ---#
#======================================================#


fhs3_race_id2 <- fhs3_e_exam_ex01_id2[,c("PID",
                                         "IDTYPE",
                                         "G3A483",
                                         "G3A484",
                                         "G3A485",
                                         "G3A486",
                                         "G3A488",
                                         "G3A489"
)]


fhs3_race_id3 <- fhs3_e_exam_ex01_id3[,c("PID",
                                         "IDTYPE",
                                         "G3A483",
                                         "G3A484",
                                         "G3A485",
                                         "G3A486",
                                         "G3A488",
                                         "G3A489"
)]


fhs3_race_id72 <- fhs3_e_exam_ex01_id72[,c("PID",
                                           "IDTYPE",
                                           "G3A483",
                                           "G3A484",
                                           "G3A485",
                                           "G3A486",
                                           "G3A488",
                                           "G3A489"
)]


fhs3_race <- rbind(fhs3_race_id2,
                   fhs3_race_id3,
                   fhs3_race_id72)


fhs3_race$race[fhs3_race$G3A483==1] <- 1
fhs3_race$race[fhs3_race$G3A489==1] <- 6
fhs3_race$race[fhs3_race$G3A485==1] <- 2
fhs3_race$race[fhs3_race$G3A486==1] <- 4
fhs3_race$race[fhs3_race$G3A488==1] <- 5
fhs3_race$race[fhs3_race$G3A484==1] <- 3

fhs3_melt_race <- melt(fhs3_race,
                       id.vars=c("PID","IDTYPE"),
                       na.rm=T,
                       stringsAsFactors=F)
fhs3_melt_race$visit <- 1
fhs3_melt_race$form <- "fhs3_race"

names(fhs3_race)[3:8] <- c("White",
                           "Hispanic",
                           "Black",
                           "Asian",
                           "Native American",
                           "Other")


#=========================================#
####  NT-proBNP, Framingham 3, Exam 1  ---#
#=========================================#

fhs3_bnp3 <- fread('~/Dropbox/BioLINCC files/Framingham third generation/datasets/CSV/BNP3_1D.csv')
fhs3_melt_bnp3 <- melt(fhs3_bnp3,
                       id.vars=c("PID",
                                 "IDTYPE"), 
                       na.rm=T, 
                       factorsAsStrings=T)

fhs3_melt_bnp3$visit <- 1
fhs3_melt_bnp3$form <- "fhs3_bnp3"

rm(fhs3_bnp3)


#=============================================================#
####  CRP, Framingham Offspring Spouse and Omni 2, Exam 1  ---#
#=============================================================#

fhs3_inflamm <- fread('~/Dropbox/BioLINCC files/Framingham third generation/datasets/CSV/l_inflamm_ex01_3b_1107d.csv')
fhs3_melt_inflamm <- melt(fhs3_inflamm,id.vars=c("PID","IDTYPE"),na.rm=T,factorsAsStrings = T)
fhs3_melt_inflamm$visit <- 1
fhs3_melt_inflamm$form <- "fhs3_inflamm"

rm(fhs3_inflamm)

#========================================================================#
####  Aldosterone/renin, Framingham 3/Offspring Spouse/Omni 2,Exam 1  ---#
#========================================================================#

fhs3_renaldo3 <- fread('~/Dropbox/BioLINCC files/Framingham third generation/datasets/CSV/RENALDO3_1D_V1.csv')
fhs3_renaldo3$aldo_renin <- (fhs3_renaldo3$ALDO/10)/fhs3_renaldo3$RENIN
fhs3_melt_renaldo3 <-  melt(fhs3_renaldo3,id.vars=c("PID","IDTYPE"),na.rm=T,factorsAsStrings = T)
fhs3_melt_renaldo3$visit <- 1
fhs3_melt_renaldo3$form <- "fhs3_renaldo3"

rm(fhs3_renaldo3)


#==================================================================#
####  Estrogen levels, Framingham 3, Exam 1/Omni 2, Exam 1      ---#
#==================================================================#

fhs3_estr_ex01 <- fread('~/Dropbox/BioLINCC files/Framingham third generation/datasets/CSV/l_estr_2005_m_0622d_v1.csv')
fhs3_melt_estr_ex01 <-  melt(fhs3_estr_ex01,id.vars=c("PID","IDTYPE"),na.rm=T,factorsAsStrings = T)
fhs3_melt_estr_ex01$visit <- 1
fhs3_melt_estr_ex01$form <- "fhs3_estr_ex01"

rm(fhs3_estr_ex01)


#==================================================================#
####  Troponin levels, Framingham 3, Exam 1/Omni 2, Exam 1      ---#
#==================================================================#

fhs3_tni_ex01 <- fread('~/Dropbox/BioLINCC files/Framingham third generation/datasets/CSV/l_hstn_ex01_3b_0832d_v1.csv')
fhs3_melt_tni_ex01 <-  melt(fhs3_tni_ex01,id.vars=c("PID","IDTYPE"),na.rm=T,factorsAsStrings = T)
fhs3_melt_tni_ex01$visit <- 1
fhs3_melt_tni_ex01$form <- "fhs3_tni_ex01"

rm(fhs3_tni_ex01)

#================================================================#
####  Male hormones, Framingham 3, Exam 1/Omni 2, Exam 1      ---#
#================================================================#

fhs3_mhorm_ex01 <- fread('~/Dropbox/BioLINCC files/Framingham third generation/datasets/CSV/l_mhorm_2005_m_0490d_v1.csv')
fhs3_melt_mhorm_ex01 <-  melt(fhs3_mhorm_ex01,id.vars=c("PID","IDTYPE"),na.rm=T,factorsAsStrings = T)
fhs3_melt_mhorm_ex01$visit <- 1
fhs3_melt_mhorm_ex01$form <- "fhs3_mhorm_ex01"

rm(fhs3_mhorm_ex01)

#=========================================#
####  Telomeres, Framingham 3, Exam 1  ---#
#=========================================#

fhs3_telomere_ex01 <- fread('~/Dropbox/BioLINCC files/Framingham third generation/datasets/CSV/l_telomere_2005_m_1161d.csv')
fhs3_melt_telomere_ex01 <-  melt(fhs3_telomere_ex01,id.vars=c("PID","IDTYPE"),na.rm=T,factorsAsStrings = T)
fhs3_melt_telomere_ex01$visit <- 1
fhs3_melt_telomere_ex01$form <- "fhs3_telomere_ex01"

rm(fhs3_telomere_ex01)

#=========================================#
####  VEGF, Framingham 3,      Exam 1  ---#
#=========================================#

fhs3_sflt3_ex01 <- fread('~/Dropbox/BioLINCC files/Framingham third generation/datasets/CSV/sflt3_1d.csv')
fhs3_melt_sflt3_ex01 <-  melt(fhs3_sflt3_ex01,id.vars=c("PID","IDTYPE"),na.rm=T,factorsAsStrings = T)
fhs3_melt_sflt3_ex01$visit <- 1
fhs3_melt_sflt3_ex01$form <- "fhs3_sflt3_ex01"

rm(fhs3_sflt3_ex01)


#================================================#
####  CT abdominal fat, Framingham 3, Exam 1  ---#
#================================================#

fhs3_ctabdom_ex01 <- fread('~/Dropbox/BioLINCC files/Framingham third generation/datasets/CSV/t_ctabdom_2005_m_0227d.csv')

fhs3_ctabdom_ex01$vat_sat_ratio <-
  fhs3_ctabdom_ex01$OB2VOL/fhs3_ctabdom_ex01$OB1VOL

fhs3_ctabdom_ex01$vat_sat_ratio_log <-
  log(fhs3_ctabdom_ex01$vat_sat_ratio)

fhs3_melt_ctabdom_ex01 <-  melt(fhs3_ctabdom_ex01,id.vars=c("PID","IDTYPE"),na.rm=T,factorsAsStrings = T)
fhs3_melt_ctabdom_ex01$visit <- 1
fhs3_melt_ctabdom_ex01$form <- "fhs3_ctabdom_ex01"

rm(fhs3_ctabdom_ex01)


#=======================================#
####  CT Lung, Framingham 3, Exam 1  ---#
#=======================================#

fhs3_ctlung_ex01 <- fread('~/Dropbox/BioLINCC files/Framingham third generation/datasets/CSV/t_ctlung_2005_m_0696d.csv')

fhs3_ctlung_ex01$global_vol_ml_avg <-
  mean(c(fhs3_ctlung_ex01$GLOBAL_VOL_ML_1,
         fhs3_ctlung_ex01$GLOBAL_VOL_ML_2),
       na.rm=T)

fhs3_melt_ctlung_ex01 <-  melt(fhs3_ctlung_ex01,id.vars=c("PID","IDTYPE"),na.rm=T,factorsAsStrings = T)
fhs3_melt_ctlung_ex01$visit <- 1
fhs3_melt_ctlung_ex01$form <- "fhs3_ctlung_ex01"

rm(fhs3_ctlung_ex01)


#======================================#
####  CAC, Framingham 3, Exam 1     ---#
#======================================#

fhs3_cac_ex01 <- fread('~/Dropbox/BioLINCC files/Framingham third generation/datasets/CSV/t_ctthrcac_ex01_3_0287d.csv')

fhs3_cac_ex01$cac_avg <-
  mean(c(fhs3_cac_ex01$CA1,
         fhs3_cac_ex01$CA2),
       na.rm=T)

fhs3_melt_cac_ex01 <-  melt(fhs3_cac_ex01,id.vars=c("PID","IDTYPE"),na.rm=T,factorsAsStrings = T)
fhs3_melt_cac_ex01$visit <- 1
fhs3_melt_cac_ex01$form <- "fhs3_cac_ex01"

rm(fhs3_cac_ex01)


#==============================================================================#
####  Vascular tonometry, Framingham 3/New Offspring Spouse/Omni 2, Exam 1  ---#
#==============================================================================#

fhs3_tonometry_ex01 <- fread('~/Dropbox/BioLINCC files/Framingham third generation/datasets/CSV/t_tonla_ex01_3b_1234d.csv')


fhs3_tonometry_ex01$e_eprime_avg <- fhs3_tonometry_ex01$MEMAX/fhs3_tonometry_ex01$DLEMAX
fhs3_tonometry_ex01$ea_ratio <- fhs3_tonometry_ex01$MEMAX/fhs3_tonometry_ex01$MAMAX

fhs3_melt_tonometry_ex01 <-  melt(fhs3_tonometry_ex01,id.vars=c("PID","IDTYPE"),na.rm=T,factorsAsStrings = T)
fhs3_melt_tonometry_ex01$visit <- 1
fhs3_melt_tonometry_ex01$form <- "fhs3_tonometry_ex01"

rm(fhs3_tonometry_ex01)


#========================================================#
####  FHS lab values, Offspring Spouse/Omni 2,Exam 1  ---#
#========================================================#

fhs3_fhslab_ex01 <- fread('~/Dropbox/BioLINCC files/Framingham third generation/datasets/CSV/L_FHSLAB_EX01_3B_0800D.csv')
fhs3_fhslab_ex01 <- merge(fhs3_fhslab_ex01,
                          fhs3_wkthru[,c("PID","IDTYPE","SEX","AGE1")],
                          by=c("PID","IDTYPE"),
                          all.x=T)
labcols <- c(names(fhs3_fhslab_ex01),"gfr_mdrd")
fhs3_fhslab_ex01 <- merge(fhs3_fhslab_ex01,
                          fhs3_race,
                          by=c("PID","IDTYPE"),
                          all.x=T)

fhs3_fhslab_ex01$gfr_mdrd <- calc_MDRD4(dat=fhs3_fhslab_ex01,
                                        cr="CREAT",
                                        age="AGE1",
                                        sex="SEX",
                                        race="Black",
                                        black=1)

fhs3_melt_fhslab_ex01 <- melt(fhs3_fhslab_ex01[,..labcols],
                              id.vars=c("PID","IDTYPE"),
                              na.rm=T,
                              factorsAsStrings = T)

fhs3_melt_fhslab_ex01$visit <- 1
fhs3_melt_fhslab_ex01$form <- "fhs3_fhslab_ex01"
rm(fhs3_fhslab_ex01)


#================================================================#
####   Male hormones in females, third generation, Exam 1     ---#
#================================================================#

fhs3_fhorm_ex01 <- fread('~/Dropbox/BioLINCC files/Framingham third generation/datasets/CSV/fhorm_2005d.csv')
fhs3_melt_fhorm_ex01 <-  melt(fhs3_fhorm_ex01,id.vars=c("PID","IDTYPE"),na.rm=T,factorsAsStrings = T)
fhs3_melt_fhorm_ex01$visit <- 1
fhs3_melt_fhorm_ex01$form <- "fhs3_fhorm_ex01"

rm(fhs3_fhorm_ex01)


#===============================================================#
####  Echocardiogram - FHS Offspring Spouse/Omni 2 - Exam 1  ---#
#===============================================================#

fhs3_t_echo_2008 <- fread('~/Dropbox/BioLINCC files/Framingham third generation/datasets/CSV/T_ECHO_2008_M_0549D_V1.csv')


fhs3_t_echo_2008 <- merge(fhs3_t_echo_2008,
                          fhs3_wkthru[,c("PID","IDTYPE","SEX","WGT1","HGT1")],
                          by=c("PID","IDTYPE"), all.x=T)

fhs3_t_echo_2008$HEIGHT <- fhs3_t_echo_2008$HGT1*2.54
fhs3_t_echo_2008$WEIGHT <- fhs3_t_echo_2008$WGT1/2.2

fhs3_t_echo_2008 <- 
  as.data.table(
    calc_hypertrophy_type(
      fhs3_t_echo_2008, 
      id = "PID",
      sex="SEX",
      lvedd="X76",
      ivsd="X62",
      lvpwtd="X69",
      height="HEIGHT",
      weight="WEIGHT"))

fhs3_melt_t_echo_2008 <- melt(fhs3_t_echo_2008,id.vars=c("PID","IDTYPE"), na.rm=T, factorsAsStrings=T)
fhs3_melt_t_echo_2008$visit <- 1

fhs3_melt_t_echo_2008$form <- "fhs3_echo_ex01"

rm(fhs3_t_echo_2008)

#================================================================================#
####         Echo Doppler Diastolic function - FHS Gen 3/Omni 2 - Exam 1      ---#
#================================================================================#

fhs3_t_doppvasc_dd_2008  <- fread('~/Dropbox/BioLINCC files/Framingham third generation/datasets/CSV/T_DOPPVASC_2008_M_0756D.csv')
names(fhs3_t_doppvasc_dd_2008)[names(fhs3_t_doppvasc_dd_2008)=="idtype"] <- "IDTYPE"
fhs3_t_doppvasc_dd_2008$visit <- 1 
fhs3_t_doppvasc_dd_2008$earatio <- fhs3_t_doppvasc_dd_2008$MEMAX/fhs3_t_doppvasc_dd_2008$MAMAX
fhs3_t_doppvasc_dd_2008$e_eprime_lateral <- fhs3_t_doppvasc_dd_2008$MEMAX/fhs3_t_doppvasc_dd_2008$DEMAX

fhs3_melt_t_doppvasc_dd_2008 <- melt(fhs3_t_doppvasc_dd_2008,
                                     id.vars=c("PID","IDTYPE","visit"), 
                                     na.rm=T, 
                                     factorsAsStrings=T)
fhs3_melt_t_doppvasc_dd_2008$form <- "fhs3_doppvasc_2008"

rm(fhs3_t_doppvasc_dd_2008)


#======================================================#
####      Echocardiogram - FHS Gen 3 - Exam 1       ---#
#======================================================#

fhs3_t_echo_ex01_id3  <- fread('~/Dropbox/BioLINCC files/Framingham third generation/datasets/CSV/T_ECHO_EX01_3_0042D.csv')

fhs3_t_echo_ex01_id3 <- merge(fhs3_t_echo_ex01_id3,
                              fhs3_wkthru[,c("PID","IDTYPE","SEX","WGT1","HGT1")],
                              by=c("PID","IDTYPE"), all.x=T)

fhs3_t_echo_ex01_id3$HEIGHT <- fhs3_t_echo_ex01_id3$HGT1*2.54
fhs3_t_echo_ex01_id3$WEIGHT <- fhs3_t_echo_ex01_id3$WGT1/2.2

fhs3_t_echo_ex01_id3 <- 
  as.data.table(
    calc_hypertrophy_type(
      fhs3_t_echo_ex01_id3, 
      id = "PID",
      sex="SEX",
      lvedd="X76",
      ivsd="X62",
      lvpwtd="X69",
      height="HEIGHT",
      weight="WEIGHT"))


fhs3_t_echo_ex01_id3$LVEDD_index <-
  fhs3_t_echo_ex01_id3$X83/fhs3_t_echo_ex01_id3$bsa

fhs3_melt_t_echo_ex01_id3 <- melt(fhs3_t_echo_ex01_id3,
                                  id.vars=c("PID","IDTYPE"),
                                  na.rm=T,
                                  factorsAsStrings=T)

fhs3_melt_t_echo_ex01_id3$visit <- 1
fhs3_melt_t_echo_ex01_id3$form <- "fhs3_echo_ex01"

rm(fhs3_t_echo_ex01_id3)

#==========================================#
####      PFT - FHS Gen 3 - Exam 1      ---#
#==========================================#

fhs3_t_pft_ex01_id3  <- fread('~/Dropbox/BioLINCC files/Framingham third generation/datasets/CSV/T_PFT_EX01_3_0170D.csv')

fhs3_melt_t_pft_ex01_id3 <- melt(fhs3_t_pft_ex01_id3,id.vars=c("PID","IDTYPE"),na.rm=T,factorsAsStrings=T)
fhs3_melt_t_pft_ex01_id3$visit <- 1
fhs3_melt_t_pft_ex01_id3$form <- "fhs3_pft_ex01"


rm(fhs3_t_pft_ex01_id3)

#===================================================================#
####  PFT Diffusion test - FHS Offspring Spouse/Omni 2 - Exam 1  ---#
#===================================================================#

fhs3_t_pftdiff_2005  <- fread('~/Dropbox/BioLINCC files/Framingham third generation/datasets/CSV/T_PFTDIFF_2005_M_0762D.csv')
fhs3_t_pftdiff_2005$DLCO_VA_mean <- rowMeans(fhs3_t_pftdiff_2005[,c('DVA1_1','DVA2_1','DVA3_1')],na.rm=T)

fhs3_melt_t_pftdiff_2005 <- melt(fhs3_t_pftdiff_2005,id.vars=c("PID","IDTYPE"),na.rm=T,factorsAsStrings=T)
fhs3_melt_t_pftdiff_2005$visit <- 1
fhs3_melt_t_pftdiff_2005$form <- "fhs3_pftdiff_2005"

rm(fhs3_t_pftdiff_2005)


#-------------------------------------------------#
####  PFT Diffusion test - FHS Gen 3 - Exam 1  ---#
#-------------------------------------------------#

fhs3_t_pftdiff_ex01_id3  <- fread('~/Dropbox/BioLINCC files/Framingham third generation/datasets/CSV/T_PFTDIFF_EX01_3_0868D.csv')
fhs3_t_pftdiff_ex01_id3$DLCO_VA_mean <- fhs3_t_pftdiff_ex01_id3$DVA1_1_3

fhs3_melt_t_pftdiff_ex01_id3 <- melt(fhs3_t_pftdiff_ex01_id3,id.vars=c("PID","IDTYPE"),na.rm=T,factorsAsStrings=T)
fhs3_melt_t_pftdiff_ex01_id3$visit <- 1
fhs3_melt_t_pftdiff_ex01_id3$form <- "fhs3_pftdiff_ex01"

rm(fhs3_t_pftdiff_ex01_id3)

#======================================================================#
####  Dietary guidelines adherence index 2010 - FHS Gen 3 - Exam 1  ---#
#======================================================================#

fhs3_vr_dgai2010_ex01_id3  <- fread('~/Dropbox/BioLINCC files/Framingham third generation/datasets/CSV/VR_DGAI2010_EX01_3_1078D.csv')

fhs3_melt_vr_dgai2010_ex01_id3 <- melt(fhs3_vr_dgai2010_ex01_id3,id.vars=c("PID","IDTYPE"),na.rm=T,factorsAsStrings=T)
fhs3_melt_vr_dgai2010_ex01_id3$visit <- 1
fhs3_melt_vr_dgai2010_ex01_id3$form <- "fhs3_dgai_ex01"

rm(fhs3_vr_dgai2010_ex01_id3)


#===========================================================#
####  Food frequency questionnaire - FHS Gen 3 - Exam 1  ---#
#===========================================================#

fhs3_vr_ffreq_ex01_id3  <- fread('~/Dropbox/BioLINCC files/Framingham third generation/datasets/CSV/VR_FFREQ_EX01_3_0587D.csv')

## Convert servings/week  to  servings/day
### Coffee  (FFD112)

fhs3_vr_ffreq_ex01_id3$FFD112_daily  <- fhs3_vr_ffreq_ex01_id3$FFD112/7 

### Decaf coffee (FFD111)
fhs3_vr_ffreq_ex01_id3$FFD111_daily  <- fhs3_vr_ffreq_ex01_id3$FFD111/7 

### Tea (FFD113)
fhs3_vr_ffreq_ex01_id3$FFD113_daily  <- fhs3_vr_ffreq_ex01_id3$FFD113/7 

fhs3_melt_vr_ffreq_ex01_id3 <- melt(fhs3_vr_ffreq_ex01_id3,id.vars=c("PID","IDTYPE"),na.rm=T,factorsAsStrings=T)
fhs3_melt_vr_ffreq_ex01_id3$visit <- 1
fhs3_melt_vr_ffreq_ex01_id3$form <- "fhs3_vr_ffreq"

rm(fhs3_vr_ffreq_ex01_id3)


#======================================================================#
####  Food frequency questionnaire - FHS Offspring Spouse - Exam 1  ---#
#======================================================================#

fhs3_vr_ffreq_ex01_id2  <- fread('~/Dropbox/BioLINCC files/Framingham third generation/datasets/CSV/VR_FFREQ_EX01_2_0984d_v1.csv')

## Convert servings/week  to  servings/day
### Coffee  (FFD112)

fhs3_vr_ffreq_ex01_id2$FFD112_daily  <- fhs3_vr_ffreq_ex01_id2$FFD112/7 

### Decaf coffee (FFD111)
fhs3_vr_ffreq_ex01_id2$FFD111_daily  <- fhs3_vr_ffreq_ex01_id2$FFD111/7 

### Tea (FFD113)
fhs3_vr_ffreq_ex01_id2$FFD113_daily  <- fhs3_vr_ffreq_ex01_id2$FFD113/7 


fhs3_melt_vr_ffreq_ex01_id2 <- melt(fhs3_vr_ffreq_ex01_id2,id.vars=c("PID","IDTYPE"),na.rm=T,factorsAsStrings=T)
fhs3_melt_vr_ffreq_ex01_id2$visit <- 1
fhs3_melt_vr_ffreq_ex01_id2$form <- "fhs3_vr_ffreq"

rm(fhs3_vr_ffreq_ex01_id2)


#============================================================#
####  Food frequency questionnaire - FHS Omni 2 - Exam 1  ---#
#============================================================#

fhs3_vr_ffreq_ex01_id72  <- fread('~/Dropbox/BioLINCC files/Framingham third generation/datasets/CSV/VR_FFREQ_EX01_72_0975d_v1.csv')

## Convert servings/week  to  servings/day
### Coffee  (FFD112)

fhs3_vr_ffreq_ex01_id72$FFD112_daily  <- fhs3_vr_ffreq_ex01_id72$FFD112/7 

### Decaf coffee (FFD111)
fhs3_vr_ffreq_ex01_id72$FFD111_daily  <- fhs3_vr_ffreq_ex01_id72$FFD111/7 

### Tea (FFD113)
fhs3_vr_ffreq_ex01_id72$FFD113_daily  <- fhs3_vr_ffreq_ex01_id72$FFD113/7 


fhs3_melt_vr_ffreq_ex01_id72 <- melt(fhs3_vr_ffreq_ex01_id72,id.vars=c("PID","IDTYPE"),na.rm=T,factorsAsStrings=T)
fhs3_melt_vr_ffreq_ex01_id72$visit <- 1
fhs3_melt_vr_ffreq_ex01_id72$form <- "fhs3_vr_ffreq"

rm(fhs3_vr_ffreq_ex01_id72)


#==========================================================#
####  FHS3: Gen 3, Offspring Spouse, Omni 2 - Menarche  ---#
#==========================================================#


fhs3_menarche <- rbind(fhs3_e_exam_ex01_id2[,c("PID","IDTYPE","G3A023")],
                       fhs3_e_exam_ex01_id3[,c("PID","IDTYPE","G3A023")],
                       fhs3_e_exam_ex01_id72[,c("PID","IDTYPE","G3A023")])

fhs3_menarche <- 
  subset(fhs3_menarche,!G3A023 %in% c(88,888)) 

names(fhs3_menarche)[3] <- "age_menarche"

rm(fhs3_e_exam_ex01_id2)
rm(fhs3_e_exam_ex01_id3)
rm(fhs3_e_exam_ex01_id72)


#==========================================================#
####  **** FHS 3/Offspring Spouse/Omni 2 - Exam 2 ****  ####
#==========================================================#

fhs3_e_exam_2011 <- fread('~/Dropbox/BioLINCC files/Framingham third generation/datasets/CSV/E_EXAM_2011_M_0017D_V3.csv')


fhs3_e_exam_2011[!is.na(fhs3_e_exam_2011$G3B0028),22:64] <- NA

fhs3_e_exam_2011$drinks_wk <- 
  fhs3_e_exam_2011$G3B0105+  # Beer
  fhs3_e_exam_2011$G3B0107+  # Wine
  fhs3_e_exam_2011$G3B0109   # Liquor

fhs3_e_exam_2011$G3B0032[!between(fhs3_e_exam_2011$G3B0032,1,8)] <- NA
fhs3_e_exam_2011$G3B0033[!between(fhs3_e_exam_2011$G3B0033,1,3)] <- NA
fhs3_e_exam_2011$G3B0081[!between(fhs3_e_exam_2011$G3B0081,17,62)] <- NA
fhs3_e_exam_2011$G3B0085[!between(fhs3_e_exam_2011$G3B0085,27,58)] <- NA
fhs3_e_exam_2011$G3B0089[!between(fhs3_e_exam_2011$G3B0089,27,65)] <- NA

fhs3_meno_ex02 <-
  fhs3_e_exam_2011[is.na(fhs3_e_exam_2011$G3B0028),
                   c("PID",
                     "IDTYPE",
                     "G3B0073",
                     "G3B0081")]


fhs3_e_exam_2011$years_menses[!fhs3_e_exam_2011$STOPAGE==0&!is.na(fhs3_e_exam_2011$STOPAGE)] <- 
  fhs3_e_exam_2011$STOPAGE[!fhs3_e_exam_2011$STOPAGE==0&!is.na(fhs3_e_exam_2011$STOPAGE)] - 
  fhs3_e_exam_2011$G3A023[!fhs3_e_exam_2011$STOPAGE==00&!is.na(fhs3_e_exam_2011$STOPAGE)]


fhs3_e_exam_2011$fhs_pai <- 
  calc_framingham_pai(fhs3_e_exam_2011,
                      slp_hrs="G3B0703",
                      sed_hrs="G3B0704",
                      slgt_hrs="G3B0705",
                      mod_hrs="G3B0706",
                      hvy_hrs="G3B0707")


fhs3_melt_e_exam_2011 <- melt(fhs3_e_exam_2011,id.vars=c("PID","IDTYPE"),na.rm=T,factorsAsStrings = T)
fhs3_melt_e_exam_2011$visit <- 2
fhs3_melt_e_exam_2011$form <- "fhs3_e_exam_2011"

rm(fhs3_e_exam_2011)


#======================================#
####  CAC, Framingham 3, Exam 2     ---#
#======================================#

fhs3_cac_ex02 <- fread('~/Dropbox/BioLINCC files/Framingham third generation/datasets/CSV/t_ctthrcac_2011_m_0682d.csv')
fhs3_melt_cac_ex02 <-  melt(fhs3_cac_ex02,id.vars=c("PID","IDTYPE"),na.rm=T,factorsAsStrings = T)
fhs3_melt_cac_ex02$visit <- 2
fhs3_melt_cac_ex02$form <- "fhs3_cac_ex02"

rm(fhs3_cac_ex02)


#======================================#
####  PASE, Framingham 3, Exam 2     ---#
#======================================#

# Physical activity score for the elderly

fhs3_pase_ex02 <- 
  fread('~/Dropbox/BioLINCC files/Framingham third generation/datasets/CSV/vr_pase_ex02_3_0642d.csv')
fhs3_melt_pase_ex02 <-  melt(fhs3_pase_ex02,id.vars=c("PID","IDTYPE"),na.rm=T,factorsAsStrings = T)
fhs3_melt_pase_ex02$visit <- 2
fhs3_melt_pase_ex02$form <- "fhs3_pase_ex02"

rm(fhs3_pase_ex02)


#======================================================================#
####  Actigraphy, Framingham 3/New Offspring Spouse/Omni 2, Exam 2  ---#
#======================================================================#

act_fields <-
  c("zmin_vdt",
    "zmin_vdtd",
    "zmin_vdtm",
    "zmin_vdtwd",
    "zmin_vdtwdd",
    "zmin_vdtwdm",
    "zmin_vdtwe",
    "zmin_vdtwed",
    "zmin_vdtwedm",
    "wstep_vdt",
    "wstep_vdtd",
    "wstep_vdtm",
    "wstep_vdtwd",
    "wstep_vdtwdd",
    "wstep_vdtwdm",
    "wstep_vdtwe",
    "wstep_vdtwed",
    "wstep_vdtwedm",
    "zb_vdt",
    "zb_vdtd",
    "zb_vdtmin",
    "zb_vdtwd",
    "zb_vdtwdmin",
    "zb_vdtwe",
    "zb_vdtwed",
    "zb_vdtwedd",
    "zb_vdtwemin",
    "vigstepvdt",
    "vigstepvdtd",
    "vigstepvdtwd",
    "vigstepvdtwdd",
    "vigstepvdtwe",
    "vigstepvdtwed",
    "vigminvdt",
    "vigminvdtd",
    "vigminvdtwd",
    "vigminvdtwdd",
    "vigminvdtwe",
    "vigminvdtwed",
    "vigcntvdt",
    "vigcntvdtd",
    "vigcntvdtwd",
    "vigcntvdtwdd",
    "vigcntvdtwe",
    "vigcntvdtwed",
    "sedcntvdt",
    "sedcntvdtd",
    "sedcntvdtwd",
    "sedcntvdtwdd",
    "sedcntvdtwe",
    "sedcntvdtwed",
    "sedminvdt",
    "sedminvdtd",
    "sedminvdtwd",
    "sedminvdtwdd",
    "sedminvdtwe",
    "sedminvdtwed",
    "sedminwed",
    "bgt_litcntvdt",
    "bgt_litcntvdtd",
    "bgt_litcntvdtwd",
    "bgt_litcntvdtwdd",
    "bgt_litcntvdtwe",
    "bgt_litcntvdtwed",
    "bgt_litminvdt",
    "bgt_litminvdtd",
    "bgt_litminvdtwd",
    "bgt_litminvdtwdd",
    "bgt_litminvdtwe",
    "bgt_litminvdtwed",
    "bgt_litstepvdt",
    "bgt_litstepvdtd",
    "bgt_litstepvdtwd",
    "bgt_litstepvdtwdd",
    "bgt_litstepvdtwe",
    "bgt_litstepvdtwed",
    "bgt_modcntvdt",
    "bgt_modcntvdtd",
    "bgt_modcntvdtwd",
    "bgt_modcntvdtwdd",
    "bgt_modcntvdtwe",
    "bgt_modcntvdtwed",
    "bgt_modminvdt",
    "bgt_modminvdtd",
    "bgt_modminvdtwd",
    "bgt_modminvdtwdd",
    "bgt_modminvdtwe",
    "bgt_modminvdtwed",
    "bgt_modstepvdt",
    "bgt_modstepvdtd",
    "bgt_modstepvdtwd",
    "bgt_modstepvdtwdd",
    "bgt_modstepvdtwe",
    "bgt_modstepvdtwed",
    "bgt_mvpcntvdt",
    "bgt_mvpcntvdtd",
    "bgt_mvpcntvdtwd",
    "bgt_mvpcntvdtwdd",
    "bgt_mvpcntvdtwe",
    "bgt_mvpcntvdtwed",
    "bgt_mvpminvdt",
    "bgt_mvpminvdtd",
    "bgt_mvpminvdtwd",
    "bgt_mvpminvdtwdd",
    "bgt_mvpminvdtwe",
    "bgt_mvpminvdtwed",
    "bgt_mvpstepvdt",
    "bgt_mvpstepvdtd",
    "bgt_mvpstepvdtwd",
    "bgt_mvpstepvdtwdd",
    "bgt_mvpstepvdtwe",
    "bgt_mvpstepvdtwed",
    "bgt_sedcntvdt",
    "bgt_sedcntvdtd",
    "bgt_sedcntvdtwd",
    "bgt_sedcntvdtwdd",
    "bgt_sedcntvdtwe",
    "bgt_sedcntvdtwed",
    "bgt_sedminvdt",
    "bgt_sedminvdtd",
    "bgt_sedminvdtwd",
    "bgt_sedminvdtwdd",
    "bgt_sedminvdtwe",
    "bgt_sedminvdtwed",
    "bgt_sedstepvdt",
    "bgt_sedstepvdtd",
    "bgt_sedstepvdtwd",
    "bgt_sedstepvdtwdd",
    "bgt_sedstepvdtwe",
    "bgt_sedstepvdtwed",
    "bgt_vigcntvdt",
    "bgt_vigcntvdtd",
    "bgt_vigcntvdtwd",
    "bgt_vigcntvdtwdd",
    "bgt_vigcntvdtwe",
    "bgt_vigcntvdtwed",
    "bgt_vigminvdt",
    "bgt_vigminvdtd",
    "bgt_vigminvdtwd",
    "bgt_vigminvdtwdd",
    "bgt_vigminvdtwe",
    "bgt_vigminvdtwed",
    "bgt_vigstepvdt",
    "bgt_vigstepvdtd",
    "bgt_vigstepvdtwd",
    "bgt_vigstepvdtwdd",
    "bgt_vigstepvdtwe",
    "bgt_vigstepvdtwed",
    "litcntvdt",
    "litcntvdtd",
    "litcntvdtwd",
    "litcntvdtwdd",
    "litcntvdtwe",
    "litcntvdtwed",
    "litminvdt",
    "litminvdtd",
    "litminvdtwd",
    "litminvdtwdd",
    "litminvdtwe",
    "litminvdtwed",
    "litstepvdt",
    "litstepvdtd",
    "litstepvdtwd",
    "litstepvdtwdd",
    "litstepvdtwe",
    "litstepvdtwed",
    "modcntvdt",
    "modcntvdtd",
    "modcntvdtwd",
    "modcntvdtwdd",
    "modcntvdtwe",
    "modcntvdtwed",
    "modminvdt",
    "modminvdtd",
    "modminvdtwd",
    "modminvdtwdd",
    "modminvdtwe",
    "modminvdtwed",
    "modstepvdt",
    "modstepvdtd",
    "modstepvdtwd",
    "modstepvdtwdd",
    "modstepvdtwe",
    "modstepvdtwed",
    "motcntvdt",
    "motcntvdtd",
    "motcntvdtwd",
    "motcntvdtwdd",
    "motcntvdtwe",
    "motcntvdtwed",
    "motminvdt",
    "motminvdtd",
    "motminvdtwd",
    "motminvdtwdd",
    "motminvdtwe",
    "motminvdtwed",
    "motstepvdt",
    "motstepvdtd",
    "motstepvdtwd",
    "motstepvdtwdd",
    "motstepvdtwe",
    "motstepvdtwed",
    "mvpcntvdt",
    "mvpcntvdtd",
    "mvpcntvdtwd",
    "mvpcntvdtwdd",
    "mvpcntvdtwe",
    "mvpcntvdtwed",
    "mvpminvdt",
    "mvpminvdtd",
    "mvpminvdtwd",
    "mvpminvdtwdd",
    "mvpminvdtwe",
    "mvpminvdtwed",
    "mvpstepvdt",
    "mvpstepvdtd",
    "mvpstepvdtwd",
    "mvpstepvdtwdd",
    "mvpstepvdtwe",
    "mvpstepvdtwed",
    "s_litcntvdt",
    "s_litcntvdtd",
    "s_litcntvdtwd",
    "s_litcntvdtwdd",
    "s_litcntvdtwe",
    "s_litcntvdtwed",
    "s_litminvdt",
    "s_litminvdtd",
    "s_litminvdtwd",
    "s_litminvdtwdd",
    "s_litminvdtwe",
    "s_litminvdtwed",
    "s_litstepvdt",
    "s_litstepvdtd",
    "s_litstepvdtwd",
    "s_litstepvdtwdd",
    "s_litstepvdtwe",
    "s_litstepvdtwed",
    "s_modcntvdt",
    "s_modcntvdtd",
    "s_modcntvdtwd",
    "s_modcntvdtwdd",
    "s_modcntvdtwe",
    "s_modcntvdtwed",
    "s_modminvdt",
    "s_modminvdtd",
    "s_modminvdtwd",
    "s_modminvdtwdd",
    "s_modminvdtwe",
    "s_modminvdtwed",
    "s_modstepvdt",
    "s_modstepvdtd",
    "s_modstepvdtwd",
    "s_modstepvdtwdd",
    "s_modstepvdtwe",
    "s_modstepvdtwed",
    "s_mvpcntvdt",
    "s_mvpcntvdtd",
    "s_mvpcntvdtwd",
    "s_mvpcntvdtwdd",
    "s_mvpcntvdtwe",
    "s_mvpcntvdtwed",
    "s_mvpminvdt",
    "s_mvpminvdtd",
    "s_mvpminvdtwd",
    "s_mvpminvdtwdd",
    "s_mvpminvdtwe",
    "s_mvpminvdtwed",
    "s_mvpstepvdt",
    "s_mvpstepvdtd",
    "s_mvpstepvdtwd",
    "s_mvpstepvdtwdd",
    "s_mvpstepvdtwe",
    "s_mvpstepvdtwed",
    "s_sedcntvdt",
    "s_sedcntvdtd",
    "s_sedcntvdtwd",
    "s_sedcntvdtwdd",
    "s_sedcntvdtwe",
    "s_sedcntvdtwed",
    "s_sedminvdt",
    "s_sedminvdtd",
    "s_sedminvdtwd",
    "s_sedminvdtwdd",
    "s_sedminvdtwe",
    "s_sedminvdtwed",
    "s_sedstepvdt",
    "s_sedstepvdtd",
    "s_sedstepvdtwd",
    "s_sedstepvdtwdd",
    "s_sedstepvdtwe",
    "s_sedstepvdtwed",
    "s_vigcntvdt",
    "s_vigcntvdtd",
    "s_vigcntvdtwd",
    "s_vigcntvdtwdd",
    "s_vigcntvdtwe",
    "s_vigcntvdtwed",
    "s_vigminvdt",
    "s_vigminvdtd",
    "s_vigminvdtwd",
    "s_vigminvdtwdd",
    "s_vigminvdtwe",
    "s_vigminvdtwed",
    "s_vigstepvdt",
    "s_vigstepvdtd",
    "s_vigstepvdtwd",
    "s_vigstepvdtwdd",
    "s_vigstepvdtwe",
    "s_vigstepvdtwed",
    "zstep_vdt",
    "zstep_vdtd",
    "zstep_vdtm",
    "zstep_vdtwd",
    "zstep_vdtwdd",
    "zstep_vdtwdm",
    "zstep_vdtwe",
    "zstep_vdtwed",
    "zstep_vdtwedm",
    "zcnt_vdt",
    "zcnt_vdtd",
    "zcnt_vdtm",
    "zcnt_vdtwd",
    "zcnt_vdtwdd",
    "zcnt_vdtwdm",
    "zcnt_vdtwe",
    "zcnt_vdtwed",
    "zcnt_vdtwedm",
    "zb_vdt",
    "zb_vdtd",
    "zb_vdtmin",
    "zb_vdtwd",
    "zb_vdtwdmin",
    "zb_vdtwe",
    "zb_vdtwed",
    "zb_vdtwedd",
    "zb_vdtwemin",
    "wb_vdt",
    "wb_vdtd",
    "wb_vdtmin",
    "wb_vdtwd",
    "wb_vdtwdmin",
    "wb_vdtwe",
    "wb_vdtwed",
    "wb_vdtwedd",
    "wb_vdtwemin",
    "wcnt_vdt",
    "wcnt_vdtd",
    "wcnt_vdtm",
    "wcnt_vdtwd",
    "wcnt_vdtwdd",
    "wcnt_vdtwdm",
    "wcnt_vdtwe",
    "wcnt_vdtwed",
    "wcnt_vdtwedm",
    "wmin_vdt",
    "wmin_vdtd",
    "wmin_vdtm",
    "wmin_vdtwd",
    "wmin_vdtwdd",
    "wmin_vdtwdm",
    "wmin_vdtwe",
    "wmin_vdtwed",
    "wmin_vdtwedm",
    "wstep_vdt",
    "wstep_vdtd",
    "wstep_vdtm",
    "wstep_vdtwd",
    "wstep_vdtwdd",
    "wstep_vdtwdm",
    "wstep_vdtwe",
    "wstep_vdtwed",
    "wstep_vdtwedm")

act_fields <- toupper(act_fields)

fhs3_actigraphy_ex02 <- 
  as.data.table(
    read_sas('~/Dropbox/BioLINCC files/Framingham third generation/datasets/t_physactf_ex02_3b_0914d.sas7bdat',
             skip=1))

fhs3_melt_actigraphy_ex02 <-  melt(fhs3_actigraphy_ex02,id.vars=c("PID","IDTYPE"),na.rm=T,factorsAsStrings = T)
fhs3_melt_actigraphy_ex02$visit <- 2
fhs3_melt_actigraphy_ex02$form <- "fhs3_actigraphy_ex02"

fhs3_melt_actigraphy_ex02 <- 
  subset(fhs3_melt_actigraphy_ex02,variable %in% act_fields)

rm(fhs3_actigraphy_ex02)


#==============================================================================#
####  Vascular tonometry, Framingham 3/New Offspring Spouse/Omni 2, Exam 2  ---#
#==============================================================================#

fhs3_tonometry_ex02 <- fread('~/Dropbox/BioLINCC files/Framingham third generation/datasets/CSV/t_tonla_ex02_3b_1235d.csv')
fhs3_melt_tonometry_ex02 <-  melt(fhs3_tonometry_ex02,id.vars=c("PID","IDTYPE"),na.rm=T,factorsAsStrings = T)
fhs3_melt_tonometry_ex02$visit <- 2
fhs3_melt_tonometry_ex02$form <- "fhs3_tonometry_ex02"

rm(fhs3_tonometry_ex02)

#=============================================#
####  Whole body DXA Framingham 3, Exam 2  ---#
#=============================================#

fhs3_wbdxa_ex02_pt1 <- fread('~/Dropbox/BioLINCC files/Framingham third generation/datasets/CSV/t_wbdxa_2010_3_0585d_v1.csv')
fhs3_wbdxa_ex02_pt2 <- fread('~/Dropbox/BioLINCC files/Framingham third generation/datasets/CSV/t_wbdxa_2011_3_0624d.csv')
fhs3_wbdxa_ex02 <-
  rbind(fhs3_wbdxa_ex02_pt1,
        fhs3_wbdxa_ex02_pt2)
fhs3_melt_wbdxa_ex02 <-  melt(fhs3_wbdxa_ex02,id.vars=c("PID","IDTYPE"),na.rm=T,factorsAsStrings = T)
fhs3_melt_wbdxa_ex02$visit <- 2
fhs3_melt_wbdxa_ex02$form <- "fhs3_wbdxa_ex02"

rm(fhs3_wbdxa_ex02)


#===============================================================#
####  FHS labs, Framingham 3/Offspring Spouse/Omni 2,Exam 2  ---#
#===============================================================#

fhs3_fhslab_2011 <- fread('~/Dropbox/BioLINCC files/Framingham third generation/datasets/CSV/L_FHSLAB_2011_M_0656D_V1.csv')
fhs3_fhslab_2011 <- merge(fhs3_fhslab_2011,
                          fhs3_wkthru[,c("PID","IDTYPE","SEX","AGE2")],
                          by=c("PID","IDTYPE"),
                          all.x=T)
labcols <- c(names(fhs3_fhslab_2011),"gfr_mdrd")
fhs3_fhslab_2011 <- merge(fhs3_fhslab_2011,
                          fhs3_race,
                          by=c("PID","IDTYPE"),
                          all.x=T)

fhs3_fhslab_2011$gfr_mdrd <- calc_MDRD4(dat=fhs3_fhslab_2011,
                                        cr="CREAT",
                                        age="AGE2",
                                        sex="SEX",
                                        race="Black",
                                        black=1)

fhs3_melt_fhslab_2011 <- melt(fhs3_fhslab_2011[,..labcols],
                              id.vars=c("PID","IDTYPE"),
                              na.rm=T,
                              factorsAsStrings = T)
fhs3_melt_fhslab_2011$visit <- 2
fhs3_melt_fhslab_2011$form <- "fhs3_fhslab_2011"

rm(fhs3_fhslab_2011)


#=========================================================================#
####  PFT Diffusion test - FHS Gen 3/Offspring Spouse/Omni 2 - Exam 2  ---#
#=========================================================================#


fhs3_t_pftdiff_2011  <- fread('~/Dropbox/BioLINCC files/Framingham third generation/datasets/CSV/T_PFTDIFF_2011_M_0648D_V2.csv')
fhs3_t_pftdiff_2011$DLCO_VA_mean <- rowMeans(fhs3_t_pftdiff_2011[,c('DVA1_2','DVA2_2','DVA3_2')],na.rm=T)

fhs3_melt_t_pftdiff_2011 <- melt(fhs3_t_pftdiff_2011,id.vars=c("PID","IDTYPE"),na.rm=T,factorsAsStrings=T)
fhs3_melt_t_pftdiff_2011$visit <- 1
fhs3_melt_t_pftdiff_2011$form <- "fhs3_pftdiff_2011"

rm(fhs3_t_pftdiff_2011)


#======================================================================#
####  Dietary guidelines adherence index 2010 - FHS Gen 3 - Exam 2  ---#
#======================================================================#

fhs3_vr_dgai2010_ex02_id3  <- fread('~/Dropbox/BioLINCC files/Framingham third generation/datasets/CSV/VR_DGAI2010_EX02_3_0996D.csv')
fhs3_melt_vr_dgai2010_ex02_id3 <- melt(fhs3_vr_dgai2010_ex02_id3,id.vars=c("PID","IDTYPE"),na.rm=T,factorsAsStrings=T)
fhs3_melt_vr_dgai2010_ex02_id3$visit <- 2
fhs3_melt_vr_dgai2010_ex02_id3$form <- "fhs3_dgai"

rm(fhs3_vr_dgai2010_ex02_id3)


#===========================================================#
####  Food frequency questionnaire - FHS Gen 3 - Exam 2  ---#
#===========================================================#

fhs3_vr_ffreq_ex02_id3  <- fread('~/Dropbox/BioLINCC files/Framingham third generation/datasets/CSV/VR_FFREQ_EX02_3b_1345D.csv')
fhs3_melt_vr_ffreq_ex02_id3 <- melt(fhs3_vr_ffreq_ex02_id3,id.vars=c("PID","IDTYPE"),na.rm=T,factorsAsStrings=T)
fhs3_melt_vr_ffreq_ex02_id3$visit <- 2
fhs3_melt_vr_ffreq_ex02_id3$form <- "fhs3_vr_ffreq"

rm(fhs3_vr_ffreq_ex02_id3)


#=========================================#
####  CT Lung, Framingham 3, Exam 2    ---#
#=========================================#

fhs3_ctlung_ex02 <- fread('~/Dropbox/BioLINCC files/Framingham third generation/datasets/CSV/t_ctlung_2011_m_0829d.csv')
fhs3_melt_ctlung_ex02 <-  melt(fhs3_ctlung_ex02,id.vars=c("PID","IDTYPE"),na.rm=T,factorsAsStrings = T)
fhs3_melt_ctlung_ex02$visit <- 2
fhs3_melt_ctlung_ex02$form <- "fhs3_ctlung_ex02"

rm(fhs3_ctlung_ex02)


#================================================#
####  CT abdominal fat, Framingham 3, Exam 2  ---#
#================================================#

fhs3_ctabdom_ex02 <- fread('~/Dropbox/BioLINCC files/Framingham third generation/datasets/CSV/t_ctabfat_2011_m_0669d.csv')
fhs3_melt_ctabdom_ex02 <-  melt(fhs3_ctabdom_ex02,id.vars=c("PID","IDTYPE"),na.rm=T,factorsAsStrings = T)
fhs3_melt_ctabdom_ex02$visit <- 2
fhs3_melt_ctabdom_ex02$form <- "fhs3_ctabdom_ex02"

rm(fhs3_ctabdom_ex02)


#==========================================================#
####  **** FHS 3/Offspring Spouse/Omni 2 - Exam 3 ****  ####
#==========================================================#

fhs3_e_exam_ex03 <- fread('~/Dropbox/BioLINCC files/Framingham third generation/datasets/CSV/e_exam_ex03_3b_1069d.csv')

fhs3_e_exam_ex03$G3C0234[!between(fhs3_e_exam_ex03$G3C0234,15,63)] <- NA
fhs3_e_exam_ex03$G3C0238[!between(fhs3_e_exam_ex03$G3C0238,30,87)] <- NA
fhs3_e_exam_ex03$G3C0242[!between(fhs3_e_exam_ex03$G3C0242,22,87)] <- NA

fhs3_e_exam_ex03$drinks_wk <- 
  fhs3_e_exam_ex03$G3C0264+  # Beer
  fhs3_e_exam_ex03$G3C0268+  # Wine
  fhs3_e_exam_ex03$G3C0272   # Liquor

fhs3_e_exam_ex03$fhs_pai <- 
  calc_framingham_pai(fhs3_e_exam_ex03,
                      slp_hrs="G3C0748",
                      sed_hrs="G3C0749",
                      slgt_hrs="G3C0750",
                      mod_hrs="G3C0751",
                      hvy_hrs="G3C0752")


fhs3_melt_e_exam_ex03 <- melt(fhs3_e_exam_ex03,id.vars=c("PID","IDTYPE"),na.rm=T,factorsAsStrings = T)
fhs3_melt_e_exam_ex03$visit <- 3


fhs3_melt_e_exam_ex03$form <- "fhs3_e_ex03"


#=========================================================#
####  Menopause/HRT - FHS 3/Offspring Spouse/Omni 2   ----#
#=========================================================#


fhs3_vr_meno_ex02_id2  <- fread('~/Dropbox/BioLINCC files/Framingham third generation/datasets/CSV/VR_MENO_EX02_2_0719D.csv')
fhs3_vr_meno_ex02_id3  <- fread('~/Dropbox/BioLINCC files/Framingham third generation/datasets/CSV/VR_MENO_EX02_3_0653D_V1.csv')
fhs3_vr_meno_ex02_id72  <- fread('~/Dropbox/BioLINCC files/Framingham third generation/datasets/CSV/VR_MENO_EX02_72_0720D.csv')

fhs3_vr_meno_ex02 <- 
  rbind(fhs3_vr_meno_ex02_id2,
        fhs3_vr_meno_ex02_id3,
        fhs3_vr_meno_ex02_id72)

fhs3_meno_ex03 <- 
  fhs3_e_exam_ex03[
    fhs3_e_exam_ex03$G3C0199 %in% c(0,1),
    c("PID",
      "IDTYPE",
      "G3C0228",
      "G3C0234")]

fhs3_menocalcs_ex01 <- 
  merge(fhs3_meno_ex01,
        fhs3_meno_ex02,
        by=c("PID","IDTYPE"),
        all.x=T)

fhs3_menocalcs_ex03 <- 
  merge(fhs3_menocalcs_ex01,
        fhs3_meno_ex03,
        by=c("PID","IDTYPE"),
        all.x=T)

fhs3_menocalcs_ex03 <-
  merge(fhs3_menocalcs_ex03,
        fhs3_vr_meno_ex02[,c("PID","IDTYPE","STOPAGE")],
        by=c("PID","IDTYPE"),
        all.x=T)

fhs3_menocalcs_ex03$STOPAGE[fhs3_menocalcs_ex03$STOPAGE==0] <- NA

fhs3_menocalcs_ex03$age_menopause <- 
  fhs3_menocalcs_ex03$STOPAGE

fhs3_menocalcs_ex03$age_menopause[is.na(fhs3_menocalcs_ex03$age_menopause)] <- 
  fhs3_menocalcs_ex03$G3C0234[is.na(fhs3_menocalcs_ex03$age_menopause)]

fhs3_menocalcs_ex03$age_menopause[is.na(fhs3_menocalcs_ex03$age_menopause)] <- 
  fhs3_menocalcs_ex03$G3B0081[is.na(fhs3_menocalcs_ex03$age_menopause)]

fhs3_menocalcs_ex03$age_menopause[is.na(fhs3_menocalcs_ex03$age_menopause)] <- 
  fhs3_menocalcs_ex03$G3A051[is.na(fhs3_menocalcs_ex03$age_menopause)]

fhs3_menocalcs_ex03$age_menarche <- fhs3_menocalcs_ex03$G3A023

fhs3_menocalcs_ex03$years_menses <- 
  fhs3_menocalcs_ex03$age_menopause-fhs3_menocalcs_ex03$age_menarche

fhs3_melt_menocalcs_ex03 <-
  melt(fhs3_menocalcs_ex03[,c("PID",
                              "IDTYPE",
                              "age_menarche",
                              "age_menopause",
                              "years_menses")],
       id.vars=c("PID",
                 "IDTYPE"),
       na.rm=T)

fhs3_melt_menocalcs_ex03$form <- "fhs3_menocalcs_ex03"
fhs3_melt_menocalcs_ex03$visit <- 3


#======================================================================#
####  Actigraphy, Framingham 3, New Offspring Spouse/Omni 2 Exam 3  ---#
#======================================================================#


fhs3_actigraphy_ex03 <- 
  as.data.table(
    read_sas('~/Dropbox/BioLINCC files/Framingham third generation/datasets/t_physactf_ex03_3b_1007d.sas7bdat',
             skip=1))

fhs3_melt_actigraphy_ex03 <-  melt(fhs3_actigraphy_ex03,id.vars=c("PID","IDTYPE"),na.rm=T,factorsAsStrings = T)
fhs3_melt_actigraphy_ex03$visit <- 3
fhs3_melt_actigraphy_ex03$form <- "fhs3_actigraphy_ex03"

fhs3_melt_actigraphy_ex03 <- 
  subset(fhs3_melt_actigraphy_ex03,variable %in% act_fields)

rm(fhs3_actigraphy_ex03)


#=====================================================================#
####  Validated education Framingham 3/New Offspring Spouse/Omni 2 ---#
#=====================================================================#

fhs3_melt_vr_educ <- fread('~/Dropbox/BioLINCC files/Framingham third generation/datasets/CSV/vr_educ_2018_a_1307d.csv')
fhs3_melt_vr_educ <- subset(fhs3_melt_vr_educ,!EDUCATION=="")
fhs3_melt_vr_educ$visit <- 1
fhs3_melt_vr_educ$form <- "fhs3_melt_vr_educ"
fhs3_melt_vr_educ$variable <- "EDUCATION"
fhs3_melt_vr_educ$value <- fhs3_melt_vr_educ$EDUCATION

#==========================================================================#
####  Validated smoking status Framingham 3/New Offspring Spouse/Omni 2 ---#
#==========================================================================#

fhs3_vr_smoke <- fread('~/Dropbox/BioLINCC files/Framingham third generation/datasets/CSV/vr_smokst_2019_m_1325d.csv')
fhs3_melt_vr_smoke <-  melt(fhs3_vr_smoke,id.vars=c("PID","IDTYPE"),na.rm=T,factorsAsStrings = T)
fhs3_melt_vr_smoke$visit <- substr(fhs3_melt_vr_smoke$variable,7,7)
fhs3_melt_vr_smoke$variable <- "smoke_status_vr"
fhs3_melt_vr_smoke$form <- "fhs3_vr_smokst"

rm(fhs3_vr_smoke)


#===============================================================================#
####  FHS Labs, Framingham 3, Exam 3/Offspring Spouse Exam 3/Omni 2, Exam 3  ---#
#===============================================================================#

fhs3_fhslab_ex03 <- fread('~/Dropbox/BioLINCC files/Framingham third generation/datasets/CSV/L_FHSLAB_EX03_3B_1170D.csv')
fhs3_fhslab_ex03 <- merge(fhs3_fhslab_ex03,
                          fhs3_wkthru[,c("PID","IDTYPE","SEX","AGE3")],
                          by=c("PID","IDTYPE"),
                          all.x=T)
labcols <- c(names(fhs3_fhslab_ex03),"gfr_mdrd")
fhs3_fhslab_ex03 <- merge(fhs3_fhslab_ex03,
                          fhs3_race,
                          by=c("PID","IDTYPE"),
                          all.x=T)

fhs3_fhslab_ex03$gfr_mdrd <- calc_MDRD4(dat=fhs3_fhslab_ex03,
                                        cr="CREATININE",
                                        age="AGE3",
                                        sex="SEX",
                                        race="Black",
                                        black=1)

fhs3_melt_fhslab_ex03 <- melt(fhs3_fhslab_ex03[,..labcols],
                              id.vars=c("PID","IDTYPE"),
                              na.rm=T,
                              factorsAsStrings = T)

fhs3_melt_fhslab_ex03$visit <- 3
fhs3_melt_fhslab_ex03$form <- "fhs3_fhslab_ex03"
rm(fhs3_fhslab_ex03)


#================================================#
####      Wkthru (reference) - FHS Gen 3      ####
#================================================#

fhs3_wkthru <-
  merge(fhs3_wkthru,
        fhs3_race[,c("PID","IDTYPE","Black")],
        by=c("PID","IDTYPE"),
        all.x=T)

fhs3_wkthru$WTCHG_PCT2 <-
  round((fhs3_wkthru$WGT2-fhs3_wkthru$WGT1)/fhs3_wkthru$WGT1 * 100,1)

fhs3_wkthru$WTCHG_PCT3 <-
  round((fhs3_wkthru$WGT3-fhs3_wkthru$WGT2)/fhs3_wkthru$WGT2 * 100,1)

fhs3_wkthru$WHR2 <- 
  fhs3_wkthru$WAIST2/fhs3_wkthru$HIP2

fhs3_wkthru$WHR3 <- 
  fhs3_wkthru$WAIST3/fhs3_wkthru$HIP3


fhs3_wkthru$gfr_mdrd1 <- calc_MDRD4(dat=fhs3_wkthru,
                                    cr="CREAT1",
                                    age="AGE1",
                                    sex="SEX",
                                    race="Black",
                                    black=1)


fhs3_melt_wkthru <- melt(fhs3_wkthru,
                         id.vars=c("PID","IDTYPE","SEX"),
                         na.rm=T,
                         factorsAsStrings=T)

fhs3_melt_wkthru$variable <-  as.character(fhs3_melt_wkthru$variable)
fhs3_melt_wkthru$visit  <- substring(fhs3_melt_wkthru$variable,
                                     nchar(fhs3_melt_wkthru$variable))
fhs3_melt_wkthru$variable <- substring(fhs3_melt_wkthru$variable,
                                       1, nchar(fhs3_melt_wkthru$variable)-1)

fhs3_melt_wkthru$value[fhs3_melt_wkthru$variable=="WAIST"] <- 
  fhs3_melt_wkthru$value[fhs3_melt_wkthru$variable=="WAIST"] * 2.54
fhs3_melt_wkthru_kg <- subset(fhs3_melt_wkthru,variable=="WGT")
fhs3_melt_wkthru_kg$value <- fhs3_melt_wkthru_kg$value/2.20462
fhs3_melt_wkthru_kg$variable <- "WGT_KG"
fhs3_melt_wkthru <- rbind(fhs3_melt_wkthru,fhs3_melt_wkthru_kg)

fhs3_melt_wkthru$value[fhs3_melt_wkthru$variable=="WAIST"] <- 
  fhs3_melt_wkthru$value[fhs3_melt_wkthru$variable=="WAIST"] * 2.54


fhs3_melt_wkthru$form <- "fhs3_wkthru"


#===================================================================#
####  Assemble FHS Gen 3/Offspring Spouse/Omni 2 cohorts         ####
#===================================================================#


fhs3_melt_all <- rbind(
  fhs3_melt_bnp3[,c("PID","IDTYPE","visit","variable","value","form")],
  fhs3_melt_inflamm[,c("PID","IDTYPE","visit","variable","value","form")],
  fhs3_melt_renaldo3[,c("PID","IDTYPE","visit","variable","value","form")],
  fhs3_melt_fhslab_2011[,c("PID","IDTYPE","visit","variable","value","form")],
  fhs3_melt_fhslab_ex01[,c("PID","IDTYPE","visit","variable","value","form")],
  fhs3_melt_fhslab_ex03[,c("PID","IDTYPE","visit","variable","value","form")],
  fhs3_melt_mhorm_ex01[,c("PID","IDTYPE","visit","variable","value","form")],
  fhs3_melt_e_exam_2011[,c("PID","IDTYPE","visit","variable","value","form")],
  fhs3_melt_e_exam_ex01_id2[,c("PID","IDTYPE","visit","variable","value","form")],
  fhs3_melt_e_exam_ex01_id3[,c("PID","IDTYPE","visit","variable","value","form")],
  fhs3_melt_e_exam_ex01_id72[,c("PID","IDTYPE","visit","variable","value","form")],
  fhs3_melt_e_exam_ex03[,c("PID","IDTYPE","visit","variable","value","form")],
  fhs3_melt_estr_ex01[,c("PID","IDTYPE","visit","variable","value","form")],
  fhs3_melt_t_echo_2008[,c("PID","IDTYPE","visit","variable","value","form")],
  fhs3_melt_t_doppvasc_dd_2008[,c("PID","IDTYPE","visit","variable","value","form")],
  fhs3_melt_t_echo_ex01_id3[,c("PID","IDTYPE","visit","variable","value","form")],
  fhs3_melt_t_pft_ex01_id3[,c("PID","IDTYPE","visit","variable","value","form")],
  fhs3_melt_t_pftdiff_2005[,c("PID","IDTYPE","visit","variable","value","form")],
  fhs3_melt_t_pftdiff_2011[,c("PID","IDTYPE","visit","variable","value","form")],
  fhs3_melt_t_pftdiff_ex01_id3[,c("PID","IDTYPE","visit","variable","value","form")],
  fhs3_melt_vr_dgai2010_ex01_id3[,c("PID","IDTYPE","visit","variable","value","form")],
  fhs3_melt_vr_dgai2010_ex02_id3[,c("PID","IDTYPE","visit","variable","value","form")],
  fhs3_melt_vr_ffreq_ex01_id3[,c("PID","IDTYPE","visit","variable","value","form")],
  fhs3_melt_vr_ffreq_ex02_id3[,c("PID","IDTYPE","visit","variable","value","form")],
  fhs3_melt_vr_ffreq_ex01_id2[,c("PID","IDTYPE","visit","variable","value","form")],
  fhs3_melt_vr_ffreq_ex01_id72[,c("PID","IDTYPE","visit","variable","value","form")],
  fhs3_melt_wkthru[,c("PID","IDTYPE","visit","variable","value","form")],
  fhs3_melt_sex[,c("PID","IDTYPE","visit","variable","value","form")],
  fhs3_melt_tni_ex01[,c("PID","IDTYPE","visit","variable","value","form")],
  fhs3_melt_telomere_ex01[,c("PID","IDTYPE","visit","variable","value","form")],
  fhs3_melt_sflt3_ex01[,c("PID","IDTYPE","visit","variable","value","form")],
  fhs3_melt_ctabdom_ex01[,c("PID","IDTYPE","visit","variable","value","form")],
  fhs3_melt_ctlung_ex01[,c("PID","IDTYPE","visit","variable","value","form")],
  fhs3_melt_ctlung_ex02[,c("PID","IDTYPE","visit","variable","value","form")],
  fhs3_melt_cac_ex01[,c("PID","IDTYPE","visit","variable","value","form")],
  fhs3_melt_cac_ex02[,c("PID","IDTYPE","visit","variable","value","form")],
  fhs3_melt_tonometry_ex01[,c("PID","IDTYPE","visit","variable","value","form")],
  fhs3_melt_tonometry_ex02[,c("PID","IDTYPE","visit","variable","value","form")],
  fhs3_melt_wbdxa_ex02[,c("PID","IDTYPE","visit","variable","value","form")],
  fhs3_melt_vr_educ[,c("PID","IDTYPE","visit","variable","value","form")],
  fhs3_melt_vr_smoke[,c("PID","IDTYPE","visit","variable","value","form")],
  fhs3_melt_actigraphy_ex02[,c("PID","IDTYPE","visit","variable","value","form")],
  fhs3_melt_actigraphy_ex03[,c("PID","IDTYPE","visit","variable","value","form")],
  fhs3_melt_menocalcs_ex03[,c("PID","IDTYPE","visit","variable","value","form")],
  fhs3_melt_pase_ex02[,c("PID","IDTYPE","visit","variable","value","form")]
)

fhs3_breastfeed <-
  dcast(
    subset(fhs3_melt_all, variable %in% c(
      "G3A035",
      "G3B0033",
      "G3C0205",
      "G3B0041",
      "G3B0042",
      "G3B0046",
      "G3B0047",
      "G3B0051",
      "G3B0052",
      "G3B0056",
      "G3B0057",
      "G3B0061",
      "G3B0062",
      "G3B0066",
      "G3B0067",
      "G3B0071",
      "G3B0072",
      "G3C0209",
      "G3C0210",
      "G3C0214",
      "G3C0215",
      "G3C0219",
      "G3C0220",
      "G3C0224",
      "G3C0225")),
    PID+IDTYPE~variable)

fhs3_breastfeed <-
  subset(fhs3_breastfeed,
         !(is.na(fhs3_breastfeed$G3A035)&
             is.na(fhs3_breastfeed$G3B0033)&
             is.na(fhs3_breastfeed$G3C0205)))

fhs3_breastfeed <- subset(fhs3_breastfeed,!G3A035=="88")
fhs3_breastfeed <- subset(fhs3_breastfeed,!G3B0033=="88")
fhs3_breastfeed <- subset(fhs3_breastfeed,!G3C0205=="88")

fhs3_breastfeed$num_breastfeed <- 
  apply(fhs3_breastfeed[,c("G3B0041","G3B0046","G3B0051","G3B0056","G3B0061","G3C0209","G3C0214")],
        MARGIN=1,
        FUN=function(x) sum(as.numeric(x),na.rm=T))

fhs3_melt_breastfeed <- 
  melt(fhs3_breastfeed,
       id=c("PID","IDTYPE"),
       na.rm=T)

fhs3_melt_breastfeed$visit <- 3
fhs3_melt_breastfeed$form <- "fhs3_breastfeed"

fhs3_melt_all <-
  rbind(fhs3_melt_all[,c("PID","IDTYPE","visit","variable","value","form")],
        fhs3_melt_breastfeed[,c("PID","IDTYPE","visit","variable","value","form")])

fhs3_no_attend <- as.data.table(sqldf("select PID,
      visit, 
      value as att 
      from fhs3_melt_wkthru 
      where variable = 'ATT'"))

fhs3_melt_all <- merge(fhs3_melt_all,fhs3_no_attend,by=c("PID","visit"))

fhs3_melt_all <- subset(fhs3_melt_all,att==1)

fhs3_melt_all$variable <- as.character(fhs3_melt_all$variable)

#===============================================================##
####              Assemble all FHS cohorts                    ####
#===============================================================##

names(fhs_vr_diab)[2] <- "IDTYPE"

fhs_melt_all <- rbind(
  fhs1_melt_all[,c("PID","IDTYPE","visit","form","variable","value")],
  fhs2_melt_all[,c("PID","IDTYPE","visit","form","variable","value")],
  fhs3_melt_all[,c("PID","IDTYPE","visit","form","variable","value")],
  fhs_vr_diab[,c("PID","IDTYPE","visit","form","variable","value")]
)

fhs_melt_all$PID <- as.character(fhs_melt_all$PID)
names(fhs_melt_all)[1:2] <- c("patientid","cohort")

fhs_dates_long$PID <- as.character(fhs_dates_long$PID)
fhs_dates_long$visit <- as.character(fhs_dates_long$visit)

fhs_melt_all <- merge(fhs_melt_all,
                      fhs_dates_long[,c("PID","cohort","visit","visit_yr","visitdays","cohort_name","age_obs")],
                      by.x=c("patientid","cohort","visit"),
                      by.y=c("PID","cohort","visit"),
                      all.x=T)

fhs_melt_all$visit <- as.integer(fhs_melt_all$visit)
fhs_melt_all$visit_yr <- as.integer(fhs_melt_all$visit_yr)

fhs_melt_all$study <- "FHS"

fhs_melt_all$form <- toupper(fhs_melt_all$form)
fhs_melt_all$variable <- toupper(fhs_melt_all$variable)

fhs_melt_all$study_field <- paste(fhs_melt_all$study,
                                  fhs_melt_all$form,
                                  fhs_melt_all$variable,sep="_")


# fhs_melt_all <- subset(fhs_melt_all,study_field %in% hdcp_cohort_fields_used$study_field&study=="FHS")

fhs_melt_all$datapoint <- 
  paste("FHS",row.names(fhs_melt_all),sep="")

fhs_melt_all$patientid <- as.character(fhs_melt_all$patientid)

write_parquet(fhs_melt_all[,..data_fields],
              "~/Dropbox/ADAPT-HF/Master HDCP files/Cohort data/alldata_fhs.parquet")

# dates_long <- fhs_dates_long[,dates_long_fields]

rm(list=ls(pattern="\\bfhs1."))
rm(list=ls(pattern="\\bfhs2."))
rm(list=ls(pattern="\\bfhs3."))


gcs_auth("~/Dropbox/ADAPT-HF/Master HDCP files/harmonization-286013-39f492122f69.json")

gcs_upload(fhs_melt_all[,..data_fields], 
           bucket="master_hdcp_files",
           name="fhs_melt_all.parquet",
           object_function = f)


rm(list=ls(pattern="\\bfhs."))
rm(offspring_30,
   offspring_35,
   omni1_10,
   omni1_5)


gc()


#### ************************ ARIC ************************ #### 

# ....%%%....%%%%%%%%..%%%%..%%%%%%.
# ...%%.%%...%%.....%%..%%..%%....%%
# ..%%...%%..%%.....%%..%%..%%......
# .%%.....%%.%%%%%%%%...%%..%%......
# .%%%%%%%%%.%%...%%....%%..%%......
# .%%.....%%.%%....%%...%%..%%....%%
# .%%.....%%.%%.....%%.%%%%..%%%%%%.


#===============================================================##
####                      ARIC dates                          ####
#===============================================================##

aric_derive1 <- fread('~/Dropbox/BioLINCC files/ARIC/Main_Study/v1/csv/derive13.csv',na.strings=c("NA","","NULL"))
aric_derive2 <- fread('~/Dropbox/BioLINCC files/ARIC/Main_Study/v2/csv/derive2_10.csv',na.strings=c("NA","","NULL"))
aric_derive3 <- fread('~/Dropbox/BioLINCC files/ARIC/Main_Study/v3/csv/derive37.csv',na.strings=c("NA","","NULL"))
aric_derive4 <- fread('~/Dropbox/BioLINCC files/ARIC/Main_Study/v4/csv/derive47.csv',na.strings=c("NA","","NULL"))
aric_derive5 <- fread('~/Dropbox/BioLINCC files/ARIC/Main_Study/v5/csv/derive51.csv',na.strings=c("NA","","NULL"))
aric_derive6 <- fread('~/Dropbox/BioLINCC files/ARIC/Main_Study/V6/csv/derive61.csv',na.strings=c("NA","","NULL"))
aric_derive7 <- fread('~/Dropbox/BioLINCC files/ARIC/Main_Study/V7/csv/derive71.csv',na.strings=c("NA","","NULL"))
aric_derive8 <- fread('~/Dropbox/BioLINCC files/ARIC/Main_Study/V8T/csv/derive8t1_np.csv',na.strings=c("NA","","NULL"))

aric_dates <- merge(aric_derive1[,c("ID_C","CENTERID","ENROLL_YR","PREVHF01","V1AGE01")],
                    aric_derive2[,c("ID_C","V2DAYS","V2AGE22")],all.x=T)
aric_dates <- merge(aric_dates,aric_derive3[,c("ID_C","V3DAYS","V3AGE31")],all.x=T)
aric_dates <- merge(aric_dates,aric_derive4[,c("ID_C","V4DAYS","V4AGE41")],all.x=T)
aric_dates <- merge(aric_dates,aric_derive5[,c("ID_C","V5DATE51_DAYS","V5AGE51")],all.x=T)
aric_dates <- merge(aric_dates,aric_derive6[,c("ID_C","V6DATE61_DAYS","V6AGE61")],all.x=T)
aric_dates <- merge(aric_dates,aric_derive7[,c("ID_C","V7DATE71_FOLLOWUPDAYS","V7AGE72")],all.x=T)
aric_dates <- merge(aric_dates,aric_derive8[,c("ID_C","V8TDATE8T1_FOLLOWUPDAYS","V8TAGE8T2")],all.x=T)

aric_dates$cohort <- aric_dates$CENTERID

aric_dates$cohort_name[aric_dates$cohort=="A"] <- "MN"
aric_dates$cohort_name[aric_dates$cohort=="B"] <- "MD"
aric_dates$cohort_name[aric_dates$cohort=="C"] <- "MI"
aric_dates$cohort_name[aric_dates$cohort=="D"] <- "NC"


aric_dates1_long <- as.data.table(sqldf("select ID_C,
                          1 as visit,
                          0 as visitdays,
                          V1AGE01 as age_obs
                          from aric_derive1"))

aric_dates2_long <- as.data.table(sqldf("select ID_C,
                          2 as visit,
                          V2DAYS as visitdays,
                          V2AGE22 as age_obs
                          from aric_derive2"))

aric_dates3_long <- as.data.table(sqldf("select ID_C,
                          3 as visit,
                          V3DAYS as visitdays,
                          V3AGE31 as age_obs
                          from aric_derive3"))

aric_dates4_long <- as.data.table(sqldf("select ID_C,
                          4 as visit,
                          V4DAYS as visitdays,
                          V4AGE41 as age_obs
                          from aric_derive4"))

aric_dates5_long <- as.data.table(sqldf("select ID_C,
                          5 as visit,
                          V5DATE51_DAYS as visitdays,
                          V5AGE51 as age_obs
                          from aric_derive5"))

aric_dates6_long <- as.data.table(sqldf("select ID_C,
                          6 as visit,
                          V6DATE61_DAYS as visitdays,
                          V6AGE61 as age_obs
                          from aric_derive6"))

aric_dates7_long <- as.data.table(sqldf("select ID_C,
                          7 as visit,
                          V7DATE71_FOLLOWUPDAYS as visitdays,
                          V7AGE72 as age_obs
                          from aric_derive7"))

aric_dates8_long <- as.data.table(sqldf("select ID_C,
                          8 as visit,
                          V8TDATE8T1_FOLLOWUPDAYS as visitdays,
                          V8TAGE8T2 as age_obs
                          from aric_derive8"))


aric_dates_long <- rbind(aric_dates1_long,
                         aric_dates2_long,
                         aric_dates3_long,
                         aric_dates4_long,
                         aric_dates5_long,
                         aric_dates6_long,
                         aric_dates7_long,
                         aric_dates8_long)


rm(aric_dates1_long,
   aric_dates2_long,
   aric_dates3_long,
   aric_dates4_long,
   aric_dates5_long,
   aric_dates6_long,
   aric_dates7_long,
   aric_dates8_long)


aric_dates_long <- merge(aric_dates_long,
                         aric_derive1[,c("ID_C","GENDER")],
                         by="ID_C",
                         all.x=T)

aric_dates_long <- merge(aric_dates_long,
                         subset(visit_yrs,
                                study=="ARIC"&
                                  cohort=="A"),
                         by="visit")

aric_dates_long$cohort <- aric_dates_long$CENTERID

aric_dates_long$cohort_name[aric_dates_long$cohort=="A"] <- "MN"
aric_dates_long$cohort_name[aric_dates_long$cohort=="B"] <- "MD"
aric_dates_long$cohort_name[aric_dates_long$cohort=="C"] <- "MI"
aric_dates_long$cohort_name[aric_dates_long$cohort=="D"] <- "NC"

aric_dates_long$study <- "ARIC"


# ==============================================================#
####                      ARIC Outcomes                      ####
# ==============================================================#


### Cohort

aric_incps <- fread('~/Dropbox/BioLINCC files/ARIC/Main_Study/cohort_incident_recurrent/csv/incps19.csv',na.strings=c("NA","","NULL"))
aric_hfcchips <- fread('~/Dropbox/BioLINCC files/ARIC/Main_Study/cohort_HF/csv/hfcchips19.csv',na.strings=c("NA","","NULL"))

aric_ccelps19 <- fread('~/Dropbox/BioLINCC files/ARIC/Main_Study/cohort_HF/csv/hfccelps19.csv',na.strings=c("NA","","NULL"))


aric_outcomes <- as.data.table(sqldf('select distinct ID_C, 
                        C7_INCHF_P_V5 as hf_after_exam5_status, 
                        C7_FT_INCHF_P_V5 as hf_after_exam5_dys,
                        ISP19 as cadhosp_status,
                        C7_FUTIMEA as cadhosp_dt,
                        PRVCHD05 as baseline_chd,
                        INCHF19 as hfhosp_status, 
                        C7_FUTIMEHF as hfhosp_dt,
                        PREVHF01 as baseline_hf,
                        DEAD19 as dth_status, 
                        FUDTH19 as dth_dt,
                        UCOD as dth_cause
                       from aric_incps'))


### CV death was defined as death with ICD‐9 code 401‐459 or ICD‐10 code I10‐I99 
# PMC4710457
# PMC6343542
# PMC8109763

aric_outcomes$cvdth_status <- 0
aric_outcomes$cvdth_status[between(as.numeric(substr(aric_outcomes$dth_cause,1,3)),401,459)|
                             substr(aric_outcomes$dth_cause,1,1)=="I"] <- 1

aric_outcomes$noncvdth_status[aric_outcomes$dth_status==0|aric_outcomes$cvdth_status==1] <- 0
aric_outcomes$noncvdth_status[aric_outcomes$cvdth_status==0&aric_outcomes$dth_status==1] <- 1

aric_outcomes$cvdth_dt <- aric_outcomes$dth_dt
aric_outcomes$noncvdth_dt <- aric_outcomes$dth_dt

aric_outcomes$hfhosp_status[aric_outcomes$baseline_hf==1&!is.na(aric_outcomes$baseline_hf)] <- 1
aric_outcomes$hfhosp_dt[aric_outcomes$baseline_hf==1&!is.na(aric_outcomes$baseline_hf)] <- 0

aric_hf_cohort <- fread('~/Dropbox/BioLINCC files/ARIC/Main_Study/cohort_HF/csv/hfcoccps19.csv',na.strings=c("NA","","NULL"))
aric_hf_cohort$visit_yr <- floor(aric_hf_cohort$HFDAYS/365)
aric_hfchfaps <- fread('~/Dropbox/BioLINCC files/ARIC/Main_Study/cohort_HF/csv/hfchfaps19.csv',na.strings=c("NA","","NULL"))

aric_hf_cohort <- 
  merge(aric_hf_cohort,
        aric_outcomes[,c("ID_C","cvdth_status","noncvdth_status")],
        by="ID_C",
        all.x=T)

aric_hfchfaps$dispo[aric_hfchfaps$HFAA0D=="D"] <- "Deceased"
aric_hfchfaps$dispo[aric_hfchfaps$HFAA0D1=="Y"] <- "Home"
aric_hfchfaps$dispo[aric_hfchfaps$HFAA0D2=="Y"] <- "Home"
aric_hfchfaps$dispo[aric_hfchfaps$HFAA0D3=="Y"] <- "Short term care"
aric_hfchfaps$dispo[aric_hfchfaps$HFAA0D4=="Y"] <- "Home"
aric_hfchfaps$dispo[aric_hfchfaps$HFAA0D5=="Y"] <- "Long term care"
aric_hfchfaps$dispo[aric_hfchfaps$HFAA0D6=="Y"] <- "Hospice"
aric_hfchfaps$dispo[aric_hfchfaps$HFAA0D7=="Y"] <- "Left AMA"

aric_hfchfaps$adm_wt_kg[aric_hfchfaps$HFAA20A1=="K"&!is.na(aric_hfchfaps$HFAA20A1)] <- 
  aric_hfchfaps$HFAA20A[aric_hfchfaps$HFAA20A1=="K"&!is.na(aric_hfchfaps$HFAA20A1)] 

aric_hfchfaps$adm_wt_kg[aric_hfchfaps$HFAA20A1=="L"&!is.na(aric_hfchfaps$HFAA20A1)] <- 
  aric_hfchfaps$HFAA20A[aric_hfchfaps$HFAA20A1=="L"&!is.na(aric_hfchfaps$HFAA20A1)] / 2.2 

aric_hfchfaps$dc_wt_kg[aric_hfchfaps$HFAA20B1=="K"&!is.na(aric_hfchfaps$HFAA20B1)] <- 
  aric_hfchfaps$HFAA20B[aric_hfchfaps$HFAA20B1=="K"&!is.na(aric_hfchfaps$HFAA20B1)] 

aric_hfchfaps$dc_wt_kg[aric_hfchfaps$HFAA20B1=="L"&!is.na(aric_hfchfaps$HFAA20B1)] <- 
  aric_hfchfaps$HFAA20B[aric_hfchfaps$HFAA20B1=="L"&!is.na(aric_hfchfaps$HFAA20B1)] / 2.2 

aric_hfchfaps$wt_chg <- 
  aric_hfchfaps$dc_wt_kg-aric_hfchfaps$adm_wt_kg

aric_hfchfaps$wt_chg_pct <- 
  (aric_hfchfaps$dc_wt_kg-aric_hfchfaps$adm_wt_kg)/aric_hfchfaps$adm_wt_kg


aric_hfchfaps$lvh_aric_grade[!aric_hfchfaps$HFAA29D1 %in% c("N","P")] <-
  aric_hfchfaps$HFAA29D1[!aric_hfchfaps$HFAA29D1 %in% c("N","P")]

aric_hfchfaps$lvh_aric_present[aric_hfchfaps$HFAA29D1 %in% c("E")] <- "No"
aric_hfchfaps$lvh_aric_present[aric_hfchfaps$HFAA29D1 %in% c("D","E","M","P","S")] <- "Yes"

aric_hfchfaps$HFAA29C1[aric_hfchfaps$ID_S0 %in% c("S111312",
                                                  "S113765",
                                                  "S114430",
                                                  "S115152",
                                                  "S116317",
                                                  "S116365",
                                                  "S117549",
                                                  "S118803",
                                                  "S120190",
                                                  "S120281",
                                                  "S122791",
                                                  "S124361",
                                                  "S125222",
                                                  "S125919",
                                                  "S127584",
                                                  "S129313",
                                                  "S130485",
                                                  "S132063",
                                                  "S135940",
                                                  "S136416",
                                                  "S136850",
                                                  "S138443",
                                                  "S138586",
                                                  "S140721",
                                                  "S141731",
                                                  "S143363",
                                                  "S143410",
                                                  "S143485",
                                                  "S144028",
                                                  "S144323",
                                                  "S144348",
                                                  "S145732",
                                                  "S146215",
                                                  "S147470",
                                                  "S148454",
                                                  "S150389",
                                                  "S152207",
                                                  "S173877",
                                                  "S185468",
                                                  "S196087",
                                                  "S196969",
                                                  "S199559",
                                                  "S203483",
                                                  "S206479")] <- 1


aric_hfchfaps$HFAA29C1[aric_hfchfaps$ID_S0 %in%  c("S112486",
                                                   "S113665",
                                                   "S114916",
                                                   "S115209",
                                                   "S124266",
                                                   "S131451",
                                                   "S132554",
                                                   "S137531",
                                                   "S140461",
                                                   "S146420",
                                                   "S150799",
                                                   "S194585",
                                                   "S204599",
                                                   "S217931",
                                                   "S218887")] <- 2


aric_hfchfaps$HFAA29C1[aric_hfchfaps$ID_S0 %in% c("S127574",
                                                  "S136289",
                                                  "S155314",
                                                  "S155682",
                                                  "S212030")] <- 3


aric_hfchfaps$ivs_cm[aric_hfchfaps$HFAA29C1==1&!is.na(aric_hfchfaps$HFAA29C1)] <-
  aric_hfchfaps$HFAA29C[aric_hfchfaps$HFAA29C1==1&!is.na(aric_hfchfaps$HFAA29C1)]

aric_hfchfaps$ivs_cm[aric_hfchfaps$HFAA29C1==2&!is.na(aric_hfchfaps$HFAA29C1)] <-
  aric_hfchfaps$HFAA29C[aric_hfchfaps$HFAA29C1==2&!is.na(aric_hfchfaps$HFAA29C1)]/10

aric_hfchfaps$ivs_cm[aric_hfchfaps$HFAA29C1==3&!is.na(aric_hfchfaps$HFAA29C1)] <-
  aric_hfchfaps$HFAA29C[aric_hfchfaps$HFAA29C1==3&!is.na(aric_hfchfaps$HFAA29C1)]/100


aric_hfchfaps$HFAA29C3[aric_hfchfaps$ID_S0 %in%  c("S111312",
                                                   "S111340",
                                                   "S113765",
                                                   "S114430",
                                                   "S114649",
                                                   "S115152",
                                                   "S116317",
                                                   "S116365",
                                                   "S116866",
                                                   "S117549",
                                                   "S117788",
                                                   "S118803",
                                                   "S119432",
                                                   "S120190",
                                                   "S120281",
                                                   "S121711",
                                                   "S122791",
                                                   "S124361",
                                                   "S124890",
                                                   "S125222",
                                                   "S125919",
                                                   "S126733",
                                                   "S127584",
                                                   "S128895",
                                                   "S129313",
                                                   "S130485",
                                                   "S132505",
                                                   "S133402",
                                                   "S134453",
                                                   "S134634",
                                                   "S135940",
                                                   "S136416",
                                                   "S136616",
                                                   "S136850",
                                                   "S138443",
                                                   "S138586",
                                                   "S140721",
                                                   "S141731",
                                                   "S142015",
                                                   "S143363",
                                                   "S143410",
                                                   "S143485",
                                                   "S144028",
                                                   "S144056",
                                                   "S144323",
                                                   "S144348",
                                                   "S145732",
                                                   "S146215",
                                                   "S147470",
                                                   "S148454",
                                                   "S148785",
                                                   "S150389",
                                                   "S152207",
                                                   "S154133",
                                                   "S154852",
                                                   "S158042",
                                                   "S169510",
                                                   "S172049",
                                                   "S181658",
                                                   "S185468",
                                                   "S185548",
                                                   "S193553",
                                                   "S201989",
                                                   "S203050",
                                                   "S203483")] <- 1


aric_hfchfaps$HFAA29C3[aric_hfchfaps$ID_S0 %in% c("S112486",
                                                  "S114916",
                                                  "S115209",
                                                  "S115893",
                                                  "S128703",
                                                  "S131451",
                                                  "S217931",
                                                  "S208111")] <- 2


aric_hfchfaps$HFAA29C3[aric_hfchfaps$ID_S0 %in% c("S127574",
                                                  "S136289",
                                                  "S155314",
                                                  "S212030")] <- 3


aric_hfchfaps$HFAA29C3[aric_hfchfaps$ID_S0 %in% c("S169510",
                                                  "S172049",
                                                  "S181658",
                                                  "S154133")] <- 4


aric_hfchfaps$lvpwd_cm[aric_hfchfaps$HFAA29C3==1&!is.na(aric_hfchfaps$HFAA29C3)] <-
  aric_hfchfaps$HFAA29C2[aric_hfchfaps$HFAA29C3==1&!is.na(aric_hfchfaps$HFAA29C3)]

aric_hfchfaps$lvpwd_cm[aric_hfchfaps$HFAA29C3==2&!is.na(aric_hfchfaps$HFAA29C3)] <-
  aric_hfchfaps$HFAA29C2[aric_hfchfaps$HFAA29C3==2&!is.na(aric_hfchfaps$HFAA29C3)]/10

aric_hfchfaps$lvpwd_cm[aric_hfchfaps$HFAA29C3==3&!is.na(aric_hfchfaps$HFAA29C3)] <-
  aric_hfchfaps$HFAA29C2[aric_hfchfaps$HFAA29C3==3&!is.na(aric_hfchfaps$HFAA29C3)]/100

aric_hfchfaps$lvpwd_cm[aric_hfchfaps$HFAA29C3==4&!is.na(aric_hfchfaps$HFAA29C3)] <-
  aric_hfchfaps$HFAA29C2[aric_hfchfaps$HFAA29C3==4&!is.na(aric_hfchfaps$HFAA29C3)] * 10


aric_hf_cohort <- merge(aric_hf_cohort, 
                        aric_hfchfaps[,c("ID_S0",
                                         "dispo",
                                         "HFAA1A",
                                         "HFAA1B",
                                         "HFAA1C",
                                         "HFAA1D",
                                         "HFAA1E",
                                         "HFAA6A",
                                         "HFAA6B",
                                         "HFAA6JNEW",
                                         "HFAA7A",
                                         "HFAA7B",
                                         "HFAA7C",
                                         "HFAA8A",
                                         "HFAA8A1",
                                         "HFAA11B1",
                                         "HFAA11B3",
                                         "HFAA11E4",
                                         "HFAA11E5",
                                         "HFAA11G",
                                         "HFAA11H",
                                         "HFAA16A",
                                         "HFAA16B",
                                         "HFAA16D",
                                         "HFAA16E",
                                         "HFAA17A",
                                         "HFAA18A",
                                         "adm_wt_kg",
                                         "dc_wt_kg",
                                         "wt_chg",
                                         "wt_chg_pct",
                                         "HFAA29B",
                                         "ivs_cm",
                                         "lvpwd_cm",
                                         "HFAA29D1",
                                         "HFAA29D2",
                                         "HFAA29D3",
                                         "HFAA29D4",
                                         "HFAA29D5",
                                         "HFAA29D6",
                                         "HFAA29D7",
                                         "HFAA29D8",
                                         "HFAA29D9",
                                         "HFAA29D9A",
                                         "HFAA29D10",
                                         "HFAA29D12",
                                         "HFAA29D13",
                                         "HFAA29D14",
                                         "HFAA32",
                                         "HFAA32B1",
                                         "HFAA32B2A",
                                         "HFAA32B2B",
                                         "HFAA32B2C",
                                         "HFAA32B2D",
                                         "HFAA32B2E",
                                         "HFAA32B3",
                                         "HFAA32B3A",
                                         "HFAA37A",
                                         "HFAA39A",
                                         "HFAA39B",
                                         "HFAA40A",
                                         "HFAA40B",
                                         "HFAA41A",
                                         "HFAA41B",
                                         "HFAA42A",
                                         "HFAA42B",
                                         "HFAA43A",
                                         "HFAA43B",
                                         "HFAA44A",
                                         "HFAA44B",
                                         "HFAA45A",
                                         "HFAA45B",
                                         "HFAA59",
                                         "HFAA59A",
                                         "HFAA60",
                                         "HFAA60A",
                                         "HFAA61A",
                                         "HFAA61A1",
                                         "HFAA61B",
                                         "HFAA61B1",
                                         "HFAA62",
                                         "HFAA62A",
                                         "HFAA64A",
                                         "HFAA64A1",
                                         "HFAA64B",
                                         "HFAA64B1",
                                         "HFAA65",
                                         "HFAA65A",
                                         "HFAA66",
                                         "HFAA66A",
                                         "HFAA71",
                                         "HFAA71A",
                                         "HFAA72",
                                         "HFAA72A")], 
                        by="ID_S0", 
                        all.x=T)


aric_hf_cohort$adm_wt_kg[aric_hf_cohort$wt_chg < -20|aric_hf_cohort$wt_chg > 20] <- NA
aric_hf_cohort$dc_wt_kg[aric_hf_cohort$wt_chg < -20|aric_hf_cohort$wt_chg > 20] <- NA
aric_hf_cohort$wt_chg[aric_hf_cohort$wt_chg < -20|aric_hf_cohort$wt_chg > 20] <- NA

aric_hf_cohort$HFAA29D2[aric_hf_cohort$HFAA29D2=="N"] <- NA
aric_hf_cohort$HFAA29D2[aric_hf_cohort$HFAA29D2==""] <- NA

aric_hf_cohort$HFAA29D3[aric_hf_cohort$HFAA29D3=="N"] <- NA
aric_hf_cohort$HFAA29D3[aric_hf_cohort$HFAA29D3==""] <- NA

aric_hf_cohort$ef_type <- aric_hf_cohort$LVEF_CUR_LOW
aric_hf_cohort$ef_type[is.na(aric_hf_cohort$ef_type)] <-
  aric_hf_cohort$LVEF_PRE_LOW[is.na(aric_hf_cohort$ef_type)]

aric_hf_cohort$ef_val <- aric_hf_cohort$LVEF_CUR
aric_hf_cohort$ef_val[is.na(aric_hf_cohort$ef_val)] <-
  aric_hf_cohort$LVEF_PRE[is.na(aric_hf_cohort$ef_val)]


aric_hf_cohort_first <- 
  as.data.table(sqldf('select * 
        from aric_hf_cohort 
        group by id_c 
        having min(HFDAYS)'))


aric_outcomes <- merge(aric_outcomes,
                       aric_hf_cohort[,c("ID_C",
                                         "HFDAYS",
                                         "ef_type",
                                         "ef_val",
                                         "HFAA29D2",
                                         "HFAA29B")],
                       by.x=c("ID_C",
                              "hfhosp_dt"),
                       by.y=c("ID_C",
                              "HFDAYS"),
                       all.x=T,
                       all.y=F)

aric_outcomes <- as.data.table(sqldf("select distinct * from aric_outcomes"))

aric_outcomes$hfhosp_ef_cat[aric_outcomes$HFAA29D2=='E'] <- 'Normal'
aric_outcomes$hfhosp_ef_cat[aric_outcomes$HFAA29D2=='M'] <- 'Mildly reduced'
aric_outcomes$hfhosp_ef_cat[aric_outcomes$HFAA29D2=='D'] <- 'Moderately reduced'
aric_outcomes$hfhosp_ef_cat[aric_outcomes$HFAA29D2=='S'] <- 'Severely reduced'
aric_outcomes$hfhosp_ef_val <- aric_outcomes$HFAA29B


aric_hfccelps <- fread('~/Dropbox/BioLINCC files/ARIC/Main_study/cohort_HF/csv/hfccelps19.csv')


names(aric_outcomes)[1] <- "patientid"
aric_outcomes$study <- "ARIC"


write_parquet(aric_outcomes,
              "~/Dropbox/ADAPT-HF/Master HDCP files/Cohort data/aric_outcomes.parquet")

#### Community ####

aric_hf_comm <- fread('~/Dropbox/BioLINCC files/aric/Main_Study/comm_HF/csv/hfsoccps14.csv',na.strings=c("NA","","NULL"))
aric_hf_comm_first <- as.data.table(sqldf('select * from aric_hf_comm group by id_c having min(HFDAYS)'))
aric_hf_comm$visit_yr <- floor(aric_hf_comm$HFDAYS/365)


aric_sdthps <- fread('~/Dropbox/BioLINCC files/ARIC/Main_Study/comm_CHD/csv/sdthps14.csv',na.strings=c("NA","","NULL"))
aric_hfschips <- fread('~/Dropbox/BioLINCC files/ARIC/Main_Study/comm_HF/csv/hfschips14.csv',na.strings=c("NA","","NULL"))

aric_sdthps$Type[
  substr(aric_sdthps$DTH2Z18,1,1) %in% c("A","B","C","D")] <- "CV" 

aric_sdthps$Type[
  substr(aric_sdthps$DTH2Z18,1,1) %in% c("E","F","G","K","L")] <- "Non-CV" 


aric_hfshfaps <- fread('~/Dropbox/BioLINCC files/ARIC/Main_Study/comm_HF/csv/hfshfaps14.csv',na.strings=c("NA","","NULL"))

aric_hfshfaps$dispo[aric_hfshfaps$HFAA0D=="D"] <- "Deceased"
aric_hfshfaps$dispo[aric_hfshfaps$HFAA0D1=="Y"] <- "Home"
aric_hfshfaps$dispo[aric_hfshfaps$HFAA0D2=="Y"] <- "Home"
aric_hfshfaps$dispo[aric_hfshfaps$HFAA0D3=="Y"] <- "Short term care"
aric_hfshfaps$dispo[aric_hfshfaps$HFAA0D4=="Y"] <- "Home"
aric_hfshfaps$dispo[aric_hfshfaps$HFAA0D5=="Y"] <- "Long term care"
aric_hfshfaps$dispo[aric_hfshfaps$HFAA0D6=="Y"] <- "Hospice"
aric_hfshfaps$dispo[aric_hfshfaps$HFAA0D7=="Y"] <- "Left AMA"


aric_hfshfaps$adm_wt_kg[aric_hfshfaps$HFAA20A1=="K"&!is.na(aric_hfshfaps$HFAA20A1)] <- 
  aric_hfshfaps$HFAA20A[aric_hfshfaps$HFAA20A1=="K"&!is.na(aric_hfshfaps$HFAA20A1)] 


aric_hfshfaps$adm_wt_kg[aric_hfshfaps$HFAA20A1=="L"&!is.na(aric_hfshfaps$HFAA20A1)] <- 
  aric_hfshfaps$HFAA20A[aric_hfshfaps$HFAA20A1=="L"&!is.na(aric_hfshfaps$HFAA20A1)] / 2.2 


aric_hfshfaps$dc_wt_kg[aric_hfshfaps$HFAA20B1=="K"&!is.na(aric_hfshfaps$HFAA20B1)] <- 
  aric_hfshfaps$HFAA20B[aric_hfshfaps$HFAA20B1=="K"&!is.na(aric_hfshfaps$HFAA20B1)] 

aric_hfshfaps$dc_wt_kg[aric_hfshfaps$HFAA20B1=="L"&!is.na(aric_hfshfaps$HFAA20B1)] <- 
  aric_hfshfaps$HFAA20B[aric_hfshfaps$HFAA20B1=="L"&!is.na(aric_hfshfaps$HFAA20B1)] / 2.2 

aric_hfshfaps$wt_chg <- 
  aric_hfshfaps$dc_wt_kg-aric_hfshfaps$adm_wt_kg

aric_hfshfaps$wt_chg_pct <- 
  (aric_hfshfaps$dc_wt_kg-aric_hfshfaps$adm_wt_kg)/aric_hfshfaps$adm_wt_kg

aric_hfshfaps$lvh_aric_grade[!aric_hfshfaps$HFAA29D1 %in% c("N","P")] <-
  aric_hfshfaps$HFAA29D1[!aric_hfshfaps$HFAA29D1 %in% c("N","P")]

aric_hfshfaps$lvh_aric_present[aric_hfshfaps$HFAA29D1 %in% c("E")] <- "No"
aric_hfshfaps$lvh_aric_present[aric_hfshfaps$HFAA29D1 %in% c("D","E","M","P","S")] <- "Yes"


## Fixing unit errors
## Units codes: 1=cm, 2=mm

aric_hfshfaps$HFAA29C1[
  aric_hfshfaps$ID_S0 %in% 
    subset(aric_hfshfaps, 
           HFAA29C<5&
             HFAA29C1==2)$ID_S0] <- 1

aric_hfshfaps$HFAA29C1[
  aric_hfshfaps$ID_S0 %in% 
    subset(aric_hfshfaps, 
           HFAA29C<5&
             HFAA29C1==1)$ID_S0] <- 2

aric_hfshfaps$ivs_cm[aric_hfshfaps$HFAA29C1==1&!is.na(aric_hfshfaps$HFAA29C1)] <-
  aric_hfshfaps$HFAA29C[aric_hfshfaps$HFAA29C1==1&!is.na(aric_hfshfaps$HFAA29C1)]

aric_hfshfaps$ivs_cm[aric_hfshfaps$HFAA29C1==2&!is.na(aric_hfshfaps$HFAA29C1)] <-
  aric_hfshfaps$HFAA29C[aric_hfshfaps$HFAA29C1==2&!is.na(aric_hfshfaps$HFAA29C1)]/10

aric_hfshfaps$ivs_cm[aric_hfshfaps$HFAA29C1==3&!is.na(aric_hfshfaps$HFAA29C1)] <-
  aric_hfshfaps$HFAA29C[aric_hfshfaps$HFAA29C1==3&!is.na(aric_hfshfaps$HFAA29C1)]/100


aric_hfshfaps$HFAA29C3[
  aric_hfshfaps$ID_S0 %in% 
    subset(aric_hfshfaps, HFAA29C2 <5&HFAA29C1==2)[,c("ID_S0")]] <- 1


aric_hfshfaps$HFAA29C3[
  aric_hfshfaps$ID_S0 %in% 
    subset(aric_hfshfaps, HFAA29C2 >7&HFAA29C1==1)[,c("ID_S0")]] <- 2


aric_hfshfaps$lvpwd_cm[aric_hfshfaps$HFAA29C3==1&!is.na(aric_hfshfaps$HFAA29C3)] <-
  aric_hfshfaps$HFAA29C2[aric_hfshfaps$HFAA29C3==1&!is.na(aric_hfshfaps$HFAA29C3)]

aric_hfshfaps$lvpwd_cm[aric_hfshfaps$HFAA29C3==2&!is.na(aric_hfshfaps$HFAA29C3)] <-
  aric_hfshfaps$HFAA29C2[aric_hfshfaps$HFAA29C3==2&!is.na(aric_hfshfaps$HFAA29C3)]/10


aric_hf_comm <-
  aric_hfshfaps[,c("ID_S0",
                   "dispo",
                   "HFAA1A",
                   "HFAA1B",
                   "HFAA1C",
                   "HFAA1D",
                   "HFAA1E",
                   "HFAA6A",
                   "HFAA6B",
                   "HFAA6JNEW",
                   "HFAA7A",
                   "HFAA7B",
                   "HFAA7C",
                   "HFAA8A",
                   "HFAA8A1",
                   "HFAA11B1",
                   "HFAA11B3",
                   "HFAA11E4",
                   "HFAA11E5",
                   "HFAA11G",
                   "HFAA11H",
                   "HFAA16A",
                   "HFAA16B",
                   "HFAA16D",
                   "HFAA16E",
                   "HFAA17A",
                   "HFAA18A",
                   "adm_wt_kg",
                   "dc_wt_kg",
                   "wt_chg",
                   "wt_chg_pct",
                   "HFAA29B",
                   "ivs_cm",
                   "lvpwd_cm",
                   "HFAA29D1",
                   "HFAA29D2",
                   "HFAA29D3",
                   "HFAA29D4",
                   "HFAA29D5",
                   "HFAA29D6",
                   "HFAA29D7",
                   "HFAA29D8",
                   "HFAA29D9",
                   "HFAA29D9A",
                   "HFAA29D10",
                   "HFAA29D12",
                   "HFAA29D13",
                   "HFAA29D14",
                   "HFAA32",
                   "HFAA32B1",
                   "HFAA32B2A",
                   "HFAA32B2B",
                   "HFAA32B2C",
                   "HFAA32B2D",
                   "HFAA32B2E",
                   "HFAA32B3",
                   "HFAA32B3A",
                   "HFAA37A",
                   "HFAA39A",
                   "HFAA39B",
                   "HFAA40A",
                   "HFAA40B",
                   "HFAA41A",
                   "HFAA41B",
                   "HFAA42A",
                   "HFAA42B",
                   "HFAA43A",
                   "HFAA43B",
                   "HFAA44A",
                   "HFAA44B",
                   "HFAA45A",
                   "HFAA45B",
                   "HFAA59",
                   "HFAA59A",
                   "HFAA60",
                   "HFAA60A",
                   "HFAA61A",
                   "HFAA61A1",
                   "HFAA61B",
                   "HFAA61B1",
                   "HFAA62",
                   "HFAA62A",
                   "HFAA64A",
                   "HFAA64A1",
                   "HFAA64B",
                   "HFAA64B1",
                   "HFAA65",
                   "HFAA65A",
                   "HFAA66",
                   "HFAA66A",
                   "HFAA71",
                   "HFAA71A",
                   "HFAA72",
                   "HFAA72A")]

aric_hf_comm$adm_wt_kg[aric_hf_comm$wt_chg < -20|aric_hf_comm$wt_chg > 20] <- NA
aric_hf_comm$dc_wt_kg[aric_hf_comm$wt_chg < -20|aric_hf_comm$wt_chg > 20] <- NA
aric_hf_comm$wt_chg[aric_hf_comm$wt_chg < -20|aric_hf_comm$wt_chg > 20] <- NA

aric_hf_comm$HFAA29D2[aric_hf_comm$HFAA29D2=="N"] <- NA
aric_hf_comm$HFAA29D2[aric_hf_comm$HFAA29D2==""] <- NA

aric_hf_comm$HFAA29D3[aric_hf_comm$HFAA29D3=="N"] <- NA
aric_hf_comm$HFAA29D3[aric_hf_comm$HFAA29D3==""] <- NA

# aric_hf_comm$ef_type <- aric_hf_comm$LVEF_CUR_LOW
# aric_hf_comm$ef_type[is.na(aric_hf_comm$ef_type)] <-
#   aric_hf_comm$LVEF_PRE_LOW[is.na(aric_hf_comm$ef_type)]
# 
# aric_hf_comm$ef_val <- aric_hf_comm$LVEF_CUR
# aric_hf_comm$ef_val[is.na(aric_hf_comm$ef_val)] <-
#   aric_hf_comm$LVEF_PRE[is.na(aric_hf_comm$ef_val)]


aric_hf_comm$hfhosp_ef_cat[aric_hf_comm$HFAA29D2=='E'] <- 'Normal'
aric_hf_comm$hfhosp_ef_cat[aric_hf_comm$HFAA29D2=='M'] <- 'Mildly reduced'
aric_hf_comm$hfhosp_ef_cat[aric_hf_comm$HFAA29D2=='D'] <- 'Moderately reduced'
aric_hf_comm$hfhosp_ef_cat[aric_hf_comm$HFAA29D2=='S'] <- 'Severely reduced'
aric_hf_comm$hfhosp_ef_val <- aric_hf_comm$HFAA29B

write_parquet(aric_hf_comm,
              "~/Dropbox/ADAPT-HF/Master HDCP files/Cohort data/aric_hf_comm.parquet")

rm(list=ls(pattern="\\baric_hf.*$"))


#===============================================================#
####             Semi-annual follow-up forms                 ####
#===============================================================#


### GEN

aric_genps1 <- fread('~/Dropbox/BioLINCC files/ARIC/Main_Study/AFU/csv/genps1.csv',
                     na.strings=c("NA","","NULL"))

aric_melt_genps1 <- melt(aric_genps1,
                         id.vars=c("ID_C",
                                   "GEN_FUDAYS",
                                   "CONTYR"),
                         na.rm=T,
                         stringsAsFactors=T)
aric_melt_genps1$form <- "genps"
names(aric_melt_genps1)[1:3] <- c("patientid","visitdays","visit_yr")

### GNB

aric_gnbps1 <- fread('~/Dropbox/BioLINCC files/ARIC/Main_Study/AFU/csv/gnbps1.csv',
                     na.strings=c("NA","","NULL"))
aric_gnbps1$CONTYR <- substr(aric_gnbps1$EVENTNAME,3,4)

aric_melt_gnbps1 <- melt(aric_gnbps1[,-c(3,38,40)],
                         id.vars=c("ID_C",
                                   "GNB_FUDAYS",
                                   "CONTYR"),
                         na.rm=T,
                         stringsAsFactors=T)
aric_melt_gnbps1$form <- "gnbps"
names(aric_melt_gnbps1)[1:3] <- c("patientid","visitdays","visit_yr")


### GNC

aric_gncps1 <- fread('~/Dropbox/BioLINCC files/ARIC/Main_Study/AFU/csv/gncps1.csv',
                     na.strings=c("NA","","NULL"))
aric_gncps1$CONTYR <- substr(aric_gncps1$EVENTNAME,3,4)


aric_gncps1$gh1 <- aric_gncps1$GNC1
aric_gncps1$pf02 <- aric_gncps1$GNC2A
aric_gncps1$pf04 <- aric_gncps1$GNC2B
aric_gncps1$rp2[aric_gncps1$GNC3A %in% c(1,2,3)] <- 1
aric_gncps1$rp2[aric_gncps1$GNC3A %in% c(4,5)] <- 0

aric_gncps1$rp3[aric_gncps1$GNC3B %in% c(1,2,3)] <- 1
aric_gncps1$rp3[aric_gncps1$GNC3B %in% c(4,5)] <- 0

aric_gncps1$re2[aric_gncps1$GNC4A %in% c(1,2,3)] <- 1
aric_gncps1$re2[aric_gncps1$GNC4A %in% c(4,5)] <- 0

aric_gncps1$re3[aric_gncps1$GNC4B %in% c(1,2,3)] <- 1
aric_gncps1$re3[aric_gncps1$GNC4B %in% c(4,5)] <- 0

aric_gncps1$bp2 <- aric_gncps1$GNC5

aric_gncps1$mh3 <- aric_gncps1$GNC6A
aric_gncps1$vt2 <- aric_gncps1$GNC6B
aric_gncps1$mh4 <- aric_gncps1$GNC6C

aric_gncps1$sf2 <- aric_gncps1$GNC7

# aric_gncps1 <- cbind(aric_gncps1,sf12(aric_gncps1[,30:41]))

aric_melt_gncps1 <- melt(aric_gncps1[,-c(3,28)],
                         id.vars=c("ID_C",
                                   "GNC_FUDAYS",
                                   "CONTYR"),
                         na.rm=T,
                         stringsAsFactors=T)
aric_melt_gncps1$form <- "gncps"
names(aric_melt_gncps1)[1:3] <- c("patientid","visitdays","visit_yr")


### GND


aric_gndps1 <- fread('~/Dropbox/BioLINCC files/ARIC/Main_Study/AFU/csv/gndps1.csv',
                     na.strings=c("NA","","NULL"))

aric_gndps1$CONTYR <- substr(aric_gndps1$EVENTNAME,4,5)

aric_melt_gndps1 <- melt(aric_gndps1[,-c(2,25)],
                         id.vars=c("ID_C",
                                   "GND_FUDAYS",
                                   "CONTYR"),
                         na.rm=T,
                         stringsAsFactors=T)


aric_melt_gndps1$form <- "gndps"
names(aric_melt_gndps1)[1:3] <- c("patientid","visitdays","visit_yr")


### GNE


aric_gneps1 <- fread('~/Dropbox/BioLINCC files/ARIC/Main_Study/AFU/csv/gneps1.csv',
                     na.strings=c("NA","","NULL"))
aric_gneps1$CONTYR <- substr(aric_gneps1$EVENTNAME,4,5)

aric_gneps1$forgetful <- 0
aric_gneps1$forgetful[aric_gneps1$GNE13==1|
                        aric_gneps1$GNE14==1|
                        aric_gneps1$GNE15==1|
                        aric_gneps1$GNE16==1|
                        aric_gneps1$GNE17==1|
                        aric_gneps1$GNE18==1|
                        aric_gneps1$GNE19==1|
                        aric_gneps1$GNE20==1|
                        aric_gneps1$GNE21==1|
                        aric_gneps1$GNE22==1] <- 1


aric_melt_gneps1 <- melt(aric_gneps1[,-c(2,32)],
                         id.vars=c("ID_C",
                                   "GNE_FUDAYS",
                                   "CONTYR"),
                         na.rm=T,
                         stringsAsFactors=T)
aric_melt_gneps1$form <- "gneps"
names(aric_melt_gneps1)[1:3] <- c("patientid","visitdays","visit_yr")


### Assemble GN

aric_melt_gnps <- rbind(aric_melt_genps1,
                        aric_melt_gnbps1,
                        aric_melt_gncps1,
                        aric_melt_gndps1,
                        aric_melt_gneps1)


rm(list=ls(pattern="\\baric_melt_g.*\\ps1$"))

#### ARIC analytes, all visits ####


aric_analytes <- fread('~/Dropbox/BioLINCC files/ARIC/Main_Study/Longitudinal/csv/v1_v5_analytes.csv',na.strings=c("NA","","NULL"))

aric_analytes_long <- melt(aric_analytes,
                           id.vars=c("ID_C"),
                           na.rm=T,
                           factorsAsStrings = T)

aric_analytes_long$variable <- as.character(aric_analytes_long$variable)

aric_analytes_long$visit[substring(aric_analytes_long$variable,nchar(aric_analytes_long$variable))==6] <- 6
aric_analytes_long$visit[substring(aric_analytes_long$variable,nchar(aric_analytes_long$variable))==5] <- 5
aric_analytes_long$visit[substring(aric_analytes_long$variable,nchar(aric_analytes_long$variable))==4] <- 4  
aric_analytes_long$visit[substring(aric_analytes_long$variable,nchar(aric_analytes_long$variable))==3] <- 3
aric_analytes_long$visit[substring(aric_analytes_long$variable,nchar(aric_analytes_long$variable))==2] <- 2
aric_analytes_long$visit[substring(aric_analytes_long$variable,nchar(aric_analytes_long$variable))==1] <- 1

analyte_points <- names(aric_analytes)[grep("_V",names(aric_analytes))]

aric_analytes_long$variable[aric_analytes_long$variable %in% analyte_points] <- 
  substring(aric_analytes_long$variable[aric_analytes_long$variable %in% analyte_points],
            1,
            str_locate(aric_analytes_long$variable[aric_analytes_long$variable %in% analyte_points],"_")[,"start"]-1)

aric_analytes_long$variable[aric_analytes_long$variable=="PRO"] <- "PRO_BNP"

aric_analytes_long <- merge(aric_analytes_long,
                            aric_dates_long[,c("ID_C","visit","visitdays","age_obs","visit_yr")],
                            by=c("ID_C","visit"),
                            all.x=T)

aric_analytes_long$form <- "analytes"


#===============================================================#
####                       ARIC Exam 1                       ####
#===============================================================#

aric_derive1$drinks_wk <- aric_derive1$ETHANL03/14

aric_anta_v1 <- fread('~/Dropbox/BioLINCC files/ARIC/Main_Study/V1/csv/anta.csv',na.strings=c("NA","","NULL"))
aric_dtia_v1 <- fread('~/Dropbox/BioLINCC files/ARIC/Main_Study/V1/csv/dtia.csv',na.strings=c("NA","","NULL"))

aric_anta_v1$weight_kg <-  as.numeric(aric_anta_v1$ANTA04)/2.2046226218

aric_ecgma03_v1 <- fread('~/Dropbox/BioLINCC files/ARIC/Main_Study/V1/csv/ecgma03.csv',na.strings=c("NA","","NULL"))

aric_rpaa_v1 <- fread('~/Dropbox/BioLINCC files/ARIC/Main_Study/V1/csv/rpaa02.csv',na.strings=c("NA","","NULL"))
aric_anut2_v1 <- fread('~/Dropbox/BioLINCC files/ARIC/Main_Study/V1/csv/anut2.csv',na.strings=c("NA","","NULL"))
aric_totnutx_v1 <- fread('~/Dropbox/BioLINCC files/ARIC/Main_Study/V1/csv/totnutx.csv',na.strings=c("NA","","NULL"))
aric_atrfib11_v1 <- fread('~/Dropbox/BioLINCC files/ARIC/Main_Study/V1/csv/atrfib11.csv',na.strings=c("NA","","NULL"))
aric_chma_v1 <- fread('~/Dropbox/BioLINCC files/ARIC/Main_Study/V1/csv/chma.csv',na.strings=c("NA","","NULL"))
aric_sbpa02_v1 <- fread('~/Dropbox/BioLINCC files/ARIC/Main_Study/V1/csv/sbpa02.csv',na.strings=c("NA","","NULL"))
aric_atrfib11_v1$AFIBFL <- 0
aric_atrfib11_v1$AFIBFL[aric_atrfib11_v1$AF==1|aric_atrfib11_v1$AFL==1] <- 1
aric_ftra02_v1 <- fread('~/Dropbox/BioLINCC files/ARIC/Main_Study/V1/csv/ftra02.csv',na.strings=c("NA","","NULL"))
aric_hom_v1 <- fread('~/Dropbox/BioLINCC files/ARIC/Main_Study/V1/csv/hom.csv',na.strings=c("NA","","NULL"))
aric_msra_v1 <- fread('~/Dropbox/BioLINCC files/ARIC/Main_Study/V1/csv/msra.csv',na.strings=c("NA","","NULL"))
aric_msrcod07_v1 <- fread('~/Dropbox/BioLINCC files/ARIC/Main_Study/V1/csv/msrcod07.csv',na.strings=c("NA","","NULL"))
aric_mhxa_v1 <- fread('~/Dropbox/BioLINCC files/ARIC/Main_Study/V1/csv/mhxa02.csv',na.strings=c("NA","","NULL"))
aric_hmta_v1 <- fread('~/Dropbox/BioLINCC files/ARIC/Main_Study/V1/csv/hmta.csv',na.strings=c("NA","","NULL"))
aric_pulm_v1_ref <- fread('~/Dropbox/BioLINCC files/ARIC/Main_Study/V1/csv/pulm.csv',na.strings=c("NA","","NULL"))
aric_pfta_v1 <- fread('~/Dropbox/BioLINCC files/ARIC/Main_Study/V1/csv/pfta.csv',na.strings=c("NA","","NULL"))
aric_phea_v1 <- fread('~/Dropbox/BioLINCC files/ARIC/Main_Study/V1/csv/phea.csv',na.strings=c("NA","","NULL"))
aric_sbpa_v1 <- fread('~/Dropbox/BioLINCC files/ARIC/Main_Study/V1/csv/sbpa02.csv',na.strings=c("NA","","NULL"))

aric_pfta_ref_v1 <- merge(aric_pfta_v1,aric_pulm_v1_ref, by="ID_C")
aric_pfta_ref_v1$FEV1PP <- aric_pfta_ref_v1$PFTA26/aric_pfta_ref_v1$FEV_101


aric_hom_v1$ed_years[aric_hom_v1$HOM54<=12&!is.na(aric_hom_v1$HOM54)] <- 
  aric_hom_v1$HOM54[aric_hom_v1$HOM54<=12&!is.na(aric_hom_v1$HOM54)]
aric_hom_v1$ed_years[aric_hom_v1$HOM54==13] <- 12
aric_hom_v1$ed_years[aric_hom_v1$HOM54==14|aric_hom_v1$HOM54==17] <- 13
aric_hom_v1$ed_years[aric_hom_v1$HOM54==15|aric_hom_v1$HOM54==18] <- 14
aric_hom_v1$ed_years[aric_hom_v1$HOM54==16|aric_hom_v1$HOM54==19] <- 15
aric_hom_v1$ed_years[aric_hom_v1$HOM54==20] <- 16
aric_hom_v1$ed_years[aric_hom_v1$HOM54==21] <- 20

aric_lipa_v1 <- fread('~/Dropbox/BioLINCC files/ARIC/Main_Study/V1/csv/lipa.csv',na.strings=c("NA","","NULL"))
aric_rhxa_v1 <- fread('~/Dropbox/BioLINCC files/ARIC/Main_Study/V1/csv/rhxa.csv',na.strings=c("NA","","NULL"), colClasses = "character")


string_to_replace <- "A"
columns_to_modify <- names(aric_rhxa_v1)[4:52]

aric_rhxa_v1 <- aric_rhxa_v1 %>%
  mutate(across(all_of(columns_to_modify), ~ na_if(., string_to_replace)))

aric_rhxa_v1$RHXA01 <- as.numeric(aric_rhxa_v1$RHXA01)

aric_rhxa_v1$RHXA08 <- as.numeric(aric_rhxa_v1$RHXA08)


aric_rhxa_v1$RHXA15 <- as.numeric(aric_rhxa_v1$RHXA15)

aric_rhxa_v1$RHXA22 <- as.numeric(aric_rhxa_v1$RHXA22)

aric_rhxa_v1$RHXA29 <- as.numeric(aric_rhxa_v1$RHXA29)

aric_rhxa_v1$RHXA36 <- as.numeric(aric_rhxa_v1$RHXA36)

aric_rhxa_v1$RHXA43 <- as.numeric(aric_rhxa_v1$RHXA43)


aric_rhxa_v1$hrt_agent1_yrs <- 
  aric_rhxa_v1$RHXA22

aric_rhxa_v1$hrt_agent2_yrs <- 
  aric_rhxa_v1$RHXA29

aric_rhxa_v1$hrt_agent3_yrs <- 
  aric_rhxa_v1$RHXA36

aric_rhxa_v1$hrt_agent4_yrs <- 
  aric_rhxa_v1$RHXA43

aric_rhxa_v1$years_on_ocp <- 
  aric_rhxa_v1$RHXA15

aric_hrt_yrs <-
  aric_rhxa_v1[,c("ID_C",
                  "hrt_agent1_yrs",
                  "hrt_agent2_yrs",
                  "hrt_agent3_yrs",
                  "hrt_agent4_yrs")]


aric_msrcod07_v1 <- fread('~/Dropbox/BioLINCC files/ARIC/Main_Study/V1/csv/msrcod07.csv',na.strings=c("NA","","NULL"))
aric_msrcod07_codes <- aric_msrcod07_v1[,c(1,19:35)]

aric_melt_msrcod07_codes <- melt(aric_msrcod07_codes,id.vars="ID_C",na.rm=T,factorsAsStrings=T)
aric_melt_msrcod07_codes$variable <- "medcode"
aric_melt_msrcod07_codes$form <- "msrcod"
aric_melt_msrcod07_codes <-  
  as.data.table(
    sqldf("select distinct * from aric_melt_msrcod07_codes"))

aric_melt_derive1_v1 <- melt(aric_derive1,id.vars=c("ID_C"),na.rm=T,factorsAsStrings=T)
aric_melt_derive1_v1$form <- "derive1"
aric_melt_anta_v1 <- melt(aric_anta_v1,id.vars=c("ID_C"),na.rm=T,factorsAsStrings=T)
aric_melt_anta_v1$form <- "anta"
aric_melt_anut2_v1 <- melt(aric_anut2_v1,id.vars=c("ID_C"),na.rm=T,factorsAsStrings=T)
aric_melt_anut2_v1$form <- "anut2"
aric_melt_totnux_v1 <- melt(aric_totnutx_v1,id.vars=c("ID_C"),na.rm=T,factorsAsStrings=T)
aric_melt_totnux_v1$form <- "totnux"
aric_melt_dtia_v1 <- melt(aric_dtia_v1,id.vars=c("ID_C"),na.rm=T,factorsAsStrings=T)
aric_melt_dtia_v1$form <- "dtia"
aric_melt_ecgma03_v1 <- melt(aric_ecgma03_v1,id.vars=c("ID_C"),na.rm=T,factorsAsStrings=T)
aric_melt_ecgma03_v1$form <- "ecgma03"
aric_melt_rpaa_v1 <- melt(aric_rpaa_v1,id.vars=c("ID_C"),na.rm=T,factorsAsStrings=T)
aric_melt_rpaa_v1$form <- "rpaa"
aric_melt_atrfib11_v1 <- melt(aric_atrfib11_v1,id.vars=c("ID_C"),na.rm=T,factorsAsStrings=T)
aric_melt_atrfib11_v1$form <- "atrfib11"
aric_melt_chma_v1 <- melt(aric_chma_v1,id.vars=c("ID_C"),na.rm=T,factorsAsStrings=T)
aric_melt_chma_v1$form <- "chma"
aric_melt_hmta_v1 <- melt(aric_hmta_v1,id.vars=c("ID_C"),na.rm=T,factorsAsStrings=T)
aric_melt_hmta_v1$form <- "hmta"
aric_melt_mhxa_v1 <- melt(aric_mhxa_v1,id.vars=c("ID_C"),na.rm=T,factorsAsStrings=T)
aric_melt_mhxa_v1$form <- "mhxa"
aric_melt_pfta_ref_v1 <- melt(aric_pfta_ref_v1,id.vars=c("ID_C"),na.rm=T,factorsAsStrings=T)
aric_melt_pfta_ref_v1$form <- "pfta"
aric_melt_phea_v1 <- melt(aric_phea_v1,id.vars=c("ID_C"),na.rm=T,factorsAsStrings=T)
aric_melt_phea_v1$form <- "phea"
aric_melt_ftra02_v1 <- melt(aric_ftra02_v1,id.vars=c("ID_C"),na.rm=T,factorsAsStrings=T)
aric_melt_ftra02_v1$form <- "ftra"
aric_melt_hom_v1 <- melt(aric_hom_v1,id.vars=c("ID_C"),na.rm=T,factorsAsStrings=T)
aric_melt_hom_v1$form <- "hom"
aric_melt_lipa_v1 <- melt(aric_lipa_v1,id.vars=c("ID_C"),na.rm=T,factorsAsStrings=T)
aric_melt_lipa_v1$form <- "lipa"
aric_melt_pulm_v1 <- melt(aric_pulm_v1_ref,id.vars=c("ID_C"),na.rm=T,factorsAsStrings=T)
aric_melt_pulm_v1$form <- "pulm"
aric_melt_rhxa_v1 <- melt(aric_rhxa_v1,id.vars=c("ID_C"),na.rm=T,factorsAsStrings=T)
aric_melt_rhxa_v1$form <- "rhxa"
aric_melt_sbpa_v1 <- melt(aric_sbpa02_v1,id.vars=c("ID_C"),na.rm=T,factorsAsStrings=T)
aric_melt_sbpa_v1$form <- "sbpa"


aric_melt_v1_all <- rbind(
  aric_melt_derive1_v1,
  aric_melt_anta_v1,
  aric_melt_anut2_v1,
  aric_melt_atrfib11_v1,
  aric_melt_chma_v1,
  aric_melt_dtia_v1,
  aric_melt_ecgma03_v1,
  aric_melt_ftra02_v1,
  aric_melt_hmta_v1,
  aric_melt_lipa_v1,
  aric_melt_hom_v1,
  aric_melt_mhxa_v1,
  aric_melt_pfta_ref_v1,
  aric_melt_phea_v1,
  aric_melt_pulm_v1,
  aric_melt_rhxa_v1,
  aric_melt_rpaa_v1,
  aric_melt_sbpa_v1,
  aric_melt_totnux_v1)


aric_melt_v1_all$visit <- 1
aric_melt_v1_all$visitdays <- 0


rm(list=ls(pattern="\\baric_.*\\_v1$"))


#===============================================================#
####                       ARIC Exam 2                       ####
#===============================================================#


aric_derive2$drinks_wk <- aric_derive2$ETHANL24/14

aric_antb_v2 <- fread('~/Dropbox/BioLINCC files/ARIC/Main_Study/V2/csv/antb.csv',na.strings=c("NA","","NULL"))
aric_antb_v2$weight_kg <- as.numeric(aric_antb_v2$ANTB01)/2.2046226218

aric_ecgmb22_v2 <- fread('~/Dropbox/BioLINCC files/ARIC/Main_Study/V2/csv/ecgmb22.csv',na.strings=c("NA","","NULL"))
aric_fhxa_v2 <- fread('~/Dropbox/BioLINCC files/ARIC/Main_Study/V2/csv/fhxa.csv',na.strings=c("NA","","NULL"))
aric_chmb_v2 <- fread('~/Dropbox/BioLINCC files/ARIC/Main_Study/V2/csv/chmb.csv',na.strings=c("NA","","NULL"))
aric_dtib_v2 <- fread('~/Dropbox/BioLINCC files/ARIC/Main_Study/V2/csv/dtib.csv',na.strings=c("NA","","NULL"))
aric_hhxb_v2 <- fread('~/Dropbox/BioLINCC files/ARIC/Main_Study/V2/csv/hhxb.csv',na.strings=c("NA","","NULL"),colClasses = "character")

aric_hhxb_v2$HHXB01A[aric_hhxb_v2$HHXB01A=="A"] <- NA
aric_hhxb_v2$HHXB01B[aric_hhxb_v2$HHXB01B=="A"] <- NA
aric_hhxb_v2$last_doc_months <- as.numeric(aric_hhxb_v2$HHXB01A)*12+as.numeric(aric_hhxb_v2$HHXB01B)

string_to_replace <- "A"
columns_to_modify <- names(aric_hhxb_v2)[4:96]

aric_hhxb_v2 <- aric_hhxb_v2 %>%
  mutate(across(all_of(columns_to_modify), ~ na_if(., string_to_replace)))

aric_hhxb_v2$hrt_agent5_yrs <-
  as.numeric(aric_hhxb_v2$HHXB28A) +
  as.numeric(aric_hhxb_v2$HHXB28B)/12


aric_hhxb_v2$hrt_agent6_yrs <-
  as.numeric(aric_hhxb_v2$HHXB35A) +
  as.numeric(aric_hhxb_v2$HHXB35B)/12


aric_hrt_yrs <-
  merge(aric_hrt_yrs,
        aric_hhxb_v2[,c("ID_C",
                        "hrt_agent5_yrs",
                        "hrt_agent6_yrs")],
        by="ID_C",
        all.x=T)


aric_hmtb_v2 <- fread('~/Dropbox/BioLINCC files/ARIC/Main_Study/V2/csv/hmtb.csv',na.strings=c("NA","","NULL"))
aric_hpaa_v2 <- fread('~/Dropbox/BioLINCC files/ARIC/Main_Study/V2/csv/hpaa.csv',na.strings=c("NA","","NULL"))


aric_hpba_v2 <- fread('~/Dropbox/BioLINCC files/ARIC/Main_Study/V2/csv/hpba.csv',na.strings=c("NA","","NULL"))


for (i in 3:23) {
  set(aric_hpba_v2, which(aric_hpba_v2[[i]] == "Y" & !is.na(aric_hpba_v2[[i]])), i, 2)
  set(aric_hpba_v2, which(aric_hpba_v2[[i]] == "D" & !is.na(aric_hpba_v2[[i]])), i, 1)
  set(aric_hpba_v2, which(aric_hpba_v2[[i]] == "N" & !is.na(aric_hpba_v2[[i]])), i, 0)
}  

aric_hpba_v2$maas_vit_exhaustm <-
  apply(aric_hpba_v2[,3:23],
        MARGIN=1,
        FUN=function(x) sum(as.numeric(x)))

aric_hpca_v2 <- fread('~/Dropbox/BioLINCC files/ARIC/Main_Study/V2/csv/hpca.csv',na.strings=c("NA","","NULL"))

for (i in 4:13) {
  set(aric_hpca_v2, which(aric_hpca_v2[[i]]=="A"&!is.na(aric_hpca_v2[[i]])),i,1)
      set(aric_hpca_v2, which(aric_hpca_v2[[i]]=="B"&!is.na(aric_hpca_v2[[i]])),i,2)
          set(aric_hpca_v2, which(aric_hpca_v2[[i]]=="C"&!is.na(aric_hpca_v2[[i]])),i,3)
              set(aric_hpca_v2, which(aric_hpca_v2[[i]]=="D"&!is.na(aric_hpca_v2[[i]])),i,4) 
              set(aric_hpca_v2, which(aric_hpca_v2[[i]]=="E"&!is.na(aric_hpca_v2[[i]])),i,NA)
}  

aric_hpca_v2$anger_spielberger <-
  apply(aric_hpca_v2[,4:13],
        MARGIN=1,
        FUN=function(x) sum(as.numeric(x)))

aric_lipb_v2 <- fread('~/Dropbox/BioLINCC files/ARIC/Main_Study/V2/csv/lipb.csv',na.strings=c("NA","","NULL"))
aric_nutv2_v2 <- fread('~/Dropbox/BioLINCC files/ARIC/Main_Study/V2/csv/nutv2.csv',na.strings=c("NA","","NULL"))
aric_pheb_v2 <- fread('~/Dropbox/BioLINCC files/ARIC/Main_Study/V2/csv/pheb.csv',na.strings=c("NA","","NULL"))
aric_sbpb_v2 <- fread('~/Dropbox/BioLINCC files/ARIC/Main_Study/V2/csv/sbpb02.csv',na.strings=c("NA","","NULL"))
aric_atrfib21_v2 <- fread('~/Dropbox/BioLINCC files/ARIC/Main_Study/V2/csv/atrfib21.csv',na.strings=c("NA","","NULL"))
aric_atrfib21_v2$AFIBFL <- 0
aric_atrfib21_v2$AFIBFL[aric_atrfib21_v2$AF==1|aric_atrfib21_v2$AFL==1] <- 1

aric_pftb_v2 <- fread('~/Dropbox/BioLINCC files/ARIC/Main_Study/V2/csv/pftb.csv',na.strings=c("NA","","NULL"))
aric_pftb_v2_ref <- merge(aric_pftb_v2,aric_pulm_v1_ref, by="ID_C")
aric_pftb_v2_ref$FEV1PP <- aric_pftb_v2_ref$PFTB26/aric_pftb_v2_ref$FEV_101

#### Hypertension


#### Diabetes


#### COPD


#### Asthma


#### Arthritis


#### CKD


#### CHF

aric_derive2 <- merge(aric_derive2,
                      aric_outcomes[,c("patientid","hfhosp_status","hfhosp_dt")],
                      by.x="ID_C",
                      by.y="patientid",
                      all.x=T)

aric_derive2$chf_frail[!is.na(aric_derive2$hfhosp_status)] <- 0
aric_derive2$chf_frail[!is.na(aric_derive2$hfhosp_status)&
                         aric_derive2$hfhosp_status==1&
                         aric_derive2$hfhosp_dt <= aric_derive2$V2DAYS] <- 1

#### CAD


#### Stroke


#### Cancer

aric_msrcod26_v2 <- fread('~/Dropbox/BioLINCC files/ARIC/Main_Study/V2/csv/msrcod26.csv',na.strings=c("NA","","NULL"))
aric_msrcod26_codes <- aric_msrcod26_v2[,c(1,19:35)]
aric_melt_msrcod26_codes <- melt(aric_msrcod26_codes,id.vars="ID_C",na.rm=T,factorsAsStrings=T)
aric_melt_msrcod26_codes$variable <- "medcode"
aric_melt_msrcod26_codes$form <- "msrcod"
aric_melt_msrcod26_codes <- 
  as.data.table(
  sqldf("select distinct * from aric_melt_msrcod26_codes"))

aric_melt_derive2_v2 <- melt(aric_derive2,id.vars=c("ID_C"),na.rm=T,factorsAsStrings=T)
aric_melt_derive2_v2$form <- "derive2"
aric_melt_antb_v2 <- melt(aric_antb_v2,id.vars=c("ID_C"),na.rm=T,factorsAsStrings=T)
aric_melt_antb_v2$form <- "antb"
aric_melt_chmb_v2 <- melt(aric_chmb_v2,id.vars=c("ID_C"),na.rm=T,factorsAsStrings=T)
aric_melt_chmb_v2$form <- "chmb"
aric_melt_dtib_v2 <- melt(aric_dtib_v2,id.vars=c("ID_C"),na.rm=T,factorsAsStrings=T)
aric_melt_dtib_v2$form <- "dtib"
aric_melt_ecgmb22_v2 <- melt(aric_ecgmb22_v2,id.vars=c("ID_C"),na.rm=T,factorsAsStrings=T)
aric_melt_ecgmb22_v2$form <- "ecgmb22"
aric_melt_fhxa_v2 <- melt(aric_fhxa_v2,id.vars=c("ID_C"),na.rm=T,factorsAsStrings=T)
aric_melt_fhxa_v2$form <- "fhxa"
aric_melt_hhxb_v2 <- melt(aric_hhxb_v2,id.vars=c("ID_C"),na.rm=T,factorsAsStrings=T)
aric_melt_hhxb_v2$form <- "hhxb"
aric_melt_hmtb_v2 <- melt(aric_hmtb_v2,id.vars=c("ID_C"),na.rm=T,factorsAsStrings=T)
aric_melt_hmtb_v2$form <- "hmtb"
aric_melt_hpaa_v2 <- melt(aric_hpaa_v2,id.vars=c("ID_C"),na.rm=T,factorsAsStrings=T)
aric_melt_hpaa_v2$form <- "hpaa"
aric_melt_hpba_v2 <- melt(aric_hpba_v2,id.vars=c("ID_C"),na.rm=T,factorsAsStrings=T)
aric_melt_hpba_v2$form <- "hpba"
aric_melt_hpca_v2 <- melt(aric_hpca_v2,id.vars=c("ID_C"),na.rm=T,factorsAsStrings=T)
aric_melt_hpca_v2$form <- "hpca"
aric_melt_lipb_v2 <- melt(aric_lipb_v2,id.vars=c("ID_C"),na.rm=T,factorsAsStrings=T)
aric_melt_lipb_v2$form <- "lipb"
aric_melt_nutv2_v2 <- melt(aric_nutv2_v2,id.vars=c("ID_C"),na.rm=T,factorsAsStrings=T)
aric_melt_nutv2_v2$form <- "nutv2"
aric_melt_pftb_ref_v2 <- melt(aric_pftb_v2_ref,id.vars=c("ID_C"),na.rm=T,factorsAsStrings=T)
aric_melt_pftb_ref_v2$form <- "pftb"
aric_melt_pheb_v2 <- melt(aric_pheb_v2,id.vars=c("ID_C"),na.rm=T,factorsAsStrings=T)
aric_melt_pheb_v2$form <- "pheb"

aric_melt_atrfib21_v2 <- melt(aric_atrfib21_v2,id.vars=c("ID_C"),na.rm=T,factorsAsStrings=T)
aric_melt_atrfib21_v2$form <- "atrfib21"
aric_melt_sbpb_v2 <- melt(aric_sbpb_v2,id.vars=c("ID_C"),na.rm=T,factorsAsStrings=T)
aric_melt_sbpb_v2$form <- "sbpb"

aric_melt_v2_all <- rbind(aric_melt_derive2_v2,
                          aric_melt_antb_v2,
                          aric_melt_chmb_v2,
                          aric_melt_dtib_v2,
                          aric_melt_ecgmb22_v2,
                          aric_melt_fhxa_v2,
                          aric_melt_hhxb_v2,
                          aric_melt_hmtb_v2,
                          aric_melt_hpaa_v2,
                          aric_melt_hpba_v2,
                          aric_melt_hpca_v2,
                          aric_melt_lipb_v2,
                          aric_melt_nutv2_v2,
                          aric_melt_pheb_v2,
                          aric_melt_pftb_ref_v2,
                          aric_melt_sbpb_v2,
                          aric_melt_atrfib21_v2
)

aric_v2_tempdays <- as.data.table(sqldf("select ID_C,
                          V2DAYS as visitdays
                          from aric_dates
                          where visitdays is not null"))

aric_melt_v2_all$visit <- 2
aric_melt_v2_all <- merge(aric_melt_v2_all,
                          aric_v2_tempdays,
                          by="ID_C")

rm(list=ls(pattern="\\baric_.*\\_v2$"))


#===============================================================#
####                       ARIC Exam 3                       ####
#===============================================================#


aric_derive3$drinks_wk <- aric_derive3$ETHANL32/14

aric_antc04_v3 <- fread('~/Dropbox/BioLINCC files/ARIC/Main_Study/V3/csv/antc04.csv',na.strings=c("NA","","NULL"))
aric_amha02_v3 <- fread('~/Dropbox/BioLINCC files/ARIC/Main_Study/V3/csv/amha02.csv',na.strings=c("NA","","NULL"))
aric_antc04_v3$weight_kg <- as.numeric(aric_antc04_v3$ANTC2)/2.2046226218
aric_atrfib31_v3 <- fread('~/Dropbox/BioLINCC files/ARIC/Main_Study/V3/csv/atrfib31.csv',na.strings=c("NA","","NULL"))
aric_atrfib31_v3$AFIBFL <- 0
aric_atrfib31_v3$AFIBFL[aric_atrfib31_v3$AF==1|aric_atrfib31_v3$AFL==1] <- 1

aric_dtic04_v3 <- fread('~/Dropbox/BioLINCC files/ARIC/Main_Study/V3/csv/dtic04.csv',na.strings=c("NA","","NULL"))
aric_ecgmc35_v3 <- fread('~/Dropbox/BioLINCC files/ARIC/Main_Study/V3/csv/ecgmc35.csv',na.strings=c("NA","","NULL"))
aric_echa_v3 <- fread('~/Dropbox/BioLINCC files/ARIC/Main_Study/V3/csv/echa04.csv',na.strings=c("NA","","NULL"))
aric_echa_v3$lvmass_ix <- aric_echa_v3$ECHA58/aric_echa_v3$ECHA6
aric_hhxc_v3 <- fread('~/Dropbox/BioLINCC files/ARIC/Main_Study/V3/csv/hhxc04.csv',na.strings=c("NA","","NULL"))
aric_hmtcv301_v3 <- fread('~/Dropbox/BioLINCC files/ARIC/Main_Study/V3/csv/hmtcv301.csv',na.strings=c("NA","","NULL"))
aric_lipc04_v3 <- fread('~/Dropbox/BioLINCC files/ARIC/Main_Study/V3/csv/lipc04.csv',na.strings=c("NA","","NULL"))
aric_nutv3_v3 <- fread('~/Dropbox/BioLINCC files/ARIC/Main_Study/V3/csv/nutv3.csv',na.strings=c("NA","","NULL"))
aric_nutv3x_v3 <- fread('~/Dropbox/BioLINCC files/ARIC/Main_Study/V3/csv/nutv3.csv',na.strings=c("NA","","NULL"))
aric_phxa04_v3 <- fread('~/Dropbox/BioLINCC files/ARIC/Main_Study/V3/csv/phxa04.csv',na.strings=c("NA","","NULL"))
aric_rhxb_v3 <- fread('~/Dropbox/BioLINCC files/ARIC/Main_Study/V3/csv/rhxb04.csv',na.strings=c("NA","","NULL"))
aric_rpac04_v3 <- fread('~/Dropbox/BioLINCC files/ARIC/Main_Study/V3/csv/rpac04.csv',na.strings=c("NA","","NULL"))
aric_sbpc_v3 <- fread('~/Dropbox/BioLINCC files/ARIC/Main_Study/V3/csv/sbpc04_02.csv',na.strings=c("NA","","NULL"))
aric_vitnut01_v3 <- fread('~/Dropbox/BioLINCC files/ARIC/Main_Study/V3/csv/vitnut01.csv',na.strings=c("NA","","NULL"))

#### Hypertension


#### Diabetes


#### COPD


#### Asthma


#### Arthritis


#### CKD


#### Reproductive history 

## Calculate total time using hormone replacement agents
## Note these are only since last ARIC visit


aric_rhxb_v3$hrt_agent7_yrs <-
  aric_rhxb_v3$RHXB16A +
  aric_rhxb_v3$RHXB16B/12

aric_rhxb_v3$hrt_agent8_yrs <-
  aric_rhxb_v3$RHXB28A +
  aric_rhxb_v3$RHXB28B/12


aric_hrt_yrs <-
  merge(aric_hrt_yrs,
        aric_rhxb_v3[,c("ID_C",
                        "hrt_agent7_yrs",
                        "hrt_agent8_yrs")],
        by="ID_C",
        all.x=T)

#### CHF

aric_derive3 <- merge(aric_derive3,
                      aric_outcomes[,c("patientid","hfhosp_status","hfhosp_dt")],
                      by.x="ID_C",
                      by.y="patientid",
                      all.x=T)

aric_derive3$chf_frail[!is.na(aric_derive3$hfhosp_status)] <- 0
aric_derive3$chf_frail[!is.na(aric_derive3$hfhosp_status)&
                         aric_derive3$hfhosp_status==1&
                         aric_derive3$hfhosp_dt <= aric_derive3$V3DAYS] <- 1


#### CAD


#### Stroke


#### Cancer


aric_msrcod36_v3 <- fread('~/Dropbox/BioLINCC files/ARIC/Main_Study/V3/csv/msrcod36.csv',na.strings=c("NA","","NULL"))
aric_msrcod36_codes <- aric_msrcod36_v3[,c(1,19:35)]
aric_melt_msrcod36_codes <- melt(aric_msrcod36_codes,id.vars="ID_C",na.rm=T,factorsAsStrings=T)
aric_melt_msrcod36_codes$variable <- "medcode"
aric_melt_msrcod36_codes$form <- "msrcod"
aric_melt_msrcod36_codes <- as.data.table(sqldf("select distinct * from aric_melt_msrcod36_codes"))

aric_melt_derive3_v3 <- melt(aric_derive3,id.vars=c("ID_C"),na.rm=T,factorsAsStrings=T)
aric_melt_derive3_v3$form <- "derive3"
aric_melt_antc_v3 <- melt(aric_antc04_v3,id.vars=c("ID_C"),na.rm=T,factorsAsStrings=T)
aric_melt_antc_v3$form <- "antc"
aric_melt_atrfib31_v3 <- melt(aric_atrfib31_v3,id.vars=c("ID_C"),na.rm=T,factorsAsStrings=T)
aric_melt_atrfib31_v3$form <- "atrfib31"
aric_melt_dtic04_v3 <- melt(aric_dtic04_v3,id.vars=c("ID_C"),na.rm=T,factorsAsStrings=T)
aric_melt_dtic04_v3$form <- "dtic04"
aric_melt_ecgmc35_v3 <- melt(aric_ecgmc35_v3,id.vars=c("ID_C"),na.rm=T,factorsAsStrings=T)
aric_melt_ecgmc35_v3$form <- "ecgmc35"
aric_melt_echa_v3 <- melt(aric_echa_v3,id.vars=c("ID_C"),na.rm=T,factorsAsStrings=T)
aric_melt_echa_v3$form <- "echa"
aric_melt_hhxc_v3 <- melt(aric_hhxc_v3,id.vars=c("ID_C"),na.rm=T,factorsAsStrings=T)
aric_melt_hhxc_v3$form <- "hhxc"
aric_melt_hmtcv301_v3 <- melt(aric_hmtcv301_v3,id.vars=c("ID_C"),na.rm=T,factorsAsStrings=T)
aric_melt_hmtcv301_v3$form <- "hmtcv301"
aric_melt_lipc04_v3 <- melt(aric_lipc04_v3,id.vars=c("ID_C"),na.rm=T,factorsAsStrings=T)
aric_melt_lipc04_v3$form <- "lipc04"
aric_melt_nutv3_v3 <- melt(aric_nutv3_v3,id.vars=c("ID_C"),na.rm=T,factorsAsStrings=T)
aric_melt_nutv3_v3$form <- "nutv3"
aric_melt_phxa04_v3 <- melt(aric_phxa04_v3,id.vars=c("ID_C"),na.rm=T,factorsAsStrings=T)
aric_melt_phxa04_v3$form <- "phxa"
aric_melt_rhxb_v3 <- melt(aric_rhxb_v3,id.vars=c("ID_C"),na.rm=T,factorsAsStrings=T)
aric_melt_rhxb_v3$form <- "rhxb"
aric_melt_sbpc_v3 <- melt(aric_sbpc_v3,id.vars=c("ID_C"),na.rm=T,factorsAsStrings=T)
aric_melt_sbpc_v3$form <- "sbpc"
aric_melt_vitnut01_v3 <- melt(aric_vitnut01_v3,id.vars=c("ID_C"),na.rm=T,factorsAsStrings=T)
aric_melt_vitnut01_v3$form <- "vitnut01"

aric_melt_amha02_v3 <- melt(aric_amha02_v3,id.vars="ID_C",na.rm=T, factorsAsStrings=T)
aric_melt_amha02_v3$form <- "amha"
aric_melt_v3_all <- rbind(aric_melt_derive3_v3,
                          aric_melt_antc_v3,
                          aric_melt_atrfib31_v3,
                          aric_melt_dtic04_v3,
                          aric_melt_ecgmc35_v3,
                          aric_melt_echa_v3,
                          aric_melt_amha02_v3,
                          aric_melt_hhxc_v3,
                          aric_melt_hmtcv301_v3,
                          aric_melt_lipc04_v3,
                          aric_melt_nutv3_v3,
                          aric_melt_phxa04_v3,
                          aric_melt_rhxb_v3,
                          aric_melt_sbpc_v3,
                          aric_melt_vitnut01_v3)

aric_melt_v3_all$variable <- as.character(aric_melt_v3_all$variable)

aric_v3_tempdays <- as.data.table(sqldf("select ID_C,
                          V3DAYS as visitdays
                          from aric_dates
                          where visitdays is not null"))

aric_melt_v3_all$visit <- 3
aric_melt_v3_all <- merge(aric_melt_v3_all,
                          aric_v3_tempdays,
                          by="ID_C")

rm(list=ls(pattern="\\baric_.*\\_v3$"))


#===============================================================#
####                       ARIC Exam 4                       ####
#===============================================================#


aric_derive4$drinks_wk <- aric_derive4$ETHANL41/14


#### CHF

aric_derive4 <- merge(aric_derive4,
                      aric_outcomes[,c("patientid","hfhosp_status","hfhosp_dt")],
                      by.x="ID_C",
                      by.y="patientid",
                      all.x=T)

aric_derive4$chf_frail[!is.na(aric_derive4$hfhosp_status)] <- 0
aric_derive4$chf_frail[!is.na(aric_derive4$hfhosp_status)&
                         aric_derive4$hfhosp_status==1&
                         aric_derive4$hfhosp_dt <= aric_derive4$V4DAYS] <- 1

#### CAD

aric_antd05_v4 <- fread('~/Dropbox/BioLINCC files/ARIC/Main_Study/V4/csv/antd05.csv',na.strings=c("NA","","NULL"))
aric_antd05_v4$weight_kg <- aric_antd05_v4$ANTD2/2.2046226218
aric_atrfib41_v4 <- fread('~/Dropbox/BioLINCC files/ARIC/Main_Study/V4/csv/atrfib41.csv',na.strings=c("NA","","NULL"))
aric_atrfib41_v4$AFIBFL <- 0
aric_atrfib41_v4$AFIBFL[aric_atrfib41_v4$AF==1|aric_atrfib41_v4$AFL==1] <- 1
aric_bpub04_v4 <- fread('~/Dropbox/BioLINCC files/ARIC/Main_Study/V4/csv/bpub04.csv',na.strings=c("NA","","NULL"))
aric_dhsa_v4 <- fread('~/Dropbox/BioLINCC files/ARIC/Main_Study/V4/csv/dhsa04.csv',na.strings=c("NA","","NULL"))

### For FIFE dental score, using history of tooth loss due to cavities, gum disease, or accident and presence of false teeh

aric_dhsa_v4$mouth_frail[!is.na(aric_dhsa_v4$DHSA2A)&
                           !is.na(aric_dhsa_v4$DHSA2B)&
                           !is.na(aric_dhsa_v4$DHSA2C)&
                           !is.na(aric_dhsa_v4$DHSA3)] <- 0

aric_dhsa_v4$mouth_frail[aric_dhsa_v4$DHSA2A=="Y"|
                           aric_dhsa_v4$DHSA2B=="Y"|
                           aric_dhsa_v4$DHSA2C=="Y"|
                           aric_dhsa_v4$DHSA3=="Y"] <- 1


aric_ecgmd41_v4 <- fread('~/Dropbox/BioLINCC files/ARIC/Main_Study/V4/csv/ecgmd41.csv',na.strings=c("NA","","NULL"))
aric_hhxd04_v4 <- fread('~/Dropbox/BioLINCC files/ARIC/Main_Study/V4/csv/hhxd04.csv',na.strings=c("NA","","NULL"))
aric_hmtcv401_v4 <- fread('~/Dropbox/BioLINCC files/ARIC/Main_Study/V4/csv/hmtcv401.csv',na.strings=c("NA","","NULL"))
aric_infa04_v4 <- fread('~/Dropbox/BioLINCC files/ARIC/Main_Study/V4/csv/infa04.csv',na.strings=c("NA","","NULL"))
aric_mhqa04_v4 <- fread('~/Dropbox/BioLINCC files/ARIC/Main_Study/V4/csv/mhqa04.csv',na.strings=c("NA","","NULL"))
aric_lipd04_v4 <- fread('~/Dropbox/BioLINCC files/ARIC/Main_Study/V4/csv/lipd04.csv',na.strings=c("NA","","NULL"))
aric_phxb04_v4 <- fread('~/Dropbox/BioLINCC files/ARIC/Main_Study/V4/csv/phxb04.csv',na.strings=c("NA","","NULL"))
aric_sbpd_v4 <- fread('~/Dropbox/BioLINCC files/ARIC/Main_Study/V4/csv/sbpd04_02.csv',na.strings=c("NA","","NULL"))

aric_phxb04_v4$health_coverage[aric_phxb04_v4$PHXB2A=="Y"|
                                 aric_phxb04_v4$PHXB2B=="Y"|
                                 aric_phxb04_v4$PHXB2C=="Y"|
                                 aric_phxb04_v4$PHXB2D=="Y"]  <- "Y"
aric_phxb04_v4$health_coverage[aric_phxb04_v4$PHXB2A=="N"|
                                 aric_phxb04_v4$PHXB2B=="N"|
                                 aric_phxb04_v4$PHXB2C=="N"|
                                 aric_phxb04_v4$PHXB2D=="N"]  <- "N"

aric_rhxc04_v4 <- fread('~/Dropbox/BioLINCC files/ARIC/Main_Study/V4/csv/rhxc04.csv',na.strings=c("NA","","NULL"))

#### Reproductive history 

## Calculate total time using hormone replacement agents
## Note these are only since last ARIC visit


aric_rhxc04_v4$hrt_agent9_yrs <-
  aric_rhxc04_v4$RHXC16A +
  aric_rhxc04_v4$RHXC16B/12

aric_rhxc04_v4$hrt_agent10_yrs <-
  aric_rhxc04_v4$RHXC28A +
  aric_rhxc04_v4$RHXC28B/12

aric_hrt_yrs <-
  merge(aric_hrt_yrs,
        aric_rhxc04_v4[,c("ID_C",
                          "hrt_agent9_yrs",
                          "hrt_agent10_yrs")],
        by="ID_C",
        all.x=T)


aric_hrt_yrs$hrt_years_sum <-
  apply(aric_hrt_yrs[,2:11],MARGIN=1,FUN=function(x) sum(x,na.rm=T))

aric_sesa04_v4 <- fread('~/Dropbox/BioLINCC files/ARIC/Main_Study/V4/csv/sesa04.csv',na.strings=c("NA","","NULL"))


aric_msrcod43_v4 <- fread('~/Dropbox/BioLINCC files/ARIC/Main_Study/V4/csv/msrcod43.csv',na.strings=c("NA","","NULL"))
aric_msrcod43_codes <- aric_msrcod43_v4[,c(1,19:35)]
aric_melt_msrcod43_codes <- melt(aric_msrcod43_codes,id.vars="ID_C",na.rm=T,factorsAsStrings=T)
aric_melt_msrcod43_codes$variable <- "medcode"
aric_melt_msrcod43_codes$form <- "msrcod"
aric_melt_msrcod43_codes <- as.data.table(sqldf("select distinct * from aric_melt_msrcod43_codes"))

aric_melt_derive4_v4 <- melt(aric_derive4,id.vars=c("ID_C"),na.rm=T,factorsAsStrings=T)
aric_melt_derive4_v4$form <- "derive4"

aric_melt_antd05_v4 <- melt(aric_antd05_v4,id.vars=c("ID_C"),na.rm=T,factorsAsStrings=T)
aric_melt_antd05_v4$form <- "antd"

aric_melt_atrfib41_v4 <- melt(aric_atrfib41_v4,id.vars=c("ID_C"),na.rm=T,factorsAsStrings=T)
aric_melt_atrfib41_v4$form <- "atrfib41"
aric_melt_ecgmd41_v4 <- melt(aric_ecgmd41_v4,id.vars=c("ID_C"),na.rm=T,factorsAsStrings=T)
aric_melt_ecgmd41_v4$form <- "ecgmd41"
aric_melt_hhxd04_v4 <- melt(aric_hhxd04_v4,id.vars=c("ID_C"),na.rm=T,factorsAsStrings=T)
aric_melt_hhxd04_v4$form <- "hhxd04"
aric_melt_hmtcv401_v4 <- melt(aric_hmtcv401_v4,id.vars=c("ID_C"),na.rm=T,factorsAsStrings=T)
aric_melt_hmtcv401_v4$form <- "hmtcv401"
aric_melt_lipd04_v4 <- melt(aric_lipd04_v4,id.vars=c("ID_C"),na.rm=T,factorsAsStrings=T)
aric_melt_lipd04_v4$form <- "lipd04"
aric_melt_phxb04_v4 <- melt(aric_phxb04_v4,id.vars=c("ID_C"),na.rm=T,factorsAsStrings=T)
aric_melt_phxb04_v4$form <- "phxb04"
aric_melt_rhxc04_v4 <- melt(aric_rhxc04_v4,id.vars=c("ID_C"),na.rm=T,factorsAsStrings=T)
aric_melt_rhxc04_v4$form <- "rhxc"
aric_melt_sesa04_v4 <- melt(aric_sesa04_v4,id.vars=c("ID_C"),na.rm=T,factorsAsStrings=T)
aric_melt_sesa04_v4$form <- "sesa"
aric_melt_sbpd04_v4 <- melt(aric_sbpd_v4,id.vars=c("ID_C"),na.rm=T,factorsAsStrings=T)
aric_melt_sbpd04_v4$form <- "sbpd"

aric_melt_hrt_yrs <- aric_hrt_yrs[,c("ID_C","hrt_years_sum")]
aric_melt_hrt_yrs$variable <- "hrt_years_total"
aric_melt_hrt_yrs$value <- aric_melt_hrt_yrs$hrt_years_sum
aric_melt_hrt_yrs$form <- "hrt_yrs"


aric_melt_v4_all <- rbind(aric_melt_derive4_v4,
                          aric_melt_antd05_v4,
                          aric_melt_atrfib41_v4,
                          aric_melt_ecgmd41_v4,
                          aric_melt_hhxd04_v4,
                          aric_melt_hmtcv401_v4,
                          aric_melt_lipd04_v4,
                          aric_melt_phxb04_v4,
                          aric_melt_rhxc04_v4,
                          aric_melt_sesa04_v4,
                          aric_melt_sbpd04_v4,
                          aric_melt_hrt_yrs[,c("ID_C","variable","value","form")])

aric_v4_tempdays <- 
as.data.table(sqldf("select ID_C,
                          V4DAYS as visitdays
                          from aric_dates
                          where visitdays is not null"))

aric_melt_v4_all$visit <- 4
aric_melt_v4_all <- merge(aric_melt_v4_all,
                          aric_v4_tempdays,
                          by="ID_C")

rm(list=ls(pattern="\\baric_.*\\_v4$"))


#===============================================================#
####                       ARIC Exam 5                       ####
#===============================================================#


aric_derive5$drinks_wk <- aric_derive5$ETHANL51/14
aric_derive5$fev1fvc_fraction <- aric_derive5$FEV1FVC51/100

aric_alc_v5 <- fread('~/Dropbox/BioLINCC files/ARIC/Main_Study/V5/csv/alc.csv',na.strings=c("NA","","NULL"))
aric_ant_v5 <- fread('~/Dropbox/BioLINCC files/ARIC/Main_Study/V5/csv/ant.csv',na.strings=c("NA","","NULL"))
aric_cbc_v5 <- fread('~/Dropbox/BioLINCC files/ARIC/Main_Study/V5/csv/cbc.csv',na.strings=c("NA","","NULL"))
aric_cdi_v5 <- fread('~/Dropbox/BioLINCC files/ARIC/Main_Study/V5/csv/cdi.csv',na.strings=c("NA","","NULL"))
aric_ces_v5 <- fread('~/Dropbox/BioLINCC files/ARIC/Main_Study/V5/csv/ces.csv',na.strings=c("NA","","NULL"))
aric_chm_v5 <- fread('~/Dropbox/BioLINCC files/ARIC/Main_Study/V5/csv/chm.csv',na.strings=c("NA","","NULL"))
aric_dcm_v5 <- fread('~/Dropbox/BioLINCC files/ARIC/Main_Study/V5/csv/dcm.csv',na.strings=c("NA","","NULL"))
aric_ecg_v5 <- fread('~/Dropbox/BioLINCC files/ARIC/Main_Study/V5/csv/ecg.csv',na.strings=c("NA","","NULL"))
aric_ech_v5 <- fread('~/Dropbox/BioLINCC files/ARIC/Main_Study/V5/csv/ech.csv',na.strings=c("NA","","NULL"))
aric_eco_v5 <- fread('~/Dropbox/BioLINCC files/ARIC/Main_Study/V5/csv/eco.csv',na.strings=c("NA","","NULL"))
aric_lip_v5 <- fread('~/Dropbox/BioLINCC files/ARIC/Main_Study/V5/csv/lip.csv',na.strings=c("NA","","NULL"))
aric_nfh_v5 <- fread('~/Dropbox/BioLINCC files/ARIC/Main_Study/V5/csv/nfh.csv',na.strings=c("NA","","NULL"))
aric_nhx_v5 <- fread('~/Dropbox/BioLINCC files/ARIC/Main_Study/V5/csv/nhx.csv',na.strings=c("NA","","NULL"))
aric_pac_v5 <- fread('~/Dropbox/BioLINCC files/ARIC/Main_Study/V5/csv/pac.csv',na.strings=c("NA","","NULL"))
aric_phx_v5 <- fread('~/Dropbox/BioLINCC files/ARIC/Main_Study/V5/csv/phx.csv',na.strings=c("NA","","NULL"))
aric_rse_v5 <- fread('~/Dropbox/BioLINCC files/ARIC/Main_Study/V5/csv/rse.csv',na.strings=c("NA","","NULL"))
aric_aqc_v5 <- fread('~/Dropbox/BioLINCC files/ARIC/Main_Study/V5/csv/aqc.csv',na.strings=c("NA","","NULL"))
aric_sbp_v5 <- fread('~/Dropbox/BioLINCC files/ARIC/Main_Study/V5/csv/sbp.csv',na.strings=c("NA","","NULL"))
aric_sfe_v5 <- fread('~/Dropbox/BioLINCC files/ARIC/Main_Study/V5/csv/sfe.csv',na.strings=c("NA","","NULL"))
aric_status51_v5 <- fread('~/Dropbox/BioLINCC files/ARIC/Main_Study/V5/csv/status51.csv',na.strings=c("NA","","NULL"))
aric_hom_v1 <- fread('~/Dropbox/BioLINCC files/ARIC/Main_Study/V1/csv/hom.csv',na.strings=c("NA","","NULL"))


aric_status51_v5$age_menopause_ex05 <-
  apply(aric_status51_v5[,c("AGENATMENOPAUSEF","AGESRGMENOPAUSEF")],
        MARGIN=1,
        FUN=function(x) min(x,na.rm=T))

aric_status51_v5$age_menopause_ex05[aric_status51_v5$age_menopause_ex05=="Inf"] <- NA


### Exam 5 FRAIL Score ###

## Fatigue

aric_derive5$fatigue_frail <- aric_derive5$EXHAUSTCOMP

## Resistance

##### Used SFE question 2b ('Does your health now limit you in climing several flights of stairs?)
##### 1: Yes, limited a lot
##### 2: Yes limited a little
##### 3: No, not limited at all

# aric_genps1$GEN10
# aric_gnbps1$GNB5
# aric_gncps1$GNC2B
# aric_gneps1$GNE4
# aric_paq[,c("PAQ2","PAQ2A")]
# aric_paqa04[,c("PAQA2")]
# aric_sfe$SFE2B


aric_derive5 <- merge(aric_derive5,
                      aric_sfe_v5[,c('ID_C','SFE2B')],
                      by="ID_C",
                      all.x=T)

aric_derive5$resistance_frail[aric_derive5$SFE2B %in% c(1,2)] <- 1
aric_derive5$resistance_frail[aric_derive5$SFE2B == 3] <- 0


## Ambulate

aric_derive5 <- merge(aric_derive5,
                      aric_genps1[,c('ID_C','GEN9')],
                      by="ID_C",
                      all.x=T)

aric_derive5 <- merge(aric_derive5,
                      aric_rse_v5[,c("ID_C","RSE7","RSE8","RSE9")],
                      by="ID_C",
                      all.x=T)


aric_derive5$ambulate_frail[aric_derive5$RSE8=="Y"|
                              aric_derive5$RSE9=="Y"|
                              aric_derive5$GEN9 %in% c("Much Difficulty",
                                                       "Some Difficulty",
                                                       "Unable to Do")] <- 1

aric_derive5$ambulate_frail[aric_derive5$RSE8=="N"&
                              aric_derive5$RSE9=="N"&
                              aric_derive5$GEN9 %in% c("No Difficulty")] <- 0


## Illness (composite)

#### Hypertension

aric_derive5$htn_frail <- aric_derive5$HYPERT54

#### Diabetes

aric_derive5$dm_frail[!aric_derive5$DIABTS54=="T"] <- as.numeric(aric_derive5$DIABTS54[!aric_derive5$DIABTS54=="T"])


#### COPD

aric_derive5 <- merge(aric_derive5,
                      aric_status51_v5[,c("ID_C","INCSELFREPLUNG51")],
                      by="ID_C",
                      all.x=T)

aric_derive5 <- merge(aric_derive5,
                      aric_rse_v5[,c("ID_C","RSE11","RSE11B")],
                      by="ID_C",
                      all.x=T)

aric_derive5 <- merge(aric_derive5,
                      aric_hom_v1[,c("ID_C","HOM10G")],
                      by="ID_C",
                      all.x=T)

aric_derive5$copd_frail[aric_derive5$INCSELFREPLUNG51==1|
                          aric_derive5$RSE11=="Y"|
                          aric_derive5$RSE11B=="Y"|
                          aric_derive5$HOM10G=="Y"] <- 1

aric_derive5$copd_frail[is.na(aric_derive5$copd_frail)] <- 0

#### Asthma

aric_derive5 <- merge(aric_derive5,
                      aric_status51_v5[,c("ID_C","INCSELFREPAST51")],
                      by="ID_C",
                      all.x=T)


aric_derive5 <- merge(aric_derive5,
                      aric_rse_v5[,c("ID_C","RSE14","RSE14C")],
                      by="ID_C",
                      all.x=T)

aric_derive5 <- merge(aric_derive5,
                      aric_hom_v1[,c("ID_C","HOM10H")],
                      by="ID_C",
                      all.x=T)


aric_derive5$asthma_frail[aric_derive5$INCSELFREPAST51==1|
                            aric_derive5$RSE14=="Y"|
                            aric_derive5$RSE14C=="Y"|
                            aric_derive5$HOM10H=="Y"] <- 1


aric_derive5$asthma_frail[is.na(aric_derive5$asthma_frail)] <- 0


#### Arthritis

aric_infa04_v4 <- fread('~/Dropbox/BioLINCC files/ARIC/Main_Study/V4/csv/infa04.csv',na.strings=c("NA","","NULL"))
aric_derive5 <- merge(aric_derive5, 
                      aric_infa04_v4,
                      by="ID_C",
                      all.x=T)

aric_derive5$djd_frail[aric_derive5$INFA3A=="Y"] <- 1
aric_derive5$djd_frail[aric_derive5$INFA3A=="N"] <- 0

#### CKD

aric_derive5 <- merge(aric_derive5,
                      aric_analytes[,c('ID_C',
                                       'EGFRSCR_V5')],
                      by="ID_C",
                      all.x=T)

aric_derive5$ckd_frail[aric_derive5$EGFRSCR_V5>=60] <- 0
aric_derive5$ckd_frail[aric_derive5$EGFRSCR_V5 <60] <- 1


#### CHF

aric_derive5$chf_frail <- aric_derive5$PREVDEFHF51


#### CAD

aric_derive5$chd_frail <- aric_derive5$PRVCHD51


#### Stroke

aric_derive5$stroke_frail <- aric_derive5$PRVSTR51


#### Cancer

aric_derive5$cancer_frail <- aric_derive5$V5CANCER51

aric_derive5$conditions_frail <- 
  aric_derive5$dm_frail+
  aric_derive5$htn_frail+
  aric_derive5$copd_frail+
  aric_derive5$asthma_frail+
  aric_derive5$djd_frail+
  aric_derive5$ckd_frail+
  aric_derive5$chf_frail+
  aric_derive5$chd_frail+
  aric_derive5$stroke_frail+
  aric_derive5$cancer_frail

aric_derive5$illness_frail[aric_derive5$conditions_frail <= 4] <- 0
aric_derive5$illness_frail[aric_derive5$conditions_frail > 4] <- 1

aric_derive5$wtloss_frail <- aric_derive5$WTLOSSCOMP10

aric_derive5$total_frail <- 
  aric_derive5$fatigue_frail+
  aric_derive5$resistance_frail+
  aric_derive5$ambulate_frail+
  aric_derive5$illness_frail+
  aric_derive5$wtloss_frail


# -------------------------------------------------- #


######### FRIED

##### Walking speed

#### Men:
# Height ≤ 173 cm (68.1 in):  ≤0.96 m/s      
# Height >173 cm (68.1 in):   ≤1.04  m/s	
# 

aric_derive5 <- merge(aric_derive5,
                      aric_ant_v5[,c("ID_C","ANT3")],
                      by="ID_C",
                      all.x=T)

aric_derive5$gaitspeed_fried[!is.na(aric_derive5$WALK4M51)] <- 0 
aric_derive5$gaitspeed_fried[aric_derive5$GENDER=="M"&
                               aric_derive5$ANT3>173&
                               aric_derive5$WALK4M51 >= 6] <- 1

aric_derive5$gaitspeed_fried[aric_derive5$GENDER=="M"&
                               aric_derive5$ANT3 <= 173&
                               aric_derive5$WALK4M51 >=7] <- 1


### Women
# Height ≤ 159 cm (62.6 in):  ≤0.90m/s
# Height >159 cm (62.6 in):   ≤1.02 m/s


aric_derive5$gaitspeed_fried[aric_derive5$GENDER=="F"&
                               aric_derive5$ANT3>159&
                               aric_derive5$WALK4M51 >= 6] <- 1

aric_derive5$gaitspeed_fried[aric_derive5$GENDER=="F"&
                               aric_derive5$ANT3 <= 159&
                               aric_derive5$WALK4M51 >= 7] <- 1

########### Grip (kg)

# Trials for each hand

## Uses BMI for normalization

##### Men
# BMI ≤ 24.0:     ≤ 29 kg
# BMI 24.1-26.0:  ≤ 30 kg
# BMI 26.1-28.0:  ≤ 30 kg
# BMI > 28.0:     ≤ 32 kg

##### Women
# BMI ≤ 23.0:     ≤ 17
# BMI 23.1-26.0:  ≤ 17.3 kg
# BMI 26.1-29.0:  ≤ 18 kg
# BMI > 29.0:     ≤ 21 kg

aric_derive5$grip_avg <- aric_derive5$GRIPMEAN51
aric_derive5$grip_max <- aric_derive5$GRIPBEST51

aric_derive5$grip_fried[!is.na(aric_derive5$grip_max)] <- 0 

# Men

aric_derive5$grip_fried[aric_derive5$BMI51<=24&
                          aric_derive5$grip_max <= 29&
                          aric_derive5$GENDER=="M"] <- 1 

aric_derive5$grip_fried[aric_derive5$BMI51>24&aric_derive5$BMI51<=26&
                          aric_derive5$grip_max <= 30&
                          aric_derive5$GENDER=="M"] <- 1 

aric_derive5$grip_fried[aric_derive5$BMI51>26&aric_derive5$BMI51<=28&
                          aric_derive5$grip_max <= 30&
                          aric_derive5$GENDER=="M"] <- 1 

aric_derive5$grip_fried[aric_derive5$BMI51>28&
                          aric_derive5$grip_max <= 32&
                          aric_derive5$GENDER=="M"] <- 1 

# Women

aric_derive5$grip_fried[aric_derive5$BMI51<=23&
                          aric_derive5$grip_max <= 17&
                          aric_derive5$GENDER=="F"] <- 1 

aric_derive5$grip_fried[aric_derive5$BMI51>23&
                          aric_derive5$BMI51<=26&
                          aric_derive5$grip_max <= 17.3&
                          aric_derive5$GENDER=="F"] <- 1 

aric_derive5$grip_fried[aric_derive5$BMI51>26&
                          aric_derive5$BMI51<=29&
                          aric_derive5$grip_max <= 18&
                          aric_derive5$GENDER=="F"] <- 1 

aric_derive5$grip_fried[aric_derive5$BMI51>29&
                          aric_derive5$grip_max <= 21&
                          aric_derive5$GENDER=="F"] <- 1 


aric_derive5$activity_fried <- aric_derive5$PACCOMP20

aric_derive5$wtloss_fried <- aric_derive5$WTLOSSCOMP10

aric_derive5$fatigue_fried <- aric_derive5$EXHAUSTCOMP


aric_derive5$total_fried <- 
  aric_derive5$wtloss_fried+
  aric_derive5$grip_fried+
  aric_derive5$fatigue_fried+
  aric_derive5$gaitspeed_fried+
  aric_derive5$activity_fried


### CES-D

aric_ces_v5$cesd_11 <- apply(aric_ces_v5[,2:12],
                             MARGIN=1,
                             function(x)
                               sum(x))

## Echo E/e'

aric_ech_v5$e_eprime_lateral <- aric_ech_v5$ECH20/aric_ech_v5$ECH24
aric_ech_v5$e_eprime_septal <- aric_ech_v5$ECH20/aric_ech_v5$ECH26
aric_ech_v5$e_eprime_avg <- (aric_ech_v5$e_eprime_lateral+aric_ech_v5$e_eprime_septal)/2

# -------------------------------------------------- #

## SF-12

### Physical functioning scale

aric_sfe_v5$SF12PF51 <-
  100*(((aric_sfe_v5$SFE2A+aric_sfe_v5$SFE2B)-2)/4)

aric_sfe_v5$SF12PFZ51 <-
  (aric_sfe_v5$SF12PF51 -81.18122)/29.10588


### Role Physical Scale

aric_sfe_v5$SF12RP51 <-
  100*(((aric_sfe_v5$SFE3A+aric_sfe_v5$SFE3B)-2)/8)

aric_sfe_v5$SF12RPZ51 <-
  (aric_sfe_v5$SF12RP51 - 80.52856)/27.13526


### Bodily Pain scale

aric_sfe_v5$SF12BP51 <-
  100*(((6+aric_sfe_v5$SFE5)-1)/4)

aric_sfe_v5$SF12BPZ51 <-
  (aric_sfe_v5$SF12BP51 - 81.74015)/24.53019


### General Health Scale

aric_sfe_v5$SFE1_ren[aric_sfe_v5$SFE1 %in% c(1,4,5)] <-
  6-aric_sfe_v5$SFE1[aric_sfe_v5$SFE1 %in% c(1,4,5)]

aric_sfe_v5$SFE1_ren[aric_sfe_v5$SFE1 %in% c(2,3)] <-
  (6-aric_sfe_v5$SFE1[aric_sfe_v5$SFE1 %in% c(2,3)])+0.4

aric_sfe_v5$SF12GH51 <- 
  100*((aric_sfe_v5$SFE1_ren=1)/4)

aric_sfe_v5$SF12GHZ51 <- 
  (aric_sfe_v5$SF12GH51-55.9090)/24.84380 


### Vitality scale

aric_sfe_v5$SF12VT51 <- 
  100*(((6-aric_sfe_v5$SFE6B)-1)/4)

aric_sfe_v5$SF12VTZ51 <- 
  (aric_sfe_v5$SF12VT51 - 55.59090)/24.84380


### Social functioning scale

aric_sfe_v5$SF12SF51 <- 
  100*(((6-aric_sfe_v5$SFE7)-1)/4)

aric_sfe_v5$SF12SFZ51 <- 
  (aric_sfe_v5$SF12SF51 - 86.41051)/22.35543


### Role Emotional Scale

aric_sfe_v5$SF12RE51 <- 
  100*(((aric_sfe_v5$SFE4A+
           aric_sfe_v5$SFE4B)-2)/8)

aric_sfe_v5$SF12REZ51 <- 
  (aric_sfe_v5$SF12RE51 - 86.41051)/22.35543


### Mental health scale

aric_sfe_v5$SF12MH51 <- 
  100*((((6-aric_sfe_v5$SFE6A)+
           aric_sfe_v5$SFE6C)-2)/8)

aric_sfe_v5$SF12MHZ51 <- 
  (aric_sfe_v5$SF12MH51 - 70.18217)/20.50597 


### Aggregate Mental Health Score

aric_sfe_v5$SF12AGGMENT51 <- 
  (aric_sfe_v5$SF12PFZ51* (-0.22999)) +
  (aric_sfe_v5$SF12RPZ51* (-0.12329)) +
  (aric_sfe_v5$SF12BPZ51* (-0.09731)) +
  (aric_sfe_v5$SF12GHZ51* (-0.01571)) +
  (aric_sfe_v5$SF12VTZ51* 0.23534) +
  (aric_sfe_v5$SF12SFZ51* 0.26876) +
  (aric_sfe_v5$SF12REZ51* 0.43407) +
  (aric_sfe_v5$SF12MHZ51* 0.48581)


### Aggregate Physical Health Score

aric_sfe_v5$SF12AGGPHYS51 <-
  (aric_sfe_v5$SF12PFZ51 * 0.42402) +
  (aric_sfe_v5$SF12RPZ51* 0.35119) +
  (aric_sfe_v5$SF12BPZ51* 0.31754) +
  (aric_sfe_v5$SF12GHZ51* 0.24954) +
  (aric_sfe_v5$SF12VTZ51* 0.02877) +
  (aric_sfe_v5$SF12SFZ51* (-0.00753)) +
  (aric_sfe_v5$SF12REZ51* (-0.19206)) +
  (aric_sfe_v5$SF12MHZ51* (-0.22069)) 

# -------------------------------------------------- #

## Medications

aric_msrcod51_v5 <- fread('~/Dropbox/BioLINCC files/ARIC/Main_Study/V5/csv/msrcod51.csv',na.strings=c("NA","","NULL"))
aric_msrcod51_codes <- aric_msrcod51_v5[,c(1,27:51)]
aric_melt_msrcod51_codes <- melt(aric_msrcod51_codes,id.vars="ID_C",na.rm=T,factorsAsStrings=T)
aric_melt_msrcod51_codes$variable <- "medcode"
aric_melt_msrcod51_codes$form <- "msrcod"
aric_melt_msrcod51_codes <- unique(aric_melt_msrcod51_codes)

aric_melt_derive5_v5 <- melt(aric_derive5,id.vars=c("ID_C"),na.rm=T,factorsAsStrings=T)
aric_melt_derive5_v5$form <- "derive5"
aric_melt_ant_v5 <- melt(aric_ant_v5,id.vars=c("ID_C"),na.rm=T,factorsAsStrings=T)
aric_melt_ant_v5$form <- "ant"
aric_melt_cbc_v5 <- melt(aric_cbc_v5,id.vars=c("ID_C"),na.rm=T,factorsAsStrings=T)
aric_melt_cbc_v5$form <- "cbc"
aric_melt_cdi_v5 <- melt(aric_cdi_v5,id.vars=c("ID_C"),na.rm=T,factorsAsStrings=T)
aric_melt_cdi_v5$form <- "cdi"
aric_melt_ces_v5 <- melt(aric_ces_v5,id.vars=c("ID_C"),na.rm=T,factorsAsStrings=T)
aric_melt_ces_v5$form <- "ces"
aric_melt_chm_v5 <- melt(aric_chm_v5,id.vars=c("ID_C"),na.rm=T,factorsAsStrings=T)
aric_melt_chm_v5$form <- "chm"
aric_melt_dcm_v5 <- melt(aric_dcm_v5,id.vars=c("ID_C"),na.rm=T,factorsAsStrings=T)
aric_melt_dcm_v5$form <- "dcm"
aric_melt_ech_v5 <- melt(aric_ech_v5,id.vars=c("ID_C"),na.rm=T,factorsAsStrings=T)
aric_melt_ech_v5$form <- "ech"
aric_melt_eco_v5 <- melt(aric_eco_v5,id.vars=c("ID_C"),na.rm=T,factorsAsStrings=T)
aric_melt_eco_v5$form <- "eco"
aric_melt_ecg_v5 <- melt(aric_ecg_v5,id.vars=c("ID_C"),na.rm=T,factorsAsStrings=T)
aric_melt_ecg_v5$form <- "ecg"
aric_melt_lip_v5 <- melt(aric_lip_v5,id.vars=c("ID_C"),na.rm=T,factorsAsStrings=T)
aric_melt_lip_v5$form <- "lip"

aric_melt_nfh_v5 <- melt(aric_nfh_v5,id.vars=c("ID_C"),na.rm=T,factorsAsStrings=T)
aric_melt_nfh_v5$form <- "nfh"
aric_melt_nhx_v5 <- melt(aric_nhx_v5,id.vars=c("ID_C"),na.rm=T,factorsAsStrings=T)
aric_melt_nhx_v5$form <- "nhx"

aric_melt_pac_v5 <- melt(aric_pac_v5,id.vars=c("ID_C"),na.rm=T,factorsAsStrings=T)
aric_melt_pac_v5$form <- "pac"

aric_melt_rse_v5 <- melt(aric_rse_v5,id.vars=c("ID_C"),na.rm=T,factorsAsStrings=T)
aric_melt_rse_v5$form <- "rse"

aric_melt_sfe_v5 <- melt(aric_sfe_v5,id.vars=c("ID_C"),na.rm=T,factorsAsStrings=T)
aric_melt_sfe_v5$form <- "sfe"

aric_melt_status51_v5 <- melt(aric_status51_v5,id.vars=c("ID_C"),na.rm=T,factorsAsStrings=T)
aric_melt_status51_v5$form <- "status51"


aric_melt_v5_all <- rbind(aric_melt_derive5_v5,
                          # aric_melt_sbp_v5,
                          aric_melt_ant_v5,
                          aric_melt_cbc_v5,
                          aric_melt_ces_v5,
                          aric_melt_chm_v5,
                          aric_melt_dcm_v5,
                          aric_melt_ecg_v5,
                          aric_melt_ech_v5,
                          aric_melt_eco_v5,
                          aric_melt_lip_v5,
                          aric_melt_nfh_v5,
                          aric_melt_nhx_v5,
                          aric_melt_pac_v5,
                          aric_melt_rse_v5,
                          aric_melt_sfe_v5,
                          aric_melt_status51_v5)


aric_v5_tempdays <- as.data.table(sqldf("select ID_C,
                          V5DATE51_DAYS as visitdays
                          from aric_dates
                          where visitdays is not null"))

aric_melt_v5_all$visit <- 5
aric_melt_v5_all <- merge(aric_melt_v5_all,
                          aric_v5_tempdays,
                          by="ID_C")


rm(list=ls(pattern="\\baric_.*\\_v5$"))

#===============================================================#
####                       ARIC Exam 6                       ####
#===============================================================#


aric_derive6$drinks_wk <- aric_derive6$ETHANL61/14
aric_derive6$hdl_mgdl <- aric_derive6$HDLSIU61 * 38.67


aric_alc_v6 <- fread('~/Dropbox/BioLINCC files/ARIC/Main_Study/V6/csv/alc.csv',na.strings=c("NA","","NULL"))
aric_ant_v6 <- fread('~/Dropbox/BioLINCC files/ARIC/Main_Study/V6/csv/ant.csv',na.strings=c("NA","","NULL"))
aric_cdi_v6 <- fread('~/Dropbox/BioLINCC files/ARIC/Main_Study/V6/csv/cdi.csv',na.strings=c("NA","","NULL"))
aric_ces_v6 <- fread('~/Dropbox/BioLINCC files/ARIC/Main_Study/V6/csv/ces.csv',na.strings=c("NA","","NULL"))
aric_chm_v6 <- fread('~/Dropbox/BioLINCC files/ARIC/Main_Study/V6/csv/chem2.csv',na.strings=c("NA","","NULL"))
aric_egfr_v6 <- fread('~/Dropbox/BioLINCC files/ARIC/Main_Study/V6/csv/egfr.csv',na.strings=c("NA","","NULL"))
aric_ess_v6 <- fread('~/Dropbox/BioLINCC files/ARIC/Main_Study/V6/csv/ess.csv',na.strings=c("NA","","NULL"))
aric_lip_v6 <- fread('~/Dropbox/BioLINCC files/ARIC/Main_Study/V6/csv/lipf.csv',na.strings=c("NA","","NULL"))
aric_nhx_v6 <- fread('~/Dropbox/BioLINCC files/ARIC/Main_Study/V6/csv/nhx.csv',na.strings=c("NA","","NULL"))
aric_pac_v6 <- fread('~/Dropbox/BioLINCC files/ARIC/Main_Study/V6/csv/pac.csv',na.strings=c("NA","","NULL"))
aric_paq_v6 <- fread('~/Dropbox/BioLINCC files/ARIC/Main_Study/V6/csv/paq.csv',na.strings=c("NA","","NULL"))
aric_pfx_v6 <- fread('~/Dropbox/BioLINCC files/ARIC/Main_Study/V6/csv/pfx.csv',na.strings=c("NA","","NULL"))
aric_phx_v6 <- fread('~/Dropbox/BioLINCC files/ARIC/Main_Study/V6/csv/phx.csv',na.strings=c("NA","","NULL"))
aric_sbp_v6 <- fread('~/Dropbox/BioLINCC files/ARIC/Main_Study/V6/csv/sbp.csv',na.strings=c("NA","","NULL"))
aric_sfe_v6 <- fread('~/Dropbox/BioLINCC files/ARIC/Main_Study/V6/csv/sfe.csv',na.strings=c("NA","","NULL"))
aric_status61_v6 <- fread('~/Dropbox/BioLINCC files/ARIC/Main_Study/V6/csv/status61.csv',na.strings=c("NA","","NULL"))
aric_tmw_v6 <- fread('~/Dropbox/BioLINCC files/ARIC/Main_Study/V6/csv/tmw.csv',na.strings=c("NA","","NULL"))

aric_status61_v6$age_menopause_ex06 <-
  apply(aric_status61_v6[,c("AGENATMENOPAUSEF","AGESRGMENOPAUSEF")],
        MARGIN=1,
        FUN=function(x) min(x,na.rm=T))

aric_status61_v6$age_menopause_ex06[aric_status61_v6$age_menopause_ex06=="Inf"] <- NA

### FRAIL Score ###

## Fatigue

aric_derive6$fatigue_frail <- aric_derive6$EXHAUST61

## Resistance

##### Used SFE question 2b ('Does your health now limit you in climing several flights of stairs?)
##### 1: Yes, limited a lot
##### 2: Yes limited a little
##### 3: No, not limited at all

# aric_genps1$GEN10
# aric_gnbps1$GNB5
# aric_gncps1$GNC2B
# aric_gneps1$GNE4
# aric_paq[,c("PAQ2","PAQ2A")]
# aric_paqa04[,c("PAQA2")]
# aric_sfe$SFE2B


aric_derive6 <- merge(aric_derive6,
                      aric_paq_v6[,c('ID_C','PAQ2')],
                      by="ID_C",
                      all.x=T)

aric_derive6$resistance_frail[aric_derive6$PAQ2 %in% c('B','C')] <- 1
aric_derive6$resistance_frail[aric_derive6$PAQ2=='A'] <- 0


## Ambulate

aric_derive6 <- merge(aric_derive6,
                      aric_paq_v6[,c('ID_C','PAQ1')],
                      by="ID_C",
                      all.x=T)

aric_derive6$ambulate_frail[aric_derive6$PAQ1 %in% c('B','C')] <- 1
aric_derive6$ambulate_frail[aric_derive6$PAQ1=='A'] <- 0


## Illness (composite)

#### Hypertension

aric_derive6$htn_frail[!aric_derive6$HYPERT64=='T'] <- 
  aric_derive6$HYPERT64[!aric_derive6$HYPERT64=='T']

#### Diabetes


aric_derive6$dm_frail[!aric_derive6$DIABTS64=="T"] <- as.numeric(aric_derive6$DIABTS64[!aric_derive6$DIABTS64=="T"])


#### COPD

aric_derive6 <- merge(aric_derive6,
                      aric_status61_v6[,c("ID_C","INCSELFREPLUNG61")],
                      by="ID_C",
                      all.x=T)


aric_derive6 <- merge(aric_derive6,
                      aric_derive5[,c("ID_C","copd_frail")],
                      by="ID_C",
                      all.x=T)


aric_derive6$copd_frail[aric_derive6$INCSELFREPLUNG61==1] <- 1


aric_derive6$copd_frail[is.na(aric_derive6$copd_frail)|
                          aric_derive6$INCSELFREPLUNG=='T'] <- 0

#### Asthma

aric_derive6 <- merge(aric_derive6,
                      aric_status61_v6[,c("ID_C","INCSELFREPAST61")],
                      by="ID_C",
                      all.x=T)

aric_derive6 <- merge(aric_derive6,
                      aric_derive5[,c("ID_C","asthma_frail")],
                      by="ID_C",
                      all.x=T)


aric_derive6$asthma_frail[aric_derive6$INCSELFREPAST61==1] <- 1


aric_derive6$asthma_frail[is.na(aric_derive6$asthma_frail)|
                            aric_derive6$INCSELFREPAST61=='T'] <- 0

#### Arthritis

aric_derive6 <- merge(aric_derive6,
                      aric_derive5[,c('ID_C','djd_frail')],
                      by="ID_C",
                      all.x=T)

#### CKD

aric_derive6$ckd_frail[aric_derive6$EGFRCR61>=60] <- 0
aric_derive6$ckd_frail[aric_derive6$EGFRCR61 <60] <- 1


#### CHF


aric_derive6$chf_frail <- aric_derive6$PREVDEFHF61


#### CAD

aric_derive6$chd_frail[!aric_derive6$PRVCHD61=='T'] <- 
  aric_derive6$PRVCHD61[!aric_derive6$PRVCHD61=='T']


#### Stroke

aric_derive6$stroke_frail[!aric_derive6$PRVSTR61=='T'] <- 
  aric_derive6$PRVSTR61[!aric_derive6$PRVSTR61=='T']


#### Cancer

aric_derive6$cancer_frail <- aric_derive6$V6CANCER61

aric_derive6$conditions_frail <-
  as.numeric(aric_derive6$dm_frail)+
  as.numeric(aric_derive6$htn_frail)+
  as.numeric(aric_derive6$copd_frail)+
  as.numeric(aric_derive6$asthma_frail)+
  as.numeric(aric_derive6$djd_frail)+
  as.numeric(aric_derive6$ckd_frail)+
  as.numeric(aric_derive6$chf_frail)+
  as.numeric(aric_derive6$chd_frail)+
  as.numeric(aric_derive6$stroke_frail)+
  as.numeric(aric_derive6$cancer_frail)

aric_derive6$illness_frail[aric_derive6$conditions_frail <= 4] <- 0
aric_derive6$illness_frail[aric_derive6$conditions_frail > 4] <- 1

aric_derive6$wtloss_frail <- aric_derive6$WTLOSSCOMPA61
aric_derive6$wtloss_frail <- aric_derive6$WTLOSSCOMPA61

aric_derive6$total_frail <-
  aric_derive6$fatigue_frail+
  aric_derive6$resistance_frail+
  aric_derive6$ambulate_frail+
  aric_derive6$illness_frail+
  aric_derive6$wtloss_frail


######### FRIED

aric_ant_v5 <- fread('~/Dropbox/BioLINCC files/ARIC/Main_Study/V5/csv/ant.csv',na.strings=c("NA","","NULL"))
aric_derive6 <- merge(aric_derive6,
                      aric_ant_v5[,c("ID_C","ANT3")],
                      by="ID_C",
                      all.x=T)

aric_derive6$gaitspeed_fried[!is.na(aric_derive6$WALK4M61)] <- 0 


##### Walking speed

#### Men:
# Height ≤ 173 cm (68.1 in):  ≤0.96 m/s      
# Height >173 cm (68.1 in):   ≤1.04  m/s	
# 


aric_derive6$gaitspeed_fried[aric_derive6$GENDER=="M"&
                               aric_derive6$ANT3>173&
                               aric_derive6$WALK4M61 >= 6] <- 1

aric_derive6$gaitspeed_fried[aric_derive6$GENDER=="M"&
                               aric_derive6$ANT3 <= 173&
                               aric_derive6$WALK4M61 >=7] <- 1


### Women
# Height ≤ 159 cm (62.6 in):  ≤0.90m/s
# Height >159 cm (62.6 in):   ≤1.02 m/s


aric_derive6$gaitspeed_fried[aric_derive6$GENDER=="F"&
```
