## Supplementary material for "Aggregating data to accelerate personalized therapy in heart failure (ADAPT-HF)": Harmonization scripts: Compile FHS data table.R

```
library(Hmisc)
library(descr)
library(dplyr)
library(relper)
library(stringr)
library(bigrquery)
library(haven)
library(googleCloudStorageR)
library(data.table)
library(arrow)
library(parquetize)
library(bigQueryR)

source("~/Dropbox/R scripts/Misc clinical scripts.R")
source("~/Dropbox/R scripts/echo scripts.R")

f <- function(input,output) {
  write.csv(input,file=output, row.names=F, na="")
}

visit_yrs <- fread('~/Dropbox/ADAPT-HF/Master HDCP files/Cohort visit yrs.csv')

data_fields <- 
  c("study",
    "patientid",
    "visit_yr",
    "visitdays",
    "form",
    "variable",
    "value",
    "datapoint",
    "study_field",
    "cohort",
    "cohort_name",
    "age_obs")

outcomes_list <- c("dth_status","dth_dt",
                   "cvdth_status","cvdth_dt",
                   "hfdth_status","hfdth_dt",
                   "hfhosp_ef_val","hfhosp_ef_cat",
                   "noncvdth_status","noncvdth_dt",
                   "hosp_status","hosp_dt",
                   "cadhosp_status","cadhosp_dt",
                   "cvhosp_status","cvhosp_dt",
                   "hfhosp_status","hfhosp_dt",
                   "noncvhosp_status","noncvhosp_dt")

dates_long_fields <-
  c("study",
    "cohort",
    "cohort_name",
    "patientid",
    "sex",
    "visit_yr",
    "visitdays",
    "age_obs")


#### ************************ FRAMINGHAM ************************ #### 

# .%%%%%%%%.%%%%%%%%.....%%%....%%.....%%.%%%%.%%....%%..%%%%%%...%%.....%%....%%%....%%.....%%
# .%%.......%%.....%%...%%.%%...%%%...%%%..%%..%%%...%%.%%....%%..%%.....%%...%%.%%...%%%...%%%
# .%%.......%%.....%%..%%...%%..%%%%.%%%%..%%..%%%%..%%.%%........%%.....%%..%%...%%..%%%%.%%%%
# .%%%%%%...%%%%%%%%..%%.....%%.%%.%%%.%%..%%..%%.%%.%%.%%...%%%%.%%%%%%%%%.%%.....%%.%%.%%%.%%
# .%%.......%%...%%...%%%%%%%%%.%%.....%%..%%..%%..%%%%.%%....%%..%%.....%%.%%%%%%%%%.%%.....%%
# .%%.......%%....%%..%%.....%%.%%.....%%..%%..%%...%%%.%%....%%..%%.....%%.%%.....%%.%%.....%%
# .%%.......%%.....%%.%%.....%%.%%.....%%.%%%%.%%....%%..%%%%%%...%%.....%%.%%.....%%.%%.....%%


#^^^^^^^^^^^^^^^^^^^^^^^^^^^^^^^^^^^^^^^^^^^^^^^^^^^^^^^^^^^^^^##
####                       FHS Outcomes                      ####
#^^^^^^^^^^^^^^^^^^^^^^^^^^^^^^^^^^^^^^^^^^^^^^^^^^^^^^^^^^^^^^##


fhs1_chfinit <- fread('~/Dropbox/BioLINCC files/Framingham cohort/datasets/CSV/vr_chfinit_2013_a_0828d.csv',na.strings=c("NA","","NULL"))
fhs1_survcvd <- fread('~/Dropbox/BioLINCC files/Framingham cohort/datasets/CSV/vr_survcvd_2018_a_1267d.csv',na.strings=c("NA","","NULL"))
fhs1_survdth <- fread('~/Dropbox/BioLINCC files/Framingham cohort/datasets/CSV/vr_survdth_2018_a_1268d.csv',na.strings=c("NA","","NULL"))
fhs1_survstk <- fread('~/Dropbox/BioLINCC files/Framingham cohort/datasets/CSV/vr_svstk_2018_a_1269d.csv',na.strings=c("NA","","NULL"))
fhs1_soechf <- fread('~/Dropbox/BioLINCC files/Framingham cohort/datasets/CSV/VR_SOECHF_2016_a_1070d.csv',na.strings=c("NA","","NULL"))
fhs1_afcum <- fread('~/Dropbox/BioLINCC files/Framingham cohort/datasets/CSV/VR_AFCUM_2019_a_1186d_v1.csv',na.strings=c("NA","","NULL")) # Cumulative Atrial fibrillation
fhs1_survaf <- fread('~/Dropbox/BioLINCC files/Framingham cohort/datasets/CSV/VR_SURVAF_2018_a_1272D.csv',na.strings=c("NA","","NULL")) # Cumulative Atrial fibrillation
fhs1_cancer <- fread('~/Dropbox/BioLINCC files/Framingham cohort/datasets/CSV/VR_CANCER_2019_A_1162D.csv',na.strings=c("NA","","NULL")) # Incident cancer
fhs1_diab <- fread('~/Dropbox/BioLINCC files/Framingham cohort/datasets/CSV/VR_DIAB_EX28_0_0601D.csv',na.strings=c("NA","","NULL")) # Diabetes status by cycle
fhs1_soe <- fread('~/Dropbox/BioLINCC files/Framingham cohort/datasets/CSV/VR_SOE_2019_A_1217D.csv',na.strings=c("NA","","NULL"))

names(fhs1_chfinit) <- toupper(names(fhs1_chfinit))
names(fhs1_survcvd) <- toupper(names(fhs1_survcvd))
names(fhs1_survdth) <- toupper(names(fhs1_survdth))
names(fhs1_soechf) <- toupper(names(fhs1_soechf))
names(fhs1_afcum) <- toupper(names(fhs1_afcum))
names(fhs1_survaf) <- toupper(names(fhs1_survaf))
names(fhs1_cancer) <- toupper(names(fhs1_cancer))
names(fhs1_diab) <- toupper(names(fhs1_diab))
names(fhs1_soe) <- toupper(names(fhs1_soe))


fhs2_chfinit <- fread('~/Dropbox/BioLINCC files/Framingham offspring/datasets/CSV/vr_chfinit_2013_a_0828d.csv',na.strings=c("NA","","NULL"))
fhs2_survcvd <- fread('~/Dropbox/BioLINCC files/Framingham offspring/datasets/CSV/vr_survcvd_2021_a_1453d.csv',na.strings=c("NA","","NULL"))
fhs2_survdth <- fread('~/Dropbox/BioLINCC files/Framingham offspring/datasets/CSV/vr_survdth_2021_a_1452d.csv',na.strings=c("NA","","NULL"))
fhs2_survstk <- fread('~/Dropbox/BioLINCC files/Framingham offspring/datasets/CSV/vr_svstk_2021_a_1455d.csv',na.strings=c("NA","","NULL"))
fhs2_soechf <- fread('~/Dropbox/BioLINCC files/Framingham offspring/datasets/CSV/VR_SOECHF_2016_A_1070D.csv',na.strings=c("NA","","NULL"))
fhs2_afcum <- fread('~/Dropbox/BioLINCC files/Framingham offspring/datasets/CSV/VR_AFCUM_2022_A_1412D.csv',na.strings=c("NA","","NULL")) # Cumulative Atrial fibrillation
fhs2_survaf <- fread('~/Dropbox/BioLINCC files/Framingham offspring/datasets/CSV/VR_SURVAF_2021_A_1456D.csv',na.strings=c("NA","","NULL")) # Cumulative Atrial fibrillation
fhs2_cancer <- fread('~/Dropbox/BioLINCC files/Framingham offspring/datasets/CSV/VR_CANCER_2019_A_1162D.csv',na.strings=c("NA","","NULL")) # Incident cancer
fhs2_diab <- fread('~/Dropbox/BioLINCC files/Framingham offspring/datasets/CSV/vr_diab_ex10_1b_1489d.csv',na.strings=c("NA","","NULL")) # Diabetes status by cycle
fhs2_soe <- fread('~/Dropbox/BioLINCC files/Framingham offspring/datasets/CSV/VR_SOE_2022_A_1424D.csv',na.strings=c("NA","","NULL"))

names(fhs2_chfinit) <- toupper(names(fhs2_chfinit))
names(fhs2_survcvd) <- toupper(names(fhs2_survcvd))
names(fhs2_survdth) <- toupper(names(fhs2_survdth))
names(fhs2_survstk) <- toupper(names(fhs2_survstk))
names(fhs2_soechf) <- toupper(names(fhs2_soechf))
names(fhs2_afcum) <- toupper(names(fhs2_afcum))
names(fhs2_survaf) <- toupper(names(fhs2_survaf))
names(fhs2_cancer) <- toupper(names(fhs2_cancer))
names(fhs2_diab) <- toupper(names(fhs2_diab))
names(fhs2_soe) <- toupper(names(fhs2_soe))

fhs2_afcum[,ORIGIN_SOURCE := NA]

fhs3_chfinit <- fread('~/Dropbox/BioLINCC files/Framingham third generation/datasets/CSV/vr_chfinit_2013_a_0828d.csv',na.strings=c("NA","","NULL"))
fhs3_survcvd <- fread('~/Dropbox/BioLINCC files/Framingham third generation/datasets/CSV/vr_survcvd_2019_a_1334d.csv',na.strings=c("NA","","NULL"))
fhs3_survdth <- fread('~/Dropbox/BioLINCC files/Framingham third generation/datasets/CSV/vr_survdth_2019_a_1337d.csv',na.strings=c("NA","","NULL"))
fhs3_survstk <- fread('~/Dropbox/BioLINCC files/Framingham third generation/datasets/CSV/vr_svstk_2019_a_1335d.csv',na.strings=c("NA","","NULL"))
fhs3_soechf <- fread('~/Dropbox/BioLINCC files/Framingham third generation/datasets/CSV/VR_SOECHF_2016_a_1070D.csv',na.strings=c("NA","","NULL"))
fhs3_afcum <- fread('~/Dropbox/BioLINCC files/Framingham third generation/datasets/CSV/VR_AFCUM_2021_A_1366D.csv',na.strings=c("NA","","NULL")) # Cumulative Atrial fibrillation
fhs3_survaf <- fread('~/Dropbox/BioLINCC files/Framingham third generation/datasets/CSV/VR_SURVAF_2019_A_1338D.csv',na.strings=c("NA","","NULL")) # Cumulative Atrial fibrillation
fhs3_cancer <- fread('~/Dropbox/BioLINCC files/Framingham third generation/datasets/CSV/VR_CANCER_2019_A_1162D.csv',na.strings=c("NA","","NULL")) # Incident cancer
fhs3_diab <- fread('~/Dropbox/BioLINCC files/Framingham third generation/datasets/CSV/VR_DIAB_EX03_3b_1312D.csv',na.strings=c("NA","","NULL")) # Diabetes status by cycle
fhs3_soe <- fread('~/Dropbox/BioLINCC files/Framingham third generation/datasets/CSV/VR_SOE_2020_A_1340D.csv',na.strings=c("NA","","NULL"))

fhs3_afcum[,ORIGIN_SOURCE := NA]


dm_names <- unique(c(names(fhs1_diab),names(fhs2_diab),names(fhs3_diab)))

fhs1_diab[,dm_names[!dm_names %in% names(fhs1_diab)]] <- NA
fhs2_diab[,dm_names[!dm_names %in% names(fhs2_diab)]] <- NA
fhs3_diab[,dm_names[!dm_names %in% names(fhs3_diab)]] <- NA


fhs_chfinit <- rbind(fhs1_chfinit,fhs2_chfinit,fhs3_chfinit)
fhs_survcvd <- rbind(fhs1_survcvd,fhs2_survcvd,fhs3_survcvd)
fhs_survdth <- rbind(fhs1_survdth,fhs2_survdth,fhs3_survdth)
fhs_survstk <- rbind(fhs1_survstk,fhs2_survstk,fhs3_survstk)
fhs_survaf <- rbind(fhs1_survaf,fhs2_survaf,fhs3_survaf)
fhs_afcum <- rbind(fhs1_afcum,fhs2_afcum,fhs3_afcum)
fhs_soechf <- rbind(fhs1_soechf,fhs2_soechf,fhs3_soechf)
fhs_soe <- rbind(fhs1_soe,fhs2_soe,fhs3_soe)
fhs_cancer <- rbind(fhs1_cancer,fhs2_cancer,fhs3_cancer)
fhs_diab <- rbind(fhs1_diab,fhs2_diab,fhs3_diab)

fhs_cancer <- 
  fhs_cancer[!TOPO==173|(TOPO==173&HIST>=8720&HIST<=8790)|HIST==8247]

fhs_survca <- fhs_cancer[
  , .SD[which.min(D_DATE)], 
  by = .(IDTYPE, PID)
]

fhs_survca_first_any <- unique(fhs_survca[,.(PID,IDTYPE,D_DATE)])

fhs_survdth[,dth_status := 0] 
fhs_survdth[!is.na(DATEDTH),dth_status := 1] 
fhs_survdth[,dth_dt := DATEDTH]
fhs_survdth[dth_status==0, dth_dt := LASTCON]

fhs_survdth[,cvdth_status := 0] 
fhs_survdth[CVDDEATH==1, cvdth_status := 1]
fhs_survdth[,cvdth_dt := dth_dt]

fhs_survdth[,noncvdth_status := 0] 
fhs_survdth[CVDDEATH==0, noncvdth_status := 1]
fhs_survdth[,noncvdth_dt := dth_dt]

names(fhs_survcvd)[c(3,4,5,6,7,8)] <- 
  c("cadhosp_status",
    "cadhosp_dt",
    "hfhosp_status",
    "hfhosp_dt",
    "cvhosp_status",
    "cvhosp_dt")

names(fhs_survstk)[3:6] <-
  c("cvahosp_status",
    "cva_type",
    "cva_lacunar",
    "cvahosp_dt")


fhs_outcomes <- 
  fhs_survcvd[,.(PID,IDTYPE,
                 cadhosp_status,cadhosp_dt,
                 cvhosp_status,cvhosp_dt,
                 hfhosp_status,hfhosp_dt)][
                   fhs_survdth[,.(PID,IDTYPE,
                                  dth_status,dth_dt,
                                  cvdth_status,cvdth_dt,
                                  noncvdth_status,noncvdth_dt)],
                   on=.(PID,IDTYPE),
                   nomatch=0]

fhs_outcomes <-
  fhs_survstk[,.(PID,
                 IDTYPE,
                 cvahosp_status,
                 cva_type,
                 cvahosp_dt)][fhs_outcomes,
        on=.(PID,IDTYPE)]


## Ischemic stroke

fhs_outcomes[,ischem_cvahosp_status := 0]
fhs_outcomes[cva_type %in% c(11,13), 
             ischem_cvahosp_status :=  1]


fhs_outcomes[ischem_cvahosp_status==1,
             ischem_cvahosp_dt := cvahosp_dt]

fhs_outcomes[ischem_cvahosp_status==0, 
             ischem_cvahosp_dt := dth_dt]


fhs_outcomes[
  cva_type %in% c(10, 16, 17, 19),
  c("ischem_cvahosp_status", "ischem_cvahosp_dt") := NA
]

## Hemorrhagic CVA

fhs_outcomes[,hemo_cvahosp_status := 0]
fhs_outcomes[cva_type %in% c(14,15), 
             hemo_cvahosp_status := 1]

fhs_outcomes[hemo_cvahosp_status==1, 
             hemo_cvahosp_dt := cvahosp_dt]

fhs_outcomes[hemo_cvahosp_status==0, 
             hemo_cvahosp_dt := dth_dt]

fhs_outcomes[
  cva_type %in% 
    c(10,16,17,19),
  c("hemo_cvahosp_status",
    "hemo_cvahosp_dt") := NA]


fhs_outcomes <-
  fhs_chfinit[,c("PID",
                 "IDTYPE",
                 "CHF011D",
                 "CHF017")][fhs_outcomes,
                            on=.(PID,IDTYPE,CHF011D = hfhosp_dt)]

setnames(fhs_outcomes,"CHF011D","hfhosp_dt")

fhs_outcomes[CHF017==0, hfhosp_ef_cat := "Normal"]
fhs_outcomes[CHF017==3, hfhosp_ef_cat := "Mildly reduced"]
fhs_outcomes[CHF017==2, hfhosp_ef_cat := "Moderately reduced"]
fhs_outcomes[CHF017==1, hfhosp_ef_cat := "Severely reduced"]


fhs_outcomes[,study := "FHS"]

write_parquet(fhs_outcomes,
              "~/Dropbox/ADAPT-HF/Master HDCP files/Cohort data/fhs_outcomes.parquet")

#=========================================================================#
####                    ** Combine FHS Dates **                        ####
#=========================================================================#

# ==============================================================#
####       Framingham Original cohort dates & outcomes       ####
# ==============================================================#

fhs1_dates <- fread('~/Dropbox/BioLINCC files/Framingham cohort/datasets/CSV/vr_dates_2014_a_0912d.csv',na.strings=c("NA","","NULL"))
fhs1_wkthru <- fread('~/Dropbox/BioLINCC files/Framingham cohort/datasets/CSV/vr_wkthru_ex32_0_0997d.csv',na.strings=c("NA","","NULL"))

fhs1_melt_sex <- 
  fhs1_dates[,.(PID,IDTYPE,visit=1,variable='sex',value=SEX, form="fhs1_dates")]


names(fhs1_dates) <- toupper(names(fhs1_dates))
names(fhs1_wkthru) <- toupper(names(fhs1_wkthru))

for (v in 1:32) {
  fhs1_wkthru[,paste("BMIC",v,sep="")] <- 
    round(calc_bmi(dat=as.data.frame(fhs1_wkthru),
                   weight=paste("WGT",v,sep=""),
                   height="HGT1",
                   metric=F),2)
}


whrs   <- 19:23
wcols  <- paste0("WAIST", whrs)
hcols  <- paste0("HIP",   whrs)
outcol <- paste0("WHR",   whrs)

fhs1_wkthru[, (outcol) :=
              round(.SD[, wcols, with = FALSE] /
                      .SD[, hcols, with = FALSE], 3)]

################################################################ FHS 2 Walkthru ############################################################

fhs2_offspring_wkthru <- fread('~/Dropbox/BioLINCC files/Framingham offspring/datasets/CSV/vr_wkthru_ex10_1b_1488d.csv',na.strings=c("NA","","NULL"))
fhs2_dates <- fread('~/Dropbox/BioLINCC files/Framingham offspring/datasets/CSV/vr_dates_2022_a_1487d.csv',na.strings=c("NA","","NULL"))
fhs2_race_1 <- fread('~/Dropbox/BioLINCC files/Framingham offspring/datasets/CSV/race_1.csv',na.strings=c("NA","","NULL")) # Physical exam questionnaire, exam 5

fhs2_offspring_wkthru[, c("att5_status",
                          "att10_status",
                          "date5_fu",
                          "date10_fu") := NULL]

names(fhs2_race_1) <- toupper(names(fhs2_race_1))

fhs2_race_1[ETHNICITY=="Hisp", RACE := "Hisp"] 
fhs2_race_1[is.na(RACE), RACE := "W"]

fhs2_melt_race <-
  melt(fhs2_race_1,
       id.vars=c("PID","IDTYPE"),
       na.rm=T)

fhs2_offspring_wkthru <-
  fhs2_race_1[fhs2_offspring_wkthru,
              on=.(PID,IDTYPE)]


fhs2_melt_race[,form := "fhs2_race_1"]
fhs2_melt_race[,visit := 1]

fhs2_melt_sex <- 
  fhs2_dates[,.(PID,IDTYPE=idtype,visit=1,variable='sex', form='fhs2_dates', value=sex)]


names(fhs2_dates) <- toupper(names(fhs2_dates))
names(fhs2_offspring_wkthru) <- toupper(names(fhs2_offspring_wkthru))

whrs   <- c(1,2,4,5,6,7,9,10)
wcols  <- paste0("WAIST", whrs)
hcols  <- paste0("HIP",   whrs)
outcol <- paste0("WHR",   whrs)

fhs2_offspring_wkthru[, (outcol) :=
              round(.SD[, wcols, with = FALSE] /
                      .SD[, hcols, with = FALSE], 3)]

for (i in 2:10) {
  fhs2_offspring_wkthru[
    , paste0("EGFR_MDRD", i) :=
      calc_MDRD4(
        dat  = as.data.frame(.SD),    # or as.data.frame(fhs2_offspring_wkthru)
        age  = paste0("AGE",   i),
        cr   = paste0("CREAT", i),
        sex  = "SEX",
        race = "RACE",
        black = "B"
      )
  ]
}


#### FHS 3 


fhs3_wkthru <- fread('~/Dropbox/BioLINCC files/Framingham third generation/datasets/CSV/vr_wkthru_ex03_3b_1191d.csv',na.strings=c("NA","","NULL"))
fhs3_dates <- fread('~/Dropbox/BioLINCC files/Framingham third generation/datasets/CSV/vr_dates_2019_a_1175d.csv',na.strings=c("NA","","NULL"))
fhs3_black <-
  fread('~/Dropbox/BioLINCC files/Framingham third generation/datasets/CSV/e_exam_ex01_3_0086d.csv')
fhs_spouse_black <-
  fread('~/Dropbox/BioLINCC files/Framingham third generation/datasets/CSV/e_exam_ex01_2_0813d.csv')
fhs_omni2_black <-
  fread('~/Dropbox/BioLINCC files/Framingham third generation/datasets/CSV/e_exam_ex01_72_0652d.csv')

names(fhs3_dates) <- toupper(names(fhs3_dates))
names(fhs3_wkthru) <- toupper(names(fhs3_wkthru))

fhs3_melt_sex <- 
  fhs3_dates[,.(PID,IDTYPE,visit=1,variable='sex', form='fhs3_dates', value=SEX)]

fhs3_black <-
  fhs3_black[,c("PID",
                "IDTYPE",
                "G3A485")]

fhs_spouse_black <-    
  fhs_spouse_black[,
                   c("PID",
                     "IDTYPE",
                     "G3A485")]


fhs_omni2_black <-    
  fhs_omni2_black[, 
                  c("PID",
                    "IDTYPE",
                    "G3A485")]

fhs3_black_all <-
  rbindlist(
    list(fhs3_black,
        fhs_spouse_black,
        fhs_omni2_black))

fhs3_wkthru <-
  fhs3_black_all[fhs3_wkthru,
        on=.(PID,IDTYPE)]

names(fhs3_wkthru)[names(fhs3_wkthru) == "G3A485"] <- "BLACK"

for (i in 1:3) {
  fhs3_wkthru[
    , paste0("EGFR_MDRD", i) :=
      calc_MDRD4(
        dat  = as.data.frame(.SD),    # or as.data.frame(fhs2_offspring_wkthru)
        age  = paste0("AGE",   i),
        cr   = paste0("CREAT", i),
        sex  = "SEX",
        race = "BLACK",
        black = 1
      )
  ]
}


#======================================================#
####  Assemble Framingham dates and outcomes files  ####
#======================================================#


fhs_dates_fields <- names(fhs1_dates)
fhs2_dates[,fhs_dates_fields[!fhs_dates_fields %in% names(fhs2_dates)]] <- NA
fhs3_dates[,fhs_dates_fields[!fhs_dates_fields %in% names(fhs3_dates)]] <- NA


fhs_dates <- 
  rbindlist(
    list(fhs1_dates[,..fhs_dates_fields],
         fhs2_dates[,..fhs_dates_fields],
         fhs3_dates[,..fhs_dates_fields]))


fhs_dates <- fhs_survcvd[fhs_dates,
                   on=.(IDTYPE,PID)]

fhs_dates[,DATE1 := 0]


for (i in 2:20) {
  thisexamdate <- paste0("DATE", i)
  fhs_dates[
    hfhosp_status == 1 & 
      hfhosp_dt > get(thisexamdate) & 
      !is.na(hfhosp_dt) & 
      !is.na(get(thisexamdate)),
    last_exam_before_hf := i
  ]
}

age_cols <- names(fhs_dates)[grep("AGE", names(fhs_dates))]
date_cols <- names(fhs_dates)[grep("DATE", names(fhs_dates))]

col_age <- c("PID","IDTYPE","SEX",age_cols)

fhs_dates_long_age <- melt(fhs_dates[,..col_age],
                           id.vars=c("PID","IDTYPE","SEX"),
                           na.rm=T,
                           factorsAsStrings = T)

fhs_dates_long_age[,variable := as.character(variable)]

fhs_dates_long_age[,visit := 
                     substring(variable,
                               str_locate(variable,
                                          "\\d")[1],length(variable))]

fhs_dates_long_age[,variable := gsub("\\d","",variable)]

###########################3

col_dates <- 
  c("PID","IDTYPE",date_cols)

fhs_dates_long_date <- 
  melt(fhs_dates[,..col_dates],
       id.vars=c("PID","IDTYPE"),
       na.rm=T,
       factorsAsStrings = T)

fhs_dates_long_date[,variable := as.character(variable)]

fhs_dates_long_date <- fhs_dates_long_date[!variable=="CHFDATE"]

fhs_dates_long_date[,visit := 
                      substring(variable,
                                str_locate(variable,
                                           "\\d")[1],length(variable))]
                    
fhs_dates_long_date[,variable := gsub("\\d","",variable)]

names(fhs_dates_long_date)[names(fhs_dates_long_date)=="value"] <- "visitdays"
names(fhs_dates_long_age)[names(fhs_dates_long_age)=="value"] <- "age_obs"

fhs_dates_long <- 
  fhs_dates_long_date[,c("PID","IDTYPE","visit","visitdays")][
  fhs_dates_long_age[,c("PID","IDTYPE","SEX","visit","age_obs")],
                        on=.(PID,IDTYPE,visit)]

fhs_dates_long[,study := "FHS"]

names(fhs_dates_long)[2] <- "cohort"

fhs_dates_long[,visit := as.integer(visit)]
fhs_dates_long[,cohort := as.character(cohort)]

fhs_dates_long <- 
  visit_yrs[study=="FHS",
            c("cohort","study","visit","visit_yr")][fhs_dates_long,
                                                    on=.(visit,study,cohort)]


fhs_dates_long[cohort==0, cohort_name := "Original"]
fhs_dates_long[cohort==1, cohort_name := "Offspring"]
fhs_dates_long[cohort==7, cohort_name := "OMNI 1"]
fhs_dates_long[cohort==3, cohort_name := "Gen III"]
fhs_dates_long[cohort==2, cohort_name := "Offspring spouse"]
fhs_dates_long[cohort==72, cohort_name := "OMNI 2"]

fhs_dates_long[,patientid := PID]
fhs_dates_long[SEX==1, sex := "Male"]
fhs_dates_long[SEX==2, sex := "Female"]

rm(fhs_dates_long_age,
   fhs_dates_long_date)

#====================================================#
####  Breakdown Framingham Original wkthru table  ####
#====================================================#

fhs1_melt_wkthru <- melt(fhs1_wkthru,
                         id.vars=c("PID","IDTYPE","SEX"),
                         na.rm=T,
                         factorsAsStrings=T)

fhs1_melt_wkthru[,variable :=  as.character(variable)]
fhs1_melt_wkthru[,visit := 
                   substr(variable,
                          str_locate(variable,"\\d")[,1],
                          nchar(variable))]

fhs1_melt_wkthru[,variable := 
                   substr(variable,
                          1, str_locate(variable,"\\d")[,1]-1)]

fhs1_melt_wkthru[variable=="WAIST", value := value[variable=="WAIST"] * 2.54]
fhs1_melt_wkthru_kg <- fhs1_melt_wkthru[variable=="WGT"]
fhs1_melt_wkthru_kg[,value := value/2.20462]
fhs1_melt_wkthru_kg[,variable := "WGT_KG"]
fhs1_melt_wkthru <- rbind(fhs1_melt_wkthru,fhs1_melt_wkthru_kg)
fhs1_melt_wkthru[,form := "fhs1_wkthru"]


# ============================================================== #
####                  Assemble Framingham DM                  ####
# ============================================================== #


fhs1_vr_diab <- fread('~/Dropbox/BioLINCC files/Framingham cohort/datasets/CSV/VR_DIAB_EX28_0_0601D.csv',
                      na.strings=c("NA","","NULL"),
                      stringsAsFactors=F)

names(fhs1_vr_diab) <- toupper(names(fhs1_vr_diab))

fhs1_melt_vr_diab <- melt(fhs1_vr_diab[,c(1,25,82:109)],
                          id.vars=c("PID",
                                    "IDTYPE"),
                          na.rm=T,
                          factorsAsStrings=T)

fhs1_melt_vr_diab[,variable := as.character(variable)]

fhs1_melt_vr_diab[,visit := 
                    str_select(variable,
                               after="BG200_HX_DIAB")]

fhs1_melt_vr_diab[,variable := "BG200_HX_DIAB"]

fhs1_melt_vr_diab[,form := "fhs1_vr_diab"]


#### Diabetes status by cycle

fhs2_vr_diab <- 
  fread('~/Dropbox/BioLINCC files/Framingham offspring/datasets/CSV/vr_diab_ex10_1b_1489d.csv',
        na.strings=c("NA","","NULL"),
        stringsAsFactors=F)

names(fhs2_vr_diab) <- toupper(names(fhs2_vr_diab))

diab_cols <-
  c("PID",
    "IDTYPE",
    "HX_DIAB1",
    "HX_DIAB2",
    "HX_DIAB3",
    "HX_DIAB4",
    "HX_DIAB5",
    "HX_DIAB6",
    "HX_DIAB7",
    "HX_DIAB8",
    "HX_DIAB9",
    "HX_DIAB10")

fhs2_melt_vr_diab <- melt(fhs2_vr_diab[,..diab_cols],
                          id.vars=c("PID",
                                    "IDTYPE"),
                          na.rm=T,
                          factorsAsStrings=T)

fhs2_melt_vr_diab[,variable := as.character(variable)]

fhs2_melt_vr_diab[,visit := str_select(variable,
                                      after="HX_DIAB")]

fhs2_melt_vr_diab[,variable := "HX_DIAB"]

fhs2_melt_vr_diab[,form := "fhs2_vr_diab"]


####

fhs3_vr_diab <- fread('~/Dropbox/BioLINCC files/Framingham third generation/datasets/CSV/VR_DIAB_EX03_3b_1312D.csv',na.strings=c("NA","","NULL")) # Diabetes status by cycle
names(fhs3_vr_diab) <- toupper(names(fhs3_vr_diab))

fhs3_melt_vr_diab <- melt(fhs3_vr_diab[,c("PID","IDTYPE","HX_DIAB1","HX_DIAB2")],
                          id.vars=c("PID",
                                    "IDTYPE"),
                          na.rm=T,
                          factorsAsStrings=T)

fhs3_melt_vr_diab[,variable := as.character(variable)]

fhs3_melt_vr_diab[,visit := 
                    str_select(variable,after="HX_DIAB")]

fhs3_melt_vr_diab[,variable := "HX_DIAB"]

fhs3_melt_vr_diab[,form := "fhs3_vr_diab"]

## For some reason FHS Omni did not was not included in vr_diab files.  
## Rederived baseline DM status per documentation (BG > 200 or fasting BG ≥ 126 or DM tx)
## Did not do Omni1 visits 2-4 due-to missing variables. 


fhs_vr_diab <- 
  rbindlist(
    list(fhs1_melt_vr_diab,
         fhs2_melt_vr_diab,
         fhs3_melt_vr_diab))

names(fhs_vr_diab)[names(fhs_vr_diab)=="IDTYPE"] <- "cohort"

fhs_vr_diab[,cohort := as.character(cohort)]

fhs_vr_diab[,visit := as.integer(visit)]

fhs_vr_diab <- 
  fhs_dates_long[,.(PID,cohort,visit,visitdays)][fhs_vr_diab,
                     on=.(PID,cohort,visit)]

fhs_vr_diab[,study := "FHS"]

rm(fhs1_melt_vr_diab,
   fhs2_melt_vr_diab,
   fhs3_melt_vr_diab)


#=========================================================================#
#                                                                         #
####          ******** FRAMINGHAM ORIGINAL DATA ********               ####
#                                                                         #
#=========================================================================#

fhs1_ecgalld <- fread('~/Dropbox/BioLINCC files/Framingham cohort/datasets/CSV/ECG_ALLD.csv',na.strings=c("NA","","NULL"))
fhs1_ecgalld[,visit := 11]
fhs1_melt_ecgalld <- melt(fhs1_ecgalld,id.vars=c("PID","IDTYPE","visit"),na.rm=T,factorsAsStrings=T)
fhs1_melt_ecgalld[,form := "fhs1_ecgall"]

# ==================================================================##
####                    FHS original, Exam 1-7                    ####
# ==================================================================##

fhs1_ex0_7d <- fread('~/Dropbox/BioLINCC files/Framingham cohort/datasets/CSV/EX0_7D.csv',na.strings=c("NA","","NULL"))  # Exams 1-7

fhs1_ex0_7d[,height_cm := MF67*2.54]
fhs1_ex0_7d[,height_m := height_cm/100]

fhs1_ex0_7d[,MF69_kg := MF69/2.2]
fhs1_ex0_7d[,MF166_kg := MF166/2.2]
fhs1_ex0_7d[,MF180_kg := MF180/2.2]
fhs1_ex0_7d[,MF216_kg := MF216/2.2]
fhs1_ex0_7d[,MF292_kg := MF292/2.2]
fhs1_ex0_7d[,MF380_kg := MF380/2.2]
fhs1_ex0_7d[,MF470_kg := MF470/2.2]

fhs1_exam1_calc_vars <- "MF69_kg"
fhs1_exam2_calc_vars <- "MF166_kg"
fhs1_exam3_calc_vars <- "MF180_kg"
fhs1_exam4_calc_vars <- "MF216_kg"
fhs1_exam5_calc_vars <- "MF292_kg"
fhs1_exam6_calc_vars <- "MF380_kg"
fhs1_exam7_calc_vars <- "MF470_kg"

fhs1_m2 <- fhs1_ex0_7d[,.(PID,IDTYPE,height_m,height_cm)]
fhs1_m2[,height_m2 := height_m^2]

fhs1_ex0_7d[,MF78_wk := MF78/4.354]
fhs1_ex0_7d[,MF253_wk := MF253/4.354]
fhs1_ex0_7d[,MF667_wk := MF667/4.354]

fhs1_exam2_calc_vars <- c(fhs1_exam2_calc_vars,"MF78_wk")
fhs1_exam4_calc_vars <- c(fhs1_exam4_calc_vars,"MF253_wk")
fhs1_exam7_calc_vars <- c(fhs1_exam7_calc_vars,"MF667_wk")

fhs1_ex0_7d[,MF63_gdL := MF63/10]
fhs1_ex0_7d[,MF161_gdL := MF161/10]
fhs1_ex0_7d[,MF168_gdL := MF168/10]
fhs1_ex0_7d[,MF209_gdL := MF209/10]
fhs1_ex0_7d[,MF312_gdL := MF312/10]
fhs1_ex0_7d[,MF392_gdL := MF392/10]

fhs1_exam1_calc_vars <- c(fhs1_exam1_calc_vars,"MF63_gdL")
fhs1_exam2_calc_vars <- c(fhs1_exam2_calc_vars,"MF161_gdL")
fhs1_exam3_calc_vars <- c(fhs1_exam3_calc_vars,"MF168_gdL")
fhs1_exam4_calc_vars <- c(fhs1_exam4_calc_vars,"MF209_gdL")
fhs1_exam5_calc_vars <- c(fhs1_exam5_calc_vars,"MF312_gdL")
fhs1_exam6_calc_vars <- c(fhs1_exam6_calc_vars,"MF392_gdL")

fhs1_ex0_7d[,MF223_cm := MF223/10]

fhs1_exam4_calc_vars <- c(fhs1_exam4_calc_vars,"MF223_cm")

fhs1_ex0_7d[,MF302_ratio := MF302/100] # FEV1/FVC
fhs1_ex0_7d[,MF299_L := MF299/10]
fhs1_ex0_7d[,FEV1_ex5 := MF302_ratio*MF299_L]

fhs1_exam5_calc_vars <- c(fhs1_exam5_calc_vars,"FEV1_ex5")
fhs1_exam5_calc_vars <- c(fhs1_exam5_calc_vars,"MF302_ratio")
fhs1_exam5_calc_vars <- c(fhs1_exam5_calc_vars,"MF299_L")


fhs1_ex0_7d[MF452==0&!is.na(MF452), MF451_dth := MF451]

fhs1_exam6_calc_vars <- c(fhs1_exam6_calc_vars,"MF451_dth")

fhs1_ex0_7d[MF452==1&!is.na(MF452), MF451_alive := MF451]

fhs1_exam6_calc_vars <- c(fhs1_exam6_calc_vars,"MF451_alive")

fhs1_ex0_7d[MF454==0&!is.na(MF454), MF453_dth := MF453]

fhs1_exam6_calc_vars <- c(fhs1_exam6_calc_vars,"MF453_dth")

fhs1_ex0_7d[MF454==1&!is.na(MF454), MF453_olive := MF453]

fhs1_exam6_calc_vars <- c(fhs1_exam6_calc_vars,"MF453_alive")

fhs1_ex0_7d[MF20<=8&!is.na(MF20), num_preg_ex1 := MF20] 

fhs1_ex0_7d[MF120==0&
    !is.na(MF120), num_preg_ex2 := 0]
    
fhs1_ex0_7d[
  MF120 %in% c(1,2)&
    !is.na(MF120), num_preg_ex2 := 1]

fhs1_ex0_7d[
  MF237==0&!is.na(MF137), num_preg_ex4 := 0]

fhs1_ex0_7d[MF237 %in% c(1,2)&
              !is.na(MF237), 
            num_preg_ex4 := 1]

fhs1_ex0_7d[MF237 == 3&
    !is.na(MF237), num_preg_ex4 := 2]

fhs1_ex0_7d[
  MF329==0&
    !is.na(MF329), num_preg_ex5 := 0]

fhs1_ex0_7d[
  MF329 %in% c(1,2)&
    !is.na(MF329), num_preg_ex5 := 1]

fhs1_ex0_7d[, tot_pregs :=
              rowSums(.SD, na.rm = TRUE),
            .SDcols = c("num_preg_ex1",
                        "num_preg_ex2",
                        "num_preg_ex4",
                        "num_preg_ex5")]

exam1_names <- colnames(fhs1_ex0_7d)[c(1:71,73:74,540,541:542,549)]
exam1_names <- c(exam1_names,fhs1_exam1_calc_vars,"height_cm","height_m")
exam2_names <- colnames(fhs1_ex0_7d)[c(72,75:154,422:427,543,546,550)]
exam2_names <- c(exam2_names,fhs1_exam2_calc_vars)
exam3_names <- colnames(fhs1_ex0_7d)[c(155:192,428:431,544,547,551)]
exam3_names <- c(exam3_names,fhs1_exam3_calc_vars)
exam4_names <- colnames(fhs1_ex0_7d)[c(193:265,432:436,552,539,545,548,552)]
exam4_names <- c(exam4_names,fhs1_exam4_calc_vars)
exam5_names <- colnames(fhs1_ex0_7d)[c(266:350,553)]
exam5_names <- c(exam5_names,fhs1_exam5_calc_vars)
exam6_names <- colnames(fhs1_ex0_7d)[c(351:421,554)]
exam6_names <- c(exam6_names,fhs1_exam6_calc_vars)
exam7_names <- colnames(fhs1_ex0_7d)[c(437:538,555:556)]
exam7_names <- c(exam7_names,fhs1_exam7_calc_vars)

fhs1_melt_ex0_7d <- melt(fhs1_ex0_7d,id.vars=c("PID","IDTYPE"),na.rm=T,factorsAsStrings=T)
fhs1_melt_ex0_7d[,form := "fhs1_ex0_7"]

fhs1_melt_ex0_7d[variable %in% exam1_names, visit := 1]
fhs1_melt_ex0_7d[variable %in% exam2_names, visit := 2]
fhs1_melt_ex0_7d[variable %in% exam3_names, visit := 2]
fhs1_melt_ex0_7d[variable %in% exam4_names, visit := 2]
fhs1_melt_ex0_7d[variable %in% exam5_names, visit := 2]
fhs1_melt_ex0_7d[variable %in% exam6_names, visit := 2]
fhs1_melt_ex0_7d[variable %in% exam7_names, visit := 2]


rm(fhs1_ex0_7d,
   fhs1_exam1_calc_vars,
   fhs1_exam2_calc_vars,
   fhs1_exam3_calc_vars,
   fhs1_exam4_calc_vars,
   fhs1_exam5_calc_vars,
   fhs1_exam6_calc_vars,
   fhs1_exam7_calc_vars)

rm(list=ls(pattern="\\bexam\\d."))

# ==============================================================##
####                     FHS original, Exam 8                 ####
# ==============================================================##

fhs1_ex0_8d <- fread('~/Dropbox/BioLINCC files/Framingham cohort/datasets/CSV/EX0_8D_V1.csv',na.strings=c("NA","","NULL")) # Exam 8
fhs1_ex0_8d[,visit := 8]

fhs1_ex0_8d[fhs1_ex0_8d==2000] <- NA
fhs1_ex0_8d[fhs1_ex0_8d==9999] <- NA
fhs1_ex0_8d[fhs1_ex0_8d==1999] <- NA

fhs1_melt_ex0_8d <- melt(fhs1_ex0_8d,id.vars=c("PID","IDTYPE","visit"),na.rm=T,factorsAsStrings=T)
fhs1_melt_ex0_8d[,form := "fhs1_ex0_8"]

rm(fhs1_ex0_8d)

# ==============================================================##
####                     FHS original, Exam 9                 ####
# ==============================================================##

fhs1_ex0_9d <- fread('~/Dropbox/BioLINCC files/Framingham cohort/datasets/CSV/EX0_9D.csv',na.strings=c("NA","","NULL")) # Exam 9
fhs1_ex0_9d[,visit := 9]
fhs1_melt_ex0_9d <- melt(fhs1_ex0_9d,id.vars=c("PID","IDTYPE","visit"),na.rm=T,factorsAsStrings=T)
fhs1_melt_ex0_9d[,form := "fhs1_ex0_9"]

rm(fhs1_ex0_9d)


# ==============================================================#
####                    FHS original, Exam 10                ####
# ==============================================================#

fhs1_ex0_10d <- fread('~/Dropbox/BioLINCC files/Framingham cohort/datasets/CSV/EX0_10D_V1.csv',na.strings=c("NA","","NULL")) # Exam 10
fhs1_ex0_10d[,visit := 10]
fhs1_melt_ex0_10d <- melt(fhs1_ex0_10d,id.vars=c("PID","IDTYPE","visit"),na.rm=T,factorsAsStrings=T)
fhs1_melt_ex0_10d[,form := "fhs1_ex0_10"]

rm(fhs1_ex0_10d)


# ==============================================================#
####                    FHS original, Exam 11                ####
# ==============================================================#

fhs1_ex0_11d <- fread('~/Dropbox/BioLINCC files/Framingham cohort/datasets/CSV/EX0_11D.csv',na.strings=c("NA","","NULL")) # Exam 11
fhs1_ex0_11d[,visit := 11]
fhs1_ex0_11d[fhs1_ex0_11d==99] <- NA

fhs1_ex0_11d[,fhs_pai := 
  calc_framingham_pai(as.data.frame(fhs1_ex0_11d),
                      slp_hrs= "FD58",
                      sed_hrs="FD60",
                      slgt_hrs= "FD61",
                      mod_hrs="FD62",
                      hvy_hrs="FD63")]

fhs1_melt_ex0_11d <- melt(fhs1_ex0_11d,id.vars=c("PID","IDTYPE","visit"),na.rm=T,factorsAsStrings=T)
fhs1_melt_ex0_11d[,form := "fhs1_ex0_11"]

rm(fhs1_ex0_11d)


# ===============================================================#
####                    FHS original, Exam 12                 ####
# ===============================================================#

fhs1_ex0_12d <- fread('~/Dropbox/BioLINCC files/Framingham cohort/datasets/CSV/EX0_12D.csv',na.strings=c("NA","","NULL")) # Exam 12
fhs1_ex0_12d[,visit := 12]

fhs1_ex0_12d[FE127==99, FE127 := NA] 
fhs1_ex0_12d[FE128==99, FE128 := NA] 
fhs1_ex0_12d[FE129==99, FE129 := NA] 
fhs1_ex0_12d[FE186==99, FE186 := NA] 

fhs1_ex0_12d[,fhs_pai := 
  calc_framingham_pai(as.data.frame(fhs1_ex0_12d),
                      slp_hrs="FE186",
                      sed_hrs="FE187",
                      slgt_hrs="FE188",
                      mod_hrs="FE189",
                      hvy_hrs="FE190")]

fhs1_ex0_12d[,drinks_wk := FE127+FE128+FE129]

fhs1_melt_ex0_12d <- melt(fhs1_ex0_12d,id.vars=c("PID","IDTYPE","visit"),na.rm=T,factorsAsStrings=T)
fhs1_melt_ex0_12d[,form := "fhs1_ex0_12"]

rm(fhs1_ex0_12d)


# ==============================================================##
##                 FHS original, lipids exam 12                 ##
# ==============================================================##

fhs1_l_lipids_ex12 <- fread('~/Dropbox/BioLINCC files/Framingham cohort/datasets/CSV/L_LIPIDS_EX12_0_0342D.csv',na.strings=c("NA","","NULL"))  # Standard lipid panel
fhs1_l_lipids_ex12[,visit := 12]

fhs1_melt_l_lipids_ex12 <- melt(fhs1_l_lipids_ex12,id.vars=c("PID","IDTYPE","visit"),na.rm=T,factorsAsStrings=T)
fhs1_melt_l_lipids_ex12[,form := "fhs1_lipids"]

rm(fhs1_l_lipids_ex12)


# ==================================================================#
####                    FHS original, Exam 13                    ####
# ==================================================================#

fhs1_ex0_13d <- fread('~/Dropbox/BioLINCC files/Framingham cohort/datasets/CSV/EX0_13D.csv',na.strings=c("NA","","NULL")) # Exam 13
fhs1_ex0_13d[,visit := 13]

fhs1_ex0_13d[fhs1_ex0_13d=="."] <- NA

fhs1_ex0_13d[,drinks_wk := FF125+FF126+FF127]


fhs1_melt_ex0_13d <- melt(fhs1_ex0_13d,id.vars=c("PID","IDTYPE","visit"),na.rm=T,factorsAsStrings=T)
fhs1_melt_ex0_13d[,form := "fhs1_ex0_13"]

rm(fhs1_ex0_13d)


# ===============================================================#
####                    FHS original, Exam 14                 ####
# ===============================================================#

fhs1_ex0_14d <- fread('~/Dropbox/BioLINCC files/Framingham cohort/datasets/CSV/EX0_14D.csv',na.strings=c("NA","","NULL")) # Exam 14
fhs1_ex0_14d[, visit := 14]

fhs1_meno <- fread('~/Dropbox/BioLINCC files/Framingham cohort/datasets/CSV/vr_meno_ex14_0_0153d.csv',na.strings=c("NA","","NULL")) # Exam 14
fhs1_meno[, visit := 14]

fhs1_meno[AM2==0, menopause := "N"]
fhs1_meno[between(AM2,18,59), menopause := "Y"]

fhs1_ex0_14d[fhs1_ex0_14d=="."] <- NA

fhs1_ex0_14d[,drinks_wk := FG118+FG119+FG120]

fhs1_ex0_14d[,FG62_kg := FG62/2.2]

fhs1_ex0_14d <- 
  fhs1_m2[,c("PID","height_m2")][fhs1_ex0_14d,,on=.(PID)]

fhs1_ex0_14d[,FG62_bmi := FG62_kg/height_m2]


fhs1_melt_ex0_14d <- melt(fhs1_ex0_14d,id.vars=c("PID","IDTYPE","visit"),na.rm=T,factorsAsStrings=T)
fhs1_melt_ex0_14d[,form := "fhs1_ex0_14"]

rm(fhs1_ex0_14d)


# ==============================================================#
####                    FHS original, Exam 15                ####
# ==============================================================#


fhs1_ex0_15d <- fread('~/Dropbox/BioLINCC files/Framingham cohort/datasets/CSV/EX0_15D.csv',na.strings=c("NA","","NULL")) # Exam 15
fhs1_ex0_15d[,visit := 15]

fhs1_ex0_15d[fhs1_ex0_15d=="."] <- NA

fhs1_ex0_15d[,drinks_wk := FH115+FH116+FH117]


fhs1_melt_ex0_15d <- melt(fhs1_ex0_15d,id.vars=c("PID","IDTYPE","visit"),na.rm=T,factorsAsStrings=T)
fhs1_melt_ex0_15d[,form := "fhs1_ex0_15"]

rm(fhs1_ex0_15d)


# ===============================================================#
####                    FHS original, Exam 16                 ####
# ===============================================================#

fhs1_ex0_16d <-fread('~/Dropbox/BioLINCC files/Framingham cohort/datasets/CSV/EX0_16D_V2.csv',na.strings=c("NA","","NULL")) # Exam 16
fhs1_ex0_16d[,visit := 16]

fhs1_ex0_16d <- 
  fhs1_dates[,c("PID","SEX")][fhs1_ex0_16d,on=.(PID)]

fhs1_ex0_16d[,FI297_cm  := FI297/10]
fhs1_ex0_16d[,FI301_cm  := FI301/10]
fhs1_ex0_16d[,FI306_cm  := FI306/10]
fhs1_ex0_16d[,FI15_cm := FI15*2.54]
fhs1_ex0_16d[,FI15_m := FI15_cm/100]
fhs1_ex0_16d[,FI14_kg := FI14/2.2]

fhs1_ex0_16d$BSA2 <- calc_bsa(dat=as.data.frame(fhs1_ex0_16d),weight_kg = "FI14_kg",height_cm = "FI15_cm")
               # 0.20247*((FI14_kg)^0.425)*((FI15_m)^0.725)]

fhs1_ex0_16d <- 
  setDT(
    calc_hypertrophy_type(
      df=as.data.frame(fhs1_ex0_16d),
      sex="SEX",
      male="1",
      female="2",
      lvedd="FI306_cm",
      ivsd="FI297_cm",
      lvpwtd="FI301_cm",
      height="FI15_cm",
      weight="FI14_kg"))


fhs1_melt_ex0_16d <- melt(fhs1_ex0_16d,id.vars=c("PID","IDTYPE","visit"),na.rm=T,factorsAsStrings=T)
fhs1_melt_ex0_16d[, form := "fhs1_ex0_16"]

rm(fhs1_ex0_16d)


# ===============================================================#
####                    FHS original, Exam 17                 ####
# ===============================================================#

fhs1_ex0_17d <-fread('~/Dropbox/BioLINCC files/Framingham cohort/datasets/CSV/EX0_17D.csv',na.strings=c("NA","","NULL")) # Exam 17
fhs1_ex0_17d[,visit := 17]

fhs1_ex0_17d[,drinks_wk := FJ59+FJ60+FJ61]


####### FRAIL score

## Fatigue, Resistance, Ambulation encoded in 'criteriaset'

fhs1_ex0_17d[FJ191==1|FJ192==1, fatigue_frail := 1]
fhs1_ex0_17d[FJ191 %in% c(0,2)|FJ192 %in% c(0,2), fatigue_frail := 0]

## Resistance

fhs1_ex0_17d[FJ175==1, resistance_frail := 0]
fhs1_ex0_17d[FJ175==0, resistance_frail := 1]

## Ambulate

fhs1_ex0_17d[FJ176==1, ambulate_frail := 0]
fhs1_ex0_17d[FJ176==0, ambulate_frail := 1]

# FRAIL Illness score counts presence of HTN, DM, cancer, chronic lung disease, MI, CHD, angina, asthma, arthritis, stroke, and ckd
# Scores for 0-4 conditions, score = 0, for ≥ 5, score=1

fhs1_ex0_17d <- fhs_dates[,c("PID","IDTYPE","DATE17")][fhs1_ex0_17d,on=.(PID,IDTYPE)]
fhs1_ex0_17d <- fhs_outcomes[fhs1_ex0_17d,on=.(PID,IDTYPE)]
fhs1_ex0_17d <- fhs_survca_first_any[,c("PID","IDTYPE","D_DATE")][fhs1_ex0_17d,on=.(PID,IDTYPE)]
fhs1_ex0_17d <- fhs1_wkthru[,c("PID","IDTYPE","WGT16","WGT17")][fhs1_ex0_17d,on=.(PID,IDTYPE)]

# DM

fhs1_ex0_17d <- 
  fhs_diab[,c("PID","BG200_HX_DIAB17")][fhs1_ex0_17d,
                                        on=.(PID)]

fhs1_ex0_17d[,dm_frail := BG200_HX_DIAB17]

# HTN

fhs1_ex0_17d[FJ318==0, htn_frail := 0]
fhs1_ex0_17d[FJ318==1, htn_frail := 1]

###------------###

# COPD (chronic lung disease)

fhs1_ex0_17d[FJ348==0, copd_frail := 0]
fhs1_ex0_17d[FJ348==1, copd_frail := 1]


###------------###


# Asthma

fhs1_ex0_17d[FJ71==0, asthma_frail := 0]
fhs1_ex0_17d[FJ71==1, asthma_frail := 1]

###------------###

# DJD (arthritis)

fhs1_ex0_17d[FJ351==0, djd_frail := 0]
fhs1_ex0_17d[FJ351==1, djd_frail := 1]


###------------###

# CKD (renal disease)


fhs1_ex0_17d[FJ346==0, ckd_frail := 0]
fhs1_ex0_17d[FJ346==1, ckd_frail := 1]

###------------###

# CHF

fhs1_ex0_17d[hfhosp_status==0|(hfhosp_status==1&DATE17 < hfhosp_dt), chf_frail := 0]
fhs1_ex0_17d[hfhosp_status==1&DATE17 >= hfhosp_dt, chf_frail := 1]

###------------###

# CAD/MI

fhs1_ex0_17d[cadhosp_status==0|(cadhosp_status==1&DATE17 < cadhosp_dt), chd_frail := 0]
fhs1_ex0_17d[cadhosp_status==1&DATE17 >= cadhosp_dt, chd_frail := 1]

###------------###


# Stroke

fhs1_ex0_17d[cvahosp_status==0|(cvahosp_status==1&DATE17 < cvahosp_dt), stroke_frail := 0]
fhs1_ex0_17d[cvahosp_status==1&cvahosp_dt <= DATE17, stroke_frail := 1]


###------------###

# Cancer

fhs1_ex0_17d[,cancer_frail := 0]
fhs1_ex0_17d[D_DATE <= DATE17, cancer_frail := 1]

###------------###

# Compile chronic illness

fhs1_ex0_17d[,conditions_frail := 
  dm_frail+
  htn_frail+
  copd_frail+
  asthma_frail+
  djd_frail+
  ckd_frail+
  chf_frail+
  chd_frail+
  stroke_frail+
  cancer_frail]

fhs1_ex0_17d[conditions_frail <= 4, illness_frail := 0]
fhs1_ex0_17d[conditions_frail > 4, illness_frail := 1]

fhs1_ex0_17d[WGT17-WGT16 <= -15, wtloss_frail := 1]
fhs1_ex0_17d[WGT17-WGT16 > -15, wtloss_frail := 0]

fhs1_ex0_17d[,total_frail := 
  fatigue_frail+
  resistance_frail+
  ambulate_frail+
  illness_frail+
  wtloss_frail]

#### ADL mod

fhs1_ex0_17d[FJ177 > 3, FJ177 := 0]
fhs1_ex0_17d[FJ178 > 3, FJ178 := 0]
fhs1_ex0_17d[FJ179 > 3, FJ179 := 0]
fhs1_ex0_17d[FJ180 > 3, FJ180 := 0]
fhs1_ex0_17d[FJ181 > 3, FJ181 := 0]
fhs1_ex0_17d[FJ182 > 3, FJ182 := 0]


fhs1_melt_ex0_17d <- melt(fhs1_ex0_17d,id.vars=c("PID","IDTYPE","visit"),na.rm=T,factorsAsStrings=T)
fhs1_melt_ex0_17d[,form := "fhs1_ex0_17"]

rm(fhs1_ex0_17d)

# ==============================================================#
####                    FHS original, Exam 18                ####
# ==============================================================#

fhs1_ex0_18d <-fread('~/Dropbox/BioLINCC files/Framingham cohort/datasets/CSV/EX0_18D_V2.csv',na.strings=c("NA","","NULL")) # Exam 18
fhs1_ex0_18d[,visit := 18]
fhs1_melt_ex0_18d <- melt(fhs1_ex0_18d,id.vars=c("PID","IDTYPE","visit"),na.rm=T,factorsAsStrings=T)
fhs1_melt_ex0_18d[,form := "fhs1_ex0_18"]

rm(fhs1_ex0_18d)


# ==============================================================#
####                    FHS original, Exam 19                ####
# ==============================================================#

fhs1_ex0_19d <-fread('~/Dropbox/BioLINCC files/Framingham cohort/datasets/CSV/EX0_19D.csv',na.strings=c("NA","","NULL")) # Exam 19
fhs1_ex0_19d[,visit := 19]

fhs1_ex0_19d[,fhs_pai := 
               calc_framingham_pai(
                 as.data.frame(fhs1_ex0_19d),
                 slp_hrs="FL089",
                 sed_hrs="FL090",
                 slgt_hrs="FL091",
                 mod_hrs="FL092",
                 hvy_hrs="FL093")]


fhs1_melt_ex0_19d <- melt(fhs1_ex0_19d,id.vars=c("PID","IDTYPE","visit"),na.rm=T,factorsAsStrings=T)
fhs1_melt_ex0_19d[,form := "fhs1_ex0_19"]

rm(fhs1_ex0_19d)


# ===============================================================#
####                  FHS original, Exam 20                   ####
# ===============================================================#

fhs1_ex0_20d <-fread('~/Dropbox/BioLINCC files/Framingham cohort/datasets/CSV/EX0_20D_V1.csv',na.strings=c("NA","","NULL")) # Exam 20
fhs1_ex0_20d[,visit := 20]

fhs1_ex0_20d[,fhs_pai := 
  calc_framingham_pai(
    as.data.frame(fhs1_ex0_20d),
    slp_hrs="FM94",
    sed_hrs="FM95",
    slgt_hrs="FM96",
    mod_hrs="FM97",
    hvy_hrs="FM98")]

fhs1_meno <- 
  fhs1_ex0_20d[,c("PID","FM201")][fhs1_meno, on=.(PID)]


fhs1_melt_ex0_20d <- melt(fhs1_ex0_20d,id.vars=c("PID","IDTYPE","visit"),na.rm=T,factorsAsStrings=T)
fhs1_melt_ex0_20d[,form := "fhs1_ex0_20"]

rm(fhs1_ex0_20d)


# ==============================================================#
#                     FHS original, Lab exam 20                 #
# ==============================================================#

fhs1_l_fhslab1_ex20 <-fread('~/Dropbox/BioLINCC files/Framingham cohort/datasets/CSV/L_FHSLAB1_EX20_0494D.csv',na.strings=c("NA","","NULL")) # Standard labs
fhs1_l_fhslab1_ex20[,visit := 20]

fhs1_ex20_demo <- 
  fhs1_wkthru[,.(PID,
                 IDTYPE,
                 SEX,
                 AGE20)]

fhs1_l_fhslab1_ex20 <- 
  fhs1_ex20_demo[fhs1_l_fhslab1_ex20,
        on=.(PID,IDTYPE)]

fhs1_l_fhslab1_ex20[,race := 1]

fhs1_l_fhslab1_ex20[,gfr_mdrd := 
                      calc_MDRD4(dat=as.data.frame(fhs1_l_fhslab1_ex20),
                                           cr="CREAT",
                                           age="AGE20",
                                           sex="SEX",
                                           race="race")]

fhs1_melt_l_fhslab1_ex20 <- melt(fhs1_l_fhslab1_ex20,id.vars=c("PID","IDTYPE","visit"),na.rm=T,factorsAsStrings=T)
fhs1_melt_l_fhslab1_ex20[,form := "fhs1_fhslab1"]

rm(fhs1_l_fhslab1_ex20)


#================================================================##
#             FHS original, echo with doppler, exam 20            #
#================================================================##

fhs1_t_echo_ex20 <- fread('~/Dropbox/BioLINCC files/Framingham cohort/datasets/CSV/T_ECHO_EX20_0_0576D.csv',na.strings=c("NA","","NULL")) # Echo dimensions
fhs1_t_echo_ex20[,visit := 20]

fhs1_t_echo_ex20 <- 
  fhs1_wkthru[,c("PID","SEX","HGT1","WGT20")][fhs1_t_echo_ex20, on = .(PID)]

fhs1_t_echo_ex20[,height_cm := HGT1*2.54]
fhs1_t_echo_ex20[,weight_kg := WGT20/2.2]

fhs1_t_echo_ex20 <- 
  setDT(
    calc_hypertrophy_type(
      df=as.data.frame(fhs1_t_echo_ex20),
      sex="SEX",
      male="1",
      female="2",
      lvedd="LVDD20",
      ivsd="IVSD20",
      lvpwtd="LVPD20",
      height="height_cm",
      weight="weight_kg"))


fhs1_melt_t_echo_ex20 <- melt(fhs1_t_echo_ex20,id.vars=c("PID","IDTYPE","visit"),na.rm=T,factorsAsStrings=T)
fhs1_melt_t_echo_ex20[,form := "fhs1_echo_ex20"]


fhs1_t_echodop_ex20 <- fread('~/Dropbox/BioLINCC files/Framingham cohort/datasets/CSV/T_ECHODOP_EX20_0_0886D.csv',na.strings=c("NA","","NULL")) # Echo doppler
fhs1_t_echodop_ex20[,visit := 20]
fhs1_t_echodop_ex20[,DEC_MM_msec := DEC_MM*1000]

fhs1_melt_t_echodop_ex20 <- melt(fhs1_t_echodop_ex20,id.vars=c("PID","IDTYPE","visit"),na.rm=T,factorsAsStrings=T)

fhs1_melt_t_echodop_ex20[,form := "fhs1_echodopp_ex20"]


rm(fhs1_t_echo_ex20,
   fhs1_t_echodop_ex20)

# ==================================================================#
#              FHS original, HR variability, exam 20                #
# ==================================================================#

fhs1_t_hrv_ex20 <- fread('~/Dropbox/BioLINCC files/Framingham cohort/datasets/CSV/T_HRV_1987_M_0118D.csv',na.strings=c("NA","","NULL")) # Heart rate variability
fhs1_t_hrv_ex20[,visit := 20]
fhs1_melt_t_hrv_ex20 <- melt(fhs1_t_hrv_ex20,id.vars=c("PID","IDTYPE","visit"),na.rm=T,factorsAsStrings=T)
fhs1_melt_t_hrv_ex20[,form := "fhs1_hrv"]

rm(fhs1_t_hrv_ex20)


#=============================================#
#     FHS original food frequency exam 20     #
#=============================================#

fhs1_vr_ffreq_ex20 <- fread('~/Dropbox/BioLINCC files/Framingham cohort/datasets/CSV/VR_FFREQ_EX20_0_0572D_v1.csv',na.strings=c("NA","","NULL")) # Food intake exam 20
fhs1_vr_ffreq_ex20[,visit := 20]
fhs1_vr_ffreq_ex20[,SCORE33_wk := SCORE33/14*7]
fhs1_melt_vr_ffreq_ex20 <- melt(fhs1_vr_ffreq_ex20,id.vars=c("PID","IDTYPE","visit"),na.rm=T,factorsAsStrings=T)
fhs1_melt_vr_ffreq_ex20[,form := "fhs1_vr_ffreq"]

rm(fhs1_vr_ffreq_ex20)


# ==============================================================#
####                    FHS original, Exam 21                ####
# ==============================================================#

fhs1_ex0_21d <-fread('~/Dropbox/BioLINCC files/Framingham cohort/datasets/CSV/EX0_21D_V2.csv',na.strings=c("NA","","NULL")) # Exam 21
fhs1_ex0_21d[,visit := 21]

fhs1_ex0_21d[!between(FN150,23,87), FN150 := NA]

fhs1_meno <- 
  fhs1_ex0_21d[,c("PID","FN151")][fhs1_meno,
                                  on=.(PID)]

fhs1_melt_ex0_21d <- melt(fhs1_ex0_21d,id.vars=c("PID","IDTYPE","visit"),na.rm=T,factorsAsStrings=T)
fhs1_melt_ex0_21d[,form := "fhs1_ex0_21"]

rm(fhs1_ex0_21d)


#=============================================#
#     FHS original food frequency exam 21     #
#=============================================#

fhs1_vr_ffreq_ex21 <- fread('~/Dropbox/BioLINCC files/Framingham cohort/datasets/CSV/VR_FFREQ_EX21_0_0573D_v1.csv',na.strings=c("NA","","NULL")) # Food intake exam 21
fhs1_vr_ffreq_ex21[,visit := 21]
fhs1_vr_ffreq_ex21[,SCORE33_wk := SCORE33/14*7]
fhs1_melt_vr_ffreq_ex21 <- melt(fhs1_vr_ffreq_ex21,id.vars=c("PID","IDTYPE","visit"),na.rm=T,factorsAsStrings=T)
fhs1_melt_vr_ffreq_ex21[,form := "fhs1_vr_ffreq"]

rm(fhs1_vr_ffreq_ex21)


# ==============================================================#
####                    FHS original, Exam 22                ####
# ==============================================================#

fhs1_ex0_22d <-fread('~/Dropbox/BioLINCC files/Framingham cohort/datasets/CSV/EX0_22D_V3.csv',na.strings=c("NA","","NULL")) # Exam 21
fhs1_ex0_22d[,visit := 22]

fhs1_meno <- 
  fhs1_ex0_22d[,c("PID","FO149")][fhs1_meno,
        on=.(PID)]

fhs1_meno[,ovaries_removed :=
  apply(fhs1_meno[,c("AM4","FM201","FN151","FO149")],
        MARGIN=1,
        FUN=function(x) max(x, na.rm=T))]

fhs1_meno[ovaries_removed %in% c("-Inf","7"), ovaries_removed := NA]

fhs1_ex0_22d[,CESD20_missing := rowSums(is.na(fhs1_ex0_22d[,c(paste0("FO0",47:66))]))]
fhs1_ex0_22d[,CESD20 := rowSums(fhs1_ex0_22d[,c(paste0("FO0",47:66))])]
fhs1_ex0_22d[CESD20_missing >= 4,CESD20 := NA]

fhs1_ex0_22d[,barthel_hygiene := 0]

fhs1_ex0_22d[!between(FO148,20,87), FO148 := NA]
fhs1_ex0_22d[!between(FO150,0,9), FO150 := NA]


fhs1_melt_ex0_22d <- melt(fhs1_ex0_22d,id.vars=c("PID","IDTYPE","visit"),na.rm=T,factorsAsStrings=T)
fhs1_melt_ex0_22d[,form := "fhs1_ex0_22"]

fhs1_melt_meno <- melt(fhs1_meno,id.vars=c("PID","IDTYPE","visit"),na.rm=T,factorsAsStrings=T)
fhs1_melt_meno[,form := "fhs1_meno"]


rm(fhs1_ex0_22d)

#=============================================#
#    FHS original food frequency exam 22     #
#=============================================#

fhs1_vr_ffreq_ex22 <- fread('~/Dropbox/BioLINCC files/Framingham cohort/datasets/CSV/VR_FFREQ_EX22_0_0574D_v1.csv',na.strings=c("NA","","NULL")) # Food intake exam 22
fhs1_vr_ffreq_ex22[,visit := 22]
fhs1_vr_ffreq_ex22[,SCORE33_wk := SCORE33/14*7]
fhs1_melt_vr_ffreq_ex22 <- melt(fhs1_vr_ffreq_ex22,id.vars=c("PID","IDTYPE","visit"),na.rm=T,factorsAsStrings=T)
fhs1_melt_vr_ffreq_ex22[,form := "fhs1_vr_ffreq"]

rm(fhs1_vr_ffreq_ex22)


# ==============================================================#
####                   FHS original, Exam 23                 ####
# ==============================================================#

fhs1_ex0_23d <-fread('~/Dropbox/BioLINCC files/Framingham cohort/datasets/CSV/EX0_23D_V3.csv',na.strings=c("NA","","NULL")) # Exam 23
fhs1_ex0_23d[,visit := 23]


fhs1_ex0_23d[,CESD20_missing := rowSums(is.na(fhs1_ex0_23d[,c(paste0("FP",499:518))]))]
fhs1_ex0_23d[,CESD20 := 
               rowSums(
                 fhs1_ex0_23d[,
                              c(
                                paste0("FP",
                                       499:518
                                       )
                                )
                              ]
                 )
             ]

fhs1_ex0_23d[CESD20_missing >= 4, CESD20 := NA]

fhs1_ex0_23d[,barthel_hygiene := 0]


fhs1_melt_ex0_23d <- melt(fhs1_ex0_23d,id.vars=c("PID","IDTYPE","visit"),na.rm=T,factorsAsStrings=T)
fhs1_melt_ex0_23d[,form := "fhs1_ex0_23"]

rm(fhs1_ex0_23d)


# ==============================================================#
####                    FHS original, Exam 24                ####
# ==============================================================#

fhs1_ex0_24d <-fread('~/Dropbox/BioLINCC files/Framingham cohort/datasets/CSV/EX0_24D_V2.csv',na.strings=c("NA","","NULL")) # Exam 24
fhs1_ex0_24d[,visit := 24]

fhs1_ex0_24d[,barthel_hygiene := 0]


fhs1_melt_ex0_24d <- melt(fhs1_ex0_24d,id.vars=c("PID","IDTYPE","visit"),na.rm=T,factorsAsStrings=T)
fhs1_melt_ex0_24d[,form := "fhs1_ex0_24"]

rm(fhs1_ex0_24d)


# ==============================================================#
####                 FHS original, Exam 25                   ####
# ==============================================================#

fhs1_ex0_25d <-fread('~/Dropbox/BioLINCC files/Framingham cohort/datasets/CSV/EX0_25D_V1.csv',na.strings=c("NA","","NULL")) # Exam 25
fhs1_ex0_25d[,visit := 25]

fhs1_ex0_25d[,CESD20_missing := rowSums(is.na(fhs1_ex0_25d[,c(paste0("FR",140:159))]))]
fhs1_ex0_25d[,CESD20 := rowSums(fhs1_ex0_25d[,c(paste0("FR",140:159))])]
fhs1_ex0_25d[CESD20_missing >= 4, CESD20 := NA]

fhs1_ex0_25d[,barthel_hygiene := 0]
fhs1_ex0_25d[,essi_chores := 0]

fhs1_melt_ex0_25d <- melt(fhs1_ex0_25d,id.vars=c("PID","IDTYPE","visit"),na.rm=T,factorsAsStrings=T)
fhs1_melt_ex0_25d[,form := "fhs1_ex0_25"]

rm(fhs1_ex0_25d)


# ==============================================================#
####                    FHS original, Exam 26                ####
# ==============================================================#

## Uses method described here https://www.ncbi.nlm.nih.gov/pmc/articles/PMC5005887/

fhs1_ex0_26d <-fread('~/Dropbox/BioLINCC files/Framingham cohort/datasets/CSV/EX0_26D_V1.csv',na.strings=c("NA","","NULL")) # Exam 26
fhs1_ex0_26d[,visit := 26]
fhs1_ex0_26d[fhs1_ex0_26d==99] <- NA

fhs1_ex0_26d[,CESD20_missing := rowSums(is.na(fhs1_ex0_26d[,c(paste0("FS",157:176))]))]
fhs1_ex0_26d[,CESD20 := rowSums(fhs1_ex0_26d[,c(paste0("FS",157:176))])]
fhs1_ex0_26d[CESD20_missing >= 4,CESD20 := NA]

fhs1_ex0_26d[,barthel_hygiene := 0]
fhs1_ex0_26d[,essi_chores := 0]


fhs1_ex0_26d <- 
  fhs1_wkthru[,.(PID,
                 SEX,
                 AGE26,
                 HGT1,
                 BMIC26,
                 DATE26,
                 WGT26,
                 WGT25,
                 DMRX26,
                 HRX26,
                 CREAT26)][fhs1_ex0_26d,
                             on=.(PID)]

fhs1_ex0_26d[,essi_chores := 0]


##### Walking speed (sec/15 ft)

fhs1_ex0_26d[,walktime_1 := FS213+FS214/100]
fhs1_ex0_26d[,walktime_2 := FS216+FS217/100]

fhs1_ex0_26d[,walktime_avg := round(apply(fhs1_ex0_26d[,c("walktime_1","walktime_2")],MARGIN=1,mean,na.rm=T),3)]
fhs1_ex0_26d[walktime_avg=="NaN",walktime_avg := NA]
fhs1_ex0_26d[,walktime_min := round(apply(fhs1_ex0_26d[,c("walktime_1","walktime_2")],MARGIN=1,min,na.rm=T),3)]
fhs1_ex0_26d[walktime_min=="-Inf",walktime_min := NA]


########### Grip (kg)

# Produces highest average by hand (assumes dominant hand is stronger)

fhs1_ex0_26d[,grip_avg_right := round(apply(fhs1_ex0_26d[,.(FS191,FS192,FS193)],MARGIN=1,mean),1)]
fhs1_ex0_26d[,grip_avg_left := round(apply(fhs1_ex0_26d[,.(FS194,FS195,FS196)],MARGIN=1,mean),1)]

fhs1_ex0_26d[,grip_max := 
  round(
    apply(
      fhs1_ex0_26d[,c("grip_avg_right","grip_avg_left")],
      MARGIN=1,max),1)]


##### Exhaustion

fhs1_ex0_26d[FS163==3|FS176==3, exhaustion_fried := 1]
fhs1_ex0_26d[FS163<3&FS176<3, exhaustion_fried := 0]

fhs1_ex0_26d[FS163==3|FS176==3, exhaustion_fried := 1]
fhs1_ex0_26d[FS163<3&FS176<3, exhaustion_fried := 0]


#### (Weight) Loss

fhs1_ex0_26d[(WGT26-WGT25) >= -10, wtloss_fried := 0]
fhs1_ex0_26d[(WGT26-WGT25) < -10, wtloss_fried := 1]


######### FRAIL Score


##### Fatigue

fhs1_ex0_26d[FS163 >=2|FS176 >= 2, fatigue_frail := 1]
fhs1_ex0_26d[FS163<2&FS176<2, fatigue_frail := 0]


#### Resistance

fhs1_ex0_26d[FS072==0, resistance_frail := 0]
fhs1_ex0_26d[FS072 %in% c(1,2,3), resistance_frail := 1]

#### Ambulation

fhs1_ex0_26d[FS071==1, ambulate_frail := 0]
fhs1_ex0_26d[FS071 %in% c(0,2), ambulate_frail := 1]


# FRAIL Illness score counts presence of HTN, DM, cancer, chronic lung disease, MI, CHD, angina, asthma, arthritis, stroke, and ckd
# Scores for 0-4 conditions, score = 0, for ≥ 5, score=1


# COPD (chronic lung disease)

fhs1_ex0_26d[FS522==2, copd_frail := 0]
fhs1_ex0_26d[FS522==1, copd_frail := 1]
fhs1_ex0_26d[FS522==0, copd_frail := 0]

# asthma

fhs1_ex0_26d[FS525==2, asthma_frail := 0]
fhs1_ex0_26d[FS525==1, asthma_frail := 1]
fhs1_ex0_26d[FS525==0, asthma_frail := 0]

# DJD (arthritis)

fhs1_ex0_26d[FS528==2, djd_frail := 0]
fhs1_ex0_26d[FS528==1, djd_frail := 1]
fhs1_ex0_26d[FS528==0, djd_frail := 0]

# CKD (renal disease)

fhs1_ex0_26d[,RACE := 1]
fhs1_ex0_26d[,crcl := calc_MDRD4(as.data.frame(fhs1_ex0_26d),cr="CREAT26",age="AGE26")]

fhs1_ex0_26d[FS521==2, ckd_frail := 0]
fhs1_ex0_26d[FS521==1|crcl < 60, ckd_frail := 1]
fhs1_ex0_26d[FS521==0, ckd_frail := 0]

# CHF/CAD

fhs1_ex0_26d <- fhs_outcomes[fhs1_ex0_26d,on=.(PID,IDTYPE)]

fhs1_ex0_26d[,chf_frail := 0]
fhs1_ex0_26d[hfhosp_status==1&hfhosp_dt <= DATE26, chf_frail := 1]

fhs1_ex0_26d[,chd_frail <- 0]
fhs1_ex0_26d[cadhosp_status==1&cadhosp_dt <= DATE26, chd_frail := 1]

# Stroke

fhs1_ex0_26d[,stroke_frail := 0]
fhs1_ex0_26d[cvahosp_status==1&cvahosp_dt <= DATE26, stroke_frail := 1]

# Cancer

fhs1_ex0_26d <- 
  fhs_survca_first_any[,c("PID","IDTYPE","D_DATE")][
    fhs1_ex0_26d,
    on=.(PID,IDTYPE)]

fhs1_ex0_26d[,cancer_frail := 0]
fhs1_ex0_26d[D_DATE <= DATE26, cancer_frail := 1]

fhs1_ex0_26d[,conditions_frail := 
  HRX26+
  DMRX26+
  copd_frail+
  asthma_frail+
  djd_frail+
  ckd_frail+
  chf_frail+
  chd_frail+
  stroke_frail+
  cancer_frail]

fhs1_ex0_26d[conditions_frail <= 4, illness_frail := 0]
fhs1_ex0_26d[conditions_frail > 4, illness_frail := 1]


#### (Weight) Loss

fhs1_ex0_26d[,wgt26_25_delta := WGT26-WGT25]


fhs1_ex0_26d[(WGT26-WGT25) > -10, wtloss_frail := 0]
fhs1_ex0_26d[(WGT26-WGT25) <= -10, wtloss_frail := 1]


#### Calculate FRAIL score

fhs1_ex0_26d[,total_frail := 
  fatigue_frail+
  resistance_frail+
  ambulate_frail+
  illness_frail+
  wtloss_frail]

fhs1_ex0_26d <- fhs1_ex0_26d[-c(354,356),]

fhs1_melt_ex0_26d <- melt(fhs1_ex0_26d,id.vars=c("PID","IDTYPE","visit"),na.rm=T,factorsAsStrings=T)
fhs1_melt_ex0_26d[,form := "fhs1_ex0_26"]

rm(fhs1_ex0_26d)


# ==============================================================#
####                  FHS original, Exam 27                  ####
# ==============================================================#

fhs1_ex0_27d <-fread('~/Dropbox/BioLINCC files/Framingham cohort/datasets/CSV/E_EXAM_EX27_0_0075D.csv',na.strings=c("NA","","NULL")) # Exam 27
fhs1_ex0_27d[,visit := 27]
fhs1_ex0_27d <- 
  fhs1_wkthru[,.(PID,
                 SEX,
                 HGT1,
                 DATE27,
                 BMIC27,
                 WGT27,
                 WGT26,
                 DMRX27,
                 HRX27,
                 CREAT27,
                 AGE27)][fhs1_ex0_27d,
                         on=.(PID)]

fhs1_ex0_27d[,essi_chores := 0]

## Walking speed

fhs1_ex0_27d[,walktime_1 := FT225+FT226/100]
fhs1_ex0_27d[,walktime_2 := FT229+FT230/100]

fhs1_ex0_27d[,walktime_avg := 
               round(
                 apply(
                   fhs1_ex0_27d[,.(walktime_1,walktime_2)],
                   MARGIN=1,
                   mean,na.rm=T),3)]

fhs1_ex0_27d[walktime_avg=="NaN", walktime_avg := NA]
fhs1_ex0_27d[,walktime_min := 
               round(
                 apply(
                   fhs1_ex0_27d[,c("walktime_1","walktime_2")],
                   MARGIN=1,min,na.rm=T),3)]

fhs1_ex0_27d[walktime_min=="-Inf", walktime_min := NA]


########### Grip (kg)

# Produces highest average by hand (assumes dominant hand is stronger)

fhs1_ex0_27d[,grip_avg_right := round(apply(fhs1_ex0_27d[,.(FT193,FT194,FT195)],MARGIN=1,mean),1)]
fhs1_ex0_27d[,grip_avg_left := round(apply(fhs1_ex0_27d[,.(FT196,FT197,FT198)],MARGIN=1,mean),1)]

fhs1_ex0_27d[,grip_max := 
               round(
                 apply(
                   fhs1_ex0_27d[,c("grip_avg_right","grip_avg_left")],
                   MARGIN=1,max),1)]


#### Exhaustion

fhs1_ex0_27d[FT032==3|FT045==3, exhaustion_fried := 1]
fhs1_ex0_27d[FT032<3&FT045<3, exhaustion_fried := 0]


#### (Weight) Loss

fhs1_ex0_27d[(WGT27-WGT26) <= - 10, wtloss_fried := 1]
fhs1_ex0_27d[(WGT27-WGT26) > 10, wtloss_fried := 0]


######### FRAIL Score

#### Fatigue

fhs1_ex0_27d[FT032==3|FT045==3, fatigue_frail := 1]
fhs1_ex0_27d[FT032<3&FT045<3, fatigue_frail := 0]

#### Resistance

fhs1_ex0_27d[FT095==0, resistance_frail := 0]
fhs1_ex0_27d[FT095 %in% c(1,2,3), resistance_frail := 1]

#### Ambulation

fhs1_ex0_27d[FT136==1, ambulate_frail := 0]
fhs1_ex0_27d[FT136 %in% c(0,2), ambulate_frail := 1]


# FRAIL Illness score counts presence of HTN, DM, cancer, chronic lung disease, MI, CHD, angina, asthma, arthritis, stroke, and ckd
# Scores for 0-4 conditions, score = 0, for ≥ 5, score=1


# COPD (chronic lung disease)

fhs1_ex0_27d[FT527==2, copd_frail := 0]
fhs1_ex0_27d[FT527==1, copd_frail := 1]
fhs1_ex0_27d[FT527==0, copd_frail := 0]

# asthma

fhs1_ex0_27d[FT530==2, asthma_frail := 0]
fhs1_ex0_27d[FT530==1, asthma_frail := 1]
fhs1_ex0_27d[FT530==0, asthma_frail := 0]

# DJD (arthritis)

fhs1_ex0_27d[FT533==2, djd_frail := 0]
fhs1_ex0_27d[FT533==1, djd_frail := 1]
fhs1_ex0_27d[FT533==0, djd_frail := 0]

# CKD (renal disease)

fhs1_ex0_27d[,RACE := 1]
fhs1_ex0_27d[,crcl := 
               calc_MDRD4( dat= (.SD),
                          cr="CREAT27",
                          age="AGE27")]


fhs1_ex0_27d[FT526==2, ckd_frail := 0]
fhs1_ex0_27d[FT526==1|crcl < 60, ckd_frail := 1]
fhs1_ex0_27d[FT526==0, ckd_frail := 0]

# CHF/MI

fhs1_ex0_27d <- fhs_outcomes[fhs1_ex0_27d,on=.(PID,IDTYPE)]

fhs1_ex0_27d[,chf_frail := 0]
fhs1_ex0_27d[hfhosp_status==1&hfhosp_dt <= DATE27, chf_frail := 1]

fhs1_ex0_27d[,chd_frail := 0]
fhs1_ex0_27d[hfhosp_status==1&hfhosp_dt <= DATE27, chd_frail := 1]

# Stroke

fhs1_ex0_27d[,stroke_frail := 0]
fhs1_ex0_27d[cvahosp_status==1&cvahosp_dt <= DATE27, stroke_fail := 1]

# Cancer

fhs1_ex0_27d <- 
  fhs_survca_first_any[,c("PID","IDTYPE","D_DATE")][fhs1_ex0_27d,
                                           on=.(PID,IDTYPE)]

fhs1_ex0_27d[,cancer_frail := 0]
fhs1_ex0_27d[D_DATE <= DATE27, cancer_frail := 1]

fhs1_ex0_27d[,conditions_frail := 
  HRX27+
  DMRX27+
  copd_frail+
  asthma_frail+
  djd_frail+
  ckd_frail+
  chf_frail+
  chd_frail+
  stroke_frail+
  cancer_frail]

fhs1_ex0_27d[conditions_frail <= 4, illness_frail := 0]
fhs1_ex0_27d[conditions_frail > 4, illness_frail := 1]


#### (Weight) Loss

fhs1_ex0_27d[,wgt27_26_delta := WGT27-WGT26]

fhs1_ex0_27d[(WGT27-WGT26) >= 10, wtloss_frail := 1]
fhs1_ex0_27d[(WGT27-WGT26) < 10, wtloss_frail := 0]


#### Calculate FRAIL score

fhs1_ex0_27d[,total_frail := 
  fatigue_frail +
  resistance_frail+
  ambulate_frail+
  illness_frail+
  wtloss_frail]

fhs1_ex0_27d[,barthel_hygiene := 0]


fhs1_melt_ex0_27d <- melt(fhs1_ex0_27d,id.vars=c("PID","IDTYPE","visit"),na.rm=T,factorsAsStrings=T)
fhs1_melt_ex0_27d[,form := "fhs1_ex0_27"]

rm(fhs1_ex0_27d)


# ==============================================================#
####                    FHS original, Exam 28                ####
# ==============================================================#

fhs1_ex0_28d <-
  fread('~/Dropbox/BioLINCC files/Framingham cohort/datasets/CSV/E_EXAM_EX28_0_0256D.csv',
        na.strings=c("NA","","NULL")) # Exam 28

fhs1_ex0_28d[,visit := 28]
fhs1_ex0_28d <- 
  fhs1_wkthru[,c("PID",
                 "DATE28",
                 "SEX",
                 "HGT1",
                 "BMIC28",
                 "WGT28",
                 "WGT27",
                 "DMRX28",
                 "HRX28",
                 "AGE28",
                 "CREAT28")][fhs1_ex0_28d,on=.(PID)]


fhs1_ex0_28d[,barthel_hygiene := 0]
fhs1_ex0_28d[,essi_chores := 0]

###### Walking speed

fhs1_ex0_28d[,walktime_1 := FU542]
fhs1_ex0_28d[,walktime_2 := FU545]

fhs1_ex0_28d[,walktime_avg := 
               round(
                 apply(
                   fhs1_ex0_28d[,c("walktime_1","walktime_2")],
                   MARGIN=1,mean,na.rm=T),3)]

fhs1_ex0_28d[walktime_avg=="NaN", walktime_avg := NA]
fhs1_ex0_28d[,walktime_min := 
               round(
                 apply(
                   fhs1_ex0_28d[,c("walktime_1","walktime_2")],
                   MARGIN=1,min,na.rm=T),3)]

fhs1_ex0_28d[walktime_min=="-Inf", walktime_min := NA]


########### Grip (kg)

# Produces highest average by hand (assumes dominant hand is stronger)

fhs1_ex0_28d[,grip_avg_right := round(apply(fhs1_ex0_28d[,.(FU516,FU517,FU518)],MARGIN=1,mean),1)]
fhs1_ex0_28d[,grip_avg_left := round(apply(fhs1_ex0_28d[,.(FU519,FU520,FU521)],MARGIN=1,mean),1)]

fhs1_ex0_28d[,grip_max := 
               round(
                 apply(
                   fhs1_ex0_28d[,c("grip_avg_right","grip_avg_left")],
                   MARGIN=1,max),1)]


#### Exhaustion


fhs1_ex0_28d[FU367==3|FU380==3, exhaustion_fried := 1]
fhs1_ex0_28d[FU367<3&FU380<3, exhaustion_fried := 0]


#### (Weight) Loss

fhs1_ex0_28d[(WGT28-WGT27) >= 10, wtloss_fried := 1]
fhs1_ex0_28d[(WGT28-WGT27) < 10, wtloss_fried := 0]


######### FRAIL Score

#### Fatigue

fhs1_ex0_28d[FU367==3|FU380==3, fatigue_frail := 1]
fhs1_ex0_28d[FU367<3&FU380<3, fatigue_frail := 0]


#### Resistance

fhs1_ex0_28d[FU403==0, resistance_frail := 0]
fhs1_ex0_28d[FU403 %in% c(1,2,3), resistance_frail := 1]


#### Ambulation

fhs1_ex0_28d[FU444==1, ambulate_frail := 0]
fhs1_ex0_28d[FU444 %in% c(0,2), ambulate_frail := 1]


# FRAIL Illness score counts presence of HTN, DM, cancer, chronic lung disease, MI, CHD, angina, asthma, arthritis, stroke, and ckd
# Scores for 0-4 conditions, score = 0, for ≥ 5, score=1

# DM and HTN

# COPD (chronic lung disease)

fhs1_ex0_28d[FU239==2, copd_frail := 0]
fhs1_ex0_28d[FU239==1, copd_frail := 1]
fhs1_ex0_28d[FU239==0, copd_frail := 0]

# asthma

fhs1_ex0_28d[FU242==2, asthma_frail := 0]
fhs1_ex0_28d[FU242==1, asthma_frail := 1]
fhs1_ex0_28d[FU242==0, asthma_frail := 0]

# DJD (arthritis)

fhs1_ex0_28d[FU245==2, djd_frail := 0]
fhs1_ex0_28d[FU245==1, djd_frail := 1]
fhs1_ex0_28d[FU245==0, djd_frail := 0]

# CKD (renal disease)

fhs1_ex0_28d[,RACE := 1]
fhs1_ex0_28d[,crcl := 
  calc_MDRD4(
    as.data.frame(fhs1_ex0_28d),cr="CREAT28",age="AGE28")]

fhs1_ex0_28d[FU238==2, ckd_frail := 0]
fhs1_ex0_28d[FU238==1|crcl < 60, ckd_frail := 1]
fhs1_ex0_28d[FU238==0, ckd_frail := 0]

# CHF/MI

fhs1_ex0_28d <- fhs_outcomes[fhs1_ex0_28d,on=.(PID,IDTYPE)]

fhs1_ex0_28d[,chf_frail := 0]
fhs1_ex0_28d[hfhosp_status==1&hfhosp_dt <= DATE28, chf_frail := 1]

fhs1_ex0_28d[,chd_frail := 0]
fhs1_ex0_28d[cadhosp_status==1&cadhosp_dt <= DATE28, chf_frail := 1]

# Stroke

fhs1_ex0_28d[,stroke_frail := 0]
fhs1_ex0_28d[cvahosp_status==1&cvahosp_dt <= DATE28, stroke_fail := 1]

# Cancer

fhs1_ex0_28d <- 
  fhs_survca_first_any[,c("PID","IDTYPE","D_DATE")][fhs1_ex0_28d,
                      on=.(PID,IDTYPE)]

fhs1_ex0_28d[,cancer_frail := 0]
fhs1_ex0_28d[D_DATE <= DATE28, cancer_frail := 1]

fhs1_ex0_28d[,conditions_frail := 
  HRX28+
  DMRX28+
  copd_frail+
  asthma_frail+
  djd_frail+
  ckd_frail+
  chf_frail+
  chd_frail+
  stroke_frail+
  cancer_frail]

fhs1_ex0_28d[conditions_frail <= 4, illness_frail := 0]
fhs1_ex0_28d[conditions_frail > 4, illness_frail := 1]

#### (Weight) Loss

fhs1_ex0_28d[(WGT28-WGT27) >= 10, wtloss_frail := 1]
fhs1_ex0_28d[(WGT28-WGT27) < 10, wtloss_frail := 0]


#### Calculate FRAIL score

fhs1_ex0_28d[,total_frail := 
  fatigue_frail+
  resistance_frail+
  ambulate_frail+
  illness_frail+
  wtloss_frail]


fhs1_melt_ex0_28d <- 
  melt(fhs1_ex0_28d,
       id.vars=c("PID","IDTYPE","visit"),
       na.rm=T,
       factorsAsStrings=T)

fhs1_melt_ex0_28d[,form := "fhs1_ex0_28"]

rm(fhs1_ex0_28d)


# ============================================================#
###          FHS original, Physical function, Exam 28       ###
# ============================================================#

fhs1_physf_2005 <-
  fread('~/Dropbox/BioLINCC files/Framingham cohort/datasets/CSV/t_physf_2005_m_0162d_v2.csv',
        na.strings=c("NA","","NULL")) # Exam 28

names(fhs1_physf_2005) <-
  toupper(names(fhs1_physf_2005))

fhs1_physf_2005[,visit := 28]

fhs1_melt_physf_2005 <- 
  melt(fhs1_physf_2005,
       id.vars=c("PID","IDTYPE","visit"),
       na.rm=T,
       factorsAsStrings=T)

fhs1_melt_physf_2005[,form := "fhs1_physf_2005"]

rm(fhs1_physf_2005)


# ==============================================================#
####                FHS original, Exam 29                    ####
# ==============================================================#

fhs1_ex0_29d <-
  fread('~/Dropbox/BioLINCC files/Framingham cohort/datasets/CSV/E_EXAM_EX29_0_0210D.csv',
        na.strings=c("NA","","NULL")) # Exam 29

fhs1_ex0_29d[,visit := 29]
fhs1_ex0_29d <- 
  fhs1_wkthru[,.(PID,
                 SEX,
                 DATE29,
                 HGT1,
                 BMIC29,
                 WGT29,
                 WGT28,
                 DMRX29,
                 HRX29)][fhs1_ex0_29d,
                           on=.(PID)]

fhs1_ex0_29d[,barthel_hygiene := 0]
fhs1_ex0_29d[,essi_chores := 0]


######### FRIED

##### Walking speed 

## Walking speed

fhs1_ex0_29d[,walktime_1 := FV315]
fhs1_ex0_29d[,walktime_2 := FV319]

fhs1_ex0_29d[,walktime_avg := 
               round(
                 apply(
                   fhs1_ex0_29d[,.(walktime_1,walktime_2)],
                   MARGIN=1,mean,na.rm=T),3)]

fhs1_ex0_29d[walktime_avg=="NaN", walktime_avg <- NA]
fhs1_ex0_29d[,walktime_min := 
               round(
                 apply(
                   fhs1_ex0_29d[,.(walktime_1,walktime_2)],
                   MARGIN=1,min,na.rm=T),3)]
fhs1_ex0_29d[walktime_min=="-Inf", walktime_min := NA]


########### Grip (kg)

# Produces highest average by hand (assumes dominant hand is stronger)

fhs1_ex0_29d[,grip_avg_right := round(apply(fhs1_ex0_29d[,.(FV288,FV289,FV290)],MARGIN=1,mean),1)]
fhs1_ex0_29d[,grip_avg_left := round(apply(fhs1_ex0_29d[,.(FV291,FV292,FV293)],MARGIN=1,mean),1)]

fhs1_ex0_29d[,grip_max := 
               round(
                 apply(
                   fhs1_ex0_29d[,.(grip_avg_right,grip_avg_left)],
                   MARGIN=1,max),1)]


##### Exhaustion - Fried

fhs1_ex0_29d[FV526==3|FV539==3, exhaustion_fried := 1]
fhs1_ex0_29d[FV526<3&FV539<3, exhaustion_fried := 0]

#### (Weight) Loss

fhs1_ex0_29d[(WGT29-WGT28) >= 10, wtloss_fried := 1]
fhs1_ex0_29d[(WGT29-WGT28) < 10, wtloss_fried <- 0]


######### FRAIL Score

#### Fatigue

fhs1_ex0_29d[FV526==3|FV539==3, fatigue_frail := 1]
fhs1_ex0_29d[FV526<3&FV539<3, fatigue_frail := 0]


#### Resistance

fhs1_ex0_29d[FV399==0, resistance_frail := 0]
fhs1_ex0_29d[FV399 %in% c(1,2,3), resistance_frail := 1]

#### Ambulation

fhs1_ex0_29d[FV386==1, ambulate_frail := 0]
fhs1_ex0_29d[FV386==0, ambulate_frail := 1]


# FRAIL Illness score counts presence of HTN, DM, cancer, 
# chronic lung disease, MI, CHD, angina, asthma, arthritis, stroke, and ckd
# Scores for 0-4 conditions, score = 0, for ≥ 5, score=1

# DM and HTN


# COPD (chronic lung disease)

fhs1_ex0_29d[FV241==2, copd_frail := 0]
fhs1_ex0_29d[FV241==1, copd_frail := 1]
fhs1_ex0_29d[FV241==0, copd_frail := 0]

# asthma

fhs1_ex0_29d[FV244==2, asthma_frail := 0]
fhs1_ex0_29d[FV244==1, asthma_frail := 1]
fhs1_ex0_29d[FV244==0, asthma_frail := 0]

# DJD (arthritis)

fhs1_ex0_29d[FV247==2, djd_frail := 0]
fhs1_ex0_29d[FV247==1, djd_frail := 1]
fhs1_ex0_29d[FV247==0, djd_frail := 0]

# CKD (renal disease)

fhs1_ex0_29d[FV240==2, ckd_frail := 0]
fhs1_ex0_29d[FV240==1, ckd_frail := 1]
fhs1_ex0_29d[FV240==0, ckd_frail := 0]

# CHF/MI

fhs1_ex0_29d <-fhs_outcomes[fhs1_ex0_29d,on=.(PID,IDTYPE)]

fhs1_ex0_29d[,chf_frail := 0]
fhs1_ex0_29d[hfhosp_status==1&hfhosp_dt <= DATE29, chf_frail := 1]

fhs1_ex0_29d[,chd_frail := 0]
fhs1_ex0_29d[cadhosp_status==1&cadhosp_dt <= DATE29, chd_frail:= 1]

# Stroke

fhs1_ex0_29d[,stroke_frail := 0]
fhs1_ex0_29d[cvahosp_status==1&cvahosp_dt <= DATE29, stroke_frail := 1]

# Cancer

fhs1_ex0_29d <- 
  fhs_survca_first_any[,.(PID,IDTYPE,D_DATE)][fhs1_ex0_29d,
                                           on=.(PID,IDTYPE)]

fhs1_ex0_29d[,cancer_frail := 0]
fhs1_ex0_29d[D_DATE <= DATE29, cancer_frail := 1]

fhs1_ex0_29d[,conditions_frail := 
  HRX29+
  DMRX29+
  copd_frail+
  asthma_frail+
  djd_frail+
  ckd_frail+
  chf_frail+
  chd_frail+
  stroke_frail+
  cancer_frail]

fhs1_ex0_29d[conditions_frail <= 4, illness_frail := 0]
fhs1_ex0_29d[conditions_frail > 4, illness_frail := 1]

#### (Weight) Loss

fhs1_ex0_29d[(WGT29-WGT28) >= 10, wtloss_frail := 1]
fhs1_ex0_29d[(WGT29-WGT28) < 10, wtloss_frail := 0]

#### Calculate FRAIL score

fhs1_ex0_29d[,total_frail := 
  fatigue_frail+
  resistance_frail+
  ambulate_frail+
  illness_frail+
  wtloss_frail]


fhs1_melt_ex0_29d <- melt(fhs1_ex0_29d,id.vars=c("PID","IDTYPE","visit"),na.rm=T,factorsAsStrings=T)
fhs1_melt_ex0_29d[,form := "fhs1_ex0_29"]

rm(fhs1_ex0_29d)

# =============================================================#
###         FHS original, Exam 29 Physical function          ###
# =============================================================#

fhs1_t_physfunc_2010 <-
  fread('~/Dropbox/BioLINCC files/Framingham cohort/datasets/CSV/t_physfunc_2010_m_0634d.csv',
        na.strings=c("NA","","NULL")) # Exam 29

fhs1_t_physfunc_2010[,visit := 29]
fhs1_melt_t_physfunc_2010 <- melt(fhs1_t_physfunc_2010,id.vars=c("PID","IDTYPE","visit"),na.rm=T,factorsAsStrings=T)
fhs1_melt_t_physfunc_2010[,form := "fhs1_physfunc_2010"]

rm(fhs1_t_physfunc_2010)


# ===============================================================#
####                    FHS original, Exam 30                 ####
# ===============================================================#

fhs1_ex0_30d <-fread('~/Dropbox/BioLINCC files/Framingham cohort/datasets/CSV/E_EXAM_EX30_0_0274D.csv',na.strings=c("NA","","NULL")) # Exam 30
fhs1_ex0_30d[,visit := 30]
fhs1_ex0_30d <- 
  fhs1_wkthru[,.(PID,
                 SEX,
                 AGE30,
                 DATE30,
                 HGT1,
                 BMIC30,
                 WGT30,
                 WGT29,
                 HRX30,
                 DMRX30)][fhs1_ex0_30d,on=.(PID)]


fhs1_ex0_30d[,barthel_hygiene := 0]
fhs1_ex0_30d[,essi_chores := 0]


## Walking speed

fhs1_ex0_30d[,walktime_1 := FW367]
fhs1_ex0_30d[,walktime_2 := FW372]

fhs1_ex0_30d[,walktime_avg := 
               round(
                 apply(
                   fhs1_ex0_30d[,c("walktime_1","walktime_2")],
                   MARGIN=1,mean,na.rm=T),3)]

fhs1_ex0_30d[walktime_avg=="NaN", walktime_avg := NA]
fhs1_ex0_30d[,walktime_min := 
               round(
                 apply(
                   fhs1_ex0_30d[,c("walktime_1","walktime_2")],
                   MARGIN=1,min,na.rm=T),3)]

fhs1_ex0_30d[walktime_min=="-Inf", walktime_min := NA]


########### Grip (kg)

# Produces highest average by hand (assumes dominant hand is stronger)

fhs1_ex0_30d[,grip_avg_right := round(apply(fhs1_ex0_30d[,.(FW336,FW337,FW338)],MARGIN=1,mean),1)]
fhs1_ex0_30d[,grip_avg_left := round(apply(fhs1_ex0_30d[,.(FW339,FW340,FW341)],MARGIN=1,mean),1)]

fhs1_ex0_30d[,grip_max := 
               round(
                 apply(
                   fhs1_ex0_30d[,c("grip_avg_right","grip_avg_left")],
                   MARGIN=1,max),1)]


#### Exhaustion

fhs1_ex0_30d[FW528==3|FW541==3, exhaustion_fried := 1]
fhs1_ex0_30d[FW528<3&FW541<3, exhaustion_fried := 0]


#### (Weight) Loss

fhs1_ex0_30d[(WGT30-WGT29)>=10, wtloss_fried := 1]
fhs1_ex0_30d[(WGT30-WGT29) < 10, wtloss_fried := 0]


##### CES-D

fhs1_ex0_30d[,CESD20_missing := rowSums(is.na(fhs1_ex0_30d[,300:319]))]
fhs1_ex0_30d[,CESD20 := rowSums(fhs1_ex0_30d[,300:319])]
fhs1_ex0_30d[CESD20_missing >= 4, CESD20 := NA]


######### FRAIL Score

#### Fatigue

fhs1_ex0_30d[FW528==3|FW541==3, fatigue_frail := 1]
fhs1_ex0_30d[FW528<3&FW541<3, fatigue_frail := 0]


#### Resistance

fhs1_ex0_30d[FW448 %in% c(0,1), resistance_frail := FW448]

#### Ambulation

fhs1_ex0_30d[FW446 %in% c(0,1), ambulate_frail := FW446]


# FRAIL Illness score counts presence of HTN, DM, cancer, chronic lung disease, MI, CHD, angina, asthma, arthritis, stroke, and ckd
# Scores for 0-4 conditions, score = 0, for ≥ 5, score=1

# DM and HTN

# COPD (chronic lung disease)

fhs1_ex0_30d[FW283==2, copd_frail := 0]
fhs1_ex0_30d[FW283==1, copd_frail := 1]
fhs1_ex0_30d[FW283==0, copd_frail := 0]

# asthma

fhs1_ex0_30d[FW286==2, asthma_frail := 0]
fhs1_ex0_30d[FW286==1, asthma_frail := 1]
fhs1_ex0_30d[FW286==0, asthma_frail := 0]

# DJD (arthritis)

fhs1_ex0_30d[FW289==2, djd_frail := 0]
fhs1_ex0_30d[FW289==1, djd_frail := 1]
fhs1_ex0_30d[FW289==0, djd_frail := 0]

# CKD (renal disease)

fhs1_ex0_30d[FW281==2, ckd_frail := 0]
fhs1_ex0_30d[FW281==1, ckd_frail := 1]
fhs1_ex0_30d[FW281==0, ckd_frail := 0]

# CHF/MI

fhs1_ex0_30d <- 
  fhs_outcomes[fhs1_ex0_30d,on=.(PID,IDTYPE)]

fhs1_ex0_30d[,chf_frail := 0]
fhs1_ex0_30d[hfhosp_status==1&hfhosp_dt <= DATE30, chf_frail := 1]

fhs1_ex0_30d[,chd_frail := 0]
fhs1_ex0_30d[cadhosp_status==1&cadhosp_dt <= DATE30, chf_frail := 1]

# Stroke

fhs1_ex0_30d[,stroke_frail := 0]
fhs1_ex0_30d[cvahosp_status==1&
               cvahosp_dt <= DATE30, 
             stroke_frail := 1]

# Cancer

fhs1_ex0_30d <- 
        fhs_survca_first_any[,.(PID,IDTYPE,D_DATE)][fhs1_ex0_30d,
        on=.(PID,IDTYPE)]

fhs1_ex0_30d[,cancer_frail := 0]
fhs1_ex0_30d[D_DATE <= DATE30, cancer_frail := 1]

fhs1_ex0_30d[,conditions_frail := 
  HRX30+
  DMRX30+
  copd_frail+
  asthma_frail+
  djd_frail+
  ckd_frail+
  chf_frail+
  chd_frail+
  stroke_frail+
  cancer_frail]

fhs1_ex0_30d[conditions_frail <= 4, illness_frail := 0]
fhs1_ex0_30d[conditions_frail > 4, illness_frail := 1]

#### (Weight) Loss

fhs1_ex0_30d[(WGT30-WGT29) <= -10, wtloss_frail := 1]
fhs1_ex0_30d[(WGT30-WGT29) > -10, wtloss_frail := 0]


#### Calculate FRAIL score

fhs1_ex0_30d[,total_frail :=
               fatigue_frail+
               resistance_frail+
               ambulate_frail+
               illness_frail+
               wtloss_frail]


fhs1_melt_ex0_30d <- melt(fhs1_ex0_30d,id.vars=c("PID","IDTYPE","visit"),na.rm=T,factorsAsStrings=T)
fhs1_melt_ex0_30d[,form := "fhs1_ex0_30"]

rm(fhs1_ex0_30d)


# =============================================================#
###      FHS original, Berkman questionnaire Exam 30         ###
# =============================================================#

fhs1_bsni_2009 <-
  fread('~/Dropbox/BioLINCC files/Framingham cohort/datasets/CSV/q_berkman_2009_m_0635d.csv',
        na.strings=c("NA","","NULL")) # Exam 30

names(fhs1_bsni_2009) <- toupper(names(fhs1_bsni_2009))

fhs1_bsni_2009[,visit := 30]

fhs1_melt_bsni_2009 <- 
  melt(fhs1_bsni_2009,
       id.vars=c("PID","IDTYPE","visit"),
       na.rm=T,
       factorsAsStrings=T)

fhs1_melt_bsni_2009[,form := "fhs1_bsni_2009"]

rm(fhs1_bsni_2009)


# ==============================================================#
####                   FHS original, Exam 31                 ####
# ==============================================================#


fhs1_ex0_31d <-
  fread('~/Dropbox/BioLINCC files/Framingham cohort/datasets/CSV/E_EXAM_EX31_0_0738D.csv',
        na.strings=c("NA","","NULL")) # Exam 31

fhs1_ex0_31d[,visit := 31]
fhs1_ex0_31d <- 
  fhs1_wkthru[,c("PID",
                 "SEX",
                 "HGT1",
                 "BMIC31",
                 "WGT31",
                 "WGT30",
                 "HRX31",
                 "DMRX31",
                 "DATE31")][fhs1_ex0_31d,on=.(PID)]

fhs1_ex0_31d[,barthel_hygiene := 0]
fhs1_ex0_31d[,essi_chores := 0]


## Walking speed

fhs1_ex0_31d[,walktime_1 := FX367]
fhs1_ex0_31d[,walktime_2 := FX372]

fhs1_ex0_31d[,walktime_avg := 
               round(
                 apply(
                   fhs1_ex0_31d[,.(walktime_1,walktime_2)],
                   MARGIN=1,mean,na.rm=T),3)]
fhs1_ex0_31d[walktime_avg=="NaN", walktime_avg := NA]
fhs1_ex0_31d[,walktime_min := 
               round(
                 apply(
                   fhs1_ex0_31d[,.(walktime_1,walktime_2)],
                   MARGIN=1,min,na.rm=T),3)]
fhs1_ex0_31d[walktime_min=="-Inf",walktime_min := NA]


########### Grip (kg)

# Produces highest average by hand (assumes dominant hand is stronger)

fhs1_ex0_31d[,grip_avg_right := 
               round(
                 apply(
                   fhs1_ex0_31d[,.(FX336,FX337,FX338)],
                   MARGIN=1,mean),1)]
fhs1_ex0_31d[,grip_avg_left := 
               round(
                 apply(
                   fhs1_ex0_31d[,.(FX339,FX340,FX341)],
                   MARGIN=1,mean),1)]

fhs1_ex0_31d[,grip_max := 
               round(
                 apply(
                   fhs1_ex0_31d[,.(grip_avg_right,grip_avg_left)],
                   MARGIN=1,max),1)]


#### Exhaustion

fhs1_ex0_31d[FX528==3|FX541==3, exhaustion_fried := 1]
fhs1_ex0_31d[FX528<3&FX541<3, exhaustion_fried := 0]


#### (Weight) Loss

fhs1_ex0_31d[(WGT31-WGT30)>=10, wtloss_fried := 1]
fhs1_ex0_31d[(WGT31-WGT30) < 10, wtloss_fried := 0]


######### FRAIL Score

fhs1_ex0_31d[FX528==3|FX541==3, fatigue_frail := 1]
fhs1_ex0_31d[FX528<3&FX541<3, fatigue_frail := 0]


#### Resistance

fhs1_ex0_31d[FX448 %in% c(0,2), resistance_frail := 1]
fhs1_ex0_31d[FX448==1, resistance_frail := 0]

#### Ambulation

fhs1_ex0_31d[FX446 %in% c(0,2), ambulate_frail := 1]
fhs1_ex0_31d[FX446==1, ambulate_frail := 0]


# FRAIL Illness score counts presence of HTN, DM, cancer, chronic lung disease, MI, CHD, angina, asthma, arthritis, stroke, and ckd
# Scores for 0-4 conditions, score = 0, for ≥ 5, score=1


# COPD (chronic lung disease)

fhs1_ex0_31d[FX283==2, copd_frail := 0]
fhs1_ex0_31d[FX283==1, copd_frail := 1]
fhs1_ex0_31d[FX283==0, copd_frail := 0]

# asthma

fhs1_ex0_31d[FX286==2, asthma_frail := 0]
fhs1_ex0_31d[FX286==1, asthma_frail := 1]
fhs1_ex0_31d[FX286==0, asthma_frail := 0]

# DJD (arthritis)

fhs1_ex0_31d[FX289==2, djd_frail := 0]
fhs1_ex0_31d[FX289==1, djd_frail := 1]
fhs1_ex0_31d[FX289==0, djd_frail := 0]

# CKD (renal disease)

fhs1_ex0_31d[FX281==2, ckd_frail := 0]
fhs1_ex0_31d[FX281==1, ckd_frail := 1]
fhs1_ex0_31d[FX281==0, ckd_frail := 0]

# CHF/MI

fhs1_ex0_31d <- fhs_outcomes[fhs1_ex0_31d,on=.(PID,IDTYPE)]

fhs1_ex0_31d[,chf_frail := 0]
fhs1_ex0_31d[hfhosp_status==1&hfhosp_dt <= DATE31, chf_frail := 1]

fhs1_ex0_31d[,chd_frail := 0]
fhs1_ex0_31d[cadhosp_status==1&cadhosp_dt <= DATE31, chd_frail := 1]

# Stroke

fhs1_ex0_31d[,stroke_frail := 0]
fhs1_ex0_31d[cvahosp_status==1&cvahosp_dt <= DATE31, stroke_frail := 1]

# Cancer

fhs1_ex0_31d <- 
  fhs_survca_first_any[,c("PID","IDTYPE","D_DATE")][
    fhs1_ex0_31d, on=.(PID,IDTYPE)]

fhs1_ex0_31d[,cancer_frail := 0]
fhs1_ex0_31d[D_DATE <= DATE31, cancer_frail := 1]

fhs1_ex0_31d[,conditions_frail := 
  HRX31+
  DMRX31+
  copd_frail+
  asthma_frail+
  djd_frail+
  ckd_frail+
  chf_frail+
  chd_frail+
  stroke_frail+
  cancer_frail]

fhs1_ex0_31d[conditions_frail <= 4, illness_frail := 0]
fhs1_ex0_31d[conditions_frail > 4, illness_frail <- 1]


#### (Weight) Loss

fhs1_ex0_31d[(WGT31-WGT30)>=10, wtloss_frail := 1]
fhs1_ex0_31d[(WGT31-WGT30) < 10, wtloss_frail := 0]


#### Calculate FRAIL score

fhs1_ex0_31d[,total_frail := 
  fatigue_frail+
  resistance_frail+
  ambulate_frail+
  illness_frail+
  wtloss_frail]


##### CES-D

fhs1_ex0_31d[,CESD20_missing := 
               rowSums(
                 is.na(
                   fhs1_ex0_31d[,
                                c(
                                  paste0(
                                    "FX",
                                    c(519,522:541)
                                  )
                                )
                   ]
                 )
               )
]

fhs1_ex0_31d[,CESD20 := 
               rowSums(
                 fhs1_ex0_31d[,
                              c(
                                paste0(
                                  "FX",
                                  c(519,522:541)
                                )
                              )
                 ]
               )
]

fhs1_ex0_31d[CESD20_missing >= 4, CESD20 := NA]


####

fhs1_melt_ex0_31d <- melt(fhs1_ex0_31d,id.vars=c("PID","IDTYPE","visit"),na.rm=T,factorsAsStrings=T)
fhs1_melt_ex0_31d[,form := "fhs1_ex0_31"]

rm(fhs1_ex0_31d)


# ==============================================================#
####                    FHS original, Exam 32                ####
# ==============================================================#

fhs1_ex0_32d <-
  fread('~/Dropbox/BioLINCC files/Framingham cohort/datasets/CSV/E_EXAM_EX32_0_0939D.csv',
        na.strings=c("NA","","NULL")) # Exam 32

fhs1_ex0_32d[,visit := 32]
fhs1_ex0_32d <-
  fhs1_wkthru[,c("PID",
                 "SEX",
                 "HGT1",
                 "BMIC32",
                 "DATE32",
                 "WGT32",
                 "WGT31",
                 "DMRX32",
                 "HRX32")][fhs1_ex0_32d,on=.(PID)]

fhs1_ex0_32d[,barthel_hygiene := 0]


## Walking speed

fhs1_ex0_32d[,walktime_1 := FY367]
fhs1_ex0_32d[,walktime_2 := FY372]

fhs1_ex0_32d[,walktime_avg := round(apply(fhs1_ex0_32d[,c("walktime_1","walktime_2")],MARGIN=1,mean,na.rm=T),3)]
fhs1_ex0_32d[walktime_avg=="NaN", walktime_avg <- NA]
fhs1_ex0_32d[,walktime_min := round(apply(fhs1_ex0_32d[,c("walktime_1","walktime_2")],MARGIN=1,min,na.rm=T),3)]
fhs1_ex0_32d[walktime_min=="-Inf", walktime_min := NA]

########### Grip (kg)


# Produces highest average by hand (assumes dominant hand is stronger)

fhs1_ex0_32d[,grip_avg_right := round(apply(fhs1_ex0_32d[,.(FY336,FY337,FY338)],MARGIN=1,mean),1)]
fhs1_ex0_32d[,grip_avg_left := round(apply(fhs1_ex0_32d[,.(FY339,FY340,FY341)],MARGIN=1,mean),1)]

fhs1_ex0_32d[,grip_max := round(apply(fhs1_ex0_32d[,c("grip_avg_right","grip_avg_left")],MARGIN=1,max),1)]


#### Exhaustion

fhs1_ex0_32d[FY528==3|FY541==3, exhaustion_fried := 1]
fhs1_ex0_32d[FY528<3&FY541<3, exhaustion_fried := 0]


#### (Weight) Loss

fhs1_ex0_32d[(WGT32-WGT31)>=10, wtloss_fried := 1]
fhs1_ex0_32d[(WGT32-WGT31) < 10, wtloss_fried := 0]


#=============================================================================#

######### FRAIL Score


#### Fatigue

fhs1_ex0_32d[FY528==3|FY541==3, fatigue_frail := 1]
fhs1_ex0_32d[FY528<3&FY541<3, fatigue_frail := 0]


#### Resistance

fhs1_ex0_32d[FY463==0, resistance_frail := 1]
fhs1_ex0_32d[FY463==1, resistance_frail := 0]

#### Ambulation

fhs1_ex0_32d[FY462==0, ambulate_frail := 1]
fhs1_ex0_32d[FY462==1, ambulate_frail := 0]


# FRAIL Illness score counts presence of HTN, DM, cancer, chronic lung disease, MI, CHD, angina, asthma, arthritis, stroke, and ckd
# Scores for 0-4 conditions, score = 0, for ≥ 5, score=1


# COPD (chronic lung disease)

fhs1_ex0_32d[FY283==2, copd_frail := 0]
fhs1_ex0_32d[FY283==1, copd_frail := 1]
fhs1_ex0_32d[FY283==0, copd_frail := 0]

# asthma

fhs1_ex0_32d[FY286==2, asthma_frail := 0]
fhs1_ex0_32d[FY286==1, asthma_frail := 1]
fhs1_ex0_32d[FY286==0, asthma_frail := 0]

# DJD (arthritis)

fhs1_ex0_32d[FY289==2, djd_frail := 0]
fhs1_ex0_32d[FY289==1, djd_frail := 1]
fhs1_ex0_32d[FY289==0, djd_frail := 0]

# CKD (renal disease)

fhs1_ex0_32d[FY281==2, ckd_frail := 0]
fhs1_ex0_32d[FY281==1, ckd_frail := 1]
fhs1_ex0_32d[FY281==0, ckd_frail := 0]

# CHF/MI

fhs1_ex0_32d <- fhs_outcomes[fhs1_ex0_32d,on=.(PID,IDTYPE)]

fhs1_ex0_32d[,chf_frail := 0]
fhs1_ex0_32d[hfhosp_status==1&hfhosp_dt <= DATE32, chf_frail := 1]

fhs1_ex0_32d[,chd_frail := 0]
fhs1_ex0_32d[cadhosp_status==1&cadhosp_dt <= DATE32, chd_frail := 1]

# Stroke

fhs1_ex0_32d <- fhs_survstk[fhs1_ex0_32d,on=.(PID,IDTYPE)]

fhs1_ex0_32d[,stroke_frail := 0]
fhs1_ex0_32d[cvahosp_status==1&cvahosp_dt <= DATE32, stroke_frail := 1]

# Cancer

fhs1_ex0_32d <- 
  fhs_survca_first_any[,c("PID","IDTYPE","D_DATE")][fhs1_ex0_32d,
                      on=.(PID,IDTYPE)]

fhs1_ex0_32d[,cancer_frail := 0]
fhs1_ex0_32d[D_DATE <= DATE32, cancer_frail := 1]

fhs1_ex0_32d[,conditions_frail := 
  HRX32+
  DMRX32+
  copd_frail+
  asthma_frail+
  djd_frail+
  ckd_frail+
  chf_frail+
  chd_frail+
  stroke_frail+
  cancer_frail]

fhs1_ex0_32d[conditions_frail <= 4, illness_frail := 0]
fhs1_ex0_32d[conditions_frail > 4, illness_frail := 1]


#### (Weight) Loss

fhs1_ex0_32d[(WGT32-WGT31)>=10, wtloss_frail := 1]
fhs1_ex0_32d[(WGT32-WGT31) < 10, wtloss_frail := 0]


#### Calculate FRAIL score

fhs1_ex0_32d[,total_frail := 
  fatigue_frail+
  resistance_frail+
  ambulate_frail+
  illness_frail+
  wtloss_frail]


##### CES-D


fhs1_ex0_32d[,CESD11_missing := rowSums(is.na(fhs1_ex0_32d[,c(paste0("FY",c(519,
                                                                            522,
                                                                            526:529,
                                                                            531:533,
                                                                            535,
                                                                            541)))]))]

fhs1_ex0_32d[,CESD11 := rowSums(fhs1_ex0_32d[,c(paste0("FY",c(519,
                                                              522,
                                                              526:529,
                                                              531:533,
                                                              535,
                                                              541)))])]
fhs1_ex0_32d[CESD11_missing >= 4, CESD11 := NA]

### 

fhs1_melt_ex0_32d <- 
  melt(fhs1_ex0_32d,
       id.vars=c("PID","IDTYPE","visit"),
       na.rm=T,
       factorsAsStrings=T)

fhs1_melt_ex0_32d[,form := "fhs1_ex0_32"]

rm(fhs1_ex0_32d)


# ===============================================================##
####              FHS original, HA1c, exam 19,23               ---#
# ===============================================================##

fhs1_l_hba1c_ex23 <-
  fread('~/Dropbox/BioLINCC files/Framingham cohort/datasets/CSV/L_HBA1C_EX23_0_0110D.csv',
        na.strings=c("NA","","NULL")) # HbA1c

fhs1_l_hba1c_ex19 <- fhs1_l_hba1c_ex23[,c("PID","IDTYPE","DATE1920","HBA1920")]
fhs1_l_hba1c_ex19[,visit := 19]
fhs1_l_hba1c_ex19[,form := "fhs1_hba1c"]
fhs1_l_hba1c_ex19[,variable := "HB1920"]
names(fhs1_l_hba1c_ex19) <- c("PID","IDTYPE","DATE","value","visit","form","variable")
fhs1_melt_l_hba1c_ex19 <- fhs1_l_hba1c_ex19


fhs1_l_hba1c_ex22 <- fhs1_l_hba1c_ex23[,c("PID","IDTYPE","DATE22","HBA22")]
fhs1_l_hba1c_ex22[,visit := 22]
fhs1_l_hba1c_ex22[,form := "fhs1_hba1c"]
fhs1_l_hba1c_ex22[,variable := "HBA22"]
names(fhs1_l_hba1c_ex22) <- c("PID","IDTYPE","DATE","value","visit","form","variable")
fhs1_melt_l_hba1c_ex22 <- fhs1_l_hba1c_ex22

fhs1_melt_l_hba1c <- 
  rbindlist(
    list(fhs1_melt_l_hba1c_ex19,
         fhs1_melt_l_hba1c_ex22))

fhs1_melt_l_hba1c <- fhs1_melt_l_hba1c[!is.na(value)]

rm(fhs1_l_hba1c_ex23,
   fhs1_l_hba1c_ex19,
   fhs1_l_hba1c_ex22,
   fhs1_melt_l_hba1c_ex19,
   fhs1_melt_l_hba1c_ex22
)


# ==============================================================#
##                 FHS original, PFTs, exam 19                 ##
# ==============================================================#

fhs1_t_pft_ex19 <- 
  fread('~/Dropbox/BioLINCC files/Framingham cohort/datasets/CSV/T_PFT_EX19_0_0169D.csv',
        na.strings=c("NA","","NULL")) # Pulmonary function tests

fhs1_t_pft_ex19[,visit := 19]

fhs1_melt_t_pft_ex19 <- 
  melt(fhs1_t_pft_ex19,
       id.vars=c("PID","IDTYPE","visit"),
       na.rm=T,
       factorsAsStrings=T)

fhs1_melt_t_pft_ex19[,form := "fhs1_t_pft_ex19"]

rm(fhs1_t_pft_ex19)

# ==============================================================#
##          FHS original, sex hormones, exam 17-21             ##
# ==============================================================#

fhs1_shorm0 <- 
  fread('~/Dropbox/BioLINCC files/Framingham cohort/datasets/CSV/shorm0_21d.csv',
        na.strings=c("NA","","NULL")) 

fhs1_melt_shorm <- 
  melt(fhs1_shorm0,
       na.rm=T,
       factorsAsStrings=T,
       id.vars=c("PID","IDTYPE"))

fhs1_melt_shorm[,var :=
  substr(variable, 1, str_locate(variable,"_")-1)]

fhs1_melt_shorm[,visit :=
  substr(variable,
         str_locate(variable,"_")+1,
         str_length(variable))]

fhs1_melt_shorm[,variable := var]

fhs1_melt_shorm[,form := "fhs1_shorm0"]

# =============================================================#
####       * FHS original, Assemble all analysis data *     ####
# =============================================================#

fhs1_melt_all <- 
  rbindlist(
    list(fhs1_melt_ex0_7d[,c("PID","IDTYPE","visit","variable","value","form")],
    fhs1_melt_ex0_8d[,c("PID","IDTYPE","visit","variable","value","form")],
    fhs1_melt_ex0_9d[,c("PID","IDTYPE","visit","variable","value","form")],
    fhs1_melt_ex0_10d[,c("PID","IDTYPE","visit","variable","value","form")],
    fhs1_melt_ex0_11d[,c("PID","IDTYPE","visit","variable","value","form")],
    fhs1_melt_ex0_12d[,c("PID","IDTYPE","visit","variable","value","form")],
    fhs1_melt_ex0_13d[,c("PID","IDTYPE","visit","variable","value","form")],
    fhs1_melt_ex0_14d[,c("PID","IDTYPE","visit","variable","value","form")],
    fhs1_melt_ex0_15d[,c("PID","IDTYPE","visit","variable","value","form")],
    fhs1_melt_ex0_16d[,c("PID","IDTYPE","visit","variable","value","form")],
    fhs1_melt_ex0_17d[,c("PID","IDTYPE","visit","variable","value","form")],
    fhs1_melt_ex0_18d[,c("PID","IDTYPE","visit","variable","value","form")],
    fhs1_melt_ex0_19d[,c("PID","IDTYPE","visit","variable","value","form")],
    fhs1_melt_ex0_20d[,c("PID","IDTYPE","visit","variable","value","form")],
    fhs1_melt_ex0_21d[,c("PID","IDTYPE","visit","variable","value","form")],
    fhs1_melt_ex0_22d[,c("PID","IDTYPE","visit","variable","value","form")],
    fhs1_melt_ex0_23d[,c("PID","IDTYPE","visit","variable","value","form")],
    fhs1_melt_ex0_24d[,c("PID","IDTYPE","visit","variable","value","form")],
    fhs1_melt_ex0_25d[,c("PID","IDTYPE","visit","variable","value","form")],
    fhs1_melt_ex0_26d[,c("PID","IDTYPE","visit","variable","value","form")],
    fhs1_melt_ex0_27d[,c("PID","IDTYPE","visit","variable","value","form")],
    fhs1_melt_ex0_28d[,c("PID","IDTYPE","visit","variable","value","form")],
    fhs1_melt_ex0_29d[,c("PID","IDTYPE","visit","variable","value","form")],
    fhs1_melt_ex0_30d[,c("PID","IDTYPE","visit","variable","value","form")],
    fhs1_melt_ex0_31d[,c("PID","IDTYPE","visit","variable","value","form")],
    fhs1_melt_ex0_32d[,c("PID","IDTYPE","visit","variable","value","form")],
    fhs1_melt_l_fhslab1_ex20[,c("PID","IDTYPE","visit","variable","value","form")],
    fhs1_melt_l_hba1c[,c("PID","IDTYPE","visit","variable","value","form")],
    fhs1_melt_l_lipids_ex12[,c("PID","IDTYPE","visit","variable","value","form")],
    fhs1_melt_t_echo_ex20[,c("PID","IDTYPE","visit","variable","value","form")],
    fhs1_melt_t_echodop_ex20[,c("PID","IDTYPE","visit","variable","value","form")],
    fhs1_melt_t_hrv_ex20[,c("PID","IDTYPE","visit","variable","value","form")],
    fhs1_melt_t_pft_ex19[,c("PID","IDTYPE","visit","variable","value","form")],
    fhs1_melt_vr_ffreq_ex20[,c("PID","IDTYPE","visit","variable","value","form")],
    fhs1_melt_vr_ffreq_ex21[,c("PID","IDTYPE","visit","variable","value","form")],
    fhs1_melt_vr_ffreq_ex22[,c("PID","IDTYPE","visit","variable","value","form")],
    fhs1_melt_wkthru[,c("PID","IDTYPE","visit","variable","value","form")],
    fhs1_melt_sex[,c("PID","IDTYPE","visit","variable","value","form")],
    fhs1_melt_meno[,c("PID","IDTYPE","visit","variable","value","form")],
    fhs1_melt_shorm[,c("PID","IDTYPE","visit","variable","value","form")],
    fhs1_melt_bsni_2009[,c("PID","IDTYPE","visit","variable","value","form")],
    fhs1_melt_physf_2005[,c("PID","IDTYPE","visit","variable","value","form")],
    fhs1_melt_t_physfunc_2010[,c("PID","IDTYPE","visit","variable","value","form")]))

fhs1_no_attend <- 
fhs1_melt_wkthru[variable=="ATT",.(PID, visit,att=value)]

fhs1_melt_all <- fhs1_no_attend[fhs1_melt_all,on=.(PID,visit)]
fhs1_melt_all <- fhs1_wkthru[,.(PID,SEX)][fhs1_melt_all,on=.(PID)]

fhs1_melt_all <- fhs1_melt_all[att==1]

fhs1_melt_all[, variable := as.character(variable)]

#=========================================================================#
#                                                                         #
####            *** FRAMINGHAM Offspring/Omni 1 DATA ***               ####
#                                                                         #
#=========================================================================#


# ================================================================#
####  ** Framingham Offspring + OMNI 1 dates & outcomes**      ####
# ================================================================#

names(fhs2_dates) <- toupper(names(fhs2_dates))
names(fhs2_offspring_wkthru) <- toupper(names(fhs2_offspring_wkthru))

fhs2_offspring_wkthru[,HGTCM1 := HGT1 * 2.54]
fhs2_offspring_wkthru[,WGTKG1 := WGT1/2.2]
fhs2_offspring_wkthru[,WGTKG2 := WGT2/2.2]
fhs2_offspring_wkthru[,WGTKG3 := WGT3/2.2]
fhs2_offspring_wkthru[,WGTKG4 := WGT4/2.2]
fhs2_offspring_wkthru[,WGTKG5 := WGT5/2.2]
fhs2_offspring_wkthru[,WGTKG6 := WGT6/2.2]
fhs2_offspring_wkthru[,WGTKG7 := WGT7/2.2]
fhs2_offspring_wkthru[,WGTKG8 := WGT8/2.2]
fhs2_offspring_wkthru[,WGTKG9 := WGT9/2.2]

fhs2_offspring_wkthru[,BSA1 := calc_bsa(fhs2_offspring_wkthru,"WGTKG1","HGTCM1")]
fhs2_offspring_wkthru[,BSA2 := calc_bsa(fhs2_offspring_wkthru,"WGTKG2","HGTCM1")]
fhs2_offspring_wkthru[,BSA3 := calc_bsa(fhs2_offspring_wkthru,"WGTKG3","HGTCM1")]
fhs2_offspring_wkthru[,BSA4 := calc_bsa(fhs2_offspring_wkthru,"WGTKG4","HGTCM1")]
fhs2_offspring_wkthru[,BSA5 := calc_bsa(fhs2_offspring_wkthru,"WGTKG5","HGTCM1")]
fhs2_offspring_wkthru[,BSA6 := calc_bsa(fhs2_offspring_wkthru,"WGTKG6","HGTCM1")]
fhs2_offspring_wkthru[,BSA7 := calc_bsa(fhs2_offspring_wkthru,"WGTKG7","HGTCM1")]
fhs2_offspring_wkthru[,BSA8 := calc_bsa(fhs2_offspring_wkthru,"WGTKG8","HGTCM1")]
fhs2_offspring_wkthru[,BSA9 := calc_bsa(fhs2_offspring_wkthru,"WGTKG9","HGTCM1")]

fhs2_melt_offspring_wkthru <- 
  melt(fhs2_offspring_wkthru,
       id.vars=c("PID","IDTYPE","SEX"),
       na.rm=T,
       factorsAsStrings=T)

fhs2_melt_offspring_wkthru[,variable :=  as.character(variable)]

fhs2_melt_offspring_wkthru[variable=="RACE", variable :=
  paste(variable,"1",sep="")]

fhs2_melt_offspring_wkthru[variable=="ETHNICITY", variable :=
  paste(variable[variable=="ETHNICITY"],"1",sep="")]


fhs2_melt_offspring_wkthru[,visit  :=  substr(variable,
         str_locate(variable,"\\d")[,1],
         nchar(variable))]

fhs2_melt_offspring_wkthru[,variable  := 
  substr(variable,
         1,
         str_locate(variable,"\\d")[,1]-1)]


fhs2_melt_wkthru <- fhs2_melt_offspring_wkthru

fhs2_melt_wkthru[variable=="WAIST", value :=  
  as.character(as.numeric(value[variable=="WAIST"]) * 2.54)]

fhs2_melt_wkthru_kg <- fhs2_melt_wkthru[variable=="WGT"]
fhs2_melt_wkthru_kg[,value := as.numeric(value)/2.20462]
fhs2_melt_wkthru_kg[,variable := "WGT_KG"]
fhs2_melt_wkthru <- 
  as.data.table(
    rbindlist(
      list(fhs2_melt_wkthru,
          fhs2_melt_wkthru_kg)))

fhs2_wkthru <- dcast(fhs2_melt_wkthru,...~variable)
fhs2_melt_wkthru[,form := "fhs2_wkthru"]


#================================#
#### *** Offspring exam 1 *** ####
#================================#

fhs2_ex1_1d <- fread('~/Dropbox/BioLINCC files/Framingham offspring/datasets/CSV/ex1_1d_v4.csv',na.strings=c("NA","","NULL"))  # Offspring, Exam 1
fhs2_ex1_1d[,visitdays := 0]
fhs2_ex1_1d[,visit := 1]
setnames(fhs2_ex1_1d, "idtype", "IDTYPE")

fhs2_ex1_1d[,drinks_wk := 
  A111+
  A112+
  A113]

fhs2_ex1_1d[!between(A89,18,55), A89 := NA]


fhs2_melt_ex1_1d <- 
  melt(fhs2_ex1_1d,
       id.vars=c("PID",
                 "IDTYPE",
                 "visit",
                 "visitdays"), 
       na.rm=T, 
       factorsAsStrings=T)

fhs2_melt_ex1_1d[,form := "fhs2_offspring_ex01"]

rm(fhs2_ex1_1d)

#==========================================#
#.    Renin activity, Offspring Exam 1     #
#==========================================#

fhs2_renin1 <- fread('~/Dropbox/BioLINCC files/Framingham offspring/datasets/CSV/renin1_6d.csv',na.strings=c("NA","","NULL"))
fhs2_renin1[,visit := 1]
fhs2_melt_renin1 <- melt(fhs2_renin1,id.vars=c("PID","IDTYPE","visit"), na.rm=T, factorsAsStrings=T)
fhs2_melt_renin1[,form := "fhs2_renin1"]

rm(fhs2_renin1)


#==================================#
####  *** Offspring exam 2 ***  ####
#==================================#

fhs2_ex1_2d <- 
  fread('~/Dropbox/BioLINCC files/Framingham offspring/datasets/CSV/ex1_2d_v4.csv',
        na.strings=c("NA","","NULL"))  # Offspring, Exam 2
setnames(fhs2_ex1_2d, "idtype", "IDTYPE")
fhs2_ex1_2d[,visit := 2]
fhs2_ex1_2d <- 
        fhs2_dates[,c("PID","IDTYPE","SEX")][fhs2_ex1_2d,
        on=.(PID,IDTYPE)]

fhs2_ex1_2d[,B439_cm := B439/10]  # IVS
fhs2_ex1_2d[,B443_cm := B443/10]  # LVPWd
fhs2_ex1_2d[,B448_cm := B448/10]  # LVIDD


fhs2_ex1_2d <- 
  as.data.table(
    calc_hypertrophy_type(
      as.data.frame(fhs2_ex1_2d), 
      id = "PID",
      sex="SEX",
      male="1",
      female="2",
      lvedd="B448_cm",
      ivsd="B439_cm",
      lvpwtd="B443_cm",
      height="B14",
      weight="B13"))

fhs2_ex1_2d[,fhs_pai := 
  calc_framingham_pai(
    as.data.frame(fhs2_ex1_2d),
    slp_hrs = "B104",
    sed_hrs = "B105",
    slgt_hrs = "B106",
    mod_hrs = "B107",
    hvy_hrs = "B108")]

fhs2_ex1_2d[,drinks_wk := 
              B117+
              B118+
              B119]

fhs2_ex1_2d[!between(B71,19,58), B71 := NA]
fhs2_ex1_2d[!between(B75,7,22), B75 := NA]
fhs2_ex1_2d[!between(B76,1,11), B76 := NA]
fhs2_ex1_2d[!between(B77,20,53), B77:= NA]
fhs2_ex1_2d[!between(B80,1,16), B80 := NA]
fhs2_ex1_2d[!between(B82,1,23), B82 := NA]


fhs2_melt_ex1_2d <- 
    melt(fhs2_ex1_2d,
         id.vars=c("PID","IDTYPE","visit"), 
         na.rm=T, 
         factorsAsStrings=T)

fhs2_melt_ex1_2d[,form := "fhs2_offspring_ex02"]

rm(fhs2_ex1_2d)

#========================#
####    CRP, Exam 2      #
#========================#


fhs2_l_crp_ex02 <- 
  fread('~/Dropbox/BioLINCC files/Framingham offspring/datasets/CSV/l_crp_ex02_1_0089d.csv',
        na.strings=c("NA","","NULL"))

setnames(fhs2_l_crp_ex02,"idtype","IDTYPE")

fhs2_l_crp_ex02[,visit := 2]
fhs2_melt_l_crp_ex02 <- 
  melt(fhs2_l_crp_ex02,
       id.vars=c("PID","IDTYPE","visit"), 
       na.rm=T, 
       factorsAsStrings=T)
fhs2_melt_l_crp_ex02[,form := "fhs2_crp_ex02"]

rm(fhs2_l_crp_ex02)


#================================#
#### *** Offspring exam 3 *** ####
#================================#

fhs2_ex1_3d <- 
  fread('~/Dropbox/BioLINCC files/Framingham offspring/datasets/CSV/ex1_3d_v1.csv',
        na.strings=c("NA","","NULL"))  # Offspring, Exam 3

setnames(fhs2_ex1_3d,"idtype","IDTYPE")

fhs2_ex1_3d[,visit := 3]

fhs2_ex1_3d[,drinks_wk := 
  C83+
  C86+
  C89]

fhs2_ex1_3d[!between(C48,19,60), C48 := NA]
fhs2_ex1_3d[!between(C50,20,57), C50 := NA]
fhs2_ex1_3d[!between(C52,1,11), C52 := NA]
fhs2_ex1_3d[!between(C53,20,51), C53 := NA]

fhs2_melt_ex1_3d <- 
  melt(fhs2_ex1_3d,
       id.vars=c("PID","IDTYPE","visit"), 
       na.rm=T, 
       factorsAsStrings=T)

fhs2_melt_ex1_3d[,form := "fhs2_offspring_ex03"]

rm(fhs2_ex1_3d)

#=========================================================#
####  Food frequency questionnaire, Offspring, Exam 3. ---#
#=========================================================#

fhs2_q_ffreq_ex03 <- fread('~/Dropbox/BioLINCC files/Framingham offspring/datasets/CSV/q_ffreq_ex03_1_0302d.csv',na.strings=c("NA","","NULL"))
setnames(fhs2_q_ffreq_ex03,"idtype","IDTYPE")
fhs2_q_ffreq_ex03[,visit := 3]

fhs2_melt_q_ffreq_ex03 <- 
  melt(fhs2_q_ffreq_ex03,
       id.vars=c("PID","IDTYPE","visit"), 
       na.rm=T, 
       factorsAsStrings=T)

fhs2_melt_q_ffreq_ex03[,form := "fhs2_q_ffreq"]

rm(fhs2_q_ffreq_ex03)

#================================================#
####  Psych questionnaire, Offspring, Exam 3  ---#
#================================================#

fhs2_q_psych_ex03<- 
  fread('~/Dropbox/BioLINCC files/Framingham offspring/datasets/CSV/q_psych_ex03_1_0167d.csv',
        na.strings=c("NA","","NULL"))

setnames(fhs2_q_psych_ex03,"idtype","IDTYPE")

fhs2_q_psych_ex03[is.na(PY109), PY109 := 0]

fhs2_q_psych_ex03[,visit := 3]
fhs2_melt_q_psych_ex03 <- 
  melt(fhs2_q_psych_ex03,
       id.vars=c("PID","IDTYPE","visit"), 
       na.rm=T, 
       factorsAsStrings=T)

fhs2_melt_q_psych_ex03[,form := "fhs2_q_psych_ex03"]

rm(fhs2_q_psych_ex03)


#====================================================#
####  Heart rate variability, Offspring - Exam 3  ---#
#====================================================#

fhs2_t_hrv <- fread('~/Dropbox/BioLINCC files/Framingham offspring/datasets/CSV/t_hrv_1987_m_0118d.csv',na.strings=c("NA","","NULL"))
names(fhs2_t_hrv) <- toupper(names(fhs2_t_hrv))
fhs2_t_hrv[,visit := HRV_EXAM]
fhs2_melt_t_hrv <- melt(fhs2_t_hrv,id.vars=c("PID","IDTYPE","visit"), na.rm=T, factorsAsStrings=T)
fhs2_melt_t_hrv[,form := "fhs2_t_hrv"]

rm(fhs2_t_hrv)


#================================================================#
####          Sex hormones, Offspring, Exam 3                 ---#
#================================================================#

fhs2_shorm_ex03 <- fread('~/Dropbox/BioLINCC files/Framingham Offspring/datasets/CSV/l_shorm_ex03_1_0177d.csv')
fhs2_melt_shorm_ex03 <-  melt(fhs2_shorm_ex03,id.vars=c("PID","IDTYPE"),na.rm=T,factorsAsStrings = T)
fhs2_melt_shorm_ex03[,visit := 3]
fhs2_melt_shorm_ex03[,form := "fhs2_shorm_ex03"]

rm(fhs2_shorm_ex03)


#==================================#
####  *** Offspring exam 4 ***  ####
#==================================#

fhs2_ex1_4d <- fread('~/Dropbox/BioLINCC files/Framingham offspring/datasets/CSV/ex1_4d_v1.csv',na.strings=c("NA","","NULL"))  # Offspring, Exam 4
setnames(fhs2_ex1_4d,"idtype","IDTYPE")
fhs2_ex1_4d[,visit := 4]

fhs2_ex1_4d[,drinks_wk := 
              D082+
              D086+
              D088]

fhs2_ex1_4d[,fhs_pai := 
  calc_framingham_pai(
    as.data.frame(fhs2_ex1_4d),
    slp_hrs = "D426",
    sed_hrs = "D427",
    slgt_hrs = "D428",
    mod_hrs = "D429",
    hvy_hrs = "D430")]


fhs2_ex1_4d[!between(D054,23,75), D054 := NA]
fhs2_ex1_4d[!between(D056,19,81), D056 := NA]
fhs2_ex1_4d[!between(D058,1,11), D058 := NA]
fhs2_ex1_4d[!between(D059,19,61), D059 := NA]


fhs2_melt_ex1_4d <- 
    melt(fhs2_ex1_4d,
         id.vars=c("PID","IDTYPE","visit"),
         na.rm=T, 
         factorsAsStrings=T)

fhs2_melt_ex1_4d[,form := "fhs2_offspring_ex04"]

rm(fhs2_ex1_4d)

#==========================#
####    Echo, Exam 4    ---#
#==========================#

fhs2_t_echo_ex04 <- fread('~/Dropbox/BioLINCC files/Framingham offspring/datasets/CSV/t_echo_ex04_1_0101d.csv',na.strings=c("NA","","NULL"))

setnames(fhs2_t_echo_ex04,"idtype","IDTYPE")

fhs2_t_echo_ex04[,visit := 4]

fhs2_meas4 <- 
  fhs2_offspring_wkthru[
    ,
    .(
      PID,
      IDTYPE,
      SEX,
      HEIGHT = HGT1 * 2.54,
      WEIGHT = WGT4 / 2.2
    )
  ]

fhs2_t_echo_ex04 <- 
  fhs2_meas4[fhs2_t_echo_ex04,
        on=.(PID,IDTYPE)]

fhs2_t_echo_ex04 <- 
  as.data.table(
    calc_hypertrophy_type(
      as.data.frame(fhs2_t_echo_ex04), 
      id = "PID",
      sex="SEX",
      lvedd="LVDD4",
      ivsd="IVSD4",
      lvpwtd="LVPD4",
      height="HEIGHT",
      weight="WEIGHT"))

fhs2_melt_t_echo_ex04 <- 
    melt(fhs2_t_echo_ex04,
         id.vars=c("PID","IDTYPE","visit"), 
         na.rm=T, 
         factorsAsStrings=T)

fhs2_melt_t_echo_ex04[,form := "fhs2_echo_ex04"]

rm(fhs2_t_echo_ex04,
   fhs2_meas4)


#================================================================#
####          Sex hormones, Offspring, Exam 4                 ---#
#================================================================#

fhs2_shorm_ex04 <- fread('~/Dropbox/BioLINCC files/Framingham Offspring/datasets/CSV/l_shorm_ex04_1_0178d.csv')

fhs2_melt_shorm_ex04 <-  
  melt(fhs2_shorm_ex04,
       id.vars=c("PID","IDTYPE"),
       na.rm=T,
       factorsAsStrings = T)

fhs2_melt_shorm_ex04[,visit := 4]
fhs2_melt_shorm_ex04[,form := "fhs2_shorm_ex04"]

rm(fhs2_shorm_ex04)


#=================================#
####  *** Offspring exam 5 *** ####
#=================================#

fhs2_ex1_5d <- fread('~/Dropbox/BioLINCC files/Framingham offspring/datasets/CSV/ex1_5d_v1.csv',na.strings=c("NA","","NULL"))  # Offspring, Exam 5
fhs2_ex1_5d[,visit := 5]

fhs2_ex1_5d[,drinks_wk := 
  E310+
  E313+
  E316]

fhs2_ex1_5d[!between(E274,19,61), E274 := NA]
fhs2_ex1_5d[!between(E276,19,67), E276 := NA]
fhs2_ex1_5d[!between(E278,0,11), E278 := NA]
fhs2_ex1_5d[!between(E279,19,51), E279 := NA]

fhs2_melt_ex1_5d <- 
  melt(fhs2_ex1_5d,
       id.vars=c("PID","IDTYPE","visit"), 
       na.rm=T, 
       factorsAsStrings=T)

fhs2_melt_ex1_5d[,form := "fhs2_offspring_ex05"]

rm(fhs2_ex1_5d)


#=============================#
####   Lab panel, exam 5   ---#
#=============================#

fhs2_l_fhslab_ex05 <- 
  fread('~/Dropbox/BioLINCC files/Framingham offspring/datasets/CSV/l_fhslaba_ex05_1_0578d.csv',
        na.strings=c("NA","","NULL"))

setnames(fhs2_l_fhslab_ex05,"idtype","IDTYPE")

fhs2_l_fhslab_ex05[,visit := 5]

fhs2_melt_l_fhslab_ex05 <- 
  melt(fhs2_l_fhslab_ex05,
       id.vars=c("PID","IDTYPE","visit"), 
       na.rm=T, 
       factorsAsStrings=T)

fhs2_melt_l_fhslab_ex05[,form := "fhs2_fhslab_ex05"]

rm(fhs2_l_fhslab_ex05)


# ============================================##
####     Physical activity, exam 5          ---#
# ============================================##

fhs2_act1_5d <- 
  fread('~/Dropbox/BioLINCC files/Framingham offspring/datasets/CSV/act1_5d_v1.csv',
        na.strings=c("NA","","NULL")) # Physical exam questionnaire, exam 5

setnames(fhs2_act1_5d, "EXAM","visit")

fhs2_act1_5d[,visit := 5]

fhs2_act1_5d[,totkcd := TOTKCW/7]

fhs2_act1_5d[,fhs_pai := 
  calc_framingham_pai(
    as.data.frame(fhs2_act1_5d),
    slp_hrs="EH_10A",
    sed_hrs="EH_10B",
    slgt_hrs="EH_10C",
    mod_hrs="EH_10D",
    hvy_hrs="EH_10E")]

fhs2_melt_act1_5d <- 
  melt(fhs2_act1_5d,
       id.vars=c("PID","IDTYPE","visit"),
       na.rm=T,
       factorsAsStrings=T)

fhs2_melt_act1_5d[,form := "fhs2_act1_5"]

rm(fhs2_act1_5d)


#===========================#
####     Echo, Exam 5    ---#
#===========================#

fhs2_t_echo_ex05 <- 
  fread('~/Dropbox/BioLINCC files/Framingham offspring/datasets/CSV/t_echo_ex05_1_0206d.csv',
        na.strings=c("NA","","NULL"))

setnames(fhs2_t_echo_ex05, "idtype","IDTYPE")
fhs2_t_echo_ex05[,visit := 5]


fhs2_meas5 <- 
  fhs2_offspring_wkthru[
    ,
    .(
      PID,
      IDTYPE,
      SEX,
      HEIGHT = HGT1 * 2.54,
      WEIGHT = WGT5 / 2.2
    )
  ]


fhs2_t_echo_ex05 <- fhs2_meas5[fhs2_t_echo_ex05,on=.(PID,IDTYPE)]

fhs2_t_echo_ex05 <- 
  as.data.table(
    calc_hypertrophy_type(
      as.data.frame(fhs2_t_echo_ex05), 
      id = "PID",
      sex="SEX",
      lvedd="LVDD5",
      ivsd="IVSD5",
      lvpwtd="LVPD5",
      height="HEIGHT",
      weight="WEIGHT"))

fhs2_melt_t_echo_ex05 <- 
  as.data.table(
    melt(
      fhs2_t_echo_ex05,
      id.vars=c("PID","IDTYPE","visit"), 
      na.rm=T, 
      factorsAsStrings=T))

fhs2_melt_t_echo_ex05[,form := "fhs2_echo_ex05"]

rm(fhs2_t_echo_ex05,
   fhs2_meas5)


#=============================================#
####  Echo mitral valve prolapse, Exam 5   ---#
#=============================================#

fhs2_t_echomvp_ex05 <- fread('~/Dropbox/BioLINCC files/Framingham offspring/datasets/CSV/t_echomvp_ex05_1_0753d.csv',na.strings=c("NA","","NULL"))
setnames(fhs2_t_echomvp_ex05, "idtype","IDTYPE")
fhs2_t_echomvp_ex05[,visit := 5]
fhs2_melt_t_echomvp_ex05 <- melt(fhs2_t_echomvp_ex05,id.vars=c("PID","IDTYPE","visit"), na.rm=T, factorsAsStrings=T)
fhs2_melt_t_echomvp_ex05[,form := "fhs2_t_echomvp_ex05"]

rm(fhs2_t_echomvp_ex05)


#=================================================#
####    Validated food frequency data, Exam 5  ---#
#=================================================#

fhs2_vr_ffreq_ex05 <- fread('~/Dropbox/BioLINCC files/Framingham offspring/datasets/CSV/vr_ffreq_ex05_1_0575d.csv',na.strings=c("NA","","NULL"))
names(fhs2_vr_ffreq_ex05) <- toupper(names(fhs2_vr_ffreq_ex05))
fhs2_vr_ffreq_ex05[,visit := 5]

fhs2_melt_vr_ffreq_ex05 <- melt(fhs2_vr_ffreq_ex05,id.vars=c("PID","IDTYPE","visit"), na.rm=T, factorsAsStrings=T)
fhs2_melt_vr_ffreq_ex05[,form := "fhs2_vr_ffreq"]

rm(fhs2_vr_ffreq_ex05)


#================================#
#### *** Offspring exam 6 *** ####
#================================#

fhs2_ex1_6d <- 
  fread('~/Dropbox/BioLINCC files/Framingham offspring/datasets/CSV/ex1_6d_v1.csv',
        na.strings=c("NA","","NULL"))  # Offspring, Exam 6

fhs2_ex1_6d[,visit := 6]

fhs2_ex1_6d[,drinks_wk := 
  F276+
  F279+
  F285]

fhs2_ex1_6d[,CESD20_missing := rowSums(is.na(fhs2_ex1_6d[,c(paste0("F",151:170))]))]
fhs2_ex1_6d[,CESD20 := rowSums(fhs2_ex1_6d[,c(paste0("F",151:170))])]
fhs2_ex1_6d[CESD20_missing >= 4, CESD20 := NA]

fhs2_ex1_6d[!between(F238,20,67), F238 := NA]
fhs2_ex1_6d[!between(F242,20,81), F242 := NA]
fhs2_ex1_6d[!between(F244,1,11), F244 := NA]
fhs2_ex1_6d[!between(F245,19,51), F245 := NA]


fhs2_melt_ex1_6d <- melt(fhs2_ex1_6d,id.vars=c("PID","IDTYPE","visit"), na.rm=T, factorsAsStrings=T)
fhs2_melt_ex1_6d[,form := "fhs2_offspring_ex06"]

rm(fhs2_ex1_6d)

#==============================#
####  Childbirth, Exam 6    ---#
#==============================#

fhs2_ex1_bwgt <- 
  fread('~/Dropbox/BioLINCC files/Framingham offspring/datasets/CSV/bwgt1_6d_v1.csv',
        na.strings=c("NA","","NULL"))  
fhs2_ex1_bwgt[,visit := 6]

fhs2_ex1_bwgt[,age_lastbirth := BW79]

fhs2_ex1_bwgt[is.na(age_lastbirth), age_lastbirth := BW79]

fhs2_ex1_bwgt[is.na(age_lastbirth), age_lastbirth := BW66]

fhs2_ex1_bwgt[is.na(age_lastbirth), age_lastbirth := BW53]

fhs2_ex1_bwgt[is.na(age_lastbirth), age_lastbirth := BW40]

fhs2_ex1_bwgt[is.na(age_lastbirth), age_lastbirth := BW27]


fhs2_melt_ex1_bwgt <- melt(fhs2_ex1_bwgt,id.vars=c("PID","IDTYPE","visit"), na.rm=T, factorsAsStrings=T)
fhs2_melt_ex1_bwgt[,form := "fhs2_offspring_bwgt"]

rm(fhs2_ex1_bwgt)


#==============================#
####  Aldosterone, Exam 6   ---#
#==============================#

fhs2_l_aldost_ex06 <- fread('~/Dropbox/BioLINCC files/Framingham offspring/datasets/CSV/l_aldost_ex06_1b_0046d_v1.csv',na.strings=c("NA","","NULL"))
fhs2_l_aldost_ex06[,visit := 6]
fhs2_melt_l_aldost_ex06 <- melt(fhs2_l_aldost_ex06,id.vars=c("PID","IDTYPE","visit"), na.rm=T, factorsAsStrings=T)
fhs2_melt_l_aldost_ex06[,form := "fhs2_offspring_aldost_ex06"]

rm(fhs2_l_aldost_ex06)

# ========================================##
####    Physical Activity, Exam 6       ---#
# ========================================##

fhs2_act1_6d <- fread('~/Dropbox/BioLINCC files/Framingham offspring/datasets/CSV/act1_6d_v1.csv',na.strings=c("NA","","NULL")) # Physical exam questionnaire, exam 6
fhs2_act1_6d[,visit := 6]
fhs2_melt_act1_6d <- 
  melt(fhs2_act1_6d,
       id.vars=c("PID",
                 "IDTYPE",
                 "visit"),
       na.rm=T,
       factorsAsStrings=T)
fhs2_melt_act1_6d[,form := "fhs2_act1_6"]

rm(fhs2_act1_6d)


#=============================#
####   Lab Panel, Exam 6   ---#
#=============================#

fhs2_l_fhslab_ex06 <- fread('~/Dropbox/BioLINCC files/Framingham offspring/datasets/CSV/l_fhslaba_ex06_1_0579d.csv',na.strings=c("NA","","NULL"))
setnames(fhs2_l_fhslab_ex06, "idtype","IDTYPE")
fhs2_l_fhslab_ex06[,visit := 6]
fhs2_melt_l_fhslab_ex06 <- melt(fhs2_l_fhslab_ex06,id.vars=c("PID","IDTYPE","visit"), na.rm=T, factorsAsStrings=T)
fhs2_melt_l_fhslab_ex06[,form := "fhs2_fhslab_ex06"]

rm(fhs2_l_fhslab_ex06)

#=======================================#
####  Natriuretic peptide, Exam 6    ---#
#=======================================#

fhs2_natpep1 <- fread('~/Dropbox/BioLINCC files/Framingham offspring/datasets/CSV/natpep1_6d_v1.csv',na.strings=c("NA","","NULL"))
setnames(fhs2_natpep1, "idtype","IDTYPE")
fhs2_natpep1[,visit := 6]
fhs2_melt_natpep1 <- melt(fhs2_natpep1,id.vars=c("PID","IDTYPE","visit"), na.rm=T, factorsAsStrings=T)
fhs2_melt_natpep1[,form := "fhs2_natpep1_ex06"]


rm(fhs2_natpep1)

# ================================================#
####               Galectin Exam 6             ---#
# ================================================#


fhs2_l_gal3_ex06 <- fread('~/Dropbox/BioLINCC files/Framingham offspring/datasets/CSV/l_gal3_ex06_1_0623d.csv',na.strings=c("NA","","NULL"))
fhs2_l_gal3_ex06[,visit := 6]

fhs2_melt_l_gal3_ex06 <- melt(fhs2_l_gal3_ex06,id.vars=c("PID","IDTYPE","visit"), na.rm=T, factorsAsStrings=T)
fhs2_melt_l_gal3_ex06[,form := 'fhs2_gal3_ex06']

rm(fhs2_l_gal3_ex06)

#==================================#
####  CKD biomarkers, Exam 6    ---#
#==================================#

fhs2_ckd_ex06_1 <- fread('~/Dropbox/BioLINCC files/Framingham offspring/datasets/CSV/l_ckd_ex06_1_0536d.csv',na.strings=c("NA","","NULL"))
setnames(fhs2_ckd_ex06_1,"idtype", "IDTYPE")
fhs2_ckd_ex06_1[,visit := 6]
fhs2_melt_l_ckd_ex06 <- melt(fhs2_ckd_ex06_1,id.vars=c("PID","IDTYPE","visit"), na.rm=T, factorsAsStrings=T)
fhs2_melt_l_ckd_ex06[,form := "fhs2_ckd_ex06"]

rm(fhs2_ckd_ex06_1)

#================================#
####  Echo, Exam 6            ---#
#================================#

fhs2_t_echo_ex06 <- fread('~/Dropbox/BioLINCC files/Framingham offspring/datasets/CSV/t_echo_ex06_1b_0207d_v2.csv',na.strings=c("NA","","NULL"))
fhs2_t_echo_ex06[,visit := 6]

fhs2_meas6 <- 
  fhs2_offspring_wkthru[
    ,
    .(
      PID,
      IDTYPE,
      SEX,
      HEIGHT = HGT1 * 2.54,
      WEIGHT = WGT6 / 2.2
    )
  ]

fhs2_t_echo_ex06 <- fhs2_meas6[fhs2_t_echo_ex06,on=.(PID,IDTYPE)]


fhs2_t_echo_ex06 <- 
  as.data.table(
    calc_hypertrophy_type(
      fhs2_t_echo_ex06, 
      id = "PID",
      sex="SEX",
      lvedd="LMLVD_DV",
      ivsd="LMIVS_DV",
      lvpwtd="LMLVP_DV",
      height="HEIGHT",
      weight="WEIGHT"))

fhs2_melt_t_echo_ex06 <- melt(fhs2_t_echo_ex06,id.vars=c("PID","IDTYPE","visit"), na.rm=T, factorsAsStrings=T)
fhs2_melt_t_echo_ex06[,form := "fhs2_echo_ex06"]

rm(fhs2_t_echo_ex06,
   fhs2_meas6)


#==============================#
####       CRP, Exam 6      ---#
#==============================#


fhs2_l_crp_ex06 <- fread('~/Dropbox/BioLINCC files/Framingham offspring/datasets/CSV/l_crp_ex06_1_0091d.csv',na.strings=c("NA","","NULL"))
setnames(fhs2_l_crp_ex06, "idtype","IDTYPE")
fhs2_l_crp_ex06[,visit := 6]
fhs2_melt_l_crp_ex06 <- melt(fhs2_l_crp_ex06,id.vars=c("PID","IDTYPE","visit"), na.rm=T, factorsAsStrings=T)
fhs2_melt_l_crp_ex06[,form := "fhs2_crp_ex06"]

rm(fhs2_l_crp_ex06)


#=================================================#
####  Validated food frequency data, Exam 6    ---#
#=================================================#

fhs2_vr_ffreq_ex06 <- fread('~/Dropbox/BioLINCC files/Framingham offspring/datasets/CSV/vr_ffreq_ex06_1_0506d.csv',na.strings=c("NA","","NULL"))
setnames(fhs2_vr_ffreq_ex06, "idtype","IDTYPE")
fhs2_vr_ffreq_ex06[,visit := 6]

fhs2_melt_vr_ffreq_ex06 <- melt(fhs2_vr_ffreq_ex06,id.vars=c("PID","IDTYPE","visit"), na.rm=T, factorsAsStrings=T)
fhs2_melt_vr_ffreq_ex06[,form := "fhs2_vr_ffreq"]

rm(fhs2_vr_ffreq_ex06)


#================================#
#### *** Offspring exam 7 *** ####
#================================#

fhs2_ex1_7d <- fread('~/Dropbox/BioLINCC files/Framingham offspring/datasets/CSV/ex1_7d_v2.csv',na.strings=c("NA","","NULL"))   # Offspring, Exam 7
names(fhs2_ex1_7d)[names(fhs2_ex1_7d)=="idtype"] <- "IDTYPE"


fhs2_obsper_2005 <- fread('~/Dropbox/BioLINCC files/Framingham offspring/datasets/CSV/t_physf_2005_m_0162d_v2.csv',na.strings=c("NA","","NULL"))  # Omni 1, Exam 1
setnames(fhs2_obsper_2005, "idtype","IDTYPE")
names(fhs2_obsper_2005)[names(fhs2_obsper_2005)=="idtype"] <- "IDTYPE"

offspring_30 <- fhs_dates_long[cohort_name=="Offspring"&visit_yr==30,.(PID,visitdays)]
offspring_35 <- fhs_dates_long[cohort_name=="Offspring"&visit_yr==35,.(PID,visitdays)]

names(offspring_30)[2] <- "visitdays_30"
names(offspring_35)[2] <- "visitdays_35"

fhs2_obsper_2005 <- 
  offspring_30[fhs2_obsper_2005,
        on=.(PID)]

fhs2_obsper_2005 <- 
  offspring_35[fhs2_obsper_2005,
        on=.(PID)]

fhs2_obsper_2005_ex30 <- 
  fhs2_obsper_2005[
    mridate >= visitdays_30 &
      mridate <  visitdays_35,
    .SD[which.min(mridate)],
    by = .(PID, IDTYPE)
  ]


fhs2_ex1_7d <- 
  fhs2_obsper_2005_ex30[fhs2_ex1_7d,
                        on=.(PID,IDTYPE)]


fhs2_survca_first <- fhs2_cancer[
  TOPO != 173,                    # filter out TOPO == 173
  .(D_DATE = min(D_DATE)),       # compute first D_DATE
  by = .(PID, IDTYPE)
]


fhs2_ex1_7d[,drinks_wk := 
  G104+
  G107+
  G110+
  G113]

fhs2_ex1_7d[,bsnq_total := 
  G645+
  G646+
  G647+
  G648+
  G649+
  G650+
  G651+
  G652+
  G653+
  G654+
  G655+
  G656+
  G657]

fhs2_ex1_7d[!between(G078,22,62), G078 := NA]
fhs2_ex1_7d[!between(G083,20,83), G083 := NA]
fhs2_ex1_7d[!between(G085,1,11), G085 := NA]
fhs2_ex1_7d[!between(G086,21,47), G086 := NA]

fhs2_ex1_7d[between(G086,1,40)&!is.na(G086), G092 := G092*12]


fhs2_ex1_7d <- 
  fhs2_offspring_wkthru[,c("PID",
                           "AGE7",
                           "SEX",
                           "DMRX7",
                           "HRX7",
                           "DATE7",
                           "HGT1",
                           "WGT7",
                           "BMI7",
                           "CREAT7",
                           "WGT6")][fhs2_ex1_7d,
                                    on=.(PID) ]


fhs2_ex1_7d <- 
  fhs_survcvd[fhs2_ex1_7d, 
              on=.(PID,IDTYPE)]

fhs2_ex1_7d <- 
  fhs_survstk[fhs2_ex1_7d, 
                     on=.(PID,IDTYPE)]

fhs2_ex1_7d <- 
  fhs2_survca_first[fhs2_ex1_7d,
                     on=.(PID,IDTYPE)]


fhs2_ex1_7d[,CESD20_missing := rowSums(is.na(fhs2_ex1_7d[,c(paste0("G",587:606))]))]
fhs2_ex1_7d[,CESD20 := rowSums(fhs2_ex1_7d[,c(paste0("G",587:606))])]
fhs2_ex1_7d[CESD20_missing >= 4, CESD20 := NA]


####### FRIED frailty score

## Walking speed

# Uses greater of 2 speeds (PMID 26695510)

### Men:
# Height ≤ 173 cm (68.1 in):  ≤0.96 m/s
# Height >173 cm (68.1 in):   ≤1.04  m/s


fhs2_ex1_7d[,gaitspeed_1 := 4/mri339]
fhs2_ex1_7d[,gaitspeed_2 := 4/mri341]

fhs2_ex1_7d[,gaitspeed_avg := 
              round(
                apply(
                  fhs2_ex1_7d[,.(gaitspeed_1,gaitspeed_2)],
                                         MARGIN=1,
                                         mean,
                                         na.rm=T),3)]

fhs2_ex1_7d[gaitspeed_avg=="NaN", gaitspeed_avg := NA]

fhs2_ex1_7d[,gaitspeed_max := 
              round(
                apply(
                  fhs2_ex1_7d[,.(gaitspeed_1,gaitspeed_2)],
                                         MARGIN=1,max),3)]

fhs2_ex1_7d[gaitspeed_max=="-Inf", gaitspeed_max := NA]

fhs2_ex1_7d[!is.na(gaitspeed_1)|
              !is.na(gaitspeed_2), gaitspeed_fried := 0]

fhs2_ex1_7d[SEX==1&
              HGT1 >68.1&
              gaitspeed_max <=1.04, gaitspeed_fried := 1]

fhs2_ex1_7d[SEX==1&
              HGT1 <= 68.1&
              gaitspeed_max <= 0.96,gaitspeed_fried := 1]


### Women
# Height ≤ 159 cm (62.6 in):  ≤0.90m/s
# Height >159 cm (62.6 in):   ≤1.02 m/s


fhs2_ex1_7d[SEX==2&
              HGT1 >62.6&
              gaitspeed_max <= 1.02, gaitspeed_fried := 1]

fhs2_ex1_7d[SEX==2&
            HGT1 <= 62.6&
              gaitspeed_max <= 0.90, gaitspeed_fried := 1]


########### Grip (kg)

# Used max of all trials per visit  (PMID 26695510)


## Uses BMI for normalization

##### Men
# BMI ≤ 24.0:     ≤ 29 kg
# BMI 24.1-26.0:  ≤ 30 kg
# BMI 26.1-28.0:  ≤ 30 kg
# BMI > 28.0:     ≤ 32 kg

##### Women
# BMI ≤ 23.0:     ≤ 17
# BMI 23.1-26.0:  ≤ 17.3 kg
# BMI 26.1-29.0:  ≤ 18 kg
# BMI > 29.0:     ≤ 21 kg

fhs2_ex1_7d[,grip_avg_right := round(apply(fhs2_ex1_7d[,.(mri356,mri357,mri358)],MARGIN=1,mean),1)]
fhs2_ex1_7d[,grip_avg_left := round(apply(fhs2_ex1_7d[,.(mri360,mri361,mri362)],MARGIN=1,mean),1)]
fhs2_ex1_7d[,grip_max := round(apply(fhs2_ex1_7d[,.(grip_avg_right,grip_avg_left)],MARGIN=1,max),1)]


##### Men

fhs2_ex1_7d[BMI7<=24&
              grip_max <= 29&
              SEX==1, grip_fried := 1]

fhs2_ex1_7d[BMI7>24&
              BMI7<=26&
              grip_max <= 30&
              SEX==1,grip_fried := 1]

fhs2_ex1_7d[BMI7>26&
              BMI7<=28&
              grip_max <= 30&
              SEX==1, grip_fried := 1]

fhs2_ex1_7d[BMI7>28&
              grip_max <= 32&
              SEX==1, grid_fried := 1]

##### Women

fhs2_ex1_7d[BMI7<=23&
              grip_max <= 17&
              SEX==2, grip_fried := 1]

fhs2_ex1_7d[BMI7>23&
              BMI7<=26&
              grip_max <= 17.3&
              SEX==2, grip_fried := 1]

fhs2_ex1_7d[BMI7>26&
              BMI7<=29&
              grip_max <= 18&
              SEX==2, grip_fried := 1]

fhs2_ex1_7d[BMI7>29&
              grip_max <= 21&
              SEX==2, grip_fried := 1]

fhs2_ex1_7d[G606 %in% c(0,1)|G593 %in% c(0,1), exhaustion_fried := 0]
fhs2_ex1_7d[G606 %in% c(2,3)|G593 %in% c(2,3), exhaustion_fried := 1]


#### Weight loss - Fried

fhs2_ex1_7d[,wt_loss_pct := round((WGT6 - WGT7)/WGT6*100,1)]
fhs2_ex1_7d[wt_loss_pct >= 5, wtloss_fried := 1]
fhs2_ex1_7d[wt_loss_pct < 5, wtloss_fried := 0]


#### Physical activity - Fried

fhs2_ex1_7d[,fhs_pai := 
  calc_framingham_pai(as.data.frame(fhs2_ex1_7d),
                      slp_hrs = "G689",
                      sed_hrs = "G690",
                      slgt_hrs = "G691",
                      mod_hrs = "G692",
                      hvy_hrs = "G693")]

fhs2_ex1_7d[SEX==1&
              fhs_pai < 30.3, 
            activity_fried := 1]

fhs2_ex1_7d[SEX==1&
              fhs_pai >= 30.3, 
            activity_fried := 0]


fhs2_ex1_7d[SEX==2&
              fhs_pai < 30.1, 
            activity_fried := 1]

fhs2_ex1_7d[SEX==2&
              fhs_pai >= 30.1, 
            activity_fried := 0]


#### Calculate Fried score

fhs2_ex1_7d[,total_fried :=
  gaitspeed_fried+
  grip_fried+
  exhaustion_fried+
  wtloss_fried+
  activity_fried]


######### FRAIL Score

#### Fatigue

fhs2_ex1_7d[G606 %in% c(0,1)|G593 %in% c(0,1), fatigue_frail := 0]
fhs2_ex1_7d[G606 %in% c(2,3)|G593 %in% c(2,3)] <- 1


#### Resistance

fhs2_ex1_7d[G525==0,resistance_frail := 0]
fhs2_ex1_7d[G525 %in% c(1,2,3,4), resistance_frail := 1]


#### Ambulation

fhs2_ex1_7d[G548==1, ambulate_frail := 0]
fhs2_ex1_7d[G548==0, ambulate_frail := 1]


# FRAIL Illness score counts presence of HTN, DM, cancer, chronic lung disease, MI, CHD, angina, asthma, arthritis, stroke, and ckd
# Scores for 0-4 conditions, score = 0, for ≥ 5, score=1

# HTN

fhs2_ex1_7d[HRX7==0, htn_frail := 0]
fhs2_ex1_7d[HRX7==1, htn_frail := 1]


# COPD (chronic lung disease)

fhs2_ex1_7d[G415 %in% c(0,2), copd_frail := 0]
fhs2_ex1_7d[G415==1, copd_frail := 1]

# Asthma

fhs2_ex1_7d[G418 %in% c(0,2), asthma_frail := 0]
fhs2_ex1_7d[G418==1, asthma_frail := 1]


# DJD (arthritis)

fhs2_ex1_7d[G421 %in% c(0,2), djd_frail := 0]
fhs2_ex1_7d[G421==1, djd_frail := 1]


# CKD (renal disease)

fhs2_ex1_7d <-
  fhs2_race_1[fhs2_ex1_7d,
        on=.(PID,IDTYPE)]

fhs2_ex1_7d[,crcl := 
  calc_MDRD4(dat=as.data.frame(fhs2_ex1_7d),
             age="AGE7",
             cr="CREAT7",
             black="B")]

fhs2_ex1_7d[crcl >= 60, ckd_frail := 0]
fhs2_ex1_7d[crcl < 60, ckd_frail := 1]


# CHF/MI

fhs2_ex1_7d <- 
  fhs2_survcvd[fhs2_ex1_7d,
        on=.(PID,
             IDTYPE)]

fhs2_ex1_7d[,chf_frail := 0]
fhs2_ex1_7d[hfhosp_status==1&hfhosp_dt <= DATE7, chf_frail := 1]

fhs2_ex1_7d[,chd_frail := 0]
fhs2_ex1_7d[cadhosp_status==1&cadhosp_dt <= DATE7, chd_frail := 1]


# Stroke

fhs2_ex1_7d[,stroke_frail := 0]
fhs2_ex1_7d[cvahosp_status==1&cvahosp_dt <= DATE7, stroke_frail := 1]


# Cancer

fhs2_ex1_7d[,cancer_frail := 0] 
fhs2_ex1_7d[D_DATE <= DATE7, cancer_frail := 1]


# Sum conditions

fhs2_ex1_7d[,conditions_frail :=
  htn_frail+
  DMRX7+
  copd_frail+
  asthma_frail+
  djd_frail+
  ckd_frail+
  chf_frail+
  chd_frail+
  stroke_frail+
  cancer_frail]

fhs2_ex1_7d[conditions_frail <= 4, illness_frail := 0]
fhs2_ex1_7d[conditions_frail > 4, illness_frail := 1]

#### (Weight) Loss

fhs2_ex1_7d[wt_loss_pct >= 5, wtloss_frail := 1]
fhs2_ex1_7d[wt_loss_pct < 5] <- 0


#### Calculate FRAIL score

fhs2_ex1_7d[,total_frail := 
  fatigue_frail+
  resistance_frail+
  ambulate_frail+
  illness_frail+
  wtloss_frail]


#### 

fhs2_melt_ex1_7d <- melt(fhs2_ex1_7d,id.vars=c("PID","IDTYPE"), na.rm=T, factorsAsStrings=T)
fhs2_melt_ex1_7d[,form := "fhs2_offspring_ex07"]

fhs2_melt_ex1_7d[,visit := 7]

rm(fhs2_ex1_7d)


#=================================#
####     Cardiac MRI exam 7    ---#
#=================================#

fhs2_cmrlvh1 <- fread('~/Dropbox/BioLINCC files/Framingham offspring/datasets/CSV/t_mrcdlvh_2006_1b_0907d_v1.csv',na.strings=c("NA","","NULL")) # Offspring cardiac MRI LVH 
fhs2_cmrlvh1[,visit := 7]

fhs2_cmrlvh1[,CMR03_cm := CMR03/10]
fhs2_cmrlvh1[,CMR04_cm := CMR04/10]
fhs2_cmrlvh1[,CMR05_cm := CMR05/10]

fhs2_cmrlvh1[,rwt := 2*CMR04/CMR05]


fhs2_meas7 <- fhs2_offspring_wkthru[,c("PID","IDTYPE","SEX","HGT1","WGT7")]

fhs2_meas7[,weight_kg := WGT7/2.2]
fhs2_meas7[,height_cm := HGT1*2.54]


fhs2_meas7[,BSA := calc_bsa(dat=fhs2_meas7)]

fhs2_cmrlvh1 <- 
  fhs2_meas7[fhs2_cmrlvh1,
             on=.(PID,IDTYPE)]

fhs2_cmrlvh1[,lvmass_ix := CMR22/BSA]

fhs2_cmrlvh1[SEX==1&lvmass_ix>115&rwt<=0.42&!is.na(lvmass_ix)&!is.na(rwt), lvh_type := "Eccentric hypertropy"]
fhs2_cmrlvh1[SEX==2&lvmass_ix>95&rwt<=0.42&!is.na(lvmass_ix)&!is.na(rwt), lvh_type := "Eccentric hypertropy"]

fhs2_cmrlvh1[SEX==1&lvmass_ix>115&rwt>0.42&!is.na(lvmass_ix)&!is.na(rwt), lvh_type := "Concentric hypertropy"]
fhs2_cmrlvh1[SEX==2&lvmass_ix>95&rwt>0.42&!is.na(lvmass_ix)&!is.na(rwt), lvh_type := "Concentric hypertropy"]

fhs2_cmrlvh1[SEX==1&lvmass_ix<=115&rwt<=0.42&!is.na(lvmass_ix)&!is.na(rwt), lvh_type := "Normal geometry"]
fhs2_cmrlvh1[SEX==2&lvmass_ix<=95&rwt<=0.42&!is.na(lvmass_ix)&!is.na(rwt), lvh_type := "Normal geometry"]

fhs2_cmrlvh1[SEX==1&lvmass_ix<=115&rwt>0.42&!is.na(lvmass_ix)&!is.na(rwt), lvh_type := "Concentric remodeling"]
fhs2_cmrlvh1[SEX==2&lvmass_ix<=95&rwt>0.42&!is.na(lvmass_ix)&!is.na(rwt), lvh_type := "Concentric remodeling"]

fhs2_melt_cmrlvh1 <- 
  melt(fhs2_cmrlvh1,id.vars=c("PID",
                              "IDTYPE",
                              "visit"),
       na.rm=T,
       factorsAsStrings=T)

fhs2_melt_cmrlvh1[,form := "fhs2_cmrlvh1"]

rm(fhs2_cmrlvh1)


### MRI wall motion abnormalities

fhs2_cmrwma1 <- fread('~/Dropbox/BioLINCC files/Framingham offspring/datasets/CSV/t_mrcdwma_2006_1b_0909d_v1.csv',na.strings=c("NA","","NULL"))  # Offspring cardiac MRI wall motion abnormalities
fhs2_cmrwma1[,visit := 7]
fhs2_melt_cmrwma1 <- melt(fhs2_cmrwma1,id.vars=c("PID","IDTYPE","visit"),na.rm=T,factorsAsStrings=T)
fhs2_melt_cmrwma1[,form := "fhs2_cmrwma1"]

rm(fhs2_cmrwma1)


#=========================#
####  DM labs, Exam 7  ---#
#=========================#

fhs2_l_dbtlab_ex07 <- fread('~/Dropbox/BioLINCC files/Framingham offspring/datasets/CSV/l_dbtlab_ex07_1b_1237d_v1.csv',na.strings=c("NA","","NULL"))
setnames(fhs2_l_dbtlab_ex07,"EXAM","visit")
fhs2_melt_l_dbtlab_ex07 <- melt(fhs2_l_dbtlab_ex07,id.vars=c("PID","IDTYPE","visit"), na.rm=T, factorsAsStrings=T)
fhs2_melt_l_dbtlab_ex07[,form := "fhs2_dbtlab_ex07"]

rm(fhs2_l_dbtlab_ex07)


#======================================#
####      Lab panel, Exam 7         ---#
#======================================#

fhs2_l_fhslab_ex07 <- fread('~/Dropbox/BioLINCC files/Framingham offspring/datasets/CSV/l_fhslaba_ex07_1_0284d.csv',na.strings=c("NA","","NULL"))
setnames(fhs2_l_fhslab_ex07, "idtype","IDTYPE")
fhs2_l_fhslab_ex07[,visit := 7]
fhs2_melt_l_fhslab_ex07 <- melt(fhs2_l_fhslab_ex07,id.vars=c("PID","IDTYPE","visit"), na.rm=T, factorsAsStrings=T)
fhs2_melt_l_fhslab_ex07[,form := "fhs2_fhslab_ex07"]

rm(fhs2_l_fhslab_ex07)

#=======================================#
####  Creatinine/cysteine, Exam 7    ---#
#=======================================#

fhs2_creacys_ex07_1 <- fread('~/Dropbox/BioLINCC files/Framingham offspring/datasets/CSV/l_creacys_ex07_1_0092d.csv',na.strings=c("NA","","NULL"))
fhs2_creacys_ex07_1[, visit := 7]
fhs2_melt_l_creacys_ex07 <- melt(fhs2_creacys_ex07_1,id.vars=c("PID","IDTYPE","visit"), na.rm=T, factorsAsStrings=T)
fhs2_melt_l_creacys_ex07[,form := "fhs2_creacys_ex07"]

rm(fhs2_creacys_ex07_1)


#====================================#
####      Walk test, Exam 7       ---#
#====================================#

fhs2_t_wktest_07 <- fread('~/Dropbox/BioLINCC files/Framingham offspring/datasets/CSV/t_wktest_ex07_1_0016d.csv',na.strings=c("NA","","NULL"))
setnames(fhs2_t_wktest_07, "idtype","IDTYPE")
fhs2_t_wktest_07[,visit := 7]
fhs2_melt_t_wktest_07 <- melt(fhs2_t_wktest_07,id.vars=c("PID","IDTYPE","visit"), na.rm=T, factorsAsStrings=T)
fhs2_melt_t_wktest_07[,form := "fhs2_t_wktest"]

rm(fhs2_t_wktest_07)


#================================================================#
####  Estrogen levels, Framingham 3, Exam 1/Omni 2, Exam 1    ---#
#================================================================#

fhs2_estr_ex7 <- fread('~/Dropbox/BioLINCC files/Framingham offspring/datasets/CSV/l_estr_2005_m_0604d.csv')
fhs2_melt_estr_ex7 <-  melt(fhs2_estr_ex7,id.vars=c("PID","IDTYPE"),na.rm=T,factorsAsStrings = T)
fhs2_melt_estr_ex7[,visit := 7]
fhs2_melt_estr_ex7[,form := "fhs2_estr_07"]

rm(fhs2_estr_ex7)


#==============================#
####       CRP, Exam 7      ---#
#==============================#


fhs2_l_inflamm_ex07 <- 
  fread('~/Dropbox/BioLINCC files/Framingham offspring/datasets/CSV/l_inflamm_ex07_1b_1105d.csv',
        na.strings=c("NA","","NULL"))

names(fhs2_l_inflamm_ex07) <- toupper(names(fhs2_l_inflamm_ex07))

fhs2_l_inflamm_ex07[IDTYPE==1, VISIT := 7]
fhs2_l_inflamm_ex07[IDTYPE==7, VISIT := 2]

fhs2_melt_inflamm_ex07 <- 
  melt(fhs2_l_inflamm_ex07,
       id.vars=c("PID","IDTYPE","VISIT"), 
       na.rm=T, 
       factorsAsStrings=T)

names(fhs2_melt_inflamm_ex07)[3] <- "visit"

fhs2_melt_inflamm_ex07[,form := "fhs2_inflamm_ex07"]

rm(fhs2_l_inflamm_ex07)


#====================================================#
####  Validated food frequency data, exam 7       ---#
#====================================================#

fhs2_vr_ffreq_ex07 <- fread('~/Dropbox/BioLINCC files/Framingham offspring/datasets/CSV/vr_ffreq_ex07_1_0460d.csv',na.strings=c("NA","","NULL"))
setnames(fhs2_vr_ffreq_ex07, "idtype","IDTYPE")
fhs2_vr_ffreq_ex07[,visit := 7]
fhs2_melt_vr_ffreq_ex07 <- melt(fhs2_vr_ffreq_ex07,id.vars=c("PID","IDTYPE","visit"), na.rm=T, factorsAsStrings=T)
fhs2_melt_vr_ffreq_ex07[,form := "fhs2_vr_ffreq"]

rm(fhs2_vr_ffreq_ex07)


#================================================================#
####   Male hormones in females, Offspring, Exam 7            ---#
#================================================================#

fhs2_fhorm_ex07 <- fread('~/Dropbox/BioLINCC files/Framingham offspring/datasets/CSV/l_fhorm_2005_m_0493d.csv')
fhs2_melt_fhorm_ex07 <-  melt(fhs2_fhorm_ex07,id.vars=c("PID","IDTYPE"),na.rm=T,factorsAsStrings = T)
fhs2_melt_fhorm_ex07[,visit := 7]
fhs2_melt_fhorm_ex07[,form := "fhs2_fhorm_ex07"]

rm(fhs2_fhorm_ex07)

#================================================================#
####  Male hormones, Offspring, Exam 7      ---#
#================================================================#

fhs2_mhorm_ex07 <- fread('~/Dropbox/BioLINCC files/Framingham offspring/datasets/CSV/l_mhorm_2005_m_0490d_v1.csv')
fhs2_melt_mhorm_ex07 <-  melt(fhs2_mhorm_ex07,id.vars=c("PID","IDTYPE"),na.rm=T,factorsAsStrings = T)
fhs2_melt_mhorm_ex07[,visit <- 7]
fhs2_melt_mhorm_ex07[,form <- "fhs2_mhorm_ex07"]

rm(fhs2_mhorm_ex07)


#==================================#
####  *** Offspring exam 8 ***  ####
#==================================#

fhs2_ex1_8d <- fread('~/Dropbox/BioLINCC files/Framingham offspring/datasets/CSV/e_exam_ex08_1_0005d.csv',na.strings=c("NA","","NULL")) # Omni 1, Exam 8
fhs2_ex1_8d[,visit := 8]
fhs2_ex1_8d <- 
  fhs2_offspring_wkthru[,c("PID",
                           "AGE8",
                           "SEX",
                           "DMRX8",
                           "HRX8",
                           "DATE8",
                           "HGT1",
                           "WGT8",
                           "BMI8",
                           "CREAT8",
                           "WGT7")][fhs2_ex1_8d,
                                    on=.(PID)]

fhs2_ex1_8d[!between(H033,22,83), H033 := NA]
fhs2_ex1_8d[!between(H046,21,77), H046 := NA]

fhs2_ex1_8d[!H047==88&!is.na(H047), hrt_years_total := as.numeric(H047)+round(as.numeric(H048)/12,1)] 

####### FRIED frailty score

## Walking speed

# Uses greater of 2 speeds (PMID 26695510)

fhs2_ex1_8d[,gaitspeed_1 := 4/H617]
fhs2_ex1_8d[,gaitspeed_2 := 4/H620]

fhs2_ex1_8d[,gaitspeed_avg := 
  round(
    apply(
      fhs2_ex1_8d[,.(gaitspeed_1,gaitspeed_2)],
                                         MARGIN=1,
                                         mean,
                                         na.rm=T),3)]

fhs2_ex1_8d[gaitspeed_avg=="NaN", gaitspeed_avg := NA]

fhs2_ex1_8d[,gaitspeed_max := 
              round(
                apply(
                  fhs2_ex1_8d[,.(gaitspeed_1,gaitspeed_2)],
                                         MARGIN=1,max),3)]

fhs2_ex1_8d[gaitspeed_max=="-Inf", gaitspeed_max := NA]

fhs2_ex1_8d[!is.na(gaitspeed_1)|
              !is.na(gaitspeed_2), gaitspeed_fried := 0]

### Men:

# Height ≤ 173 cm (68.1 in):  ≤0.96 m/s
# Height >173 cm (68.1 in):   ≤1.04  m/s

fhs2_ex1_8d[SEX==1&
              HGT1 >68.1&
              gaitspeed_max <=1.04, 
            gaitspeed_fried := 1]

fhs2_ex1_8d[SEX==1&
              HGT1 <= 68.1&
              gaitspeed_max <= 0.96, 
            gaitspeed_fried := 1]

### Women
# Height ≤ 159 cm (62.6 in):  ≤0.90m/s
# Height >159 cm (62.6 in):   ≤1.02 m/s


fhs2_ex1_8d[SEX==2&
              HGT1 >62.6&
              gaitspeed_max <= 1.02, 
            gaitspeed_fried := 1]

fhs2_ex1_8d[SEX==2&
              HGT1 <= 62.6&
              gaitspeed_max <= 0.90, 
            gaitspeed_fried := 1]


########### Grip (kg)

# Used max of all trials per visit  (PMID 26695510)


## Uses BMI for normalization

##### Men
# BMI ≤ 24.0:     ≤ 29 kg
# BMI 24.1-26.0:  ≤ 30 kg
# BMI 26.1-28.0:  ≤ 30 kg
# BMI > 28.0:     ≤ 32 kg

##### Women
# BMI ≤ 23.0:     ≤ 17
# BMI 23.1-26.0:  ≤ 17.3 kg
# BMI 26.1-29.0:  ≤ 18 kg
# BMI > 29.0:     ≤ 21 kg

fhs2_ex1_8d[,grip_avg := round(apply(fhs2_ex1_8d[,.(H605,H606,H607,H608,H609,H610)],MARGIN=1,mean),1)]
fhs2_ex1_8d[,grip_max := round(apply(fhs2_ex1_8d[,.(H605,H606,H607,H608,H609,H610)],MARGIN=1,max),1)]

fhs2_ex1_8d[!is.na(grip_max), grip_fried := 0]


##### Men

fhs2_ex1_8d[BMI8<=24&
              grip_max <= 29&
              SEX==1, 
            grip_fried := 1]

fhs2_ex1_8d[BMI8>24&
              BMI8<=26&
              grip_max <= 30&
              SEX==1, 
            grip_fried := 1]

fhs2_ex1_8d[BMI8>26&
              BMI8<=28&
              grip_max <= 30&
              SEX==1, 
            grip_fried := 1]

fhs2_ex1_8d[BMI8>28&
              grip_max <= 32&
              SEX==1, 
            grip_fried := 1]

##### Women

fhs2_ex1_8d[BMI8<=23&
              grip_max <= 17&
              SEX==2, 
            grip_fried := 1]

fhs2_ex1_8d[BMI8>23&
              BMI8<=26&
              grip_max <= 17.3&
              SEX==2, 
            grip_fried := 1]

fhs2_ex1_8d[BMI8>26&
              BMI8<=29&
              grip_max <= 18&
              SEX==2, 
            grip_fried := 1]

fhs2_ex1_8d[BMI8>29&
              grip_max <= 21&
              SEX==2, 
            grip_fried := 1]

### Fried exhaustion

fhs2_ex1_8d[H471 %in% c(0,1)&H472 %in% c(0,1), exhaustion_fried := 0]
fhs2_ex1_8d[H471 %in% c(2,3)|H472 %in% c(2,3)] <- 1


#### Weight loss - Fried

fhs2_ex1_8d[, wt_loss_pct := round((WGT7 - WGT8)/WGT7*100,1)]
fhs2_ex1_8d[wt_loss_pct >= 5, wtloss_fried := 1]
fhs2_ex1_8d[wt_loss_pct < 5, wtloss_fried := 0]


#### Physical activity - Fried

fhs2_ex1_8d[,fhs_pai := 
  calc_framingham_pai(
    as.data.frame(fhs2_ex1_8d),
    slp_hrs = "H480",
    sed_hrs = "H481",
    slgt_hrs = "H482",
    mod_hrs = "H483",
    hvy_hrs = "H484")]

fhs2_ex1_8d[SEX==1&
              fhs_pai < 30.3, 
            activity_fried := 1]

fhs2_ex1_8d[SEX==1&
              fhs_pai >= 30.3, 
            activity_fried := 0]


fhs2_ex1_8d[SEX==2&
              fhs_pai < 30.1, 
            activity_fried := 1]

fhs2_ex1_8d[SEX==2&
              fhs_pai >= 30.1, 
            activity_fried := 0]


#### Calculate Fried score

fhs2_ex1_8d[,total_fried :=
              gaitspeed_fried+
              grip_fried+
              exhaustion_fried+
              wtloss_fried+
              activity_fried]


fhs2_ex1_8d[,BMIC8 := 
  calc_bmi(dat=as.data.frame(fhs2_ex1_8d), 
           weight="WGT8",
           height="HGT1",
           metric=F)]

fhs2_ex1_8d <- fhs_survcvd[fhs2_ex1_8d, 
                     on=.(PID,IDTYPE)]

fhs2_ex1_8d <- fhs_survstk[fhs2_ex1_8d, 
                     on=.(PID,IDTYPE)]


fhs2_ex1_8d <- 
  fhs_survca_first_any[,.(PID,IDTYPE,D_DATE)][fhs2_ex1_8d,
                     on=.(PID,IDTYPE)]

fhs2_ex1_8d[,drinks_wk := 
  H072+
  H075+
  H078]

fhs2_ex1_8d[,epworth_ss_nas := 
  is.na(H734)+
  is.na(H735)+
  is.na(H736)+
  is.na(H737)+
  is.na(H738)+
  is.na(H739)+
  is.na(H740)+
  is.na(H741)]

fhs2_ex1_8d[,epworth_ss := 
  H734+
  H735+
  H736+
  H737+
  H738+
  H739+
  H740+
  H741]


## Denormalize

fhs2_melt_ex1_8d <- melt(fhs2_ex1_8d,
                         id.vars=c("PID","IDTYPE","visit"), 
                         na.rm=T, 
                         factorsAsStrings=T)

fhs2_melt_ex1_8d[,form := "fhs2_offspring_ex08"]

rm(fhs2_ex1_8d)


#================================#
####     Lab panel, Exam 8    ---#
#================================#

fhs2_l_fhslab_ex08 <- fread('~/Dropbox/BioLINCC files/Framingham offspring/datasets/CSV/l_fhslab_ex08_1_0257d.csv',na.strings=c("NA","","NULL"))
fhs2_l_fhslab_ex08[,visit := 8]
fhs2_melt_l_fhslab_ex08 <- melt(fhs2_l_fhslab_ex08,id.vars=c("PID","IDTYPE","visit"), na.rm=T, factorsAsStrings=T)
fhs2_melt_l_fhslab_ex08[,form := "fhs2_fhslab_ex08"]

rm(fhs2_l_fhslab_ex08)


#==========================================#
####     Echo cardiac strain, Exam 8    ---#
#==========================================#

fhs2_meas8 <- fhs2_offspring_wkthru[,c("PID","IDTYPE","SEX","HGT1","WGT8")]

fhs2_meas8[,weight_kg := WGT8/2.2]
fhs2_meas8[,height_cm := HGT1*2.54]

fhs2_meas8[,BSA := calc_bsa(dat=fhs2_meas8)]


fhs2_t_echocs_ex08 <- fread('~/Dropbox/BioLINCC files/Framingham offspring/datasets/CSV/t_echocs_ex08_1_0705d.csv',na.strings=c("NA","","NULL"))
setnames(fhs2_t_echocs_ex08,"idtype","IDTYPE")
names(fhs2_t_echocs_ex08) <-
  sub("^_","",names(fhs2_t_echocs_ex08))

fhs2_t_echocs_ex08[,visit := 8]
fhs2_melt_t_echocs_ex08 <- melt(fhs2_t_echocs_ex08,id.vars=c("PID","IDTYPE","visit"), na.rm=T, factorsAsStrings=T)
fhs2_melt_t_echocs_ex08[,form := "fhs2_echocs_ex08"]

rm(fhs2_t_echocs_ex08)


#==========================================#
####     Echo RV, Exam 8                ---#
#==========================================#

fhs2_t_echorv_ex08 <- fread('~/Dropbox/BioLINCC files/Framingham offspring/datasets/CSV/t_echorv_ex08_1b_0830d_v1.csv',na.strings=c("NA","","NULL"))
setnames(fhs2_t_echorv_ex08, "idtype","IDTYPE")

fhs2_t_echorv_ex08[IDTYPE==1, visit := 8]
fhs2_t_echorv_ex08[IDTYPE==7, visit := 3]
fhs2_t_echorv_ex08 <-
  fhs2_offspring_wkthru[,.(PID,IDTYPE,BSA8)][fhs2_t_echorv_ex08, on=.(PID,IDTYPE)]

fhs2_t_echorv_ex08[,RVEDA_ix := RVEDA/BSA8]

fhs2_melt_t_echorv_ex08 <- 
  melt(fhs2_t_echorv_ex08,id.vars=c("PID","IDTYPE","visit"), 
       na.rm=T, 
       factorsAsStrings=T)

fhs2_melt_t_echorv_ex08[,form := "fhs2_echorv_ex08"]

rm(fhs2_t_echorv_ex08)


#==========================================#
####     Echo LA, Exam 8                ---#
#==========================================#

fhs2_t_echola_ex08 <- fread('~/Dropbox/BioLINCC files/Framingham offspring/datasets/CSV/t_echola_ex08_1b_1084d.csv',na.strings=c("NA","","NULL"))
setnames(fhs2_t_echola_ex08, "idtype","IDTYPE")

fhs2_t_echola_ex08[IDTYPE==1, visit := 8]
fhs2_t_echola_ex08[IDTYPE==7, visit := 3]


fhs2_t_echola_ex08 <-
  fhs2_meas8[fhs2_t_echola_ex08,
        on=.(PID,IDTYPE)]

fhs2_t_echola_ex08[,lav_ix :=
  Left_atrial_volume_max_average/BSA]

fhs2_melt_t_echola_ex08 <- 
  melt(fhs2_t_echola_ex08,id.vars=c("PID","IDTYPE","visit"), 
       na.rm=T, 
       factorsAsStrings=T)

fhs2_melt_t_echola_ex08[,form := "fhs2_echola_ex08"]

rm(fhs2_t_echola_ex08)


#==========================================#
####     Echo Doppler, Exam 8                ---#
#==========================================#

fhs2_t_echodop_ex08 <- fread('~/Dropbox/BioLINCC files/Framingham offspring/datasets/CSV/t_echodop_ex08_1b_0988d.csv',na.strings=c("NA","","NULL"))

fhs2_t_echodop_ex08[IDTYPE==1, visit := 8]
fhs2_t_echodop_ex08[IDTYPE==7, visit := 3]

fhs2_melt_t_echodop_ex08 <- 
  melt(fhs2_t_echodop_ex08,
       id.vars=c("PID","IDTYPE","visit"), 
       na.rm=T, 
       factorsAsStrings=T)

fhs2_melt_t_echodop_ex08[,form := "fhs2_echodop_ex08"]

rm(fhs2_t_echodop_ex08)


#================================================#
####     Vasc Tonometry, Offspring Exam 8     ---#
#================================================#

fhs2_t_tonla_ex08 <- fread('~/Dropbox/BioLINCC files/Framingham offspring/datasets/CSV/t_tonla_ex08_1b_1232d.csv',na.strings=c("NA","","NULL"))
setnames(fhs2_t_tonla_ex08, "EXAM","visit")

fhs2_t_tonla_ex08[,e_eprime_avg := memax/dlemax]
fhs2_t_tonla_ex08[,ea_ratio := memax/mamax]

fhs2_melt_t_tonla_ex08 <-  
  melt(fhs2_t_tonla_ex08,
       id.vars=c("PID","IDTYPE","visit"),
       na.rm=T,
       factorsAsStrings = T)

fhs2_melt_t_tonla_ex08[,visit := 8]
fhs2_melt_t_tonla_ex08[,form := "fhs2_tonla_ex08"]

rm(fhs2_t_tonla_ex08)


#================================================#
####     Vasc Tonometry, Offspring Exam 9     ---#
#================================================#

fhs2_t_tonla_ex09 <- fread('~/Dropbox/BioLINCC files/Framingham offspring/datasets/CSV/t_tonla_ex09_1b_1233d.csv',na.strings=c("NA","","NULL"))
setnames(fhs2_t_tonla_ex09, "EXAM","visit")

fhs2_t_tonla_ex09[,e_eprime_avg := memax/dlemax]
fhs2_t_tonla_ex09[,ea_ratio := memax/mamax]

fhs2_melt_t_tonla_ex09 <-  
  melt(fhs2_t_tonla_ex09,
       id.vars=c("PID","IDTYPE","visit"),
       na.rm=T,
       factorsAsStrings = T)

fhs2_melt_t_tonla_ex09[,visit := 9]
fhs2_melt_t_tonla_ex09[,form := "fhs2_tonla_ex09"]

rm(fhs2_t_tonla_ex09)


#=========================================================#
####  Validated food frequency data, Exam 8            ---#
#=========================================================#

fhs2_vr_ffreq_ex08 <- fread('~/Dropbox/BioLINCC files/Framingham offspring/datasets/CSV/vr_ffreq_ex08_1_0615d_v1.csv',na.strings=c("NA","","NULL"))
setnames(fhs2_vr_ffreq_ex08, "idtype","IDTYPE")
fhs2_vr_ffreq_ex08[,visit := 8]
fhs2_melt_vr_ffreq_ex08 <- melt(fhs2_vr_ffreq_ex08,id.vars=c("PID","IDTYPE","visit"), na.rm=T, factorsAsStrings=T)
fhs2_melt_vr_ffreq_ex08[,form := "fhs2_vr_ffreq"]

rm(fhs2_vr_ffreq_ex08)


#================================#
####  *** OMNI 1, exam 1 ***  ####
#================================#

fhs2_omni1_ex01 <- fread('~/Dropbox/BioLINCC files/Framingham offspring/datasets/CSV/e_exam_ex01_7_0020d.csv',na.strings=c("NA","","NULL"))  # Omni 1, Exam 1

names(fhs2_omni1_ex01) <- toupper(names(fhs2_omni1_ex01))

fhs2_omni1_ex01[,drinks_wk := 
                  E310+
                  E313+
                  E316]

fhs2_omni1_ex01[,visit := 1]
fhs2_melt_omni1_ex01 <- melt(fhs2_omni1_ex01,id.vars=c("PID","IDTYPE","visit"), na.rm=T, factorsAsStrings=T)
fhs2_melt_omni1_ex01[,form := "fhs2_omni1_ex01"]

rm(fhs2_omni1_ex01)


#===================================#
####   Lab panel, Exam 1         ---#
#===================================#

fhs2_l_fhslab_ex01 <- fread('~/Dropbox/BioLINCC files/Framingham offspring/datasets/CSV/l_fhslab_ex01_7_0270d_v1.csv',na.strings=c("NA","","NULL"))
setnames(fhs2_l_fhslab_ex01, "EXAM","visit")
names(fhs2_l_fhslab_ex01)[names(fhs2_l_fhslab_ex01)=="idtype"] <- "IDTYPE"
fhs2_l_fhslab_ex01[,visit := 1]
fhs2_melt_l_fhslab_ex01 <- melt(fhs2_l_fhslab_ex01,id.vars=c("PID","IDTYPE","visit"), na.rm=T, factorsAsStrings=T)
fhs2_melt_l_fhslab_ex01[,form := "fhs2_fhslab_ex01_7"]

rm(fhs2_l_fhslab_ex01)


#==============================#
#### *** OMNI 1, exam 2 *** ####
#==============================#

fhs2_omni1_ex02 <- fread('~/Dropbox/BioLINCC files/Framingham offspring/datasets/CSV/e_exam_ex02_7_0003d.csv',na.strings=c("NA","","NULL"))  # Omni 1, Exam 2
names(fhs2_omni1_ex02) <- toupper(names(fhs2_omni1_ex02))

fhs2_omni1_ex02 <- 
  fhs2_race_1[,c('PID',
                 'IDTYPE',
                 'RACE',
                 'ETHNICITY')][fhs2_omni1_ex02,
                               on=.(PID,IDTYPE)]

fhs2_omni1_ex02[,drinks_wk := 
  G104+
  G107+
  G110+
  G113]


fhs2_omni1_ex02[,bsnq_total := 
  G645+
  G646+
  G647+
  G648+
  G649+
  G650+
  G651+
  G652+
  G653+
  G654+
  G655+
  G656+
  G657]


fhs2_omni1_ex02[,CESD20_missing := rowSums(is.na(fhs2_omni1_ex02[,c(paste0("G",587:606))]))]
fhs2_omni1_ex02[,CESD20 := rowSums(fhs2_omni1_ex02[,c(paste0("G",587:606))])]
fhs2_omni1_ex02[CESD20_missing >= 4, CESD20 := NA]

fhs2_obsper_2005 <- 
  fread('~/Dropbox/BioLINCC files/Framingham offspring/datasets/CSV/t_physf_2005_m_0162d_v2.csv',
        na.strings=c("NA","","NULL"))  # Omni 1, Exam 1
setnames(fhs2_obsper_2005, "idtype","IDTYPE")

omni1_5 <- fhs_dates_long[cohort_name=="OMNI 1"&visit_yr==5,.(PID,visitdays)]
omni1_10 <- fhs_dates_long[cohort_name=="OMNI 1"&visit_yr==10,.(PID,visitdays)]

setnames(omni1_5, "visitdays", "visitdays_omni1_5")
setnames(omni1_10, "visitdays", "visitdays_omni1_10")

fhs2_obsper_2005 <- 
  omni1_5[fhs2_obsper_2005,
          on=.(PID)]

fhs2_obsper_2005 <- 
  omni1_10[fhs2_obsper_2005,
           on=.(PID)]


fhs2_obsper_2005_omni1_ex02 <- 
  fhs2_obsper_2005[
    mridate >= visitdays_omni1_5 &
      mridate <  visitdays_omni1_10,
    .SD[which.min(mridate)],
    by = .(PID, IDTYPE)
  ]

fhs2_omni1_ex02 <- 
  fhs2_obsper_2005_omni1_ex02[fhs2_omni1_ex02,
                         on=.(PID,IDTYPE)]

fhs2_omni1_ex02 <- 
  fhs2_offspring_wkthru[,c("PID",
                           "IDTYPE",
                           "AGE2",
                           "SEX",
                           "DMRX2",
                           "HRX2",
                           "DATE2",
                           "HGT1",
                           "WGT2",
                           "CREAT2",
                           "WGT1")][fhs2_omni1_ex02,
                         on=.(PID,IDTYPE)]

fhs2_omni1_ex02[,BMI2 := 
  calc_bmi(dat=as.data.frame(fhs2_omni1_ex02), 
           weight="WGT2",
           height="HGT1",
           metric=F)]


fhs2_omni1_ex02 <- 
  fhs_survcvd[fhs2_omni1_ex02,
              on=.(PID,IDTYPE)]

fhs2_omni1_ex02 <- 
  fhs_survstk[fhs2_omni1_ex02, 
        on=.(PID,IDTYPE)]


fhs2_omni1_ex02 <- 
  fhs2_survca_first[fhs2_omni1_ex02,
        on=.(PID,IDTYPE)]


####### FRIED frailty score

## Walking speed

# Uses greater of 2 speeds (PMID 26695510)

### Men:
# Height ≤ 173 cm (68.1 in):  ≤0.96 m/s
# Height >173 cm (68.1 in):   ≤1.04  m/s


fhs2_omni1_ex02[,gaitspeed_1 := 4/(mri339)]
fhs2_omni1_ex02[,gaitspeed_2 := 4/(mri341)]

fhs2_omni1_ex02[,gaitspeed_avg := 
  round(apply(fhs2_omni1_ex02[,.(gaitspeed_1,gaitspeed_2)],
              MARGIN=1,
              mean,
              na.rm=T),3)]

fhs2_omni1_ex02[gaitspeed_avg=="NaN", gaitspeed_avg := NA]

fhs2_omni1_ex02[,gaitspeed_max := 
  round(
    apply(
      fhs2_omni1_ex02[,.(gaitspeed_1,gaitspeed_2)],
      MARGIN=1,max),3)]

fhs2_omni1_ex02[gaitspeed_max=="-Inf", gaitspeed_max := NA]

fhs2_omni1_ex02[
  !is.na(gaitspeed_1)|
    !is.na(gaitspeed_2), gaitspeed_fried := 0]

fhs2_omni1_ex02[SEX==1&
                  HGT1>68.1&
                  gaitspeed_max <=1.04, 
                gaitspeed_fried := 1]

fhs2_omni1_ex02[SEX==1&
                  HGT1 <= 68.1&
                  gaitspeed_max <= 0.96, gaitspeed_fried := 1]


### Women
# Height ≤ 159 cm (62.6 in):  ≤0.90m/s
# Height >159 cm (62.6 in):   ≤1.02 m/s


fhs2_omni1_ex02[SEX==2&
                HGT1>62.6&
                  gaitspeed_max <= 1.02, 
                gaitspeed_fried := 1]

fhs2_omni1_ex02[SEX==2&
                  HGT1 <= 62.6&
                  gaitspeed_max <= 0.90, 
                gaitspeed_fried := 1]


########### Grip (kg)

# Used max of all trials per visit  (PMID 26695510)


## Uses BMI for normalization

##### Men
# BMI ≤ 24.0:     ≤ 29 kg
# BMI 24.1-26.0:  ≤ 30 kg
# BMI 26.1-28.0:  ≤ 30 kg
# BMI > 28.0:     ≤ 32 kg

##### Women
# BMI ≤ 23.0:     ≤ 17
# BMI 23.1-26.0:  ≤ 17.3 kg
# BMI 26.1-29.0:  ≤ 18 kg
# BMI > 29.0:     ≤ 21 kg

fhs2_omni1_ex02[,grip_avg := 
                  round(
                    apply(
                      fhs2_omni1_ex02[,c("mri356",
                                         "mri357",
                                         "mri358",
                                         "mri360",
                                         "mri361",
                                         "mri362")],
                      MARGIN=1,
                      mean),1)]

fhs2_omni1_ex02[,grip_max := round(apply(fhs2_omni1_ex02[,c("mri359","mri363")],MARGIN=1,max),1)]

fhs2_omni1_ex02[!is.na(grip_max), grip_fried := 0]


##### Men

fhs2_omni1_ex02[BMI2<=24&
                  grip_max <= 29&
                  SEX==1, grip_fried := 1]

fhs2_omni1_ex02[BMI2>24&
                  BMI2<=26&
                  grip_max <= 30&
                  SEX==1, grip_fried := 1]

fhs2_omni1_ex02[BMI2>26&
                  BMI2<=28&
                  grip_max <= 30&
                  SEX==1, grip_fried := 1]

fhs2_omni1_ex02[BMI2>28&
                  grip_max <= 32&
                  SEX==1, grip_fried := 1]

##### Women

fhs2_omni1_ex02[BMI2<=23&
                  grip_max <= 17&
                  SEX==2, grip_fried := 1]

fhs2_omni1_ex02[BMI2>23&
                  BMI2<=26&
                  grip_max <= 17.3&
                  SEX==2, grip_fried := 1]

fhs2_omni1_ex02[BMI2>26&
                  BMI2<=29&
                  grip_max <= 18&
                  SEX==2, grip_fried := 1]

fhs2_omni1_ex02[BMI2>29&
                  grip_max <= 21&
                  SEX==2, grip_fried := 1]


fhs2_omni1_ex02[G606 %in% c(0,1)|G593 %in% c(0,1), exhaustion_fried := 0]
fhs2_omni1_ex02[G606 %in% c(2,3)|G593 %in% c(2,3), exhaustion_fried := 1]


#### Weight loss - Fried

fhs2_omni1_ex02[,wt_loss_pct := round((WGT1 - WGT2)/WGT2*100,1)]
fhs2_omni1_ex02[wt_loss_pct >= 5, wtloss_fried := 1]
fhs2_omni1_ex02[wt_loss_pct < 5] <- 0


#### Physical activity - Fried

fhs2_omni1_ex02[,fhs_pai := 
  calc_framingham_pai(
    as.data.frame(fhs2_omni1_ex02),
    slp_hrs = "G689",
    sed_hrs = "G690",
    slgt_hrs = "G691",
    mod_hrs = "G692",
    hvy_hrs = "G693")]

fhs2_omni1_ex02[SEX==1&
                  fhs_pai < 30.3, 
                activity_fried := 1]

fhs2_omni1_ex02[SEX==1&
                  fhs_pai >= 30.3, 
                activity_fried := 0]


fhs2_omni1_ex02[SEX==2&
                  fhs_pai < 30.1, 
                activity_fried := 1]

fhs2_omni1_ex02[SEX==2&
                  fhs_pai >= 30.1, 
                activity_fried := 0]


#### Calculate Fried score

fhs2_omni1_ex02[, total_fried :=
  gaitspeed_fried+
  grip_fried+
  exhaustion_fried+
  wtloss_fried+
  activity_fried]


######### FRAIL Score

#### Fatigue

fhs2_omni1_ex02[G606 %in% c(0,1)|G593 %in% c(0,1), fatigue_frail := 0]
fhs2_omni1_ex02[G606 %in% c(2,3)|G593 %in% c(2,3), fatigue_frail := 1]


#### Resistance

fhs2_omni1_ex02[G525==0, resistance_frail := 0]
fhs2_omni1_ex02[G525 %in% c(1,2,3,4), resistance_frail := 1]


#### Ambulation

fhs2_omni1_ex02[G548==1, ambulate_frail := 0]
fhs2_omni1_ex02[G548==0, ambulate_frail := 1]


# FRAIL Illness score counts presence of HTN, DM, cancer, chronic lung disease, MI, CHD, angina, asthma, arthritis, stroke, and ckd
# Scores for 0-4 conditions, score = 0, for ≥ 5, score=1

# HTN

fhs2_omni1_ex02[HRX2==0, htn_frail := 0]
fhs2_omni1_ex02[HRX2==1, htn_frail := 1]

# COPD (chronic lung disease)

fhs2_omni1_ex02[G415 %in% c(0,2), copd_frail := 0]
fhs2_omni1_ex02[G415==1, copd_frail := 1]

# Asthma

fhs2_omni1_ex02[G418 %in% c(0,2), asthma_frail := 0]
fhs2_omni1_ex02[G418==1, asthma_frail := 1]

# DJD (arthritis)

fhs2_omni1_ex02[G421 %in% c(0,2), djd_frail := 0]
fhs2_omni1_ex02[G421==1, djd_frail := 1]


# CKD (renal disease)


fhs2_omni1_ex02[,crcl := 
  calc_MDRD4(
    dat=as.data.frame(fhs2_omni1_ex02),
    age="AGE2",
    cr="CREAT2",
    race="RACE",
    black="B")]

fhs2_omni1_ex02[crcl >= 60, ckd_frail := 0]
fhs2_omni1_ex02[crcl < 60, ckd_frail := 1]


# CHF/MI

fhs2_omni1_ex02 <- 
  fhs2_survcvd[fhs2_omni1_ex02,
               on=.(PID,IDTYPE)]

fhs2_omni1_ex02[,chf_frail := 0]
fhs2_omni1_ex02[hfhosp_status==1&hfhosp_dt <= DATE2, chf_frail := 1]

fhs2_omni1_ex02[,chd_frail := 0]
fhs2_omni1_ex02[cadhosp_status==1&cadhosp_dt <= DATE2, chd_frail := 1]


# Stroke

fhs2_omni1_ex02[,stroke_frail := 0]
fhs2_omni1_ex02[cvahosp_status==1&cvahosp_dt <= DATE2, stroke_frail := 1]


# Cancer

fhs2_omni1_ex02[,cancer_frail := 0] 
fhs2_omni1_ex02[D_DATE <= DATE2, cancer_frail := 1]


# Sum conditions

fhs2_omni1_ex02[,conditions_frail :=
  htn_frail+
  DMRX2+
  copd_frail+
  asthma_frail+
  djd_frail+
  ckd_frail+
  chf_frail+
  chd_frail+
  stroke_frail+
  cancer_frail]

fhs2_omni1_ex02[conditions_frail <= 4, illness_frail := 0]
fhs2_omni1_ex02[conditions_frail > 4, illness_frail := 1]

#### (Weight) Loss

fhs2_omni1_ex02[wt_loss_pct >= 5, wtloss_frail := 1]
fhs2_omni1_ex02[wt_loss_pct < 5, wtloss_frail := 0]

#### Calculate FRAIL score

fhs2_omni1_ex02[,total_frail := 
  fatigue_frail+
  resistance_frail+
  ambulate_frail+
  illness_frail+
  wtloss_frail]


fhs2_omni1_ex02[,visit := 2]

fhs2_melt_omni1_ex02 <- 
  melt(
    fhs2_omni1_ex02,
    id.vars=c("PID","IDTYPE","visit"), 
    na.rm=T, 
    factorsAsStrings=T)

fhs2_melt_omni1_ex02[,form := "fhs2_omni1_ex02"]

rm(fhs2_omni1_ex02)


#===================================#
####     Lab panel, Exam 2       ---#
#===================================#

fhs2_l_fhslab_ex02 <- fread('~/Dropbox/BioLINCC files/Framingham offspring/datasets/CSV/l_fhslab_ex02_7_0561d.csv',na.strings=c("NA","","NULL"))
fhs2_l_fhslab_ex02[,visit := 2]
fhs2_melt_l_fhslab_ex02 <- melt(fhs2_l_fhslab_ex02,id.vars=c("PID","IDTYPE","visit"), na.rm=T, factorsAsStrings=T)
fhs2_melt_l_fhslab_ex02[,form := "fhs2_fhslab_ex02_7"]

rm(fhs2_l_fhslab_ex02)


#===================================#
####     Food freq, Exam 2       ---#
#===================================#

fhs2_vr_ffreq_omni1_ex02 <- fread('~/Dropbox/BioLINCC files/Framingham offspring/datasets/CSV/vr_ffreq_ex02_7_1301d.csv',na.strings=c("NA","","NULL"))
setnames(fhs2_vr_ffreq_omni1_ex02,'IDType','IDTYPE')
fhs2_vr_ffreq_omni1_ex02[,visit := 2]
fhs2_melt_vr_ffreq_omni1_ex02 <- melt(fhs2_vr_ffreq_omni1_ex02,id.vars=c("PID","IDTYPE","visit"), na.rm=T, factorsAsStrings=T)
fhs2_melt_vr_ffreq_omni1_ex02[,form := "fhs2_vr_ffreq"]

rm(fhs2_vr_ffreq_omni1_ex02)


#===============================#
#### *** OMNI 1, exam 3 ***  ####
#===============================#

fhs2_omni1_ex03 <- fread('~/Dropbox/BioLINCC files/Framingham offspring/datasets/CSV/e_exam_ex03_7_0426d.csv',na.strings=c("NA","","NULL")) # Omni 1, Exam 3
names(fhs2_omni1_ex03) <- toupper(names(fhs2_omni1_ex03))
fhs2_omni1_ex03[,visit := 3]

fhs2_omni1_ex03 <- 
    fhs2_offspring_wkthru[,c("PID",
                             "DMRX3",
                             "HRX3",
                             "DATE3",
                             "AGE3",
                             "WGT3",
                             "WGT2",
                             "SEX",
                             "CREAT3",
                             "BMI3")][fhs2_omni1_ex03,
    on=.(PID)]


fhs2_omni1_ex03[,drinks_wk := 
  H072+
  H075+
  H078]

fhs2_omni1_ex03[,epworth_ss_nas := 
  is.na(H734)+
  is.na(H735)+
  is.na(H736)+
  is.na(H737)+
  is.na(H738)+
  is.na(H739)+
  is.na(H740)+
  is.na(H741)]

fhs2_omni1_ex03[!between(H033,22,83),H033 := NA]
fhs2_omni1_ex03[!between(H037,18,83), H037 := NA]
fhs2_omni1_ex03[!between(H046,21,77), H046 := NA]

fhs2_omni1_ex03[H047==88,H047 := NA]
fhs2_omni1_ex03[H048==88,H048 := NA]

fhs2_omni1_ex03[between(H047,1,42)&!is.na(H047), hrt_years_total := H047]

fhs2_omni1_ex03[between(H048,1,10)&!is.na(H048), hrt_years_total := 
   hrt_years_total + H048/12]


fhs2_omni1_ex03[,epworth_ss := 
  H734+
  H735+
  H736+
  H737+
  H738+
  H739+
  H740+
  H741]


########### FRIED

## Walking speed

# Uses greater of 2 speeds (PMID 26695510)


## Walking speed

fhs2_omni1_ex03[,walktime_1 := H617]
fhs2_omni1_ex03[,walktime_2 := H620]

fhs2_omni1_ex03[,walktime_avg := 
  round(apply(fhs2_omni1_ex03[,.(walktime_1,walktime_2)],
                                            MARGIN=1,
                                            mean,
                                            na.rm=T),3)]

fhs2_omni1_ex03[walktime_avg=="NaN", walktime_avg := NA]

fhs2_omni1_ex03[,walktime_min := 
                  round(apply(fhs2_omni1_ex03[,.(walktime_1,walktime_2)],
                              MARGIN=1,min),3)]

fhs2_omni1_ex03[walktime_min=="-Inf", walktime_min := NA]

fhs2_omni1_ex03[!is.na(walktime_1)|
                  !is.na(walktime_2), 
                walktime_min := 0] 


########### Grip (kg)

# Produces highest average by hand (assumes dominant hand is stronger)

fhs2_omni1_ex03[,grip_avg_right := round(apply(fhs2_omni1_ex03[,.(H605,H606,H607)],MARGIN=1,mean),1)]
fhs2_omni1_ex03[,grip_avg_left := round(apply(fhs2_omni1_ex03[,.(H608,H609,H610)],MARGIN=1,mean),1)]

fhs2_omni1_ex03[,grip_max := round(apply(fhs2_omni1_ex03[,.(grip_avg_right,grip_avg_left)],MARGIN=1,max),1)]


#### Exhaustion

fhs2_omni1_ex03[H471==3|H472==3, exhaustion_fried := 1]
fhs2_omni1_ex03[H471<3&H472<3, exhaustion_fried := 0]


#### Weight loss

fhs2_omni1_ex03[H401==1, wtloss_fried := 1]
fhs2_omni1_ex03[H401 %in% c(0,2), wtloss_fried := 1]


#### Physical activity

fhs2_omni1_ex03[,fhs_pai := 
  calc_framingham_pai(
    as.data.frame(fhs2_omni1_ex03),
    slp_hrs="H480",
    sed_hrs="H481",
    slgt_hrs="H482",
    mod_hrs="H483",
    hvy_hrs="H484")]

fhs2_omni1_ex03[SEX==1&
                  fhs_pai < 30.3, 
                physical_activity_fried := 1]

fhs2_omni1_ex03[SEX==1&
                  fhs_pai >= 30.3, 
                physical_activity_fried := 0]


fhs2_omni1_ex03[SEX==2&
                  fhs_pai < 30.1, 
                physical_activity_fried := 1]

fhs2_omni1_ex03[SEX==2&
                  fhs_pai >= 30.1, 
                physical_activity_fried := 0]


######### FRAIL Score

#### Fatigue

fhs2_omni1_ex03[H723 <=3, fatigue_frail := 0]
fhs2_omni1_ex03[H723 > 3, fatigue_frail := 1]


#### Resistance

fhs2_omni1_ex03[H470==1, resistance_frail := 0]
fhs2_omni1_ex03[H470==0, resistance_frail := 1]

#### Ambulation

fhs2_omni1_ex03[H469==1, ambulate := 0]
fhs2_omni1_ex03[H469==0, ambulate := 1]


# FRAIL Illness score counts presence of HTN, DM, cancer, chronic lung disease, MI, CHD, angina, asthma, arthritis, stroke, and ckd
# Scores for 0-4 conditions, score = 0, for ≥ 5, score=1


# HTN

fhs2_omni1_ex03[H014 %in% c(0,2), htn_frail := 0]
fhs2_omni1_ex03[H014==1] <- 1

# COPD (chronic lung disease)

fhs2_omni1_ex03[H357 %in% c(0,2), copd_frail := 0]
fhs2_omni1_ex03[H357==1, copd_frail := 1]

# asthma

fhs2_omni1_ex03[H359 %in% c(0,2), asthma_frail := 0]
fhs2_omni1_ex03[H359==1] <- 1

# DJD (arthritis)

fhs2_omni1_ex03[H362 %in% c(0,2), djd_frail := 0]
fhs2_omni1_ex03[H362==1, djd_frail := 1]

# CKD (renal disease)

fhs2_omni1_ex03[,RACE := 2]

fhs2_omni1_ex03[,crcl := 
  calc_MDRD4(
    dat=as.data.frame(fhs2_omni1_ex03),
    cr="CREAT3",
    age="AGE3")]

fhs2_omni1_ex03[H354 %in% c(0,2), ckd_frail := 0]
fhs2_omni1_ex03[H354==1|crcl < 60, ckd_frail := 1]

# CHF/MI

fhs2_omni1_ex03 <- fhs_survcvd[fhs2_omni1_ex03,on=.(PID,IDTYPE)]

fhs2_omni1_ex03[,chf_frail := 0]
fhs2_omni1_ex03[hfhosp_status==1&hfhosp_dt <= DATE3, chf_frail := 1]

fhs2_omni1_ex03[,chd_frail := 0]
fhs2_omni1_ex03[cadhosp_status==1&cadhosp_dt <= DATE3, chd_frail := 1]

# Stroke

fhs2_omni1_ex03 <- fhs_survstk[fhs2_omni1_ex03,on=.(PID,IDTYPE)]

fhs2_omni1_ex03[,stroke_frail := 0]
fhs2_omni1_ex03[cvahosp_status==1&cvahosp_dt <= DATE3, stroke_frail := 1]


# Cancer

fhs2_omni1_ex03 <- 
  fhs2_survca_first[,c("PID","IDTYPE","D_DATE")][fhs2_omni1_ex03,
                                                 on=.(PID,IDTYPE)]

fhs2_omni1_ex03[,cancer_frail := 0]
fhs2_omni1_ex03[D_DATE <= DATE3, cancer_frail := 1]

fhs2_omni1_ex03[,conditions_frail := 
  htn_frail+
  DMRX3+
  copd_frail+
  asthma_frail+
  djd_frail+
  ckd_frail+
  chf_frail+
  chd_frail+
  stroke_frail+
  cancer_frail]

fhs2_omni1_ex03[conditions_frail <= 4, illness_frail := 0]
fhs2_omni1_ex03[conditions_frail > 4, illness_frail := 1]

#### (Weight) Loss

fhs2_omni1_ex03[H401==1, wtloss_frail := 1]
fhs2_omni1_ex03[H401 %in% c(0,2), illness_frail := 0]

fhs2_omni1_ex03[,wgt_delta := WGT3 - WGT2]


fhs2_melt_omni1_ex03 <- melt(fhs2_omni1_ex03,id.vars=c("PID","IDTYPE","visit"), na.rm=T, factorsAsStrings=T)
fhs2_melt_omni1_ex03[,form := "fhs2_omni1_ex03"]

rm(fhs2_omni1_ex03)

#===================================#
####     Lab Panel, Exam 3       ---#
#===================================#

fhs2_l_fhslab_ex03 <- fread('~/Dropbox/BioLINCC files/Framingham offspring/datasets/CSV/l_fhslab_ex03_7_0267d.csv',na.strings=c("NA","","NULL"))
fhs2_l_fhslab_ex03[,visit := 3]
fhs2_melt_l_fhslab_ex03 <- melt(fhs2_l_fhslab_ex03,id.vars=c("PID","IDTYPE","visit"), na.rm=T, factorsAsStrings=T)
fhs2_melt_l_fhslab_ex03[,form := "fhs2_fhslab_ex03_7"]

rm(fhs2_l_fhslab_ex03)

#===================================#
####     Food freq, Exam 3       ---#
#===================================#

fhs2_vr_ffreq_omni1_ex03 <- fread('~/Dropbox/BioLINCC files/Framingham offspring/datasets/CSV/vr_ffreq_ex03_7_0973d.csv',na.strings=c("NA","","NULL"))
setnames(fhs2_vr_ffreq_omni1_ex03, "idtype","IDTYPE")
fhs2_vr_ffreq_omni1_ex03[,visit := 3]
fhs2_melt_vr_ffreq_omni1_ex03 <- melt(fhs2_vr_ffreq_omni1_ex03,id.vars=c("PID","IDTYPE","visit"), na.rm=T, factorsAsStrings=T)
fhs2_melt_vr_ffreq_omni1_ex03[,form := "fhs2_vr_ffreq"]

rm(fhs2_vr_ffreq_omni1_ex03)


# ===============================================##
####  CRP, Offspring Exam 8, Omni 1 Exam 3     ---#
# ===============================================##

fhs2_inflamm_ex08 <- fread('~/Dropbox/BioLINCC files/Framingham offspring/datasets/CSV/l_inflamm_ex08_1b_1106d.csv',na.strings=c("NA","","NULL"))

names(fhs2_inflamm_ex08) <- toupper(names(fhs2_inflamm_ex08))

fhs2_inflamm_ex08[IDTYPE==1, visit := 8]
fhs2_inflamm_ex08[IDTYPE==7, visit := 3]

fhs2_melt_inflamm_ex08 <- 
  melt(fhs2_inflamm_ex08,
       id.vars=c("PID","IDTYPE","visit"), 
       na.rm=T, 
       factorsAsStrings=T)

fhs2_melt_inflamm_ex08[,form := "fhs2_inflamm_ex08"]

rm(fhs2_inflamm_ex08)


#==============================================#
####  CES-D Offspring exam 8, OMNI1 exam 3  ---#
#==============================================#


fhs2_q_cesd_ex09 <- fread('~/Dropbox/BioLINCC files/Framingham offspring/datasets/CSV/q_cesd_2009_m_0570sd.csv',na.strings=c("NA","","NULL"))
fhs2_q_cesd_ex09[IDTYPE==1, visit := 8]
fhs2_q_cesd_ex09[IDTYPE==7, visit := 3]
fhs2_q_cesd_ex09[,CESD20_missing := rowSums(is.na(fhs2_q_cesd_ex09[,2:21]))]
fhs2_q_cesd_ex09[,CESD20 := rowSums(fhs2_q_cesd_ex09[,2:21])]
fhs2_q_cesd_ex09[CESD20_missing >= 4, CESD20 := NA]

fhs2_q_cesd_ex09 <- fhs2_q_cesd_ex09[
  !is.na(CESD20),                        # WHERE CESD20 is not null
  .SD[which.min(testdate)],             # row with minimum testdate
  by = .(PID, IDTYPE, visit)            # GROUP BY PID, IDTYPE, VISIT
]


fhs2_melt_q_cesd_ex09 <- 
  melt(fhs2_q_cesd_ex09,
       id.vars=c("PID","IDTYPE","visit"), 
       na.rm=T, 
       factorsAsStrings=T)

fhs2_melt_q_cesd_ex09[,form :="fhs2_q_cesd"]

rm(fhs2_q_cesd_ex09)


#========================================================#
####  Echo Doppler - Offspring Exam 8, OMNI 1 Exam 3  ---#
#========================================================#

fhs2_t_doppvasc_2008 <- fread('~/Dropbox/BioLINCC files/Framingham offspring/datasets/CSV/t_doppvasc_2008_m_0756d.csv',na.strings=c("NA","","NULL"))
setnames(fhs2_t_doppvasc_2008, "idtype","IDTYPE")
fhs2_t_doppvasc_2008[IDTYPE==1, visit := 8]
fhs2_t_doppvasc_2008[IDTYPE==7, visit := 3]

fhs2_t_doppvasc_2008[,earatio := MEMAX/MAMAX]
fhs2_t_doppvasc_2008[,e_eprime_lateral := MEMAX/DEMAX]

fhs2_melt_t_doppvasc_2008 <- melt(fhs2_t_doppvasc_2008,id.vars=c("PID","IDTYPE","visit"), na.rm=T, factorsAsStrings=T)
fhs2_melt_t_doppvasc_2008[,form := "fhs2_doppvasc_2008"]

rm(fhs2_t_doppvasc_2008)


#================================================#
####  Echo - Offspring Exam 8, OMNI 1 Exam 3  ---#
#=========s=======================================#

fhs2_t_echo_2008 <- fread('~/Dropbox/BioLINCC files/Framingham offspring 2022a/datasets/CSV/t_echo_2008_m_0549d_v1.csv',na.strings=c("NA","","NULL"))
names(fhs2_t_echo_2008) <- toupper(names(fhs2_t_echo_2008))
setnames(fhs2_t_echo_2008, "EXAM","visit")

fhs2_t_echo_2008[,visit := as.character(visit)]

fhs2_t_echo_2008 <- 
  fhs2_wkthru[,.(PID,IDTYPE,SEX,visit,WGT,HGT)][fhs2_t_echo_2008,
                          on=.(PID,IDTYPE,visit)]

fhs2_t_echo_2008[,HEIGHT := as.numeric(HGT)*2.54]
fhs2_t_echo_2008[,WEIGHT := as.numeric(WGT)/2.2]

fhs2_t_echo_2008 <- 
  as.data.table(
    calc_hypertrophy_type(
      as.data.frame(fhs2_t_echo_2008), 
      id = "PID",
      sex="SEX",
      lvedd="X76",
      ivsd="X62",
      lvpwtd="X69",
      height="HEIGHT",
      weight="WEIGHT"))

fhs2_melt_t_echo_2008 <- melt(fhs2_t_echo_2008,id.vars=c("PID","IDTYPE","visit"), na.rm=T, factorsAsStrings=T)

fhs2_melt_t_echo_2008[,form := "fhs2_echo_2008"]

rm(fhs2_t_echo_2008)


#==========================================#
####     Echo cardiac strain, Omni 3    ---#
#==========================================#

fhs2_t_echocs_ex03 <- 
  fread('~/Dropbox/BioLINCC files/Framingham offspring/datasets/CSV/t_echocs_ex03_7_0901d.csv',
        na.strings=c("NA","","NULL"))
setnames(fhs2_t_echocs_ex03,"idtype","IDTYPE")
fhs2_t_echocs_ex03[,visit := 3]

names(fhs2_t_echocs_ex03) <-
  sub("^_","",names(fhs2_t_echocs_ex03))

fhs2_melt_t_echocs_ex03 <- 
  melt(fhs2_t_echocs_ex03,id.vars=c("PID","IDTYPE","visit"), 
                                na.rm=T, factorsAsStrings=T)

fhs2_melt_t_echocs_ex03[,form := "fhs2_omni1_echocs_ex03"]

rm(fhs2_t_echocs_ex03)


#=================================================#
####  *** Offspring exam 9, Omni 1 exam 4 ***  ####
#=================================================#

fhs2_offspring_ex09_omni1_ex04 <- 
  fread('~/Dropbox/BioLINCC files/Framingham offspring/datasets/CSV/e_exam_ex09_1b_0844d.csv',
        na.strings=c("NA","","NULL")) # Omni1, Exam 9

names(fhs2_offspring_ex09_omni1_ex04) <- 
  toupper(names(fhs2_offspring_ex09_omni1_ex04))

fhs2_offspring_ex09_omni1_ex04 <-
  fhs2_wkthru[,.(PID,IDTYPE,DATE)][fhs2_offspring_ex09_omni1_ex04, on=.(PID,IDTYPE)]

fhs2_offspring_ex09_omni1_ex04[,DATE := as.integer(DATE)] 

fhs2_offspring_ex09_omni1_ex04[,CESD20_missing := 
  rowSums(is.na(fhs2_offspring_ex09_omni1_ex04[,c(paste0("J",
                                                         c(724,
                                                         727:746)))]))]

fhs2_offspring_ex09_omni1_ex04[,CESD20 := 
  rowSums(fhs2_offspring_ex09_omni1_ex04[,c(paste0("J",c(724,
                                                         727:746)))])]

fhs2_offspring_ex09_omni1_ex04[CESD20_missing >= 4, CESD20 := NA]

fhs2_offspring_ex09_omni1_ex04[IDTYPE==1, visit := 9]
fhs2_offspring_ex09_omni1_ex04[IDTYPE==7, visit := 4]

fhs2_offspring_ex09_omni1_ex04[,drinks_wk := 
                                 J075+
                                 J077+
                                 J079]
                               
fhs2_offspring_ex09_omni1_ex04[!between(J055,39,79), J055 := NA]


fhs2_offspring_ex09_omni1_ex04 <- 
  fhs2_wkthru[visit==1,
                    c("PID",
                      "IDTYPE",
                      "SEX",
                      "HGT")][fhs2_offspring_ex09_omni1_ex04,
        on=.(PID,IDTYPE)]

fhs2_omni1_ex04_wgt3 <- 
  fhs2_wkthru[visit==3&
                IDTYPE==7,
              c("PID",
                "IDTYPE",
                "WGT")]

fhs2_offspring_ex09_wgt8 <- 
  fhs2_wkthru[visit==8&
                IDTYPE==1,
              c("PID",
                "IDTYPE",
                "WGT")]

fhs2_offspring_ex09_omni1_ex04_lastwgt <- 
  rbindlist(list(fhs2_omni1_ex04_wgt3,
        fhs2_offspring_ex09_wgt8))

names(fhs2_offspring_ex09_omni1_ex04_lastwgt)[3] <- "last_wgt"

fhs2_omni1_ex04_wgt4 <- 
  fhs2_wkthru[visit==4&
                IDTYPE==7,
              c("PID",
                "IDTYPE",
                "WGT")]

fhs2_offspring_ex09_wgt9 <- 
  fhs2_wkthru[visit==9&
                IDTYPE==1,
              c("PID",
                "IDTYPE",
                "WGT")]


fhs2_offspring_ex09_omni1_ex04_wgt <- 
  rbind(fhs2_omni1_ex04_wgt4,
        fhs2_offspring_ex09_wgt9)

fhs2_offspring_ex09_omni1_ex04 <- 
  fhs2_offspring_ex09_omni1_ex04_wgt[
    fhs2_offspring_ex09_omni1_ex04,
        on=.(PID,IDTYPE)]

fhs2_offspring_ex09_omni1_ex04 <- 
  fhs2_offspring_ex09_omni1_ex04_lastwgt[
    fhs2_offspring_ex09_omni1_ex04,
    on=.(PID,IDTYPE)]


fhs2_offspring_ex09_bmi <- 
  fhs2_wkthru[visit==9&
                IDTYPE==1&
                !is.na(BMI),
              c("PID",
                "IDTYPE",
                "BMI",
                "CREAT",
                "DMRX",
                "HRX",
                "AGE")]

fhs2_omni1_ex04_bmi <- 
  fhs2_wkthru[visit==4&
                IDTYPE==7&
                !is.na(BMI),
              c("PID",
                "IDTYPE",
                "BMI",
                "CREAT",
                "DMRX",
                "HRX",
                "AGE")]

fhs2_offspring9_omni4_bmi <- 
  rbind(fhs2_offspring_ex09_bmi,
        fhs2_omni1_ex04_bmi)

fhs2_offspring_ex09_omni1_ex04 <- 
  fhs2_offspring9_omni4_bmi[
    fhs2_offspring_ex09_omni1_ex04,
        on=.(PID,IDTYPE)]


fhs2_offspring_ex09_omni1_ex04 <- 
  fhs_outcomes[fhs2_offspring_ex09_omni1_ex04,
        on=.(PID,
             IDTYPE)]


fhs2_offspring_ex09_omni1_ex04 <- 
  fhs2_survcvd[fhs2_offspring_ex09_omni1_ex04,
        on=.(PID,
             IDTYPE)]


fhs2_offspring_ex09_omni1_ex04 <- 
  fhs2_survstk[fhs2_offspring_ex09_omni1_ex04,
               on=.(PID,
                    IDTYPE)]


fhs2_offspring_ex09_omni1_ex04 <- 
  fhs_survca_first_any[,c("PID",
                          "IDTYPE",
                          "D_DATE")][fhs2_offspring_ex09_omni1_ex04,
                                     on=.(PID,
                                          IDTYPE)]


## Walking speed - Fried

# Uses greater of 2 speeds (PMID 26695510)

fhs2_offspring_ex09_omni1_ex04[,walktime_1 := J785]
fhs2_offspring_ex09_omni1_ex04[,walktime_2 := J790]

fhs2_offspring_ex09_omni1_ex04[,walktime_avg := 
  round(
    apply(
      fhs2_offspring_ex09_omni1_ex04[,.(walktime_1,walktime_2)],
                                                           MARGIN=1,
                                                           mean,
                                                           na.rm=T),3)]

fhs2_offspring_ex09_omni1_ex04[walktime_avg=="NaN", walktime_avg := NA]

fhs2_offspring_ex09_omni1_ex04[,walktime_min := 
                                 round(
                                   apply(
                                     fhs2_offspring_ex09_omni1_ex04[,.(walktime_1,walktime_2)],
                                                           MARGIN=1,min),3)]

fhs2_offspring_ex09_omni1_ex04[walktime_min=="-Inf", walktime_min := NA]

fhs2_offspring_ex09_omni1_ex04[!is.na(walktime_1)|
                                 !is.na(walktime_2), walktime_min := 0] 


########### Grip (kg)

# Produces highest average by hand (assumes dominant hand is stronger)

fhs2_offspring_ex09_omni1_ex04[,grip_avg_right := 
                                 round(apply(fhs2_offspring_ex09_omni1_ex04[,.(J765,J766,J767)],MARGIN=1,mean),1)]

fhs2_offspring_ex09_omni1_ex04[,grip_avg_left := round(apply(fhs2_offspring_ex09_omni1_ex04[,.(J768,J769,J770)],MARGIN=1,mean),1)]

fhs2_offspring_ex09_omni1_ex04[,grip_max := round(apply(fhs2_offspring_ex09_omni1_ex04[,c("grip_avg_right","grip_avg_left")],MARGIN=1,max),1)]


#### Physical activity index - Fried

fhs2_offspring_ex09_omni1_ex04[,fhs_pai := 
  calc_framingham_pai(
    as.data.frame(fhs2_offspring_ex09_omni1_ex04),
    slp_hrs="J628",
    sed_hrs="J629",
    slgt_hrs="J630",
    mod_hrs="J631",
    hvy_hrs="J632")]

fhs2_offspring_ex09_omni1_ex04[SEX==1&
                                 fhs_pai < 30.3, 
                               physical_activity_fried := 1]

fhs2_offspring_ex09_omni1_ex04[SEX==1&
                                 fhs_pai >= 30.3,
                               physical_activity_fried := 0]


fhs2_offspring_ex09_omni1_ex04[SEX==2&
                                 fhs_pai < 30.1, 
                               physical_activity_fried := 1]

fhs2_offspring_ex09_omni1_ex04[SEX==2&
                                 fhs_pai >= 30.1, 
                               physical_activity_fried := 0]


#### Exhaustion - Fried

fhs2_offspring_ex09_omni1_ex04[J733==3|
                                 J746==3, 
                               exhaustion_fried := 1]

fhs2_offspring_ex09_omni1_ex04[J733<3&
                                 J746<3, 
                               exhaustion_fried := 0]

#### Weight loss - Fried

fhs2_offspring_ex09_omni1_ex04[,wgt_delta :=
  as.numeric(WGT) - 
  as.numeric(last_wgt)]

fhs2_offspring_ex09_omni1_ex04[wgt_delta <= -10, wtloss_fried := 1]
fhs2_offspring_ex09_omni1_ex04[wgt_delta > -10, wtloss := 0]


######### FRAIL Score

#### Fatigue

fhs2_offspring_ex09_omni1_ex04[J900 <=3, fatigue_frail := 0]
fhs2_offspring_ex09_omni1_ex04[J900 > 3, fatigue_frail := 0]


#### Resistance

fhs2_offspring_ex09_omni1_ex04[J611==1, resistance_frail := 0]
fhs2_offspring_ex09_omni1_ex04[J611==0, resistance_frail := 1]

#### Ambulation

fhs2_offspring_ex09_omni1_ex04[J610==0, ambulate_frail := 1]
fhs2_offspring_ex09_omni1_ex04[J610==1, ambulate_frail := 0]


# FRAIL Illness score counts presence of HTN, DM, cancer, chronic lung disease, MI, CHD, angina, asthma, arthritis, stroke, and ckd
# Scores for 0-4 conditions, score = 0, for ≥ 5, score=1


# COPD (chronic lung disease)

fhs2_offspring_ex09_omni1_ex04[J419 %in% c(0,2), copd_frail := 0]
fhs2_offspring_ex09_omni1_ex04[J419==1, copd_frail := 1]

# asthma

fhs2_offspring_ex09_omni1_ex04[J421 %in% c(0,2), asthma_frail := 0]
fhs2_offspring_ex09_omni1_ex04[J421==1, ashtma_frail := 1]

# DJD (arthritis)

fhs2_offspring_ex09_omni1_ex04[J426 %in% c(0,2), djd_frail := 0]
fhs2_offspring_ex09_omni1_ex04[J426==1, djd_frail := 1]

# CKD (renal disease)

fhs2_offspring_ex09_omni1_ex04[IDTYPE==1, RACE := 1]
fhs2_offspring_ex09_omni1_ex04[IDTYPE==7, RACE := 2]

crcl <- 
  calc_MDRD4(
    dat=as.data.frame(fhs2_offspring_ex09_omni1_ex04),
    cr="CREAT")

fhs2_offspring_ex09_omni1_ex04[J414 %in% c(0,2), ckd_frail := 0]
fhs2_offspring_ex09_omni1_ex04[J414==1, ckd_frail := 1]

# CHF/MI


fhs2_offspring_ex09_omni1_ex04[,chf_frail := 0]
fhs2_offspring_ex09_omni1_ex04[hfhosp_status==1&
                                           hfhosp_dt <= DATE, chf_frail := 1]

fhs2_offspring_ex09_omni1_ex04[,chd_frail := 0]
fhs2_offspring_ex09_omni1_ex04[cadhosp_status==1&
                                           cadhosp_dt <= DATE, chd_frail := 1]

# Stroke

fhs2_offspring_ex09_omni1_ex04[,stroke_frail := 0]
fhs2_offspring_ex09_omni1_ex04[STROKE==1&STROKEDATE <= DATE, stroke_frail := 1]

# Cancer

fhs2_offspring_ex09_omni1_ex04[,cancer_frail := 0]
fhs2_offspring_ex09_omni1_ex04[D_DATE <= DATE, cancer_frail := 1]


#### Calculate FRAIL illness score
fhs2_offspring_ex09_omni1_ex04[,conditions_frail := 
  as.numeric(HRX)+
  as.numeric(DMRX)+
  copd_frail+
  asthma_frail+
  djd_frail+
  ckd_frail+
  chf_frail+
  chd_frail+
  stroke_frail+
  cancer_frail]

fhs2_offspring_ex09_omni1_ex04[conditions_frail <= 4, illness_frail := 0]
fhs2_offspring_ex09_omni1_ex04[conditions_frail > 4, illness_frail := 1]

#### (Weight) Loss

fhs2_offspring_ex09_omni1_ex04[wgt_delta <= -10, wtloss_frail := 1]
fhs2_offspring_ex09_omni1_ex04[wgt_delta > -10, wtloss_frail := 0]


######### Epworth Sleepiness scale. 

fhs2_offspring_ex09_omni1_ex04[, epworth_ss_nas := 
  is.na(J904)+
  is.na(J905)+
  is.na(J906)+
  is.na(J907)+
  is.na(J908)+
  is.na(J909)+
  is.na(J910)+
  is.na(J911)]

fhs2_offspring_ex09_omni1_ex04[,epworth_ss := 
  J904+
  J905+
  J906+
  J907+
  J908+
  J909+
  J910+
  J911]

## DENORMALIZE

fhs2_melt_offspring_ex09_omni1_ex04 <- melt(fhs2_offspring_ex09_omni1_ex04,id.vars=c("PID","IDTYPE","visit"), na.rm=T, factorsAsStrings=T)

fhs2_melt_offspring_ex09_omni1_ex04[,form := "fhs2_offspring_ex09_omni1_ex04"]

rm(fhs2_offspring_ex09_omni1_ex04)


#======================================================================#
####  Actigraphy, Framingham Offspring Exam 9 - Omni 1, Exam 4      ---#
#======================================================================#

act_fields <-
  c("zmin_vdt",
    "zmin_vdtd",
    "zmin_vdtm",
    "zmin_vdtwd",
    "zmin_vdtwdd",
    "zmin_vdtwdm",
    "zmin_vdtwe",
    "zmin_vdtwed",
    "zmin_vdtwedm",
    "wstep_vdt",
    "wstep_vdtd",
    "wstep_vdtm",
    "wstep_vdtwd",
    "wstep_vdtwdd",
    "wstep_vdtwdm",
    "wstep_vdtwe",
    "wstep_vdtwed",
    "wstep_vdtwedm",
    "zb_vdt",
    "zb_vdtd",
    "zb_vdtmin",
    "zb_vdtwd",
    "zb_vdtwdmin",
    "zb_vdtwe",
    "zb_vdtwed",
    "zb_vdtwedd",
    "zb_vdtwemin",
    "vigstepvdt",
    "vigstepvdtd",
    "vigstepvdtwd",
    "vigstepvdtwdd",
    "vigstepvdtwe",
    "vigstepvdtwed",
    "vigminvdt",
    "vigminvdtd",
    "vigminvdtwd",
    "vigminvdtwdd",
    "vigminvdtwe",
    "vigminvdtwed",
    "vigcntvdt",
    "vigcntvdtd",
    "vigcntvdtwd",
    "vigcntvdtwdd",
    "vigcntvdtwe",
    "vigcntvdtwed",
    "sedcntvdt",
    "sedcntvdtd",
    "sedcntvdtwd",
    "sedcntvdtwdd",
    "sedcntvdtwe",
    "sedcntvdtwed",
    "sedminvdt",
    "sedminvdtd",
    "sedminvdtwd",
    "sedminvdtwdd",
    "sedminvdtwe",
    "sedminvdtwed",
    "sedminwed",
    "bgt_litcntvdt",
    "bgt_litcntvdtd",
    "bgt_litcntvdtwd",
    "bgt_litcntvdtwdd",
    "bgt_litcntvdtwe",
    "bgt_litcntvdtwed",
    "bgt_litminvdt",
    "bgt_litminvdtd",
    "bgt_litminvdtwd",
    "bgt_litminvdtwdd",
    "bgt_litminvdtwe",
    "bgt_litminvdtwed",
    "bgt_litstepvdt",
    "bgt_litstepvdtd",
    "bgt_litstepvdtwd",
    "bgt_litstepvdtwdd",
    "bgt_litstepvdtwe",
    "bgt_litstepvdtwed",
    "bgt_modcntvdt",
    "bgt_modcntvdtd",
    "bgt_modcntvdtwd",
    "bgt_modcntvdtwdd",
    "bgt_modcntvdtwe",
    "bgt_modcntvdtwed",
    "bgt_modminvdt",
    "bgt_modminvdtd",
    "bgt_modminvdtwd",
    "bgt_modminvdtwdd",
    "bgt_modminvdtwe",
    "bgt_modminvdtwed",
    "bgt_modstepvdt",
    "bgt_modstepvdtd",
    "bgt_modstepvdtwd",
    "bgt_modstepvdtwdd",
    "bgt_modstepvdtwe",
    "bgt_modstepvdtwed",
    "bgt_mvpcntvdt",
    "bgt_mvpcntvdtd",
    "bgt_mvpcntvdtwd",
    "bgt_mvpcntvdtwdd",
    "bgt_mvpcntvdtwe",
    "bgt_mvpcntvdtwed",
    "bgt_mvpminvdt",
    "bgt_mvpminvdtd",
    "bgt_mvpminvdtwd",
    "bgt_mvpminvdtwdd",
    "bgt_mvpminvdtwe",
    "bgt_mvpminvdtwed",
    "bgt_mvpstepvdt",
    "bgt_mvpstepvdtd",
    "bgt_mvpstepvdtwd",
    "bgt_mvpstepvdtwdd",
    "bgt_mvpstepvdtwe",
    "bgt_mvpstepvdtwed",
    "bgt_sedcntvdt",
    "bgt_sedcntvdtd",
    "bgt_sedcntvdtwd",
    "bgt_sedcntvdtwdd",
    "bgt_sedcntvdtwe",
    "bgt_sedcntvdtwed",
    "bgt_sedminvdt",
    "bgt_sedminvdtd",
    "bgt_sedminvdtwd",
    "bgt_sedminvdtwdd",
    "bgt_sedminvdtwe",
    "bgt_sedminvdtwed",
    "bgt_sedstepvdt",
    "bgt_sedstepvdtd",
    "bgt_sedstepvdtwd",
    "bgt_sedstepvdtwdd",
    "bgt_sedstepvdtwe",
    "bgt_sedstepvdtwed",
    "bgt_vigcntvdt",
    "bgt_vigcntvdtd",
    "bgt_vigcntvdtwd",
    "bgt_vigcntvdtwdd",
    "bgt_vigcntvdtwe",
    "bgt_vigcntvdtwed",
    "bgt_vigminvdt",
    "bgt_vigminvdtd",
    "bgt_vigminvdtwd",
    "bgt_vigminvdtwdd",
    "bgt_vigminvdtwe",
    "bgt_vigminvdtwed",
    "bgt_vigstepvdt",
    "bgt_vigstepvdtd",
    "bgt_vigstepvdtwd",
    "bgt_vigstepvdtwdd",
    "bgt_vigstepvdtwe",
    "bgt_vigstepvdtwed",
    "litcntvdt",
    "litcntvdtd",
    "litcntvdtwd",
    "litcntvdtwdd",
    "litcntvdtwe",
    "litcntvdtwed",
    "litminvdt",
    "litminvdtd",
    "litminvdtwd",
    "litminvdtwdd",
    "litminvdtwe",
    "litminvdtwed",
    "litstepvdt",
    "litstepvdtd",
    "litstepvdtwd",
    "litstepvdtwdd",
    "litstepvdtwe",
    "litstepvdtwed",
    "modcntvdt",
    "modcntvdtd",
    "modcntvdtwd",
    "modcntvdtwdd",
    "modcntvdtwe",
    "modcntvdtwed",
    "modminvdt",
    "modminvdtd",
    "modminvdtwd",
    "modminvdtwdd",
    "modminvdtwe",
    "modminvdtwed",
    "modstepvdt",
    "modstepvdtd",
    "modstepvdtwd",
    "modstepvdtwdd",
    "modstepvdtwe",
    "modstepvdtwed",
    "motcntvdt",
    "motcntvdtd",
    "motcntvdtwd",
    "motcntvdtwdd",
    "motcntvdtwe",
    "motcntvdtwed",
    "motminvdt",
    "motminvdtd",
    "motminvdtwd",
    "motminvdtwdd",
    "motminvdtwe",
    "motminvdtwed",
    "motstepvdt",
    "motstepvdtd",
    "motstepvdtwd",
    "motstepvdtwdd",
    "motstepvdtwe",
    "motstepvdtwed",
    "mvpcntvdt",
    "mvpcntvdtd",
    "mvpcntvdtwd",
    "mvpcntvdtwdd",
    "mvpcntvdtwe",
    "mvpcntvdtwed",
    "mvpminvdt",
    "mvpminvdtd",
    "mvpminvdtwd",
    "mvpminvdtwdd",
    "mvpminvdtwe",
    "mvpminvdtwed",
    "mvpstepvdt",
    "mvpstepvdtd",
    "mvpstepvdtwd",
    "mvpstepvdtwdd",
    "mvpstepvdtwe",
    "mvpstepvdtwed",
    "s_litcntvdt",
    "s_litcntvdtd",
    "s_litcntvdtwd",
    "s_litcntvdtwdd",
    "s_litcntvdtwe",
    "s_litcntvdtwed",
    "s_litminvdt",
    "s_litminvdtd",
    "s_litminvdtwd",
    "s_litminvdtwdd",
    "s_litminvdtwe",
    "s_litminvdtwed",
    "s_litstepvdt",
    "s_litstepvdtd",
    "s_litstepvdtwd",
    "s_litstepvdtwdd",
    "s_litstepvdtwe",
    "s_litstepvdtwed",
    "s_modcntvdt",
    "s_modcntvdtd",
    "s_modcntvdtwd",
    "s_modcntvdtwdd",
    "s_modcntvdtwe",
    "s_modcntvdtwed",
    "s_modminvdt",
    "s_modminvdtd",
    "s_modminvdtwd",
    "s_modminvdtwdd",
    "s_modminvdtwe",
    "s_modminvdtwed",
    "s_modstepvdt",
    "s_modstepvdtd",
    "s_modstepvdtwd",
    "s_modstepvdtwdd",
    "s_modstepvdtwe",
    "s_modstepvdtwed",
    "s_mvpcntvdt",
    "s_mvpcntvdtd",
    "s_mvpcntvdtwd",
    "s_mvpcntvdtwdd",
    "s_mvpcntvdtwe",
    "s_mvpcntvdtwed",
    "s_mvpminvdt",
    "s_mvpminvdtd",
    "s_mvpminvdtwd",
    "s_mvpminvdtwdd",
    "s_mvpminvdtwe",
    "s_mvpminvdtwed",
    "s_mvpstepvdt",
    "s_mvpstepvdtd",
    "s_mvpstepvdtwd",
    "s_mvpstepvdtwdd",
    "s_mvpstepvdtwe",
    "s_mvpstepvdtwed",
    "s_sedcntvdt",
    "s_sedcntvdtd",
    "s_sedcntvdtwd",
    "s_sedcntvdtwdd",
    "s_sedcntvdtwe",
    "s_sedcntvdtwed",
    "s_sedminvdt",
    "s_sedminvdtd",
    "s_sedminvdtwd",
    "s_sedminvdtwdd",
    "s_sedminvdtwe",
    "s_sedminvdtwed",
    "s_sedstepvdt",
    "s_sedstepvdtd",
    "s_sedstepvdtwd",
    "s_sedstepvdtwdd",
    "s_sedstepvdtwe",
    "s_sedstepvdtwed",
    "s_vigcntvdt",
    "s_vigcntvdtd",
    "s_vigcntvdtwd",
    "s_vigcntvdtwdd",
    "s_vigcntvdtwe",
    "s_vigcntvdtwed",
    "s_vigminvdt",
    "s_vigminvdtd",
    "s_vigminvdtwd",
    "s_vigminvdtwdd",
    "s_vigminvdtwe",
    "s_vigminvdtwed",
    "s_vigstepvdt",
    "s_vigstepvdtd",
    "s_vigstepvdtwd",
    "s_vigstepvdtwdd",
    "s_vigstepvdtwe",
    "s_vigstepvdtwed",
    "zstep_vdt",
    "zstep_vdtd",
    "zstep_vdtm",
    "zstep_vdtwd",
    "zstep_vdtwdd",
    "zstep_vdtwdm",
    "zstep_vdtwe",
    "zstep_vdtwed",
    "zstep_vdtwedm",
    "zcnt_vdt",
    "zcnt_vdtd",
    "zcnt_vdtm",
    "zcnt_vdtwd",
    "zcnt_vdtwdd",
    "zcnt_vdtwdm",
    "zcnt_vdtwe",
    "zcnt_vdtwed",
    "zcnt_vdtwedm",
    "zb_vdt",
    "zb_vdtd",
    "zb_vdtmin",
    "zb_vdtwd",
    "zb_vdtwdmin",
    "zb_vdtwe",
    "zb_vdtwed",
    "zb_vdtwedd",
    "zb_vdtwemin",
    "wb_vdt",
    "wb_vdtd",
    "wb_vdtmin",
    "wb_vdtwd",
    "wb_vdtwdmin",
    "wb_vdtwe",
    "wb_vdtwed",
    "wb_vdtwedd",
    "wb_vdtwemin",
    "wcnt_vdt",
    "wcnt_vdtd",
    "wcnt_vdtm",
    "wcnt_vdtwd",
    "wcnt_vdtwdd",
    "wcnt_vdtwdm",
    "wcnt_vdtwe",
    "wcnt_vdtwed",
    "wcnt_vdtwedm",
    "wmin_vdt",
    "wmin_vdtd",
    "wmin_vdtm",
    "wmin_vdtwd",
    "wmin_vdtwdd",
    "wmin_vdtwdm",
    "wmin_vdtwe",
    "wmin_vdtwed",
    "wmin_vdtwedm",
    "wstep_vdt",
    "wstep_vdtd",
    "wstep_vdtm",
    "wstep_vdtwd",
    "wstep_vdtwdd",
    "wstep_vdtwdm",
    "wstep_vdtwe",
    "wstep_vdtwed",
    "wstep_vdtwedm")

act_fields <- toupper(act_fields)

fhs2_actigraphy_offspring_ex09_omni1_ex04 <- 
  as.data.table(
    read_sas('~/Dropbox/BioLINCC files/Framingham Offspring/datasets/t_physactf_ex09_1b_0833d.sas7bdat'))


names(fhs2_actigraphy_offspring_ex09_omni1_ex04) <- 
  toupper(names(fhs2_actigraphy_offspring_ex09_omni1_ex04))

fhs2_melt_actigraphy_offspring_ex09_omni1_ex04 <-  
  melt(fhs2_actigraphy_offspring_ex09_omni1_ex04,id.vars=c("PID","IDTYPE"),na.rm=T,factorsAsStrings = T)

fhs2_melt_actigraphy_offspring_ex09_omni1_ex04[IDTYPE==1, visit := 9]
fhs2_melt_actigraphy_offspring_ex09_omni1_ex04[IDTYPE==7, visit := 4]

fhs2_melt_actigraphy_offspring_ex09_omni1_ex04[,form := "fhs2_actigraphy_ex09_ex04"]

fhs2_melt_actigraphy_offspring_ex09_omni1_ex04 <- 
  fhs2_melt_actigraphy_offspring_ex09_omni1_ex04[variable %in% act_fields]

rm(fhs2_actigraphy_offspring_ex09_omni1_ex04)


#=========================================================#
####  Validated food frequency data, exam 9            ---#
#=========================================================#

fhs2_vr_ffreq_offspring_ex09_omni1_ex04 <- fread('~/Dropbox/BioLINCC files/Framingham offspring/datasets/CSV/vr_ffreq_ex09_1b_1636d_v1.csv',na.strings=c("NA","","NULL"))
fhs2_vr_ffreq_offspring_ex09_omni1_ex04[IDTYPE==1, visit := 9]
fhs2_vr_ffreq_offspring_ex09_omni1_ex04[IDTYPE==7, visit := 4]

fhs2_melt_vr_ffreq_offspring_ex09_omni1_ex04 <- melt(fhs2_vr_ffreq_offspring_ex09_omni1_ex04,
                                                     id.vars=c("PID","IDTYPE","visit"),
                                                     na.rm=T, 
                                                     factorsAsStrings=T)
fhs2_melt_vr_ffreq_offspring_ex09_omni1_ex04[,form := "fhs2_vr_ffreq"]

rm(fhs2_vr_ffreq_offspring_ex09_omni1_ex04)


#==============================#
####  Lab panel, Exam 9     ---#
#==============================#

fhs2_l_fhslab_ex09 <- fread('~/Dropbox/BioLINCC files/Framingham offspring/datasets/CSV/l_fhslab_ex09_1b_0658d.csv',na.strings=c("NA","","NULL"))
fhs2_l_fhslab_ex09[IDTYPE==1, visit := 9]
fhs2_l_fhslab_ex09[IDTYPE==7, visit := 4]
fhs2_melt_l_fhslab_ex09 <- melt(fhs2_l_fhslab_ex09,id.vars=c("PID","IDTYPE","visit"), na.rm=T, factorsAsStrings=T)
fhs2_melt_l_fhslab_ex09[,form := "fhs2_fhslab_ex09"]

rm(fhs2_l_fhslab_ex09)


#================================================#
####  Menses                                  ---#
#================================================#

fhs2_menarche <- fread('~/Dropbox/BioLINCC files/Framingham offspring/datasets/CSV/menarche1_7d.csv',na.strings=c("NA","","NULL"))

names(fhs2_menarche) <- toupper(names(fhs2_menarche))

fhs2_offspring_menopause <- fread('~/Dropbox/BioLINCC files/Framingham offspring/datasets/CSV/meno1_8d.csv',
                                  na.strings=c("NA","","NULL"))


fhs2_offspring_menopause[,num_ovaries_removed :=
  apply(fhs2_offspring_menopause[,c(3:9,17)],
        MARGIN=1,
        function(x) max(x, na.rm=T))]

fhs2_offspring_menopause[,STOPAGE := STOP_AGE]

fhs2_offspring_menopause[num_ovaries_removed=="-Inf", num_ovaries_removed := NA]
fhs2_offspring_menopause[num_ovaries_removed > 0, age_oophorectomy := STOPAGE] 

fhs2_omni1_menopause <- fread('~/Dropbox/BioLINCC files/Framingham offspring/datasets/CSV/vr_meno_ex03_7_0916d.csv',
                              na.strings=c("NA","","NULL"))

fhs2_omni1_menopause[,num_ovaries_removed :=
  apply(fhs2_omni1_menopause[,11:13],
        MARGIN=1,
        function(x) max(x, na.rm=T))]

fhs2_omni1_menopause[num_ovaries_removed=="-Inf", num_ovaries_removed := NA]
fhs2_omni1_menopause[num_ovaries_removed > 0, age_oophorectomy := STOPAGE] 


fhs2_menopause <- 
  rbindlist(
    list(fhs2_offspring_menopause[,c("PID","IDTYPE","STOPAGE","CAUSE","num_ovaries_removed","age_oophorectomy")],
         fhs2_omni1_menopause[,c("PID","IDTYPE","STOPAGE","CAUSE","num_ovaries_removed","age_oophorectomy")])
  )

fhs2_menopause[STOPAGE==0, STOPAGE := NA]

fhs2_menses <- fhs2_menopause[fhs2_menarche,
                     on=.(PID,IDTYPE)]

fhs2_menses[,years_menses := STOPAGE-IVQ1AGE]

fhs2_melt_menses <- melt(fhs2_menses,id.vars=c("PID","IDTYPE"), na.rm=T, factorsAsStrings=T)

fhs2_melt_menses[,form := "fhs2_menses"]
fhs2_melt_menses[,visit := 8]

rm(fhs2_menses)

#===============================================#
#### *** Offspring exam 10/Omni 1 exam 5 *** ####
#===============================================#

fhs2_offspring_ex10_omni1_ex05 <- 
  fread('~/Dropbox/BioLINCC files/Framingham offspring/datasets/CSV/e_exam_ex10_1b_1409d.csv',
        na.strings=c("NA","","NULL")) # Omni1 5  Offspring Exam 10

fhs2_offspring_ex10_omni1_ex05 <-
        fhs_dates[IDTYPE==1,c("PID","DATE10")][fhs2_offspring_ex10_omni1_ex05,
        on=.(PID)]

fhs2_offspring_ex10_omni1_ex05 <-
        fhs_dates[IDTYPE==7,c("PID","DATE5")][fhs2_offspring_ex10_omni1_ex05,
        on=.(PID)]

fhs2_offspring_ex10_omni1_ex05[IDTYPE==7, 
                               DATE := DATE5]

fhs2_offspring_ex10_omni1_ex05[IDTYPE==1,
                               DATE := DATE10]


fhs2_offspring_ex10_omni1_ex05[,CESD20_missing := 
  rowSums(is.na(fhs2_offspring_ex10_omni1_ex05[,c(paste0("K",1197:1216))]))]

fhs2_offspring_ex10_omni1_ex05[,CESD20 := 
  rowSums(
    fhs2_offspring_ex10_omni1_ex05[,c(paste0("K",1197:1216))])]

fhs2_offspring_ex10_omni1_ex05[CESD20_missing >= 4,CESD20 := NA]

fhs2_offspring_ex10_omni1_ex05[IDTYPE==1, visit := 10]
fhs2_offspring_ex10_omni1_ex05[IDTYPE==7, visit := 5]

fhs2_offspring_ex10_omni1_ex05[,drinks_wk := 
  K0321+
  K0325+
  K0329]

fhs2_offspring_ex10_omni1_ex05[
  !between(K0295,37,80), K0295 := NA]


fhs2_offspring_ex10_omni1_ex05 <- 
        fhs2_wkthru[visit==1,
                    c("PID",
                      "IDTYPE",
                      "SEX",
                      "HGT")][fhs2_offspring_ex10_omni1_ex05,
        on=.(PID,IDTYPE)]

fhs2_omni1_ex05_wgt4 <- 
  fhs2_wkthru[visit==4&
                IDTYPE==7,
              .(PID,
                IDTYPE,
                WGT)]

fhs2_offspring_ex10_wgt9 <- 
  fhs2_wkthru[visit==9&
              IDTYPE==1,
              .(PID,
                IDTYPE,
                WGT)]

fhs2_offspring_ex10_omni1_ex05_lastwgt <- 
  rbindlist(
    list(fhs2_omni1_ex05_wgt4,
         fhs2_offspring_ex10_wgt9))

names(fhs2_offspring_ex10_omni1_ex05_lastwgt)[3] <- "last_wgt"

fhs2_offspring_ex10_omni1_ex05 <- 
  fhs2_offspring_ex10_omni1_ex05_lastwgt[
    fhs2_offspring_ex10_omni1_ex05,
        on=.(PID,IDTYPE)]

fhs2_offspring_ex10_bmi <- 
  fhs2_wkthru[visit==10&
              IDTYPE==1&
                !is.na(BMI),
              c("PID",
                "IDTYPE",
                "BMI",
                "WGT",
                "CREAT",
                "DMRX",
                "HRX",
                "AGE")]

fhs2_omni1_ex05_bmi <- 
  fhs2_wkthru[visit==5&
                IDTYPE==7&
                !is.na(BMI),
              c("PID",
                "IDTYPE",
                "BMI",
                "WGT",
                "CREAT",
                "DMRX",
                "HRX",
                "AGE")]

fhs2_off10_omni5_bmi <- 
  rbindlist(
    list(fhs2_offspring_ex10_bmi,
        fhs2_omni1_ex05_bmi))

fhs2_offspring_ex10_omni1_ex05 <- 
  fhs2_off10_omni5_bmi[
    fhs2_offspring_ex10_omni1_ex05,
        on=.(PID,IDTYPE)]


fhs2_offspring_ex10_omni1_ex05 <- 
  fhs_outcomes[fhs2_offspring_ex10_omni1_ex05,
        on=.(PID,IDTYPE)]


fhs2_offspring_ex10_omni1_ex05 <- 
        fhs_survca_first_any[,c("PID",
                                "IDTYPE",
                                "D_DATE")][fhs2_offspring_ex10_omni1_ex05,
        on=.(PID,
             IDTYPE)]


## Walking speed - Fried

# Uses greater of 2 speeds (PMID 26695510)

fhs2_offspring_ex10_omni1_ex05[,walktime_1 := K1593]
fhs2_offspring_ex10_omni1_ex05[,walktime_2 := K1595]

fhs2_offspring_ex10_omni1_ex05[,walktime_avg := 
  round(apply(fhs2_offspring_ex10_omni1_ex05[,.(walktime_1,walktime_2)],
              MARGIN=1,
              mean,
              na.rm=T),3)]

fhs2_offspring_ex10_omni1_ex05[walktime_avg=="NaN", walktime_avg := NA]

fhs2_offspring_ex10_omni1_ex05[,walktime_min :=
  round(apply(fhs2_offspring_ex10_omni1_ex05[,.(walktime_1,walktime_2)],
              MARGIN=1,min),3)]

fhs2_offspring_ex10_omni1_ex05[walktime_min=="-Inf", walktime_min := NA]

fhs2_offspring_ex10_omni1_ex05[
  !is.na(walktime_1)|!is.na(walktime_2), walktime := 0] 


########### Grip (kg)

# Produces highest average by hand (assumes dominant hand is stronger)

fhs2_offspring_ex10_omni1_ex05[,grip_avg_right := 
  round(apply(fhs2_offspring_ex10_omni1_ex05[,.(K1182,K1183,K1184)],MARGIN=1,mean),1)]

fhs2_offspring_ex10_omni1_ex05[,grip_avg_left :=
  round(apply(fhs2_offspring_ex10_omni1_ex05[,.(K1185,K1186,K1187)],MARGIN=1,mean),1)]

fhs2_offspring_ex10_omni1_ex05[,grip_max := 
  round(apply(fhs2_offspring_ex10_omni1_ex05[,c("grip_avg_right","grip_avg_left")],MARGIN=1,max),1)]


#### Physical activity index - Fried

fhs2_offspring_ex10_omni1_ex05[,fhs_pai := 
  calc_framingham_pai(
    as.data.frame(fhs2_offspring_ex10_omni1_ex05),
    slp_hrs="K1234",
    sed_hrs="K1235",
    slgt_hrs="K1236",
    mod_hrs="K1237",
    hvy_hrs="K1238")]

fhs2_offspring_ex10_omni1_ex05[SEX==1&fhs_pai < 30.3, 
                               physical_activity_fried := 1]

fhs2_offspring_ex10_omni1_ex05[SEX==1&fhs_pai >= 30.3, 
                               physical_activity_fried := 0]


fhs2_offspring_ex10_omni1_ex05[
  SEX==2&
    fhs_pai < 30.1, 
  physical_activity_fried := 1]

fhs2_offspring_ex10_omni1_ex05[
  SEX==2&
    fhs_pai >= 30.1, 
  physical_activity_fried := 0]


#### Exhaustion - Fried

fhs2_offspring_ex10_omni1_ex05[
  K1203==3|
    K1216==3, 
  exhaustion_fried := 1]

fhs2_offspring_ex10_omni1_ex05[
  K1203<3&
    K1216<3, 
  exhaustion_fried := 0]

#### Weight loss - Fried

fhs2_offspring_ex10_omni1_ex05[,wgt_delta := as.numeric(WGT) - as.numeric(last_wgt)]

fhs2_offspring_ex10_omni1_ex05[
  wgt_delta <= -10, 
  wtloss_fried := 1]

fhs2_offspring_ex10_omni1_ex05[
  wgt_delta > -10, 
  wtloss_fried := 0]


######### FRAIL Score

#### Fatigue

fhs2_offspring_ex10_omni1_ex05[K1739 <=3, fatigue_frail := 0]

fhs2_offspring_ex10_omni1_ex05[K1739 > 3, fatigue_frail := 1]


#### Resistance

fhs2_offspring_ex10_omni1_ex05[K1224==1, resistance_frail := 0]
fhs2_offspring_ex10_omni1_ex05[K1224==0, resistance_frail := 1]

#### Ambulation

fhs2_offspring_ex10_omni1_ex05[K1222==0, ambulate_frail := 1]
fhs2_offspring_ex10_omni1_ex05[K1222==1, ambulate_frail := 0]


# FRAIL Illness score counts presence of HTN, DM, cancer, chronic lung disease, MI, CHD, angina, asthma, arthritis, stroke, and ckd
# Scores for 0-4 conditions, score = 0, for ≥ 5, score=1


# COPD (chronic lung disease)

fhs2_offspring_ex10_omni1_ex05[K0995 %in% c(0,2), copd_frail := 0]
fhs2_offspring_ex10_omni1_ex05[K0995==1, copd_frail := 1]

# asthma

fhs2_offspring_ex10_omni1_ex05[K0997 %in% c(0,2), asthma_frail := 0]
fhs2_offspring_ex10_omni1_ex05[K0997==1, asthma_frail := 1]

# DJD (arthritis)

fhs2_offspring_ex10_omni1_ex05[K1003 %in% c(0,2), djd_frail := 0]
fhs2_offspring_ex10_omni1_ex05[K1003==1, djd_frail := 1]

# CKD (renal disease)

fhs2_offspring_ex10_omni1_ex05[IDTYPE==1, RACE := 1]
fhs2_offspring_ex10_omni1_ex05[IDTYPE==7, RACE := 2]

fhs2_offspring_ex10_omni1_ex05[,crcl := 
                                 calc_MDRD4(
                                   dat=as.data.frame(fhs2_offspring_ex10_omni1_ex05),
                                   cr="CREAT")]

fhs2_offspring_ex10_omni1_ex05[K0990 %in% c(0,2), ckd_frail := 0]
fhs2_offspring_ex10_omni1_ex05[K0990==1, ckd_frail := 1]

# CHF/MI


fhs2_offspring_ex10_omni1_ex05[,chf_frail := 0]
fhs2_offspring_ex10_omni1_ex05[hfhosp_status==1&
                                 hfhosp_dt <= DATE, chf_frail := 1]

fhs2_offspring_ex10_omni1_ex05[,chd_frail := 0]
fhs2_offspring_ex10_omni1_ex05[cadhosp_status==1&
                                 cadhosp_dt <= DATE, chd_frail := 1]

# Stroke

fhs2_offspring_ex10_omni1_ex05[,stroke_frail := 0]
fhs2_offspring_ex10_omni1_ex05[
  cvahosp_status==1&
    cvahosp_dt <= DATE, 
  stroke_frail := 1]

# Cancer

fhs2_offspring_ex10_omni1_ex05[,cancer_frail := 0]
fhs2_offspring_ex10_omni1_ex05[D_DATE <= DATE, cancer_frail := 1]


#### Calculate FRAIL illness score
fhs2_offspring_ex10_omni1_ex05[,conditions_frail := 
  as.numeric(HRX)+
  as.numeric(DMRX)+
  copd_frail+
  asthma_frail+
  djd_frail+
  ckd_frail+
  chf_frail+
  chd_frail+
  stroke_frail+
  cancer_frail]

fhs2_offspring_ex10_omni1_ex05[conditions_frail <= 4, illness_frail := 0]
fhs2_offspring_ex10_omni1_ex05[conditions_frail > 4, illness_frail := 1]

#### (Weight) Loss

fhs2_offspring_ex10_omni1_ex05[wgt_delta <= -10, wtloss_frail := 1]
fhs2_offspring_ex10_omni1_ex05[wgt_delta > -10, wtloss_frail := 0]


## DENORMALIZE

fhs2_melt_offspring_ex10_omni1_ex05 <- 
  melt(fhs2_offspring_ex10_omni1_ex05,
       id.vars=c("PID","IDTYPE","visit"), 
       na.rm=T, 
       factorsAsStrings=T)

fhs2_melt_offspring_ex10_omni1_ex05[,form := "fhs2_offspring_ex10_omni1_ex05"]

rm(fhs2_offspring_ex10_omni1_ex05)


#==============================#
####  Lab panel, Exam 10     ---#
#==============================#

fhs2_l_fhslab_ex10 <- 
  fread('~/Dropbox/BioLINCC files/Framingham offspring/datasets/CSV/l_fhslab_ex10_1b_1401d.csv',na.strings=c("NA","","NULL"))
names(fhs2_l_fhslab_ex10)[names(fhs2_l_fhslab_ex10)=="idtype"] <- "IDTYPE"
fhs2_l_fhslab_ex10[IDTYPE==1, visit := 10]
fhs2_l_fhslab_ex10[IDTYPE==7, visit := 5]
fhs2_melt_l_fhslab_ex10 <- melt(fhs2_l_fhslab_ex10,id.vars=c("PID","IDTYPE","visit"), na.rm=T, factorsAsStrings=T)
fhs2_melt_l_fhslab_ex10[,form := "fhs2_fhslab_ex10"]

rm(fhs2_l_fhslab_ex10)


#=================================#
####  MMSE, Offspring/Omni     ---#
#=================================#

fhs2_vr_mmse_ex10 <- 
  fread('~/Dropbox/BioLINCC files/Framingham offspring/datasets/CSV/vr_mmse_ex10_1b_1395d.csv',
        na.strings=c("NA","","NULL"))

names(fhs2_vr_mmse_ex10) <- toupper(names(fhs2_vr_mmse_ex10))
setnames(fhs2_vr_mmse_ex10,"EXAM","visit")

fhs2_vr_mmse_ex10[,COGSCR_diff := MAXCOG-COGSCR]

fhs2_melt_vr_mmse_ex10 <- 
  melt(fhs2_vr_mmse_ex10,
       id.vars=c("PID","IDTYPE","visit"), 
       na.rm=T, 
       factorsAsStrings=T)

fhs2_melt_vr_mmse_ex10[,form := "fhs2_mmse_ex10"]

rm(fhs2_vr_mmse_ex10)


#======================================#
####  Fibroscan, Offspring 10/Omni 5     ---#
#======================================#

fhs2_t_livrvcte_ex10 <- 
  fread('~/Dropbox/BioLINCC files/Framingham offspring/datasets/CSV/t_livrvcte_ex10_1b_1413d.csv',
        na.strings=c("NA","","NULL"))

names(fhs2_t_livrvcte_ex10) <- toupper(names(fhs2_t_livrvcte_ex10))
fhs2_t_livrvcte_ex10[IDTYPE==1, visit := 10]
fhs2_t_livrvcte_ex10[IDTYPE==7, visit := 5]

fhs2_melt_t_livrvcte_ex10 <- 
  melt(fhs2_t_livrvcte_ex10,
       id.vars=c("PID","IDTYPE","visit"), 
       na.rm=T, 
       factorsAsStrings=T)

fhs2_melt_t_livrvcte_ex10[,form := "fhs2_livrvcte_ex10"]

rm(fhs2_t_livrvcte_ex10)


# ==============================================================##
####               Assemble Offspring and Omni 1 data         ####
# ==============================================================##


fhs2_melt_all <- rbindlist(
  list(fhs2_melt_act1_5d[,c("PID","IDTYPE","visit","variable","value","form")],
                       fhs2_melt_act1_6d[,c("PID","IDTYPE","visit","variable","value","form")],
                       fhs2_melt_cmrlvh1[,c("PID","IDTYPE","visit","variable","value","form")],
                       fhs2_melt_cmrwma1[,c("PID","IDTYPE","visit","variable","value","form")],
                       fhs2_melt_ex1_1d[,c("PID","IDTYPE","visit","variable","value","form")],
                       fhs2_melt_ex1_2d[,c("PID","IDTYPE","visit","variable","value","form")],
                       fhs2_melt_ex1_3d[,c("PID","IDTYPE","visit","variable","value","form")],
                       fhs2_melt_ex1_4d[,c("PID","IDTYPE","visit","variable","value","form")],
                       fhs2_melt_ex1_5d[,c("PID","IDTYPE","visit","variable","value","form")],
                       fhs2_melt_ex1_6d[,c("PID","IDTYPE","visit","variable","value","form")],
                       fhs2_melt_ex1_7d[,c("PID","IDTYPE","visit","variable","value","form")],
                       fhs2_melt_ex1_8d[,c("PID","IDTYPE","visit","variable","value","form")],
                       fhs2_melt_offspring_ex09_omni1_ex04[,c("PID","IDTYPE","visit","variable","value","form")],
                       fhs2_melt_offspring_ex10_omni1_ex05[,c("PID","IDTYPE","visit","variable","value","form")],
                       fhs2_melt_l_gal3_ex06[,c("PID","IDTYPE","visit","variable","value","form")],
                       fhs2_melt_l_aldost_ex06[,c("PID","IDTYPE","visit","variable","value","form")],
                       fhs2_melt_l_creacys_ex07[,c("PID","IDTYPE","visit","variable","value","form")],
                       fhs2_melt_l_ckd_ex06[,c("PID","IDTYPE","visit","variable","value","form")],
                       fhs2_melt_l_fhslab_ex01[,c("PID","IDTYPE","visit","variable","value","form")],  
                       fhs2_melt_l_fhslab_ex02[,c("PID","IDTYPE","visit","variable","value","form")],
                       fhs2_melt_l_fhslab_ex03[,c("PID","IDTYPE","visit","variable","value","form")],
                       fhs2_melt_l_fhslab_ex05[,c("PID","IDTYPE","visit","variable","value","form")],
                       fhs2_melt_l_fhslab_ex06[,c("PID","IDTYPE","visit","variable","value","form")],
                       fhs2_melt_l_fhslab_ex07[,c("PID","IDTYPE","visit","variable","value","form")], 
                       fhs2_melt_l_fhslab_ex08[,c("PID","IDTYPE","visit","variable","value","form")],
                       fhs2_melt_l_fhslab_ex09[,c("PID","IDTYPE","visit","variable","value","form")],
                       fhs2_melt_l_fhslab_ex10[,c("PID","IDTYPE","visit","variable","value","form")],
                       fhs2_melt_natpep1[,c("PID","IDTYPE","visit","variable","value","form")],
                       fhs2_melt_omni1_ex01[,c("PID","IDTYPE","visit","variable","value","form")],
                       fhs2_melt_omni1_ex02[,c("PID","IDTYPE","visit","variable","value","form")],    
                       fhs2_melt_omni1_ex03[,c("PID","IDTYPE","visit","variable","value","form")],
                       fhs2_melt_q_cesd_ex09[,c("PID","IDTYPE","visit","variable","value","form")],
                       fhs2_melt_q_psych_ex03[,c("PID","IDTYPE","visit","variable","value","form")],
                       fhs2_melt_renin1[,c("PID","IDTYPE","visit","variable","value","form")],
                       fhs2_melt_t_echo_2008[,c("PID","IDTYPE","visit","variable","value","form")],
                       fhs2_melt_t_echo_ex04[,c("PID","IDTYPE","visit","variable","value","form")],
                       fhs2_melt_t_echo_ex05[,c("PID","IDTYPE","visit","variable","value","form")],    
                       fhs2_melt_t_echo_ex06[,c("PID","IDTYPE","visit","variable","value","form")],       
                       fhs2_melt_t_echocs_ex03[,c("PID","IDTYPE","visit","variable","value","form")],
                       fhs2_melt_t_echocs_ex08[,c("PID","IDTYPE","visit","variable","value","form")],
                       fhs2_melt_t_echomvp_ex05[,c("PID","IDTYPE","visit","variable","value","form")],
                       fhs2_melt_t_echodop_ex08[,c("PID","IDTYPE","visit","variable","value","form")],
                       fhs2_melt_t_echola_ex08[,c("PID","IDTYPE","visit","variable","value","form")],
                       fhs2_melt_t_echorv_ex08[,c("PID","IDTYPE","visit","variable","value","form")],
                       fhs2_melt_t_doppvasc_2008[,c("PID","IDTYPE","visit","variable","value","form")],
                       fhs2_melt_t_tonla_ex08[,c("PID","IDTYPE","visit","variable","value","form")],
                       fhs2_melt_t_tonla_ex09[,c("PID","IDTYPE","visit","variable","value","form")],
                       fhs2_melt_t_wktest_07[,c("PID","IDTYPE","visit","variable","value","form")],
                       fhs2_melt_q_ffreq_ex03[,c("PID","IDTYPE","visit","variable","value","form")],
                       fhs2_melt_vr_ffreq_ex05[,c("PID","IDTYPE","visit","variable","value","form")],
                       fhs2_melt_vr_ffreq_ex06[,c("PID","IDTYPE","visit","variable","value","form")],
                       fhs2_melt_vr_ffreq_ex07[,c("PID","IDTYPE","visit","variable","value","form")],
                       fhs2_melt_vr_ffreq_ex08[,c("PID","IDTYPE","visit","variable","value","form")],
                       fhs2_melt_vr_ffreq_omni1_ex02[,c("PID","IDTYPE","visit","variable","value","form")],
                       fhs2_melt_vr_ffreq_omni1_ex03[,c("PID","IDTYPE","visit","variable","value","form")],
                       fhs2_melt_vr_ffreq_offspring_ex09_omni1_ex04[,c("PID","IDTYPE","visit","variable","value","form")],
                       fhs2_melt_wkthru[,c("PID","IDTYPE","visit","variable","value","form")],
                       fhs2_melt_l_crp_ex02[,c("PID","IDTYPE","visit","variable","value","form")],
                       fhs2_melt_l_crp_ex06[,c("PID","IDTYPE","visit","variable","value","form")],
                       fhs2_melt_inflamm_ex07[,c("PID","IDTYPE","visit","variable","value","form")],
                       fhs2_melt_inflamm_ex08[,c("PID","IDTYPE","visit","variable","value","form")],
                       fhs2_melt_l_dbtlab_ex07[,c("PID","IDTYPE","visit","variable","value","form")],
                       fhs2_melt_sex[,c("PID","IDTYPE","visit","variable","value","form")],
                       fhs2_melt_race[,c("PID","IDTYPE","visit","variable","value","form")],
                       fhs2_melt_menses[,c("PID","IDTYPE","visit","variable","value","form")],
                       fhs2_melt_actigraphy_offspring_ex09_omni1_ex04[,c("PID","IDTYPE","visit","variable","value","form")],
                       fhs2_melt_fhorm_ex07[,c("PID","IDTYPE","visit","variable","value","form")],
                       fhs2_melt_shorm_ex03[,c("PID","IDTYPE","visit","variable","value","form")],
                       fhs2_melt_shorm_ex04[,c("PID","IDTYPE","visit","variable","value","form")],
                       fhs2_melt_vr_mmse_ex10[,c("PID","IDTYPE","visit","variable","value","form")],
                       fhs2_melt_t_livrvcte_ex10[,c("PID","IDTYPE","visit","variable","value","form")])
)


fhs2_melt_all <- 
  fhs2_wkthru[,c("PID",
                 "visit",
                 "ATT",
                 "SEX")][fhs2_melt_all,
                         on=.(PID,visit)]

fhs2_melt_all <- fhs2_melt_all[ATT==1]

fhs2_melt_all[,variable := as.character(variable)]

# ============================================================ #
####                  Assemble Framingham DM                ####
# ============================================================ #

fhs1_vr_diab <- fread('~/Dropbox/BioLINCC files/Framingham cohort/datasets/CSV/VR_DIAB_EX28_0_0601D.csv',
                      na.strings=c("NA","","NULL"),
                      stringsAsFactors=F)

names(fhs1_vr_diab) <- toupper(names(fhs1_vr_diab))

fhs1_melt_vr_diab <- melt(fhs1_vr_diab[,c(1,25,82:109)],
                          id.vars=c("PID",
                                    "IDTYPE"),
                          na.rm=T,
                          factorsAsStrings=T)

fhs1_melt_vr_diab[,variable := as.character(variable)]

fhs1_melt_vr_diab[,visit := 
                    str_select(variable,
                                      after="BG200_HX_DIAB")]

fhs1_melt_vr_diab[,variable := "BG200_HX_DIAB"]

fhs1_melt_vr_diab[,form := "fhs1_vr_diab"]

rm(fhs1_vr_diab)

#### Diabetes status by cycle

fhs2_vr_diab <- fread('~/Dropbox/BioLINCC files/Framingham offspring/datasets/CSV/VR_DIAB_EX10_1b_1489D.csv',
                      na.strings=c("NA","","NULL"),
                      stringsAsFactors=F)

names(fhs2_vr_diab) <- toupper(names(fhs2_vr_diab))

fhs2_vr_diab <- 
  fhs2_vr_diab[,c(1,12:22)]

fhs2_melt_vr_diab <- melt(fhs2_vr_diab,
                          id.vars=c("PID",
                                    "IDTYPE"),
                          na.rm=T,
                          factorsAsStrings=T)

fhs2_melt_vr_diab[,variable := as.character(variable)]

fhs2_melt_vr_diab[,visit := 
                    str_select(variable,
                               after="HX_DIAB")]

fhs2_melt_vr_diab[,variable := "HX_DIAB"]

fhs2_melt_vr_diab[,form := "fhs2_vr_diab"]

rm(fhs2_vr_diab)

####

fhs3_vr_diab <- 
  fread('~/Dropbox/BioLINCC files/Framingham third generation/datasets/CSV/VR_DIAB_EX03_3b_1312D.csv',
        na.strings=c("NA","","NULL")) # Diabetes status by cycle

names(fhs3_vr_diab) <- toupper(names(fhs3_vr_diab))

fhs3_melt_vr_diab <- melt(fhs3_vr_diab[,c("PID","IDTYPE","HX_DIAB1","HX_DIAB2")],
                          id.vars=c("PID",
                                    "IDTYPE"),
                          na.rm=T,
                          factorsAsStrings=T)

fhs3_melt_vr_diab[,variable := as.character(variable)]

fhs3_melt_vr_diab[,visit := str_select(variable,
                                       after="HX_DIAB")]

fhs3_melt_vr_diab[,variable := "HX_DIAB"]

fhs3_melt_vr_diab[,form := "fhs3_vr_diab"]

rm(fhs3_vr_diab)

fhs_vr_diab <- 
  rbindlist(
    list(fhs1_melt_vr_diab,
         fhs2_melt_vr_diab,
         fhs3_melt_vr_diab))

names(fhs_vr_diab)[2] <- "cohort"
fhs_vr_diab[,cohort := as.character(cohort)]
fhs_vr_diab[,visit := as.integer(visit)]

fhs_vr_diab <- fhs_dates_long[,c("PID","cohort","visit","visitdays")][fhs_vr_diab,
                     on=.(PID,cohort,visit)]

fhs_vr_diab[,study := "FHS"]

rm(fhs1_melt_vr_diab,
   fhs2_melt_vr_diab,
   fhs3_melt_vr_diab)


# ==========================================================================#
##                                                                         ##
####                  ******** FRAMINGHAM GEN III ********               ####
##                                                                         ##
# ==========================================================================#


#=======================================#
####    *** FHS3: Gen 3 - Exam 1 *** ####
#=======================================#

fhs3_e_exam_ex01_id3 <- 
  fread('~/Dropbox/BioLINCC files/Framingham third generation/datasets/CSV/E_EXAM_EX01_3_0086D.csv')

## Calculate drinks/week by adding beers/wk + white wine/week + red wine/week + liquor/week + other/week

fhs3_e_exam_ex01_id3[,drinks_wk := 
G3A115+
G3A119+
G3A123+
G3A127+
G3A131]

fhs3_e_exam_ex01_id3[,CESD20_missing := 
  rowSums(is.na(fhs3_e_exam_ex01_id3[,402:421]))]

fhs3_e_exam_ex01_id3[,CESD20 := 
  rowSums(fhs3_e_exam_ex01_id3[,402:421])]

fhs3_e_exam_ex01_id3[CESD20_missing >= 4, CESD20 := NA]

fhs3_e_exam_ex01_id3[,ever_smoke := 0]
fhs3_e_exam_ex01_id3[G3A070==1|    # Cigarettes
                       G3A079==1| # Pipes
                       G3A088==1, ever_smoke := 1]  # Cigars

fhs3_e_exam_ex01_id3[!between(G3A023,7,25), G3A023 := NA]
fhs3_e_exam_ex01_id3[!between(G3A032,1,30), G3A032 := NA]
fhs3_e_exam_ex01_id3[!between(G3A034,1,11), G3A034 := NA]
fhs3_e_exam_ex01_id3[!between(G3A035,1,10), G3A035 := NA]
fhs3_e_exam_ex01_id3[!between(G3A036,15,43), G3A036 := NA]
fhs3_e_exam_ex01_id3[!between(G3A037,16,45), G3A037 := NA]
fhs3_e_exam_ex01_id3[!between(G3A040,19,57), G3A040 := NA]
fhs3_e_exam_ex01_id3[!between(G3A044,9,57), G3A044 := NA]
fhs3_e_exam_ex01_id3[!between(G3A051,16,51), G3A051 := NA]
fhs3_e_exam_ex01_id3[!between(G3A055,1,32), G3A055 := NA]
fhs3_e_exam_ex01_id3[!between(G3A056,1,12), G3A056 := NA]

fhs3_e_exam_ex01_id3[,fhs_pai := 
  calc_framingham_pai(fhs3_e_exam_ex01_id3,
                      slp_hrs="G3A596",
                      sed_hrs="G3A597",
                      slgt_hrs="G3A598",
                      mod_hrs="G3A599",
                      hvy_hrs="G3A600")]


fhs3_e_exam_ex01_id3[,hrt_years_total := G3A055 + G3A056/12]

fhs3_e_exam_ex01_id3[G3A483==1, race := "W"]
fhs3_e_exam_ex01_id3[G3A484==1, race := "Hisp"]
fhs3_e_exam_ex01_id3[G3A485==1, race := "B"]
fhs3_e_exam_ex01_id3[G3A486==1, race := "A"]
fhs3_e_exam_ex01_id3[G3A488==1, race := "N"]
fhs3_e_exam_ex01_id3[G3A489==1, race := "O"]


fhs3_meno_ex01_id3 <- 
  fhs3_e_exam_ex01_id3[G3A033 %in% c(0,1),
                       c("PID",
                         "IDTYPE",
                         "G3A023",
                         "G3A045",
                         "G3A051")]


fhs3_melt_e_exam_ex01_id3 <- melt(fhs3_e_exam_ex01_id3,id.vars=c("PID","IDTYPE"),na.rm=T,factorsAsStrings=T)
fhs3_melt_e_exam_ex01_id3[,visit := 1]
fhs3_melt_e_exam_ex01_id3[,form := "fhs3_e_ex01"]


#===================================#
####    FHS3: Omni 2 - Exam 1    ---#
#===================================#

fhs3_e_exam_ex01_id72 <- fread('~/Dropbox/BioLINCC files/Framingham third generation/datasets/CSV/E_EXAM_EX01_72_0652D.csv')

## Calculate drinks/week by adding beers/wk + white wine/week + red wine/week + liquor/week + other/week

fhs3_e_exam_ex01_id72[,drinks_wk := 
  G3A115+
  G3A119+
  G3A123+
  G3A127+
  G3A131]

fhs3_e_exam_ex01_id72[,CESD20_missing := rowSums(is.na(fhs3_e_exam_ex01_id72[,c(paste0("G3A",518:537))]))]
fhs3_e_exam_ex01_id72[,CESD20 := rowSums(fhs3_e_exam_ex01_id72[,c(paste0("G3A",518:537))])]
fhs3_e_exam_ex01_id72[CESD20_missing >= 4, CESD20 := NA]

fhs3_e_exam_ex01_id72[,ever_smoke := 0]
fhs3_e_exam_ex01_id72[G3A070==1|    # Cigarettes
                        G3A079==1| # Pipes
                        G3A088==1, # Cigars 
                      ever_smoke := 1]  


fhs3_e_exam_ex01_id72[,fhs_pai := 
  calc_framingham_pai(fhs3_e_exam_ex01_id72,
                      slp_hrs="G3A596",
                      sed_hrs="G3A597",
                      slgt_hrs="G3A598",
                      mod_hrs="G3A599",
                      hvy_hrs="G3A600")]


fhs3_e_exam_ex01_id72[G3A483==1, race := "W"]
fhs3_e_exam_ex01_id72[G3A484==1, race := "Hisp"]
fhs3_e_exam_ex01_id72[G3A485==1, race := "B"]
fhs3_e_exam_ex01_id72[G3A486==1, race := "A"]
fhs3_e_exam_ex01_id72[G3A488==1, race := "N"]
fhs3_e_exam_ex01_id72[G3A489==1, race := "O"]


fhs3_meno_ex01_id72 <- 
  fhs3_e_exam_ex01_id72[G3A033 %in% c(0,1),c("PID",
                                             "IDTYPE",
                                             "G3A023",
                                             "G3A045",
                                             "G3A051")]


fhs3_melt_e_exam_ex01_id72 <- 
  melt(fhs3_e_exam_ex01_id72,
       id.vars=c("PID","IDTYPE"),
       na.rm=T,
       factorsAsStrings=T)

fhs3_melt_e_exam_ex01_id72[,visit := 1]
fhs3_melt_e_exam_ex01_id72[,form := "fhs3_e_ex01"]


#=======================================#
####  FHS Offspring Spouse - Exam 1  ---#
#=======================================#

fhs3_e_exam_ex01_id2 <- fread('~/Dropbox/BioLINCC files/Framingham third generation/datasets/CSV/E_EXAM_EX01_2_0813D.csv')

## Calculate drinks/week by adding beers/wk + white wine/week + red wine/week + liquor/week + other/week

fhs3_e_exam_ex01_id2[,drinks_wk := 
  G3A115+
  G3A119+
  G3A123+
  G3A127+
  G3A131]

fhs3_e_exam_ex01_id2[,CESD20_missing := rowSums(is.na(fhs3_e_exam_ex01_id2[,c(paste0("G3A",518:537))]))]
fhs3_e_exam_ex01_id2[,CESD20 := rowSums(fhs3_e_exam_ex01_id2[,c(paste0("G3A",518:537))])]
fhs3_e_exam_ex01_id2[CESD20_missing >= 4, CESD20 := NA]

fhs3_e_exam_ex01_id2[,ever_smoke := 0]
fhs3_e_exam_ex01_id2[G3A070==1|    # Cigarettes
                                  G3A079==1| # Pipes
                                  G3A088==1, ever_smoke := 1]  # Cigars

fhs3_e_exam_ex01_id2[,fhs_pai := 
  calc_framingham_pai(fhs3_e_exam_ex01_id2,
                      slp_hrs="G3A596",
                      sed_hrs="G3A597",
                      slgt_hrs="G3A598",
                      mod_hrs="G3A599",
                      hvy_hrs="G3A600")]


fhs3_e_exam_ex01_id2[G3A483==1, race := "W"]
fhs3_e_exam_ex01_id2[G3A484==1, race := "Hisp"]
fhs3_e_exam_ex01_id2[G3A485==1, race := "B"]
fhs3_e_exam_ex01_id2[G3A486==1, race := "A"]
fhs3_e_exam_ex01_id2[G3A488==1, race := "N"]
fhs3_e_exam_ex01_id2[G3A489==1, race := "O"]


fhs3_meno_ex01_id2 <- 
  fhs3_e_exam_ex01_id2[G3A033 %in% c(0,1),
                       c("PID",
                         "IDTYPE",
                         "G3A023",
                         "G3A045",
                         "G3A051")]

fhs3_meno_ex01 <- 
  rbindlist(
    list(fhs3_meno_ex01_id2,
        fhs3_meno_ex01_id3,
        fhs3_meno_ex01_id72))


fhs3_melt_e_exam_ex01_id2 <- melt(fhs3_e_exam_ex01_id2,id.vars=c("PID","IDTYPE"),na.rm=T,factorsAsStrings=T)
fhs3_melt_e_exam_ex01_id2[,visit := 1]
fhs3_melt_e_exam_ex01_id2[,form := "fhs3_e_ex01"]

fhs3_menarche <-
  rbindlist(
    list(fhs3_melt_e_exam_ex01_id2[variable=="G3A023", .(PID,IDTYPE,value)], 
         fhs3_melt_e_exam_ex01_id3[variable=="G3A023", .(PID,IDTYPE,value)], 
         fhs3_melt_e_exam_ex01_id72[variable=="G3A023", .(PID,IDTYPE,value)])
  )


fhs3_menarche <-
  fhs3_menarche[value < 25]

setnames(fhs3_menarche,"value","age_menarche")

#======================================================#
####  FHS3: Gen 3, Offspring Spouse, Omni 2 - Race  ---#
#======================================================#


fhs3_race_id2 <- fhs3_e_exam_ex01_id2[,c("PID",
                                         "IDTYPE",
                                         "G3A483",
                                         "G3A484",
                                         "G3A485",
                                         "G3A486",
                                         "G3A488",
                                         "G3A489"
)]


fhs3_race_id3 <- fhs3_e_exam_ex01_id3[,c("PID",
                                         "IDTYPE",
                                         "G3A483",
                                         "G3A484",
                                         "G3A485",
                                         "G3A486",
                                         "G3A488",
                                         "G3A489"
)]


fhs3_race_id72 <- fhs3_e_exam_ex01_id72[,c("PID",
                                           "IDTYPE",
                                           "G3A483",
                                           "G3A484",
                                           "G3A485",
                                           "G3A486",
                                           "G3A488",
                                           "G3A489"
)]


fhs3_race <- 
  rbindlist(
    list(fhs3_race_id2,
         fhs3_race_id3,
         fhs3_race_id72))


fhs3_race[G3A483==1, race := 1]
fhs3_race[G3A489==1, race := 6]
fhs3_race[G3A485==1, race := 2]
fhs3_race[G3A486==1, race := 4]
fhs3_race[G3A488==1, race := 5]
fhs3_race[G3A484==1, race := 3]

fhs3_melt_race <- melt(fhs3_race,
                       id.vars=c("PID","IDTYPE"),
                       na.rm=T,
                       stringsAsFactors=F)
fhs3_melt_race[,visit := 1]
fhs3_melt_race[,form <- "fhs3_race"]

names(fhs3_race)[3:8] <- c("White",
                           "Hispanic",
                           "Black",
                           "Asian",
                           "Native American",
                           "Other")


#=========================================#
####  NT-proBNP, Framingham 3, Exam 1  ---#
#=========================================#

fhs3_bnp3 <- fread('~/Dropbox/BioLINCC files/Framingham third generation/datasets/CSV/BNP3_1D.csv')
fhs3_melt_bnp3 <- melt(fhs3_bnp3,
                       id.vars=c("PID",
                                 "IDTYPE"), 
                       na.rm=T, 
                       factorsAsStrings=T)

fhs3_melt_bnp3[,visit := 1]
fhs3_melt_bnp3[,form := "fhs3_bnp3"]

rm(fhs3_bnp3)


#=============================================================#
####  CRP, Framingham Offspring Spouse and Omni 2, Exam 1  ---#
#=============================================================#

fhs3_inflamm <- fread('~/Dropbox/BioLINCC files/Framingham third generation/datasets/CSV/l_inflamm_ex01_3b_1107d.csv')
fhs3_melt_inflamm <- melt(fhs3_inflamm,id.vars=c("PID","IDTYPE"),na.rm=T,factorsAsStrings = T)
fhs3_melt_inflamm[,visit := 1]
fhs3_melt_inflamm[,form := "fhs3_inflamm"]

rm(fhs3_inflamm)

#========================================================================#
####  Aldosterone/renin, Framingham 3/Offspring Spouse/Omni 2,Exam 1  ---#
#========================================================================#

fhs3_renaldo3 <- fread('~/Dropbox/BioLINCC files/Framingham third generation/datasets/CSV/RENALDO3_1D_V1.csv')
fhs3_renaldo3[,aldo_renin := (ALDO/10)/RENIN]
fhs3_melt_renaldo3 <-  melt(fhs3_renaldo3,id.vars=c("PID","IDTYPE"),na.rm=T,factorsAsStrings = T)
fhs3_melt_renaldo3[,visit := 1]
fhs3_melt_renaldo3[,form := "fhs3_renaldo3"]

rm(fhs3_renaldo3)


#==================================================================#
####  Estrogen levels, Framingham 3, Exam 1/Omni 2, Exam 1      ---#
#==================================================================#

fhs3_estr_ex01 <- fread('~/Dropbox/BioLINCC files/Framingham third generation/datasets/CSV/l_estr_2005_m_0622d_v1.csv')
fhs3_melt_estr_ex01 <-  melt(fhs3_estr_ex01,id.vars=c("PID","IDTYPE"),na.rm=T,factorsAsStrings = T)
fhs3_melt_estr_ex01[,visit := 1]
fhs3_melt_estr_ex01[,form := "fhs3_estr_ex01"]

rm(fhs3_estr_ex01)


#==================================================================#
####  Troponin levels, Framingham 3, Exam 1/Omni 2, Exam 1      ---#
#==================================================================#

fhs3_tni_ex01 <- fread('~/Dropbox/BioLINCC files/Framingham third generation/datasets/CSV/l_hstn_ex01_3b_0832d_v1.csv')
fhs3_melt_tni_ex01 <-  melt(fhs3_tni_ex01,id.vars=c("PID","IDTYPE"),na.rm=T,factorsAsStrings = T)
fhs3_melt_tni_ex01[,visit := 1]
fhs3_melt_tni_ex01[,form := "fhs3_tni_ex01"]

rm(fhs3_tni_ex01)

#================================================================#
####  Male hormones, Framingham 3, Exam 1/Omni 2, Exam 1      ---#
#================================================================#

fhs3_mhorm_ex01 <- fread('~/Dropbox/BioLINCC files/Framingham third generation/datasets/CSV/l_mhorm_2005_m_0490d_v1.csv')
fhs3_melt_mhorm_ex01 <-  melt(fhs3_mhorm_ex01,id.vars=c("PID","IDTYPE"),na.rm=T,factorsAsStrings = T)
fhs3_melt_mhorm_ex01[,visit := 1]
fhs3_melt_mhorm_ex01[,form := "fhs3_mhorm_ex01"]

rm(fhs3_mhorm_ex01)

#=========================================#
####  Telomeres, Framingham 3, Exam 1  ---#
#=========================================#

fhs3_telomere_ex01 <- fread('~/Dropbox/BioLINCC files/Framingham third generation/datasets/CSV/l_telomere_2005_m_1161d.csv')
fhs3_melt_telomere_ex01 <-  melt(fhs3_telomere_ex01,id.vars=c("PID","IDTYPE"),na.rm=T,factorsAsStrings = T)
fhs3_melt_telomere_ex01[,visit := 1]
fhs3_melt_telomere_ex01[,form := "fhs3_telomere_ex01"]

rm(fhs3_telomere_ex01)

#=========================================#
####  VEGF, Framingham 3,      Exam 1  ---#
#=========================================#

fhs3_sflt3_ex01 <- fread('~/Dropbox/BioLINCC files/Framingham third generation/datasets/CSV/sflt3_1d.csv')
fhs3_melt_sflt3_ex01 <-  melt(fhs3_sflt3_ex01,id.vars=c("PID","IDTYPE"),na.rm=T,factorsAsStrings = T)
fhs3_melt_sflt3_ex01[,visit := 1]
fhs3_melt_sflt3_ex01[,form := "fhs3_sflt3_ex01"]

rm(fhs3_sflt3_ex01)


#================================================#
####  CT abdominal fat, Framingham 3, Exam 1  ---#
#================================================#

fhs3_ctabdom_ex01 <- fread('~/Dropbox/BioLINCC files/Framingham third generation/datasets/CSV/t_ctabdom_2005_m_0227d.csv')

fhs3_ctabdom_ex01[,vat_sat_ratio := OB2VOL/OB1VOL]

fhs3_ctabdom_ex01[, vat_sat_ratio_log := log(vat_sat_ratio)]

fhs3_melt_ctabdom_ex01 <-  melt(fhs3_ctabdom_ex01,id.vars=c("PID","IDTYPE"),na.rm=T,factorsAsStrings = T)
fhs3_melt_ctabdom_ex01[,visit := 1]
fhs3_melt_ctabdom_ex01[,form := "fhs3_ctabdom_ex01"]

rm(fhs3_ctabdom_ex01)


#=======================================#
####  CT Lung, Framingham 3, Exam 1  ---#
#=======================================#

fhs3_ctlung_ex01 <- fread('~/Dropbox/BioLINCC files/Framingham third generation/datasets/CSV/t_ctlung_2005_m_0696d.csv')

fhs3_ctlung_ex01[
  ,
  global_vol_ml_avg :=
    rowMeans(
      cbind(GLOBAL_VOL_ML_1, GLOBAL_VOL_ML_2),
      na.rm = TRUE
    )
]
fhs3_melt_ctlung_ex01 <-  melt(fhs3_ctlung_ex01,id.vars=c("PID","IDTYPE"),na.rm=T,factorsAsStrings = T)
fhs3_melt_ctlung_ex01[,visit := 1]
fhs3_melt_ctlung_ex01[,form := "fhs3_ctlung_ex01"]

rm(fhs3_ctlung_ex01)


#======================================#
####  CAC, Framingham 3, Exam 1     ---#
#======================================#

fhs3_cac_ex01 <- fread('~/Dropbox/BioLINCC files/Framingham third generation/datasets/CSV/t_ctthrcac_ex01_3_0287d.csv')

  fhs3_cac_ex01[
    ,
    cac_avg :=
      rowMeans(
        cbind(CA1, CA2),
        na.rm = TRUE
      )]
  
fhs3_melt_cac_ex01 <-  melt(fhs3_cac_ex01,id.vars=c("PID","IDTYPE"),na.rm=T,factorsAsStrings = T)
fhs3_melt_cac_ex01[,visit := 1]
fhs3_melt_cac_ex01[,form := "fhs3_cac_ex01"]

rm(fhs3_cac_ex01)


#==============================================================================#
####  Vascular tonometry, Framingham 3/New Offspring Spouse/Omni 2, Exam 1  ---#
#==============================================================================#

fhs3_tonometry_ex01 <- fread('~/Dropbox/BioLINCC files/Framingham third generation/datasets/CSV/t_tonla_ex01_3b_1234d.csv')


fhs3_tonometry_ex01[,e_eprime_avg := MEMAX/DLEMAX]
fhs3_tonometry_ex01[,ea_ratio := MEMAX/MAMAX]

fhs3_melt_tonometry_ex01 <-  melt(fhs3_tonometry_ex01,id.vars=c("PID","IDTYPE"),na.rm=T,factorsAsStrings = T)
fhs3_melt_tonometry_ex01[,visit := 1]
fhs3_melt_tonometry_ex01[,form := "fhs3_tonometry_ex01"]

rm(fhs3_tonometry_ex01)


#========================================================#
####  FHS lab values, Offspring Spouse/Omni 2,Exam 1  ---#
#========================================================#

fhs3_fhslab_ex01 <- fread('~/Dropbox/BioLINCC files/Framingham third generation/datasets/CSV/L_FHSLAB_EX01_3B_0800D.csv')
fhs3_fhslab_ex01 <- 
  fhs3_wkthru[,c("PID","IDTYPE","SEX","AGE1")][fhs3_fhslab_ex01,
                          on=.(PID,IDTYPE)]

labcols <- c(names(fhs3_fhslab_ex01),"gfr_mdrd")

fhs3_fhslab_ex01 <- fhs3_race[fhs3_fhslab_ex01,
                          on=.(PID,IDTYPE)]

fhs3_fhslab_ex01[,gfr_mdrd := 
  calc_MDRD4(dat=fhs3_fhslab_ex01,
             cr="CREAT",
             age="AGE1",
             sex="SEX",
             race="Black",
             black=1)]

fhs3_melt_fhslab_ex01 <- melt(fhs3_fhslab_ex01[,..labcols],
                              id.vars=c("PID","IDTYPE"),
                              na.rm=T,
                              factorsAsStrings = T)

fhs3_melt_fhslab_ex01[,visit := 1]
fhs3_melt_fhslab_ex01[,form := "fhs3_fhslab_ex01"]
rm(fhs3_fhslab_ex01)


#================================================================#
####   Male hormones in females, third generation, Exam 1     ---#
#================================================================#

fhs3_fhorm_ex01 <- fread('~/Dropbox/BioLINCC files/Framingham third generation/datasets/CSV/fhorm_2005d.csv')
fhs3_melt_fhorm_ex01 <-  melt(fhs3_fhorm_ex01,id.vars=c("PID","IDTYPE"),na.rm=T,factorsAsStrings = T)
fhs3_melt_fhorm_ex01[,visit := 1]
fhs3_melt_fhorm_ex01[,form := "fhs3_fhorm_ex01"]

rm(fhs3_fhorm_ex01)


#===============================================================#
####  Echocardiogram - FHS Offspring Spouse/Omni 2 - Exam 1  ---#
#===============================================================#

fhs3_t_echo_2008 <- fread('~/Dropbox/BioLINCC files/Framingham third generation/datasets/CSV/T_ECHO_2008_M_0549D_V1.csv')


fhs3_t_echo_2008 <- 
  fhs3_wkthru[,c("PID","IDTYPE","SEX","WGT1","HGT1")][fhs3_t_echo_2008,
on=.(PID,IDTYPE)]

fhs3_t_echo_2008[,HEIGHT := HGT1*2.54]
fhs3_t_echo_2008[,WEIGHT := WGT1/2.2]

fhs3_t_echo_2008 <- 
  as.data.table(
    calc_hypertrophy_type(
      fhs3_t_echo_2008, 
      id = "PID",
      sex="SEX",
      lvedd="X76",
      ivsd="X62",
      lvpwtd="X69",
      height="HEIGHT",
      weight="WEIGHT"))

fhs3_melt_t_echo_2008 <- melt(fhs3_t_echo_2008,id.vars=c("PID","IDTYPE"), na.rm=T, factorsAsStrings=T)
fhs3_melt_t_echo_2008[,visit := 1]

fhs3_melt_t_echo_2008[,form := "fhs3_echo_ex01"]

rm(fhs3_t_echo_2008)

#================================================================================#
####         Echo Doppler Diastolic function - FHS Gen 3/Omni 2 - Exam 1      ---#
#================================================================================#

fhs3_t_doppvasc_dd_2008  <- fread('~/Dropbox/BioLINCC files/Framingham third generation/datasets/CSV/T_DOPPVASC_2008_M_0756D.csv')
fhs3_t_doppvasc_dd_2008[,visit := 1]
fhs3_t_doppvasc_dd_2008[,earatio := MEMAX/MAMAX]
fhs3_t_doppvasc_dd_2008[,e_eprime_lateral := MEMAX/DEMAX]

fhs3_melt_t_doppvasc_dd_2008 <- melt(fhs3_t_doppvasc_dd_2008,
                                     id.vars=c("PID","IDTYPE","visit"), 
                                     na.rm=T, 
                                     factorsAsStrings=T)
fhs3_melt_t_doppvasc_dd_2008[,form := "fhs3_doppvasc_2008"]

rm(fhs3_t_doppvasc_dd_2008)


#======================================================#
####      Echocardiogram - FHS Gen 3 - Exam 1       ---#
#======================================================#

fhs3_t_echo_ex01_id3  <- fread('~/Dropbox/BioLINCC files/Framingham third generation/datasets/CSV/T_ECHO_EX01_3_0042D.csv')

fhs3_t_echo_ex01_id3 <-
  fhs3_wkthru[,c("PID","IDTYPE","SEX","WGT1","HGT1")][fhs3_t_echo_ex01_id3,
                                                      on=.(PID,IDTYPE)]

fhs3_t_echo_ex01_id3[,HEIGHT := HGT1*2.54]
fhs3_t_echo_ex01_id3[,WEIGHT := WGT1/2.2]

fhs3_t_echo_ex01_id3 <- 
  as.data.table(
    calc_hypertrophy_type(
      fhs3_t_echo_ex01_id3, 
      id = "PID",
      sex="SEX",
      lvedd="X76",
      ivsd="X62",
      lvpwtd="X69",
      height="HEIGHT",
      weight="WEIGHT"))


fhs3_t_echo_ex01_id3[,LVEDD_index :=
  X83/bsa]

fhs3_melt_t_echo_ex01_id3 <- melt(fhs3_t_echo_ex01_id3,
                                  id.vars=c("PID","IDTYPE"),
                                  na.rm=T,
                                  factorsAsStrings=T)

fhs3_melt_t_echo_ex01_id3[,visit := 1]
fhs3_melt_t_echo_ex01_id3[,form := "fhs3_echo_ex01"]

rm(fhs3_t_echo_ex01_id3)

#==========================================#
####      PFT - FHS Gen 3 - Exam 1      ---#
#==========================================#

fhs3_t_pft_ex01_id3  <- fread('~/Dropbox/BioLINCC files/Framingham third generation/datasets/CSV/T_PFT_EX01_3_0170D.csv')

fhs3_melt_t_pft_ex01_id3 <- melt(fhs3_t_pft_ex01_id3,id.vars=c("PID","IDTYPE"),na.rm=T,factorsAsStrings=T)
fhs3_melt_t_pft_ex01_id3[,visit := 1]
fhs3_melt_t_pft_ex01_id3[,form := "fhs3_pft_ex01"]


rm(fhs3_t_pft_ex01_id3)

#===================================================================#
####  PFT Diffusion test - FHS Offspring Spouse/Omni 2 - Exam 1  ---#
#===================================================================#

fhs3_t_pftdiff_2005  <- fread('~/Dropbox/BioLINCC files/Framingham third generation/datasets/CSV/T_PFTDIFF_2005_M_0762D.csv')
fhs3_t_pftdiff_2005[,DLCO_VA_mean := rowMeans(fhs3_t_pftdiff_2005[,c('DVA1_1','DVA2_1','DVA3_1')],na.rm=T)]

fhs3_melt_t_pftdiff_2005 <- melt(fhs3_t_pftdiff_2005,id.vars=c("PID","IDTYPE"),na.rm=T,factorsAsStrings=T)
fhs3_melt_t_pftdiff_2005[,visit := 1]
fhs3_melt_t_pftdiff_2005[,form := "fhs3_pftdiff_2005"]

rm(fhs3_t_pftdiff_2005)


#-------------------------------------------------#
####  PFT Diffusion test - FHS Gen 3 - Exam 1  ---#
#-------------------------------------------------#

fhs3_t_pftdiff_ex01_id3  <- fread('~/Dropbox/BioLINCC files/Framingham third generation/datasets/CSV/T_PFTDIFF_EX01_3_0868D.csv')
fhs3_t_pftdiff_ex01_id3[,DLCO_VA_mean := DVA_1_3]

fhs3_melt_t_pftdiff_ex01_id3 <- melt(fhs3_t_pftdiff_ex01_id3,id.vars=c("PID","IDTYPE"),na.rm=T,factorsAsStrings=T)
fhs3_melt_t_pftdiff_ex01_id3[,visit := 1]
fhs3_melt_t_pftdiff_ex01_id3[,form := "fhs3_pftdiff_ex01"]

rm(fhs3_t_pftdiff_ex01_id3)

#======================================================================#
####  Dietary guidelines adherence index 2010 - FHS Gen 3 - Exam 1  ---#
#======================================================================#

fhs3_vr_dgai2010_ex01_id3  <- fread('~/Dropbox/BioLINCC files/Framingham third generation/datasets/CSV/VR_DGAI2010_EX01_3_1078D.csv')

fhs3_melt_vr_dgai2010_ex01_id3 <- melt(fhs3_vr_dgai2010_ex01_id3,id.vars=c("PID","IDTYPE"),na.rm=T,factorsAsStrings=T)
fhs3_melt_vr_dgai2010_ex01_id3[,visit := 1]
fhs3_melt_vr_dgai2010_ex01_id3[,form := "fhs3_dgai_ex01"]

rm(fhs3_vr_dgai2010_ex01_id3)


#===========================================================#
####  Food frequency questionnaire - FHS Gen 3 - Exam 1  ---#
#===========================================================#

fhs3_vr_ffreq_ex01_id3  <- fread('~/Dropbox/BioLINCC files/Framingham third generation/datasets/CSV/VR_FFREQ_EX01_3_0587D.csv')

## Convert servings/week  to  servings/day
### Coffee  (FFD112)

fhs3_vr_ffreq_ex01_id3[,FFD112_daily  := FFD112/7] 

### Decaf coffee (FFD111)
fhs3_vr_ffreq_ex01_id3[,FFD111_daily  :=  FFD111/7]

### Tea (FFD113)
fhs3_vr_ffreq_ex01_id3[,FFD113_daily  :=  FFD113/7]

fhs3_melt_vr_ffreq_ex01_id3 <- melt(fhs3_vr_ffreq_ex01_id3,id.vars=c("PID","IDTYPE"),na.rm=T,factorsAsStrings=T)
fhs3_melt_vr_ffreq_ex01_id3[,visit := 1]
fhs3_melt_vr_ffreq_ex01_id3[,form := "fhs3_vr_ffreq"]

rm(fhs3_vr_ffreq_ex01_id3)


#======================================================================#
####  Food frequency questionnaire - FHS Offspring Spouse - Exam 1  ---#
#======================================================================#

fhs3_vr_ffreq_ex01_id2  <- fread('~/Dropbox/BioLINCC files/Framingham third generation/datasets/CSV/VR_FFREQ_EX01_2_0984d_v1.csv')

## Convert servings/week  to  servings/day
### Coffee  (FFD112)

fhs3_vr_ffreq_ex01_id2[,FFD112_daily  := FFD112/7]

### Decaf coffee (FFD111)
fhs3_vr_ffreq_ex01_id2[,FFD111_daily  := FFD111/7]

### Tea (FFD113)
fhs3_vr_ffreq_ex01_id2[,FFD113_daily  := FFD113/7] 


fhs3_melt_vr_ffreq_ex01_id2 <- melt(fhs3_vr_ffreq_ex01_id2,id.vars=c("PID","IDTYPE"),na.rm=T,factorsAsStrings=T)
fhs3_melt_vr_ffreq_ex01_id2[,visit := 1]
fhs3_melt_vr_ffreq_ex01_id2[,form := "fhs3_vr_ffreq"]

rm(fhs3_vr_ffreq_ex01_id2)


#============================================================#
####  Food frequency questionnaire - FHS Omni 2 - Exam 1  ---#
#============================================================#

fhs3_vr_ffreq_ex01_id72  <- fread('~/Dropbox/BioLINCC files/Framingham third generation/datasets/CSV/VR_FFREQ_EX01_72_0975d_v1.csv')

## Convert servings/week  to  servings/day
### Coffee  (FFD112)

fhs3_vr_ffreq_ex01_id72[,FFD112_daily  := FFD112/7]

### Decaf coffee (FFD111)
fhs3_vr_ffreq_ex01_id72[,FFD111_daily  := FFD111/7]

### Tea (FFD113)
fhs3_vr_ffreq_ex01_id72[,FFD113_daily  := FFD113/7]


fhs3_melt_vr_ffreq_ex01_id72 <- melt(fhs3_vr_ffreq_ex01_id72,id.vars=c("PID","IDTYPE"),na.rm=T,factorsAsStrings=T)
fhs3_melt_vr_ffreq_ex01_id72[,visit := 1]
fhs3_melt_vr_ffreq_ex01_id72[,form := "fhs3_vr_ffreq"]

rm(fhs3_vr_ffreq_ex01_id72)


#==========================================================#
####  FHS3: Gen 3, Offspring Spouse, Omni 2 - Menarche  ---#
#==========================================================#


fhs3_menarche <- 
  rbindlist(
    list(
      fhs3_e_exam_ex01_id2[,.(PID,IDTYPE,G3A023)],
                       fhs3_e_exam_ex01_id3[,.(PID,IDTYPE,G3A023)],
                       fhs3_e_exam_ex01_id72[,.(PID,IDTYPE,G3A023)]))

fhs3_menarche <- 
  fhs3_menarche[!G3A023 %in% c(88,888)]

names(fhs3_menarche)[3] <- "age_menarche"

rm(fhs3_e_exam_ex01_id2)
rm(fhs3_e_exam_ex01_id3)
rm(fhs3_e_exam_ex01_id72)


#### ********************************************** Continue here ***************************************************#####

#==========================================================#
####  **** FHS 3/Offspring Spouse/Omni 2 - Exam 2 ****  ####
#==========================================================#

fhs3_e_exam_2011 <- fread('~/Dropbox/BioLINCC files/Framingham third generation/datasets/CSV/E_EXAM_2011_M_0017D_V3.csv')


fhs3_e_exam_2011[!is.na(G3B0028), (22:64) := NA]
fhs3_e_exam_2011 <-
  fhs3_menarche[fhs3_e_exam_2011,on=.(PID,IDTYPE)]

fhs3_e_exam_2011[,drinks_wk := 
  G3B0105+  # Beer
  G3B0107+  # Wine
  G3B0109]   # Liquor

fhs3_e_exam_2011[!between(G3B0032,1,8), G3B0032 := NA]
fhs3_e_exam_2011[!between(G3B0033,1,3), G3B0033 := NA]
fhs3_e_exam_2011[!between(G3B0081,17,62), G3B0081 := NA]
fhs3_e_exam_2011[!between(G3B0085,27,58), G3B0085 := NA]
fhs3_e_exam_2011[!between(G3B0089,27,65), G3B0089 := NA]

fhs3_meno_ex02 <-
  fhs3_e_exam_2011[is.na(G3B0028),
                   c("PID",
                     "IDTYPE",
                     "G3B0073",
                     "G3B0081")]


fhs3_e_exam_2011[!G3B0081==0&!is.na(G3B0081), years_menses := G3B0081 - age_menarche]


fhs3_e_exam_2011[,fhs_pai := 
  calc_framingham_pai(fhs3_e_exam_2011,
                      slp_hrs="G3B0703",
                      sed_hrs="G3B0704",
                      slgt_hrs="G3B0705",
                      mod_hrs="G3B0706",
                      hvy_hrs="G3B0707")]


fhs3_melt_e_exam_2011 <- melt(fhs3_e_exam_2011,id.vars=c("PID","IDTYPE"),na.rm=T,factorsAsStrings = T)
fhs3_melt_e_exam_2011[,visit := 2]
fhs3_melt_e_exam_2011[,form := "fhs3_e_exam_2011"]

rm(fhs3_e_exam_2011)


#======================================#
####  CAC, Framingham 3, Exam 2     ---#
#======================================#

fhs3_cac_ex02 <- fread('~/Dropbox/BioLINCC files/Framingham third generation/datasets/CSV/t_ctthrcac_2011_m_0682d.csv')
fhs3_melt_cac_ex02 <-  melt(fhs3_cac_ex02,id.vars=c("PID","IDTYPE"),na.rm=T,factorsAsStrings = T)
fhs3_melt_cac_ex02[,visit := 2]
fhs3_melt_cac_ex02[,form := "fhs3_cac_ex02"]

rm(fhs3_cac_ex02)


#======================================#
####  PASE, Framingham 3, Exam 2     ---#
#======================================#

# Physical activity score for the elderly

fhs3_pase_ex02 <- 
  fread('~/Dropbox/BioLINCC files/Framingham third generation/datasets/CSV/vr_pase_ex02_3_0642d.csv')
fhs3_melt_pase_ex02 <-  melt(fhs3_pase_ex02,id.vars=c("PID","IDTYPE"),na.rm=T,factorsAsStrings = T)
fhs3_melt_pase_ex02[,visit := 2]
fhs3_melt_pase_ex02[,form := "fhs3_pase_ex02"]

rm(fhs3_pase_ex02)


#======================================================================#
####  Actigraphy, Framingham 3/New Offspring Spouse/Omni 2, Exam 2  ---#
#======================================================================#

act_fields <-
  c("zmin_vdt",
    "zmin_vdtd",
    "zmin_vdtm",
    "zmin_vdtwd",
    "zmin_vdtwdd",
    "zmin_vdtwdm",
    "zmin_vdtwe",
    "zmin_vdtwed",
    "zmin_vdtwedm",
    "wstep_vdt",
    "wstep_vdtd",
    "wstep_vdtm",
    "wstep_vdtwd",
    "wstep_vdtwdd",
    "wstep_vdtwdm",
    "wstep_vdtwe",
    "wstep_vdtwed",
    "wstep_vdtwedm",
    "zb_vdt",
    "zb_vdtd",
    "zb_vdtmin",
    "zb_vdtwd",
    "zb_vdtwdmin",
    "zb_vdtwe",
    "zb_vdtwed",
    "zb_vdtwedd",
    "zb_vdtwemin",
    "vigstepvdt",
    "vigstepvdtd",
    "vigstepvdtwd",
    "vigstepvdtwdd",
    "vigstepvdtwe",
    "vigstepvdtwed",
    "vigminvdt",
    "vigminvdtd",
    "vigminvdtwd",
    "vigminvdtwdd",
    "vigminvdtwe",
    "vigminvdtwed",
    "vigcntvdt",
    "vigcntvdtd",
    "vigcntvdtwd",
    "vigcntvdtwdd",
    "vigcntvdtwe",
    "vigcntvdtwed",
    "sedcntvdt",
    "sedcntvdtd",
    "sedcntvdtwd",
    "sedcntvdtwdd",
    "sedcntvdtwe",
    "sedcntvdtwed",
    "sedminvdt",
    "sedminvdtd",
    "sedminvdtwd",
    "sedminvdtwdd",
    "sedminvdtwe",
    "sedminvdtwed",
    "sedminwed",
    "bgt_litcntvdt",
    "bgt_litcntvdtd",
    "bgt_litcntvdtwd",
    "bgt_litcntvdtwdd",
    "bgt_litcntvdtwe",
    "bgt_litcntvdtwed",
    "bgt_litminvdt",
    "bgt_litminvdtd",
    "bgt_litminvdtwd",
    "bgt_litminvdtwdd",
    "bgt_litminvdtwe",
    "bgt_litminvdtwed",
    "bgt_litstepvdt",
    "bgt_litstepvdtd",
    "bgt_litstepvdtwd",
    "bgt_litstepvdtwdd",
    "bgt_litstepvdtwe",
    "bgt_litstepvdtwed",
    "bgt_modcntvdt",
    "bgt_modcntvdtd",
    "bgt_modcntvdtwd",
    "bgt_modcntvdtwdd",
    "bgt_modcntvdtwe",
    "bgt_modcntvdtwed",
    "bgt_modminvdt",
    "bgt_modminvdtd",
    "bgt_modminvdtwd",
    "bgt_modminvdtwdd",
    "bgt_modminvdtwe",
    "bgt_modminvdtwed",
    "bgt_modstepvdt",
    "bgt_modstepvdtd",
    "bgt_modstepvdtwd",
    "bgt_modstepvdtwdd",
    "bgt_modstepvdtwe",
    "bgt_modstepvdtwed",
    "bgt_mvpcntvdt",
    "bgt_mvpcntvdtd",
    "bgt_mvpcntvdtwd",
    "bgt_mvpcntvdtwdd",
    "bgt_mvpcntvdtwe",
    "bgt_mvpcntvdtwed",
    "bgt_mvpminvdt",
    "bgt_mvpminvdtd",
    "bgt_mvpminvdtwd",
    "bgt_mvpminvdtwdd",
    "bgt_mvpminvdtwe",
    "bgt_mvpminvdtwed",
    "bgt_mvpstepvdt",
    "bgt_mvpstepvdtd",
    "bgt_mvpstepvdtwd",
    "bgt_mvpstepvdtwdd",
    "bgt_mvpstepvdtwe",
    "bgt_mvpstepvdtwed",
    "bgt_sedcntvdt",
    "bgt_sedcntvdtd",
    "bgt_sedcntvdtwd",
    "bgt_sedcntvdtwdd",
    "bgt_sedcntvdtwe",
    "bgt_sedcntvdtwed",
    "bgt_sedminvdt",
    "bgt_sedminvdtd",
    "bgt_sedminvdtwd",
    "bgt_sedminvdtwdd",
    "bgt_sedminvdtwe",
    "bgt_sedminvdtwed",
    "bgt_sedstepvdt",
    "bgt_sedstepvdtd",
    "bgt_sedstepvdtwd",
    "bgt_sedstepvdtwdd",
    "bgt_sedstepvdtwe",
    "bgt_sedstepvdtwed",
    "bgt_vigcntvdt",
    "bgt_vigcntvdtd",
    "bgt_vigcntvdtwd",
    "bgt_vigcntvdtwdd",
    "bgt_vigcntvdtwe",
    "bgt_vigcntvdtwed",
    "bgt_vigminvdt",
    "bgt_vigminvdtd",
    "bgt_vigminvdtwd",
    "bgt_vigminvdtwdd",
    "bgt_vigminvdtwe",
    "bgt_vigminvdtwed",
    "bgt_vigstepvdt",
    "bgt_vigstepvdtd",
    "bgt_vigstepvdtwd",
    "bgt_vigstepvdtwdd",
    "bgt_vigstepvdtwe",
    "bgt_vigstepvdtwed",
    "litcntvdt",
    "litcntvdtd",
    "litcntvdtwd",
    "litcntvdtwdd",
    "litcntvdtwe",
    "litcntvdtwed",
    "litminvdt",
    "litminvdtd",
    "litminvdtwd",
    "litminvdtwdd",
    "litminvdtwe",
    "litminvdtwed",
    "litstepvdt",
    "litstepvdtd",
    "litstepvdtwd",
    "litstepvdtwdd",
    "litstepvdtwe",
    "litstepvdtwed",
    "modcntvdt",
    "modcntvdtd",
    "modcntvdtwd",
    "modcntvdtwdd",
    "modcntvdtwe",
    "modcntvdtwed",
    "modminvdt",
    "modminvdtd",
    "modminvdtwd",
    "modminvdtwdd",
    "modminvdtwe",
    "modminvdtwed",
    "modstepvdt",
    "modstepvdtd",
    "modstepvdtwd",
    "modstepvdtwdd",
    "modstepvdtwe",
    "modstepvdtwed",
    "motcntvdt",
    "motcntvdtd",
    "motcntvdtwd",
    "motcntvdtwdd",
    "motcntvdtwe",
    "motcntvdtwed",
    "motminvdt",
    "motminvdtd",
    "motminvdtwd",
    "motminvdtwdd",
    "motminvdtwe",
    "motminvdtwed",
    "motstepvdt",
    "motstepvdtd",
    "motstepvdtwd",
    "motstepvdtwdd",
    "motstepvdtwe",
    "motstepvdtwed",
    "mvpcntvdt",
    "mvpcntvdtd",
    "mvpcntvdtwd",
    "mvpcntvdtwdd",
    "mvpcntvdtwe",
    "mvpcntvdtwed",
    "mvpminvdt",
    "mvpminvdtd",
    "mvpminvdtwd",
    "mvpminvdtwdd",
    "mvpminvdtwe",
    "mvpminvdtwed",
    "mvpstepvdt",
    "mvpstepvdtd",
    "mvpstepvdtwd",
    "mvpstepvdtwdd",
    "mvpstepvdtwe",
    "mvpstepvdtwed",
    "s_litcntvdt",
    "s_litcntvdtd",
    "s_litcntvdtwd",
    "s_litcntvdtwdd",
    "s_litcntvdtwe",
    "s_litcntvdtwed",
    "s_litminvdt",
    "s_litminvdtd",
    "s_litminvdtwd",
    "s_litminvdtwdd",
    "s_litminvdtwe",
    "s_litminvdtwed",
    "s_litstepvdt",
    "s_litstepvdtd",
    "s_litstepvdtwd",
    "s_litstepvdtwdd",
    "s_litstepvdtwe",
    "s_litstepvdtwed",
    "s_modcntvdt",
    "s_modcntvdtd",
    "s_modcntvdtwd",
    "s_modcntvdtwdd",
    "s_modcntvdtwe",
    "s_modcntvdtwed",
    "s_modminvdt",
    "s_modminvdtd",
    "s_modminvdtwd",
    "s_modminvdtwdd",
    "s_modminvdtwe",
    "s_modminvdtwed",
    "s_modstepvdt",
    "s_modstepvdtd",
    "s_modstepvdtwd",
    "s_modstepvdtwdd",
    "s_modstepvdtwe",
    "s_modstepvdtwed",
    "s_mvpcntvdt",
    "s_mvpcntvdtd",
    "s_mvpcntvdtwd",
    "s_mvpcntvdtwdd",
    "s_mvpcntvdtwe",
    "s_mvpcntvdtwed",
    "s_mvpminvdt",
    "s_mvpminvdtd",
    "s_mvpminvdtwd",
    "s_mvpminvdtwdd",
    "s_mvpminvdtwe",
    "s_mvpminvdtwed",
    "s_mvpstepvdt",
    "s_mvpstepvdtd",
    "s_mvpstepvdtwd",
    "s_mvpstepvdtwdd",
    "s_mvpstepvdtwe",
    "s_mvpstepvdtwed",
    "s_sedcntvdt",
    "s_sedcntvdtd",
    "s_sedcntvdtwd",
    "s_sedcntvdtwdd",
    "s_sedcntvdtwe",
    "s_sedcntvdtwed",
    "s_sedminvdt",
    "s_sedminvdtd",
    "s_sedminvdtwd",
    "s_sedminvdtwdd",
    "s_sedminvdtwe",
    "s_sedminvdtwed",
    "s_sedstepvdt",
    "s_sedstepvdtd",
    "s_sedstepvdtwd",
    "s_sedstepvdtwdd",
    "s_sedstepvdtwe",
    "s_sedstepvdtwed",
    "s_vigcntvdt",
    "s_vigcntvdtd",
    "s_vigcntvdtwd",
    "s_vigcntvdtwdd",
    "s_vigcntvdtwe",
    "s_vigcntvdtwed",
    "s_vigminvdt",
    "s_vigminvdtd",
    "s_vigminvdtwd",
    "s_vigminvdtwdd",
    "s_vigminvdtwe",
    "s_vigminvdtwed",
    "s_vigstepvdt",
    "s_vigstepvdtd",
    "s_vigstepvdtwd",
    "s_vigstepvdtwdd",
    "s_vigstepvdtwe",
    "s_vigstepvdtwed",
    "zstep_vdt",
    "zstep_vdtd",
    "zstep_vdtm",
    "zstep_vdtwd",
    "zstep_vdtwdd",
    "zstep_vdtwdm",
    "zstep_vdtwe",
    "zstep_vdtwed",
    "zstep_vdtwedm",
    "zcnt_vdt",
    "zcnt_vdtd",
    "zcnt_vdtm",
    "zcnt_vdtwd",
    "zcnt_vdtwdd",
    "zcnt_vdtwdm",
    "zcnt_vdtwe",
    "zcnt_vdtwed",
    "zcnt_vdtwedm",
    "zb_vdt",
    "zb_vdtd",
    "zb_vdtmin",
    "zb_vdtwd",
    "zb_vdtwdmin",
    "zb_vdtwe",
    "zb_vdtwed",
    "zb_vdtwedd",
    "zb_vdtwemin",
    "wb_vdt",
    "wb_vdtd",
    "wb_vdtmin",
    "wb_vdtwd",
    "wb_vdtwdmin",
    "wb_vdtwe",
    "wb_vdtwed",
    "wb_vdtwedd",
    "wb_vdtwemin",
    "wcnt_vdt",
    "wcnt_vdtd",
    "wcnt_vdtm",
    "wcnt_vdtwd",
    "wcnt_vdtwdd",
    "wcnt_vdtwdm",
    "wcnt_vdtwe",
    "wcnt_vdtwed",
    "wcnt_vdtwedm",
    "wmin_vdt",
    "wmin_vdtd",
    "wmin_vdtm",
    "wmin_vdtwd",
    "wmin_vdtwdd",
    "wmin_vdtwdm",
    "wmin_vdtwe",
    "wmin_vdtwed",
    "wmin_vdtwedm",
    "wstep_vdt",
    "wstep_vdtd",
    "wstep_vdtm",
    "wstep_vdtwd",
    "wstep_vdtwdd",
    "wstep_vdtwdm",
    "wstep_vdtwe",
    "wstep_vdtwed",
    "wstep_vdtwedm")

act_fields <- toupper(act_fields)

fhs3_actigraphy_ex02 <- 
  as.data.table(
    read_sas('~/Dropbox/BioLINCC files/Framingham third generation/datasets/t_physactf_ex02_3b_0914d.sas7bdat',
             skip=1))

fhs3_melt_actigraphy_ex02 <-  melt(fhs3_actigraphy_ex02,id.vars=c("PID","IDTYPE"),na.rm=T,factorsAsStrings = T)
fhs3_melt_actigraphy_ex02[,visit := 2]
fhs3_melt_actigraphy_ex02[,form := "fhs3_actigraphy_ex02"]

fhs3_melt_actigraphy_ex02 <- 
  fhs3_melt_actigraphy_ex02[variable %in% act_fields]

rm(fhs3_actigraphy_ex02)


#==============================================================================#
####  Vascular tonometry, Framingham 3/New Offspring Spouse/Omni 2, Exam 2  ---#
#==============================================================================#

fhs3_tonometry_ex02 <- fread('~/Dropbox/BioLINCC files/Framingham third generation/datasets/CSV/t_tonla_ex02_3b_1235d.csv')
fhs3_melt_tonometry_ex02 <-  melt(fhs3_tonometry_ex02,id.vars=c("PID","IDTYPE"),na.rm=T,factorsAsStrings = T)
fhs3_melt_tonometry_ex02[,visit := 2]
fhs3_melt_tonometry_ex02[,form := "fhs3_tonometry_ex02"]

rm(fhs3_tonometry_ex02)

#=============================================#
####  Whole body DXA Framingham 3, Exam 2  ---#
#=============================================#

fhs3_wbdxa_ex02_pt1 <- fread('~/Dropbox/BioLINCC files/Framingham third generation/datasets/CSV/t_wbdxa_2010_3_0585d_v1.csv')
fhs3_wbdxa_ex02_pt2 <- fread('~/Dropbox/BioLINCC files/Framingham third generation/datasets/CSV/t_wbdxa_2011_3_0624d.csv')
fhs3_wbdxa_ex02 <-
  rbindlist(
    list(fhs3_wbdxa_ex02_pt1,
        fhs3_wbdxa_ex02_pt2))
fhs3_melt_wbdxa_ex02 <-  melt(fhs3_wbdxa_ex02,id.vars=c("PID","IDTYPE"),na.rm=T,factorsAsStrings = T)
fhs3_melt_wbdxa_ex02[,visit := 2]
fhs3_melt_wbdxa_ex02[,form := "fhs3_wbdxa_ex02"]

rm(fhs3_wbdxa_ex02)


#===============================================================#
####  FHS labs, Framingham 3/Offspring Spouse/Omni 2,Exam 2  ---#
#===============================================================#

fhs3_fhslab_2011 <- fread('~/Dropbox/BioLINCC files/Framingham third generation/datasets/CSV/L_FHSLAB_2011_M_0656D_V1.csv')
fhs3_fhslab_2011 <- 
  fhs3_wkthru[,.(PID,IDTYPE,SEX,AGE2)][fhs3_fhslab_2011,
                                               on=.(PID,IDTYPE)]
labcols <- c(names(fhs3_fhslab_2011),"gfr_mdrd")
fhs3_fhslab_2011 <- 
  fhs3_race[fhs3_fhslab_2011,
            on=.(PID,IDTYPE)]

fhs3_fhslab_2011[,gfr_mdrd := 
                   calc_MDRD4(dat=fhs3_fhslab_2011,
                              cr="CREAT",
                              age="AGE2",
                              sex="SEX",
                              race="Black",
                              black=1)]

fhs3_melt_fhslab_2011 <- melt(fhs3_fhslab_2011[,..labcols],
                              id.vars=c("PID","IDTYPE"),
                              na.rm=T,
                              factorsAsStrings = T)
fhs3_melt_fhslab_2011[,visit := 2]
fhs3_melt_fhslab_2011[,form := "fhs3_fhslab_2011"]

rm(fhs3_fhslab_2011)


#=========================================================================#
####  PFT Diffusion test - FHS Gen 3/Offspring Spouse/Omni 2 - Exam 2  ---#
#=========================================================================#


fhs3_t_pftdiff_2011  <- fread('~/Dropbox/BioLINCC files/Framingham third generation/datasets/CSV/T_PFTDIFF_2011_M_0648D_V2.csv')
fhs3_t_pftdiff_2011[,DLCO_VA_mean := 
                      rowMeans(fhs3_t_pftdiff_2011[,c('DVA1_2','DVA2_2','DVA3_2')],na.rm=T)]

fhs3_melt_t_pftdiff_2011 <- melt(fhs3_t_pftdiff_2011,id.vars=c("PID","IDTYPE"),na.rm=T,factorsAsStrings=T)
fhs3_melt_t_pftdiff_2011[,visit := 1]
fhs3_melt_t_pftdiff_2011[,form := "fhs3_pftdiff_2011"]

rm(fhs3_t_pftdiff_2011)


#======================================================================#
####  Dietary guidelines adherence index 2010 - FHS Gen 3 - Exam 2  ---#
#======================================================================#

fhs3_vr_dgai2010_ex02_id3  <- fread('~/Dropbox/BioLINCC files/Framingham third generation/datasets/CSV/VR_DGAI2010_EX02_3_0996D.csv')
fhs3_melt_vr_dgai2010_ex02_id3 <- melt(fhs3_vr_dgai2010_ex02_id3,id.vars=c("PID","IDTYPE"),na.rm=T,factorsAsStrings=T)
fhs3_melt_vr_dgai2010_ex02_id3[,visit := 2]
fhs3_melt_vr_dgai2010_ex02_id3[,form := "fhs3_dgai"]

rm(fhs3_vr_dgai2010_ex02_id3)


#===========================================================#
####  Food frequency questionnaire - FHS Gen 3 - Exam 2  ---#
#===========================================================#

fhs3_vr_ffreq_ex02_id3  <- fread('~/Dropbox/BioLINCC files/Framingham third generation/datasets/CSV/VR_FFREQ_EX02_3b_1345D.csv')
fhs3_melt_vr_ffreq_ex02_id3 <- melt(fhs3_vr_ffreq_ex02_id3,id.vars=c("PID","IDTYPE"),na.rm=T,factorsAsStrings=T)
fhs3_melt_vr_ffreq_ex02_id3[,visit := 2]
fhs3_melt_vr_ffreq_ex02_id3[,form := "fhs3_vr_ffreq"]

rm(fhs3_vr_ffreq_ex02_id3)


#=========================================#
####  CT Lung, Framingham 3, Exam 2    ---#
#=========================================#

fhs3_ctlung_ex02 <- fread('~/Dropbox/BioLINCC files/Framingham third generation/datasets/CSV/t_ctlung_2011_m_0829d.csv')
fhs3_melt_ctlung_ex02 <-  melt(fhs3_ctlung_ex02,id.vars=c("PID","IDTYPE"),na.rm=T,factorsAsStrings = T)
fhs3_melt_ctlung_ex02[,visit := 2]
fhs3_melt_ctlung_ex02[,form := "fhs3_ctlung_ex02"]

rm(fhs3_ctlung_ex02)


#================================================#
####  CT abdominal fat, Framingham 3, Exam 2  ---#
#================================================#

fhs3_ctabdom_ex02 <- fread('~/Dropbox/BioLINCC files/Framingham third generation/datasets/CSV/t_ctabfat_2011_m_0669d.csv')
fhs3_melt_ctabdom_ex02 <-  melt(fhs3_ctabdom_ex02,id.vars=c("PID","IDTYPE"),na.rm=T,factorsAsStrings = T)
fhs3_melt_ctabdom_ex02[,visit := 2]
fhs3_melt_ctabdom_ex02[,form := "fhs3_ctabdom_ex02"]

rm(fhs3_ctabdom_ex02)


#==========================================================#
####  **** FHS 3/Offspring Spouse/Omni 2 - Exam 3 ****  ####
#==========================================================#

fhs3_e_exam_ex03 <- fread('~/Dropbox/BioLINCC files/Framingham third generation/datasets/CSV/e_exam_ex03_3b_1069d.csv')

fhs3_e_exam_ex03[!between(G3C0234,15,63), G3C0234 := NA]
fhs3_e_exam_ex03[!between(G3C0238,30,87), G3C0238 := NA]
fhs3_e_exam_ex03[!between(G3C0242,22,87), G3C0242 := NA]

fhs3_e_exam_ex03[,drinks_wk := 
  G3C0264+  # Beer
  G3C0268+  # Wine
  G3C0272]  # Liquor

fhs3_e_exam_ex03[,fhs_pai := 
  calc_framingham_pai(fhs3_e_exam_ex03,
                      slp_hrs="G3C0748",
                      sed_hrs="G3C0749",
                      slgt_hrs="G3C0750",
                      mod_hrs="G3C0751",
                      hvy_hrs="G3C0752")]


fhs3_melt_e_exam_ex03 <- melt(fhs3_e_exam_ex03,id.vars=c("PID","IDTYPE"),na.rm=T,factorsAsStrings = T)
fhs3_melt_e_exam_ex03[,visit := 3]
fhs3_melt_e_exam_ex03[,form := "fhs3_e_ex03"]


#=========================================================#
####  Menopause/HRT - FHS 3/Offspring Spouse/Omni 2   ----#
#=========================================================#

fhs3_women <-
  fhs3_wkthru[SEX==2,.(PID,IDTYPE)]

fhs3_vr_meno_ex02_id2  <- fread('~/Dropbox/BioLINCC files/Framingham third generation/datasets/CSV/VR_MENO_EX02_2_0719D.csv')
fhs3_vr_meno_ex02_id3  <- fread('~/Dropbox/BioLINCC files/Framingham third generation/datasets/CSV/VR_MENO_EX02_3_0653D_V1.csv')
fhs3_vr_meno_ex02_id72  <- fread('~/Dropbox/BioLINCC files/Framingham third generation/datasets/CSV/VR_MENO_EX02_72_0720D.csv')

fhs3_vr_meno_ex02 <- 
  rbindlist(
    list(fhs3_vr_meno_ex02_id2,
        fhs3_vr_meno_ex02_id3,
        fhs3_vr_meno_ex02_id72))

fhs3_women <-
  fhs3_vr_meno_ex02[fhs3_women,on=.(PID,IDTYPE)]

fhs3_meno_ex03 <- 
  fhs3_e_exam_ex03[!G3C0234==88,
    c("PID",
      "IDTYPE",
      "G3C0234",
      "G3C0235",
      "G3C0237",
      "G3C0238",
      "G3C0241",
      "G3C0243")]

fhs3_menocalcs_ex03 <-
  fhs3_meno_ex03[fhs3_women,
                 on=.(PID,IDTYPE)]

fhs3_menocalcs_ex03 <-
  fhs3_menarche[fhs3_menocalcs_ex03,
        on=.(PID,IDTYPE)]

setnames(fhs3_menocalcs_ex03,
         c("G3C0234","G3C0235","G3C0241","G3C0243"),
         c("STOPAGE3","CAUSE3","OOPHOR3","OVREM3"))

fhs3_menocalcs_ex03[STOPAGE==0, STOPAGE := NA]

fhs3_menocalcs_ex03[,age_menopause := STOPAGE3]
fhs3_menocalcs_ex03[is.na(age_menopause),age_menopause := STOPAGE]


fhs3_menocalcs_ex03[,meno := "N"]
fhs3_menocalcs_ex03[!is.na(age_menopause),meno := "Y"]

fhs3_menocalcs_ex03[CAUSE==2|CAUSE3==2,meno_cause := 2]
fhs3_menocalcs_ex03[CAUSE==3|CAUSE3==3,meno_cause := 3]
fhs3_menocalcs_ex03[is.na(meno_cause)&(CAUSE==1|CAUSE3==1),meno_cause := 1]


fhs3_menocalcs_ex03[age_menopause > 0, years_menses := age_menopause-age_menarche]
fhs3_menocalcs_ex03[age_menopause == 0, age_menopause := 0]


fhs3_menocalcs_ex03[,num_ovaries_removed :=
  apply(fhs3_menocalcs_ex03[,c("OVREM1","OVREM2","OVREM3")],
        MARGIN=1,
        function(x) max(x, na.rm=T))]

fhs3_menocalcs_ex03[num_ovaries_removed %in% c("-Inf",0),num_ovaries_removed := NA]

fhs3_menocalcs_ex03[meno=="Y",oophorectomy := "N"]
fhs3_menocalcs_ex03[num_ovaries_removed %in% c(1,2),oophorectomy := "Y"]

fhs3_menocalcs_ex03[oophorectomy=="Y",age_oophorectomy := age_menopause] 

fhs3_melt_menocalcs_ex03 <-
  melt(fhs3_menocalcs_ex03[,c("PID",
                              "IDTYPE",
                              "age_menarche",
                              "meno",
                              "age_menopause",
                              "years_menses",
                              "meno_cause",
                              "oophorectomy",
                              "num_ovaries_removed",
                              "age_oophorectomy")],
       id.vars=c("PID",
                 "IDTYPE"),
       na.rm=T)

fhs3_melt_menocalcs_ex03[,form := "fhs3_menocalcs_ex03"]
fhs3_melt_menocalcs_ex03[,visit := 3]


#======================================================================#
####  Actigraphy, Framingham 3, New Offspring Spouse/Omni 2 Exam 3  ---#
#======================================================================#


fhs3_actigraphy_ex03 <- 
  as.data.table(
    read_sas('~/Dropbox/BioLINCC files/Framingham third generation/datasets/t_physactf_ex03_3b_1007d.sas7bdat',
             skip=1))

fhs3_melt_actigraphy_ex03 <-  melt(fhs3_actigraphy_ex03,id.vars=c("PID","IDTYPE"),na.rm=T,factorsAsStrings = T)
fhs3_melt_actigraphy_ex03[,visit := 3]
fhs3_melt_actigraphy_ex03[,form := "fhs3_actigraphy_ex03"]

fhs3_melt_actigraphy_ex03 <- 
  fhs3_melt_actigraphy_ex03[variable %in% act_fields]

rm(fhs3_actigraphy_ex03)


#=====================================================================#
####  Validated education Framingham 3/New Offspring Spouse/Omni 2 ---#
#=====================================================================#

fhs3_melt_vr_educ <- fread('~/Dropbox/BioLINCC files/Framingham third generation/datasets/CSV/vr_educ_2018_a_1307d.csv')
fhs3_melt_vr_educ <- fhs3_melt_vr_educ[!EDUCATION==""]
fhs3_melt_vr_educ[,visit := 1]
fhs3_melt_vr_educ[,form := "fhs3_melt_vr_educ"]
fhs3_melt_vr_educ[,variable := "EDUCATION"]
fhs3_melt_vr_educ[,value := EDUCATION]

#==========================================================================#
####  Validated smoking status Framingham 3/New Offspring Spouse/Omni 2 ---#
#==========================================================================#

fhs3_vr_smoke <- fread('~/Dropbox/BioLINCC files/Framingham third generation/datasets/CSV/vr_smokst_2019_m_1325d.csv')
fhs3_melt_vr_smoke <-  melt(fhs3_vr_smoke,id.vars=c("PID","IDTYPE"),na.rm=T,factorsAsStrings = T)
fhs3_melt_vr_smoke[,visit := substr(variable,7,7)]
fhs3_melt_vr_smoke[,variable := "smoke_status_vr"]
fhs3_melt_vr_smoke[,form := "fhs3_vr_smokst"]

rm(fhs3_vr_smoke)


#===============================================================================#
####  FHS Labs, Framingham 3, Exam 3/Offspring Spouse Exam 3/Omni 2, Exam 3  ---#
#===============================================================================#

fhs3_fhslab_ex03 <- fread('~/Dropbox/BioLINCC files/Framingham third generation/datasets/CSV/L_FHSLAB_EX03_3B_1170D.csv')
fhs3_fhslab_ex03 <- 
  fhs3_wkthru[,c("PID","IDTYPE","SEX","AGE3")][fhs3_fhslab_ex03,
                                               on=.(PID,IDTYPE)]
labcols <- c(names(fhs3_fhslab_ex03),"gfr_mdrd")

fhs3_fhslab_ex03 <- 
  fhs3_race[fhs3_fhslab_ex03,
            on=.(PID,IDTYPE)]

fhs3_fhslab_ex03[,gfr_mdrd := 
                   calc_MDRD4(dat=fhs3_fhslab_ex03,
                              cr="CREATININE",
                              age="AGE3",
                              sex="SEX",
                              race="Black",
                              black=1)]

fhs3_melt_fhslab_ex03 <- 
  melt(fhs3_fhslab_ex03[,..labcols],
       id.vars=c("PID","IDTYPE"),
       na.rm=T,
       factorsAsStrings = T)

fhs3_melt_fhslab_ex03[,visit := 3]
fhs3_melt_fhslab_ex03[,form := "fhs3_fhslab_ex03"]
rm(fhs3_fhslab_ex03)


#================================================#
####      Wkthru (reference) - FHS Gen 3      ####
#================================================#

fhs3_wkthru <-
        fhs3_race[,c("PID","IDTYPE","Black")][fhs3_wkthru,
        on=.(PID,IDTYPE)]

fhs3_wkthru[,WTCHG_PCT2 :=
  round((WGT2-WGT1)/WGT1 * 100,1)]

fhs3_wkthru[,WTCHG_PCT3 :=
  round((WGT3-WGT2)/WGT2 * 100,1)]

fhs3_wkthru[,WHR2 := WAIST2/HIP2]

fhs3_wkthru[,WHR3 := WAIST3/HIP3]


fhs3_wkthru[,gfr_mdrd1 := 
              calc_MDRD4(dat=fhs3_wkthru,
                         cr="CREAT1",
                         age="AGE1",
                         sex="SEX",
                         race="Black",
                         black=1)]

fhs3_wkthru[,HGT1_cm := HGT1*2.54]
fhs3_wkthru[,HGT2_cm := HGT2*2.54]
fhs3_wkthru[,HGT3_cm := HGT3*2.54]

fhs3_wkthru[,WGT1_kg := WGT1/2.2]
fhs3_wkthru[,WGT2_kg := WGT2/2.2]
fhs3_wkthru[,WGT3_kg := WGT3/2.2]

fhs3_wkthru[,bsa1 := calc_bsa(fhs3_wkthru,weight_kg="WGT1_kg",height_cm = "HGT1_cm")]

fhs3_wkthru[,bsa2 := calc_bsa(fhs3_wkthru,weight_kg="WGT2_kg",height_cm = "HGT2_cm")]

fhs3_wkthru[,bsa3 := calc_bsa(fhs3_wkthru,weight_kg="WGT3_kg",height_cm = "HGT3_cm")]

fhs3_melt_wkthru <- 
  melt(fhs3_wkthru,
       id.vars=c("PID","IDTYPE","SEX"),
       na.rm=T,
       factorsAsStrings=T)

fhs3_melt_wkthru[,variable := as.character(variable)]
fhs3_melt_wkthru[,visit := 
                   substring(variable,
                             nchar(variable))]

fhs3_melt_wkthru[,variable := 
                   substring(variable,
                             1, nchar(variable)-1)]

fhs3_melt_wkthru[variable=="WAIST", value :=  value * 2.54]

fhs3_melt_wkthru_kg <- fhs3_melt_wkthru[variable=="WGT"]

fhs3_melt_wkthru_kg[,value := value/2.20462]

fhs3_melt_wkthru_kg[,variable := "WGT_KG"]

fhs3_melt_wkthru <- 
  rbindlist(
    list(
      fhs3_melt_wkthru,
      fhs3_melt_wkthru_kg))

fhs3_melt_wkthru[variable=="WAIST", value := value * 2.54]

fhs3_melt_wkthru[,form := "fhs3_wkthru"]


#===================================================================#
####  Assemble FHS Gen 3/Offspring Spouse/Omni 2 cohorts         ####
#===================================================================#


fhs3_melt_all <- 
  rbindlist(
    list(
      fhs3_melt_bnp3[,c("PID","IDTYPE","visit","variable","value","form")],
      fhs3_melt_inflamm[,c("PID","IDTYPE","visit","variable","value","form")],
      fhs3_melt_renaldo3[,c("PID","IDTYPE","visit","variable","value","form")],
      fhs3_melt_fhslab_2011[,c("PID","IDTYPE","visit","variable","value","form")],
      fhs3_melt_fhslab_ex01[,c("PID","IDTYPE","visit","variable","value","form")],
      fhs3_melt_fhslab_ex03[,c("PID","IDTYPE","visit","variable","value","form")],
      fhs3_melt_mhorm_ex01[,c("PID","IDTYPE","visit","variable","value","form")],
      fhs3_melt_e_exam_2011[,c("PID","IDTYPE","visit","variable","value","form")],
      fhs3_melt_e_exam_ex01_id2[,c("PID","IDTYPE","visit","variable","value","form")],
      fhs3_melt_e_exam_ex01_id3[,c("PID","IDTYPE","visit","variable","value","form")],
      fhs3_melt_e_exam_ex01_id72[,c("PID","IDTYPE","visit","variable","value","form")],
      fhs3_melt_e_exam_ex03[,c("PID","IDTYPE","visit","variable","value","form")],
      fhs3_melt_estr_ex01[,c("PID","IDTYPE","visit","variable","value","form")],
      fhs3_melt_t_echo_2008[,c("PID","IDTYPE","visit","variable","value","form")],
      fhs3_melt_t_doppvasc_dd_2008[,c("PID","IDTYPE","visit","variable","value","form")],
      fhs3_melt_t_echo_ex01_id3[,c("PID","IDTYPE","visit","variable","value","form")],
      fhs3_melt_t_pft_ex01_id3[,c("PID","IDTYPE","visit","variable","value","form")],
      fhs3_melt_t_pftdiff_2005[,c("PID","IDTYPE","visit","variable","value","form")],
      fhs3_melt_t_pftdiff_2011[,c("PID","IDTYPE","visit","variable","value","form")],
      fhs3_melt_t_pftdiff_ex01_id3[,c("PID","IDTYPE","visit","variable","value","form")],
      fhs3_melt_vr_dgai2010_ex01_id3[,c("PID","IDTYPE","visit","variable","value","form")],
      fhs3_melt_vr_dgai2010_ex02_id3[,c("PID","IDTYPE","visit","variable","value","form")],
      fhs3_melt_vr_ffreq_ex01_id3[,c("PID","IDTYPE","visit","variable","value","form")],
      fhs3_melt_vr_ffreq_ex02_id3[,c("PID","IDTYPE","visit","variable","value","form")],
      fhs3_melt_vr_ffreq_ex01_id2[,c("PID","IDTYPE","visit","variable","value","form")],
      fhs3_melt_vr_ffreq_ex01_id72[,c("PID","IDTYPE","visit","variable","value","form")],
      fhs3_melt_wkthru[,c("PID","IDTYPE","visit","variable","value","form")],
      fhs3_melt_sex[,c("PID","IDTYPE","visit","variable","value","form")],
      fhs3_melt_tni_ex01[,c("PID","IDTYPE","visit","variable","value","form")],
      fhs3_melt_telomere_ex01[,c("PID","IDTYPE","visit","variable","value","form")],
      fhs3_melt_sflt3_ex01[,c("PID","IDTYPE","visit","variable","value","form")],
      fhs3_melt_ctabdom_ex01[,c("PID","IDTYPE","visit","variable","value","form")],
      fhs3_melt_ctlung_ex01[,c("PID","IDTYPE","visit","variable","value","form")],
      fhs3_melt_ctlung_ex02[,c("PID","IDTYPE","visit","variable","value","form")],
      fhs3_melt_cac_ex01[,c("PID","IDTYPE","visit","variable","value","form")],
      fhs3_melt_cac_ex02[,c("PID","IDTYPE","visit","variable","value","form")],
      fhs3_melt_tonometry_ex01[,c("PID","IDTYPE","visit","variable","value","form")],
      fhs3_melt_tonometry_ex02[,c("PID","IDTYPE","visit","variable","value","form")],
      fhs3_melt_wbdxa_ex02[,c("PID","IDTYPE","visit","variable","value","form")],
      fhs3_melt_vr_educ[,c("PID","IDTYPE","visit","variable","value","form")],
      fhs3_melt_vr_smoke[,c("PID","IDTYPE","visit","variable","value","form")],
      fhs3_melt_actigraphy_ex02[,c("PID","IDTYPE","visit","variable","value","form")],
      fhs3_melt_actigraphy_ex03[,c("PID","IDTYPE","visit","variable","value","form")],
      fhs3_melt_menocalcs_ex03[,c("PID","IDTYPE","visit","variable","value","form")],
      fhs3_melt_pase_ex02[,c("PID","IDTYPE","visit","variable","value","form")]
    )
  )

fhs3_breastfeed <-
  dcast(
    subset(fhs3_melt_all, variable %in% c(
      "G3A035",
      "G3B0033",
      "G3C0205",
      "G3B0041",
      "G3B0042",
      "G3B0046",
      "G3B0047",
      "G3B0051",
      "G3B0052",
      "G3B0056",
      "G3B0057",
      "G3B0061",
      "G3B0062",
      "G3B0066",
      "G3B0067",
      "G3B0071",
      "G3B0072",
      "G3C0209",
      "G3C0210",
      "G3C0214",
      "G3C0215",
      "G3C0219",
      "G3C0220",
      "G3C0224",
      "G3C0225")),
    PID+IDTYPE~variable)

fhs3_breastfeed <-
  fhs3_breastfeed[
         !(is.na(G3A035)&
             is.na(G3B0033)&
             is.na(G3C0205))]

fhs3_breastfeed <- fhs3_breastfeed[!G3A035=="88"]
fhs3_breastfeed <- fhs3_breastfeed[!G3B0033=="88"]
fhs3_breastfeed <- fhs3_breastfeed[!G3C0205=="88"]

fhs3_breastfeed[,num_breastfeed := 
  apply(fhs3_breastfeed[,c("G3B0041","G3B0046","G3B0051","G3B0056","G3B0061","G3C0209","G3C0214")],
        MARGIN=1,
        FUN=function(x) sum(as.numeric(x),na.rm=T))]

fhs3_melt_breastfeed <- 
  melt(fhs3_breastfeed,
       id=c("PID","IDTYPE"),
       na.rm=T)

fhs3_melt_breastfeed[,visit := 3]
fhs3_melt_breastfeed[,form := "fhs3_breastfeed"]

fhs3_melt_all <-
  rbindlist(
    list(fhs3_melt_all[,c("PID","IDTYPE","visit","variable","value","form")],
        fhs3_melt_breastfeed[,c("PID","IDTYPE","visit","variable","value","form")]))

fhs3_no_attend <-
  fhs3_melt_wkthru[variable=='ATT',.(PID,visit,att=value)]

fhs3_melt_all <- fhs3_no_attend[fhs3_melt_all,on=.(PID,visit)]

fhs3_melt_all <- fhs3_melt_all[att==1]

fhs3_melt_all[,variable := as.character(variable)]

#===============================================================##
####              Assemble all FHS cohorts                    ####
#===============================================================##

names(fhs_vr_diab)[2] <- "IDTYPE"

fhs_melt_all <- 
  rbindlist(
    list(
      fhs1_melt_all[,c("PID","IDTYPE","visit","form","variable","value")],
      fhs2_melt_all[,c("PID","IDTYPE","visit","form","variable","value")],
      fhs3_melt_all[,c("PID","IDTYPE","visit","form","variable","value")],
      fhs_vr_diab[,c("PID","IDTYPE","visit","form","variable","value")]
    )
  )

fhs_melt_all[,PID := as.character(PID)]
names(fhs_melt_all)[1:2] <- c("patientid","cohort")

fhs_dates_long[,PID := as.character(PID)]
fhs_dates_long[,visit := as.character(visit)]

fhs_melt_all <- 
  fhs_dates_long[,c("PID","cohort","visit","visit_yr","visitdays","cohort_name","age_obs")][fhs_melt_all,
        on=.(PID=patientid,cohort,visit)]

fhs_melt_all[,visit := as.integer(visit)]
fhs_melt_all[,visit_yr := as.integer(visit_yr)]

fhs_melt_all[,study := "FHS"]

fhs_melt_all[,form := toupper(form)]
fhs_melt_all[,variable := toupper(variable)]

fhs_melt_all[,study_field := 
               paste(study,
                     form,
                     variable,
                     sep="_")]

fhs_melt_all[,datapoint := 
  paste("FHS",row.names(fhs_melt_all),sep="")]

fhs_melt_all[,patientid := as.character(PID)]

write_parquet(fhs_melt_all[,..data_fields],
              "~/Dropbox/ADAPT-HF/Master HDCP files/Cohort data/alldata_fhs.parquet")

# dates_long <- fhs_dates_long[,dates_long_fields]

rm(list=ls(pattern="\\bfhs1."))
rm(list=ls(pattern="\\bfhs2."))
rm(list=ls(pattern="\\bfhs3."))


# gcs_auth(email="")
gcs_auth("~/Dropbox/ADAPT-HF/Master HDCP files/harmonization-286013-39f492122f69.json")

gcs_upload(fhs_melt_all[,..data_fields], 
           bucket="master_hdcp_files",
           name="fhs_melt_all.parquet",
           object_function = f)


rm(list=ls(pattern="\\bfhs."))
rm(offspring_30,
   offspring_35,
   omni1_10,
   omni1_5)


gc()
```
