## Supplementary material for "Aggregating data to accelerate personalized therapy in heart failure (ADAPT-HF)": Harmonization scripts: Compile JHS.R

```
library(sqldf)
library(Hmisc)
library(plotrix)
library(descr)
library(dplyr)
library(relper)
library(stringr)
library(bigrquery)
library(haven)
library(googleCloudStorageR)
library(data.table)
library(arrow)
library(parquetize)
library(bigQueryR)

source("~/Dropbox/R scripts/Misc clinical scripts.R")
source("~/Dropbox/R scripts/echo scripts.R")

f <- function(input,output) {
  write.csv(input,file=output, row.names=F, na="")
}

visit_yrs <- fread('~/Dropbox/ADAPT-HF/Master HDCP files/Cohort visit yrs.csv')

data_fields <- 
  c("study",
    "patientid",
    "visit_yr",
    "visitdays",
    "form",
    "variable",
    "value",
    "datapoint",
    "study_field",
    "cohort",
    "cohort_name",
    "age_obs")

outcomes_list <- c("dth_status","dth_dt",
                   "cvdth_status","cvdth_dt",
                   "hfdth_status","hfdth_dt",
                   "hfhosp_ef_val","hfhosp_ef_cat",
                   "noncvdth_status","noncvdth_dt",
                   "hosp_status","hosp_dt",
                   "cadhosp_status","cadhosp_dt",
                   "cvhosp_status","cvhosp_dt",
                   "hfhosp_status","hfhosp_dt",
                   "noncvhosp_status","noncvhosp_dt")

dates_long_fields <-
  c("study",
    "cohort",
    "cohort_name",
    "patientid",
    "sex",
    "visit_yr",
    "visitdays",
    "age_obs")


# .......%%.%%.....%%..%%%%%%.
# .......%%.%%.....%%.%%....%%
# .......%%.%%.....%%.%%......
# .......%%.%%%%%%%%%..%%%%%%.
# .%%....%%.%%.....%%.......%%
# .%%....%%.%%.....%%.%%....%%
# ..%%%%%%..%%.....%%..%%%%%%.


#================================================================#
###                     JHS dates tables                       ###
#================================================================#

jhs_analysis1 <- fread("~/Dropbox/BioLINCC files/JHS/Data/Analysis_Data/CSV/analysis1.csv",na.strings=c("","NA","NULL"))
jhs_analysis2 <- fread("~/Dropbox/BioLINCC files/JHS/Data/Analysis_Data/CSV/analysis2.csv",na.strings=c("","NA","NULL"))
jhs_analysis3 <- fread("~/Dropbox/BioLINCC files/JHS/Data/Analysis_Data/CSV/analysis3.csv",na.strings=c("","NA","NULL"))

jhs_v2date <- sqldf('select newid,DaysFromV1 as DATE6 from jhs_analysis2')
jhs_v3date <- sqldf('select newid, DaysFromV1 as DATE9 from jhs_analysis3')


jhs_hfdates <- fread("~/Dropbox/BioLINCC files/JHS/Data/Events/CSV/incevthfder.csv",na.strings=c("","NA","NULL"))

jhs_analysis1$enroll_yr <- gsub(".*/","",jhs_analysis1$VisitDate)

jhs_dates <- merge(jhs_analysis1[,c("newid","ARIC","age","enroll_yr","sex")],jhs_v2date,by="newid",all.x=T)
jhs_dates <- merge(jhs_dates,jhs_v3date,by="newid",all.x=T)
jhs_dates <- merge(jhs_dates,jhs_hfdates[,c("newid","days","Status","HF")],by="newid",all.x=T)


jhs_dates$v6_age <- jhs_dates$age+floor(jhs_dates$DATE6/365)
jhs_dates$v9_age <- jhs_dates$age+floor(jhs_dates$DATE9/365)
jhs_dates$hf_age <- jhs_dates$age+floor(jhs_dates$days/365)

# jhs_dates$enroll_yr <- as.numeric(jhs_dates$enroll_yr)

names(jhs_dates)[names(jhs_dates)=="newid"] <- "patientid"


## Create a dataset starting at a set age for all patients who survive to that age by adding the observation day to baseline age.
## From there,  choose the most recent value available for each patient/variable pair going all the way back to baseline.
## This will be accomplished by subsetting on age at observation (age_obs) being
## Will use cutoff of age_obs <= specified baseline age to capture baseline observations (visitdays = 0, so age_obs==baseline age)
## Then will subset again to select highest available age for each patient/variable pair (i.e. most recent)
## Need to check on methdologic issues here - should be a sub-analysis until validity/robustness proven.


jhs_dates$cohort <- jhs_dates$ARIC

jhs_dates$cohort_name[jhs_dates$cohort==1] <- "ARIC"
jhs_dates$cohort_name[jhs_dates$cohort==0] <- "Not ARIC"


# ==============================================================##
####                       JHS Outcomes                       ####
# ==============================================================##

jhs_allevtchd <- fread("~/Dropbox/BioLINCC files/JHS/Data/Events/csv/allevtchd.csv")
jhs_allevthf <- fread("~/Dropbox/BioLINCC files/JHS/Data/Events/csv/allevthf.csv")
jhs_allevtstroke <- fread("~/Dropbox/BioLINCC files/JHS/Data/Events/csv/allevtstroke.csv")
jhs_incevtchd <- fread("~/Dropbox/BioLINCC files/JHS/Data/Events/csv/incevtchd.csv")
jhs_incevthfder <- fread("~/Dropbox/BioLINCC files/JHS/Data/Events/csv/incevthfder.csv")
jhs_incevtstroke <- fread("~/Dropbox/BioLINCC files/JHS/Data/Events/csv/incevtstroke.csv")

jhs_allevtchd <- merge(jhs_allevtchd,
                       jhs_incevtchd[,c("newid","V1date")],
                       by="newid")

jhs_allevtchd$days_to_event <- as.numeric(as.Date(jhs_allevtchd$eventDate, format="%m/%d/%Y") - 
                                            as.Date(jhs_allevtchd$V1date, 
                                                    format="%m/%d/%Y"))

jhs_incevthfder$HF[jhs_incevthfder$examdate==""] <- 0
jhs_incevthfder$days[jhs_incevthfder$examdate==""] <- 0
jhs_incevthfder$examdate[jhs_incevthfder$examdate==""] <- 
  jhs_incevthfder$V1date[jhs_incevthfder$examdate==""]

jhs_allevthf <- merge(jhs_allevthf,
                      jhs_incevthfder[,c("newid","examdate")],
                      by="newid")

jhs_allevthf$days_to_event <- as.numeric(as.Date(jhs_allevthf$eventDate, format="%m/%d/%Y") - 
                                           as.Date(jhs_allevthf$examdate, format="%m/%d/%Y"))

jhs_allevtstroke <- merge(jhs_allevtstroke,
                          jhs_incevthfder[,c("newid","V1date")],
                          by="newid")

jhs_allevtstroke$days_to_event <- as.numeric(as.Date(jhs_allevtstroke$eventdate, format="%m/%d/%Y") - 
                                               as.Date(jhs_allevtstroke$V1date, format="%m/%d/%Y"))

jhs_outcomes_hf <- sqldf("select newid,
                      days as hfhosp_dt,
                      HF as hfhosp_status
                      from jhs_incevthfder")

jhs_outcomes_cad <- sqldf("select newid,
                      days as cadhosp_dt,
                      CHD as cadhosp_status
                      from jhs_incevtchd")

jhs_outcomes <- merge(jhs_outcomes_hf,
                      jhs_outcomes_cad,
                      by="newid",
                      all.x=T)


jhs_outcomes$study <- "JHS"


names(jhs_outcomes)[1] <- "patientid"
outcomes_to_fill <- outcomes_list[!outcomes_list %in% names(jhs_outcomes)]
outcomes_to_fill <- outcomes_to_fill[!is.na(outcomes_to_fill)]
jhs_outcomes[,outcomes_to_fill] <- NA


outcomes_list_2 <- c("patientid","study",outcomes_list)

write_parquet(jhs_outcomes,
              "~/Dropbox/ADAPT-HF/Master HDCP files/Cohort data/jhs_outcomes.parquet")


#===============================================================#
####                  *** JHS Visit 1 ***                    ####
#===============================================================#


#### Analysis ### 

jhs_analysis1$HOMA_B[jhs_analysis1$HOMA_B < 0] <- NA
jhs_analysis1$fev1_fvc <- jhs_analysis1$FEV1/jhs_analysis1$FVC
jhs_analysis1$aldo_renin <- jhs_analysis1$ALDOSTERONE/jhs_analysis1$reninRIA

jhs_melt_analysis1 <- melt(jhs_analysis1,c(id.vars=c("newid","visit")),na.rm=T,stringsAsFactor=T)
jhs_melt_analysis1$visitdays <- 0
jhs_melt_analysis1$visit_yr <- 0
jhs_melt_analysis1$form <- "analysis1"


#### Alcohol + drug use (ADRA) ---#

jhs_adra <- fread("~/Dropbox/BioLINCC files/JHS/Data/Visit1/CSV/adra.csv",na.strings=c("","NA","NULL"))
jhs_adra$drinks_days_per_week[jhs_adra$ADRA2B=="M"&!is.na(jhs_adra$ADRA2B)]  <- jhs_adra$ADRA2A[jhs_adra$ADRA2B=="M"&!is.na(jhs_adra$ADRA2B)]*4.333
jhs_adra$drinks_days_per_week[jhs_adra$ADRA2B=="Y"&!is.na(jhs_adra$ADRA2B)]  <- jhs_adra$ADRA2A[jhs_adra$ADRA2B=="Y"&!is.na(jhs_adra$ADRA2B)]*52
jhs_adra$drink_days_per_week[jhs_adra$ADRA2B=="W"&!is.na(jhs_adra$ADRA2B)]  <- jhs_adra$ADRA2A[jhs_adra$ADRA2B=="W"&!is.na(jhs_adra$ADRA2B)]
jhs_adra$drinks_week <- jhs_adra$drink_days_per_week * jhs_adra$ADRA3
jhs_adra$visitdays <- 0

jhs_melt_adra_v1 <- melt(jhs_adra,c(id.vars=c("newid","VISIT","visitdays")),na.rm=T,stringsAsFactor=T)
jhs_melt_adra_v1$form <- "adra"


### Central lab results (CENA) ---#

jhs_cena_v1 <- fread("~/Dropbox/BioLINCC files/JHS/Data/Visit1/CSV/cena.csv",na.strings=c("","NA","NULL"))
jhs_cena_v1$visitdays <- 0

jhs_melt_cena_v1 <- melt(jhs_cena_v1,
                         c(id.vars=c("newid","VISIT","visitdays")),
                         na.rm=T,
                         stringsAsFactor=T)
jhs_melt_cena_v1$form <- "cena_v1"


### CESD (CESA) ####


### ISL (ISLA) ####


#### Echo data (ECHA) ---#

jhs_echa_v1 <- fread("~/Dropbox/BioLINCC files/JHS/Data/Visit1/CSV/echa.csv",
                     na.strings=c("","NA","NULL"))
jhs_echa_v1$visitdays <- 0
jhs_echa_v1$lvmass_ix <- jhs_echa_v1$ECHA58/jhs_echa_v1$ECHA13
jhs_echa_v1$rwt <- 2*jhs_echa_v1$ECHA54/jhs_echa_v1$ECHA52
jhs_echa_v1 <- merge(jhs_echa_v1,jhs_analysis1[,c("newid","sex")],all.x=T)

jhs_echa_v1[,ECHA41 := ECHA41/10]
jhs_echa_v1[,ECHA43 := ECHA43/10]
jhs_echa_v1[,ECHA44 := ECHA44/10]
jhs_echa_v1[,ECHA45 := ECHA45/10]
jhs_echa_v1[,ECHA47 := ECHA47/10]
jhs_echa_v1[,ECHA48 := ECHA48/10]
jhs_echa_v1[,ECHA50 := ECHA50/10]
jhs_echa_v1[,ECHA52 := ECHA52/10]
jhs_echa_v1[,ECHA53 := ECHA53/10]
jhs_echa_v1[,ECHA54 := ECHA54/10]
jhs_echa_v1[,ECHA56 := ECHA56/10]
jhs_echa_v1[,ECHA57 := ECHA57/10]


jhs_echa_v1$lvh_type[jhs_echa_v1$sex=="Male"&
                       jhs_echa_v1$lvmass_ix>115&
                       jhs_echa_v1$rwt<=0.42&
                       !is.na(jhs_echa_v1$lvmass_ix)&
                       !is.na(jhs_echa_v1$rwt)] <- "Eccentric hypertrophy"

jhs_echa_v1$lvh_type[jhs_echa_v1$sex=="Female"&
                       jhs_echa_v1$lvmass_ix>95&
                       jhs_echa_v1$rwt<=0.42&
                       !is.na(jhs_echa_v1$lvmass_ix)&
                       !is.na(jhs_echa_v1$rwt)] <- "Eccentric hypertrophy"

jhs_echa_v1$lvh_type[jhs_echa_v1$sex=="Male"&
                       jhs_echa_v1$lvmass_ix>115&
                       jhs_echa_v1$rwt>0.42&
                       !is.na(jhs_echa_v1$lvmass_ix)&
                       !is.na(jhs_echa_v1$rwt)] <- "Concentric hypertrophy"

jhs_echa_v1$lvh_type[jhs_echa_v1$sex=="Female"&
                       jhs_echa_v1$lvmass_ix>95&
                       jhs_echa_v1$rwt>0.42&
                       !is.na(jhs_echa_v1$lvmass_ix)&
                       !is.na(jhs_echa_v1$rwt)] <- "Concentric hypertrophy"

jhs_echa_v1$lvh_type[jhs_echa_v1$sex=="Male"&
                       jhs_echa_v1$lvmass_ix<=115&
                       jhs_echa_v1$rwt<=0.42&
                       !is.na(jhs_echa_v1$lvmass_ix)&
                       !is.na(jhs_echa_v1$rwt)] <- "Normal geometry"

jhs_echa_v1$lvh_type[jhs_echa_v1$sex=="Female"&
                       jhs_echa_v1$lvmass_ix<=95&
                       jhs_echa_v1$rwt<=0.42&
                       !is.na(jhs_echa_v1$lvmass_ix)&
                       !is.na(jhs_echa_v1$rwt)] <- "Normal geometry"

jhs_echa_v1$lvh_type[jhs_echa_v1$sex=="Male"&
                       jhs_echa_v1$lvmass_ix<=115&
                       jhs_echa_v1$rwt>0.42&
                       !is.na(jhs_echa_v1$lvmass_ix)&
                       !is.na(jhs_echa_v1$rwt)] <- "Concentric remodeling"

jhs_echa_v1$lvh_type[jhs_echa_v1$sex=="Female"&
                       jhs_echa_v1$lvmass_ix<=95&
                       jhs_echa_v1$rwt>0.42&
                       !is.na(jhs_echa_v1$lvmass_ix)&
                       !is.na(jhs_echa_v1$rwt)] <- "Concentric remodeling"

jhs_melt_echa_v1 <- melt(jhs_echa_v1,c(id.vars=c("newid","VISIT","visitdays")),
                         na.rm=T,
                         stringsAsFactor=T)
jhs_melt_echa_v1$form <- "echa_v1"


#### ECGa ---#

jhs_ecga_adj_v1 <- fread("~/Dropbox/BioLINCC files/JHS/Data/Visit1/CSV/ecga_adj.csv",na.strings=c("","NA","NULL"))
jhs_ecga_adj_v1$visitdays <- 0

jhs_melt_ecga_adj_v1 <- melt(jhs_ecga_adj_v1,c(id.vars=c("newid","VISIT","visitdays")),na.rm=T,stringsAsFactor=T)
jhs_melt_ecga_adj_v1$form <- "ecga_adj_v1"


#### Local lab results (LOCA) ---#

jhs_loca_v1 <- fread("~/Dropbox/BioLINCC files/JHS/Data/Visit1/CSV/loca.csv",na.strings=c("","NA","NULL"))
jhs_loca_v1$visitdays <- 0

jhs_melt_loca_v1 <- melt(jhs_loca_v1,c(id.vars=c("newid","VISIT","visitdays")),na.rm=T,stringsAsFactor=T)
jhs_melt_loca_v1$form <- "loca_v1"


#### Medical History (MHXA) ---#

jhs_mhxa_v1 <- fread("~/Dropbox/BioLINCC files/JHS/Data/Visit1/CSV/mhxa.csv",na.strings=c("","NA","NULL"))
jhs_mhxa_v1$visitdays <- 0

jhs_melt_mhxa_v1 <- melt(jhs_mhxa_v1,c(id.vars=c("newid","VISIT","visitdays")),na.rm=T,stringsAsFactor=T)
jhs_melt_mhxa_v1$form <- "mhxa_v1"


#### Medications (MSRA) ---#

jhs_msra_v1 <- fread("~/Dropbox/BioLINCC files/JHS/Data/Visit1/CSV/msra.csv",na.strings=c("","NA","NULL"))
jhs_msra_v1$visitdays <- 0

jhs_msra_v1$unintent_nonadhere[
  jhs_msra_v1$MSRA31A==1|
    jhs_msra_v1$MSRA31J==1] <- "Yes"

jhs_msra_v1$unintent_nonadhere[
  jhs_msra_v1$MSRA31A==2&
    jhs_msra_v1$MSRA31J==2] <- "No"

jhs_msra_v1$intent_nonadhere[
  jhs_msra_v1$MSRA31B==1|
    jhs_msra_v1$MSRA31C==1|
    jhs_msra_v1$MSRA31D==1|
    jhs_msra_v1$MSRA31E==1|
    jhs_msra_v1$MSRA31F==1|
    jhs_msra_v1$MSRA31G==1|
    jhs_msra_v1$MSRA31H==1|
    jhs_msra_v1$MSRA31K==1] <- "Yes"

jhs_msra_v1$intent_nonadhere[
  jhs_msra_v1$MSRA31B==2&
    jhs_msra_v1$MSRA31C==2&
    jhs_msra_v1$MSRA31D==2&
    jhs_msra_v1$MSRA31E==2&
    jhs_msra_v1$MSRA31F==2&
    jhs_msra_v1$MSRA31G==2&
    jhs_msra_v1$MSRA31H==2&
    jhs_msra_v1$MSRA31K==2] <- "No"

jhs_msra_v1$med_nonadhere[
  jhs_msra_v1$intent_nonadhere=="No"&
    jhs_msra_v1$unintent_nonadhere=="No"] <- "No"

jhs_msra_v1$med_nonadhere[
  jhs_msra_v1$intent_nonadhere=="Yes"&
    jhs_msra_v1$unintent_nonadhere=="No"] <- "Intentional"

jhs_msra_v1$med_nonadhere[
  jhs_msra_v1$intent_nonadhere=="No"&
    jhs_msra_v1$unintent_nonadhere=="Yes"] <- "Unintentional"

jhs_msra_v1$med_nonadhere[
  jhs_msra_v1$intent_nonadhere=="Yes"&
    jhs_msra_v1$unintent_nonadhere=="Yes"] <- "Both"


jhs_melt_msra_v1 <- melt(jhs_msra_v1,c(id.vars=c("newid","VISIT","visitdays")),na.rm=T,stringsAsFactor=T)
jhs_melt_msra_v1$form <- "msra_v1"


#### Occupation (occcode_dv) ---#

jhs_occode_v1  <- fread("~/Dropbox/BioLINCC files/JHS/Data/Visit1/CSV/occode_dv.csv",na.strings=c("","NA","NULL"))
jhs_occode_v1$visitdays <- 0
jhs_occode_v1$VISIT <- 1

jhs_melt_occode_v1 <- melt(jhs_occode_v1,c(id.vars=c("newid","VISIT","visitdays")),
                           na.rm=T,
                           stringsAsFactor=T)
jhs_melt_occode_v1$form <- "occode_v1"


#### Physical activity (PACA) ---#

# ARIC assessed physical activity using a questionnaire developed by Baecke et al. (4). 
# Baecke et al. (4) defined three semicontinuous indices ranging from 1 (low) to 5 (high) for physical activity at work, in sports, and during leisure time. 
# The eight questions related to work ask about the participant's main occupation; 
#     1. Work's vigor compared with others of same age) (PACA9)
#     2. Frequency of following at work:
# a) Sitting, (PACA11A)
# b) Standing (PACA11B)
# c) Walking  (PACA11C)
# d) Lifting  (PACA11D)
# e) Sweating (PACA11E) 
#     3. Frequency of fatigue after work (PACA10)
# 
# The sports score is a function of 
#     1) Frequency, duration, and an assigned intensity of the reported sports  
# a) PACA21B,22,23
# b) PACA24B,25,26 
# c) PACA27B,28,29
#     2) Three additional questions on 
#           a) Frequency of sweating, (PACA5) 
#           b) General frequency of playing sports (PACA20)
#           c) Amount of leisure time physical activity compared with others of the same age. (PACA30)
# 
# The four leisure questions ask about frequency of
#     1) Watching television (PACA6)
#     2) Walking (PACA3)
#     3) Bicycling (PACA4)
#     4) Minutes walking/biking to work or shopping. (PACA1)
# 
# The ARIC investigators made some minor modifications to the original questionnaire. 
#     1) Interviewers administered the questionnaire, and they specified the time reference as the “past year.” 
#     2) ARIC coded occupations using the Labor Department's Dictionary of Occupational Titles; two exercise physiology research assistants with help from an industrial hygienist coded occupational activity level as low, medium, or high. 
#        We created two work indices: one that excluded respondents who answered they did not work, and one that assigned the lowest value to the work index to these non-working respondents. 
#     3) Third, captured up to four sports or types of exercise (instead of two) in descending order of frequency of participation. 
#        The exercise physiology research assistants assigned intensity codes to the sports/exercises based on standard references. 
#     4) Fourth, we multiplied the“simple sports score” (I9) by 1.25, because the original reference (4) had typographically omitted this factor.
# 
# 
# Excerpt From
# Physical activity and incidence of coronary heart disease in middle-aged women and men
# FOLSOM, AARON R.; ARNETT, DONNA K.; HUTCHINSON, RICHARD G.; LIAO, FANGZI; CLEGG, LIMIN X.; COOPER, LAWTON S.


LETTER_position <- function(let) {
  LETTER <- toupper(let)
  position <- match(LETTER, LETTERS)
  return(position)
}


jhs_paca_v1  <- fread("~/Dropbox/BioLINCC files/JHS/Data/Visit1/CSV/paca.csv",
                      na.strings=c("","NA","NULL"))
jhs_paca_v1$visitdays <- 0


### Work index

jhs_paca_v1$aric_pai_work_heavier <- LETTER_position(jhs_paca_v1$PACA9)
jhs_paca_v1$aric_pai_work_tired <- LETTER_position(jhs_paca_v1$PACA10)
jhs_paca_v1$aric_pai_sit <- LETTER_position(jhs_paca_v1$PACA11A)
jhs_paca_v1$aric_pai_stand <- LETTER_position(jhs_paca_v1$PACA11B)
jhs_paca_v1$aric_pai_walk <- LETTER_position(jhs_paca_v1$PACA11C)
jhs_paca_v1$aric_pai_heavy_load <- LETTER_position(jhs_paca_v1$PACA11D)
jhs_paca_v1$aric_pai_work_sweat <- LETTER_position((jhs_paca_v1$PACA11E))

jhs_paca_v1$work_index <-
  jhs_paca_v1$aric_pai_work_tired+
  jhs_paca_v1$aric_pai_sit+
  jhs_paca_v1$aric_pai_stand+
  jhs_paca_v1$aric_pai_walk+
  jhs_paca_v1$aric_pai_heavy_load+
  jhs_paca_v1$aric_pai_work_sweat+
  jhs_paca_v1$aric_pai_work_heavier


## Leisure index

jhs_paca_v1$aric_pai_leisure_walkrun <- LETTER_position(jhs_paca_v1$PACA1)
jhs_paca_v1$aric_pai_leisure_walk <- LETTER_position(jhs_paca_v1$PACA3)
jhs_paca_v1$aric_pai_leisure_cycle <- LETTER_position(jhs_paca_v1$PACA4)
jhs_paca_v1$aric_pai_leisure_tv <- LETTER_position(jhs_paca_v1$PACA6)

jhs_paca_v1$active_index <-
  jhs_paca_v1$aric_pai_leisure_walkrun+
  jhs_paca_v1$aric_pai_leisure_walk+
  jhs_paca_v1$aric_pai_leisure_cycle+
  jhs_paca_v1$aric_pai_leisure_tv

## Sport index

jhs_paca_v1$aric_pai_sport_sweat <- LETTER_position(jhs_paca_v1$PACA5)
jhs_paca_v1$aric_pai_sport_freq <- LETTER_position(jhs_paca_v1$PACA20)
jhs_paca_v1$aric_pai_sport_to_others <- LETTER_position(jhs_paca_v1$PACA30)

### Coefficients for calculating sport values are from JHS Analysis1 Exam Manual

jhs_paca_intensity <- fread("~/Dropbox/BioLINCC files/JHS/Data/Visit1/CSV/sport intensity.csv")

jhs_sport_time_coefs <-
  data.frame(let=c("A","B","C","D","E"),
             prop_yr=c(0.5,1.3,2.5,3.5,4.5),
             time_wk=c(0.04,0.17,0.42,0.67,0.92))

jhs_paca_v1 <- 
  merge(jhs_paca_v1,
        jhs_paca_intensity,
        by.x = "PACA21B",
        by.y="code",
        all.x=T)

names(jhs_paca_v1)[ncol(jhs_paca_v1)-2] <- "sport1"
names(jhs_paca_v1)[ncol(jhs_paca_v1)-1] <- "intensity_label1"
names(jhs_paca_v1)[ncol(jhs_paca_v1)] <- "intensity_num1"

jhs_paca_v1 <- 
  merge(jhs_paca_v1,
        jhs_sport_time_coefs[,c("let","prop_yr")],
        by.x="PACA22",
        by.y="let",
        all.x=T)

jhs_paca_v1 <- 
  merge(jhs_paca_v1,
        jhs_sport_time_coefs[,c("let","time_wk")],
        by.x="PACA23",
        by.y="let",
        all.x=T)

names(jhs_paca_v1)[ncol(jhs_paca_v1)-1] <- "prop_yr1"
names(jhs_paca_v1)[ncol(jhs_paca_v1)] <- "time_wk1"

jhs_paca_v1$aric_pai_sport1 <- 
  jhs_paca_v1$intensity_num1*
  jhs_paca_v1$prop_yr1*
  jhs_paca_v1$time_wk1

# Sport 2


jhs_paca_v1 <- 
  merge(jhs_paca_v1,
        jhs_paca_intensity,
        by.x = "PACA24B",
        by.y="code",
        all.x=T)

names(jhs_paca_v1)[ncol(jhs_paca_v1)-2] <- "sport2"
names(jhs_paca_v1)[ncol(jhs_paca_v1)-1] <- "intensity_label2"
names(jhs_paca_v1)[ncol(jhs_paca_v1)] <- "intensity_num2"

jhs_paca_v1 <- 
  merge(jhs_paca_v1,
        jhs_sport_time_coefs[,c("let","prop_yr")],
        by.x="PACA25",
        by.y="let",
        all.x=T)

jhs_paca_v1 <- 
  merge(jhs_paca_v1,
        jhs_sport_time_coefs[,c("let","time_wk")],
        by.x="PACA26",
        by.y="let",
        all.x=T)

names(jhs_paca_v1)[ncol(jhs_paca_v1)-1] <- "prop_yr2"
names(jhs_paca_v1)[ncol(jhs_paca_v1)] <- "time_wk2"

jhs_paca_v1$aric_pai_sport2 <- 
  jhs_paca_v1$intensity_num2*
  jhs_paca_v1$prop_yr2*
  jhs_paca_v1$time_wk2


# Sport 3


jhs_paca_v1 <- 
  merge(jhs_paca_v1,
        jhs_paca_intensity,
        by.x = "PACA27B",
        by.y="code",
        all.x=T)

names(jhs_paca_v1)[ncol(jhs_paca_v1)-2] <- "sport3"
names(jhs_paca_v1)[ncol(jhs_paca_v1)-1] <- "intensity_label3"
names(jhs_paca_v1)[ncol(jhs_paca_v1)] <- "intensity_num3"

jhs_paca_v1 <- 
  merge(jhs_paca_v1,
        jhs_sport_time_coefs[,c("let","prop_yr")],
        by.x="PACA28",
        by.y="let",
        all.x=T)

jhs_paca_v1 <- 
  merge(jhs_paca_v1,
        jhs_sport_time_coefs[,c("let","time_wk")],
        by.x="PACA29",
        by.y="let",
        all.x=T)

names(jhs_paca_v1)[ncol(jhs_paca_v1)-1] <- "prop_yr3"
names(jhs_paca_v1)[ncol(jhs_paca_v1)] <- "time_wk3"

jhs_paca_v1$aric_pai_sport3 <- 
  jhs_paca_v1$intensity_num3*
  jhs_paca_v1$prop_yr3*
  jhs_paca_v1$time_wk3


jhs_paca_v1$sport_index <-
  jhs_paca_v1$aric_pai_sport1+
  jhs_paca_v1$aric_pai_sport2+
  jhs_paca_v1$aric_pai_sport3+
  jhs_paca_v1$aric_pai_sport_sweat+
  jhs_paca_v1$aric_pai_sport_freq+ 
  jhs_paca_v1$aric_pai_sport_to_others 


jhs_melt_paca_v1 <- melt(jhs_paca_v1,
                         c(id.vars=c("newid","VISIT","visitdays")),
                         na.rm=T,
                         stringsAsFactor=T)
jhs_melt_paca_v1$form <- "paca_v1"


#### Personal situation/SES (PDSA) ---#

jhs_pdsa_v1  <- fread("~/Dropbox/BioLINCC files/JHS/Data/Visit1/CSV/pdsa.csv",
                      na.strings=c("","NA","NULL"))
jhs_pdsa_v1$visitdays <- 0

jhs_melt_pdsa_v1 <- melt(jhs_pdsa_v1,
                         c(id.vars=c("newid","VISIT","visitdays")),
                         na.rm=T,
                         stringsAsFactor=T)
jhs_melt_pdsa_v1$form <- "pdsa_v1"


#### Personal + Family Hx (PFHA) ---#

jhs_pfha_v1  <- fread("~/Dropbox/BioLINCC files/JHS/Data/Visit1/CSV/pfha.csv",
                      na.strings=c("","NA","NULL"))
jhs_pfha_v1$visitdays <- 0

jhs_pfha_v1$natural_children <- 
  apply(jhs_pfha_v1[,c("PFHA37A","PFHA37D")],
        MARGIN=1,
        FUN=function(x) sum(x, na.rm=T))

jhs_melt_pfha_v1 <- melt(jhs_pfha_v1,c(id.vars=c("newid","VISIT","visitdays")),
                         na.rm=T,
                         stringsAsFactor=T)
jhs_melt_pfha_v1$form <- "pfha_v1"


#### Reproductive hx (RHXA) ---#

jhs_rhxa_v1  <- fread("~/Dropbox/BioLINCC files/JHS/Data/Visit1/CSV/rhxa.csv",
                      na.strings=c("","NA","NULL"))
jhs_rhxa_v1$visitdays <- 0

jhs_rhxa_v1[jhs_rhxa_v1==99] <- 0

jhs_rhxa_v1$RHXA22A[is.na(jhs_rhxa_v1$RHXA22A)] <- 0
jhs_rhxa_v1$RHXA22B[is.na(jhs_rhxa_v1$RHXA22B)] <- 0

jhs_rhxa_v1$hrt_agent1_yrs <-
  jhs_rhxa_v1$RHXA22A +
  jhs_rhxa_v1$RHXA22B/12


jhs_rhxa_v1$RHXA28A[is.na(jhs_rhxa_v1$RHXA28A)] <- 0
jhs_rhxa_v1$RHXA28B[is.na(jhs_rhxa_v1$RHXA28B)] <- 0

jhs_rhxa_v1$hrt_agent2_yrs <-
  jhs_rhxa_v1$RHXA28A +
  jhs_rhxa_v1$RHXA28B/12


jhs_rhxa_v1$RHXA34A[is.na(jhs_rhxa_v1$RHXA34A)] <- 0
jhs_rhxa_v1$RHXA34B[is.na(jhs_rhxa_v1$RHXA34B)] <- 0

jhs_rhxa_v1$hrt_agent3_yrs <-
  jhs_rhxa_v1$RHXA34A +
  jhs_rhxa_v1$RHXA34B/12


jhs_rhxa_v1$RHXA40A[is.na(jhs_rhxa_v1$RHXA40A)] <- 0
jhs_rhxa_v1$RHXA40B[is.na(jhs_rhxa_v1$RHXA40B)] <- 0

jhs_rhxa_v1$hrt_agent4_yrs <-
  jhs_rhxa_v1$RHXA40A +
  jhs_rhxa_v1$RHXA40B/12


jhs_rhxa_v1$hrt_years_sum <-
  apply(jhs_rhxa_v1[,c("hrt_agent1_yrs",
                       "hrt_agent2_yrs",
                       "hrt_agent3_yrs",
                       "hrt_agent4_yrs")],
        MARGIN=1,
        FUN=function(x) sum(x, na.rm=T))

jhs_rhxa_v1$hrt_years_sum[jhs_rhxa_v1$hrt_years_sum==0] <- NA


jhs_rhxa_v1$hrt_start_age_min <- 
  apply(jhs_rhxa_v1[,c(
    "RHXA19",
    "RHXA25",
    "RHXA31",
    "RHXA37")],
    MARGIN = 1,
    FUN = function(x) min(x, na.rm=T))

jhs_rhxa_v1$hrt_stop_age_max <- 
  apply(jhs_rhxa_v1[,c(
    "RHXA21",
    "RHXA27",
    "RHXA33",
    "RHXA39")],
    MARGIN = 1,
    FUN = function(x) max(x, na.rm=T))

jhs_rhxa_v1[jhs_rhxa_v1=="Inf"] <- NA
jhs_rhxa_v1[jhs_rhxa_v1=="-Inf"] <- NA

jhs_rhxa_v1$hrt_years_age <-
  jhs_rhxa_v1$hrt_stop_age_max - jhs_rhxa_v1$hrt_start_age_min


jhs_melt_rhxa_v1 <- melt(jhs_rhxa_v1,c(id.vars=c("newid","VISIT","visitdays")),
                         na.rm=T,
                         stringsAsFactor=T)
jhs_melt_rhxa_v1$form <- "rhxa_v1"


#### Social support (SOCA) ---#

jhs_soca_v1  <- fread("~/Dropbox/BioLINCC files/JHS/Data/Visit1/CSV/soca.csv",na.strings=c("","NA","NULL"))
jhs_soca_v1$visitdays <- 0

jhs_melt_soca_v1 <- melt(jhs_soca_v1,c(id.vars=c("newid","VISIT","visitdays")),
                         na.rm=T,
                         stringsAsFactor=T)
jhs_melt_soca_v1$form <- "soca_v1"


#### Stress (STSA) ---#

jhs_stsa_v1  <- fread("~/Dropbox/BioLINCC files/JHS/Data/Visit1/CSV/stsa.csv",
                      na.strings=c("","NA","NULL"))
jhs_stsa_v1$visitdays <- 0

jhs_melt_stsa_v1 <- melt(jhs_stsa_v1,
                         c(id.vars=c("newid","VISIT","visitdays")),
                         na.rm=T,
                         stringsAsFactor=T)
jhs_melt_stsa_v1$form <- "stsa_v1"


#### Tobacco use (TOBA) ---#

jhs_toba_v1  <- fread("~/Dropbox/BioLINCC files/JHS/Data/Visit1/CSV/toba.csv",na.strings=c("","NA","NULL"))
jhs_toba_v1$visitdays <- 0

jhs_melt_toba_v1 <- melt(jhs_toba_v1,c(id.vars=c("newid","VISIT","visitdays")),na.rm=T,stringsAsFactor=T)
jhs_melt_toba_v1$form <- "toba_v1"


#### Health care access and utilization (HCAA) ---#

jhs_hcaa_v1  <- fread("~/Dropbox/BioLINCC files/JHS/Data/Visit1/CSV/hcaa.csv",na.strings=c("","NA","NULL"))
jhs_hcaa_v1$visitdays <- 0

jhs_melt_hcaa_v1 <- melt(jhs_hcaa_v1,c(id.vars=c("newid","VISIT","visitdays")),na.rm=T,stringsAsFactor=T)
jhs_melt_hcaa_v1$form <- "hcaa_v1"


#### Assemble JHS visit 1  ---#

jhs_melt_others_v1 <- rbind(jhs_melt_adra_v1,
                            jhs_melt_cena_v1,
                            jhs_melt_ecga_adj_v1,
                            jhs_melt_loca_v1,
                            jhs_melt_echa_v1,
                            jhs_melt_mhxa_v1,
                            jhs_melt_msra_v1,
                            jhs_melt_occode_v1,
                            jhs_melt_pdsa_v1,
                            jhs_melt_pfha_v1,
                            jhs_melt_soca_v1,
                            jhs_melt_stsa_v1,
                            jhs_melt_toba_v1,
                            jhs_melt_hcaa_v1,
                            jhs_melt_rhxa_v1)

jhs_melt_others_v1$visit_yr <- 0

names(jhs_melt_others_v1)[2] <- "visit"


# ==============================================================#
####                   *** JHS Visit 2 ***                   ####
# ==============================================================#


#### Analysis ---#


jhs_melt_analysis2 <- melt(jhs_analysis2,c(id.vars=c("newid","visit","DaysFromV1")),na.rm=T,stringsAsFactor=T)
names(jhs_melt_analysis2)[names(jhs_melt_analysis2)=="DaysFromV1"] <- "visitdays"
jhs_melt_analysis2$form <- "analysis2"
jhs_melt_analysis2$visit_yr <- 6


#### Body composition (BCFA) ---# 

jhs_bcfa_v2  <- fread("~/Dropbox/BioLINCC files/JHS/Data/Visit2/CSV/bcfa.csv",na.strings=c("","NA","NULL"))
jhs_bcfa_v2 <- merge(jhs_bcfa_v2,jhs_v2date,by="newid")
jhs_melt_bcfa_v2 <- melt(jhs_bcfa_v2, c(id.vars=c("newid","VISIT","DATE6")), na.rm=T, stringsAsFactor=T)
jhs_melt_bcfa_v2$form <- "bcfa_v2"


#### Central lab results (CENB) ---# 

jhs_cenb_v2  <- fread("~/Dropbox/BioLINCC files/JHS/Data/Visit2/CSV/cenb.csv",na.strings=c("","NA","NULL"))
jhs_cenb_v2 <- merge(jhs_cenb_v2,jhs_v2date,by="newid")
jhs_melt_cenb_v2 <- melt(jhs_cenb_v2, c(id.vars=c("newid","VISIT","DATE6")), na.rm=T, stringsAsFactor=T)
jhs_melt_cenb_v2$form <- "cenb_v2"


#### Medical History form (MHXB) ---# 

jhs_mhxb_v2  <- fread("~/Dropbox/BioLINCC files/JHS/Data/Visit2/CSV/mhxb.csv",na.strings=c("","NA","NULL"))
jhs_mhxb_v2 <- merge(jhs_mhxb_v2,jhs_v2date,by="newid")
jhs_melt_mhxb_v2 <- melt(jhs_mhxb_v2,c(id.vars=c("newid","VISIT","DATE6")),na.rm=T,stringsAsFactor=T)
jhs_melt_mhxb_v2$form <- "mhxb_v2"


#### Health History form (HHXA) ---# 

jhs_hhxa_v2  <- fread("~/Dropbox/BioLINCC files/JHS/Data/Visit2/CSV/hhxa.csv",na.strings=c("","NA","NULL"))
jhs_hhxa_v2 <- merge(jhs_hhxa_v2,jhs_v2date,by="newid")

jhs_hhxa_v2$last_doc_cat <- 
  apply(jhs_hhxa_v2[,c("HHXA25","HHXA26")],
        MARGIN=1,
        FUN=function(x) min(x,na.rm=T))


jhs_melt_hhxa_v2 <- melt(jhs_hhxa_v2,c(id.vars=c("newid","VISIT","DATE6")),na.rm=T,stringsAsFactor=T)
jhs_melt_hhxa_v2$form <- "hhxa_v2"


#### MRIB ---# 

jhs_mrib_v2  <- fread("~/Dropbox/BioLINCC files/JHS/Data/Visit2/CSV/mrib.csv",na.strings=c("","NA","NULL"))
jhs_mrib_v2 <- merge(jhs_mrib_v2,jhs_v2date,by="newid")
jhs_mrib_v2 <- merge(jhs_mrib_v2,jhs_analysis2[,c("newid","bsa")])
jhs_mrib_v2$lvmass_ix <- jhs_mrib_v2$MRIB16/jhs_mrib_v2$bsa
jhs_melt_mrib_v2 <- melt(jhs_mrib_v2,c(id.vars=c("newid","VISIT","DATE6")),na.rm=T,stringsAsFactor=T)
jhs_melt_mrib_v2$form <- "mrib_v2"
jhs_melt_mrib_v2$VISIT <- 2


#### Medication Survey (MSRB) ---# 

jhs_msrb_v2  <- fread("~/Dropbox/BioLINCC files/JHS/Data/Visit2/CSV/msrb.csv",na.strings=c("","NA","NULL"))
jhs_msrb_v2 <- merge(jhs_msrb_v2,jhs_v2date,by="newid")


### JHS nonadherence

jhs_msrb_v2$unintent_nonadhere[
  jhs_msrb_v2$MSRB30A==1|
    jhs_msrb_v2$MSRB30J==1] <- "Yes"

jhs_msrb_v2$unintent_nonadhere[
  jhs_msrb_v2$MSRB30A==2&
    jhs_msrb_v2$MSRB30J==2] <- "No"

jhs_msrb_v2$intent_nonadhere[
  jhs_msrb_v2$MSRB30B==1|
    jhs_msrb_v2$MSRB30C==1|
    jhs_msrb_v2$MSRB30D==1|
    jhs_msrb_v2$MSRB30E==1|
    jhs_msrb_v2$MSRB30F==1|
    jhs_msrb_v2$MSRB30G==1|
    jhs_msrb_v2$MSRB30H==1|
    jhs_msrb_v2$MSRB30K==1] <- "Yes"

jhs_msrb_v2$intent_nonadhere[
  jhs_msrb_v2$MSRB30B==2&
    jhs_msrb_v2$MSRB30C==2&
    jhs_msrb_v2$MSRB30D==2&
    jhs_msrb_v2$MSRB30E==2&
    jhs_msrb_v2$MSRB30F==2&
    jhs_msrb_v2$MSRB30G==2&
    jhs_msrb_v2$MSRB30H==2&
    jhs_msrb_v2$MSRB30K==2] <- "No"

jhs_msrb_v2$med_nonadhere[
  jhs_msrb_v2$intent_nonadhere=="No"&
    jhs_msrb_v2$unintent_nonadhere=="No"] <- "No"

jhs_msrb_v2$med_nonadhere[
  jhs_msrb_v2$intent_nonadhere=="Yes"&
    jhs_msrb_v2$unintent_nonadhere=="No"] <- "Intentional"

jhs_msrb_v2$med_nonadhere[
  jhs_msrb_v2$intent_nonadhere=="No"&
    jhs_msrb_v2$unintent_nonadhere=="Yes"] <- "Unintentional"

jhs_msrb_v2$med_nonadhere[
  jhs_msrb_v2$intent_nonadhere=="Yes"&
    jhs_msrb_v2$unintent_nonadhere=="Yes"] <- "Both"


jhs_melt_msrb_v2 <- melt(jhs_msrb_v2,c(id.vars=c("newid","VISIT","DATE6")),na.rm=T,stringsAsFactor=T)
jhs_melt_msrb_v2$form <- "msrb_v2"


#### Assemble JHS visit 2 ---#

jhs_melt_others_v2 <- rbind(jhs_melt_bcfa_v2,
                            jhs_melt_cenb_v2,
                            jhs_melt_hhxa_v2,
                            jhs_melt_mhxb_v2,
                            jhs_melt_mrib_v2,
                            jhs_melt_msrb_v2)

names(jhs_melt_others_v2)[1:3] <- c("newid","visit","visitdays")

jhs_melt_others_v2$visit_yr <- 6


# ==============================================================#
####                   *** JHS Visit 3 ***                   ####
# ==============================================================#


#### Analysis ---#

jhs_melt_analysis3 <- melt(jhs_analysis3,c(id.vars=c("newid","visit","DaysFromV1")),na.rm=T,stringsAsFactor=T)
names(jhs_melt_analysis3)[names(jhs_melt_analysis3)=="DaysFromV1"] <- "visitdays"
jhs_melt_analysis3$form <- "analysis3"
jhs_melt_analysis3$visit_yr <- 9


#### Body composition (BCFV) ---#

jhs_bcfv_v3  <- fread("~/Dropbox/BioLINCC files/JHS/Data/Visit3/CSV/bcfv.csv",na.strings=c("","NA","NULL"))
jhs_bcfv_v3 <- merge(jhs_bcfv_v3,jhs_v3date,by="newid")
jhs_melt_bcfv_v3 <- melt(jhs_bcfv_v3,
                         c(id.vars=c("newid","VISIT","DATE9")),
                         na.rm=T,
                         stringsAsFactor=T)
jhs_melt_bcfv_v3$form <- "bcfv_v3"


#### Personal + Family Hx (PFHB) ---#

jhs_pfhb_v3  <- fread("~/Dropbox/BioLINCC files/JHS/Data/Visit3/CSV/pfhb.csv",
                      na.strings=c("","NA","NULL"))

jhs_pfhb_v3 <- merge(jhs_pfhb_v3,
                     jhs_v3date,
                     by="newid",
                     all.x=T)

jhs_melt_pfhb_v3 <- melt(jhs_pfhb_v3,c(id.vars=c("newid","VISIT","DATE9")),
                         na.rm=T,
                         stringsAsFactor=T)
jhs_melt_pfhb_v3$form <- "pfhb_v3"


#### Cantral lab (CENC) ---#

jhs_cenc_v3  <- fread("~/Dropbox/BioLINCC files/JHS/Data/Visit3/CSV/cenc.csv",na.strings=c("","NA","NULL"))
jhs_cenc_v3 <- merge(jhs_cenc_v3,jhs_v3date,by="newid")

jhs_cenc_v3 <- merge(jhs_cenc_v3,jhs_analysis3[,c('newid','age','male')],by='newid')
jhs_cenc_v3$gfr <- calc_MDRD4(dat=jhs_cenc_v3,
                              cr='creatr',
                              age='age',
                              race=2,
                              sex='male',
                              male=1,
                              black=2)
jhs_cenc_v3$VISIT <- 3
jhs_melt_cenc_v3 <- melt(jhs_cenc_v3, c(id.vars=c("newid","VISIT","DATE9")), na.rm=T, stringsAsFactor=T)
jhs_melt_cenc_v3$form <- "cenc_v3"


#### ECGb ---#

jhs_ecgb_v3  <- fread("~/Dropbox/BioLINCC files/JHS/Data/Visit3/CSV/ecgb.csv",na.strings=c("","NA","NULL"))
jhs_ecgb_v3 <- merge(jhs_ecgb_v3,jhs_v3date,by="newid")
jhs_melt_ecgb_v3 <- melt(jhs_ecgb_v3,c(id.vars=c("newid","VISIT","DATE9")),na.rm=T, stringsAsFactor=T)
jhs_melt_ecgb_v3$form <- "ecgb_v3"


#### HCTA ---#

jhs_hcta_v3  <- fread("~/Dropbox/BioLINCC files/JHS/Data/Visit3/CSV/hcta.csv",na.strings=c("","NA","NULL"))
jhs_hcta_v3 <- merge(jhs_hcta_v3,jhs_v3date,by="newid")
jhs_melt_hcta_v3 <- melt(jhs_hcta_v3,c(id.vars=c("newid","VISIT","DATE9")),na.rm=T, stringsAsFactor=T)
jhs_melt_hcta_v3$form <- "hcta_v3"


#### MHXC ---#

jhs_mhxc_v3  <- fread("~/Dropbox/BioLINCC files/JHS/Data/Visit3/CSV/mhxc.csv",na.strings=c("","NA","NULL"))
jhs_mhxc_v3 <- merge(jhs_mhxc_v3,jhs_v3date,by="newid")
jhs_melt_mhxc_v3 <- melt(jhs_mhxc_v3, c(id.vars=c("newid","VISIT","DATE9")), na.rm=T, stringsAsFactor=T)
jhs_melt_mhxc_v3$form <- "mhxc_v3"


#### MRIB ---#

jhs_mrib_v3  <- fread("~/Dropbox/BioLINCC files/JHS/Data/Visit3/CSV/mrib.csv",na.strings=c("","NA","NULL"))
jhs_mrib_v3 <- merge(jhs_mrib_v3,jhs_v3date,by="newid")
jhs_mrib_v3 <- merge(jhs_mrib_v3,jhs_analysis3[,c("newid","bsa")])
jhs_mrib_v3$lvmass_ix <- jhs_mrib_v3$MRIB16/jhs_mrib_v3$bsa
jhs_melt_mrib_v3 <- melt(jhs_mrib_v3, c(id.vars=c("newid","VISIT","DATE9")), na.rm=T,                          stringsAsFactor=T)
jhs_melt_mrib_v3$form <- "mrib_v3"


#### Medication Survey (MSRC) ---#

jhs_msrc_v3  <- fread("~/Dropbox/BioLINCC files/JHS/Data/Visit3/CSV/msrc.csv",na.strings=c("","NA","NULL"))
jhs_msrc_v3 <- merge(jhs_msrc_v3,jhs_v3date,by="newid")


### JHS nonadherence

jhs_msrc_v3$unintent_nonadhere[
  jhs_msrc_v3$MSRC30A==1|
    jhs_msrc_v3$MSRC30J==1] <- "Yes"

jhs_msrc_v3$unintent_nonadhere[
  jhs_msrc_v3$MSRC30A==2&
    jhs_msrc_v3$MSRC30J==2] <- "No"

jhs_msrc_v3$intent_nonadhere[
  jhs_msrc_v3$MSRC30B==1|
    jhs_msrc_v3$MSRC30C==1|
    jhs_msrc_v3$MSRC30D==1|
    jhs_msrc_v3$MSRC30E==1|
    jhs_msrc_v3$MSRC30F==1|
    jhs_msrc_v3$MSRC30G==1|
    jhs_msrc_v3$MSRC30H==1|
    jhs_msrc_v3$MSRC30K==1] <- "Yes"

jhs_msrc_v3$intent_nonadhere[
  jhs_msrc_v3$MSRC30B==2&
    jhs_msrc_v3$MSRC30C==2&
    jhs_msrc_v3$MSRC30D==2&
    jhs_msrc_v3$MSRC30E==2&
    jhs_msrc_v3$MSRC30F==2&
    jhs_msrc_v3$MSRC30G==2&
    jhs_msrc_v3$MSRC30H==2&
    jhs_msrc_v3$MSRC30K==2] <- "No"

jhs_msrc_v3$med_nonadhere[
  jhs_msrc_v3$intent_nonadhere=="No"&
    jhs_msrc_v3$unintent_nonadhere=="No"] <- "No"

jhs_msrc_v3$med_nonadhere[
  jhs_msrc_v3$intent_nonadhere=="Yes"&
    jhs_msrc_v3$unintent_nonadhere=="No"] <- "Intentional"

jhs_msrc_v3$med_nonadhere[
  jhs_msrc_v3$intent_nonadhere=="No"&
    jhs_msrc_v3$unintent_nonadhere=="Yes"] <- "Unintentional"

jhs_msrc_v3$med_nonadhere[
  jhs_msrc_v3$intent_nonadhere=="Yes"&
    jhs_msrc_v3$unintent_nonadhere=="Yes"] <- "Both"

### Morisky score

jhs_msrc_v3$MSRC31A[jhs_msrc_v3$MSRC31A==2] <- 0
jhs_msrc_v3$MSRC31A[jhs_msrc_v3$MSRC31A > 2] <- NA

jhs_msrc_v3$MSRC31B[jhs_msrc_v3$MSRC31B==2] <- 0
jhs_msrc_v3$MSRC31B[jhs_msrc_v3$MSRC31B > 2] <- NA

jhs_msrc_v3$MSRC31C[jhs_msrc_v3$MSRC31C==2] <- 0
jhs_msrc_v3$MSRC31C[jhs_msrc_v3$MSRC31C > 2] <- NA

jhs_msrc_v3$MSRC31D[jhs_msrc_v3$MSRC31D==2] <- 0
jhs_msrc_v3$MSRC31D[jhs_msrc_v3$MSRC31D > 2] <- NA


jhs_msrc_v3$morisky_score <- 
  jhs_msrc_v3$MSRC31A+
  jhs_msrc_v3$MSRC31B+
  jhs_msrc_v3$MSRC31C+
  jhs_msrc_v3$MSRC31D


jhs_melt_msrc_v3 <- melt(jhs_msrc_v3, c(id.vars=c("newid","VISIT","DATE9")), na.rm=T, stringsAsFactor=T)
jhs_melt_msrc_v3$form <- "msrc_v3"

#### Personal data - SES (PDSB) ---#

jhs_pdsb_v3  <- fread("~/Dropbox/BioLINCC files/JHS/Data/Visit3/CSV/pdsb.csv",na.strings=c("","NA","NULL"))
jhs_pdsb_v3 <- merge(jhs_pdsb_v3,jhs_v3date,by="newid")
jhs_melt_pdsb_v3 <- melt(jhs_pdsb_v3, c(id.vars=c("newid","VISIT","DATE9")), na.rm=T, stringsAsFactor=T)
jhs_melt_pdsb_v3$form <- "pdsb_v3"


#### Physical activity (PACB) ---#

jhs_pacb_v3  <- fread("~/Dropbox/BioLINCC files/JHS/Data/Visit3/CSV/pacb.csv",
                      na.strings=c("","NA","NULL"))
jhs_pacb_v3 <- merge(jhs_pacb_v3,jhs_v3date,by="newid")

jhs_pacb_v3$PACB20 <- LETTERS[jhs_pacb_v3$PACB20]
jhs_pacb_v3$PACB21 <- LETTERS[jhs_pacb_v3$PACB21]

jhs_pacb_v3$PACB23 <- LETTERS[jhs_pacb_v3$PACB23]
jhs_pacb_v3$PACB24 <- LETTERS[jhs_pacb_v3$PACB24]

jhs_pacb_v3$PACB26 <- LETTERS[jhs_pacb_v3$PACB26]
jhs_pacb_v3$PACB27 <- LETTERS[jhs_pacb_v3$PACB27]

### Leisure index

jhs_pacb_v3$active_index <-
  (jhs_pacb_v3$PACB1+
     jhs_pacb_v3$PACB2+
     jhs_pacb_v3$PACB3+
     (6-jhs_pacb_v3$PACB5))/4


### Work index

jhs_pacb_v3$work_index <-
  (jhs_pacb_v3$PACB7+
     jhs_pacb_v3$PACB8+
     jhs_pacb_v3$PACB9A+
     jhs_pacb_v3$PACB9B+
     jhs_pacb_v3$PACB9C+
     jhs_pacb_v3$PACB9D+
     jhs_pacb_v3$PACB9E)/8

### Home & yard index

jhs_pacb_v3$hy_index <- 
  (jhs_pacb_v3$PACB10+
     jhs_pacb_v3$PACB11+
     jhs_pacb_v3$PACB12+
     jhs_pacb_v3$PACB13+
     jhs_pacb_v3$PACB14+
     jhs_pacb_v3$PACB15+
     jhs_pacb_v3$PACB16)/7

## Sport index

jhs_pacb_v3$aric_pai_sport_sweat <- LETTER_position(jhs_pacb_v3$PACB4)
jhs_pacb_v3$aric_pai_sport_freq <- LETTER_position(jhs_pacb_v3$PACB18)
jhs_pacb_v3$aric_pai_sport_to_others <- LETTER_position(jhs_pacb_v3$PACB28)

### Coefficients for calculating sport values are from JHS Analysis1 Exam Manual

jhs_sport_time_coefs <-
  data.frame(let=c("A","B","C","D","E"),
             prop_yr=c(0.5,1.3,2.5,3.5,4.5),
             time_wk=c(0.04,0.17,0.42,0.67,0.92))

jhs_pacb_v3 <- 
  merge(jhs_pacb_v3,
        jhs_paca_intensity,
        by.x = "PACB19B",
        by.y="code",
        all.x=T)

names(jhs_pacb_v3)[ncol(jhs_pacb_v3)-2] <- "sport1"
names(jhs_pacb_v3)[ncol(jhs_pacb_v3)-1] <- "intensity_label1"
names(jhs_pacb_v3)[ncol(jhs_pacb_v3)] <- "intensity_num1"

jhs_pacb_v3 <- 
  merge(jhs_pacb_v3,
        jhs_sport_time_coefs[,c("let","prop_yr")],
        by.x="PACB20",
        by.y="let",
        all.x=T)

jhs_pacb_v3 <- 
  merge(jhs_pacb_v3,
        jhs_sport_time_coefs[,c("let","time_wk")],
        by.x="PACB21",
        by.y="let",
        all.x=T)

names(jhs_pacb_v3)[ncol(jhs_pacb_v3)-1] <- "prop_yr1"
names(jhs_pacb_v3)[ncol(jhs_pacb_v3)] <- "time_wk1"

jhs_pacb_v3$aric_pai_sport1 <- 
  jhs_pacb_v3$intensity_num1*
  jhs_pacb_v3$prop_yr1*
  jhs_pacb_v3$time_wk1

# Sport 2


jhs_pacb_v3 <- 
  merge(jhs_pacb_v3,
        jhs_paca_intensity,
        by.x = "PACB22B",
        by.y="code",
        all.x=T)

names(jhs_pacb_v3)[ncol(jhs_pacb_v3)-2] <- "sport2"
names(jhs_pacb_v3)[ncol(jhs_pacb_v3)-1] <- "intensity_label2"
names(jhs_pacb_v3)[ncol(jhs_pacb_v3)] <- "intensity_num2"

jhs_pacb_v3 <- 
  merge(jhs_pacb_v3,
        jhs_sport_time_coefs[,c("let","prop_yr")],
        by.x="PACB23",
        by.y="let",
        all.x=T)

jhs_pacb_v3 <- 
  merge(jhs_pacb_v3,
        jhs_sport_time_coefs[,c("let","time_wk")],
        by.x="PACB24",
        by.y="let",
        all.x=T)

names(jhs_pacb_v3)[ncol(jhs_pacb_v3)-1] <- "prop_yr2"
names(jhs_pacb_v3)[ncol(jhs_pacb_v3)] <- "time_wk2"

jhs_pacb_v3$aric_pai_sport2 <- 
  jhs_pacb_v3$intensity_num2*
  jhs_pacb_v3$prop_yr2*
  jhs_pacb_v3$time_wk2


# Sport 3


jhs_pacb_v3 <- 
  merge(jhs_pacb_v3,
        jhs_paca_intensity,
        by.x = "PACB25B",
        by.y="code",
        all.x=T)

names(jhs_pacb_v3)[ncol(jhs_pacb_v3)-2] <- "sport3"
names(jhs_pacb_v3)[ncol(jhs_pacb_v3)-1] <- "intensity_label3"
names(jhs_pacb_v3)[ncol(jhs_pacb_v3)] <- "intensity_num3"

jhs_pacb_v3 <- 
  merge(jhs_pacb_v3,
        jhs_sport_time_coefs[,c("let","prop_yr")],
        by.x="PACB26",
        by.y="let",
        all.x=T)

jhs_pacb_v3 <- 
  merge(jhs_pacb_v3,
        jhs_sport_time_coefs[,c("let","time_wk")],
        by.x="PACB27",
        by.y="let",
        all.x=T)

names(jhs_pacb_v3)[ncol(jhs_pacb_v3)-1] <- "prop_yr3"
names(jhs_pacb_v3)[ncol(jhs_pacb_v3)] <- "time_wk3"

jhs_pacb_v3$aric_pai_sport3 <- 
  jhs_pacb_v3$intensity_num3*
  jhs_pacb_v3$prop_yr3*
  jhs_pacb_v3$time_wk3


jhs_pacb_v3$sportindex <-
  jhs_pacb_v3$aric_pai_sport1+
  jhs_pacb_v3$aric_pai_sport2+
  jhs_pacb_v3$aric_pai_sport3+
  jhs_pacb_v3$aric_pai_sport_sweat+
  jhs_pacb_v3$aric_pai_sport_freq+ 
  jhs_pacb_v3$aric_pai_sport_to_others 


jhs_melt_pacb_v3 <- melt(jhs_pacb_v3,
                         c(id.vars=c("newid","VISIT","DATE9")),
                         na.rm=T,
                         stringsAsFactor=T)
jhs_melt_pacb_v3$form <- "pacb_v3"


#### Sleep history form (SLEA) ---#

jhs_slea_v3  <- fread("~/Dropbox/BioLINCC files/JHS/Data/Visit3/CSV/slea.csv",na.strings=c("","NA","NULL"))
jhs_slea_v3 <- merge(jhs_slea_v3,jhs_v3date,by="newid")
jhs_melt_slea_v3 <- melt(jhs_slea_v3, c(id.vars=c("newid","VISIT","DATE9")), na.rm=T, stringsAsFactor=T)
jhs_melt_slea_v3$form <- "slea_v3"


#### Tobacco use (TOBB) ---#

jhs_tobb_v3  <- fread("~/Dropbox/BioLINCC files/JHS/Data/Visit3/CSV/tobb.csv",na.strings=c("","NA","NULL"))
jhs_tobb_v3 <- merge(jhs_tobb_v3,jhs_v3date,by="newid")
jhs_melt_tobb_v3 <- melt(jhs_tobb_v3, c(id.vars=c("newid","VISIT","DATE9")),na.rm=T, stringsAsFactor=T)
jhs_melt_tobb_v3$form <- "tobb_v3"

#### Years of menses ---#

jhs_rhxb_v3 <- fread("~/Dropbox/BioLINCC files/JHS/Data/Visit3/CSV/rhxb.csv",na.strings=c("","NA","NULL"))

jhs_rhxb_v3$VISIT <- 9

jhs_rhxb_v3 <-
  merge(jhs_rhxb_v3,
        jhs_rhxa_v1[,c("newid","RHXA1","RHXA9")],
        by="newid",
        all.x=T)

jhs_rhxb_v3$RHXA9[jhs_rhxb_v3$RHXA9 < jhs_rhxb_v3$RHXA1+3] <- NA
jhs_rhxb_v3$RHXB6[jhs_rhxb_v3$RHXB6 < jhs_rhxb_v3$RHXA1+3] <- NA

jhs_rhxb_v3$age_menopause <- jhs_rhxb_v3$RHXB6
jhs_rhxb_v3$age_menopause[is.na(jhs_rhxb_v3$RHXB6)] <-
  jhs_rhxb_v3$RHXA9[is.na(jhs_rhxb_v3$RHXB6)]

jhs_rhxb_v3$years_menses <-
  jhs_rhxb_v3$age_menopause - jhs_rhxb_v3$RHXA1

jhs_rhxb_v3 <- merge(jhs_rhxb_v3,jhs_v3date,by="newid")

jhs_melt_rhxb_v3 <- melt(jhs_rhxb_v3, c(id.vars=c("newid","VISIT","DATE9")),na.rm=T, stringsAsFactor=T)
jhs_melt_rhxb_v3$form <- "rhxb_v3"

#### Assemble JHS visit 3 ---#

jhs_melt_others_v3 <- rbind(
  jhs_melt_bcfv_v3,
  jhs_melt_cenc_v3,
  jhs_melt_ecgb_v3,
  jhs_melt_mhxc_v3,
  jhs_melt_mrib_v3,
  jhs_melt_msrc_v3,
  jhs_melt_pdsb_v3,
  jhs_melt_slea_v3,
  jhs_melt_tobb_v3,
  jhs_melt_rhxb_v3,
  jhs_melt_hcta_v3)

names(jhs_melt_others_v3)[1:3] <- c("newid","visit","visitdays")

jhs_melt_others_v3$visit_yr <- 9


# ==============================================================#
####             *** JHS longitudinal visits ***             ####
# ==============================================================#

## Loading and preparing annual longitudinal visits here
## Currently Visit 1 = baseline, Visit 2  = 6 years, Visit 3 = 9 years.
## afulong visit assigned by taking time since baseline rounded to nearest year (jhs_afulong$time) +1
## 1 added because Baseline visit = Visit 1
## Then remove any afulong record with assigned visit 1,6,or 9 to eliminate clashing with main study visits.


jhs_afulong <- fread("~/Dropbox/BioLINCC files/JHS/Data/AFU/CSV/afulong.csv",na.strings=c("","NA","NULL"))

jhs_afulong$visit_yr <- 
  round(jhs_afulong$time)

jhs_afulong$visitdays <- 
  as.numeric(
    difftime(
      as.Date(jhs_afulong$date,"%m/%d/%Y"),
      as.Date(jhs_afulong$v1date,"%m/%d/%Y"),
      units="days"))

jhs_afulong$visit2days <- 
  as.numeric(
    difftime(
      as.Date(jhs_afulong$date,"%m/%d/%Y"),
      as.Date(jhs_afulong$V2date,"%m/%d/%Y"),
      units="days"))

jhs_afulong$visit2years <- round(jhs_afulong$visit2days/365,0)+1

jhs_afulong$visit3days <- 
  as.numeric(
    difftime(
      as.Date(jhs_afulong$date,"%m/%d/%Y"),
      as.Date(jhs_afulong$V3date,"%m/%d/%Y"),
      units="days"))

jhs_afulong$visit3years <- round(jhs_afulong$visit3days/365,0)+1

jhs_afulong$days_from_v2 <-   
  as.numeric(
    difftime(
      as.Date(jhs_afulong$date,"%m/%d/%Y"),
      as.Date(jhs_afulong$V2date,"%m/%d/%Y"),
      units="days"))

jhs_afulong$days_from_v3 <-   
  as.numeric(
    difftime(
      as.Date(jhs_afulong$date,"%m/%d/%Y"),
      as.Date(jhs_afulong$V3date,"%m/%d/%Y"),
      units="days"))

jhs_afulong <- subset(jhs_afulong,
                      !visit_yr %in% c(-2,-1,0,6,9)&
                        abs(days_from_v2)>180&
                        abs(days_from_v3)&
                        abs(visitdays)>180)

## This statement handles a situation where multiple studies round to the same visit, resulting in multiple separate lines for that visit.
## This solution takes the MAX (or latest) observation - this can be changed to the earliest, etc.

jhs_afulong <- setDT(sqldf("select * 
                      from jhs_afulong
                      group by newid,visit_yr
                      having max(visitdays)"))

jhs_melt_afulong <- melt(jhs_afulong,
                         c(id.vars=c("newid",
                                     "visit_yr",
                                     "visitdays")),
                         na.rm=T,
                         stringsAsFactor=T)

jhs_melt_afulong$form <-"afulong"

#### Parental SES (AF1) ---# 

jhs_af1 <- fread("~/Dropbox/BioLINCC files/JHS/Data/AFU/CSV/af1.csv",
                 na.strings=c("","NA","NULL"))
jhs_melt_af1 <- melt(jhs_af1,
                     c(id.vars=c("newid")),
                     na.rm=T,
                     stringsAsFactor=T)

jhs_melt_af1$visit_yr <- 0
jhs_melt_af1$visitdays <- 0
jhs_melt_af1$form <- "af1"


#### Life Orientation, Job Situation (AF2) ---#

jhs_af2 <- fread("~/Dropbox/BioLINCC files/JHS/Data/AFU/CSV/af2.csv",
                 na.strings=c("","NA","NULL"))

lotr_convert <- data.frame(let=c("A","B","C","D"),
                           num=c(4,3,2,1))

lotr_convert_inverse <-
  data.frame(let=c("A","B","C","D"),
             num=c(1,2,3,4))

jhs_af2 <- merge(jhs_af2,
                 lotr_convert,
                 by.x="AF2v1",
                 by.y="let",
                 all.x=T)

names(jhs_af2)[ncol(jhs_af2)] <- "lotr_1"

jhs_af2 <- merge(jhs_af2,
                 lotr_convert_inverse,
                 by.x="AF2v3",
                 by.y="let",
                 all.x=T)

names(jhs_af2)[ncol(jhs_af2)] <- "lotr_3"


jhs_af2 <- merge(jhs_af2,
                 lotr_convert,
                 by.x="AF2v4",
                 by.y="let",
                 all.x=T)

names(jhs_af2)[ncol(jhs_af2)] <- "lotr_4"


jhs_af2 <- merge(jhs_af2,
                 lotr_convert_inverse,
                 by.x="AF2v7",
                 by.y="let",
                 all.x=T)

names(jhs_af2)[ncol(jhs_af2)] <- "lotr_7"


jhs_af2 <- merge(jhs_af2,
                 lotr_convert_inverse,
                 by.x="AF2v9",
                 by.y="let",
                 all.x=T)

names(jhs_af2)[ncol(jhs_af2)] <- "lotr_9"


jhs_af2 <- merge(jhs_af2,
                 lotr_convert,
                 by.x="AF2v10",
                 by.y="let",
                 all.x=T)

names(jhs_af2)[ncol(jhs_af2)] <- "lotr_10"


jhs_af2$lotr_total <-
  apply(jhs_af2[,c("lotr_1",
                   "lotr_3",
                   "lotr_4",
                   "lotr_7",
                   "lotr_9",
                   "lotr_10")],
        MARGIN=1,
        FUN=function(x) sum(x))


jhs_af2 <- merge(jhs_af2,
                 lotr_convert,
                 by.x="AF2v1",
                 by.y="let",
                 all.x=T)

names(jhs_af2)[ncol(jhs_af2)] <- "lotr_1"


jhs_melt_af2 <- melt(jhs_af2,
                     c(id.vars=c("newid")),
                     na.rm=T,
                     stringsAsFactor=T)

jhs_melt_af2$visit_yr <- 0
jhs_melt_af2$visitdays <- 0
jhs_melt_af2$form <- "af2"


#### Stress & Major events (AF3) ---# 

jhs_af3 <- fread("~/Dropbox/BioLINCC files/JHS/Data/AFU/CSV/af3.csv",
                 na.strings=c("","NA","NULL"))


# jhs_af3[,c("jhac_1",
#            "jhac_2",
#            "jhac_3",
#            "jhac_4",
#            "jhac_5",
#            "jhac_6",
#            "jhac_7",
#            "jhac_8",
#            "jhac_9",
#            "jhac_10",
#            "jhac_11",
#            "jhac_12")] <-
#   apply(jhs_af3[,3:14],
#         MARGIN=2,
#         FUN = function(x) match(x,LETTERS))

jhs_af3[
  ,
  paste0("jhac_", 1:12) := lapply(.SD, function(x) match(x, LETTERS)),
  .SDcols = 3:14
]

jhs_af3[
  ,
  jhac_total := rowSums(.SD),
  .SDcols = paste0("jhac_", 1:12)
]

jhs_melt_af3 <- melt(jhs_af3,
                     c(id.vars=c("newid")),
                     na.rm=T,
                     stringsAsFactor=T)

jhs_melt_af3$visit_yr <- 0
jhs_melt_af3$visitdays <- 0
jhs_melt_af3$form <- "af3"

rm(lotr_convert,
   lotr_convert_inverse)

# ==============================================================--#
####                ** Assemble all JHS tables **              ####
# ==============================================================--#

jhs_melt_all <- rbind(jhs_melt_analysis1[,c("newid","visit_yr","visitdays","variable","value","form")],
                      jhs_melt_analysis2[,c("newid","visit_yr","visitdays","variable","value","form")],
                      jhs_melt_analysis3[,c("newid","visit_yr","visitdays","variable","value","form")],
                      jhs_melt_others_v1[,c("newid","visit_yr","visitdays","variable","value","form")],
                      jhs_melt_others_v2[,c("newid","visit_yr","visitdays","variable","value","form")],
                      jhs_melt_others_v3[,c("newid","visit_yr","visitdays","variable","value","form")],
                      jhs_melt_afulong[,c("newid","visit_yr","visitdays","variable","value","form")],
                      jhs_melt_af1[,c("newid","visit_yr","visitdays","variable","value","form")],
                      jhs_melt_af2[,c("newid","visit_yr","visitdays","variable","value","form")],
                      jhs_melt_af3[,c("newid","visit_yr","visitdays","variable","value","form")])


names(jhs_melt_all)[1] <- "patientid"
jhs_melt_all$study <- "JHS"

jhs_melt_all$form <- toupper(jhs_melt_all$form)
jhs_melt_all$variable <- toupper(jhs_melt_all$variable)
jhs_melt_all$study_field <- paste(jhs_melt_all$study,
                                  jhs_melt_all$form,
                                  jhs_melt_all$variable,sep="_")

##  Add age at observation for assembling cohorts of same starting age

jhs_melt_all <- merge(jhs_melt_all,jhs_dates[,c("patientid","enroll_yr","age")], by="patientid")

names(jhs_melt_all)[ncol(jhs_melt_all)] <- "baseline_age"

jhs_melt_all$age_obs <- floor(jhs_melt_all$baseline_age+jhs_melt_all$visitdays/365)
# jhs_melt_all$year_obs <- round(jhs_melt_all$enroll_yr+jhs_melt_all$visitdays/365)


jhs_dates_long <- sqldf("select distinct
                        patientid,
                        age_obs,
                        visitdays,
                        visit_yr,
                        study
                        from jhs_melt_all")


jhs_dates_long <- merge(jhs_dates_long,
                        jhs_dates[,c("patientid","cohort","cohort_name","sex")],
                        by="patientid")

jhs_melt_all <- merge(jhs_melt_all,
                      jhs_dates[,c("patientid","cohort","cohort_name")],
                      by="patientid",
                      all.x=T)

# jhs_melt_all <- subset(jhs_melt_all,study_field %in% hdcp_cohort_fields_used$study_field)

jhs_melt_all$datapoint <-
  paste("JHS",row.names(jhs_melt_all),sep="")

jhs_melt_all$patientid <- as.character(jhs_melt_all$patientid)
jhs_melt_all$cohort <- as.character(jhs_melt_all$cohort)


# dates_long <-
#   rbind(dates_long[,dates_long_fields],
#         jhs_dates_long[,dates_long_fields])


# bq_table_upload(x="harmonization-286013.cohorts.cohort_dates",
#                 dates_long,quiet=T,
#                 create_disposition="CREATE_IF_NEEDED",
#                 write_disposition="WRITE_TRUNCATE")


gcs_auth("~/Dropbox/ADAPT-HF/Master HDCP files/harmonization-286013-39f492122f69.json")

write_parquet(jhs_melt_all[,..data_fields],
              "~/Dropbox/ADAPT-HF/Master HDCP files/Cohort data/alldata_jhs.parquet")
gcs_upload(jhs_melt_all[,..data_fields], 
           bucket="master_hdcp_files",
           name="jhs_melt_all.parquet",
           object_function = f)

rm(list=ls(pattern="\\bjhs."))

gc()
```
