## Supplementary material for "Aggregating data to accelerate personalized therapy in heart failure (ADAPT-HF)": Harmonization scripts: Compile MESA.R

```
library(sqldf)
library(Hmisc)
library(plotrix)
library(descr)
library(dplyr)
library(relper)
library(stringr)
library(bigrquery)
library(haven)
library(googleCloudStorageR)
library(data.table)
library(arrow)
library(parquetize)
library(bigQueryR)

source("~/Dropbox/R scripts/Misc clinical scripts.R")
source("~/Dropbox/R scripts/echo scripts.R")


outcomes_list <- c("dth_status","dth_dt",
                   "cvdth_status","cvdth_dt",
                   "hfdth_status","hfdth_dt",
                   "hfhosp_ef_val","hfhosp_ef_cat",
                   "noncvdth_status","noncvdth_dt",
                   "hosp_status","hosp_dt",
                   "cadhosp_status","cadhosp_dt",
                   "cvhosp_status","cvhosp_dt",
                   "hfhosp_status","hfhosp_dt",
                   "noncvhosp_status","noncvhosp_dt")


# f <- function(input,output) {
#   write.csv(input,file=output, row.names=F, na="")
# }

visit_yrs <- fread('~/Dropbox/ADAPT-HF/Master HDCP files/Cohort visit yrs.csv')

data_fields <- 
  c("study",
    "patientid",
    "visit_yr",
    "visitdays",
    "form",
    "variable",
    "value",
    "datapoint",
    "study_field",
    "cohort",
    "cohort_name",
    "age_obs")

outcomes_list <- c("dth_status","dth_dt",
                   "cvdth_status","cvdth_dt",
                   "hfdth_status","hfdth_dt",
                   "hfhosp_ef_val","hfhosp_ef_cat",
                   "noncvdth_status","noncvdth_dt",
                   "hosp_status","hosp_dt",
                   "cadhosp_status","cadhosp_dt",
                   "cvhosp_status","cvhosp_dt",
                   "hfhosp_status","hfhosp_dt",
                   "noncvhosp_status","noncvhosp_dt")

dates_long_fields <-
  c("study",
    "cohort",
    "cohort_name",
    "patientid",
    "sex",
    "visit_yr",
    "visitdays",
    "age_obs")


#### ************************ MESA ************************ #### 

# .%%.....%%.%%%%%%%%..%%%%%%.....%%%...
# .%%%...%%%.%%.......%%....%%...%%.%%..
# .%%%%.%%%%.%%.......%%........%%...%%.
# .%%.%%%.%%.%%%%%%....%%%%%%..%%.....%%
# .%%.....%%.%%.............%%.%%%%%%%%%
# .%%.....%%.%%.......%%....%%.%%.....%%
# .%%.....%%.%%%%%%%%..%%%%%%..%%.....%%


# ==============================================================#
##                MESA dates and major outcomes                ##
# ==============================================================#

mesa_exam1 <- fread("~/Dropbox/BioLINCC files/MESA/Primary/Exam1/Data/mesae1dres20220813.csv",na.strings=c("NA","","NULL"))
mesa_exam2 <- fread("~/Dropbox/BioLINCC files/MESA/Primary/Exam2/Data/mesae2dres06222012.csv",na.strings=c("NA","","NULL"))
mesa_exam3 <- fread("~/Dropbox/BioLINCC files/MESA/Primary/Exam3/Data/mesae3dres06222012.csv",na.strings=c("NA","","NULL"))
mesa_exam4 <- fread("~/Dropbox/BioLINCC files/MESA/Primary/Exam4/Data/mesae4dres06222012.csv",na.strings=c("NA","","NULL"))
mesa_exam5 <- fread("~/Dropbox/BioLINCC files/MESA/Primary/Exam5/Data/mesae5_drepos_20220820.csv",na.strings=c("NA","","NULL"))
mesa_exam6 <- fread("~/Dropbox/BioLINCC files/MESA/Primary/Exam6/Data/mesae6_drepos_20250102.csv",na.strings=c("NA","","NULL"))

names(mesa_exam1) <- tolower(names(mesa_exam1))
names(mesa_exam2) <- tolower(names(mesa_exam2))
names(mesa_exam3) <- tolower(names(mesa_exam3))
names(mesa_exam4) <- tolower(names(mesa_exam4))
names(mesa_exam5) <- tolower(names(mesa_exam5))
names(mesa_exam6) <- tolower(names(mesa_exam6))


mesa_efevents <- fread("~/Dropbox/BioLINCC files/MESA/Primary/Events/CVD/Data/mesaevefthru2015_drepos_20200330.csv",na.strings=c("NA","","NULL"))
mesa_events <- fread("~/Dropbox/BioLINCC files/MESA/Primary/Events/CVD/Data/mesaevthr2020_drepos_20241120.csv",na.strings=c("NA","","NULL"))
mesa_site <- fread("~/Dropbox/BioLINCC files/MESA/Primary/Site/mesa_site_drepos_20181106.csv")

mesa_site$cohort_name[mesa_site$site1c=="A"] <- "MD" 
mesa_site$cohort_name[mesa_site$site1c=="B"] <- "MN" 
mesa_site$cohort_name[mesa_site$site1c=="C"] <- "IL" 
mesa_site$cohort_name[mesa_site$site1c=="D"] <- "CA" 
mesa_site$cohort_name[mesa_site$site1c=="E"] <- "NC" 
mesa_site$cohort_name[mesa_site$site1c=="F"] <- "NY" 

names(mesa_efevents) <- tolower(names(mesa_efevents))
names(mesa_events) <- tolower(names(mesa_events))

mesa_dates <- mesa_exam1[,c("mesaid","age1c")]
mesa_dates <- merge(mesa_dates, mesa_exam2[,c("mesaid","age2c","e12dyc")],all.x=T)
mesa_dates <- merge(mesa_dates, mesa_exam3[,c("mesaid","age3c","e13dyc")],all.x=T)
mesa_dates <- merge(mesa_dates, mesa_exam4[,c("mesaid","age4c","e14dyc")],all.x=T)
mesa_dates <- merge(mesa_dates, mesa_exam5[,c("mesaid","age5c","e15dyc")],all.x=T)
mesa_dates <- merge(mesa_dates, mesa_exam6[,c("mesaid","age6c","e16dyc")],all.x=T)

mesa_sex <- mesa_exam1[,c("mesaid","gender1")]

mesa_dates_1 <- mesa_exam1[,c("mesaid","age1c")]
names(mesa_dates_1)[names(mesa_dates_1)=="age1c"] <- "age_obs"
mesa_dates_1$visitdays <- 0
mesa_dates_1$visit <- 1
mesa_dates_1$visit_yr <- 0

mesa_dates_2 <-mesa_exam2[,c("mesaid","age2c","e12dyc")]
names(mesa_dates_2)[names(mesa_dates_2)=="age2c"] <- "age_obs"
names(mesa_dates_2)[names(mesa_dates_2)=="e12dyc"] <- "visitdays"
mesa_dates_2$visit_yr <- 3
mesa_dates_2$visit <- 2

mesa_dates_3 <-mesa_exam3[,c("mesaid","age3c","e13dyc")]
names(mesa_dates_3)[names(mesa_dates_3)=="age3c"] <- "age_obs"
names(mesa_dates_3)[names(mesa_dates_3)=="e13dyc"] <- "visitdays"
mesa_dates_3$visit <- 3
mesa_dates_3$visit_yr <- 5

mesa_dates_4 <-mesa_exam4[,c("mesaid","age4c","e14dyc")]
names(mesa_dates_4)[names(mesa_dates_4)=="age4c"] <- "age_obs"
names(mesa_dates_4)[names(mesa_dates_4)=="e14dyc"] <- "visitdays"
mesa_dates_4$visit <- 4
mesa_dates_4$visit_yr <- 6

mesa_dates_5 <-mesa_exam5[,c("mesaid","age5c","e15dyc")]
names(mesa_dates_5)[names(mesa_dates_5)=="age5c"] <- "age_obs"
names(mesa_dates_5)[names(mesa_dates_5)=="e15dyc"] <- "visitdays"
mesa_dates_5$visit <- 5
mesa_dates_5$visit_yr <- 10

mesa_dates_6 <-mesa_exam6[,c("mesaid","age6c","e16dyc")]
names(mesa_dates_6)[names(mesa_dates_6)=="age6c"] <- "age_obs"
names(mesa_dates_6)[names(mesa_dates_6)=="e16dyc"] <- "visitdays"
mesa_dates_6$visit <- 6
mesa_dates_6$visit_yr <- 16


mesa_dates_long <- rbind(mesa_dates_1,
                         mesa_dates_2,
                         mesa_dates_3,
                         mesa_dates_4,
                         mesa_dates_5,
                         mesa_dates_6)

mesa_dates_long <- subset(mesa_dates_long,!is.na(visitdays))

mesa_dates_long <- merge(mesa_dates_long,
                         mesa_sex,
                         by="mesaid")

mesa_dates <- merge(mesa_dates,mesa_events,by="mesaid")
mesa_dates <- merge(mesa_dates,
                    mesa_site[,c("MESAID","site1c","cohort_name")],
                    by.x="mesaid",
                    by.y="MESAID",
                    all.x=T)

mesa_dates_long <- merge(mesa_dates_long,
                         mesa_site[,c("MESAID","site1c","cohort_name")],
                         by.x="mesaid",
                         by.y="MESAID",
                         all.x=T)

mesa_dates_long$study <- "MESA"

names(mesa_dates_long)[names(mesa_dates_long)=="site1c"] <- "cohort"


mesa_dates$age_hf <- round(mesa_dates$age1c+mesa_dates$chftt/365)

mesa_dates$last_exam_before_hf[mesa_dates$chf==1&mesa_dates$chftt>0] <- 1
mesa_dates$last_exam_before_hf[mesa_dates$chf==1&mesa_dates$chftt>=mesa_dates$e12dyc] <- 3
mesa_dates$last_exam_before_hf[mesa_dates$chf==1&mesa_dates$chftt>=mesa_dates$e13dyc] <- 5
mesa_dates$last_exam_before_hf[mesa_dates$chf==1&mesa_dates$chftt>=mesa_dates$e14dyc] <- 6
mesa_dates$last_exam_before_hf[mesa_dates$chf==1&mesa_dates$chftt>=mesa_dates$e15dyc] <- 10
mesa_dates$last_exam_before_hf[mesa_dates$chf==1&mesa_dates$chftt>=mesa_dates$e16dyc] <- 10

mesa_dates$last_exam_date_before_hf[mesa_dates$last_exam_before_hf==1&!is.na(mesa_dates$last_exam_before_hf)] <-  0
mesa_dates$last_exam_date_before_hf[mesa_dates$last_exam_before_hf==2&!is.na(mesa_dates$last_exam_before_hf)] <-  mesa_dates$e12dyc[mesa_dates$last_exam_before_hf==2&!is.na(mesa_dates$last_exam_before_hf)]
mesa_dates$last_exam_date_before_hf[mesa_dates$last_exam_before_hf==3&!is.na(mesa_dates$last_exam_before_hf)] <-  mesa_dates$e13dyc[mesa_dates$last_exam_before_hf==3&!is.na(mesa_dates$last_exam_before_hf)]
mesa_dates$last_exam_date_before_hf[mesa_dates$last_exam_before_hf==4&!is.na(mesa_dates$last_exam_before_hf)] <-  mesa_dates$e14dyc[mesa_dates$last_exam_before_hf==4&!is.na(mesa_dates$last_exam_before_hf)]
mesa_dates$last_exam_date_before_hf[mesa_dates$last_exam_before_hf==5&!is.na(mesa_dates$last_exam_before_hf)] <-  mesa_dates$e15dyc[mesa_dates$last_exam_before_hf==5&!is.na(mesa_dates$last_exam_before_hf)]
mesa_dates$last_exam_date_before_hf[mesa_dates$last_exam_before_hf==6&!is.na(mesa_dates$last_exam_before_hf)] <-  mesa_dates$e16dyc[mesa_dates$last_exam_before_hf==6&!is.na(mesa_dates$last_exam_before_hf)]

mesa_dates$hf_dys_last_exam <- mesa_dates$chftt-mesa_dates$last_exam_before_hf
mesa_dates$time_hf_to_dth <- mesa_dates$dthtt-mesa_dates$chftt


# ==============================================================#
####                      MESA Outcomes                      ####
# ==============================================================#

mesa_hfevents <- fread("~/Dropbox/BioLINCC files/MESA/Primary/Events/CVD/Data/mesaevefthru2015_drepos_20200330.csv")
mesa_afevents <- fread("~/Dropbox/BioLINCC files/MESA/Primary/Events/CVD/Data/mesaevaffu10_drepos_2015416.csv")
mesa_cvevents <- fread("~/Dropbox/BioLINCC files/MESA/Primary/Events/CVD/Data/mesaevthr2020_drepos_20241120.csv")
mesa_noncvevents <- fread("~/Dropbox/BioLINCC files/MESA/Primary/Events/NonCVD/Data/mesaevnoncvddres06192012.csv")

names(mesa_cvevents) <- toupper(names(mesa_cvevents))
mesa_cvevents$dth_status <- mesa_cvevents$DTH
mesa_cvevents$dth_dt <- mesa_cvevents$DTHTT

mesa_cvevents$cvdth_status[mesa_cvevents$DTHTYPE %in% c(1,2,3,4)] <- 1
mesa_cvevents$cvdth_status[is.na(mesa_cvevents$cvdth_status)] <- 0
mesa_cvevents$cvdth_dt <- mesa_cvevents$DTHTT

mesa_cvevents$noncvdth_status[mesa_cvevents$DTHTYPE %in% c(5,9)] <- 1
mesa_cvevents$noncvdth_status[is.na(mesa_cvevents$noncvdth_status)] <- 0
mesa_cvevents$noncvdth_dt <- mesa_cvevents$DTHTT

mesa_cvevents$cvhosp_status <- mesa_cvevents$CVDA
mesa_cvevents$cvhosp_dt <- mesa_cvevents$CVDATT

mesa_cvevents$hfhosp_status <- mesa_cvevents$CHF
mesa_cvevents$hfhosp_dt <- mesa_cvevents$CHFTT

mesa_cvevents$cadhosp_status <- mesa_cvevents$CHDA
mesa_cvevents$cadhosp_dt <- mesa_cvevents$CHDATT

mesa_noncvevents$noncvhosp_dt <- apply(mesa_noncvevents[,c(2,
                                                           4,
                                                           6,
                                                           8,
                                                           10,
                                                           12,
                                                           14,
                                                           16,
                                                           18,
                                                           20,
                                                           22,
                                                           24,
                                                           26,
                                                           28,
                                                           30)],
                                       MARGIN=1,
                                       FUN=min,
                                       na.rm=T)


mesa_noncvevents$noncvhosp_status[mesa_noncvevents$noncvhosp_dt==mesa_noncvevents$fuptt] <- 0
mesa_noncvevents$noncvhosp_status[!mesa_noncvevents$noncvhosp_dt==mesa_noncvevents$fuptt] <- 1


mesa_outcomes <- merge(
  mesa_cvevents[,c("MESAID",
                   "dth_status","dth_dt",
                   "cvdth_status","cvdth_dt",
                   "noncvdth_status","noncvdth_dt",
                   "cvhosp_status","cvhosp_dt",
                   "cadhosp_status","cadhosp_dt",
                   "hfhosp_status","hfhosp_dt")],
  mesa_noncvevents[,c("MESAID",
                      "noncvhosp_status",
                      "noncvhosp_dt")],
  by="MESAID",
  all.x=T
)

mesa_outcomes <- merge(mesa_outcomes,
                       mesa_hfevents[,c("MESAID","TTCHF","EFMEAS","EFCLASS")],
                       by.x=c("MESAID","hfhosp_dt"),
                       by.y=c("MESAID","TTCHF"),
                       all.x=T)

mesa_outcomes$hfhosp_ef_val <- mesa_outcomes$EFMEAS

mesa_outcomes$hfhosp_ef_cat[mesa_outcomes$hfhosp_ef_val <= 35] <- "Severely reduced"
mesa_outcomes$hfhosp_ef_cat[mesa_outcomes$hfhosp_ef_val > 35&
                              mesa_outcomes$hfhosp_ef_val <= 45] <- "Moderately reduced"
mesa_outcomes$hfhosp_ef_cat[mesa_outcomes$hfhosp_ef_val > 45&
                              mesa_outcomes$hfhosp_ef_val <= 55] <- "Mildly reduced"

mesa_outcomes$hfhosp_ef_cat[mesa_outcomes$hfhosp_ef_val > 55] <- "Normal"

mesa_outcomes$hosp_status <- 0
mesa_outcomes$hosp_dt <- mesa_outcomes$dth_dt
mesa_outcomes$hosp_status[mesa_outcomes$cvhosp_status==1|
                            mesa_outcomes$noncvhosp_status==1] <- 1

mesa_outcomes$hosp_dt[mesa_outcomes$hosp_status==1] <-
  min(mesa_outcomes$cvhosp_dt[mesa_outcomes$hosp_status==1],
      mesa_outcomes$noncvhosp_dt[mesa_outcomes$hosp_status==1])

mesa_outcomes$study <- "MESA"

names(mesa_outcomes)[1] <- "patientid"
outcomes_to_fill <- outcomes_list[!outcomes_list %in% names(mesa_outcomes)]
outcomes_to_fill <- outcomes_to_fill[!is.na(outcomes_to_fill)]
mesa_outcomes[,outcomes_to_fill] <- NA

# write_parquet(mesa_outcomes,
#               '~/Dropbox/ADAPT-HF/Master HDCP files/Cohort data/mesa_outcomes.parquet')

#===============================================================#
####                       MESA Events                       ####
#===============================================================#


# mesa_cvevents <- fread("~/Dropbox/BioLINCC files/MESA/Primary/Events/CVD/Data/mesaevthr2020_drepos_20241120.csv",
#                        na.strings=c("NA","","NULL"))
# mesa_noncvevents <- fread('~/Dropbox/BioLINCC files/MESA/Primary/Events/NonCVD/Data/mesaevnoncvddres06192012.csv',na.strings=c("NA","","NULL"))
# 
# mesa_hfevents <- fread("~/Dropbox/BioLINCC files/MESA/Primary/Events/CVD/Data/mesaevefthru2015_drepos_20200330.csv",
#                        na.strings=c("NA","","NULL"))


#===============================================================#
####                       MESA Exam 1                       ####
#===============================================================#

## QRS

mesa_qrs1 <- fread("~/Dropbox/BioLINCC files/MESA/Primary/Exam1/Data/mesae1ecgcw_drepos_20201102.csv",
                   na.strings=c("NA","","NULL"))[,c("MESAID","QRSDUR1")]
names(mesa_qrs1) <- tolower(names(mesa_qrs1))

mesa_exam1$visit <- 1
mesa_exam1$visitdays <- 0
mesa_exam1$visit_yr <- 0
mesa_exam1 <- merge(mesa_exam1,mesa_qrs1,all.x=T)
mesa_exam1$weight_kg <- mesa_exam1$wtlb1/2.2046226218
mesa_exam1$lvmass_ix <- mesa_exam1$olvedm1/mesa_exam1$bsa1c

## HRT

mesa_hrt <-
  mesa_exam1[!is.na(mesa_exam1$hrmage1c),
             c("mesaid",
               "age1c",
               "hrmage1c",
               "hrmqage1",
               "hrmtyp1",
               "hrmrep1")]

mesa_exam1$emot_soc_supp_ix <-
  mesa_exam1$talkto1+
  mesa_exam1$advice1+
  mesa_exam1$affectn1+
  mesa_exam1$hlpchr1+
  mesa_exam1$emospt1+
  mesa_exam1$confide1


######.MESA Physical activity 

# Will define 'sedentary' activities as:
# 'Transportation' (trnmn1c)
# 'Light Leisure TV/read' (leismn1c)
# 'Light work sitting' (q20jlcn1)
# 'Light work standing' (q21jlcn1)

mesa_exam1$sed_act_hr_day <- 
  (mesa_exam1$trnmn1c +
     mesa_exam1$leismn1c +
     mesa_exam1$q20jlcn1 +
     mesa_exam1$q21jlcn1)/60/7

mesa_exam1$sed_act_met_hr_wk <- 
  (mesa_exam1$trnmt1c +
     mesa_exam1$leismt1c +
     mesa_exam1$q20jlcm1 +
     mesa_exam1$q21jlcm1)/60

### Light activity

mesa_exam1$light_act_hr_day <- 
  (mesa_exam1$q01hlcn1 +
     mesa_exam1$q05olcn1 +
     mesa_exam1$q25ulcn1)/60/7

mesa_exam1$light_act_met_hr_wk <- 
  (mesa_exam1$q01hlcm1 +
     mesa_exam1$q05olcm1 +
     mesa_exam1$q25ulcm1)/60

### Moderate activity

mesa_exam1$mod_act_hr_day <- 
  (mesa_exam1$q02hmcn1 +
     mesa_exam1$q03ymcn1 +
     mesa_exam1$q06omcn1 +
     mesa_exam1$q08wmcn1 +
     mesa_exam1$q09wmcn1 +
     mesa_exam1$q10smcn1 +
     mesa_exam1$q13smcn1 +
     mesa_exam1$q14cmcn1 +
     mesa_exam1$q22jmcn1 +
     mesa_exam1$q26umcn1)/60/7


mesa_exam1$mod_act_met_hr_wk <- 
  (mesa_exam1$q02hmcm1 +
     mesa_exam1$q03ymcm1 +
     mesa_exam1$q06omcm1 +
     mesa_exam1$q08wmcm1 +
     mesa_exam1$q09wmcm1 +
     mesa_exam1$q10smcm1 +
     mesa_exam1$q13smcm1 +
     mesa_exam1$q14cmcm1 +
     mesa_exam1$q22jmcm1 +
     mesa_exam1$q26umcm1)/60

### Vigorous activity

mesa_exam1$vig_act_hr_day <- 
  (mesa_exam1$q04yvcn1 +
     mesa_exam1$q11svcn1 +
     mesa_exam1$q12svcn1 +
     mesa_exam1$q15cvcn1 +
     mesa_exam1$q23jvcn1 +
     mesa_exam1$q27uvcn1)/60/7

mesa_exam1$vig_act_met_hr_wk <- 
  (mesa_exam1$q04yvcm1 +
     mesa_exam1$q11svcm1 +
     mesa_exam1$q12svcm1 +
     mesa_exam1$q15cvcm1 +
     mesa_exam1$q23jvcm1 +
     mesa_exam1$q27uvcm1)/60


mesa_exam1$exer_hr_day <-
  (mesa_exam1$q09wmcn1 +
     mesa_exam1$q10smcn1 +
     mesa_exam1$q11svcn1 +
     mesa_exam1$q12svcn1 +
     mesa_exam1$q13smcn1 +
     mesa_exam1$q14cmcn1 +
     mesa_exam1$q15cvcn1)/60/7

mesa_exam1$exer_met_hr_wk <-
  (mesa_exam1$q09wmcm1 +
     mesa_exam1$q10smcm1 +
     mesa_exam1$q11svcm1 +
     mesa_exam1$q12svcm1 +
     mesa_exam1$q13smcm1 +
     mesa_exam1$q14cmcm1 +
     mesa_exam1$q15cvcm1)/60

### Total activity/week

mesa_exam1$total_act_hr_day <-
  mesa_exam1$sed_act_hr_day +
  mesa_exam1$light_act_hr_day +
  mesa_exam1$mod_act_hr_day +
  mesa_exam1$vig_act_hr_day

mesa_exam1$total_act_hr_day[between(mesa_exam1$total_act_hr_day,0,24)] <- NA

mesa_exam1$sleep_hr_day <- 
  24-mesa_exam1$total_act_hr_day


mesa_exam1$total_met_hr_wk <-
  mesa_exam1$sed_act_met_hr_wk +
  mesa_exam1$light_act_met_hr_wk +
  mesa_exam1$mod_act_met_hr_wk +
  mesa_exam1$vig_act_met_hr_wk

mesa_exam1$total_act_kcal_day <-
  mesa_exam1$total_met_hr_wk/7 * 
  mesa_exam1$weight_kg * 3.5

mesa_exam1$exer_met_hr_wk <-
  round(mesa_exam1$exer_met_hr_wk/60)

mesa_exam1$exer_met_hr_day <-
  round(mesa_exam1$exer_met_hr_wk/7)

mesa_exam1$exer_act_kcal_day <-
  mesa_exam1$exer_met_hr_day * 
  mesa_exam1$weight_kg * 3.5


## Using Framingham PAI

mesa_exam1$fhs_pai <- 
  mesa_exam1$sleep_hr_day +
  mesa_exam1$sed_act_hr_day*1.1 + 
  mesa_exam1$light_act_hr_day*1.5 + 
  mesa_exam1$mod_act_hr_day*2.4 + 
  mesa_exam1$vig_act_hr_day*5


## These yield 20%ile cutoff of 119 for men and 121 for women

mesa_diet_exam1 <- fread("~/Dropbox/BioLINCC files/MESA/Primary/Exam1/Data/mesae1dietdres06192012.csv")
names(mesa_diet_exam1) <- tolower(names(mesa_diet_exam1))
mesa_diet_exam1$visit <- 1
mesa_diet_exam1$visitdays <- 0
mesa_diet_exam1$visit_yr <- 0
mesa_diet_exam1_melt <- melt(mesa_diet_exam1,id.vars=c("mesaid","visit","visitdays","visit_yr"),na.rm=T)
mesa_diet_exam1_melt$form <- "mesa1dietres"

mesa_exam1_melt <- melt(mesa_exam1,
                        id.vars=c("mesaid","visit","visitdays","visit_yr"),
                        na.rm=T)
mesa_exam1_melt$form <- "mesa_exam1"


#===============================================================#
####                       MESA Exam 2                       ####
#===============================================================#


names(mesa_exam2) <- tolower(names(mesa_exam2))
mesa_exam2$visit <- 2
mesa_exam2$visit_yr <- 3
mesa_exam2$visitdays <- mesa_exam2$e12dyc
mesa_exam2$weight_kg <- mesa_exam2$wtlb2/2.2046226218

mesa_exam2$brthslp2[mesa_exam2$stpbrfr2 > 0] <- 1

mesa_hrt <- 
  merge(mesa_hrt,
        mesa_exam2[,c("mesaid",
                      "hrmage2c",
                      "hrmqage2",
                      "hrmtyp2")],
        by="mesaid",
        all.x = T)

mesa_exam2 <- merge(mesa_exam2,mesa_exam1[,c("mesaid","wtlb1","diabet1","cancer1","arthrit1")],by="mesaid",all.x=T)
mesa_exam2 <- merge(mesa_exam2,mesa_cvevents,by.x="mesaid",by.y="MESAID",all.x=T)
mesa_exam2 <- merge(mesa_exam2,mesa_noncvevents[,c(1,3:30)],by.x="mesaid",by.y="MESAID",all.x=T)

mesa_exam2$drinks_wk <- 
  mesa_exam2$rwinewk2+
  mesa_exam2$wwinewk2+
  mesa_exam2$beerwk2+
  mesa_exam2$highalc2

mesa_exam2$loud_snoring <- NA
mesa_exam2$loud_snoring <- paste(mesa_exam2$snrfrq2,mesa_exam2$snrloud2,sep = "_")


###### FRAIL Score, Exam 2

##### Fatigue (using FHS def - CESD everything an effort OR could not get going)

mesa_exam2$fatigue_frail[mesa_exam2$energy2 < 5] <- 1
mesa_exam2$fatigue_frail[mesa_exam2$energy2 >=5 ] <- 0


#### Resistance

mesa_exam2$resistance_frail[mesa_exam2$hilwalk2 %in% c(0,1)] <- mesa_exam2$hilwalk2[mesa_exam2$hilwalk2 %in% c(0,1)]

#### Ambulation

mesa_exam2$ambulate_frail[mesa_exam2$levwalk2 %in% c(0,1)] <- mesa_exam2$levwalk2[mesa_exam2$levwalk2 %in% c(0,1)]


# FRAIL Illness score counts presence of HTN, DM, cancer, chronic lung disease, MI, CHD, angina, asthma, arthritis, stroke, and ckd
# Scores for 0-4 conditions, score = 0, for ≥ 5, score=1


# DM

mesa_exam2$dm_frail <- 0
mesa_exam2$dm_frail[mesa_exam2$dm032c %in% c(2,3)|
                      mesa_exam2$diabins2==1|
                      mesa_exam2$diabet1==1|
                      (mesa_exam2$diab==1&mesa_exam2$diabtt&mesa_exam2$e12dyc)] <- 1

# HTN

mesa_exam2$htn_frail <- 0
mesa_exam2$htn_frail[mesa_exam2$htn2c ==1|
                       mesa_exam2$highbp1==1|
                       mesa_exam2$htnmed2c==1] <- 1

# COPD (chronic lung disease)

mesa_exam2$copd_frail <- 0
mesa_exam2$copd_frail[mesa_exam2$emphys2==1|
                        (mesa_exam2$copd==1&mesa_exam2$copdtt <= mesa_exam2$e12dyc)] <- 1

# asthma

mesa_exam2$asthma_frail <- 0
mesa_exam2$asthma_frail[mesa_exam2$asthma2==1|
                          (mesa_exam2$asthma==1&mesa_exam2$asthmatt <= mesa_exam2$e12dyc)] <- 1

# DJD (arthritis)

mesa_exam2$djd_frail <- 0
mesa_exam2$djd_frail[mesa_exam2$arth2wk2==1|
                       mesa_exam2$arthrit2==1] <- 1

# CKD (renal disease)

mesa_exam2$ckd_frail <- 0
mesa_exam2$ckd_frail[mesa_exam2$kdnydis2==1|
                       (mesa_exam2$chkdds==1&mesa_exam2$chkddstt <= mesa_exam2$e12dyc)] <- 1

# CHF/MI


mesa_exam2$chf_frail <- 0
mesa_exam2$chf_frail[mesa_exam2$CHF==1&mesa_exam2$CHFTT <= mesa_exam2$e12dyc] <- 1

# CAD

# Using 'all' CHD events from MESA as this includes angina which is specified in the FRAIL criteria

mesa_exam2$chd_frail <- 0
mesa_exam2$chd_frail[mesa_exam2$CHDA==1&mesa_exam2$CHDATT <= mesa_exam2$e12dyc] <- 1

# Stroke

mesa_exam2$stroke_frail <- 0
mesa_exam2$stroke_frail[mesa_exam2$STRK==1&mesa_exam2$STRKTYPE <= mesa_exam2$e12dyc] <- 1

# Cancer

mesa_exam2$cancer_frail <- 0
mesa_exam2$cancer_frail[mesa_exam2$cancer1==1|
                          mesa_exam2$cancer==1&mesa_exam2$cancertt <= mesa_exam2$e12dyc] <- 1

# Compile chronic illness

mesa_exam2$conditions_frail <- mesa_exam2$dm_frail+
  mesa_exam2$htn_frail+
  mesa_exam2$copd_frail+
  mesa_exam2$asthma_frail+
  mesa_exam2$djd_frail+
  mesa_exam2$ckd_frail+
  mesa_exam2$chf_frail+
  mesa_exam2$chd_frail+
  mesa_exam2$stroke_frail+
  mesa_exam2$cancer_frail

mesa_exam2$illness_frail[mesa_exam2$conditions_frail <= 4] <- 0
mesa_exam2$illness_frail[mesa_exam2$conditions_frail > 4] <- 1

#### weight Loss

# Supposed to be ~ 10 lbs in past year, but mesa visits are further apart
# Instead will use lowest quintile for each interval, since each interval is different length

mesa_exam2$wtlb2_1_delta <- mesa_exam2$wtlb2-mesa_exam1$wtlb1
quantile(mesa_exam2$wtlb2_1_delta,probs=seq(0,1,0.2),na.rm=T)

# For exam 2 to 1, lowest quintile is ~ -5.8 lbs

mesa_exam2$wtloss_frail[mesa_exam2$wtlb2_1_delta >= -5.8] <- 0
mesa_exam2$wtloss_frail[mesa_exam2$wtlb2_1_delta < -5.8] <- 1

#### Calculate FRAIL score

mesa_exam2$total_frail <- mesa_exam2$fatigue_frail+
  mesa_exam2$resistance_frail+
  mesa_exam2$ambulate_frail+
  mesa_exam2$illness_frail+
  mesa_exam2$wtloss_frail


######.MESA Physical activity 

mesa_exam2$sed_act_hr_day <- 
  (mesa_exam2$q16ilcn2+         # LEISURE TV MIN/WK
     mesa_exam2$q17ilcn2)/60/7    # LEISURE READING MIN/WK

mesa_exam2$sed_act_met_hr_wk <- 
  (mesa_exam2$q16ilcm2+         # LEISURE TV MET MIN/WK
     mesa_exam2$q17ilcm2)/60       # LEISURE READING MIN/WK

mesa_exam2$light_act_hr_day <-
  (mesa_exam2$q01hlcn2+         # LIGHT HOUSEHOLD CHORES MIN/WK
     mesa_exam2$q05olcn2 +      # LIGHT CARE OTHERS MIN/WK
     mesa_exam2$q20jlcn2 +      # LIGHT WORK SITTING MIN/WK
     mesa_exam2$q21jlcn2)/60/7  # LIGHT WORK STANDING MIN/WK


mesa_exam2$light_act_met_hr_wk <-
  (mesa_exam2$q01hlcm2+         # LIGHT HOUSEHOLD CHORES MIN/WK
     mesa_exam2$q05olcm2 +         # LIGHT CARE OTHERS MIN/WK
     mesa_exam2$q20jlcm2 +       # LIGHT WORK SITTING MIN/WK
     mesa_exam2$q21jlcm2)/60     # LIGHT WORK STANDING MIN/WK


mesa_exam2$mod_act_hr_day <-
  (mesa_exam2$q02hmcn2 +        # MODERATE HOUSEHOLD CHORES MIN/WK
     mesa_exam2$q03ymcn2 +      # MODERATE YARD WORK MIN/WK
     mesa_exam2$q06omcn2 +      # MODERATE CARE OTHERS MIN/WK
     mesa_exam2$q08wmcn2 +      # MODERATE WALKING MIN/WK
     mesa_exam2$q09wmcn2 +      # MODERATE WALKING EXERCISE MIN/WK
     mesa_exam2$q10smcn2 +      # MODERATE DANCE MIN/WK
     mesa_exam2$q13smcn2 +      # MODERATE INDIVIDUAL ACTIVITIES MIN/WK
     mesa_exam2$q14cmcn2 +      # MODERATE CONDITIONING MIN/WK
     mesa_exam2$q22jmcn2)/60/7  # MODERATE WORK MIN/WK


mesa_exam2$mod_act_met_hr_wk <-
  (mesa_exam2$q02hmcm2 +        # MODERATE HOUSEHOLD CHORES MET MIN/WK
     mesa_exam2$q03ymcm2 +         # MODERATE YARD WORK MET MIN/WK
     mesa_exam2$q06omcm2 +         # MODERATE CARE OTHERS MET MIN/WK
     mesa_exam2$q08wmcm2 +         # MODERATE WALKING MET MIN/WK
     mesa_exam2$q09wmcm2 +         # MODERATE WALKING EXERCISE MET MIN/WK
     mesa_exam2$q10smcm2 +         # MODERATE DANCE MET MIN/WK
     mesa_exam2$q13smcm2 +         # MODERATE INDIVIDUAL MET ACTIVITIES MIN/WK
     mesa_exam2$q14cmcm2 +         # MODERATE CONDITIONING MET MIN/WK
     mesa_exam2$q22jmcm2)/60       # MODERATE WORK MET MIN/WK


mesa_exam2$vig_act_hr_day <-
  (mesa_exam2$q04yvcn2 +        # VIGOROUS YARD WORK MIN/WK
     mesa_exam2$q11svcn2 +      # VIGOROUS TEAM SPORTS MIN/WK
     mesa_exam2$q12svcn2 +      # VIGOROUS DUAL SPORTS MIN/WK
     mesa_exam2$q15cvcn2 +      # VIGOROUS CONDITIONING MIN/WK
     mesa_exam2$q23jvcn2)/60/7  # VIGOROUS WORK MIN/WK


mesa_exam2$vig_act_met_hr_wk <-
  (mesa_exam2$q04yvcm2 +        # VIGOROUS YARD WORK MIN/WK
     mesa_exam2$q11svcm2 +         # VIGOROUS TEAM SPORTS MIN/WK
     mesa_exam2$q12svcm2 +         # VIGOROUS DUAL SPORTS MIN/WK
     mesa_exam2$q15cvcm2 +         # VIGOROUS CONDITIONING MIN/WK
     mesa_exam2$q23jvcm2)/60       # VIGOROUS WORK MIN/WK

mesa_exam2$exer_met_hr_wk <- 
  (mesa_exam2$q09wmcm2 +        # MODERATE WALKING EXERCISE MET MIN/WK
     mesa_exam2$q10smcm2 +         # MODERATE DANCE MET MIN/WK
     mesa_exam2$q11svcm2 +         # VIGOROUS TEAM SPORTS MET MIN/WK
     mesa_exam2$q12svcm2 +         # VIGOROUS DUAL SPORTS MET MIN/WK
     mesa_exam2$q13smcm2 +         # MODERATE INDIVIDUAL ACTIVITIES MET MIN/WK
     mesa_exam2$q14cmcm2 +         # MODERATE CONDITIONING MET MIN/WK
     mesa_exam2$q15cvcm2)/60       # VIGOROUS CONDITIOINING MET MIN/WK

mesa_exam2$total_met_hr_wk <-
  mesa_exam2$light_act_met_hr_wk +
  mesa_exam2$mod_act_met_hr_wk +
  mesa_exam2$vig_act_met_hr_wk

mesa_exam2$total_act_hr_day <-
  mesa_exam2$sed_act_hr_day+
  mesa_exam2$light_act_hr_day+
  mesa_exam2$mod_act_hr_day+
  mesa_exam2$vig_act_hr_day

mesa_exam2$total_act_hr_day[!between(mesa_exam2$total_act_hr_day,0,24)] <- NA


mesa_exam2$sleep_hr_day <-
  24-mesa_exam2$total_act_hr_day

mesa_exam2$total_met_hr_wk[mesa_exam2$total_met_hr_wk>450] <- NA
mesa_exam2$total_met_hr_wk[mesa_exam2$total_met_hr_wk<15] <- NA

mesa_exam2$total_met_hr_day <-
  mesa_exam2$total_met_hr_wk/7

mesa_exam2$total_act_kcal_day <-
  mesa_exam2$total_met_hr_day * 
  mesa_exam2$weight_kg * 3.5

mesa_exam2$exer_met_hr_day <-
  round(mesa_exam2$exer_met_hr_wk/7)

mesa_exam2$exer_act_kcal_day <-
  (mesa_exam2$exer_met_hr_wk * 
     mesa_exam2$weight_kg) * 3.5*7


mesa_exam2$total_met_hr_wk <- 
  mesa_exam2$light_act_met_hr_wk +
  mesa_exam2$mod_act_met_hr_wk +
  mesa_exam2$vig_act_met_hr_wk

mesa_exam2$total_met_hr_day <-
  round(mesa_exam2$total_met_hr_wk/7)

mesa_exam2$total_act_kcal_day <-
  mesa_exam2$total_met_hr_day * 
  mesa_exam2$weight_kg * 3.5

mesa_exam2$exer_met_hr_day <-
  round(mesa_exam2$exer_met_hr_wk/7)

mesa_exam2$exer_act_kcal_day <-
  mesa_exam2$exer_met_hr_day * 
  mesa_exam2$weight_kg * 3.5


mesa_exam2$fhs_pai <- 
  mesa_exam2$sleep_hr_day+
  mesa_exam2$sed_act_hr_day*1.1 + 
  mesa_exam2$light_act_hr_day*1.5 + 
  mesa_exam2$mod_act_hr_day*2.4 + 
  mesa_exam2$vig_act_hr_day*5


mesa_exam2_melt <- melt(subset(mesa_exam2,!is.na(visitdays)),
                        id.vars=c("mesaid","visit","visitdays","visit_yr"),
                        na.rm=T)

mesa_exam2_melt$form <- "mesa_exam2"


#===============================================================#
####                       MESA Exam 3                       ####
#===============================================================#


names(mesa_exam3) <- tolower(names(mesa_exam3))
mesa_exam3$visit <- 3
mesa_exam3$visitdays <- mesa_exam3$e13dyc
mesa_exam3$visit_yr <- 5
mesa_exam3$weight_kg <- mesa_exam3$wtlb3/2.2046226218


mesa_exam3 <- merge(mesa_exam3,mesa_exam1[,c("mesaid","diabet1","cancer1","arthrit1")],by="mesaid",all.x=T)
mesa_exam3 <- merge(mesa_exam3,mesa_exam2[,c("mesaid","wtlb2")],by="mesaid",all.x=T)
mesa_exam3 <- merge(mesa_exam3,mesa_cvevents,by.x="mesaid",by.y="MESAID",all.x=T)
mesa_exam3 <- merge(mesa_exam3,mesa_noncvevents[,c(1,3:30)],by.x="mesaid",by.y="MESAID",all.x=T)

mesa_exam3$drinks_wk <- 
  mesa_exam3$rwinewk3+
  mesa_exam3$wwinewk3+
  mesa_exam3$beerwk3+
  mesa_exam3$highalc3


mesa_hrt <-
  merge(mesa_hrt,
        mesa_exam3[,c("mesaid",
                      "hrmage3c",
                      "hrmqage3",
                      "hrmtyp3")],
        by="mesaid",
        all.x=T)

mesa_exam3$job_stress_derive[mesa_exam3$job1prb3==0] <- 0
mesa_exam3$job_stress_derive[!is.na(mesa_exam3$job3prb3)] <- mesa_exam3$job3prb3[!is.na(mesa_exam3$job3prb3)]

mesa_exam3$med_stress_derive[mesa_exam3$hprb3pt3==0] <- 0
mesa_exam3$med_stress_derive[!is.na(mesa_exam3$hprb3pt3)] <- mesa_exam3$hprb3pt3[!is.na(mesa_exam3$hprb3pt3)]

mesa_exam3$rel_stress_derive[mesa_exam3$rel1prb3==0] <- 0
mesa_exam3$rel_stress_derive[!is.na(mesa_exam3$rel1prb3)] <- mesa_exam3$rel1prb3[!is.na(mesa_exam3$rel1prb3)]


mesa_exam3$emot_soc_supp_ix <-
  mesa_exam3$talkto3+
  mesa_exam3$advice3+
  mesa_exam3$affectn3+
  mesa_exam3$hlpchr3+
  mesa_exam3$emospt3+
  mesa_exam3$confide3


###### FRAIL Score, Exam 3

##### Fatigue (using FHS def - CESD everything an effort OR could not get going)

mesa_exam3$exhaustion_fried[mesa_exam3$effort3==4|mesa_exam3$getgoin3==4] <- 1
mesa_exam3$exhaustion_fried[mesa_exam3$effort3<4&mesa_exam3$getgoin3<4] <- 0


#### Resistance

mesa_exam3$resistance_frail[mesa_exam3$hilwalk3 %in% c(0,1)] <- mesa_exam3$hilwalk3[mesa_exam3$hilwalk3 %in% c(0,1)]

#### Ambulation

mesa_exam3$ambulate_frail[mesa_exam3$levwalk3 %in% c(0,1)] <- mesa_exam3$levwalk3[mesa_exam3$levwalk3 %in% c(0,1)]


# FRAIL Illness score counts presence of HTN, DM, cancer, chronic lung disease, MI, CHD, angina, asthma, arthritis, stroke, and ckd
# Scores for 0-4 conditions, score = 0, for ≥ 5, score=1


# DM

mesa_exam3$dm_frail <- 0
mesa_exam3$dm_frail[mesa_exam3$dm033c %in% c(2,3)|
                      mesa_exam3$diabins3==1|
                      mesa_exam3$diabet1==1|
                      (mesa_exam3$diab==1&mesa_exam3$diabtt&mesa_exam3$e13dyc)] <- 1

# HTN

mesa_exam3$htn_frail <- 0
mesa_exam3$htn_frail[mesa_exam3$htn3c ==1|
                       mesa_exam3$highbp1==1|
                       mesa_exam3$htnmed3c==1] <- 1

# COPD (chronic lung disease)

mesa_exam3$copd_frail <- 0
mesa_exam3$copd_frail[mesa_exam3$emphys3==1|
                        (mesa_exam3$copd==1&mesa_exam3$copdtt <= mesa_exam3$e13dyc)] <- 1

# asthma

mesa_exam3$asthma_frail <- 0
mesa_exam3$asthma_frail[mesa_exam3$asthma3==1|
                          (mesa_exam3$asthma==1&mesa_exam3$asthmatt <= mesa_exam3$e13dyc)] <- 1

# DJD (arthritis)

mesa_exam3$djd_frail <- 0
mesa_exam3$djd_frail[mesa_exam3$arth2wk3==1|
                       mesa_exam3$arthrit3==1] <- 1

# CKD (renal disease)

mesa_exam3$ckd_frail <- 0
mesa_exam3$ckd_frail[mesa_exam3$kdnydis3==1|
                       (mesa_exam3$chkdds==1&mesa_exam3$chkddstt <= mesa_exam3$e13dyc)] <- 1

# CHF/MI


mesa_exam3$chf_frail <- 0
mesa_exam3$chf_frail[mesa_exam3$CHF==1&mesa_exam3$CHFTT <= mesa_exam3$e13dyc] <- 1

# CAD

# Using 'all' CHD events from MESA as this includes angina which is specified in the FRAIL criteria

mesa_exam3$chd_frail <- 0
mesa_exam3$chd_frail[mesa_exam3$CHDA==1&mesa_exam3$CHDATT <= mesa_exam3$e13dyc] <- 1

# Stroke

mesa_exam3$stroke_frail <- 0
mesa_exam3$stroke_frail[mesa_exam3$STRK==1&mesa_exam3$STRKTYPE <= mesa_exam3$e13dyc] <- 1

# Cancer

mesa_exam3$cancer_frail <- 0
mesa_exam3$cancer_frail[mesa_exam3$cancer==1&mesa_exam3$cancertt <= mesa_exam3$e13dyc] <- 1

# Compile chronic illness

mesa_exam3$conditions_frail <- mesa_exam3$dm_frail+
  mesa_exam3$htn_frail+
  mesa_exam3$copd_frail+
  mesa_exam3$asthma_frail+
  mesa_exam3$djd_frail+
  mesa_exam3$ckd_frail+
  mesa_exam3$chf_frail+
  mesa_exam3$chd_frail+
  mesa_exam3$stroke_frail+
  mesa_exam3$cancer_frail

mesa_exam3$illness_frail[mesa_exam3$conditions_frail <= 4] <- 0
mesa_exam3$illness_frail[mesa_exam3$conditions_frail > 4] <- 1

#### weight Loss

# Supposed to be ~ 10 lbs in past year, but mesa visits are further apart
# Instead will use lowest quintile for each interval, since each interval is different length


mesa_exam3$wtlb3_2_delta <- mesa_exam3$wtlb3-mesa_exam3$wtlb2
quantile(mesa_exam3$wtlb3_2_delta,probs=seq(0,1,0.2),na.rm=T)

# For exam 2 to 3, lowest quintile is ~ -6 lbs

mesa_exam3$wtloss_frail[mesa_exam3$wtlb3_2_delta >= -6] <- 0
mesa_exam3$wtloss_frail[mesa_exam3$wtlb3_2_delta < -6] <- 1

#### Calculate FRAIL score

mesa_exam3$total_frail <- mesa_exam3$exhaustion_fried+
  mesa_exam3$resistance_frail+
  mesa_exam3$ambulate_frail+
  mesa_exam3$illness_frail+
  mesa_exam3$wtloss_frail


######.MESA Physical activity 

mesa_exam3$sed_act_hr_day <- 
  (mesa_exam3$q16ilcn3+         # LEISURE TV MIN/WK
     mesa_exam3$q17ilcn3)/60/7    # LEISURE READING MIN/WK

mesa_exam3$sed_act_met_hr_wk <- 
  (mesa_exam3$q16ilcm3+         # LEISURE TV MET MIN/WK
     mesa_exam3$q17ilcm3)/60       # LEISURE READING MIN/WK

mesa_exam3$light_act_hr_day <-
  (mesa_exam3$q01hlcn3+         # LIGHT HOUSEHOLD CHORES MIN/WK
     mesa_exam3$q05olcn3 +      # LIGHT CARE OTHERS MIN/WK
     mesa_exam3$q20jlcn3 +      # LIGHT WORK SITTING MIN/WK
     mesa_exam3$q21jlcn3)/60/7  # LIGHT WORK STANDING MIN/WK


mesa_exam3$light_act_met_hr_wk <-
  (mesa_exam3$q01hlcm3+         # LIGHT HOUSEHOLD CHORES MIN/WK
     mesa_exam3$q05olcm3 +         # LIGHT CARE OTHERS MIN/WK
     mesa_exam3$q20jlcm3 +       # LIGHT WORK SITTING MIN/WK
     mesa_exam3$q21jlcm3)/60     # LIGHT WORK STANDING MIN/WK


mesa_exam3$mod_act_hr_day <-
  (mesa_exam3$q02hmcn3 +        # MODERATE HOUSEHOLD CHORES MIN/WK
     mesa_exam3$q03ymcn3 +      # MODERATE YARD WORK MIN/WK
     mesa_exam3$q06omcn3 +      # MODERATE CARE OTHERS MIN/WK
     mesa_exam3$q08wmcn3 +      # MODERATE WALKING MIN/WK
     mesa_exam3$q09wmcn3 +      # MODERATE WALKING EXERCISE MIN/WK
     mesa_exam3$q10smcn3 +      # MODERATE DANCE MIN/WK
     mesa_exam3$q13smcn3 +      # MODERATE INDIVIDUAL ACTIVITIES MIN/WK
     mesa_exam3$q14cmcn3 +      # MODERATE CONDITIONING MIN/WK
     mesa_exam3$q22jmcn3)/60/7  # MODERATE WORK MIN/WK


mesa_exam3$mod_act_met_hr_wk <-
  (mesa_exam3$q02hmcm3 +        # MODERATE HOUSEHOLD CHORES MET MIN/WK
     mesa_exam3$q03ymcm3 +         # MODERATE YARD WORK MET MIN/WK
     mesa_exam3$q06omcm3 +         # MODERATE CARE OTHERS MET MIN/WK
     mesa_exam3$q08wmcm3 +         # MODERATE WALKING MET MIN/WK
     mesa_exam3$q09wmcm3 +         # MODERATE WALKING EXERCISE MET MIN/WK
     mesa_exam3$q10smcm3 +         # MODERATE DANCE MET MIN/WK
     mesa_exam3$q13smcm3 +         # MODERATE INDIVIDUAL MET ACTIVITIES MIN/WK
     mesa_exam3$q14cmcm3 +         # MODERATE CONDITIONING MET MIN/WK
     mesa_exam3$q22jmcm3)/60       # MODERATE WORK MET MIN/WK


mesa_exam3$vig_act_hr_day <-
  (mesa_exam3$q04yvcn3 +        # VIGOROUS YARD WORK MIN/WK
     mesa_exam3$q11svcn3 +      # VIGOROUS TEAM SPORTS MIN/WK
     mesa_exam3$q12svcn3 +      # VIGOROUS DUAL SPORTS MIN/WK
     mesa_exam3$q15cvcn3 +      # VIGOROUS CONDITIONING MIN/WK
     mesa_exam3$q23jvcn3)/60/7  # VIGOROUS WORK MIN/WK


mesa_exam3$vig_act_met_hr_wk <-
  (mesa_exam3$q04yvcm3 +        # VIGOROUS YARD WORK MIN/WK
     mesa_exam3$q11svcm3 +         # VIGOROUS TEAM SPORTS MIN/WK
     mesa_exam3$q12svcm3 +         # VIGOROUS DUAL SPORTS MIN/WK
     mesa_exam3$q15cvcm3 +         # VIGOROUS CONDITIONING MIN/WK
     mesa_exam3$q23jvcm3)/60       # VIGOROUS WORK MIN/WK

mesa_exam3$exer_met_hr_wk <- 
  (mesa_exam3$q09wmcm3 +        # MODERATE WALKING EXERCISE MET MIN/WK
     mesa_exam3$q10smcm3 +         # MODERATE DANCE MET MIN/WK
     mesa_exam3$q11svcm3 +         # VIGOROUS TEAM SPORTS MET MIN/WK
     mesa_exam3$q12svcm3 +         # VIGOROUS DUAL SPORTS MET MIN/WK
     mesa_exam3$q13smcm3 +         # MODERATE INDIVIDUAL ACTIVITIES MET MIN/WK
     mesa_exam3$q14cmcm3 +         # MODERATE CONDITIONING MET MIN/WK
     mesa_exam3$q15cvcm3)/60       # VIGOROUS CONDITIOINING MET MIN/WK

mesa_exam3$total_met_hr_wk <-
  mesa_exam3$light_act_met_hr_wk +
  mesa_exam3$mod_act_met_hr_wk +
  mesa_exam3$vig_act_met_hr_wk

mesa_exam3$total_act_hr_day <-
  mesa_exam3$sed_act_hr_day+
  mesa_exam3$light_act_hr_day+
  mesa_exam3$mod_act_hr_day+
  mesa_exam3$vig_act_hr_day

mesa_exam3$total_act_hr_day[!between(mesa_exam3$total_act_hr_day,0,24)] <- NA


mesa_exam3$sleep_hr_day <-
  24-mesa_exam3$total_act_hr_day

mesa_exam3$total_met_hr_wk[mesa_exam3$total_met_hr_wk>450] <- NA
mesa_exam3$total_met_hr_wk[mesa_exam3$total_met_hr_wk<15] <- NA

mesa_exam3$total_met_hr_day <-
  mesa_exam3$total_met_hr_wk/7

mesa_exam3$total_act_kcal_day <-
  mesa_exam3$total_met_hr_day * 
  mesa_exam3$weight_kg * 3.5

mesa_exam3$exer_met_hr_day <-
  round(mesa_exam3$exer_met_hr_wk/7)

mesa_exam3$exer_act_kcal_day <-
  (mesa_exam3$exer_met_hr_wk * 
     mesa_exam3$weight_kg) * 3.5*7


mesa_exam3$total_met_hr_wk <- 
  mesa_exam3$light_act_met_hr_wk +
  mesa_exam3$mod_act_met_hr_wk +
  mesa_exam3$vig_act_met_hr_wk

mesa_exam3$total_met_hr_day <-
  round(mesa_exam3$total_met_hr_wk/7)

mesa_exam3$total_act_kcal_day <-
  mesa_exam3$total_met_hr_day * 
  mesa_exam3$weight_kg * 3.5

mesa_exam3$exer_met_hr_day <-
  round(mesa_exam3$exer_met_hr_wk/7)

mesa_exam3$exer_act_kcal_day <-
  mesa_exam3$exer_met_hr_day * 
  mesa_exam3$weight_kg * 3.5


mesa_exam3$fhs_pai <- 
  mesa_exam3$sleep_hr_day+
  mesa_exam3$sed_act_hr_day*1.1 + 
  mesa_exam3$light_act_hr_day*1.5 + 
  mesa_exam3$mod_act_hr_day*2.4 + 
  mesa_exam3$vig_act_hr_day*5


mesa_exam3_melt <- melt(subset(mesa_exam3,!is.na(visitdays)),
                        id.vars=c("mesaid","visit","visitdays","visit_yr"),
                        na.rm=T)
mesa_exam3_melt$form <- "mesa_exam3"


#===============================================================#
####                       MESA Exam 4                       ####
#===============================================================#

names(mesa_exam4) <- tolower(names(mesa_exam4))
mesa_exam4$visit <- 4
mesa_exam4$visitdays <- mesa_exam4$e14dyc
mesa_exam4$visit_yr <- 6
mesa_exam4$weight_kg <- mesa_exam4$wtlb4/2.2046226218


mesa_exam4 <- merge(mesa_exam4,mesa_exam1[,c("mesaid","diabet1","cancer1","arthrit1")],by="mesaid",all.x=T)
mesa_exam4 <- merge(mesa_exam4,mesa_exam3[,c("mesaid","wtlb3")],by="mesaid",all.x=T)
mesa_exam4 <- merge(mesa_exam4,mesa_cvevents,by.x="mesaid",by.y="MESAID",all.x=T)
mesa_exam4 <- merge(mesa_exam4,mesa_noncvevents[,c(1,3:30)],by.x="mesaid",by.y="MESAID",all.x=T)

mesa_exam4$drinks_wk <- 
  mesa_exam4$rwinewk4+
  mesa_exam4$wwinewk4+
  mesa_exam4$beerwk4+
  mesa_exam4$liqwk4

mesa_hrt <- 
  merge(mesa_hrt,
        mesa_exam4[,c("mesaid",
                      "hrmage4c",
                      "hrmqage4",
                      "hrmtyp4")],
        by="mesaid",
        all.x=T)

mesa_exam4$emot_soc_supp_ix <-
  mesa_exam4$talkto4+
  mesa_exam4$advice4+
  mesa_exam4$affectn4+
  mesa_exam4$hlpchr4+
  mesa_exam4$emospt4+
  mesa_exam4$confide

###### FRAIL Score, Exam 4

##### Fatigue (using FHS def - CESD everything an effort OR could not get going)

mesa_exam4$exhaustion_fried[mesa_exam4$effort4==4|mesa_exam4$getgoin4==4] <- 1
mesa_exam4$exhaustion_fried[mesa_exam4$effort4<4&mesa_exam4$getgoin4<4] <- 0


#### Resistance

mesa_exam4$resistance_frail[mesa_exam4$hilwalk4 %in% c(0,1)] <- mesa_exam4$hilwalk4[mesa_exam4$hilwalk4 %in% c(0,1)]

#### Ambulation

mesa_exam4$ambulate_frail[mesa_exam4$levwalk4 %in% c(0,1)] <- mesa_exam4$levwalk4[mesa_exam4$levwalk4 %in% c(0,1)]


# FRAIL Illness score counts presence of HTN, DM, cancer, chronic lung disease, MI, CHD, angina, asthma, arthritis, stroke, and ckd
# Scores for 0-4 conditions, score = 0, for ≥ 5, score=1


# DM

mesa_exam4$dm_frail <- 0
mesa_exam4$dm_frail[mesa_exam4$dm034c %in% c(2,3)|
                      mesa_exam4$diabins4==1|
                      mesa_exam4$diabet1==1|
                      (mesa_exam4$diab==1&mesa_exam4$diabtt&mesa_exam4$e14dyc)] <- 1

# HTN

mesa_exam4$htn_frail <- 0
mesa_exam4$htn_frail[mesa_exam4$htn4c==1|
                       mesa_exam4$highbp1==1|
                       mesa_exam4$htnmed4c==1] <- 1


# COPD (chronic lung disease)

mesa_exam4$copd_frail <- 0
mesa_exam4$copd_frail[mesa_exam4$emphys4==1|
                        (mesa_exam4$copd==1&
                           mesa_exam4$copdtt <= mesa_exam4$e14dyc)] <- 1

# asthma

mesa_exam4$asthma_frail <- 0
mesa_exam4$asthma_frail[mesa_exam4$asthma4==1|
                          (mesa_exam4$asthma==1&
                             mesa_exam4$asthmatt <= mesa_exam4$e14dyc)] <- 1

# DJD (arthritis)

mesa_exam4$djd_frail <- 0
mesa_exam4$djd_frail[mesa_exam4$arth2wk4==1|
                       mesa_exam4$arthrit4==1] <- 1

# CKD (renal disease)

mesa_exam4$ckd_frail <- 0
mesa_exam4$ckd_frail[mesa_exam4$kdnydis4==1|
                       (mesa_exam4$chkdds==1&
                          mesa_exam4$chkddstt <= mesa_exam4$e14dyc)] <- 1

# CHF/MI


mesa_exam4$chf_frail <- 0
mesa_exam4$chf_frail[mesa_exam4$CHF==1&mesa_exam4$CHFTT <= mesa_exam4$e14dyc] <- 1

# CAD

# Using 'all' CHD events from MESA as this includes angina which is specified in the FRAIL criteria

mesa_exam4$chd_frail <- 0
mesa_exam4$chd_frail[mesa_exam4$CHDA==1&
                       mesa_exam4$CHDATT <= mesa_exam4$e14dyc] <- 1

# Stroke

mesa_exam4$stroke_frail <- 0
mesa_exam4$stroke_frail[mesa_exam4$STRK==1&
                          mesa_exam4$STRKTYPE <= mesa_exam4$e14dyc] <- 1

# Cancer

mesa_exam4$cancer_frail <- 0
mesa_exam4$cancer_frail[mesa_exam4$cancer==1&
                          mesa_exam4$cancertt <= mesa_exam4$e14dyc] <- 1

# Compile chronic illness

mesa_exam4$conditions_frail <- mesa_exam4$dm_frail+
  mesa_exam4$htn_frail+
  mesa_exam4$copd_frail+
  mesa_exam4$asthma_frail+
  mesa_exam4$djd_frail+
  mesa_exam4$ckd_frail+
  mesa_exam4$chf_frail+
  mesa_exam4$chd_frail+
  mesa_exam4$stroke_frail+
  mesa_exam4$cancer_frail

mesa_exam4$illness_frail[mesa_exam4$conditions_frail <= 4] <- 0
mesa_exam4$illness_frail[mesa_exam4$conditions_frail > 4] <- 1

#### weight Loss

# Supposed to be ~ 10 lbs in past year, but mesa visits are further apart
# Instead will use lowest quintile for each interval, since each interval is different length


mesa_exam4$wtlb4_3_delta <- mesa_exam4$wtlb4-mesa_exam4$wtlb3
quantile(mesa_exam4$wtlb4_3_delta,probs=seq(0,1,0.2),na.rm=T)

# For exam 4 to 5, lowest quintile is ~ -5.5 lbs

mesa_exam4$wtloss_frail[mesa_exam4$wtlb4_3_delta >= -5.5] <- 0
mesa_exam4$wtloss_frail[mesa_exam4$wtlb4_3_delta < -5.5] <- 1

#### Calculate FRAIL score

mesa_exam4$total_frail <- mesa_exam4$exhaustion_fried+
  mesa_exam4$resistance_frail+
  mesa_exam4$ambulate_frail+
  mesa_exam4$illness_frail+
  mesa_exam4$wtloss_frail


######.MESA Physical activity 


mesa_exam4_melt <- melt(subset(mesa_exam4,
                               !is.na(visitdays)),
                        id.vars=c("mesaid","visit","visitdays","visit_yr"),
                        na.rm=T)
mesa_exam4_melt$form <- "mesa_exam4"


#===============================================================#
####                       MESA Exam 5                       ####
#===============================================================#

mesa_exam5 <- fread("~/Dropbox/BioLINCC files/MESA/Primary/Exam5/Data/mesae5_drepos_20220820.csv",na.strings=c("NA","","NULL"))
names(mesa_exam5) <- tolower(names(mesa_exam5))
mesa_exam5$visit <- 5
mesa_exam5$visit_yr <- 10
mesa_exam5$visitdays <- mesa_exam5$e15dyc
mesa_exam5$weight_kg <- mesa_exam5$wtlb5/2.2046226218
mesa_exam5$lvmass_ix <- mesa_exam5$olvedm5t/mesa_exam5$bsa5c

mesa_exam5 <- merge(mesa_exam5,mesa_exam4[,c("mesaid","wtlb4")],by="mesaid",all.x=T)
mesa_exam5 <- merge(mesa_exam5,mesa_cvevents,by.x="mesaid",by.y="MESAID",all.x=T)
mesa_exam5 <- merge(mesa_exam5,mesa_noncvevents,
                    by.x="mesaid",
                    by.y="MESAID",
                    all.x=T)

mesa_hrt <- 
  merge(mesa_hrt,
        mesa_exam5[,c("mesaid",
                      "hrmage5",
                      "hrmqage5",
                      "hrmsage5",
                      "hrmtyp5")],
        by="mesaid",
        all.x=T)


mesa_hrt$hrmage5[is.na(mesa_hrt$hrmage5)&!is.na(mesa_hrt$hrmsage5)] <-
  mesa_hrt$hrmsage5[is.na(mesa_hrt$hrmage5)&!is.na(mesa_hrt$hrmsage5)]

mesa_hrt$hrt_start_age_min <-
  apply(mesa_hrt[,c("hrmage1c",
                    "hrmage2c",
                    "hrmage3c",
                    "hrmage4c",
                    "hrmage5")],
        MARGIN=1,
        FUN=function(x) min(x,na.rm=T))

mesa_hrt$hrt_stop_age_max <-
  apply(mesa_hrt[,c("hrmqage1",
                    "hrmqage2",
                    "hrmqage3",
                    "hrmqage4",
                    "hrmqage5")],
        MARGIN=1,
        FUN=function(x) min(x,na.rm=T))

mesa_hrt[mesa_hrt=="Inf"] <- NA
mesa_hrt[mesa_hrt=="-Inf"] <- NA

mesa_hrt$hrt_stop_age_max[!is.na(mesa_hrt$hrt_start_age_min)&
                            is.na(mesa_hrt$hrt_stop_age_max)&
                            !is.na(mesa_hrt$age_eos)] <-
  mesa_hrt$age_eos[!is.na(mesa_hrt$hrt_start_age_min)&
                     is.na(mesa_hrt$hrt_stop_age_max)&
                     !is.na(mesa_hrt$age_eos)]

mesa_hrt$hrt_years_age <-
  mesa_hrt$hrt_stop_age_max - mesa_hrt$hrt_start_age_min

mesa_exam5$casisum[mesa_exam5$FLAGCASI5C==0] <- 
  mesa_exam5$CASISUM5C[mesa_exam5$FLAGCASI5C==0]

mesa_exam5$drinks_wk <- 
  mesa_exam5$rwinewk5+
  mesa_exam5$wwinewk5+
  mesa_exam5$beerwk5+
  mesa_exam5$liqwk5

mesa_exam5$fam_support <-
  mesa_exam5$frely5+
  mesa_exam5$fopen5+
  mesa_exam5$fdemand5+
  mesa_exam5$fldown5

mesa_exam5$pss4 <- 
  mesa_exam5$ucontrl5+
  mesa_exam5$confid5+
  mesa_exam5$goingyw5+
  mesa_exam5$pileup5

mesa_exam5 <- merge(mesa_exam5,
                    mesa_noncvevents,
                    by.x="mesaid",
                    by.y ="MESAID",
                    all.x=T,
                    all.y=F)

mesa_exam5$dementia_dx <- 0
mesa_exam5$dementia_dx[mesa_exam5$demen==1&mesa_exam5$dementt <= mesa_exam5$E15DYC] <- 1

######### FRAIL Score, Exam 5

##### Fatigue (using FHS def - CESD everything an effort OR could not get going)

mesa_exam5$exhaustion_fried[mesa_exam5$effort5==4|mesa_exam5$getgoin5==4] <- 1
mesa_exam5$exhaustion_fried[mesa_exam5$effort5<4&mesa_exam5$getgoin5<4] <- 0


#### Resistance

mesa_exam5$resistance_frail[mesa_exam5$hilwalk5 %in% c(0,1)] <- mesa_exam5$hilwalk5[mesa_exam5$hilwalk5 %in% c(0,1)]

#### Ambulation

mesa_exam5$ambulate_frail[mesa_exam5$levwalk5 %in% c(0,1)] <- mesa_exam5$levwalk5[mesa_exam5$levwalk5 %in% c(0,1)]


# FRAIL Illness score counts presence of HTN, DM, cancer, chronic lung disease, MI, CHD, angina, asthma, arthritis, stroke, and ckd
# Scores for 0-4 conditions, score = 0, for ≥ 5, score=1

# DM

mesa_exam5$dm_frail <- 0
mesa_exam5$dm_frail[mesa_exam5$dm035c %in% c(2,3)|
                      mesa_exam5$diab5==1|
                      mesa_exam5$diabhx5==1|
                      (mesa_exam5$diab==1&mesa_exam5$diabtt&mesa_exam5$e15dyc)] <- 1

# HTN

mesa_exam5$htn_frail <- 0
mesa_exam5$htn_frail[mesa_exam5$htn5c ==1|
                       mesa_exam5$highbp1==1|
                       mesa_exam5$htnmed5c==1] <- 1

# COPD (chronic lung disease)

mesa_exam5$copd_frail <- 0
mesa_exam5$copd_frail[mesa_exam5$emphys5==1|
                        (mesa_exam5$copd==1&mesa_exam5$copdtt <= mesa_exam5$e15dyc)] <- 1

# asthma

mesa_exam5$asthma_frail <- 0
mesa_exam5$asthma_frail[mesa_exam5$asthma5==1|
                          (mesa_exam5$asthma==1&mesa_exam5$asthmatt <= mesa_exam5$e15dyc)] <- 1

# DJD (arthritis)

mesa_exam5$djd_frail <- 0
mesa_exam5$djd_frail[mesa_exam5$arth2wk5==1|
                       mesa_exam5$arthrit1==1] <- 1

# CKD (renal disease)

mesa_exam5$ckd_frail <- 0
mesa_exam5$ckd_frail[mesa_exam5$kdnydis5==1|
                       (mesa_exam5$chkdds==1&mesa_exam5$chkddstt <= mesa_exam5$e15dyc)] <- 1


# CHF/MI

mesa_exam5$chf_frail <- 0
mesa_exam5$chf_frail[mesa_exam5$CHF==1&mesa_exam5$CHFTT <= mesa_exam5$e15dyc] <- 1

# CAD

# Using 'all' CHD events from MESA as this includes angina which is specified in the FRAIL criteria

mesa_exam5$chd_frail <- 0
mesa_exam5$chd_frail[mesa_exam5$CHDA==1&mesa_exam5$CHDATT <= mesa_exam5$e15dyc] <- 1

# Stroke

mesa_exam5$stroke_frail <- 0
mesa_exam5$stroke_frail[mesa_exam5$STRK==1&mesa_exam5$STRKTYPE <= mesa_exam5$e15dyc] <- 1

# Cancer

mesa_exam5$cancer_frail <- 0
mesa_exam5$cancer_frail[mesa_exam5$cancer1==1|
                          mesa_exam5$cancer==1&mesa_exam5$cancertt <= mesa_exam5$e15dyc] <- 1

# Compile chronic illness

mesa_exam5$conditions_frail <- mesa_exam5$dm_frail+
  mesa_exam5$htn_frail+
  mesa_exam5$copd_frail+
  mesa_exam5$asthma_frail+
  mesa_exam5$djd_frail+
  mesa_exam5$ckd_frail+
  mesa_exam5$chf_frail+
  mesa_exam5$chd_frail+
  mesa_exam5$stroke_frail+
  mesa_exam5$cancer_frail

mesa_exam5$illness_frail[mesa_exam5$conditions_frail <= 4] <- 0
mesa_exam5$illness_frail[mesa_exam5$conditions_frail > 4] <- 1

#### weight Loss

# Supposed to be ~ 10 lbs in past year, but mesa visits are further apart
# Instead will use lowest quintile for each interval, since each interval is different length


mesa_exam5$wtlb5_4_delta <- mesa_exam5$wtlb5-mesa_exam5$wtlb4
quantile(mesa_exam5$wtlb5_4_delta,probs=seq(0,1,0.2),na.rm=T)

# For exam 4 to 5, lowest quintile is ~ -10 lbs

mesa_exam5$wtloss_frail[mesa_exam5$wtlb5_4_delta >= -10] <- 0
mesa_exam5$wtloss_frail[mesa_exam5$wtlb5_4_delta < -10] <- 1

#### Calculate FRAIL score

mesa_exam5$total_frail <- mesa_exam5$exhaustion_fried+
  mesa_exam5$resistance_frail+
  mesa_exam5$ambulate_frail+
  mesa_exam5$illness_frail+
  mesa_exam5$wtloss_frail

######.MESA Physical activity 

mesa_exam5$sed_act_hr_day <- 
  (mesa_exam5$q16ilcn5+         # LEISURE TV MIN/WK
     mesa_exam5$q17ilcn5)/60/7    # LEISURE READING MIN/WK

mesa_exam5$sed_act_met_hr_wk <- 
  (mesa_exam5$q16ilcm5+         # LEISURE TV MET MIN/WK
     mesa_exam5$q17ilcm5)/60       # LEISURE READING MIN/WK

mesa_exam5$light_act_hr_day <-
  (mesa_exam5$q01hlcn5+         # LIGHT HOUSEHOLD CHORES MIN/WK
     mesa_exam5$q05olcn5 +      # LIGHT CARE OTHERS MIN/WK
     mesa_exam5$q20jlcn5 +      # LIGHT WORK SITTING MIN/WK
     mesa_exam5$q21jlcn5)/60/7  # LIGHT WORK STANDING MIN/WK


mesa_exam5$light_act_met_hr_wk <-
  (mesa_exam5$q01hlcm5+         # LIGHT HOUSEHOLD CHORES MIN/WK
     mesa_exam5$q05olcm5 +         # LIGHT CARE OTHERS MIN/WK
     mesa_exam5$q20jlcm5 +       # LIGHT WORK SITTING MIN/WK
     mesa_exam5$q21jlcm5)/60     # LIGHT WORK STANDING MIN/WK


mesa_exam5$mod_act_hr_day <-
  (mesa_exam5$q02hmcn5 +        # MODERATE HOUSEHOLD CHORES MIN/WK
     mesa_exam5$q03ymcn5 +      # MODERATE YARD WORK MIN/WK
     mesa_exam5$q06omcn5 +      # MODERATE CARE OTHERS MIN/WK
     mesa_exam5$q08wmcn5 +      # MODERATE WALKING MIN/WK
     mesa_exam5$q09wmcn5 +      # MODERATE WALKING EXERCISE MIN/WK
     mesa_exam5$q10smcn5 +      # MODERATE DANCE MIN/WK
     mesa_exam5$q13smcn5 +      # MODERATE INDIVIDUAL ACTIVITIES MIN/WK
     mesa_exam5$q14cmcn5 +      # MODERATE CONDITIONING MIN/WK
     mesa_exam5$q22jmcn5)/60/7  # MODERATE WORK MIN/WK


mesa_exam5$mod_act_met_hr_wk <-
  (mesa_exam5$q02hmcm5 +        # MODERATE HOUSEHOLD CHORES MET MIN/WK
     mesa_exam5$q03ymcm5 +         # MODERATE YARD WORK MET MIN/WK
     mesa_exam5$q06omcm5 +         # MODERATE CARE OTHERS MET MIN/WK
     mesa_exam5$q08wmcm5 +         # MODERATE WALKING MET MIN/WK
     mesa_exam5$q09wmcm5 +         # MODERATE WALKING EXERCISE MET MIN/WK
     mesa_exam5$q10smcm5 +         # MODERATE DANCE MET MIN/WK
     mesa_exam5$q13smcm5 +         # MODERATE INDIVIDUAL MET ACTIVITIES MIN/WK
     mesa_exam5$q14cmcm5 +         # MODERATE CONDITIONING MET MIN/WK
     mesa_exam5$q22jmcm5)/60       # MODERATE WORK MET MIN/WK


mesa_exam5$vig_act_hr_day <-
  (mesa_exam5$q04yvcn5 +        # VIGOROUS YARD WORK MIN/WK
     mesa_exam5$q11svcn5 +      # VIGOROUS TEAM SPORTS MIN/WK
     mesa_exam5$q12svcn5 +      # VIGOROUS DUAL SPORTS MIN/WK
     mesa_exam5$q15cvcn5 +      # VIGOROUS CONDITIONING MIN/WK
     mesa_exam5$q23jvcn5)/60/7  # VIGOROUS WORK MIN/WK


mesa_exam5$vig_act_met_hr_wk <-
  (mesa_exam5$q04yvcm5 +        # VIGOROUS YARD WORK MIN/WK
     mesa_exam5$q11svcm5 +         # VIGOROUS TEAM SPORTS MIN/WK
     mesa_exam5$q12svcm5 +         # VIGOROUS DUAL SPORTS MIN/WK
     mesa_exam5$q15cvcm5 +         # VIGOROUS CONDITIONING MIN/WK
     mesa_exam5$q23jvcm5)/60       # VIGOROUS WORK MIN/WK

mesa_exam5$exer_met_hr_wk <- 
  (mesa_exam5$q09wmcm5 +        # MODERATE WALKING EXERCISE MET MIN/WK
     mesa_exam5$q10smcm5 +         # MODERATE DANCE MET MIN/WK
     mesa_exam5$q11svcm5 +         # VIGOROUS TEAM SPORTS MET MIN/WK
     mesa_exam5$q12svcm5 +         # VIGOROUS DUAL SPORTS MET MIN/WK
     mesa_exam5$q13smcm5 +         # MODERATE INDIVIDUAL ACTIVITIES MET MIN/WK
     mesa_exam5$q14cmcm5 +         # MODERATE CONDITIONING MET MIN/WK
     mesa_exam5$q15cvcm5)/60       # VIGOROUS CONDITIOINING MET MIN/WK

mesa_exam5$total_met_hr_wk <-
  mesa_exam5$light_act_met_hr_wk +
  mesa_exam5$mod_act_met_hr_wk +
  mesa_exam5$vig_act_met_hr_wk

mesa_exam5$total_act_hr_day <-
  mesa_exam5$sed_act_hr_day+
  mesa_exam5$light_act_hr_day+
  mesa_exam5$mod_act_hr_day+
  mesa_exam5$vig_act_hr_day

mesa_exam5$total_act_hr_day[!between(mesa_exam5$total_act_hr_day,0,24)] <- NA


mesa_exam5$sleep_hr_day <-
  24-mesa_exam5$total_act_hr_day

mesa_exam5$total_met_hr_wk[mesa_exam5$total_met_hr_wk>450] <- NA
mesa_exam5$total_met_hr_wk[mesa_exam5$total_met_hr_wk<15] <- NA

mesa_exam5$total_met_hr_day <-
  mesa_exam5$total_met_hr_wk/7

mesa_exam5$total_act_kcal_day <-
  mesa_exam5$total_met_hr_day * 
  mesa_exam5$weight_kg * 3.5

mesa_exam5$exer_met_hr_day <-
  round(mesa_exam5$exer_met_hr_wk/7)

mesa_exam5$exer_act_kcal_day <-
  mesa_exam5$exer_met_hr_day * 
  mesa_exam5$weight_kg * 3.5


mesa_exam5$total_met_hr_wk <- 
  mesa_exam5$light_act_met_hr_wk +
  mesa_exam5$mod_act_met_hr_wk +
  mesa_exam5$vig_act_met_hr_wk

mesa_exam5$total_met_hr_day <-
  round(mesa_exam5$total_met_hr_wk/7)

mesa_exam5$total_act_kcal_day <-
  mesa_exam5$total_met_hr_day * 
  mesa_exam5$weight_kg * 3.5

mesa_exam5$exer_met_hr_day <-
  round(mesa_exam5$exer_met_hr_wk/7)

mesa_exam5$exer_act_kcal_day <-
  mesa_exam5$exer_met_hr_day * 
  mesa_exam5$weight_kg * 3.5


mesa_exam5$fhs_pai <- 
  mesa_exam5$sleep_hr_day+
  mesa_exam5$sed_act_hr_day*1.1 + 
  mesa_exam5$light_act_hr_day*1.5 + 
  mesa_exam5$mod_act_hr_day*2.4 + 
  mesa_exam5$vig_act_hr_day*5

mesa_exam5$brim_fss <-
  (5-mesa_exam5$frely5)+
  (5-mesa_exam5$fopen5)+
  mesa_exam5$fldown5+
  mesa_exam5$fdemand5


mesa_exam5_melt <- melt(subset(mesa_exam5,!is.na(visitdays)),
                        id.vars=c("mesaid","visit","visitdays","visit_yr"),
                        na.rm=T)

mesa_exam5_melt$form <- "mesa_exam5"


#===============================================================#
####                       MESA Exam 6                       ####
#===============================================================#

mesa_exam6 <- fread("~/Dropbox/BioLINCC files/MESA/Primary/Exam6/Data/mesae6_drepos_20250102.csv",na.strings=c("NA","","NULL"))


names(mesa_exam6) <- tolower(names(mesa_exam6))
mesa_exam6$visit <- 6
mesa_exam6$visit_yr <- 16
mesa_exam6$visitdays <- mesa_exam6$e16dyc
mesa_exam6$weight_kg <- mesa_exam6$wtlb6/2.2046226218

mesa_exam6 <- merge(mesa_exam6,mesa_exam5[,c("mesaid","wtlb5","arth2wk5")],by="mesaid",all.x=T)
mesa_exam6 <- merge(mesa_exam6,mesa_cvevents,by.x="mesaid",by.y="MESAID",all.x=T)
mesa_exam6 <- merge(mesa_exam6,mesa_noncvevents,by.x="mesaid",by.y="MESAID",all.x=T)

mesa_hrt <- 
  merge(mesa_hrt,
        mesa_exam6[,c("mesaid",
                      "hrmage6",
                      "hrmsage6",
                      "hrmqage6",
                      "hrmtyp6")],
        by="mesaid",
        all.x=T)

mesa_hrt$hrmage6[is.na(mesa_hrt$hrmage6)&!is.na(mesa_hrt$hrmsage6)] <-
  mesa_hrt$hrmsage6[is.na(mesa_hrt$hrmage6)&!is.na(mesa_hrt$hrmsage6)]


mesa_hrt$hrt_start_age_min <-
  apply(mesa_hrt[,c("hrmage1c",
                    "hrmage2c",
                    "hrmage3c",
                    "hrmage4c",
                    "hrmage5",
                    "hrmage6")],
        MARGIN=1,
        FUN=function(x) min(x,na.rm=T))

mesa_hrt$hrt_stop_age_max <-
  apply(mesa_hrt[,c("hrmqage1",
                    "hrmqage2",
                    "hrmqage3",
                    "hrmqage4",
                    "hrmqage5",
                    "hrmqage6")],
        MARGIN=1,
        FUN=function(x) min(x,na.rm=T))

mesa_hrt[mesa_hrt=="Inf"] <- NA
mesa_hrt[mesa_hrt=="-Inf"] <- NA

mesa_hrt$hrt_stop_age_max[!is.na(mesa_hrt$hrt_start_age_min)&
                            is.na(mesa_hrt$hrt_stop_age_max)&
                            !is.na(mesa_hrt$age_eos)] <-
  mesa_hrt$age_eos[!is.na(mesa_hrt$hrt_start_age_min)&
                     is.na(mesa_hrt$hrt_stop_age_max)&
                     !is.na(mesa_hrt$age_eos)]

mesa_hrt$hrt_years_age <-
  mesa_hrt$hrt_stop_age_max - mesa_hrt$hrt_start_age_min


mesa_exam6$dementia_dx <- 0
mesa_exam6$dementia_dx[mesa_exam6$demen==1&mesa_exam6$dementt <= mesa_exam6$e16dyc] <- 1

######### FRAIL Score, Exam 6

##### Fatigue (using FHS def - CESD everything an effort OR could not get going)

mesa_exam6$exhaustion_fried[mesa_exam6$effort5==4|mesa_exam6$getgoin5==4] <- 1
mesa_exam6$exhaustion_fried[mesa_exam6$effort5<4&mesa_exam6$getgoin5<4] <- 0


#### Resistance

mesa_exam6$resistance_frail[mesa_exam6$stairs6 %in% c(3)] <- 0
mesa_exam6$resistance_frail[mesa_exam6$stairs6 %in% c(1,2)] <- 1

#### Ambulation

mesa_exam6$ambulate_frail[mesa_exam6$levwalk6 %in% c(0,1)] <- mesa_exam6$levwalk6[mesa_exam6$levwalk6 %in% c(0,1)]


# FRAIL Illness score counts presence of HTN, DM, cancer, chronic lung disease, MI, CHD, angina, asthma, arthritis, stroke, and ckd
# Scores for 0-4 conditions, score = 0, for ≥ 5, score=1

# DM

mesa_exam6$dm_frail <- 0
mesa_exam6$dm_frail[mesa_exam6$dm036c %in% c(2,3)|
                      mesa_exam6$diab6==1|
                      mesa_exam6$diabhx6==1|
                      mesa_exam6$diabet6==1|
                      (mesa_exam6$diab6==1&mesa_exam6$diabtt < mesa_exam6$e16dyc)] <- 1

# HTN

mesa_exam6$htn_frail <- 0
mesa_exam6$htn_frail[mesa_exam6$htn5c ==1|
                       mesa_exam6$highbp1==1|
                       mesa_exam6$htnmed5c==1] <- 1

# COPD (chronic lung disease)

mesa_exam6$copd_frail <- 0
mesa_exam6$copd_frail[mesa_exam6$emphys5==1|
                        (mesa_exam6$copd==1&mesa_exam6$copdtt <= mesa_exam6$e15dyc)] <- 1

# asthma

mesa_exam6$asthma_frail <- 0
mesa_exam6$asthma_frail[mesa_exam6$asthma6==1|
                          (mesa_exam6$asthma==1&mesa_exam6$asthmatt <= mesa_exam6$e16dyc)] <- 1

# DJD (arthritis)

mesa_exam6$djd_frail <- 0
mesa_exam6$djd_frail[mesa_exam6$arth2wk5==1] <- 1

# CKD (renal disease)

mesa_exam6$ckd_frail <- 0
mesa_exam6$ckd_frail[mesa_exam6$kdnydis6==1|
                       (mesa_exam6$chkdds==1&mesa_exam6$chkddstt <= mesa_exam6$e16dyc)] <- 1

# CHF/MI

mesa_exam6$chf_frail <- 0
mesa_exam6$chf_frail[mesa_exam6$CHF==1&mesa_exam6$CHFTT <= mesa_exam6$e16dyc] <- 1

# CAD

# Using 'all' CHD events from MESA as this includes angina which is specified in the FRAIL criteria

mesa_exam6$chd_frail <- 0
mesa_exam6$chd_frail[mesa_exam6$CHDA==1&mesa_exam6$CHDATT <= mesa_exam6$e16dyc] <- 1

# Stroke

mesa_exam6$stroke_frail <- 0
mesa_exam6$stroke_frail[mesa_exam6$STRK==1&mesa_exam6$STRKTYPE <= mesa_exam6$e16dyc] <- 1

# Cancer

mesa_exam6$cancer_frail <- 0
mesa_exam6$cancer_frail[mesa_exam6$cancer==1&mesa_exam6$cancertt <= mesa_exam6$e16dyc] <- 1

# Compile chronic illness

mesa_exam6$conditions_frail <- 
  mesa_exam6$dm_frail+
  mesa_exam6$htn_frail+
  mesa_exam6$copd_frail+
  mesa_exam6$asthma_frail+
  mesa_exam6$djd_frail+
  mesa_exam6$ckd_frail+
  mesa_exam6$chf_frail+
  mesa_exam6$chd_frail+
  mesa_exam6$stroke_frail+
  mesa_exam6$cancer_frail

mesa_exam6$illness_frail[mesa_exam6$conditions_frail <= 4] <- 0
mesa_exam6$illness_frail[mesa_exam6$conditions_frail > 4] <- 1

#### weight Loss

# Supposed to be ~ 10 lbs in past year, but mesa visits are further apart
# Instead will use lowest quintile for each interval, since each interval is different length


mesa_exam6$wtlb6_5_delta <- mesa_exam6$wtlb6-mesa_exam6$wtlb5
quantile(mesa_exam6$wtlb6_5_delta,probs=seq(0,1,0.2),na.rm=T)

# For exam 5 to 6, lowest quintile is ~ -10 lbs

mesa_exam6$wtloss_frail[mesa_exam6$wtlb6_5_delta >= -10] <- 0
mesa_exam6$wtloss_frail[mesa_exam6$wtlb6_5_delta < -10] <- 1

#### Calculate FRAIL score

mesa_exam6$total_frail <- mesa_exam6$exhaustion_fried+
  mesa_exam6$resistance_frail+
  mesa_exam6$ambulate_frail+
  mesa_exam6$illness_frail+
  mesa_exam6$wtloss_frail

######.MESA Physical activity 

mesa_exam6$sed_act_hr_day <- 
  (mesa_exam6$q16ilcn6+         # LEISURE TV MIN/WK
     mesa_exam6$q17ilcn6)/60/7    # LEISURE READING MIN/WK

mesa_exam6$sed_act_met_hr_wk <- 
  (mesa_exam6$q16ilcm6+         # LEISURE TV MET MIN/WK
     mesa_exam6$q17ilcm6)/60       # LEISURE READING MIN/WK

mesa_exam6$light_act_hr_day <-
  (mesa_exam6$q01hlcn6+         # LIGHT HOUSEHOLD CHORES MIN/WK
     mesa_exam6$q05olcn6 +      # LIGHT CARE OTHERS MIN/WK
     mesa_exam6$q20jlcn6 +      # LIGHT WORK SITTING MIN/WK
     mesa_exam6$q21jlcn6)/60/7  # LIGHT WORK STANDING MIN/WK


mesa_exam6$light_act_met_hr_wk <-
  (mesa_exam6$q01hlcm6+         # LIGHT HOUSEHOLD CHORES MIN/WK
     mesa_exam6$q05olcm6 +         # LIGHT CARE OTHERS MIN/WK
     mesa_exam6$q20jlcm6 +       # LIGHT WORK SITTING MIN/WK
     mesa_exam6$q21jlcm6)/60     # LIGHT WORK STANDING MIN/WK


mesa_exam6$mod_act_hr_day <-
  (mesa_exam6$q02hmcn6 +        # MODERATE HOUSEHOLD CHORES MIN/WK
     mesa_exam6$q03ymcn6 +      # MODERATE YARD WORK MIN/WK
     mesa_exam6$q06omcn6 +      # MODERATE CARE OTHERS MIN/WK
     mesa_exam6$q08wmcn6 +      # MODERATE WALKING MIN/WK
     mesa_exam6$q09wmcn6 +      # MODERATE WALKING EXERCISE MIN/WK
     mesa_exam6$q10smcn6 +      # MODERATE DANCE MIN/WK
     mesa_exam6$q13smcn6 +      # MODERATE INDIVIDUAL ACTIVITIES MIN/WK
     mesa_exam6$q14cmcn6 +      # MODERATE CONDITIONING MIN/WK
     mesa_exam6$q22jmcn6)/60/7  # MODERATE WORK MIN/WK


mesa_exam6$mod_act_met_hr_wk <-
  (mesa_exam6$q02hmcm6 +        # MODERATE HOUSEHOLD CHORES MET MIN/WK
     mesa_exam6$q03ymcm6 +         # MODERATE YARD WORK MET MIN/WK
     mesa_exam6$q06omcm6 +         # MODERATE CARE OTHERS MET MIN/WK
     mesa_exam6$q08wmcm6 +         # MODERATE WALKING MET MIN/WK
     mesa_exam6$q09wmcm6 +         # MODERATE WALKING EXERCISE MET MIN/WK
     mesa_exam6$q10smcm6 +         # MODERATE DANCE MET MIN/WK
     mesa_exam6$q13smcm6 +         # MODERATE INDIVIDUAL MET ACTIVITIES MIN/WK
     mesa_exam6$q14cmcm6 +         # MODERATE CONDITIONING MET MIN/WK
     mesa_exam6$q22jmcm6)/60       # MODERATE WORK MET MIN/WK


mesa_exam6$vig_act_hr_day <-
  (mesa_exam6$q04yvcn5 +        # VIGOROUS YARD WORK MIN/WK
     mesa_exam6$q11svcn6 +      # VIGOROUS TEAM SPORTS MIN/WK
     mesa_exam6$q12svcn6 +      # VIGOROUS DUAL SPORTS MIN/WK
     mesa_exam6$q15cvcn6 +      # VIGOROUS CONDITIONING MIN/WK
     mesa_exam6$q23jvcn6)/60/7  # VIGOROUS WORK MIN/WK


mesa_exam6$vig_act_met_hr_wk <-
  (mesa_exam6$q04yvcm6 +        # VIGOROUS YARD WORK MIN/WK
     mesa_exam6$q11svcm6 +         # VIGOROUS TEAM SPORTS MIN/WK
     mesa_exam6$q12svcm6 +         # VIGOROUS DUAL SPORTS MIN/WK
     mesa_exam6$q15cvcm6 +         # VIGOROUS CONDITIONING MIN/WK
     mesa_exam6$q23jvcm6)/60       # VIGOROUS WORK MIN/WK

mesa_exam6$exer_met_hr_wk <- 
  (mesa_exam6$q09wmcm6 +        # MODERATE WALKING EXERCISE MET MIN/WK
     mesa_exam6$q10smcm6 +         # MODERATE DANCE MET MIN/WK
     mesa_exam6$q11svcm6 +         # VIGOROUS TEAM SPORTS MET MIN/WK
     mesa_exam6$q12svcm6 +         # VIGOROUS DUAL SPORTS MET MIN/WK
     mesa_exam6$q13smcm6 +         # MODERATE INDIVIDUAL ACTIVITIES MET MIN/WK
     mesa_exam6$q14cmcm6 +         # MODERATE CONDITIONING MET MIN/WK
     mesa_exam6$q15cvcm6)/60       # VIGOROUS CONDITIOINING MET MIN/WK

mesa_exam6$total_met_hr_wk <-
  mesa_exam6$light_act_met_hr_wk +
  mesa_exam6$mod_act_met_hr_wk +
  mesa_exam6$vig_act_met_hr_wk

mesa_exam6$total_act_hr_day <-
  mesa_exam6$sed_act_hr_day+
  mesa_exam6$light_act_hr_day+
  mesa_exam6$mod_act_hr_day+
  mesa_exam6$vig_act_hr_day

mesa_exam6$total_act_hr_day[!between(mesa_exam6$total_act_hr_day,0,24)] <- NA


mesa_exam6$sleep_hr_day <-
  24-mesa_exam6$total_act_hr_day

mesa_exam6$total_met_hr_wk[mesa_exam6$total_met_hr_wk>450] <- NA
mesa_exam6$total_met_hr_wk[mesa_exam6$total_met_hr_wk<15] <- NA

mesa_exam6$total_met_hr_day <-
  mesa_exam6$total_met_hr_wk/7

mesa_exam6$total_act_kcal_day <-
  mesa_exam6$total_met_hr_day * 
  mesa_exam6$weight_kg * 3.5

mesa_exam6$exer_met_hr_day <-
  round(mesa_exam6$exer_met_hr_wk/7)

mesa_exam6$exer_act_kcal_day <-
  mesa_exam6$exer_met_hr_day * 
  mesa_exam6$weight_kg * 3.5


mesa_exam6$total_met_hr_wk <- 
  mesa_exam6$light_act_met_hr_wk +
  mesa_exam6$mod_act_met_hr_wk +
  mesa_exam6$vig_act_met_hr_wk

mesa_exam6$total_met_hr_day <-
  round(mesa_exam6$total_met_hr_wk/7)

mesa_exam6$total_act_kcal_day <-
  mesa_exam6$total_met_hr_day * 
  mesa_exam6$weight_kg * 3.5

mesa_exam6$exer_met_hr_day <-
  round(mesa_exam6$exer_met_hr_wk/7)

mesa_exam6$exer_act_kcal_day <-
  mesa_exam6$exer_met_hr_day * 
  mesa_exam6$weight_kg * 3.5


mesa_exam6$fhs_pai <- 
  mesa_exam6$sleep_hr_day+
  mesa_exam6$sed_act_hr_day*1.1 + 
  mesa_exam6$light_act_hr_day*1.5 + 
  mesa_exam6$mod_act_hr_day*2.4 + 
  mesa_exam6$vig_act_hr_day*5

mesa_exam6_melt <- melt(subset(mesa_exam6,!is.na(visitdays)),
                        id.vars=c("mesaid","visit","visitdays","visit_yr"),
                        na.rm=T)

mesa_exam6_melt$form <- "mesa_exam6"


mesa_hrt_melt <- melt(subset(mesa_hrt),
                      id.vars=c("mesaid"),
                      na.rm=T)

mesa_hrt_melt$visit <- 1
mesa_hrt_melt$visitdays <- 0
mesa_hrt_melt$visit_yr <- 0

mesa_hrt_melt$form <- "mesa_hrt"

########################## Ancillary studies #############################


# MesaAS023RaceSeg_DS_20220111  - race segragation stats. 

mesa_raceseg <- fread("~/Dropbox/BioLINCC files/MESA/Ancillary_Studies/Ancillary_023_Neighborhood_RacialSeg/MesaAS023RaceSeg_DS_20220111.csv",
                    na.strings=c("NA","","NULL"))

names(mesa_raceseg) <- tolower(names(mesa_raceseg))

mesa_raceseg_melt <-
  melt(subset(mesa_raceseg),
     id.vars=c("mesaid","exam"),
     na.rm=T)

names(mesa_raceseg_melt)[2] <- "visit"

mesa_raceseg_melt <-
  merge(mesa_raceseg_melt,
        mesa_dates_long[,c("mesaid","visit","visit_yr","visitdays")],
        by=c("mesaid","visit"))

mesa_raceseg_melt$form <- "mesa_raceseg"

########## Air exposure 1 ##############

# mesa_airexpos_ds_20231211 - air exposure

mesa_airexpose1 <- 
  fread("~/Dropbox/BioLINCC files/MESA/Ancillary_Studies/Ancillary_039_AirExposure/mesa_airexpos_ds_20231211.csv",
        na.strings=c("NA","","NULL"))

names(mesa_airexpose1) <- tolower(names(mesa_airexpose1))

mesa_airexpose1_melt <-
  melt(subset(mesa_airexpose1),
       id.vars=c("mesaid","exam"),
       na.rm=T)

names(mesa_airexpose1_melt)[2] <- "visit"

mesa_airexpose1_melt <-
  merge(mesa_airexpose1_melt,
        mesa_dates_long[,c("mesaid","visit","visit_yr","visitdays")],
        by=c("mesaid","visit"))

mesa_airexpose1_melt$form <- "mesa_airexpose1"

########## RV function Exam 1 ##############

mesa_rvfunction <- 
  fread("~/Dropbox/BioLINCC files/MESA/Ancillary_Studies/Ancillary_067_RVfunction/mesa_20161011.csv",
        na.strings=c("NA","","NULL"))

names(mesa_rvfunction) <- tolower(names(mesa_rvfunction))

mesa_rvfunction_melt <-
  melt(subset(mesa_rvfunction),
       id.vars=c("mesaid"),
       na.rm=T)

mesa_rvfunction_melt$visit <- 1
mesa_rvfunction_melt$visit_yr <- 0
mesa_rvfunction_melt$visitdays <- 0
mesa_rvfunction_melt$form <- "mesa_rvfunction"

########## NT-proBNP Exam 1,3  ##############

mesa_ntbnp <- 
  fread("~/Dropbox/BioLINCC files/MESA/Ancillary_Studies/Ancillary_079_NTProBNP/mesaas079_drepos_20151118.csv",
        na.strings=c("NA","","NULL"))

names(mesa_ntbnp) <- tolower(names(mesa_ntbnp))

mesa_ntbnp_melt <-
  melt(subset(mesa_ntbnp),
       id.vars=c("mesaid"),
       na.rm=T)

mesa_ntbnp_melt$visit[str_detect(mesa_ntbnp_melt$variable,"1")] <- 1
mesa_ntbnp_melt$visit[str_detect(mesa_ntbnp_melt$variable,"3")] <- 3

mesa_ntbnp_melt <-
  merge(mesa_ntbnp_melt,
        mesa_dates_long[,c("mesaid","visit","visit_yr","visitdays")],
        by=c("mesaid","visit"))


mesa_ntbnp_melt$form <- "mesa_ntbnp"

########## PSG Exam 5  ##############

mesa_psg <- 
  fread("~/Dropbox/BioLINCC files/MESA/Ancillary_Studies/Ancillary_113_SleepPolysomnography/mesaas113_ps_drepos_20190301.csv",
        na.strings=c("NA","","NULL"))

names(mesa_psg) <- tolower(names(mesa_psg))

mesa_psg_melt <-
  melt(subset(mesa_psg[,c(1,46:55,79:313)]),
       id.vars=c("mesaid"),
       na.rm=T)

mesa_psg_melt$visit <- 5

mesa_psg_melt <-
  merge(mesa_psg_melt,
        mesa_dates_long[,c("mesaid","visit","visit_yr","visitdays")],
        by=c("mesaid","visit"))


mesa_psg_melt$form <- "mesa_psg"


########## Total FFA Exam 1  ##############

mesa_totffa <- 
  fread("~/Dropbox/BioLINCC files/MESA/Ancillary_Studies/Ancillary_200_TotalFFA/mesaas200_drepos_20190816.csv",
        na.strings=c("NA","","NULL"))

names(mesa_totffa) <- tolower(names(mesa_totffa))

mesa_totffa_melt <-
  melt(subset(mesa_totffa),
       id.vars=c("mesaid"),
       na.rm=T)

mesa_totffa_melt$visit <- 1
mesa_totffa_melt$visit_yr <- 0
mesa_totffa_melt$visitdays <- 0
mesa_totffa_melt$form <- "mesa_totffa"


########## FFA complete Exam 1  ##############

mesa_ffa <- 
  fread("~/Dropbox/BioLINCC files/MESA/Ancillary_Studies/Ancillary_195_FattyAcid/mesaas195_drepos_20190816.csv",
        na.strings=c("NA","","NULL"))

names(mesa_ffa) <- tolower(names(mesa_ffa))

mesa_ffa_melt <-
  melt(subset(mesa_ffa),
       id.vars=c("mesaid"),
       na.rm=T)

mesa_ffa_melt$visit <- 1
mesa_ffa_melt$visit_yr <- 0
mesa_ffa_melt$visitdays <- 0
mesa_ffa_melt$form <- "mesa_ffa"

########## LpA complete Exam 1  ##############

mesa_lpa <- 
  fread("~/Dropbox/BioLINCC files/MESA/Ancillary_Studies/Ancillary_324_LpA/mesaas324_drepos_20210910.csv",
        na.strings=c("NA","","NULL"))

names(mesa_lpa) <- tolower(names(mesa_lpa))

mesa_lpa_melt <-
  melt(subset(mesa_lpa),
       id.vars=c("mesaid"),
       na.rm=T)

mesa_lpa_melt$visit <- 1
mesa_lpa_melt$visit_yr <- 0
mesa_lpa_melt$visitdays <- 0
mesa_lpa_melt$form <- "mesa_lpa"


########## NT-proBNP HS troponin T, Exam 1,3  ##############

mesa_hstnt_ntbnp <- 
  fread("~/Dropbox/BioLINCC files/MESA/Ancillary_Studies/Ancillary_244_HScTnT_NTpBNP/mesaAS244_drepos_20161011.csv",
        na.strings=c("NA","","NULL"))

names(mesa_hstnt_ntbnp) <- tolower(names(mesa_hstnt_ntbnp))

mesa_hstnt_ntbnp_melt <-
  melt(subset(mesa_hstnt_ntbnp[,c(1:2,5,8,10)]),
       id.vars=c("mesaid"),
       na.rm=T)

mesa_hstnt_ntbnp_melt$visit[str_detect(mesa_hstnt_ntbnp_melt$variable,"1")] <- 1
mesa_hstnt_ntbnp_melt$visit[str_detect(mesa_hstnt_ntbnp_melt$variable,"3")] <- 3

mesa_hstnt_ntbnp_melt <-
  merge(mesa_hstnt_ntbnp_melt,
        mesa_dates_long[,c("mesaid","visit","visit_yr","visitdays")],
        by=c("mesaid","visit"))


mesa_hstnt_ntbnp_melt$form <- "mesa_hstnt_ntbnp"


##### **** Assemble MESA data **** #####

mesa_melt_all <- rbind(mesa_exam1_melt,
                       mesa_diet_exam1_melt,
                       mesa_exam2_melt,
                       mesa_exam3_melt,
                       mesa_exam4_melt,
                       mesa_exam5_melt,
                       mesa_hrt_melt,
                       mesa_airexpose1_melt,
                       mesa_rvfunction_melt,
                       mesa_totffa_melt,
                       mesa_ffa_melt,
                       mesa_lpa_melt,
                       mesa_psg_melt,
                       mesa_ntbnp_melt,
                       mesa_hstnt_ntbnp_melt)
                       
                       

mesa_melt_all$study <- "MESA"
names(mesa_melt_all)[1] <- "patientid"

mesa_melt_all$form <- toupper(mesa_melt_all$form)
mesa_melt_all$variable <- toupper(mesa_melt_all$variable)

mesa_melt_all$study_field <- paste(mesa_melt_all$study,
                                   mesa_melt_all$form,
                                   mesa_melt_all$variable,
                                   sep="_")

names(mesa_dates_long)[1] <- "patientid"

mesa_melt_all <-
  merge(mesa_melt_all,
        mesa_dates_long[,c("patientid","visit","age_obs","cohort","cohort_name")],
        by=c("patientid","visit"))

# mesa_melt_all <- subset(mesa_melt_all,
#                         study_field %in% 
#                           hdcp_cohort_fields_used$study_field[hdcp_cohort_fields_used$study=="MESA"])


mesa_melt_all$datapoint <-
  paste("MESA",row.names(mesa_melt_all),sep="")

mesa_melt_all$patientid <- as.character(mesa_melt_all$patientid)

library(arrow)

f <- function(input,output) {
  write.csv(input,file=output, row.names=F, na="")
}

write_parquet(mesa_melt_all[,..data_fields],
              "~/Dropbox/ADAPT-HF/Master HDCP files/Cohort data/alldata_mesa.parquet")

gcs_auth("~/Dropbox/ADAPT-HF/Master HDCP files/harmonization-286013-39f492122f69.json")
gcs_upload(mesa_melt_all[,..data_fields], 
           bucket="master_hdcp_files",
           name="mesa_melt_all.parquet",
           object_function = f)


rm(list=ls(pattern="\\bmesa."))

gc()
```
