## Supplementary material for "Aggregating data to accelerate personalized therapy in heart failure (ADAPT-HF)": Harmonization scripts: Compiled cohort dates.R

```
library(sqldf)
library(reshape2)
library(descr)
library(dplyr)
library(relper)
library(stringr)
library(bigrquery)
library(haven)
library(googleCloudStorageR)
library(parquetize)

visit_yrs <- read.csv('~/Dropbox/ADAPT-HF/Master HDCP files/Cohort visit yrs.csv')

data_fields <- 
  c("study",
    "patientid",
    "visit",
    "visit_yr",
    "visitdays",
    "form",
    "variable",
    "value",
    "datapoint",
    "study_field",
    "cohort",
    "cohort_name",
    "age_obs")

outcomes_list <- c("dth_status","dth_dt",
                   "cvdth_status","cvdth_dt",
                   "hfdth_status","hfdth_dt",
                   "hfhosp_ef_val","hfhosp_ef_cat",
                   "noncvdth_status","noncvdth_dt",
                   "hosp_status","hosp_dt",
                   "cadhosp_status","cadhosp_dt",
                   "cvhosp_status","cvhosp_dt",
                   "hfhosp_status","hfhosp_dt",
                   "noncvhosp_status","noncvhosp_dt")

dates_long_fields <-
  c("study",
    "cohort",
    "cohort_name",
    "patientid",
    "sex",
    "visit_yr",
    "visitdays",
    "age_obs")


#### FHS Dates ####


fhs1_dates <- read.csv('~/Dropbox/BioLINCC files/Framingham cohort/datasets/CSV/vr_dates_2014_a_0912d.csv',na.strings=c("NA","","NULL"))
fhs1_wkthru <- read.csv('~/Dropbox/BioLINCC files/Framingham cohort/datasets/CSV/vr_wkthru_ex32_0_0997d.csv',na.strings=c("NA","","NULL"))
fhs1_survcvd <- read.csv('~/Dropbox/BioLINCC files/Framingham cohort/datasets/CSV/vr_survcvd_2018_a_1267d.csv',na.strings=c("NA","","NULL"))

fhs1_melt_sex <- sqldf("select PID,
                       IDTYPE,
                       1 as visit,
                       'sex' as variable,
                       SEX as value,
                       'fhs1_dates' as form
                       from fhs1_dates")

names(fhs1_dates) <- toupper(names(fhs1_dates))
names(fhs1_wkthru) <- toupper(names(fhs1_wkthru))
names(fhs1_survcvd) <- toupper(names(fhs1_survcvd))


fhs2_wkthru <- read.csv('~/Dropbox/BioLINCC files/Framingham offspring/datasets/CSV/vr_wkthru_ex10_1b_1488d.csv',na.strings=c("NA","","NULL"))
fhs2_dates <- read.csv('~/Dropbox/BioLINCC files/Framingham offspring/datasets/CSV/vr_dates_2022_a_1487d.csv',na.strings=c("NA","","NULL"))
fhs2_survcvd <- read.csv('~/Dropbox/BioLINCC files/Framingham offspring/datasets/CSV/vr_survcvd_2021_a_1453d.csv',na.strings=c("NA","","NULL"))

fhs2_melt_sex <- 
  sqldf("select PID,
                IDTYPE as IDTYPE,
                1 as visit,
                'sex' as variable,
                SEX as value,
                'fhs2_dates' as form
                from fhs2_dates")


names(fhs2_dates) <- toupper(names(fhs2_dates))
names(fhs2_wkthru) <- toupper(names(fhs2_wkthru))
names(fhs2_survcvd) <- toupper(names(fhs2_survcvd))

fhs3_wkthru <- read.csv('~/Dropbox/BioLINCC files/Framingham third generation/datasets/CSV/vr_wkthru_ex03_3b_1191d.csv',na.strings=c("NA","","NULL"))
fhs3_dates <- read.csv('~/Dropbox/BioLINCC files/Framingham third generation/datasets/CSV/vr_dates_2019_a_1175d.csv',na.strings=c("NA","","NULL"))
fhs3_survcvd <- read.csv('~/Dropbox/BioLINCC files/Framingham third generation/datasets/CSV/vr_survcvd_2019_a_1334d.csv',na.strings=c("NA","","NULL"))

names(fhs3_dates) <- toupper(names(fhs3_dates))
names(fhs3_wkthru) <- toupper(names(fhs3_wkthru))
names(fhs3_survcvd) <- toupper(names(fhs3_survcvd))

fhs3_melt_sex <- sqldf("select PID,
                       IDTYPE,
                       1 as visit,
                       'sex' as variable,
                       SEX as value,
                       'fhs3_dates' as form
                       from fhs3_dates")

fhs_dates_fields <- names(fhs1_dates)
fhs2_dates[,fhs_dates_fields[!fhs_dates_fields %in% names(fhs2_dates)]] <- NA
fhs3_dates[,fhs_dates_fields[!fhs_dates_fields %in% names(fhs3_dates)]] <- NA


fhs_dates <- rbind(fhs1_dates[fhs_dates_fields],
                   fhs2_dates[fhs_dates_fields],
                   fhs3_dates[fhs_dates_fields])

fhs_survcvd <-
  rbind(fhs1_survcvd,
        fhs2_survcvd,
        fhs3_survcvd)

fhs_dates <- merge(fhs_dates,
                   fhs_survcvd,
                   by=c("IDTYPE","PID"),
                   all.x=T)
fhs_dates$DATE1 <- 0

for (i in 2:20) {
  thisexamdate <- paste("DATE",i,sep="")
  fhs_dates[(fhs_dates$hfhosp_status==1&
               fhs_dates$hfhosp_dt > fhs_dates[,thisexamdate]&
               !is.na(fhs_dates$hfhosp_dt)&
               !is.na(fhs_dates[,thisexamdate])),'last_exam_before_hf']  <- i
}

age_cols <- names(fhs_dates)[grep("AGE", names(fhs_dates))]
date_cols <- names(fhs_dates)[grep("DATE", names(fhs_dates))]

fhs_dates_long_age <- melt(fhs_dates[,c("PID","IDTYPE","SEX",age_cols)],
                           id.vars=c("PID","IDTYPE","SEX"),
                           na.rm=T,
                           factorsAsStrings = T)

fhs_dates_long_age$variable <- as.character(fhs_dates_long_age$variable)

fhs_dates_long_age$visit <- 
  substring(fhs_dates_long_age$variable,
            str_locate(fhs_dates_long_age$variable,
                       "\\d")[1],length(fhs_dates_long_age$variable))

fhs_dates_long_age$variable <- 
  gsub("\\d","",fhs_dates_long_age$variable)

###########################3


fhs_dates_long_date <- 
  melt(fhs_dates[,c("PID","IDTYPE",date_cols)],
       id.vars=c("PID","IDTYPE"),
       na.rm=T,
       factorsAsStrings = T)

fhs_dates_long_date$variable <- as.character(fhs_dates_long_date$variable)

fhs_dates_long_date <- subset(fhs_dates_long_date,!variable=="CHFDATE")

fhs_dates_long_date$visit <- 
  substring(fhs_dates_long_date$variable,
            str_locate(fhs_dates_long_date$variable,
                       "\\d")[1],length(fhs_dates_long_date$variable))

fhs_dates_long_date$variable <- gsub("\\d","",fhs_dates_long_date$variable)

names(fhs_dates_long_date)[names(fhs_dates_long_date)=="value"] <- "visitdays"
names(fhs_dates_long_age)[names(fhs_dates_long_age)=="value"] <- "age_obs"

fhs_dates_long <- merge(fhs_dates_long_age[,c("PID","IDTYPE","SEX","visit","age_obs")],
                        fhs_dates_long_date[,c("PID","IDTYPE","visit","visitdays")],
                        by=c("PID","IDTYPE","visit"))

fhs_dates_long$study <- "FHS"

names(fhs_dates_long)[2] <- "cohort"

fhs_dates_long <- 
  merge(fhs_dates_long,
        visit_yrs[visit_yrs$study=="FHS",
                  c("cohort","cohort_name","visit","visit_yr")])


names(fhs_dates_long)[3:4]<-
  c("patientid","sex")

fhs_dates_long$sex[fhs_dates_long$sex==1] <- "Male"
fhs_dates_long$sex[fhs_dates_long$sex==2] <- "Female"

rm(fhs_dates_long_age,
   fhs_dates_long_date)


#### ARIC Dates ####


aric_derive1 <- read.csv('~/Dropbox/BioLINCC files/ARIC/Main_Study/V1/csv/derive13.csv',na.strings=c("NA","","NULL"))
aric_derive2 <- read.csv('~/Dropbox/BioLINCC files/ARIC/Main_Study/V2/csv/derive2_10.csv',na.strings=c("NA","","NULL"))
aric_derive3 <- read.csv('~/Dropbox/BioLINCC files/ARIC/Main_Study/V3/csv/derive37.csv',na.strings=c("NA","","NULL"))
aric_derive4 <- read.csv('~/Dropbox/BioLINCC files/ARIC/Main_Study/V4/csv/derive47.csv',na.strings=c("NA","","NULL"))
aric_derive5 <- read.csv('~/Dropbox/BioLINCC files/ARIC/Main_Study/V5/csv/derive51.csv',na.strings=c("NA","","NULL"))
aric_derive6 <- read.csv('~/Dropbox/BioLINCC files/ARIC/Main_Study/V6/csv/derive61.csv',na.strings=c("NA","","NULL"))
aric_derive7 <- read.csv('~/Dropbox/BioLINCC files/ARIC/Main_Study/V7/csv/derive71.csv',na.strings=c("NA","","NULL"))
aric_derive8 <- read.csv('~/Dropbox/BioLINCC files/ARIC/Main_Study/V8T/csv/derive8t1_np.csv',na.strings=c("NA","","NULL"))

aric_dates <- merge(aric_derive1[,c("ID_C","CENTERID","ENROLL_YR","PREVHF01","V1AGE01")],
                    aric_derive2[,c("ID_C","V2DAYS","V2AGE22")],all.x=T)
aric_dates <- merge(aric_dates,aric_derive3[,c("ID_C","V3DAYS","V3AGE31")],all.x=T)
aric_dates <- merge(aric_dates,aric_derive4[,c("ID_C","V4DAYS","V4AGE41")],all.x=T)
aric_dates <- merge(aric_dates,aric_derive5[,c("ID_C","V5DATE51_DAYS","V5AGE51")],all.x=T)
aric_dates <- merge(aric_dates,aric_derive6[,c("ID_C","V6DATE61_DAYS","V6AGE61")],all.x=T)
aric_dates <- merge(aric_dates,aric_derive7[,c("ID_C","V7DATE71_FOLLOWUPDAYS","V7AGE72")],all.x=T)
aric_dates <- merge(aric_dates,aric_derive8[,c("ID_C","V8TDATE8T1_FOLLOWUPDAYS","V8TAGE8T1")],all.x=T)

aric_dates$cohort <- aric_dates$CENTERID

aric_dates$cohort_name[aric_dates$cohort=="A"] <- "MN"
aric_dates$cohort_name[aric_dates$cohort=="B"] <- "MD"
aric_dates$cohort_name[aric_dates$cohort=="C"] <- "MI"
aric_dates$cohort_name[aric_dates$cohort=="D"] <- "NC"


aric_dates1_long <- sqldf("select ID_C,
                          1 as visit,
                          0 as visitdays,
                          V1AGE01 as age_obs
                          from aric_derive1")

aric_dates2_long <- sqldf("select ID_C,
                          2 as visit,
                          V2DAYS as visitdays,
                          V2AGE22 as age_obs
                          from aric_derive2")

aric_dates3_long <- sqldf("select ID_C,
                          3 as visit,
                          V3DAYS as visitdays,
                          V3AGE31 as age_obs
                          from aric_derive3")

aric_dates4_long <- sqldf("select ID_C,
                          4 as visit,
                          V4DAYS as visitdays,
                          V4AGE41 as age_obs
                          from aric_derive4")

aric_dates5_long <- sqldf("select ID_C,
                          5 as visit,
                          V5DATE51_DAYS as visitdays,
                          V5AGE51 as age_obs
                          from aric_derive5")

aric_dates6_long <- sqldf("select ID_C,
                          6 as visit,
                          V6DATE61_DAYS as visitdays,
                          V6AGE61 as age_obs
                          from aric_derive6")

aric_dates7_long <- sqldf("select ID_C,
                          7 as visit,
                          V7DATE71_FOLLOWUPDAYS as visitdays,
                          V7AGE72 as age_obs
                          from aric_derive7")

aric_dates8_long <- sqldf("select ID_C,
                          8 as visit,
                          V8TDATE8T1_FOLLOWUPDAYS as visitdays,
                          V8TAGE8T1 as age_obs
                          from aric_derive8")


aric_dates_long <- rbind(aric_dates1_long,
                         aric_dates2_long,
                         aric_dates3_long,
                         aric_dates4_long,
                         aric_dates5_long,
                         aric_dates6_long,
                         aric_dates7_long,
                         aric_dates8_long)


rm(aric_dates1_long,
   aric_dates2_long,
   aric_dates3_long,
   aric_dates4_long,
   aric_dates5_long,
   aric_dates6_long,
   aric_dates7_long,
   aric_dates8_long)


aric_dates_long <- merge(aric_dates_long,
                         aric_derive1[,c("ID_C",
                                         "CENTERID",
                                         "GENDER")],
                         by="ID_C",
                         all.x=T)

names(aric_dates_long) <-
  c("patientid",
    "visit",
    "visitdays",
    "age_obs",
    "cohort",
    "sex")

aric_dates_long <- 
  merge(aric_dates_long,
        subset(visit_yrs,
               study=="ARIC"&
                 cohort=="A"))

aric_dates_long$sex[aric_dates_long$sex=="M"] <- "Male"
aric_dates_long$sex[aric_dates_long$sex=="F"] <- "Female"


#### CHS Dates ####

#### Baseboth ###

chs_baseboth <- 
  read.csv("~/Dropbox/BioLINCC files/CHS/Main_Study/BASELINE/baseboth.csv",
           na.strings=c("NA","","NULL"))

names(chs_baseboth) <- toupper(names(chs_baseboth))

chs_baseboth$agey <- 2*(chs_baseboth$AGE2-1)+64

chs_melt_baseboth <- unique(chs_baseboth[,c("NEWID","PERSTAT")])
chs_melt_baseboth$STDYTIME <- 0
chs_melt_baseboth$visit <- 1


#### CHS Year 3 ###

chs_yr3 <- read.csv("~/Dropbox/BioLINCC files/CHS/Main_Study/YEAR3/yr3.csv",na.strings=c("NA","","NULL"))
names(chs_yr3) <- toupper(names(chs_yr3))

chs_yr3$PERSTAT <- 1
chs_melt_yr3 <- 
  unique(chs_yr3[
    !is.na(chs_yr3$STDYTIME),
    c("NEWID","STDYTIME","PERSTAT")])
# names(chs_melt_yr3)[1:2] <- c("patientid","visitdays")
chs_melt_yr3$visit <- 3


#-----------------------------------------------------------------------------------#

#### CHS Year 4 ###

chs_yr4 <- read.csv("~/Dropbox/BioLINCC files/CHS/Main_Study/YEAR4/yr4.csv",na.strings=c("NA","","NULL"))
names(chs_yr4) <- toupper(names(chs_yr4))

chs_yr4$PERSTAT <- 1
chs_melt_yr4 <- 
  unique(chs_yr4[
    !is.na(chs_yr4$STDYTIME),
    c("NEWID","STDYTIME","PERSTAT")])

# names(chs_melt_yr4)[1:2] <- c("patientid","visitdays")
chs_melt_yr4$visit <- 4


#----------------------------------------------------------------#

#### CHS Year 5 - AA PERSTAT ###

chs_yr5new <- read.csv("~/Dropbox/BioLINCC files/CHS/Main_Study/YEAR5/yr5new.csv",
                       na.strings=c("NA","","NULL"))
names(chs_yr5new) <- toupper(names(chs_yr5new))

chs_yr5new$STDYTIME <- 0

chs_melt_yr5new <- 
  unique(chs_yr5new[
    !is.na(chs_yr5new$PERSTAT),
    c("NEWID","STDYTIME","PERSTAT")])

# names(chs_melt_yr5new)[1:2] <- c("patientid","visitdays")

chs_melt_yr5new$visit <- 1


#### CHS Year 5 - Original PERSTAT ###

chs_yr5old <- read.csv("~/Dropbox/BioLINCC files/CHS/Main_Study/YEAR5/yr5old.csv",na.strings=c("NA","","NULL"))
names(chs_yr5old) <- toupper(names(chs_yr5old))


chs_melt_yr5old <- 
  unique(chs_yr5old[
    !is.na(chs_yr5old$STDYTIME),
    c("NEWID","STDYTIME","PERSTAT")])

# names(chs_melt_yr5old)[1:2] <- c("patientid","visitdays")
chs_melt_yr5old$visit <- 5


#### CHS Year 6 ###


chs_yr6 <- read.csv("~/Dropbox/BioLINCC files/CHS/Main_Study/YEAR6/yr6.csv",na.strings=c("NA","","NULL"))
names(chs_yr6) <- toupper(names(chs_yr6))

chs_melt_yr6 <- 
  unique(chs_yr6[
    !is.na(chs_yr6$STDYTIME),
    c("NEWID","STDYTIME","PERSTAT")])

# names(chs_melt_yr6)[1:2] <- c("patientid","visitdays")
chs_melt_yr6$visit <- 6


#### CHS Year 7 ###


chs_yr7 <- read.csv("~/Dropbox/BioLINCC files/CHS/Main_Study/YEAR7/yr7.csv",na.strings=c("NA","","NULL"))
names(chs_yr7) <- toupper(names(chs_yr7))


chs_melt_yr7 <- 
  unique(chs_yr7[
    !is.na(chs_yr7$STDYTIME),
    c("NEWID","STDYTIME","PERSTAT")])

# names(chs_melt_yr7)[1:2] <- c("patientid","visitdays")
chs_melt_yr7$visit <- 7


#----------------------------------------------------------------------------------------------------------------------#

#### CHS Year 8 ###

chs_yr8 <- read.csv("~/Dropbox/BioLINCC files/CHS/Main_Study/YEAR8/yr8.csv",na.strings=c("NA","","NULL"))
names(chs_yr8) <- toupper(names(chs_yr8))


chs_melt_yr8 <- 
  unique(chs_yr8[
    !is.na(chs_yr8$STDYTIME),
    c("NEWID","STDYTIME","PERSTAT")])

chs_melt_yr8$visit <- 8


#----------------------------------------------------------------------------------------------------------------------#

#### CHS Year 9 ###

chs_yr9 <- read.csv("~/Dropbox/BioLINCC files/CHS/Main_Study/YEAR9/yr9.csv",na.strings=c("NA","","NULL"))
names(chs_yr9) <- toupper(names(chs_yr9))

chs_melt_yr9 <- 
  unique(chs_yr9[
    !is.na(chs_yr9$STDYTIME),
    c("NEWID","STDYTIME","PERSTAT")])

chs_melt_yr9$visit <- 9

#----------------------------------------------------------------------------------------------------------------------#

#### CHS Year 10 ###

chs_yr10 <- read.csv("~/Dropbox/BioLINCC files/CHS/Main_Study/YEAR10/yr10.csv",na.strings=c("NA","","NULL"))
names(chs_yr10) <- toupper(names(chs_yr10))

chs_melt_yr10 <- 
  unique(chs_yr10[
    !is.na(chs_yr10$STDYTIME),
    c("NEWID","STDYTIME","PERSTAT")])

chs_melt_yr10$visit <- 10


#----------------------------------------------------------------------------------------------------------------------#

#### CHS Year 11 ###

chs_yr11<- read.csv("~/Dropbox/BioLINCC files/CHS/Main_Study/YEAR11/yr11.csv",na.strings=c("NA","","NULL"))
names(chs_yr11) <- toupper(names(chs_yr11))

chs_melt_yr11 <- 
  unique(chs_yr11[
    !is.na(chs_yr11$STDYTIME),
    c("NEWID","STDYTIME","PERSTAT")])

chs_melt_yr11$visit <- 11


#----------------------------------------------------------------------------------------------------------------------#

#### CHS Year 12 ###

chs_yr12 <- read.csv("~/Dropbox/BioLINCC files/CHS/Main_Study/YEAR12/yr12ann.csv",na.strings=c("NA","","NULL"))
names(chs_yr12) <- toupper(names(chs_yr12))


chs_melt_yr12 <- 
  unique(chs_yr12[
    !is.na(chs_yr12$STDYTIME),
    c("NEWID","STDYTIME","PERSTAT")])

chs_melt_yr12$visit <- 12


#----------------------------------------------------------------------------------------------------------------------#

#### CHS Year 13 ###

chs_yr13 <- read.csv("~/Dropbox/BioLINCC files/CHS/Main_Study/YEAR13/yr13ann.csv",na.strings=c("NA","","NULL"))
names(chs_yr13) <- toupper(names(chs_yr13))

chs_melt_yr13 <- 
  unique(chs_yr13[
    !is.na(chs_yr13$STDYTIME),
    c("NEWID","STDYTIME","PERSTAT")])

chs_melt_yr13$visit <- 13


#----------------------------------------------------------------------------------------------------------------------#

#### CHS year 14 ###

chs_yr14 <- read.csv("~/Dropbox/BioLINCC files/CHS/Main_Study/YEAR14/yr14ann.csv",na.strings=c("NA","","NULL"))
names(chs_yr14) <- toupper(names(chs_yr14))

chs_melt_yr14 <- 
  unique(chs_yr14[
    !is.na(chs_yr14$STDYTIME),
    c("NEWID","STDYTIME","PERSTAT")])

chs_melt_yr14$visit <- 14


#----------------------------------------------------------------------------------------------------------------------#

#### CHS year 15 ###


chs_yr15 <- read.csv("~/Dropbox/BioLINCC files/CHS/Main_Study/YEAR15/yr15ann.csv",na.strings=c("NA","","NULL"))

names(chs_yr15) <- toupper(names(chs_yr15))


chs_melt_yr15 <- 
  unique(chs_yr15[
    !is.na(chs_yr15$STDYTIME),
    c("NEWID","STDYTIME","PERSTAT")])

chs_melt_yr15$visit <- 15


#----------------------------------------------------------------------------------------------------------------------#


#### CHS year 16 ###

chs_yr16 <- read.csv("~/Dropbox/BioLINCC files/CHS/Main_Study/YEAR16/yr16ann.csv",na.strings=c("NA","","NULL"))
names(chs_yr16) <- toupper(names(chs_yr16))


chs_melt_yr16 <- 
  unique(chs_yr16[
    !is.na(chs_yr16$STDYTIME),
    c("NEWID","STDYTIME","PERSTAT")])

chs_melt_yr16$visit <- 16


#----------------------------------------------------------------------------------------------------------------------#

#### CHS year 17 ###


chs_yr17 <- read.csv("~/Dropbox/BioLINCC files/CHS/Main_Study/YEAR17/yr17ann.csv",na.strings=c("NA","","NULL"))
names(chs_yr17) <- toupper(names(chs_yr17))


chs_melt_yr17 <- 
  unique(chs_yr17[
    !is.na(chs_yr17$STDYTIME),
    c("NEWID","STDYTIME","PERSTAT")])

chs_melt_yr17$visit <- 17


#----------------------------------------------------------------------------------------------------------------------#

#### CHS year 18 ###

chs_yr18 <- read.csv("~/Dropbox/BioLINCC files/CHS/Main_Study/YEAR18/yr18ann.csv",na.strings=c("NA","","NULL"))
names(chs_yr18) <- toupper(names(chs_yr18))

chs_melt_yr18 <- 
  unique(chs_yr18[
    !is.na(chs_yr18$STDYTIME),
    c("NEWID","STDYTIME","PERSTAT")])

chs_melt_yr18$visit <- 18


#----------------------------------------------------------------------------------------------------------------------#

#### CHS year 19 ###

chs_yr19 <- read.csv("~/Dropbox/BioLINCC files/CHS/Main_Study/YEAR19/yr19ann.csv",na.strings=c("NA","","NULL"))
names(chs_yr19) <- toupper(names(chs_yr19))

chs_melt_yr19 <- 
  unique(chs_yr19[
    !is.na(chs_yr19$STDYTIME),
    c("NEWID","STDYTIME","PERSTAT")])

chs_melt_yr19$visit <- 19


#----------------------------------------------------------------------------------------------------------------------#

#### CHS year 20 ###

chs_yr20 <- read.csv("~/Dropbox/BioLINCC files/CHS/Main_Study/YEAR20/yr20ann.csv",na.strings=c("NA","","NULL"))
names(chs_yr20) <- toupper(names(chs_yr20))

chs_melt_yr20 <- 
  unique(chs_yr20[
    !is.na(chs_yr20$STDYTIME),
    c("NEWID","STDYTIME","PERSTAT")])

chs_melt_yr20$visit <- 20


#----------------------------------------------------------------------------------------------------------------------#

#### CHS year 21 ###

chs_yr21 <- read.csv("~/Dropbox/BioLINCC files/CHS/Main_Study/YEAR21/yr21ann.csv",na.strings=c("NA","","NULL"))
names(chs_yr21) <- toupper(names(chs_yr21))

chs_melt_yr21 <- 
  unique(chs_yr21[
    !is.na(chs_yr21$STDYTIME),
    c("NEWID","STDYTIME","PERSTAT")])

chs_melt_yr21$visit <- 21


#----------------------------------------------------------------------------------------------------------------------#


### CHS year 22 ###

chs_yr22 <- read.csv("~/Dropbox/BioLINCC files/CHS/Main_Study/YEAR22/yr22ann.csv",na.strings=c("NA","","NULL"))
names(chs_yr22) <- toupper(names(chs_yr22))

chs_melt_yr22 <- 
  unique(chs_yr22[
    !is.na(chs_yr22$STDYTIME),
    c("NEWID","STDYTIME","PERSTAT")])

chs_melt_yr22$visit <- 22

#----------------------------------------------------------------------------------------------------------------------#

### CHS year 23 ###

chs_yr23 <- read.csv("~/Dropbox/BioLINCC files/CHS/Main_Study/YEAR23/yr23ann.csv",na.strings=c("NA","","NULL"))
names(chs_yr23) <- toupper(names(chs_yr23))

chs_melt_yr23 <- 
  unique(chs_yr23[
    !is.na(chs_yr23$STDYTIME),
    c("NEWID","STDYTIME","PERSTAT")])

chs_melt_yr23$visit <- 23


#----------------------------------------------------------------------------------------------------------------------#

### CHS year 24 ###

chs_yr24 <- read.csv("~/Dropbox/BioLINCC files/CHS/Main_Study/YEAR24/yr24ann.csv",na.strings=c("NA","","NULL"))
names(chs_yr24) <- toupper(names(chs_yr24))

chs_melt_yr24 <- 
  unique(chs_yr24[
    !is.na(chs_yr24$STDYTIME),
    c("NEWID","STDYTIME","PERSTAT")])

chs_melt_yr24$visit <- 24


#----------------------------------------------------------------------------------------------------------------------#


### CHS year 25 ###

chs_yr25 <- read.csv("~/Dropbox/BioLINCC files/CHS/Main_Study/YEAR25/yr25ann.csv",na.strings=c("NA","","NULL"))
names(chs_yr25) <- toupper(names(chs_yr25))

chs_melt_yr25 <- 
  unique(chs_yr25[
    !is.na(chs_yr25$STDYTIME),
    c("NEWID","STDYTIME","PERSTAT")])

chs_melt_yr25$visit <- 25


#----------------------------------------------------------------------------------------------------------------------#

### CHS year 26 ###

chs_yr26 <- read.csv("~/Dropbox/BioLINCC files/CHS/Main_Study/YEAR26/yr26ann.csv",na.strings=c("NA","","NULL"))
names(chs_yr26) <- toupper(names(chs_yr26))

chs_melt_yr26 <- 
  unique(chs_yr26[
    !is.na(chs_yr26$STDYTIME),
    c("NEWID","STDYTIME","PERSTAT")])

chs_melt_yr26$visit <- 26


#----------------------------------------------------------------------------------------------------------------------#

### CHS year 27 ###

chs_yr27 <- read.csv("~/Dropbox/BioLINCC files/CHS/Main_Study/YEAR27/yr27ann.csv",na.strings=c("NA","","NULL"))
names(chs_yr27) <- toupper(names(chs_yr27))

chs_melt_yr27 <- 
  unique(chs_yr27[
    !is.na(chs_yr27$STDYTIME),
    c("NEWID","STDYTIME","PERSTAT")])

chs_melt_yr27$visit <- 27


#----------------------------------------------------------------------------------------------------------------------#

### CHS year 28 ###

chs_yr28 <- read.csv("~/Dropbox/BioLINCC files/CHS/Main_Study/YEAR28/yr28ann.csv",na.strings=c("NA","","NULL"))
names(chs_yr28) <- toupper(names(chs_yr28))

chs_melt_yr28 <- 
  unique(chs_yr28[
    !is.na(chs_yr28$STDYTIME),
    c("NEWID","STDYTIME","PERSTAT")])

chs_melt_yr28$visit <- 28


#----------------------------------------------------------------------------------------------------------------------#


### CHS year 29 ###

chs_yr29 <- read.csv("~/Dropbox/BioLINCC files/CHS/Main_Study/YEAR29/yr29ann.csv",na.strings=c("NA","","NULL"))
names(chs_yr29) <- toupper(names(chs_yr29))

chs_melt_yr29 <- 
  unique(chs_yr29[
    !is.na(chs_yr29$STDYTIME),
    c("NEWID","STDYTIME","PERSTAT")])

chs_melt_yr29$visit <- 29


#----------------------------------------------------------------------------------------------------------------------#

### CHS year 30 ###

chs_yr30 <- read.csv("~/Dropbox/BioLINCC files/CHS/Main_Study/YEAR30/yr30ann.csv",na.strings=c("NA","","NULL"))
names(chs_yr30) <- toupper(names(chs_yr30))

chs_melt_yr30 <- 
  unique(chs_yr30[
    !is.na(chs_yr30$STDYTIME),
    c("NEWID","STDYTIME","PERSTAT")])

chs_melt_yr30$visit <- 30


#----------------------------------------------------------------------------------------------------------------------#

### CHS year 31 ###

chs_yr31 <- read.csv("~/Dropbox/BioLINCC files/CHS/Main_Study/YEAR31/yr31ann.csv",na.strings=c("NA","","NULL"))
names(chs_yr31) <- toupper(names(chs_yr31))

chs_melt_yr31 <- 
  unique(chs_yr31[
    !is.na(chs_yr31$STDYTIME),
    c("NEWID","STDYTIME","PERSTAT")])

chs_melt_yr31$visit <- 31


#----------------------------------------------------------------------------------------------------------------------#


### CHS year 32 ###

chs_yr32 <- read.csv("~/Dropbox/BioLINCC files/CHS/Main_Study/YEAR32/yr32ann.csv",na.strings=c("NA","","NULL"))
names(chs_yr32) <- toupper(names(chs_yr32))

chs_melt_yr32 <- 
  unique(chs_yr32[
    !is.na(chs_yr32$STDYTIME),
    c("NEWID","STDYTIME","PERSTAT")])

chs_melt_yr32$visit <- 32


#----------------------------------------------------------------------------------------------------------------------#


### CHS year 33 ###

chs_yr33 <- read.csv("~/Dropbox/BioLINCC files/CHS/Main_Study/YEAR33/yr33ann.csv",na.strings=c("NA","","NULL"))
names(chs_yr33) <- toupper(names(chs_yr33))

chs_melt_yr33 <- 
  unique(chs_yr33[
    !is.na(chs_yr33$STDYTIME),
    c("NEWID","STDYTIME","PERSTAT")])

chs_melt_yr33$visit <- 33


#----------------------------------------------------------------------------------------------------------------------#

### CHS year 34 ###

chs_yr34 <- read.csv("~/Dropbox/BioLINCC files/CHS/Main_Study/YEAR34/yr34ann.csv",na.strings=c("NA","","NULL"))
names(chs_yr34) <- toupper(names(chs_yr34))

chs_melt_yr34 <- 
  unique(chs_yr34[
    !is.na(chs_yr34$STDYTIME),
    c("NEWID","STDYTIME","PERSTAT")])

chs_melt_yr34$visit <- 34


#----------------------------------------------------------------------------------------------------------------------#


### CHS year 35 ###

chs_yr35 <- read.csv("~/Dropbox/BioLINCC files/CHS/Main_Study/YEAR35/yr35ann.csv",na.strings=c("NA","","NULL"))
names(chs_yr35) <- toupper(names(chs_yr35))

chs_melt_yr35 <- 
  unique(chs_yr35[
    !is.na(chs_yr35$STDYTIME),
    c("NEWID","STDYTIME","PERSTAT")])

chs_melt_yr35$visit <- 35


#----------------------------------------------------------------------------------------------------------------------#


### *** CHS final assembly *** ###

chs_melt_all <- rbind(
  chs_melt_baseboth[,c("NEWID","PERSTAT","STDYTIME","visit")],
  chs_melt_yr3[,c("NEWID","PERSTAT","STDYTIME","visit")],
  chs_melt_yr4[,c("NEWID","PERSTAT","STDYTIME","visit")],
  chs_melt_yr5old[,c("NEWID","PERSTAT","STDYTIME","visit")],
  chs_melt_yr6[,c("NEWID","PERSTAT","STDYTIME","visit")],
  chs_melt_yr7[,c("NEWID","PERSTAT","STDYTIME","visit")],
  chs_melt_yr8[,c("NEWID","PERSTAT","STDYTIME","visit")],
  chs_melt_yr9[,c("NEWID","PERSTAT","STDYTIME","visit")],
  chs_melt_yr10[,c("NEWID","PERSTAT","STDYTIME","visit")],
  chs_melt_yr11[,c("NEWID","PERSTAT","STDYTIME","visit")],
  chs_melt_yr12[,c("NEWID","PERSTAT","STDYTIME","visit")],
  chs_melt_yr13[,c("NEWID","PERSTAT","STDYTIME","visit")],
  chs_melt_yr14[,c("NEWID","PERSTAT","STDYTIME","visit")],
  chs_melt_yr15[,c("NEWID","PERSTAT","STDYTIME","visit")],
  chs_melt_yr16[,c("NEWID","PERSTAT","STDYTIME","visit")],
  chs_melt_yr17[,c("NEWID","PERSTAT","STDYTIME","visit")],
  chs_melt_yr18[,c("NEWID","PERSTAT","STDYTIME","visit")],
  chs_melt_yr19[,c("NEWID","PERSTAT","STDYTIME","visit")],
  chs_melt_yr20[,c("NEWID","PERSTAT","STDYTIME","visit")],
  chs_melt_yr21[,c("NEWID","PERSTAT","STDYTIME","visit")],
  chs_melt_yr22[,c("NEWID","PERSTAT","STDYTIME","visit")],
  chs_melt_yr23[,c("NEWID","PERSTAT","STDYTIME","visit")],
  chs_melt_yr24[,c("NEWID","PERSTAT","STDYTIME","visit")],
  chs_melt_yr25[,c("NEWID","PERSTAT","STDYTIME","visit")],
  chs_melt_yr26[,c("NEWID","PERSTAT","STDYTIME","visit")],
  chs_melt_yr27[,c("NEWID","PERSTAT","STDYTIME","visit")],
  chs_melt_yr28[,c("NEWID","PERSTAT","STDYTIME","visit")],
  chs_melt_yr29[,c("NEWID","PERSTAT","STDYTIME","visit")],
  chs_melt_yr30[,c("NEWID","PERSTAT","STDYTIME","visit")],
  chs_melt_yr31[,c("NEWID","PERSTAT","STDYTIME","visit")],
  chs_melt_yr32[,c("NEWID","PERSTAT","STDYTIME","visit")],
  chs_melt_yr33[,c("NEWID","PERSTAT","STDYTIME","visit")],
  chs_melt_yr34[,c("NEWID","PERSTAT","STDYTIME","visit")],
  chs_melt_yr35[,c("NEWID","PERSTAT","STDYTIME","visit")]
)

chs_melt_all <-
  merge(chs_melt_all,
        chs_baseboth[,c("NEWID","agey","GEND01")],
        by="NEWID",
        all.x=T)

chs_melt_all$age_obs <-
  floor(chs_melt_all$agey+chs_melt_all$STDYTIME/365)

chs_melt_all$study <- "CHS"

chs_melt_all <-
  merge(chs_melt_all,
        visit_yrs,
        by.x=c("study","PERSTAT","visit"),
        by.y=c("study","cohort","visit"),
        all.x=T)

names(chs_melt_all) <-
  c("study",
    "cohort",
    "visit",
    "patientid",
    "visitdays",
    "agey",
    "sex",
    "age_obs",
    "cohort_name",
    "visit_yr")

chs_dates_long <-
  unique(chs_melt_all[,c(
    "study",
    "cohort",
    "cohort_name",
    "patientid",
    "age_obs",
    "sex",
    "visit",
    "visit_yr",
    "visitdays")])

chs_dates_long$sex[chs_dates_long$sex=="1"] <- "Male"
chs_dates_long$sex[chs_dates_long$sex=="0"] <- "Female"


#### MESA Dates ####


mesa_exam1 <- read.csv("~/Dropbox/BioLINCC files/MESA/Primary/Exam1/Data/mesae1dres20220813.csv",na.strings=c("NA","","NULL"))
mesa_exam2 <- read.csv("~/Dropbox/BioLINCC files/MESA/Primary/Exam2/Data/mesae2dres06222012.csv",na.strings=c("NA","","NULL"))
mesa_exam3 <- read.csv("~/Dropbox/BioLINCC files/MESA/Primary/Exam3/Data/mesae3dres06222012.csv",na.strings=c("NA","","NULL"))
mesa_exam4 <- read.csv("~/Dropbox/BioLINCC files/MESA/Primary/Exam4/Data/mesae4dres06222012.csv",na.strings=c("NA","","NULL"))
mesa_exam5 <- read.csv("~/Dropbox/BioLINCC files/MESA/Primary/Exam5/Data/mesae5_drepos_20220820.csv",na.strings=c("NA","","NULL"))
mesa_exam6 <- read.csv("~/Dropbox/BioLINCC files/MESA/Primary/Exam6/Data/mesae6_drepos_20250102.csv",na.strings=c("NA","","NULL"))

names(mesa_exam1) <- tolower(names(mesa_exam1))
names(mesa_exam2) <- tolower(names(mesa_exam2))
names(mesa_exam3) <- tolower(names(mesa_exam3))
names(mesa_exam4) <- tolower(names(mesa_exam4))
names(mesa_exam5) <- tolower(names(mesa_exam5))
names(mesa_exam6) <- tolower(names(mesa_exam6))


mesa_efevents <- read.csv("~/Dropbox/BioLINCC files/MESA/Primary/Events/CVD/Data/mesaevefthru2015_drepos_20200330.csv",na.strings=c("NA","","NULL"))
mesa_events <- read.csv("~/Dropbox/BioLINCC files/MESA/Primary/Events/CVD/Data/mesaevthr2020_drepos_20241120.csv",na.strings=c("NA","","NULL"))
mesa_site <- read.csv("~/Dropbox/BioLINCC files/MESA/Primary/Site/mesa_site_drepos_20181106.csv")

mesa_site$cohort_name[mesa_site$site1c=="A"] <- "MD" 
mesa_site$cohort_name[mesa_site$site1c=="B"] <- "MN" 
mesa_site$cohort_name[mesa_site$site1c=="C"] <- "IL" 
mesa_site$cohort_name[mesa_site$site1c=="D"] <- "CA" 
mesa_site$cohort_name[mesa_site$site1c=="E"] <- "NC" 
mesa_site$cohort_name[mesa_site$site1c=="F"] <- "NY" 

names(mesa_efevents) <- tolower(names(mesa_efevents))
names(mesa_events) <- tolower(names(mesa_events))

mesa_dates <- mesa_exam1[,c("mesaid","age1c")]
mesa_dates <- merge(mesa_dates, mesa_exam2[,c("mesaid","age2c","e12dyc")],all.x=T)
mesa_dates <- merge(mesa_dates, mesa_exam3[,c("mesaid","age3c","e13dyc")],all.x=T)
mesa_dates <- merge(mesa_dates, mesa_exam4[,c("mesaid","age4c","e14dyc")],all.x=T)
mesa_dates <- merge(mesa_dates, mesa_exam5[,c("mesaid","age5c","e15dyc")],all.x=T)
mesa_dates <- merge(mesa_dates, mesa_exam6[,c("mesaid","age6c","e16dyc")],all.x=T)

mesa_sex <- mesa_exam1[,c("mesaid","gender1")]

mesa_dates_1 <- mesa_exam1[,c("mesaid","age1c")]
names(mesa_dates_1)[names(mesa_dates_1)=="age1c"] <- "age_obs"
mesa_dates_1$visitdays <- 0
mesa_dates_1$visit <- 1
mesa_dates_1$visit_yr <- 0

mesa_dates_2 <-mesa_exam2[,c("mesaid","age2c","e12dyc")]
names(mesa_dates_2)[names(mesa_dates_2)=="age2c"] <- "age_obs"
names(mesa_dates_2)[names(mesa_dates_2)=="e12dyc"] <- "visitdays"
mesa_dates_2$visit_yr <- 3
mesa_dates_2$visit <- 2

mesa_dates_3 <-mesa_exam3[,c("mesaid","age3c","e13dyc")]
names(mesa_dates_3)[names(mesa_dates_3)=="age3c"] <- "age_obs"
names(mesa_dates_3)[names(mesa_dates_3)=="e13dyc"] <- "visitdays"
mesa_dates_3$visit <- 3
mesa_dates_3$visit_yr <- 5

mesa_dates_4 <-mesa_exam4[,c("mesaid","age4c","e14dyc")]
names(mesa_dates_4)[names(mesa_dates_4)=="age4c"] <- "age_obs"
names(mesa_dates_4)[names(mesa_dates_4)=="e14dyc"] <- "visitdays"
mesa_dates_4$visit <- 4
mesa_dates_4$visit_yr <- 6

mesa_dates_5 <-mesa_exam5[,c("mesaid","age5c","e15dyc")]
names(mesa_dates_5)[names(mesa_dates_5)=="age5c"] <- "age_obs"
names(mesa_dates_5)[names(mesa_dates_5)=="e15dyc"] <- "visitdays"
mesa_dates_5$visit <- 5
mesa_dates_5$visit_yr <- 10

mesa_dates_6 <-mesa_exam5[,c("mesaid","age6c","e16dyc")]
names(mesa_dates_6)[names(mesa_dates_6)=="age6c"] <- "age_obs"
names(mesa_dates_6)[names(mesa_dates_6)=="e16dyc"] <- "visitdays"
mesa_dates_6$visit <- 6
mesa_dates_6$visit_yr <- 16

mesa_dates_long <- rbind(mesa_dates_1,
                         mesa_dates_2,
                         mesa_dates_3,
                         mesa_dates_4,
                         mesa_dates_5,
                         mesa_dates_6)

mesa_dates_long <- subset(mesa_dates_long,!is.na(visitdays))

mesa_dates_long <- merge(mesa_dates_long,
                         mesa_sex,
                         by="mesaid")

mesa_dates <- merge(mesa_dates,mesa_events,by="mesaid")
mesa_dates <- merge(mesa_dates,
                    mesa_site[,c("MESAID","site1c","cohort_name")],
                    by.x="mesaid",
                    by.y="MESAID",
                    all.x=T)

mesa_dates_long <- merge(mesa_dates_long,
                         mesa_site[,c("MESAID","site1c","cohort_name")],
                         by.x="mesaid",
                         by.y="MESAID",
                         all.x=T)

mesa_dates_long$study <- "MESA"

names(mesa_dates_long)[names(mesa_dates_long)=="site1c"] <- "cohort"
names(mesa_dates_long)[names(mesa_dates_long)=="mesaid"] <- "patientid"
names(mesa_dates_long)[names(mesa_dates_long)=="gender1"] <- "sex"

mesa_dates_long$sex[mesa_dates_long$sex==1] <- "Male"
mesa_dates_long$sex[mesa_dates_long$sex==0] <- "Female"

mesa_dates$age_hf <- round(mesa_dates$age1c+mesa_dates$chftt/365)

mesa_dates$last_exam_before_hf[mesa_dates$chf==1&mesa_dates$chftt>0] <- 1
mesa_dates$last_exam_before_hf[mesa_dates$chf==1&mesa_dates$chftt>=mesa_dates$e12dyc] <- 2
mesa_dates$last_exam_before_hf[mesa_dates$chf==1&mesa_dates$chftt>=mesa_dates$e13dyc] <- 3
mesa_dates$last_exam_before_hf[mesa_dates$chf==1&mesa_dates$chftt>=mesa_dates$e14dyc] <- 4
mesa_dates$last_exam_before_hf[mesa_dates$chf==1&mesa_dates$chftt>=mesa_dates$e15dyc] <- 5
mesa_dates$last_exam_before_hf[mesa_dates$chf==1&mesa_dates$chftt>=mesa_dates$e16dyc] <- 6

mesa_dates$last_year_before_hf[mesa_dates$chf==1&mesa_dates$chftt>0] <- 0
mesa_dates$last_year_before_hf[mesa_dates$chf==1&mesa_dates$chftt>=mesa_dates$e12dyc] <- 3
mesa_dates$last_year_before_hf[mesa_dates$chf==1&mesa_dates$chftt>=mesa_dates$e13dyc] <- 4
mesa_dates$last_year_before_hf[mesa_dates$chf==1&mesa_dates$chftt>=mesa_dates$e14dyc] <- 6
mesa_dates$last_year_before_hf[mesa_dates$chf==1&mesa_dates$chftt>=mesa_dates$e15dyc] <- 10
mesa_dates$last_year_before_hf[mesa_dates$chf==1&mesa_dates$chftt>=mesa_dates$e16dyc] <- 16


mesa_dates$last_exam_date_before_hf[mesa_dates$last_exam_before_hf==1&!is.na(mesa_dates$last_exam_before_hf)] <-  0
mesa_dates$last_exam_date_before_hf[mesa_dates$last_exam_before_hf==2&!is.na(mesa_dates$last_exam_before_hf)] <-  
  mesa_dates$e12dyc[mesa_dates$last_exam_before_hf==2&!is.na(mesa_dates$last_exam_before_hf)]
mesa_dates$last_exam_date_before_hf[mesa_dates$last_exam_before_hf==3&!is.na(mesa_dates$last_exam_before_hf)] <-  
  mesa_dates$e13dyc[mesa_dates$last_exam_before_hf==3&!is.na(mesa_dates$last_exam_before_hf)]
mesa_dates$last_exam_date_before_hf[mesa_dates$last_exam_before_hf==4&!is.na(mesa_dates$last_exam_before_hf)] <-  
  mesa_dates$e14dyc[mesa_dates$last_exam_before_hf==4&!is.na(mesa_dates$last_exam_before_hf)]
mesa_dates$last_exam_date_before_hf[mesa_dates$last_exam_before_hf==5&!is.na(mesa_dates$last_exam_before_hf)] <-  
  mesa_dates$e15dyc[mesa_dates$last_exam_before_hf==5&!is.na(mesa_dates$last_exam_before_hf)]
mesa_dates$last_exam_date_before_hf[mesa_dates$last_exam_before_hf==6&!is.na(mesa_dates$last_exam_before_hf)] <-  
  mesa_dates$e16dyc[mesa_dates$last_exam_before_hf==6&!is.na(mesa_dates$last_exam_before_hf)]

mesa_dates$hf_dys_last_exam <- mesa_dates$chftt-mesa_dates$last_exam_before_hf
mesa_dates$time_hf_to_dth <- mesa_dates$dthtt-mesa_dates$chftt


#### JHS Dates ####


jhs_analysis1 <- read.csv("~/Dropbox/BioLINCC files/JHS/Data/Analysis_Data/CSV/analysis1.csv",na.strings=c("","NA","NULL"))
jhs_analysis2 <- read.csv("~/Dropbox/BioLINCC files/JHS/Data/Analysis_Data/CSV/analysis2.csv",na.strings=c("","NA","NULL"))
jhs_analysis3 <- read.csv("~/Dropbox/BioLINCC files/JHS/Data/Analysis_Data/CSV/analysis3.csv",na.strings=c("","NA","NULL"))

jhs_v1date <- 
  sqldf('select newid as patientid, 
        0 as visitdays, 
        1 as visit, 
        0 as visit_yr, 
        age as age_obs,
        ARIC as cohort
        from jhs_analysis1')

jhs_v2date <- 
  sqldf('select newid as patientid,
        DaysFromV1 as visitdays, 
        2 as visit, 
        6 as visit_yr, 
        age as age_obs,
        ARIC as cohort
        from jhs_analysis2')


jhs_v3date <- 
  sqldf('select newid as patientid, 
        DaysFromV1 as visitdays, 
        3 as visit, 
        9 as visit_yr, 
        age as age_obs,
        ARIC as cohort 
        from jhs_analysis3')

jhs_dates_long <-
  rbind(jhs_v1date,
        jhs_v2date,
        jhs_v3date)


# ==============================================================#
####             *** JHS longitudinal visits ***             ####
# ==============================================================#

## Loading and preparing annual longitudinal visits here
## Currently Visit 1 = baseline, Visit 2  = 6 years, Visit 3 = 9 years.
## afulong visit assigned by taking time since baseline rounded to nearest year (jhs_afulong$time) +1
## 1 added because Baseline visit = Visit 1
## Then remove any afulong record with assigned visit 1,6,or 9 to eliminate clashing with main study visits.


jhs_afulong <- 
  read.csv("~/Dropbox/BioLINCC files/JHS/Data/AFU/CSV/afulong.csv",
           na.strings=c("","NA","NULL"))

jhs_afulong <-
  merge(jhs_afulong[,c("newid","date","v1date","time","V2date","V3date")],
        jhs_analysis1[,c("newid","ARIC","sex","age")],
        by="newid",
        all.x=T)

jhs_afulong$visit_yr <- 
  round(jhs_afulong$time)

jhs_afulong$visitdays <- 
  as.numeric(
    difftime(
      as.Date(jhs_afulong$date,"%m/%d/%Y"),
      as.Date(jhs_afulong$v1date,"%m/%d/%Y"),
      units="days"))

jhs_afulong$age_obs <-
  floor(jhs_afulong$age + jhs_afulong$time)

jhs_afulong$visit2days <- 
  as.numeric(
    difftime(
      as.Date(jhs_afulong$date,"%m/%d/%Y"),
      as.Date(jhs_afulong$V2date,"%m/%d/%Y"),
      units="days"))

jhs_afulong$visit2years <- round(jhs_afulong$visit2days/365,0)+1

jhs_afulong$visit3days <- 
  as.numeric(
    difftime(
      as.Date(jhs_afulong$date,"%m/%d/%Y"),
      as.Date(jhs_afulong$V3date,"%m/%d/%Y"),
      units="days"))

jhs_afulong$visit3years <- round(jhs_afulong$visit3days/365,0)+1

jhs_afulong$days_from_v2 <- jhs_afulong$visitdays-jhs_afulong$visit2days
jhs_afulong$days_from_v3 <- jhs_afulong$visitdays-jhs_afulong$visit3days

jhs_afulong <- subset(jhs_afulong,
                      !visit_yr %in% c(-2,-1,0,6,9)&
                        abs(days_from_v2)>180&
                        abs(days_from_v3)>180&
                        abs(visitdays)>180)

## This statement handles a situation where multiple studies round to the same visit, resulting in multiple separate lines for that visit.
## This solution takes the MAX (or latest) observation - this can be changed to the earliest, etc.

jhs_dates_long <- sqldf("select newid as patientid,
                      age_obs,
                      visitdays,
                      visit_yr,
                      NULL as visit,
                      sex,
                      ARIC as cohort,
                      'JHS' as study
                      from jhs_afulong
                      group by newid,visit_yr
                      having max(visitdays)")

jhs_dates_long$cohort_name[jhs_dates_long$cohort==1] <- "ARIC"
jhs_dates_long$cohort_name[jhs_dates_long$cohort==0] <- "JHS"

jhs_dates_long <- 
  unique(jhs_dates_long[,c("patientid",
                        "age_obs",
                        "visitdays",
                        "visit",
                        "visit_yr",
                        "sex",
                        "cohort",
                        "cohort_name",
                        "study")])
         

all_dates_long <-
  rbind(fhs_dates_long[,dates_long_fields],
        aric_dates_long[,dates_long_fields],
        chs_dates_long[,dates_long_fields],
        mesa_dates_long[,dates_long_fields],
        jhs_dates_long[,dates_long_fields])

### Upload to BQ and GCS

bq_auth(path="harmonization-286013-39f492122f69.json")

my_bq <- dbConnect(
  bigrquery::bigquery(),
  project="harmonization",
  dataset="cohorts",
  billing="harmonization"
)


if (bq_table_exists("harmonization-286013.cohorts_update.ptlist"))
{bq_table_delete("harmonization-286013.cohorts_update.ptlist")}

bq_table_upload(x="harmonization-286013.cohorts_update.ptlist",
                all_dates_long,
                fields=all_dates_long,
                quiet=T,
                create_disposition="CREATE_IF_NEEDED",
                write_disposition="WRITE_TRUNCATE")

library(arrow)
write_parquet(all_dates_long,"Cohort data/ptlist.parquet")

gcs_auth("~/Dropbox/ADAPT-HF/Master HDCP files/harmonization-286013-39f492122f69.json")
gcs_upload("Cohort data/ptlist.parquet", 
           bucket="master_hdcp_files")
```
