## Supplementary material for "Aggregating data to accelerate personalized therapy in heart failure (ADAPT-HF)": Harmonization scripts: Transform trial acute outcomes.R

```
library(plyr)
library(readstata13)
library(Hmisc)
library(reshape2)
library(descr)
library(rmeta)
library(sas7bdat)
library(sqldf)
library(DescTools)
library(dplyr)
library(nephro)
library(googleCloudStorageR)


source("~/Dropbox/R scripts/echo scripts.R")
source("~/Dropbox/R scripts/Misc clinical scripts.R")


### outcome columns specified here for convenience.

acute_analysis_outcomes <- c('patientid',
                             'study',
                             'wt_kg_bl','wt_kg_1d', 'wt_kg_2d','wt_kg_3d','wt_kg_4d','wt_kg_last',
                            'gfr_bl','gfr_1d','gfr_2d','gfr_3d','gfr_4d','gfr_last',
                            'io_balance_1d','io_balance_2d','io_balance_3d','io_balance_4d','io_balance_5d',
                            'io_cum_2d','io_cum_3d','io_cum_4d','io_cum_5d','io_cum_6d','io_cum_7d',
                            'los',
                            'inhosp_dth_status')
                            
                            
# #######################################################################################################################################
# #######################################################################################################################################
# #######################################################################################################################################

##### ******************************************** OUTCOMES ******************************************** ####
#
# ..%%%%%%%..%%.....%%.%%%%%%%%..%%%%%%...%%%%%%%..%%.....%%.%%%%%%%%..%%%%%%.
# .%%.....%%.%%.....%%....%%....%%....%%.%%.....%%.%%%...%%%.%%.......%%....%%
# .%%.....%%.%%.....%%....%%....%%.......%%.....%%.%%%%.%%%%.%%.......%%......
# .%%.....%%.%%.....%%....%%....%%.......%%.....%%.%%.%%%.%%.%%%%%%....%%%%%%.
# .%%.....%%.%%.....%%....%%....%%.......%%.....%%.%%.....%%.%%.............%%
# .%%.....%%.%%.....%%....%%....%%....%%.%%.....%%.%%.....%%.%%.......%%....%%
# ..%%%%%%%...%%%%%%%.....%%.....%%%%%%...%%%%%%%..%%.....%%.%%%%%%%%..%%%%%%.
#
# 

# 
# ######################### ******* CARRESS ******* ############################
# 
# ..%%%%%%.....%%%....%%%%%%%%..%%%%%%%%..%%%%%%%%..%%%%%%...%%%%%%.
# .%%....%%...%%.%%...%%.....%%.%%.....%%.%%.......%%....%%.%%....%%
# .%%........%%...%%..%%.....%%.%%.....%%.%%.......%%.......%%......
# .%%.......%%.....%%.%%%%%%%%..%%%%%%%%..%%%%%%....%%%%%%...%%%%%%.
# .%%.......%%%%%%%%%.%%...%%...%%...%%...%%.............%%.......%%
# .%%....%%.%%.....%%.%%....%%..%%....%%..%%.......%%....%%.%%....%%
# ..%%%%%%..%%.....%%.%%.....%%.%%.....%%.%%%%%%%%..%%%%%%...%%%%%%.
# 


thisfolder <- "~/Dropbox/BioLINCC files/CARRESS/CARRESS_2017a/main_study/data/csv/"

carress_endpts <- read.csv(paste(thisfolder,"a_endpts.csv",sep=""),na.strings=c("","NULL","NA",NA))
carress_rehosptl <- read.csv(paste(thisfolder,"rehosptl.csv",sep=""),na.strings=c("","NULL","NA",NA))
carress_assessmt <- read.csv(paste(thisfolder,"assessmt.csv",sep=""),na.strings=c("","NULL","NA",NA))
carress_visitsumm <- read.csv(paste(thisfolder,"a_visitsumm.csv",sep=""),na.strings=c("","NULL","NA",NA))
carress_deathpag <- read.csv(paste(thisfolder,"deathpag.csv",sep=""),na.strings=c("","NULL","NA",NA))
carress_term <- read.csv(paste(thisfolder,"term.csv",sep=""),na.strings=c("","NULL","NA",NA))
carress_crfluid <- read.csv(paste(thisfolder,"crfluid.csv",sep=""),na.strings=c("","NULL","NA",NA))
carress_vas <- read.csv(paste(thisfolder,"vas.csv",sep=""),na.strings=c("","NULL","NA",NA))

carress_deathpag <- subset(carress_deathpag,!is.na(DEATHDT))

acute_carress_outcomes <- sqldf('select PATNUMB as patientid, 
                                 DCALIVE1 as dc_alive, 
                                 DRNDIS as los 
                                 from carress_endpts')

acute_carress_outcomes$inhosp_dth_status[acute_carress_outcomes$dc_alive==1] <- 0
acute_carress_outcomes$inhosp_dth_status[acute_carress_outcomes$dc_alive==0] <- 1


carress_visitsumm_bl <- sqldf('select PATNUMB as patientid, 
                              WTLBS/2.2 as wt_kg_bl,
                              LL_GFR as gfr_bl, 
                              CL_NTPRO as ntbnp_bl
                              from carress_visitsumm 
                              where FORM = "BASELINE"')

carress_visitsumm_1d <- sqldf('select PATNUMB as patientid, 
                              WTLBS/2.2 as wt_kg_1d,
                              LL_GFR as gfr_1d
                              from carress_visitsumm 
                              where FORM = "DAY1"')

carress_visitsumm_2d <- sqldf('select PATNUMB as patientid, 
                              WTLBS/2.2 as wt_kg_2d,
                              LL_GFR as gfr_2d
                              from carress_visitsumm 
                              where FORM = "DAY2"')

carress_visitsumm_3d <- sqldf('select PATNUMB as patientid, 
                              WTLBS/2.2 as wt_kg_3d,
                              LL_GFR as gfr_3d                              
                              from carress_visitsumm 
                              where FORM = "DAY3"')

carress_visitsumm_4d <- sqldf('select PATNUMB as patientid, 
                              WTLBS/2.2 as wt_kg_4d,
                              LL_GFR as gfr_4d, 
                              CL_NTPRO as ntbnp_4d 
                              from carress_visitsumm 
                              where FORM = "DAY4"')

carress_visitsumm_5d <- sqldf('select PATNUMB as patientid, 
                              WTLBS/2.2 as wt_kg_5d
                              from carress_visitsumm 
                              where FORM = "DAY5"')

carress_visitsumm_6d <- sqldf('select PATNUMB as patientid, 
                              WTLBS/2.2 as wt_kg_6d
                              from carress_visitsumm 
                              where FORM = "DAY6"')

carress_visitsumm_7d <- sqldf('select PATNUMB as patientid, 
                                WTLBS/2.2 as wt_kg_7d,
                                LL_GFR as gfr_7d, 
                                CL_NTPRO as ntbnp_7d 
                                from carress_visitsumm 
                                where FORM = "DAY7"')

acute_carress_outcomes <- merge(acute_carress_outcomes,carress_visitsumm_bl,all.x=T)
acute_carress_outcomes <- merge(acute_carress_outcomes,carress_visitsumm_1d,all.x=T)
acute_carress_outcomes <- merge(acute_carress_outcomes,carress_visitsumm_2d,all.x=T)
acute_carress_outcomes <- merge(acute_carress_outcomes,carress_visitsumm_3d,all.x=T)
acute_carress_outcomes <- merge(acute_carress_outcomes,carress_visitsumm_4d,all.x=T)
acute_carress_outcomes <- merge(acute_carress_outcomes,carress_visitsumm_7d,all.x=T)


####. LOCF weight

acute_carress_outcomes$wt_kg_last <- acute_carress_outcomes$wt_kg_7d

acute_carress_outcomes$wt_kg_last[is.na(acute_carress_outcomes$wt_kg_last)&
                                    !is.na(acute_carress_outcomes$wt_kg_6d)] <-
  acute_carress_outcomes$wt_kg_6d[is.na(acute_carress_outcomes$wt_kg_last)&
                                      !is.na(acute_carress_outcomes$wt_kg_6d)]

acute_carress_outcomes$wt_kg_last[is.na(acute_carress_outcomes$wt_kg_last)&
                                    !is.na(acute_carress_outcomes$wt_kg_5d)] <-
  acute_carress_outcomes$wt_kg_5d[is.na(acute_carress_outcomes$wt_kg_last)&
                                    !is.na(acute_carress_outcomes$wt_kg_5d)]

acute_carress_outcomes$wt_kg_last[is.na(acute_carress_outcomes$wt_kg_last)&
                                    !is.na(acute_carress_outcomes$wt_kg_4d)] <-
  acute_carress_outcomes$wt_kg_4d[is.na(acute_carress_outcomes$wt_kg_last)&
                                    !is.na(acute_carress_outcomes$wt_kg_4d)]

acute_carress_outcomes$wt_kg_last[is.na(acute_carress_outcomes$wt_kg_last)&
                                    !is.na(acute_carress_outcomes$wt_kg_3d)] <-
  acute_carress_outcomes$wt_kg_3d[is.na(acute_carress_outcomes$wt_kg_last)&
                                    !is.na(acute_carress_outcomes$wt_kg_3d)]

acute_carress_outcomes$wt_kg_last[is.na(acute_carress_outcomes$wt_kg_last)&
                                    !is.na(acute_carress_outcomes$wt_kg_2d)] <-
  acute_carress_outcomes$wt_kg_2d[is.na(acute_carress_outcomes$wt_kg_last)&
                                    !is.na(acute_carress_outcomes$wt_kg_2d)]

acute_carress_outcomes$wt_kg_last[is.na(acute_carress_outcomes$wt_kg_last)&
                                    !is.na(acute_carress_outcomes$wt_kg_1d)] <-
  acute_carress_outcomes$wt_kg_1d[is.na(acute_carress_outcomes$wt_kg_last)&
                                    !is.na(acute_carress_outcomes$wt_kg_1d)]

acute_carress_outcomes$wt_kg_bl_last <- 
  acute_carress_outcomes$wt_kg_last-
  acute_carress_outcomes$wt_kg_bl


acute_carress_outcomes$wt_kg_bl_last_pct <- 
  acute_carress_outcomes$wt_kg_bl_last/
  acute_carress_outcomes$wt_kg_bl * 100


#### GFR LOCF

acute_carress_outcomes$gfr_last <- acute_carress_outcomes$gfr_7d

acute_carress_outcomes$gfr_last[is.na(acute_carress_outcomes$gfr_last)&
                                    !is.na(acute_carress_outcomes$gfr_4d)] <-
  acute_carress_outcomes$gfr_4d[is.na(acute_carress_outcomes$gfr_last)&
                                    !is.na(acute_carress_outcomes$gfr_4d)]

acute_carress_outcomes$gfr_last[is.na(acute_carress_outcomes$gfr_last)&
                                    !is.na(acute_carress_outcomes$gfr_3d)] <-
  acute_carress_outcomes$gfr_3d[is.na(acute_carress_outcomes$gfr_last)&
                                    !is.na(acute_carress_outcomes$gfr_3d)]

acute_carress_outcomes$gfr_last[is.na(acute_carress_outcomes$gfr_last)&
                                    !is.na(acute_carress_outcomes$gfr_2d)] <-
  acute_carress_outcomes$gfr_2d[is.na(acute_carress_outcomes$gfr_last)&
                                    !is.na(acute_carress_outcomes$gfr_2d)]

acute_carress_outcomes$gfr_last[is.na(acute_carress_outcomes$gfr_last)&
                                    !is.na(acute_carress_outcomes$gfr_1d)] <-
  acute_carress_outcomes$gfr_1d[is.na(acute_carress_outcomes$gfr_last)&
                                    !is.na(acute_carress_outcomes$gfr_1d)]


#### NT-BNP LOCF

acute_carress_outcomes$ntbnp_last <- acute_carress_outcomes$ntbnp_7d


acute_carress_outcomes$ntbnp_last[is.na(acute_carress_outcomes$ntbnp_last)&
                                  !is.na(acute_carress_outcomes$ntbnp_4d)] <-
  acute_carress_outcomes$ntbnp_4d[is.na(acute_carress_outcomes$ntbnp_last)&
                                  !is.na(acute_carress_outcomes$ntbnp_4d)]


# ### Vitals
# 
# carress_assessmt_bl <-
#   sqldf('select PATNUMB as patientid,
#                        HRATE as hr_bl,
#                        BPSYS as sbp_bl,
#                        BPDIA as dbp_bl,
#                        SPO2 as o2_bl,
#                        JVP as jvp_bl,
#                        RALES as rales_bl,
#                        AUSCULTN as s3_bl,
#                        ASCITES as ascites_bl,
#                        PEREDEMA as edema_bl,
#                        NYHA as nyha_bl,
#                        ORTHPNEA as orthopnea_bl
#                        from carress_assessmt
#                        where FORM = "BASELINE"
#                        AND HRATE is not null')
# 
# carress_assessmt_1d <-
#   sqldf('select PATNUMB as patientid,
#                        HRATE as hr_1d,
#                        BPSYS as sbp_1d,
#                        BPDIA as dbp_1d,
#                        SPO2 as o2_1d,
#                        JVP as jvp_1d,
#                        RALES as rales_1d,
#                        AUSCULTN as s3_1d,
#                        ASCITES as ascites_1d,
#                        PEREDEMA as edema_1d,
#                        NYHA as nyha_1d,
#                        ORTHPNEA as orthopnea_1d
#                        from carress_assessmt
#                        where FORM = "DAY1"
#                        AND HRATE is not null')
# 
# carress_assessmt_2d <-
#   sqldf('select PATNUMB as patientid,
#                        HRATE as hr_2d,
#                        BPSYS as sbp_2d,
#                        BPDIA as dbp_2d,
#                        SPO2 as o2_2d,
#                        JVP as jvp_2d,
#                        RALES as rales_2d,
#                        AUSCULTN as s3_2d,
#                        ASCITES as ascites_2d,
#                        PEREDEMA as edema_2d,
#                        NYHA as nyha_2d,
#                        ORTHPNEA as orthopnea_2d
#                        from carress_assessmt
#                        where FORM = "DAY2"
#                        AND HRATE is not null')
# 
# 
# carress_assessmt_3d <-
#   sqldf('select PATNUMB as patientid,
#                        HRATE as hr_3d,
#                        BPSYS as sbp_3d,
#                        BPDIA as dbp_3d,
#                        SPO2 as o2_3d,
#                        JVP as jvp_3d,
#                        RALES as rales_3d,
#                        AUSCULTN as s3_3d,
#                        ASCITES as ascites_3d,
#                        PEREDEMA as edema_3d,
#                        NYHA as nyha_3d,
#                        ORTHPNEA as orthopnea_3d
#                        from carress_assessmt
#                        where FORM = "DAY3"
#                        AND HRATE is not null')
# 
# carress_assessmt_4d <-
#   sqldf('select PATNUMB as patientid,
#                        HRATE as hr_4d,
#                        BPSYS as sbp_4d,
#                        BPDIA as dbp_4d,
#                        SPO2 as o2_4d,
#                        JVP as jvp_4d,
#                        RALES as rales_4d,
#                        AUSCULTN as s3_4d,
#                        ASCITES as ascites_4d,
#                        PEREDEMA as edema_4d,
#                        NYHA as nyha_4d,
#                        ORTHPNEA as orthopnea_4d
#                        from carress_assessmt
#                        where FORM = "DAY4"
#                        AND HRATE is not null')
# 
# 
# carress_assessmt_7d <-
#   sqldf('select PATNUMB as patientid,
#                        HRATE as hr_7d,
#                        BPSYS as sbp_7d,
#                        BPDIA as dbp_7d,
#                        SPO2 as o2_7d,
#                        JVP as jvp_7d,
#                        RALES as rales_7d,
#                        AUSCULTN as s3_7d,
#                        ASCITES as ascites_7d,
#                        PEREDEMA as edema_7d,
#                        NYHA as nyha_7d,
#                        ORTHPNEA as orthopnea_7d
#                        from carress_assessmt
#                        where FORM = "DAY7"
#                        AND HRATE is not null')
# 
# 
# carress_exam <-
#   merge(carress_assessmt_bl,
#         carress_assessmt_1d,
#         all=T)
# 
# carress_exam <-
#   merge(carress_exam,
#         carress_assessmt_2d,
#         all=T)
# 
# carress_exam <-
#   merge(carress_exam,
#         carress_assessmt_3d,
#         all=T)
# 
# carress_exam <-
#   merge(carress_exam,
#         carress_assessmt_4d,
#         all=T)
# 
# carress_exam <-
#   merge(carress_exam,
#         carress_assessmt_7d,
#         all=T)
# 


### Fluid balance

carress_crfluid$CRIVIN[is.na(carress_crfluid$CRIVIN)&carress_crfluid$CRFNONE==1] <- 0
carress_crfluid$CRORALIN[is.na(carress_crfluid$CRORALIN)&carress_crfluid$CRFNONE==1] <- 0
carress_crfluid$CRULTOUT[is.na(carress_crfluid$CRULTOUT)&carress_crfluid$CRFNONE==1] <- 0
carress_crfluid$CRURINOT[is.na(carress_crfluid$CRURINOT)&carress_crfluid$CRFNONE==1] <- 0

carress_crfluid$io_balance <- carress_crfluid$CRIVIN+carress_crfluid$CRORALIN-carress_crfluid$CRULTOUT-carress_crfluid$CRURINOT

carress_io_1d <- sqldf('select PATNUMB as patientid, 
                       io_balance as io_balance_1d 
                       from carress_crfluid 
                       where FORM = "DAY1"') 

carress_io_2d <- sqldf('select PATNUMB as patientid, 
                       io_balance as io_balance_2d 
                       from carress_crfluid 
                       where FORM = "DAY2"')

carress_io_3d <- sqldf('select PATNUMB as patientid, 
                       io_balance as io_balance_3d 
                       from carress_crfluid 
                       where FORM = "DAY3"') 

carress_io_4d <- sqldf('select PATNUMB as patientid, 
                       io_balance as io_balance_4d 
                       from carress_crfluid 
                       where FORM = "DAY4"') 

carress_io_5d <- sqldf('select PATNUMB as patientid, 
                       io_balance as io_balance_5d 
                       from carress_crfluid 
                       where FORM = "DAY5"') 

carress_io_6d <- sqldf('select PATNUMB as patientid, 
                       io_balance as io_balance_6d 
                       from carress_crfluid 
                       where FORM = "DAY6"') 

carress_io_7d <- sqldf('select PATNUMB as patientid, 
                         io_balance as io_balance_7d 
                         from carress_crfluid 
                         where FORM = "DAY7"') 


acute_carress_io <- merge(carress_io_1d,carress_io_2d,all.x=T)
acute_carress_io <- merge(acute_carress_io,carress_io_3d,all.x=T)
acute_carress_io <- merge(acute_carress_io,carress_io_4d,all.x=T)
acute_carress_io <- merge(acute_carress_io,carress_io_5d,all.x=T)
acute_carress_io <- merge(acute_carress_io,carress_io_6d,all.x=T)
acute_carress_io <- merge(acute_carress_io,carress_io_7d,all.x=T)

acute_carress_io$io_cum_2d <- apply(acute_carress_io[,2:3],MARGIN=1,function(x) sum(x))
acute_carress_io$io_cum_3d <- apply(acute_carress_io[,2:4],MARGIN=1,function(x) sum(x))
acute_carress_io$io_cum_4d <- apply(acute_carress_io[,2:5],MARGIN=1,function(x) sum(x))
acute_carress_io$io_cum_5d <- apply(acute_carress_io[,2:6],MARGIN=1,function(x) sum(x))
acute_carress_io$io_cum_6d <- apply(acute_carress_io[,2:7],MARGIN=1,function(x) sum(x))
acute_carress_io$io_cum_7d <- apply(acute_carress_io[,2:8],MARGIN=1,function(x) sum(x))


acute_carress_io$io_cum_last <- acute_carress_io$io_cum_7d

acute_carress_io$io_cum_last[is.na(acute_carress_io$io_cum_last)&
                                  !is.na(acute_carress_io$io_cum_6d)] <-
  acute_carress_io$io_cum_6d[is.na(acute_carress_io$io_cum_last)&
                                  !is.na(acute_carress_outcomes$io_cum_6d)]

acute_carress_io$io_cum_last[is.na(acute_carress_io$io_cum_last)&
                                  !is.na(acute_carress_io$io_cum_5d)] <-
  acute_carress_io$io_cum_5d[is.na(acute_carress_io$io_cum_last)&
                                  !is.na(acute_carress_io$io_cum_5d)]

acute_carress_io$io_cum_last[is.na(acute_carress_io$io_cum_last)&
                                  !is.na(acute_carress_io$io_cum_4d)] <-
  acute_carress_io$io_cum_4d[is.na(acute_carress_io$io_cum_last)&
                                  !is.na(acute_carress_io$io_cum_4d)]

acute_carress_io$io_cum_last[is.na(acute_carress_io$io_cum_last)&
                                  !is.na(acute_carress_io$io_cum_3d)] <-
  acute_carress_io$io_cum_3d[is.na(acute_carress_io$io_cum_last)&
                                  !is.na(acute_carress_io$io_cum_3d)]

acute_carress_io$io_cum_last[is.na(acute_carress_io$io_cum_last)&
                                  !is.na(acute_carress_io$io_cum_2d)] <-
  acute_carress_io$io_cum_2d[is.na(acute_carress_io$io_cum_last)&
                                  !is.na(acute_carress_io$io_cum_2d)]

acute_carress_io$io_cum_last[is.na(acute_carress_io$io_cum_last)&
                                  !is.na(acute_carress_io$io_cum_1d)] <-
  acute_carress_io$io_cum_1d[is.na(acute_carress_io$io_cum_last)&
                                  !is.na(acute_carress_io$io_cum_1d)]


#### VAS

carress_vas_bl <- sqldf('select PATNUMB as patientid, 
                        DYSPVAS as dyspnea_vas_bl, 
                        GLOBLVAS as global_vas_bl 
                        from carress_vas 
                        where FORM = "BASELINE"')


carress_vas_4d <- sqldf('select PATNUMB as patientid, 
                        DYSPVAS as dyspnea_vas_4d, 
                        GLOBLVAS as global_vas_4d 
                        from carress_vas 
                        where FORM = "DAY4"')

carress_vas_7d <- sqldf('select PATNUMB as patientid, 
                        DYSPVAS as dyspnea_vas_7d,
                        GLOBLVAS as global_vas_7d 
                        from carress_vas 
                        where FORM = "DAY7"')

acute_carress_vas <- merge(carress_vas_bl,carress_vas_4d,all.x=T)
acute_carress_vas <- merge(acute_carress_vas,carress_vas_7d,all.x=T)

## Global VAS

acute_carress_vas$global_vas_bl_4d <- acute_carress_vas$global_vas_4d - acute_carress_vas$global_vas_bl
acute_carress_vas$global_vas_bl_7d <- acute_carress_vas$global_vas_7d - acute_carress_vas$global_vas_bl
acute_carress_vas$global_vas_4d_7d <- acute_carress_vas$global_vas_7d - acute_carress_vas$global_vas_4d

acute_carress_vas$global_vas_last <- acute_carress_vas$global_vas_7d

acute_carress_vas$global_vas_last[!is.na(acute_carress_vas$global_vas_4d)&is.na(acute_carress_vas$global_vas_last)] <- 
  acute_carress_vas$global_vas_4d[!is.na(acute_carress_vas$global_vas_4d)&is.na(acute_carress_vas$global_vas_last)]

acute_carress_vas$global_vas_bl_last <- acute_carress_vas$global_vas_last - acute_carress_vas$global_vas_bl


## Dyspnea VAS

acute_carress_vas$dyspnea_vas_bl_4d <- acute_carress_vas$dyspnea_vas_4d - acute_carress_vas$dyspnea_vas_bl
acute_carress_vas$dyspnea_vas_bl_7d <- acute_carress_vas$dyspnea_vas_7d - acute_carress_vas$dyspnea_vas_bl
acute_carress_vas$dyspnea_vas_4d_7d <- acute_carress_vas$dyspnea_vas_7d - acute_carress_vas$dyspnea_vas_4d

acute_carress_vas$dyspnea_vas_last <- acute_carress_vas$dyspnea_vas_7d

acute_carress_vas$dyspnea_vas_last[!is.na(acute_carress_vas$dyspnea_vas_4d)&is.na(acute_carress_vas$dyspnea_vas_last)] <- 
  acute_carress_vas$dyspnea_vas_4d[!is.na(acute_carress_vas$dyspnea_vas_4d)&is.na(acute_carress_vas$dyspnea_vas_last)]

acute_carress_vas$global_vas_bl_last <- acute_carress_vas$global_vas_last - acute_carress_vas$global_vas_bl

### Combined

acute_carress_outcomes <- merge(acute_carress_outcomes,acute_carress_io, by="patientid", all.x=T)
acute_carress_outcomes <- merge(acute_carress_outcomes,acute_carress_vas, by="patientid", all.x=T)


missing_acute_outcomes_carress <- acute_analysis_outcomes[!acute_analysis_outcomes %in% names(acute_carress_outcomes)]
acute_carress_outcomes[,missing_acute_outcomes_carress] <- NA
acute_analysis_outcomes_carress <- acute_carress_outcomes[,acute_analysis_outcomes]

acute_carress_outcomes$study <- "CARRESS"
acute_analysis_outcomes_carress$study <- "CARRESS"

all_acute_carress_outcomes <- acute_carress_outcomes

rm(list=ls(pattern="\\bcarress_."))


# 
######################### ******* DOSE ******* ###########################
# 
# .%%%%%%%%...%%%%%%%...%%%%%%..%%%%%%%%
# .%%.....%%.%%.....%%.%%....%%.%%......
# .%%.....%%.%%.....%%.%%.......%%......
# .%%.....%%.%%.....%%..%%%%%%..%%%%%%..
# .%%.....%%.%%.....%%.......%%.%%......
# .%%.....%%.%%.....%%.%%....%%.%%......
# .%%%%%%%%...%%%%%%%...%%%%%%..%%%%%%%%
# 


thisfolder <- "~/Dropbox/BioLINCC files/DOSE/data/"

dose_endpts <- read.csv(paste(thisfolder,"analysis/csv/a_endpts.csv",sep=""),stringsAsFactors = F,na.strings=c("NA","NULL","",NA))
dose_visitsumm <- read.csv(paste(thisfolder,"analysis/csv/a_visitsumm.csv",sep=""),na.strings=c("NA","NULL","",NA))
dose_deathpag <- read.csv(paste(thisfolder,"data/csv/deathpag.csv",sep=""),na.strings=c("NA","NULL","",NA))
dose_rehosptl <- read.csv(paste(thisfolder,"data/csv/rehosptl.csv",sep=""),na.strings=c("NA","NULL","",NA))
dose_assessmt <- read.csv(paste(thisfolder,"data/csv/assessmt.csv",sep=""),na.strings=c("NA","NULL","",NA))
dose_fluid <- read.csv(paste(thisfolder,"data/csv/fluid.csv",sep=""),na.strings=c("NA","NULL","",NA))
dose_subjsymp <- read.csv(paste(thisfolder,"data/csv/subjsymp.csv",sep=""),na.strings=c("NA","NULL","",NA))
dose_term <- read.csv(paste(thisfolder,"data/csv/term.csv",sep=""),na.strings=c("NA","NULL","",NA))

dose_deathpag <- subset(dose_deathpag,!is.na(DEATHDT))


acute_dose_outcomes <- sqldf('select PATNUMB as patientid, 
                              DCALIVE as dc_alive,
                              DRNDIS as los from dose_endpts')

acute_dose_outcomes$inhosp_dth_status[acute_dose_outcomes$dc_alive==1] <- 0
acute_dose_outcomes$inhosp_dth_status[acute_dose_outcomes$dc_alive==0] <- 1


#### DOSE Visit summary ####

dose_visitsumm_bl <- sqldf('select PATNUMB as patientid, 
                           WTLBS/2.2 as wt_kg_bl,
                           LL_BUN as bun_bl,
                           LL_GFR as gfr_bl, 
                           LL_BNP as bnp_bl, 
                           CL_NTPRO as ntbnp_bl,
                           CL_ALDOST as aldo_bl
                           from dose_visitsumm 
                           where FORM = "BASELINE"')

dose_visitsumm_1d <- sqldf('select PATNUMB as patientid, 
                           WTLBS/2.2 as wt_kg_1d,
                           LL_BUN as bun_bl,
                           LL_GFR as gfr_1d 
                           from dose_visitsumm 
                           where FORM = "24HOUR"')

dose_visitsumm_2d <- sqldf('select PATNUMB as patientid, 
                           WTLBS/2.2 as wt_kg_2d,
                           LL_BUN as bun_bl,
                           LL_GFR as gfr_2d 
                           from dose_visitsumm 
                           where FORM = "48HOUR"')

dose_visitsumm_3d <- sqldf('select PATNUMB as patientid, 
                           WTLBS/2.2 as wt_kg_3d,
                           LL_BUN as bun_bl,
                           LL_GFR as gfr_3d, 
                           CL_NTPRO as ntbnp_3d, 
                           CL_ALDOST as aldo_3d 
                           from dose_visitsumm 
                           where FORM = "72HOUR"')

dose_visitsumm_4d <- sqldf('select PATNUMB as patientid, 
                           WTLBS/2.2 as wt_kg_4d,
                           LL_BUN as bun_4d 
                           from dose_visitsumm 
                           where FORM = "96HOUR"')

dose_visitsumm_7d <- sqldf('select PATNUMB as patientid, 
                           WTLBS/2.2 as wt_kg_7d,
                           LL_BUN as bun_7d,
                           LL_GFR as gfr_7d, 
                           CL_NTPRO as ntbnp_7d,
                           CL_ALDOST as aldo_7d 
                           from dose_visitsumm 
                           where FORM = "DAY7"')


acute_dose_visitsumm <- merge(dose_visitsumm_bl,dose_visitsumm_1d,all.x=T)
acute_dose_visitsumm <- merge(acute_dose_visitsumm,dose_visitsumm_2d,all.x=T)
acute_dose_visitsumm <- merge(acute_dose_visitsumm,dose_visitsumm_3d,all.x=T)
acute_dose_visitsumm <- merge(acute_dose_visitsumm,dose_visitsumm_4d,all.x=T)
acute_dose_visitsumm <- merge(acute_dose_visitsumm,dose_visitsumm_7d,all.x=T)


#### NT-BNP LOCF ####

acute_dose_visitsumm$ntbnp_last <- acute_dose_visitsumm$ntbnp_7d

acute_dose_visitsumm$ntbnp_last[is.na(acute_dose_visitsumm$ntbnp_last)&
                               !is.na(acute_dose_visitsumm$ntbnp_3d)] <-
  acute_dose_visitsumm$ntbnp_3d[is.na(acute_dose_visitsumm$ntbnp_last)&
                               !is.na(acute_dose_visitsumm$ntbnp_3d)]


##### Aldosterone LOCF ####

acute_dose_visitsumm$aldo_last <- acute_dose_visitsumm$aldo_7d

acute_dose_visitsumm$aldo_last[is.na(acute_dose_visitsumm$aldo_last)&
                                 !is.na(acute_dose_visitsumm$aldo_3d)] <-
  acute_dose_visitsumm$aldo_3d[is.na(acute_dose_visitsumm$aldo_last)&
                                 !is.na(acute_dose_visitsumm$aldo_3d)]

acute_dose_visitsumm$aldo_bl_last <- acute_dose_visitsumm$aldo_last - acute_dose_visitsumm$aldo_bl


#### I/O balance ####

dose_fluid$io_balance <- dose_fluid$TOTALIN-dose_fluid$TOTALOUT

dose_io_1d <- sqldf('select PATNUMB as patientid, 
                    io_balance as io_balance_1d 
                    from dose_fluid 
                    where FORM = "24HOUR"') 

dose_io_2d <- sqldf('select PATNUMB as patientid, 
                    io_balance as io_balance_2d 
                    from dose_fluid 
                    where FORM = "48HOUR"') 

dose_io_3d <- sqldf('select PATNUMB as patientid, 
                    io_balance as io_balance_3d 
                    from dose_fluid 
                    where FORM = "72HOUR"') 

dose_io_4d <- sqldf('select PATNUMB as patientid, 
                    io_balance as io_balance_4d 
                    from dose_fluid 
                    where FORM = "96HOUR"') 


acute_dose_io <- merge(dose_io_1d,dose_io_2d,all.x=T)
acute_dose_io <- merge(acute_dose_io,dose_io_3d,all.x=T)
acute_dose_io <- merge(acute_dose_io,dose_io_4d,all.x=T)

acute_dose_io$io_cum_2d <- acute_dose_io$io_balance_2d + acute_dose_io$io_balance_1d
acute_dose_io$io_cum_3d <- acute_dose_io$io_cum_2d + acute_dose_io$io_balance_3d
acute_dose_io$io_cum_4d <- acute_dose_io$io_cum_3d + acute_dose_io$io_balance_4d

acute_dose_io$io_cum_last <- acute_dose_io$io_cum_last 

acute_dose_io$io_cum_last[is.na(acute_dose_io$io_cum_last)] <- 
  acute_dose_io$io_cum_3d[is.na(acute_dose_io$io_cum_last)]

acute_dose_io$io_cum_last[is.na(acute_dose_io$io_cum_last)] <- 
  acute_dose_io$io_cum_2d[is.na(acute_dose_io$io_cum_last)]

acute_dose_io$io_cum_last[is.na(acute_dose_io$io_cum_last)] <- 
  acute_dose_io$io_balance_1d[is.na(acute_dose_io$io_cum_last)]


#### Combine DOSE

acute_dose_outcomes <- merge(acute_dose_outcomes,
                             acute_dose_visitsumm,
                             by="patientid",
                             all.x=T)

# acute_dose_outcomes <- merge(acute_dose_outcomes,
#                              acute_dose_io,
#                              by="patientid",
#                              all.x=T)


missing_outcomes_dose <- acute_analysis_outcomes[!acute_analysis_outcomes %in% names(acute_dose_outcomes)]
acute_dose_outcomes[,missing_outcomes_dose] <- NA
analysis_outcomes_dose <- acute_dose_outcomes[,acute_analysis_outcomes]

missing_acute_outcomes_dose <- acute_analysis_outcomes[!acute_analysis_outcomes %in% names(acute_dose_outcomes)]
acute_dose_outcomes[,missing_acute_outcomes_dose] <- NA
acute_analysis_outcomes_dose <- acute_dose_outcomes[,acute_analysis_outcomes]

acute_analysis_outcomes_dose$study <- "DOSE"

all_acute_dose_outcomes <- acute_dose_outcomes  


 
# 
####################### ******* ESCAPE ******* ###########################
# 
# .%%%%%%%%..%%%%%%...%%%%%%.....%%%....%%%%%%%%..%%%%%%%%
# .%%.......%%....%%.%%....%%...%%.%%...%%.....%%.%%......
# .%%.......%%.......%%........%%...%%..%%.....%%.%%......
# .%%%%%%....%%%%%%..%%.......%%.....%%.%%%%%%%%..%%%%%%..
# .%%.............%%.%%.......%%%%%%%%%.%%........%%......
# .%%.......%%....%%.%%....%%.%%.....%%.%%........%%......
# .%%%%%%%%..%%%%%%...%%%%%%..%%.....%%.%%........%%%%%%%%
# 

escape_patient <- read.csv("~/Dropbox/BioLINCC files/ESCAPE/data/main/analdata/patient.csv",stringsAsFactor=F)
escape_exercise_all <- read.csv("~/Dropbox/BioLINCC files/ESCAPE/data/main/analdata/exercise.csv",stringsAsFactor=F)
escape_walk <- read.csv("~/Dropbox/BioLINCC files/ESCAPE/data/main/sasdata/walk.csv",stringsAsFactor=F)
escape_quality <- read.csv("~/Dropbox/BioLINCC files/ESCAPE/data/main/analdata/quality.csv",stringsAsFactor=F)
escape_hemo <- read.csv("~/Dropbox/BioLINCC files/ESCAPE/data/main/analdata/hemo.csv",stringsAsFactor=F)
escape_physical <- read.csv("~/Dropbox/BioLINCC files/ESCAPE/data/main/analdata/physical.csv",stringsAsFactor=F)
escape_physexam <- read.csv("~/Dropbox/BioLINCC files/ESCAPE/data/main/sasdata/physexam.csv",stringsAsFactor=F)
escape_volume <- read.csv("~/Dropbox/BioLINCC files/ESCAPE/data/main/sasdata/volume.csv",stringsAsFactor=F)
escape_visual <- read.csv("~/Dropbox/BioLINCC files/ESCAPE/data/main/sasdata/visual.csv",stringsAsFactor=F)
escape_mechd <- read.csv("~/Dropbox/BioLINCC files/ESCAPE/data/mechanistic/analdata/mechd.csv",stringsAsFactor=F)
escape_merged <- read.csv("~/Dropbox/BioLINCC files/ESCAPE/data/big/analdata/merged.csv",stringsAsFactors = F)
escape_labs <- read.csv("~/Dropbox/BioLINCC files/ESCAPE/data/main/analdata/labs.csv",stringsAsFactor=F)
escape_lab <- read.csv("~/Dropbox/BioLINCC files/ESCAPE/data/main/sasdata/lab.csv",stringsAsFactor=F)
escape_enzymes <- read.csv("~/Dropbox/BioLINCC files/ESCAPE/data/main/sasdata/enzymes.csv",stringsAsFactor=F)


#######


acute_escape_outcomes <- sqldf('select DEIDNUM as patientid, 
                                DTH_INIT as inhosp_dth_status, 
                                HOSPDAY as los from escape_patient')

acute_escape_outcomes$inhosp_dth_status[is.na(acute_escape_outcomes$inhosp_dth_status)] <- 0

#####

acute_escape_mechd <- sqldf("select DEIDNUM as patientid,
                            NOREPIB as norepi_bl,
                            EPIB as epi_bl,
                            DOPAMINB as dopa_bl,
                            NOREPID as norepi_last,
                            EPID as epi_last,
                            DOPAMIND as dopa_last,
                            ANPB as anp_bl,
                            BNPB as bnp_bl,
                            ANPD as anp_last,
                            BNPD as bnp_last
                            from escape_mechd")


acute_escape_mechd$norepi_bl_last <- acute_escape_mechd$norepi_last - acute_escape_mechd$norepi_bl
acute_escape_mechd$epi_bl_last <- acute_escape_mechd$epi_last - acute_escape_mechd$epi_bl
acute_escape_mechd$dopa_bl_last <- acute_escape_mechd$dopa_last - acute_escape_mechd$dopa_bl
acute_escape_mechd$anp_bl_last <- acute_escape_mechd$anp_last - acute_escape_mechd$anp_bl
acute_escape_mechd$bnp_bl_last <- acute_escape_mechd$bnp_last - acute_escape_mechd$bnp_bl

######


# acute_escape_renal <- sqldf('select DEIDNUM as patientid, 
#                       CRTB as creatinine_bl, 
#                       CRTD3 as creatinine_3d, 
#                       CRTD5 as creatinine_5d, 
#                       CRTD7 as creatinine_7d 
#                       from escape_labs')
# 
# acute_escape_renal <- merge(acute_escape_renal,
#                       escape_patient[,c('DEIDNUM',
#                                         'AGE',
#                                         'GENDER',
#                                         'RACE')],
#                       by.x="patientid",
#                       by.y="DEIDNUM",
#                       all.x=T)
# 
# acute_escape_renal$gfr_bl <- calc_MDRD4(dat=acute_escape_renal,
#                                   cr="creatinine_bl",
#                                   sex="GENDER",
#                                   race="RACE",
#                                   male=1,
#                                   black=2)
# 
# acute_escape_renal$gfr_3d <- calc_MDRD4(dat=acute_escape_renal,
#                                   cr="creatinine_3d",
#                                   sex="GENDER",
#                                   race="RACE",
#                                   male=1,
#                                   black=2)
# 
# acute_escape_renal$gfr_5d <- calc_MDRD4(dat=acute_escape_renal,
#                                   cr="creatinine_5d",
#                                   sex="GENDER",
#                                   race="RACE",
#                                   male=1,
#                                   black=2)
# 
# acute_escape_renal$gfr_7d <- calc_MDRD4(dat=acute_escape_renal,
#                                   cr="creatinine_7d",
#                                   sex="GENDER",
#                                   race="RACE",
#                                   male=1,
#                                   black=2)
# 
# 
# acute_escape_renal$gfr_last <- acute_escape_renal$gfr_7d
# 
# acute_escape_renal$gfr_last[is.na(acute_escape_renal$gfr_last)&
#                               !is.na(acute_escape_renal$gfr_6d)] <-
#   acute_escape_renal$gfr_6d[is.na(acute_escape_renal$gfr_last)&
#                               !is.na(acute_escape_renal$gfr_6d)]
# acute_escape_renal$gfr_last[is.na(acute_escape_renal$gfr_last)&
#                               !is.na(acute_escape_renal$gfr_5d)] <-
#   acute_escape_renal$gfr_5d[is.na(acute_escape_renal$gfr_last)&
#                               !is.na(acute_escape_renal$gfr_5d)]
# 
# acute_escape_renal$gfr_last[is.na(acute_escape_renal$gfr_last)&
#                               !is.na(acute_escape_renal$gfr_4d)] <-
#   acute_escape_renal$gfr_4d[is.na(acute_escape_renal$gfr_last)&
#                               !is.na(acute_escape_renal$gfr_4d)]
# 
# acute_escape_renal$gfr_last[is.na(acute_escape_renal$gfr_last)&
#                               !is.na(acute_escape_renal$gfr_3d)] <-
#   acute_escape_renal$gfr_3d[is.na(acute_escape_renal$gfr_last)&
#                               !is.na(acute_escape_renal$gfr_3d)]
# 
# acute_escape_renal$gfr_last[is.na(acute_escape_renal$gfr_last)&
#                               !is.na(acute_escape_renal$gfr_2d)] <-
#   acute_escape_renal$gfr_2d[is.na(acute_escape_renal$gfr_last)&
#                               !is.na(acute_escape_renal$gfr_2d)]
# 
# acute_escape_renal$gfr_last[is.na(acute_escape_renal$gfr_last)&
#                               !is.na(acute_escape_renal$gfr_1d)] <-
#   acute_escape_renal$gfr_1d[is.na(acute_escape_renal$gfr_last)&
#                               !is.na(acute_escape_renal$gfr_1d)]
# 
# 
# acute_escape_renal$gfr_bl_3d <- acute_escape_renal$gfr_3d - acute_escape_renal$gfr_bl
# acute_escape_renal$gfr_bl_5d <- acute_escape_renal$gfr_5d - acute_escape_renal$gfr_bl
# acute_escape_renal$gfr_bl_7d <- acute_escape_renal$gfr_7d - acute_escape_renal$gfr_bl
# acute_escape_renal$gfr_bl_last <- acute_escape_renal$gfr_last - acute_escape_renal$gfr_bl
# 
# 
# acute_escape_renal$gfr_bl_3d_pct <- (acute_escape_renal$gfr_3d - acute_escape_renal$gfr_bl)/acute_escape_renal$gfr_bl
# acute_escape_renal$gfr_bl_5d_pct <- (acute_escape_renal$gfr_5d - acute_escape_renal$gfr_bl)/acute_escape_renal$gfr_bl
# acute_escape_renal$gfr_bl_7d_pct <- (acute_escape_renal$gfr_7d - acute_escape_renal$gfr_bl)/acute_escape_renal$gfr_bl
# acute_escape_renal$gfr_bl_last_pct <- (acute_escape_renal$gfr_last - acute_escape_renal$gfr_bl)/acute_escape_renal$gfr_bl
# 

#### I/O balance ####

escape_volume <- subset(escape_volume, VOLDT >= 0)

escape_volume$io_balance[escape_volume$VOLSIGN==1&!is.na(escape_volume$VOLSIGN)] <-
  escape_volume$VOLNET[escape_volume$VOLSIGN==1&!is.na(escape_volume$VOLSIGN)]

escape_volume$io_balance[escape_volume$VOLSIGN==2&!is.na(escape_volume$VOLSIGN)] <-
  -escape_volume$VOLNET[escape_volume$VOLSIGN==2&!is.na(escape_volume$VOLSIGN)]

escape_volume <- subset(escape_volume,!is.na(VOLDT))


escape_volume_melt <- sqldf("select DEIDNUM as patientid,
                            VOLDT,
                            'io_balance' as variable,
                            io_balance as value
                            from escape_volume")

escape_io_bl <- sqldf('select patientid,
                      value as io_balance_bl
                      from escape_volume_melt
                      where VOLDT = 0')

escape_io_1d <- sqldf('select patientid,
                      value as io_balance_1d
                      from escape_volume_melt
                      where VOLDT = 1')

escape_io_2d <- sqldf('select patientid,
                      value as io_balance_2d
                      from escape_volume_melt
                      where VOLDT = 2')

escape_io_3d <- sqldf('select patientid,
                      value as io_balance_3d
                      from escape_volume_melt
                      where VOLDT = 3')

escape_io_4d <- sqldf('select patientid,
                      value as io_balance_4d
                      from escape_volume_melt
                      where VOLDT = 4')

escape_io_5d <- sqldf('select patientid,
                      value as io_balance_5d
                      from escape_volume_melt
                      where VOLDT = 5')

escape_io_6d <- sqldf('select patientid,
                      value as io_balance_6d
                      from escape_volume_melt
                      where VOLDT = 6')

escape_io_7d <- sqldf('select patientid,
                      value as io_balance_7d
                      from escape_volume_melt
                      where VOLDT = 7')


acute_escape_io <- merge(escape_io_bl,escape_io_1d,by="patientid",all=T)
acute_escape_io <- merge(acute_escape_io,escape_io_2d,by="patientid",all=T)
acute_escape_io <- merge(acute_escape_io,escape_io_3d,by="patientid",all=T)
acute_escape_io <- merge(acute_escape_io,escape_io_4d,by="patientid",all=T)
acute_escape_io <- merge(acute_escape_io,escape_io_5d,by="patientid",all=T)
acute_escape_io <- merge(acute_escape_io,escape_io_6d,by="patientid",all=T)
acute_escape_io <- merge(acute_escape_io,escape_io_7d,by="patientid",all=T)

acute_escape_io$io_cum_1d <- apply(acute_escape_io[,c("io_balance_bl","io_balance_1d")],FUN=function(x) sum(x,na.rm=T), MARGIN=1)
acute_escape_io$io_cum_2d <- acute_escape_io$io_cum_1d+acute_escape_io$io_balance_2d
acute_escape_io$io_cum_3d <- acute_escape_io$io_cum_1d+acute_escape_io$io_balance_3d
acute_escape_io$io_cum_4d <- acute_escape_io$io_cum_1d+acute_escape_io$io_balance_4d
acute_escape_io$io_cum_5d <- acute_escape_io$io_cum_1d+acute_escape_io$io_balance_5d
acute_escape_io$io_cum_6d <- acute_escape_io$io_cum_1d+acute_escape_io$io_balance_6d
acute_escape_io$io_cum_7d <- acute_escape_io$io_cum_1d+acute_escape_io$io_balance_7d

acute_escape_io$io_cum_last <- acute_escape_io$io_cum_7d
acute_escape_io$io_cum_last[is.na(acute_escape_io$io_cum_last)] <-
  acute_escape_io$io_cum_6d[is.na(acute_escape_io$io_cum_last)]

acute_escape_io$io_cum_last[is.na(acute_escape_io$io_cum_last)] <-
  acute_escape_io$io_cum_5d[is.na(acute_escape_io$io_cum_last)]

acute_escape_io$io_cum_last[is.na(acute_escape_io$io_cum_last)] <-
  acute_escape_io$io_cum_4d[is.na(acute_escape_io$io_cum_last)]

acute_escape_io$io_cum_last[is.na(acute_escape_io$io_cum_last)] <-
  acute_escape_io$io_cum_3d[is.na(acute_escape_io$io_cum_last)]

acute_escape_io$io_cum_last[is.na(acute_escape_io$io_cum_last)] <-
  acute_escape_io$io_cum_2d[is.na(acute_escape_io$io_cum_last)]

acute_escape_io$io_cum_last[is.na(acute_escape_io$io_cum_last)] <-
  acute_escape_io$io_cum_1d[is.na(acute_escape_io$io_cum_last)]

#### Physical exam/vital signs ####

escape_physexam$wt_kg[escape_physexam$WTUNIT==2&!is.na(escape_physexam$WTUNIT)] <- 
  escape_physexam$WT[escape_physexam$WTUNIT==2&!is.na(escape_physexam$WTUNIT)]

escape_physexam$wt_kg[escape_physexam$WTUNIT==1&!is.na(escape_physexam$WTUNIT)] <- 
  escape_physexam$WT[escape_physexam$WTUNIT==1&!is.na(escape_physexam$WTUNIT)]/2.2

escape_physexam$PHYSDAY[escape_physexam$FORM=="Baseline"] <- 0 
escape_physexam$PHYSDAY[escape_physexam$FORM=="Discharge"] <- 7 

escape_physexam$studyvisit[!is.na(escape_physexam$PHYSDAY)] <- paste("DAY",escape_physexam$PHYSDAY[!is.na(escape_physexam$PHYSDAY)] ,sep="")
escape_physexam$studyvisit[is.na(escape_physexam$PHYSDAY)] <- escape_physexam$FORM[is.na(escape_physexam$PHYSDAY)]


escape_vitals_bl <- sqldf('select DEIDNUM as patientid, 
                          wt_kg as wt_kg_bl, 
                          HRSUP as hr_bl, 
                          SUPSYSBP as sbp_bl, 
                          SUPDIABP as dbp_bl
                          from escape_physexam 
                          where FORM = "Baseline" ')

escape_vitals_bl$pupr_bl <- escape_vitals_bl$sbp_bl-escape_vitals_bl$dbp_bl

escape_vitals_bl$map_bl <- 
  calc_map(diabp=escape_vitals_bl$dbp_bl,
           sysbp=escape_vitals_bl$sbp_bl,
           hr=escape_vitals_bl$hr_bl)

escape_vitals_1d <- sqldf('select DEIDNUM as patientid, 
                          wt_kg as wt_kg_1d, 
                          HRSUP as hr_1d, 
                          SUPSYSBP as sbp_1d, 
                          SUPDIABP as dbp_1d 
                          from escape_physexam 
                          where PHYSDAY = 1')


escape_vitals_1d$pupr_1d <- escape_vitals_1d$sbp_1d-escape_vitals_1d$dbp_1d

escape_vitals_1d$map_1d <- 
  calc_map(diabp=escape_vitals_1d$dbp_1d,
           sysbp=escape_vitals_1d$sbp_1d,
           hr=escape_vitals_1d$hr_1d)


escape_vitals_2d <- sqldf('select DEIDNUM as patientid, 
                          wt_kg as wt_kg_2d, 
                          HRSUP as hr_2d, 
                          SUPSYSBP as sbp_2d, 
                          SUPDIABP as dbp_2d 
                          from escape_physexam 
                          where PHYSDAY = 2 ')

escape_vitals_2d$pupr_2d <- 
  escape_vitals_2d$sbp_2d-
  escape_vitals_2d$dbp_2d


escape_vitals_2d$map_2d <- 
  calc_map(diabp=escape_vitals_2d$dbp_2d,
           sysbp=escape_vitals_2d$sbp_2d,
           hr=escape_vitals_2d$hr_2d)


escape_vitals_3d <- sqldf('select DEIDNUM as patientid, 
                          wt_kg as wt_kg_3d, 
                          HRSUP as hr_3d, 
                          SUPSYSBP as sbp_3d, 
                          SUPDIABP as dbp_3d 
                          from escape_physexam 
                          where PHYSDAY = 3 ')

escape_vitals_3d$pupr_3d <- escape_vitals_3d$sbp_3d-escape_vitals_3d$dbp_3d

escape_vitals_3d$map_3d <- 
  calc_map(diabp=escape_vitals_3d$dbp_3d,
           sysbp=escape_vitals_3d$sbp_3d,
           hr=escape_vitals_3d$hr_3d)


escape_vitals_4d <- sqldf('select DEIDNUM as patientid, 
                          wt_kg as wt_kg_4d, 
                          HRSUP as hr_4d, 
                          SUPSYSBP as sbp_4d, 
                          SUPDIABP as dbp_4d 
                          from escape_physexam 
                          where PHYSDAY = 4 ')

escape_vitals_4d$pupr_4d <- escape_vitals_4d$sbp_4d-escape_vitals_4d$dbp_4d

escape_vitals_4d$map_4d <- 
  calc_map(diabp=escape_vitals_4d$dbp_4d,
           sysbp=escape_vitals_4d$sbp_4d,
           hr=escape_vitals_4d$hr_4d)


escape_vitals_last <- sqldf('select DEIDNUM as patientid, 
                          wt_kg as wt_kg_last, 
                          HRSUP as hr_last, 
                          SUPSYSBP as sbp_last, 
                          SUPDIABP as dbp_last
                          from escape_physexam 
                          where FORM = "Discharge" ')


escape_vitals_last$pupr_last <- escape_vitals_last$sbp_last-escape_vitals_last$dbp_last

escape_vitals_last$map_last <- 
  calc_map(diabp=escape_vitals_last$dbp_last,
           sysbp=escape_vitals_last$sbp_last,
           hr=escape_vitals_last$hr_last)


escape_vitals_2w <- sqldf('select DEIDNUM as patientid, 
                          wt_kg as wt_kg_2w, 
                          HRSUP as hr_2w, 
                          SUPSYSBP as sbp_2w, 
                          SUPDIABP as dbp_2w
                          from escape_physexam 
                          where FORM = "2-Week Follow-Up" ')

escape_vitals_2w$pupr_2w <- escape_vitals_2w$sbp_2w-escape_vitals_2w$dbp_2w

escape_vitals_2w$map_2w <- 
  calc_map(diabp=escape_vitals_2w$dbp_2w,
           sysbp=escape_vitals_2w$sbp_2w,
           hr=escape_vitals_2w$hr_2w)


escape_vitals_8w <- sqldf('select DEIDNUM as patientid, 
                          wt_kg as wt_kg_8w, 
                          HRSUP as hr_8w, 
                          SUPSYSBP as sbp_8w, 
                          SUPDIABP as dbp_8w 
                          from escape_physexam 
                          where FORM = "2-Month Follow-Up" ')

escape_vitals_8w$pupr_8w <- 
  escape_vitals_8w$sbp_8w-
  escape_vitals_8w$dbp_8w

escape_vitals_8w$map_8w <- 
  calc_map(diabp=escape_vitals_8w$dbp_8w,
           sysbp=escape_vitals_8w$sbp_8w,
           hr=escape_vitals_8w$hr_8w)


escape_vitals_12w <- sqldf('select DEIDNUM as patientid, 
                           wt_kg as wt_kg_12w, 
                           HRSUP as hr_12w, 
                           SUPSYSBP as sbp_12w, 
                           SUPDIABP as dbp_12w 
                           from escape_physexam 
                           where FORM = "3-Month Follow-Up" ')


escape_vitals_12w$pupr_12w <- 
  escape_vitals_12w$sbp_12w-
  escape_vitals_12w$dbp_12w

escape_vitals_12w$map_12w <- 
  calc_map(diabp=escape_vitals_12w$dbp_12w,
           sysbp=escape_vitals_12w$sbp_12w,
           hr=escape_vitals_12w$hr_12w)

escape_vitals_24w <- sqldf('select DEIDNUM as patientid, 
                           wt_kg as wt_kg_24w, 
                           HRSUP as hr_24w, 
                           SUPSYSBP as sbp_24w, 
                           SUPDIABP as dbp_24w from escape_physexam 
                           where FORM = "6-Month Follow-Up" ')

escape_vitals_24w$pupr_24w <- 
  escape_vitals_24w$sbp_24w-
  escape_vitals_24w$dbp_24w

escape_vitals_24w$map_24w <- 
  calc_map(diabp=escape_vitals_24w$dbp_24w,
           sysbp=escape_vitals_24w$sbp_24w,
           hr=escape_vitals_24w$hr_24w)


acute_escape_vitals <- merge(escape_vitals_bl,escape_vitals_1d,by="patientid",all.x=T)
acute_escape_vitals <- merge(acute_escape_vitals,escape_vitals_2d,by="patientid",all.x=T)
acute_escape_vitals <- merge(acute_escape_vitals,escape_vitals_3d,by="patientid",all.x=T)
acute_escape_vitals <- merge(acute_escape_vitals,escape_vitals_4d,by="patientid",all.x=T)
acute_escape_vitals <- merge(acute_escape_vitals,escape_vitals_last,by="patientid",all.x=T)
acute_escape_vitals <- merge(acute_escape_vitals,escape_vitals_2w,by="patientid",all.x=T)
acute_escape_vitals <- merge(acute_escape_vitals,escape_vitals_8w,by="patientid",all.x=T)
acute_escape_vitals <- merge(acute_escape_vitals,escape_vitals_12w,by="patientid",all.x=T)
acute_escape_vitals <- merge(acute_escape_vitals,escape_vitals_24w,by="patientid",all.x=T)


acute_escape_vitals$wt_kg_bl_1d <- acute_escape_vitals$wt_kg_1d - acute_escape_vitals$wt_kg_bl
acute_escape_vitals$wt_kg_bl_2d <- acute_escape_vitals$wt_kg_2d - acute_escape_vitals$wt_kg_bl
acute_escape_vitals$wt_kg_bl_3d <- acute_escape_vitals$wt_kg_3d - acute_escape_vitals$wt_kg_bl
acute_escape_vitals$wt_kg_bl_4d <- acute_escape_vitals$wt_kg_4d - acute_escape_vitals$wt_kg_bl
acute_escape_vitals$wt_kg_bl_last <- acute_escape_vitals$wt_kg_last - acute_escape_vitals$wt_kg_bl
acute_escape_vitals$wt_kg_bl_last_pct <- acute_escape_vitals$wt_kg_bl_last/acute_escape_vitals$wt_kg_bl*100


### ESCAPE exercise

acute_escape_exercise <- escape_exercise_all[,c("DEIDNUM",
                                                "ILLWLKB",
                                                "ILLWLKD",
                                                "ILLVO2B",
                                                "ILLVO2D",
                                                "FTWLKB",
                                                "FTWLKD",
                                                "VO2B",
                                                "VO2D")]

acute_escape_exercise$FTWLKB[acute_escape_exercise$ILLWLKB==1] <- NA
acute_escape_exercise$FTWLKD[acute_escape_exercise$ILLWLKD==1] <- NA
acute_escape_exercise$ILLVO2B[acute_escape_exercise$ILLVO2B==1] <- NA
acute_escape_exercise$ILLVO2D[acute_escape_exercise$ILLVO2D==1] <- NA

names(acute_escape_exercise)[8:9] <- c("vo2_bl","vo2_dc")

acute_escape_exercise$sixmw_dist_m_bl_ac <- acute_escape_exercise$FTWLKB * 0.3048
acute_escape_exercise$sixmw_dist_m_dc_ac <- acute_escape_exercise$FTWLKD * 0.3048

acute_escape_exercise$vo2_bl_dc_ac <- acute_escape_exercise$vo2_dc - acute_escape_exercise$vo2_bl
acute_escape_exercise$sixmw_dist_m_bl_dc_ac <- acute_escape_exercise$sixmw_dist_m_dc - acute_escape_exercise$sixmw_dist_m_bl


acute_escape_outcomes <- merge(acute_escape_outcomes,
                               acute_escape_mechd,
                               by="patientid",
                               all.x=T)


acute_escape_outcomes <- merge(acute_escape_outcomes,
                               acute_escape_vitals,
                               by="patientid",
                               all.x=T)


# acute_escape_outcomes <- merge(acute_escape_outcomes,
#                                acute_escape_renal[,c(1:5,9:21)],
#                                by="patientid",
#                                all.x=T)

acute_escape_outcomes <- merge(acute_escape_outcomes,
                               acute_escape_io,
                               by="patientid",
                               all.x=T)

acute_escape_outcomes <- merge(acute_escape_outcomes,
                               acute_escape_vas,
                               by="patientid",
                               all.x=T)

acute_escape_outcomes <- merge(acute_escape_outcomes,
                               acute_escape_exercise,
                               by.x="patientid",
                               by.y="DEIDNUM",
                               all.x=T)

acute_escape_outcomes$study <- "ESCAPE"


####

missing_outcomes_escape <- acute_analysis_outcomes[!acute_analysis_outcomes %in% names(acute_escape_outcomes)]
acute_escape_outcomes[,missing_outcomes_escape] <- NA
acute_analysis_outcomes_escape <- acute_escape_exercise$vo2_bl_last[,acute_analysis_outcomes]

missing_acute_outcomes_escape <- acute_analysis_outcomes[!acute_analysis_outcomes %in% names(acute_escape_outcomes)]
acute_escape_outcomes[,missing_acute_outcomes_escape] <- NA
acute_analysis_outcomes_escape <- acute_escape_outcomes[,acute_analysis_outcomes]

rm(list=ls(pattern="\\bescape_."))


 
########################## ******* ROSE ******* ##################################
# 
# .%%%%%%%%...%%%%%%%...%%%%%%..%%%%%%%%
# .%%.....%%.%%.....%%.%%....%%.%%......
# .%%.....%%.%%.....%%.%%.......%%......
# .%%%%%%%%..%%.....%%..%%%%%%..%%%%%%..
# .%%...%%...%%.....%%.......%%.%%......
# .%%....%%..%%.....%%.%%....%%.%%......
# .%%.....%%..%%%%%%%...%%%%%%..%%%%%%%%
# 


thisfolder <- "~/Dropbox/BioLINCC files/ROSE/ROSE_2016a/Data/"

rose_base <- read.sas7bdat(paste(thisfolder,"a_base.sas7bdat",sep=""))
rose_endpts <- read.sas7bdat(paste(thisfolder,"a_endpts.sas7bdat",sep=""))
rose_visitsumm <- read.sas7bdat(paste(thisfolder,"a_visitsumm.sas7bdat",sep=""))
rose_assessmt <- read.sas7bdat(paste(thisfolder,"assessmt.sas7bdat",sep=""))
rose_discharg <- read.sas7bdat(paste(thisfolder,"discharg.sas7bdat",sep=""))
rose_rsfluid <- read.sas7bdat(paste(thisfolder,"rsfluid.sas7bdat",sep=""))
rose_vas <- read.sas7bdat(paste(thisfolder,"vas.sas7bdat",sep=""))
rose_fatigue <- read.sas7bdat(paste(thisfolder,"fatigue.sas7bdat",sep=""))
rose_swelling <- read.sas7bdat(paste(thisfolder,"swelling.sas7bdat",sep=""))
rose_wsymptom <- read.sas7bdat(paste(thisfolder,"wsymptom.sas7bdat",sep=""))

rose_endpts[rose_endpts=="NaN"] <- NA
rose_assessmt[rose_assessmt=="NaN"] <- NA
rose_visitsumm[rose_visitsumm=="NaN"] <- NA
rose_rsfluid[rose_rsfluid=="NaN"] <- NA
rose_vas[rose_vas=="NaN"] <- NA
rose_fatigue[rose_fatigue=="NaN"] <- NA
rose_swelling[rose_swelling=="NaN"] <- NA
rose_wsymptom[rose_wsymptom=="NaN"] <- NA


acute_rose_outcomes <- sqldf('select patnumb as patientid, DCALIVE=0 as inhosp_dth_status, DISCHDT as los from rose_discharg')
acute_rose_outcomes$inhosp_dth_status[is.na(acute_rose_outcomes$inhosp_dth_status)] <- 0

rose_visitsumm_bl <- sqldf('select PATNUMB as patientid, 
                           WTLBS/2.2 as wt_kg_bl,
                           LL_GFR as gfr_bl,
                           LL_BUN as bun_bl,
                           CL_NTPRO as ntbnp_bl
                           from rose_visitsumm 
                           where FORM = "BASELINE"')

rose_visitsumm_1d <- sqldf('select PATNUMB as patientid, 
                           WTLBS/2.2 as wt_kg_1d,
                           LL_GFR as gfr_1d, 
                           LL_BUN as bun_1d,
                           CL_NTPRO as ntbnp_1d
                           from rose_visitsumm 
                           where FORM = "24 HOURS"')

rose_visitsumm_2d <- sqldf('select PATNUMB as patientid, 
                           WTLBS/2.2 as wt_kg_2d,
                           LL_BUN as bun_2d,
                           CL_NTPRO as ntbnp_2d,
                           LL_GFR as gfr_2d 
                           from rose_visitsumm 
                           where FORM = "48 HOURS"')

rose_visitsumm_3d <- sqldf('select PATNUMB as patientid, 
                           WTLBS/2.2 as wt_kg_3d,
                           LL_BUN as bun_3d,
                           CL_NTPRO as ntbnp_3d,
                           LL_GFR as gfr_3d 
                           from rose_visitsumm 
                           where FORM = "72 HOURS"')

rose_visitsumm_7d <- sqldf('select PATNUMB as patientid, 
                             WTLBS/2.2 as wt_kg_7d,
                             LL_GFR as gfr_7d, 
                             LL_BUN as bun_7d
                             from rose_visitsumm 
                             where FORM = "DAY 7"')


####

acute_rose_labs <- merge(rose_visitsumm_bl,
                         rose_visitsumm_1d,
                         by="patientid",
                         all.x=T)

acute_rose_labs <- merge(acute_rose_labs,
                         rose_visitsumm_2d,
                         by="patientid",
                         all.x=T)

acute_rose_labs <- merge(acute_rose_labs,
                         rose_visitsumm_3d,
                         by="patientid",
                         all.x=T)

acute_rose_labs <- merge(acute_rose_labs,
                         rose_visitsumm_7d,
                         by="patientid",
                         all.x=T)


acute_rose_labs$gfr_last <- acute_rose_labs$gfr_7d

acute_rose_labs$gfr_last[is.na(acute_rose_labs$gfr_last)&
                              !is.na(acute_rose_labs$gfr_3d)] <-
  acute_rose_labs$gfr_3d[is.na(acute_rose_labs$gfr_last)&
                              !is.na(acute_rose_labs$gfr_3d)]

acute_rose_labs$gfr_last[is.na(acute_rose_labs$gfr_last)&
                              !is.na(acute_rose_labs$gfr_2d)] <-
  acute_rose_labs$gfr_2d[is.na(acute_rose_labs$gfr_last)&
                              !is.na(acute_rose_labs$gfr_2d)]

acute_rose_labs$gfr_last[is.na(acute_rose_labs$gfr_last)&
                              !is.na(acute_rose_labs$gfr_1d)] <-
  acute_rose_labs$gfr_1d[is.na(acute_rose_labs$gfr_last)&
                              !is.na(acute_rose_labs$gfr_1d)]


acute_rose_labs$bun_last <- acute_rose_labs$bun_7d

acute_rose_labs$bun_last[is.na(acute_rose_labs$bun_last)&
                           !is.na(acute_rose_labs$bun_3d)] <-
  acute_rose_labs$bun_3d[is.na(acute_rose_labs$bun_last)&
                           !is.na(acute_rose_labs$bun_3d)]

acute_rose_labs$bun_last[is.na(acute_rose_labs$bun_last)&
                           !is.na(acute_rose_labs$bun_2d)] <-
  acute_rose_labs$bun_2d[is.na(acute_rose_labs$bun_last)&
                           !is.na(acute_rose_labs$bun_2d)]

acute_rose_labs$bun_last[is.na(acute_rose_labs$bun_last)&
                           !is.na(acute_rose_labs$bun_1d)] <-
  acute_rose_labs$bun_1d[is.na(acute_rose_labs$bun_last)&
                           !is.na(acute_rose_labs$bun_1d)]


acute_rose_labs$wt_kg_last <- acute_rose_labs$wt_kg_7d

acute_rose_labs$wt_kg_last[is.na(acute_rose_labs$wt_kg_last)&
                           !is.na(acute_rose_labs$wt_kg_3d)] <-
  acute_rose_labs$wt_kg_3d[is.na(acute_rose_labs$wt_kg_last)&
                           !is.na(acute_rose_labs$wt_kg_3d)]

acute_rose_labs$wt_kg_last[is.na(acute_rose_labs$wt_kg_last)&
                           !is.na(acute_rose_labs$wt_kg_2d)] <-
  acute_rose_labs$wt_kg_2d[is.na(acute_rose_labs$wt_kg_last)&
                           !is.na(acute_rose_labs$wt_kg_2d)]

acute_rose_labs$wt_kg_last[is.na(acute_rose_labs$wt_kg_last)&
                           !is.na(acute_rose_labs$wt_kg_1d)] <-
  acute_rose_labs$wt_kg_1d[is.na(acute_rose_labs$wt_kg_last)&
                           !is.na(acute_rose_labs$wt_kg_1d)]


acute_rose_labs$ntbnp_last <- acute_rose_labs$ntbnp_3d

acute_rose_labs$ntbnp_last[is.na(acute_rose_labs$ntbnp_last)&
                             !is.na(acute_rose_labs$ntbnp_2d)] <-
  acute_rose_labs$ntbnp_2d[is.na(acute_rose_labs$ntbnp_last)&
                             !is.na(acute_rose_labs$ntbnp_2d)]

acute_rose_labs$ntbnp_last[is.na(acute_rose_labs$ntbnp_last)&
                             !is.na(acute_rose_labs$ntbnp_1d)] <-
  acute_rose_labs$ntbnp_1d[is.na(acute_rose_labs$ntbnp_last)&
                             !is.na(acute_rose_labs$ntbnp_1d)]

acute_rose_labs$ntbnp_bl_last <- acute_rose_labs$ntbnp_last - acute_rose_labs$ntbnp_bl
acute_rose_labs$wt_kg_bl_last <- acute_rose_labs$wt_kg_last - acute_rose_labs$wt_kg_bl
acute_rose_labs$wt_kg_bl_last_pct <- acute_rose_labs$wt_kg_bl_last/acute_rose_labs$wt_kg_bl * 100
acute_rose_labs$gfr_bl_last <- acute_rose_labs$gfr_last - acute_rose_labs$gfr_bl
acute_rose_labs$gfr_bl_last_pct <- acute_rose_labs$gfr_bl_last/acute_rose_labs$gfr_bl * 100

acute_rose_labs$bun_bl_last <- acute_rose_labs$bun_last - acute_rose_labs$bun_bl

### ROSE vital signs

rose_assessmt$MAP <- calc_map(diabp=rose_assessmt$BPDIA,
                              sysbp=rose_assessmt$BPSYS,
                              hr=rose_assessmt$HRATE)

rose_assessmt$PUPR <- rose_assessmt$BPSYS - rose_assessmt$BPDIA

rose_assessmt_bl <- sqldf('select PATNUMB as patientid, 
                           HRATE as hr_bl,
                           BPSYS as sbp_bl,
                           BPDIA as dbp_bl,
                           MAP as map_bl,
                           PUPR as pupr_bl,
                           JVP as jvp_bl,
                           RALES as rales_bl,
                           AUSCULTN as s3_bl,
                           ASCITES as ascites_bl,
                           PERIEDMA as edema_bl,
                           NYHA as nyha_bl,
                           ORTHPNEA as orthopnea_bl,
                           DYSPNEA as dyspnea_bl
                           from rose_assessmt 
                           where FORM = "BASELINE"')

rose_assessmt_1d <- sqldf('select PATNUMB as patientid, 
                           HRATE as hr_1d,
                           BPSYS as sbp_1d,
                           BPDIA as dbp_1d,
                           MAP as map_1d,
                           PUPR as pupr_1d,
                           JVP as jvp_1d,
                           RALES as rales_1d,
                           AUSCULTN as s3_1d,
                           ASCITES as ascites_1d,
                           PERIEDMA as edema_1d,
                           NYHA as nyha_1d,
                           ORTHPNEA as orthopnea_1d,
                           DYSPNEA as dyspnea_1d
                           from rose_assessmt 
                           where FORM = "24 HOURS"')

rose_assessmt_2d <- sqldf('select PATNUMB as patientid, 
                           HRATE as hr_2d,
                           BPSYS as sbp_2d,
                           BPDIA as dbp_2d,
                           MAP as map_2d,
                           PUPR as pupr_2d,
                           JVP as jvp_2d,
                           RALES as rales_2d,
                           AUSCULTN as s3_2d,
                           ASCITES as ascites_2d,
                           PERIEDMA as edema_2d,
                           NYHA as nyha_2d,
                           ORTHPNEA as orthopnea_2d,
                           DYSPNEA as dyspnea_2d
                           from rose_assessmt 
                           where FORM = "48 HOURS"')

rose_assessmt_3d <- sqldf('select PATNUMB as patientid, 
                           HRATE as hr_3d,
                           BPSYS as sbp_3d,
                           BPDIA as dbp_3d,
                           MAP as map_3d,
                           PUPR as pupr_3d,
                           JVP as jvp_3d,
                           RALES as rales_3d,
                           AUSCULTN as s3_3d,
                           ASCITES as ascites_3d,
                           PERIEDMA as edema_3d,
                           NYHA as nyha_3d,
                           ORTHPNEA as orthopnea_3d,
                           DYSPNEA as dyspnea_3d
                           from rose_assessmt 
                           where FORM = "72 HOURS"')

rose_assessmt_7d <- sqldf('select PATNUMB as patientid, 
                           HRATE as hr_7d,
                           BPSYS as sbp_7d,
                           BPDIA as dbp_7d,
                           MAP as map_7d,
                           PUPR as pupr_7d,
                           JVP as jvp_7d,
                           RALES as rales_7d,
                           AUSCULTN as s3_7d,
                           ASCITES as ascites_7d,
                           PERIEDMA as edema_7d,
                           NYHA as nyha_7d,
                           ORTHPNEA as orthopnea_7d,
                           DYSPNEA as dyspnea_7d
                           from rose_assessmt 
                             where FORM = "DAY 7"')


rose_phys_exam <-
  merge(rose_assessmt_bl,
        rose_assessmt_1d)

rose_phys_exam <-
  merge(rose_phys_exam,
        rose_assessmt_2d)

rose_phys_exam <-
  merge(rose_phys_exam,
        rose_assessmt_3d)

rose_phys_exam <-
  merge(rose_phys_exam,
        rose_assessmt_7d)


### ROSE input/output

rose_rsfluid$io_balance <- 
  rose_rsfluid$RSIVIN+
  rose_rsfluid$RSORALIN-
  rose_rsfluid$RSUROUT-
  rose_rsfluid$RSNUROUT


rose_io_1d <- sqldf('select PATNUMB as patientid, 
                    io_balance as io_cum_1d 
                    from rose_rsfluid 
                    where FORM = "24 HOURS"') 


rose_io_2d <- sqldf('select PATNUMB as patientid, 
                    io_balance as io_balance_2d
                    from rose_rsfluid 
                    where FORM = "48 HOURS"') 

rose_io_cum_2d <- sqldf('select PATNUMB as patientid, 
                    sum(io_balance) as io_cum_2d
                    from rose_rsfluid 
                    where FORM in ("24 HOURS","48 HOURS")
                        group by PATNUMB') 


rose_io_3d <- sqldf('select PATNUMB as patientid, 
                    io_balance as io_balance_3d 
                    from rose_rsfluid 
                    where FORM = "72 HOURS"') 


rose_io_cum_3d <- sqldf('select PATNUMB as patientid, 
                    sum(io_balance) as io_cum_3d
                    from rose_rsfluid 
                    where FORM in ("24 HOURS","48 HOURS","72 HOURS")
                        group by PATNUMB') 


rose_io_total <- sqldf('select PATNUMB as patientid, 
                      sum(io_balance) as io_cum_total
                      from rose_rsfluid
                      group by PATNUMB') 


acute_rose_io <- merge(rose_io_1d,rose_io_2d,by="patientid",all.x=T)
acute_rose_io <- merge(acute_rose_io,rose_io_cum_2d,by="patientid",all.x=T)
acute_rose_io <- merge(acute_rose_io,rose_io_3d,by="patientid",all.x=T)
acute_rose_io <- merge(acute_rose_io,rose_io_cum_3d,by="patientid",all.x=T)
acute_rose_io <- merge(acute_rose_io,rose_io_total,by="patientid",all.x=T)

### ROSE VAS

rose_vas$FATVAS <- rose_fatigue$FATVAS
rose_vas$BSVAS <- rose_swelling$BSVAS

rose_vas_bl <- sqldf('select PATNUMB as patientid, 
                     DYSPVAS as dyspnea_vas_bl, 
                     GLOBLVAS as global_vas_bl,
                     FATVAS as fatigue_vas_bl,
                     BSVAS as swelling_vas_bl
                     from rose_vas 
                     where FORM = "BASELINE"')

rose_vas_1d <- sqldf('select PATNUMB as patientid, 
                     DYSPVAS as dyspnea_vas_1d, 
                     GLOBLVAS as global_vas_1d,
                     FATVAS as fatigue_vas_1d,
                     BSVAS as swelling_vas_1d
                     from rose_vas 
                     where FORM = "24 HOURS"')

rose_vas_2d <- sqldf('select PATNUMB as patientid, 
                     DYSPVAS as dyspnea_vas_2d, 
                     GLOBLVAS as global_vas_2d, 
                     FATVAS as fatigue_vas_2d,
                     BSVAS as swelling_vas_2d
                     from rose_vas 
                     where FORM = "48 HOURS"')

rose_vas_3d <- sqldf('select PATNUMB as patientid, 
                     DYSPVAS as dyspnea_vas_3d, 
                     GLOBLVAS as global_vas_3d,
                     FATVAS as fatigue_vas_3d,
                     BSVAS as swelling_vas_3d
                     from rose_vas
                     where FORM = "72 HOURS"')

acute_rose_vas <- merge(rose_vas_bl,rose_vas_1d,all.x=T)
acute_rose_vas <- merge(acute_rose_vas,rose_vas_2d,all.x=T)
acute_rose_vas <- merge(acute_rose_vas,rose_vas_3d,all.x=T)


acute_rose_vas$global_vas_last <- acute_rose_vas$global_vas_3d

acute_rose_vas$global_vas_last[is.na(acute_rose_vas$global_vas_last)&
                                 !is.na(acute_rose_vas$global_vas_2d)] <- 
  acute_rose_vas$global_vas_2d[is.na(acute_rose_vas$global_vas_last)&
                                 !is.na(acute_rose_vas$global_vas_2d)]

acute_rose_vas$global_vas_last[is.na(acute_rose_vas$global_vas_last)&
                                 !is.na(acute_rose_vas$global_vas_1d)] <- 
  acute_rose_vas$global_vas_1d[is.na(acute_rose_vas$global_vas_last)&
                                 !is.na(acute_rose_vas$global_vas_1d)]


acute_rose_vas$dyspnea_vas_last <- acute_rose_vas$dyspnea_vas_3d

acute_rose_vas$dyspnea_vas_last[is.na(acute_rose_vas$dyspnea_vas_last)&
                                 !is.na(acute_rose_vas$dyspnea_vas_2d)] <- 
  acute_rose_vas$dyspnea_vas_2d[is.na(acute_rose_vas$dyspnea_vas_last)&
                                 !is.na(acute_rose_vas$dyspnea_vas_2d)]

acute_rose_vas$dyspnea_vas_last[is.na(acute_rose_vas$dyspnea_vas_last)&
                                 !is.na(acute_rose_vas$dyspnea_vas_1d)] <- 
  acute_rose_vas$dyspnea_vas_1d[is.na(acute_rose_vas$dyspnea_vas_last)&
                                 !is.na(acute_rose_vas$dyspnea_vas_1d)]


acute_rose_vas$fatigue_vas_last <- acute_rose_vas$fatigue_vas_3d

acute_rose_vas$fatigue_vas_last[is.na(acute_rose_vas$fatigue_vas_last)&
                                  !is.na(acute_rose_vas$fatigue_vas_2d)] <- 
  acute_rose_vas$fatigue_vas_2d[is.na(acute_rose_vas$fatigue_vas_last)&
                                  !is.na(acute_rose_vas$fatigue_vas_2d)]

acute_rose_vas$fatigue_vas_last[is.na(acute_rose_vas$fatigue_vas_last)&
                                  !is.na(acute_rose_vas$fatigue_vas_1d)] <- 
  acute_rose_vas$fatigue_vas_1d[is.na(acute_rose_vas$fatigue_vas_last)&
                                  !is.na(acute_rose_vas$fatigue_vas_1d)]


acute_rose_vas$swelling_vas_last <- acute_rose_vas$swelling_vas_3d

acute_rose_vas$swelling_vas_last[is.na(acute_rose_vas$swelling_vas_last)&
                                  !is.na(acute_rose_vas$swelling_vas_2d)] <- 
  acute_rose_vas$swelling_vas_2d[is.na(acute_rose_vas$swelling_vas_last)&
                                  !is.na(acute_rose_vas$swelling_vas_2d)]

acute_rose_vas$swelling_vas_last[is.na(acute_rose_vas$swelling_vas_last)&
                                  !is.na(acute_rose_vas$swelling_vas_1d)] <- 
  acute_rose_vas$swelling_vas_1d[is.na(acute_rose_vas$swelling_vas_last)&
                                  !is.na(acute_rose_vas$swelling_vas_1d)]


acute_rose_vas$global_vas_bl_last <- acute_rose_vas$global_vas_last - acute_rose_vas$global_vas_bl
acute_rose_vas$dyspnea_vas_bl_last <- acute_rose_vas$dyspnea_vas_last - acute_rose_vas$dyspnea_vas_bl
acute_rose_vas$fatigue_vas_bl_last <- acute_rose_vas$fatigue_vas_last - acute_rose_vas$fatigue_vas_bl
acute_rose_vas$swelling_vas_bl_last <- acute_rose_vas$swelling_vas_last - acute_rose_vas$swelling_vas_bl


acute_rose_vas$wsymptom[rose_wsymptom$WORST==1] <- "Dyspnea"
acute_rose_vas$wsymptom[rose_wsymptom$WORST==2] <- "Fatigue"
acute_rose_vas$wsymptom[rose_wsymptom$WORST==3] <- "Swelling"

acute_rose_outcomes <- merge(acute_rose_outcomes,acute_rose_labs,by="patientid",all.x=T)
acute_rose_outcomes <- merge(acute_rose_outcomes,acute_rose_vas,by="patientid",all.x=T)
acute_rose_outcomes <- merge(acute_rose_outcomes,acute_rose_io,by="patientid",all.x=T)


########

acute_rose_outcomes$study <- "ROSE"

missing_acute_outcomes_rose <- acute_analysis_outcomes[!acute_analysis_outcomes %in% names(acute_rose_outcomes)]
acute_rose_outcomes[,missing_acute_outcomes_rose] <- NA
acute_analysis_outcomes_rose <- acute_rose_outcomes[,acute_analysis_outcomes]

rm(list=ls(pattern="\\brose_."))


#----------------------------------------------------------------------------------------#
#----------------------------------------------------------------------------------------#
########################### ********** ATHENA ********** ####################################

# ....%%%....%%%%%%%%.%%.....%%.%%%%%%%%.%%....%%....%%%...
# ...%%.%%......%%....%%.....%%.%%.......%%%...%%...%%.%%..
# ..%%...%%.....%%....%%.....%%.%%.......%%%%..%%..%%...%%.
# .%%.....%%....%%....%%%%%%%%%.%%%%%%...%%.%%.%%.%%.....%%
# .%%%%%%%%%....%%....%%.....%%.%%.......%%..%%%%.%%%%%%%%%
# .%%.....%%....%%....%%.....%%.%%.......%%...%%%.%%.....%%
# .%%.....%%....%%....%%.....%%.%%%%%%%%.%%....%%.%%.....%%

athena_endpts <- read.csv("~/Dropbox/BioLINCC files/ATHENA/data/csv/a_endpts.csv",
                          na.strings=c("","NaN","NULL"),
                          stringsAsFactors = F)

athena_endpts$inhosp_dth_status[athena_endpts$DISCHARGED==2] <- 1
athena_endpts$inhosp_dth_status[athena_endpts$DISCHARGED %in% c(0,1)] <- 0

athena_visitsumm <- read.csv("~/Dropbox/BioLINCC files/ATHENA/data/csv/a_visitsumm.csv",
                          na.strings=c("","NaN","NULL"),
                          stringsAsFactors = F)


athena_visitsumm_bl <- sqldf("select PATNUMB as patientid,
                          WTLBS/2.2 as wt_kg_bl,
                          CL_NTPRO as ntbnp_bl
                          from athena_visitsumm
                          where VISIT = 'BASE'")

athena_visitsumm_1d <- sqldf("select PATNUMB as patientid,
                          WTLBS/2.2 as wt_kg_1d,
                          CL_NTPRO as ntbnp_1d
                          from athena_visitsumm
                          where VISIT = '24H'")

athena_visitsumm_2d <- sqldf("select PATNUMB as patientid,
                          WTLBS/2.2 as wt_kg_2d,
                          CL_NTPRO as ntbnp_2d
                          from athena_visitsumm
                          where VISIT = '48H'")

athena_visitsumm_3d <- sqldf("select PATNUMB as patientid,
                          WTLBS/2.2 as wt_kg_3d,
                          CL_NTPRO as ntbnp_3d
                          from athena_visitsumm
                          where VISIT = '72H'")

athena_visitsumm_4d <- sqldf("select PATNUMB as patientid,
                          WTLBS/2.2 as wt_kg_4d,
                          CL_NTPRO as ntbnp_4d
                          from athena_visitsumm
                          where VISIT = '96H'")

athena_visitsumm_dc <- sqldf("select PATNUMB as patientid,
                          WTLBS/2.2 as wt_kg_dc,
                          CL_NTPRO as ntbnp_dc
                          from athena_visitsumm
                          where VISIT = 'DISCHRGE'")


acute_athena_visitsumm <- merge(athena_visitsumm_bl,
                                athena_visitsumm_1d,
                              by="patientid",
                              all.x=T)

acute_athena_visitsumm <- merge(acute_athena_visitsumm,
                                athena_visitsumm_2d,
                                by="patientid",
                                all.x=T)

acute_athena_visitsumm <- merge(acute_athena_visitsumm,
                                athena_visitsumm_3d,
                                by="patientid",
                                all.x=T)

acute_athena_visitsumm <- merge(acute_athena_visitsumm,
                                athena_visitsumm_4d,
                                by="patientid",
                                all.x=T)


acute_athena_visitsumm <- merge(acute_athena_visitsumm,
                                athena_visitsumm_dc,
                                by="patientid",
                                all.x=T)


acute_athena_visitsumm$wt_kg_last <- acute_athena_visitsumm$wt_kg_dc

acute_athena_visitsumm$wt_kg_last[is.na(acute_athena_visitsumm$wt_kg_last)&
                                     !is.na(acute_athena_visitsumm$wt_kg_4d)] <-
  acute_athena_visitsumm$wt_kg_4d[is.na(acute_athena_visitsumm$wt_kg_last)&
                                     !is.na(acute_athena_visitsumm$wt_kg_4d)]

acute_athena_visitsumm$wt_kg_last[is.na(acute_athena_visitsumm$wt_kg_last)&
                                     !is.na(acute_athena_visitsumm$wt_kg_3d)] <-
  acute_athena_visitsumm$wt_kg_3d[is.na(acute_athena_visitsumm$wt_kg_last)&
                                     !is.na(acute_athena_visitsumm$wt_kg_3d)]

acute_athena_visitsumm$wt_kg_last[is.na(acute_athena_visitsumm$wt_kg_last)&
                                     !is.na(acute_athena_visitsumm$wt_kg_2d)] <-
  acute_athena_visitsumm$wt_kg_2d[is.na(acute_athena_visitsumm$wt_kg_last)&
                                     !is.na(acute_athena_visitsumm$wt_kg_2d)]

acute_athena_visitsumm$wt_kg_last[is.na(acute_athena_visitsumm$wt_kg_last)&
                                     !is.na(acute_athena_visitsumm$wt_kg_1d)] <-
  acute_athena_visitsumm$wt_kg_1d[is.na(acute_athena_visitsumm$wt_kg_last)&
                                     !is.na(acute_athena_visitsumm$wt_kg_1d)]


acute_athena_visitsumm$wt_kg_bl_last <- 
  acute_athena_visitsumm$wt_kg_last - 
  acute_athena_visitsumm$wt_kg_bl

acute_athena_visitsumm$wt_kg_bl_last_pct <- 
  acute_athena_visitsumm$wt_kg_bl_last/
  acute_athena_visitsumm$wt_kg_bl * 100


#### ATHENA exam

athena_exam <- read.csv("~/Dropbox/BioLINCC files/ATHENA/data/csv/exam.csv",
                         na.strings=c("","NaN","NULL"),
                         stringsAsFactors = F)

athena_exam$DYSLIKRT[athena_exam$DYSLIKRT==97] <- NA
athena_exam$DYSVISDN[athena_exam$DYSVISDN==97] <- NA
athena_exam$NYHA[athena_exam$NYHA==97] <- NA

athena_exam$MAP <- calc_map(diabp=athena_exam$BPDIA,
                            sysbp=athena_exam$BPSYS,
                            hr=athena_exam$HRATEVAL)

athena_exam$PUPR <- athena_exam$BPSYS - athena_exam$BPDIA


athena_exam_bl <- sqldf("select PATNUMB as patientid,
                        HRATEVAL as hr_bl,
                        BPSYS as sbp_bl,
                        BPDIA as dbp_bl,
                        MAP as map_bl,
                        PUPR as pupr_bl,
                        DYSANAVL as dyspnea_vas_bl
                        from athena_exam
                        where VISIT = 'BASE'")

athena_exam_1d <- sqldf("select PATNUMB as patientid,
                        HRATEVAL as hr_1d,
                        BPSYS as sbp_1d,
                        BPDIA as dbp_1d,
                        MAP as map_1d,
                        PUPR as pupr_1d,
                        DYSANAVL as dyspnea_vas_1d
                        from athena_exam
                        where VISIT = '24H'")

athena_exam_2d <- sqldf("select PATNUMB as patientid,
                        HRATEVAL as hr_2d,
                        BPSYS as sbp_2d,
                        BPDIA as dbp_2d,
                        MAP as map_2d,
                        PUPR as pupr_2d,
                        DYSANAVL as dyspnea_vas_2d
                        from athena_exam
                        where VISIT = '48H'")

athena_exam_3d <- sqldf("select PATNUMB as patientid,
                        HRATEVAL as hr_3d,
                        BPSYS as sbp_3d,
                        BPDIA as dbp_3d,
                        MAP as map_3d,
                        PUPR as pupr_3d,
                        DYSANAVL as dyspnea_vas_3d
                        from athena_exam
                        where VISIT = '72H'")

athena_exam_4d <- sqldf("select PATNUMB as patientid,
                        HRATEVAL as hr_4d,
                        BPSYS as sbp_4d,
                        BPDIA as dbp_4d,
                        MAP as map_4d,
                        PUPR as pupr_4d,
                        DYSANAVL as dyspnea_vas_4d
                        from athena_exam
                        where VISIT = '96H'")

athena_exam_dc <- sqldf("select PATNUMB as patientid,
                        HRATEVAL as hr_dc,
                        BPSYS as sbp_dc,
                        BPDIA as dbp_dc,
                        MAP as map_dc,
                        PUPR as pupr_dc,
                        DYSANAVL as dyspnea_vas_dc
                        from athena_exam
                        where VISIT = 'DISCHRGE'")

acute_athena_exam <- merge(athena_exam_bl,
                           athena_exam_1d,
                           by="patientid",
                           all.x=T)

acute_athena_exam <- merge(acute_athena_exam,
                           athena_exam_2d,
                           by="patientid",
                           all.x=T)

acute_athena_exam <- merge(acute_athena_exam,
                           athena_exam_3d,
                           by="patientid",
                           all.x=T)

acute_athena_exam <- merge(acute_athena_exam,
                           athena_exam_4d,
                           by="patientid",
                           all.x=T)

acute_athena_exam <- merge(acute_athena_exam,
                           athena_exam_dc,
                           by="patientid",
                           all.x=T)


####

acute_athena_exam$dyspnea_vas_last <- acute_athena_exam$dyspnea_vas_dc

acute_athena_exam$dyspnea_vas_last[is.na(acute_athena_exam$dyspnea_vas_last)&
                              !is.na(acute_athena_exam$dyspnea_vas_4d)] <-
  acute_athena_exam$dyspnea_vas_4d[is.na(acute_athena_exam$dyspnea_vas_last)&
                              !is.na(acute_athena_exam$dyspnea_vas_4d)]

acute_athena_exam$dyspnea_vas_last[is.na(acute_athena_exam$dyspnea_vas_last)&
                                      !is.na(acute_athena_exam$dyspnea_vas_3d)] <-
  acute_athena_exam$dyspnea_vas_3d[is.na(acute_athena_exam$dyspnea_vas_last)&
                                      !is.na(acute_athena_exam$dyspnea_vas_3d)]

acute_athena_exam$dyspnea_vas_last[is.na(acute_athena_exam$dyspnea_vas_last)&
                              !is.na(acute_athena_exam$dyspnea_vas_2d)] <-
  acute_athena_exam$dyspnea_vas_2d[is.na(acute_athena_exam$dyspnea_vas_last)&
                              !is.na(acute_athena_exam$dyspnea_vas_2d)]

acute_athena_exam$dyspnea_vas_last[is.na(acute_athena_exam$dyspnea_vas_last)&
                              !is.na(acute_athena_exam$dyspnea_vas_1d)] <-
  acute_athena_exam$dyspnea_vas_1d[is.na(acute_athena_exam$dyspnea_vas_last)&
                              !is.na(acute_athena_exam$dyspnea_vas_1d)]


acute_athena_exam$hr_last <- acute_athena_exam$hr_dc

acute_athena_exam$hr_last[is.na(acute_athena_exam$hr_last)&
                                      !is.na(acute_athena_exam$hr_4d)] <-
  acute_athena_exam$hr_4d[is.na(acute_athena_exam$hr_last)&
                                      !is.na(acute_athena_exam$hr_4d)]

acute_athena_exam$hr_last[is.na(acute_athena_exam$hr_last)&
                                      !is.na(acute_athena_exam$hr_3d)] <-
  acute_athena_exam$hr_3d[is.na(acute_athena_exam$hr_last)&
                                      !is.na(acute_athena_exam$hr_3d)]

acute_athena_exam$hr_last[is.na(acute_athena_exam$hr_last)&
                                      !is.na(acute_athena_exam$hr_2d)] <-
  acute_athena_exam$hr_2d[is.na(acute_athena_exam$hr_last)&
                                      !is.na(acute_athena_exam$hr_2d)]

acute_athena_exam$hr_last[is.na(acute_athena_exam$hr_last)&
                                      !is.na(acute_athena_exam$hr_1d)] <-
  acute_athena_exam$hr_1d[is.na(acute_athena_exam$hr_last)&
                                      !is.na(acute_athena_exam$hr_1d)]


athena_exam_melt <- melt(athena_exam,id.vars = c("PATNUMB","VISIT"))
athena_exam_melt <- subset(athena_exam_melt,!is.na(value)&!variable %in% c("FORM"))


#### ATHENA labs

athena_labs1 <- read.csv("~/Dropbox/BioLINCC files/ATHENA/data/csv/labs1.csv",
                         na.strings=c("","NaN","NULL"),
                         stringsAsFactors = F)


athena_labs2 <- read.csv("~/Dropbox/BioLINCC files/ATHENA/data/csv/labs2.csv",
                         na.strings=c("","NaN","NULL"),
                         stringsAsFactors = F)

athena_labs1$BUNVAL[athena_labs1$BUNUNT==1&!is.na(athena_labs1$BUNUNT)] <- athena_labs1$BUNVAL[athena_labs1$BUNUNT==1&!is.na(athena_labs1$BUNVAL)] * 2.8
athena_labs2$BUNVAL2[athena_labs2$BUNUNT2==1&!is.na(athena_labs2$BUNUNT2)] <- athena_labs2$BUNVAL2[athena_labs2$BUNUNT2==1&!is.na(athena_labs2$BUNUNT2)] * 2.8

athena_labs_bl <- sqldf("select PATNUMB as patientid,
                         BUNVAL as bun_bl,
                         BNPVAL as bnp_bl
                         from athena_labs1
                         where VISIT = 'BASE'")


athena_labs_1d <- sqldf("select PATNUMB as patientid,
                         BUNVAL2 as bun_1d
                         from athena_labs2
                         where VISIT = '24H'")

athena_labs_2d <- sqldf("select PATNUMB as patientid,
                         BUNVAL2 as bun_2d
                         from athena_labs2
                         where VISIT = '48H'")


athena_labs_3d <- sqldf("select PATNUMB as patientid,
                         BUNVAL2 as bun_3d
                         from athena_labs2
                         where VISIT = '72H'")

athena_labs_4d <- sqldf("select PATNUMB as patientid,
                         BUNVAL2 as bun_4d
                         from athena_labs2
                         where VISIT = '96H'")

athena_labs_dc <- sqldf("select PATNUMB as patientid,
                         BUNVAL as bun_dc,
                         BNPVAL as bnp_dc
                         from athena_labs1
                         where VISIT = 'DISCHRGE'")

acute_athena_labs <- merge(athena_labs_bl,athena_labs_1d, by="patientid",all.x=T)
acute_athena_labs <- merge(acute_athena_labs,athena_labs_2d, by="patientid",all.x=T)
acute_athena_labs <- merge(acute_athena_labs,athena_labs_3d, by="patientid",all.x=T)
acute_athena_labs <- merge(acute_athena_labs,athena_labs_4d, by="patientid",all.x=T)
acute_athena_labs <- merge(acute_athena_labs,athena_labs_dc, by="patientid",all.x=T)


acute_athena_labs$bun_last <- acute_athena_labs$bun_dc

acute_athena_labs$bun_last[is.na(acute_athena_labs$bun_last)&
                              !is.na(acute_athena_labs$bun_3d)] <-
  acute_athena_labs$bun_3d[is.na(acute_athena_labs$bun_last)&
                              !is.na(acute_athena_labs$bun_3d)]

acute_athena_labs$bun_last[is.na(acute_athena_labs$bun_last)&
                              !is.na(acute_athena_labs$bun_2d)] <-
  acute_athena_labs$bun_2d[is.na(acute_athena_labs$bun_last)&
                              !is.na(acute_athena_labs$bun_2d)]

acute_athena_labs$bun_last[is.na(acute_athena_labs$bun_last)&
                              !is.na(acute_athena_labs$bun_1d)] <-
  acute_athena_labs$bun_1d[is.na(acute_athena_labs$bun_last)&
                              !is.na(acute_athena_labs$bun_1d)]


#### Renal function

athena_labs1_kcr <- sqldf("select PATNUMB, 
                          VISIT,
                          CREATVAL as CREATVL3, 
                          POTASVAL as POTASVL3
                          from athena_labs1") 

athena_kcreat <- read.csv("~/Dropbox/BioLINCC files/ATHENA/data/csv/kcreat.csv",
                         na.strings=c("","NaN","NULL"),
                         stringsAsFactors = F)[,c("PATNUMB","VISIT","CREATVL3","POTASVL3")]

athena_kcreat <- rbind(athena_labs1_kcr,
                       athena_kcreat)

athena_demog <- read.csv("~/Dropbox/BioLINCC files/ATHENA/data/csv/demog.csv",
                         na.strings=c("","NaN","NULL"),
                         stringsAsFactors = F)

athena_base <- read.csv("~/Dropbox/BioLINCC files/ATHENA/data/csv/a_base.csv",
         na.strings=c("","NaN","NULL"),
         stringsAsFactors = F)

athena_kcreat <- merge(athena_kcreat,athena_base[,c("PATNUMB","AGE","RACE")],by="PATNUMB",al.x=T)
athena_kcreat <- merge(athena_kcreat,athena_demog[,c("PATNUMB","SEX")],by="PATNUMB",al.x=T)

athena_kcreat$gfr <- calc_MDRD4(athena_kcreat, cr="CREATVL3", black=3)


athena_renal_bl <- sqldf("select PATNUMB as patientid,
                         CREATVL3 as creatinine_bl,
                         POTASVL3 as k_bl,
                         gfr as gfr_bl
                         from athena_kcreat
                         where VISIT = 'BASE'")

athena_renal_1d <- sqldf("select PATNUMB as patientid,
                         CREATVL3 as creatinine_1d,
                         POTASVL3 as k_1d,
                         gfr as gfr_1d
                         from athena_kcreat
                         where VISIT = '24H'")

athena_renal_2d <- sqldf("select PATNUMB as patientid,
                         CREATVL3 as creatinine_2d,
                         POTASVL3 as k_2d,
                         gfr as gfr_2d
                         from athena_kcreat
                         where VISIT = '48H'")

athena_renal_3d <- sqldf("select PATNUMB as patientid,
                         CREATVL3 as creatinine_3d,
                         POTASVL3 as k_3d,
                         gfr as gfr_3d
                         from athena_kcreat
                         where VISIT = '72H'")

athena_renal_4d <- sqldf("select PATNUMB as patientid,
                         CREATVL3 as creatinine_4d,
                         POTASVL3 as k_4d,
                         gfr as gfr_4d
                         from athena_kcreat
                         where VISIT = '96H'")

athena_renal_dc <- sqldf("select PATNUMB as patientid,
                         CREATVL3 as creatinine_dc,
                         POTASVL3 as k_dc,
                         gfr as gfr_dc
                         from athena_kcreat
                         where VISIT = 'DISCHRGE'")


acute_athena_renal <- merge(athena_renal_bl,
                            athena_renal_1d,
                            by=c("patientid"),
                            all.x=T)


acute_athena_renal <- merge(acute_athena_renal,
                            athena_renal_2d,
                            by=c("patientid"),
                            all.x=T)

acute_athena_renal <- merge(acute_athena_renal,
                            athena_renal_3d,
                            by=c("patientid"),
                            all.x=T)

acute_athena_renal <- merge(acute_athena_renal,
                            athena_renal_4d,
                            by=c("patientid"),
                            all.x=T)

acute_athena_renal <- merge(acute_athena_renal,
                            athena_renal_dc,
                            by=c("patientid"),
                            all.x=T)


acute_athena_renal$gfr_last <- acute_athena_renal$gfr_dc

acute_athena_renal$gfr_last[is.na(acute_athena_renal$gfr_last)&
                           !is.na(acute_athena_renal$gfr_3d)] <-
  acute_athena_renal$gfr_3d[is.na(acute_athena_renal$gfr_last)&
                           !is.na(acute_athena_renal$gfr_3d)]

acute_athena_renal$gfr_last[is.na(acute_athena_renal$gfr_last)&
                           !is.na(acute_athena_renal$gfr_2d)] <-
  acute_athena_renal$gfr_2d[is.na(acute_athena_renal$gfr_last)&
                           !is.na(acute_athena_renal$gfr_2d)]

acute_athena_renal$gfr_last[is.na(acute_athena_renal$gfr_last)&
                           !is.na(acute_athena_renal$gfr_1d)] <-
  acute_athena_renal$gfr_1d[is.na(acute_athena_renal$gfr_last)&
                           !is.na(acute_athena_renal$gfr_1d)]


acute_athena_renal$gfr_bl_1d <- acute_athena_renal$gfr_1d - acute_athena_renal$gfr_bl
acute_athena_renal$gfr_bl_2d <- acute_athena_renal$gfr_2d - acute_athena_renal$gfr_bl
acute_athena_renal$gfr_bl_3d <- acute_athena_renal$gfr_3d - acute_athena_renal$gfr_bl
acute_athena_renal$gfr_bl_4d <- acute_athena_renal$gfr_4d - acute_athena_renal$gfr_bl
acute_athena_renal$gfr_bl_dc <- acute_athena_renal$gfr_dc - acute_athena_renal$gfr_bl

acute_athena_renal$gfr_bl_last <- acute_athena_renal$gfr_last - acute_athena_renal$gfr_bl


acute_athena_renal$gfr_bl_1d_pct <- acute_athena_renal$gfr_bl_1d/acute_athena_renal$gfr_bl * 100
acute_athena_renal$gfr_bl_2d_pct <- acute_athena_renal$gfr_bl_2d/acute_athena_renal$gfr_bl * 100
acute_athena_renal$gfr_bl_3d_pct <- acute_athena_renal$gfr_bl_3d/acute_athena_renal$gfr_bl * 100
acute_athena_renal$gfr_bl_4d_pct <- acute_athena_renal$gfr_bl_4d/acute_athena_renal$gfr_bl * 100
acute_athena_renal$gfr_bl_dc_pct <- acute_athena_renal$gfr_bl_dc/acute_athena_renal$gfr_bl * 100

acute_athena_renal$gfr_bl_last_pct <- acute_athena_renal$gfr_bl_last/acute_athena_renal$gfr_bl * 100


### Summary endpoints

acute_athena_outcomes <- sqldf('select PATNUMB as patientid, 
                          inhosp_dth_status,
                          TIME2DC as los,
                          CONGSCORE_BL as congestion_bl,
                          CONGSCORE_BLPOST as congestion_bl_dc,
                          DYSPVAS_BL as dyspnea_vas_bl,
                          DYSPVAS_BLPOST as dyspnea_vas_bl_dc,
                          WTLBS_BL/2.2 as wt_kg_bl,
                          WTLBS_POST/2.2 as wt_kg_dc,
                          WTLBS_BLPOST/2.2 as wt_kg_bl_dc,
                          (WTLBS_BLPOST/2.2/WTLBS_BL)*100 as wt_kg_bl_dc_pct,
                          FEDIURDS_PH as daily_furo_equiv_bl,
                          CREAT_BL24 as cr_bl_1d,
                          CREAT_BL48 as cr_bl_2d,
                          CREAT_BL72 as cr_bl_3d,
                          CREAT_BL96 as cr_bl_4d,
                          -NFL_24 as io_cum_1d,
                          -NFL_24 as io_balance_1d,
                          -NFL_2448 as io_cum_2d,
                          -(NFL_2448-NFL_24) as io_balance_2d,
                          -(NFL_244872) as io_cum_3d,
                          -(NFL_244872-NFL_2448) as io_balance_3d,
                          -NFL_24487296 as io_cum_4d,
                          -(NFL_24487296-NFL_244872) as io_balance_4d,
                          LOG_CL_NTPRO_BL as log_ntbnp_bl,
                          EXP(LOG_CL_NTPRO_BL) as ntbnp_bl,
                          LOG_CL_NTPRO_POST as log_ntbnp_dc,
                          EXP(LOG_CL_NTPRO_POST) as ntbnp_dc,
                          LOG_CL_NTPRO_BLPOST as ratio_ntbnp_bl_dc
                          from athena_endpts')

acute_athena_outcomes <- merge(acute_athena_outcomes,
                               acute_athena_exam[,c("patientid",
                                                    "hr_bl",
                                                    "hr_2d",
                                                    "hr_4d",
                                                    "hr_dc",
                                                    "hr_last",
                                                    "dyspnea_vas_2d",
                                                    "dyspnea_vas_4d",
                                                    "dyspnea_vas_last")],
                               by="patientid",
                               all.x=T)

acute_athena_outcomes <- merge(acute_athena_outcomes,
                               acute_athena_visitsumm[,c("patientid",
                                                         "wt_kg_1d",
                                                         "wt_kg_2d",
                                                         "wt_kg_3d",
                                                         "wt_kg_4d",
                                                         "wt_kg_last",
                                                         "ntbnp_1d",
                                                         "ntbnp_2d",
                                                         "ntbnp_3d",
                                                         "ntbnp_4d")],
                               by="patientid",
                               all.x=T)


acute_athena_outcomes <- merge(acute_athena_outcomes,
                           acute_athena_labs[,c("patientid",
                                                "bun_bl",
                                                "bun_1d",
                                                "bun_2d",
                                                "bun_3d",
                                                "bun_4d",
                                                "bun_dc",
                                                "bnp_bl",
                                                "bnp_dc")],
                           by="patientid",
                           all.x=T)

acute_athena_outcomes <- merge(acute_athena_outcomes,
                               acute_athena_renal[,c("patientid",
                                                     "gfr_bl_1d",
                                                     "gfr_bl_1d_pct",
                                                     "gfr_bl_2d",
                                                     "gfr_bl_2d_pct",
                                                     "gfr_bl_3d",
                                                     "gfr_bl_3d_pct",
                                                     "gfr_bl_4d",
                                                     "gfr_bl_4d_pct",
                                                     "gfr_bl_last_pct")],
                               by="patientid",
                               all.x=T)


acute_athena_outcomes$study <- "ATHENA"

missing_acute_outcomes_athena <- acute_analysis_outcomes[!acute_analysis_outcomes %in% names(acute_athena_outcomes)]
acute_athena_outcomes[,missing_acute_outcomes_athena] <- NA
acute_analysis_outcomes_athena <- acute_athena_outcomes[,acute_analysis_outcomes]

rm(list=ls(pattern="\\bathena_."))


########################################### **************** COMBINE OUTCOMES ***************** #############################################
#
# ..%%%%%%...%%%%%%%..%%.....%%.%%%%%%%%..%%%%.%%....%%.%%%%%%%%.....%%%%%%%..%%.....%%.%%%%%%%%..%%%%%%...%%%%%%%..%%.....%%.%%%%%%%%..%%%%%%.
# .%%....%%.%%.....%%.%%%...%%%.%%.....%%..%%..%%%...%%.%%..........%%.....%%.%%.....%%....%%....%%....%%.%%.....%%.%%%...%%%.%%.......%%....%%
# .%%.......%%.....%%.%%%%.%%%%.%%.....%%..%%..%%%%..%%.%%..........%%.....%%.%%.....%%....%%....%%.......%%.....%%.%%%%.%%%%.%%.......%%......
# .%%.......%%.....%%.%%.%%%.%%.%%%%%%%%...%%..%%.%%.%%.%%%%%%......%%.....%%.%%.....%%....%%....%%.......%%.....%%.%%.%%%.%%.%%%%%%....%%%%%%.
# .%%.......%%.....%%.%%.....%%.%%.....%%..%%..%%..%%%%.%%..........%%.....%%.%%.....%%....%%....%%.......%%.....%%.%%.....%%.%%.............%%
# .%%....%%.%%.....%%.%%.....%%.%%.....%%..%%..%%...%%%.%%..........%%.....%%.%%.....%%....%%....%%....%%.%%.....%%.%%.....%%.%%.......%%....%%
# ..%%%%%%...%%%%%%%..%%.....%%.%%%%%%%%..%%%%.%%....%%.%%%%%%%%.....%%%%%%%...%%%%%%%.....%%.....%%%%%%...%%%%%%%..%%.....%%.%%%%%%%%..%%%%%%.


### Acute outcomes

hdcp_acute_outcomes <- rbind(acute_analysis_outcomes_carress,
                             acute_analysis_outcomes_dose,
                             acute_analysis_outcomes_rose,
                             acute_analysis_outcomes_escape,
                             acute_analysis_outcomes_athena)

hdcp_acute_outcomes$wt_kg_bl_last <- hdcp_acute_outcomes$wt_kg_last - hdcp_acute_outcomes$wt_kg_bl
hdcp_acute_outcomes$wt_kg_bl_last_pct <- hdcp_acute_outcomes$wt_kg_bl_last/hdcp_acute_outcomes$wt_kg_bl*100


write.csv(hdcp_acute_outcomes,'~/Dropbox/ADAPT-HF/Master HDCP files/hdcp_acute_outcomes.csv',row.names=F)
write.csv(acute_escape_outcomes,'~/Dropbox/ADAPT-HF/Master HDCP files/acute_escape_outcomes.csv',row.names=F)
write.csv(acute_carress_outcomes,'~/Dropbox/ADAPT-HF/Master HDCP files/acute_carress_outcomes.csv',row.names=F)
write.csv(acute_dose_outcomes,'~/Dropbox/ADAPT-HF/Master HDCP files/acute_dose_outcomes.csv',row.names=F)
write.csv(acute_rose_outcomes,'~/Dropbox/ADAPT-HF/Master HDCP files/acute_rose_outcomes.csv',row.names=F)
write.csv(acute_athena_outcomes,'~/Dropbox/ADAPT-HF/Master HDCP files/acute_athena_outcomes.csv',row.names=F)


gcs_auth(email="")
gcs_upload(hdcp_acute_outcomes[,acute_analysis_outcomes], 
           bucket="master_hdcp_files",
           name="trial_acute_outcomes_all.csv",
           object_function = f)

gcs_upload(acute_escape_outcomes, 
           bucket="master_hdcp_files",
           name="trial_acute_outcomes_escape.csv",
           object_function = f)

gcs_upload(acute_carress_outcomes, 
           bucket="master_hdcp_files",
           name="trial_acute_outcomes_carress.csv",
           object_function = f)

gcs_upload(acute_dose_outcomes, 
           bucket="master_hdcp_files",
           name="trial_acute_outcomes_dose.csv",
           object_function = f)

gcs_upload(acute_rose_outcomes, 
           bucket="master_hdcp_files",
           name="trial_acute_outcomes_rose.csv",
           object_function = f)

gcs_upload(acute_athena_outcomes, 
           bucket="master_hdcp_files",
           name="trial_acute_outcomes_athena.csv",
           object_function = f)
```
