## Supplementary material for "Aggregating data to accelerate personalized therapy in heart failure (ADAPT-HF)": Harmonization scripts: Transform trial characteristics.R

```
######### ************README************* #########

# This is version 4.0 of a script for loading clinical trial data. Major updates from the previous versions are:
# - Intended for HDCP analysis completely within R
# - Provides consistency with observational study data code (FHS, CHS, ARIC, MESA, JHS), which are much larger
# - Moves calculated/derived values from SQL code to within this file load and transformation code.
#     - Because I have not had opportunity to make a generalizable formula implementation framework, these 
#     derived values were hardcoded into SQL anyway - they are now included here instead.
# - Now creates dataset including all major timepoints for each study (i.e. not ad hoc encounters)
#     - This is done to enable both phenotyping and longitudinal analysis from this single dataset, which was
#     which was the original intent for this platform.  This allows explicit harmonization mapping at
#     all timepoints
# - Now includes all patient reported outcomes data (KCCQ, MLHG, VAS, EQ5D, HRQOL) in single unified codebank
# - Includes all randomized clinical trial data available as of 10.24.2024.  This includes the following studies:
# - Loads denormalized raw and harmonized tables into Google BigQuery (schema `harmonization-286013.trials`).
# - Will also write to DB for now given legacy analyses. 
# 
# ATHENA (BioLINCC, full)
# BEST (MRB, full)
# CARRESS (BioLINCC, full)
# CORONA (Glasgow, *sample only - for testing only*)
# DIG (BioLINCC, full)
# DOSE (BioLINCC, full)
# ESCAPE (BioLINCC, full)
# EXACT (BioLINCC, full)
# FIGHT (BioLINCC, full)
# GUIDE-IT (BioLINCC, full)
# HF-ACTION (BioLINCC, full)
# INDIE (BioLINCC, full)
# IPRESERVE (Glasgow, full)
# IRONOUT (BioLINCC, full)
# LIFE (BioLINCC, full)
# MOCHA (MRB, full)
# NEAT-HFpEF (BioLINCC, full)
# PARADIGM (Glasgow, *sample only - for testing only*)
# RELAX (BioLINCC, full)
# ROSE (BioLINCC, full)
# SCD-HEFT (BioLINCC, full)
# SOLVD Prevent (BioLINCC, full)
# SOLVD Treat (BioLINCC, full)
# SOLVD Registry (BioLINCC, full)
# STICH + STICHES (BioLINCC, full)
# TOPCAT (BioLINCC, full)


# This will produce a denormalized data set in the fashion of the previous "alldata"  This will continue
# to be the foundation of other phenotyping method code sets but primarily LCA

#########  ***********Required Libraries*********** ##########

zpz <- Sys.time()

if (!require("pacman")) install.packages("pacman")

pacman::p_load("foreign",
               "plyr",
               "gmodels",
               "rmeta",
               "poLCA",
               "descr",
               "reshape2",
               "Hmisc",
               "readstata13",
               "sas7bdat")

if (!require("relper")) remotes::install_github("vbfelix/relper")

library(plyr)
library(readstata13)
library(Hmisc)
library(reshape2)
library(descr)
library(sas7bdat)
library(sqldf)
library(DescTools)
library(dplyr)
library(stringr)
library(relper)
library(googleCloudStorageR)
library(haven)
source("~/Dropbox/R scripts/echo scripts.R")
source("~/Dropbox/R scripts/Misc clinical scripts.R")


## Study names made using textkool.com/en/ascii-art-generator, font = Banner4

# Use this code to make subsection names:
  # library(bannerCommenter)
  # print(block("ATHENA: a_visitsumm",
  # leftSideHashes = 4,
  # rightSideHashes=4,
  # bandChar="#",snug=T))


athena_folder <- "~/Dropbox/BioLINCC files/ATHENA/data/csv/"
best_folder <- "~/Dropbox/BioLINCC files/BEST/BEST CSVs/"
carress_folder <- "~/Dropbox/BioLINCC files/CARRESS/CARRESS_2017a/main_study/data/csv/"
corona_folder <- "~/Dropbox/BioLINCC files/CORONA/"
dig_folder <- "~/Dropbox/BioLINCC files/DIG/DIG_2015a/ASCII data/"
dose_folder <- "~/Dropbox/BioLINCC files/DOSE/data/"
escape_folder <- "~/Dropbox/BioLINCC files/ESCAPE/data/" 
exact_folder <- "~/Dropbox/BioLINCC files/EXACT-HF/EXACT_2017a/data/csv/"
fight_folder <- "~/Dropbox/BioLINCC files/FIGHT/data/csv/"
guideit_folder <- "~/Dropbox/BioLINCC files/GUIDE-IT/GUIDE_IT_2020a/data/csv/"
hfaction_folder <- "~/Dropbox/BioLINCC files/HF-ACTION/Data_DR/"
indie_folder <- "~/Dropbox/BioLINCC files/INDIE-HFpEF/data/CSV/"
ipreserve_folder <- "~/Dropbox/BioLINCC files/IPRESERVE/"
ironout_folder <- "~/Dropbox/BioLINCC files/IRONOUT/data/CSV/"
life_folder <- "~/Dropbox/BioLINCC files/LIFE/data/CSV/"
mocha_folder <- "~/Dropbox/BioLINCC files/MOCHA/CSVs/"
neat_folder <- "~/Dropbox/BioLINCC files/NEAT-HFpEF/datasets/csv/"
paradigm_folder <- "~/Dropbox/BioLINCC files/PARADIGM/"
relax_folder <- "~/Dropbox/BioLINCC files/RELAX/hfn_relax/data/"
rose_folder <- "~/Dropbox/BioLINCC files/ROSE/ROSE_2016a/Data/"
scdheft_folder <- "~/Dropbox/BioLINCC files/SCD-HeFT/data/"
solvd_folder <- "~/Dropbox/BioLINCC files/SOLVD/data/Datasets/"
solvdreg_folder <- "~/Dropbox/BioLINCC files/SOLVD/data/Registry/Datasets/"
stich_folder <- "~/Dropbox/BioLINCC files/STICH/STICH_2018b/STICH (main study)/data/csv/"
stiches_folder <- "~/Dropbox/BioLINCC files/STICH/STICH_2018b/STICHES (ancillary)/data/csv/"
topcat_folder <- "~/Dropbox/BioLINCC files/TOPCAT/data/sas_data/"


###############  **********HF-ACTION**********  ###############


# .%%.....%%.%%%%%%%%............%%%.....%%%%%%..%%%%%%%%.%%%%..%%%%%%%..%%....%%
# .%%.....%%.%%.................%%.%%...%%....%%....%%.....%%..%%.....%%.%%%...%%
# .%%.....%%.%%................%%...%%..%%..........%%.....%%..%%.....%%.%%%%..%%
# .%%%%%%%%%.%%%%%%...%%%%%%%.%%.....%%.%%..........%%.....%%..%%.....%%.%%.%%.%%
# .%%.....%%.%%...............%%%%%%%%%.%%..........%%.....%%..%%.....%%.%%..%%%%
# .%%.....%%.%%...............%%.....%%.%%....%%....%%.....%%..%%.....%%.%%...%%%
# .%%.....%%.%%...............%%.....%%..%%%%%%.....%%....%%%%..%%%%%%%..%%....%%


##########  baseline  ##########


hfaction_analysis <- read_sas(paste(hfaction_folder,"analysis.sas7bdat",sep=""))
hfaction_analysis[hfaction_analysis=="NaN"] <- NA

hfaction_analysis$bsa <- 0.20247*(hfaction_analysis$weightkg^0.425)*(hfaction_analysis$heightcm/100)^0.725
hfaction_analysis$pupr <- hfaction_analysis$sbp-hfaction_analysis$dbp

hfaction_analysis$lvmass_ix <- hfaction_analysis$lftvmass/hfaction_analysis$bsa
hfaction_analysis$LVEDD_index <- hfaction_analysis$lftvdiad/hfaction_analysis$bsa
hfaction_analysis$eeprime_lat <- hfaction_analysis$mvpkevel/hfaction_analysis$ewavlat
hfaction_analysis$eeprime_sept <- hfaction_analysis$mvpkevel/hfaction_analysis$ewavsept
hfaction_analysis$eeprime_avg <- apply(hfaction_analysis[,c("eeprime_lat","eeprime_sept")], MARGIN=1, FUN=mean)

hfaction_analysis_melt <- melt(hfaction_analysis,id.vars=c("newid"),na.rm=T) 
hfaction_analysis_melt$form <- "analysis"
hfaction_analysis_melt$studyvisit <- "BASELINE"


##########  cpx  ##########

### Need to watch the logic on cpx if you want to extract other CPX parameters, since protocol stated use the first baseline VO2 rather than second (n=385)

hfaction_cpx <- read_sas(paste(hfaction_folder,"cpx.sas7bdat",sep=""))

hfaction_cpx <- subset(hfaction_cpx,CPXVIS >= 2)

hfaction_cpx_melt <- 
  melt(data=hfaction_cpx, id.vars=c("newid","CPXVIS"),na.rm=T) 

hfaction_cpx_melt$form <- "cpx"

colnames(hfaction_cpx_melt)[colnames(hfaction_cpx_melt)=="CPXVIS"] <- "studyvisit"


#### kccq ####


hfaction_kccq1 <- read_sas(paste(hfaction_folder,"kccq1.sas7bdat",sep=""))
hfaction_kccq2 <- read_sas(paste(hfaction_folder,"kccq2.sas7bdat",sep=""))
hfaction_kccq3 <- read_sas(paste(hfaction_folder,"kccq3.sas7bdat",sep=""))

hfaction_kccq <- sqldf('select
                       newid,
                       FORM as studyvisit, 
                       KCCQ1A, 
                       KCCQ1B, 
                       KCCQ1C, 
                       KCCQ1D,
                       KCCQ1E,
                       KCCQ1F,
                       KCCQ2,
                       KCCQ3,
                       KCCQ4,
                       KCCQ5,
                       KCCQ6,
                       KCCQ7,
                       KCCQ8,
                       KCCQ9,
                       KCCQ10,
                       KCCQ11,
                       KCCQ12,
                       KCCQ13,
                       KCCQ14,
                       KCCQ15A,
                       KCCQ15B,
                       KCCQ15C,
                       KCCQ15D
                       from hfaction_kccq1 left join hfaction_kccq2 using (newid,FORM)
                       join hfaction_kccq3 using (newid,FORM)')

hfaction_kccq <- score_kccq(dat=hfaction_kccq,
                            q1a="KCCQ1A",
                            q1b="KCCQ1B",
                            q1c="KCCQ1C",
                            q1d="KCCQ1D",
                            q1e="KCCQ1E",
                            q1f="KCCQ1F",
                            q2="KCCQ2",
                            q3="KCCQ3",
                            q4="KCCQ4",
                            q5="KCCQ5",
                            q6="KCCQ6",
                            q7="KCCQ7",
                            q8="KCCQ8",
                            q9="KCCQ9",
                            q10="KCCQ10",
                            q11="KCCQ11",
                            q12="KCCQ12",
                            q13="KCCQ13",
                            q14="KCCQ14",
                            q15a="KCCQ15A",
                            q15b="KCCQ15B",
                            q15c="KCCQ15C",
                            q15d="KCCQ15D")

hfaction_kccq_melt <- melt(data=hfaction_kccq,
                          id.vars=c("newid","studyvisit"),
                          na.rm=T)

hfaction_kccq_melt$form <- "kccq"


##########  survey  ##########

hfaction_survey <- read_sas(paste(hfaction_folder,"survey.sas7bdat",sep=""))

hfaction_survey_melt <- 
  melt(data=hfaction_survey, id.vars=c("newid","FORM"),na.rm=T) 

hfaction_survey_melt$form <- "survey"

colnames(hfaction_survey_melt)[colnames(hfaction_survey_melt)=="FORM"] <- "studyvisit"


##########  euro  ##########

hfaction_euro <- read_sas(paste(hfaction_folder,"euro.sas7bdat",sep=""))

hfaction_euro_melt <- 
  melt(data=hfaction_euro, id.vars=c("newid","FORM"),na.rm=T) 

hfaction_euro_melt$form <- "euro"

colnames(hfaction_euro_melt)[colnames(hfaction_euro_melt)=="FORM"] <- "studyvisit"


##########  walkt  ##########

hfaction_walkt <- read_sas(paste(hfaction_folder,"walkt.sas7bdat",sep=""))

hfaction_walkt$sixmw_meters[hfaction_walkt$DISTMU==2&!is.na(hfaction_walkt$DISTMU)] <-
  hfaction_walkt$DISTM[hfaction_walkt$DISTMU==2&!is.na(hfaction_walkt$DISTMU)]

hfaction_walkt$sixmw_meters[hfaction_walkt$DISTMU==1&!is.na(hfaction_walkt$DISTMU)] <-
  hfaction_walkt$DISTM[hfaction_walkt$DISTMU==1&!is.na(hfaction_walkt$DISTMU)]*0.3048


hfaction_walkt_melt <- 
  melt(data=hfaction_walkt, id.vars=c("newid","FORM"),na.rm=T) 

hfaction_walkt_melt$form <- "walkt"

colnames(hfaction_walkt_melt)[colnames(hfaction_walkt_melt)=="FORM"] <- "studyvisit"


##########  bdi  ##########

hfaction_bdi1 <- read_sas(paste(hfaction_folder,"bdi1.sas7bdat",sep=""))
hfaction_bdi2 <- read_sas(paste(hfaction_folder,"bdi2.sas7bdat",sep=""))

hfaction_bdi1 <- subset(hfaction_bdi1, select = -c(PAGEID,NODATA,com_use,noncom_use,GENDER,visitdays))
hfaction_bdi2 <- subset(hfaction_bdi2, select = -c(PAGEID,NODATA,com_use,noncom_use,GENDER,visitdays))

hfaction_bdi <- merge(hfaction_bdi1,hfaction_bdi2,by=c("newid","FORM"),all=T)
hfaction_bdi[hfaction_bdi=="NaN"] <- NA

colnames(hfaction_bdi)[colnames(hfaction_bdi)=="FORM"] <- "studyvisit"


hfaction_bdi$bdi_total <- apply(X=hfaction_bdi[3:23],
                                MARGIN=1,
                                FUN=function(x) sum(x))

hfaction_bdi_melt <- melt(data=hfaction_bdi,
                          id.vars=c("newid","studyvisit"),
                          na.rm=T)

hfaction_bdi_melt$form <- "bdi"

##########  labs  ##########

### Load labs using labtest as data_field

hfaction_labs <- read_sas(paste(hfaction_folder,"labs.sas7bdat",sep=""))

hfaction_labs$hct_derive[hfaction_labs$HCT > 1] <- hfaction_labs$data_string
hfaction_labs$hct_derive[hfaction_labs$HCT > 1] <- hfaction_labs$data_string*100


hfaction_labs$studyvisit <- hfaction_labs$FORM

hfaction_labs$form <- "labs"
hfaction_labs$variable <- hfaction_labs$LABTEST
hfaction_labs$value <- hfaction_labs$LABVAL

hfaction_labs$variable <- as.character(hfaction_labs$variable)


##########  Furosemide equivalent  ##########

hfaction_furo_equiv <- read_sas(paste(hfaction_folder,"cmed.sas7bdat",sep=""))

hfaction_furo_equiv$form <- "furo_equiv"
hfaction_furo_equiv$studyvisit <- hfaction_furo_equiv$FORM

hfaction_furo_equiv$FURODOSE[hfaction_furo_equiv$YESLOOP==1&
                               !is.na(hfaction_furo_equiv$YESLOOP)] <- 
  hfaction_furo_equiv$LOOPVAL[hfaction_furo_equiv$YESLOOP==1&
                                !is.na(hfaction_furo_equiv$YESLOOP)]

hfaction_furo_equiv$BUMDOSE[hfaction_furo_equiv$YESLOOP==2&
                              !is.na(hfaction_furo_equiv$YESLOOP)] <- 
  hfaction_furo_equiv$LOOPVAL[hfaction_furo_equiv$YESLOOP==2&
                                !is.na(hfaction_furo_equiv$YESLOOP)] * 40

hfaction_furo_equiv$TORSDOSE[hfaction_furo_equiv$YESLOOP==3&
                               !is.na(hfaction_furo_equiv$YESLOOP)] <- 
  hfaction_furo_equiv$LOOPVAL[hfaction_furo_equiv$YESLOOP==3&
                                !is.na(hfaction_furo_equiv$YESLOOP)] * 2


hfaction_furo_equiv$value <- apply(hfaction_furo_equiv
                        [,c("FURODOSE",
                            "TORSDOSE",
                            "BUMDOSE")],
                        MARGIN=1,
                        FUN=diur_calc)

hfaction_furo_equiv$variable <- "daily_furo_eq_derive"

hfaction_furo_equiv$form <- "daily_furo"


##########  cmed  ##########


hfaction_cmed <- read_sas(paste(hfaction_folder,"cmed.sas7bdat",sep=""))

hfaction_cmed_melt <- 
  melt(data=hfaction_cmed, id.vars=c("newid","FORM"),na.rm=T) 

hfaction_cmed_melt$form <- "cmed"

colnames(hfaction_cmed_melt)[colnames(hfaction_cmed_melt)=="FORM"] <- "studyvisit"


##########  cmeds  ##########

### Load cmeds using curmeds as a data field

hfaction_cmeds <- read_sas(paste(hfaction_folder,"cmeds.sas7bdat",sep=""))

hfaction_cmeds$form <- "cmeds"
hfaction_cmeds$studyvisit <- hfaction_cmeds$FORM

colnames(hfaction_cmeds)[c(4,5)] <- c("variable","value")
hfaction_cmeds$variable <- as.character(hfaction_cmeds$variable)


##########  compile  ##########

hfaction_melt <- rbind(hfaction_analysis_melt[,c("newid","form","studyvisit","variable","value")],
                       hfaction_bdi_melt[,c("newid","form","studyvisit","variable","value")],
                       hfaction_kccq_melt[,c("newid","form","studyvisit","variable","value")],
                       hfaction_euro_melt[,c("newid","form","studyvisit","variable","value")],
                       hfaction_cpx_melt[,c("newid","form","studyvisit","variable","value")],
                       hfaction_survey_melt[,c("newid","form","studyvisit","variable","value")],
                       hfaction_walkt_melt[,c("newid","form","studyvisit","variable","value")],
                       hfaction_furo_equiv[,c("newid","form","studyvisit","variable","value")],
                       hfaction_labs[,c("newid","form","studyvisit","variable","value")],
                       hfaction_cmed_melt[,c("newid","form","studyvisit","variable","value")],
                       hfaction_cmeds[,c("newid","form","studyvisit","variable","value")])
                       
hfaction_melt <- subset(hfaction_melt,!value %in% c(NA,"NaN",""))

hfaction_melt <- subset(hfaction_melt,!is.na(value)&!value=="NaN"&!value=="")
colnames(hfaction_melt)[c(1)] <- c("patientid") 
hfaction_melt$study <- "HF-ACTION"


##############################  **********BEST**********  ##############################


# .%%%%%%%%..%%%%%%%%..%%%%%%..%%%%%%%%
# .%%.....%%.%%.......%%....%%....%%...
# .%%.....%%.%%.......%%..........%%...
# .%%%%%%%%..%%%%%%....%%%%%%.....%%...
# .%%.....%%.%%.............%%....%%...
# .%%.....%%.%%.......%%....%%....%%...
# .%%%%%%%%..%%%%%%%%..%%%%%%.....%%...


#### pcsf ####

best_pcsf <- read.csv(paste(best_folder,"pcsf.csv",sep=""),na.strings=c("",NA,"NULL"))
best_pcsf_melt <- melt(data=best_pcsf,id.vars=c("ID","VISIT"),na.rm=T)
best_pcsf_melt$form <- "pcsf"
best_pcsf_melt$VISIT <- 1

##########  t (treatment arm)  ##########

best_t_melt <- read.csv(paste(best_folder,"t.csv",sep=""),head=T)
best_t_melt$VISIT <- "1"
best_t_melt$variable <- "group"
best_t_melt$value <- best_t_melt$GROUP
best_t_melt$form <- "t"


##########  lab1  ##########

best_lab1 <- read.csv(paste(best_folder,"lab1",".csv",sep=""),na.strings=c("",NA,"NULL"))
best_clab1 <- read.csv(paste(best_folder,"clab1",".csv",sep=""),na.strings=c("",NA,"NULL"))

best_lab1 <- merge(best_lab1,best_pcsf[,c("ID","PCAGE","PCSEX","PCRACE")],by="ID")
best_clab1 <- merge(best_clab1,best_pcsf[,c("ID","PCAGE","PCSEX","PCRACE")],by="ID")

names(best_lab1)[35:37] <- c("lab_age","lab_sex","lab_race")
names(best_clab1)[42:44] <- c("lab_age","lab_sex","lab_race")

best_lab1$CrCl <- calc_MDRD4(dat=best_lab1, cr="LACRE", age="lab_age",sex="lab_sex",race="lab_race",male=1,black=2)
best_clab1$CrCl <- calc_MDRD4(dat=best_clab1, cr="CLACREX", age="lab_age",sex="lab_sex",race="lab_race",male=1,black=2)

best_lab1_melt <- melt(data=best_lab1,id.vars=c("ID","VISIT"),na.rm=T)
best_lab1_melt$form <- "lab1"

best_clab1_melt <- melt(data=best_clab1,id.vars=c("ID","VISIT"),na.rm=T)
best_clab1_melt$form <- "clab1"


##########  lab2  ##########

best_lab2 <- read.csv(paste(best_folder,"lab2.csv",sep=""),head=T)
best_lab2_melt <- melt(data=best_lab2,id.vars=c("ID","VISIT"),na.rm=T)
best_lab2_melt$form <- "lab2"

best_clab2 <- read.csv(paste(best_folder,"clab2.csv",sep=""),head=T)
best_clab2_melt <- melt(data=best_clab2,id.vars=c("ID","VISIT"),na.rm=T)
best_clab2_melt$form <- "clab2"


##########  cvh1  ##########

best_cvh1 <- read.csv(paste(best_folder,"cvh1.csv",sep=""),head=T)
best_cvh1_melt <- melt(data=best_cvh1,id.vars=c("ID","VISIT"),na.rm=T)
best_cvh1_melt$form <- "cvh1"


##########  cvh2  ##########

best_cvh2 <- read.csv(paste(best_folder,"cvh2.csv",sep=""),head=T)

best_cvh2[best_cvh2==2] <- 0


best_cvh2$num_etiol <- apply(X=best_cvh2[12:20],
                             MARGIN=1,
                             function (x)
                               sum(x,na.rm=T))

best_cvh2[best_cvh2$num_etiol>1,c("CVCAD",
                                  "CVMITRAL",
                                  "CVAORTIC",
                                  "CVETOH",
                                  "CVDRUG",
                                  "CVHYPRIN",
                                  "CVFAMIL",
                                  "CVVIRAL",
                                  "CVIDIOP",
                                  "CVOTHER")]<- 0

best_cvh2_melt <- melt(data=best_cvh2,id.vars=c("ID","VISIT"),na.rm=T)
best_cvh2_melt$form <- "cvh2"


##########  ecg  ##########

best_ecg <- read.csv(paste(best_folder,"ecg.csv",sep=""),head=T)
best_ecg_melt <- melt(data=best_ecg,id.vars=c("ID","VISIT"),na.rm=T)
best_ecg_melt$form <- "ecg"


##########  diab  ##########

best_diab <- read.csv(paste(best_folder,"diab.csv",sep=""),head=T)
best_diab_melt <- melt(data=best_diab,id.vars=c("ID","VISIT"),na.rm=T)
best_diab_melt$form <- "diab"


##########  pe  ##########

best_pe <- read.csv(paste(best_folder,"pe.csv",sep=""),head=T)
best_ht <- best_pe[best_pe$VISIT==1&!is.na(best_pe$PEHT),c("ID","PEHT")]
best_pe <- merge(best_pe,best_ht,by="ID", all.x=T)
best_pe$ht_bl <- best_pe$PEHT.y

best_pe$weight_kg <- round(best_pe$PEWT/2.2,1)

best_pe <- best_pe[,!names(best_pe) %in% c("PEHT.x","PEHT.y")]


# Calculate BMI

best_pe$bmi <- round((best_pe$PEWT*703)/(best_pe$ht_bl^2),1)


# Calculate PE

best_pe$pupr <- best_pe$PEBPS - best_pe$PEBPD

best_pe_melt <- melt(data=best_pe,id.vars=c("ID","VISIT"),na.rm=T)
best_pe_melt$form <- "pe"


##########  muga  ##########

best_muga <- read.csv(paste(best_folder,"muga.csv",sep=""),head=T)
best_muga_melt <- melt(data=best_muga,id.vars=c("ID","VISIT"),na.rm=T)
best_muga_melt$form <- "muga"


##########  cvs  ##########

best_cvs <- read.csv(paste(best_folder,"cvs.csv",sep=""),head=T)
best_cvs_melt <- melt(data=best_cvs,id.vars=c("ID","VISIT"),na.rm=T)
best_cvs_melt$form <- "cvs"


######### qol  ##########


best_qol2 <- read.csv(paste(best_folder,"qol2.csv",sep=""),header=T,na.strings="")
best_qol3 <- read.csv(paste(best_folder,"qol3.csv",sep=""),header=T,na.strings="")

best_mlhf <- sqldf('select ID,
              VISIT,
              QL_1-1 as MLHFQ1, 
              QL_4-1 as MLHFQ2,
              QL_7-1 as MLHFQ3,
              QL_2-1 as MLHFQ4, 
              QL_11-1 as MLHFQ5,
              QL_9-1 as MLHFQ6,
              QL_3-1 as MLHFQ7,
              QL_6-1 as MLHFQ8,
              QL_13-1 as MLHFQ9,
              QL_12-1 as MLHFQ10,
              QL_10-1 as MLHFQ11,
              QL_8-1 as MLHFQ12,
              QL_5-1 as MLHFQ13,
              QL_20-1 as MLHFQ14,
              QL_18-1 as MLHFQ15,
              QL_15-1 as MLHFQ16,
              QL_21-1 as MLHFQ17,
              QL_19-1 as MLHFQ18,
              QL_16-1 as MLHFQ19,
              QL_14-1 as MLHFQ20,
              QL_17-1 as MLHFQ21
              from best_qol2 join best_qol3 using (ID,VISIT)
              ')

best_mlhf <- score_mlhf(dat=best_mlhf,
                        swelling = "MLHFQ1", 
                        day_rest="MLHFQ2",
                        stairs = "MLHFQ3", 
                        housework="MLHFQ4",
                        go_places="MLHFQ5",
                        sleep = "MLHFQ6",
                        friendfam = "MLHFQ7",
                        earn_living = "MLHFQ8",
                        hobbies = "MLHFQ9",
                        sex = "MLHFQ10",
                        food = "MLHFQ11",
                        sob = "MLHFQ12",
                        fatigue = "MLHFQ13",
                        hospital = "MLHFQ14",
                        cost = "MLHFQ15",
                        sideeffect = "MLHFQ16",
                        burden = "MLHFQ17",
                        nocontrol = "MLHFQ18",
                        worry="MLHFQ19",
                        concentrate="MLHFQ20",
                        depressed="MLHFQ21", 
                        total=c("Y"))

best_mlhf$VISIT[best_mlhf$VISIT==2] <- 1

best_mlhf_melt <- melt(data=best_mlhf, id.vars=c("ID","VISIT"), na.rm=T)
best_mlhf_melt$form <- "qol"


##########  sct (meds)  ##########

best_sct1 <- read.csv(paste(best_folder,"sct1.csv",sep=""),stringsAsFactors = F,na.strings=c("NA","","NULL"))

best_sct1_med1 <- best_sct1[,c('ID','VISIT','SCDRUG1','SCCODE1','SCDOSE1','SCUNIT1','SCCAT1','SCROUT1')]
best_sct1_med2 <- best_sct1[,c('ID','VISIT','SCDRUG2','SCCODE2','SCDOSE2','SCUNIT2','SCCAT2','SCROUT2')]
best_sct1_med3 <- best_sct1[,c('ID','VISIT','SCDRUG3','SCCODE3','SCDOSE3','SCUNIT3','SCCAT3','SCROUT3')]
best_sct1_med4 <- best_sct1[,c('ID','VISIT','SCDRUG4','SCCODE4','SCDOSE4','SCUNIT4','SCCAT4','SCROUT4')]
best_sct1_med5 <- best_sct1[,c('ID','VISIT','SCDRUG5','SCCODE5','SCDOSE5','SCUNIT5','SCCAT5','SCROUT5')]
best_sct1_med6 <- best_sct1[,c('ID','VISIT','SCDRUG6','SCCODE6','SCDOSE6','SCUNIT6','SCCAT6','SCROUT6')]
best_sct1_med7 <- best_sct1[,c('ID','VISIT','SCDRUG7','SCCODE7','SCDOSE7','SCUNIT7','SCCAT7','SCROUT7')]
best_sct1_med8 <- best_sct1[,c('ID','VISIT','SCDRUG8','SCCODE8','SCDOSE8','SCUNIT8','SCCAT8','SCROUT8')]
best_sct1_med9 <- best_sct1[,c('ID','VISIT','SCDRUG9','SCCODE9','SCDOSE9','SCUNIT9','SCCAT9','SCROUT9')]
best_sct1_med10 <- best_sct1[,c('ID','VISIT','SCDRUG10','SCCODE10','SCDOSE10','SCUNIT10','SCCAT10','SCROUT10')]
best_sct1_med11 <- best_sct1[,c('ID','VISIT','SCDRUG11','SCCODE11','SCDOSE11','SCUNIT11','SCCAT11','SCROUT11')]
best_sct1_med12 <- best_sct1[,c('ID','VISIT','SCDRUG12','SCCODE12','SCDOSE12','SCUNIT12','SCCAT12','SCROUT12')]
best_sct1_med13 <- best_sct1[,c('ID','VISIT','SCDRUG13','SCCODE13','SCDOSE13','SCUNIT13','SCCAT13','SCROUT13')]
best_sct1_med14 <- best_sct1[,c('ID','VISIT','SCDRUG14','SCCODE14','SCDOSE14','SCUNIT14','SCCAT14','SCROUT14')]

names(best_sct1_med1) <- c('ID','VISIT','med_name','med_code','med_dose','med_units','med_cat','med_route')
names(best_sct1_med2) <- c('ID','VISIT','med_name','med_code','med_dose','med_units','med_cat','med_route')
names(best_sct1_med3) <- c('ID','VISIT','med_name','med_code','med_dose','med_units','med_cat','med_route')
names(best_sct1_med4) <- c('ID','VISIT','med_name','med_code','med_dose','med_units','med_cat','med_route')
names(best_sct1_med5) <- c('ID','VISIT','med_name','med_code','med_dose','med_units','med_cat','med_route')
names(best_sct1_med6) <- c('ID','VISIT','med_name','med_code','med_dose','med_units','med_cat','med_route')
names(best_sct1_med7) <- c('ID','VISIT','med_name','med_code','med_dose','med_units','med_cat','med_route')
names(best_sct1_med8) <- c('ID','VISIT','med_name','med_code','med_dose','med_units','med_cat','med_route')
names(best_sct1_med9) <- c('ID','VISIT','med_name','med_code','med_dose','med_units','med_cat','med_route')
names(best_sct1_med10) <- c('ID','VISIT','med_name','med_code','med_dose','med_units','med_cat','med_route')
names(best_sct1_med11) <- c('ID','VISIT','med_name','med_code','med_dose','med_units','med_cat','med_route')
names(best_sct1_med12) <- c('ID','VISIT','med_name','med_code','med_dose','med_units','med_cat','med_route')
names(best_sct1_med13) <- c('ID','VISIT','med_name','med_code','med_dose','med_units','med_cat','med_route')
names(best_sct1_med14) <- c('ID','VISIT','med_name','med_code','med_dose','med_units','med_cat','med_route')

best_sct1_op_melt <- rbind(best_sct1_med1,
                           best_sct1_med2,
                           best_sct1_med3,
                           best_sct1_med4,
                           best_sct1_med5,
                           best_sct1_med6,
                           best_sct1_med7,
                           best_sct1_med8,
                           best_sct1_med9,
                           best_sct1_med10,
                           best_sct1_med11,
                           best_sct1_med12,
                           best_sct1_med13,
                           best_sct1_med14)


best_sct1_op_melt$med_units_derive[best_sct1_op_melt$med_units==1] <- "mg"

best_sct1_op_melt$form <- "sct1"
best_sct1_op_melt$variable <- "med_code"
best_sct1_op_melt$value <- best_sct1_op_melt$med_code


best_sct1_op_melt <- subset(best_sct1_op_melt,!is.na(med_code))


best_sct1_op_furo <- sqldf("select ID, VISIT,med_dose as FURODOSE from best_sct1_op_melt where med_code=='JHG'")
best_sct1_op_bum <- sqldf("select ID, VISIT,med_dose as BUMDOSE from best_sct1_op_melt where med_code=='JLA'")
best_sct1_op_tors <- sqldf("select ID, VISIT,med_dose as TORSDOSE from best_sct1_op_melt where med_code=='JLB'")

best_sct1_op_daily_furo <- best_sct1_op_furo

best_sct1_op_daily_furo <- merge(best_sct1_op_daily_furo,
                                 best_sct1_op_bum,
                                 by=c("ID","VISIT"),
                                 all.x=T)

best_sct1_op_daily_furo <- merge(best_sct1_op_daily_furo,
                                 best_sct1_op_tors,
                                 by=c("ID","VISIT"),
                                 all.x=T)

best_sct1_op_daily_furo$form <- "sct1"

best_sct1_op_daily_furo$daily_furo_eq_derive <- apply(best_sct1_op_daily_furo
                                                      [,c("FURODOSE",
                                                          "TORSDOSE",
                                                          "BUMDOSE")],
                                                      MARGIN=1,
                                                      FUN=diur_calc)

best_sct1_op_daily_furo$variable <- "daily_furo_eq_derive"
best_sct1_op_daily_furo$value <- best_sct1_op_daily_furo$daily_furo_eq_derive


best_sct1_op_melt <- rbind(best_sct1_op_melt[,c('ID',
                                                'form',
                                                'VISIT',
                                                "variable",
                                                "value")],
                           best_sct1_op_daily_furo[,c('ID',
                                                      'form',
                                                      'VISIT',
                                                      "variable",
                                                      "value")])


##########  COTX  ##########

best_cotx1 <- read.csv(paste(best_folder,"cotx1.csv",sep=""),stringsAsFactors = F,na.strings=c("NA","","NULL"))

best_cotx1_med1 <- best_cotx1[,c('ID','VISIT','CON1','COO1','COD1','COU1','COT1')]
best_cotx1_med2 <- best_cotx1[,c('ID','VISIT','CON2','COO2','COD2','COU2','COT2')]
best_cotx1_med3 <- best_cotx1[,c('ID','VISIT','CON3','COO3','COD3','COU3','COT3')]
best_cotx1_med4 <- best_cotx1[,c('ID','VISIT','CON4','COO4','COD4','COU4','COT4')]
best_cotx1_med5 <- best_cotx1[,c('ID','VISIT','CON5','COO5','COD5','COU5','COT5')]
best_cotx1_med6 <- best_cotx1[,c('ID','VISIT','CON6','COO6','COD6','COU6','COT6')]
best_cotx1_med7 <- best_cotx1[,c('ID','VISIT','CON7','COO7','COD7','COU7','COT7')]
best_cotx1_med8 <- best_cotx1[,c('ID','VISIT','CON8','COO8','COD8','COU8','COT8')]
best_cotx1_med9 <- best_cotx1[,c('ID','VISIT','CON9','COO9','COD9','COU9','COT9')]
best_cotx1_med10 <- best_cotx1[,c('ID','VISIT','CON10','COO10','COD10','COU10','COT10')]
best_cotx1_med11 <- best_cotx1[,c('ID','VISIT','CON11','COO11','COD11','COU11','COT11')]
best_cotx1_med12 <- best_cotx1[,c('ID','VISIT','CON12','COO12','COD12','COU12','COT12')]

names(best_cotx1_med1) <- c('ID','VISIT','med_name','med_code','med_dose','med_units','med_cat')
names(best_cotx1_med2) <- c('ID','VISIT','med_name','med_code','med_dose','med_units','med_cat')
names(best_cotx1_med3) <- c('ID','VISIT','med_name','med_code','med_dose','med_units','med_cat')
names(best_cotx1_med4) <- c('ID','VISIT','med_name','med_code','med_dose','med_units','med_cat')
names(best_cotx1_med5) <- c('ID','VISIT','med_name','med_code','med_dose','med_units','med_cat')
names(best_cotx1_med6) <- c('ID','VISIT','med_name','med_code','med_dose','med_units','med_cat')
names(best_cotx1_med7) <- c('ID','VISIT','med_name','med_code','med_dose','med_units','med_cat')
names(best_cotx1_med8) <- c('ID','VISIT','med_name','med_code','med_dose','med_units','med_cat')
names(best_cotx1_med9) <- c('ID','VISIT','med_name','med_code','med_dose','med_units','med_cat')
names(best_cotx1_med10) <- c('ID','VISIT','med_name','med_code','med_dose','med_units','med_cat')
names(best_cotx1_med11) <- c('ID','VISIT','med_name','med_code','med_dose','med_units','med_cat')
names(best_cotx1_med12) <- c('ID','VISIT','med_name','med_code','med_dose','med_units','med_cat')

best_cotx1_op_melt <- rbind(best_cotx1_med1,
                            best_cotx1_med2,
                            best_cotx1_med3,
                            best_cotx1_med4,
                            best_cotx1_med5,
                            best_cotx1_med6,
                            best_cotx1_med7,
                            best_cotx1_med8,
                            best_cotx1_med9,
                            best_cotx1_med10,
                            best_cotx1_med11,
                            best_cotx1_med12)


best_cotx1_op_melt <- subset(best_cotx1_op_melt,!is.na(med_code)&VISIT %in% c(1,30,60,120,180,240,300,360))

best_pt_visits <- sqldf("select ID, VISIT 
                        from best_cotx1_op_melt 
                        group by ID,VISIT")

best_cotx1_op_furo <- sqldf("select ID,VISIT, 
                            med_dose as FURODOSE 
                            from best_cotx1_op_melt 
                            where med_code=='JHG'")

best_cotx1_op_bum <- sqldf("select ID,VISIT,med_dose as BUMDOSE
                           from best_cotx1_op_melt 
                           where med_code=='JLA'")

best_cotx1_op_tors <- sqldf("select ID,VISIT,med_dose as TORSDOSE 
                            from best_cotx1_op_melt 
                            where med_code=='JLB'")


best_cotx1_op_daily_furo <- merge(best_pt_visits,
                                  best_cotx1_op_furo,
                                  by=c("ID","VISIT"),
                                  all.x=T)


best_cotx1_op_daily_furo <- merge(best_cotx1_op_daily_furo,
                                  best_cotx1_op_bum,
                                  by=c("ID","VISIT"),
                                  all.x=T)

best_cotx1_op_daily_furo <- merge(best_cotx1_op_daily_furo,
                                  best_cotx1_op_tors,
                                  by=c("ID","VISIT"),
                                  all.x=T)


best_cotx1_op_daily_furo$daily_furo_eq_derive <- apply(best_cotx1_op_daily_furo
                                                       [,c("FURODOSE",
                                                           "TORSDOSE",
                                                           "BUMDOSE")],
                                                       MARGIN=1,
                                                       FUN=diur_calc)

best_cotx1_op_daily_furo <- subset(best_cotx1_op_daily_furo,
                                   !is.na(best_cotx1_op_daily_furo$daily_furo_eq_derive))

best_cotx1_op_daily_furo$variable <- "daily_furo_eq_derive"
best_cotx1_op_daily_furo$form <- "cotx1"
best_cotx1_op_daily_furo$value <- best_cotx1_op_daily_furo$daily_furo_eq_derive


best_cotx1_op_melt$form <- "cotx1"
best_cotx1_op_melt$variable <- "med_code"
best_cotx1_op_melt$value <- best_cotx1_op_melt$med_code

best_cotx1_op_melt <- rbind(best_cotx1_op_melt[,c('ID',
                                                  'form',
                                                  'VISIT',
                                                  "variable",
                                                  "value")],
                            best_cotx1_op_daily_furo[,c('ID',
                                                        'form',
                                                        'VISIT',
                                                        "variable",
                                                        "value")])

best_meds_op_melt <- rbind(best_sct1_op_melt[c('ID',
                                              'form',
                                              'VISIT',
                                              "variable",
                                              "value")],
                           best_cotx1_op_melt[c('ID',
                                               'form',
                                               'VISIT',
                                               "variable",
                                               "value")])

best_meds_op_melt$value[best_meds_op_melt$value==0] <- NA


##########  compile  ##########

var_order <- c('ID',
                'form',
                'VISIT',
                "variable",
                "value")

best_melt <- rbind(best_pcsf_melt[,var_order],
                   best_t_melt[,var_order],
                   best_pe_melt[,var_order],
                   best_meds_op_melt[,var_order],
                   best_lab1_melt[,var_order],
                   best_clab1_melt[,var_order],
                   best_lab2_melt[,var_order],
                   best_clab2_melt[,var_order],
                   best_cvs_melt[,var_order],
                   best_muga_melt[,var_order],
                   best_mlhf_melt[,var_order],
                   best_cvh1_melt[,var_order],
                   best_cvh2_melt[,var_order],
                   best_ecg_melt[,var_order],
                   best_diab_melt[,var_order])


best_melt <- subset(best_melt,!value %in% c(NA,"NaN",""))


names(best_melt)[names(best_melt)=="ID"] <- "patientid"
names(best_melt)[names(best_melt)=="VISIT"] <- "studyvisit"
best_melt$study <- "BEST"


##############################  **********SCD-HeFT**********  ##############################


# ..%%%%%%...%%%%%%..%%%%%%%%..........%%.....%%.%%%%%%%%.%%%%%%%%.%%%%%%%%
# .%%....%%.%%....%%.%%.....%%.........%%.....%%.%%.......%%..........%%...
# .%%.......%%.......%%.....%%.........%%.....%%.%%.......%%..........%%...
# ..%%%%%%..%%.......%%.....%%.%%%%%%%.%%%%%%%%%.%%%%%%...%%%%%%......%%...
# .......%%.%%.......%%.....%%.........%%.....%%.%%.......%%..........%%...
# .%%....%%.%%....%%.%%.....%%.........%%.....%%.%%.......%%..........%%...
# ..%%%%%%...%%%%%%..%%%%%%%%..........%%.....%%.%%%%%%%%.%%..........%%...


##########  basecrf  ##########

scdheft_basecrf <- read_sas(paste(scdheft_folder,"basecrf.sas7bdat",sep=""))
scdheft_basecrf[scdheft_basecrf=="NaN"] <- NA

scdheft_basecrf$six_mw_distm <- round(scdheft_basecrf$CWALKDIS/3.28,1)
scdheft_basecrf$pupr <- scdheft_basecrf$CPESYS-scdheft_basecrf$CPEDIAS

scdheft_basecrf$CPEPEL_max <-
  apply(scdheft_basecrf[,c("CPEPELA","CPEPELL","CPEPERA","CPEPERL")],
        MARGIN=1,function(x) max(x,na.rm=T))

a <- subset(read.csv(paste(scdheft_folder,"scdheft sched furo.csv",sep=""), na.strings=c("","NA","NULL")))
b <- subset(read.csv(paste(scdheft_folder,"scdheft sched bum.csv",sep=""), na.strings=c("","NA","NULL")))
c <- subset(read.csv(paste(scdheft_folder,"scdheft sched tors.csv",sep=""), na.strings=c("","NA","NULL")))

scdheft_basecrf <- merge(scdheft_basecrf,a[a$FORM=="CRF",c("PID","FURO_DAILY")], by=c("PID"), all.x=T)
scdheft_basecrf <- merge(scdheft_basecrf,b[b$FORM=="CRF",c("PID","bum_daily")], by=c("PID"), all.x=T)
scdheft_basecrf <- merge(scdheft_basecrf,c[c$FORM=="CRF",c("PID","tors_daily")], by=c("PID"), all.x=T)

scdheft_basecrf$weight_kg <- round(scdheft_basecrf$CPEWGHT/2.2,1)

scdheft_basecrf$daily_furo_eq_derive <- apply(scdheft_basecrf
                                               [,c("FURO_DAILY",
                                                   "tors_daily",
                                                   "bum_daily")],
                                               MARGIN=1,
                                               FUN=diur_calc)


scdheft_basecrf_melt <- melt(scdheft_basecrf,id.vars=c("PID"),na.rm=T) 
scdheft_basecrf_melt$studyvisit <- "0"
scdheft_basecrf_melt$form <- "basecrf"


##########  walk  ##########

scdheft_walk <- read_sas(paste(scdheft_folder,"walk.sas7bdat",sep=""))

names(scdheft_walk)[names(scdheft_walk)=="FORM"] <- "studyvisit"
scdheft_walk_melt <- melt(data=scdheft_walk,id.vars=c("PID","studyvisit"),na.rm=T) 
scdheft_walk_melt$form <- "walk"


##########  baseline_new  ##########

scdheft_baseline_new <- read_sas(paste(scdheft_folder,"baseline_new.sas7bdat",sep=""))
scdheft_baseline_new_melt <- melt(data=scdheft_baseline_new,
                                  id.vars=c("PID"),
                                  na.rm=T)
scdheft_baseline_new_melt$form <- "baseline_new"
scdheft_baseline_new_melt$studyvisit <- "0"


##########  ecg  ##########

scdheft_ecg <- read_sas(paste(scdheft_folder,"ecg.sas7bdat",sep=""))
scdheft_ecg_melt <- melt(data=scdheft_ecg,id.vars=c("PID"),na.rm=T)
scdheft_ecg_melt$form <- "ecg"
scdheft_ecg_melt$studyvisit <- "0"

  

##########  rdemog  ##########

scdheft_rdemog <- read_sas(paste(scdheft_folder,"rdemog.sas7bdat",sep=""))
scdheft_rdemog_melt <- melt(data=scdheft_rdemog,
                            id.vars=c("PID"),
                            na.rm=T)

scdheft_rdemog_melt$form <- "rdemog"
scdheft_rdemog_melt$studyvisit <- "0"


##########  valvdz  ##########

scdheft_valvdz <- read_sas(paste(scdheft_folder,"valvdz.sas7bdat",sep=""))
scdheft_valvdz_melt <- melt(data=scdheft_valvdz,id.vars=c("PID"),na.rm=T)
scdheft_valvdz_melt$form <- "valvdz"
scdheft_valvdz_melt$studyvisit <- "0"


##########  btreat  ##########

scdheft_btreat <- read_sas(paste(scdheft_folder,"btreat.sas7bdat",sep=""))
scdheft_btreat_melt <- melt(data=scdheft_btreat,id.vars=c("PID"),na.rm=T)
scdheft_btreat_melt$form <- "btreat"
scdheft_btreat_melt$studyvisit <- "0"


#########  meds  ##########

scdheft_med_visits <- c("CRF", 
                        "03 MONTH FOLLOW-UP",
                        "06 MONTH FOLLOW-UP",
                        "12 MONTH FOLLOW-UP",
                        "18 MONTH FOLLOW-UP",
                        "24 MONTH FOLLOW-UP",
                        "30 MONTH FOLLOW-UP",
                        "36 MONTH FOLLOW-UP",
                        "42 MONTH FOLLOW-UP",
                        "48 MONTH FOLLOW-UP")

scdheft_meds1 <- subset(read_sas(paste(scdheft_folder,"meds1.sas7bdat",sep="")),
                        FORM %in% scdheft_med_visits)
scdheft_meds2 <- subset(read_sas(paste(scdheft_folder,"meds2.sas7bdat",sep="")),
                        FORM %in% scdheft_med_visits)
scdheft_meds3 <- subset(read_sas(paste(scdheft_folder,"meds3.sas7bdat",sep="")),
                        FORM %in% scdheft_med_visits)

scdheft_meds4 <- subset(read_sas(paste(scdheft_folder,"meds4.sas7bdat",sep="")),
                        FORM %in% scdheft_med_visits)


scdheft_meds4 <- merge(scdheft_meds4,a[,c("PID","FORM","FURO_DAILY")], by=c("PID","FORM"), all.x=T)
scdheft_meds4 <- merge(scdheft_meds4,b[,c("PID","FORM","bum_daily")], by=c("PID","FORM"), all.x=T)
scdheft_meds4 <- merge(scdheft_meds4,c[,c("PID","FORM","tors_daily")], by=c("PID","FORM"), all.x=T)

scdheft_meds4$daily_furo_eq_derive <- apply(scdheft_meds4
                                            [,c("FURO_DAILY",
                                                "tors_daily",
                                                "bum_daily")],
                                            MARGIN=1,
                                            FUN=diur_calc)

scdheft_meds5 <- subset(read_sas(paste(scdheft_folder,"meds5.sas7bdat",sep="")),
                        FORM %in% scdheft_med_visits)
scdheft_meds6 <- subset(read_sas(paste(scdheft_folder,"meds6.sas7bdat",sep="")),
                        FORM %in% scdheft_med_visits)
scdheft_meds7 <- subset(read_sas(paste(scdheft_folder,"meds7.sas7bdat",sep="")),
                        FORM %in% scdheft_med_visits)
scdheft_meds8 <- subset(read_sas(paste(scdheft_folder,"meds8.sas7bdat",sep="")),
                        FORM %in% scdheft_med_visits)
scdheft_meds9 <- subset(read_sas(paste(scdheft_folder,"meds9.sas7bdat",sep="")),
                        FORM %in% scdheft_med_visits)
scdheft_meds10 <- subset(read_sas(paste(scdheft_folder,"meds10.sas7bdat",sep="")),
                         FORM %in% scdheft_med_visits)
scdheft_meds11 <- subset(read_sas(paste(scdheft_folder,"meds11.sas7bdat",sep="")),
                         FORM %in% scdheft_med_visits)
scdheft_meds12 <- subset(read_sas(paste(scdheft_folder,"meds12.sas7bdat",sep="")),
                         FORM %in% scdheft_med_visits)

scdheft_meds <- merge(scdheft_meds1[,c("PID",
                                       "FORM",
                                       "VISDT_DAYS",
                                       "ACE",     # On any ACE inhibitor
                                       "ENAL",    # On enalapril
                                       "ENALDS",    # Enalapril dose
                                       "ENALSC",    # Enalapril schedule
                                       "CAPT",    # On captopril
                                       "CAPTDS",    # Captopril dose
                                       "CAPTSC",    # Captopril schedule
                                       "LISN",    # On lisinopril
                                       "LISNDS",    # Lisinopril dose
                                       "LISNSC",    # Lisinopril schedule
                                       "QUIN",    # On quinapril
                                       "QUINDS",    # Quinapril dose
                                       "QUINSC",    # Quinapril schedule
                                       "BENZ",    # On benazapril
                                       "BENZDS",    # Benazapril dose
                                       "BENZSC",    # Benaapril schedule
                                       "FOSN",    # On fosinopril
                                       "FOSNDS",    # Fosinopril dose
                                       "FOSNSC",    # Fosinopril schedule
                                       "AOTH",    # On other ACE inhibitor
                                       "ACEOTH",    # Other ACE inhibitor text
                                       "AOTHDS",    # Other ACE inhibitor dose
                                       "AOTHSC")],  # Other ACE inhibitor schedule
                      scdheft_meds2[,c("PID",
                                       "FORM",
                                       "LOSARTA", # On any ARB
                                       "LOSAR",   # On losartan
                                       "LOSARDS",   # Losartan dose
                                       "LOSARSC",   # Losartan schedule
                                       "ANGI",    # Other ARB
                                       "ANGIOTH",   # Other ARB text
                                       "ANGIDS",    # Other ARB dose
                                       "ANGISC")],  # Other ARB schedule
                      by=c("PID",
                          "FORM"))

scdheft_meds <- merge(scdheft_meds,
                      scdheft_meds3[,c("PID",
                                       "FORM",
                                       "DIG",     # On digoxin
                                       "DIGLVL")],# Digoxin levevl
                      by=c("PID",
                           "FORM"))


scdheft_meds <- merge(scdheft_meds,
                      scdheft_meds4[,c("PID",
                                       "FORM",
                                       "LDIUR",       # On any loop diuretic
                                       "FURO",        # On furosemide
                                       "FURODS",        # Furosemide dose
                                       "FUROSC",        # Furosemide schedule
                                       "BUMT",        # On bumetanide
                                       "BUMTDS",        # Bumetanide dose
                                       "BUMTSC",        # Bumetanide schedule
                                       "TORS",        # On torsemide
                                       "TORSDS",        # Torsemide dose
                                       "TORSSC",        # Torsemide schedule
                                       "ETHC",        # On ethacrynic acid
                                       "ETHCDS",        # Ethacrynic acid dose
                                       "ETHCSC",        # Ethacrynic acid schedule
                                       "LDOTH",       # On other loop diuretic
                                       "LDIUROT",       # Other loop diuretic text
                                       "LDOTHDS",       # Other loop diuretic dose
                                       "LDOTHSC",       # Other loop diuretic schedule
                                       "FURO_DAILY",  # Total daily furosemide dose 
                                       "bum_daily",   # Total daily bumetanide dose
                                       "tors_daily",  # Total daily torsemide dose
                                       "daily_furo_eq_derive")],  # Total furosemide equivalent daily dose
                      by=c("PID",
                           "FORM"))

scdheft_meds <- merge(scdheft_meds,
                      scdheft_meds5[,c("PID",
                                       "FORM",
                                       "PSDIUR",
                                       "SPIR",        # On spironolactone
                                       "SPIRDS",        # Spironolactone dose
                                       "SPIRSC",        # Spironolactone schedule
                                       "TRIA",        # On triamterene
                                       "TRIADS",        # Triamterene dose
                                       "TRIASC",        # Triamterene schedule
                                       "AMIL",        # On amiloride
                                       "AMILDS",        # Amiloride dose
                                       "AMILSC",        # Amiloride schedule
                                       "PSDOTH",      # On other K-sparing diuretic
                                       "PSDIURO",       # Other K-sparing diuretic text
                                       "PSDOTDS",       # Other K-sparing diuretic dose
                                       "PSDOTSC")],     # Other K-sparing diuretic schedule
                      by=c("PID",
                           "FORM"))

scdheft_meds <- merge(scdheft_meds,
                      scdheft_meds6[,c("PID",
                                       "FORM",
                                       "TDIUR",       # On thiazide diuretic
                                       "TDIUROT",       # Thiazide diuretic name
                                       "TDIURDS",       # Thiazide diuretic dose
                                       "TDIURSC")],     # Thiazide diuretic schedule
                      by=c("PID",
                           "FORM"))

scdheft_meds <- merge(scdheft_meds,
                      scdheft_meds7[,c("PID",
                                       "FORM",
                                       "WARF",        # On warfarin
                                       "WARFINR")],     # INR on warfarin
                      by=c("PID",
                           "FORM"))

scdheft_meds <- merge(scdheft_meds,
                      scdheft_meds8[,c("PID",
                                       "FORM",
                                       "ASA",         # On aspirin
                                       "AMLDP",       # On amlodipine
                                       "AMLDPDS",       # Amlodipine dose
                                       "AMLDPSC")],     # Amlodipine schedule
                      by=c("PID",
                           "FORM"))

scdheft_meds <- merge(scdheft_meds,
                      scdheft_meds9[,c(21,2,6:8,10:20)],
                      by=c("PID",
                           "FORM"))

scdheft_meds <- merge(scdheft_meds,
                      scdheft_meds10[,c(24,2,6:9,11:23)],
                      by=c("PID",
                           "FORM"))

scdheft_meds <- merge(scdheft_meds,
                      scdheft_meds11[,c(18,2,6:8,10:17)],
                      by=c("PID",
                           "FORM"))

scdheft_meds <- merge(scdheft_meds,
                      scdheft_meds12[,c(35,2,6:9,11:34)],
                      by=c("PID",
                           "FORM"))


scdheft_meds[scdheft_meds=="NaN"] <- NA

scdheft_meds_melt <- melt(scdheft_meds,id.vars=c("PID","FORM","VISDT_DAYS"),na.rm=T)

names(scdheft_meds_melt)[names(scdheft_meds_melt) %in% c("FORM")] <- c("studyvisit")

scdheft_meds_melt$form <- "meds"


########## follow-up form  ##########

scdheft_followup <- subset(read_sas(paste(scdheft_folder,"followup.sas7bdat",sep="")))

scdheft_followup <- merge(scdheft_followup,
                         scdheft_baseline_new[,c("PID","AGE","GENDER")],
                         by="PID",
                         all.x=T)

scdheft_followup <- merge(scdheft_followup,
                         scdheft_rdemog[,c("PID","RACE_REDACT")],
                         by="PID",
                         all.x=T)

scdheft_followup[scdheft_followup=="NaN"|scdheft_followup==""] <- NA

scdheft_followup$FPEPEL_max <-
  apply(scdheft_followup[,c("FPEPELA","FPEPELL","FPEPERA","FPEPERL")],
        MARGIN=1,function(x) max(x,na.rm=T))

scdheft_followup$FPEPEL_max[scdheft_followup$FPEPEL_max == "-Inf"] <- NA

scdheft_followup <- merge(scdheft_followup,a[,c("PID","FORM","FURO_DAILY")], by=c("PID","FORM"), all.x=T)
scdheft_followup <- merge(scdheft_followup,b[,c("PID","FORM","bum_daily")], by=c("PID","FORM"), all.x=T)
scdheft_followup <- merge(scdheft_followup,c[,c("PID","FORM","tors_daily")], by=c("PID","FORM"), all.x=T)

scdheft_followup$daily_furo_eq_derive <- apply(scdheft_followup
                                            [,c("FURO_DAILY",
                                                "tors_daily",
                                                "bum_daily")],
                                            MARGIN=1,
                                            FUN=diur_calc)


scdheft_followup$pupr <- scdheft_followup$FPESYS-scdheft_followup$FPEDIAS
scdheft_followup$weight_kg <- round(scdheft_followup$FPEWGHT/2.2,1)

scdheft_followup$bun[scdheft_followup$FCHBUNU==1&
                       !is.na(scdheft_followup$FCHBUNU)] <- 
  scdheft_followup$FCHBUN[scdheft_followup$FCHBUNU==1&
                            !is.na(scdheft_followup$FCHBUNU)]

scdheft_followup$bun[scdheft_followup$FCHBUNU==2&
                       scdheft_followup$FCHBUNUO=="MG/DL"&
                       !is.na(scdheft_followup$FCHBUNU)&
                       !is.na(scdheft_followup$FCHBUNUO)] <- 
  scdheft_followup$FCHBUN[scdheft_followup$
                            FCHBUNU==2&
                            scdheft_followup$
                            FCHBUNUO=="MG/DL"&
                            !is.na(scdheft_followup$FCHBUNU)&
                            !is.na(scdheft_followup$FCHBUNUO)]

scdheft_followup$bun[scdheft_followup$FCHBUNU==2&
                       scdheft_followup$FCHBUNUO=="MMOL/DL"&
                       !is.na(scdheft_followup$FCHBUNU)&
                       !is.na(scdheft_followup$FCHBUNUO)] <- 
  scdheft_followup$FCHBUN[scdheft_followup$FCHBUNU==2&
                            scdheft_followup$FCHBUNUO=="MMOL/L"&
                            !is.na(scdheft_followup$FCHBUNU)&
                            !is.na(scdheft_followup$FCHBUNUO)] /0.3571


scdheft_followup$creatinine[scdheft_followup$FCHCRU==1&
                              !is.na(scdheft_followup$FCHCRU)] <- 
  scdheft_followup$FCHCREAT[scdheft_followup$FCHCRU==1&
                              !is.na(scdheft_followup$FCHCRU)]

scdheft_followup$creatinine[scdheft_followup$FCHCRU==2&
                              !is.na(scdheft_followup$FCHCRU)&
                             scdheft_followup$FCHCRUO=="MG/DL"] <- 
  scdheft_followup$FCHCREAT[scdheft_followup$FCHCRU==2&
                              !is.na(scdheft_followup$FCHCRU)&
                             scdheft_followup$FCHCRUO=="MG/DL"]

scdheft_followup$creatinine[scdheft_followup$FCHCRU==2&
                              !is.na(scdheft_followup$FCHCRU)&
                              scdheft_followup$FCHCRUO=="UMOL/L"] <- 
  scdheft_followup$FCHCREAT[scdheft_followup$FCHCRU==2&
                              !is.na(scdheft_followup$FCHCRU)&
                              scdheft_followup$FCHCRUO=="UMOL/L"] / 88.42 

scdheft_followup$creatinine[scdheft_followup$FCHCRU==2&
                              !is.na(scdheft_followup$FCHCRU)&
                             scdheft_followup$FCHCRUO=="MMOL/L"] <- 
  scdheft_followup$FCHCREAT[scdheft_followup$FCHCRU==2&
                              !is.na(scdheft_followup$FCHCRU)&
                             scdheft_followup$FCHCRUO=="MMOL/L"] / 0.08842 


scdheft_followup$gfr <- calc_MDRD4(dat=scdheft_followup,
                               cr="creatinine",
                               sex="GENDER",
                               male=1,
                               race="RACE_REDACT",
                               black="African American",
                               age="AGE")


scdheft_followup$magnesium[scdheft_followup$FCHMGU==1&
                             !is.na(scdheft_followup$FCHMGU)] <- 
  scdheft_followup$FCHMG[scdheft_followup$FCHMGU==1&
                           !is.na(scdheft_followup$FCHMGU)]

scdheft_followup$magnesium[scdheft_followup$FCHMGU==2&
                             scdheft_followup$FCHMGUO=="MG/DL"&
                             !is.na(scdheft_followup$FCHMGU)&
                             !is.na(scdheft_followup$FCHMGUO)] <- 
  scdheft_followup$FCHMG[scdheft_followup$FCHMGU==2&
                           scdheft_followup$FCHMGUO=="MG/DL"&
                           !is.na(scdheft_followup$FCHMGU)&
                           !is.na(scdheft_followup$FCHMGUO)]

scdheft_followup$magnesium[scdheft_followup$FCHMGU==2&
                             scdheft_followup$FCHMGUO=="MEQ/L"&
                             !is.na(scdheft_followup$FCHMGU)&
                             !is.na(scdheft_followup$FCHMGUO)] <- 
  scdheft_followup$FCHMG[scdheft_followup$FCHMGU==2&
                           scdheft_followup$FCHMGUO=="MEQ/L"&
                           !is.na(scdheft_followup$FCHMGU)&
                           !is.na(scdheft_followup$FCHMGUO)]*2.43

scdheft_followup$magnesium[scdheft_followup$FCHMGU==2&
                             scdheft_followup$FCMGUO=="MMOL/L"&
                             !is.na(scdheft_followup$FCHMGU)&
                             !is.na(scdheft_followup$FCHMGUO)] <- 
  scdheft_followup$FCHMG[scdheft_followup$FCHMGU==2&
                           scdheft_followup$FCHMGUO=="MMOL/L"&
                           !is.na(scdheft_followup$FCHMGU)&
                           !is.na(scdheft_followup$FCHMGUO)]*2.43


####### 6 MW convert#####

scdheft_followup$six_mw_distm <- round(scdheft_followup$FWALKDIS/3.28,1)


scdheft_followup_melt <- melt(subset(scdheft_followup,
                                     FORM %in% scdheft_med_visits),
                                     id.vars=c("PID","FORM"),
                                     na.rm=T)

names(scdheft_followup_melt)[names(scdheft_followup_melt)=="FORM"] <- "studyvisit"

scdheft_followup_melt$form <- "followup"


#### lwhfques ####

scdheft_lwhfques <- read_sas(paste(scdheft_folder,"lwhfques.sas7bdat",sep=""))
scdheft_lwhfques[scdheft_lwhfques=="NaN"] <- NA

scdheft_mlhf <- score_mlhf(dat=scdheft_lwhfques,
                           swelling = "SWELL", 
                           day_rest="REST",
                           stairs = "WALKC", 
                           housework="WORK",
                           go_places="AWAY",
                           sleep = "SLEEP",
                           friendfam = "FRIENDS",
                           earn_living = "DIFF",
                           hobbies = "HOBBIE",
                           sex = "SEX",
                           food = "EAT",
                           sob = "BREATH",
                           fatigue = "ENERGY",
                           hospital = "STAY",
                           cost = "COST",
                           sideeffect = "EFFECT",
                           burden = "BURDEN",
                           nocontrol = "LOSS",
                           worry="WORRY",
                           concentrate="REMEM",
                           depressed="DEPRS", 
                           total=c("Y"))

scdheft_mlhf$studyvisit <- scdheft_mlhf$FORM

scdheft_mlhf_melt <- melt(scdheft_mlhf,
                          id.vars=c("PID","studyvisit"),
                          na.rm=T)

scdheft_mlhf_melt <- subset(scdheft_mlhf_melt,!value==""&!is.na(value))
scdheft_mlhf_melt$form <- "lwhfques"
scdheft_mlhf_melt$study <- "SCD-HeFT"


##########  compile  ##########

scdheft_melt <- rbind(scdheft_basecrf_melt[,c("PID","studyvisit","form","variable","value")],
                      scdheft_baseline_new_melt[,c("PID","studyvisit","form","variable","value")],
                      scdheft_btreat_melt[,c("PID","studyvisit","form","variable","value")],
                      scdheft_ecg_melt[,c("PID","studyvisit","form","variable","value")],
                      scdheft_followup_melt[,c("PID","studyvisit","form","variable","value")],
                      scdheft_mlhf_melt[,c("PID","studyvisit","form","variable","value")],
                      scdheft_meds_melt[,c("PID","studyvisit","form","variable","value")],
                      scdheft_rdemog_melt[,c("PID","studyvisit","form","variable","value")],
                      scdheft_valvdz_melt[,c("PID","studyvisit","form","variable","value")],
                      scdheft_walk_melt[,c("PID","studyvisit","form","variable","value")])

scdheft_melt <- subset(scdheft_melt,!value %in% c(NA,"NaN",""))

colnames(scdheft_melt)[1] <- "patientid"
scdheft_melt$study <- "SCD-HeFT"


#######################  ********TOPCAT*******  #############################

# .%%%%%%%%..%%%%%%%..%%%%%%%%...%%%%%%.....%%%....%%%%%%%%
# ....%%....%%.....%%.%%.....%%.%%....%%...%%.%%......%%...
# ....%%....%%.....%%.%%.....%%.%%........%%...%%.....%%...
# ....%%....%%.....%%.%%%%%%%%..%%.......%%.....%%....%%...
# ....%%....%%.....%%.%%........%%.......%%%%%%%%%....%%...
# ....%%....%%.....%%.%%........%%....%%.%%.....%%....%%...
# ....%%.....%%%%%%%..%%.........%%%%%%..%%.....%%....%%...


######  t003 - Demographics ####

topcat_t003 <- read_sas(paste(topcat_folder,"t003.sas7bdat",sep=""))


##  Here we override the race categorization with Hispanic ethnicity if Hispanic ethnicity is specified

topcat_t003$race_cat[topcat_t003$ETHNICITY==1] <- 8

topcat_t003_melt <- melt(data=topcat_t003,id.vars=c("ID","VISIT"),na.rm=T)
topcat_t003_melt$form <- "t003"


######  t006 - Baseline physical exam ####

thisfile <- paste(topcat_folder,"t006.sas7bdat",sep="")
topcat_t006 <- read.sas7bdat(thisfile)

topcat_t006$pupr <- topcat_t006$SBP-topcat_t006$DBP
topcat_t006$bsa <- calc_bsa(topcat_t006,weight_kg="weight",height_cm="height")
topcat_t006$bmi <- calc_bmi(topcat_t006,weight="weight",height="height")


topcat_t006_melt <- melt(data=topcat_t006,id.vars=c("ID","VISIT"),na.rm=T)
topcat_t006_melt$form <- "t006"


##########  t002 - eligibility version D  ##########

topcat_t002 <- read.sas7bdat(paste(topcat_folder,"t002.sas7bdat",sep=""))
topcat_t002$BNP_VALUE[topcat_t002$BNP_TYPE==1] <- topcat_t002$BNP_VAL[topcat_t002$BNP_TYPE==1] 
topcat_t002$PROBNP_VALUE[topcat_t002$BNP_TYPE==2] <- topcat_t002$BNP_VAL[topcat_t002$BNP_TYPE==2] 

topcat_t002_melt <- 
  melt(data=topcat_t002,id.vars=c("ID","VISIT"),na.rm=T) 
topcat_t002_melt$form <- "t002"


###### t004 - Clinical evidence of HF ####

topcat_t004 <- read.sas7bdat(paste(topcat_folder,"t004.sas7bdat",sep=""))
topcat_t004[topcat_t004=="NaN"|topcat_t004<0] <- NA


topcat_t004_melt <- melt(data=topcat_t004,id.vars=c("ID","VISIT"),na.rm=T)
topcat_t004_melt$form <- "t004"


###### t013 - Follow-up physical exam ####

topcat_t013 <- read.sas7bdat(paste(topcat_folder,"t013.sas7bdat",sep=""))
topcat_t013[topcat_t013=="NaN"|topcat_t013<0] <- NA

topcat_t013 <- merge(topcat_t013,topcat_t006[c("ID","height")])

topcat_t013$pupr <- topcat_t013$SBP-topcat_t013$DBP
topcat_t013$bsa <- calc_bsa(topcat_t013,weight_kg="weight",height_cm="height")
topcat_t013$bmi <- calc_bmi(topcat_t013,weight="weight",height="height")

topcat_t013_melt <- melt(data=topcat_t013,id.vars=c("ID","VISIT"),na.rm=T)
topcat_t013_melt$form <- "t013"


##########  echo_baseline  ##########

topcat_echo_baseline <- read.sas7bdat(paste(topcat_folder,"echo_baseline.sas7bdat",sep=""))
topcat_echo_baseline[topcat_echo_baseline=="NaN"] <- NA


topcat_echo_baseline <- merge(topcat_echo_baseline,topcat_t004[,c("ID","JVP_CUR")],by="ID",all.x=T)
topcat_echo_baseline <- merge(topcat_echo_baseline,topcat_t003[,c("ID","GENDER")],by="ID",all.x=T)
topcat_echo_baseline <- merge(topcat_echo_baseline,topcat_t006[,c("ID","bsa","weight","height")],by="ID",all.x=T)

topcat_echo_baseline$tr_ms <- topcat_echo_baseline$tr/100
topcat_echo_baseline$RVSP <- 5*(topcat_echo_baseline$JVP_CUR+1) + 4*(topcat_echo_baseline$tr_ms)^2

topcat_echo_baseline <- calc_hypertrophy_type(df=topcat_echo_baseline,
                                              id="ID",
                                              sex="GENDER",
                                              lvedd_cm = "lvdd",
                                              ivsd_cm = "lvs",
                                              lvpwtd_cm = "lvpw",
                                              height_cm = "height",
                                              weight_kg = "weight")


topcat_echo_baseline$LVEDD_index <- topcat_echo_baseline$lvdd/topcat_echo_baseline$bsa
topcat_echo_baseline$LVESD_index <- topcat_echo_baseline$lvesd/topcat_echo_baseline$bsa
topcat_echo_baseline$LVEDV_index <- topcat_echo_baseline$lvedv/topcat_echo_baseline$bsa
topcat_echo_baseline$LAV_index <- topcat_echo_baseline$lav/topcat_echo_baseline$bsa
topcat_echo_baseline$RVEDA_index <- topcat_echo_baseline$rveda/topcat_echo_baseline$bsa
topcat_echo_baseline$eeprime_avg <- apply(topcat_echo_baseline[,c("eeprime_lat",
                                                                  "eeprime_sept")],
                                          MARGIN=1,
                                          FUN=mean)

topcat_echo_bl_pulse <-
  merge(topcat_echo_baseline[,c("ID","occurred_on1","lvedv","lvesv")],
        topcat_t006[,c("ID", "visit_dt1", "SBP", "DBP", "HR", "pupr","bsa")],
        by="ID")

topcat_echo_bl_pulse$time_diff <- 
  round(abs(topcat_echo_bl_pulse$occurred_on1-
        topcat_echo_bl_pulse$visit_dt1)*365,0)

topcat_echo_bl_pulse <- subset(topcat_echo_bl_pulse, time_diff < 30)

topcat_echo_bl_pulse <- 
  calc_pulsatility(dat=topcat_echo_bl_pulse,
                   sbp = "SBP",
                   dbp = "DBP",
                   edv = "lvedv",
                   esv = "lvesv",
                   bsa = "bsa",
                   hr = "HR",
                   lvot_d = NA,
                   vti = NA)

topcat_echo_baseline <-
  merge(topcat_echo_baseline,
        topcat_echo_bl_pulse[,c("ID",
                                "sv_pulsatile",
                                "svi_pulsatile",
                                "co_pulsatile",
                                "ci_pulsatile",
                                "map",
                                "arterial_elastance_ix_pulsatile",
                                "syst_art_comp_pulsatile",
                                "svr_pulsatile",
                                "svr_index_pulsatile")],
        by="ID")

topcat_echo_baseline_melt <- 
  melt(data=topcat_echo_baseline,id.vars=c("ID","visit1"),na.rm=T) 
names(topcat_echo_baseline_melt)[names(topcat_echo_baseline_melt)=="visit1"] <- "VISIT"

topcat_echo_baseline_melt$form <- "echo_baseline"


##########  echo_followup  ##########

topcat_echo_followup <- read.sas7bdat(paste(topcat_folder,"echo_followup.sas7bdat",sep=""))
topcat_echo_followup[topcat_echo_followup=="NaN"] <- NA

topcat_echo_followup$VISIT[topcat_echo_followup$visit1=="12 Months"] <- "MN12"
topcat_echo_followup$VISIT[topcat_echo_followup$visit1=="18 Months"] <- "MN18"

topcat_echo_followup <- 
  merge(topcat_echo_followup,
        topcat_t013[,c("ID",
                       "VISIT",
                       "JVP",
                       "SBP",
                       "DBP",
                       "HR",
                       "bsa")],
        by=c("ID","VISIT"),
        all.x=T)

topcat_echo_followup <- merge(topcat_echo_followup,topcat_t003[,c("ID","GENDER")],by="ID",all.x=T)
topcat_echo_followup <- merge(topcat_echo_followup,topcat_t006[,c("ID","bsa","weight","height")],by="ID",all.x=T)

topcat_echo_followup$tr_ms <- topcat_echo_followup$tr/100
topcat_echo_followup$RVSP <- 5*(topcat_echo_followup$JVP+1) + 4*(topcat_echo_followup$tr_ms)^2

topcat_echo_followup <- calc_hypertrophy_type(df=topcat_echo_followup,
                                              id="ID",
                                              sex="GENDER",
                                              lvedd_cm = "lvdd",
                                              ivsd_cm = "lvs",
                                              lvpwtd_cm = "lvpw",
                                              height_cm = "height",
                                              weight_kg = "weight")


topcat_echo_followup$LVEDD_index <- topcat_echo_followup$lvdd/topcat_echo_followup$bsa
topcat_echo_followup$LVESD_index <- topcat_echo_followup$lvesd/topcat_echo_followup$bsa
topcat_echo_followup$LVEDV_index <- topcat_echo_followup$lvedv/topcat_echo_followup$bsa
topcat_echo_followup$LAV_index <- topcat_echo_followup$lav/topcat_echo_followup$bsa
topcat_echo_followup$RVEDA_index <- topcat_echo_followup$rveda/topcat_echo_followup$bsa
topcat_echo_followup$eeprime_avg <- apply(topcat_echo_followup[,c("eeprime_lat",
                                                                  "eeprime_sept")],
                                          MARGIN=1,
                                          FUN=mean)

topcat_echo_followup <- 
  calc_pulsatility(dat=topcat_echo_followup,
                   sbp = "SBP",
                   dbp = "SBP",
                   edv = "lvedv",
                   esv = "lvesv",
                   bsa = "bsa",
                   hr = "HR")


topcat_echo_followup_melt <- 
  melt(data=topcat_echo_followup,id.vars=c("ID","VISIT"),na.rm=T) 

topcat_echo_followup_melt$form <- "echo_followup"


######  t008 - Baseline lab tests ####


topcat_t008 <- read.sas7bdat(paste(topcat_folder,"t008.sas7bdat",sep=""))
topcat_t008[topcat_t008=="NaN"] <- NA
topcat_t008[topcat_t008 < 0] <- NA

topcat_t008 <- merge(topcat_t008,
                          topcat_t003[,c("ID","age_entry","GENDER","RACE_BLACK")],
                          by="ID",
                          all.x=T)


topcat_t008$gfr <- calc_MDRD4(dat=topcat_t008,
                                   cr="CR_mgdl",
                                   age="age_entry",
                                   sex = "GENDER",
                                   race= "RACE_BLACK",
                                   male=1,
                                   black=1)

topcat_t008_melt <- melt(data=topcat_t008,id.vars=c("ID","VISIT"),na.rm=T)
topcat_t008_melt$form <- "t008"


######  t016 - Follow-up lab tests ####

topcat_t016 <- 
  read.sas7bdat(
    paste(
      topcat_folder,
      "t016.sas7bdat",
      sep=""))

topcat_t016[topcat_t016=="NaN"|topcat_t016 < 0] <- NA

topcat_t016 <- 
  sqldf('select *
        from topcat_t016
        where VISIT <> "BASE"
        group by ID, VISIT
        having min(labs_dt1)')

topcat_t016 <- 
  merge(topcat_t016,
        topcat_t003[,c("ID",
                       "age_entry",
                       "GENDER",
                       "RACE_BLACK")],
        by="ID",
        all.x=T)


topcat_t016$gfr <- calc_MDRD4(dat=topcat_t016,
                                   cr="CR_mgdL",
                                   age="age_entry",
                                   sex = "GENDER",
                                   race= "RACE_BLACK",
                                   male=1,
                                   black=1)

topcat_t016_melt <- melt(data=topcat_t016,id.vars=c("ID","VISIT"),na.rm=T)
topcat_t016_melt$form <- "t016"


##### q002 - KCCQ #####

topcat_q002 <- read.sas7bdat(paste(topcat_folder,"q002.sas7bdat",sep=""))
topcat_q002[topcat_q002=="NaN"] <- NA


topcat_kccq <- sqldf('select "TOPCAT" as study,
                        ID,
                        VISIT,
                        DRESS as KCCQ1A,
                        SHOWER as KCCQ1B,
                        WALK as KCCQ1C,
                        YARDWORK as KCCQ1D,
                        CLIMB as KCCQ1E,
                        HURRY as KCCQ1F,
                        SYMP_CHANGE as KCCQ2,
                        SWELL_NUM as KCCQ3,
                        SWELL_FEEL as KCCQ4,
                        FATIGUE_NUM as KCCQ5,
                        FATIGUE_FEEL as KCCQ6,
                        BREATH_NUM as KCCQ7,
                        BREATH_FEEL as KCCQ8,
                        BREATH_SLEEP as KCCQ9,
                        TO_DO as KCCQ10,
                        UNDERSTAND as KCCQ11,
                        LIMIT_LIFE as KCCQ12,
                        REST_LIFE as KCCQ13,
                        DISCOURAGED as KCCQ14,
                        HOBBIES as KCCQ15A,
                        WORKING as KCCQ15B,
                        VISITING as KCCQ15C,
                        INTIMATE as KCCQ15D
                        from topcat_q002')

topcat_kccq <- score_kccq(dat=topcat_kccq,
                          q1a="KCCQ1A",
                          q1b="KCCQ1B",
                          q1c="KCCQ1C",
                          q1d="KCCQ1D",
                          q1e="KCCQ1E",
                          q1f="KCCQ1F",
                          q2="KCCQ2",
                          q3="KCCQ3",
                          q4="KCCQ4",
                          q5="KCCQ5",
                          q6="KCCQ6",
                          q7="KCCQ7",
                          q8="KCCQ8",
                          q9="KCCQ9",
                          q10="KCCQ10",
                          q11="KCCQ11",
                          q12="KCCQ12",
                          q13="KCCQ13",
                          q14="KCCQ14",
                          q15a="KCCQ15A",
                          q15b="KCCQ15B",
                          q15c="KCCQ15C",
                          q15d="KCCQ15D")

topcat_kccq_melt <- melt(data=topcat_kccq,
                         id.vars=c("ID","study","VISIT"),
                         na.rm=T)

topcat_kccq_melt$form <- "kccq"

#### q003 Visual Analog Scale global ####

topcat_q003_melt <- read.sas7bdat(paste(topcat_folder,"q003.sas7bdat",sep=""))
topcat_q003_melt[topcat_q003_melt=="NaN"|topcat_q003_melt < 0] <- NA

names(topcat_q003_melt)[3] <- "value"
topcat_q003_melt$variable <- "HEALTH_SCALE"
topcat_q003_melt$form <- "q003"


######  t005 - Medical history ####

topcat_t005 <- read.sas7bdat(paste(topcat_folder,"t005.sas7bdat",sep=""))
topcat_t005[topcat_t005 == -2] <- NA 

## Where both paroxysmal and chronic AF are indicated, select chronic

topcat_t005$aftype[topcat_t005$AFIB==1&topcat_t005$AFIB_PAROX==1] <- "paroxysmal"
topcat_t005$aftype[topcat_t005$AFIB==1&topcat_t005$AFIB_CHRON==1] <- "chronic"


topcat_t005_melt <- melt(data=topcat_t005,id.vars=c("ID","VISIT"),na.rm=T)
topcat_t005_melt$form <- "t005"


######  t0010 - Electrocardiogram ####

topcat_t010 <- read.sas7bdat(paste(topcat_folder,"t010.sas7bdat",sep=""))
topcat_t010_melt <- melt(data=topcat_t010,id.vars=c("ID","VISIT"),na.rm=T)
topcat_t010_melt$form <- "t010"


######  t007_allvisits_bysub - Medications by class ####

topcat_t007_allvisits_bysub <- read.sas7bdat(paste(topcat_folder,"t007_allvisits_bysub.sas7bdat",sep=""))
topcat_t007_allvisits_bysub_melt <- melt(data=topcat_t007_allvisits_bysub,id.vars=c("ID","VISIT"),na.rm=T)
topcat_t007_allvisits_bysub_melt$form <- "t007_allvisits_bysub"


###### t011 - Randomization visit ####

topcat_t011 <- read.sas7bdat(paste(topcat_folder,"t011.sas7bdat",sep=""))
topcat_t011_melt <- melt(data=topcat_t011,id.vars=c("ID","VISIT"),na.rm=T)
topcat_t011_melt$form <- "t011"


##########  t007_allvisits_bymed - Medications by individual med  ##########

topcat_t007_allvisits_bymed <- read.sas7bdat(paste(topcat_folder,"t007_allvisits_bymed.sas7bdat",sep=""))
topcat_t007_allvisits_bymed$variable <- "MEDCAT_WHODDE"
topcat_t007_allvisits_bymed$study <- "TOPCAT"
topcat_t007_allvisits_bymed$form <- "t007_allvisits_bymed"
topcat_t007_allvisits_bymed$value <- as.character(topcat_t007_allvisits_bymed$MEDCAT_WHODDE)


topcat_t007_allvisits_bymed2 <- read.sas7bdat(paste(topcat_folder,"t007_allvisits_bymed.sas7bdat",sep=""))
topcat_t007_allvisits_bymed2$variable <- "CODED_MED_NAME"
topcat_t007_allvisits_bymed2$study <- "TOPCAT"
topcat_t007_allvisits_bymed2$form <- "t007_allvisits_bymed"
topcat_t007_allvisits_bymed2$value <- as.character(topcat_t007_allvisits_bymed2$CODED_MED_NAME)

#### t007 diuretic dose ####

topcat_t007_allvisits_bymed$FURODOSE[
  topcat_t007_allvisits_bymed$CODED_MED_NAME %in% 
    c("FUROSEMMIDE",
      "FUROSEMIDA",
      "FUROSEMID",
      "FUROSE",
      "FURO",
      "FUROSEMIDUM",
      "FURSEMID",
      "FURSEMIDA",
      "LASIX",
      "APO FUROSEMIDE",
      "APO-FUROSEMIDE",
      "NOVOSEMIDE",
      "NOVO-SEMIDE",
      "NURIBAN")] <- 
  topcat_t007_allvisits_bymed$DOSE[
    topcat_t007_allvisits_bymed$CODED_MED_NAME %in%
      c("FUROSEMMIDE",
        "FUROSEMIDA",
        "FUROSEMID",
        "FUROSE",
        "FURO",
        "FUROSEMIDUM",
        "FURSEMID",
        "FURSEMIDA",
        "LASIX",
        "APO FUROSEMIDE",
        "APO-FUROSEMIDE",
        "NOVOSEMIDE",
        "NOVO-SEMIDE",
        "NURIBAN")]


topcat_t007_allvisits_bymed$TORSDOSE[
  topcat_t007_allvisits_bymed$CODED_MED_NAME %in% 
    c("DEMADEX", 
      "TORASEMID",
      "TORASEMIDE",
      "TORSEMIDE",
      "DIUVER")] <- 
  topcat_t007_allvisits_bymed$DOSE[
    topcat_t007_allvisits_bymed$CODED_MED_NAME %in% 
      c("DEMADEX", 
        "TORASEMID",
        "TORASEMIDE",
        "TORSEMIDE",
        "DIUVER")]  


topcat_t007_allvisits_bymed$BUMDOSE[
  topcat_t007_allvisits_bymed$CODED_MED_NAME %in% 
    c("BUMETANIDE",
      "BUMEX",
      "BURINEX")] <- 
  
  topcat_t007_allvisits_bymed$DOSE[
    topcat_t007_allvisits_bymed$CODED_MED_NAME %in% 
      c("BUMETANIDE",
        "BUMEX",
        "BURINEX")] 


topcat_t007_allvisits_bymed$ETHADOSE[
  topcat_t007_allvisits_bymed$CODED_MED_NAME %in% 
    c("EDECRIN",
      "ETACRYNIC ACID",
      "ETHACRYNIC ACID")] <- 
  topcat_t007_allvisits_bymed$DOSE[
    topcat_t007_allvisits_bymed$CODED_MED_NAME %in% 
      c("EDECRIN",
        "ETACRYNIC ACID",
        "ETHACRYNIC ACID")]  


topcat_t007_allvisits_bymed$daily_furo_eq_derive <- 
  apply(topcat_t007_allvisits_bymed
        [,c("FURODOSE",
            "TORSDOSE",
            "BUMDOSE",
            "ETHADOSE")],
        MARGIN=1,
        FUN=diur_calc)


topcat_t007_diur_melt <- sqldf("select ID, 
                               't007_allvisits_bymed' as form,
                               VISIT,
                               'daily_furo_eq_derive' as variable,
                               sum(daily_furo_eq_derive) as value
                               from topcat_t007_allvisits_bymed
                               where daily_furo_eq_derive >= 0
                               group by ID, form, visit")


##########  compile  ##########

topcat_melt <- rbind(topcat_q003_melt[,c("ID","form","VISIT","variable","value")],
                     topcat_t002_melt[,c("ID","form","VISIT","variable","value")],
                     topcat_t003_melt[,c("ID","form","VISIT","variable","value")],
                     topcat_t004_melt[,c("ID","form","VISIT","variable","value")],
                     topcat_t005_melt[,c("ID","form","VISIT","variable","value")],
                     topcat_t006_melt[,c("ID","form","VISIT","variable","value")],
                     topcat_t007_allvisits_bymed[,c("ID","form","VISIT","variable","value")],
                     topcat_t007_allvisits_bymed2[,c("ID","form","VISIT","variable","value")],
                     topcat_t007_allvisits_bysub_melt[,c("ID","form","VISIT","variable","value")],
                     topcat_t007_diur_melt[,c("ID","form","VISIT","variable","value")],
                     topcat_t008_melt[,c("ID","form","VISIT","variable","value")],
                     topcat_t010_melt[,c("ID","form","VISIT","variable","value")],
                     topcat_t011_melt[,c("ID","form","VISIT","variable","value")],
                     topcat_t013_melt[,c("ID","form","VISIT","variable","value")],
                     topcat_t016_melt[,c("ID","form","VISIT","variable","value")],
                     topcat_kccq_melt[,c("ID","form","VISIT","variable","value")],
                     topcat_echo_baseline_melt[,c("ID","form","VISIT","variable","value")],
                     topcat_echo_followup_melt[,c("ID","form","VISIT","variable","value")])

topcat_melt <- subset(topcat_melt,!is.na(value)&!value=="NaN")
topcat_melt <- subset(topcat_melt,value >=0)

colnames(topcat_melt)[colnames(topcat_melt)=="ID"] <- "patientid"
colnames(topcat_melt)[colnames(topcat_melt)=="VISIT"] <- "studyvisit"

topcat_melt$study <- "TOPCAT"


############################# **********PARADIGM**********  ################################

# .%%%%%%%%.....%%%....%%%%%%%%.....%%%....%%%%%%%%..%%%%..%%%%%%...%%.....%%
# .%%.....%%...%%.%%...%%.....%%...%%.%%...%%.....%%..%%..%%....%%..%%%...%%%
# .%%.....%%..%%...%%..%%.....%%..%%...%%..%%.....%%..%%..%%........%%%%.%%%%
# .%%%%%%%%..%%.....%%.%%%%%%%%..%%.....%%.%%.....%%..%%..%%...%%%%.%%.%%%.%%
# .%%........%%%%%%%%%.%%...%%...%%%%%%%%%.%%.....%%..%%..%%....%%..%%.....%%
# .%%........%%.....%%.%%....%%..%%.....%%.%%.....%%..%%..%%....%%..%%.....%%
# .%%........%%.....%%.%%.....%%.%%.....%%.%%%%%%%%..%%%%..%%%%%%...%%.....%%


paradigm_sample <- as.matrix(read.dta13(paste(paradigm_folder,
                                              'paradigmhf sample with labs and vitals.dta',
                                              sep=""),
                              convert.factors=F))

paradigm_sample <- as.data.frame(paradigm_sample)

paradigm_sample_melt <- melt(paradigm_sample[,c(1,3:441)],id.vars=c("sid1a"),na.rm=T)
paradigm_sample_melt$sid1a <- as.character(paradigm_sample_melt$sid1a)
paradigm_sample_melt$variable <- as.character(paradigm_sample_melt$variable)
paradigm_sample_melt$visit <- 5
paradigm_sample_melt$form <- "sample"

paradigm_long <- paradigm_sample[,c(1,442:881)]
paradigm_long_melt <- melt(paradigm_long,id.vars="sid1a",na.rm=T)
paradigm_long_melt$variable1 <- str_select(paradigm_long_melt$variable,before="_")
paradigm_long_melt$visit <- str_select(paradigm_long_melt$variable,after="_")
paradigm_long_melt$variable <- paradigm_long_melt$variable1

paradigm_long_melt$value <- as.numeric(paradigm_long_melt$value)

paradigm_long_melt$value[paradigm_long_melt$variable=="hct"] <- 
  paradigm_long_melt$value[paradigm_long_melt$variable=="hct"] * 100


paradigm_long_melt$value[paradigm_long_melt$variable=="crea"] <- 
  round(paradigm_long_melt$value[paradigm_long_melt$variable=="crea"]/88.42,2)


paradigm_long_melt$value[paradigm_long_melt$variable=="sglucf"] <- 
  round(paradigm_long_melt$value[paradigm_long_melt$variable=="sglucf"] * 18,0)

paradigm_long_melt$value[paradigm_long_melt$variable=="pglucf"] <- 
  round(paradigm_long_melt$value[paradigm_long_melt$variable=="pglucf"] * 18,0)

paradigm_neu_pct <- subset(paradigm_long_melt,variable=="neu")[,c('sid1a','value','visit')]
names(paradigm_neu_pct)[2] <- "neu"
paradigm_lym_pct <- subset(paradigm_long_melt,variable=="lym")[,c('sid1a','value','visit')]
names(paradigm_lym_pct)[2] <- "lym"
paradigm_wbc <- subset(paradigm_long_melt,variable=="wbc")[,c('sid1a','value','visit')]
names(paradigm_wbc)[2] <- "wbc"

paradigm_abs_leuk_counts <- merge(paradigm_wbc,paradigm_neu_pct,by=c('sid1a','visit'))
paradigm_abs_leuk_counts <- merge(paradigm_abs_leuk_counts,paradigm_lym_pct,by=c('sid1a','visit'))

paradigm_abs_leuk_counts$alc <- round(paradigm_abs_leuk_counts$wbc*paradigm_abs_leuk_counts$lym/100,1)
paradigm_abs_leuk_counts$anc <- round(paradigm_abs_leuk_counts$wbc*paradigm_abs_leuk_counts$neu/100,1)

paradigm_abs_counts <- melt(paradigm_abs_leuk_counts[,c("sid1a","visit","alc","anc")],
                                                     id.vars=c("sid1a","visit"),
                            na.rm=T)

paradigm_long_melt <- rbind(paradigm_long_melt[,c("sid1a","visit","variable","value")],
                            paradigm_abs_counts[,c("sid1a","visit","variable","value")])

## Start labs at randomization rather than baseline for convenience
## PARADIGM-HF sample dataset appears to include labs up to 1 year/12 months.

paradigm_long_melt <- subset(paradigm_long_melt,visit %in% c(5,7,9,10,11))
paradigm_long_melt$form <- "long"


paradigm_pglucf <- subset(paradigm_long_melt,variable=="pglucf")[,c("sid1a","visit","value")]
paradigm_sglucf <- subset(paradigm_long_melt,variable=="sglucf")[,c("sid1a","visit","value")]

names(paradigm_pglucf)[3] <- "pglucf"
names(paradigm_sglucf)[3] <- "sglucf"

paradigm_glu <- merge(paradigm_pglucf,
                      paradigm_sglucf,
                      by=c("sid1a","visit"),
                      all=T)

paradigm_glu$value <- paradigm_glu$pglucf
paradigm_glu$value[!is.na(paradigm_glu$sglucf)] <- paradigm_glu$sglucf[!is.na(paradigm_glu$sglucf)]
paradigm_glu$variable <- "glucf"
paradigm_glu$form <- "long"

paradigm_melt <- rbind(paradigm_sample_melt[,c("sid1a","visit","variable","value","form")],
                  paradigm_long_melt[,c("sid1a","visit","variable","value","form")],
                  paradigm_glu[,c("sid1a","visit","variable","value","form")])

paradigm_melt <- subset(paradigm_melt, !value %in% c("NaN", NA, ""))

colnames(paradigm_melt)[1:2] <-c("patientid","studyvisit")

paradigm_melt$study <- "PARADIGM"


########################### **********CORONA********** #################################

# % ..%%%%%%...%%%%%%%..%%%%%%%%...%%%%%%%..%%....%%....%%%...
# % .%%....%%.%%.....%%.%%.....%%.%%.....%%.%%%...%%...%%.%%..
# % .%%.......%%.....%%.%%.....%%.%%.....%%.%%%%..%%..%%...%%.
# % .%%.......%%.....%%.%%%%%%%%..%%.....%%.%%.%%.%%.%%.....%%
# % .%%.......%%.....%%.%%...%%...%%.....%%.%%..%%%%.%%%%%%%%%
# % .%%....%%.%%.....%%.%%....%%..%%.....%%.%%...%%%.%%.....%%
# % ..%%%%%%...%%%%%%%..%%.....%%..%%%%%%%..%%....%%.%%.....%%


corona_sample <- read.csv('~/Dropbox/BioLINCC files/CORONA/CORONA sample.csv', 
                     na.strings=c(".N","","NA","NULL"),stringsAsFactors = F)

corona_melt <- melt(corona_sample,id.vars=c("usubjid"),na.rm=T)

corona_melt <- subset(corona_melt, !value %in% c("NaN", NA, ""))

corona_melt$form <- "analysis"
corona_melt$studyvisit <- "BASE"
names(corona_melt)[names(corona_melt)=="usubjid"] <- c("patientid")

corona_melt$study <- "CORONA"


############################### **********I-PRESERVE********** ###############################

# % .%%%%.........%%%%%%%%..%%%%%%%%..%%%%%%%%..%%%%%%..%%%%%%%%.%%%%%%%%..%%.....%%.%%%%%%%%
# % ..%%..........%%.....%%.%%.....%%.%%.......%%....%%.%%.......%%.....%%.%%.....%%.%%......
# % ..%%..........%%.....%%.%%.....%%.%%.......%%.......%%.......%%.....%%.%%.....%%.%%......
# % ..%%..%%%%%%%.%%%%%%%%..%%%%%%%%..%%%%%%....%%%%%%..%%%%%%...%%%%%%%%..%%.....%%.%%%%%%..
# % ..%%..........%%........%%...%%...%%.............%%.%%.......%%...%%....%%...%%..%%......
# % ..%%..........%%........%%....%%..%%.......%%....%%.%%.......%%....%%....%%.%%...%%......
# % .%%%%.........%%........%%.....%%.%%%%%%%%..%%%%%%..%%%%%%%%.%%.....%%....%%%....%%%%%%%


ipreserve_analysis <- read.csv('~/Dropbox/BioLINCC files/IPRESERVE/ip_rawdata.csv')


ipreserve_analysis$pupr <- ipreserve_analysis$sbp0-ipreserve_analysis$dbp0

ipreserve_analysis$LVEDD_index <- ipreserve_analysis$lvdidm/ipreserve_analysis$bsa
ipreserve_analysis$LVESD_index <- ipreserve_analysis$lvsydm/ipreserve_analysis$bsa
ipreserve_analysis$LVEDV_index <- ipreserve_analysis$lvedvl/ipreserve_analysis$bsa


ipreserve_analysis <- calc_hypertrophy_type(df=ipreserve_analysis,
                                            male="M",
                                            female="F",
                                            lvedd_cm = "lvdidm",
                                            ivsd_cm = "slwltk",
                                            lvpwtd_cm = "lvpwdi",
                                            height_cm = "entryht",
                                            weight_kg = "entrywt")


ipreserve_analysis <- calc_lav(dat=ipreserve_analysis,
                     laa_4c="laesaf", 
                     laa_2c="laesat", 
                     lal_4c="lasfdm", 
                     lal_2c="lasfdm", 
                     height_cm="entryht", 
                     weight_kg="entrywt")
  

ipreserve_analysis$eeprime_avg <- mean(c(ipreserve_analysis$elrt,ipreserve_analysis$esrt),na.rm=T)

ipreserve_analysis$RVSP_new <- (ipreserve_analysis$jvp)*5 +
                            4*(ipreserve_analysis$tcpkvo/100)^2


ipreserve_analysis$qrs_ecg_ms[ipreserve_analysis$qrs_ecg < 0.3&!is.na(ipreserve_analysis$qrs_ecg)] <- 
  ipreserve_analysis$qrs_ecg[ipreserve_analysis$qrs_ecg < 0.3&!is.na(ipreserve_analysis$qrs_ecg)]*1000

ipreserve_analysis$qrs_ecg_ms[ipreserve_analysis$qrs_ecg > 0.3&!is.na(ipreserve_analysis$qrs_ecg)] <- 
  ipreserve_analysis$qrs_ecg[ipreserve_analysis$qrs_ecg > 0.3&!is.na(ipreserve_analysis$qrs_ecg)]*100

names(ipreserve_analysis)[1] <- "patientid"
ipreserve_analysis$VISIT <- 0

ip <- 
  assign_diastolic_function(dat=ipreserve_analysis,
                            pid="patientid",
                            vst="VISIT",
                            ep_lat="ltmevo",
                            ep_sept="slmevo",
                            earat="ekrt",
                            e_vel="mipkvo",
                            dt="mddu",
                            def="TOPCAT",
                            verbose=F) 

ipreserve_analysis <- 
  merge(ipreserve_analysis,
        ip,
        by=c("patientid","VISIT"),
        all.x=T)

ipreserve_melt <- melt(ipreserve_analysis,id.vars=c("patientid"),na.rm=T)

ipreserve_melt <- subset(ipreserve_melt, !value %in% c("NaN", NA,""))

ipreserve_melt$studyvisit <- "BASE"
ipreserve_melt$form <- "analysis"
ipreserve_melt$study <- "IPRESERVE"

names(ipreserve_melt)[1] <- "patientid"


################  **********SOLVD**********  ####################      

# % ..%%%%%%...%%%%%%%..%%.......%%.....%%.%%%%%%%%.
# % .%%....%%.%%.....%%.%%.......%%.....%%.%%.....%%
# % .%%.......%%.....%%.%%.......%%.....%%.%%.....%%
# % ..%%%%%%..%%.....%%.%%.......%%.....%%.%%.....%%
# % .......%%.%%.....%%.%%........%%...%%..%%.....%%
# % .%%....%%.%%.....%%.%%.........%%.%%...%%.....%%
# % ..%%%%%%...%%%%%%%..%%%%%%%%....%%%....%%%%%%%%.

solvd_sbf <- read.sas7bdat(paste(solvd_folder,"sbf_lad2.sas7bdat",sep=""))
solvd_sbf[solvd_sbf==""|solvd_sbf=="NaN"] <- NA
solvd_sef <- read.sas7bdat(paste(solvd_folder,"sef_lad2.sas7bdat",sep=""))
solvd_sef[solvd_sef == ""|solvd_sef == "NaN"] <- NA
solvd_sfe <- read.sas7bdat(paste(solvd_folder,"sfe_lad2.sas7bdat",sep=""))
solvd_sfe[solvd_sfe == ""|solvd_sfe == "NaN"] <- NA
solvd_tx <- read.sas7bdat(paste(solvd_folder,"sep_lad2.sas7bdat",sep=""))[,c("ID_SOL","TRTMENT")]
solvd_tx[solvd_tx == ""|solvd_tx == "NaN"] <- NA
solvd_sql <- read.sas7bdat(paste(solvd_folder,"sql_lad2.sas7bdat",sep=""))
solvd_sql[solvd_sql == ""|solvd_sql=="NaN"] <- NA


#### SBF (Baseline form) ####


sbf_strings <- names(solvd_sbf)[!is.na(str_match(names(solvd_sbf),"SBF"))]
sbf_strings <- c("ID_SOL",
                 "FORMCODE",
                 "VISIT",
                 "TRIAL",
                 sbf_strings)
solvd_sbf <- solvd_sbf[,sbf_strings]

solvd_sbf$SBF33Z2[is.na(solvd_sbf$SBF33Z2)] <- solvd_sbf$SBF33Z1[is.na(solvd_sbf$SBF33Z2)] * 0.453592
solvd_sbf$pupr <- solvd_sbf$SBF35Z1-solvd_sbf$SBF35Z2

solvd_sbf_melt <- melt(solvd_sbf,
                       id.vars=c("ID_SOL",
                                 "FORMCODE",
                                 "VISIT",
                                 "TRIAL"),
                       na.rm=T)

solvd_sbf <- merge(solvd_sbf,
                   solvd_tx,
                   by="ID_SOL")

solvd_sbf$qrs_120[!is.na(solvd_sbf$SBF28C)|!is.na(solvd_sbf$SBF30) ] <- "N"
solvd_sbf$qrs_120[solvd_sbf$SBF28C=="N"|solvd_sbf$SBF30=="Y"] <- "Y"


solvd_sbf_melt <- melt(solvd_sbf,
                       id.vars=c("ID_SOL",
                                 "FORMCODE",
                                 "VISIT",
                                 "TRIAL"),
                       na.rm=T)


#### SQL (quality of life) ####

# Per manuscript PMID: 8475871 , supposedly MLHF is represented, but I cannot find all 23 questions.
# Also included are Rand Mental Health Inventory, Functional Status Scale, Dyspnea Scale, Current Health Perceptions
# Perceived Social Support, Vocabulary, Digit Span (attention).  References are all in the PMID above.
# Although the SQL form is included, it doesn't seem that all of these scales are complete.  However there may
# be additional datasets missing.

# I have not yet tried to recreate official QOL scores, but this would be nice in the future.

sql_strings <- names(solvd_sql)[!is.na(str_match(names(solvd_sql),"SQL"))]
sql_strings <- c("ID_SOL",
                 "FORMCODE",
                 "VISIT",
                 "TRIAL",
                 sql_strings)
solvd_sql <- solvd_sql[,sql_strings]

solvd_sql$vas_global <- solvd_sql$SQL35A * 10
solvd_sql$poms <- sum(solvd_sql[,14:45],na.rm=T)

solvd_sql$poms <- apply(solvd_sql[,14:45],MARGIN=1,FUN=function (x) sum(x,na.rm=T))

solvd_sql_melt <- melt(solvd_sql,
                       id.vars=c("ID_SOL",
                                 "FORMCODE",
                                 "VISIT",
                                 "TRIAL"),
                       na.rm=T)


#### SEF (Eligibility visit) ####


solvd_sef$CrCl <- calc_MDRD4(dat=solvd_sef,
                             age="SEF_AGE",
                             cr="SEF26",
                             race="SEF10",
                             sex="SEF9",
                             male="M",
                             black="B")

solvd_sef$alc <- solvd_sef$SEF22Z3/100*
  solvd_sef$SEF22Z1


solvd_sef$anc <- solvd_sef$SEF22Z2/100*
  solvd_sef$SEF22Z1


sef_strings <- names(solvd_sef)[!is.na(str_match(names(solvd_sef),"SEF"))]
sef_strings <- c("ID_SOL",
                 "FORMCODE",
                 "VISIT",
                 "TRIAL",
                 sef_strings,
                 "CrCl",
                 "alc",
                 "anc")
solvd_sef <- solvd_sef[,sef_strings]

solvd_sef_melt <- melt(solvd_sef,
                       id.vars=c("ID_SOL",
                                 "FORMCODE",
                                 "VISIT",
                                 "TRIAL"),
                       na.rm=T)


#### SFE (Follow-up interview-exam form) ####

sfe_strings <- names(solvd_sfe)[!is.na(str_match(names(solvd_sfe),"SFE"))]
sfe_strings <- c("ID_SOL",
                 "FORMCODE",
                 "VISIT",
                 "TRIAL",
                 sfe_strings)
solvd_sfe <- solvd_sfe[,sfe_strings]

solvd_sfe$SFE24Z2[is.na(solvd_sfe$SFE24Z2)] <- solvd_sfe$SFE24Z1[is.na(solvd_sfe$SFE24Z2)] * 0.453592
solvd_sfe$pupr <- solvd_sfe$SFE26Z1 - solvd_sfe$SFE26Z2

solvd_sfe <- merge(solvd_sfe,solvd_sef[,c("ID_SOL",
                                          "SEF_AGE",
                                          "SEF9",
                                          "SEF10",
                                          "SEF26")],
                   all.x=T)

solvd_sfe$visit_age[solvd_sfe$VISIT>7] <- solvd_sfe$SEF_AGE[solvd_sfe$VISIT>7] + (solvd_sfe$VISIT[solvd_sfe$VISIT>7]-7)

solvd_sfe$CrCl <- calc_MDRD4(dat=solvd_sfe,
                             age="visit_age",
                             cr="SFE37",
                             race="SEF10",
                             sex="SEF9",
                             male="M",
                             black="B")


solvd_sfe$anc <- solvd_sfe$SFE33Z2/100*
                  solvd_sfe$SFE33Z1


solvd_sfe$alc <- solvd_sfe$SFE33Z3/100*
  solvd_sfe$SFE33Z1

solvd_sfe_melt <- melt(solvd_sfe,
                       id.vars=c("ID_SOL",
                                 "FORMCODE",
                                 "VISIT",
                                 "TRIAL"),
                       na.rm=T)


##########  compile  ##########


solvd_melt <- rbind(solvd_sbf_melt,
                     solvd_sef_melt,
                     solvd_sql_melt,
                     solvd_sfe_melt)

solvd_melt <- subset(solvd_melt, !value %in% c("NaN",NA,""))

names(solvd_melt)[c(1:3)] <- c("patientid","form","studyvisit")

solvd_melt$study[solvd_melt$TRIAL=="P"] <- "SOLVD Prevent"
solvd_melt$study[solvd_melt$TRIAL=="T"] <- "SOLVD Treat"


##############  **********SOLVD-Registry**********  ####################      

# % ..%%%%%%...%%%%%%%..%%.......%%.....%%.%%%%%%%%.....%%%%%%%%..%%%%%%%%..%%%%%%...%%%%..%%%%%%..%%%%%%%%.%%%%%%%%..%%....%%
# % .%%....%%.%%.....%%.%%.......%%.....%%.%%.....%%....%%.....%%.%%.......%%....%%...%%..%%....%%....%%....%%.....%%..%%..%%.
# % .%%.......%%.....%%.%%.......%%.....%%.%%.....%%....%%.....%%.%%.......%%.........%%..%%..........%%....%%.....%%...%%%%..
# % ..%%%%%%..%%.....%%.%%.......%%.....%%.%%.....%%....%%%%%%%%..%%%%%%...%%...%%%%..%%...%%%%%%.....%%....%%%%%%%%.....%%...
# % .......%%.%%.....%%.%%........%%...%%..%%.....%%....%%...%%...%%.......%%....%%...%%........%%....%%....%%...%%......%%...
# % .%%....%%.%%.....%%.%%.........%%.%%...%%.....%%....%%....%%..%%.......%%....%%...%%..%%....%%....%%....%%....%%.....%%...
# % ..%%%%%%...%%%%%%%..%%%%%%%%....%%%....%%%%%%%%.....%%.....%%.%%%%%%%%..%%%%%%...%%%%..%%%%%%.....%%....%%.....%%....%%...


solvd_rbf <- read.sas7bdat(paste(solvdreg_folder,"rbf_reg.sas7bdat",sep=""))
solvd_reg_tx <- read.sas7bdat(paste(solvd_folder,"sep_lad2.sas7bdat",sep=""))[,c("ID_SOL","TRTMENT")]
solvd_rbf <- merge(solvd_rbf,solvd_reg_tx,all.x=T,by="ID_SOL")

solvd_rec <- read.sas7bdat(paste(solvdreg_folder,"rec_reg.sas7bdat",sep=""))
solvd_rsb <- read.sas7bdat(paste(solvdreg_folder,"rsb_reg.sas7bdat",sep=""))


#### RBF (Registry baseline form) ####

solvd_rbf[solvd_rbf=="NaN"|solvd_rbf==""] <- NA
rbf_strings <- names(solvd_rbf)[!is.na(str_match(names(solvd_rbf),"RBF"))]
rbf_strings <- c("id_reg",
                 "FORMCODE",
                 "VISIT",
                 "age",
                 "weight",
                 "height",
                 "bmi",
                 "TRTMENT",
                 rbf_strings)
solvd_rbf <- solvd_rbf[,rbf_strings]

solvd_rbf$CrCl <- calc_MDRD4(dat=solvd_rbf,
                             cr="RBF60",
                             age="age",
                             race="RBF7",
                             sex="RBF6",
                             black=3,
                             male="M")

solvd_rbf$pupr <- solvd_rbf$RBF41-solvd_rbf$RBF42

solvd_rbf_melt <- melt(data=solvd_rbf,id.vars=c("id_reg","VISIT","FORMCODE"),na.rm=T) 


#### RSB (Registry substudy baseline form) ####

solvd_rsb[solvd_rsb=="NaN"|solvd_rsb==""] <- NA
rsb_strings <- names(solvd_rsb)[!is.na(str_match(names(solvd_rsb),"RSB"))]
rsb_strings <- c("id_reg",
                 "FORMCODE",
                 "VISIT",
                 "TIME",
                 "DISTANCE",
                 "age",
                 "weight",
                 "height",
                 "bmi",
                 rsb_strings)
solvd_rsb <- solvd_rsb[,rsb_strings]

solvd_rsb$bsa <- 0.20247*solvd_rsb$weight^0.425*(solvd_rsb$height/100)^0.725

solvd_rsb_melt <- melt(data=solvd_rsb,id.vars=c("id_reg","VISIT","FORMCODE"),na.rm=T) 

####  REC (Registry echo form)  ####

solvd_rec[solvd_rec=="NaN"|solvd_rec==""] <- NA

solvd_rec <- merge(solvd_rec,solvd_rsb[,c("id_reg","height","weight")], all.x=T)
solvd_rec <- merge(solvd_rec,solvd_rbf[,c("id_reg","RBF6")], all.x=T)

solvd_rec <- calc_hypertrophy_type(df=solvd_rec,
                      sex="RBF6",
                      male="M",
                      female="F",
                      lvedd_cm="REC4",
                      ivsd_cm="REC6",
                      lvpwtd_cm="REC7",
                      height_cm="height",
                      weight_kg="weight")

solvd_rec$LVEDD_index <- solvd_rec$REC4/solvd_rec$bsa
solvd_rec$LVESD_index <- solvd_rec$REC5/solvd_rec$bsa
solvd_rec$LVEDV_index <- solvd_rec$REC10/solvd_rec$bsa

solvd_rec_melt <- melt(solvd_rec,id.vars=c("id_reg","VISIT","FORMCODE"),na.rm=T)


##########  compile  ##########

solvd_reg_melt <- rbind(solvd_rbf_melt,
                  solvd_rsb_melt,
                  solvd_rec_melt)

names(solvd_reg_melt)[c(1:3)] <- c("patientid",
                             "studyvisit",
                             "form")

solvd_reg_melt <- subset(solvd_reg_melt, !value %in% c("NaN", NA,""))

solvd_reg_melt$study <- "SOLVD Registry"


  
################  **********RELAX********** ############################

# .%%%%%%%%..%%%%%%%%.%%..........%%%....%%.....%%
# .%%.....%%.%%.......%%.........%%.%%....%%...%%.
# .%%.....%%.%%.......%%........%%...%%....%%.%%..
# .%%%%%%%%..%%%%%%...%%.......%%.....%%....%%%...
# .%%...%%...%%.......%%.......%%%%%%%%%...%%.%%..
# .%%....%%..%%.......%%.......%%.....%%..%%...%%.
# .%%.....%%.%%%%%%%%.%%%%%%%%.%%.....%%.%%.....%%


####  a_base  ####

relax_a_base <- read.sas7bdat(paste(relax_folder,"a_base.sas7bdat",sep=""))

relax_a_base$yihf_months <- relax_a_base$YIHF*12

relax_a_base_melt <- 
  melt(data=relax_a_base,
       id.vars=c("PATNUMB"),
       na.rm=T) 

relax_a_base_melt$studyvisit <- "BASELINE"
colnames(relax_a_base_melt)[1] <- "studyid"
relax_a_base_melt$form <- "a_base"


####  cpx  ####

relax_cpx <- read.sas7bdat(paste(relax_folder,"cpx.sas7bdat",sep=""))
relax_cpx[relax_cpx=="NaN"] <- NA

relax_cpx_melt <- 
  subset(melt(data=relax_cpx,id.vars=c("patnumb","TEST"),na.rm=T))
relax_cpx_melt$studyvisit <- relax_cpx_melt$TEST
colnames(relax_cpx_melt)[1] <- "studyid"
relax_cpx_melt$form <- "cpx"

####  a_visitsumm  ####

relax_visitsumm <- read.sas7bdat(paste(relax_folder,"a_visitsumm.sas7bdat",sep=""))
relax_visitsumm[relax_visitsumm=="NaN"] <- NA

relax_visitsumm$weight_kg <- round(relax_visitsumm$WTLBS/2.2,1)
relax_visitsumm$height_cm <- round(relax_visitsumm$HEIGHTIN*2.54,1)

relax_visitsumm$hct_calc <- round(relax_visitsumm$LL_HMG/0.34,1)
relax_visitsumm$bsa <- round(0.20247*(relax_visitsumm$WTLBS/2.2)^0.425*(relax_visitsumm$HEIGHTIN*2.54/100)^0.725,2)
relax_visitsumm$bmi <- round(703*relax_visitsumm$WTLBS/relax_visitsumm$HEIGHTIN^2,1)

relax_visitsumm$alc <- relax_visitsumm$LL_LYMPH/100*relax_visitsumm$LL_WBC

relax_visitsumm$CL_TROPI_ngml <- relax_visitsumm$CL_TROPI/1000

relax_visitsumm_melt <- subset(melt(data=relax_visitsumm,
                                      id.vars=c("PATNUMB","FORM"),
                                      na.rm=T))

relax_visitsumm_melt$studyvisit <- relax_visitsumm_melt$FORM
colnames(relax_visitsumm_melt)[1] <- "studyid"
relax_visitsumm_melt$form <- "a_visitsumm"


#### echo ####

relax_echo <- read.sas7bdat(paste(relax_folder,"echo.sas7bdat",sep=""))
relax_echo[relax_echo=="NaN"] <- NA

relax_echo <- merge(relax_echo,relax_a_base[,c("PATNUMB","SEX")],by.x="patnumb",by.y="PATNUMB", all.x=T)
relax_echo <- merge(relax_echo,
                    relax_visitsumm[relax_visitsumm$FORM=="BASELINE",
                                    c("PATNUMB","bsa")],
                    by.x="patnumb",
                    by.y="PATNUMB",
                    all.x=T)


relax_echo$earatio <- relax_echo$EVELOC/relax_echo$AVELOC
relax_echo$DTILATE_cmsec <- relax_echo$DTILATE*100
relax_echo$DTIMEDE_cmsec <- relax_echo$DTIMEDE*100
relax_echo$EVELOC_cmsec <- relax_echo$EVELOC*100
relax_echo$AVELOC_cmsec <- relax_echo$AVELOC*100
relax_echo$LVED_best <- relax_echo$LVEDCH
relax_echo$LVED_best[is.na(relax_echo$LVED_best)] <- relax_echo$LVEDVOL[is.na(relax_echo$LVED_best)]

relax_echo$LVEDD_index <- relax_echo$LVDD/relax_echo$bsa
relax_echo$LVEDV_index <- relax_echo$LVED_best/relax_echo$bsa
relax_echo$LAV_ix_calc <- relax_echo$LAVOLUME/relax_echo$bsa

relax_echo$eeprime_lat <- round(relax_echo$EVELOC/relax_echo$DTILATE,1)
relax_echo$eeprime_sept <- round(relax_echo$EVELOC/relax_echo$DTIMEDE,1)
relax_echo$eeprime_avg <- round((relax_echo$eeprime_lat+relax_echo$eeprime_sept)/2,1)

relax_echo$pvf_ratio <- relax_echo$SYSFWRD/relax_echo$DIAFWRD

## Note that RELAX hypertrophy type cannot be determined due to fact that RELAX investigators inexplicably 
## did not include echo-determined septal or posterior wall thickness in the dataset despite it being
## present in the data documentation and listed formula for determining LV mass (also not included in echo data)
## Verified  in dataset downloaded June 24, 2022.

## The point is somewhat moot as most patients who had cardiac MRI end up not meeting criteria for LVH.  
## However, remodeling type (normal vs. concentric remodeling") cannt be determined

relax_echo$eprime_abnl[relax_echo[,"DTIMEDE_cmsec"] < 8|relax_echo[,"DTILATE_cmsec"]<10] <- 1
relax_echo$eprime_abnl[relax_echo[,"DTIMEDE_cmsec"] >= 8|relax_echo[,"DTILATE_cmsec"] >= 10] <- 0


relax_echo$patientid <- relax_echo$patnumb

rech  <-  
  assign_diastolic_function(
    dat=relax_echo,
    pid="patnumb",
    vst="SCHDTIME",
    ep_lat="DTILATE_cmsec",
    ep_sept="DTIMEDE_cmsec",
    earat="earatio",
    e_vel="EVELOC_cmsec",
    dt="DECELTM",
    def="TOPCAT",
    verbose=F)

relax_echo <-
  merge(relax_echo,
        rech,
        by=c("patnumb","SCHDTIME"))

relax_echo_melt <- subset(melt(data=relax_echo,
                        id.vars=c("patnumb","SCHDTIME"),
                        na.rm=T))

relax_echo_melt$studyvisit <- relax_echo_melt$SCHDTIME
colnames(relax_echo_melt)[1] <- "studyid"
relax_echo_melt$form <- "echo"


####  mri  ####

relax_mri <- read.sas7bdat(paste(relax_folder,"mri.sas7bdat",sep=""))


relax_mri <- merge(relax_mri, 
                   relax_visitsumm[relax_visitsumm$FORM=="BASELINE",c("PATNUMB","bsa")],
                   by.x=c("patnumb"),
                   by.y=c('PATNUMB'),
                   all.x=T)

relax_mri$lvmass_ix <- round(relax_mri$MRILVMAS/relax_mri$bsa,1)


relax_mri <- 
  calc_pulsatility(dat=relax_mri,
                   sbp = "MRISBP",
                   dbp = "MRIDBP",
                   edv = "MRIEDV",
                   esv = "MRIESV",
                   bsa = "bsa",
                   hr = "MRIHR")

relax_mri_melt <- subset(melt(data=relax_mri,
              id.vars=c("patnumb",
                        "MRIVIS"),
              na.rm=T))

relax_mri_melt$studyvisit <- relax_mri_melt$MRIVIS
colnames(relax_mri_melt)[1] <- "studyid"
relax_mri_melt$form <- "mri"


####  medhist1  ####

## Add zeros for types of ischemic heart disease and nonischemic cardiomyopathy

relax_medhist1 <- read.sas7bdat(paste(relax_folder,"medhist1.sas7bdat",sep=""))
relax_medhist1[relax_medhist1=="NaN"] <- NA

relax_cms <- names(relax_medhist1) %in% c("ALCOHOLC",
                                        "CYTOTXC",
                                        "FAMILIAL",
                                        "HCM",
                                        "HYPERTEN",
                                        "DILATED",
                                        "RESTRICT",
                                        "OTHCONT",
                                        "PERIPAR",
                                        "VAL")


relax_medhist1[relax_medhist1$ISCHEMIC==1,relax_cms] <- NA


relax_medhist1$num_etiol <- apply(relax_medhist1[relax_cms],
                                 MARGIN=1,
                                 FUN=function(x)
                                   sum(x,na.rm=T))

relax_medhist1[relax_medhist1$num_etiol>1,relax_cms] <- NA


relax_medhist1_melt <- 
  subset(melt(data=relax_medhist1,id.vars=c("patnumb",
                                            "FORM"),
              na.rm=T))

names(relax_medhist1_melt)[1] <- "studyid"
relax_medhist1_melt$studyvisit <- relax_medhist1_melt$FORM
relax_medhist1_melt$form <- "medhist1"


####  medhist2  #####

relax_medhist2 <- read.sas7bdat(paste(relax_folder,"medhist2.sas7bdat",sep=""))
relax_medhist2[relax_medhist2=="NaN"] <- NA


make_zeros <- c("MSTENOS",
                "MREGURG",
                "ATSTENOS",
                "AREGURG",
                "TSTENOS",
                "TREGURG",
                "NONSURG",
                "MITSURG",
                "AORSURG",
                "TRISURG",
                "PULSURG",
                "ATRIALFB",
                "FIBFLUTR",
                "SUSVTVF",
                "ARREST",
                "PACETYPE",
                "ICDTYPE",
                "DIABTYPE")


for (i in 1:length(make_zeros)) {
  thiscol <- make_zeros[i]
  print(thiscol)
  relax_medhist2[is.na(relax_medhist2[,thiscol]),thiscol] <- 0
}

relax_medhist2_melt <- subset(melt(data=relax_medhist2,
                        id.vars=c("patnumb","FORM"),
                        na.rm=T))

relax_medhist2_melt$studyvisit <- relax_medhist2_melt$FORM
colnames(relax_medhist2_melt)[1] <- "studyid"
relax_medhist2_melt$form <- "medhist2"


#### assessmt ####

relax_assessmt <- read.sas7bdat(paste(relax_folder,"assessmt.sas7bdat",sep=""))
relax_assessmt[relax_assessmt=="NaN"] <- NA

relax_assessmt$pupr <- relax_assessmt$BPSYS-relax_assessmt$BPDIA

relax_assessmt_melt <- subset(melt(data=relax_assessmt,
                          id.vars=c("patnumb","FORM"),
                          na.rm=T))

colnames(relax_assessmt_melt)[1] <- "studyid"
relax_assessmt_melt$form <- "assessmt"
relax_assessmt_melt$studyvisit <- relax_assessmt_melt$FORM


#### walktest ####

relax_walktest <- read.sas7bdat(paste(relax_folder,"walktest.sas7bdat",sep=""))
relax_walktest[relax_walktest=="NaN"] <- NA

relax_walktest$time_walked <- round(relax_walktest$WLKMIN + relax_walktest$WLKSEC/60,2)


relax_walktest_melt <- subset(melt(data=relax_walktest,
                                   id.vars=c("patnumb","FORM"),
                                   na.rm=T))

colnames(relax_walktest_melt)[1] <- "studyid"
relax_walktest_melt$form <- "walktest"
relax_walktest_melt$studyvisit <- relax_walktest_melt$FORM


####  ecg  ####

relax_ecg <- read.sas7bdat(paste(relax_folder,"ecg.sas7bdat",sep=""))
relax_ecg[relax_ecg=="NaN"] <- NA

relax_ecg_melt <- subset(melt(data=relax_ecg,
                                   id.vars=c("patnumb","FORM"),
                                   na.rm=T))
relax_ecg_melt$form <- "ecg"
colnames(relax_ecg_melt)[1] <- "studyid"
relax_ecg_melt$studyvisit <- relax_ecg_melt$FORM


####  meds  ####

relax_meds  <- read.sas7bdat(paste(relax_folder,"meds.sas7bdat",sep=""))
relax_meds[relax_meds=="NaN"] <- NA
relax_medmap  <- read.csv(paste(relax_folder,"medmap.csv",sep=""),
                          stringsAsFactors = F)
relax_meds <- merge(relax_meds,relax_medmap)

relax_meds$TAKING[relax_meds$FORM=="BASELINE"] <- relax_meds$MEDRAND[relax_meds$FORM=="BASELINE"] 
relax_meds$TAKING[!relax_meds$FORM=="BASELINE"] <- relax_meds$MEDSANS[!relax_meds$FORM=="BASELINE"] 


names(relax_meds)[names(relax_meds)=="FORM"] <- "studyvisit"
names(relax_meds)[names(relax_meds)=="patnumb"] <- "studyid"
names(relax_meds)[names(relax_meds)=="brief_name"] <- "variable"
names(relax_meds)[names(relax_meds)=="TAKING"] <- "value"
relax_meds$study <- "RELAX"
relax_meds$form <- "meds"

relax_meds$studyvisit <- as.character(relax_meds$studyvisit)
relax_meds$studyid <- as.character(relax_meds$studyid)


####  diuretic  ####


relax_diuretic  <- read.sas7bdat(paste(relax_folder,"diuretic.sas7bdat",sep=""))
relax_diuretic[relax_diuretic=="NaN"] <- NA


relax_diuretic$brief_name[relax_diuretic$DIUMEDS==1] <- "furosemide"
relax_diuretic$brief_name[relax_diuretic$DIUMEDS==2] <- "torsemide"
relax_diuretic$brief_name[relax_diuretic$DIUMEDS==3] <- "bumetanide"
relax_diuretic$brief_name[relax_diuretic$DIUMEDS==4] <- "metolazone"
relax_diuretic$brief_name[relax_diuretic$DIUMEDS==5] <- "hctz"

relax_diuretic$variable <- relax_diuretic$brief_name
relax_diuretic$value <- relax_diuretic$DIURANS

relax_diuretic$furo_equiv[relax_diuretic$brief_name=="furosemide"] <- relax_diuretic$DIURDOSE[relax_diuretic$brief_name=="furosemide"]
relax_diuretic$furo_equiv[relax_diuretic$brief_name=="torsemide"] <- relax_diuretic$DIURDOSE[relax_diuretic$brief_name=="torsemide"]*2
relax_diuretic$furo_equiv[relax_diuretic$brief_name=="bumetanide"] <- relax_diuretic$DIURDOSE[relax_diuretic$brief_name=="bumetanide"]*40

names(relax_diuretic)[4] <- "studyvisit"

relax_diuretic$form <- "diuretic"

relax_diuretic_melt <- sqldf("select patnumb as studyid,
                             studyvisit,
                             brief_name as variable,
                             DIURANS as value,
                             'diuretic' as form,
                             'RELAX' as study
                             from relax_diuretic")

relax_furo_equiv <- sqldf("select patnumb as studyid,
                                  studyvisit,
                                  'furo_equiv' as form,
                                  'daily_furo_equiv_derive' as variable,
                                  sum(furo_equiv) as value,
                                  'RELAX' as study
                          from relax_diuretic 
                          group by patnumb,FORM")


###### MLHFQ #####

relax_mlhfq <- read.sas7bdat(paste(relax_folder,"mlhfq.sas7bdat",sep=""))
relax_mlhfq[relax_mlhfq=="NaN"] <- NA
relax_mlhfq$MLWHF <- paste("MLHFQ",relax_mlhfq$MLWHF,sep="")
relax_mlhf <- as.data.frame(acast(relax_mlhfq,patnumb+FORM~MLWHF,value.var="MLWHFANS"))

relax_mlhf$patientid <- strtrim(rownames(relax_mlhf),6)
relax_mlhf$studyvisit[str_detect(rownames(relax_mlhf),"BASELINE")==T] <- "BASELINE"
relax_mlhf$studyvisit[str_detect(rownames(relax_mlhf),"WEEK12")==T] <- "WEEK12"
relax_mlhf$studyvisit[str_detect(rownames(relax_mlhf),"WEEK24")==T] <- "WEEK24"

relax_mlhf <- score_mlhf(dat=relax_mlhf,
                         swelling = "MLHFQ1", 
                         day_rest="MLHFQ2",
                         stairs = "MLHFQ3", 
                         housework="MLHFQ4",
                         go_places="MLHFQ5",
                         sleep = "MLHFQ6",
                         friendfam = "MLHFQ7",
                         earn_living = "MLHFQ8",
                         hobbies = "MLHFQ9",
                         sex = "MLHFQ10",
                         food = "MLHFQ11",
                         sob = "MLHFQ12",
                         fatigue = "MLHFQ13",
                         hospital = "MLHFQ14",
                         cost = "MLHFQ15",
                         sideeffect = "MLHFQ16",
                         burden = "MLHFQ17",
                         nocontrol = "MLHFQ18",
                         worry="MLHFQ19",
                         concentrate="MLHFQ20",
                         depressed="MLHFQ21", 
                         total=c("Y"))


relax_mlhf_melt <- melt(relax_mlhf,id.vars=c("patientid","studyvisit"))
relax_mlhf_melt$studyid <- relax_mlhf_melt$patientid
relax_mlhf_melt$form <- "mlhfq"


#### compile  ####

relax_melt <- rbind(relax_a_base_melt[,c('studyid','form','studyvisit','variable','value')],
                  relax_medhist1_melt[,c('studyid','form','studyvisit','variable','value')],
                  relax_medhist2_melt[,c('studyid','form','studyvisit','variable','value')],
                  relax_cpx_melt[,c('studyid','form','studyvisit','variable','value')],
                  relax_echo_melt[,c('studyid','form','studyvisit','variable','value')],
                  relax_ecg_melt[,c('studyid','form','studyvisit','variable','value')],
                  relax_visitsumm_melt[,c('studyid','form','studyvisit','variable','value')],
                  relax_assessmt_melt[,c('studyid','form','studyvisit','variable','value')],
                  relax_furo_equiv[,c('studyid','form','studyvisit','variable','value')],
                  relax_diuretic_melt[,c('studyid','form','studyvisit','variable','value')],
                  relax_meds[,c('studyid','form','studyvisit','variable','value')],
                  relax_mlhf_melt[,c('studyid','form','studyvisit','variable','value')],
                  relax_mri_melt[,c('studyid','form','studyvisit','variable','value')],
                  relax_walktest_melt[,c('studyid','form','studyvisit','variable','value')])

relax_melt <- subset(relax_melt, studyvisit %in% c("BASELINE",
                                           "Baseline",
                                           "1",
                                           "10",
                                           "12Weeks",
                                           "WEEK12",
                                           "24Weeks",
                                           "WEEK24"))


relax_melt <- subset(relax_melt,!value %in% c("NaN", NA, ""))

names(relax_melt)[1] <- "patientid"

relax_melt$study <- "RELAX"


########################### **********NEAT-HFpEF********** ##########################

# .%%....%%.%%%%%%%%....%%%....%%%%%%%%.........%%.....%%.%%%%%%%%.%%%%%%%%..%%%%%%%%.%%%%%%%%
# .%%%...%%.%%.........%%.%%......%%............%%.....%%.%%.......%%.....%%.%%.......%%......
# .%%%%..%%.%%........%%...%%.....%%............%%.....%%.%%.......%%.....%%.%%.......%%......
# .%%.%%.%%.%%%%%%...%%.....%%....%%....%%%%%%%.%%%%%%%%%.%%%%%%...%%%%%%%%..%%%%%%...%%%%%%..
# .%%..%%%%.%%.......%%%%%%%%%....%%............%%.....%%.%%.......%%........%%.......%%......
# .%%...%%%.%%.......%%.....%%....%%............%%.....%%.%%.......%%........%%.......%%......
# .%%....%%.%%%%%%%%.%%.....%%....%%............%%.....%%.%%.......%%........%%%%%%%%.%%......


#### a_base ####

neat_base <- read.csv(paste(neat_folder,"a_base.csv",sep=""))

neat_base$CrCl <- calc_MDRD4(dat=neat_base,
                             cr="CREAT_BL",
                             age="AGE",
                             race="RACE",
                             sex="SEX",
                             male=1,
                             black=3)

neat_base$hfage_months <- neat_base$HFAGE

neat_base$RACE[neat_base$ETHNIC==1] <- 8

neat_base_melt <- 
  melt(data=neat_base,
       stringsAsFactors=F,
       na.strings=c("","NA","NaN"),
       id.vars=c("PATNUMB"),
       na.rm=T) 

neat_base_melt$studyvisit <- "BASE"
colnames(neat_base_melt)[1] <- "patnumb"
neat_base_melt$form <- "a_base"


#### a_visitsumm

neat_visitsumm <- read.csv(paste(neat_folder,
                                 "a_visitsumm.csv",
                                 sep=""),
                                 stringsAsFactors=F,
                                 na.strings=c("","NA","NaN"))

neat_visitsumm <- merge(neat_visitsumm,
                        neat_base[,c("PATNUMB",
                                     "HEIGHTIN_BL",
                                     "SEX")],
                        by="PATNUMB",
                        all.x=T)

neat_visitsumm$bmi <- calc_bmi(dat=neat_visitsumm,
                               weight="WTLBS",
                               height="HEIGHTIN_BL",
                               metric=F)

neat_visitsumm$weight_kg <- neat_visitsumm$WTLBS/2.2


neat_visitsumm_melt <- 
  melt(data=neat_visitsumm,
       id.vars=c("PATNUMB","VISIT"),
       na.rm=T) 

colnames(neat_visitsumm_melt)[1] <- "patnumb"
names(neat_visitsumm_melt)[2] <- "studyvisit"
neat_visitsumm_melt$form <- "a_visitsumm"


##########  echo  ##########


neat_echo <- read.csv(paste(neat_folder,"echo.csv",sep=""),
                      stringsAsFactors = F,
                      na.strings=c("NA","NaN","","NULL"))

neat_echo <- merge(neat_echo,
                   neat_visitsumm[neat_visitsumm$VISIT=="BASE",c("PATNUMB",
                                                                 "HEIGHTIN_BL",
                                                                 "WTLBS",
                                                                 "SEX")],
                   by.x="patnumb",
                   by.y="PATNUMB")

neat_echo$weight_kg <- neat_echo$WTLBS/2.2
neat_echo$height_cm <- neat_echo$HEIGHTIN_BL*2.54

neat_echo <- calc_lav(dat=neat_echo,
                      laa_4c="LACAREA",
                      laa_2c="LACHAREA",
                      lal_4c="LACLEN",
                      lal_2c="LALEN",
                      height_cm="height_cm",
                      weight_kg="weight_kg")

neat_echo <- calc_hypertrophy_type(df=neat_echo,
                                   sex="SEX",
                                   lvedd_cm="LVDD",
                                   ivsd_cm="IVDD",
                                   lvpwtd_cm="PWDD",
                                   height_cm = "height_cm",
                                   weight_kg="weight_kg")
neat_echo$LVEDD_index <- neat_echo$LVDD/neat_echo$bsa
neat_echo$RVSP <- 4*neat_echo$PEAKVELC^2+neat_echo$RAPRESS
neat_echo$EARATIO <- neat_echo$EVELOC/neat_echo$AVELOC
neat_echo$eeprime_sept <- neat_echo$DTIMEDE/neat_echo$EVELOC
neat_echo$eeprime_lat <- neat_echo$DTILATE/neat_echo$EVELOC
neat_echo$eeprime_avg <- (neat_echo$eeprime_sept+neat_echo$eeprime_lat)/2
neat_echo$EVELOC_cmsec <- neat_echo$EVELOC*100

## Assigning TOPCAT diastolic dysfunction to NEAT-HFpEF

neat_echo$DTILATE_cmsec <- neat_echo$DTILATE*100
neat_echo$DTIMEDE_cmsec <- neat_echo$DTIMEDE*100

nech <- 
  assign_diastolic_function(dat=neat_echo,
                            pid="patnumb",
                            vst="VISIT",
                            ep_lat="DTILATE_cmsec",
                            ep_sept="DTIMEDE_cmsec",
                            earat="EARATIO",
                            e_vel="EVELOC_cmsec",
                            dt="DECELTM",
                            def="TOPCAT",
                            verbose=F) 

neat_echo <-
  merge(neat_echo,
        nech,
        by=c("patnumb","VISIT"),
        all.x=T)

neat_echo_melt <- melt(neat_echo,
                       id.vars=c("patnumb","VISIT"),
                       na.rm=T) 

names(neat_echo_melt)[2] <- "studyvisit"
neat_echo_melt$form <- "echo"


##########  medhist1  ##########

neat_medhist1 <- read.csv(paste(neat_folder,"medhist1.csv",sep=""),
                          stringsAsFactors = F,
                          na.strings=c("NA","NaN","","NULL"))


neat_medhist1 <- merge(neat_medhist1,neat_base[,c("PATNUMB","HFETIOLOGY")],by.x="patnumb",by.y="PATNUMB")


neat_cms <- names(neat_medhist1) %in% c("ALCOHOLC",
                                          "CYTOTOX",
                                          "FAMILIAL",
                                          "HCM",
                                          "HYPERTN",
                                          "IDIODIL",
                                          "IDIORES",
                                          "MYOOTH",
                                          "PERIPRT",
                                          "VALVUL")


neat_medhist1[neat_medhist1$HFETIOLOGY==1,neat_cms] <- NA

neat_medhist1$num_etiol <- apply(neat_medhist1[neat_cms],
                                  MARGIN=1,
                                  FUN=function(x)
                                    sum(x,na.rm=T))

neat_medhist1[neat_medhist1$num_etiol>1,neat_cms] <- NA

neat_medhist1_melt <- melt(data=neat_medhist1,
                           id.vars=c("patnumb","VISIT"),
                           na.rm=T)

neat_medhist1_melt$form <- "medhist1"
names(neat_medhist1_melt)[2] <- "studyvisit"


##########  medhist2  ##########

neat_medhist2 <- read.csv(paste(neat_folder,"medhist2.csv",sep=""),
                          stringsAsFactors = F,
                          na.strings=c("NA","NaN","","NULL"))

neat_medhist2_melt <- melt(data=neat_medhist2,
                           id.vars=c("patnumb","VISIT","FORM"),
                           na.rm=T)

neat_medhist2_melt$form <- "medhist2"
names(neat_medhist2_melt)[2] <- "studyvisit"


##########  exam  ##########

neat_exam <- read.csv(paste(neat_folder,"exam.csv",sep=""),
                      stringsAsFactors = F,
                      na.strings=c("NA","NaN","","NULL"))

neat_exam$pupr <- neat_exam$BPSYS-neat_exam$BPDIA
neat_exam_melt <- melt(data=neat_exam,
                       id.vars=c("patnumb","VISIT","FORM"),
                       na.rm=T)

neat_exam_melt$form <- "exam"
names(neat_exam_melt)[2] <- "studyvisit"

##########  meds  ##########

neat_meds <- read.csv(paste(neat_folder,"meds.csv",sep=""),
                      stringsAsFactors = F,
                      na.strings=c("NA","NaN","","NULL"))


neat_meds$daily_furo_eq_derive <- apply(neat_meds
                                   [,c("FURODOSE",
                                       "TORSDOSE",
                                       "BUMDOSE")],
                                   MARGIN=1,
                                   FUN=diur_calc)


neat_meds_melt <- melt(data=neat_meds,
                       id.vars=c("patnumb","VISIT","FORM"),
                       na.rm=T)

neat_meds_melt$form <- "meds"
names(neat_meds_melt)[2] <- "studyvisit"

##########  ecg  ##########

neat_ecg <- read.csv(paste(neat_folder,"ecg.csv",sep=""),
                     stringsAsFactors = F,
                     na.strings=c("NA","NaN","","NULL"))

neat_ecg_melt <- melt(data=neat_ecg,
                      id.vars=c("patnumb","VISIT","FORM"),
                      na.rm=T)

neat_ecg_melt$form <- "ecg"
names(neat_ecg_melt)[2] <- "studyvisit"


#### kccq ####

neat_kccq <- read.csv(paste(neat_folder,"kccq.csv",sep=""))
neat_mlhf <- read.csv(paste(neat_folder,"mlhfq.csv",sep=""))

# Set missing response (99) to NA

neat_kccq[neat_kccq==99] <- NA
neat_mlhf[neat_mlhf==99] <- NA


## Note that NEAT uses the official KCCQ question numbering and order

neat_kccq <- score_kccq(dat=neat_kccq,
                        q1a="KCCQ1A",
                        q1b="KCCQ1B",
                        q1c="KCCQ1C",
                        q1d="KCCQ1D",
                        q1e="KCCQ1E",
                        q1f="KCCQ1F",
                        q2="KCCQ2",
                        q3="KCCQ3",
                        q4="KCCQ4",
                        q5="KCCQ5",
                        q6="KCCQ6",
                        q7="KCCQ7",
                        q8="KCCQ8",
                        q9="KCCQ9",
                        q10="KCCQ10",
                        q11="KCCQ11",
                        q12="KCCQ12",
                        q13="KCCQ13",
                        q14="KCCQ14",
                        q15a="KCCQ15A",
                        q15b="KCCQ15B",
                        q15c="KCCQ15C",
                        q15d="KCCQ15D")

names(neat_kccq)[names(neat_kccq)=="VISIT"] <- "studyvisit"


neat_kccq_melt <- melt(data=neat_kccq,
                       id.vars=c("patnumb","studyvisit"),
                       na.rm=T)

neat_kccq_melt$form <- "kccq"


#### mlhfq ####

neat_mlhf <- score_mlhf(dat=neat_mlhf,
                        swelling = "MLHFQ1", 
                        day_rest="MLHFQ2",
                        stairs = "MLHFQ3", 
                        housework="MLHFQ4",
                        go_places="MLHFQ5",
                        sleep = "MLHFQ6",
                        friendfam = "MLHFQ7",
                        earn_living = "MLHFQ8",
                        hobbies = "MLHFQ9",
                        sex = "MLHFQ10",
                        food = "MLHFQ11",
                        sob = "MLHFQ12",
                        fatigue = "MLHFQ13",
                        hospital = "MLHFQ14",
                        cost = "MLHFQ15",
                        sideeffect = "MLHFQ16",
                        burden = "MLHFQ17",
                        nocontrol = "MLHFQ18",
                        worry="MLHFQ19",
                        concentrate="MLHFQ20",
                        depressed="MLHFQ21", 
                        total=c("Y"))


names(neat_mlhf)[names(neat_mlhf)=="VISIT"] <- "studyvisit"


neat_mlhfq_melt <- melt(data=neat_mlhf,
                       id.vars=c("patnumb","studyvisit"),
                       na.rm=T)

neat_mlhfq_melt$form <- "mlhfq"


##########  sixmwt  ##########

neat_sixmwt <- read.csv(paste(neat_folder,"sixmwt.csv",sep=""),
                        stringsAsFactors = F,
                        na.strings=c("NA","NaN","","NULL"))

names(neat_sixmwt)[names(neat_sixmwt)=="VISIT"] <- "studyvisit"


neat_sixmwt_melt <- melt(data=neat_sixmwt,
                                       id.vars=c("patnumb","studyvisit","FORM"),
                                       na.rm=T)

neat_sixmwt_melt$form <- "sixmt"


##########  compile  ##########

var_order <- c('patnumb','form','studyvisit','variable','value')

neat_melt <- rbind(neat_base_melt[,var_order],
                   neat_ecg_melt[,var_order],
                   neat_echo_melt[,var_order],
                   neat_exam_melt[,var_order],
                   neat_kccq_melt[,var_order],
                   neat_medhist1_melt[,var_order],
                   neat_medhist2_melt[,var_order],
                   neat_meds_melt[,var_order],
                   neat_mlhfq_melt[,var_order],
                   neat_sixmwt_melt[,var_order],
                   neat_visitsumm_melt[,var_order])


neat_melt <- subset(neat_melt,!value %in% c("NA","","NULL",NA))
colnames(neat_melt)[1] <- c("patientid")
neat_melt$study <- "NEAT-HFpEF"


################################ **********DIG********** ########################################

# .%%%%%%%%..%%%%..%%%%%%..
# .%%.....%%..%%..%%....%%.
# .%%.....%%..%%..%%.......
# .%%.....%%..%%..%%...%%%%
# .%%.....%%..%%..%%....%%.
# .%%.....%%..%%..%%....%%.
# .%%%%%%%%..%%%%..%%%%%%..


##########  form01  ##########

dig_form01 <- read.csv(paste(dig_folder,"form01.csv",sep=""),
                       na.strings=c(NA,"","NULL"),
                       stringsAsFactors = F)

dig_form01$Q9_MG[!is.na(dig_form01$Q9_MOL)&is.na(dig_form01$Q9_MG)] <- 
  round(dig_form01$Q9_MOL[!is.na(dig_form01$Q9_MOL)&is.na(dig_form01$Q9_MG)]*0.0113,1)

dig_form01$CrCl <- calc_MDRD4(dat=dig_form01,
                              cr="Q9_MG",
                              sex="Q4",
                              race="RACE",
                              age="AGE",
                              male=1,
                              black=2)

dig_form01$pupr <- dig_form01$Q22_SYS-dig_form01$Q22_DIA

dig_form01_melt <- melt(data=dig_form01,id.vars=c("PATIENT"),na.rm=T) 
dig_form01_melt$VISIT <- "0"
dig_form01_melt$studyvisit <- dig_form01_melt$VISIT
dig_form01_melt$form <- "form01"


##########  form10  ##########

dig_form10 <- read.csv(paste(dig_folder,"form10.csv",sep=""),
                       na.strings=c(NA,"","NULL"),
                       stringsAsFactors = F)


dig_form10 <- score_mlhf(dat=dig_form10,
                         swelling = "Q13", 
                         day_rest="Q16",
                         stairs = "Q19", 
                         housework="Q14",
                         go_places="Q23",
                         sleep = "Q21",
                         friendfam = "Q15",
                         earn_living = "Q18",
                         hobbies = "Q25",
                         sex = "Q24",
                         food = "Q22",
                         sob = "Q20",
                         fatigue = "Q17",
                         hospital = "Q32",
                         cost = "Q30",
                         sideeffect = "Q27",
                         burden = "Q33",
                         nocontrol = "Q31",
                         worry="Q28",
                         concentrate="Q26",
                         depressed="Q29", 
                         total=c("Y")) 


dig_form10$stai_y1_total <- apply(dig_form10, MARGIN=1, FUN=function(x) sum(x[53:72]))

dig_form10$vas_global <- dig_form10$Q67*10 

dig_form10$cesd_10 <- apply(dig_form10,
                            MARGIN=1,
                            function(x)
                              sum(x[c(42,
                                      44:52)]))


dig_form10_melt <- melt(data=dig_form10,id.vars=c("PATIENT","VISIT"),na.rm=T) 
dig_form10_melt$studyvisit <- dig_form10_melt$VISIT
dig_form10_melt$form <- "form10"


##########  form12  ##########

dig_form12 <- read.csv(paste(dig_folder,"form12.csv",sep=""),
                       na.strings=c(NA,"","NULL"),
                       stringsAsFactors = F)

dig_form12$Q2_KG[is.na(dig_form12$Q2_KG)&!is.na(dig_form12$Q2_LBS)] <- 
  dig_form12$Q2_LBS[is.na(dig_form12$Q2_KG)&!is.na(dig_form12$Q2_LBS)]/2.2

dig_form12 <- subset(dig_form12,!(PATIENT==2626&DATETIME==0))
dig_form12 <- subset(dig_form12,!(PATIENT==2793&DATETIME==85))
dig_form12 <- subset(dig_form12,!(PATIENT==2956&DATETIME==81))
dig_form12 <- subset(dig_form12,!(PATIENT==3621&DATETIME==49))
dig_form12 <- subset(dig_form12,!(PATIENT==4291&DATETIME==49))
dig_form12 <- subset(dig_form12,!(PATIENT==5276&DATETIME==118))
dig_form12 <- subset(dig_form12,!(PATIENT==5504&DATETIME==318))
dig_form12 <- subset(dig_form12,!(PATIENT==7220&DATETIME==17))
dig_form12 <- subset(dig_form12,!(PATIENT==7686&DATETIME==80))

dig_form12_melt <- melt(data=dig_form12,
                        id.vars=c("PATIENT","VISIT"),na.rm=T) 
dig_form12_melt$studyvisit <- dig_form12_melt$VISIT
dig_form12_melt$form <- "form12"


#### form16 ####

dig_form16 <- read.csv(paste(dig_folder,"form16.csv",sep=""),
                       na.strings=c(NA,"","NULL"),
                       stringsAsFactors = F)

dig_form16_melt <- melt(data=dig_form16,id.vars=c("PATIENT","VISIT"),na.rm=T) 
dig_form16_melt$studyvisit <- dig_form16_melt$VISIT
dig_form16_melt$form <- "form10"


#### status ####

dig_bmi_melt <- read.csv(paste(dig_folder,"status.csv",sep=""),
                         na.strings=c(NA,"","NULL"),
                         stringsAsFactors = F)[,c("PATIENT","BMI")]

dig_bmi_melt$studyvisit <- 0
dig_bmi_melt$value <- round(dig_bmi_melt$BMI,1)
dig_bmi_melt$variable <- "bmi"
dig_bmi_melt$form <- "status"


##########  compile  ##########

dig_melt <- rbind(dig_form01_melt[,c("PATIENT","studyvisit","variable","value","form")],
                  dig_form10_melt[,c("PATIENT","studyvisit","variable","value","form")],
                  dig_form12_melt[,c("PATIENT","studyvisit","variable","value","form")],
                  dig_form16_melt[,c("PATIENT","studyvisit","variable","value","form")],
                  dig_bmi_melt[,c("PATIENT","studyvisit","variable","value","form")])

dig_melt <- subset(dig_melt,!is.na(value)&!value=="NaN")
colnames(dig_melt)[c(1)] <- c("patientid")
dig_melt$study <- "DIG"


########################## **********MOCHA********** ##########################

# .%%.....%%..%%%%%%%...%%%%%%..%%.....%%....%%%...
# .%%%...%%%.%%.....%%.%%....%%.%%.....%%...%%.%%..
# .%%%%.%%%%.%%.....%%.%%.......%%.....%%..%%...%%.
# .%%.%%%.%%.%%.....%%.%%.......%%%%%%%%%.%%.....%%
# .%%.....%%.%%.....%%.%%.......%%.....%%.%%%%%%%%%
# .%%.....%%.%%.....%%.%%....%%.%%.....%%.%%.....%%
# .%%.....%%..%%%%%%%...%%%%%%..%%.....%%.%%.....%%


##########  demog  ##########

mocha_demog <- read.csv(paste(mocha_folder,"demog.csv",sep=""), 
                        na.strings = c("","NA","NULL"),
                        stringsAsFactors = F)


mocha_demog_melt <-   melt(data=mocha_demog,id.vars=c("PATNO","VISITC"),na.rm=T)
mocha_demog_melt$form <- "demog"
mocha_demog_melt$studyvisit <- 1.1


##########  core  ##########

mocha_core <- read.csv(paste(mocha_folder,"core.csv",sep=""), 
                       na.strings = c("","NA","NULL"),
                       stringsAsFactors = F)

mocha_core$hfage_months <- mocha_core$DCHF*12
mocha_core$bmi <- round(mocha_core$B_WGT/((mocha_core$B_HGT/100)^2),1)


mocha_core_melt <-   melt(data=mocha_core,id.vars=c("PATNO"),na.rm=T)
mocha_core_melt$form <- "core"

mocha_randomized <- mocha_core$PATNO[!is.na(mocha_core$GROUP)]

mocha_core_melt$studyvisit <- 1.1


##########  lab_core  ##########

mocha_labcore <- read.csv(paste(mocha_folder,
                                "lab_core.csv",
                                sep=""),
                          na.strings = c("","NULL"),
                          stringsAsFactors = F)[,c("PATNO",
                                                   "PHASE",
                                                   "VISIT",
                                                   "LPARM",
                                                   "LVAL")]


mocha_labcore$studyvisit <- paste(mocha_labcore$PHASE,
                                  floor(mocha_labcore$VISIT),
                                  sep=".")


# mocha_labcore$value <- as.numeric(mocha_labcore$LVAL)
# mocha_labcore$variable <- as.character(mocha_labcore$LPARM)


mocha_labcore_melt <- sqldf("select PATNO,studyvisit,LPARM as variable, avg(LVAL) as value from mocha_labcore
                            group by PATNO,studyvisit,variable")

mocha_labcore_melt$value <- round(mocha_labcore_melt$value,1)

mocha_alc <- sqldf("select PATNO,
                     studyvisit,
                     'alc' as variable,
                     lymph,
                     wbc,
                     lymph*wbc as value
                     from (select PATNO, studyvisit, value/100 as lymph from mocha_labcore_melt where variable = 'LYMPH') as l 
                     join (select PATNO, studyvisit, value as wbc from mocha_labcore_melt where variable = 'WBC') as w
                     using (PATNO, studyvisit)")


mocha_anc <- sqldf("select PATNO,
                     studyvisit,
                     'anc' as variable,
                     neut,
                     wbc,
                     neut*wbc as value
                     from (select PATNO, studyvisit, value/100 as neut from mocha_labcore_melt where variable = 'NEUT') as n 
                     join (select PATNO, studyvisit, value as wbc from mocha_labcore_melt where variable = 'WBC') as w
                     using (PATNO, studyvisit)")

mocha_crcl_temp <- merge(subset(mocha_labcore_melt,variable=="CREAT"),
                    mocha_core[,c("PATNO","AGE","RACE","SEX")],
                    by="PATNO",
                    all.x=T)

mocha_crcl_temp$CrCl <- calc_MDRD4(dat=mocha_crcl_temp,
                              cr="value")


mocha_crcl <- sqldf("select PATNO,studyvisit,'CrCl' as variable,CrCl as value from mocha_crcl_temp")

mocha_labcore_melt <- rbind(mocha_labcore_melt[,c("PATNO","studyvisit","variable","value")],
                            mocha_alc[,c("PATNO","studyvisit","variable","value")],
                            mocha_anc[,c("PATNO","studyvisit","variable","value")],
                            mocha_crcl[,c("PATNO","studyvisit","variable","value")])

mocha_labcore_melt$form <- "lab_core"


##########  cmed  ##########


mocha_cmed <- read.csv(paste(mocha_folder,"cmed.csv",sep=""),
                       na.strings = c("","NA","NULL"),
                       stringsAsFactors = F)

mocha_vbm <- read.csv(paste(mocha_folder,"mocha_vbmdk.csv",
                            sep=""),
                      na.strings = c("","NA","NULL"),
                      stringsAsFactors = F)

mocha_vbm <- sqldf("select distinct VERBAT,ATC as ATC_DK,CM_INGRED_DK
                   from mocha_vbm")

mocha_cmed$studyvisit <- paste(mocha_cmed$PHASE,floor(mocha_cmed$VISIT),sep=".")
mocha_cmed$studyvisit[str_detect(mocha_cmed$studyvisit, "1.*")] <- "1.1"
mocha_cmed$studyvisit[str_detect(mocha_cmed$studyvisit, "2.*")] <- "2"
mocha_cmed$studyvisit[str_detect(mocha_cmed$studyvisit, "3.*")] <- "3"


mocha_cmed <- merge(mocha_cmed,
                    mocha_vbm,
                    by=c("VERBAT"),
                    all.x=T)

mocha_ingred_melt <- sqldf("select PATNO, 
                         studyvisit,
                         'CM_INGRED_DK' as variable,
                           CM_INGRED_DK as value
                           from mocha_cmed")

mocha_atc_melt <- sqldf("select PATNO, 
                         studyvisit,
                         'ATC' as variable,
                           ATC as value
                           from mocha_cmed")


####### Daily furosemide dose #######

mocha_cmed$CM_INGRED_DK[mocha_cmed$CM_INGRED_DK=="ETHACRYNIC ACID"] <- "FUROSEMIDE"
mocha_cmed$DOSEN[mocha_cmed$CM_INGRED_DK=="ETHACRYNIC ACID"] <- mocha_cmed$DOSEN[mocha_cmed$CM_INGRED_DK=="ETHACRYNIC ACID"]*0.8

mocha_furo <- sqldf("select PATNO, 
                    studyvisit, 
                    DOSEN as FURODOSE 
                    from mocha_cmed 
                    where CM_INGRED_DK = 'FUROSEMIDE' 
                    group by PATNO,studyvisit 
                    having max(DOSEN)")

mocha_bum <- sqldf("select PATNO, 
                    studyvisit, 
                    DOSEN as BUMDOSE 
                    from mocha_cmed 
                    where CM_INGRED_DK = 'BUMETANIDE' 
                    group by PATNO,studyvisit 
                    having max(DOSEN)")


mocha_tors <- sqldf("select PATNO, 
                    studyvisit, 
                    DOSEN as TORSEDOSE 
                    from mocha_cmed 
                    where CM_INGRED_DK = 'TORSEMIDE' 
                    group by PATNO,studyvisit 
                    having max(DOSEN)")


mocha_furo_equiv <- merge(mocha_furo,
                       mocha_bum,
                       by=c("PATNO","studyvisit"),
                       all.x=T)

mocha_furo_equiv <- merge(mocha_furo_equiv,
                       mocha_tors,
                       by=c("PATNO","studyvisit"),
                       all.x=T)


mocha_furo_equiv$value <- apply(mocha_furo_equiv
                             [,c("FURODOSE",
                                 "TORSEDOSE",
                                 "BUMDOSE")],
                             MARGIN=1,
                             FUN=diur_calc)

mocha_furo_equiv$variable <- "daily_furo_eq_derive"

mocha_cmed_melt <- rbind(mocha_ingred_melt,
                         mocha_atc_melt,
                         mocha_furo_equiv[,c("PATNO","studyvisit","variable","value")])

mocha_cmed_melt$form <- "cmed"

##########  medhx  ##########


mocha_medhx <- read.csv(paste(mocha_folder,"medhx.csv",sep=""),
                       na.strings = c("","NA","NULL"),
                       stringsAsFactors = F)


mocha_medhx$studyvisit <- paste(mocha_medhx$PHASE,
                                  floor(mocha_medhx$VISIT),
                                  sep=".")

mocha_medhx_melt <-   melt(data=mocha_medhx,id.vars=c("PATNO","studyvisit"),na.rm=T)


mocha_medhx_melt$form <- "medhx"


##########  chfhx  ##########


mocha_chfhx <- read.csv(paste(mocha_folder,"chfhx.csv",sep=""),
                        na.strings = c("","NA","NULL"),
                        stringsAsFactors = F)
mocha_chfhx$studyvisit <- "1.1"

mocha_chfhx_melt <-   melt(data=mocha_chfhx,id.vars=c("PATNO","studyvisit"),na.rm=T)


mocha_chfhx_melt$form <- "chfhx"


##########  vsign  ##########


mocha_vsign <- read.csv(paste(mocha_folder,"vsign.csv",sep=""),
                        na.strings = c("","NA","NULL"),
                        stringsAsFactors = F)

mocha_vsign$studyvisit <- paste(mocha_vsign$PHASE,
                                mocha_vsign$VISIT,
                                sep=".")

mocha_vsign$rnme <- rownames(mocha_vsign)

mocha_vsign <- subset(mocha_vsign,POSITION==1)

mocha_vsign$pupr <- mocha_vsign$SYST-mocha_vsign$DIAS

mocha_vsign <- merge(mocha_vsign,mocha_core[,c("PATNO","B_HGT")], by="PATNO", all.x=T)
mocha_vsign$bmi <- round(mocha_vsign$WGT/(mocha_vsign$B_HGT/100)^2,1)


mocha_vsign_melt <-   melt(data=mocha_vsign,id.vars=c("PATNO",
                                                      "studyvisit"),na.rm=T)


mocha_vsign_melt$form <- "vsign"

##########  radnucve  ##########

mocha_radnucve <- read.csv(paste(mocha_folder,"radnucve.csv",sep=""), 
                       na.strings = c("","NA","NULL"),
                       stringsAsFactors = F)

mocha_radnucve$studyvisit <- paste(mocha_radnucve$PHASE,
                                floor(mocha_radnucve$VISIT),
                                sep=".")

mocha_radnucve$LVEJECT_pct <- mocha_radnucve$LVEJECT*100

mocha_radnucve_melt <-   melt(data=mocha_radnucve,id.vars=c("PATNO","studyvisit"),na.rm=T)
mocha_radnucve_melt$form <- "radnucve"


##########  cpex  ##########


mocha_cpex <- read.csv(paste(mocha_folder,"cpex.csv",sep=""),
                        na.strings = c("","NA","NULL"),
                        stringsAsFactors = F)

mocha_cpex$studyvisit <- paste(mocha_cpex$PHASE,
                               mocha_cpex$VISIT,
                                sep=".")

mocha_cpex_melt <-   melt(data=mocha_cpex,id.vars=c("PATNO","studyvisit"),na.rm=T)

mocha_cpex_melt$form <- "cpex"


##########  walktst  ##########

mocha_walktst <- read.csv(paste(mocha_folder,"walktst.csv",sep=""),
                        na.strings = c("","NA","NULL"),
                        stringsAsFactors = F)

mocha_walktst$studyvisit <- paste(mocha_walktst$PHASE,
                                floor(mocha_walktst$VISIT),
                                sep=".")

mocha_walktst$walk_time <- mocha_walktst$TOT_MIN + mocha_walktst$TOT_SEC/60

mocha_walktst_mean <- sqldf("select PATNO, studyvisit, avg(DISTANCE) as dist, avg(walk_time) as wlk
                             from mocha_walktst 
                             group by PATNO,studyvisit")


mocha_walktst_melt <-   melt(data=mocha_walktst_mean,id.vars=c("PATNO","studyvisit"),na.rm=T)
mocha_walktst_melt$form <- "walktst"


##########  symas  ##########

mocha_symas <- read.csv(paste(mocha_folder,"symas.csv",sep=""),
                          na.strings = c("","NA","NULL"),
                          stringsAsFactors = F)

mocha_symas$studyvisit <- paste(mocha_symas$PHASE,
                                  mocha_symas$VISIT,
                                  sep=".")

mocha_symas_totals <- sqldf("select PATNO, 
                            PHASE, 
                            VISITC, 
                            VDATE, 
                            'global_assessmt' as variable,
                            SUM(ANSWER) as tot 
                            from mocha_symas 
                            group by PATNO,PHASE,VISITC,VDATE")

mocha_symas_melt <-   melt(data=mocha_symas,id.vars=c("PATNO"),na.rm=T)
mocha_symas_melt$form <- "symas"


##########  nyha  ##########


mocha_nyha <- read.csv(paste(mocha_folder,"nyha.csv",sep=""),
                          na.strings = c("","NA","NULL"),
                          stringsAsFactors = F)

mocha_nyha$studyvisit <- paste(mocha_nyha$PHASE,mocha_nyha$VISIT,sep=".")

mocha_nyha_melt <-   melt(data=mocha_nyha,id.vars=c("PATNO","studyvisit"),na.rm=T)
mocha_nyha_melt$form <- "nyha"


##########  ecg  ##########

mocha_ecg <- read.csv(paste(mocha_folder,"ecg.csv",sep=""),
                       na.strings = c("","NA","NULL"),
                       stringsAsFactors = F)

mocha_ecg$studyvisit <- paste(mocha_ecg$PHASE,mocha_ecg$VISIT,sep=".")

mocha_ecg_melt <-   melt(data=mocha_ecg,id.vars=c("PATNO","studyvisit"),na.rm=T)
mocha_ecg_melt$form <- "ecg"


##########  a_hosp  ##########


mocha_ahosp <- read.csv(paste(mocha_folder,"a_hosp.csv",sep=""),
                        na.strings = c("","NA","NULL"),
                        stringsAsFactors = F)

mocha_ahosp_melt <-   melt(data=mocha_ahosp,id.vars=c("PATNO"),na.rm=T)

mocha_ahosp_melt$studyvisit <- "1.1"
mocha_ahosp_melt$form <- "a_hosp"


#############  LIHFE  ################

mocha_lihfe <- read.csv(paste(mocha_folder,"lihfe.csv",sep=""),
                        na.strings=c("NA","","NULL"))

mocha_lihfe$QUESTION <- paste("MLHF",mocha_lihfe$QUESTION,sep="")
mocha_mlhf <- as.data.frame(acast(mocha_lihfe,PATNO+PHASE+VISITC~QUESTION,value.var="ANSWER"))


mocha_mlhf <- score_mlhf(dat=mocha_mlhf,
                         swelling = "MLHFQ1", 
                         day_rest="MLHFQ2",
                         stairs = "MLHFQ3", 
                         housework="MLHFQ4",
                         go_places="MLHFQ5",
                         sleep = "MLHFQ6",
                         friendfam = "MLHFQ7",
                         earn_living = "MLHFQ8",
                         hobbies = "MLHFQ9",
                         sex = "MLHFQ10",
                         food = "MLHFQ11",
                         sob = "MLHFQ12",
                         fatigue = "MLHFQ13",
                         hospital = "MLHFQ14",
                         cost = "MLHFQ15",
                         sideeffect = "MLHFQ16",
                         burden = "MLHFQ17",
                         nocontrol = "MLHFQ18",
                         worry="MLHFQ19",
                         concentrate="MLHFQ20",
                         depressed="MLHFQ21", 
                         total=c("Y"))

mocha_mlhf$PATNO <- str_split_fixed(rownames(mocha_mlhf),"_",n=2)[,1]
mocha_mlhf$studyvisit <- str_split_fixed(rownames(mocha_mlhf),"_",n=2)[,2]
mocha_mlhf$studyvisit <-str_replace(mocha_mlhf$studyvisit,"_",".")

mocha_mlhf_melt <- melt(data=mocha_mlhf, id.vars=c("PATNO","studyvisit"),na.rm=T)
mocha_mlhf_melt$form <- "lihfe"

##########  compile  ##########

mocha_melt <- rbind(mocha_core_melt[,c("PATNO","studyvisit","variable","value","form")],
                    mocha_chfhx_melt[,c("PATNO","studyvisit","variable","value","form")],
                    mocha_cpex_melt[,c("PATNO","studyvisit","variable","value","form")],
                    mocha_medhx_melt[,c("PATNO","studyvisit","variable","value","form")],
                    mocha_radnucve_melt[,c("PATNO","studyvisit","variable","value","form")],
                    mocha_ahosp_melt[,c("PATNO","studyvisit","variable","value","form")],
                    mocha_cmed_melt[,c("PATNO","studyvisit","variable","value","form")],
                    mocha_demog_melt[,c("PATNO","studyvisit","variable","value","form")],
                    mocha_nyha_melt[,c("PATNO","studyvisit","variable","value","form")],
                    mocha_vsign_melt[,c("PATNO","studyvisit","variable","value","form")],
                    mocha_walktst_melt[,c("PATNO","studyvisit","variable","value","form")],
                    mocha_labcore_melt[,c("PATNO","studyvisit","variable","value","form")],
                    mocha_mlhf_melt[,c("PATNO","studyvisit","variable","value","form")])

mocha_melt <- subset(mocha_melt,!is.na(value)&!value=="<NA>")
mocha_melt <- subset(mocha_melt,str_count(studyvisit,"..") < 2)
mocha_melt <- subset(mocha_melt,PATNO %in% mocha_randomized)
names(mocha_melt)[1] <- "patientid"

mocha_melt$study <- "MOCHA"


######################### **********CARRESS********** ############################

# ..%%%%%%.....%%%....%%%%%%%%..%%%%%%%%..%%%%%%%%..%%%%%%...%%%%%%.
# .%%....%%...%%.%%...%%.....%%.%%.....%%.%%.......%%....%%.%%....%%
# .%%........%%...%%..%%.....%%.%%.....%%.%%.......%%.......%%......
# .%%.......%%.....%%.%%%%%%%%..%%%%%%%%..%%%%%%....%%%%%%...%%%%%%.
# .%%.......%%%%%%%%%.%%...%%...%%...%%...%%.............%%.......%%
# .%%....%%.%%.....%%.%%....%%..%%....%%..%%.......%%....%%.%%....%%
# ..%%%%%%..%%.....%%.%%.....%%.%%.....%%.%%%%%%%%..%%%%%%...%%%%%%.


##########  a_base  ##########

carress_base <- read.csv(paste(carress_folder,"a_base.csv",sep=""),na.strings=c(NA,"NULL",""))
carress_base$yihf_months <- carress_base$YIHF*12

carress_base$RACE[carress_base$ETHNIC==1] <- 8

carress_base_melt <- melt(data=carress_base,id.vars=c("PATNUMB"),na.rm=T) 
carress_base_melt$studyvisit <- "BASELINE"
carress_base_melt$form <- "a_base"

names(carress_base_melt)[1] <- "patnumb"
carress_base_melt$variable <- as.character(carress_base_melt$variable)

##########  meds  ##########

carress_meds_melt <- read.csv(paste(carress_folder,"meds.csv",sep=""),na.strings=c(NA,"NULL",""))
carress_meds_melt[carress_meds_melt=="NaN"] <- NA
carress_meds_melt$studyvisit <- carress_meds_melt$FORM

carress_meds_melt$variable <- carress_meds_melt$HFMEDS
carress_meds_melt$value <- carress_meds_melt$MEDSANS
carress_meds_melt$form <- "meds"

##### diuretic  ####

carress_diuretic <- read.csv(paste(carress_folder,"diuretic.csv",sep=""),na.strings=c(NA,"NULL",""))
carress_diuretic[carress_diuretic=="NaN"] <- NA

carress_diuretic_melt <- sqldf("select patnumb,
                               FORM as studyvisit,
                               DIUMEDS as variable,
                               DIURANS as value,
                               'diuretic' as form
                               from carress_diuretic")

##### Daily furosemide equivalent #####

# Daily furosemide equivalent dose at each timepoint is included in the 'endpts' table
# Verified that calculation of dose equivalents in 'endpts' using my formulas

carress_endpts <- read.csv(paste(carress_folder,"a_endpts.csv",sep=""),na.strings=c(NA,"NULL",""))
carress_endpts[carress_endpts=="NaN"] <- NA

carress_endpts_furo_eq <- carress_endpts[,c(1,114,116:117,119)]

carress_furo_eq_melt <- melt(data=carress_endpts_furo_eq,
                             id.vars='PATNUMB',
                             na.rm=T)

carress_furo_eq_melt$studyvisit[carress_furo_eq_melt$variable=="FURODSEQ_PH"] <- "BASELINE"
carress_furo_eq_melt$studyvisit[carress_furo_eq_melt$variable=="FURODSEQ_DC"] <- "DAY7"
carress_furo_eq_melt$studyvisit[carress_furo_eq_melt$variable=="FURODSEQ_30D"] <- "DAY30"
carress_furo_eq_melt$studyvisit[carress_furo_eq_melt$variable=="FURODSEQ_60D"] <- "DAY60"

carress_furo_eq_melt$variable <- "daily_furo_eq_derive"

carress_furo_eq_melt$form <- "endpts"
names(carress_furo_eq_melt)[1] <- "patnumb"

##########  medhist1  ##########

carress_medhist1 <- read.csv(paste(carress_folder,"medhist1.csv",sep=""),na.strings=c(NA,"NULL",""))
carress_medhist1[carress_medhist1=="NaN"] <- NA                            


carress_cms <- names(carress_medhist1) %in% c("ALCOHOLC",
                                            "CYTOTOXC",
                                            "FAMILIAL",
                                            "HCM",
                                            "HYPERTEN",
                                            "DILATED",
                                            "RESTRICT",
                                            "OTHCONT",
                                            "PERIPAR",
                                            "VAL")


carress_medhist1[carress_medhist1$ISCHEMIC==1,carress_cms] <- NA

carress_medhist1$num_etiol <- apply(carress_medhist1[carress_cms],
                                   MARGIN=1,
                                   FUN=function(x)
                                     sum(x,na.rm=T))

carress_medhist1[carress_medhist1$num_etiol>1,carress_cms] <- NA


carress_medhist1_melt <- subset(melt(data=carress_medhist1,
                                     id.vars=c("patnumb","FORM"),
                                     na.rm=T))

carress_medhist1_melt$form <- "medhist1"
carress_medhist1_melt$studyvisit  <- carress_medhist1_melt$FORM
carress_medhist1_melt$variable <- as.character(carress_medhist1_melt$variable)


##########  medhist2  ##########

carress_medhist2 <- read.csv(paste(carress_folder,"medhist2.csv",sep=""),na.strings=c(NA,"NULL",""))
carress_medhist2[carress_medhist2=="NaN"] <- NA                            

carress_medhist2_melt <- subset(melt(data=carress_medhist2,
                                     id.vars=c("patnumb","FORM"),
                                     na.rm=T))

carress_medhist2_melt$form <- "medhist2"
carress_medhist2_melt$studyvisit  <- carress_medhist2_melt$FORM
carress_medhist2_melt$variable <- as.character(carress_medhist2_melt$variable)


#####  a_visitsumm  #####

carress_visitsumm <- read.csv(paste(carress_folder,"a_visitsumm.csv",sep=""),na.strings=c(NA,"NULL",""))
carress_visitsumm[carress_visitsumm=="NaN"] <- NA                            

carress_ht <- sqldf("select PATNUMB,
                    HEIGHTIN as height_in
                    from carress_visitsumm
                    where FORM=='BASELINE'
                    ")

carress_visitsumm <- merge(carress_visitsumm,
                           carress_ht,
                           by="PATNUMB",
                           all.x=T)

carress_visitsumm$alc <- carress_visitsumm$LL_LYMPH/100*carress_visitsumm$LL_WBC
carress_visitsumm$hct_calc <- carress_visitsumm$LL_HMG*3
carress_visitsumm$tni_ng_ml <- carress_visitsumm$CL_TROPI/1000
carress_visitsumm$weight_kg <- carress_visitsumm$WTLBS/2.2

carress_visitsumm$BMI <- round(carress_visitsumm$WTLBS*703/(carress_visitsumm$height_in)^2,1)

carress_visitsumm_melt <- melt(data=carress_visitsumm,
                                     id.vars=c("PATNUMB","FORM"),
                                     na.rm=T)

carress_visitsumm_melt$form <- "a_visitsumm"
carress_visitsumm_melt$studyvisit <- carress_visitsumm_melt$FORM

names(carress_visitsumm_melt)[1] <- "patnumb"
carress_visitsumm_melt$variable <- as.character(carress_visitsumm_melt$variable)


#####  assessmt #####

carress_assessmt <- read.csv(paste(carress_folder,"assessmt.csv",sep=""),na.strings=c(NA,"NULL",""))
carress_assessmt[carress_assessmt=="NaN"] <- NA                            

carress_assessmt$pupr <- carress_assessmt$BPSYS-carress_assessmt$BPDIA

carress_assessmt_melt <- subset(melt(data=carress_assessmt,
                                     id.vars=c("patnumb","FORM"),
                                     na.rm=T))


carress_assessmt_melt$studyvisit <- carress_assessmt_melt$FORM
carress_assessmt_melt$form <- "assessmt"
carress_assessmt_melt$variable <- as.character(carress_assessmt_melt$variable)


##########  ecg  ##########

carress_ecg <- read.csv(paste(carress_folder,"ecg.csv",sep=""),na.strings=c(NA,"NULL",""))
carress_ecg[carress_ecg=="NaN"] <- NA                            

carress_ecg_melt <- subset(melt(data=carress_ecg,
                                     id.vars=c("patnumb","FORM"),
                                     na.rm=T))

carress_ecg_melt$studyvisit <- carress_ecg_melt$FORM
carress_ecg_melt$form <- "ecg"
carress_ecg_melt$variable <- as.character(carress_ecg_melt$variable)


##########  crfluid  ##########

carress_crfluid <- read.csv(paste(carress_folder,"crfluid.csv",sep=""),na.strings=c(NA,"NULL",""))
carress_crfluid[carress_crfluid=="NaN"] <- NA

carress_fluid_start <- carress_base[,c("PATNUMB","TREATMENT")]

names(carress_fluid_start)[1] <- "patnumb"

carress_crfluid <-subset(carress_crfluid,!is.na(CRURINOT))

carress_crfluid$CRULTOUT[is.na(carress_crfluid$CRULTOUT)&carress_crfluid$CRFNONE==1] <- 0

carress_crfluid_sum <- sqldf('select patnumb,
                                       FORM,
                                       CRIVIN+CRORALIN-CRULTOUT-CRURINOT as io_balance
                                       from carress_crfluid')


carress_fluid_cum_d1 <- subset(carress_crfluid_sum,FORM=="DAY1"&!is.na(io_balance))[,c("patnumb","io_balance")]
names(carress_fluid_cum_d1)[2] <- "io_balancexDAY1"

carress_fluid_cum_d2 <- subset(carress_crfluid_sum,FORM=="DAY2"&!is.na(io_balance))[,c("patnumb","io_balance")]
names(carress_fluid_cum_d2)[2] <- "io_balancexDAY2"

carress_fluid_cum_d3 <- subset(carress_crfluid_sum,FORM=="DAY3"&!is.na(io_balance))[,c("patnumb","io_balance")]
names(carress_fluid_cum_d3)[2] <- "io_balancexDAY3"

carress_fluid_cum_d4 <- subset(carress_crfluid_sum,FORM=="DAY4"&!is.na(io_balance))[,c("patnumb","io_balance")]
names(carress_fluid_cum_d4)[2] <- "io_balancexDAY4"

carress_fluid_cum_d5 <- subset(carress_crfluid_sum,FORM=="DAY5"&!is.na(io_balance))[,c("patnumb","io_balance")]
names(carress_fluid_cum_d5)[2] <- "io_balancexDAY5"

carress_fluid_cum_d6 <- subset(carress_crfluid_sum,FORM=="DAY6"&!is.na(io_balance))[,c("patnumb","io_balance")]
names(carress_fluid_cum_d6)[2] <- "io_balancexDAY6"

carress_fluid_cum_d7 <- subset(carress_crfluid_sum,FORM=="DAY7"&!is.na(io_balance))[,c("patnumb","io_balance")]
names(carress_fluid_cum_d7)[2] <- "io_balancexDAY7"


carress_fluid_cum <- merge(carress_fluid_start[,c("patnumb","TREATMENT")],
                           carress_fluid_cum_d1,
                           by="patnumb",
                           all.x=T)

carress_fluid_cum <- merge(carress_fluid_cum,carress_fluid_cum_d2,by="patnumb",all.x=T)
carress_fluid_cum <- merge(carress_fluid_cum,carress_fluid_cum_d3,by="patnumb",all.x=T)
carress_fluid_cum <- merge(carress_fluid_cum,carress_fluid_cum_d4,by="patnumb",all.x=T)
carress_fluid_cum <- merge(carress_fluid_cum,carress_fluid_cum_d5,by="patnumb",all.x=T)
carress_fluid_cum <- merge(carress_fluid_cum,carress_fluid_cum_d6,by="patnumb",all.x=T)
carress_fluid_cum <- merge(carress_fluid_cum,carress_fluid_cum_d7,by="patnumb",all.x=T)

carress_fluid_cum$io_cumxDAY1 <- carress_fluid_cum$io_balancexDAY1
carress_fluid_cum$io_cumxDAY2 <- carress_fluid_cum$io_balancexDAY1+carress_fluid_cum$io_balancexDAY2
carress_fluid_cum$io_cumxDAY3 <- carress_fluid_cum$io_cumxDAY2+carress_fluid_cum$io_balancexDAY3
carress_fluid_cum$io_cumxDAY4 <- carress_fluid_cum$io_cumxDAY3+carress_fluid_cum$io_balancexDAY4
carress_fluid_cum$io_cumxDAY5 <- carress_fluid_cum$io_cumxDAY4+carress_fluid_cum$io_balancexDAY5
carress_fluid_cum$io_cumxDAY6 <- carress_fluid_cum$io_cumxDAY5+carress_fluid_cum$io_balancexDAY6
carress_fluid_cum$io_cumxDAY7 <- carress_fluid_cum$io_cumxDAY6+carress_fluid_cum$io_balancexDAY7

carress_crfluid_melt <- melt(carress_fluid_cum,
                             id.vars=c("patnumb","TREATMENT"),
                             na.rm=T)
carress_crfluid_melt$variable <- as.character(carress_crfluid_melt$variable)
carress_crfluid_melt$studyvisit <- str_select(carress_crfluid_melt$variable,after="x")
carress_crfluid_melt$variable <- str_select(carress_crfluid_melt$variable,before="x")

carress_crfluid_melt$form <- "crfluid"


##### CARRESS VAS

carress_vas <- read.csv(paste(carress_folder,"vas.csv",sep=""),na.strings=c(NA,"NULL",""))
carress_vas[carress_vas=="NaN"] <- NA
carress_vas_melt <- melt(carress_vas, id.vars=c("patnumb","FORM"), na.rm=T)
carress_vas_melt$studyvisit <- carress_vas_melt$FORM
carress_vas_melt$form <- "vas"


##########  compile  ##########

carress_melt <- rbind(carress_assessmt_melt[,c("patnumb","form","studyvisit","variable","value")],
                      carress_base_melt[,c("patnumb","form","studyvisit","variable","value")],
                      carress_crfluid_melt[,c("patnumb","form","studyvisit","variable","value")],
                      carress_diuretic_melt[,c("patnumb","form","studyvisit","variable","value")],
                      carress_ecg_melt[,c("patnumb","form","studyvisit","variable","value")],
                      carress_furo_eq_melt[,c("patnumb","form","studyvisit","variable","value")],
                      carress_medhist1_melt[,c("patnumb","form","studyvisit","variable","value")],
                      carress_medhist2_melt[,c("patnumb","form","studyvisit","variable","value")],
                      carress_meds_melt[,c("patnumb","form","studyvisit","variable","value")],
                      carress_visitsumm_melt[,c("patnumb","form","studyvisit","variable","value")],
                      carress_vas_melt[,c("patnumb","form","studyvisit","variable","value")])

carress_melt <- subset(carress_melt,!is.na(value))
names(carress_melt)[1] <- "patientid"
carress_melt$study <- "CARRESS"


########################### **********DOSE********** ################################

# .%%%%%%%%...%%%%%%%...%%%%%%..%%%%%%%%
# .%%.....%%.%%.....%%.%%....%%.%%......
# .%%.....%%.%%.....%%.%%.......%%......
# .%%.....%%.%%.....%%..%%%%%%..%%%%%%..
# .%%.....%%.%%.....%%.......%%.%%......
# .%%.....%%.%%.....%%.%%...%%.%%......
# .%%%%%%%%...%%%%%%%...%%%%%%..%%%%%%%%


##########  analysis/a_base  ##########

dose_base <- read.csv(paste(dose_folder,"analysis/csv/a_base.csv",sep=""),na.strings=c(NA,"NULL",""))
dose_base %>% mutate_if(is.factor,as.character) -> dose_base

dose_base$yihf_months <- dose_base$YIHF*12
dose_base$RACE[dose_base$ETHNIC==1] <- 8

dose_base_melt <- melt(data=dose_base,id.vars=c("PATNUMB"),na.rm=T) 
dose_base_melt %>% mutate_if(is.factor,as.character) -> dose_base_melt

dose_base_melt$studyvisit <- "BASELINE"
colnames(dose_base_melt)[1] <- "patnumb"
dose_base_melt$form <- "a_base"


##### analysis/sas/a_visitsumm #####

dose_visitsumm <- read.csv(paste(dose_folder,"analysis/csv/a_visitsumm.csv",sep=""),na.strings=c(NA,"NULL",""))
dose_visitsumm %>% mutate_if(is.factor,as.character) -> dose_visitsumm
dose_visitsumm$studyvisit <- dose_visitsumm$FORM

dose_hgt <- dose_visitsumm[dose_visitsumm$FORM=="BASELINE", c("PATNUMB","HEIGHTIN")]

dose_hgt$height_cm <- round(dose_hgt$HEIGHTIN * 2.54,1)

dose_visitsumm$alc <- dose_visitsumm$LL_LYMPH/dose_visitsumm$LL_WBC
dose_visitsumm$hct_calc <- dose_visitsumm$LL_HMG*3
dose_visitsumm$tni_ng_ml <- dose_visitsumm$CL_TROPI/1000
dose_visitsumm$weight_kg <- dose_visitsumm$WTLBS/2.2
dose_visitsumm <- merge(dose_visitsumm,dose_hgt[,c('PATNUMB','height_cm')])

dose_visitsumm$bmi <- round(dose_visitsumm$weight_kg/((dose_visitsumm$height_cm/100)^2),1)

dose_visitsumm_melt <- subset(melt(data=dose_visitsumm,id.vars=c("PATNUMB","studyvisit"),na.rm=T,stringsAsFactors=F))
dose_visitsumm_melt %>% mutate_if(is.factor,as.character) -> dose_visitsumm_melt

names(dose_visitsumm_melt)[1] <- "patnumb"
dose_visitsumm_melt$form <- "a_visitsumm"


##########  data/sas/meds  ##########

dose_meds <- read.csv(paste(dose_folder,"data/csv/meds.csv",sep=""),na.strings=c(NA,"NULL",""))
dose_meds %>% mutate_if(is.factor,as.character) -> dose_meds

dose_meds_adm <- sqldf("select patnumb,
                       'BASELINE' as studyvisit,
                       HFMEDS as variable,
                       MEDRAND as value
                       from dose_meds
                       where FORM = 'INDEXHOSP'")

dose_meds_dc <- sqldf("select patnumb,
                       'DAY7' as studyvisit,
                       HFMEDS as variable,
                       MEDDSCG as value
                       from dose_meds
                       where FORM = 'INDEXHOSP'")

dose_meds_day60 <- sqldf("select patnumb,
                       'DAY60' as studyvisit,
                       HFMEDS as variable,
                       MEDSANS as value
                       from dose_meds
                       where FORM = 'DAY60'")


dose_meds_melt <- rbind(dose_meds_adm,
                        dose_meds_dc,
                        dose_meds_day60)

dose_meds_melt %>% mutate_if(is.factor,as.character) -> dose_meds_melt


dose_meds_melt$form <- "meds"


##########  data/sas/diuretic  ##########

dose_diuretic <- read.csv(paste(dose_folder,"data/csv/diuretic.csv",sep=""),na.strings=c(NA,"NULL",""))
dose_diuretic %>% mutate_if(is.factor,as.character) -> dose_diuretic
dose_diuretic[dose_diuretic=="NaN"] <- NA

dose_diuretic$furo_equiv[dose_diuretic$DIUMEDS==1] <- dose_diuretic$DIURDOSE[dose_diuretic$DIUMEDS==1]*1
dose_diuretic$furo_equiv[dose_diuretic$DIUMEDS==2] <- dose_diuretic$DIURDOSE[dose_diuretic$DIUMEDS==2]*2
dose_diuretic$furo_equiv[dose_diuretic$DIUMEDS==3] <- dose_diuretic$DIURDOSE[dose_diuretic$DIUMEDS==3]*40

dose_diuretic %>% mutate_if(is.factor,as.character) -> dose_diuretic

dose_diuretic$variable <- dose_diuretic$DIUMEDS
dose_diuretic$value <- dose_diuretic$DIURANS
dose_diuretic$studyvisit <- dose_diuretic$FORM


##### Daily furosemide dose #####

dose_daily_furo_melt <- sqldf("select patnumb,
                         FORM,
                         sum(furo_equiv) as value
                         from dose_diuretic
                         group by patnumb, FORM")

dose_daily_furo_melt %>% mutate_if(is.factor,as.character) -> dose_daily_furo_melt

dose_daily_furo_melt$studyvisit <- dose_daily_furo_melt$FORM
dose_daily_furo_melt$variable <- "daily_furo_eq_derive"
dose_daily_furo_melt$form <- "diuretic"
dose_diuretic$form <- "diuretic"


##### data/sas/medhist1 #####

dose_medhist1 <- read.csv(paste(dose_folder,"data/csv/medhist1.csv",sep=""),na.strings=c(NA,"NULL",""))
dose_medhist1 %>% mutate_if(is.factor,as.character) -> dose_medhist1

dose_cms <- names(dose_medhist1) %in% c("ALCOHOLC",
                                          "CYTOTOXC",
                                          "FAMILIAL",
                                          "HCM",
                                          "HYPERTEN",
                                          "DILATED",
                                          "RESTRICT",
                                          "OTHCONT",
                                          "PERIPAR",
                                          "VAL")

dose_medhist1[dose_medhist1$ISCHEMIC==1,dose_cms] <- NA


dose_medhist1$num_etiol <- apply(dose_medhist1[dose_cms],
                                  MARGIN=1,
                                  FUN=function(x)
                                    sum(x,na.rm=T))

dose_medhist1[dose_medhist1$num_etiol>1,dose_cms] <- NA


dose_medhist1_melt <- subset(melt(data=dose_medhist1,id.vars=c("patnumb"),na.rm=T))

dose_medhist1_melt %>% mutate_if(is.factor,as.character) -> dose_medhist1_melt

dose_medhist1_melt$studyvisit <- "BASELINE"
dose_medhist1_melt$form <- "medhist1"


##########  data/sas/medhist2  ##########

dose_medhist2 <- read.csv(paste(dose_folder,"data/csv/medhist2.csv",sep=""))
dose_medhist2 %>% mutate_if(is.factor,as.character) -> dose_medhist2

dose_medhist2_melt <- subset(melt(data=dose_medhist2,id.vars=c("patnumb"),na.rm=T))
dose_medhist2_melt %>% mutate_if(is.factor,as.character) -> dose_medhist2_melt
dose_medhist2_melt$studyvisit <- "BASELINE"
dose_medhist2_melt$form <- "medhist2"


##### data/sas/assessmt #####

dose_assessmt <- read.csv(paste(dose_folder,"data/csv/assessmt.csv",sep=""),na.strings=c(NA,"NULL",""))
dose_assessmt %>% mutate_if(is.factor,as.character) -> dose_assessmt

dose_assessmt$pupr <- dose_assessmt$BPSYS - dose_assessmt$BPDIA

dose_assessmt_melt <- subset(melt(data=dose_assessmt,id.vars=c("patnumb","FORM"),na.rm=T))
dose_assessmt_melt %>% mutate_if(is.factor,as.character) -> dose_assessmt_melt

dose_assessmt_melt$studyvisit <- dose_assessmt_melt$FORM
dose_assessmt_melt$form <- "assessmt"


##### data/sas/ecg #####

dose_ecg <- read.csv(paste(dose_folder,"data/csv/ecg.csv",sep=""),na.strings=c(NA,"NULL",""),stringsAsFactors = F)
dose_ecg %>% mutate_if(is.factor,as.character) -> dose_ecg

dose_ecg_melt <- subset(melt(data=dose_ecg,id.vars=c("patnumb"),na.rm=T))
dose_ecg_melt %>% mutate_if(is.factor,as.character) -> dose_ecg_melt

dose_ecg_melt$studyvisit <- "BASELINE"
dose_ecg_melt$form <- "ecg"


####### data/sas/subjsymp #####

dose_subjsymp <- read.csv(paste(dose_folder,"data/csv/subjsymp.csv",sep=""),na.strings=c(NA,"NULL",""),stringsAsFactors = F)
dose_subjsymp[dose_subjsymp=="NaN"] <- NA

dose_subjsymp$studyvisit[dose_subjsymp$SELFREPT==1] <- "BASELINE"
dose_subjsymp$studyvisit[dose_subjsymp$SELFREPT==2] <- "6HOUR"
dose_subjsymp$studyvisit[dose_subjsymp$SELFREPT==3] <- "12HOUR"
dose_subjsymp$studyvisit[dose_subjsymp$SELFREPT==4] <- "24HOUR"
dose_subjsymp$studyvisit[dose_subjsymp$SELFREPT==5] <- "48HOUR"
dose_subjsymp$studyvisit[dose_subjsymp$SELFREPT==6] <- "72HOUR"
dose_subjsymp$studyvisit[dose_subjsymp$SELFREPT==7] <- "96HOUR"


dose_subjsymp_melt <- melt(dose_subjsymp[,c("patnumb","studyvisit","VASDYSPN","VASGLOBL")],
                           id.vars=c("patnumb","studyvisit"),
                           na.rm=T)

dose_subjsymp_melt$form <- "subjsymp"


####### data/sas/fluid.sas7bdat ######

dose_fluid <- read.csv(paste(dose_folder,"data/csv/fluid.csv",sep=""),na.strings=c(NA,"NULL",""),stringsAsFactors = F)
dose_fluid[dose_fluid=="NaN"] <- NA


dose_fluid$io_balance <- dose_fluid$TOTALIN-dose_fluid$TOTALOUT

dose_fluid_cum_d1 <- sqldf('select patnumb, 
                    io_balance as io_balancex24HOUR 
                    from dose_fluid 
                    where FORM = "24HOUR"') 

dose_fluid_cum_d2 <- sqldf('select patnumb, 
                    io_balance as io_balancex48HOUR 
                    from dose_fluid 
                    where FORM = "48HOUR"') 

dose_fluid_cum_d3 <- sqldf('select patnumb, 
                    io_balance as io_balancex72HOUR 
                    from dose_fluid 
                    where FORM = "72HOUR"') 

dose_fluid_cum_d4 <- sqldf('select patnumb, 
                    io_balance as io_balancex96HOUR 
                    from dose_fluid 
                    where FORM = "96HOUR"') 

dose_fluid_cum <- merge(dose_fluid_cum_d1,dose_fluid_cum_d2,by="patnumb",all.x=T)
dose_fluid_cum <- merge(dose_fluid_cum,dose_fluid_cum_d3,by="patnumb",all.x=T)
dose_fluid_cum <- merge(dose_fluid_cum,dose_fluid_cum_d4,by="patnumb",all.x=T)

dose_fluid_cum$io_cumx24HOUR <- dose_fluid_cum$io_balancex24HOUR
dose_fluid_cum$io_cumx48HOUR <- dose_fluid_cum$io_balancex24HOUR+dose_fluid_cum$io_balancex48HOUR
dose_fluid_cum$io_cumx72HOUR <- dose_fluid_cum$io_cumx48HOUR+dose_fluid_cum$io_balancex72HOUR
dose_fluid_cum$io_cumx96HOUR <- dose_fluid_cum$io_cumx72HOUR+dose_fluid_cum$io_balancex96HOUR

dose_fluid_cum_melt <- melt(dose_fluid_cum,
                              id.vars=c("patnumb"),
                              na.rm=T)

dose_fluid_cum_melt$studyvisit <- str_select(dose_fluid_cum_melt$variable,after="x")
dose_fluid_cum_melt$variable <- str_select(dose_fluid_cum_melt$variable,before="x")

dose_fluid_cum_melt$form <- "fluid"


##########  compile  ##########

dose_melt <- rbind(dose_base_melt[,c("patnumb","studyvisit","variable","value","form")],
                   dose_visitsumm_melt[,c("patnumb","studyvisit","variable","value","form")],
                   dose_medhist1_melt[,c("patnumb","studyvisit","variable","value","form")],
                   dose_medhist2_melt[,c("patnumb","studyvisit","variable","value","form")],
                   dose_ecg_melt[,c("patnumb","studyvisit","variable","value","form")],
                   dose_assessmt_melt[,c("patnumb","studyvisit","variable","value","form")],
                   dose_diuretic[,c("patnumb","studyvisit","variable","value","form")],
                   dose_daily_furo_melt[,c("patnumb","studyvisit","variable","value","form")],
                   dose_meds_melt[,c("patnumb","studyvisit","variable","value","form")],
                   dose_fluid_cum_melt[,c("patnumb","studyvisit","variable","value","form")],
                   dose_subjsymp_melt[,c("patnumb","studyvisit","variable","value","form")])

names(dose_melt)[1] <- "patientid"
dose_melt$study <- "DOSE"


########################## **********ESCAPE********** ################################

# .%%%%%%%%..%%%%%%...%%%%%%.....%%%....%%%%%%%%..%%%%%%%%
# .%%.......%%....%%.%%....%%...%%.%%...%%.....%%.%%......
# .%%.......%%.......%%........%%...%%..%%.....%%.%%......
# .%%%%%%....%%%%%%..%%.......%%.....%%.%%%%%%%%..%%%%%%..
# .%%.............%%.%%.......%%%%%%%%%.%%........%%......
# .%%.......%%....%%.%%....%%.%%.....%%.%%........%%......
# .%%%%%%%%..%%%%%%...%%%%%%..%%.....%%.%%........%%%%%%%%


##########  analdata/patient  ##########

escape_patient <- read.csv(paste(escape_folder,"main/analdata/patient.csv",sep=""),stringsAsFactors = F)

escape_patient$height_cm <- escape_patient$HGHTM*100

escape_patient_melt <- melt(data=escape_patient,id.vars=c("DEIDNUM"),na.rm=T) 
escape_patient_melt$studyvisit <- "Baseline"
escape_patient_melt$variable <- as.character(escape_patient_melt$variable)
escape_patient_melt$form <- "patient"

########## sasdata/symptoms #########

escape_symptoms <- read.csv(paste(escape_folder,"main/sasdata/symptoms.csv",sep=""),
                         stringsAsFactors = F,
                         na.strings=c("","NA","NULL"))

escape_symptoms$fatigue_congest <- 
  escape_symptoms$FANYACT + 
  escape_symptoms$FDAYACT + 
  escape_symptoms$FREST

escape_symptoms$dyspnea_congest <- 
  escape_symptoms$DWALKRM + 
  escape_symptoms$DWALKBK + 
  escape_symptoms$DREST

escape_symptoms$PHYSDAY[escape_symptoms$FORM=="Baseline"] <- 0 
escape_symptoms$PHYSDAY[escape_symptoms$FORM=="Discharge"] <- 7 

escape_symptoms$studyvisit[!is.na(escape_symptoms$PHYSDAY)] <- paste("DAY",escape_symptoms$PHYSDAY[!is.na(escape_symptoms$PHYSDAY)] ,sep="")
escape_symptoms$studyvisit[is.na(escape_symptoms$PHYSDAY)] <- escape_symptoms$FORM[is.na(escape_symptoms$PHYSDAY)]


escape_symptoms_melt <- melt(data=escape_symptoms,
                          id.vars=c("DEIDNUM","FORM","studyvisit"),
                          na.rm=T,
                          stringsAsFactors=F)

escape_symptoms_melt$variable <- as.character(escape_symptoms_melt$variable)
escape_symptoms_melt$form <- "symptoms"


##########  sasdata/socio  ##########

escape_socio <- read.csv(paste(escape_folder,"main/sasdata/socio.csv",sep=""),
                           stringsAsFactors = F,
                           na.strings=c("","NA","NULL"))


escape_socio_melt <- melt(data=escape_socio,
                            id.vars=c("DEIDNUM","FORM"),
                            na.rm=T,
                            stringsAsFactors=F)

escape_socio_melt$variable <- as.character(escape_socio_melt$variable)
escape_socio_melt$studyvisit <- escape_socio_melt$FORM
escape_socio_melt$form <- "socio"


##########  analdata/mhist  ##########

escape_mhist <- read.csv(paste(escape_folder,"main/analdata/mhist.csv",sep=""),stringsAsFactors = F,na.strings=c(NA,"NULL",""))

escape_mhist_melt <- melt(data=escape_mhist,id.vars=c("DEIDNUM"),na.rm=T) 
escape_mhist_melt$studyvisit <- "Baseline"
escape_mhist_melt$variable <- as.character(escape_mhist_melt$variable)
escape_mhist_melt$form <- "mhist"


##########  sasdata/lhcath  ##########

escape_lhcath <- read.csv(paste(escape_folder,"main/sasdata/lhcath.csv",sep=""),stringsAsFactors = F)

escape_lhcath$STENLM[is.na(escape_lhcath$STENLM)] <- 0
escape_lhcath$STENLAD[is.na(escape_lhcath$STENLAD)] <- 0
escape_lhcath$STENLCX[is.na(escape_lhcath$STENLCX)] <- 0
escape_lhcath$STENRCA[is.na(escape_lhcath$STENRCA)] <- 0

escape_lhcath_melt <- melt(data=escape_lhcath,id.vars=c("DEIDNUM","FORM","LSTCTHDT"),na.rm=T) 
escape_lhcath_melt$studyvisit <- "Baseline"
escape_lhcath_melt$variable <- as.character(escape_lhcath_melt$variable)
escape_lhcath_melt$form <- "lhcath"


##########  sasdata/physexam  ##########

escape_physexam <- read.csv(paste(escape_folder,"main/sasdata/physexam.csv",sep=""),stringsAsFactors = F,na.strings=c(NA,"NULL",""))

escape_physexam <- merge(escape_physexam,
                         escape_patient[,c("DEIDNUM","HGHTM","height_cm")],
                         by="DEIDNUM",
                         all.x=T)

escape_physexam$pupr <- escape_physexam$SUPSYSBP-escape_physexam$SUPDIABP

escape_physexam$weight_kg[!is.na(escape_physexam$WTUNIT)&escape_physexam$WTUNIT==1] <- 
  escape_physexam$WT[!is.na(escape_physexam$WTUNIT)&escape_physexam$WTUNIT==1]/2.2

escape_physexam$weight_kg[!is.na(escape_physexam$WTUNIT)&escape_physexam$WTUNIT==2] <- 
  escape_physexam$WT[!is.na(escape_physexam$WTUNIT)&escape_physexam$WTUNIT==2]

escape_physexam$bmi <- round(escape_physexam$weight_kg/(escape_physexam$HGHTM^2),1)

escape_physexam$bsa <- 
  calc_bsa(
    dat=escape_physexam,
    weight_kg="weight_kg",
    height_cm = "height_cm")

escape_physexam$PHYSDAY[escape_physexam$FORM=="Baseline"] <- 0 
escape_physexam$PHYSDAY[escape_physexam$FORM=="Discharge"] <- 7 

escape_physexam$studyvisit[!is.na(escape_physexam$PHYSDAY)] <- paste("DAY",escape_physexam$PHYSDAY[!is.na(escape_physexam$PHYSDAY)] ,sep="")
escape_physexam$studyvisit[is.na(escape_physexam$PHYSDAY)] <- escape_physexam$FORM[is.na(escape_physexam$PHYSDAY)]


escape_physexam_melt <- melt(data=escape_physexam,id.vars=c("DEIDNUM","studyvisit"),na.rm=T) 
escape_physexam_melt$variable <- as.character(escape_physexam_melt$variable)
escape_physexam_melt$form <- "physexam"


############ lab parts ##############


##########  sasdata/lab  ##########

escape_lab <- read.csv(paste(escape_folder,"main/sasdata/lab.csv",sep=""),stringsAsFactor=F,na.strings=c(NA,"NULL",""))

escape_lab <- subset(escape_lab,!is.na(LABVAL))

escape_lab$value[escape_lab$LAB==1&
                    escape_lab$LABUNIT==1&
                    escape_lab$LABVAL > 50&
                    !is.na(escape_lab$LABUNIT)&
                    !is.na(escape_lab$LABVAL)] <- 
  escape_lab$LABVAL[escape_lab$LAB==1&
                      escape_lab$LABUNIT==1&
                      escape_lab$LABVAL > 50&
                      !is.na(escape_lab$LABUNIT)&
                      !is.na(escape_lab$LABVAL)]/10


escape_lab$value[escape_lab$LAB==3&
                    escape_lab$LABVAL<1&
                    !is.na(escape_lab$LABVAL)] <- 
  escape_lab$LABVAL[escape_lab$LAB==3&
                      escape_lab$LABVAL <1&
                      !is.na(escape_lab$LABVAL)]*100

escape_lab$value[escape_lab$LAB==8&
                    !is.na(escape_lab$LAB)&
                    escape_lab$LABVAL > 10&
                    !is.na(escape_lab$LABVAL)] <- 
  escape_lab$LABVAL[escape_lab$LAB==8&
                      !is.na(escape_lab$LAB)&
                      escape_lab$LABVAL>10&
                      !is.na(escape_lab$LABVAL)]/88.42

escape_lab$value[escape_lab$LAB==11&
                    escape_lab$LABVAL>20&
                    !is.na(escape_lab$LABVAL)] <- 
  escape_lab$LABVAL[escape_lab$LAB==11&
                      escape_lab$LABVAL>20&
                      !is.na(escape_lab$LABVAL)]/10


escape_lab$value[escape_lab$LAB==12&
                  escape_lab$LABVAL>10&
                  !is.na(escape_lab$LABVAL)] <- 
  escape_lab$LABVAL[escape_lab$LAB==12&
                      escape_lab$LABVAL>10&
                      !is.na(escape_lab$LABVAL)]/10


escape_lab$value[escape_lab$LAB==13&
                    escape_lab$LABUNIT==11&
                    !is.na(escape_lab$LABUNIT)&
                    !is.na(escape_lab$LABVAL)] <- 
  escape_lab$LABVAL[escape_lab$LAB==13&
                      escape_lab$LABUNIT==11&
                      !is.na(escape_lab$LABUNIT)&
                      !is.na(escape_lab$LABVAL)]/17


escape_lab$value[escape_lab$LAB==14&
                    escape_lab$LABUNIT==11&
                    !is.na(escape_lab$LABUNIT)&
                    !is.na(escape_lab$LABVAL)] <- 
  escape_lab$LABVAL[escape_lab$LAB==14&
                      escape_lab$LABUNIT==11&
                      !is.na(escape_lab$LABUNIT)&
                      !is.na(escape_lab$LABVAL)]/17


escape_lab$value[is.na(escape_lab$value)] <- escape_lab$LABVAL[is.na(escape_lab$value)]


escape_lab$value[escape_lab$LAB==3&escape_lab$LABVAL>70] <- NA 

escape_lab$variable <- escape_lab$LAB

escape_lab$studyvisit[is.na(escape_lab$LABDAY)] <- escape_lab$FORM[is.na(escape_lab$LABDAY)]

escape_lab$studyvisit[!is.na(escape_lab$LABDAY)&escape_lab$PERIOD==2&escape_lab$LABDAY==1] <- "DAY3" 
escape_lab$studyvisit[!is.na(escape_lab$LABDAY)&escape_lab$PERIOD==2&escape_lab$LABDAY==2] <- "DAY5" 
escape_lab$studyvisit[!is.na(escape_lab$LABDAY)&escape_lab$PERIOD==2&escape_lab$LABDAY==3] <- "DAY7" 
escape_lab$studyvisit[!is.na(escape_lab$LABDAY)&escape_lab$LABDAY==4] <- "DAYOPT" 


escape_lab <- merge(escape_lab,
                      escape_patient[,c('DEIDNUM',
                                        'AGE',
                                        'GENDER',
                                        'RACE')],
                      by="DEIDNUM",
                      all.x=T)


escape_renal <- subset(escape_lab,LAB==8&LABUNIT==9&!is.na(LABVAL))[,c("DEIDNUM","studyvisit","LAB","LABVAL")]


escape_renal <- merge(escape_renal,
                    escape_patient[,c('DEIDNUM',
                                      'AGE',
                                      'GENDER',
                                      'RACE')],
                    by="DEIDNUM",
                    all.x=T)

escape_renal$value <- calc_MDRD4(dat=escape_renal,
                               cr="LABVAL",
                               age="AGE",
                               sex="GENDER",
                               race="RACE")

escape_renal$value[escape_renal$value > 200] <- 200

escape_renal$variable <- "CrCl"

escape_lab_melt <- rbind(escape_lab[,c("DEIDNUM","studyvisit","variable","value")],
                         escape_renal[,c("DEIDNUM","studyvisit","variable","value")])


escape_lab_melt$form <- "lab"

##########  sasdata/enzymes  ##########

escape_enzymes <- read.csv(paste(escape_folder,"main/sasdata/enzymes.csv",sep=""),
                           stringsAsFactor=F)


escape_enzymes$trop_i[escape_enzymes$TROPTYP==1&
                        !is.na(escape_enzymes$TROPTYP)] <- 
  escape_enzymes$TROPVAL[escape_enzymes$TROPTYP==1&
                           !is.na(escape_enzymes$TROPTYP)]

escape_enzymes$trop_t[escape_enzymes$TROPTYP==2&
                        !is.na(escape_enzymes$TROPTYP)] <- 
  escape_enzymes$TROPVAL[escape_enzymes$TROPTYP==2&
                           !is.na(escape_enzymes$TROPTYP)]

escape_enzymes$ckmb_ngml[escape_enzymes$CKMBUNT==12&
                           !is.na(escape_enzymes$CKMBUNT)] <- 
  escape_enzymes$CKMBVAL[escape_enzymes$CKMBUNT==12&
                           !is.na(escape_enzymes$CKMBUNT)]

escape_enzymes$ckmb_ngml[escape_enzymes$CKMBUNT==7&
                           !is.na(escape_enzymes$CKMBUNT)] <- 
  escape_enzymes$CKMBVAL[escape_enzymes$CKMBUNT==7&
                           !is.na(escape_enzymes$CKMBUNT)] * 
  escape_enzymes$CKVAL[escape_enzymes$CKMBUNT==7&
                         !is.na(escape_enzymes$CKMBUNT)]/100

escape_enzymes$ckmb_index <- escape_enzymes$ckmb_ngml/escape_enzymes$CKVAL

escape_enzymes_melt <- melt(escape_enzymes[,c("DEIDNUM",
                                              "FORM",
                                              "CKVAL",
                                              "trop_i",
                                              "trop_t",
                                              "ckmb_ngml",
                                              "ckmb_index")],
                            id.vars=c("DEIDNUM","FORM"),
                            na.rm=T)

escape_enzymes_melt$form <- "enzymes"
escape_enzymes_melt$studyvisit <- escape_enzymes_melt$FORM


##########  mechanistic/sasdata/mayodata (natriuretic peptides)  ##########

escape_mayodata <- read.csv(paste(escape_folder,"mechanistic/sasdata/mayodata.csv",sep=""),
                            stringsAsFactor=F,
                            na.strings=c(NA,"NULL",""))
escape_mayodata[escape_mayodata==9999] <- NA


escape_mayodata_melt <- melt(data=escape_mayodata,
                             id.vars=c("DEIDNUM","VISIT","DRAWDT"),
                             na.rm=T,
                             stringsAsFactors=F)

escape_mayodata_melt$studyvisit[escape_mayodata_melt$VISIT==1] <- "Baseline"
escape_mayodata_melt$studyvisit[escape_mayodata_melt$VISIT==2] <- "Discharge"
escape_mayodata_melt$studyvisit[escape_mayodata_melt$VISIT==3] <- "3-Month Follow-up"
escape_mayodata_melt$studyvisit[escape_mayodata_melt$VISIT==4] <- "6-Month Follow-up"

escape_mayodata_melt$studyvisit[escape_mayodata_melt$DEIDNUM==28750&
                                  escape_mayodata_melt$DRAWDT==2] <- "DAY2"

escape_mayodata_melt$form <- "mayodata"


##########  sasdata/meds  ##########

escape_meds <- read.csv(paste(escape_folder,"main/sasdata/meds.csv",sep=""),
                     stringsAsFactors = F,
                     na.strings=c(NA,"NULL",""))

escape_meds_melt <- sqldf("select DEIDNUM,
                          FORM as studyvisit,
                          MED as variable,
                          MEDRSP as value
                          from escape_meds")


escape_meds_melt$variable <- as.character(escape_meds_melt$variable)
escape_meds_melt$form <- "meds"


##########  sasdata/bbantiar  ##########

escape_bbantiar <- read.csv(paste(escape_folder,"main/sasdata/bbantiar.csv",sep=""),
                        stringsAsFactors = F,
                        na.strings=c("","NA","NULL"))


escape_bbantiar_melt <- melt(data=escape_bbantiar,
                             id.vars=c("DEIDNUM","FORM"),
                             na.rm=T,
                             stringsAsFactors=F)

escape_bbantiar_melt$variable <- as.character(escape_bbantiar_melt$variable)
escape_bbantiar_melt$studyvisit <- escape_bbantiar_melt$FORM
escape_bbantiar_melt$form <- "bbantiar"


##########  sasdata/diurccb  ##########

escape_diurccb <- read.csv(paste(escape_folder,"main/sasdata/diurccb.csv",sep=""),
                            stringsAsFactors = F,
                            na.strings=c("","NA","NULL"))


escape_diurccb_melt <- melt(data=escape_diurccb,
                             id.vars=c("DEIDNUM","FORM"),
                             na.rm=T,
                             stringsAsFactors=F)

escape_diurccb_melt$variable <- as.character(escape_diurccb_melt$variable)
escape_diurccb_melt$studyvisit <- escape_diurccb_melt$FORM
escape_diurccb_melt$form <- "diurccb"


##########  sasdata/ace1  ##########

escape_ace1 <- read.csv(paste(escape_folder,"main/sasdata/ace1.csv",sep=""),
                           stringsAsFactors = F,
                           na.strings=c("","NA","NULL"))


escape_ace1_melt <- melt(data=escape_ace1,
                            id.vars=c("DEIDNUM","FORM"),
                            na.rm=T,
                            stringsAsFactors=F)

escape_ace1_melt$variable <- as.character(escape_ace1_melt$variable)
escape_ace1_melt$studyvisit <- escape_ace1_melt$FORM
escape_ace1_melt$form <- "ace1"


##########  sasdata/ace2  ##########

escape_ace2 <- read.csv(paste(escape_folder,"main/sasdata/ace2.csv",sep=""),
                        stringsAsFactors = F,
                        na.strings=c("","NA","NULL"))


escape_ace2_melt <- melt(data=escape_ace2,
                         id.vars=c("DEIDNUM","FORM"),
                         na.rm=T,
                         stringsAsFactors=F)

escape_ace2_melt$variable <- as.character(escape_ace2_melt$variable)
escape_ace2_melt$studyvisit <- escape_ace2_melt$FORM
escape_ace2_melt$form <- "ace2"


##########  sasdata/nihypot  ##########

escape_nihypot <- read.csv(paste(escape_folder,"main/sasdata/nihypot.csv",sep=""),
                            stringsAsFactors = F,
                            na.strings=c("","NA","NULL"))


escape_nihypot_melt <- melt(data=escape_nihypot,
                             id.vars=c("DEIDNUM","FORM"),
                             na.rm=T,
                             stringsAsFactors=F)

escape_nihypot_melt$variable <- as.character(escape_nihypot_melt$variable)
escape_nihypot_melt$studyvisit <- escape_nihypot_melt$FORM
escape_nihypot_melt$form <- "nihypot"


##########  sasdata/angiodig  ##########

escape_angiodig <- read.csv(paste(escape_folder,"main/sasdata/angiodig.csv",sep=""),
                           stringsAsFactors = F,
                           na.strings=c("","NA","NULL"))


escape_angiodig_melt <- melt(data=escape_angiodig,
                            id.vars=c("DEIDNUM","FORM"),
                            na.rm=T,
                            stringsAsFactors=F)

escape_angiodig_melt$variable <- as.character(escape_angiodig_melt$variable)
escape_angiodig_melt$studyvisit <- escape_angiodig_melt$FORM
escape_angiodig_melt$form <- "angiodig"


##########  sasdata/diur1  ##########

escape_diur1 <- read.csv(paste(escape_folder,"main/sasdata/diur1.csv",sep=""),
                        stringsAsFactors = F,
                        na.strings=c(NA,"NULL",""))

escape_diur1$FUROSEDS[is.na(escape_diur1$FUROSEDS)] <- 0
escape_diur1$TORSEDS[is.na(escape_diur1$TORSEDS)] <- 0
escape_diur1$BUMETADS[is.na(escape_diur1$BUMETADS)] <- 0
escape_diur1$ETHACRDS[is.na(escape_diur1$ETHACRDS)] <- 0

escape_diur1$daily_furo_eq_derive <- escape_diur1$FUROSEDS+
                                     escape_diur1$TORSEDS*2+
                                     escape_diur1$BUMETADS*40+
                                     escape_diur1$ETHACRDS*0.8

## Set outliers to max recommended 600 mg/day 

escape_diur1$daily_furo_eq_derive[escape_diur1$daily_furo_eq_derive>600] <- 600


escape_diur1_melt <- melt(escape_diur1,
                          id.vars=c("DEIDNUM",
                                   "FORM"),
                          na.rm=T)

escape_diur1_melt <- subset(escape_diur1_melt,!is.na(value))
escape_diur1_melt$studyvisit <- escape_diur1_melt$FORM

escape_diur1_melt$form <- "diur1"


##########  sasdata/echo_dat  ##########

escape_echo_dat <- read.csv(paste(escape_folder,"main/sasdata/echo_dat.csv",sep=""),
                            stringsAsFactors = F,
                            na.strings=c("","NA","NULL",999))


escape_echo <- read.csv(paste(escape_folder,"main/sasdata/echo.csv",sep=""),
                            stringsAsFactors = F,
                            na.strings=c("","NA","NULL",999))


escape_echo_dat$studyvisit[escape_echo_dat$TIME==0] <- "Baseline"
escape_echo_dat$studyvisit[escape_echo_dat$TIME==1] <- "Discharge"
escape_echo_dat$studyvisit[escape_echo_dat$TIME==2] <- "3-Month Follow-Up"

escape_echo_dat <- merge(escape_echo_dat,escape_patient[,c("DEIDNUM","GENDER","height_cm")],by="DEIDNUM",all.x=T)
escape_echo_dat <- merge(escape_echo_dat,
                         escape_physexam[,c("DEIDNUM","FORM","weight_kg")],
                         by.x=c("DEIDNUM","studyvisit"), 
                         by.y=c("DEIDNUM","FORM"),
                         all.x=T,
                         all.y=F)

escape_echo_dat$RAP[escape_echo_dat$IVCMAXE<=2.1&escape_echo_dat$IVCPULS>50] <- 3
escape_echo_dat$RAP[escape_echo_dat$IVCMAXE<=2.1&escape_echo_dat$IVCPULS<50] <- 8
escape_echo_dat$RAP[escape_echo_dat$IVCMAXE>2.1&escape_echo_dat$IVCPULS>50] <- 8
escape_echo_dat$RAP[escape_echo_dat$IVCMAXE>2.1&escape_echo_dat$IVCPULS<50] <- 13

escape_echo_dat$TRV[escape_echo_dat$TRV > 5&
               !is.na(escape_echo_dat$TRV)] <- 
  round(escape_echo_dat$TRV[escape_echo_dat$TRV > 5&
                 !is.na(escape_echo_dat$TRV)]/100,2)

escape_echo_dat$RVSP <- escape_echo_dat$RAP+4*escape_echo_dat$TRV^2

escape_echo_dat <- calc_hypertrophy_type(df=escape_echo_dat,
                                       id="DEIDNUM",
                                       sex="GENDER",
                                       male=1,
                                       female=2,
                                       lvedd_cm="EDD",
                                       ivsd_cm = "IVST",
                                       lvpwtd_cm = "PWT",
                                       height_cm="height_cm",
                                       weight_kg="weight_kg")


escape_echo_dat <- merge(escape_echo_dat,
                         escape_echo,
                         by.x=c("DEIDNUM","studyvisit","ECHODT"),
                         by.y=c("DEIDNUM","FORM","ECHODT"),
                         all.x=T)

escape_echo_dat$rveda_index <- escape_echo_dat$RVD/escape_echo_dat$bsa

escape_echo_dat$rnme <- rownames(escape_echo_dat)

escape_echo_dat$EF[!is.na(escape_echo_dat$EF)&escape_echo_dat$EF==0] <- NA

## This bit removes echos performed on the same day that cannot otherwise be distinguished.  
## It will favor studies with fewer missing values.

rmv_rws <- c(9,
             23,
             25,
             26,
             37,
             41,
             92,
             99,
             102,
             103,
             184,
             255,
             268,
             285,
             309,
             358,
             387,
             460,
             466,
             468,
             469,
             512,
             528,
             552,
             571,
             572)

escape_echo_dat <- subset(escape_echo_dat, !(DEIDNUM==13100&EDD==8.50))

escape_echo_dat[escape_echo_dat$DEIDNUM==3459&
                  escape_echo_dat$studyvisit=="Discharge"&
                  escape_echo_dat$MEANE==93.67,
                c("IVST",
                     "EDD",
                     "PWT",
                     "ESD",
                     "lvmass",
                     "lvmass_ix",
                     "rwt",
                     "lvh_type")] <- 
  escape_echo_dat[escape_echo_dat$DEIDNUM==3459&
                    escape_echo_dat$studyvisit=="Discharge"&
                    escape_echo_dat$MEANE==45.67,
                  c("IVST",
                       "EDD",
                       "PWT",
                       "ESD",
                       "lvmass",
                       "lvmass_ix",
                       "rwt",
                       "lvh_type")]


escape_echo_dat <- subset(escape_echo_dat,!rnme %in% rmv_rws)

escape_echo_dat_melt <- 
  melt(data=escape_echo_dat,id.vars=c("DEIDNUM","studyvisit","TIME"),na.rm=T) 
escape_echo_dat_melt$variable <- as.character(escape_echo_dat_melt$variable)
escape_echo_dat_melt$form <- "echo_dat"


##########  sasdata/clinhis1  ##########

escape_clinhis1 <- read.csv(paste(escape_folder,"main/sasdata/clinhis1.csv",sep=""),
                            stringsAsFactors = F,
                            na.strings=c("","NA","NULL"))

escape_clinhis1$YIHF_months <- abs(round(escape_clinhis1$INDIAGDT/-30.42,1))

escape_clinhis1_melt <- melt(data=escape_clinhis1,
                             id.vars=c("DEIDNUM","FORM"),
                             na.rm=T,
                             stringsAsFactors=F)

escape_clinhis1_melt$variable <- as.character(escape_clinhis1_melt$variable)
escape_clinhis1_melt$studyvisit <- escape_clinhis1_melt$FORM
escape_clinhis1_melt$form <- "clinhis1"


##########  sasdata/clinhis2  ##########

escape_clinhis2 <- read.csv(paste(escape_folder,"main/sasdata/clinhis2.csv",sep=""),
                            stringsAsFactors = F,
                            na.strings=c("","NA","NULL"))

escape_clinhis2$variable <- escape_clinhis2$ETIOHF
escape_clinhis2$value <- escape_clinhis2$ETIOHFRK
escape_clinhis2$studyvisit <- escape_clinhis2$FORM
escape_clinhis2$form <- "clinhis2"

escape_clinhis2_melt <- escape_clinhis2[,c("DEIDNUM","studyvisit","form","variable","value")]


##########  sasdata/clinhis3  ##########

escape_clinhis3 <- read.csv(paste(escape_folder,"main/sasdata/clinhis3.csv",sep=""),
                            stringsAsFactors = F,
                            na.strings=c(NA,"NULL",""))
escape_clinhis3[escape_clinhis3==""|escape_clinhis3=="NaN"] <- NA

escape_clinhis3_melt <- sqldf("select DEIDNUM,
                              FORM,
                              DOCHIST as variable,
                              DOCHISYN as value
                              from escape_clinhis3")

escape_clinhis3_melt$form <- "clinhis3"
escape_clinhis3_melt$studyvisit <- escape_clinhis3_melt$FORM


##########  sasdata/ecg ##########

escape_ecg <- read.csv(paste(escape_folder,"main/sasdata/ecg.csv",sep=""),
                       stringsAsFactors = F,
                       na.strings=c(NA,"NULL",""))
escape_ecg[escape_ecg==""|escape_ecg=="NaN"] <- NA

escape_ecg_melt <- melt(data=escape_ecg,
                             id.vars=c("DEIDNUM","FORM"),
                             na.rm=T,
                             stringsAsFactors=F)

escape_ecg_melt$variable <- as.character(escape_ecg_melt$variable)
escape_ecg_melt$studyvisit <- escape_ecg_melt$FORM
escape_ecg_melt$form <- "ecg"


##########  sasdata/cpx ##########

escape_cpx <- read.csv(paste(escape_folder,
                             "main/sasdata/cpx.csv",sep=""),
                       stringsAsFactors = F,
                       na.strings=c(NA,"NULL",""))

escape_cpx[escape_cpx==""|escape_cpx=="NaN"] <- NA

escape_cpx_melt <- melt(data=escape_cpx,
                        id.vars=c("DEIDNUM","FORM"),
                        na.rm=T,
                        stringsAsFactors=F)

escape_cpx_melt$variable <- as.character(escape_cpx_melt$variable)
escape_cpx_melt$studyvisit <- escape_cpx_melt$FORM
escape_cpx_melt$form <- "cpx"


##### sasdata/visual ######

escape_visual <- read.csv(paste(escape_folder,"main/sasdata/visual.csv",sep=""))
escape_visual[escape_visual=="NaN"] <- NA
escape_visual[escape_visual < 0] <- NA

escape_visual_melt <- melt(escape_visual,id.vars=c("DEIDNUM","FORM","VISUALDT"),na.rm=T)
escape_visual_melt$DEIDNUM <- as.numeric(escape_visual_melt$DEIDNUM)

escape_visual_melt$form <- "visual"
escape_visual_melt$studyvisit <- escape_visual_melt$FORM


##### sasdata/walk ######

escape_walk <- read.csv(paste(escape_folder,"main/sasdata/walk.csv",sep=""))
escape_walk[escape_walk=="NaN"] <- NA
escape_walk[escape_walk < 0] <- NA

escape_walk$sixmw_dist_m[!is.na(escape_walk$WALKUNIT)&escape_walk$WALKUNIT==1] <- escape_walk$WALKDIS[!is.na(escape_walk$WALKUNIT)&escape_walk$WALKUNIT==1]*0.3048
escape_walk$sixmw_dist_m[!is.na(escape_walk$WALKUNIT)&escape_walk$WALKUNIT==2] <- escape_walk$WALKDIS[!is.na(escape_walk$WALKUNIT)&escape_walk$WALKUNIT==2]

escape_walk_melt <- melt(escape_walk[,c("DEIDNUM",
                                        "FORM",
                                        "BORGSCOR",
                                        "sixmw_dist_m")],
                         id.vars=c("DEIDNUM",
                                   "FORM"),na.rm=T)
escape_walk_melt$DEIDNUM <- as.numeric(escape_walk_melt$DEIDNUM)

escape_walk_melt$form <- "walk"
escape_walk_melt$studyvisit <- escape_walk_melt$FORM


##### big/anal/merged ######

escape_merged <- read.csv(paste(escape_folder,"big/analdata/merged.csv",sep=""), 
                          stringsAsFactors=F,
                          na.strings=c(NA,"NULL",""))
escape_merged[escape_merged=="NaN"] <- NA
escape_merged[escape_merged < 0] <- NA
escape_merged$DEIDNUM <- as.numeric(escape_merged$DEIDNUM)

escape_merged$PCWP_mean_min <- pmax(escape_merged$PCWPAMN,
                                    escape_merged$PCWPMN,na.rm=T)

escape_merged$PVR <- (escape_merged$PAMEAN-escape_merged$PCWP_mean_min)/escape_merged$P_CO


escape_merged <- sqldf('select *
                        from escape_merged
                        group by DEIDNUM, CONTDT
                        having min(PCWP_mean_min)')


escape_merged <- sqldf('select *
                        from escape_merged
                        group by DEIDNUM, CONTDT
                        having max(P_CO)')


escape_merged$studyvisit[escape_merged$CONTDT==0] <- "DAY0" 
escape_merged$studyvisit[escape_merged$CONTDT==1] <- "DAY1" 
escape_merged$studyvisit[escape_merged$CONTDT==2] <- "DAY2" 
escape_merged$studyvisit[escape_merged$CONTDT==3] <- "DAY3" 
escape_merged$studyvisit[escape_merged$CONTDT==4] <- "DAY4" 
escape_merged$studyvisit[escape_merged$CONTDT==5] <- "DAY5" 
escape_merged$studyvisit[escape_merged$CONTDT==6] <- "DAY6" 
escape_merged$studyvisit[escape_merged$CONTDT==7] <- "DAY7" 

escape_merged$studyvisit[escape_merged$CONTACT=="2 week follow up"] <- "2-Week Follow-Up"
escape_merged$studyvisit[escape_merged$CONTACT=="1 month follow up"] <- "1-Month Follow-Up"
escape_merged$studyvisit[escape_merged$CONTACT=="2 month follow up"] <- "2-Month Follow-Up"
escape_merged$studyvisit[escape_merged$CONTACT=="3 month follow up"] <- "3-Month Follow-Up"
escape_merged$studyvisit[escape_merged$CONTACT=="6 month follow up"] <- "6-Month Follow-Up"

escape_merged <- escape_merged[,c("DEIDNUM",
                                  "studyvisit",
                                  "LCW",
                                  "LCWI",
                                  "PAMEAN",
                                  "PCWPAMN",
                                  "PCWPMN",
                                  "RAP",
                                  "P_CO",
                                  "P_CI",
                                  "MIXED",
                                  "P_SVR",
                                  "SI",
                                  "SV",
                                  "TFC",
                                  "TFI",
                                  "PCWP_mean_min")]

escape_merged$TFC[escape_merged$TFC>60] <- NA

escape_merged_melt <- melt(escape_merged,
                             id.vars=c("DEIDNUM","studyvisit"),
                             na.rm=T)

escape_merged_melt$form <- "merged"


##### sasdata/visual ######

escape_visual <- read.csv(paste(escape_folder,"main/sasdata/visual.csv",sep=""),
                          stringsAsFactors = F,
                          na.strings=c(NA,"NULL",""))
escape_visual[escape_visual=="NaN"] <- NA
escape_visual[escape_visual < 0] <- NA

escape_visual_melt <- melt(escape_visual,id.vars=c("DEIDNUM","FORM","VISUALDT"),na.rm=T)
escape_visual_melt$DEIDNUM <- as.numeric(escape_visual_melt$DEIDNUM)

escape_visual_melt$form <- "visual"
escape_visual_melt$studyvisit <- escape_visual_melt$FORM


##########  sasdata/hemo  ##########

escape_hemo <- read.csv(paste(escape_folder,"main/analdata/hemo.csv",sep=""),
                            stringsAsFactors = F,
                            na.strings=c("","NA","NULL"))


escape_hemo$PVRB <- round((escape_hemo$PAMNB-escape_hemo$PCWPB)/escape_hemo$COB,2)
escape_hemo$PVRND <- round((escape_hemo$PAMNND-escape_hemo$PCWPND)/escape_hemo$COND,2)
escape_hemo$PVRO <- round((escape_hemo$PAMNO-escape_hemo$PCWPO)/escape_hemo$COO,2)
escape_hemo$PVRL <- round((escape_hemo$PAMNL-escape_hemo$PCWPL)/escape_hemo$COL,2)

escape_hemo$PPB <- escape_hemo$BPSYSB - escape_hemo$BPDIASB
escape_hemo$PPND <- escape_hemo$BPSYSND - escape_hemo$BPDIASND
escape_hemo$PPO <- escape_hemo$BPSYSO - escape_hemo$BPDIASO
escape_hemo$PPL <- escape_hemo$BPSYSL - escape_hemo$BPDIASL

escape_hemo$MAPB <- round(escape_hemo$BPDIASB + 
                            0.3333 * escape_hemo$PPB,0)

escape_hemo$MAPND <- round(escape_hemo$BPDIASND + 
                            0.3333 * escape_hemo$PPND,0)

escape_hemo$MAPO <- round(escape_hemo$BPDIASO + 
                            0.3333 * escape_hemo$PPO)

escape_hemo$MAPL <- round(escape_hemo$BPDIASL + 
                            0.3333 * escape_hemo$PPL)

escape_hemo <- 
  merge(escape_hemo,
        escape_physexam[escape_physexam$FORM=="Baseline",
                        c("DEIDNUM","bsa")])

escape_hemo$SVIB <- escape_hemo$CIB/escape_hemo$HRTRTB
escape_hemo$SVIND <- escape_hemo$CIND/escape_hemo$HRTRTND
escape_hemo$SVIO <- escape_hemo$CIO/escape_hemo$HRTRTO
escape_hemo$SVIL <- escape_hemo$CIL/escape_hemo$HRTRTL

escape_hemo$arterial_elastance_ixB <- 0.9*escape_hemo$BPSYSB/escape_hemo$SVIB
escape_hemo$arterial_elastance_ixND <- 0.9*escape_hemo$BPSYSB/escape_hemo$SVIND
escape_hemo$arterial_elastance_ixO <- 0.9*escape_hemo$BPSYSB/escape_hemo$SVIO
escape_hemo$arterial_elastance_ixL <- 0.9*escape_hemo$BPSYSB/escape_hemo$SVIL

escape_hemo$syst_art_compB <- escape_hemo$SVIB/escape_hemo$PPB
escape_hemo$syst_art_compND <- escape_hemo$SVIND/escape_hemo$PPND
escape_hemo$syst_art_compO <- escape_hemo$SVIO/escape_hemo$PPO
escape_hemo$syst_art_compL <- escape_hemo$SVIL/escape_hemo$PPL

escape_hemo$SVRIB <- escape_hemo$SVRB/escape_hemo$bsa
escape_hemo$SVRIND <- escape_hemo$SVRND/escape_hemo$bsa
escape_hemo$SVRIO <- escape_hemo$SVRO/escape_hemo$bsa
escape_hemo$SVRIL <- escape_hemo$SVRL/escape_hemo$bsa


escape_hemo_melt <- melt(data=escape_hemo[,c(1,10:106)],
                             id.vars=c("DEIDNUM"),
                             na.rm=T,
                             stringsAsFactors=F)


escape_hemo_melt$variable <- as.character(escape_hemo_melt$variable)
escape_hemo_melt$var_temp[substr(escape_hemo_melt$variable,1,6)=="BPDIAS"] <- "BPDIAS"
escape_hemo_melt$var_temp[substr(escape_hemo_melt$variable,1,5)=="BPSYS"] <- "BPSYS"
escape_hemo_melt$var_temp[substr(escape_hemo_melt$variable,1,2)=="CI"] <- "CI"
escape_hemo_melt$var_temp[substr(escape_hemo_melt$variable,1,2)=="CO"] <- "CO"
escape_hemo_melt$var_temp[substr(escape_hemo_melt$variable,1,5)=="HRTRT"] <- "HRTRT"
escape_hemo_melt$var_temp[substr(escape_hemo_melt$variable,1,5)=="MIXED"] <- "MIXED"
escape_hemo_melt$var_temp[substr(escape_hemo_melt$variable,1,3)=="RAP"] <- "RAP"
escape_hemo_melt$var_temp[substr(escape_hemo_melt$variable,1,3)=="PAD"] <- "PAD"
escape_hemo_melt$var_temp[substr(escape_hemo_melt$variable,1,4)=="PAMN"] <- "PAMN"
escape_hemo_melt$var_temp[substr(escape_hemo_melt$variable,1,3)=="PAS"] <- "PAS"
escape_hemo_melt$var_temp[substr(escape_hemo_melt$variable,1,4)=="PCWP"] <- "PCWP"
escape_hemo_melt$var_temp[substr(escape_hemo_melt$variable,1,5)=="PCWPA"] <- "PCWPA"
escape_hemo_melt$var_temp[substr(escape_hemo_melt$variable,1,6)=="PCWPMN"] <- "PCWPMN"
escape_hemo_melt$var_temp[substr(escape_hemo_melt$variable,1,3)=="SVR"] <- "SVR"
escape_hemo_melt$var_temp[substr(escape_hemo_melt$variable,1,3)=="PVR"] <- "PVR"

escape_hemo_melt$studyvisit[substring(escape_hemo_melt$variable,
                                      nchar(escape_hemo_melt$variable),
                                      nchar(escape_hemo_melt$variable))=="B"] <- "Baseline"
escape_hemo_melt$studyvisit[substring(escape_hemo_melt$variable,
                                      nchar(escape_hemo_melt$variable)-1,
                                      nchar(escape_hemo_melt$variable))=="ND"] <- "DAY1"

escape_hemo_melt$studyvisit[substring(escape_hemo_melt$variable,
                                      nchar(escape_hemo_melt$variable),
                                      nchar(escape_hemo_melt$variable))=="O"] <- "DAYOPT"

escape_hemo_melt$studyvisit[substring(escape_hemo_melt$variable,
                                      nchar(escape_hemo_melt$variable),
                                      nchar(escape_hemo_melt$variable))=="L"] <- "DAYLAST"


escape_hemo_melt$variable <- escape_hemo_melt$var_temp

escape_hemo_melt$form <- "hemo"


##########  sasdata/hemodyna  ##########


escape_hemodyna <- read.csv("~/Dropbox/BioLINCC files/ESCAPE/data/main/sasdata/hemodyna.csv",
                            stringsAsFactor=F,
                            na.strings=c(NA,"NULL",""))


escape_hemodyna$PCWP_mean_min <- pmax(escape_hemodyna$PCWPAMN,escape_hemodyna$PCWPMN,na.rm=T)

escape_hemodyna <- sqldf('select *
                        from escape_hemodyna
                        group by DEIDNUM, HEMDT
                        having min(PCWP_mean_min)')


escape_hemodyna <- sqldf('select *
                        from escape_hemodyna
                        group by DEIDNUM, HEMDT
                        having max(COA)')


escape_hemodyna <- sqldf('select *
                        from escape_hemodyna
                        group by DEIDNUM, HEMDT
                        having min(HEMSEQ)')

escape_hemodyna$BPMEAN <- 
  escape_hemodyna$BPDIA+
  0.33333*(escape_hemodyna$BPSYS-escape_hemodyna$BPDIA)


escape_hemodyna <- escape_hemodyna[,c("DEIDNUM",
                                      "HEMDT",
                                      "PAD",
                                      "PAMEAN",
                                      "PCWPAMN",
                                      "PCWPMN",
                                      "RAP",
                                      "SVR",
                                      "COA",
                                      "CIA",
                                      "MIXED",
                                      "PCWP_mean_min",
                                      "BPMEAN",
                                      "HRTRT")]


escape_hemodyna_melt <- melt(escape_hemodyna,
                             id.vars=c("DEIDNUM","HEMDT"),
                             na.rm=T)
escape_hemodyna_melt <- subset(escape_hemodyna_melt,!is.na(HEMDT))

escape_hemodyna_melt$studyvisit[escape_hemodyna_melt$HEMDT >= 0] <- 
  paste("DAY",escape_hemodyna_melt$HEMDT[escape_hemodyna_melt$HEMDT >= 0],sep="")
escape_hemodyna_melt$form <- "hemodyna"


##### main/sasdata/lwhfques #######

escape_lwhfques <- read.csv(paste(escape_folder,"main/sasdata/lwhfques.csv",sep=""),
                            stringsAsFactors = F,
                            na.strings=c("NA",""))

escape_mlhf <- score_mlhf(dat=escape_lwhfques,
                          swelling = "SWELL", 
                          day_rest="REST",
                          stairs = "WALKC", 
                          housework="WORK",
                          go_places="AWAY",
                          sleep = "SLEEP",
                          friendfam = "FRIENDS",
                          earn_living = "DIFF",
                          hobbies = "HOBBIE",
                          sex = "SEX",
                          food = "EAT",
                          sob = "BREATH",
                          fatigue = "ENERGY",
                          hospital = "STAY",
                          cost = "COST",
                          sideeffect = "EFFECT",
                          burden = "BURDEN",
                          nocontrol = "LOSS",
                          worry="WORRY",
                          concentrate="REMEM",
                          depressed="DEPRS", 
                          total=c("Y"))


escape_mlhf_melt <- melt(data=escape_mlhf,
                         id.vars=c("DEIDNUM","FORM"),
                         na.rm=T)

escape_mlhf_melt$studyvisit <- escape_mlhf_melt$FORM
escape_mlhf_melt$form <- "lwhfques"


#### main/sasdata/volume ####

escape_volume <- read.csv("~/Dropbox/BioLINCC files/ESCAPE/data/main/sasdata/volume.csv",
                          stringsAsFactor=F,
                          na.strings=c(NA,"NULL",""))

escape_volume <- subset(escape_volume, VOLDT >= 0)

escape_volume$io_balance[escape_volume$VOLSIGN==1&
                           !is.na(escape_volume$VOLSIGN)] <- 
  escape_volume$VOLNET[escape_volume$VOLSIGN==1&
                         !is.na(escape_volume$VOLSIGN)]

escape_volume$io_balance[escape_volume$VOLSIGN==2&
                           !is.na(escape_volume$VOLSIGN)] <- 
  -escape_volume$VOLNET[escape_volume$VOLSIGN==2&
                          !is.na(escape_volume$VOLSIGN)]

escape_volume <- subset(escape_volume, !is.na(VOLDT))
escape_volume_melt <- sqldf("select DEIDNUM,
                            VOLDT,
                            'io_balance' as variable,
                            io_balance as value
                            from escape_volume")

escape_io_bl <- sqldf('select DEIDNUM,
                              value as io_balance_bl
                              from escape_volume_melt
                              where VOLDT = 0')

escape_io_1d <- sqldf('select DEIDNUM,
                              value as io_balance_1d
                              from escape_volume_melt
                              where VOLDT = 1')

escape_io_2d <- sqldf('select DEIDNUM,
                              value as io_balance_2d
                              from escape_volume_melt
                              where VOLDT = 2')

escape_io_3d <- sqldf('select DEIDNUM,
                              value as io_balance_3d
                              from escape_volume_melt
                              where VOLDT = 3')

escape_io_4d <- sqldf('select DEIDNUM,
                              value as io_balance_4d
                              from escape_volume_melt
                              where VOLDT = 4')

escape_io_5d <- sqldf('select DEIDNUM,
                              value as io_balance_5d
                              from escape_volume_melt
                              where VOLDT = 5')

escape_io_6d <- sqldf('select DEIDNUM,
                              value as io_balance_6d
                              from escape_volume_melt
                              where VOLDT = 6')

escape_io_7d <- sqldf('select DEIDNUM,
                              value as io_balance_7d
                              from escape_volume_melt
                              where VOLDT = 7')


acute_escape_io <- merge(escape_io_bl,escape_io_1d,by="DEIDNUM",all=T)
acute_escape_io <- merge(acute_escape_io,escape_io_2d,by="DEIDNUM",all=T)
acute_escape_io <- merge(acute_escape_io,escape_io_3d,by="DEIDNUM",all=T)
acute_escape_io <- merge(acute_escape_io,escape_io_4d,by="DEIDNUM",all=T)
acute_escape_io <- merge(acute_escape_io,escape_io_5d,by="DEIDNUM",all=T)
acute_escape_io <- merge(acute_escape_io,escape_io_6d,by="DEIDNUM",all=T)
acute_escape_io <- merge(acute_escape_io,escape_io_7d,by="DEIDNUM",all=T)

acute_escape_io$io_cum_1d <- apply(acute_escape_io[,c("io_balance_bl","io_balance_1d")],FUN=function(x) sum(x,na.rm=T), MARGIN=1)
acute_escape_io$io_cum_2d <- acute_escape_io$io_cum_1d+acute_escape_io$io_balance_2d
acute_escape_io$io_cum_3d <- acute_escape_io$io_cum_2d+acute_escape_io$io_balance_3d
acute_escape_io$io_cum_4d <- acute_escape_io$io_cum_3d+acute_escape_io$io_balance_4d
acute_escape_io$io_cum_5d <- acute_escape_io$io_cum_4d+acute_escape_io$io_balance_5d
acute_escape_io$io_cum_6d <- acute_escape_io$io_cum_5d+acute_escape_io$io_balance_6d
acute_escape_io$io_cum_7d <- acute_escape_io$io_cum_6d+acute_escape_io$io_balance_7d

acute_escape_io$io_cum_last <- acute_escape_io$io_cum_7d
acute_escape_io$io_cum_last[is.na(acute_escape_io$io_cum_last)] <-
  acute_escape_io$io_cum_6d[is.na(acute_escape_io$io_cum_last)]

acute_escape_io$io_cum_last[is.na(acute_escape_io$io_cum_last)] <-
  acute_escape_io$io_cum_5d[is.na(acute_escape_io$io_cum_last)]

acute_escape_io$io_cum_last[is.na(acute_escape_io$io_cum_last)] <-
  acute_escape_io$io_cum_4d[is.na(acute_escape_io$io_cum_last)]

acute_escape_io$io_cum_last[is.na(acute_escape_io$io_cum_last)] <-
  acute_escape_io$io_cum_3d[is.na(acute_escape_io$io_cum_last)]

acute_escape_io$io_cum_last[is.na(acute_escape_io$io_cum_last)] <-
  acute_escape_io$io_cum_2d[is.na(acute_escape_io$io_cum_last)]

acute_escape_io$io_cum_last[is.na(acute_escape_io$io_cum_last)] <-
  acute_escape_io$io_cum_1d[is.na(acute_escape_io$io_cum_last)]

escape_volume_cum_melt <- melt(acute_escape_io,
                            id.vars=c("DEIDNUM"),
                            na.rm=T)


escape_volume_cum_melt$variable1[str_detect(escape_volume_cum_melt$variable,"io_balance")] <- "io_balance"
escape_volume_cum_melt$variable1[str_detect(escape_volume_cum_melt$variable,"io_cum")] <- "io_cum"

escape_volume_cum_melt$studyvisit[str_detect(escape_volume_cum_melt$variable,"_bl")] <- "Baseline"
escape_volume_cum_melt$studyvisit[str_detect(escape_volume_cum_melt$variable,"_1d")] <- "DAY1"
escape_volume_cum_melt$studyvisit[str_detect(escape_volume_cum_melt$variable,"_2d")] <- "DAY2"
escape_volume_cum_melt$studyvisit[str_detect(escape_volume_cum_melt$variable,"_3d")] <- "DAY3"
escape_volume_cum_melt$studyvisit[str_detect(escape_volume_cum_melt$variable,"_4d")] <- "DAY4"
escape_volume_cum_melt$studyvisit[str_detect(escape_volume_cum_melt$variable,"_5d")] <- "DAY5"
escape_volume_cum_melt$studyvisit[str_detect(escape_volume_cum_melt$variable,"_6d")] <- "DAY6"
escape_volume_cum_melt$studyvisit[str_detect(escape_volume_cum_melt$variable,"_7d")] <- "DAY7"


escape_volume_cum_melt$form <- "fluid"

escape_volume_cum_melt$variable <- escape_volume_cum_melt$variable1

##########  compile  ##########

escape_melt <- rbind(escape_patient_melt[,c("DEIDNUM","form","studyvisit","variable","value")],
                     escape_angiodig_melt[,c("DEIDNUM","form","studyvisit","variable","value")],
                     escape_bbantiar_melt[,c("DEIDNUM","form","studyvisit","variable","value")],
                     escape_clinhis1_melt[,c("DEIDNUM","form","studyvisit","variable","value")],
                     escape_clinhis2_melt[,c("DEIDNUM","form","studyvisit","variable","value")],
                     escape_clinhis3_melt[,c("DEIDNUM","form","studyvisit","variable","value")],
                     escape_cpx_melt[,c("DEIDNUM","form","studyvisit","variable","value")],
                     escape_diurccb_melt[,c("DEIDNUM","form","studyvisit","variable","value")],
                     escape_diur1_melt[,c("DEIDNUM","form","studyvisit","variable","value")],
                     escape_meds_melt[,c("DEIDNUM","form","studyvisit","variable","value")],
                     escape_ecg_melt[,c("DEIDNUM","form","studyvisit","variable","value")],
                     escape_echo_dat_melt[,c("DEIDNUM","form","studyvisit","variable","value")],
                     escape_enzymes_melt[,c("DEIDNUM","form","studyvisit","variable","value")],
                     escape_hemo_melt[,c("DEIDNUM","form","studyvisit","variable","value")],
                     escape_hemodyna_melt[,c("DEIDNUM","form","studyvisit","variable","value")],
                     escape_lab_melt[,c("DEIDNUM","form","studyvisit","variable","value")],
                     escape_mayodata_melt[,c("DEIDNUM","form","studyvisit","variable","value")],
                     escape_mhist_melt[,c("DEIDNUM","form","studyvisit","variable","value")],
                     escape_merged_melt[,c("DEIDNUM","form","studyvisit","variable","value")],
                     escape_nihypot_melt[,c("DEIDNUM","form","studyvisit","variable","value")],
                     escape_physexam_melt[,c("DEIDNUM","form","studyvisit","variable","value")],
                     escape_socio_melt[,c("DEIDNUM","form","studyvisit","variable","value")],
                     escape_symptoms_melt[,c("DEIDNUM","form","studyvisit","variable","value")],
                     escape_mlhf_melt[,c("DEIDNUM","form","studyvisit","variable","value")],
                     escape_walk_melt[,c("DEIDNUM","form","studyvisit","variable","value")],
                     escape_visual_melt[,c("DEIDNUM","form","studyvisit","variable","value")],
                     escape_volume_cum_melt[,c("DEIDNUM","form","studyvisit","variable","value")])

escape_melt <- subset(escape_melt,!value %in% c(NA,"NaN","NULL"))

names(escape_melt)[1] <- "patientid"
escape_melt$study <- "ESCAPE"


############################## **********STICH**********  ############################

# ..%%%%%%..%%%%%%%%.%%%%..%%%%%%..%%.....%%
# .%%....%%....%%.....%%..%%....%%.%%.....%%
# .%%..........%%.....%%..%%.......%%.....%%
# ..%%%%%%.....%%.....%%..%%.......%%%%%%%%%
# .......%%....%%.....%%..%%.......%%.....%%
# .%%....%%....%%.....%%..%%....%%.%%.....%%
# ..%%%%%%.....%%....%%%%..%%%%%%..%%.....%%


##########  sourcedata/h1/demog  ##########

stich_demog <- read.csv(paste(stich_folder,"SourceData/h1/demog.csv",sep=""),
                        stringsAsFactors = F,
                        na.strings=c(".","","NA"))

stich_demog$race_new <- stich_demog$race
stich_demog$race_new[stich_demog$ETHNIC==1] <- "Hispanic"
stich_demog$pupr <- stich_demog$BPSYS-stich_demog$BPDIA

stich_demog_melt <- melt(data=stich_demog,id.vars=c("deidnum"),na.rm=T) 
stich_demog_melt$studyvisit <- "BASELINE"
stich_demog_melt$form <- "demog"


##########  analysisdata/h1/baseline  ##########

stich_baseline <- read.csv(paste(stich_folder,"AnalysisData/h1/baseline.csv",sep=""),
                     stringsAsFactors = F,
                     na.strings=c(".","","NA"))


stich_baseline2 <- read.csv(paste(stich_folder,"AnalysisData/h2/baseline.csv",sep=""),
                           stringsAsFactors = F,
                           na.strings=c(".","","NA"))


stich_baseline <- merge(stich_baseline,stich_demog[,c("deidnum","race")])

stich_baseline$hct_calc <- stich_baseline$hgb/0.34
stich_baseline$CrCl <- calc_MDRD4(stich_baseline,
                                  cr="crt",
                                  age="age",
                                  sex="female",
                                  male="0",
                                  race="race",
                                  black = "Black")

stich_baseline_melt <- melt(data=stich_baseline,id.vars=c("deidnum"),na.rm=T) 
stich_baseline_melt$studyvisit <- "BASELINE"
stich_baseline_melt$form <- "baseline"


##########  sourcedata/h1/medhist  ##########

stich_medhist <- read.csv(paste(stich_folder,"SourceData/h1/medhist.csv",sep=""),
                     stringsAsFactors = F,
                     na.strings=c(".","","NA"))

stich_medhist_melt <- melt(data=stich_medhist,id.vars=c("deidnum"),na.rm=T) 
stich_medhist_melt$studyvisit <- "BASELINE"
stich_medhist_melt$form <- "medhist"


##########  sourcedata/h1/angclass  ##########

stich_angclass <- read.csv(paste(stich_folder,"SourceData/h1/angclass.csv",sep=""),
                          stringsAsFactors = F,
                          na.strings=c(".","","NA"))

stich_angclass$pupr <- stich_angclass$HBPSYS-stich_angclass$HBPDIA

stich_angclass$studyvisit <- stich_angclass$FORM
stich_angclass_melt <- melt(data=stich_angclass,id.vars=c("deidnum","studyvisit"),na.rm=T) 
stich_angclass_melt$form <- "angclass"


##########  sourcedata/h1/corelab  ##########

stich_corelab <- read.csv(paste(stich_folder,"AnalysisData/h1/corelab.csv",sep=""),
                          stringsAsFactors = F,
                          na.strings=c(".","","NA"))

stich_corelab_melt <- melt(data=stich_corelab,id.vars=c("deidnum"),na.rm=T) 
stich_corelab_melt$studyvisit <- "BASELINE"
stich_corelab_melt$form <- "corelab"


##########  sourcedata/h1/cardcath  ##########

stich_cardcath <- read.csv(paste(stich_folder,"sourcedata/h1/cardcath.csv",sep=""),
                          stringsAsFactors = F,
                          na.strings=c(".","","NA"))

stich_cardcath_melt <- melt(data=stich_cardcath,id.vars=c("deidnum"),na.rm=T) 
stich_cardcath_melt$studyvisit <- "BASELINE"
stich_cardcath_melt$form <- "cardcath"


##########  sourcedata/h1/echodata  ##########


stich_echodata <- read.csv(paste(stich_folder,"SourceData/h1/echodata.csv",sep=""),
                     stringsAsFactors = F,
                     na.strings=c(".","","NA"))

stich_echodata <- merge(stich_echodata,
                        stich_baseline[,c("deidnum",
                                       "female",
                                       "weightkg",
                                       "heightcm")],
                        by="deidnum",all.x=T)

stich_echodata$ep_lat <- stich_echodata$ST_71*100
stich_echodata$ep_sept <- stich_echodata$ST_70*100
stich_echodata$ewave_val_cmsec <- stich_echodata$ST_67*100

stich_echodata$eeprime_lat <- round(stich_echodata$ST_67/stich_echodata$ST_71,1)
stich_echodata$eeprime_sept <- round(stich_echodata$ST_67/stich_echodata$ST_70,1)

stich_echodata$pvf_ratio <- stich_echodata$ST_72/stich_echodata$ST_73

stich_dd <- assign_diastolic_function(dat=stich_echodata, 
                                                      pid="deidnum",
                                                      vst="VISIT",
                                                ep_lat="ep_lat", 
                                                ep_sept="ep_sept", 
                                                earat="EA_RATIO", 
                                                dt="ST_69",
                                                def="TOPCAT")


stich_echodata <- merge(stich_echodata,
                        stich_dd,
                        by=c("deidnum","VISIT"),
                        all.x=T)

stich_echodata <- calc_hypertrophy_type(df=stich_echodata,
                                        sex="female",
                                        male="0",
                                        female="1",
                                        lvedd_cm="ST_21",
                                        ivsd_cm="ST_19",
                                        lvpwtd_cm="ST_20",
                                        height_cm="ST_4",
                                        weight_kg="ST_6")

stich_echodata$lvmass_echo <- stich_echodata$lvmass
stich_echodata$lvmass_ix_echo <- stich_echodata$lvmass_ix
stich_echodata$lav_ix_echo <- stich_echodata$ST_55/stich_echodata$bsa
stich_echodata$lvedd_ix_echo <- stich_echodata$ST_21/stich_echodata$bsa
stich_echodata$lvesd_ix_echo <- stich_echodata$ST_22/stich_echodata$bsa
stich_echodata$visualef_echo <- stich_echodata$ST_30 * 100
stich_echodata$qef_echo <- stich_echodata$ST_48 * 100
stich_echodata$lvedv_ix_echo <- stich_echodata$ST_46/stich_echodata$ST_8

stich_echodata$rveda_index_echo <- stich_echodata$ST_49/stich_echodata$bsa
stich_echodata$rvfac_echo <- round((stich_echodata$ST_49-stich_echodata$ST_50)/stich_echodata$ST_49,3)

stich_echodata <- calc_pulsatility(dat=stich_echodata,
                               sbp = "ST_9",
                               dbp = "ST_8",
                               edv = "ST_46",
                               esv = "ST_47",
                               bsa = "ST_8",
                               hr = "ST_10A",
                               lvot_d = "ST_62" ,
                               vti = "ST_61")

stich_echodata$studyvisit[stich_echodata$VISIT==1] <- "BASELINE"
stich_echodata$studyvisit[stich_echodata$VISIT==2] <- "4MONTH"
stich_echodata$studyvisit[stich_echodata$VISIT==3] <- "24MONTH"


stich_echodata_melt <- melt(data=stich_echodata,id.vars=c("deidnum","studyvisit"),na.rm=T) 
stich_echodata_melt$studyvisit <- stich_echodata_melt$studyvisit

stich_echodata_melt$form <- "echodata"


##########  cmrdata  ##########

stich_cmrdata <- read.csv(paste(stich_folder,"sourcedata/h1/cmrdata.csv",sep=""),
                          stringsAsFactor=F,
                          na.strings=c(".","NA",""))


stich_cmrdata$qef_mri <- stich_cmrdata$EF*100
stich_cmrdata$lvmass_mri <- stich_cmrdata$LVMASS
stich_cmrdata$lvmass_ix_mri <- stich_cmrdata$LVMASS/stich_cmrdata$BSA
stich_cmrdata$lvedv_mri <- stich_cmrdata$LV_EDV
stich_cmrdata$lvedv_ix_mri <- stich_cmrdata$LVEDVI

stich_cmrdata <- subset(stich_cmrdata,!(deidnum==969082&STDYDTDY==5))
stich_cmrdata <- subset(stich_cmrdata,!(deidnum==923023&STVOL==66))
stich_cmrdata <- subset(stich_cmrdata,!(deidnum==856349&is.na(LVMASS)))


stich_cmrdata$studyvisit[stich_cmrdata$VISIT==1] <- "BASELINE"
stich_cmrdata$studyvisit[stich_cmrdata$VISIT==2] <- "4MONTH"
stich_cmrdata$studyvisit[stich_cmrdata$VISIT==3] <- "24MONTH"


stich_cmrdata_melt <- melt(stich_cmrdata,
                          id.vars=c("deidnum","studyvisit"),
                          na.rm=T)

stich_cmrdata_melt$form <- "cmrdata"

##########  rndata  ##########

stich_rndata <- read.csv(paste(stich_folder,"sourcedata/h1/rndata.csv",sep=""),
                         stringsAsFactor=F,
                         na.strings=c(".","NA",""))


stich_rndata <- merge(stich_rndata,
                      stich_baseline[,c("deidnum","bsa")],
                      by=c("deidnum"),
                      all.x=T)

stich_rndata$qef_rn <- stich_rndata$QLVEF
stich_rndata$lvedv_ix_rn <- stich_rndata$QLVEDV/stich_rndata$bsa


stich_rndata <- subset(stich_rndata,select=-c(bsa))

stich_rndata$studyvisit[stich_rndata$VISIT==1] <- "BASELINE"
stich_rndata$studyvisit[stich_rndata$VISIT==2] <- "4MONTH"
stich_rndata$studyvisit[stich_rndata$VISIT==3] <- "24MONTH"

stich_rndata_melt <- melt(stich_rndata,
                          id.vars=c("deidnum","studyvisit"),
                          na.rm=T)

stich_rndata_melt$form <- "rndata"

##########  *Compile cardiac imaging*  ##########

stich_combined_imaging <- merge(stich_echodata[,c("deidnum","studyvisit","visualef_echo","qef_echo","ST_46","lvedv_ix_echo","lvmass_echo","lvmass_ix_echo")],
                    stich_rndata[,c("deidnum","studyvisit","qef_rn","QLVEDV","lvedv_ix_rn")],
                    all=T,
                    by=c("deidnum","studyvisit"))

stich_combined_imaging <- merge(stich_combined_imaging,
                    stich_cmrdata[,c("deidnum","studyvisit","qef_mri","LV_EDV","LVEDVI","lvmass_mri","lvmass_ix_mri")],
                    all=T,by=c("deidnum","studyvisit"))


####### Find best LVEF value for each timepoint

### Assumes MRI best, RN 2nd, quantitative echo, then visual echo.  

stich_combined_imaging$best_ef <- stich_combined_imaging$qef_mri
stich_combined_imaging$best_ef[is.na(stich_combined_imaging$qef_mri)] <- stich_combined_imaging$qef_rn[is.na(stich_combined_imaging$qef_mri)]

stich_combined_imaging$best_ef[is.na(stich_combined_imaging$qef_mri)&
                     is.na(stich_combined_imaging$qef_rn)&
                     !is.na(stich_combined_imaging$qef_echo)] <- 
  stich_combined_imaging$qef_echo[is.na(stich_combined_imaging$qef_mri)&
                             is.na(stich_combined_imaging$qef_rn)&
                             !is.na(stich_combined_imaging$qef_echo)]

stich_combined_imaging$best_ef[is.na(stich_combined_imaging$best_ef)&
                     !is.na(stich_combined_imaging$visualef_echo)] <- 
  stich_combined_imaging$visualef_echo[is.na(stich_combined_imaging$best_ef)&
                                  !is.na(stich_combined_imaging$visualef_echo)]

#### LVEDV #### 


stich_combined_imaging$best_lvedv_ix <- stich_combined_imaging$lvedv_ix_mri
stich_combined_imaging$best_lvedv_ix[is.na(stich_combined_imaging$best_lvedv_ix)] <- 
  stich_combined_imaging$lvedv_ix_rn[is.na(stich_combined_imaging$best_lvedv_ix)]

stich_combined_imaging$best_lvedv_ix[is.na(stich_combined_imaging$best_lvedv_ix)&
                                       !is.na(stich_combined_imaging$lvedv_ix_echo)] <- 
  stich_combined_imaging$lvedv_echo[is.na(stich_combined_imaging$best_lvedv_ix)&
                                         !is.na(stich_combined_imaging$lvedv_ix_echo)]


#### Same for LVEDV index 


stich_combined_imaging$best_lvedv_ix <- stich_combined_imaging$lvedv_ix_mri
stich_combined_imaging$best_lvedv_ix[is.na(stich_combined_imaging$best_lvedv_ix)] <- 
  stich_combined_imaging$lvedv_ix_rn[is.na(stich_combined_imaging$best_lvedv_ix)]

stich_combined_imaging$best_lvedv_ix[is.na(stich_combined_imaging$best_lvedv_ix)&
                     !is.na(stich_combined_imaging$lvedv_ix_echo)] <- 
  stich_combined_imaging$lvedv_ix_echo[is.na(stich_combined_imaging$best_lvedv_ix)&
                        !is.na(stich_combined_imaging$lvedv_ix_echo)]

### Same for LV mass and index (echo + MRI only)

### Note that correlation between mri and echo LV mass estimates is not that great, so this is for completeness.

stich_combined_imaging$best_lvmass <- stich_combined_imaging$lvmass_mri
stich_combined_imaging$best_lvmass[is.na(stich_combined_imaging$best_lvmass)] <- stich_combined_imaging$lvmass_echo[is.na(stich_combined_imaging$best_lvmass)]


stich_combined_imaging$best_lvmass_ix <- stich_combined_imaging$lvmass_ix_mri
stich_combined_imaging$best_lvmass_ix[is.na(stich_combined_imaging$best_lvmass_ix)] <- stich_combined_imaging$lvmass_ix_echo[is.na(stich_combined_imaging$best_lvmass_ix)]

stich_combined_imaging_melt <- melt(stich_combined_imaging,
                        id.vars=c("deidnum",
                                  "studyvisit"))

stich_combined_imaging_melt$form <- "combined_imaging"


##########  sourcedata/h1/bnpdata  ##########

stich_bnpdata <- read.csv(paste(stich_folder,"SourceData/h1/bnpdata.csv",sep=""),
                     stringsAsFactors = F,
                     na.strings=c(".","","NA"))

stich_bnpdata$bnp <- stich_bnpdata$jbnp
stich_bnpdata$bnp[is.na(stich_bnpdata$bnp)&!is.na(stich_bnpdata$sdbnp)] <- stich_bnpdata$sdbnp[is.na(stich_bnpdata$bnp)&!is.na(stich_bnpdata$sdbnp)]*1.471+133.194

stich_bnpdata <- subset(stich_bnpdata,!visit %in% c(4,NA))

stich_bnpdata_melt <- melt(data=stich_bnpdata,id.vars=c("deidnum","visit"),na.rm=T) 
stich_bnpdata_melt$studyvisit[stich_bnpdata_melt$visit==1] <- "BASELINE"
stich_bnpdata_melt$studyvisit[stich_bnpdata_melt$visit==2] <- "4MONTH"
stich_bnpdata_melt$form <- "bnpdata"


##########  sourcedata/h1/meds  ##########

stich_meds <- read.csv(paste(stich_folder,"sourcedata/h1/meds.csv",sep=""),
                      stringsAsFactors = F,
                      na.strings=c(".","","NA"))


stich_meds$studyvisit <- stich_meds$FORM
stich_meds$variable <- as.character(stich_meds$MEDTYPE)
stich_meds$value <- stich_meds$MEDTAKEN

stich_meds_melt <- stich_meds[,c("deidnum","studyvisit","variable","value")]

stich_meds_melt$form <- "meds"


##########  sourcedata/h1/disposit  ##########

stich_disposit <- read.csv(paste(stich_folder,"SourceData/h1/disposit.csv",sep=""),
                     stringsAsFactors = F,
                     na.strings=c(".","","NA"))

stich_disposit_melt <- melt(data=stich_disposit,id.vars=c("deidnum"),na.rm=T) 
stich_disposit_melt$studyvisit <- "BASELINE"
stich_disposit_melt$form <- "disposit"


##### Walk test data

stich_walktest <- read.csv(paste(stich_folder,"sourcedata/h1/walktest.csv",sep=""),
                           stringsAsFactor=F,
                           na.strings=c(".","NA",""))

stich_walktest$six_mw_distm[stich_walktest$DISUNT==2&!is.na(stich_walktest$DISUNT)] <- 
  stich_walktest$DISTWLK[stich_walktest$DISUNT==2&!is.na(stich_walktest$DISUNT)]

stich_walktest$six_mw_distm[is.na(stich_walktest$DISUNT)] <- 
  stich_walktest$DISTWLK[is.na(stich_walktest$DISUNT)]

stich_walktest$six_mw_distm[stich_walktest$DISUNT==1&!is.na(stich_walktest$DISUNT)] <- 
  stich_walktest$DISTWLK[stich_walktest$DISUNT==1&!is.na(stich_walktest$DISUNT)] * 0.3048

stich_walktest_melt <- melt(stich_walktest,
                            id.vars=c("deidnum"),
                            na.rm=T)

stich_walktest_melt$studyvisit <- "BASELINE"
stich_walktest_melt$form <- "walktest"


###### analysisdata/followup ####

stich_followup <- read.csv(paste(stich_folder,"sourcedata/h1/followup.csv",sep=""),
                                 stringsAsFactor=F,na.strings=c(".","NA",""))

stich_followup$six_mw_distm[stich_followup$FUDISTY==2&!is.na(stich_followup$FUDISTY)] <- 
  stich_followup$FUDISWLK[stich_followup$FUDISTY==2&!is.na(stich_followup$FUDISTY)]

stich_followup$six_mw_distm[stich_followup$FUDISTY==1&!is.na(stich_followup$FUDISTY)] <- 
  stich_followup$FUDISWLK[stich_followup$FUDISTY==1&!is.na(stich_followup$FUDISTY)] * 0.3048

stich_followup$six_mw_distm[is.na(stich_followup$FUDISTY)&stich_followup$FUDISWLK<700&!is.na(stich_followup$FUDISWLK)] <- 
  stich_followup$FUDISWLK[is.na(stich_followup$FUDISTY)&stich_followup$FUDISWLK<700&!is.na(stich_followup$FUDISWLK)]

stich_followup$studyvisit <- stich_followup$FORM

stich_followup_melt <- melt(stich_followup,
                            id.vars=c("deidnum","studyvisit"),
                            na.rm=T)

stich_followup_melt$form <- "followup"


###### *** STICHES *** #####

## dem ##


stiches_dem <- read.csv(paste(stiches_folder,"analysisdata/dem.csv",sep=""),
                              stringsAsFactor=F,
                        na.strings=c(".","NA",""))

stiches_dem$studyvisit <- stiches_dem$VISIT

stiches_dem_melt <- melt(stiches_dem,
                            id.vars=c("deidnum","studyvisit"),
                            na.rm=T)

stiches_dem_melt$form <- "stiches_dem"


## patients ##

stiches_patients <- read.csv(paste(stiches_folder,"sourcedata/patients.csv",sep=""),
                                   stringsAsFactor=F,
                             na.strings=c(".","NA",""))

stiches_patients_melt <- melt(stiches_patients,
                         id.vars=c("deidnum"),
                         na.rm=T)

stiches_patients_melt$form <- "stiches_patient"


## vitals ##

stiches_vitals <- read.csv(paste(stiches_folder,"analysisdata/vitals.csv",sep=""),
                                 stringsAsFactor=F,
                           na.strings=c(".","NA",""))

stiches_vitals$pupr <- stiches_vitals$HBPSYS-stiches_vitals$HBPDIA
stiches_vitals$studyvisit <- stiches_vitals$VISIT

stiches_vitals_melt <- melt(stiches_vitals,
                               id.vars=c("deidnum","studyvisit"),
                               na.rm=T)

stiches_vitals_melt$form <- "stiches_vitals"


### STICHES walk test

stiches_walk <-  read.csv(paste(stiches_folder,"analysisdata/walk.csv",sep=""),
                          stringsAsFactor=F,
                          na.strings=c(".","NA",""))

stiches_walk$six_mw_distm <- stiches_walk$FUDISWLK_NORM/3.28

stiches_walk$studyvisit <- stiches_walk$VISIT

stiches_walk_melt <- melt(stiches_walk,
                          id.var=c("deidnum","studyvisit"),
                          na.rm=T)

stiches_walk_melt$form <- "stiches_walk"


### STICHES meds

stiches_meds <-  read.csv(paste(stiches_folder,"analysisdata/meds.csv",sep=""),
                          stringsAsFactor=F,
                          na.strings=c(".","NA",""))

stiches_meds$studyvisit <- stiches_meds$VISIT

stiches_meds_melt <- melt(stiches_meds,
                          id.vars=c("deidnum",
                                    "studyvisit"),
                          na.rm=T)

stiches_meds_melt$form <- "stiches_meds"


### STICH QOL forms include 

# KCCQ, 
# Cardiac Self Efficacy Score, 
# 12-Item Short Form health survey (SF-12), 
# Seattle Angina Questionnaire
# CESD-20 + 10

stich_bqol <- read.csv(paste(stich_folder,"SourceData/h1/stich_nih_bqol.csv",sep=""),
                             na.strings=c("NA","NULL","","."))

stich_fqol <- read.csv(paste(stich_folder,"SourceData/h1/stich_nih_fqol.csv",sep=""),
                       na.strings=c("NA","NULL","","."))

stich_bqol[stich_bqol < 0|stich_bqol==""] <- NA
stich_fqol[stich_fqol < 0|stich_fqol==""] <- NA

stich_bqol$studyvisit <- "BASELINE"

stich_fqol$studyvisit[stich_fqol$intervl>0] <- paste(stich_bqol$intervl[stich_bqol$intervl>0],"MONTH",sep="")


#### KCCQ

stich_bqol$kchfworsen <- NA
stich_bqol$kcundsymp <- NA

stich_bqol <- score_kccq(dat=stich_bqol,
                         q1a="kcdress",
                         q1b="kcshowr",
                         q1c="kcwalk",
                         q1d="kchouswk",
                         q1e="kcstrs",
                         q1f="kchuryng",
                         q2="kchfsymp",
                         q3="kcswln",
                         q4="kcswlamt",
                         q5="kcftgn",
                         q6="kcftgamt",
                         q7="kcsbn",
                         q8="kcsbamt",
                         q9="kcsleep",
                         q10="kchfworsen",
                         q11="kcundsymp",
                         q12="kcenjoy",
                         q13="kcsatisf",
                         q14="kcdumps",
                         q15a="kchobby",
                         q15b="kcchores",
                         q15c="kcvisits",
                         q15d="kcrelat")


stich_fqol$kchfworsen <- NA
stich_fqol$kcundsymp <- NA

stich_fqol <- score_kccq(dat=stich_fqol,
                         q1a="kcdress",
                         q1b="kcshowr",
                         q1c="kcwalk",
                         q1d="kchouswk",
                         q1e="kcstrs",
                         q1f="kchuryng",
                         q2="kchfsymp",
                         q3="kcswln",
                         q4="kcswlamt",
                         q5="kcftgn",
                         q6="kcftgamt",
                         q7="kcsbn",
                         q8="kcsbamt",
                         q9="kcsleep",
                         q10="kchfworsen",
                         q11="kcundsymp",
                         q12="kcenjoy",
                         q13="kcsatisf",
                         q14="kcdumps",
                         q15a="kchobby",
                         q15b="kcchores",
                         q15c="kcvisits",
                         q15d="kcrelat")


### CES-D20


stich_bqol$cesd_20 <- apply(stich_bqol,
                            MARGIN=1,
                            FUN=function(x)
                              sum(as.numeric(x[88:107])))

stich_bqol$cesd_10 <- apply(stich_bqol,
                            MARGIN=1,
                            FUN=function(x)
                              sum(as.numeric(x[c(88,92,93,94,95,97,98,99,101,107)])))


stich_fqol$cesd_20 <- apply(stich_fqol,
                            MARGIN=1,
                            FUN=function(x)
                              sum(as.numeric(x[81:100])))


stich_fqol$cesd_10 <- apply(stich_fqol,
                            MARGIN=1,
                            FUN=function(x)
                              sum(as.numeric(x[c(81,85,86,87,88,90,91,92,94,100)])))


stich_bqol$studyvisit[stich_bqol$intervl==0] <- "BASELINE"
stich_bqol$studyvisit[stich_bqol$intervl>0] <- paste(stich_bqol$intervl[stich_bqol$intervl>0],"MONTH",sep="")

stich_fqol$studyvisit[stich_fqol$intervl==0] <- "BASELINE"
stich_fqol$studyvisit[stich_fqol$intervl>0] <- paste(stich_fqol$intervl[stich_fqol$intervl>0],"MONTH",sep="")


stich_bqol_melt <- melt(stich_bqol,
                        id.vars=c("deidnum","studyvisit"),
                        na.rm=T)

stich_bqol_melt$form <- "bqol"

stich_fqol_melt <- melt(stich_fqol,
                        id.vars=c("deidnum","studyvisit"),
                        na.rm=T)

stich_fqol_melt$form <- "fqol"


### EURO-QOL

stich_eq5d <- read.csv(paste(stich_folder,"SourceData/h1/stich_nih_eq5d.csv",sep=""),
                       na.strings=c("NA","NULL","","."))


stich_eq5d[stich_eq5d<0] <- NA

stich_eq5d$studyvisit[stich_eq5d$intervl==0] <- "BASELINE"
stich_eq5d$studyvisit[stich_eq5d$intervl>0] <- paste(stich_eq5d$intervl[stich_eq5d$intervl>0],"MONTH",sep="")

stich_eq5d_melt <- melt(stich_eq5d,
                        id.vars=c("deidnum","studyvisit"),
                        na.rm=T)

stich_eq5d_melt$form <- "eq5d"
stich_eq5d_melt$variable <- as.character(stich_eq5d_melt$variable)


## STICHES Euro QOL


## STICHES records same EQ5D scales as STICH and should be able to merge.

stiches_euroqol <-  read.csv(paste(stiches_folder,"analysisdata/euroqol.csv",sep=""),
                             na.strings=c("NA","NULL","","."))

stiches_euroqol[stiches_euroqol < 0] <- NA
stiches_euroqol$studyvisit <- stiches_euroqol$VISIT

stiches_euroqol_melt <- melt(stiches_euroqol,
                             id.vars=c("deidnum",
                                       "studyvisit"),
                             na.rm=T,
                             factorsAsStrings=F)

stiches_euroqol_melt$form <- "stiches_euroqol"
stiches_euroqol_melt$variable <- as.character(stiches_euroqol_melt$variable)


######### Compile ##########

stich_melt <- rbind(stich_baseline_melt[,c("deidnum","form","studyvisit","variable","value")],
                    stich_angclass_melt[,c("deidnum","form","studyvisit","variable","value")],
                    stich_bnpdata_melt[,c("deidnum","form","studyvisit","variable","value")],
                    stich_corelab_melt[,c("deidnum","form","studyvisit","variable","value")],
                    stich_cmrdata_melt[,c("deidnum","form","studyvisit","variable","value")],
                    stich_rndata_melt[,c("deidnum","form","studyvisit","variable","value")],
                    stich_echodata_melt[,c("deidnum","form","studyvisit","variable","value")],
                    stich_meds_melt[,c("deidnum","form","studyvisit","variable","value")],
                    stich_medhist_melt[,c("deidnum","form","studyvisit","variable","value")],
                    stich_cardcath_melt[,c("deidnum","form","studyvisit","variable","value")],
                    stich_combined_imaging_melt[,c("deidnum","form","studyvisit","variable","value")],
                    stich_demog_melt[,c("deidnum","form","studyvisit","variable","value")],
                    stich_disposit_melt[,c("deidnum","form","studyvisit","variable","value")],
                    stich_followup_melt[,c("deidnum","form","studyvisit","variable","value")],
                    stiches_meds_melt[,c("deidnum","form","studyvisit","variable","value")],
                    stiches_vitals_melt[,c("deidnum","form","studyvisit","variable","value")],
                    stich_walktest_melt[,c("deidnum","form","studyvisit","variable","value")],
                    stiches_walk_melt[,c("deidnum","form","studyvisit","variable","value")],
                    stich_bqol_melt[,c("deidnum","form","studyvisit","variable","value")],
                    stich_fqol_melt[,c("deidnum","form","studyvisit","variable","value")],
                    stich_eq5d_melt[,c("deidnum","form","studyvisit","variable","value")],
                    stiches_euroqol_melt[,c("deidnum","form","studyvisit","variable","value")])

stich_melt$studyvisit <- gsub(" ","",stich_melt$studyvisit)

stich_melt <- subset(stich_melt,!variable=="VISIT")

names(stich_melt)[1] <- "patientid"

stich_melt1 <- subset(stich_melt,!patientid %in% stiches_dem$deidnum)
stiches_melt <- subset(stich_melt,patientid %in% stiches_dem$deidnum&
                         !studyvisit %in% c("66MONTH",
                                            "72MONTH",
                                            "78MONTH",
                                            "84MONTH"))

stich_melt <- rbind(stich_melt1,stiches_melt)

stich_melt$study <- "STICH"


############################# **********EXACT********** ######################################


# .%%%%%%%%.%%.....%%....%%%.....%%%%%%..%%%%%%%%
# .%%........%%...%%....%%.%%...%%....%%....%%...
# .%%.........%%.%%....%%...%%..%%..........%%...
# .%%%%%%......%%%....%%.....%%.%%..........%%...
# .%%.........%%.%%...%%%%%%%%%.%%..........%%...
# .%%........%%...%%..%%.....%%.%%....%%....%%...
# .%%%%%%%%.%%.....%%.%%.....%%..%%%%%%.....%%...
# 


##########  a_base  ##########

exact_base <- read.csv(paste(exact_folder,"a_base.csv",sep=""),
                       stringsAsFactors = F,
                       na.strings=c(".","","NA"))

exact_base$YIHF_months <- exact_base$YIHF*12

exact_base_melt <- melt(data=exact_base,id.vars=c("PATNUMB"),na.rm=T,factorsAsStrings = T) 
exact_base_melt$variable <- as.character(exact_base_melt$variable)

names(exact_base_melt)[1] <- "patnumb"
exact_base_melt$studyvisit <- "BASELINE"
exact_base_melt$form <- "a_base"


##########  a_visitsumm  ##########

exact_visitsumm <- read.csv(paste(exact_folder,"a_visitsumm.csv",sep=""),
                     stringsAsFactors = F,
                     na.strings=c(".","","NA"))

exact_visitsumm$studyvisit <- exact_visitsumm$FORM

exact_visitsumm$weight_kg <- round(exact_visitsumm$WTLBS/2.2,1)

exact_hgt <- exact_visitsumm[exact_visitsumm$FORM=="SCREENING",
                             c("PATNUMB","HEIGHTIN")]

exact_hgt$height_cm <- round(exact_hgt$HEIGHTIN*2.54,0)

exact_visitsumm <- merge(exact_visitsumm,
                         exact_hgt[,c("PATNUMB","height_cm")],
                         na.rm=T)

exact_visitsumm$bmi <- round(exact_visitsumm$weight_kg/(exact_visitsumm$height_cm/100)^2,1)


exact_visitsumm$alc <- round(exact_visitsumm$LL_WBC*exact_visitsumm$LL_LYMPH/100,2)

exact_visitsumm_melt <- melt(data=exact_visitsumm,id.vars=c("PATNUMB","studyvisit"),na.rm=T) 
exact_visitsumm_melt$variable <- as.character(exact_visitsumm_melt$variable)

exact_visitsumm_melt$studyvisit[exact_visitsumm_melt$studyvisit=="SCREENING"&
                                  exact_visitsumm_melt$variable %in% c("LL_SODIUM",
                                                                       "LL_POTASS",
                                                                       "LL_BUN",
                                                                       "LL_BICARB",
                                                                       "LL_CREAT",
                                                                       "LL_GFR",
                                                                       "LL_MAGNES",
                                                                       "LL_GLUC",
                                                                       "LL_CHLOR",
                                                                       "LL_CALC",
                                                                       "LL_TCHOL",
                                                                       "LL_AST",
                                                                       "LL_ALT",
                                                                       "LL_ALKPHOS",
                                                                       "LL_BILIR",
                                                                       "LL_PROT",
                                                                       "LL_ALBUM",
                                                                       "LL_WBC",
                                                                       "LL_LYMPH",
                                                                       "LL_HMG",
                                                                       "LL_HCT",
                                                                       "LL_PLAT",
                                                                       "LL_RDW",
                                                                       "LL_BNP",
                                                                       "LL_NTPBNP",
                                                                       "LL_URIC")] <- "BASELINE"

colnames(exact_visitsumm_melt)[1] <- "patnumb"
exact_visitsumm_melt$form <- "a_visitsumm"


##########  medhist1  ##########

exact_medhist1 <- read.csv(paste(exact_folder,"medhist1.csv",sep=""),
                           stringsAsFactors = F,
                           na.strings=c(".","","NA"))


exact_cms <- names(exact_medhist1) %in% c("ALCOHOLC",
                                            "CYTOTOXC",
                                            "FAMILIAL",
                                            "HCM",
                                            "HYPERTEN",
                                            "DILATED",
                                            "RESTRICT",
                                            "OTHCONT",
                                            "PERIPART",
                                            "VAL")


exact_medhist1[exact_medhist1$ISCHEMIC==1,exact_cms] <- NA

exact_medhist1$num_etiol <- apply(exact_medhist1[exact_cms],
                                   MARGIN=1,
                                   FUN=function(x)
                                     sum(x,na.rm=T))

exact_medhist1[exact_medhist1$num_etiol>1,exact_cms] <- NA


exact_medhist1_melt <- melt(data=exact_medhist1,
                            id.vars=c("patnumb","FORM"),
                            na.rm=T) 

exact_medhist1_melt$variable <- as.character(exact_medhist1_melt$variable)

exact_medhist1_melt$studyvisit <- "BASELINE"
exact_medhist1_melt$form <- "medhist1"


##########  medhist2  ##########

exact_medhist2 <- read.csv(paste(exact_folder,"medhist2.csv",sep=""),
                           stringsAsFactors = F,
                           na.strings=c(".","","NA"))

exact_medhist2_melt <- melt(data=exact_medhist2,id.vars=c("patnumb","FORM"),na.rm=T) 

exact_medhist2_melt$variable <- as.character(exact_medhist2_melt$variable)

exact_medhist2_melt$studyvisit <- "BASELINE"
exact_medhist2_melt$form <- "medhist2"


##########  assessmt  ##########

exact_assessmt <- read.csv(paste(exact_folder,"assessmt.csv",sep=""),
                     stringsAsFactors = F,
                     na.strings=c(".","","NA"))

exact_assessmt$pupr <- exact_assessmt$BPSYS-exact_assessmt$BPDIA

exact_assessmt_melt <- melt(data=exact_assessmt,id.vars=c("patnumb","FORM"),na.rm=T) 
exact_assessmt_melt$variable <- as.character(exact_assessmt_melt$variable)
exact_assessmt_melt$studyvisit <- exact_assessmt_melt$FORM
exact_assessmt_melt$form <- "assessmt"


##########  echo  ##########

exact_echo <- read.csv(paste(exact_folder,"echo.csv",sep=""),
                       stringsAsFactors = F,
                       na.strings=c(".","","NA"))

exact_echo$studyvisit[exact_echo$SCHDTIME==1] <- "BASELINE"
exact_echo$studyvisit[exact_echo$SCHDTIME==10] <- "WEEK 24"

exact_echo <- merge(exact_echo,
                    exact_base[,c("PATNUMB","SEX")],
                    by.x="patnumb",
                    by.y="PATNUMB")

exact_echo <- merge(exact_echo,exact_visitsumm[exact_visitsumm$studyvisit=="BASELINE",
                                               c("PATNUMB","studyvisit","weight_kg","height_cm")],
                    by.x=c("patnumb","studyvisit"),
                    by.y=c("PATNUMB","studyvisit"),
                    all.x=T)

exact_echo$LVED_best[!is.na(exact_echo$LVEDCH)] <- exact_echo$LVEDCH[!is.na(exact_echo$LVEDCH)]
exact_echo$LVED_best[is.na(exact_echo$LVEDCH)] <- exact_echo$LVEDVOL[is.na(exact_echo$LVEDCH)]

exact_echo$EA_RATIO <- exact_echo$EVELOC/exact_echo$AVELOC

exact_echo$EVELOC_cmsec <- exact_echo$EVELOC*100
exact_echo$AVELOC_cmsec <- exact_echo$AVELOC*100

exact_echo$DTILATE_cmsec <- exact_echo$DTILATE*100
exact_echo$DTIMEDE_cmsec <- exact_echo$DTIMEDE*100


exact_echo$eeprime_lat <- round(exact_echo$EVELOC/exact_echo$DTILATE,1)
exact_echo$eeprime_sept <- round(exact_echo$EVELOC/exact_echo$DTIMEDE,1)
exact_echo$eeprime_avg <- round((exact_echo$eeprime_sept+exact_echo$eeprime_lat)/2,1)

exact_echo$pvf_ratio <- exact_echo$SYSFWRD/exact_echo$DIAFWRD

exact_dd <-  assign_diastolic_function(dat=exact_echo,
                                                   pid="patnumb",
                                                   vst="studyvisit",
                                                 ep_lat="DTILATE_cmsec",
                                                 ep_sept="DTIMEDE_cmsec",
                                                 earat="EA_RATIO",
                                                 e_vel="EVELOC_cmsec",
                                                 dt = "DECELTM",
                                                 tr_vel = "PEAKVELC",
                                                 def="TOPCAT") 

exact_echo <- merge(exact_echo,
                  exact_dd,
                  by=c("patnumb",
                       "studyvisit"),
                  all.x=T)

exact_echo <- calc_hypertrophy_type(df=exact_echo,
                                  sex="SEX",
                                  male="1",
                                  female="2",
                                  lvedd_cm="LVDD",
                                  ivsd_cm="IVSD",
                                  lvpwtd_cm="PWSD",
                                  height_cm="height_cm",
                                  weight_kg="weight_kg")


exact_echo <- calc_lav(exact_echo,
                     laa_4c = "LACAREA",
                     laa_2c = "LACHAREA",
                     lal_4c = "LACLEN",
                     lal_2c = "LALEN",
                     height_cm = "height_cm",
                     weight_kg = "weight_kg")


exact_echo$LVEDD_index <- exact_echo$LVDD/exact_echo$bsa
exact_echo$lvedv_ix <- exact_echo$LVED_best/exact_echo$bsa


exact_echo <- calc_pulsatility(dat=exact_echo,
                                     sbp = "ECHOSYS",
                                     dbp = "ECHODIA",
                                     edv = "LVEDCH",
                                     esv = "LVECHCH",
                                     bsa = "bsa",
                                     hr = "ECHOHR",
                                     lvot_d = "LVOT" ,
                                     vti = "TVI")

exact_echo <- subset(exact_echo,select = -c(LVMASS))

exact_echo_melt <- 
  melt(data=exact_echo,id.vars=c("patnumb","studyvisit"),na.rm=T) 

exact_echo_melt$variable <- as.character(exact_echo_melt$variable)

exact_echo_melt$form <- "echo"


##########  meds  ##########

exact_meds_melt <- read.csv(paste(exact_folder,"meds.csv",sep=""),
                            stringsAsFactors = F,
                            na.strings=c(".","","NA"))

exact_meds_melt$variable <- exact_meds_melt$HFMEDS
exact_meds_melt$value <- exact_meds_melt$MEDRAND
exact_meds_melt$studyvisit <- exact_meds_melt$FORM
exact_meds_melt$form <- "meds"


##########  diuretic  ##########

exact_diuretic <- read.csv(paste(exact_folder,"diuretic.csv",sep=""),
                           stringsAsFactors = F,
                           na.strings=c(".","","NA"))


exact_furo <- sqldf("select patnumb, 
                    FORM as studyvisit, 
                    DIURDOSE as FURODOSE 
                    from exact_diuretic 
                    where DIUMEDS = 1")

exact_bum <- sqldf("select patnumb, 
                    FORM as studyvisit, 
                    DIURDOSE as BUMDOSE 
                    from exact_diuretic 
                    where DIUMEDS = 3")

exact_tors <- sqldf("select patnumb, 
                    FORM as studyvisit, 
                    DIURDOSE as TORSEDOSE 
                    from exact_diuretic 
                    where DIUMEDS = 2")


exact_furo_equiv <- merge(exact_furo,
                          exact_bum,
                          by=c("patnumb","studyvisit"),
                          all.x=T)

exact_furo_equiv <- merge(exact_furo_equiv,
                          exact_tors,
                          by=c("patnumb","studyvisit"),
                          all.x=T)


exact_furo_equiv$value <- apply(exact_furo_equiv
                                [,c("FURODOSE",
                                    "TORSEDOSE",
                                    "BUMDOSE")],
                                MARGIN=1,
                                FUN=diur_calc)

exact_furo_equiv$variable <- "daily_furo_eq_derive"

names(exact_diuretic)[c(1,2,4)] <- c("variable","value","studyvisit")

exact_diuretic_melt <- rbind(exact_diuretic[,c("patnumb","studyvisit","variable","value")],
                             exact_furo_equiv[,c("patnumb","studyvisit","variable","value")])

exact_diuretic_melt$form <- "diuretic"


##########  ecg  ##########

exact_ecg <- read.csv(paste(exact_folder,"ecg.csv",sep=""),
                      stringsAsFactors = F,
                      na.strings=c(".","","NA"))


exact_ecg_melt <- melt(data=exact_ecg,id.vars=c("patnumb","FORM"),na.rm=T) 
exact_ecg_melt$variable <- as.character(exact_ecg_melt$variable)
exact_ecg_melt$studyvisit <- exact_ecg_melt$FORM
exact_ecg_melt$form <- "ecg"


## Load and score EXACT-HF

exact_kccq1 <- read.csv(paste(exact_folder,"kccq1.csv",sep=""),na.strings=c("NA","NULL",""))
exact_kccq2 <- read.csv(paste(exact_folder,"kccq2.csv",sep=""),na.strings=c("NA","NULL",""))
exact_kccq3 <- read.csv(paste(exact_folder,"kccq3.csv",sep=""),na.strings=c("NA","NULL",""))


exact_kccq <- sqldf('select "EXACT" as study,
                       patnumb,
                       FORM as studyvisit, 
                       KCCQ1A, 
                       KCCQ1B, 
                       KCCQ1C, 
                       KCCQ1D,
                       KCCQ1E,
                       KCCQ1F,
                       KCCQ2,
                       KCCQ3,
                       KCCQ4,
                       KCCQ5,
                       KCCQ6,
                       KCCQ7,
                       KCCQ8,
                       KCCQ9,
                       KCCQ10,
                       KCCQ11,
                       KCCQ12,
                       KCCQ13,
                       KCCQ14,
                       KCCQ15A,
                       KCCQ15B,
                       KCCQ15C,
                       KCCQ15D
                       from exact_kccq1 left join exact_kccq2 using (patnumb,FORM)
                       join exact_kccq3 using (patnumb,FORM)')

exact_kccq <- score_kccq(dat=exact_kccq,
                         q1a="KCCQ1A",
                         q1b="KCCQ1B",
                         q1c="KCCQ1C",
                         q1d="KCCQ1D",
                         q1e="KCCQ1E",
                         q1f="KCCQ1F",
                         q2="KCCQ2",
                         q3="KCCQ3",
                         q4="KCCQ4",
                         q5="KCCQ5",
                         q6="KCCQ6",
                         q7="KCCQ7",
                         q8="KCCQ8",
                         q9="KCCQ9",
                         q10="KCCQ10",
                         q11="KCCQ11",
                         q12="KCCQ12",
                         q13="KCCQ13",
                         q14="KCCQ14",
                         q15a="KCCQ15A",
                         q15b="KCCQ15B",
                         q15c="KCCQ15C",
                         q15d="KCCQ15D")


exact_kccq_melt <- melt(exact_kccq,id.vars=c("patnumb","studyvisit"))
exact_kccq_melt$variable <- as.character(exact_kccq_melt$variable)
exact_kccq_melt$form <- "kccq"

##########  walktest  ##########

exact_walktest <- read.csv(paste(exact_folder,"walktest.csv",sep=""),
                           stringsAsFactors = F)

exact_walktest$total_time <- exact_walktest$WLKMIN + exact_walktest$WLKSEC/60

exact_walktest_melt <- melt(exact_walktest, id.vars=c("patnumb","FORM"),na.rm=T)
exact_walktest_melt$variable <- as.character(exact_walktest_melt$variable)
names(exact_walktest_melt)[2] <- "studyvisit"
exact_walktest_melt$form <- "walktest"


exact_melt <- rbind(exact_assessmt_melt[,c("patnumb","studyvisit","form","variable","value")],
                    exact_base_melt[,c("patnumb","studyvisit","form","variable","value")],
                    exact_diuretic_melt[,c("patnumb","studyvisit","form","variable","value")],
                    exact_ecg_melt[,c("patnumb","studyvisit","form","variable","value")],
                    exact_echo_melt[,c("patnumb","studyvisit","form","variable","value")],
                    exact_kccq_melt[,c("patnumb","studyvisit","form","variable","value")],
                    exact_medhist1_melt[,c("patnumb","studyvisit","form","variable","value")],
                    exact_medhist2_melt[,c("patnumb","studyvisit","form","variable","value")],
                    exact_meds_melt[,c("patnumb","studyvisit","form","variable","value")],
                    exact_visitsumm_melt[,c("patnumb","studyvisit","form","variable","value")],
                    exact_walktest_melt[,c("patnumb","studyvisit","form","variable","value")])

names(exact_melt)[1] <- "patientid"
exact_melt$study <- "EXACT"


############################# **********ROSE********** #######################################

# .%%%%%%%%...%%%%%%%...%%%%%%..%%%%%%%%
# .%%.....%%.%%.....%%.%%....%%.%%......
# .%%.....%%.%%.....%%.%%.......%%......
# .%%%%%%%%..%%.....%%..%%%%%%..%%%%%%..
# .%%...%%...%%.....%%.......%%.%%......
# .%%....%%..%%.....%%.%%....%%.%%......
# .%%.....%%..%%%%%%%...%%%%%%..%%%%%%%%


##########  a_base  ##########

rose_base <- read.sas7bdat(paste(rose_folder,"a_base.sas7bdat",sep=""))

rose_base$YIHF_months <- rose_base$YIHF*12

rose_base$RACE[rose_base$ETHNIC==1] <- 8

rose_base_melt <- 
  melt(data=rose_base,id.vars=c("PATNUMB"),na.rm=T) 

rose_base_melt$variable <- as.character(rose_base_melt$variable)

rose_base_melt$studyvisit <- "BASELINE"
colnames(rose_base_melt)[1] <- "patnumb"
rose_base_melt$form <- "a_base"


##########  a_visitsumm  ##########

rose_visitsumm <- read.sas7bdat(paste(rose_folder,"a_visitsumm.sas7bdat",sep=""))
rose_visitsumm[rose_visitsumm=="NaN"] <- NA

rose_visitsumm$weight_kg <- rose_visitsumm$WTLBS/2.2

rose_ht <- rose_visitsumm[rose_visitsumm$FORM=="BASELINE", c("PATNUMB", "HEIGHTIN")]
rose_ht$height_cm <- rose_ht$HEIGHTIN*2.54

rose_visitsumm <- merge(rose_visitsumm,rose_ht[,c("PATNUMB","height_cm")],by="PATNUMB")
rose_visitsumm$bmi <- round((rose_visitsumm$weight_kg)/(rose_visitsumm$height_cm/100)^2,1)

rose_visitsumm$alc <- rose_visitsumm$LL_LYMPH/100*rose_visitsumm$LL_WBC

rose_visitsumm_melt <- 
  melt(data=rose_visitsumm,id.vars=c("PATNUMB","FORM"),na.rm=T) 

rose_visitsumm_melt$variable <- as.character(rose_visitsumm_melt$variable)

rose_visitsumm_melt$studyvisit <- rose_visitsumm_melt$FORM
colnames(rose_visitsumm_melt)[1] <- "patnumb"
rose_visitsumm_melt$form <- "a_visitsumm"


##########  fatigue  ##########

rose_fatigue <- read.sas7bdat(paste(rose_folder,"fatigue.sas7bdat",sep=""))

rose_fatigue_melt <- 
  melt(data=rose_fatigue,id.vars=c("patnumb","FORM"),na.rm=T) 

rose_fatigue_melt$variable <- as.character(rose_fatigue_melt$variable)

rose_fatigue_melt$studyvisit <- rose_fatigue_melt$FORM
rose_fatigue_melt$form <- "fatigue"


##########  swelling  ##########

rose_swelling <- read.sas7bdat(paste(rose_folder,"swelling.sas7bdat",sep=""))

rose_swelling_melt <- 
  melt(data=rose_swelling,id.vars=c("patnumb","FORM"),na.rm=T) 

rose_swelling_melt$variable <- as.character(rose_swelling_melt$variable)

rose_swelling_melt$studyvisit <- rose_swelling_melt$FORM
rose_swelling_melt$form <- "swelling"


##########  pdss  ##########

rose_pdss <- read.sas7bdat(paste(rose_folder,"pdss.sas7bdat",sep=""))

rose_pdss_melt <- 
  melt(data=rose_pdss,id.vars=c("patnumb","FORM"),na.rm=T) 

rose_pdss_melt$variable <- as.character(rose_pdss_melt$variable)

rose_pdss_melt$studyvisit <- rose_pdss_melt$FORM
rose_pdss_melt$form <- "pdss"


##########  pdss2  ##########

rose_pdss2 <- read.sas7bdat(paste(rose_folder,"pdss2.sas7bdat",sep=""))

rose_pdss2_melt <- 
  melt(data=rose_pdss2,id.vars=c("patnumb","FORM"),na.rm=T) 

rose_pdss2_melt$variable <- as.character(rose_pdss2_melt$variable)

rose_pdss2_melt$studyvisit <- rose_pdss2_melt$FORM
rose_pdss2_melt$form <- "pdss2"


##########  medhist1  ##########

rose_medhist1 <- read.sas7bdat(paste(rose_folder,"medhist1.sas7bdat",sep=""))


rose_cms <- names(rose_medhist1) %in% c("ALCOHOLC",
                                          "CYTOTXC",
                                          "FAMILIAL",
                                          "HCM",
                                          "HYPERTEN",
                                          "DILATED",
                                          "RESTRICT",
                                          "OTHCONT",
                                          "PERIPAR",
                                          "VAL")

rose_medhist1[rose_medhist1$ISCHEMIC==1,rose_cms] <- NA


rose_medhist1$num_etiol <- apply(rose_medhist1[rose_cms],
                                  MARGIN=1,
                                  FUN=function(x)
                                    sum(x,na.rm=T))

rose_medhist1[rose_medhist1$num_etiol>1,rose_cms] <- NA


rose_medhist1_melt <- 
  melt(data=rose_medhist1,id.vars=c("patnumb","FORM"),na.rm=T) 

rose_medhist1_melt$variable <- as.character(rose_medhist1_melt$variable)

rose_medhist1_melt$studyvisit <- rose_medhist1_melt$FORM
colnames(rose_medhist1_melt)[1] <- "patnumb"
rose_medhist1_melt$form <- "medhist1"


##########  medhist2  ##########

rose_medhist2 <- read.sas7bdat(paste(rose_folder,"medhist2.sas7bdat",sep=""))


rose_medhist2_melt <- 
  melt(data=rose_medhist2,id.vars=c("patnumb","FORM"),na.rm=T) 

rose_medhist2_melt$variable <- as.character(rose_medhist2_melt$variable)

rose_medhist2_melt$studyvisit <- rose_medhist2_melt$FORM
rose_medhist2_melt$form <- "medhist2"


##########  assessmt  ##########

rose_assessmt <- read.sas7bdat(paste(rose_folder,"assessmt.sas7bdat",sep=""))

rose_assessmt$pupr <- rose_assessmt$BPSYS-rose_assessmt$BPDIA

rose_assessmt_melt <- 
  melt(data=rose_assessmt,id.vars=c("patnumb","FORM"),na.rm=T) 

rose_assessmt_melt$variable <- as.character(rose_assessmt_melt$variable)

rose_assessmt_melt$studyvisit <- rose_assessmt_melt$FORM
rose_assessmt_melt$form <- "assessmt"


##########  meds  ##########

rose_meds_melt <- read.sas7bdat(paste(rose_folder,"meds.sas7bdat",sep=""))

rose_meds_melt$studyvisit <- rose_meds_melt$FORM
rose_meds_melt$variable <- as.character(rose_meds_melt$HFMEDS)
rose_meds_melt$value <- rose_meds_melt$MEDSANS
rose_meds_melt$form <- "meds"


##########  diuretic  ##########

rose_diuretic <- read.sas7bdat(paste(rose_folder,"diuretic.sas7bdat",sep=""))


rose_furo <- sqldf("select patnumb, 
                    FORM as studyvisit, 
                    DIURDOSE as FURODOSE 
                    from rose_diuretic 
                    where DIUMEDS = 1")

rose_bum <- sqldf("select patnumb, 
                    FORM as studyvisit, 
                    DIURDOSE as BUMDOSE 
                    from rose_diuretic 
                    where DIUMEDS = 3")

rose_tors <- sqldf("select patnumb, 
                    FORM as studyvisit, 
                    DIURDOSE as TORSEDOSE 
                    from rose_diuretic 
                    where DIUMEDS = 2")

rose_furo_equiv <- merge(rose_furo,
                          rose_bum,
                          by=c("patnumb","studyvisit"),
                          all.x=T)

rose_furo_equiv <- merge(rose_furo_equiv,
                          rose_tors,
                          by=c("patnumb","studyvisit"),
                          all.x=T)


rose_furo_equiv$value <- apply(rose_furo_equiv
                                [,c("FURODOSE",
                                    "TORSEDOSE",
                                    "BUMDOSE")],
                                MARGIN=1,
                                FUN=diur_calc)

rose_furo_equiv$variable <- "daily_furo_eq_derive"

names(rose_diuretic)[c(1,2,4)] <- c("variable","value","studyvisit")

rose_diuretic_melt <- rbind(rose_diuretic[,c("patnumb","studyvisit","variable","value")],
                             rose_furo_equiv[,c("patnumb","studyvisit","variable","value")])

rose_diuretic_melt$form <- "diuretic"


##########  ecg  ##########

rose_ecg <- read.sas7bdat(paste(rose_folder,"ecg.sas7bdat",sep=""))


rose_ecg_melt <- 
  melt(data=rose_ecg,id.vars=c("patnumb","FORM"),na.rm=T) 

rose_ecg_melt$variable <- as.character(rose_ecg_melt$variable)
rose_ecg_melt$studyvisit <- rose_ecg_melt$FORM
rose_ecg_melt$form <- "ecg"


######### vas #############

rose_vas <- read.sas7bdat(paste(rose_folder,"vas.sas7bdat",sep=""))
rose_vas[rose_vas=="NaN"] <- NA

rose_vas_melt <- melt(rose_vas,id.vars=c("patnumb","FORM"), na.rm=T)
rose_vas_melt$studyvisit <- rose_vas_melt$FORM
rose_vas_melt$form <- "vas"


##########  rsfluid  ##########

rose_rsfluid <- read.sas7bdat(paste(rose_folder,"rsfluid.sas7bdat",sep=""))
rose_rsfluid[rose_rsfluid=="NaN"] <- NA
rose_rsfluid$io_balance <-  rose_rsfluid$RSIVIN+
  rose_rsfluid$RSORALIN-
  rose_rsfluid$RSUROUT-
  rose_rsfluid$RSNUROUT


rose_rsfluid <- subset(rose_rsfluid,!is.na(io_balance))


rose_rsfluid_cum_d1 <- subset(rose_rsfluid,FORM=="24 HOURS"&!is.na(io_balance))[,c("patnumb","io_balance")]
names(rose_rsfluid_cum_d1)[2] <- "io_balancex24HOURS"

rose_rsfluid_cum_d2 <- subset(rose_rsfluid,FORM=="48 HOURS"&!is.na(io_balance))[,c("patnumb","io_balance")]
names(rose_rsfluid_cum_d2)[2] <- "io_balancex48HOURS"

rose_rsfluid_cum_d3 <- subset(rose_rsfluid,FORM=="72 HOURS"&!is.na(io_balance))[,c("patnumb","io_balance")]
names(rose_rsfluid_cum_d3)[2] <- "io_balancex72HOURS"


rose_base_list <- rose_base[,c("PATNUMB","TREATMENT")]
names(rose_base_list)[1] <- "patnumb"

rose_rsfluid_cum <- merge(rose_base_list[,c("patnumb","TREATMENT")],rose_rsfluid_cum_d1,by="patnumb",all.x=T)
rose_rsfluid_cum <- merge(rose_rsfluid_cum,rose_rsfluid_cum_d2,by="patnumb",all.x=T)
rose_rsfluid_cum <- merge(rose_rsfluid_cum,rose_rsfluid_cum_d3,by="patnumb",all.x=T)

rose_rsfluid_cum$io_cumx24HOURS <- rose_rsfluid_cum$io_balancex24HOURS
rose_rsfluid_cum$io_cumx48HOURS <- rose_rsfluid_cum$io_balancex24HOURS+rose_rsfluid_cum$io_balancex48HOURS
rose_rsfluid_cum$io_cumx72HOURS <- rose_rsfluid_cum$io_cumx48HOURS+rose_rsfluid_cum$io_balancex72HOURS

rose_rsfluid_cum_melt <- melt(rose_rsfluid_cum,
                          id.vars=c("patnumb"),
                          na.rm=T)

rose_rsfluid_cum_melt <- subset(rose_rsfluid_cum_melt,!variable=="TREATMENT")
rose_rsfluid_cum_melt$studyvisit <- str_select(rose_rsfluid_cum_melt$variable,after="x")
rose_rsfluid_cum_melt$variable <- str_select(rose_rsfluid_cum_melt$variable,before="x")

rose_rsfluid_cum_melt$form <- "rsfluid"

##########  walktest  ##########

rose_walktest <- read.sas7bdat(paste(rose_folder,"walktest.sas7bdat",sep=""))
rose_walktest[rose_walktest=="NaN"] <- NA


rose_walktest_melt <- melt(rose_walktest,
                           id.vars=c("patnumb","FORM"),
                           na.rm=T)

rose_walktest_melt$variable <- as.character(rose_walktest_melt$variable)
rose_walktest_melt$studyvisit <- rose_walktest_melt$FORM
rose_walktest_melt$form <- "walktest"


##### wsymptom ####

rose_wsymptom_melt <- read.sas7bdat(paste(rose_folder,"wsymptom.sas7bdat",sep=""))

rose_wsymptom_melt$value <- rose_wsymptom_melt$WORST
rose_wsymptom_melt$variable <- "WORST"
rose_wsymptom_melt$studyvisit <- rose_wsymptom_melt$FORM
rose_wsymptom_melt$form <- "wsymptom"


##### Compile ####

rose_melt <- rbind(rose_base_melt[,c("patnumb","form","studyvisit","variable","value")],
                   rose_assessmt_melt[,c("patnumb","form","studyvisit","variable","value")],
                   rose_diuretic_melt[,c("patnumb","form","studyvisit","variable","value")],
                   rose_ecg_melt[,c("patnumb","form","studyvisit","variable","value")],
                   rose_fatigue_melt[,c("patnumb","form","studyvisit","variable","value")],
                   rose_diuretic_melt[,c("patnumb","form","studyvisit","variable","value")],
                   rose_medhist1_melt[,c("patnumb","form","studyvisit","variable","value")],
                   rose_medhist2_melt[,c("patnumb","form","studyvisit","variable","value")],
                   rose_meds_melt[,c("patnumb","form","studyvisit","variable","value")],
                   rose_pdss_melt[,c("patnumb","form","studyvisit","variable","value")],
                   rose_pdss2_melt[,c("patnumb","form","studyvisit","variable","value")],
                   rose_rsfluid_cum_melt[,c("patnumb","form","studyvisit","variable","value")],
                   rose_swelling_melt[,c("patnumb","form","studyvisit","variable","value")],
                   rose_vas_melt[,c("patnumb","form","studyvisit","variable","value")],
                   rose_visitsumm_melt[,c("patnumb","form","studyvisit","variable","value")],
                   rose_walktest_melt[,c("patnumb","form","studyvisit","variable","value")],
                   rose_wsymptom_melt[,c("patnumb","form","studyvisit","variable","value")])

rose_melt <- subset(rose_melt, !value %in% c("NA","NaN",""))
rose_melt <- subset(rose_melt, !is.na(value))
names(rose_melt)[1] <- "patientid"

rose_melt$study <- "ROSE"

############################# **********GUIDE-IT********** ###################################

# ..%%%%%%...%%.....%%.%%%%.%%%%%%%%..%%%%%%%%.........%%%%.%%%%%%%%
# .%%....%%..%%.....%%..%%..%%.....%%.%%................%%.....%%...
# .%%........%%.....%%..%%..%%.....%%.%%................%%.....%%...
# .%%...%%%%.%%.....%%..%%..%%.....%%.%%%%%%...%%%%%%%..%%.....%%...
# .%%....%%..%%.....%%..%%..%%.....%%.%%................%%.....%%...
# .%%....%%..%%.....%%..%%..%%.....%%.%%................%%.....%%...
# ..%%%%%%....%%%%%%%..%%%%.%%%%%%%%..%%%%%%%%.........%%%%....%%...


##########  analysis_data/demog_ads  ##########

guideit_demog_ads <- read.csv(paste(guideit_folder,"analysis_data/demog_ads.csv",sep=""),
                     stringsAsFactors = F, 
                     na.strings=c(".","","NA"))


guideit_demog_ads_melt <- melt(data=guideit_demog_ads,id.vars=c("deidnum"),na.rm=T) 

guideit_demog_ads_melt$variable <- as.character(guideit_demog_ads_melt$variable)
guideit_demog_ads_melt$studyvisit <- "BASELINE"
guideit_demog_ads_melt$form <- "demog_ads"


##########  raw_data/base  ##########

guideit_base <- read.csv(paste(guideit_folder,"raw_data/base.csv",sep=""),
                              stringsAsFactors = F, 
                              na.strings=c(".","","NA"))


guideit_base_melt <- melt(data=guideit_base,id.vars=c("deidnum"),na.rm=T) 
guideit_base_melt$variable <- as.character(guideit_base_melt$variable)
guideit_base_melt$studyvisit <- "BASELINE"
guideit_base_melt$form <- "base"


##########  analysis_data/adh_corelabs_ads  ##########

guideit_corelab <- read.csv(paste(guideit_folder,"analysis_data/adh_corelab_ads.csv",sep=""),
                     stringsAsFactors = F,
                     na.strings=c(".","","NA"))

guideit_corelab_melt <- melt(data=guideit_corelab,id.vars=c("deidnum","visit"),na.rm=T) 
guideit_corelab_melt$studyvisit <- guideit_corelab_melt$visit
guideit_corelab_melt$form <- "adh_corelab_ads"


##########  raw_data/dem  ##########

guideit_dem <- read.csv(paste(guideit_folder,"raw_data/dem.csv",sep=""),
                     stringsAsFactors = F,
                     na.strings=c(".","","NA"))

### Normalize race field

guideit_dem$race_derived[guideit_dem$WHITE==1] <- 1
guideit_dem$race_derived[guideit_dem$BLACK==1] <- 2
guideit_dem$race_derived[guideit_dem$ASIAN==1] <- 4
guideit_dem$race_derived[guideit_dem$other==1] <- 6
guideit_dem$race_derived[guideit_dem$ETHNIC==1] <- 3

guideit_dem_melt <- 
  melt(data=guideit_dem,id.vars=c("deidnum","VISIT"),na.rm=T) 
guideit_dem_melt$variable <- as.character(guideit_dem_melt$variable)
guideit_dem_melt$studyvisit <- guideit_dem_melt$VISIT
guideit_dem_melt$form <- "dem"


##########  raw_data/medhx  ##########

guideit_medhx <- read.csv(paste(guideit_folder,"raw_data/medhx.csv",sep=""),
                     stringsAsFactors = F,
                     na.strings=c(".","","NA"))

guideit_medhx_melt <- melt(data=guideit_medhx,id.vars=c("deidnum","VISIT"),na.rm=T) 
guideit_medhx_melt$studyvisit <- guideit_medhx_melt$VISIT
guideit_medhx_melt$form <- "medhx"


##########  raw_data/exam  ##########

guideit_exam <- read.csv(paste(guideit_folder,"raw_data/exam.csv",sep=""),
                     stringsAsFactors = F,
                     na.strings=c(".","","NA"))

guideit_exam <- merge(guideit_exam,guideit_demog_ads[,c("deidnum","HEIGHTM")],by="deidnum",all.x=T)
guideit_exam$weight_kg <- guideit_exam$WEIGHTNR/2.2
guideit_exam$bmi <- guideit_exam$weight_kg/(guideit_exam$HEIGHTM^2)

guideit_exam$pupr <- guideit_exam$BPSYS-guideit_exam$BPDIA

guideit_exam_melt <- melt(data=guideit_exam,id.vars=c("deidnum","VISIT"),na.rm=T) 
guideit_exam_melt$variable <- as.character(guideit_exam_melt$variable)
guideit_exam_melt$studyvisit <- guideit_exam_melt$VISIT
guideit_exam_melt$form <- "exam"


##########  analysis_data/labs_ads  ##########

guideit_labs <- read.csv(paste(guideit_folder,"analysis_data/labs_ads.csv",sep=""),
                         stringsAsFactors = F,
                         na.strings=c(".","","NA"))

guideit_labs <- merge(guideit_labs, guideit_dem[,c("deidnum","GENDER","race_derived")],by="deidnum",all.x=T)
guideit_labs$uric_mgdl <- round(guideit_labs$URICRSLT*16.8,1)
guideit_labs$alc <- guideit_labs$LYMPRSLT/100*guideit_labs$WBCRSLT


guideit_labs$CrCl <- round(calc_MDRD4(guideit_labs,
                                cr="CRTRSLT",
                                age="age",
                                sex="GENDER",
                                race="race_derived",
                                black="2"),1)


guideit_labs_melt <- melt(data=guideit_labs,id.vars=c("deidnum","VISIT"),na.rm=T) 
guideit_labs_melt$variable <- as.character(guideit_labs_melt$variable)
guideit_labs_melt$studyvisit <- guideit_labs_melt$VISIT
guideit_labs_melt$form <- "labs_ads"


##########  raw_data/probnp  ##########

guideit_probnp <- read.csv(paste(guideit_folder,"raw_data/probnp.csv",sep=""),
                         stringsAsFactors = F,
                         na.strings=c(".","","NA"))
guideit_probnp_melt <- melt(data=guideit_probnp,id.vars=c("deidnum","VISIT"),na.rm=T) 
guideit_probnp_melt$variable <- as.character(guideit_probnp_melt$variable)
guideit_probnp_melt$studyvisit <- guideit_probnp_melt$VISIT
guideit_probnp_melt$form <- "probnp"


##########  raw_data/meds  ##########

guideit_meds <- read.csv(paste(guideit_folder,"raw_data/meds.csv",sep=""),
                     stringsAsFactors = F,
                     na.strings=c(".","","NA"))


guideit_meds$daily_furo_eq_derive <- apply(guideit_meds[,c("FUROSE","TORSEDD","BUMETA")],
                                           MARGIN=1,
                                           FUN=diur_calc)

guideit_meds_melt <- melt(data=guideit_meds,id.vars=c("deidnum","VISIT"),na.rm=T) 
guideit_meds_melt$studyvisit <- guideit_meds_melt$VISIT
guideit_meds_melt$form <- "meds"


##########  raw_data/ef  ##########

guideit_ef <- read.csv(paste(guideit_folder,"raw_data/ef.csv",sep=""),
                     stringsAsFactors = F,
                     na.strings=c(".","","NA"))

guideit_ef_melt <- melt(data=guideit_ef,id.vars=c("deidnum","VISIT"),na.rm=T) 
guideit_ef_melt$studyvisit <- guideit_ef_melt$VISIT
guideit_ef_melt$form <- "ef"


##########  qol_data/qolbaseline  ##########

guideit_qolbaseline <- read.csv(paste(guideit_folder,"qol_data/qolbaseline.csv",sep=""),
                       stringsAsFactors = F,
                       na.strings=c(".","","NA"))

guideit_qolbaseline[guideit_qolbaseline < 0] <- NA


guideit_qolbaseline$cesd_10 <- apply(guideit_qolbaseline[c(39:48)],
                                     MARGIN=1,
                                     FUN=function(x)
                                       sum(x))


guideit_qolbaseline <- score_kccq(dat=guideit_qolbaseline,
                         q1a="kcdress",
                         q1b="kcshowr",
                         q1c="kcwalk",
                         q1d="kchouswk",
                         q1e="kcstrs",
                         q1f="kchuryng",
                         q2="kchfsymp",
                         q3="kcswln",
                         q4="kcswlamt",
                         q5="kcftgn",
                         q6="kcftgamt",
                         q7="kcsbn",
                         q8="kcsbamt",
                         q9="kcsleep",
                         q10="kchfworsen",
                         q11="kcundsymp",
                         q12="kcenjoy",
                         q13="kcsatist",
                         q14="kcdumps",
                         q15a="kchobby",
                         q15b="kcchores",
                         q15c="kcvisits",
                         q15d="kcrelat")


guideit_qolbaseline_melt <- melt(data=guideit_qolbaseline,id.vars=c("deidnum"),na.rm=T) 
guideit_qolbaseline$variable <- guideit_qolbaseline$variable
guideit_qolbaseline_melt$studyvisit <- "evBLN"
guideit_qolbaseline_melt$form <- "qolbaseline"


##########  qol_data/qolfup  ##########

guideit_qolfup <- read.csv(paste(guideit_folder,"qol_data/qolfup.csv",sep=""),
                                stringsAsFactors = F,
                                na.strings=c(".","","NA"))

guideit_qolfup[guideit_qolfup < 0] <- NA


guideit_qolfup$cesd_10 <- apply(guideit_qolfup[c(39:48)],
                                     MARGIN=1,
                                     FUN=function(x)
                                       sum(x))


guideit_qolfup <- score_kccq(dat=guideit_qolfup,
                                  q1a="kcdress",
                                  q1b="kcshowr",
                                  q1c="kcwalk",
                                  q1d="kchouswk",
                                  q1e="kcstrs",
                                  q1f="kchuryng",
                                  q2="kchfsymp",
                                  q3="kcswln",
                                  q4="kcswlamt",
                                  q5="kcftgn",
                                  q6="kcftgamt",
                                  q7="kcsbn",
                                  q8="kcsbamt",
                                  q9="kcsleep",
                                  q10="kchfworsen",
                                  q11="kcundsymp",
                                  q12="kcenjoy",
                                  q13="kcsatist",
                                  q14="kcdumps",
                                  q15a="kchobby",
                                  q15b="kcchores",
                                  q15c="kcvisits",
                                  q15d="kcrelat")


guideit_qolfup$studyvisit[guideit_qolfup$intervl==3] <- "evMON3"
guideit_qolfup$studyvisit[guideit_qolfup$intervl==6] <- "evMON6"
guideit_qolfup$studyvisit[guideit_qolfup$intervl==12] <- "evMON12"
guideit_qolfup$studyvisit[guideit_qolfup$intervl==24] <- "evMON24"

guideit_qolfup_melt <- melt(data=guideit_qolfup,id.vars=c("deidnum","studyvisit"),na.rm=T) 
guideit_qolfup_melt$variable <- guideit_qolfup_melt$variable
guideit_qolfup_melt$form <- "qolfup"


guideit_melt <- rbind(guideit_base_melt[,c("deidnum","studyvisit","form","variable","value")],
                      guideit_corelab_melt[,c("deidnum","studyvisit","form","variable","value")],
                      guideit_dem_melt[,c("deidnum","studyvisit","form","variable","value")],
                      guideit_demog_ads_melt[,c("deidnum","studyvisit","form","variable","value")],
                      guideit_ef_melt[,c("deidnum","studyvisit","form","variable","value")],
                      guideit_exam_melt[,c("deidnum","studyvisit","form","variable","value")],
                      guideit_labs_melt[,c("deidnum","studyvisit","form","variable","value")],
                      guideit_medhx_melt[,c("deidnum","studyvisit","form","variable","value")],
                      guideit_meds_melt[,c("deidnum","studyvisit","form","variable","value")],
                      guideit_qolbaseline_melt[,c("deidnum","studyvisit","form","variable","value")],
                      guideit_qolfup_melt[,c("deidnum","studyvisit","form","variable","value")])

guideit_melt$study <- "GUIDE-IT"
names(guideit_melt)[1] <- "patientid"


#----------------------------------------------------------------------------------------#
########################### **********ATHENA********** ####################################

# ....%%%....%%%%%%%%.%%.....%%.%%%%%%%%.%%....%%....%%%...
# ...%%.%%......%%....%%.....%%.%%.......%%%...%%...%%.%%..
# ..%%...%%.....%%....%%.....%%.%%.......%%%%..%%..%%...%%.
# .%%.....%%....%%....%%%%%%%%%.%%%%%%...%%.%%.%%.%%.....%%
# .%%%%%%%%%....%%....%%.....%%.%%.......%%..%%%%.%%%%%%%%%
# .%%.....%%....%%....%%.....%%.%%.......%%...%%%.%%.....%%
# .%%.....%%....%%....%%.....%%.%%%%%%%%.%%....%%.%%.....%%

# Butler J, Anstrom KJ, Felker GM, Givertz MM, Kalogeropoulos AP, Konstam MA, Mann DL, Margulies KB, McNulty SE, 
# Mentz RJ, Redfield MM, Tang WHW, Whellan DJ, Shah M, Desvigne-Nickens P, Hernandez AF, Braunwald E. 
# Efficacy and safety of spironolactone in acute heart failure: The ATHENA-HF randomized clinical trial. 
# JAMA Cardiology. 2017;2(9): 950–958. https://doi.org/10.1001/jamacardio.2017.2198


##########  a_base  ##########

athena_base <- read.csv(paste(athena_folder,"a_base.csv",sep=""),
                        stringsAsFactors = F,
                        na.strings=c(".","","NA"))

athena_base$HFAGE_months <- athena_base$HFAGE*12
athena_base$height_cm <- athena_base$HEIGHTIN_BL*2.54

athena_base_melt <- melt(data=athena_base,id.vars=c("PATNUMB"),na.rm=T) 
athena_base_melt$VISIT <- "BASE"
athena_base_melt$form <- "a_base"


##########  a_visitsumm  ##########

athena_visitsumm <- read.csv(paste(athena_folder,"a_visitsumm.csv",sep=""),stringsAsFactors = F,na.strings=c(".","","NA"))

athena_visitsumm$weight_kg <- athena_visitsumm$WTLBS/2.2
athena_visitsumm <- merge(athena_visitsumm,athena_base[,c("PATNUMB","height_cm")])

athena_visitsumm$bmi <- athena_visitsumm$weight_kg/((athena_visitsumm$height_cm/100)^2)

athena_visitsumm_melt <- melt(data=athena_visitsumm,id.vars=c("PATNUMB","VISIT"),na.rm=T) 
athena_visitsumm_melt$form <- "a_visitsumm"


##########  medhist1  ##########

athena_medhist1 <- read.csv(paste(athena_folder,"medhist1.csv",sep=""),
                            stringsAsFactors = F,
                            na.strings=c(".","","NA"))

athena_medhist1 <- merge(athena_medhist1,athena_base[,c("PATNUMB","HFETIOLOGY")],by="PATNUMB",all.x=T)

# athena_medhist1$HFETIOLOGY[athena_medhist1$HFETIOLOGY==2] <- NA

athena_cms <- names(athena_medhist1) %in% c(
                              "ALCOHOLC",
                              "CYTOTOX",
                              "FAMILIAL",
                              "HCM",
                              "HYPERTN",
                              "IDIODIL",
                              "IDIORES",
                              "MYOOTH",
                              "PERIPRT",
                              "VALVUL")


athena_medhist1[athena_medhist1$HFETIOLOGY==1,athena_cms] <- NA

athena_medhist1$num_etiol <- apply(athena_medhist1[athena_cms],
                                  MARGIN=1,
                                  FUN=function(x)
                                    sum(x,na.rm=T))


athena_medhist1[athena_medhist1$num_etiol>1,c("ALCOHOLC",
                                            "CYTOTOX",
                                            "FAMILIAL",
                                            "HCM",
                                            "HYPERTN",
                                            "IDIODIL",
                                            "IDIORES",
                                            "MYOOTH",
                                            "PERIPRT",
                                            "VALVUL")] <- NA


athena_medhist1_melt <- melt(data=athena_medhist1,id.vars=c("PATNUMB","VISIT","FORM"),na.rm=T) 
athena_medhist1_melt$form <- athena_medhist1_melt$FORM
athena_medhist1_melt$VISIT <- "BASE"


##########  medhist2  ##########


athena_medhist2 <- read.csv(paste(athena_folder,"medhist2.csv",sep=""),
                            stringsAsFactors = F,
                            na.strings=c(".","","NA"))

athena_medhist2_melt <- melt(data=athena_medhist2,id.vars=c("PATNUMB","VISIT","FORM"),na.rm=T) 
athena_medhist2_melt$form <- athena_medhist2_melt$FORM


##########  demog  ##########

athena_demog <- read.csv(paste(athena_folder,"demog.csv",sep=""),
                         stringsAsFactors = F,
                         na.strings=c(".","","NA"))


athena_demog_melt <- melt(data=athena_demog,id.vars=c("PATNUMB","VISIT","FORM"),na.rm=T) 

athena_demog_melt$form <- athena_demog_melt$FORM

athena_meas <- merge(athena_base[,c("PATNUMB","AGE","RACE","height_cm")], 
                     athena_demog[,c("PATNUMB","SEX")], by=c("PATNUMB"))


##########  exam  ##########

athena_exam <- read.csv(paste(athena_folder,"exam.csv",sep=""),
                        stringsAsFactors = F,
                        na.strings=c(".","","NA"))

athena_exam$pupr <- athena_exam$BPSYS-athena_exam$BPDIA


athena_exam_melt <- melt(data=athena_exam,id.vars=c("PATNUMB","VISIT","FORM"),na.rm=T) 
athena_exam_melt$form <- athena_exam_melt$FORM


##########  labs1  ##########

athena_labs1 <- read.csv(paste(athena_folder,"labs1.csv",sep=""),
                         stringsAsFactors = F,
                         na.strings=c(".","","NA"))

athena_labs1$BUN_mgdL[athena_labs1$BUNUNT==3&!is.na(athena_labs1$BUNUNT)] <- 
  athena_labs1$BUNVAL[athena_labs1$BUNUNT==3&!is.na(athena_labs1$BUNUNT)]

athena_labs1$BUN_mgdL[athena_labs1$BUNUNT==1&!is.na(athena_labs1$BUNUNT)] <- 
  athena_labs1$BUNVAL[athena_labs1$BUNUNT==1&!is.na(athena_labs1$BUNUNT)] * 0.3571

athena_labs1 <- merge(athena_labs1,athena_meas,by=c("PATNUMB"), all.x=T)

athena_labs1$CrCl <- calc_MDRD4(dat=athena_labs1,
                                cr="CREATVAL",
                                black=3)

athena_labs1_melt <- melt(data=athena_labs1,id.vars=c("PATNUMB","VISIT","FORM"),na.rm=T) 
athena_labs1_melt$form <- athena_labs1_melt$FORM


##########  labs2  ##########

athena_labs2 <- read.csv(paste(athena_folder,"labs2.csv",sep=""),stringsAsFactors = F,na.strings=c(".","","NA"))

athena_labs2$BUN_mgdL[athena_labs2$BUNUNT2==3&!is.na(athena_labs2$BUNUNT2)] <- athena_labs2$BUNVAL2[athena_labs2$BUNUNT2==3&!is.na(athena_labs2$BUNUNT2)]
athena_labs2$BUN_mgdL[athena_labs2$BUNUNT2==1&!is.na(athena_labs2$BUNUNT2)] <- athena_labs2$BUNVAL2[athena_labs2$BUNUNT2==1&!is.na(athena_labs2$BUNUNT2)] * 0.3571

athena_labs2_melt <- melt(data=athena_labs2,id.vars=c("PATNUMB","VISIT","FORM"),na.rm=T) 
athena_labs2_melt$form <- athena_labs2_melt$FORM


###### kcreat  #####

# There are 2 additional tables kcreat3 and kcreat5 that represent repeat lab values.  There are relatively few of these.
# I elected to just keep the original table kcreat 3 rather than trying to make substitution rules.

athena_kcreat <- read.csv(paste(athena_folder,"kcreat.csv",sep=""),stringsAsFactors = F,na.strings=c(".","","NA"))

athena_kcreat <- merge(athena_kcreat,athena_meas,by=c("PATNUMB"), all.x=T)

athena_kcreat$CrCl <- calc_MDRD4(dat=athena_kcreat,
                                 cr="CREATVL3",
                                black=3)


athena_kcreat_melt <- melt(data=athena_kcreat,
                           id.vars=c("PATNUMB","VISIT","FORM"),
                           na.rm=T)

athena_kcreat_melt$form <- "kcreat"

##########  meds  ##########

athena_meds <- read.csv(paste(athena_folder,"meds.csv",sep=""),
                        stringsAsFactors = F,
                        na.strings=c(".","","NA"))

athena_meds_melt <- melt(data=athena_meds,id.vars=c("PATNUMB","VISIT","FORM"),na.rm=T) 
athena_meds_melt$form <- "med"


##########  premeds  ##########

athena_premeds <- read.csv(paste(athena_folder,"premeds.csv",sep=""),
                           stringsAsFactors = F,
                           na.strings=c(".","","NA"))

athena_premeds$daily_furo_eq_derive <- apply(athena_premeds[,c("FURODOSE","TORSDOSE","BUMDOSE")],
                                             MARGIN=1,
                                             FUN=diur_calc) 


athena_premeds_melt <- melt(data=athena_premeds,id.vars=c("PATNUMB","VISIT","FORM"),na.rm=T) 
athena_premeds_melt$form <- "premeds"


##########  dc  ##########

athena_dcmeds <- read.csv(paste(athena_folder,"dcmeds.csv",sep=""),
                          stringsAsFactors = F,
                          na.strings=c(".","","NA"))

athena_dcmeds$daily_furo_eq_derive <- apply(athena_dcmeds[,c("FURODOSD","TORSDOSD","BUMDOSED")],
                                             MARGIN=1,
                                             FUN=diur_calc) 

athena_dcmeds_melt <- melt(data=athena_dcmeds,id.vars=c("PATNUMB","VISIT","FORM"),na.rm=T) 
athena_dcmeds_melt$form <- "dcmeds"


##########  stratify  ##########

athena_stratify <- read.csv(paste(athena_folder,"stratify.csv",sep=""),
                            stringsAsFactors = F,
                            na.strings=c(".","","NA"))

athena_stratify_melt <- melt(data=athena_stratify,id.vars=c("PATNUMB","VISIT","FORM"),na.rm=T) 
athena_stratify_melt$form <- athena_stratify_melt$FORM


##########  diureti2  ##########

athena_diuret2 <- read.csv(paste(athena_folder,"diureti2.csv",sep=""),
                           stringsAsFactors = F,
                           na.strings=c(".","","NA"))

athena_diuret2_melt <- melt(data=athena_diuret2,id.vars=c("PATNUMB","VISIT","FORM"),na.rm=T) 
athena_diuret2_melt$form <- athena_diuret2_melt$FORM


###### day30med #####

athena_day30med <- read.csv(paste(athena_folder,"day30med.csv",sep=""),
                            stringsAsFactors=F,
                            na.strings=c(".","","NA","NULL"))

athena_day30med$daily_furo_eq_derive <- apply(athena_day30med[,c("FURODOS1","TORSDOS1","BUMDOSE1")],
                                                   MARGIN=1,
                                                   FUN=diur_calc)

athena_day30med_melt <- melt(athena_day30med,
                        id.vars=c("PATNUMB","VISIT","FORM"),
                        na.rm=T)

athena_day30med_melt$form <- "day30med"


######## fluid #########

athena_fluid <- read.csv(paste(athena_folder,"fluid.csv",sep=""),stringsAsFactors=F,na.strings=c(".","","NA","NULL"))

athena_fluid$IVIN[athena_fluid$IVIN==97] <- 0
athena_fluid$ORALIN[athena_fluid$ORALIN==97] <- 0

athena_fluid$UROUT[athena_fluid$UROUT==97] <- 0
athena_fluid$NONUOUT[athena_fluid$NONUOUT==97] <- 0


athena_fluid$checks <- athena_fluid$IVIN+
  athena_fluid$ORALIN+
  athena_fluid$UROUT+
  athena_fluid$NONUOUT


athena_fluid$io_balance <- round((athena_fluid$IVINVAL+athena_fluid$ORLINVAL) -
                                   (athena_fluid$UROUTVAL-athena_fluid$NONUOTVL),0)


athena_fluid$io_balance[athena_fluid$checks < 4] <- NA


athena_fluid_cum_d1 <- subset(athena_fluid,VISIT=="24H"&!is.na(io_balance))[,c("PATNUMB","io_balance")]
names(athena_fluid_cum_d1)[2] <- "io_balancex24H"

athena_fluid_cum_d2 <- subset(athena_fluid,VISIT=="48H"&!is.na(io_balance))[,c("PATNUMB","io_balance")]
names(athena_fluid_cum_d2)[2] <- "io_balancex48H"

athena_fluid_cum_d3 <- subset(athena_fluid,VISIT=="72H"&!is.na(io_balance))[,c("PATNUMB","io_balance")]
names(athena_fluid_cum_d3)[2] <- "io_balancex72H"

athena_fluid_cum_d4 <- subset(athena_fluid,VISIT=="96H"&!is.na(io_balance))[,c("PATNUMB","io_balance")]
names(athena_fluid_cum_d4)[2] <- "io_balancex96H"

athena_fluid_cum <- merge(athena_base[,c("PATNUMB","TREATMENT")],athena_fluid_cum_d1,by="PATNUMB",all.x=T)
athena_fluid_cum <- merge(athena_fluid_cum,athena_fluid_cum_d2,by="PATNUMB",all.x=T)
athena_fluid_cum <- merge(athena_fluid_cum,athena_fluid_cum_d3,by="PATNUMB",all.x=T)
athena_fluid_cum <- merge(athena_fluid_cum,athena_fluid_cum_d4,by="PATNUMB",all.x=T)

athena_fluid_cum$io_cumx24H <- athena_fluid_cum$io_balancex24H
athena_fluid_cum$io_cumx48H <- athena_fluid_cum$io_balancex24H+athena_fluid_cum$io_balancex48H
athena_fluid_cum$io_cumx72H <- athena_fluid_cum$io_cumx48H+athena_fluid_cum$io_balancex72H
athena_fluid_cum$io_cumx96H <- athena_fluid_cum$io_cumx72H+athena_fluid_cum$io_balancex96H

athena_fluid_cum_melt <- melt(athena_fluid_cum,
                              id.vars=c("PATNUMB","TREATMENT"),
                              na.rm=T)

athena_fluid_cum_melt$VISIT <- str_select(athena_fluid_cum_melt$variable,after="x")
athena_fluid_cum_melt$variable <- str_select(athena_fluid_cum_melt$variable,before="x")

athena_fluid_cum_melt$form <- "fluid"


#### Compile ATHENA ####

athena_melt <- rbind(athena_base_melt[,c("PATNUMB","VISIT","form","variable","value")],
                     athena_visitsumm_melt[,c("PATNUMB","VISIT","form","variable","value")],
                     athena_stratify_melt[,c("PATNUMB","VISIT","form","variable","value")],
                     athena_premeds_melt[,c("PATNUMB","VISIT","form","variable","value")],
                     athena_demog_melt[,c("PATNUMB","VISIT","form","variable","value")],
                     athena_day30med_melt[,c("PATNUMB","VISIT","form","variable","value")],
                     athena_kcreat_melt[,c("PATNUMB","VISIT","form","variable","value")],
                     athena_fluid_cum_melt[,c("PATNUMB","VISIT","form","variable","value")],
                     athena_labs1_melt[,c("PATNUMB","VISIT","form","variable","value")],
                     athena_labs2_melt[,c("PATNUMB","VISIT","form","variable","value")],
                     athena_exam_melt[,c("PATNUMB","VISIT","form","variable","value")],
                     athena_medhist1_melt[,c("PATNUMB","VISIT","form","variable","value")],
                     athena_medhist2_melt[,c("PATNUMB","VISIT","form","variable","value")])

colnames(athena_melt)[1:2] <- c("patientid","studyvisit")

athena_melt$study <- "ATHENA"


############################ **********FIGHT********** ########################################

# .%%%%%%%%.%%%%..%%%%%%...%%.....%%.%%%%%%%%
# .%%........%%..%%....%%..%%.....%%....%%...
# .%%........%%..%%........%%.....%%....%%...
# .%%%%%%....%%..%%...%%%%.%%%%%%%%%....%%...
# .%%........%%..%%....%%..%%.....%%....%%...
# .%%........%%..%%....%%..%%.....%%....%%...
# .%%.......%%%%..%%%%%%...%%.....%%....%%...

# Margulies KB, Hernandez AF, Redfield MM, Givertz MM, Oliveira GH, Cole R, Mann DL, 
# Whellan DJ, Kiernan MS, Felker GM, McNulty SE, Anstrom KJ, Shah MR, Braunwald E, Cappola TP, 
# for the NHLBI Heart Failure Clinical Research Network. 
# Effects of Liraglutide on Clinical Stability Among Patients With Advanced Heart Failure
# and Reduced Ejection Fraction A Randomized Clinical Trial JAMA. 2016;316(5):500–508. 
# https://doi.org/10.1001/jama.2016.10260


##########  a_base  ##########

fight_base <- read.csv(paste(fight_folder,"a_base.csv",sep=""),
                     stringsAsFactors = F,
                     na.strings=c(".","","NA"))

fight_base$RACE[fight_base$ETHNIC==1] <- 8
fight_base$HFAGE_months <- fight_base$HFAGE * 12

fight_base_melt <- melt(data=fight_base,id.vars=c("PATNUMB"),na.rm=T) 
fight_base_melt$studyvisit <- "BASE"
fight_base_melt$form <- "a_base"

fight_meas <- fight_base[,c("PATNUMB","AGE","SEX","RACE","HEIGHTIN_BL")]
fight_meas$height_cm <- round(fight_meas$HEIGHTIN_BL * 2.54,1)


##########  a_visitsumm  ##########

fight_visitsumm <- read.csv(paste(fight_folder,"a_visitsumm.csv",sep=""),
                     stringsAsFactors = F,
                     na.strings=c(".","","NA"))

fight_visitsumm <- merge(fight_visitsumm,fight_meas,by=c("PATNUMB"),all.x=T)
fight_visitsumm$weight_kg <- round(fight_visitsumm$WTLBS * 0.453592,1)
fight_visitsumm$bmi <- round(fight_visitsumm$weight_kg/((fight_visitsumm$height_cm/100)^2),1)

fight_visitsumm$CrCl <- calc_MDRD4(dat=fight_visitsumm,
                          cr="CREAT", 
                          age="AGE", 
                          sex="SEX", 
                          race="RACE", 
                          male=1, 
                          black=3 )

fight_visitsumm_melt <- melt(data=fight_visitsumm,
                 id.vars=c("PATNUMB","VISIT"),
                 na.rm=T) 

fight_visitsumm_melt$studyvisit <- fight_visitsumm_melt$VISIT
fight_visitsumm_melt$form <- "a_visitsumm"


fight_meas <- merge(fight_meas,
                    fight_visitsumm[fight_visitsumm$VISIT=="BASE",
                              c("PATNUMB",
                                "AGE",
                                "SEX",
                                "RACE",
                                "HEIGHTIN_BL",
                                "height_cm",
                                "WTLBS",
                                "weight_kg")])


##########  echolab1+2  ##########

fight_echo <- read.csv(paste(fight_folder,"echo.csv",sep=""),
                           stringsAsFactors = F,
                           na.strings=c(".","","NA"))

fight_echolab1 <- read.csv(paste(fight_folder,"echolab1.csv",sep=""),
                     stringsAsFactors = F,
                     na.strings=c(".","","NA"))

fight_echolab2 <- read.csv(paste(fight_folder,"echolab2.csv",sep=""),
                     stringsAsFactors = F,
                     na.strings=c(".","","NA"))

fight_echolab <- merge(fight_echo,
                       fight_echolab1[,2:ncol(fight_echolab1)],
                       by=c("PATNUMB","VISIT"),
                       all=T)

fight_echolab <- merge(fight_echolab,
                        fight_echolab2[,2:ncol(fight_echolab2)],
                  by=c("PATNUMB","VISIT"),
                  all=T)

fight_echolab <- merge(fight_echolab, 
                  fight_meas, 
                  by="PATNUMB", 
                  all.x=T)

rm(fight_echo,
  fight_echolab1,
   fight_echolab2)

fight_echolab$DTILATE_cmsec <- fight_echolab$DTILATE*100
fight_echolab$DTIMEDE_cmsec <- fight_echolab$DTIMEDE*100
fight_echolab$EVELOC_cmsec <- fight_echolab$EVELOC*100
fight_echolab$AVELOC_cmsec <- fight_echolab$AVELOC*100
fight_echolab$earatio <- fight_echolab$EVELOC_cmsec/fight_echolab$AVELOC_cmsec
fight_echolab$RVSP <- 4*(fight_echolab$PEAKVELC^2) + fight_echolab$RAPRESS

fight_echolab$pvf_ratio <- fight_echolab$SYSFWRD/fight_echolab$DYSFWRD

fight_echolab$LVED_best <- fight_echolab$LVEDCH
fight_echolab$LVED_best[is.na(fight_echolab$LVED_best)&!is.na(fight_echolab$LVEDVOL)] <-
  fight_echolab$LVEDVOL[is.na(fight_echolab$LVED_best)&!is.na(fight_echolab$LVEDVOL)]


fight_echolab$LVES_best <- fight_echolab$LVECHCH
fight_echolab$LVES_best[is.na(fight_echolab$LVES_best)&!is.na(fight_echolab$LVESVOL)] <-
  fight_echolab$LVESVOL[is.na(fight_echolab$LVES_best)&!is.na(fight_echolab$LVESVOL)]


fight_dd <-  assign_diastolic_function(dat=fight_echolab,
                                                      pid="PATNUMB",
                                                      vst="VISIT",
                                                   ep_lat="DTILATE_cmsec",
                                                   ep_sept="DTIMEDE_cmsec",
                                                   earat="earatio",
                                                   e_vel="EVELOC_cmsec",
                                                   dt = "DECELTM",
                                                   tr_vel = "PEAKVELC",
                                                   def="TOPCAT") 

fight_echolab <-
  merge(fight_echolab,
        fight_dd,
        by=c("PATNUMB",
             "VISIT"),
        all.x=T)

fight_echolab <- calc_hypertrophy_type(df=fight_echolab,
                                  sex="SEX",
                                  male="1",
                                  female="2",
                                  lvedd_cm="LVDD",
                                  ivsd_cm="IVSD",
                                  lvpwtd_cm="PWSD",
                                  height_cm="height_cm",
                                  weight_kg="weight_kg")


fight_echolab <- calc_lav(fight_echolab,
                     laa_4c = "LACAREA",
                     laa_2c = "LACHAREA",
                     lal_4c = "LACLEN",
                     lal_2c = "LALEN",
                     height_cm = "height_cm",
                     weight_kg = "weight_kg")


fight_echolab$LVEDD_index <- fight_echolab$LVDD/fight_echolab$bsa
fight_echolab$LVESD_index <- fight_echolab$LVSD/fight_echolab$bsa

fight_echolab$eeprime_lat <- round(fight_echolab$EVELOC/fight_echolab$DTILATE,1)
fight_echolab$eeprime_sept <- round(fight_echolab$EVELOC/fight_echolab$DTIMEDE,1)
fight_echolab$eeprime_avg <- round((fight_echolab$eeprime_lat+fight_echolab$eeprime_sept)/2,1)

fight_echolab <- calc_pulsatility(dat=fight_echolab,
                               sbp = "ECHOSYS",
                               dbp = "ECHODIA",
                               edv = "LVED_best",
                               esv = "LVES_best",
                               bsa = "bsa",
                               hr = "ECHOHRVL",
                               lvot_d = "LVOT" ,
                               vti = "TVI")


fight_echolab_melt <-  melt(data=fight_echolab,id.vars=c("PATNUMB","VISIT"),na.rm=T) 

fight_echolab_melt$studyvisit <- fight_echolab_melt$VISIT
fight_echolab_melt$form <- "echolab"


##########  medhist1  ##########

fight_medhist1 <- read.csv(paste(fight_folder,"medhist1.csv",sep=""),
                     stringsAsFactors = F,
                     na.strings=c(".","","NA"))

fight_medhist1 <- merge(fight_medhist1,fight_base[,c("PATNUMB","HFETIOLOGY")],by="PATNUMB")

fight_medhist1$HFETIOLOGY[fight_medhist1$HFETIOLOGY==2] <- 0

fight_medhist1[fight_medhist1$HFETIOLOGY==1,c("ALCOHOLC",
                                              "CYTOTOX",
                                              "FAMILIAL",
                                              "HCM",
                                              "HYPERTN",
                                              "IDIODIL",
                                              "IDIORES",
                                              "MYOOTH",
                                              "PERIPRT",
                                              "VALVUL")] <- NA

fight_medhist1$num_etiol <- apply(fight_medhist1[c("HFETIOLOGY",
  "ALCOHOLC",
                                                   "CYTOTOX",
                                                   "FAMILIAL",
                                                   "HCM",
                                                   "HYPERTN",
                                                   "IDIODIL",
                                                   "IDIORES",
                                                   "MYOOTH",
                                                   "PERIPRT",
                                                   "VALVUL")],
                                  MARGIN=1,
                                  FUN=function(x)
                                  sum(x,na.rm=T))

fight_medhist1[fight_medhist1$num_etiol>1,c("ALCOHOLC",
                                            "CYTOTOX",
                                            "FAMILIAL",
                                            "HCM",
                                            "HYPERTN",
                                            "IDIODIL",
                                            "IDIORES",
                                            "MYOOTH",
                                            "PERIPRT",
                                            "VALVUL")] <- NA

fight_medhist1_melt <- melt(data=fight_medhist1,id.vars=c("PATNUMB","VISIT","FORM"),na.rm=T) 
fight_medhist1_melt$studyvisit <- fight_medhist1_melt$VISIT
fight_medhist1_melt$form <- "medhist1"


##########  medhist2  ##########

fight_medhist2 <- read.csv(paste(fight_folder,"medhist2.csv",sep=""),
                           stringsAsFactors = F,
                           na.strings=c(".","","NA"))

fight_medhist2_melt <- melt(data=fight_medhist2,id.vars=c("PATNUMB","VISIT","FORM"),na.rm=T) 
fight_medhist2_melt$studyvisit <- fight_medhist2_melt$VISIT
fight_medhist2_melt$form <- fight_medhist2_melt$form <- "medhist2"


##########  ecg  ##########

fight_ecg <- read.csv(paste(fight_folder,"ecg.csv",sep=""),
                     stringsAsFactors = F,
                     na.strings=c(".","","NA"))

fight_ecg_melt <- melt(data=fight_ecg,
                 id.vars=c("PATNUMB","VISIT","FORM"),
                 na.rm=T)

fight_ecg_melt$studyvisit <- fight_ecg_melt$VISIT
fight_ecg_melt$form <- "ecg"


##########  exam  ##########

fight_exam <- read.csv(paste(fight_folder,"exam.csv",sep=""),
                     stringsAsFactors = F,
                     na.strings=c(".","","NA"))

fight_exam$pupr <- fight_exam$BPSYS-fight_exam$BPDIA


fight_exam_melt <- melt(data=fight_exam,
                 id.vars=c("PATNUMB","VISIT","FORM"),
                 na.rm=T)

fight_exam_melt$studyvisit <- fight_exam_melt$VISIT
fight_exam_melt$form <- "exam"


##########  sixmwt  ##########

fight_sixmwt <- read.csv(paste(fight_folder,"sixmwt.csv",sep=""),
                       stringsAsFactors = F,
                       na.strings=c(".","","NA"))

fight_sixmwt$time_completed <- fight_sixmwt$WLKDURM + (fight_sixmwt$WLKDURS/60)


fight_sixmwt_melt <- melt(data=fight_sixmwt,
                        id.vars=c("PATNUMB","VISIT","FORM"),
                        na.rm=T)


fight_sixmwt_melt$studyvisit <- fight_sixmwt_melt$VISIT
fight_sixmwt_melt$form <- "sixmwt"


##########  global  ##########

fight_global <- read.csv(paste(fight_folder,
                           "global.csv",
                           sep=""),
                     stringsAsFactors = F,
                     na.strings=c(".","","NA"))

fight_global_melt <- sqldf("select PATNUMB,
                           VISIT as studyvisit,
                           'GLOBALSC' as variable,
                           GLOBALSC as value,
                           'global' as form
                           from fight_global")


##########  meds  ##########

fight_meds <- read.csv(paste(fight_folder,"meds.csv",sep=""),
                     stringsAsFactors = F,
                     na.strings=c(".","","NA"))


fight_meds$daily_furo_eq_derive <- apply(fight_meds
                                     [,c("FURODOSE",
                                         "TORSDOSE",
                                         "BUMDOSE")],
                                     MARGIN=1,
                                     FUN=diur_calc)

fight_meds_melt <- melt(data=fight_meds,
                 id.vars=c("PATNUMB","VISIT","FORM"),
                 na.rm=T) 

fight_meds_melt$studyvisit <- fight_meds_melt$VISIT
fight_meds_melt$form <- "meds"


########## KCCQ ############

fight_kccq <- read.csv(paste(fight_folder,"kccq.csv",sep=""),
                       stringsAsFactors = F,
                       na.strings=c("","NA",99))


fight_kccq <- score_kccq(dat=fight_kccq,
                         q1a="KCCQ1A",
                         q1b="KCCQ1B",
                         q1c="KCCQ1C",
                         q1d="KCCQ1D",
                         q1e="KCCQ1E",
                         q1f="KCCQ1F",
                         q2="KCCQ2",
                         q3="KCCQ3",
                         q4="KCCQ4",
                         q5="KCCQ5",
                         q6="KCCQ6",
                         q7="KCCQ7",
                         q8="KCCQ8",
                         q9="KCCQ9",
                         q10="KCCQ10",
                         q11="KCCQ11",
                         q12="KCCQ12",
                         q13="KCCQ13",
                         q14="KCCQ14",
                         q15a="KCCQ15A",
                         q15b="KCCQ15B",
                         q15c="KCCQ15C",
                         q15d="KCCQ15D")


fight_kccq_melt <- melt(fight_kccq, id.vars=c("PATNUMB","VISIT","FORM"), na.rm=T)
fight_kccq_melt$studyvisit <- fight_kccq_melt$VISIT
fight_kccq_melt$form <- "kccq"


##########  Compile ##########

fight_melt <-  rbind(fight_base_melt[,c("PATNUMB","studyvisit","form","variable","value")],
                     fight_ecg_melt[,c("PATNUMB","studyvisit","form","variable","value")],
                     fight_echolab_melt[,c("PATNUMB","studyvisit","form","variable","value")],
                     fight_exam_melt[,c("PATNUMB","studyvisit","form","variable","value")],
                     fight_global_melt[,c("PATNUMB","studyvisit","form","variable","value")],
                     fight_kccq_melt[,c("PATNUMB","studyvisit","form","variable","value")],
                     fight_medhist1_melt[,c("PATNUMB","studyvisit","form","variable","value")],
                     fight_medhist2_melt[,c("PATNUMB","studyvisit","form","variable","value")],
                     fight_meds_melt[,c("PATNUMB","studyvisit","form","variable","value")],
                     fight_visitsumm_melt[,c("PATNUMB","studyvisit","form","variable","value")],
                     fight_sixmwt_melt[,c("PATNUMB","studyvisit","form","variable","value")])

names(fight_melt)[1] <- "patientid"
fight_melt$study <- "FIGHT"


########################################### **************** IRONOUT ***************** #############################################
#
# .%%%%.%%%%%%%%...%%%%%%%..%%....%%..%%%%%%%..%%.....%%.%%%%%%%%
# ..%%..%%.....%%.%%.....%%.%%%...%%.%%.....%%.%%.....%%....%%...
# ..%%..%%.....%%.%%.....%%.%%%%..%%.%%.....%%.%%.....%%....%%...
# ..%%..%%%%%%%%..%%.....%%.%%.%%.%%.%%.....%%.%%.....%%....%%...
# ..%%..%%...%%...%%.....%%.%%..%%%%.%%.....%%.%%.....%%....%%...
# ..%%..%%....%%..%%.....%%.%%...%%%.%%.....%%.%%.....%%....%%...
# .%%%%.%%.....%%..%%%%%%%..%%....%%..%%%%%%%...%%%%%%%.....%%...

# Lewis GD, Malhotra R, Hernandez AF, McNulty SE, Smith A, Felker GM, Tang WHW, LaRue SJ, Redfield MM, Semigran MJ, 
# Givertz MM, Van Buren P, Whellan D, Anstrom KJ, Shah MR, Desvigne-Nickens P, Butler J, Braunwald E; 
# NHLBI Heart Failure Clinical Research Network. Effect of Oral Iron Repletion on Exercise Capacity in Patients With Heart Failure 
# With Reduced Ejection Fraction and Iron Deficiency: The IRONOUT HF Randomized Clinical Trial. JAMA. 2017 May 16;317(19):1958-1966. 
# [PMC5703044](https://www.ncbi.nlm.nih.gov/pmc/articles/PMC5703044/) 


##########  demog  ##########

ironout_demog <- read.csv(paste(ironout_folder,"demog.csv",sep=""),
                         stringsAsFactors = F,
                         na.strings=c(".","","NA"))

ironout_demog_melt <-
  ironout_demog[,c("PATNUMB","VISIT","SEX")]

names(ironout_demog_melt)[2:3] <- c("studyvisit","value")
ironout_demog_melt$form <- "demog"
ironout_demog_melt$variable <- "SEX"

ironout_meas <- ironout_demog[,c("PATNUMB","SEX")]


##########  a_base  ##########

ironout_base <- read.csv(paste(ironout_folder,"a_base.csv",sep=""),
                       stringsAsFactors = F,
                       na.strings=c(".","","NA"))

ironout_base$RACE[ironout_base$ETHNIC==1] <- 8
ironout_base$HFAGE_months <- ironout_base$HFAGE * 12

ironout_base_melt <- melt(data=ironout_base,id.vars=c("PATNUMB"),na.rm=T) 
ironout_base_melt$studyvisit <- "BASELINE"
ironout_base_melt$form <- "a_base"

ironout_meas <-
  merge(ironout_meas,
        ironout_base[,c("PATNUMB","AGE","RACE")],
        by="PATNUMB",
        all=T)


##########  a_visitsumm  ##########

ironout_visitsumm <- read.csv(paste(ironout_folder,"a_visitsumm.csv",sep=""),
                            stringsAsFactors = F,
                            na.strings=c(".","","NA"))

ironout_visitsumm <- merge(ironout_visitsumm,ironout_meas,by=c("PATNUMB"),all.x=T)
ironout_visitsumm$weight_kg <- round(ironout_visitsumm$WTLBS * 0.453592,1)

ironout_crcl <- ironout_visitsumm

ironout_crcl$crcl <- calc_MDRD4(dat=ironout_crcl,
                                   cr="CREAT", 
                                   age="AGE", 
                                   sex="SEX", 
                                   race="RACE", 
                                   male=1, 
                                   black=3 )

ironout_crcl <- 
  ironout_crcl[,c("PATNUMB","VISIT","crcl")]

ironout_crcl$variable <- "crcl"
ironout_crcl$value <- ironout_crcl$crcl
ironout_crcl$studyvisit <- ironout_crcl$VISIT
ironout_crcl$form <- "a_visitsumm"

ironout_visitsumm_melt <- melt(data=ironout_visitsumm,
                             id.vars=c("PATNUMB","VISIT"),
                             na.rm=T) 

ironout_visitsumm_melt$studyvisit <- ironout_visitsumm_melt$VISIT
ironout_visitsumm_melt$form <- "a_visitsumm"

ironout_visitsumm_melt <-
  rbind(ironout_visitsumm_melt[,c("PATNUMB","VISIT","variable","value","studyvisit","form")],
        ironout_crcl[,c("PATNUMB","VISIT","variable","value","studyvisit","form")])


##########  medhist1  ##########

ironout_medhist1 <- read.csv(paste(ironout_folder,"medhist1.csv",sep=""),
                           stringsAsFactors = F,
                           na.strings=c(".","","NA"))

ironout_medhist1 <- 
  merge(ironout_medhist1,
        ironout_base[,c("PATNUMB","HFETIOLOGY")],
        by="PATNUMB")

ironout_medhist1$HFETIOLOGY[
  ironout_medhist1$HFETIOLOGY==2] <- 0

ironout_medhist1[
  ironout_medhist1$HFETIOLOGY==1,
  c("ALCOHOLC",
    "CYTOTOX",
    "FAMILIAL",
    "HCM",
    "HYPERTN",
    "IDIODIL",
    "IDIORES",
    "MYOOTH",
    "PERIPRT",
    "VALVUL")] <- NA

ironout_medhist1$num_etiol <- 
  apply(ironout_medhist1[c("HFETIOLOGY",
                           "ALCOHOLC",
                           "CYTOTOX",
                           "FAMILIAL",
                           "HCM",
                           "HYPERTN",
                           "IDIODIL",
                           "IDIORES",
                           "MYOOTH",
                           "PERIPRT",
                           "VALVUL")],
        MARGIN=1,
        FUN=function(x)
          sum(x,na.rm=T))

ironout_medhist1[
  ironout_medhist1$num_etiol>1,
  c("ALCOHOLC",
    "CYTOTOX",
    "FAMILIAL",
    "HCM",
    "HYPERTN",
    "IDIODIL",
    "IDIORES",
    "MYOOTH",
    "PERIPRT",
    "VALVUL")] <- NA

ironout_medhist1_melt <- 
  melt(data=ironout_medhist1,
       id.vars=c("PATNUMB","VISIT","FORM"),
       na.rm=T) 

ironout_medhist1_melt$studyvisit <- ironout_medhist1_melt$VISIT
ironout_medhist1_melt$form <- "medhist1"


##########  medhist2  ##########

ironout_medhist2 <- 
  read.csv(paste(ironout_folder,"medhist2.csv",
                 sep=""),
           stringsAsFactors = F,
           na.strings=c(".","","NA"))

ironout_medhist2_melt <- 
  melt(data=ironout_medhist2,
       id.vars=c("PATNUMB","VISIT","FORM"),
       na.rm=T) 

ironout_medhist2_melt$studyvisit <- ironout_medhist2_melt$VISIT
ironout_medhist2_melt$form <- ironout_medhist2_melt$form <- "medhist2"


##########  exam  ##########

ironout_exam <- 
  read.csv(paste(ironout_folder,"exam.csv",sep=""),
           stringsAsFactors = F,
           na.strings=c(".","","NA"))

ironout_exam$pupr <- ironout_exam$BPSYS-ironout_exam$BPDIA


ironout_exam_melt <- melt(data=ironout_exam,
                        id.vars=c("PATNUMB","VISIT","FORM"),
                        na.rm=T)

ironout_exam_melt$studyvisit <- ironout_exam_melt$VISIT
ironout_exam_melt$form <- "exam"


##########  sixmwt  ##########

ironout_sixmwt <- 
  read.csv(paste(ironout_folder,"sixmwt.csv",sep=""),
           stringsAsFactors = F,
           na.strings=c(".","","NA"))

ironout_sixmwt$time_completed <- 
  ironout_sixmwt$WLKDURM + (ironout_sixmwt$WLKDURS/60)


ironout_sixmwt_melt <- 
  melt(data=ironout_sixmwt,
       id.vars=c("PATNUMB","VISIT","FORM"),
       na.rm=T)


ironout_sixmwt_melt$studyvisit <- ironout_sixmwt_melt$VISIT
ironout_sixmwt_melt$form <- "sixmwt"


##########  meds  ##########

ironout_meds <- 
  read.csv(paste(ironout_folder,"meds.csv",sep=""),
           stringsAsFactors = F,
           na.strings=c(".","","NA"))


ironout_meds$daily_furo_eq_derive <- 
  apply(ironout_meds
        [,c("FURODOSE",
            "TORSDOSE",
            "BUMDOSE")],
        MARGIN=1,
        FUN=diur_calc)

ironout_meds_melt <- 
  melt(data=ironout_meds,
       id.vars=c("PATNUMB","VISIT","FORM"),
       na.rm=T) 

ironout_meds_melt$studyvisit <- ironout_meds_melt$VISIT
ironout_meds_melt$form <- "meds"


########## KCCQ ############

ironout_kccq <- read.csv(paste(ironout_folder,"kccq.csv",sep=""),
                       stringsAsFactors = F,
                       na.strings=c("","NA",99))

ironout_kccq <- score_kccq(dat=ironout_kccq,
                         q1a="KCCQ1A",
                         q1b="KCCQ1B",
                         q1c="KCCQ1C",
                         q1d="KCCQ1D",
                         q1e="KCCQ1E",
                         q1f="KCCQ1F",
                         q2="KCCQ2",
                         q3="KCCQ3",
                         q4="KCCQ4",
                         q5="KCCQ5",
                         q6="KCCQ6",
                         q7="KCCQ7",
                         q8="KCCQ8",
                         q9="KCCQ9",
                         q10="KCCQ10",
                         q11="KCCQ11",
                         q12="KCCQ12",
                         q13="KCCQ13",
                         q14="KCCQ14",
                         q15a="KCCQ15A",
                         q15b="KCCQ15B",
                         q15c="KCCQ15C",
                         q15d="KCCQ15D")

ironout_kccq_melt <- melt(ironout_kccq, id.vars=c("PATNUMB","VISIT","FORM"), na.rm=T)
ironout_kccq_melt$studyvisit <- ironout_kccq_melt$VISIT
ironout_kccq_melt$form <- "kccq"


##########  Compile ##########

ironout_melt <-  
  rbind(ironout_base_melt[,c("PATNUMB","studyvisit","form","variable","value")],
        ironout_demog_melt[,c("PATNUMB","studyvisit","form","variable","value")],
        ironout_exam_melt[,c("PATNUMB","studyvisit","form","variable","value")],
        ironout_kccq_melt[,c("PATNUMB","studyvisit","form","variable","value")],
        ironout_medhist1_melt[,c("PATNUMB","studyvisit","form","variable","value")],
        ironout_medhist2_melt[,c("PATNUMB","studyvisit","form","variable","value")],
        ironout_meds_melt[,c("PATNUMB","studyvisit","form","variable","value")],
        ironout_visitsumm_melt[,c("PATNUMB","studyvisit","form","variable","value")],
        ironout_sixmwt_melt[,c("PATNUMB","studyvisit","form","variable","value")])

names(ironout_melt)[1] <- "patientid"
ironout_melt$study <- "IRONOUT"


########################################### ***************** INDIE-HFpEF ************ #############################################
#
# .%%%%.%%....%%.%%%%%%%%..%%%%.%%%%%%%%.........%%.....%%.%%%%%%%%.%%%%%%%%..%%%%%%%%.%%%%%%%%
# ..%%..%%%...%%.%%.....%%..%%..%%...............%%.....%%.%%.......%%.....%%.%%.......%%......
# ..%%..%%%%..%%.%%.....%%..%%..%%...............%%.....%%.%%.......%%.....%%.%%.......%%......
# ..%%..%%.%%.%%.%%.....%%..%%..%%%%%%...%%%%%%%.%%%%%%%%%.%%%%%%...%%%%%%%%..%%%%%%...%%%%%%..
# ..%%..%%..%%%%.%%.....%%..%%..%%...............%%.....%%.%%.......%%........%%.......%%......
# ..%%..%%...%%%.%%.....%%..%%..%%...............%%.....%%.%%.......%%........%%.......%%......
# .%%%%.%%....%%.%%%%%%%%..%%%%.%%%%%%%%.........%%.....%%.%%.......%%........%%%%%%%%.%%......

# Borlaug BA, Anstrom KJ, Lewis GD, Shah SJ, Levine JA, Koepp GA, Givertz MM, Felker GM, LeWinter MM, Mann DL, Margulies KB, 
# Smith AL, Tang WHW, Whellan DJ, Chen HH, Davila-Roman VG, McNulty S, Desvigne-Nickens P, Hernandez AF, Braunwald E, 
# Redfield MM; National Heart, Lung, and Blood Institute Heart Failure Clinical Research Network. 
# Effect of Inorganic Nitrite vs Placebo on Exercise Capacity Among Patients With Heart Failure With Preserved Ejection Fraction: 
# The INDIE-HFpEF Randomized Clinical Trial. JAMA. 2018;320(17):1764-1773. [PMC6248105](https://www.ncbi.nlm.nih.gov/pmc/articles/PMC6248105/)


##########  a_base  ##########

indie_base <- read.csv(paste(indie_folder,"a_base.csv",sep=""),
                       stringsAsFactors = F,
                       na.strings=c(".","","NA"))

indie_demog <- read.csv(paste(indie_folder,"demog.csv",sep=""),
                       stringsAsFactors = F,
                       na.strings=c(".","","NA"))

indie_base <- merge(indie_base,indie_demog[,c("PATNUMB","SEX")],by="PATNUMB",all.x=T)

indie_base$RACE[indie_base$ETHNIC==1] <- 8
indie_base$HFAGE_months <- indie_base$HFAGE * 12

indie_base_melt <- melt(data=indie_base,id.vars=c("PATNUMB"),na.rm=T) 
indie_base_melt$studyvisit <- "BASE"
indie_base_melt$form <- "a_base"

indie_meas <- indie_base[,c("PATNUMB","AGE","SEX","RACE","HEIGHTIN_BL")]
indie_meas$height_cm <- round(indie_meas$HEIGHTIN_BL * 2.54,1)


##########  a_visitsumm  ##########

indie_visitsumm <- read.csv(paste(indie_folder,"a_visitsumm.csv",sep=""),
                            stringsAsFactors = F,
                            na.strings=c(".","","NA"))

indie_visitsumm <- merge(indie_visitsumm,indie_meas,by=c("PATNUMB"),all.x=T)
indie_visitsumm$weight_kg <- round(indie_visitsumm$WTLBS * 0.453592,1)
indie_visitsumm$bmi <- calc_bmi(indie_visitsumm)
indie_visitsumm$bsa <- calc_bsa(indie_visitsumm)


indie_visitsumm$CrCl <- calc_MDRD4(dat=indie_visitsumm,
                                   cr="CREAT", 
                                   age="AGE", 
                                   sex="SEX", 
                                   race="RACE", 
                                   male=1, 
                                   black=3 )

indie_visitsumm_melt <- melt(data=indie_visitsumm,
                             id.vars=c("PATNUMB","VISIT"),
                             na.rm=T) 

indie_visitsumm_melt$studyvisit <- indie_visitsumm_melt$VISIT
indie_visitsumm_melt$form <- "a_visitsumm"


indie_meas <- merge(indie_meas,
                    indie_visitsumm[indie_visitsumm$VISIT=="BASE",
                                    c("PATNUMB",
                                      "AGE",
                                      "SEX",
                                      "RACE",
                                      "HEIGHTIN_BL",
                                      "height_cm",
                                      "WTLBS",
                                      "weight_kg")])


##########  echolab  ##########

indie_echolab <- read.csv(paste(indie_folder,"echolab.csv",sep=""),
                           stringsAsFactors = F,
                           na.strings=c(".","","NA"))

indie_echolab <- merge(indie_echolab, 
                       indie_meas, 
                       by="PATNUMB", 
                       all.x=T)

indie_echolab$LATMIAE_cmsec <- indie_echolab$LATMIAE*100
indie_echolab$MEDMIE_cmsec <- indie_echolab$MEDMIE*100
indie_echolab$EVELOC_cmsec <- indie_echolab$EVELOC*100
indie_echolab$RVSP <- 4*(indie_echolab$PEAKVELC^2) + indie_echolab$RAPRESS

indie_echolab$LVED_best <- indie_echolab$LVEDCH
indie_echolab$LVED_best[is.na(indie_echolab$LVED_best)&!is.na(indie_echolab$LVEDVOL)] <-
  indie_echolab$LVEDVOL[is.na(indie_echolab$LVED_best)&!is.na(indie_echolab$LVEDVOL)]


# Retaining code for diastolic dyfunction here, but A-wave velocity not available in present data from BioLINCC

# indie_echolab$AVELOC_cmsec <- indie_echolab$AVELOC*100
# indie_echolab$earatio <- indie_echolab$EVELOC_cmsec/indie_echolab$AVELOC_cmsec
# indie_echolab$topcat_dd <-  assign_diastolic_function(dat=indie_echolab,
#                                                       ep_lat="LATMIAE_cmsec",
#                                                       ep_sept="MEDMIE_cmsec",
#                                                       earat="earatio",
#                                                       e_vel="EVELOC_cmsec",
#                                                       dt = "DECELTM",
#                                                       tr_vel = "PEAKVELC",
#                                                       def="TOPCAT",
#                                                       outpt="num")

indie_echolab <- calc_hypertrophy_type(df=indie_echolab,
                                       sex="SEX",
                                       male="1",
                                       female="2",
                                       lvedd_cm="LVDD",
                                       ivsd_cm="IVSDDD",
                                       lvpwtd_cm="PWSD",
                                       height_cm="height_cm",
                                       weight_kg="weight_kg")


indie_echolab <- calc_lav(indie_echolab,
                          laa_4c = "LACAREA",
                          laa_2c = "LACHAREA",
                          lal_4c = "LACLEN",
                          lal_2c = "LALEN",
                          height_cm = "height_cm",
                          weight_kg = "weight_kg")


indie_echolab$LVEDD_index <- indie_echolab$LVDD/indie_echolab$bsa
indie_echolab$LVESD_index <- indie_echolab$LVSD/indie_echolab$bsa

indie_echolab$eeprime_lat <- round(indie_echolab$EVELOC/indie_echolab$LATMIAE,1)
indie_echolab$eeprime_sept <- round(indie_echolab$EVELOC/indie_echolab$MEDMIE,1)
indie_echolab$eeprime_avg <- round((indie_echolab$eeprime_lat+indie_echolab$eeprime_sept)/2,1)


indie_echolab_melt <-  melt(data=indie_echolab,id.vars=c("PATNUMB","VISIT"),na.rm=T) 

indie_echolab_melt$studyvisit <- indie_echolab_melt$VISIT
indie_echolab_melt$form <- "echolab"


##########  medhist1  ##########

indie_medhist1 <- read.csv(paste(indie_folder,"medhist1.csv",sep=""),
                           stringsAsFactors = F,
                           na.strings=c(".","","NA"))

indie_medhist1 <- merge(indie_medhist1,indie_base[,c("PATNUMB","HFETIOLOGY")],by="PATNUMB")

indie_medhist1$HFETIOLOGY[indie_medhist1$HFETIOLOGY==2] <- 0

indie_medhist1[indie_medhist1$HFETIOLOGY==1,c("ALCOHOLC",
                                              "CYTOTOX",
                                              "FAMILIAL",
                                              "HCM",
                                              "HYPERTN",
                                              "IDIODIL",
                                              "IDIORES",
                                              "MYOOTH",
                                              "PERIPRT",
                                              "VALVUL")] <- NA

indie_medhist1$num_etiol <- apply(indie_medhist1[c("HFETIOLOGY",
                                                   "ALCOHOLC",
                                                   "CYTOTOX",
                                                   "FAMILIAL",
                                                   "HCM",
                                                   "HYPERTN",
                                                   "IDIODIL",
                                                   "IDIORES",
                                                   "MYOOTH",
                                                   "PERIPRT",
                                                   "VALVUL")],
                                  MARGIN=1,
                                  FUN=function(x)
                                    sum(x,na.rm=T))

indie_medhist1[indie_medhist1$num_etiol>1,c("ALCOHOLC",
                                            "CYTOTOX",
                                            "FAMILIAL",
                                            "HCM",
                                            "HYPERTN",
                                            "IDIODIL",
                                            "IDIORES",
                                            "MYOOTH",
                                            "PERIPRT",
                                            "VALVUL")] <- NA

indie_medhist1_melt <- melt(data=indie_medhist1,id.vars=c("PATNUMB","VISIT","FORM"),na.rm=T) 
indie_medhist1_melt$studyvisit <- indie_medhist1_melt$VISIT
indie_medhist1_melt$form <- "medhist1"


##########  medhist2  ##########

indie_medhist2 <- read.csv(paste(indie_folder,"medhist2.csv",sep=""),
                           stringsAsFactors = F,
                           na.strings=c(".","","NA"))

indie_medhist2_melt <- melt(data=indie_medhist2,id.vars=c("PATNUMB","VISIT","FORM"),na.rm=T) 
indie_medhist2_melt$studyvisit <- indie_medhist2_melt$VISIT
indie_medhist2_melt$form <- indie_medhist2_melt$form <- "medhist2"


##########  ecg  ##########

indie_ecg <- read.csv(paste(indie_folder,"ecg.csv",sep=""),
                      stringsAsFactors = F,
                      na.strings=c(".","","NA"))

indie_ecg_melt <- melt(data=indie_ecg,
                       id.vars=c("PATNUMB","VISIT","FORM"),
                       na.rm=T)

indie_ecg_melt$studyvisit <- indie_ecg_melt$VISIT
indie_ecg_melt$form <- "ecg"


##########  exam  ##########

indie_exam <- read.csv(paste(indie_folder,"exam.csv",sep=""),
                       stringsAsFactors = F,
                       na.strings=c(".","","NA"))

indie_exam$pupr <- indie_exam$BPSYS-indie_exam$BPDIA


indie_exam_melt <- melt(data=indie_exam,
                        id.vars=c("PATNUMB","VISIT","FORM"),
                        na.rm=T)

indie_exam_melt$studyvisit <- indie_exam_melt$VISIT
indie_exam_melt$form <- "exam"


##########  meds  ##########

indie_meds <- read.csv(paste(indie_folder,"meds.csv",sep=""),
                       stringsAsFactors = F,
                       na.strings=c(".","","NA"))


indie_meds$daily_furo_eq_derive <- apply(indie_meds
                                         [,c("FURODOSE",
                                             "TORSDOSE",
                                             "BUMDOSE")],
                                         MARGIN=1,
                                         FUN=diur_calc)

indie_meds_melt <- melt(data=indie_meds,
                        id.vars=c("PATNUMB","VISIT","FORM"),
                        na.rm=T) 

indie_meds_melt$studyvisit <- indie_meds_melt$VISIT
indie_meds_melt$form <- "meds"


####  cpet  ####

indie_cpet <- read.csv(paste(indie_folder,"cpet.csv",sep=""))
indie_cpet[indie_cpet=="NaN"] <- NA

indie_cpet_melt <- 
  subset(melt(data=indie_cpet,id.vars=c("PATNUMB","VISIT","FORM"),na.rm=T))
indie_cpet_melt$studyvisit <- indie_cpet_melt$VISIT
indie_cpet_melt$form <- "cpet"


########## KCCQ ############

indie_kccq <- read.csv(paste(indie_folder,"kccq.csv",sep=""),
                       stringsAsFactors = F,
                       na.strings=c("","NA",99))


indie_kccq <- score_kccq(dat=indie_kccq,
                         q1a="KCCQ1A",
                         q1b="KCCQ1B",
                         q1c="KCCQ1C",
                         q1d="KCCQ1D",
                         q1e="KCCQ1E",
                         q1f="KCCQ1F",
                         q2="KCCQ2",
                         q3="KCCQ3",
                         q4="KCCQ4",
                         q5="KCCQ5",
                         q6="KCCQ6",
                         q7="KCCQ7",
                         q8="KCCQ8",
                         q9="KCCQ9",
                         q10="KCCQ10",
                         q11="KCCQ11",
                         q12="KCCQ12",
                         q13="KCCQ13",
                         q14="KCCQ14",
                         q15a="KCCQ15A",
                         q15b="KCCQ15B",
                         q15c="KCCQ15C",
                         q15d="KCCQ15D")


indie_kccq_melt <- melt(indie_kccq, id.vars=c("PATNUMB","VISIT","FORM"), na.rm=T)
indie_kccq_melt$studyvisit <- indie_kccq_melt$VISIT
indie_kccq_melt$form <- "kccq"


##########  Compile ##########

indie_melt <-  rbind(indie_base_melt[,c("PATNUMB","studyvisit","form","variable","value")],
                     indie_ecg_melt[,c("PATNUMB","studyvisit","form","variable","value")],
                     indie_echolab_melt[,c("PATNUMB","studyvisit","form","variable","value")],
                     indie_exam_melt[,c("PATNUMB","studyvisit","form","variable","value")],
                     indie_kccq_melt[,c("PATNUMB","studyvisit","form","variable","value")],
                     indie_medhist1_melt[,c("PATNUMB","studyvisit","form","variable","value")],
                     indie_medhist2_melt[,c("PATNUMB","studyvisit","form","variable","value")],
                     indie_meds_melt[,c("PATNUMB","studyvisit","form","variable","value")],
                     indie_visitsumm_melt[,c("PATNUMB","studyvisit","form","variable","value")])

names(indie_melt)[1] <- "patientid"
indie_melt$study <- "INDIE-HFpEF"
indie_melt$variable <- as.character(indie_melt$variable)


########################################### ************ LIFE ************ #############################################
#
# .&&.......&&&&.&&&&&&&&.&&&&&&&&
# .&&........&&..&&.......&&......
# .&&........&&..&&.......&&......
# .&&........&&..&&&&&&...&&&&&&..
# .&&........&&..&&.......&&......
# .&&........&&..&&.......&&......
# .&&&&&&&&.&&&&.&&.......&&&&&&&&

# Mann DL, Givertz MM, Vader JM, Starling RC, Shah P, Mcnulty, SE, Anstrom KJ, Margulies KB, 
# Kiernan MS, Mahr C, Gupta D, Redfield MM, Lala A, Lewis GD, Devore AD, Desvigne-Nickens P, 
# Hernandez AF, Braunwald E. Effect of Treatment With Sacubitril/Valsartan in Patients With Advanced 
# Heart Failure and Reduced Ejection Fraction A Randomized Clinical Trial JAMA Cardiol. 2021;7(1):17–25. 
# https://doi.org/10.1001/jamacardio.2021.4567


########## base  ##########

life_base <- read.csv(paste(life_folder,"base.csv",sep=""),
                       stringsAsFactors = F,
                       na.strings=c(".","","NA"))

life_base <- subset(life_base,is.na(COVID19_RAND))

life_visitsumm <- read.csv(paste(life_folder,"visitsum.csv",sep=""),
                           stringsAsFactors = F,
                           na.strings=c(".","","NA"))

life_base$race[life_base$ethnic=="Hispanic or Latino"] <- "Hispanic"
life_base$HFAGE_months <- life_base$HFAGE * 12

life_base <- merge(life_base,
                   life_visitsumm[life_visitsumm$visit=="Screening",c("patnumb",'heightin')],
                   by='patnumb',
                   all.x=T)

life_base$height_cm <- life_base$heightin * 2.54

life_base$MYOOTH[life_base$HFETIOLOGY=="Ischemic"] <- NA

life_base$MYOOTH[life_base$patnumb %in% c("LF0256","LF0304","LF0310")] <- NA

lb1 <- life_base[,c("HFETIOLOGY","ALCOHOLC","CYTOTOX","FAMILIAL","HYPERTN",
                    "IDIODIL","PERIPRT","VALVUL")]

lb1$HFETIOLOGY[lb1$HFETIOLOGY=="Ischemic"] <- 1 
lb1$HFETIOLOGY[lb1$HFETIOLOGY=="Non-Ischemic"] <- 0

lb1[lb1=="Yes"] <- 1
lb1[is.na(lb1)] <- 0

life_base$num_etiol <- apply(X=lb1,
                             MARGIN=1,
                             function (x)
                               sum(as.numeric(x),na.rm=T))

## If multiple etiologies, set all other etiologies to NA

life_base[life_base$num_etiol>1,c("HFETIOLOGY","ALCOHOLC","CYTOTOX","FAMILIAL","HYPERTN",
                                  "IDIODIL","PERIPRT","VALVUL")] <- NA

life_base_melt <- melt(data=life_base,id.vars=c("patnumb"),na.rm=T) 
life_base_melt$studyvisit <- "Baseline"
life_base_melt$form <- "base"

life_meas <- life_base[,c("patnumb","AGE","SEX","race","height_cm")]


##########  a_visitsumm  ##########

life_visitsumm <- read.csv(paste(life_folder,"visitsum.csv",sep=""),
                            stringsAsFactors = F,
                            na.strings=c(".","","NA"))

life_visitsumm <- merge(life_visitsumm,life_meas,by=c("patnumb"),all.x=T)
life_visitsumm$weight_kg <- round(life_visitsumm$wtlbs * 0.453592,1)
life_visitsumm$bsa <- calc_bsa(life_visitsumm)

life_base <- merge(life_base,
                        life_visitsumm[life_visitsumm$visit=="Baseline",
                                       c("patnumb","insulin")])
life_base$insulin[life_base$DIABETES=="No"] <- NA

life_visitsumm$pupr <- 
  life_visitsumm$sbp-
  life_visitsumm$dbp

life_visitsumm_melt <- melt(data=life_visitsumm,
                             id.vars=c("patnumb","visit"),
                             na.rm=T) 

life_visitsumm_melt$visit[
  life_visitsumm_melt$variable %in% c("phylim",
                                      "symstab",
                                      "symfreq",
                                      "symburd",
                                      "tsympt",
                                      "selfeff",
                                      "qualife",
                                      "soclim",
                                      "ovsumm",
                                      "clsumm")&
    life_visitsumm_melt$visit=="Screening"] <- "Baseline"

life_visitsumm_melt$studyvisit <- life_visitsumm_melt$visit
life_visitsumm_melt$form <- "visitsum"

##########  Compile ##########

life_melt <-  rbind(life_base_melt[,c("patnumb","studyvisit","form","variable","value")],
                     life_visitsumm_melt[,c("patnumb","studyvisit","form","variable","value")])

life_melt$variable <- as.character(life_melt$variable)

names(life_melt)[1] <- "patientid"
life_melt$study <- "LIFE"


#%%%%%%%%%%%%%%%%%%%%%%%%%%%%%%%%%%%%%%%%%%%%%%%%%%%%%%%%%%%%%%%%%
####                    COMPILE ALL STUDIES                   ####
#%%%%%%%%%%%%%%%%%%%%%%%%%%%%%%%%%%%%%%%%%%%%%%%%%%%%%%%%%%%%%%%%%

alldata <- rbind(athena_melt[,c("patientid","study","studyvisit","form","variable","value")],
                 best_melt[,c("patientid","study","studyvisit","form","variable","value")],
                 carress_melt[,c("patientid","study","studyvisit","form","variable","value")],
                 dig_melt[,c("patientid","study","studyvisit","form","variable","value")],
                 dose_melt[,c("patientid","study","studyvisit","form","variable","value")],
                 escape_melt[,c("patientid","study","studyvisit","form","variable","value")],
                 exact_melt[,c("patientid","study","studyvisit","form","variable","value")],
                 fight_melt[,c("patientid","study","studyvisit","form","variable","value")],
                 guideit_melt[,c("patientid","study","studyvisit","form","variable","value")],
                 hfaction_melt[,c("patientid","study","studyvisit","form","variable","value")],
                 indie_melt[,c("patientid","study","studyvisit","form","variable","value")],
                 ipreserve_melt[,c("patientid","study","studyvisit","form","variable","value")],
                 ironout_melt[,c("patientid","study","studyvisit","form","variable","value")],
                 life_melt[,c("patientid","study","studyvisit","form","variable","value")],
                 mocha_melt[,c("patientid","study","studyvisit","form","variable","value")],
                 neat_melt[,c("patientid","study","studyvisit","form","variable","value")],
                 paradigm_melt[,c("patientid","study","studyvisit","form","variable","value")],
                 relax_melt[,c("patientid","study","studyvisit","form","variable","value")],
                 rose_melt[,c("patientid","study","studyvisit","form","variable","value")],
                 scdheft_melt[,c("patientid","study","studyvisit","form","variable","value")],
                 solvd_melt[,c("patientid","study","studyvisit","form","variable","value")],
                 solvd_reg_melt[,c("patientid","study","studyvisit","form","variable","value")],
                 stich_melt[,c("patientid","study","studyvisit","form","variable","value")],
                 topcat_melt[,c("patientid","study","studyvisit","form","variable","value")])

alldata$variable <- as.character(alldata$variable)
alldata$value <- trimws(alldata$value)

b <- Sys.time()

b-zpz
#%%%%%%%%%%%%%%%%%%%%%%%%%%%%%%%%%%%%%%%%%%%%%%%%%%%%%%%%%%%%%%%%%
####              ******CLEAN-UP******                        ####
#%%%%%%%%%%%%%%%%%%%%%%%%%%%%%%%%%%%%%%%%%%%%%%%%%%%%%%%%%%%%%%%%%


alldata <- subset(alldata, !value %in% c(NA,"NaN","NULL"))
alldata <- subset(alldata,!variable %in% c("PAGEID","VISDT_DAYS","FORM","VDATE","visit_dt1",
                                           "VER_ID","noncom_use","com_use","VISIT","VISIT_DT","PERIOD","HOSPCODE",
                                           "FORMVER","FWKVISIT","WKNOTEST","",NA))

visit_wks <- read.csv('~/Dropbox/ADAPT-HF/Master HDCP files/visit_weeks.csv')

visit_wks <- subset(visit_wks,!is.na(study_wks))

alldata <- merge(alldata,visit_wks,by=c("study","studyvisit"),all.x=T)


alldata <- subset(alldata, !is.na(alldata$study_wks))
alldata <- sqldf("select distinct patientid,
                 form,
                 studyvisit,
                 variable,
                 value,
                 study,
                 study_wks 
                 from alldata")

alldata$datapoint <- rownames(alldata)
alldata$study_field <- toupper(paste(alldata$study,alldata$form,alldata$variable,sep="_"))


rm(list=ls(pattern="\\bhfaction_."))
rm(list=ls(pattern="\\bbest_."))
rm(list=ls(pattern="\\bscdheft_."))
rm(list=ls(pattern="\\btopcat_."))
rm(list=ls(pattern="\\bparadigm_."))
rm(list=ls(pattern="\\bcorona_."))
rm(list=ls(pattern="\\bipreserve_."))
rm(list=ls(pattern="\\brelax_."))
rm(list=ls(pattern="\\bneat_."))
rm(list=ls(pattern="\\bdig_."))
rm(list=ls(pattern="\\bescape_."))
rm(list=ls(pattern="\\brose_."))
rm(list=ls(pattern="\\bexact_."))
rm(list=ls(pattern="\\bdose_."))
rm(list=ls(pattern="\\bcarress_."))
rm(list=ls(pattern="\\bmocha_."))
rm(list=ls(pattern="\\bstich."))
rm(list=ls(pattern="\\bsolvd."))
rm(list=ls(pattern="\\bguideit_."))
rm(list=ls(pattern="\\bfight_."))
rm(list=ls(pattern="\\bathena_."))
rm(list=ls(pattern="\\indie_."))
rm(list=ls(pattern="\\ironout_."))


#=======================================================================#
#=======================================================================#
##                                                                     ##
####            BEGIN HIGH-DIMENSIONAL CLINICAL PHENOTYPING          ####
##                                                                     ##
#=======================================================================#
#=======================================================================#


hdcp_trial_criteria_load <- read.csv("~/Dropbox/ADAPT-HF/Master HDCP files/Criteriaset_multistudy 2023.csv",
                                      na.strings=c("NA","","NULL","\\N"),
                                      stringsAsFactors = F)

hdcp_names <- read.csv("~/Dropbox/ADAPT-HF/Master HDCP files/phenotypelist_trial.csv",
                       na.strings=c("","NA",NA,"NULL"))


hdcp_trial_criteria_load$rule_num <- rownames(hdcp_trial_criteria_load)

hdcp_trial_criteria <- hdcp_trial_criteria_load
hdcp_trial_criteria$lower[is.na(hdcp_trial_criteria$lower)&hdcp_trial_criteria$rule_type=="range"] <- -1000000
hdcp_trial_criteria$upper[is.na(hdcp_trial_criteria$upper)&hdcp_trial_criteria$rule_type=="range"] <- 1000000


## This uses the 'include_upper' and 'include_lower' fields in the 'criteriaset...' input file to determine whether ranges are functionally e.g. < vs. <=
## This is done since ranges may vary in inclusion of their bounds.
## Note that it is only necessary to indicate "Y" in 'criteriaset.'  Again this is for ease of reading.
## I fudge this by just adding small values to increase or decrease the bounds so that the upper or lower limits are included in the < vs. > statements
## I added this so that the 'criteriaset' file would be easier to read rather than adjusting all of the bounds in a similar fashion.

hdcp_trial_criteria$upper[!is.na(hdcp_trial_criteria$include_upper)&hdcp_trial_criteria$include_upper=="Y"] <- 
  hdcp_trial_criteria$upper[!is.na(hdcp_trial_criteria$include_upper)&hdcp_trial_criteria$include_upper=="Y"] + 0.000001
hdcp_trial_criteria$lower[!is.na(hdcp_trial_criteria$include_lower)&hdcp_trial_criteria$include_lower=="Y"] <- 
  hdcp_trial_criteria$lower[!is.na(hdcp_trial_criteria$include_lower)&hdcp_trial_criteria$include_lower=="Y"] - 0.000001

hdcp_trial_criteria$study_field <- paste(hdcp_trial_criteria$study,
                                         hdcp_trial_criteria$form,
                                         hdcp_trial_criteria$field,
                                          sep="_")

hdcp_trial_criteria$study_field <- toupper(hdcp_trial_criteria$study_field)

hdcp_trial_fields_used <- sqldf("select distinct study, 
                                  form,
                                  field,
                                  study_field
                                  from hdcp_trial_criteria")


##### Make unique patient list with sex  ####

ptlist <- sqldf("select distinct
      j.study,
      patientid,
      phenotype,
      value,
      string_value
      from alldata as j
      join hdcp_trial_criteria as c
      using (study_field) 
      where value = c.match_string and
      phenotype = 'Sex'
      ")

ptlist$sex <- gsub("([0-9])\\.\\s+",
                   "",
                   ptlist$string_value)

ptlist <- ptlist[,c("study",
                    "patientid",
                    "sex")]

####    Apply 'match' rules  ####
##      Compares match_string with 'value' field

m <- sqldf("select 
      patientid,
      j.study,
      variable,
      value,
      phenotype,
      rank,
      string_value,
      studyvisit,
      study_wks,
      rule_num,
      datapoint
      from alldata as j
      join hdcp_trial_criteria as c
      using (study_field)
      where c.rule_type = 'match' and
      value = c.match_string 
      ", stringsAsFactors = F)


#### Apply 'range' rules ####
## Compare value with upper and lower bounds
## For convenience, set lower bound to -1000000 and upper bound to 1000000 where either is not specified.
## THis allows application of range criteria with only a single expression

r <- sqldf("select 
      patientid,
      j.study,
      j.form,
      variable,
      lower,
      value,
      upper,
      phenotype,
      rank,
      string_value,
      studyvisit,
      study_wks,
      rule_num,
      datapoint
      from alldata as j
      join hdcp_trial_criteria as c
      using (study_field)
      where c.rule_type = 'range' and
      sex_specific=='no' and
      value > lower and
      value < upper",stringsAsFactors = F)

r[r==-1000000] <- NA
r[r==1000000] <- NA

r_sex <- sqldf("select 
      patientid,
      sex_specific,
      sex,
      j.study,
      j.form,
      variable,
      lower,
      value,
      upper,
      phenotype,
      rank,
      string_value,
      studyvisit,
      study_wks,
      rule_num,
      datapoint
      from alldata as j
      join hdcp_trial_criteria as c
      using (study_field)
      join ptlist using (patientid,study)
      where c.rule_type = 'range' and
      sex_specific='yes' and
      criteria_sex = sex and
      value > lower and
      value < upper",stringsAsFactors = F)

r_sex[r_sex==-1000000] <- NA
r_sex[r_sex==1000000] <- NA

#### Apply identity rules ####
## Pass value through unchanged for rules marked 'value'.  This effectively just assigns a phenotype name.
## Units are not considered here so be careful.

v <- sqldf("select patientid,
      j.study,
      j.form,
      studyvisit,
      study_wks,
      variable,
      phenotype,
      value as string_value,
      rule_num,
      datapoint
      from alldata as j
      join hdcp_trial_criteria as c
      using (study_field)
      where c.rule_type = 'value'
           ",stringsAsFactors = F)

v$rank <-  as.numeric(v$string_value)

## Now assemble everything
## Only some columns will be included in the combined datasets.  However the rule number will be retained for auditing if necessary.

merged_phenotypedata <- rbind(m[,c("study",
                                   "patientid",
                                   "studyvisit",
                                   "study_wks",
                                   "phenotype",
                                   "string_value",
                                   "rank",
                                   "rule_num",
                                   "datapoint")],
                              r[,c("study",
                                   "patientid",
                                   "studyvisit",
                                   "study_wks",
                                   "phenotype",
                                   "string_value",
                                   "rank",
                                   "rule_num",
                                   "datapoint")],
                              r_sex[,c("study",
                                       "patientid",
                                       "studyvisit",
                                       "study_wks",
                                       "phenotype",
                                       "string_value",
                                       "rank",
                                       "rule_num",
                                       "datapoint")],
                              v[,c("study",
                                   "patientid",
                                   "studyvisit",
                                   "study_wks",
                                   "phenotype",
                                   "string_value",
                                   "rank",
                                   "rule_num",
                                   "datapoint")])

merged_phenotypedata <- merge(merged_phenotypedata,
                              hdcp_names[,c("phenotype","brief_name")])


analysis_phenotypes <- sqldf("select distinct 
                    study,
                    patientid,
                    studyvisit,
                    study_wks,
                    phenotype,
                    brief_name,
                    string_value,
                    rank
                    from merged_phenotypedata
                    where brief_name is not null
                    and study_wks=0")

b <- sqldf("select study,patientid,study_wks,brief_name,count(rank) as num from analysis_phenotypes
      group by study, patientid, study_wks, brief_name having num >= 2
      order by num desc")

hdcp_data_read <- dcast(analysis_phenotypes[,c("study",
                                               "patientid",
                                               "study_wks",
                                               "brief_name",
                                               "string_value")],
                        study+patientid+study_wks~brief_name,
                        value.var = "string_value")

hdcp_data <- dcast(analysis_phenotypes[,c("study",
                                          "patientid",
                                          "study_wks",
                                          "brief_name",
                                          "rank")],
                   study+patientid+study_wks~brief_name,
                   value.var = "rank")


post_harmonized_phenotypes <- unique(hdcp_trial_criteria$phenotype[hdcp_trial_criteria$form=="r_derive"])

default_vals <- hdcp_names[!is.na(hdcp_names$default_rank)&
                             !(hdcp_names$phenotype %in% post_harmonized_phenotypes)&
                                 hdcp_names$brief_name %in% names(hdcp_data_read),
                           c("brief_name","default_rank","default_text")]

  
for (i in 1:nrow(default_vals)) {
  this_briefname <- default_vals[i,"brief_name"]
  this_default_text <- default_vals[i,"default_text"]
  for (j in 1:nrow(hdcp_data_read)) {
  if (is.na(hdcp_data_read[j,this_briefname])) {
    hdcp_data_read[j,this_briefname] <- this_default_text
     }
  }
}

for (i in 1:nrow(default_vals)) {
  this_briefname <- default_vals[i,"brief_name"]
  this_default_rank <- default_vals[i,"default_rank"]
  for (j in 1:nrow(hdcp_data)) {
    if (is.na(hdcp_data[j,this_briefname])) {
      hdcp_data[j,this_briefname] <- this_default_rank
    }
  }
}

cont_vars <- hdcp_names$brief_name[hdcp_names$var_type=="continuous"&hdcp_names$brief_name %in% names(hdcp_data_read)]

class(hdcp_data_read[cont_vars]) <- "Numeric"

#### Set unused fields to NULL ####
#### Set all fields that do not have a specified code in 'Criteriaset' to NULL (study + field name) 

studies <- unique(hdcp_data_read$study)
hdcp_trial_criteria <- merge(hdcp_trial_criteria,hdcp_names,by="phenotype",all.x=T)
hdcp_trial_criteria <- subset(hdcp_trial_criteria,!is.na(brief_name))

all_fields <- merge(studies,hdcp_names$brief_name,all.x=T,all.y=F)
names(all_fields)[1:2] <- c('study','brief_name')
all_fields$study <- as.character(all_fields$study)
all_fields$brief_name <- as.character(all_fields$brief_name)
all_fields$combined <- paste(all_fields$study,all_fields$brief_name,sep="_")
study_field_pairs <- sqldf('select distinct study,brief_name from hdcp_trial_criteria')
study_field_pairs$combined <- paste(study_field_pairs$study,study_field_pairs$brief_name,sep="_")
noncoded_fields <- subset(all_fields, !combined %in% study_field_pairs$combined)

for (j in 1:nrow(noncoded_fields)) {
  thisstud <- noncoded_fields[j,'study']
  thisfield <- noncoded_fields[j,'brief_name']
  hdcp_data_read[hdcp_data_read$study==thisstud,thisfield] <- NA
  hdcp_data[hdcp_data$study==thisstud,thisfield] <- NA
}

#### Export datasets ####

now <- Sys.time()
today_dt <- paste(Month(now),
                  Day(now),
                  Year(now),
                  sep=".")

write.csv(alldata,
          paste('~/Dropbox/ADAPT-HF/Master HDCP files/hdcp long dataset - ',today_dt,'.csv',sep=''),
          row.names=F)


write.csv(hdcp_data_read, 
          '~/Dropbox/ADAPT-HF/Master HDCP files/hdcp phenotype data text.csv',
          row.names=F,
          na='')

write.csv(hdcp_data,
          '~/Dropbox/ADAPT-HF/Master HDCP files/hdcp phenotype data numeric.csv',
          row.names=F,
          na='')

write.csv(merged_phenotypedata,
          '~/Dropbox/ADAPT-HF/Master HDCP files/longitudinal phenotype data.csv',
          row.names=F,
          na='')


#### Write to BigQuery ####

library(bigrquery)
# library(bigQueryR)

bq_auth(email="")

my_bq <- dbConnect(
  bigrquery::bigquery(),
  project="harmonization",
  dataset="trials",
  billing="harmonization"
)


if (bq_table_exists("harmonization-286013.pipeline.trial_criteria")) 
{bq_table_delete("harmonization-286013.pipeline.trial_criteria")}
bq_table_upload(x="harmonization-286013.pipeline.trial_criteria", 
                hdcp_trial_criteria,quiet=T,
                create_disposition="CREATE_IF_NEEDED",
                write_disposition="WRITE_TRUNCATE")

if (bq_table_exists("harmonization-286013.pipeline.phenotypes_trial")) 
{bq_table_delete("harmonization-286013.pipeline.phenotypes_trial")}
bq_table_upload(x="harmonization-286013.pipeline.phenotypes_trial", 
                hdcp_names,quiet=T,
                create_disposition="CREATE_IF_NEEDED",
                write_disposition="WRITE_TRUNCATE")

if (bq_table_exists("harmonization-286013.trials.alldata")) 
{bq_table_delete("harmonization-286013.trials.alldata")}
bq_table_upload(x="harmonization-286013.trials.alldata", 
                alldata,quiet=T,
                create_disposition="CREATE_IF_NEEDED",
                write_disposition="WRITE_TRUNCATE")


## Load large tables into Google Cloud Storage first

gcs_auth("~/Dropbox/ADAPT-HF/Master HDCP files/harmonization-286013-39f492122f69.json")
gcs_upload(alldata, 
           bucket="master_hdcp_files",
           name="alldata_trial.csv")

if (!bq_table_exists("harmonization-286013.trials.alldata_trial"))
{bq_table_create(x="harmonization-286013.trials.alldata_trial",
                fields=alldata)}

gcs_upload(merged_phenotypedata, 
           bucket="master_hdcp_files",
           name="merged_phenotypedata_trial.csv")

if (!bq_table_exists(
  "harmonization-286013.trials.harmonized_data_trial")) {
  bq_table_create(
    x="harmonization-286013.trials.harmonized_data_trial",
    fields=merged_phenotypedata)}

bq_perform_load(
  x="harmonization-286013.trials.alldata_trial2",
alldata,
source_format = 'CSV',
 fields = alldata,
 create_disposition="CREATE_IF_NEEDED",
 write_disposition = "WRITE_TRUNCATE"
)


gcs_upload(hdcp_data_read, 
           bucket="master_hdcp_files",
           name="hdcp_data_read_trial.csv")

if (!bq_table_exists(
  "harmonization-286013.trials.baseline_data_trial")) {
  bq_table_create(
    x="harmonization-286013.trials.baseline_data_trial",
    fields=hdcp_data_read)}


bqbq <- "ALTER TABLE `trials.alldata_trial` RENAME COLUMN study to study2"

bq_perform_query("harmonization-286013",bqbq)

gcs_upload(hdcp_data, 
           bucket="master_hdcp_files",
           name="hdcp_data_trial.csv")

if (!bq_table_exists(
  "harmonization-286013.trials.baseline_data_trial_num")) {
  bq_table_create(
    x="harmonization-286013.trials.baseline_data_trial_num",
    fields=hdcp_data)}
```
