## Supplementary material for "Aggregating data to accelerate personalized therapy in heart failure (ADAPT-HF)": Harmonization scripts: Transform trial long-term outcomes v3.0.R

```
library(plyr)
library(readstata13)
library(Hmisc)
library(reshape2)
library(descr)
library(rmeta)
library(sas7bdat)
library(sqldf)
library(DescTools)
library(dplyr)
library(nephro)
library(haven)
library(bigrquery)
library(googleCloudStorageR)

source("~/Dropbox/R scripts/poLCA iterate2.R")
# source("~/Dropbox/R scripts/echo scripts.R")
source("~/Dropbox/R scripts/Misc clinical scripts.R")

#### ^^^^ Study folders ^^^^ ####

topcat_folder <- "~/Dropbox/BioLINCC files/TOPCAT/data/sas_data/"
fight_folder <- "~/Dropbox/BioLINCC files/FIGHT/data/csv/"
hfaction_folder <- "~/Dropbox/BioLINCC files/HF-ACTION/Data_DR/"
scdheft_folder <- "~/Dropbox/BioLINCC files/SCD-HeFT/data/"
best_folder <- "~/Dropbox/BioLINCC files/BEST/BEST CSVs/"
paradigm_folder <- "~/Dropbox/BioLINCC files/PARADIGM/"
ipreserve_folder <- "~/Dropbox/BioLINCC files/IPRESERVE/"
relax_folder <- "~/Dropbox/BioLINCC files/RELAX/hfn_relax/data/"
solvd_folder <- "~/Dropbox/BioLINCC files/SOLVD/data/Datasets/"
solvdreg_folder <- "~/Dropbox/BioLINCC files/SOLVD/data/Registry/Datasets/"
dig_folder <- "~/Dropbox/BioLINCC files/DIG/DIG_2015a/ASCII data/"
neat_folder <- "~/Dropbox/BioLINCC files/NEAT-HFpEF/datasets/csv/"
mocha_folder <- "~/Dropbox/BioLINCC files/MOCHA/CSVs/"
corona_folder <- "~/Dropbox/BioLINCC files/CORONA/"
carress_folder <- "~/Dropbox/BioLINCC files/CARRESS/CARRESS_2017a/main_study/data/csv/"
dose_folder <- "~/Dropbox/BioLINCC files/DOSE/data/"
escape_folder <- "~/Dropbox/BioLINCC files/ESCAPE/data/" 
stich_folder <- "~/Dropbox/BioLINCC files/STICH/STICH_2018b/STICH (main study)/data/csv/"
stiches_folder <- "~/Dropbox/BioLINCC files/STICH/STICH_2018b/STICHES (ancillary)/data/csv/"
exact_folder <- "~/Dropbox/BioLINCC files/EXACT-HF/EXACT_2017a/data/csv/"
rose_folder <- "~/Dropbox/BioLINCC files/ROSE/ROSE_2016a/Data/"
guideit_folder <- "~/Dropbox/BioLINCC files/GUIDE-IT/GUIDE_IT_2020a/data/csv/"
athena_folder <- "~/Dropbox/BioLINCC files/ATHENA/data/csv/"
fight_folder <- "~/Dropbox/BioLINCC files/FIGHT/data/CSV/"
indie_folder <- "~/Dropbox/BioLINCC files/INDIE-HFpEF/data/CSV/"
ironout_folder <- "~/Dropbox/BioLINCC files/IRONOUT/data/CSV/"
life_folder <- "~/Dropbox/BioLINCC files/LIFE/data/CSV/"


### outcome columns specified here for convenience.

analysis_outcomes <- c('patientid','study',
                       'cvdth_hfhosp_status','cvdth_hfhosp_dt',
                       'dth_hosp_status', 'dth_hosp_dt',
                       'dth_cvhosp_status','dth_cvhosp_dt',
                       'dth_status','dth_dt',
                       'cvdth_status','cvdth_dt',
                       'noncvdth_status','noncvdth_dt',
                       'hosp_status','hosp_dt',
                       'cvhosp_status','cvhosp_dt',
                       'hfhosp_status','hfhosp_dt',
                       'noncvhosp_status','noncvhosp_dt')

acute_analysis_outcomes <- c('patientid',
                             'study',
                             'wt_kg_bl','wt_kg_1d', 'wt_kg_2d','wt_kg_3d','wt_kg_4d','wt_kg_last',
                            'gfr_bl','gfr_1d','gfr_2d','gfr_3d','gfr_4d','gfr_last',
                            'io_balance_1d','io_balance_2d','io_balance_3d','io_balance_4d','io_balance_5d','io_balance_last',
                            'los',
                            'inhosp_dth_status')
                            
                            
# #######################################################################################################################################
# #######################################################################################################################################
# #######################################################################################################################################

##### ******************************************** OUTCOMES ******************************************** ####
#
# ..%%%%%%%..%%.....%%.%%%%%%%%..%%%%%%...%%%%%%%..%%.....%%.%%%%%%%%..%%%%%%.
# .%%.....%%.%%.....%%....%%....%%....%%.%%.....%%.%%%...%%%.%%.......%%....%%
# .%%.....%%.%%.....%%....%%....%%.......%%.....%%.%%%%.%%%%.%%.......%%......
# .%%.....%%.%%.....%%....%%....%%.......%%.....%%.%%.%%%.%%.%%%%%%....%%%%%%.
# .%%.....%%.%%.....%%....%%....%%.......%%.....%%.%%.....%%.%%.............%%
# .%%.....%%.%%.....%%....%%....%%....%%.%%.....%%.%%.....%%.%%.......%%....%%
# ..%%%%%%%...%%%%%%%.....%%.....%%%%%%...%%%%%%%..%%.....%%.%%%%%%%%..%%%%%%.
#
# 
# 
# 
# ######################## ********* TOPCAT ********* #############################
# 
# 
# .%%%%%%%%..%%%%%%%..%%%%%%%%...%%%%%%.....%%%....%%%%%%%%
# ....%%....%%.....%%.%%.....%%.%%....%%...%%.%%......%%...
# ....%%....%%.....%%.%%.....%%.%%........%%...%%.....%%...
# ....%%....%%.....%%.%%%%%%%%..%%.......%%.....%%....%%...
# ....%%....%%.....%%.%%........%%.......%%%%%%%%%....%%...
# ....%%....%%.....%%.%%........%%....%%.%%.....%%....%%...
# ....%%.....%%%%%%%..%%.........%%%%%%..%%.....%%....%%...
# 

topcat_death <- read.sas7bdat(paste(topcat_folder,'t079.sas7bdat',sep=""))
topcat_hosp <- read.sas7bdat(paste(topcat_folder,'t027.sas7bdat',sep=""))
topcat_af_adj <- read.sas7bdat(paste(topcat_folder,'t075.sas7bdat',sep=""))
topcat_outcomes <- read.sas7bdat('~/Dropbox/BioLINCC files/TOPCAT/data/Outcomes/outcomes.sas7bdat')

#### Mortality outcomes ####

# All-cause mortality, CV mortality, all-cause hospitalization, HF hospitalization,
# Aborted cardiac arrest, MI, Stroke, composite TOPCAT primary endpoint are now given in
# a single file with TOPCAT dataset.
# For completeness wrt other datasets, we will derive non-CV mortality, 
# CV hospitalization and non-CV hospitalization as well as corresponding composite outcomes.

#### Mortality outcomes ####

## All-cause mortality

topcat_outcomes$dth_status <- topcat_outcomes$death
topcat_outcomes$dth_dt <- topcat_outcomes$time_death


## CV mortality

topcat_outcomes$cvdth_status <- topcat_outcomes$cvd_death
topcat_outcomes$cvdth_dt <- topcat_outcomes$time_death

## Non-CV mortality

topcat_outcomes$noncvdth_status <- 0
topcat_outcomes$noncvdth_status[topcat_outcomes$death==1&topcat_outcomes$cvd_death==0] <- 1
topcat_outcomes$noncvdth_dt <- topcat_outcomes$dth_dt


#### Hospitalization outcomes ####

## All-cause hospitalization

topcat_outcomes$hosp_status <- topcat_outcomes$anyhosp
topcat_outcomes$hosp_dt <- topcat_outcomes$time_anyhosp


## CV hospitalization

topcat_hosp$cvhosp_dt[topcat_hosp$HOSP_REAS==1] <- 
  topcat_hosp$hospital_dt3[topcat_hosp$HOSP_REAS==1]

topcat_outcomes <- merge(topcat_outcomes,sqldf('select ID,
                                                 cvhosp_dt 
                                                 from topcat_hosp 
                                                 group by ID 
                                                 having min(cvhosp_dt)'), by='ID',all.x=T)

topcat_outcomes$cvhosp_status[!is.na(topcat_outcomes$cvhosp_dt)] <- 1
topcat_outcomes$cvhosp_status[is.na(topcat_outcomes$cvhosp_dt)] <- 0
topcat_outcomes$cvhosp_dt[is.na(topcat_outcomes$cvhosp_dt)] <- 
  topcat_outcomes$dth_dt[topcat_outcomes$cvhosp_status==0]


## HF hospitalization

topcat_outcomes$hfhosp_status <- topcat_outcomes$hfhosp
topcat_outcomes$hfhosp_dt <- topcat_outcomes$time_hfhosp


## Non-CV hospitalization

topcat_hosp$noncvhosp_dt[topcat_hosp$HOSP_REAS==2] <- 
  topcat_hosp$hospital_dt3[topcat_hosp$HOSP_REAS==2]

topcat_outcomes <- merge(topcat_outcomes,sqldf('select ID,
                                                 noncvhosp_dt 
                                                 from topcat_hosp 
                                                 group by ID 
                                                 having min(noncvhosp_dt)'), by='ID',all.x=T)

topcat_outcomes$noncvhosp_status[!is.na(topcat_outcomes$noncvhosp_dt)] <- 1
topcat_outcomes$noncvhosp_status[is.na(topcat_outcomes$noncvhosp_dt)] <- 0
topcat_outcomes$noncvhosp_dt[is.na(topcat_outcomes$noncvhosp_dt)] <- 
  topcat_outcomes$dth_dt[topcat_outcomes$noncvhosp_status==0]


#### Composite endpoints ####

## Composite TOPCAT endpoint - CV death + HF hospitalization

topcat_outcomes$cvdth_hfhosp_status <- topcat_outcomes$primary_ep
topcat_outcomes$cvdth_hfhosp_dt <- topcat_outcomes$time_primary_ep


## Composite IPRESERVE endpoint - all-cause death + CV hospitalization

topcat_outcomes$dth_cvhosp_status <- 0
topcat_outcomes$dth_cvhosp_status[topcat_outcomes$dth_status==1|topcat_outcomes$cvhosp_status==1] <- 1
topcat_outcomes$dth_cvhosp_dt <- pmin(topcat_outcomes$dth_dt,topcat_outcomes$cvhosp_dt)

## Composite HF-ACTION - all-cause death + all-cause hospitalization

topcat_outcomes$dth_hosp_status <- 0
topcat_outcomes$dth_hosp_status[topcat_outcomes$dth_status==1|topcat_outcomes$hosp_status==1] <- 1
topcat_outcomes$dth_hosp_dt <- pmin(topcat_outcomes$dth_dt,topcat_outcomes$hosp_dt)

names(topcat_outcomes)[names(topcat_outcomes)=="ID"] <- "patientid"

topcat_outcomes[topcat_outcomes < 0] <- NA
topcat_outcomes[topcat_outcomes=="NaN"] <- NA

## convert years to days

topcat_outcomes$cvdth_hfhosp_dt <- round(topcat_outcomes$cvdth_hfhosp_dt*365)
topcat_outcomes$dth_cvhosp_dt <- round(topcat_outcomes$dth_cvhosp_dt*365)
topcat_outcomes$dth_hosp_dt <- round(topcat_outcomes$dth_hosp_dt*365)
topcat_outcomes$dth_dt <- round(topcat_outcomes$dth_dt*365)
topcat_outcomes$cvdth_dt <- round(topcat_outcomes$cvdth_dt*365)
topcat_outcomes$noncvdth_dt <- round(topcat_outcomes$noncvdth_dt*365)
topcat_outcomes$hosp_dt <- round(topcat_outcomes$hosp_dt*365)
topcat_outcomes$cvhosp_dt <- round(topcat_outcomes$cvhosp_dt*365)
topcat_outcomes$hfhosp_dt <- round(topcat_outcomes$hfhosp_dt*365)
topcat_outcomes$noncvhosp_dt <- round(topcat_outcomes$noncvhosp_dt*365)


#### Clean-up ####

topcat_outcomes$study <- "TOPCAT"
missing_outcomes_topcat <- analysis_outcomes[!analysis_outcomes %in% names(topcat_outcomes)]
topcat_outcomes[,missing_outcomes_topcat] <- NA
topcat_outcomes <- topcat_outcomes[,analysis_outcomes]


# 
# ########################### *******  HF-ACTION ********  ############################

# .%%.....%%.%%%%%%%%............%%%.....%%%%%%..%%%%%%%%.%%%%..%%%%%%%..%%....%%
# .%%.....%%.%%.................%%.%%...%%....%%....%%.....%%..%%.....%%.%%%...%%
# .%%.....%%.%%................%%...%%..%%..........%%.....%%..%%.....%%.%%%%..%%
# .%%%%%%%%%.%%%%%%...%%%%%%%.%%.....%%.%%..........%%.....%%..%%.....%%.%%.%%.%%
# .%%.....%%.%%...............%%%%%%%%%.%%..........%%.....%%..%%.....%%.%%..%%%%
# .%%.....%%.%%...............%%.....%%.%%....%%....%%.....%%..%%.....%%.%%...%%%
# .%%.....%%.%%...............%%.....%%..%%%%%%.....%%....%%%%..%%%%%%%..%%....%%

hfaction_analysis <- read.sas7bdat(paste(hfaction_folder,"analysis.sas7bdat",sep=""))
hfaction_cecdeath <- read.sas7bdat(paste(hfaction_folder,"cecdeath.sas7bdat",sep=""))
hfaction_death <- read.sas7bdat(paste(hfaction_folder,"death.sas7bdat",sep=""))
hfaction_cechosp <- read.sas7bdat(paste(hfaction_folder,"cechosp.sas7bdat",sep=""))
hfaction_hospital <- read.sas7bdat(paste(hfaction_folder,"hospital.sas7bdat",sep=""))
hfaction_studcomp <- read.sas7bdat(paste(hfaction_folder,"studcomp.sas7bdat",sep=""))
hfaction_postfvc <- read.sas7bdat(paste(hfaction_folder,"postfvc.sas7bdat",sep=""))

hfaction_analysis$newid <- as.character(hfaction_analysis$newid)
hfaction_cecdeath$newid <- as.character(hfaction_cecdeath$newid)
hfaction_death$newid <- as.character(hfaction_death$newid)
hfaction_cechosp$newid <- as.character(hfaction_cechosp$newid)
hfaction_hospital$newid <- as.character(hfaction_hospital$newid)

hfaction_outcomes <- merge(hfaction_analysis[,c('newid',
                                                'death',
                                                'deathfu',
                                                'dthhosp',
                                                'dhfu')],
                           hfaction_cecdeath[,c('newid',
                                                'CECDTHCA',
                                                'CECCVDTH',
                                                'dthdays')],
                           by="newid",
                           all.x=T)

#### Mortality outcomes ####

## All-cause mortality

colnames(hfaction_outcomes)[2:5] <- c('dth_status',
                                      'dth_dt',
                                      'dth_hosp_status',
                                      'dth_hosp_dt')


## CV mortality

hfaction_outcomes$cvdth_status[hfaction_outcomes$dth_status==1&hfaction_outcomes$CECDTHCA==1] <- 1
hfaction_outcomes$cvdth_status[is.na(hfaction_outcomes$cvdth_status)] <- 0
hfaction_outcomes$cvdth_dt <- hfaction_outcomes$dth_dt


## Non-CV mortality

hfaction_outcomes$noncvdth_status[hfaction_outcomes$dth_status==1&hfaction_outcomes$CECDTHCA==2] <- 1
hfaction_outcomes$noncvdth_status[is.na(hfaction_outcomes$noncvdth_status)] <- 0
hfaction_outcomes$noncvdth_dt <- hfaction_outcomes$dth_dt


#### Hospitalization outcomes ####

hfaction_outcomes <- merge(hfaction_outcomes,hfaction_postfvc[,c('newid',
                                                                 'BEENHOSP',
                                                                 'NMBRHOSP',
                                                                 'CURALIVE',
                                                                 'CVDEATH',
                                                                 'CAUDEATH',
                                                                 'followdays',
                                                                 'lstalvdays',
                                                                 'daysdeath')],by="newid",all.x=T)


## All-cause hospitalization

hfaction_outcomes <- merge(hfaction_outcomes,sqldf('select newid,
                                                   min(hspdays) as hosp_dt 
                                                   from hfaction_cechosp 
                                                   group by newid'),by='newid',all.x=T)

hfaction_outcomes$hosp_status[!is.na(hfaction_outcomes$hosp_dt)] <- 1
hfaction_outcomes$hosp_status[is.na(hfaction_outcomes$hosp_dt)] <- 0
hfaction_outcomes$hosp_dt[hfaction_outcomes$hosp_status==0] <- hfaction_outcomes$dth_dt[hfaction_outcomes$hosp_status==0]


## CV hospitalization

hfaction_outcomes <- merge(hfaction_outcomes,sqldf('select newid,
                                                   min(hspdays) as cvhosp_dt 
                                                   from hfaction_cechosp 
                                                   where CECHSPCA in (1,2,3,4,5,6,7) 
                                                   group by newid'),by='newid',all.x=T)

hfaction_outcomes$cvhosp_status[!is.na(hfaction_outcomes$cvhosp_dt)] <- 1
hfaction_outcomes$cvhosp_status[is.na(hfaction_outcomes$cvhosp_dt)] <- 0
hfaction_outcomes$cvhosp_dt[hfaction_outcomes$cvhosp_status==0] <- hfaction_outcomes$dth_dt[hfaction_outcomes$cvhosp_status==0]


## HF hospitalization

hfaction_outcomes <- merge(hfaction_outcomes,sqldf('select newid,
                                                   min(hspdays) as hfhosp_dt 
                                                   from hfaction_cechosp 
                                                   where CECHSPCA=1  
                                                   group by newid'),by='newid',all.x=T)

hfaction_outcomes$hfhosp_status[!is.na(hfaction_outcomes$hfhosp_dt)] <- 1
hfaction_outcomes$hfhosp_status[is.na(hfaction_outcomes$hfhosp_dt)] <- 0
hfaction_outcomes$hfhosp_dt[hfaction_outcomes$hfhosp_status==0] <- hfaction_outcomes$dth_dt[hfaction_outcomes$hfhosp_status==0]


## Non-CV hospitalization

hfaction_outcomes <- merge(hfaction_outcomes,sqldf('select newid,
                                                   min(hspdays) as noncvhosp_dt 
                                                   from hfaction_cechosp 
                                                   where CECHSPCA=8  
                                                   group by newid'),by='newid',all.x=T)

hfaction_outcomes$noncvhosp_status[!is.na(hfaction_outcomes$noncvhosp_dt)] <- 1
hfaction_outcomes$noncvhosp_status[is.na(hfaction_outcomes$noncvhosp_dt)] <- 0
hfaction_outcomes$noncvhosp_dt[hfaction_outcomes$noncvhosp_status==0] <- hfaction_outcomes$dth_dt[hfaction_outcomes$noncvhosp_status==0]


#### Composite outcomes ####


## Composite TOPCAT endpoint - CV death + HF hospitalization

hfaction_outcomes$cvdth_hfhosp_status <- 0
hfaction_outcomes$cvdth_hfhosp_status[hfaction_outcomes$cvdth_status==1|hfaction_outcomes$hfhosp_status==1] <- 1
hfaction_outcomes$cvdth_hfhosp_dt <- pmin(hfaction_outcomes$cvdth_dt,hfaction_outcomes$hfhosp_dt)

## Composite IPRESERVE endpoint - all-cause death + HF hospitalization

hfaction_outcomes$dth_cvhosp_status <- 0
hfaction_outcomes$dth_cvhosp_status[hfaction_outcomes$dth_status==1|hfaction_outcomes$cvhosp_status==1] <- 1
hfaction_outcomes$dth_cvhosp_dt <- pmin(hfaction_outcomes$dth_dt,hfaction_outcomes$cvhosp_dt)

## Composite HF-ACTION - all-cause death + all-cause hospitalization

hfaction_outcomes$dth_hosp_status <- 0
hfaction_outcomes$dth_hosp_status[hfaction_outcomes$dth_status==1|hfaction_outcomes$hosp_status==1] <- 1
hfaction_outcomes$dth_hosp_dt <- pmin(hfaction_outcomes$dth_dt,hfaction_outcomes$hosp_dt)


#### Clean-up ####

hfaction_outcomes$study <- "HF-ACTION"
colnames(hfaction_outcomes)[colnames(hfaction_outcomes)=="newid"] <- "patientid"
missing_outcomes_hfaction <- analysis_outcomes[!analysis_outcomes %in% names(hfaction_outcomes)]
hfaction_outcomes[,missing_outcomes_hfaction] <- NA
hfaction_outcomes <- hfaction_outcomes[,analysis_outcomes]


# 
# ################## ******* SCD-HeFT ******* ####################
 
# ..%%%%%%...%%%%%%..%%%%%%%%..........%%.....%%.%%%%%%%%.%%%%%%%%.%%%%%%%%
# .%%....%%.%%....%%.%%.....%%.........%%.....%%.%%.......%%..........%%...
# .%%.......%%.......%%.....%%.........%%.....%%.%%.......%%..........%%...
# ..%%%%%%..%%.......%%.....%%.%%%%%%%.%%%%%%%%%.%%%%%%...%%%%%%......%%...
# .......%%.%%.......%%.....%%.........%%.....%%.%%.......%%..........%%...
# .%%....%%.%%....%%.%%.....%%.........%%.....%%.%%.......%%..........%%...
# ..%%%%%%...%%%%%%..%%%%%%%%..........%%.....%%.%%%%%%%%.%%..........%%...

scdheft_endpt <- read.sas7bdat(paste(scdheft_folder,"endpt_new.sas7bdat",sep=""))
scdheft_death <- read.sas7bdat(paste(scdheft_folder,"death.sas7bdat",sep=""))
scdheft_uniquehosp <- read.sas7bdat(paste(scdheft_folder,"uniqhosp.sas7bdat",sep=""))


scdheft_endpt[scdheft_endpt=="NaN"] <- NA
scdheft_death[scdheft_death=="NaN"] <- NA
scdheft_uniquehosp[scdheft_uniquehosp=="NaN"] <- NA

#### Mortality outcomes ####

scdheft_endpt$dth_status <- scdheft_endpt$DEAD
scdheft_endpt$dth_dt <- scdheft_endpt$LASTDT_DAYS


scdheft_endpt$cvdth_status[scdheft_endpt$DEAD2==1] <- 1
scdheft_endpt$cvdth_status[scdheft_endpt$DEAD2==0] <- 0
scdheft_endpt$cvdth_dt <- scdheft_endpt$dth_dt


scdheft_endpt$noncvdth_status <- 0

scdheft_endpt$noncvdth_status[
  scdheft_endpt$cvdth_status==0&
    scdheft_endpt$dth_status==1] <- 1

scdheft_endpt$noncvdth_dt <- scdheft_endpt$dth_dt


scdheft_firsthosp <- sqldf('select PID,
                           FHADMDT_DAYS 
                           from scdheft_uniquehosp 
                           group by PID having min(FHADMDT_DAYS)')

scdheft_outcomes <- merge(scdheft_endpt[,c("PID",
                                           "DEAD",
                                           "DEAD1",
                                           "DEAD2",
                                           "SCD",
                                           "LASTDT_DAYS",
                                           "dth_status",
                                           "dth_dt",
                                           "cvdth_status",
                                           "cvdth_dt",
                                           "noncvdth_status",
                                           "noncvdth_dt")],
                          scdheft_firsthosp[,c("PID",
                                               "FHADMDT_DAYS")],all.x=T)


#### Hospitalization outcomes ####

scdheft_outcomes$hosp_status[as.character(scdheft_outcomes$PID) %in% as.character(unique(scdheft_uniquehosp$PID))] <- 1
scdheft_outcomes$hosp_status[!as.character(scdheft_outcomes$PID) %in% as.character(unique(scdheft_uniquehosp$PID))] <- 0
scdheft_outcomes$hosp_dt[scdheft_outcomes$hosp_status==1] <- scdheft_outcomes$FHADMDT_DAYS[scdheft_outcomes$hosp_status==1]
scdheft_outcomes$hosp_dt[scdheft_outcomes$hosp_status==0] <- scdheft_outcomes$LASTDT_DAYS[scdheft_outcomes$hosp_status==0]


#### Composite outcomes ####

## Composite HF-ACTION - all-cause death + all-cause hospitalization

scdheft_outcomes$dth_hosp_status <- 0
scdheft_outcomes$dth_hosp_status[scdheft_outcomes$dth_status==1|scdheft_outcomes$hosp_status==1] <- 1
scdheft_outcomes$dth_hosp_dt <- pmin(scdheft_outcomes$dth_dt,scdheft_outcomes$hosp_dt)


#### Clean-up ####

names(scdheft_outcomes)[1] <- "patientid"
missing_outcomes_scdheft <- analysis_outcomes[!analysis_outcomes %in% names(scdheft_outcomes)]
scdheft_outcomes[,missing_outcomes_scdheft] <- NA
scdheft_outcomes <- scdheft_outcomes[,analysis_outcomes]
scdheft_outcomes$study <- "SCD-HeFT"


# 
# ################ ******* BEST *******  #####################
# 
# .%%%%%%%%..%%%%%%%%..%%%%%%..%%%%%%%%
# .%%.....%%.%%.......%%....%%....%%...
# .%%.....%%.%%.......%%..........%%...
# .%%%%%%%%..%%%%%%....%%%%%%.....%%...
# .%%.....%%.%%.............%%....%%...
# .%%.....%%.%%.......%%....%%....%%...
# .%%%%%%%%..%%%%%%%%..%%%%%%.....%%...
# 


best_br <- read.csv(paste(best_folder,"br.csv",sep=""),colClasses = "character")
best_wd <- read.csv(paste(best_folder,"wd.csv",sep=""),colClasses = "character")
best_eos <- read.csv(paste(best_folder,"eos.csv",sep=""),colClasses = "character")
best_adju <- read.csv(paste(best_folder,"adju.csv",sep=""),colClasses = "character")
best_mort1 <- read.csv(paste(best_folder,"mort1.csv",sep=""),colClasses = "character")
best_hv <- read.csv(paste(best_folder,"hv.csv",sep=""),colClasses = "character")
best_ame <- read.csv(paste(best_folder,"ame.csv",sep=""),colClasses = "character")


best_outcomes <- best_br[,c('ID','RANDATE')]
best_outcomes <- merge(best_outcomes,best_adju[,c('ID','ADDATE','ADCAUSE')],by="ID",all.x=T)
best_outcomes <- merge(best_outcomes,best_eos[,c('ID','EOSTATUS','EOS_DT')],by="ID",all.x=T)
best_outcomes <- merge(best_outcomes,best_wd[,c('ID','WDREPORT','WDDATE')],by="ID",all.x=T)

## There were 28 patients who died shortly after the trial was stopped 7/29/1999.  Here we treat these as alive at final observation dated 7.29.1999
## and assume they are on meds (EOSTATUS==1)

best_outcomes$EOS_DT[is.na(best_outcomes$ADDATE)&is.na(best_outcomes$EOS_DT)] <- "1999-07-29 00:00:00"
best_outcomes$EOSTATUS[is.na(best_outcomes$ADDATE)&is.na(best_outcomes$EOS_DT)] <- 1

## There were 8 patients who become inactive.  We will treat these as alive at the time of their withdrawal date.

best_outcomes$EOS_DT[best_outcomes$WDREPORT==1&!is.na(best_outcomes$WDREPORT)] <- best_outcomes$WDDATE[best_outcomes$WDREPORT==1&!is.na(best_outcomes$WDREPORT)]
best_outcomes$EOSTATUS[best_outcomes$WDREPORT==1&!is.na(best_outcomes$WDREPORT)] <-1

#### Mortality outcomes ####

best_outcomes$dth_dt <- as.numeric(as.Date(best_outcomes$ADDATE))-as.numeric(as.Date(best_outcomes$RANDATE))
best_outcomes$dth_status[!is.na(best_outcomes$ADDATE)] <- 1
best_outcomes$dth_status[is.na(best_outcomes$ADDATE)] <- 0
best_outcomes$dth_dt[is.na(best_outcomes$ADDATE)] <- as.numeric(as.Date(best_outcomes$EOS_DT[is.na(best_outcomes$ADDATE)]))-as.numeric(as.Date(best_outcomes$RANDATE[is.na(best_outcomes$ADDATE)]))

## 36 patients had cause of death given as 'no information', so CV dth and no CV dth do not add up to all-cause death

best_outcomes$cvdth_dt <- best_outcomes$dth_dt
best_outcomes$cvdth_status[best_outcomes$ADCAUSE %in% c('1','2','3','4','5','6')] <- 1
best_outcomes$cvdth_status[best_outcomes$ADCAUSE %in% c(7,NA)] <- 0

best_outcomes$noncvdth_dt <- best_outcomes$dth_dt
best_outcomes$noncvdth_status[best_outcomes$ADCAUSE %in% c('7')] <- 1
best_outcomes$noncvdth_status[best_outcomes$ADCAUSE %in% c('1','2','3','4','5','6',NA)] <- 0


#### Hospital outcomes ####

best_br$bl_date <- as.Date(best_br$RANDATE)
best_hosp <- merge(best_hv,best_br[,c('ID','bl_date')],by="ID",all.x=T,all.y=F)
best_hosp$hosp_date <- as.Date(best_hosp$HVDAT)
best_hosp$hosp_dt <- as.numeric(best_hosp$hosp_date-best_hosp$bl_date)

best_hosp_outcomes <- sqldf('select ID, 1 as hosp_status,
                            hosp_dt from best_hosp 
                            where HVTYP=1 and hosp_dt > 0 
                            group by ID 
                            having min(hosp_dt)')

best_hosp_outcomes <- merge(best_hosp_outcomes, 
                            sqldf('select ID, 
                                  1 as hfhosp_status, 
                                  hosp_dt as hfhosp_dt 
                                  from best_hosp 
                                  where HVTYP=1 and HVHFR=1 and hosp_dt > 0 
                                  group by ID having min(hosp_dt)'),
                            all.x=T)

best_outcomes <- merge(best_outcomes,best_hosp_outcomes,by="ID",all.x=T)

best_outcomes$hosp_status[is.na(best_outcomes$hosp_status)] <- 0
best_outcomes$hosp_dt[best_outcomes$hosp_status==0] <- best_outcomes$dth_dt[best_outcomes$hosp_status==0]

best_outcomes$hfhosp_status[is.na(best_outcomes$hfhosp_status)] <- 0
best_outcomes$hfhosp_dt[best_outcomes$hfhosp_status==0] <- best_outcomes$dth_dt[best_outcomes$hfhosp_status==0]


#### Composite outcomes ####


## Composite TOPCAT endpoint - CV death + HF hospitalization

best_outcomes$cvdth_hfhosp_status <- 0
best_outcomes$cvdth_hfhosp_status[best_outcomes$cvdth_status==1|best_outcomes$hfhosp_status==1] <- 1
best_outcomes$cvdth_hfhosp_dt <- pmin(best_outcomes$cvdth_dt,best_outcomes$hfhosp_dt)

## Composite IPRESERVE endpoint - all-cause death + CV hospitalization

# Could not determine as hv form only specifies HF-related hospitalization, not CV-related.

## Composite HF-ACTION endpoint - all-cause death + all-cause hospitalization

best_outcomes$dth_hosp_status <- 0
best_outcomes$dth_hosp_status[best_outcomes$dth_status==1|best_outcomes$hosp_status==1] <- 1
best_outcomes$dth_hosp_dt <- pmin(best_outcomes$dth_dt,best_outcomes$hosp_dt)


#### Clean-up ####

colnames(best_outcomes)[colnames(best_outcomes)=="ID"] <- "patientid"
missing_outcomes_best <- analysis_outcomes[!analysis_outcomes %in% names(best_outcomes)]
best_outcomes[,missing_outcomes_best] <- NA
best_outcomes <- best_outcomes[,analysis_outcomes]
best_outcomes$study <- "BEST"


# ############################# ******* PARADIGM *******  ################################
# 
# .%%%%%%%%.....%%%....%%%%%%%%.....%%%....%%%%%%%%..%%%%..%%%%%%...%%.....%%
# .%%.....%%...%%.%%...%%.....%%...%%.%%...%%.....%%..%%..%%....%%..%%%...%%%
# .%%.....%%..%%...%%..%%.....%%..%%...%%..%%.....%%..%%..%%........%%%%.%%%%
# .%%%%%%%%..%%.....%%.%%%%%%%%..%%.....%%.%%.....%%..%%..%%...%%%%.%%.%%%.%%
# .%%........%%%%%%%%%.%%...%%...%%%%%%%%%.%%.....%%..%%..%%....%%..%%.....%%
# .%%........%%.....%%.%%....%%..%%.....%%.%%.....%%..%%..%%....%%..%%.....%%
# .%%........%%.....%%.%%.....%%.%%.....%%.%%%%%%%%..%%%%..%%%%%%...%%.....%%
# 


paradigm_outcomes <- as.matrix(read.dta13('~/Dropbox/BioLINCC files/PARADIGM/paradigmhf sample with labs and vitals.dta',
                                        convert.factors=F))
paradigm_outcomes <- as.data.frame(paradigm_outcomes)


paradigm_outcomes <- paradigm_outcomes[,c("sid1a",
                                        "c4cnpt2r",
                                        "t2cnpt2r",
                                        "c4dth",
                                        "c4cvdth",
                                        "t2dth",
                                        "t2cvdth",
                                        "c4hsp",
                                        "t2hsp",
                                        "c4hcv",
                                        "t2hcv",
                                        "c4hhf1",
                                        "t2hhf1",
                                        "c4hncv",
                                        "t2hncv",
                                        "c4noaf",
                                        "t2noaf",
                                        "c4nodm",
                                        "t2nodm")]


#### Mortality outcomes ####

## All-cause mortality

paradigm_outcomes$dth_status[paradigm_outcomes$c4dth==0] <- 1
paradigm_outcomes$dth_status[paradigm_outcomes$c4dth==1] <- 0
paradigm_outcomes$dth_dt <- paradigm_outcomes$t2dth


## CV mortality

paradigm_outcomes$cvdth_status[paradigm_outcomes$c4cvdth==0] <- 1
paradigm_outcomes$cvdth_status[paradigm_outcomes$c4cvdth==1] <- 0
paradigm_outcomes$cvdth_dt <- paradigm_outcomes$t2cvdth


## Non-CV mortality
paradigm_outcomes$noncvdth_status <- 0
paradigm_outcomes$noncvdth_status[paradigm_outcomes$dth_status==1&paradigm_outcomes$cvdth_status==0] <- 1
paradigm_outcomes$noncvdth_dt <- paradigm_outcomes$t2dth


#### Hospitalization outcomes

## All-cause hospitalization

paradigm_outcomes$hosp_status[paradigm_outcomes$c4hsp==1] <- 0
paradigm_outcomes$hosp_status[paradigm_outcomes$c4hsp==0] <- 1
paradigm_outcomes$hosp_dt  <- paradigm_outcomes$t2hsp

## CV hospitalization

paradigm_outcomes$cvhosp_status[paradigm_outcomes$c4hcv==1] <- 0
paradigm_outcomes$cvhosp_status[paradigm_outcomes$c4hcv==0] <- 1
paradigm_outcomes$cvhosp_dt  <- paradigm_outcomes$t2hcv

## HF hospitalization

paradigm_outcomes$hfhosp_status[paradigm_outcomes$c4hhf1==1] <- 0
paradigm_outcomes$hfhosp_status[paradigm_outcomes$c4hhf1==0] <- 1
paradigm_outcomes$hfhosp_dt  <- paradigm_outcomes$t2hhf1

## Non-CV hospitalization
paradigm_outcomes$noncvhosp_status[paradigm_outcomes$c4hncv==1] <- 0
paradigm_outcomes$noncvhosp_status[paradigm_outcomes$c4hncv==0] <- 1
paradigm_outcomes$noncvhosp_dt  <- paradigm_outcomes$t2hncv


#### Composite outcomes

## Composite TOPCAT endpoint - CV death + HF hospitalization

paradigm_outcomes$cvdth_hfhosp_status[paradigm_outcomes$c4cnpt2r == 0] <- 1
paradigm_outcomes$cvdth_hfhosp_status[paradigm_outcomes$c4cnpt2r == 1] <- 0
paradigm_outcomes$cvdth_hfhosp_dt <- paradigm_outcomes$t2cnpt2r


## Composite IPRESERVE endpoint - all-cause death + HF hospitalization

paradigm_outcomes$dth_cvhosp_status <- 0
paradigm_outcomes$dth_cvhosp_status[paradigm_outcomes$dth_status==1|paradigm_outcomes$cvhosp_status==1] <- 1
paradigm_outcomes$dth_cvhosp_dt <- pmin(paradigm_outcomes$dth_dt,paradigm_outcomes$cvhosp_dt)


## Composite HF-ACTION - all-cause death + all-cause hospitalization
paradigm_outcomes$dth_hosp_status <- 0
paradigm_outcomes$dth_hosp_status[paradigm_outcomes$dth_status==1|paradigm_outcomes$hosp_status==1] <- 1
paradigm_outcomes$dth_hosp_dt <- pmin(paradigm_outcomes$dth_dt,paradigm_outcomes$hosp_dt)


#### Clean-up ####

paradigm_outcomes$study <- "PARADIGM"
paradigm_outcomes$patientid <- paradigm_outcomes$sid1a
missing_outcomes_paradigm <- analysis_outcomes[!analysis_outcomes %in% names(paradigm_outcomes)]
paradigm_outcomes[,missing_outcomes_paradigm] <- NA
paradigm_outcomes <- paradigm_outcomes[,analysis_outcomes]


# 
# ############################### ******* I-PRESERVE ******* ###############################
# 
# .%%%%.........%%%%%%%%..%%%%%%%%..%%%%%%%%..%%%%%%..%%%%%%%%.%%%%%%%%..%%.....%%.%%%%%%%%
# ..%%..........%%.....%%.%%.....%%.%%.......%%....%%.%%.......%%.....%%.%%.....%%.%%......
# ..%%..........%%.....%%.%%.....%%.%%.......%%.......%%.......%%.....%%.%%.....%%.%%......
# ..%%..%%%%%%%.%%%%%%%%..%%%%%%%%..%%%%%%....%%%%%%..%%%%%%...%%%%%%%%..%%.....%%.%%%%%%..
# ..%%..........%%........%%...%%...%%.............%%.%%.......%%...%%....%%...%%..%%......
# ..%%..........%%........%%....%%..%%.......%%....%%.%%.......%%....%%....%%.%%...%%......
# .%%%%.........%%........%%.....%%.%%%%%%%%..%%%%%%..%%%%%%%%.%%.....%%....%%%....%%%%%%%
#

ip_outcomes <- read.csv(paste(ipreserve_folder,'ip_rawdata.csv',sep=""))[,c('usubjid',
                                                              'prmevnt','prmtime',
                                                              'dthevnt', 'dthtime',
                                                              'cvdeath','dthtime',
                                                              'any_hosp','time_anyhosp',
                                                              'cv_hosp','time_cvhosp',
                                                              'noncv_hosp','time_noncv',
                                                              'hfevnt','hftime',
                                                              'hfhosp','hftime')]

ip_outcomes$dth_status <- ip_outcomes$dthevnt
ip_outcomes$dth_dt <- ip_outcomes$dthtime

ip_outcomes$cvdth_status <- ip_outcomes$cvdeath
ip_outcomes$cvdth_dt <- ip_outcomes$dthtime

ip_outcomes$noncvdth_status <- 0
ip_outcomes$noncvdth_status[ip_outcomes$dth_status==1&
                              ip_outcomes$cvdth_status==0] <- 1
ip_outcomes$noncvdth_dt <- ip_outcomes$dthtime


#### Hospitalization outcomes ####

## All-cause hospitalization

ip_outcomes$hosp_status <- ip_outcomes$any_hosp
ip_outcomes$hosp_dt <- ip_outcomes$time_anyhosp


## CV hospitalization

ip_outcomes$cvhosp_status <- ip_outcomes$cv_hosp
ip_outcomes$cvhosp_dt <- ip_outcomes$time_cvhosp


## HF hospitalization

ip_outcomes$hfevnt_status <- ip_outcomes$hfevnt
ip_outcomes$hfevnt_dt <- ip_outcomes$hftime

ip_outcomes$hfhosp_status <- ip_outcomes$hfhosp
ip_outcomes$hfhosp_dt[ip_outcomes$hfhosp==1] <- ip_outcomes$hftime[ip_outcomes$hfhosp==1]
ip_outcomes$hfhosp_dt[ip_outcomes$hfhosp==0] <- ip_outcomes$dth_dt[ip_outcomes$hfhosp==0]


## Non-CV hospitalization

ip_outcomes$noncvhosp_status <- ip_outcomes$noncv_hosp
ip_outcomes$noncvhosp_dt <- ip_outcomes$time_noncv


#### Composite outcomes ####

## Composite TOPCAT endpoints - CV death + HF hospitalization

ip_outcomes$cvdth_hfhosp_status <- 0
ip_outcomes$cvdth_hfhosp_status[ip_outcomes$cvdth_status==1|ip_outcomes$hfhosp_status==1] <- 1
ip_outcomes$cvdth_hfhosp_dt <- pmin(ip_outcomes$cvdth_dt,ip_outcomes$hfhosp_dt)


## Composite IPRESERVE endpoint - all-cause death + CV hospitalization

ip_outcomes$dth_cvhosp_status <- ip_outcomes$prmevnt
ip_outcomes$dth_cvhosp_dt <- ip_outcomes$prmtime


## Composite HF-ACTION endpoint - all-cause death + hospitalization

ip_outcomes$dth_hosp_status <- 0
ip_outcomes$dth_hosp_status[ip_outcomes$dth_status==1|ip_outcomes$hosp_status==1] <- 1
ip_outcomes$dth_hosp_dt <- pmin(ip_outcomes$dth_dt,ip_outcomes$hosp_dt)


#### Clean-up ####

names(ip_outcomes)[1] <- "patientid"
ip_outcomes$study <- "IPRESERVE"
missing_outcomes_ip <- analysis_outcomes[!analysis_outcomes %in% names(ip_outcomes)]
ip_outcomes[,missing_outcomes_ip] <- NA
ip_outcomes <- ip_outcomes[,analysis_outcomes]


# 
# ################  ******* RELAX *******  ############################
# 
# .%%%%%%%%..%%%%%%%%.%%..........%%%....%%.....%%
# .%%.....%%.%%.......%%.........%%.%%....%%...%%.
# .%%.....%%.%%.......%%........%%...%%....%%.%%..
# .%%%%%%%%..%%%%%%...%%.......%%.....%%....%%%...
# .%%...%%...%%.......%%.......%%%%%%%%%...%%.%%..
# .%%....%%..%%.......%%.......%%.....%%..%%...%%.
# .%%.....%%.%%%%%%%%.%%%%%%%%.%%.....%%.%%.....%%
# 

relax_death  <- read.sas7bdat(paste(relax_folder,"deathpag.sas7bdat",sep=""))
relax_death[relax_death=="NaN"] <- NA
relax_hosp  <- read.sas7bdat(paste(relax_folder,"rehosptl.sas7bdat",sep=""))
relax_hosp[relax_hosp=="NaN"] <- NA
relax_hosp$LOS <- relax_hosp$REDCHGDT-relax_hosp$REHOSPDT
relax_eos <- read.sas7bdat(paste(relax_folder,"rxterm.sas7bdat",sep=""))
relax_eos[relax_eos=="NaN"] <- NA
relax_endpts <- read.sas7bdat(paste(relax_folder,"a_endpts.sas7bdat",sep=""))
relax_endpts[relax_endpts=="NaN"] <- NA
relax_base <- read.sas7bdat(paste(relax_folder,"a_base.sas7bdat",sep=""))
relax_base[relax_base=="NaN"] <- NA

relax_outcomes <- merge(relax_eos,relax_death[,c('patnumb','DEATHCAU')],by.x="patnumb",by.y="patnumb",all.x=T)
relax_outcomes$eos_dt[!is.na(relax_outcomes$RXTERMDT)] <- relax_outcomes$RXTERMDT[!is.na(relax_outcomes$RXTERMDT)]
relax_outcomes$eos_dt[is.na(relax_outcomes$RXTERMDT)] <- relax_outcomes$RXSTPDT[is.na(relax_outcomes$RXTERMDT)]

relax_outcomes$dth_status <- 0
relax_outcomes$dth_status[relax_outcomes$RXTERMRE==4] <- 1
relax_outcomes$dth_dt <- relax_outcomes$eos_dt

relax_outcomes$cvdth_status <- 0
relax_outcomes$cvdth_status[relax_outcomes$RXTERMRE==4&relax_outcomes$DEATHCAU %in% c(1,2,3,4,5,6)] <- 1
relax_outcomes$cvdth_dt <- relax_outcomes$eos_dt

relax_outcomes$noncvdth_status <- 0
relax_outcomes$noncvdth_status[relax_outcomes$RXTERMRE==4&relax_outcomes$DEATHCAU %in% c(7,8)] <- 1
relax_outcomes$noncvdth_dt <- relax_outcomes$eos_dt


#### Hospitalization outcomes ####

## All-cause hospitalization

relax_hosp$hosp_status[!is.na(relax_hosp$REHOSPDT)] <- 1
relax_hosp$hosp_dt <- relax_hosp$REHOSPDT

relax_firsthosp <- sqldf('select patnumb, 
                         hosp_dt, 
                         hosp_status 
                         from relax_hosp 
                         where hosp_status=1 
                         group by patnumb 
                         having min(hosp_dt)')


relax_outcomes <- merge(relax_outcomes,relax_firsthosp,by.x="patnumb",by.y="patnumb",all.x=T)
relax_outcomes$hosp_status[is.na(relax_outcomes$hosp_status)] <- 0
relax_outcomes$hosp_dt[relax_outcomes$hosp_status==0] <- relax_outcomes$eos_dt[relax_outcomes$hosp_status==0]


## CV hospitalization

relax_hosp$cvhosp_status[relax_hosp$PRIMCAUS %in% c(1:29)] <- 1
relax_hosp$cvhosp_dt[relax_hosp$PRIMCAUS %in% c(1:29)] <- relax_hosp$REHOSPDT[relax_hosp$PRIMCAUS %in% c(1:29)]


relax_firstcvhosp <- sqldf('select patnumb, 
                           cvhosp_dt, 
                           cvhosp_status 
                           from relax_hosp 
                           where cvhosp_status=1 
                           group by patnumb 
                           having min(cvhosp_dt)')

relax_outcomes <- merge(relax_outcomes,relax_firstcvhosp,by.x="patnumb",by.y="patnumb",all.x=T)
relax_outcomes$cvhosp_status[is.na(relax_outcomes$cvhosp_status)] <- 0
relax_outcomes$cvhosp_dt[relax_outcomes$cvhosp_status==0] <- relax_outcomes$eos_dt[relax_outcomes$cvhosp_status==0]


## HF hospitalization

relax_hosp$hfhosp_status[relax_hosp$PRIMCAUS==1] <- 1
relax_hosp$hfhosp_dt[relax_hosp$PRIMCAUS %in% c(1)] <- relax_hosp$REHOSPDT[relax_hosp$PRIMCAUS %in% c(1)]


relax_firsthfhosp <- sqldf('select patnumb, 
                           hfhosp_dt, 
                           hfhosp_status 
                           from relax_hosp 
                           where hfhosp_status=1 
                           group by patnumb 
                           having min(hfhosp_dt)')

relax_outcomes <- merge(relax_outcomes,relax_firsthfhosp,by.x="patnumb",by.y="patnumb",all.x=T)
relax_outcomes$hfhosp_status[is.na(relax_outcomes$hfhosp_status)] <- 0
relax_outcomes$hfhosp_dt[relax_outcomes$hfhosp_status==0] <- relax_outcomes$eos_dt[relax_outcomes$hfhosp_status==0]


## Non-CV hospitalization

relax_hosp$noncvhosp_status[relax_hosp$PRIMCAUS %in% c(31:49)] <- 1
relax_hosp$noncvhosp_dt[relax_hosp$PRIMCAUS %in% c(31:49)] <- relax_hosp$REHOSPDT[relax_hosp$PRIMCAUS %in% c(31:49)]


relax_firstnoncvhosp <- sqldf('select patnumb, 
                              noncvhosp_dt, 
                              noncvhosp_status 
                              from relax_hosp 
                              where noncvhosp_status=1 
                              group by patnumb 
                              having min(noncvhosp_dt)')


relax_outcomes <- merge(relax_outcomes,relax_firstnoncvhosp,by.x="patnumb",by.y="patnumb",all.x=T)
relax_outcomes$noncvhosp_status[is.na(relax_outcomes$noncvhosp_status)] <- 0
relax_outcomes$noncvhosp_dt[relax_outcomes$noncvhosp_status==0] <- relax_outcomes$eos_dt[relax_outcomes$noncvhosp_status==0]


#### Composite outcomes ####

## Composite TOPCAT endpoint - CV-cause death + HF hospitalization

relax_outcomes$cvdth_hfhosp_status <- 0
relax_outcomes$cvdth_hfhosp_status[relax_outcomes$cvdth_status==1|relax_outcomes$hfhosp_status==1] <- 1
relax_outcomes$cvdth_hfhosp_dt <- pmin(relax_outcomes$cvdth_dt,relax_outcomes$hfhosp_dt)


## Composite IPRESERVE endpoint - all-cause death + CV hospitalization

relax_outcomes$dth_cvhosp_status <- 0
relax_outcomes$dth_cvhosp_status[relax_outcomes$dth_status==1|relax_outcomes$cvhosp_status==1] <- 1
relax_outcomes$dth_cvhosp_dt <- pmin(relax_outcomes$dth_dt,relax_outcomes$cvhosp_dt)


## Composite HF-ACTION endpoint - all-cause death +  hospitalization

relax_outcomes$dth_hosp_status <- 0
relax_outcomes$dth_hosp_status[relax_outcomes$dth_status==1|relax_outcomes$hosp_status==1] <- 1
relax_outcomes$dth_hosp_dt <- pmin(relax_outcomes$dth_dt,relax_outcomes$hosp_dt)


#### Clean-up ####

relax_outcomes$study <- "RELAX"
relax_outcomes$patientid <- relax_outcomes$patnumb
missing_outcomes_relax <- analysis_outcomes[!analysis_outcomes %in% names(relax_outcomes)]
relax_outcomes[,missing_outcomes_relax] <- NA
relax_outcomes <- relax_outcomes[,analysis_outcomes]  


# ##############  ******* SOLVD *******  ####################      
# 
# ..%%%%%%...%%%%%%%..%%.......%%.....%%.%%%%%%%%.
# .%%....%%.%%.....%%.%%.......%%.....%%.%%.....%%
# .%%.......%%.....%%.%%.......%%.....%%.%%.....%%
# ..%%%%%%..%%.....%%.%%.......%%.....%%.%%.....%%
# .......%%.%%.....%%.%%........%%...%%..%%.....%%
# .%%....%%.%%.....%%.%%.........%%.%%...%%.....%%
# ..%%%%%%...%%%%%%%..%%%%%%%%....%%%....%%%%%%%%.
# 

solvd_outcomes <- read.sas7bdat(paste(solvd_folder,"sep_lad2.sas7bdat",sep=""))
solvd_outcomes[solvd_outcomes=="NaN"] <- NA
solvd_outcomes[solvd_outcomes==""] <- NA
solvd_sbf <- read.sas7bdat(paste(solvd_folder,"sbf_lad2.sas7bdat",sep=""))
solvd_sbf[solvd_sbf==""|solvd_sbf=="NaN"] <- NA


solvd_outcomes$patientid <- solvd_outcomes$ID_SOL


solvd_allhosp <- read.sas7bdat(paste(solvd_folder,"shf_lad2.sas7bdat",sep=""))[,c("ID_SOL",
                                                                                  "TRIAL",
                                                                                  "SHF4Z1",
                                                                                  "SHF4Z2",
                                                                                  "SHF5",
                                                                                  "SHF7",
                                                                                  "SHF7A",
                                                                                  "SHF7B",
                                                                                  "SHF7BH",
                                                                                  "SHF7BM",
                                                                                  "SHF7C",
                                                                                  "SHF9Z1",
                                                                                  "SHF11",
                                                                                  "SHF13",
                                                                                  "SHF14",
                                                                                  "SHF15",
                                                                                  "SHF16Z1",
                                                                                  "SHF16Z2",
                                                                                  "SHF16Z3",
                                                                                  "SHF17",
                                                                                  "SHF18",
                                                                                  "SHF19",
                                                                                  "SHF20",
                                                                                  "SHF21",
                                                                                  "SHF22Z1",
                                                                                  "SHF22Z2")]
solvd_allhosp[solvd_allhosp =="NaN"] <- NA
solvd_allhosp[solvd_allhosp ==""] <- NA

#### Mortality outcomes ####

solvd_outcomes$dth_status <- solvd_outcomes$EP2
solvd_outcomes$dth_dt <- solvd_outcomes$FUTIME
solvd_outcomes$cvdth_status <- solvd_outcomes$EP3
solvd_outcomes$cvdth_dt <- solvd_outcomes$FUTIME
solvd_outcomes$noncvdth_status <- solvd_outcomes$EP13
solvd_outcomes$noncvdth_dt <- solvd_outcomes$FUTIME


#### Hospitalization outcomes ####

## All-cause hospitalization

solvd_firsthosp <- sqldf('select ID_SOL as patientid, 
                                  1 as hosp_status, 
                                  SHF4Z1 as hosp_dt 
                                  from solvd_allhosp 
                                  where hosp_dt > 0 
                                  group by patientid 
                                  having min(hosp_dt)')


solvd_outcomes <- merge(solvd_outcomes,solvd_firsthosp,by="patientid",all.x=T)
solvd_outcomes$hosp_status[is.na(solvd_outcomes$hosp_status)] <- 0
solvd_outcomes$hosp_dt[solvd_outcomes$hosp_status==0] <- solvd_outcomes$dth_dt[solvd_outcomes$hosp_status==0]


## CV hospitalization

solvd_firstcvhosp <- sqldf('select ID_SOL as patientid, 
                                    1 as cvhosp_status, 
                                    SHF4Z1 as cvhosp_dt 
                                    from solvd_allhosp 
                                    where SHF5 = "C" and cvhosp_dt > 0 
                                    group by patientid 
                                    having min(cvhosp_dt)')

solvd_outcomes <- merge(solvd_outcomes,solvd_firstcvhosp,by="patientid",all.x=T)
solvd_outcomes$cvhosp_status[is.na(solvd_outcomes$cvhosp_status)] <- 0
solvd_outcomes$cvhosp_dt[solvd_outcomes$cvhosp_status==0] <- solvd_outcomes$dth_dt[solvd_outcomes$cvhosp_status==0]


## HF hospitalization

solvd_outcomes$hfhosp_status <- solvd_outcomes$EPY
solvd_outcomes$hfhosp_dt <- solvd_outcomes$EPYTIME
solvd_outcomes$patientid <- solvd_outcomes$ID_SOL

## Non-CV hospitalization

solvd_firstnoncvhosp <- sqldf('select ID_sol as patientid, 
                                       1 as noncvhosp_status, 
                                       SHF4Z1 as noncvhosp_dt 
                                       from solvd_allhosp 
                                       where SHF5 = "N" and noncvhosp_dt > 0 
                                       group by patientid 
                                       having min(noncvhosp_dt)')

solvd_outcomes <- merge(solvd_outcomes,solvd_firstnoncvhosp,by="patientid",all.x=T)
solvd_outcomes$noncvhosp_status[is.na(solvd_outcomes$noncvhosp_status)] <- 0
solvd_outcomes$noncvhosp_dt[solvd_outcomes$noncvhosp_status==0] <- solvd_outcomes$dth_dt[solvd_outcomes$noncvhosp_status==0]


#### Composite outcomes ####

## Composite TOPCAT endpoint - CV-cause death + HF hospitalization

solvd_outcomes$cvdth_hfhosp_status <- 0
solvd_outcomes$cvdth_hfhosp_status[solvd_outcomes$cvdth_status==1|solvd_outcomes$hfhosp_status==1] <- 1
solvd_outcomes$cvdth_hfhosp_dt <- pmin(solvd_outcomes$cvdth_dt,solvd_outcomes$hfhosp_dt)


## Composite IPRESERVE endpoint - all-cause death + CV hospitalization

solvd_outcomes$dth_cvhosp_status <- 0
solvd_outcomes$dth_cvhosp_status[solvd_outcomes$dth_status==1|solvd_outcomes$cvhosp_status==1] <- 1
solvd_outcomes$dth_cvhosp_dt <- pmin(solvd_outcomes$dth_dt,solvd_outcomes$cvhosp_dt)


## Composite HF-ACTION endpoint - all-cause death +  hospitalization

solvd_outcomes$dth_hosp_status <- 0
solvd_outcomes$dth_hosp_status[solvd_outcomes$dth_status==1|solvd_outcomes$hosp_status==1] <- 1
solvd_outcomes$dth_hosp_dt <- pmin(solvd_outcomes$dth_dt,solvd_outcomes$hosp_dt)


#### Clean-up ####

solvd_outcomes <- merge(solvd_outcomes,solvd_sbf[,c("ID_SOL","TRIAL")],by="ID_SOL")
solvd_outcomes$study[solvd_outcomes$TRIAL=="T"] <- "SOLVD Treat"
solvd_outcomes$study[solvd_outcomes$TRIAL=="P"] <- "SOLVD Prevent"

missing_outcomes_solvd <- analysis_outcomes[!analysis_outcomes %in% names(solvd_outcomes)]
solvd_outcomes[,missing_outcomes_solvd] <- NA
solvd_outcomes <- solvd_outcomes[,c(analysis_outcomes)]  


############## ******* SOLVD-Registry ******* ####################      

# ..%%%%%%...%%%%%%%..%%.......%%.....%%.%%%%%%%%.....%%%%%%%%..%%%%%%%%..%%%%%%...%%%%..%%%%%%..%%%%%%%%.%%%%%%%%..%%....%%
# .%%....%%.%%.....%%.%%.......%%.....%%.%%.....%%....%%.....%%.%%.......%%....%%...%%..%%....%%....%%....%%.....%%..%%..%%.
# .%%.......%%.....%%.%%.......%%.....%%.%%.....%%....%%.....%%.%%.......%%.........%%..%%..........%%....%%.....%%...%%%%..
# ..%%%%%%..%%.....%%.%%.......%%.....%%.%%.....%%....%%%%%%%%..%%%%%%...%%...%%%%..%%...%%%%%%.....%%....%%%%%%%%.....%%...
# .......%%.%%.....%%.%%........%%...%%..%%.....%%....%%...%%...%%.......%%....%%...%%........%%....%%....%%...%%......%%...
# .%%....%%.%%.....%%.%%.........%%.%%...%%.....%%....%%....%%..%%.......%%....%%...%%..%%....%%....%%....%%....%%.....%%...
# ..%%%%%%...%%%%%%%..%%%%%%%%....%%%....%%%%%%%%.....%%.....%%.%%%%%%%%..%%%%%%...%%%%..%%%%%%.....%%....%%.....%%....%%...


## Registry hospitalization form

solvd_registry_rhf <- read_sas(paste(solvdreg_folder,"rhf_reg.sas7bdat",sep=""))
solvd_registry_rhf[solvd_registry_rhf=="#"] <- NA
solvd_registry_rhf[solvd_registry_rhf==""] <- NA

## Registration Designation of Death Form

solvd_registry_rdd <- read_sas(paste(solvdreg_folder,"rdd_reg.sas7bdat",sep=""))
solvd_registry_rdd[solvd_registry_rdd=="#"] <- NA
solvd_registry_rdd[solvd_registry_rdd==""] <- NA

## Registry baseline form

solvd_registry_subj <- read_sas(paste(solvdreg_folder,"rbf_reg.sas7bdat",sep=""))[,c('id_reg','ID_SOL','RBF3Z1')]
solvd_registry_subj[solvd_registry_subj=="#"] <- NA
solvd_registry_subj[solvd_registry_subj==""] <- NA

## Registry 

solvd_registry_rfu <- read_sas(paste(solvdreg_folder,"rfu_reg.sas7bdat",sep=""))
solvd_registry_rfu[solvd_registry_rfu=="#"] <- NA
solvd_registry_subj[solvd_registry_subj==""] <- NA


## Registry follow-up form

solvd_registry_rff <- read_sas(paste(solvdreg_folder,"rff_reg.sas7bdat",sep=""))
solvd_registry_rff[solvd_registry_rff=="#"] <- NA
solvd_registry_rff[solvd_registry_rff==""] <- NA
solvd_registry_rff$eos_dt <- solvd_registry_rff$rff3end
solvd_registry_rff$eos_dt[solvd_registry_rff$eos_dt>366] <- 366

solvd_registry_outcomes <- merge(solvd_registry_subj,solvd_registry_rff[,c('id_reg','eos_dt')], by="id_reg")
solvd_registry_outcomes <- merge(solvd_registry_outcomes,solvd_registry_rdd,by='id_reg',all.x=T)


#### Mortality outcomes ####

solvd_registry_outcomes$dth_dt[solvd_registry_outcomes$rdd2days < 365&!is.na(solvd_registry_outcomes$rdd2days)] <- 
  solvd_registry_outcomes$rdd2days[solvd_registry_outcomes$rdd2days < 365&!is.na(solvd_registry_outcomes$rdd2days)]
solvd_registry_outcomes$dth_status[!is.na(solvd_registry_outcomes$dth_dt)] <- 1
solvd_registry_outcomes$dth_status[is.na(solvd_registry_outcomes$dth_status)|solvd_registry_outcomes$dth_dt > 366|solvd_registry_outcomes$dth_dt < 0] <- 0
solvd_registry_outcomes$dth_dt[solvd_registry_outcomes$dth_status==0] <- solvd_registry_outcomes$eos_dt[solvd_registry_outcomes$dth_status==0]

solvd_registry_outcomes$cvdth_status[solvd_registry_outcomes$RDD5=="C"] <- 1
solvd_registry_outcomes$cvdth_dt[solvd_registry_outcomes$RDD5=="C"&!is.na(solvd_registry_outcomes$RDD5)] <- 
  solvd_registry_outcomes$rdd2days[solvd_registry_outcomes$RDD5=="C"&!is.na(solvd_registry_outcomes$RDD5)]
solvd_registry_outcomes$cvdth_status[is.na(solvd_registry_outcomes$cvdth_status)|
                                       solvd_registry_outcomes$dth_dt > 366|
                                       solvd_registry_outcomes$dth_dt > 366|
                                       solvd_registry_outcomes$dth_dt < 0] <- 0
solvd_registry_outcomes$cvdth_dt[solvd_registry_outcomes$cvdth_status==0] <- solvd_registry_outcomes$eos_dt[solvd_registry_outcomes$cvdth_status==0]

solvd_registry_outcomes$noncvdth_status[solvd_registry_outcomes$RDD5=="N"] <- 1
solvd_registry_outcomes$noncvdth_dt[solvd_registry_outcomes$RDD5=="N"&!is.na(solvd_registry_outcomes$RDD5)] <- 
  solvd_registry_outcomes$rdd2days[solvd_registry_outcomes$RDD5=="N"&!is.na(solvd_registry_outcomes$RDD5)]
solvd_registry_outcomes$noncvdth_status[is.na(solvd_registry_outcomes$noncvdth_status)|
                                          solvd_registry_outcomes$dth_dt > 366|
                                          solvd_registry_outcomes$dth_dt < 0] <- 0

solvd_registry_outcomes$noncvdth_dt[solvd_registry_outcomes$noncvdth_status==0] <- 
  solvd_registry_outcomes$eos_dt[solvd_registry_outcomes$noncvdth_status==0]


#### Hospitalization outcomes

## All-cause hospitalization

solvd_registry_firsthosp <- sqldf('select id_reg, 
                                  1 as hosp_status, 
                                  admtdays as hosp_dt 
                                  from solvd_registry_rhf 
                                  where hosp_dt > 0 
                                  group by id_reg 
                                  having min(hosp_dt)')
solvd_registry_outcomes <- merge(solvd_registry_outcomes,solvd_registry_firsthosp[,c('id_reg','hosp_status','hosp_dt')],by='id_reg',all.x=T)
solvd_registry_outcomes$hosp_status[is.na(solvd_registry_outcomes$hosp_status)|solvd_registry_outcomes$hosp_dt > 366|solvd_registry_outcomes$hosp_dt < 0] <- 0
solvd_registry_outcomes$hosp_dt[solvd_registry_outcomes$hosp_status==0] <- solvd_registry_outcomes$eos_dt[solvd_registry_outcomes$hosp_status==0]


## CV hospitalization

solvd_registry_firstcvhosp <- sqldf('select id_reg, 
                                    1 as cvhosp_status, 
                                    admtdays as cvhosp_dt 
                                    from solvd_registry_rhf 
                                    where RHF5 = "C" and cvhosp_dt > 0 
                                    group by id_reg having min(cvhosp_dt)')
solvd_registry_outcomes <- merge(solvd_registry_outcomes,solvd_registry_firstcvhosp[,c('id_reg','cvhosp_status','cvhosp_dt')],by='id_reg',all.x=T)
solvd_registry_outcomes$cvhosp_status[is.na(solvd_registry_outcomes$cvhosp_status)|solvd_registry_outcomes$cvhosp_dt > 366|solvd_registry_outcomes$cvhosp_dt < 0] <- 0
solvd_registry_outcomes$cvhosp_dt[solvd_registry_outcomes$cvhosp_status==0] <- solvd_registry_outcomes$eos_dt[solvd_registry_outcomes$cvhosp_status==0]


## HF hospitalization

solvd_registry_firsthfhosp <- sqldf('select id_reg, 
                                    1 as hfhosp_status, 
                                    admtdays as hfhosp_dt 
                                    from solvd_registry_rhf 
                                    where RHF7 in ("A","B") and hfhosp_dt > 0 
                                    group by id_reg having min(hfhosp_dt)')
solvd_registry_outcomes <- merge(solvd_registry_outcomes,solvd_registry_firsthfhosp[,c('id_reg','hfhosp_status','hfhosp_dt')],by='id_reg',all.x=T)
solvd_registry_outcomes$hfhosp_status[is.na(solvd_registry_outcomes$hfhosp_status)|solvd_registry_outcomes$hfhosp_dt > 366|solvd_registry_outcomes$hfhosp_dt < 0] <- 0
solvd_registry_outcomes$hfhosp_dt[solvd_registry_outcomes$hfhosp_status==0] <- solvd_registry_outcomes$eos_dt[solvd_registry_outcomes$hfhosp_status==0]


## Non-CV hospitalization

solvd_registry_firstnoncvhosp <- sqldf('select id_reg, 
                                       1 as noncvhosp_status, 
                                       admtdays as noncvhosp_dt 
                                       from solvd_registry_rhf 
                                       where RHF5 = "N" and noncvhosp_dt > 0 
                                       group by id_reg having min(noncvhosp_dt)')
solvd_registry_outcomes <- merge(solvd_registry_outcomes,solvd_registry_firstnoncvhosp[,c('id_reg','noncvhosp_status','noncvhosp_dt')],by='id_reg',all.x=T)
solvd_registry_outcomes$noncvhosp_status[is.na(solvd_registry_outcomes$noncvhosp_status)|solvd_registry_outcomes$noncvhosp_dt > 366|solvd_registry_outcomes$noncvhosp_dt < 0] <- 0
solvd_registry_outcomes$noncvhosp_dt[solvd_registry_outcomes$noncvhosp_status==0] <- solvd_registry_outcomes$eos_dt[solvd_registry_outcomes$noncvhosp_status==0]


#### Composite outcomes ####

## Composite TOPCAT endpoint - CV-cause death + HF hospitalization

solvd_registry_outcomes$cvdth_hfhosp_status <- 0
solvd_registry_outcomes$cvdth_hfhosp_status[solvd_registry_outcomes$cvdth_status==1|solvd_registry_outcomes$hfhosp_status==1] <- 1
solvd_registry_outcomes$cvdth_hfhosp_dt <- pmin(solvd_registry_outcomes$cvdth_dt,solvd_registry_outcomes$hfhosp_dt)

## Composite IPRESERVE endpoint - all-cause death + CV hospitalization

solvd_registry_outcomes$dth_cvhosp_status <- 0
solvd_registry_outcomes$dth_cvhosp_status[solvd_registry_outcomes$dth_status==1|solvd_registry_outcomes$cvhosp_status==1] <- 1
solvd_registry_outcomes$dth_cvhosp_dt <- pmin(solvd_registry_outcomes$dth_dt,solvd_registry_outcomes$cvhosp_dt)

## Composite HF-ACTION endpoint - all-cause death + all-cause hospitalization

solvd_registry_outcomes$dth_hosp_status <- 0
solvd_registry_outcomes$dth_hosp_status[solvd_registry_outcomes$dth_status==1|solvd_registry_outcomes$hosp_status==1] <- 1
solvd_registry_outcomes$dth_hosp_dt <- pmin(solvd_registry_outcomes$dth_dt,solvd_registry_outcomes$hosp_dt)


#### Clean-up ####

solvd_registry_outcomes$study <- 'SOLVD Registry'
solvd_registry_outcomes$patientid <- solvd_registry_outcomes$id_reg
missing_outcomes_solvd_registry <- analysis_outcomes[!analysis_outcomes %in% names(solvd_registry_outcomes)]
solvd_registry_outcomes[,missing_outcomes_solvd_registry] <- NA
solvd_registry_outcomes <- solvd_registry_outcomes[,c(analysis_outcomes)]  


# 
################## ******* DIG ******* ######################
# 
# .%%%%%%%%..%%%%..%%%%%%..
# .%%.....%%..%%..%%....%%.
# .%%.....%%..%%..%%.......
# .%%.....%%..%%..%%...%%%%
# .%%.....%%..%%..%%....%%.
# .%%.....%%..%%..%%....%%.
# .%%%%%%%%..%%%%..%%%%%%..
# 

dig_outcomes <- read.csv(paste(dig_folder,"form02_3.csv",sep=""),na.strings=c("",NA,"NA","NULL"))

dig_outcomes[dig_outcomes=="NaN"] <- NA


#### Mortality outcomes ####

dig_outcomes$dth_status <- 0
dig_outcomes$dth_status[!is.na(dig_outcomes$REASON)] <- 1
dig_outcomes$dth_dt <- dig_outcomes$DEATHDAY

dig_outcomes$cvdth_status <- 0
dig_outcomes$cvdth_status[dig_outcomes$REASON == 1] <- 1
dig_outcomes$cvdth_dt <- dig_outcomes$DEATHDAY

dig_outcomes$noncvdth_status <- 0
dig_outcomes$noncvdth_status[dig_outcomes$REASON==5] <- 1
dig_outcomes$noncvdth_dt <- dig_outcomes$DEATHDAY

dig_outcomes$hfdth_status <- 0
dig_outcomes$hfdth_status[dig_outcomes$REASON==2] <- 1
dig_outcomes$hfdth_dt <- dig_outcomes$DEATHDAY


#### Hospitalization outcomes ####
dig_outcomes$hosp_status <- dig_outcomes$STATUS16
dig_outcomes$hosp_dt <- pmin(dig_outcomes$DAYS16,dig_outcomes$dth_dt)

dig_outcomes$cvhosp_status <- dig_outcomes$STATUS1
dig_outcomes$cvhosp_dt <- pmin(dig_outcomes$DAYS1,dig_outcomes$dth_dt)

dig_outcomes$hfhosp_status <- dig_outcomes$STATUS2
dig_outcomes$hfhosp_dt <- pmin(dig_outcomes$DAYS2,dig_outcomes$dth_dt)

dig_outcomes$noncvhosp_status <- pmax(dig_outcomes$STATUS13,dig_outcomes$STATUS14)
dig_outcomes$noncvhosp_dt <- pmin(dig_outcomes$DAYS13,dig_outcomes$DAYS14,dig_outcomes$dth_dt)

dig_outcomes$arrest_status <- dig_outcomes$STATUS3
dig_outcomes$arrest_dt <- pmin(dig_outcomes$DAYS3,dig_outcomes$dth_dt)


#### Composite outcomes ####

## Composite TOPCAT endpoint - CV-cause death + HF hospitalization

dig_outcomes$cvdth_hfhosp_status <- 0
dig_outcomes$cvdth_hfhosp_status[dig_outcomes$cvdth_status==1|dig_outcomes$hfhosp_status==1] <- 1
dig_outcomes$cvdth_hfhosp_dt <- pmin(dig_outcomes$cvdth_dt,dig_outcomes$hfhosp_dt)


## Composite IPRESERVE endpoint - all-cause death + CV hospitalization

dig_outcomes$dth_cvhosp_status <- 0
dig_outcomes$dth_cvhosp_status[dig_outcomes$dth_status==1|dig_outcomes$cvhosp_status==1] <- 1
dig_outcomes$dth_cvhosp_dt <- pmin(dig_outcomes$dth_dt,dig_outcomes$cvhosp_dt)


## Composite HF-ACTION endpoint - all-cause death +  hospitalization

dig_outcomes$dth_hosp_status <- 0
dig_outcomes$dth_hosp_status[dig_outcomes$dth_status==1|dig_outcomes$hosp_status==1] <- 1
dig_outcomes$dth_hosp_dt <- pmin(dig_outcomes$dth_dt,dig_outcomes$hosp_dt)


#### Clean-up ####

dig_outcomes$patientid <- dig_outcomes$PATIENT
missing_outcomes_dig <- analysis_outcomes[!analysis_outcomes %in% names(dig_outcomes)]
dig_outcomes[,missing_outcomes_dig] <- NA
dig_outcomes <- dig_outcomes[,analysis_outcomes]  
dig_outcomes$study <- 'DIG'


########################### ******* NEAT-HFpEF ******* ##########################
# 
# .%%....%%.%%%%%%%%....%%%....%%%%%%%%.........%%.....%%.%%%%%%%%.%%%%%%%%..%%%%%%%%.%%%%%%%%
# .%%%...%%.%%.........%%.%%......%%............%%.....%%.%%.......%%.....%%.%%.......%%......
# .%%%%..%%.%%........%%...%%.....%%............%%.....%%.%%.......%%.....%%.%%.......%%......
# .%%.%%.%%.%%%%%%...%%.....%%....%%....%%%%%%%.%%%%%%%%%.%%%%%%...%%%%%%%%..%%%%%%...%%%%%%..
# .%%..%%%%.%%.......%%%%%%%%%....%%............%%.....%%.%%.......%%........%%.......%%......
# .%%...%%%.%%.......%%.....%%....%%............%%.....%%.%%.......%%........%%.......%%......
# .%%....%%.%%%%%%%%.%%.....%%....%%............%%.....%%.%%.......%%........%%%%%%%%.%%......
# 

##  NEAT-HFpEF had no death/hospitalization outcomes
## Continuous measurement outcomes
## Labs, PRO, exercise

## TREATMENT 1 = ISMN first then placebo.  2 = Placebo first then ISMN

neat_base <- read.csv(paste(neat_folder,"a_base.csv",sep=""))
neat_outcomes <- read.csv(paste(neat_folder,"a_endpts.csv",sep=""),stringsAsFactors = F)
neat_visitsumm <- read.csv(paste(neat_folder,"a_visitsumm.csv",sep=""),stringsAsFactors = F)
neat_rehosp <- read.csv(paste(neat_folder,"rehosp.csv",sep=""),stringsAsFactors = F)
neat_eos <- read.csv(paste(neat_folder,"eos.csv",sep=""),stringsAsFactors = F)

names(neat_outcomes)[1] <- "patientid"

neat_outcomes <- merge(neat_outcomes,neat_eos, by.x="patientid", by.y="patnumb", all.x=T)
neat_outcomes$dth_dt <- neat_outcomes$LSTCON_D
neat_outcomes$dth_status <- 0

neat_outcomes$cvdth_dt <- neat_outcomes$dth_dt
neat_outcomes$cvdth_status <- 0

neat_outcomes$noncvdth_dt <- neat_outcomes$dth_dt
neat_outcomes$noncvdth_status <- 0


## All-cause hospitalization

neat_firsthosp <- sqldf('select PATNUMB as patientid, 
                          1 as hosp_status,
                          ADMITD_D as hosp_dt 
                          from neat_rehosp
                          where ELECTAD=1
                          group by PATNUMB 
                          having min(hosp_dt)')
neat_outcomes <- merge(neat_outcomes,neat_firsthosp,by="patientid",all.x=T)
neat_outcomes$hosp_status[is.na(neat_outcomes$hosp_status)] <- 0
neat_outcomes$hosp_dt[neat_outcomes$hosp_status==0] <- 
  neat_outcomes$dth_dt[neat_outcomes$hosp_status==0]


## CV hospitalization

neat_firstcvhosp <- sqldf('select PATNUMB as patientid, 
                          1 as cvhosp_status, 
                          ADMITD_D as cvhosp_dt 
                          from neat_rehosp 
                          where HOSPRS in (1,2,3,4,5,6,7)
                          and ELECTAD=1
                          group by PATNUMB 
                          having min(cvhosp_dt)')
neat_outcomes <- merge(neat_outcomes,neat_firstcvhosp,by="patientid",all.x=T)
neat_outcomes$cvhosp_status[is.na(neat_outcomes$cvhosp_status)] <- 0
neat_outcomes$cvhosp_dt[neat_outcomes$cvhosp_status==0] <- 
  neat_outcomes$dth_dt[neat_outcomes$cvhosp_status==0]


## HF hospitalization

neat_firsthfhosp <- sqldf('select PATNUMB as patientid, 
                          1 as hfhosp_status, 
                          ADMITD_D as hfhosp_dt 
                          from neat_rehosp 
                          where HOSPRS in (1)
                          and ELECTAD=1
                          group by PATNUMB 
                          having min(hfhosp_dt)')

neat_outcomes <- merge(neat_outcomes,neat_firsthfhosp,by="patientid",all.x=T)
neat_outcomes$hfhosp_status[is.na(neat_outcomes$hfhosp_status)] <- 0
neat_outcomes$hfhosp_dt[neat_outcomes$hfhosp_status==0] <- 
  neat_outcomes$dth_dt[neat_outcomes$hfhosp_status==0]


## Non-CV hospitalization

neat_firstnoncvhosp <- sqldf('select PATNUMB as patientid, 
                          1 as noncvhosp_status, 
                          ADMITD_D as noncvhosp_dt 
                          from neat_rehosp 
                          where HOSPRS in (8)
                          and ELECTAD=1
                          group by PATNUMB 
                          having min(noncvhosp_dt)')
neat_outcomes <- merge(neat_outcomes,neat_firstnoncvhosp,by="patientid",all.x=T)
neat_outcomes$noncvhosp_status[is.na(neat_outcomes$noncvhosp_status)] <- 0
neat_outcomes$noncvhosp_dt[neat_outcomes$noncvhosp_status==0] <- 
  neat_outcomes$dth_dt[neat_outcomes$noncvhosp_status==0]


#### Composite endpoints #### 

## Composite TOPCAT endpoint - CV death + HF hospitalization

neat_outcomes$cvdth_hfhosp_status <- 0
neat_outcomes$cvdth_hfhosp_status[neat_outcomes$cvdth_status==1|neat_outcomes$hfhosp_status==1] <- 1
neat_outcomes$cvdth_hfhosp_dt <- pmin(neat_outcomes$hfhosp_dt)

## Composite IPRESERVE endpoint - all-cause death + CV hospitalization

neat_outcomes$dth_cvhosp_status <- 0
neat_outcomes$dth_cvhosp_status[neat_outcomes$dth_status==1|neat_outcomes$cvhosp_status==1] <- 1
neat_outcomes$dth_cvhosp_dt <- pmin(neat_outcomes$cvhosp_dt)

## Composite HF-ACTION - all-cause death + all-cause hospitalization

neat_outcomes$dth_hosp_status <- 0
neat_outcomes$dth_hosp_status[neat_outcomes$dth_status==1|neat_outcomes$hosp_status==1] <- 1
neat_outcomes$dth_hosp_dt <- pmin(neat_outcomes$hosp_dt)


#### Clean-up ####

missing_outcomes_neat <- analysis_outcomes[!analysis_outcomes %in% names(neat_outcomes)]
neat_outcomes[,missing_outcomes_neat] <- NA
neat_outcomes <- neat_outcomes[,analysis_outcomes]
neat_outcomes$study <- "NEAT-HFpEF"


############## ******* MOCHA ******* ######################
# 
# .%%.....%%..%%%%%%%...%%%%%%..%%.....%%....%%%...
# .%%%...%%%.%%.....%%.%%....%%.%%.....%%...%%.%%..
# .%%%%.%%%%.%%.....%%.%%.......%%.....%%..%%...%%.
# .%%.%%%.%%.%%.....%%.%%.......%%%%%%%%%.%%.....%%
# .%%.....%%.%%.....%%.%%.......%%.....%%.%%%%%%%%%
# .%%.....%%.%%.....%%.%%....%%.%%.....%%.%%.....%%
# .%%.....%%..%%%%%%%...%%%%%%..%%.....%%.%%.....%%
# 


mocha_death <- read.csv(paste(mocha_folder,"death.csv",sep=""),na.strings=c("NA","NULL",""),stringsAsFactors = F)
mocha_hospcore <- read.csv(paste(mocha_folder,"hospcore.csv",sep=""),na.strings=c("NA","NULL",""),stringsAsFactors = F)
mocha_mort <- read.csv(paste(mocha_folder,"mort.csv",sep=""),na.strings=c("NA","NULL",""),stringsAsFactors = F)

mocha_mort <- merge(mocha_mort,mocha_death[,c('PATNO','DCARDIO')], all.x=T,by="PATNO")

#### Mortality outcomes ####

mocha_mort$dth_dt <- mocha_mort$DAYS
mocha_mort$cvdth_dt <- mocha_mort$DAYS
mocha_mort$noncvdth_dt <- mocha_mort$DAYS
mocha_mort$dth_status <- 0
mocha_mort$dth_status[mocha_mort$DIED==1] <- 1
mocha_mort$cvdth_status <- 0
mocha_mort$cvdth_status[mocha_mort$DIED==1&mocha_mort$DCARDIO==1] <- 1
mocha_mort$noncvdth_status <- 0
mocha_mort$noncvdth_status[mocha_mort$DIED==1&mocha_mort$DCARDIO==2] <- 1


#### Hospitalization outcomes ####

mocha_hospcore$hosp_dt <- as.double(as.Date(mocha_hospcore$VDATE,"%Y-%m-%d")-as.Date(mocha_hospcore$M0DATE,"%Y-%m-%d"))
mocha_hospcore$hosp_status <- 1
mocha_hospcore$hfhosp_status <- 0
mocha_hospcore$hfhosp_status[mocha_hospcore$PRIMARY==1] <- 1
mocha_hospcore$hfhosp_dt[mocha_hospcore$hfhosp_status==1] <- mocha_hospcore$hosp_dt[mocha_hospcore$hfhosp_status==1]
mocha_hospcore$cvhosp_status <- 0
mocha_hospcore$cvhosp_status[mocha_hospcore$PRIMARY %in% c(1,2,3)] <- 1

mocha_hospcore$cvhosp_status[mocha_hospcore$PRIMARY==9&mocha_hospcore$PC_DESC %in% 
                               c("SYNCOPE DUE TO ARRHYTHMIA",
                                 "A-FLUTTER AYMPTOMATIC",
                                 "CABG",
                                 "ORTHOTIC HEART TRANSPLANT",
                                 "ORTHOTOPIC HEART TRANSPLANT",
                                 "UNSTABLE ANGINA",
                                 "SYNCOPE VENTRIC TACHYCARDIA",
                                 "BRADYCARDIA",
                                 "VT/VF",
                                 "CARDIOVERSION FOR A-FIB",
                                 "ATRIAL FIBRILLATION",
                                 "SEVERE BRADYCARDIA")] <- 1

mocha_hospcore$noncvhosp_status <- 0
mocha_hospcore$noncvhosp_status[mocha_hospcore$cvhosp_status==0] <- 1
mocha_hospcore$cvhosp_dt[mocha_hospcore$cvhosp_status==1] <- mocha_hospcore$hosp_dt[mocha_hospcore$cvhosp_status==1]
mocha_hospcore$noncvhosp_dt[mocha_hospcore$noncvhosp_status==1] <- mocha_hospcore$hosp_dt[mocha_hospcore$noncvhosp_status==1]

mocha_firsthosp <- sqldf("select PATNO, 
                         hosp_status,
                         hosp_dt 
                         from mocha_hospcore 
                         where hosp_status=1 
                         group by PATNO having min(hosp_dt)")

mocha_firstcvhosp <- sqldf("select PATNO, 
                           cvhosp_status,
                           cvhosp_dt 
                           from mocha_hospcore 
                           where cvhosp_status=1 
                           group by PATNO having min(cvhosp_dt)")

mocha_firsthfhosp <- sqldf("select PATNO, 
                           hfhosp_status,
                           hfhosp_dt 
                           from mocha_hospcore 
                           where hfhosp_status=1 
                           group by PATNO 
                           having min(hfhosp_dt)")

mocha_firstnoncvhosp <- sqldf("select PATNO, 
                              noncvhosp_status,
                              noncvhosp_dt 
                              from mocha_hospcore 
                              where noncvhosp_status=1 
                              group by PATNO having min(noncvhosp_dt)")

mocha_hosp_outcomes <- merge(mocha_firsthosp,mocha_firstcvhosp,all.x=T,by="PATNO")
mocha_hosp_outcomes <- merge(mocha_hosp_outcomes,mocha_firsthfhosp,all.x=T,by="PATNO")
mocha_hosp_outcomes <- merge(mocha_hosp_outcomes,mocha_firstnoncvhosp,all.x=T,by="PATNO")

mocha_outcomes <- merge(mocha_mort,mocha_hosp_outcomes,by="PATNO",all.x=T)
mocha_outcomes$study <- "MOCHA"

mocha_outcomes$hosp_status[is.na(mocha_outcomes$hosp_status)] <- 0
mocha_outcomes$hosp_dt[mocha_outcomes$hosp_status==0] <- mocha_outcomes$DAYS[mocha_outcomes$hosp_status==0]
mocha_outcomes$cvhosp_status[is.na(mocha_outcomes$cvhosp_status)] <- 0
mocha_outcomes$cvhosp_dt[mocha_outcomes$cvhosp_status==0] <- mocha_outcomes$DAYS[mocha_outcomes$cvhosp_status==0]
mocha_outcomes$hfhosp_status[is.na(mocha_outcomes$hfhosp_status)] <- 0
mocha_outcomes$hfhosp_dt[mocha_outcomes$hfhosp_status==0] <- mocha_outcomes$DAYS[mocha_outcomes$hfhosp_status==0]
mocha_outcomes$noncvhosp_status[is.na(mocha_outcomes$noncvhosp_status)] <- 0
mocha_outcomes$noncvhosp_dt[mocha_outcomes$noncvhosp_status==0] <- mocha_outcomes$DAYS[mocha_outcomes$noncvhosp_status==0]


## Composite TOPCAT endpoint - CV death + HF hospitalization

mocha_outcomes$cvdth_hfhosp_status <- 0
mocha_outcomes$cvdth_hfhosp_status[mocha_outcomes$cvdth_status==1|mocha_outcomes$hfhosp_status==1] <- 1
mocha_outcomes$cvdth_hfhosp_dt <- pmin(mocha_outcomes$cvdth_dt,mocha_outcomes$hfhosp_dt)


## Composite IPRESERVE endpoint - all-cause death + HF hospitalization

mocha_outcomes$dth_cvhosp_status <- 0
mocha_outcomes$dth_cvhosp_status[mocha_outcomes$dth_status==1|mocha_outcomes$cvhosp_status==1] <- 1
mocha_outcomes$dth_cvhosp_dt <- pmin(mocha_outcomes$dth_dt,mocha_outcomes$cvhosp_dt)


## Composite HF-ACTION - all-cause death + all-cause hospitalization

mocha_outcomes$dth_hosp_status <- 0
mocha_outcomes$dth_hosp_status[mocha_outcomes$dth_status==1|mocha_outcomes$hosp_status==1] <- 1
mocha_outcomes$dth_hosp_dt <- pmin(mocha_outcomes$dth_dt,mocha_outcomes$hosp_dt)


#### Clean-up ####

mocha_outcomes$patientid <- mocha_outcomes$PATNO
mocha_outcomes$study <- "MOCHA"
missing_outcomes_mocha <- analysis_outcomes[!analysis_outcomes %in% names(mocha_outcomes)]
mocha_outcomes[,missing_outcomes_mocha] <- NA
mocha_outcomes <- mocha_outcomes[,analysis_outcomes]


# ########################### ******* CORONA ******* #################################
# 
# ..%%%%%%...%%%%%%%..%%%%%%%%...%%%%%%%..%%....%%....%%%...
# .%%....%%.%%.....%%.%%.....%%.%%.....%%.%%%...%%...%%.%%..
# .%%.......%%.....%%.%%.....%%.%%.....%%.%%%%..%%..%%...%%.
# .%%.......%%.....%%.%%%%%%%%..%%.....%%.%%.%%.%%.%%.....%%
# .%%.......%%.....%%.%%...%%...%%.....%%.%%..%%%%.%%%%%%%%%
# .%%....%%.%%.....%%.%%....%%..%%.....%%.%%...%%%.%%.....%%
# ..%%%%%%...%%%%%%%..%%.....%%..%%%%%%%..%%....%%.%%.....%%
# 

corona_analysis <- read.csv(paste(corona_folder,'CORONA sample.csv',sep=""),
                            na.strings=c("","NA","NULL"))

corona_analysis$patientid <- corona_analysis$usubjid
corona_analysis$study <- "CORONA"

corona_outcomes <- corona_analysis[,c('patientid','study','death','cv_death','d_days','cvd_days','d_hf','d_sudd','hfhosp','hftime')]

corona_outcomes$dth_status <- corona_outcomes$death
corona_outcomes$dth_dt <- corona_outcomes$d_days
corona_outcomes$cvdth_status <- corona_outcomes$cv_death
corona_outcomes$cvdth_dt <- corona_outcomes$d_days
corona_outcomes$noncvdth_status[corona_outcomes$dth_status==1&corona_outcomes$cvdth_status==0] <- 1
corona_outcomes$noncvdth_status[is.na(corona_outcomes$noncvdth_status)] <- 0
corona_outcomes$noncvdth_dt <- corona_outcomes$d_days


corona_outcomes$hfhosp_status <- corona_outcomes$hfhosp
corona_outcomes$hfhosp_status[is.na(corona_outcomes$hfhosp)] <- 0
corona_outcomes$hfhosp_dt[is.na(corona_outcomes$hfhosp)] <- corona_outcomes$d_days[is.na(corona_outcomes$hfhosp)]
corona_outcomes$hfhosp_dt[corona_outcomes$hfhosp_status==1] <- corona_outcomes$hftime[corona_outcomes$hfhosp_status==1]

## Composite TOPCAT endpoint - CV death + HF hospitalization

corona_outcomes$cvdth_hfhosp_status <- 0
corona_outcomes$cvdth_hfhosp_status[corona_outcomes$cvdth_status==1|corona_outcomes$hfhosp_status==1] <- 1
corona_outcomes$cvdth_hfhosp_dt <- pmin(corona_outcomes$cvdth_dt,corona_outcomes$hfhosp_dt)


#### Clean-up ####

missing_outcomes_corona <- analysis_outcomes[!analysis_outcomes %in% names(corona_outcomes)]
corona_outcomes[,missing_outcomes_corona] <- NA
corona_outcomes <- corona_outcomes[,analysis_outcomes]
corona_outcomes$study <- "CORONA"


# 
# ######################### ******* CARRESS ******* ############################
# 
# ..%%%%%%.....%%%....%%%%%%%%..%%%%%%%%..%%%%%%%%..%%%%%%...%%%%%%.
# .%%....%%...%%.%%...%%.....%%.%%.....%%.%%.......%%....%%.%%....%%
# .%%........%%...%%..%%.....%%.%%.....%%.%%.......%%.......%%......
# .%%.......%%.....%%.%%%%%%%%..%%%%%%%%..%%%%%%....%%%%%%...%%%%%%.
# .%%.......%%%%%%%%%.%%...%%...%%...%%...%%.............%%.......%%
# .%%....%%.%%.....%%.%%....%%..%%....%%..%%.......%%....%%.%%....%%
# ..%%%%%%..%%.....%%.%%.....%%.%%.....%%.%%%%%%%%..%%%%%%...%%%%%%.
# 


carress_endpts <- read.csv(paste(carress_folder,"a_endpts.csv",sep=""))
carress_rehosptl <- read.csv(paste(carress_folder,"rehosptl.csv",sep=""))
carress_deathpag <- read.csv(paste(carress_folder,"deathpag.csv",sep=""))

carress_deathpag[carress_deathpag=="NaN"] <- NA
carress_deathpag <- subset(carress_deathpag,!is.na(DEATHDT))

carress_endpts[carress_endpts=="NaN"] <- NA
carress_rehosptl[carress_rehosptl=="NaN"] <- NA

#### Mortality outcomes ####

carress_acm <- sqldf('select PATNUMB as patientid, 
                     PTDIED as dth_status, 
                     TIMETODTH as dth_dt, 
                     TIMETODTH as eos_dt 
                     from carress_endpts')

carress_cvm <- sqldf('select PATNUMB as patientid, 
                     1 as cvdth_status, 
                     DEATHDT as cvdth_dt 
                     from carress_deathpag 
                     where DEATHCAU in (1,2,3,4,5,6)')

carress_noncvm <- sqldf('select PATNUMB as patientid, 
                        1 as noncvdth_status, 
                        DEATHDT as noncvdth_dt 
                        from carress_deathpag 
                        where DEATHCAU in (7,8,9)')


carress_indhosp_outcome <- sqldf('select PATNUMB as patientid, 
                                 DCALIVE1 as dc_alive, 
                                 DRNDIS as los 
                                 from carress_endpts')

carress_indhosp_outcome$inhosp_dth_status[carress_indhosp_outcome$dc_alive==1] <- 0
carress_indhosp_outcome$inhosp_dth_status[carress_indhosp_outcome$dc_alive==0] <- 1

carress_outcomes <- merge(carress_acm,carress_cvm,all.x=T)
carress_outcomes <- merge(carress_outcomes,carress_noncvm,all.x=T)
carress_outcomes <- merge(carress_outcomes,carress_indhosp_outcome,all.x=T)

## CV mortality

carress_outcomes$cvdth_status[is.na(carress_outcomes$cvdth_dt)] <- 0
carress_outcomes$cvdth_dt <- carress_outcomes$dth_dt

## Non-CV mortality

carress_outcomes$noncvdth_status[is.na(carress_outcomes$noncvdth_dt)] <- 0
carress_outcomes$noncvdth_dt <- carress_outcomes$dth_dt


#### Hospitalization outcomes ####

## All-cause hospitalization

carress_firsthosp <- sqldf('select PATNUMB as patientid, 
                           1 as hosp_status, 
                           REHOSPDT as hosp_dt 
                           from carress_rehosptl 
                           group by  patientid having min(hosp_dt)')

carress_outcomes <- merge(carress_outcomes,carress_firsthosp,by="patientid",all.x=T)
carress_outcomes$hosp_status[!is.na(carress_outcomes$hosp_dt)] <- 1
carress_outcomes$hosp_status[is.na(carress_outcomes$hosp_dt)] <- 0
carress_outcomes$hosp_dt[carress_outcomes$hosp_status==0] <- carress_outcomes$dth_dt[carress_outcomes$hosp_status==0]


## CV hospitalization

carress_firstcvhosp <- sqldf('select PATNUMB as patientid, 
                            1 as cvhosp_status, 
                            REHOSPDT as cvhosp_dt 
                            from carress_rehosptl 
                            where PRIMCAUS in (1,2,3,4,5,6,7,8,9,10,11,12,28,29) 
                             group by patientid having min(cvhosp_dt)')

carress_outcomes <- merge(carress_outcomes,carress_firstcvhosp,by="patientid",all.x=T)
carress_outcomes$cvhosp_status[!is.na(carress_outcomes$cvhosp_dt)] <- 1
carress_outcomes$cvhosp_status[is.na(carress_outcomes$cvhosp_dt)] <- 0
carress_outcomes$cvhosp_dt[carress_outcomes$cvhosp_status==0] <- carress_outcomes$dth_dt[carress_outcomes$cvhosp_status==0]


## HF hospitalization

carress_firsthfhosp <- sqldf('select PATNUMB as patientid, 
                            1 as hfhosp_status, 
                            REHOSPDT as hfhosp_dt 
                            from carress_rehosptl 
                            where PRIMCAUS in (1) 
                             group by patientid 
                             having min(hfhosp_dt)')

carress_outcomes <- merge(carress_outcomes,carress_firsthfhosp,by="patientid",all.x=T)
carress_outcomes$hfhosp_status[!is.na(carress_outcomes$hfhosp_dt)] <- 1
carress_outcomes$hfhosp_status[is.na(carress_outcomes$hfhosp_dt)] <- 0
carress_outcomes$hfhosp_dt[carress_outcomes$hfhosp_status==0] <- carress_outcomes$dth_dt[carress_outcomes$hfhosp_status==0]


## Non-CV hospitalization

carress_firstnoncvhosp <- sqldf('select PATNUMB as patientid, 
                              1 as noncvhosp_status, 
                              REHOSPDT as noncvhosp_dt 
                              from carress_rehosptl 
                              where PRIMCAUS in (31,32,33,34,48,49) 
                              group by patientid 
                              having min(noncvhosp_dt)')

carress_outcomes <- merge(carress_outcomes,carress_firstnoncvhosp,by="patientid",all.x=T)
carress_outcomes$noncvhosp_status[!is.na(carress_outcomes$noncvhosp_dt)] <- 1
carress_outcomes$noncvhosp_status[is.na(carress_outcomes$noncvhosp_dt)] <- 0
carress_outcomes$noncvhosp_dt[carress_outcomes$noncvhosp_status==0] <- carress_outcomes$dth_dt[carress_outcomes$noncvhosp_status==0]


#### Composite outcomes ####

## Composite TOPCAT endpoint - CV death + HF hospitalization

carress_outcomes$cvdth_hfhosp_status <- 0
carress_outcomes$cvdth_hfhosp_status[carress_outcomes$cvdth_status==1|carress_outcomes$hfhosp_status==1] <- 1
carress_outcomes$cvdth_hfhosp_dt <- pmin(carress_outcomes$cvdth_dt,carress_outcomes$hfhosp_dt)

## Composite IPRESERVE endpoint - all-cause death + CV hospitalization

carress_outcomes$dth_cvhosp_status <- 0
carress_outcomes$dth_cvhosp_status[carress_outcomes$dth_status==1|carress_outcomes$cvhosp_status==1] <- 1
carress_outcomes$dth_cvhosp_dt <- pmin(carress_outcomes$dth_dt,carress_outcomes$cvhosp_dt)

## Composite HF-ACTION - all-cause death + all-cause hospitalization

carress_outcomes$dth_hosp_status <- 0
carress_outcomes$dth_hosp_status[carress_outcomes$dth_status==1|carress_outcomes$hosp_status==1] <- 1
carress_outcomes$dth_hosp_dt <- pmin(carress_outcomes$dth_dt,carress_outcomes$hosp_dt)


#### Clean-up ####

missing_outcomes_carress <- analysis_outcomes[!analysis_outcomes %in% names(carress_outcomes)]
carress_outcomes[,missing_outcomes_carress] <- NA
carress_outcomes <- carress_outcomes[,analysis_outcomes]
carress_outcomes$study <- "CARRESS"


# 
######################### ******* DOSE ******* ###########################
# 
# .%%%%%%%%...%%%%%%%...%%%%%%..%%%%%%%%
# .%%.....%%.%%.....%%.%%....%%.%%......
# .%%.....%%.%%.....%%.%%.......%%......
# .%%.....%%.%%.....%%..%%%%%%..%%%%%%..
# .%%.....%%.%%.....%%.......%%.%%......
# .%%.....%%.%%.....%%.%%....%%.%%......
# .%%%%%%%%...%%%%%%%...%%%%%%..%%%%%%%%
# 


dose_endpts <- read.csv(paste(dose_folder,"analysis/csv/a_endpts.csv",sep=""),na.strings=c("NULL","",NA))
dose_deathpag <- read.csv(paste(dose_folder,"data/csv/deathpag.csv",sep=""),na.strings=c("NULL","",NA))
dose_rehosptl <- read.csv(paste(dose_folder,"data/csv/rehosptl.csv",sep=""),na.strings=c("NULL","",NA))

dose_deathpag <- subset(dose_deathpag,!is.na(DEATHDT))


#### Mortality outcomes ####

## In-hospital mortality

dose_indhosp_outcome <- sqldf('select PATNUMB as patientid, 
                              DCALIVE as dc_alive,
                              DRNDIS as los from dose_endpts')

dose_indhosp_outcome$inhosp_dth_status[dose_indhosp_outcome$dc_alive==1] <- 0
dose_indhosp_outcome$inhosp_dth_status[dose_indhosp_outcome$dc_alive==0] <- 1


## All-cause mortality

dose_acm <- sqldf('select PATNUMB as patientid, 
                  1 as dth_status, 
                  DEATHDT as dth_dt 
                  from dose_deathpag')
dose_outcomes <- merge(dose_indhosp_outcome,dose_acm,all.x=T)
dose_outcomes$dth_status[is.na(dose_outcomes$dth_status)] <- 0
dose_outcomes$dth_dt[dose_outcomes$dth_status==0] <- 60


## CV mortality

dose_cvm <- sqldf('select PATNUMB as patientid, 
                  1 as cvdth_status, 
                  DEATHDT as cvdth_dt 
                  from dose_deathpag 
                  where DEATHCAU in (1,2,3,4,5,6)')


dose_outcomes <- merge(dose_outcomes,dose_cvm,all.x=T)
dose_outcomes$cvdth_status[is.na(dose_outcomes$cvdth_dt)] <- 0
dose_outcomes$cvdth_dt <- dose_outcomes$dth_dt


## Non-CV mortality

dose_noncvm <- sqldf('select PATNUMB as patientid, 
                     1 as noncvdth_status, 
                     DEATHDT as noncvdth_dt 
                     from dose_deathpag 
                     where DEATHCAU in (7,8,9)')
dose_outcomes <- merge(dose_outcomes,dose_noncvm,all.x=T)
dose_outcomes$noncvdth_status[is.na(dose_outcomes$noncvdth_dt)] <- 0
dose_outcomes$noncvdth_dt <- dose_outcomes$dth_dt


#### Hospitalization outcomes ####


## All-cause hospitalization

dose_firsthosp <- sqldf('select PATNUMB as patientid, 
                        1 as hosp_status, 
                        REHOSPDT as hosp_dt 
                        from dose_rehosptl 
                        group by patientid having min(hosp_dt)')
dose_outcomes <- merge(dose_outcomes,dose_firsthosp,all.x=T)
dose_outcomes$hosp_status[is.na(dose_outcomes$hosp_dt)] <- 0
dose_outcomes$hosp_dt[dose_outcomes$hosp_status==0] <- dose_outcomes$dth_dt[dose_outcomes$hosp_status==0]


## CV hospitalization

dose_firstcvhosp <- sqldf('select PATNUMB as patientid,
                          1 as cvhosp_status, 
                          REHOSPDT as cvhosp_dt 
                          from dose_rehosptl 
                          where PRIMCAUS in (1,2,3,4,5,6,7,8,9,10,11,12,28,29) 
                          group by patientid 
                          having min(cvhosp_dt)')
dose_outcomes <- merge(dose_outcomes,dose_firstcvhosp,all.x=T)
dose_outcomes$cvhosp_status[is.na(dose_outcomes$cvhosp_dt)] <- 0
dose_outcomes$cvhosp_dt[dose_outcomes$cvhosp_status==0] <- dose_outcomes$dth_dt[dose_outcomes$cvhosp_status==0]


## HF hospitalization

dose_firsthfhosp <- sqldf('select PATNUMB as patientid, 
                        1 as hfhosp_status, 
                        REHOSPDT as hfhosp_dt 
                        from dose_rehosptl 
                        where PRIMCAUS in (1) 
                        group by patientid 
                        having min(hfhosp_dt)')
dose_outcomes <- merge(dose_outcomes,dose_firsthfhosp,all.x=T)
dose_outcomes$hfhosp_status[is.na(dose_outcomes$hfhosp_dt)] <- 0
dose_outcomes$hfhosp_dt[dose_outcomes$hfhosp_status==0] <- dose_outcomes$dth_dt[dose_outcomes$hfhosp_status==0]


## Non-CV hospitalization

dose_firstnoncvhosp <- sqldf('select PATNUMB as patientid, 
                              1 as noncvhosp_status, 
                              REHOSPDT as noncvhosp_dt 
                              from dose_rehosptl 
                              where PRIMCAUS in (31,32,33,34,48,49) 
                              group by patientid 
                              having min(noncvhosp_dt)')
dose_outcomes <- merge(dose_outcomes,dose_firstnoncvhosp,all.x=T)
dose_outcomes$noncvhosp_status[is.na(dose_outcomes$noncvhosp_dt)] <- 0
dose_outcomes$noncvhosp_dt[dose_outcomes$noncvhosp_status==0] <- dose_outcomes$dth_dt[dose_outcomes$noncvhosp_status==0]


#### Composite outcomes ####

## Composite TOPCAT endpoint - CV death + HF hospitalization

dose_outcomes$cvdth_hfhosp_status <- 0
dose_outcomes$cvdth_hfhosp_status[dose_outcomes$cvdth_status==1|dose_outcomes$hfhosp_status==1] <- 1
dose_outcomes$cvdth_hfhosp_dt <- pmin(dose_outcomes$cvdth_dt,dose_outcomes$hfhosp_dt)

## Composite IPRESERVE endpoint - all-cause death + CV hospitalization

dose_outcomes$dth_cvhosp_status <- 0
dose_outcomes$dth_cvhosp_status[dose_outcomes$dth_status==1|dose_outcomes$cvhosp_status==1] <- 1
dose_outcomes$dth_cvhosp_dt <- pmin(dose_outcomes$dth_dt,dose_outcomes$cvhosp_dt)

## Composite HF-ACTION - all-cause death + all-cause hospitalization

dose_outcomes$dth_hosp_status <- 0
dose_outcomes$dth_hosp_status[dose_outcomes$dth_status==1|dose_outcomes$hosp_status==1] <- 1
dose_outcomes$dth_hosp_dt <- pmin(dose_outcomes$dth_dt,dose_outcomes$hosp_dt)


#### Clean-up ####

missing_outcomes_dose <- analysis_outcomes[!analysis_outcomes %in% names(dose_outcomes)]
dose_outcomes[,missing_outcomes_dose] <- NA
dose_outcomes <- dose_outcomes[,analysis_outcomes]
dose_outcomes$study <- "DOSE"


 
# 
####################### ******* ESCAPE ******* ###########################
# 
# .%%%%%%%%..%%%%%%...%%%%%%.....%%%....%%%%%%%%..%%%%%%%%
# .%%.......%%....%%.%%....%%...%%.%%...%%.....%%.%%......
# .%%.......%%.......%%........%%...%%..%%.....%%.%%......
# .%%%%%%....%%%%%%..%%.......%%.....%%.%%%%%%%%..%%%%%%..
# .%%.............%%.%%.......%%%%%%%%%.%%........%%......
# .%%.......%%....%%.%%....%%.%%.....%%.%%........%%......
# .%%%%%%%%..%%%%%%...%%%%%%..%%.....%%.%%........%%%%%%%%
# 

escape_death <- read.csv(paste(escape_folder,"main/sasdata/death.csv",sep=""),stringsAsFactor=F)
escape_patient <- read.csv(paste(escape_folder,"main/analdata/patient.csv",sep=""),stringsAsFactor=F)
escape_rehosp1 <- read.csv(paste(escape_folder,"main/sasdata/rehosp1.csv",sep=""),stringsAsFactor=F)
escape_rehosp2 <- read.csv(paste(escape_folder,"main/sasdata/rehosp2.csv",sep=""),stringsAsFactor=F)

escape_indhosp_outcome <- sqldf('select DEIDNUM as patientid, DTH_INIT as inhosp_dth_status, HOSPDAY as los from escape_patient')
escape_indhosp_outcome$inhosp_dth_status[is.na(escape_indhosp_outcome$inhosp_dth_status)] <- 0

escape_outcomes <- sqldf('select DEIDNUM as patientid, 
                         LASTFUDT as eos_dt 
                         from escape_patient')


#### Mortality outcomes ####


## All-cause mortality

escape_dth <- sqldf('select DEIDNUM as patientid, 
                    1 as dth_status, 
                    DTHDT as dth_dt 
                    from escape_death')
escape_outcomes <- merge(escape_outcomes,escape_dth, by="patientid", all.x=T)
escape_outcomes$dth_status[is.na(escape_outcomes$dth_status)] <- 0
escape_outcomes$dth_dt[escape_outcomes$dth_status==0] <- escape_outcomes$eos_dt[escape_outcomes$dth_status==0]


## CV mortality

escape_cvdth <- sqldf('select DEIDNUM as patientid, 
                      1 as cvdth_status, 
                      DTHDT as cvdth_dt 
                      from escape_death 
                      where DTHCAUS in (1,2,3,4)')
escape_outcomes <- merge(escape_outcomes,escape_cvdth, by="patientid", all.x=T)
escape_outcomes$cvdth_status[is.na(escape_outcomes$cvdth_status)] <- 0
escape_outcomes$cvdth_dt[escape_outcomes$cvdth_status==0] <- escape_outcomes$eos_dt[escape_outcomes$cvdth_status==0]


## Non-CV mortality

escape_noncvdth <- sqldf('select DEIDNUM as patientid, 
                         1 as noncvdth_status, 
                         DTHDT as noncvdth_dt 
                         from escape_death 
                         where DTHCAUS in (5,6)')


escape_outcomes <- merge(escape_outcomes,escape_noncvdth, by="patientid", all.x=T)
escape_outcomes$noncvdth_status[is.na(escape_outcomes$noncvdth_status)] <- 0
escape_outcomes$noncvdth_dt[escape_outcomes$noncvdth_status==0] <- escape_outcomes$eos_dt[escape_outcomes$noncvdth_status==0]


#### Hospitalization outcomes ####

escape_rehosps <- merge(escape_rehosp1[,c('DEIDNUM','PAGEREP','HSPADMDT','HSPDISDT')],
                        escape_rehosp2[,c("DEIDNUM","PAGEREP",'HOSPRE')],
                        by=c("DEIDNUM","PAGEREP"),all.x=T)


## All-cause hospitalization

escape_hosp <- sqldf('select DEIDNUM as patientid, 
                           1 as hosp_status, 
                           HSPADMDT as hosp_dt 
                           from escape_rehosps 
                           group by patientid 
                           having min(hosp_dt)')
escape_outcomes <- merge(escape_outcomes,escape_hosp, by="patientid", all.x=T)
escape_outcomes$hosp_status[is.na(escape_outcomes$hosp_status)] <- 0
escape_outcomes$hosp_dt[escape_outcomes$hosp_status==0] <- escape_outcomes$eos_dt[escape_outcomes$hosp_status==0]


## CV hospitalization

escape_cvhosp <- sqldf('select DEIDNUM as patientid, 
                          1 as cvhosp_status, 
                          HSPADMDT as cvhosp_dt 
                          from escape_rehosps 
                          where HOSPRE in (1,2,3,4) 
                          group by patientid 
                          having min(cvhosp_dt)')
escape_outcomes <- merge(escape_outcomes,escape_cvhosp, by="patientid", all.x=T)
escape_outcomes$cvhosp_status[is.na(escape_outcomes$cvhosp_status)] <- 0
escape_outcomes$cvhosp_dt[escape_outcomes$cvhosp_status==0] <- escape_outcomes$eos_dt[escape_outcomes$cvhosp_status==0]


## HF hospitalization

escape_hfhosp <- sqldf('select DEIDNUM as patientid, 
                          1 as hfhosp_status, 
                          HSPADMDT as hfhosp_dt 
                          from escape_rehosps 
                          where HOSPRE = 1 
                          group by patientid 
                          having min(hfhosp_dt)')
escape_outcomes <- merge(escape_outcomes,escape_hfhosp, by="patientid", all.x=T)
escape_outcomes$hfhosp_status[is.na(escape_outcomes$hfhosp_status)] <- 0
escape_outcomes$hfhosp_dt[escape_outcomes$hfhosp_status==0] <- escape_outcomes$eos_dt[escape_outcomes$hfhosp_status==0]


## Non-CV hospitalization

escape_noncvhosp <- sqldf('select DEIDNUM as patientid, 
                             1 as noncvhosp_status, 
                             HSPADMDT as noncvhosp_dt 
                             from escape_rehosps 
                             where HOSPRE in (5,6) 
                             group by patientid 
                             having min(noncvhosp_dt)')
escape_outcomes <- merge(escape_outcomes,escape_noncvhosp, by="patientid", all.x=T)
escape_outcomes$noncvhosp_status[is.na(escape_outcomes$noncvhosp_status)] <- 0
escape_outcomes$noncvhosp_dt[escape_outcomes$noncvhosp_status==0] <- escape_outcomes$eos_dt[escape_outcomes$noncvhosp_status==0]


#### Composite outcomes ####

## Composite CV death + HF hospitalization

escape_outcomes$cvdth_hfhosp_status <- 0
escape_outcomes$cvdth_hfhosp_status[escape_outcomes$cvdth_status==1|escape_outcomes$hfhosp_status==1] <- 1
escape_outcomes$cvdth_hfhosp_dt <- pmin(escape_outcomes$cvdth_dt,escape_outcomes$hfhosp_dt)


## Composite IPRESERVE endpoint - all-cause death + CV hospitalization

escape_outcomes$dth_cvhosp_status <- 0
escape_outcomes$dth_cvhosp_status[escape_outcomes$dth_status==1|escape_outcomes$cvhosp_status==1] <- 1
escape_outcomes$dth_cvhosp_dt <- pmin(escape_outcomes$dth_dt,escape_outcomes$cvhosp_dt)

## Composite HF-ACTION - all-cause death + all-cause hospitalization

escape_outcomes$dth_hosp_status <- 0
escape_outcomes$dth_hosp_status[escape_outcomes$dth_status==1|escape_outcomes$hosp_status==1] <- 1
escape_outcomes$dth_hosp_dt <- pmin(escape_outcomes$dth_dt,escape_outcomes$hosp_dt)


#### Clean-up ####

missing_outcomes_escape <- analysis_outcomes[!analysis_outcomes %in% names(escape_outcomes)]
escape_outcomes[,missing_outcomes_escape] <- NA
escape_outcomes <- escape_outcomes[,analysis_outcomes]
escape_outcomes$study <- "ESCAPE"


# ############################## ******* STICH/STICHES ******* ############################
# 
# ..%%%%%%..%%%%%%%%.%%%%..%%%%%%..%%.....%%.......%%..%%%%%%..%%%%%%%%.%%%%..%%%%%%..%%.....%%.%%%%%%%%..%%%%%%.
# .%%....%%....%%.....%%..%%....%%.%%.....%%......%%..%%....%%....%%.....%%..%%....%%.%%.....%%.%%.......%%....%%
# .%%..........%%.....%%..%%.......%%.....%%.....%%...%%..........%%.....%%..%%.......%%.....%%.%%.......%%......
# ..%%%%%%.....%%.....%%..%%.......%%%%%%%%%....%%.....%%%%%%.....%%.....%%..%%.......%%%%%%%%%.%%%%%%....%%%%%%.
# .......%%....%%.....%%..%%.......%%.....%%...%%...........%%....%%.....%%..%%.......%%.....%%.%%.............%%
# .%%....%%....%%.....%%..%%....%%.%%.....%%..%%......%%....%%....%%.....%%..%%....%%.%%.....%%.%%.......%%....%%
# ..%%%%%%.....%%....%%%%..%%%%%%..%%.....%%.%%........%%%%%%.....%%....%%%%..%%%%%%..%%.....%%.%%%%%%%%..%%%%%%.
#

stich_cec <- read.csv(paste(stich_folder,"analysisdata/h1/cec.csv",sep=""),
                      stringsAsFactor=F,
                      na.strings=c(".","NA",""))


stich_cechosp <- read.csv(paste(stich_folder,"sourcedata/h1/cechosp.csv",sep=""),
                                  stringsAsFactor=F,
                                  na.strings=c(".","NA",""))


stich_death <- read.csv(paste(stich_folder,"sourcedata/h1/death.csv",sep=""),
                          stringsAsFactor=F,
                          na.strings=c(".","NA",""))


#### Mortality outcomes ####

stich_outcomes <- sqldf('select deidnum as patientid, 
                        deathc as dth_status, dthfuc as dth_dt,
                        carddthc as cvdth_status, cdthfuc as cvdth_dt,
                        rehospc as hosp_status, rhpdaysc as hosp_dt,
                        cardhspc as cvhosp_status, cardaysc as cvhosp_dt,
                        hfhospc as hfhosp_status, hfdaysc as hfhosp_dt,
                        dthcardc as dth_cvhosp_status, dcfuc as dth_cvhosp_dt
                        from stich_cec')


## Non-CV mortality

stich_noncvdth <- sqldf('select deidnum as patientid,
                                        1 as noncvdth_status,
                                        DETHDY as noncvdth_dt
                                        from stich_death
                                        where DTHCAU=2')

stich_outcomes <- merge(stich_outcomes,stich_noncvdth,
                        by="patientid",
                        all.x=T)
stich_outcomes$noncvdth_status[is.na(stich_outcomes$noncvdth_dt)] <- 0
stich_outcomes$noncvdth_dt <- stich_outcomes$dth_dt


#### Hospitalization outcomes ####


## All-cause hospitalization

stich_outcomes$hosp_dt[stich_outcomes$hosp_status==0&is.na(stich_outcomes$hosp_dt)] <-
  stich_outcomes$dth_dt[stich_outcomes$hosp_status==0&is.na(stich_outcomes$hosp_dt)]


## CV hospitalization

stich_outcomes$cvhosp_dt[stich_outcomes$cvhosp_status==0&is.na(stich_outcomes$cvhosp_dt)] <-
  stich_outcomes$dth_dt[stich_outcomes$cvhosp_status==0&is.na(stich_outcomes$cvhosp_dt)]


## HF hospitalization

stich_outcomes$hfhosp_dt[stich_outcomes$hfhosp_status==0&is.na(stich_outcomes$hfhosp_dt)] <-
  stich_outcomes$dth_dt[stich_outcomes$hfhosp_status==0&is.na(stich_outcomes$hfhosp_dt)]


## Non-CV hospitalization

stich_noncvhosp <- sqldf('select deidnum as patientid,
                                        1 as noncvhosp_status,
                                        HOSPDY as noncvhosp_dt
                                        from stich_cechosp
                                        where REASON=2
                                        group by (patientid)
                          having min(noncvhosp_dt)')
stich_outcomes <- merge(stich_outcomes,stich_noncvhosp,
                        by="patientid",
                        all.x=T)
stich_outcomes$noncvhosp_status[is.na(stich_outcomes$noncvhosp_dt)] <- 0
stich_outcomes$noncvhosp_dt[stich_outcomes$noncvhosp_status==0&is.na(stich_outcomes$noncvhosp_dt)] <-
  stich_outcomes$dth_dt[stich_outcomes$noncvhosp_status==0&is.na(stich_outcomes$noncvhosp_dt)]


#### Composite outcomes ####

## Composite TOPCAT endpoint - CV death + HF hospitalization

stich_outcomes$cvdth_hfhosp_status <- 0
stich_outcomes$cvdth_hfhosp_status[stich_outcomes$cvdth_status==1|stich_outcomes$hfhosp_status==1] <- 1
stich_outcomes$cvdth_hfhosp_dt <- pmin(stich_outcomes$cvdth_dt,stich_outcomes$hfhosp_dt)


## Composite HF-ACTION endpoint - all-cause death + all-cause hospitalization

stich_outcomes$dth_hosp_status <- 0
stich_outcomes$dth_hosp_status[stich_outcomes$dth_status==1|stich_outcomes$hosp_status==1] <- 1
stich_outcomes$dth_hosp_dt <- pmin(stich_outcomes$dth_dt,stich_outcomes$hosp_dt)


#### Clean-up ####

stich_outcomes$study <- "STICH"
missing_outcomes_stich <- analysis_outcomes[!analysis_outcomes %in% names(stich_outcomes)]
stich_outcomes[,missing_outcomes_stich] <- NA
stich_outcomes <- stich_outcomes[,analysis_outcomes]


# ################### --- STICHES --- ########################
# 
# ..######..########.####..######..##.....##.########..######.
# .##....##....##.....##..##....##.##.....##.##.......##....##
# .##..........##.....##..##.......##.....##.##.......##......
# ..######.....##.....##..##.......#########.######....######.
# .......##....##.....##..##.......##.....##.##.............##
# .##....##....##.....##..##....##.##.....##.##.......##....##
# ..######.....##....####..######..##.....##.########..######.


stiches_eos <- read.csv(paste(stiches_folder,"AnalysisData/endstudy.csv",sep=""),
                              stringsAsFactor=F,
                              na.strings=c(".","NA",""))

stiches_hosp <- read.csv(paste(stiches_folder,"analysisdata/hosp.csv",sep=""),
                               stringsAsFactor=F,
                               na.strings=c(".","NA",""))


stiches_cecevent <- read.csv(paste(stiches_folder,"analysisdata/cecevent.csv",sep=""),
                         stringsAsFactor=F,
                         na.strings=c(".","NA",""))


stiches_event_es <- read.csv(paste(stiches_folder,"sourcedata/event_es.csv",sep=""),
                         stringsAsFactor=F,
                         na.strings=c(".","NA",""))

stiches_patients <- read.csv(paste(stiches_folder,"sourcedata/patients.csv",sep=""),
                             stringsAsFactor=F,
                             na.strings=c(".","NA",""))

stiches_outcomes <- merge(stiches_patients[,c("deidnum","death1")],
                        stiches_event_es,
                        by="deidnum",
                        all.x=T)


#### Mortality outcomes #### 

## All-cause mortality

stiches_outcomes$dth_status <- stiches_outcomes$death1
stiches_outcomes$dth_dt <- stiches_outcomes$dthfu1


## CV mortality

stiches_outcomes$cvdth_status <- stiches_outcomes$carddth1
stiches_outcomes$cvdth_dt <- stiches_outcomes$dthfu1


## Non-CV mortality

stiches_outcomes$noncvdth_status <- 0
stiches_outcomes$noncvdth_status[stiches_outcomes$dthcauc0==1|stiches_outcomes$dthcauc2==1] <- 1
stiches_outcomes$noncvdth_dt <- stiches_outcomes$dthfu1


#### Hospitalization outcomes #### 

## All-cause hospitalization

stiches_outcomes$hosp_status <- stiches_outcomes$rehosp1
stiches_outcomes$hosp_dt <- stiches_outcomes$rhspday1


## CV hospitalization

stiches_outcomes$cvhosp_status <- stiches_outcomes$cvhosp1
stiches_outcomes$cvhosp_dt <- stiches_outcomes$cvday1


## HF hospitalization

stiches_outcomes$hfhosp_status <- stiches_outcomes$hfhosp1
stiches_outcomes$hfhosp_dt <- stiches_outcomes$hfday1
stiches_outcomes$hfhosp_dt[stiches_outcomes$hfhosp_status==0] <- stiches_outcomes$dthfu1[stiches_outcomes$hfhosp_status==0]


## Non-CV hospitalization

# Here we define a hospitalization as 'non-CV' if there is a non-CV diagnosis AND non CV diagnosis

stiches_hosp$cvdx <- apply(X=stiches_hosp[c("NEWMR",
                                            "HRTF",
                                            "ARRHY",
                                            "UNSTAANG",
                                            "AMI",
                                            "MONPICD",
                                            "STRK",
                                            "OTHCAR",
                                            "CARCATH",
                                            "PACEKHR",
                                            "PACEKSYN",
                                            "HPPCI",
                                            "ICDIMP",
                                            "CABG",
                                            "LVAD",
                                            "HRTRAN",
                                            "ICDGCH",
                                            "PPMBCH")],
                           MARGIN=1,
                           FUN=function(x) max(x,na.rm=T))

stiches_hosp <- subset(stiches_hosp,!cvdx=="-Inf")

stiches_hosp$noncvdx <- apply(X=stiches_hosp[c("GASTRO","INFECT","MALIG","PULM","RENAL","OTHNCAR")],
                           MARGIN=1,
                           FUN=function(x) max(x,na.rm=T))
stiches_hosp <- subset(stiches_hosp,!noncvdx=="-Inf")

stiches_hosp$noncvhosp_status[stiches_hosp$cvdx==0&stiches_hosp$noncvdx==1] <- 1
stiches_hosp$noncvhosp_status[stiches_hosp$cvdx==1] <- 0

stiches_hosp$noncvhosp_dt[stiches_hosp$noncvhosp_status==1&!is.na(stiches_hosp$noncvhosp_status)] <- 
  stiches_hosp$HOSDY[stiches_hosp$noncvhosp_status==1&!is.na(stiches_hosp$noncvhosp_status)]

stiches_firstnoncvhosp <- sqldf('select deidnum, 
                                noncvhosp_status, 
                                noncvhosp_dt 
                                from stiches_hosp 
                                group by deidnum 
                                having min(noncvhosp_dt)')
stiches_outcomes <- merge(stiches_outcomes,stiches_firstnoncvhosp,by="deidnum",all.x=T)
stiches_outcomes$noncvhosp_status[is.na(stiches_outcomes$noncvhosp_dt)] <- 0
stiches_outcomes$noncvhosp_dt[stiches_outcomes$noncvhosp_status==0] <-
  stiches_outcomes$dth_dt[stiches_outcomes$noncvhosp_status==0]


#### Composite outcomes ####

## Composite TOPCAT endpoint - CV death + HF hospitalization

stiches_outcomes$cvdth_hfhosp_status <- 0
stiches_outcomes$cvdth_hfhosp_status[stiches_outcomes$cvdth_status==1|stiches_outcomes$hfhosp_status==1] <- 1
stiches_outcomes$cvdth_hfhosp_dt <- pmin(stiches_outcomes$cvdth_dt,stiches_outcomes$hfhosp_dt)


## Composite IPRESERVE endpoint - all-cause death + HF hospitalization

stiches_outcomes$dth_cvhosp_status <- stiches_outcomes$dthcard1
stiches_outcomes$dth_cvhosp_dt <- stiches_outcomes$dcfu1


## Composite HF-ACTION - all-cause death + all-cause hospitalization

stiches_outcomes$dth_hosp_status <- stiches_outcomes$dthall1
stiches_outcomes$dth_hosp_dt <- stiches_outcomes$dallfu1


#### Clean-up ####

stiches_outcomes$patientid <- stiches_outcomes$deidnum
stiches_outcomes$study <- "STICHES"

missing_outcomes_stiches <- analysis_outcomes[!analysis_outcomes %in% names(stiches_outcomes)]
stiches_outcomes[,missing_outcomes_stiches] <- NA
stiches_outcomes <- stiches_outcomes[,analysis_outcomes]


# ##########################  ******* EXACT ******* ################################
# 
# .%%%%%%%%.%%.....%%....%%%.....%%%%%%..%%%%%%%%
# .%%........%%...%%....%%.%%...%%....%%....%%...
# .%%.........%%.%%....%%...%%..%%..........%%...
# .%%%%%%......%%%....%%.....%%.%%..........%%...
# .%%.........%%.%%...%%%%%%%%%.%%..........%%...
# .%%........%%...%%..%%.....%%.%%....%%....%%...
# .%%%%%%%%.%%.....%%.%%.....%%..%%%%%%.....%%...

exact_endpts <- read.csv(paste(exact_folder,'a_endpts.csv',sep=""),stringsAsFactors =F )
exact_base <- read.csv(paste(exact_folder,'a_base.csv',sep=""), stringsAsFactors = F)
exact_rehosptl <- read.csv(paste(exact_folder,'rehosptl.csv',sep=""),stringsAsFactors = F)


#### Mortality outcomes

exact_outcomes <- sqldf('select patnumb as patientid, 
                        PTDIED as dth_status, TIMETODTH*7 as dth_dt, 
                        CARDIOVDTH as cvdth_status, TIMETOCVDTH*7 as cvdth_dt,
                        ANYHFHOSP as hfhosp_status, TIMETOHFHOSP*7 as hfhosp_dt
                        from exact_endpts')

exact_outcomes$noncvdth_status <- 0
exact_outcomes$noncvdth_status[exact_outcomes$dth_status==1&exact_outcomes$cvdth_status==0] <- 1
exact_outcomes$noncvdth_dt <- exact_outcomes$dth_dt


#### Hospitalization outcomes ####

## All-cause hospitaliation

exact_hosps <- sqldf('select patnumb as patientid, 
                     1 as hosp_status, 
                     primcaus as primary_cause, 
                     rehospdt as hosp_dt 
                     from exact_rehosptl  
                     group by patientid 
                     having min(hosp_dt)')

exact_outcomes <- merge(exact_outcomes,exact_hosps[,c("patientid","hosp_status","hosp_dt")],by="patientid",all.x=T)
exact_outcomes$hosp_status[is.na(exact_outcomes$hosp_status)] <- 0
exact_outcomes$hosp_dt[exact_outcomes$hosp_status==0] <- 
  exact_outcomes$dth_dt[exact_outcomes$hosp_status==0]


## CV hospitalization

exact_cvhosps <- sqldf('select patnumb as patientid, 
                       1 as cvhosp_status, 
                       primcaus as primary_cause, 
                       rehospdt as cvhosp_dt 
                       from exact_rehosptl 
                       where primary_cause < 30 
                       group by patientid 
                       having min(cvhosp_dt)')

exact_outcomes <- merge(exact_outcomes,exact_cvhosps[,c("patientid","cvhosp_status","cvhosp_dt")],by="patientid",all.x=T)
exact_outcomes$cvhosp_status[is.na(exact_outcomes$cvhosp_status)] <- 0
exact_outcomes$cvhosp_dt[exact_outcomes$cvhosp_status==0] <- 
  exact_outcomes$dth_dt[exact_outcomes$cvhosp_status==0]


## Non-CV hospitalization

exact_noncvhosps <- sqldf('select patnumb as patientid, 
                          1 as noncvhosp_status, 
                          primcaus as primary_cause, 
                          rehospdt as noncvhosp_dt 
                          from exact_rehosptl 
                          where primary_cause > 30 
                          group by patientid 
                          having min(noncvhosp_dt)')

exact_outcomes <- merge(exact_outcomes,exact_noncvhosps[,c("patientid","noncvhosp_status","noncvhosp_dt")],by="patientid",all.x=T)
exact_outcomes$noncvhosp_status[is.na(exact_outcomes$noncvhosp_status)] <- 0
exact_outcomes$noncvhosp_dt[exact_outcomes$noncvhosp_status==0] <- 
  exact_outcomes$dth_dt[exact_outcomes$noncvhosp_status==0]


#### Composite outcomes ####

## Composite TOPCAT endpoint - CV death + HF hospitalization

exact_outcomes$cvdth_hfhosp_status <- 0
exact_outcomes$cvdth_hfhosp_status[exact_outcomes$cvdth_status==1|exact_outcomes$hfhosp_status==1] <- 1
exact_outcomes$cvdth_hfhosp_dt <- pmin(exact_outcomes$cvdth_dt,exact_outcomes$hfhosp_dt)


## Composite IPRESERVE endpoint - all-cause death + HF hospitalization

exact_outcomes$dth_cvhosp_status <- 0
exact_outcomes$dth_cvhosp_status[exact_outcomes$dth_status==1|exact_outcomes$cvhosp_status==1] <- 1
exact_outcomes$dth_cvhosp_dt <- pmin(exact_outcomes$dth_dt,exact_outcomes$cvhosp_dt)

## Composite HF-ACTION - all-cause death + all-cause hospitalization

exact_outcomes$dth_hosp_status <- 0
exact_outcomes$dth_hosp_status[exact_outcomes$dth_status==1|exact_outcomes$hosp_status==1] <- 1
exact_outcomes$dth_hosp_dt <- pmin(exact_outcomes$dth_dt,exact_outcomes$hosp_dt)


#### Clean-up #### 

missing_outcomes_exact <- analysis_outcomes[!analysis_outcomes %in% names(exact_outcomes)]
exact_outcomes[,missing_outcomes_exact] <- NA
exact_outcomes <- exact_outcomes[,analysis_outcomes]
exact_outcomes$study <- "EXACT"


 
########################## ******* ROSE ******* ##################################
# 
# .%%%%%%%%...%%%%%%%...%%%%%%..%%%%%%%%
# .%%.....%%.%%.....%%.%%....%%.%%......
# .%%.....%%.%%.....%%.%%.......%%......
# .%%%%%%%%..%%.....%%..%%%%%%..%%%%%%..
# .%%...%%...%%.....%%.......%%.%%......
# .%%....%%..%%.....%%.%%....%%.%%......
# .%%.....%%..%%%%%%%...%%%%%%..%%%%%%%%
# 


rose_endpts <- read.sas7bdat(paste(rose_folder,"a_endpts.sas7bdat",sep=""))
rose_deathpag <- read.sas7bdat(paste(rose_folder,"deathpag.sas7bdat",sep=""))
rose_discharg <- read.sas7bdat(paste(rose_folder,"discharg.sas7bdat",sep=""))
rose_rehosptl <- read.sas7bdat(paste(rose_folder,"rehosptl.sas7bdat",sep=""))
rose_rsterm <- read.sas7bdat(paste(rose_folder,"rsterm.sas7bdat",sep=""))

rose_deathpag[rose_deathpag=="NaN"] <- NA
rose_deathpag <- subset(rose_deathpag,!is.na(DEATHDT))

rose_endpts[rose_endpts=="NaN"] <- NA
rose_rehosptl[rose_rehosptl=="NaN"] <- NA
rose_rsterm[rose_rsterm=="NaN"] <- NA

rose_indhosp_outcome <- sqldf('select patnumb as patientid, DCALIVE=0 as inhosp_dth_status, DISCHDT as los from rose_discharg')
rose_indhosp_outcome$inhosp_dth_status[is.na(rose_indhosp_outcome$inhosp_dth_status)] <- 0


rose_acm <- sqldf('select PATNUMB as patientid, 
                  PTDIED6MO as dth_status, 
                  TIMETODTH6MO as dth_dt 
                  from rose_endpts')
rose_outcomes <- merge(rose_indhosp_outcome,rose_acm,all.x=T)


## CV mortality

rose_cvm <- sqldf('select PATNUMB as patientid, 
                  1 as cvdth_status, 
                  DEATHDT as cvdth_dt 
                  from rose_deathpag 
                  where DEATHCAU in (1,2,3,4,5,6)')
rose_outcomes <- merge(rose_outcomes,rose_cvm,all.x=T)
rose_outcomes$cvdth_status[is.na(rose_outcomes$cvdth_dt)] <- 0
rose_outcomes$cvdth_dt <- rose_outcomes$dth_dt


## Non-CV mortality

rose_noncvm <- sqldf('select PATNUMB as patientid, 
                     1 as noncvdth_status, 
                     DEATHDT as noncvdth_dt 
                     from rose_deathpag 
                     where DEATHCAU in (7,8,9)')
rose_outcomes <- merge(rose_outcomes,rose_noncvm,all.x=T)
rose_outcomes$noncvdth_status[is.na(rose_outcomes$noncvdth_dt)] <- 0
rose_outcomes$noncvdth_dt <- rose_outcomes$dth_dt


#### Hospitalization outcomes

## All-cause hospitalization

rose_firsthosp <- sqldf('select PATNUMB as patientid, 
                        1 as hosp_status, 
                        REHOSPDT as hosp_dt 
                        from rose_rehosptl 
                        group by patientid 
                        having min(hosp_dt)')
rose_outcomes <- merge(rose_outcomes,rose_firsthosp,all.x=T)
rose_outcomes$hosp_status[is.na(rose_outcomes$hosp_dt)] <- 0
rose_outcomes$hosp_dt[rose_outcomes$hosp_status==0] <- 
  rose_outcomes$dth_dt[rose_outcomes$hosp_status==0]


## CV hospitalization

rose_firstcvhosp <- sqldf('select PATNUMB as patientid, 
                          1 as cvhosp_status, 
                          REHOSPDT as cvhosp_dt 
                          from rose_rehosptl 
                          where PRIMCAUS in (1,2,3,4,5,6,7,8,9,10,11,12,28,29) 
                          group by patientid 
                          having min(cvhosp_dt)')
rose_outcomes <- merge(rose_outcomes,rose_firstcvhosp,all.x=T)
rose_outcomes$cvhosp_status[is.na(rose_outcomes$cvhosp_dt)] <- 0
rose_outcomes$cvhosp_dt[rose_outcomes$cvhosp_status==0] <- 
  rose_outcomes$dth_dt[rose_outcomes$cvhosp_status==0]


## HF hospitalization

rose_firsthfhosp <- sqldf('select PATNUMB as patientid, 
                          1 as hfhosp_status, 
                          REHOSPDT as hfhosp_dt 
                          from rose_rehosptl 
                          where PRIMCAUS in (1) 
                          group by patientid 
                          having min(hfhosp_dt)')
rose_outcomes <- merge(rose_outcomes,rose_firsthfhosp,all.x=T)
rose_outcomes$hfhosp_status[is.na(rose_outcomes$hfhosp_dt)] <- 0
rose_outcomes$hfhosp_dt[rose_outcomes$hfhosp_status==0] <- 
  rose_outcomes$dth_dt[rose_outcomes$hfhosp_status==0]


## Non-CV hospitalization

rose_firstnoncvhosp <- sqldf('select PATNUMB as patientid, 
                              1 as noncvhosp_status, 
                              REHOSPDT as noncvhosp_dt 
                              from rose_rehosptl 
                              where PRIMCAUS in (31,32,33,34,48,49) 
                             group by patientid 
                             having min(noncvhosp_dt)')
rose_outcomes <- merge(rose_outcomes,rose_firstnoncvhosp,all.x=T)
rose_outcomes$noncvhosp_status[is.na(rose_outcomes$noncvhosp_dt)] <- 0
rose_outcomes$noncvhosp_dt[rose_outcomes$noncvhosp_status==0] <- 
  rose_outcomes$dth_dt[rose_outcomes$noncvhosp_status==0]


#### Composite outcomes ####

## Composite TOPCAT endpoint - CV death + HF hospitalization

rose_outcomes$cvdth_hfhosp_status <- 0
rose_outcomes$cvdth_hfhosp_status[rose_outcomes$cvdth_status==1|rose_outcomes$hfhosp_status==1] <- 1
rose_outcomes$cvdth_hfhosp_dt <- pmin(rose_outcomes$cvdth_dt,rose_outcomes$hfhosp_dt)

## Composite IPRESERVE endpoint - all-cause death + CV hospitalization

rose_outcomes$dth_cvhosp_status <- 0
rose_outcomes$dth_cvhosp_status[rose_outcomes$dth_status==1|rose_outcomes$cvhosp_status==1] <- 1
rose_outcomes$dth_cvhosp_dt <- pmin(rose_outcomes$dth_dt,rose_outcomes$cvhosp_dt)

## Composite HF-ACTION - all-cause death + all-cause hospitalization

rose_outcomes$dth_hosp_status <- 0
rose_outcomes$dth_hosp_status[rose_outcomes$dth_status==1|rose_outcomes$hosp_status==1] <- 1
rose_outcomes$dth_hosp_dt <- pmin(rose_outcomes$dth_dt,rose_outcomes$hosp_dt)


#### Clean-up ####

rose_outcomes$study <- "ROSE"
missing_outcomes_rose <- analysis_outcomes[!analysis_outcomes %in% names(rose_outcomes)]
rose_outcomes[,missing_outcomes_rose] <- NA
rose_outcomes <- rose_outcomes[,analysis_outcomes]


# ############################# ******* GUIDE-IT ******* ###################################
# 
# ..%%%%%%...%%.....%%.%%%%.%%%%%%%%..%%%%%%%%.........%%%%.%%%%%%%%
# .%%....%%..%%.....%%..%%..%%.....%%.%%................%%.....%%...
# .%%........%%.....%%..%%..%%.....%%.%%................%%.....%%...
# .%%...%%%%.%%.....%%..%%..%%.....%%.%%%%%%...%%%%%%%..%%.....%%...
# .%%....%%..%%.....%%..%%..%%.....%%.%%................%%.....%%...
# .%%....%%..%%.....%%..%%..%%.....%%.%%................%%.....%%...
# ..%%%%%%....%%%%%%%..%%%%.%%%%%%%%..%%%%%%%%.........%%%%....%%...
#

guideit_endpoints <- read.csv(paste(guideit_folder,'analysis_data/best_endpoints_ads.csv',sep=""),stringsAsFactors=F,na.strings=c("","NA","."))
guideit_hosp1 <- read.csv(paste(guideit_folder,'raw_data/hosp1.csv',sep=""),stringsAsFactors=F,na.strings=c("","NA","."))
guideit_death1 <- read.csv(paste(guideit_folder,'raw_data/death1.csv',sep=""),stringsAsFactors=F,na.strings=c("","NA","."))
guideit_ceceam <- read.csv(paste(guideit_folder,'raw_data/ceceam.csv',sep=""),stringsAsFactors=F,na.strings=c("","NA","."))


guideit_outcomes <- sqldf('select deidnum as patientid, 
                          DEATHADJ as dth_status, dadjdy as dth_dt,
                          dthcvadj as cvdth_status, dcvadjdy as cvdth_dt,
                          dthncadj as noncvdth_status, dncadjdy as noncvdth_dt,
                          hoshfadj as hfhosp_status, hfadjdy as hfhosp_dt,
                          hospall as hosp_status, d2hosp as hosp_dt,
                          dthhfadj as cvdth_hfhosp_status, dhfadjdy as cvdth_hfhosp_dt,
                          hospcvnum as num_cvhosps, hospnum as num_hfhosp 
                          from guideit_endpoints')


##  Might be possible to do CVH and non-CVH using the hosp1 table, but HFH counts don't match with endpts table - probably due to adjudication.

guideit_cvhosp <- sqldf('select deidnum as patientid, 
                       1 as cvhosp_status ,
                       HSPADM_DDY as cvhosp_dt 
                       from guideit_hosp1
                       where ENCOUNTR=3
                       and HSPREAS in (1,2,3)
                       group by patientid 
                       having min(cvhosp_dt)')

guideit_noncvhosp <- sqldf('select deidnum as patientid, 
                       1 as noncvhosp_status,
                       HSPADM_DDY as noncvhosp_dt 
                       from guideit_hosp1
                       where ENCOUNTR=3
                       and HSPREAS=4
                       group by patientid 
                       having min(noncvhosp_dt)')


guideit_outcomes <- merge(guideit_outcomes,guideit_cvhosp,by="patientid",all.x=T)
guideit_outcomes <- merge(guideit_outcomes,guideit_noncvhosp,by="patientid",all.x=T)


guideit_outcomes$cvhosp_dt[guideit_outcomes$hfhosp_status==1&is.na(guideit_outcomes$cvhosp_status)] <- guideit_outcomes$hfhosp_dt[guideit_outcomes$hfhosp_status==1&is.na(guideit_outcomes$cvhosp_status)]
guideit_outcomes$cvhosp_status[guideit_outcomes$hfhosp_status==1&is.na(guideit_outcomes$cvhosp_status)] <- 1

guideit_outcomes$cvhosp_status[is.na(guideit_outcomes$cvhosp_dt)] <- 0
guideit_outcomes$cvhosp_dt[guideit_outcomes$cvhosp_status==0] <- 
  guideit_outcomes$dth_dt[guideit_outcomes$cvhosp_status==0]

guideit_outcomes$noncvhosp_status[is.na(guideit_outcomes$noncvhosp_dt)] <- 0
guideit_outcomes$noncvhosp_dt[guideit_outcomes$noncvhosp_status==0] <- 
  guideit_outcomes$dth_dt[guideit_outcomes$noncvhosp_status==0]


##  Composite IPRESERVE endpoint: death + CV hosp 

guideit_outcomes$dth_cvhosp_status <- 0
guideit_outcomes$dth_cvhosp_status[guideit_outcomes$dth_status==1|guideit_outcomes$cvhosp_status==1] <- 1
guideit_outcomes$dth_cvhosp_dt <- pmin(guideit_outcomes$dth_dt,guideit_outcomes$cvhosp_dt)


## Composite HF-ACTION - all-cause death + all-cause hospitalization

guideit_outcomes$dth_hosp_status <- 0
guideit_outcomes$dth_hosp_status[guideit_outcomes$dth_status==1|guideit_outcomes$hosp_status==1] <- 1
guideit_outcomes$dth_hosp_dt <- pmin(guideit_outcomes$dth_dt,guideit_outcomes$hosp_dt)


#### Clean-up ####

guideit_outcomes$study <- "GUIDE-IT"
missing_outcomes_guideit <- analysis_outcomes[!analysis_outcomes %in% names(guideit_outcomes)]
guideit_outcomes[,missing_outcomes_guideit] <- NA
guideit_outcomes <- guideit_outcomes[,analysis_outcomes]


########################### ********** ATHENA ********** ####################################

# ....%%%....%%%%%%%%.%%.....%%.%%%%%%%%.%%....%%....%%%...
# ...%%.%%......%%....%%.....%%.%%.......%%%...%%...%%.%%..
# ..%%...%%.....%%....%%.....%%.%%.......%%%%..%%..%%...%%.
# .%%.....%%....%%....%%%%%%%%%.%%%%%%...%%.%%.%%.%%.....%%
# .%%%%%%%%%....%%....%%.....%%.%%.......%%..%%%%.%%%%%%%%%
# .%%.....%%....%%....%%.....%%.%%.......%%...%%%.%%.....%%
# .%%.....%%....%%....%%.....%%.%%%%%%%%.%%....%%.%%.....%%

athena_endpts <- read.csv(paste(athena_folder,"a_endpts.csv",sep=""),
                          na.strings=c("","NaN","NULL"),
                          stringsAsFactors = F)

athena_rehosp <- read.csv(paste(athena_folder,"rehosp.csv",sep=""),
                          na.strings=c("","NaN","NULL"),
                          stringsAsFactors = F)

athena_death <- read.csv(paste(athena_folder,"death.csv",sep=""),
                         na.strings=c("","NaN","NULL"),
                         stringsAsFactors = F)


athena_endpts <- merge(athena_endpts,athena_death,by="PATNUMB",all.x=T)

#### Mortality outcomes ####

athena_outcomes <- sqldf('select PATNUMB as patientid, 
                          PTDIED60 as dth_status, 
                          TIME2DTH60 as dth_dt,
                          DISCHARGED,
                          TIME2DC as los
                          from athena_endpts')

athena_outcomes$inhospdth_status[athena_outcomes$DISCHARGED==2] <- 1
athena_outcomes$inhospdth_status[athena_outcomes$DISCHARGED %in% c(0,1)] <- 0


## CV mortality

athena_cvm <- sqldf('select PATNUMB as patientid, 
                          1 as cvdth_status, 
                          DEATHD_D as cvdth_dt 
                          from athena_death 
                          where DTHCAUSE=1')

athena_outcomes <- merge(athena_outcomes,athena_cvm,by="patientid",all.x=T)
athena_outcomes$cvdth_status[is.na(athena_outcomes$cvdth_dt)] <- 0
athena_outcomes$cvdth_dt <- athena_outcomes$dth_dt


## Non-CV mortality

athena_noncvm <- sqldf('select PATNUMB as patientid, 
                          1 as noncvdth_status, 
                          DEATHD_D as noncvdth_dt 
                          from athena_death 
                          where DTHCAUSE=2')
athena_outcomes <- merge(athena_outcomes,athena_noncvm,by="patientid",all.x=T)
athena_outcomes$noncvdth_status[is.na(athena_outcomes$noncvdth_dt)] <- 0
athena_outcomes$noncvdth_dt <- athena_outcomes$dth_dt


#### Hospitalization outcomes ####

## All-cause hospitalization

athena_hosp <- sqldf('select PATNUMB as patientid, 
                          ANYHOSP as hosp_status, 
                          TIME2HOSP as hosp_dt 
                          from athena_endpts 
                          group by patientid 
                          having min(hosp_dt)')
athena_outcomes <- merge(athena_outcomes,athena_hosp,by="patientid",all.x=T)


## CV hospitalization


athena_firstcvhosp <- sqldf('select PATNUMB as patientid, 
                          1 as cvhosp_status, 
                          ADMITD_D as cvhosp_dt 
                          from athena_rehosp 
                          where HOSPRS in (1,2,3,4,5,6,7) 
                          group by patientid 
                          having min(cvhosp_dt)')
athena_outcomes <- merge(athena_outcomes,athena_firstcvhosp,by="patientid",all.x=T)
athena_outcomes$cvhosp_status[is.na(athena_outcomes$cvhosp_dt)] <- 0
athena_outcomes$cvhosp_dt[athena_outcomes$cvhosp_status==0] <- athena_outcomes$dth_dt[athena_outcomes$cvhosp_status==0]


## HF hospitalization

athena_firsthfhosp <- sqldf('select PATNUMB as patientid, 
                          1 as hfhosp_status, 
                          ADMITD_D as hfhosp_dt 
                          from athena_rehosp 
                          where HOSPRS in (1) 
                          group by patientid 
                          having min(hfhosp_dt)')
athena_outcomes <- merge(athena_outcomes,athena_firsthfhosp,by="patientid",all.x=T)
athena_outcomes$hfhosp_status[is.na(athena_outcomes$hfhosp_dt)] <- 0
athena_outcomes$hfhosp_dt[athena_outcomes$hfhosp_status==0] <- athena_outcomes$dth_dt[athena_outcomes$hfhosp_status==0]


## Non-CV hospitalization

athena_firstnoncvhosp <- sqldf('select PATNUMB as patientid, 
                          1 as noncvhosp_status, 
                          ADMITD_D as noncvhosp_dt 
                          from athena_rehosp 
                          where HOSPRS in (8) 
                          group by patientid 
                          having min(noncvhosp_dt)')
athena_outcomes <- merge(athena_outcomes,athena_firstnoncvhosp,by="patientid",all.x=T)
athena_outcomes$noncvhosp_status[is.na(athena_outcomes$noncvhosp_dt)] <- 0
athena_outcomes$noncvhosp_dt[athena_outcomes$noncvhosp_status==0] <- athena_outcomes$dth_dt[athena_outcomes$noncvhosp_status==0]


#### Composite outcomes #### 

## Composite TOPCAT endpoint - CV death + HF hospitalization

athena_outcomes$cvdth_hfhosp_status <- 0
athena_outcomes$cvdth_hfhosp_status[athena_outcomes$cvdth_status==1|athena_outcomes$hfhosp_status==1] <- 1
athena_outcomes$cvdth_hfhosp_dt <- pmin(athena_outcomes$cvdth_dt,athena_outcomes$hfhosp_dt)

## Composite IPRESERVE endpoint - all-cause death + CV hospitalization

athena_outcomes$dth_cvhosp_status <- 0
athena_outcomes$dth_cvhosp_status[athena_outcomes$dth_status==1|athena_outcomes$cvhosp_status==1] <- 1
athena_outcomes$dth_cvhosp_dt <- pmin(athena_outcomes$dth_dt,athena_outcomes$cvhosp_dt)

## Composite HF-ACTION - all-cause death + all-cause hospitalization

athena_outcomes$dth_hosp_status <- 0
athena_outcomes$dth_hosp_status[athena_outcomes$dth_status==1|athena_outcomes$hosp_status==1] <- 1
athena_outcomes$dth_hosp_dt <- pmin(athena_outcomes$dth_dt,athena_outcomes$hosp_dt)


#### Clean-up ####

athena_outcomes$study <- "ATHENA"
missing_outcomes_athena <- analysis_outcomes[!analysis_outcomes %in% names(athena_outcomes)]
athena_outcomes[,missing_outcomes_athena] <- NA
athena_outcomes <- athena_outcomes[,analysis_outcomes]


############################ ********** LIFE ********** ########################################

#
# .&&.......&&&&.&&&&&&&&.&&&&&&&&
# .&&........&&..&&.......&&......
# .&&........&&..&&.......&&......
# .&&........&&..&&&&&&...&&&&&&..
# .&&........&&..&&.......&&......
# .&&........&&..&&.......&&......
# .&&&&&&&&.&&&&.&&.......&&&&&&&&

# Mann DL, Givertz MM, Vader JM, Starling RC, Shah P, Mcnulty, SE, Anstrom KJ, Margulies KB, 
# Kiernan MS, Mahr C, Gupta D, Redfield MM, Lala A, Lewis GD, Devore AD, Desvigne-Nickens P, 
# Hernandez AF, Braunwald E. Effect of Treatment With Sacubitril/Valsartan in Patients With Advanced 
# Heart Failure and Reduced Ejection Fraction A Randomized Clinical Trial JAMA Cardiol. 2021;7(1):17–25. 
# https://doi.org/10.1001/jamacardio.2021.4567

life_endpts335 <- read.csv(paste(life_folder,"endpt335.csv",sep=""),
                         stringsAsFactors = F,
                         na.strings=c(".","","NA"))

life_endpts365 <- read.csv(paste(life_folder,"endpt365.csv",sep=""),
                           stringsAsFactors = F,
                           na.strings=c(".","","NA"))

life_rehosp <- read.csv(paste(life_folder,"rehosp.csv",sep=""),
                           stringsAsFactors = F,
                           na.strings=c(".","","NA"))


#### Mortality outcomes ####

life_outcomes <- 
  sqldf("select patnumb as patientid,
        PTDIED24 as dth_status,
        TIME2DTH24 as dth_dt,
        CVDTH24_ADJ as cvdth_status,
        TIME2CVDTH24_ADJ as cvdth_dt,
        ANYCVDTHHFHOSP_POST24 AS cvdth_hfhosp_status,
        TIME2CVDTHHFHOSP24_ADJ as cvdth_hfhosp_dt,
        ANYHFHOSP_POST24 as hfhosp_status,
        TIME2HFHOSP24 as hfhosp_dt
        from life_endpts365")
  
life_outcomes$noncvdth_status <- 0
life_outcomes$noncvdth_status[life_outcomes$dth_status==1&life_outcomes$cvdth_status==0] <- 1
life_outcomes$noncvdth_dt <- life_outcomes$dth_dt

life_hosp <-
  sqldf("select patnumb as patientid,
        1 as hosp_status,
        admitd_d as hosp_dt
        from life_rehosp
        where hosp_dt > 0
        group by patientid
        having min(hosp_dt)")


life_cvhosp <-
  sqldf("select patnumb as patientid,
        1 as cvhosp_status,
        admitd_d as cvhosp_dt
        from life_rehosp
        where cvhosp_dt > 0
        and HOSPRS in (1,2,3,4,5,6,7)
        group by patientid
        having min(cvhosp_dt)")

life_noncvhosp <-
  sqldf("select patnumb as patientid,
        1 as noncvhosp_status,
        admitd_d as noncvhosp_dt
        from life_rehosp
        where noncvhosp_dt > 0
        and HOSPRS = 8
        group by patientid
        having min(noncvhosp_dt)")

life_outcomes <-
  merge(life_outcomes,
        life_hosp,
        by="patientid",
        all.x=T)

life_outcomes$hosp_status[is.na(life_outcomes$hosp_status)] <- 0
life_outcomes$hosp_dt[life_outcomes$hosp_status==0] <- 
  life_outcomes$dth_dt[life_outcomes$hosp_status==0]

life_outcomes <-
  merge(life_outcomes,
        life_cvhosp,
        by="patientid",
        all.x=T)

life_outcomes$cvhosp_status[is.na(life_outcomes$cvhosp_status)] <- 0
life_outcomes$cvhosp_dt[life_outcomes$cvhosp_status==0] <- 
  life_outcomes$dth_dt[life_outcomes$cvhosp_status==0]


life_outcomes <-
  merge(life_outcomes,
        life_noncvhosp,
        by="patientid",
        all.x=T)

life_outcomes$noncvhosp_status[is.na(life_outcomes$noncvhosp_status)] <- 0
life_outcomes$noncvhosp_dt[life_outcomes$noncvhosp_status==0] <- 
  life_outcomes$dth_dt[life_outcomes$noncvhosp_status==0]

life_outcomes$dth_cvhosp_status <- 0
life_outcomes$dth_cvhosp_status[life_outcomes$dth_status==1|life_outcomes$cvhosp_status==1] <- 1
life_outcomes$dth_cvhosp_dt <- pmin(life_outcomes$dth_dt,life_outcomes$cvhosp_dt)


life_outcomes$study <- "LIFE"
missing_outcomes_life <- analysis_outcomes[!analysis_outcomes %in% names(life_outcomes)]
life_outcomes[,missing_outcomes_life] <- NA
life_outcomes <- life_outcomes[,analysis_outcomes]


############################ **********FIGHT********** ########################################

# .%%%%%%%%.%%%%..%%%%%%...%%.....%%.%%%%%%%%
# .%%........%%..%%....%%..%%.....%%....%%...
# .%%........%%..%%........%%.....%%....%%...
# .%%%%%%....%%..%%...%%%%.%%%%%%%%%....%%...
# .%%........%%..%%....%%..%%.....%%....%%...
# .%%........%%..%%....%%..%%.....%%....%%...
# .%%.......%%%%..%%%%%%...%%.....%%....%%...

# Margulies KB, Hernandez AF, Redfield MM, Givertz MM, Oliveira GH, Cole R, Mann DL, 
# Whellan DJ, Kiernan MS, Felker GM, McNulty SE, Anstrom KJ, Shah MR, Braunwald E, Cappola TP, 
# for the NHLBI Heart Failure Clinical Research Network. 
# Effects of Liraglutide on Clinical Stability Among Patients With Advanced Heart Failure
# and Reduced Ejection Fraction A Randomized Clinical Trial JAMA. 2016;316(5):500–508. 
# https://doi.org/10.1001/jama.2016.10260


fight_endpts <- read.csv(paste(fight_folder,"a_endpts.csv",sep=""),
                         stringsAsFactors = F,
                         na.strings=c(".","","NA"))

fight_adhasmt <- read.csv(paste(fight_folder,"adhasmt.csv",sep=""),
                          stringsAsFactors = F,
                          na.strings=c(".","","NA"))

fight_death <- read.csv(paste(fight_folder,"death.csv",sep=""),
                        stringsAsFactors = F,
                        na.strings=c(".","","NA"))

fight_rehosp <- read.csv(paste(fight_folder,"rehosp.csv",sep=""),
                         stringsAsFactors = F,
                         na.strings=c(".","","NA"))

fight_edvisit <- read.csv(paste(fight_folder,"edvisit.csv",sep=""),
                          stringsAsFactors = F,
                          na.strings=c(".","","NA"))

fight_eos <-  read.csv(paste(fight_folder,"eos.csv",sep=""),
                       stringsAsFactors = F,
                       na.strings=c(".","","NA"))


#### Mortality outcomes ####

fight_death <- subset(fight_death, 
                      PATNUMB %in% fight_endpts$PATNUMB[fight_endpts$PTDIED180==1])

fight_endpts <- merge(fight_endpts,fight_death[,c("PATNUMB",
                                                  "DTHCARD",
                                                  "DTHCAUSE")],
                      by="PATNUMB",
                      all.x=T)

fight_outcomes <- fight_endpts[,c("PATNUMB",
                                  "PTDIED180",
                                  "DTHCAUSE",
                                  "DTHCARD",
                                  "TIME2DTH180",
                                  "ANYHFHOSP_BA",
                                  "TIME2HFHOSP_BA")]


#### Mortality outcomes

## All-cause mortality

fight_outcomes$dth_status <- fight_outcomes$PTDIED180
fight_outcomes$dth_dt <- fight_outcomes$TIME2DTH180


## CV mortality

fight_outcomes$cvdth_status[fight_outcomes$DTHCAUSE==1] <- 1
fight_outcomes$cvdth_status[fight_outcomes$DTHCAUSE==2|is.na(fight_outcomes$DTHCAUSE)] <- 0
fight_outcomes$cvdth_dt <- fight_outcomes$TIME2DTH180


## Non-CV mortality

fight_outcomes$noncvdth_status[fight_outcomes$DTHCAUSE==2] <- 1
fight_outcomes$noncvdth_status[fight_outcomes$DTHCAUSE==1|is.na(fight_outcomes$DTHCAUSE)] <- 0
fight_outcomes$noncvdth_dt <- fight_outcomes$TIME2DTH180


#### Hospitalization outcomes ####


## All-cause hospitalization

fight_firsthosp <- sqldf('select PATNUMB, 
                          1 as hosp_status,
                          ADMITD_D as hosp_dt 
                          from fight_rehosp
                          where ELECTAD=1
                          group by PATNUMB 
                          having min(hosp_dt)')
fight_outcomes <- merge(fight_outcomes,fight_firsthosp,by="PATNUMB",all.x=T)
fight_outcomes$hosp_status[is.na(fight_outcomes$hosp_status)] <- 0
fight_outcomes$hosp_dt[fight_outcomes$hosp_status==0] <- 
  fight_outcomes$dth_dt[fight_outcomes$hosp_status==0]


## CV hospitalization

fight_firstcvhosp <- sqldf('select PATNUMB, 
                          1 as cvhosp_status, 
                          ADMITD_D as cvhosp_dt 
                          from fight_rehosp 
                          where HOSPRS in (1,2,3,4,5,6,7)
                          and ELECTAD=1
                          group by PATNUMB 
                          having min(cvhosp_dt)')
fight_outcomes <- merge(fight_outcomes,fight_firstcvhosp,by="PATNUMB",all.x=T)
fight_outcomes$cvhosp_status[is.na(fight_outcomes$cvhosp_status)] <- 0
fight_outcomes$cvhosp_dt[fight_outcomes$cvhosp_status==0] <- 
  fight_outcomes$dth_dt[fight_outcomes$cvhosp_status==0]


## HF hospitalization

fight_outcomes$hfhosp_status <- fight_outcomes$ANYHFHOSP_BA
fight_outcomes$hfhosp_dt <- fight_outcomes$TIME2HFHOSP_BA


## Non-CV hospitalization

fight_firstnoncvhosp <- sqldf('select PATNUMB, 
                          1 as noncvhosp_status, 
                          ADMITD_D as noncvhosp_dt 
                          from fight_rehosp 
                          where HOSPRS in (8)
                          and ELECTAD=1
                          group by PATNUMB 
                          having min(noncvhosp_dt)')
fight_outcomes <- merge(fight_outcomes,fight_firstnoncvhosp,by="PATNUMB",all.x=T)
fight_outcomes$noncvhosp_status[is.na(fight_outcomes$noncvhosp_status)] <- 0
fight_outcomes$noncvhosp_dt[fight_outcomes$noncvhosp_status==0] <- 
  fight_outcomes$dth_dt[fight_outcomes$noncvhosp_status==0]


#### Composite endpoints #### 

## Composite TOPCAT endpoint - CV death + HF hospitalization

fight_outcomes$cvdth_hfhosp_status <- 0
fight_outcomes$cvdth_hfhosp_status[fight_outcomes$cvdth_status==1|fight_outcomes$hfhosp_status==1] <- 1
fight_outcomes$cvdth_hfhosp_dt <- pmin(fight_outcomes$cvdth_dt,fight_outcomes$hfhosp_dt)

## Composite IPRESERVE endpoint - all-cause death + CV hospitalization

fight_outcomes$dth_cvhosp_status <- 0
fight_outcomes$dth_cvhosp_status[fight_outcomes$dth_status==1|fight_outcomes$cvhosp_status==1] <- 1
fight_outcomes$dth_cvhosp_dt <- pmin(fight_outcomes$dth_dt,fight_outcomes$cvhosp_dt)

## Composite HF-ACTION - all-cause death + all-cause hospitalization

fight_outcomes$dth_hosp_status <- 0
fight_outcomes$dth_hosp_status[fight_outcomes$dth_status==1|fight_outcomes$hosp_status==1] <- 1
fight_outcomes$dth_hosp_dt <- pmin(fight_outcomes$dth_dt,fight_outcomes$hosp_dt)


#### Clean-up ####

names(fight_outcomes)[1] <- "patientid"
fight_outcomes$study <- "FIGHT"
missing_outcomes_fight <- analysis_outcomes[!analysis_outcomes %in% names(fight_outcomes)]
fight_outcomes[,missing_outcomes_fight] <- NA
fight_outcomes <- fight_outcomes[,analysis_outcomes]


########################################### **************** IRONOUT ***************** #############################################
#
# .%%%%.%%%%%%%%...%%%%%%%..%%....%%..%%%%%%%..%%.....%%.%%%%%%%%
# ..%%..%%.....%%.%%.....%%.%%%...%%.%%.....%%.%%.....%%....%%...
# ..%%..%%.....%%.%%.....%%.%%%%..%%.%%.....%%.%%.....%%....%%...
# ..%%..%%%%%%%%..%%.....%%.%%.%%.%%.%%.....%%.%%.....%%....%%...
# ..%%..%%...%%...%%.....%%.%%..%%%%.%%.....%%.%%.....%%....%%...
# ..%%..%%....%%..%%.....%%.%%...%%%.%%.....%%.%%.....%%....%%...
# .%%%%.%%.....%%..%%%%%%%..%%....%%..%%%%%%%...%%%%%%%.....%%...


ironout_endpts <- read.csv(paste(ironout_folder,"a_endpts.csv",sep=""),
                         stringsAsFactors = F,
                         na.strings=c(".","","NA"))


ironout_death <- read.csv(paste(ironout_folder,"death.csv",sep=""),
                        stringsAsFactors = F,
                        na.strings=c(".","","NA"))

ironout_rehosp <- read.csv(paste(ironout_folder,"rehosp.csv",sep=""),
                         stringsAsFactors = F,
                         na.strings=c(".","","NA"))

ironout_edvisit <- read.csv(paste(ironout_folder,"edvisit.csv",sep=""),
                          stringsAsFactors = F,
                          na.strings=c(".","","NA"))

ironout_eos <-  read.csv(paste(ironout_folder,"eos.csv",sep=""),
                       stringsAsFactors = F,
                       na.strings=c(".","","NA"))


#### Mortality outcomes ####


ironout_endpts <- merge(ironout_endpts,ironout_death[,c("PATNUMB",
                                                  "DTHCARD",
                                                  "DTHCAUSE")],
                      by="PATNUMB",
                      all.x=T)

ironout_outcomes <- ironout_endpts[,c("PATNUMB",
                                  "PTDIEDW16",
                                  "DTHCAUSE",
                                  "DTHCARD",
                                  "TIME2DTH",
                                  "ANYHFHOSP",
                                  "TIME2HFHOSP")]


#### Mortality outcomes

## All-cause mortality

ironout_outcomes$dth_status <- ironout_outcomes$PTDIEDW16
ironout_outcomes$dth_dt <- ironout_outcomes$TIME2DTH


## CV mortality

ironout_outcomes$cvdth_status[ironout_outcomes$DTHCAUSE==1] <- 1
ironout_outcomes$cvdth_status[ironout_outcomes$DTHCAUSE==2|ironout_outcomes$PTDIEDW16==0] <- 0
ironout_outcomes$cvdth_dt <- ironout_outcomes$TIME2DTH


## Non-CV mortality

ironout_outcomes$noncvdth_status[ironout_outcomes$DTHCAUSE==2] <- 1
ironout_outcomes$noncvdth_status[ironout_outcomes$DTHCAUSE==1|ironout_outcomes$PTDIEDW16==0] <- 0
ironout_outcomes$noncvdth_dt <- ironout_outcomes$TIME2DTH


#### Hospitalization outcomes ####


## All-cause hospitalization

ironout_firsthosp <- sqldf('select PATNUMB, 
                          1 as hosp_status,
                          ADMITD_D as hosp_dt 
                          from ironout_rehosp
                          where ELECTAD=1
                          group by PATNUMB 
                          having min(hosp_dt)')
ironout_outcomes <- merge(ironout_outcomes,ironout_firsthosp,by="PATNUMB",all.x=T)
ironout_outcomes$hosp_status[is.na(ironout_outcomes$hosp_status)] <- 0
ironout_outcomes$hosp_dt[ironout_outcomes$hosp_status==0] <- 
  ironout_outcomes$dth_dt[ironout_outcomes$hosp_status==0]


## CV hospitalization

ironout_firstcvhosp <- sqldf('select PATNUMB, 
                          1 as cvhosp_status, 
                          ADMITD_D as cvhosp_dt 
                          from ironout_rehosp 
                          where HOSPRS in (1,2,3,4,5,6,7)
                          and ELECTAD=1
                          group by PATNUMB 
                          having min(cvhosp_dt)')
ironout_outcomes <- merge(ironout_outcomes,ironout_firstcvhosp,by="PATNUMB",all.x=T)
ironout_outcomes$cvhosp_status[is.na(ironout_outcomes$cvhosp_status)] <- 0
ironout_outcomes$cvhosp_dt[ironout_outcomes$cvhosp_status==0] <- 
  ironout_outcomes$dth_dt[ironout_outcomes$cvhosp_status==0]


## HF hospitalization

ironout_outcomes$hfhosp_status <- ironout_outcomes$ANYHFHOSP
ironout_outcomes$hfhosp_dt <- ironout_outcomes$TIME2HFHOSP


## Non-CV hospitalization

ironout_firstnoncvhosp <- sqldf('select PATNUMB, 
                          1 as noncvhosp_status, 
                          ADMITD_D as noncvhosp_dt 
                          from ironout_rehosp 
                          where HOSPRS in (8)
                          and ELECTAD=1
                          group by PATNUMB 
                          having min(noncvhosp_dt)')
ironout_outcomes <- merge(ironout_outcomes,ironout_firstnoncvhosp,by="PATNUMB",all.x=T)
ironout_outcomes$noncvhosp_status[is.na(ironout_outcomes$noncvhosp_status)] <- 0
ironout_outcomes$noncvhosp_dt[ironout_outcomes$noncvhosp_status==0] <- 
  ironout_outcomes$dth_dt[ironout_outcomes$noncvhosp_status==0]


#### Composite endpoints #### 

## Composite TOPCAT endpoint - CV death + HF hospitalization

ironout_outcomes$cvdth_hfhosp_status <- 0
ironout_outcomes$cvdth_hfhosp_status[ironout_outcomes$cvdth_status==1|ironout_outcomes$hfhosp_status==1] <- 1
ironout_outcomes$cvdth_hfhosp_dt <- pmin(ironout_outcomes$cvdth_dt,ironout_outcomes$hfhosp_dt)

## Composite IPRESERVE endpoint - all-cause death + CV hospitalization

ironout_outcomes$dth_cvhosp_status <- 0
ironout_outcomes$dth_cvhosp_status[ironout_outcomes$dth_status==1|ironout_outcomes$cvhosp_status==1] <- 1
ironout_outcomes$dth_cvhosp_dt <- pmin(ironout_outcomes$dth_dt,ironout_outcomes$cvhosp_dt)

## Composite HF-ACTION - all-cause death + all-cause hospitalization

ironout_outcomes$dth_hosp_status <- 0
ironout_outcomes$dth_hosp_status[ironout_outcomes$dth_status==1|ironout_outcomes$hosp_status==1] <- 1
ironout_outcomes$dth_hosp_dt <- pmin(ironout_outcomes$dth_dt,ironout_outcomes$hosp_dt)


#### Clean-up ####

names(ironout_outcomes)[1] <- "patientid"
ironout_outcomes$study <- "IRONOUT"
missing_outcomes_ironout <- analysis_outcomes[!analysis_outcomes %in% names(ironout_outcomes)]
ironout_outcomes[,missing_outcomes_ironout] <- NA
ironout_outcomes <- ironout_outcomes[,analysis_outcomes]


########################################### ***************** INDIE-HFpEF ************ #############################################
#
# .%%%%.%%....%%.%%%%%%%%..%%%%.%%%%%%%%.........%%.....%%.%%%%%%%%.%%%%%%%%..%%%%%%%%.%%%%%%%%
# ..%%..%%%...%%.%%.....%%..%%..%%...............%%.....%%.%%.......%%.....%%.%%.......%%......
# ..%%..%%%%..%%.%%.....%%..%%..%%...............%%.....%%.%%.......%%.....%%.%%.......%%......
# ..%%..%%.%%.%%.%%.....%%..%%..%%%%%%...%%%%%%%.%%%%%%%%%.%%%%%%...%%%%%%%%..%%%%%%...%%%%%%..
# ..%%..%%..%%%%.%%.....%%..%%..%%...............%%.....%%.%%.......%%........%%.......%%......
# ..%%..%%...%%%.%%.....%%..%%..%%...............%%.....%%.%%.......%%........%%.......%%......
# .%%%%.%%....%%.%%%%%%%%..%%%%.%%%%%%%%.........%%.....%%.%%.......%%........%%%%%%%%.%%......


indie_endpts <- read.csv(paste(indie_folder,"a_endpts.csv",sep=""),
                         stringsAsFactors = F,
                         na.strings=c(".","","NA"))


indie_death <- read.csv(paste(indie_folder,"death.csv",sep=""),
                        stringsAsFactors = F,
                        na.strings=c(".","","NA"))


indie_rehosp <- read.csv(paste(indie_folder,"rehosp.csv",sep=""),
                         stringsAsFactors = F,
                         na.strings=c(".","","NA"))


indie_edvisit <- read.csv(paste(indie_folder,"edvisit.csv",sep=""),
                          stringsAsFactors = F,
                          na.strings=c(".","","NA"))

indie_eos <-  read.csv(paste(indie_folder,"eos.csv",sep=""),
                       stringsAsFactors = F,
                       na.strings=c(".","","NA"))


indie_outcomes <- merge(indie_endpts,indie_death[,c("PATNUMB","DTHCAUSE","DTHCARD")], all.x=T)
indie_outcomes <- merge(indie_outcomes,indie_eos[,c("PATNUMB","LSTCON_D")], all.x=T)


#### Mortality outcomes

## All-cause mortality

indie_outcomes$dth_status <- 0
indie_outcomes$dth_status[!is.na(indie_outcomes$DTHCAUSE)] <- 1
indie_outcomes$dth_dt <- indie_outcomes$LSTCON_D


## CV mortality

indie_outcomes$cvdth_status[indie_outcomes$DTHCAUSE==1] <- 1
indie_outcomes$cvdth_status[indie_outcomes$DTHCAUSE==2|is.na(indie_outcomes$DTHCAUSE)] <- 0
indie_outcomes$cvdth_dt <- indie_outcomes$dth_dt


## Non-CV mortality

indie_outcomes$noncvdth_status[indie_outcomes$DTHCAUSE==2] <- 1
indie_outcomes$noncvdth_status[indie_outcomes$DTHCAUSE==1|is.na(indie_outcomes$DTHCAUSE)] <- 0
indie_outcomes$noncvdth_dt <- indie_outcomes$dth_dt


#### Hospitalization outcomes ####


## All-cause hospitalization

indie_firsthosp <- sqldf('select PATNUMB, 
                          1 as hosp_status,
                          ADMITD_D as hosp_dt 
                          from indie_rehosp
                          where ELECTAD=1
                          group by PATNUMB 
                          having min(hosp_dt)')
indie_outcomes <- merge(indie_outcomes,indie_firsthosp,by="PATNUMB",all.x=T)
indie_outcomes$hosp_status[is.na(indie_outcomes$hosp_status)] <- 0
indie_outcomes$hosp_dt[indie_outcomes$hosp_status==0] <- 
  indie_outcomes$dth_dt[indie_outcomes$hosp_status==0]


## CV hospitalization

indie_firstcvhosp <- sqldf('select PATNUMB, 
                          1 as cvhosp_status, 
                          ADMITD_D as cvhosp_dt 
                          from indie_rehosp 
                          where HOSPRS in (1,2,3,4,5,6,7)
                          and ELECTAD=1
                          group by PATNUMB 
                          having min(cvhosp_dt)')
indie_outcomes <- merge(indie_outcomes,indie_firstcvhosp,by="PATNUMB",all.x=T)
indie_outcomes$cvhosp_status[is.na(indie_outcomes$cvhosp_status)] <- 0
indie_outcomes$cvhosp_dt[indie_outcomes$cvhosp_status==0] <- 
  indie_outcomes$dth_dt[indie_outcomes$cvhosp_status==0]


## HF hospitalization

indie_firsthfhosp <- sqldf('select PATNUMB, 
                          1 as hfhosp_status, 
                          ADMITD_D as hfhosp_dt 
                          from indie_rehosp 
                          where HOSPRS in (1)
                          and ELECTAD=1
                          group by PATNUMB 
                          having min(hfhosp_dt)')

indie_outcomes <- merge(indie_outcomes,indie_firsthfhosp,by="PATNUMB",all.x=T)
indie_outcomes$hfhosp_status[is.na(indie_outcomes$hfhosp_status)] <- 0
indie_outcomes$hfhosp_dt[indie_outcomes$hfhosp_status==0] <- 
  indie_outcomes$dth_dt[indie_outcomes$hfhosp_status==0]


## Non-CV hospitalization

indie_firstnoncvhosp <- sqldf('select PATNUMB, 
                          1 as noncvhosp_status, 
                          ADMITD_D as noncvhosp_dt 
                          from indie_rehosp 
                          where HOSPRS in (8)
                          and ELECTAD=1
                          group by PATNUMB 
                          having min(noncvhosp_dt)')
indie_outcomes <- merge(indie_outcomes,indie_firstnoncvhosp,by="PATNUMB",all.x=T)
indie_outcomes$noncvhosp_status[is.na(indie_outcomes$noncvhosp_status)] <- 0
indie_outcomes$noncvhosp_dt[indie_outcomes$noncvhosp_status==0] <- 
  indie_outcomes$dth_dt[indie_outcomes$noncvhosp_status==0]


#### Composite endpoints #### 

## Composite TOPCAT endpoint - CV death + HF hospitalization

indie_outcomes$cvdth_hfhosp_status <- 0
indie_outcomes$cvdth_hfhosp_status[indie_outcomes$cvdth_status==1|indie_outcomes$hfhosp_status==1] <- 1
indie_outcomes$cvdth_hfhosp_dt <- pmin(indie_outcomes$cvdth_dt,indie_outcomes$hfhosp_dt)

## Composite IPRESERVE endpoint - all-cause death + CV hospitalization

indie_outcomes$dth_cvhosp_status <- 0
indie_outcomes$dth_cvhosp_status[indie_outcomes$dth_status==1|indie_outcomes$cvhosp_status==1] <- 1
indie_outcomes$dth_cvhosp_dt <- pmin(indie_outcomes$dth_dt,indie_outcomes$cvhosp_dt)

## Composite HF-ACTION - all-cause death + all-cause hospitalization

indie_outcomes$dth_hosp_status <- 0
indie_outcomes$dth_hosp_status[indie_outcomes$dth_status==1|indie_outcomes$hosp_status==1] <- 1
indie_outcomes$dth_hosp_dt <- pmin(indie_outcomes$dth_dt,indie_outcomes$hosp_dt)


#### Clean-up ####

names(indie_outcomes)[1] <- "patientid"
indie_outcomes$study <- "INDIE"
missing_outcomes_indie <- analysis_outcomes[!analysis_outcomes %in% names(indie_outcomes)]
indie_outcomes[,missing_outcomes_indie] <- NA
indie_outcomes <- indie_outcomes[,analysis_outcomes]


########################################### **************** COMBINE OUTCOMES ***************** #############################################
#
# ..%%%%%%...%%%%%%%..%%.....%%.%%%%%%%%..%%%%.%%....%%.%%%%%%%%.....%%%%%%%..%%.....%%.%%%%%%%%..%%%%%%...%%%%%%%..%%.....%%.%%%%%%%%..%%%%%%.
# .%%....%%.%%.....%%.%%%...%%%.%%.....%%..%%..%%%...%%.%%..........%%.....%%.%%.....%%....%%....%%....%%.%%.....%%.%%%...%%%.%%.......%%....%%
# .%%.......%%.....%%.%%%%.%%%%.%%.....%%..%%..%%%%..%%.%%..........%%.....%%.%%.....%%....%%....%%.......%%.....%%.%%%%.%%%%.%%.......%%......
# .%%.......%%.....%%.%%.%%%.%%.%%%%%%%%...%%..%%.%%.%%.%%%%%%......%%.....%%.%%.....%%....%%....%%.......%%.....%%.%%.%%%.%%.%%%%%%....%%%%%%.
# .%%.......%%.....%%.%%.....%%.%%.....%%..%%..%%..%%%%.%%..........%%.....%%.%%.....%%....%%....%%.......%%.....%%.%%.....%%.%%.............%%
# .%%....%%.%%.....%%.%%.....%%.%%.....%%..%%..%%...%%%.%%..........%%.....%%.%%.....%%....%%....%%....%%.%%.....%%.%%.....%%.%%.......%%....%%
# ..%%%%%%...%%%%%%%..%%.....%%.%%%%%%%%..%%%%.%%....%%.%%%%%%%%.....%%%%%%%...%%%%%%%.....%%.....%%%%%%...%%%%%%%..%%.....%%.%%%%%%%%..%%%%%%.


#### Long term/chronic/outpatient outcomes ####

hdcp_outcomes <- rbind(topcat_outcomes,hfaction_outcomes)
hdcp_outcomes <- rbind(hdcp_outcomes, scdheft_outcomes)
hdcp_outcomes <- rbind(hdcp_outcomes, best_outcomes)
hdcp_outcomes <- rbind(hdcp_outcomes, ip_outcomes)
hdcp_outcomes <- rbind(hdcp_outcomes, relax_outcomes)
hdcp_outcomes <- rbind(hdcp_outcomes, solvd_outcomes)
hdcp_outcomes <- rbind(hdcp_outcomes, solvd_registry_outcomes)
hdcp_outcomes <- rbind(hdcp_outcomes, dig_outcomes)
hdcp_outcomes <- rbind(hdcp_outcomes, neat_outcomes)
hdcp_outcomes <- rbind(hdcp_outcomes, mocha_outcomes)
hdcp_outcomes <- rbind(hdcp_outcomes, corona_outcomes)
hdcp_outcomes <- rbind(hdcp_outcomes, paradigm_outcomes)
hdcp_outcomes <- rbind(hdcp_outcomes, escape_outcomes)
hdcp_outcomes <- rbind(hdcp_outcomes, stich_outcomes)
hdcp_outcomes <- rbind(hdcp_outcomes, stiches_outcomes)
hdcp_outcomes <- rbind(hdcp_outcomes, exact_outcomes)
hdcp_outcomes <- rbind(hdcp_outcomes, guideit_outcomes)
hdcp_outcomes <- rbind(hdcp_outcomes, carress_outcomes)
hdcp_outcomes <- rbind(hdcp_outcomes, dose_outcomes)
hdcp_outcomes <- rbind(hdcp_outcomes, rose_outcomes)
hdcp_outcomes <- rbind(hdcp_outcomes, athena_outcomes)
hdcp_outcomes <- rbind(hdcp_outcomes, fight_outcomes)
hdcp_outcomes <- rbind(hdcp_outcomes, indie_outcomes)
hdcp_outcomes <- rbind(hdcp_outcomes, ironout_outcomes)
hdcp_outcomes <- rbind(hdcp_outcomes, life_outcomes)

write.csv(hdcp_outcomes,
          '~/Dropbox/ADAPT-HF/Master HDCP files/hdcp_outcomes.csv',
          row.names=F)


gcs_auth("~/Dropbox/ADAPT-HF/Master HDCP files/harmonization-286013-39f492122f69.json")
gcs_upload(hdcp_outcomes, 
           bucket="master_hdcp_files",
           name="trial_outcomes.csv")
```
