## Supplementary material for "Aggregating data to accelerate personalized therapy in heart failure (ADAPT-HF)": Harmonization scripts: Upload outcomes and criteria.R

```
library(bigrquery)
library(readr)
library(data.table)
library(arrow)
library(stringr)
library(readxl)

### Upload phenotyping criteria

bq_auth(path="harmonization-286013-39f492122f69.json")

my_bq <- dbConnect(
  bigrquery::bigquery(),
  project="harmonization",
  dataset="cohorts",
  billing="harmonization"
)


hdcp_cohort_criteria <- 
  read_xlsx("~/Dropbox/ADAPT-HF/Master HDCP files/Criteriaset_cohort study 5.1.xlsx")

hdcp_cohort_criteria$rule_num <-
  rownames(hdcp_cohort_criteria)

phenotypes_cohort <- 
  read.csv("~/Dropbox/ADAPT-HF/Master HDCP files/phenotypelist_cohort.csv",
        na.strings=c("NA","","NULL","\\N"))

phenos <- unique(hdcp_cohort_criteria[,c("phenotype")])

miss_phenos <- phenos[!phenos$phenotype %in% phenotypes_cohort$phenotype,]

hdcp_cohort_criteria$string_value <- str_replace(hdcp_cohort_criteria$string_value,"\xb2 ","≤ ")
hdcp_cohort_criteria$string_value <- str_replace(hdcp_cohort_criteria$string_value,"\xb3","≥ ")
hdcp_cohort_criteria$rule_num <- rownames(hdcp_cohort_criteria)

hdcp_cohort_criteria$lower_bound[is.na(hdcp_cohort_criteria$lower_bound)&hdcp_cohort_criteria$rule_type=="range"] <- -1000000
hdcp_cohort_criteria$upper_bound[is.na(hdcp_cohort_criteria$upper_bound)&hdcp_cohort_criteria$rule_type=="range"] <- 1000000

hdcp_cohort_criteria$form <- toupper(hdcp_cohort_criteria$form)


hdcp_cohort_criteria$field <- toupper(hdcp_cohort_criteria$field)

unique(hdcp_cohort_criteria$field[hdcp_cohort_criteria$study=="JHS"])

hdcp_cohort_criteria$study_field <-
  paste(hdcp_cohort_criteria$study,
        hdcp_cohort_criteria$form,
        hdcp_cohort_criteria$field,
        sep="_")

if (bq_table_exists("harmonization-286013.pipeline.cohort_criteria"))
{bq_table_delete("harmonization-286013.pipeline.cohort_criteria")}


bq_table_upload(x="harmonization-286013.pipeline.cohort_criteria",
                hdcp_cohort_criteria,quiet=T,
                create_disposition="CREATE_IF_NEEDED",
                write_disposition="WRITE_TRUNCATE")


if (bq_table_exists("harmonization-286013.pipeline.phenotypes_cohort"))
{bq_table_delete("harmonization-286013.pipeline.phenotypes_cohort")}
bq_table_upload(x="harmonization-286013.pipeline.phenotypes_cohort",
                phenotypes_cohort,
                quiet=T,
                create_disposition="CREATE_IF_NEEDED",
                write_disposition="WRITE_TRUNCATE")


### Combine all outcomes

aric_outcomes <- 
  read_parquet("~/Dropbox/ADAPT-HF/Master HDCP files/Cohort data/aric_outcomes.parquet",
               na.strings = c(NA,"NA","NULL",""))

chs_outcomes <- 
  read_parquet("~/Dropbox/ADAPT-HF/Master HDCP files/Cohort data/chs_outcomes.parquet",
               na.strings = c(NA,"NA","NULL",""))


fhs_outcomes <- 
  read_parquet("~/Dropbox/ADAPT-HF/Master HDCP files/Cohort data/fhs_outcomes.parquet",
               na.strings = c(NA,"NA","NULL",""))

jhs_outcomes <- 
  read_parquet("~/Dropbox/ADAPT-HF/Master HDCP files/Cohort data/jhs_outcomes.parquet",
               na.strings = c(NA,"NA","NULL",""))

mesa_outcomes <- 
  read_parquet("~/Dropbox/ADAPT-HF/Master HDCP files/Cohort data/mesa_outcomes.parquet",
               na.strings = c(NA,"NA","NULL",""))

aric_outcomes[,outcomes_list_2[!outcomes_list_2 %in% names(aric_outcomes)]] <- NA
chs_outcomes[,outcomes_list_2[!outcomes_list_2 %in% names(chs_outcomes)]] <- NA
fhs_outcomes[,outcomes_list_2[!outcomes_list_2 %in% names(fhs_outcomes)]] <- NA
jhs_outcomes[,outcomes_list_2[!outcomes_list_2 %in% names(jhs_outcomes)]] <- NA
mesa_outcomes[,outcomes_list_2[!outcomes_list_2 %in% names(mesa_outcomes)]] <- NA

all_outcomes <-
  rbind(aric_outcomes[,outcomes_list_2],
        chs_outcomes[,outcomes_list_2],
        fhs_outcomes[,outcomes_list_2],
        jhs_outcomes[,outcomes_list_2],
        mesa_outcomes[,outcomes_list_2])

if (bq_table_exists("harmonization-286013.cohorts.outcomes"))
{bq_table_delete("harmonization-286013.cohorts.outcomes")}

bq_table_upload(x="harmonization-286013.cohorts.outcomes",
                all_outcomes,
                quiet=T,
                create_disposition="CREATE_IF_NEEDED",
                write_disposition="WRITE_TRUNCATE")
```
